# Mutation Pathogenicity Prediction by a Biology Based Explainable AI Multi-Modal Algorithm

**DOI:** 10.1101/2024.06.05.24308476

**Authors:** Raizy Kellerman, Omri Nayshool, Ortal Barel, Sharon Paz, Ninette Amariglio, Eyal Klang, Gideon Rechavi

**Author notes:** These authors contributed equally to this work (Raizy Kellerman, Omri Nayshool).

## Abstract

Most known pathogenic mutations occur in protein-coding regions of DNA and change the way proteins are made. Deciphering the protein structure therefore provides great insight into the molecular mechanisms underlying biological functions in human disease. While there have recently been major advances in the artificial intelligence-based prediction of protein structure, the determination of the biological and clinical relevance of specific mutations is not yet up to clinical standards. This challenge is of utmost medical importance when decisions, as critical as suggesting termination of pregnancy or recommending cancer-directed rational drugs, depend on the accuracy of prediction of the effect of the specific mutation. Currently, available tools are aiming to characterize the effect of a mutation on the functionality of the protein according to biochemical criteria, independent of the biological context. A specific change in protein structure can result either in loss of function (LOF) or gain-of-function (GOF) and the ability to identify the directionality of effect needs to be taken into consideration when interpreting the biological outcome of the mutation. Here we describe <u>Tri</u>ple-modalities <u>V</u>ariant Interpretation and <u>A</u>nalysis (TriVIAI), a tool incorporating three complementing modalities for improved prediction of missense mutations pathogenicity: protein language model (pLM), graph neural network (GNN) and a tabular model incorporating physical properties from the protein structure. The TriVIAl ensemble’s predictions compare favorably with the existing tools across various metrics, achieving an AUC-ROC of 0.887, a precision-recall curve (PRC) score of 0.68, and a Brier score of 0.16. The TriVIAI ensemble is also endowed with two major advantages compared to other available tools. The first is the incorporation of biological insights which allow to differentiate between GOF mutations that tend to cluster in specific hotspots and affect structure in a specific functional way versus LOF mutations that are usually dispersed and can cripple the protein in a variety of different ways. Importantly, the advantage over other available tools is more noticeable with GOF mutations as their effect on the protein structure is less disruptive and can be misinterpreted by current variant prioritization strategies. Until now available AI-based pathogenicity predicting algorithms were a black box for the users. The second significant advantage of TriVIAI is the explainability of the ensemble which contrasts the other available AI-based pathogenicity predicting algorithms which constitute a black box for the users. This explainability feature is of major importance considering the clinical responsibility of the medical decision-makers using AI-based pathogenicity predictors.

## Introduction

Predicting the pathogenicity of missense mutations is crucial for both understanding underlying genetic processes and enabling more precise clinical interventions. Despite its importance, this task remains challenging due to the diverse molecular mechanisms and functional consequences that can arise from such mutations. The prevalent use of next generation sequencing in recent years made the interpretation of Variants of Uncertain Significance (VUS) a major challenge in genetics and molecular oncology and further underscored the necessity of precise interpretation within the realm of personalized medicine.

Several computational tools have been developed to address this challenge, with methods ranging from simple linear regression models to more complex machine learning algorithms, incorporating energy, functional, conservational and statistical features.^1–3^ While effective to some extent, these predictors often rely on limited types of data and fail to capture the full complexity of biological systems. To address this challenge, we describe a versatile machine learning framework that adopts a multi-modal approach. This framework integrates tabular features, sequence-based information, and three dimensional (3D) structural data to provide a holistic view of protein alterations. Our results demonstrate that the proposed model not only achieves improved predictive accuracy but also generalizes effectively, capturing nuanced representations of the biological principles governing protein function. This unified approach provides a robust and meaningful framework for the next generation of missense mutations pathogenicity prediction, offering new avenues for research and clinical applications.

Understanding the functional consequences of genetic mutations is at the core of both fundamental biological research and translational clinical medicine. Among various types of genetic mutations, missense variations, where a single nucleotide change results in a different amino acid in the protein sequence, stand out as particularly significant. Predicting the pathogenicity of these missense variations is crucial for diagnosing genetic diseases and somatic disorders including cancer, informing therapeutic interventions, as well as advancing the understanding of functional genomics and proteomics. However, these variations can have myriad effects on protein structure, function, and interaction, making them challenging to analyze and interpret.

Recent advancements in deep learning have resulted in improving the accuracy of predictive models. Specifically, protein language transformer models^4–9^ —inspired by breakthroughs in natural language processing—have been successfully applied to predict the pathogenicity of missense variations, or more generally variant effect prediction (VEP) by analyzing amino acid sequences. Concurrently, structure-based graph neural networks have emerged as a powerful tool for understanding protein interactions and conformations. The critical role played by AlphaFold^10^ in advancing protein structure prediction has opened new doors for employing graph neural networks over predicted protein structural data in various protein-related tasks, including missense mutation pathogenicity prediction^11,12^.

Our predictor framework also incorporates traditional tools such as FoldX to capture the energy change caused by the mutations to further increase accuracy of the predictions of the effect caused by specific mutations. The present study aims to build upon these advances by proposing a multi-modal machine learning framework. Our framework, TriVIAl, stands for <u>Tri</u>ple-modalities <u>V</u>ariant Interpretation and <u>A</u>nalysis, integrating tabular, sequence-based, and structure-based data. Beyond achieving state-of-the-art predictive accuracy, it provides a deeper understanding of the biological intricacies underlying protein functionality.

As a consequence of developing a pathogenicity classification model, the next step involves fine-tuning of the model weights, making it capable of distinguishing between Gain of Function (GOF) and Loss of Function (LOF) variants in proteins. GOF refers to mutations that enhance or introduce new functions, while LOF pertains to changes resulting in partial or complete loss of function. This distinction is vital for drug discovery, personalized medicine, and understanding disease mechanisms.

Furthermore, in addition to providing classification labels along with their confidence, we propose an explainability framework that enables us to visualize the input residues that contribute the most to the output prediction. This enables a deeper understanding of the mechanisms involved in making the mutation pathogenic or benign. This understanding is especially important in GOF mutations, where the changes in the protein structure and stability are usually subtler and may depend on environment-related features such as binding sites and interaction with other proteins.

## Methods

### Principles and Rational

#### Overview of TriVIAl

TriVIAl is a hybrid deep learning model that combines three distinct data types. *A fully connected neural network (FCN)* which models the tabular features encapsulating numeric attributes, such as difference in free energy, the root mean square deviation (RMSD) of the proteins’ atoms, BLOSUM substitution values, and additional parameters, as detailed in **Supplementary Table 1**; *A transformer-based Protein Language Model (PLM)* that has been pre-trained on a massive set of sequences; and a *Graph Neural Network (GNN)* that captures and maps the molecular 3D aspects that were introduced by the mutation. Together, these modalities complement each other to gain synergistic effect.

Tabular features, such as energy and molecular properties, as well as population distribution and co-evolutionary information, are core components of many missense pathogenicity prediction tools. While it is clear that tabular features cannot capture the complex context of the proteins, they offer well-established features that can easily be computed and offer clear prediction interpretability. For instance, a significant change in ΔΔG can be directly linked to destabilization of a protein, providing reasoning for pathogenicity prediction. We assume that ΔΔG also enables the distinction of the nature of the pathogenic mutation, especially if it Gain-of-Function or Loss-of-Function^13^.

Protein language Models (PLMs), successors of general Large Language Models (LLMs), are powerful models that learn meaningful representations with biophysical quantitative features of proteins in unsupervised process. Protein sequence data is vastly more abundant than structural or energy and co-evolution data, enabling PLMs to learn through the largest sets of known sequences such as UniProt and UniRef datasets, without homology information or labeled properties. However, the current PLMs have several limitations: First, most of them are limited to process sequences with maximal length of ∼1K amino acids, raising the question of their utility for proteins with longer sequences, especially where the mutation occurs after the length limitation. Another concern is raised from the training procedure – LLMs are trained by masking specific tokens in a sequence (words in natural language, residues in protein sequences) and learning to predict the masked token; in other words, they are trained to predict what amino acid may appear in a specific location with what likelihood; hence, their ability to fully represent all of the physical and chemical interaction is not fully proven.

Structure-based models operate on proteins represented as augmented 3D graphs with features attached to each node – either in residue level^14,15^, atom level^16,17^ or combined hierarchical representations – and to each edge between near nodes – either predefined *k* nearest neighbors^18^ or all neighbors under certain distance thresholds, usually 4.5-10 Angstrom. Structures are richer in information than sequences and allow models to derive insights directly from local chemical interactions. This enables structure-based models to utilize broader, deeper and richer knowledge encapsulated in atomic- or residual level of interactions and by comparing wild-type to mutation structures they can learn how the physicochemical state of the mutation is implicitly expressed by the backdrop of structure itself, rather than against phylogenetic sequence data that merely implies structure. ^15^

To take advantage of all worlds, TriVIAl is designed as a triple-backbone neural network with a unified head. An FCN backbone operates on the tabular features; a pre-trained PLM transformer backbone processes the sequence information; and a GNN is deployed on the structural data.

An overview of the system is depicted in **Figure 1**.

**Figure 1:**
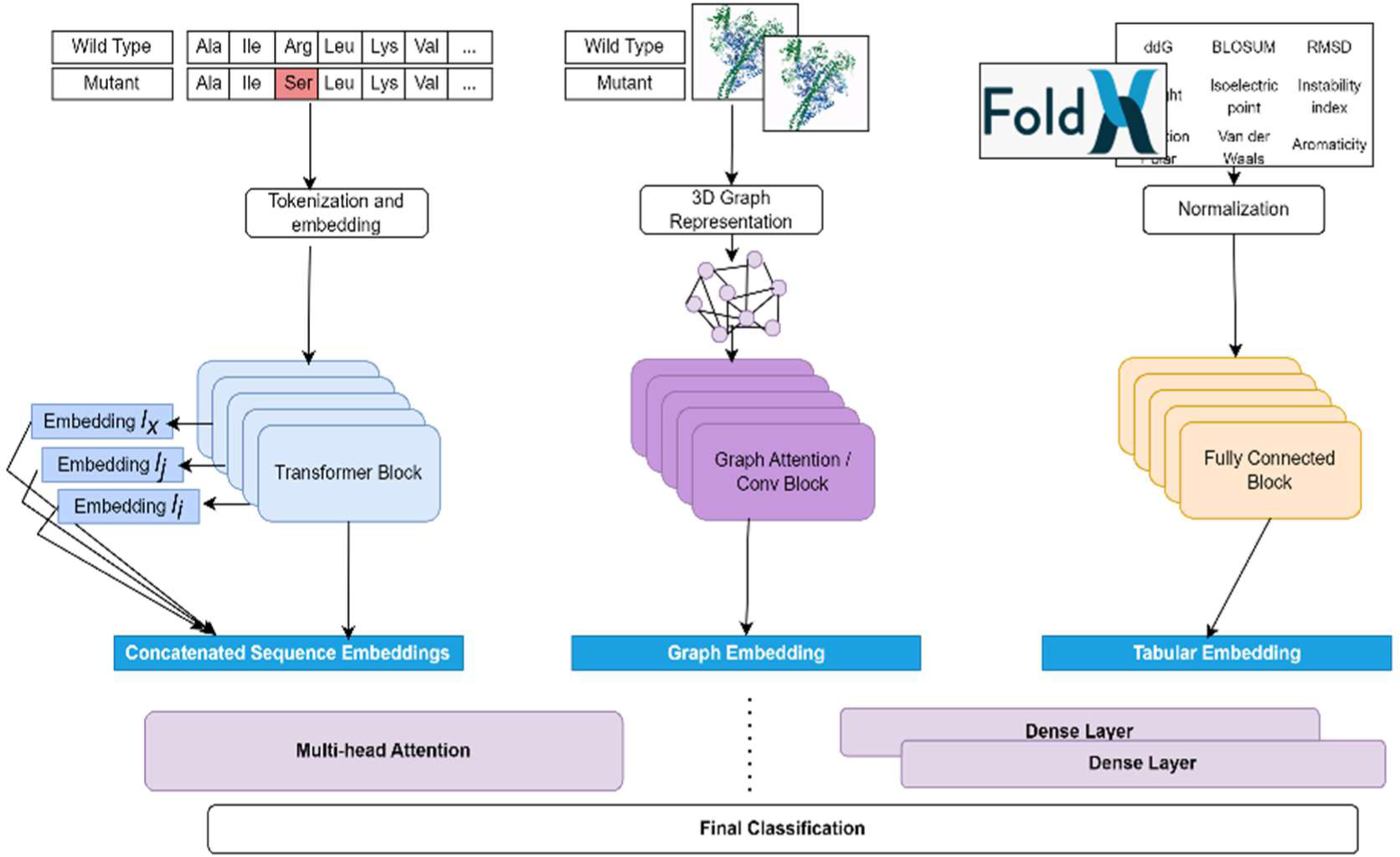
A systematic overview of the TriVIAl missense pathogenicity ensemble classifier. TriVIAl is built upon three inputs: sequences, structures and tabular physical properties, and forwards them to three independent models: pLM, GNN, and fully-connected, respectively. These models create a latent representation with a unified dimension that are then concatenated and passed through either a stack of dense layers or a multi-head attention network towards a final classification layer.

### Classification Framework Design

#### Tabular model

To properly evaluate the tabular features’ prediction-based utility three independent models were evaluated: Random Forest, gradient-boosting (XGBoost), and a fully-connected 7-layers neural network. All hyper-parameters such as number of trees and shape and number of layers were determined per hyper-parameters grid search of 5-fold cross validation. The full configuration details are described in **appendix A1**.

Although classic ML models, namely random forest and XGBoost on their own perform better than the NN model, for the combined multi-modality model which is trained by back-propagation, the neural network was integrated.

#### Sequence model

We evaluated five flavors of Protein Language Models: ProtBert-BFD^4^, ProteinBert^6^, ESM1v^8^, ESM1b^7^, and Ankh^19^. For each model, we froze the learning on all but the top 3 layers, maintaining the original configuration of layers.

During the sequence model training, each pair of wild-type and mutation sequences were forwarded through the layers, producing a final embedded representation vector. Then, inspired by the concept of feature pyramid networks^20^ ^21^, we pooled the representation of the amino acid in the mutation position from several layers. This approach enabled it to capture both lower-level, higher-resolution as well as higher-level, lower-resolution embedded representations. The global (full-sequence) representation was concatenated to the amino acid representation for both the wild-type and the mutation and were forwarded to final linear classification layers. For mutations occurring past the model limit (1022 amino acids for ProtBert-BFD and ESM), a randomized window of the maximal tokens limit, containing the mutation position, was generated per forward, enabling mild data augmentation.

#### Structure model

For balancing between precision and complexity we followed ProNet’s backbone-level representation^22^ with additional residue-level features. The backbone is represented by the Alpha Carbon (*Cα*) atom, with the corresponding amino (*-NH*_2_) group and carboxyl (*-COOH*) group. We tested both the radius-based edges (An edge between nodes was defined for all residues with any atom existing in a radius of 6 Angstrom) and *k*-nearest neighbors with *k*=30. The node features were augmented, in addition to residue ID as a one-hot encoding of *1×20* vector, with polarity as *1×4* vector (polar, apolar, negatively charged and positively charged), and residue charge (floating point single value), similar to Réau et al.^15^. As edge features, we followed Ingraham et al.^23^ and used relative distance, direction and orientation based on the spherical coordinates between neighboring backbone nodes, in addition to relative positional embedding, effectively combining structural and positional encoding.

We investigated two flavors of Graph Neural Networks: Equivariant Convolution Graph Network^24^ and Graph Attention networks^25^.The rotation- and translation-equivariant convolution or the attention layers were used as interaction blocks that update the nodes based on the edge features and mean-based summation function with several fully-connected layers used as the global readout function. The global representation is augmented, similarly to the sequence model implementation, with node-position pooling operation which accumulates the hidden representation of the mutation position over sampled subset of the intermediate layers, followed by attention pooling layer for adaptive and contextual aggregation. The full architecture details are described in **Appendix A2**.

For each implementation, similar to the sequence-based model, different levels of feature embedding were obtained for the mutation node and edges, through pooling from different layers. The mutation specific embedding was concatenated to the global protein-wide readout feature before the final classification layers. The Graph Attention Network model performed slightly better, though being more computationally-heavy.

### Multiple Modalities Integration Considerations

Numerous strategies exist for combining several representations of data. Specifically, for protein-related tasks, existing approaches vary from simple all-numeric features to sophisticated consolidated representation deep learning. Some methods create unified tabular features, leveraging the embedding from learned deep learning models such as PLMs either as-is or after pre-processing such as principal components analysis (PCA), retaining the top *N* principal components^26^. Ensemble learning offers another straight-forward tactic, with the adjustment of the weights of the predictions retrieved from the discrete models by e.g. logistic regression. A more recent approach that is successfully applied to PPI^14,27^ and protein function prediction^28,29^ adopts PLM’s embedding for each amino acid as residue-level node feature. While generalizable and showing competitive results, the inherent limitations of PLMs such as sequence length restriction apply to some extent. An additional contemporary technique integrates protein sequence with Gene Ontology (GO) annotations, fed as either graph-based features or language model embedding^6,30,31^.

A recent promising method, AlphaMissense^51^, reverses the multi-input approach and instead uses sequence, sampled variants and multiple sequence alignment (MSA) data to create a multi-tasking model that predicts both the mutated structure and a missense pathogenicity score.

However, our method, though conceptually straightforward, highlights the advantages of using diverse data sources to deeply comprehend the multifaceted origins of pathogenicity, as expressed by each modality. When combined with explainability techniques outlined in the explainability section, we can delve into the diverse mechanisms and underlying processes responsible for the predicted result.

To demonstrate the importance of combining various modalities, we calculated the error rate, defined as the mean absolute difference between predictions and ground truth values, in relation to both mutation position and the mean value of AlphaFold’s pLDDT (Predicted Local Distance Difference Test) metric. The pLDDT metric estimates the per-residue confidence in the agreement between AlphaFold predictions and experimental structures. 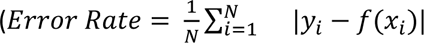, where *y* is the ground-truth label, *x* is the input and *f* is the prediction function)

Our findings illustrate that a model relying solely on sequence information performs worse when mutations occur outside of the model’s context window limitation, e.g., ProtBert has an actual context window of 1022 amino acids, making it challenging in dealing with proteins with longer sequences (**Figure 2**). Similarly, the performance of a structure-only model degrades as pLDDT values decrease (**Figure 3**). This trend may be attributed to the increased complexity of the structural context, which appears to underlie the decreased performance observed in both AlphaFold and the GNN models. Alternatively, it illustrates that the model’s quality is directly affected by the input’s quality.

**Figure 2:**
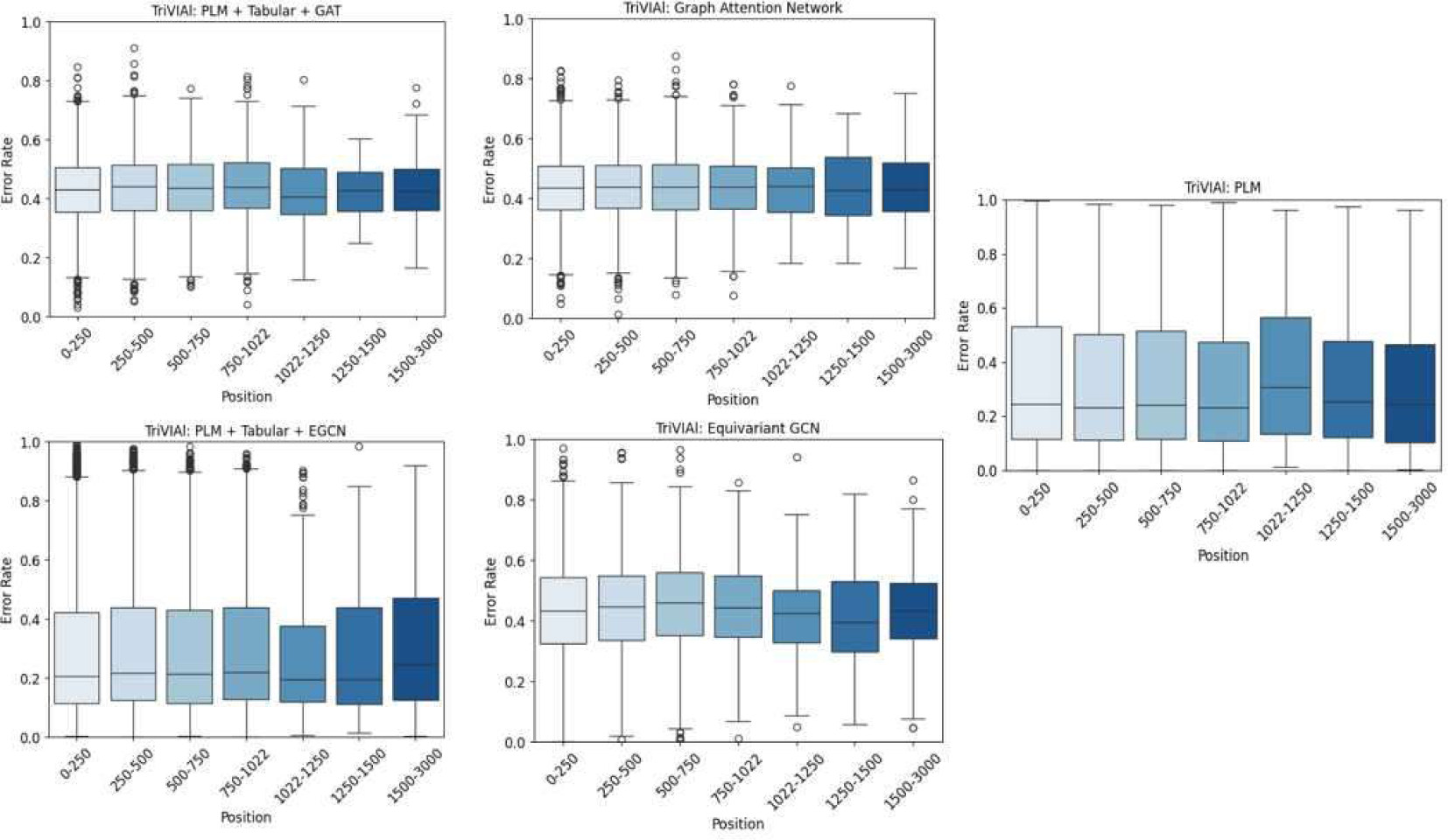
A box plot of prediction error rate by mutation position, for each of the five model flavors. The X axis represents the mutation position, in 250 bin size. The Y axis shows the distribution of the error rate for each bucket. The sequence-only classifier has a higher error rate when the mutation occurs beyond the model’s context window of 1022 residues.

**Figure 3:**
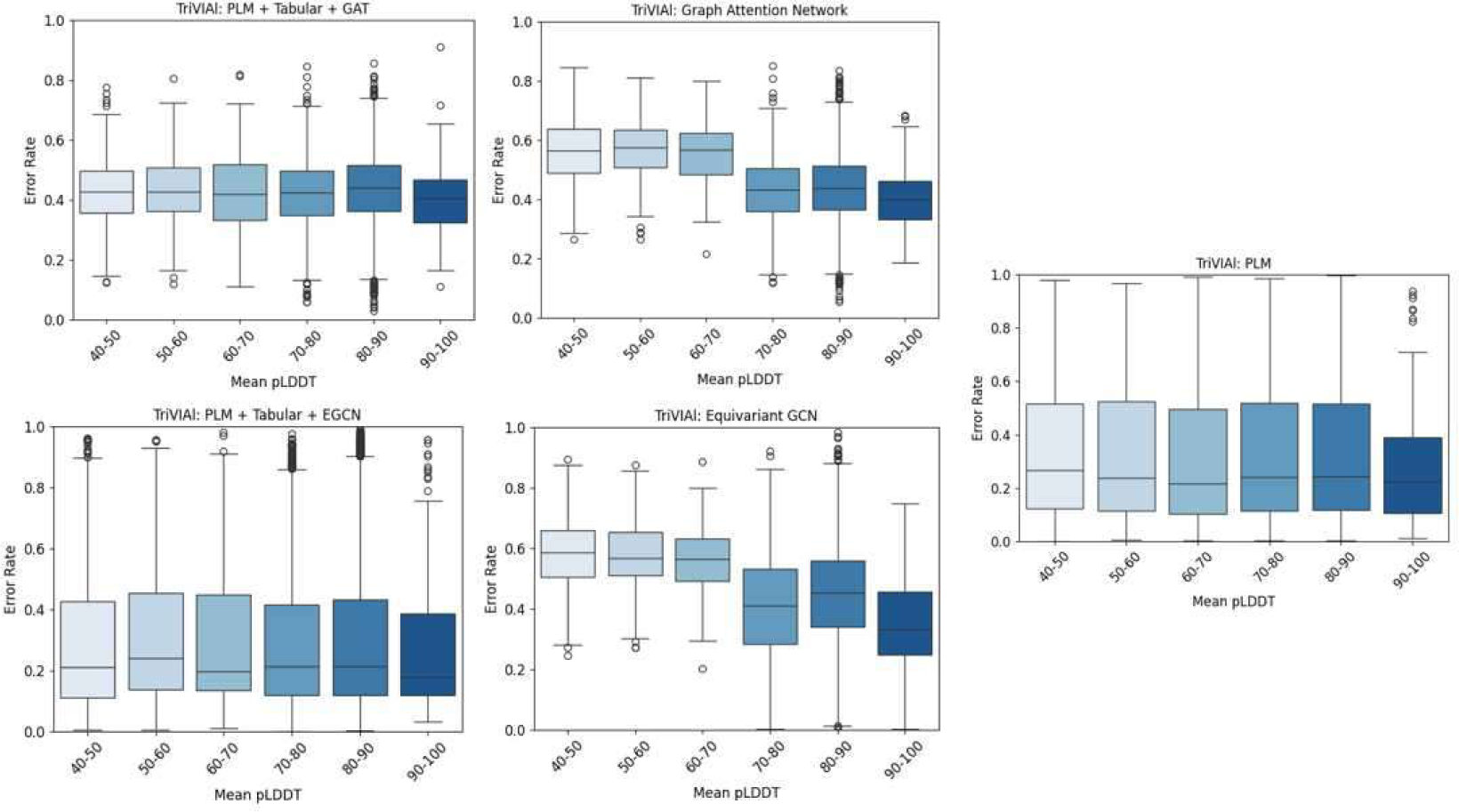
A box plot of prediction error rate by mean AlphaFold’s pLDDT. The structure-only classifiers have a higher error rate where the overall mean pLDDT is lower, i.e. there is opposite correlation between the mean pLDDT and the error rate of the structure models.

### Data Annotation and Preparation

#### Pathogenicity Classification Task

The training and validation data samples were collected from ClinVar^32^ (obtained in 07/22/2022) and HGMD^33^ Pro 2022.2 hg38. We used only the pathogenic/damaging and benign samples and opted to exclude the likely benign/likely pathogenic variants to ensure higher quality of data. The wild-type PDB structures were obtained from **AlphaFold Protein Structure Database** ^10,34^ Proteome UP000005640, and **FoldX software**^35,36^ (https://foldxsuite.crg.eu/) was used to create the mutated structures.

All VEP algorithms rank scores were annotated using dbNSFP4.4^37^. Pre-calculated AlphaMissense ranks cores were downloaded and annotated separately. Next, the tabular features were computed either directly by FoldX or computed using BioPython package, after superimposing the wild-type and mutation structure.

The test sets were obtained from Zhang et. al.^38^ and include deep mutation scan (DMS) ensemble of four genes from different sources: BRCA1^39^, PTEN^40^, *TP53*^41^ and MSH2^42^. Supp. table 2

##### Loss of Function (LOF) and Gain of Function (GOF) Classification Task

We developed a dataset derived from the COSMIC - Cancer Mutation Census ^43^ ^44^, focusing on gain-of-function (GOF) and loss-of-function (LOF) variants in cancer genes. Guided by the understanding that GOF mutations are prevalent in oncogenes and LOF mutations in tumor suppressor genes^45^, we implemented specific selection criteria. Our analysis was limited to missense mutations (’Substitution - Missense’) not found in the general population (’GNOMAD_GENOMES_AF’ == 0.0) and reported at least three times in the COSMIC dataset (’COSMIC_SAMPLE_MUTATED’ > 2). We excluded genes with overlapping oncogenic and tumor suppressor functions. This approach yielded a dataset of 2970 TSG-associated LOF and 3323 oncogene-linked GOF variants across 430 distinct genes.

#### Training Procedure

For the combined model, the curated ClinVar + HGMD dataset was divided into a stratified 80%-20% train/validation sets, respectively. The split was done at the gene level to make sure that there is no data contamination.

Each model for each data modality is initially trained as a standalone model with larger number of epochs and higher initial learning rate. Next, a unified fine-tuning is conducted on all backbones together for a smaller number of epochs and with lower learning rate. At the unified training, the final classification layer of the original standalone models is removed, and instead the last embedding layer outputs a tensor with unified predefined dimensions is then concatenated to the other backbones’ outputs and processed through the final classification layers.

All experiments were done on Nvidia DGX platform with 4 A100 GPUs, each having 80GB of memory. A simple parallelization approach of running each modality on a different GPU was applied.

## Results

### Missense Pathogenicity Prediction Metrics

A summary of Area Under the Curve Receiver Operating Characteristics (AUC ROC) and Precision Recall Curve (PRC AUC) of the TriVIAl model, compared to other state-of-the-art models, is presented in **Figures 4-7**. The TriVIAl combined model performs on par with AlphaMissense, and achieves the best PRC AUC, making it specifically well-suited for clinical decisions support.

**Figure 4:**
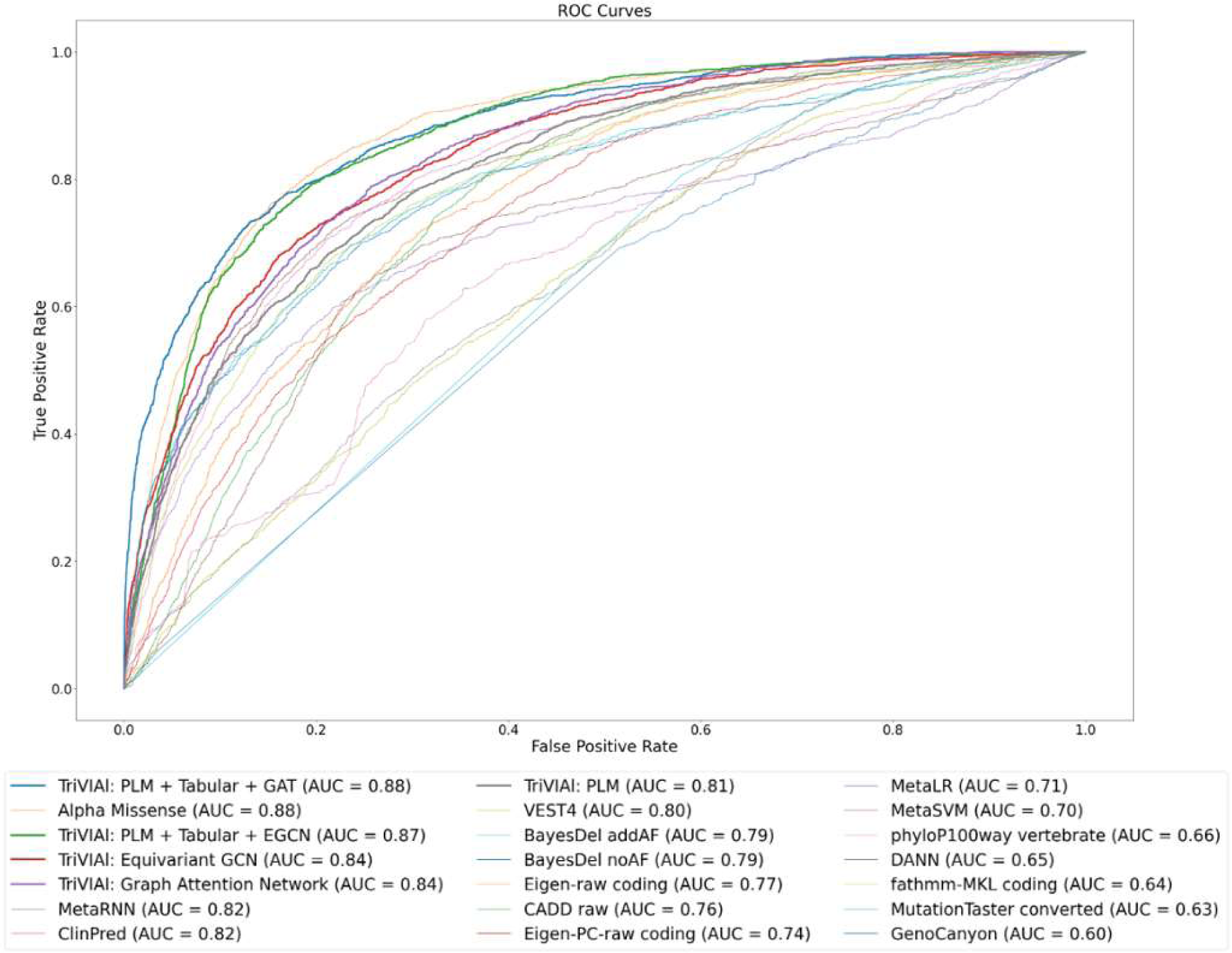
Validation ROC curves for TriVIAl and other leading classifiers on the validation set (Based on 9352 DMS variants). TriVIAl results are drawn with bolder line. TriVIAl and AlphaMissense yield the top performance of 0.88 AUC while other tools reach notably lower results.

**Figure 5:**
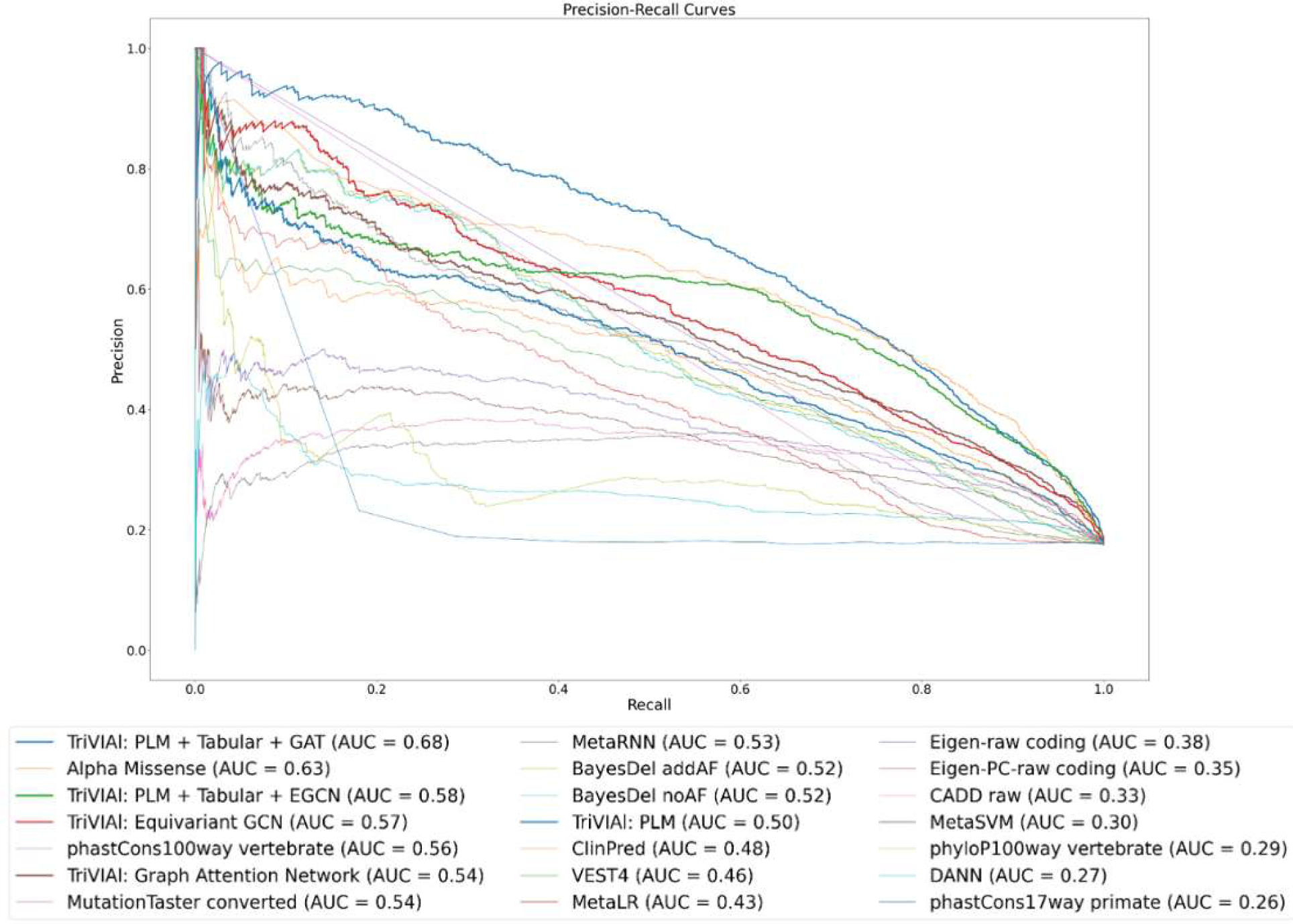
Precision-Recall curves plot on the validation set, for TriVIAl and the other top-20 classifiers. TriVIAl results are drawn with bolder lines. The scores were taken from dbNSFP.

**Figure 6:**
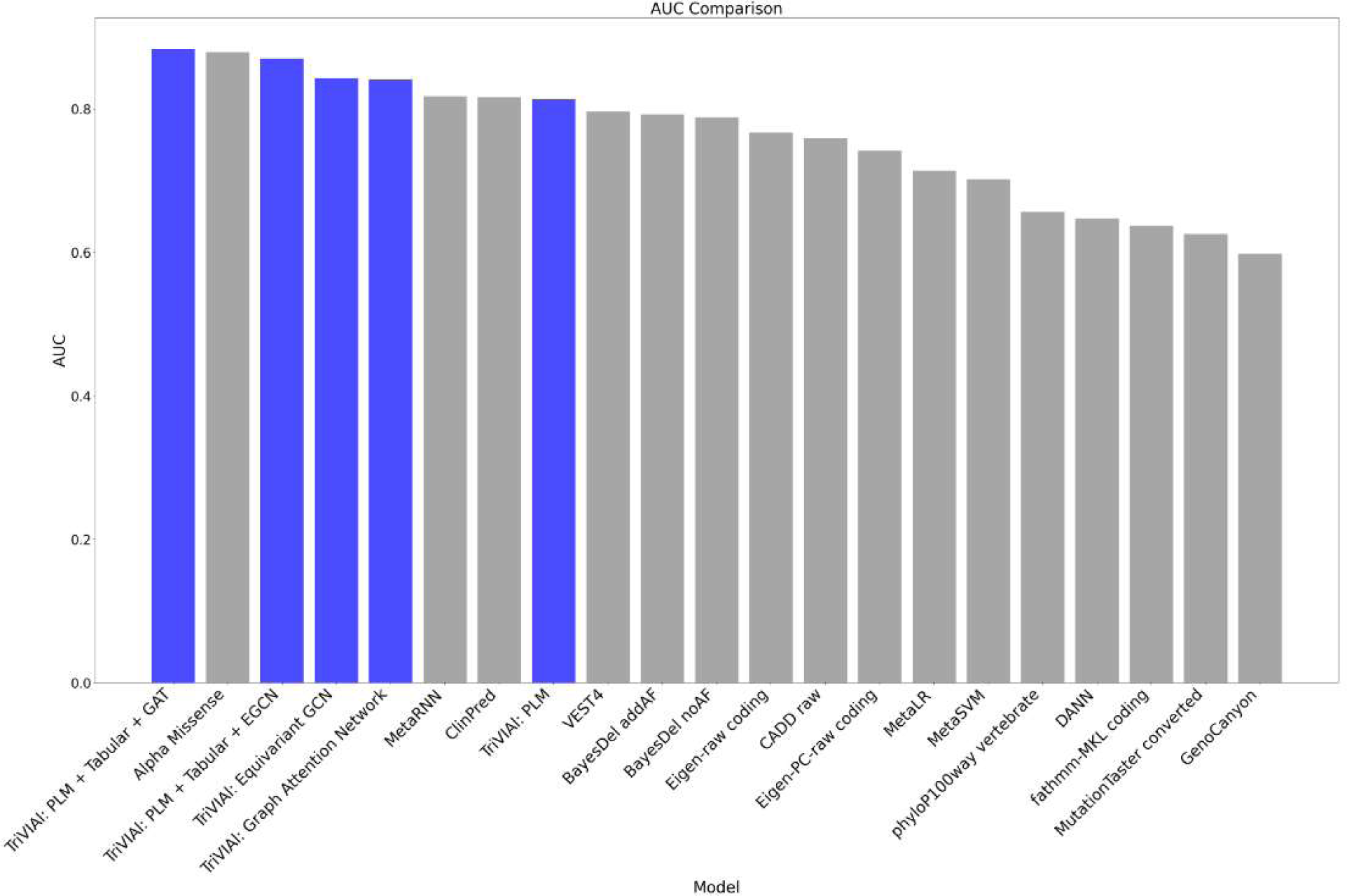
ROC AUC bar comparison. TriVIAl bars are in blue, other leading classifiers bars in grey.

**Figure 7:**
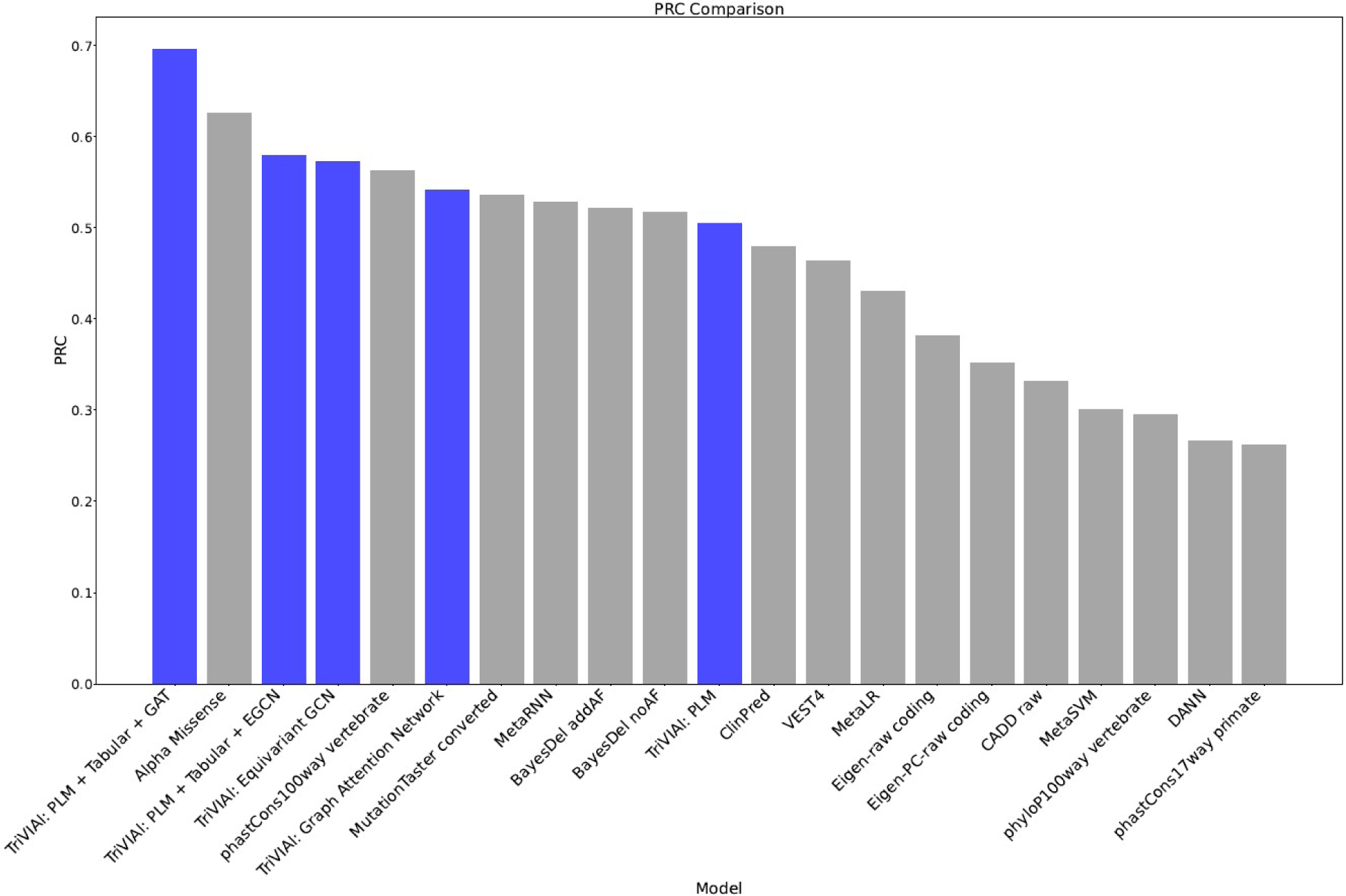
PRC AUC bar comparison. TriVIAl bars are in blue, other leading classifiers bars in grey.

An important aspect of classification models is calibration: Deep learning model calibration refers to the process of aligning the confidence of a model’s predictions with the actual likelihood of those predictions being correct. In an ideally calibrated model, if it assigns a confidence level of 80% to a series of predictions, then approximately 80% of these predictions should be correct. Calibration is crucial because deep learning models, especially those in critical applications like healthcare or autonomous driving, not only need to be accurate but also trustworthy in their uncertainty estimates. Without proper calibration, a model’s confidence levels may not accurately reflect the true probabilities of outcomes. This means a model could be overconfident, assigning high probabilities to predictions that are not as likely as indicated, or underconfident, assigning lower probabilities to outcomes that are more likely; both can lead to misguided decisions based on the model’s output.

Calibration techniques involve adjusting the output layer or applying post-processing methods like temperature scaling, where the confidence scores are systematically adjusted to better reflect reality. This adjustment helps in ensuring that the confidence levels are meaningful and reliable, making the model more useful and trustworthy for decision-making processes.

The Brier Score, also known as the Brier Proper Score, is a scoring or performance metric used in the evaluation of probabilistic predictions made by classification models, particularly in the context of binary classification tasks. It assesses the accuracy of these predicted probabilities.

In binary classification, there are typically two possible outcomes: positive and negative – pathogenic and benign, in our case. The Brier Score measures the mean squared difference between the predicted probabilities and the actual labels.

A lower Brier Score indicates better performance, as it means that the predicted probabilities are closer to the actual outcomes. Again, TriVIAl achieves the best Brier score amongst the other algorithms, showing that it is well-calibrated (**Figure 8**).

**Figure 8:**
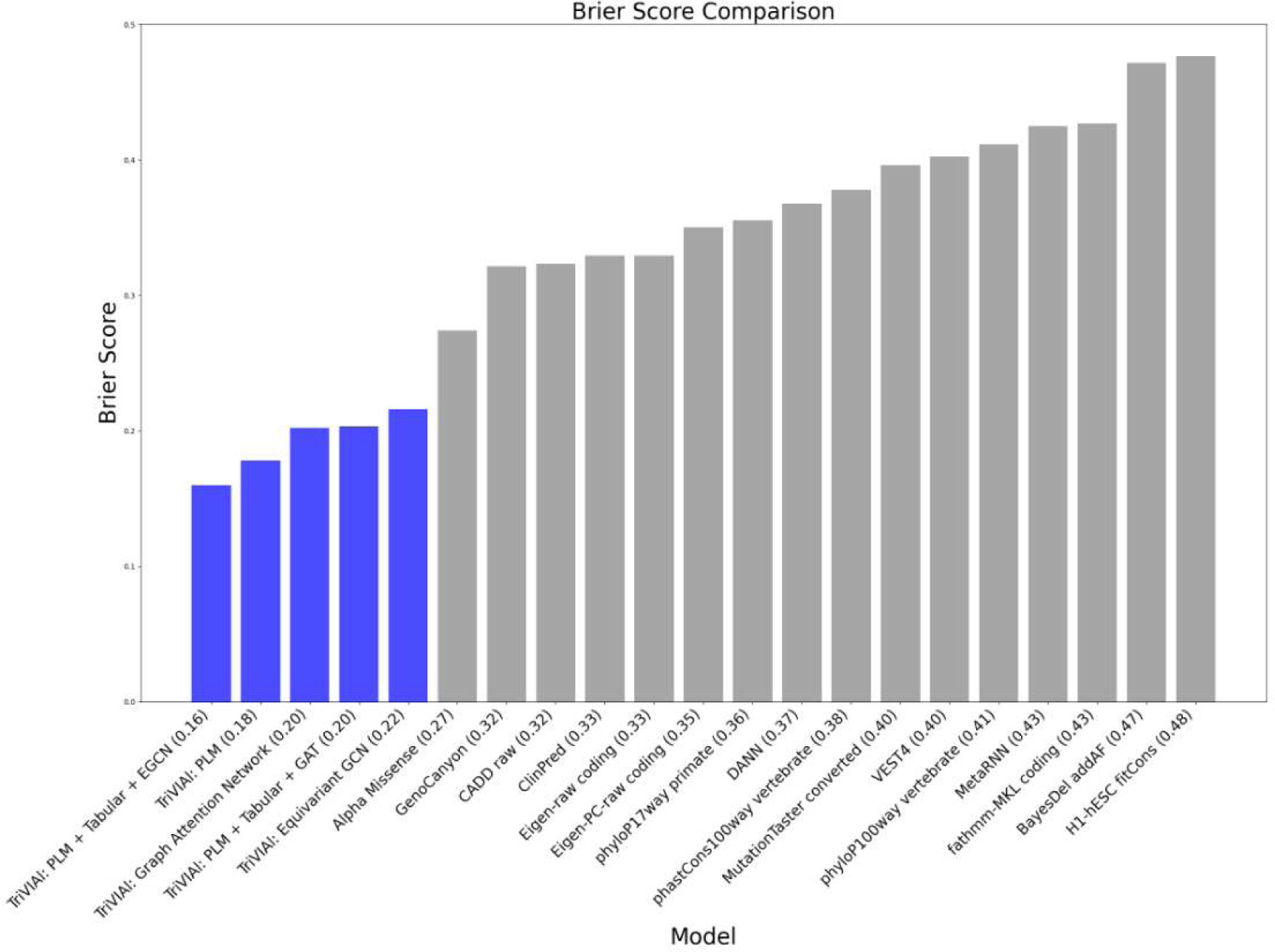
Brier score bar comparison of TriVIAl with other leading classifiers. TriVIAl bars are in blue.

A pivotal aspect of understanding the implications of a given mutation, is distinguishing between GOF and LOF mutations. Distinguishing between these two types is a challenging task that remains a largely unsolved issue with active research efforts^46^.

To assess our model’s ability to differentiate between GOF and LOF mutations, we initially applied t-Stochastic Neighbor Embedding (t-SNE) to project the embedding from the last classification layer into 2D space and to color them by their LOF or GOF label (**Figure 9**).

**Figure 9:**
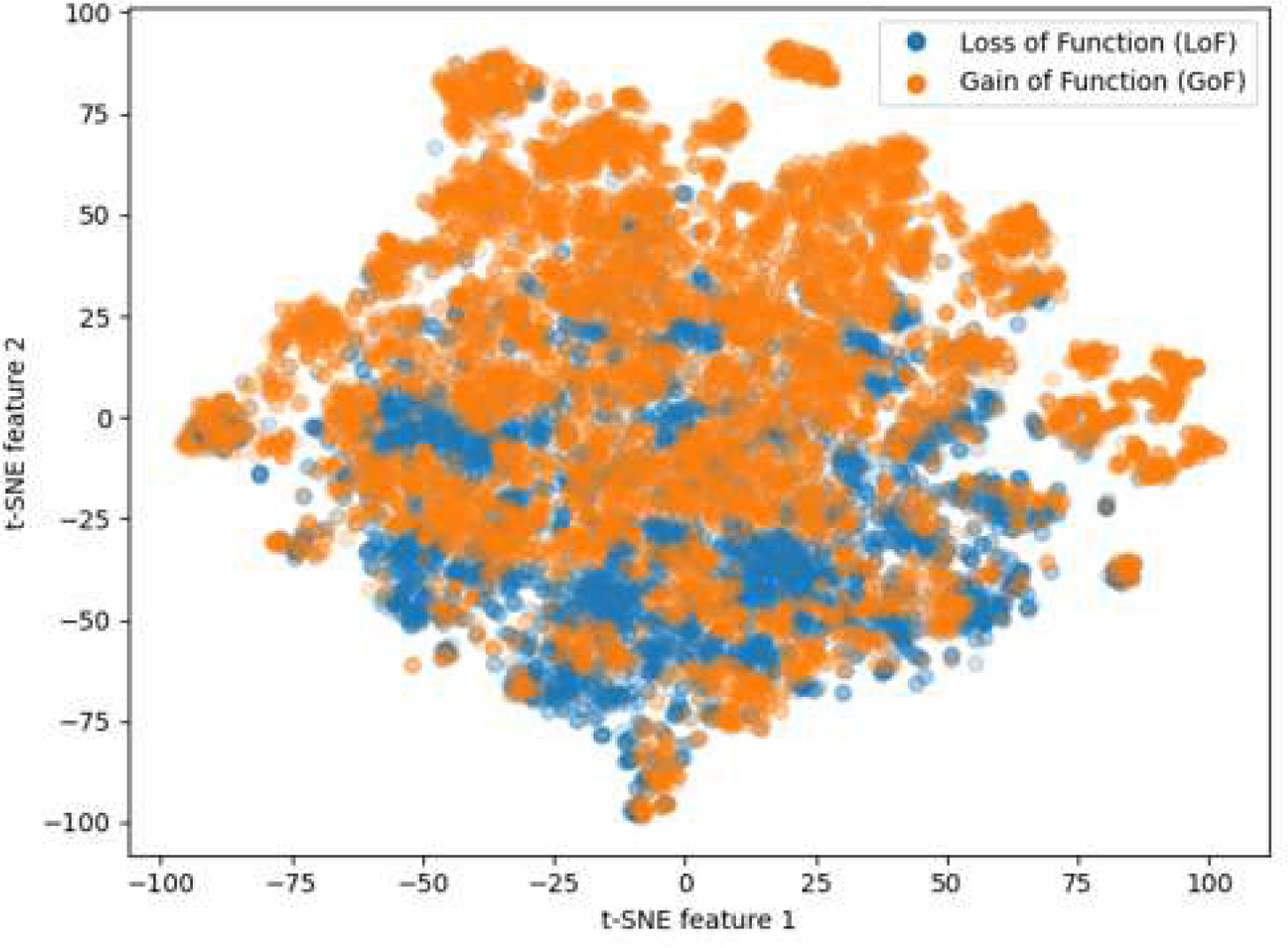
t-stochastic neighbor embedding (t-SNE) 2D plot of the pathogenicity classifier’s hidden representation, colored by LOF (blue) or GOF (orange) label. The hidden representation is taken from the last layer of the final classification, after the fusion blocks.

The t-SNE plot indicated that, to some extent, our model is capable of distinguishing between LOF and GOF mutations. We then proceeded to further fine-tune the classifier specifically for the task of LOF vs. GOF separation.

### LOF and GOF Prediction Metrics

The following four consequent experiments were performed in order to discriminate gain vs loss effects of the analyzed mutations. Experiments were designed to: (1) Fine-tune the pathogenicity classifier on LOF vs. GOF labels on the subset from ClinVar and HGMD, with annotations taken from Sevim Bayrak et al.^47,48^ (1150 samples, with 80%-20% split for train and validation respectively)(2)fine-tune the pathogenicity classifier on LoF vs. GoF labels on the data derived from COSMIC (4791 samples, with 80%-20% split for train and validation respectively) (3) Test the classifier obtained from step 1 on the COSMIC derived dataset, without additional training and (4)Test the classifier obtained from step 2 on the ClinVar and HGMD derived dataset, without additional training

These results as described in Table 1 demonstrate the capability of our model to discriminate between LOF and GOF mutations, and suggest that both germline and somatic mutations exhibit molecular properties with fundamental similarity that aids with this distinction.

**Table 1:** LOF vs. GOF training experiments results.

|  | Accuracy | Precision | Recall | AUC |
| --- | --- | --- | --- | --- |
| Step 1 (ClinVar/HGMD only) | 0.825 | 0.905 | 0.6 | 0.781 |
| Step 2 (COSMIC only) | 0.790 | 0.971 | 0.594 | 0.801 |
| Step 3 (ClinVar/HGMD train, COSMIC validation) | 0.745 | 0.952 | 0.512 | 0.74 |
| Step 4 (COSMIC train, ClinVar/HGMD validation) | 0.666 | 0.616 | 0.866 | 0.667 |

### Explainability

To facilitate more comprehensive understanding of our predictions from both a clinical and structural biology perspective, we utilized model explainability techniques. These techniques allowed us to identify the most impactful residues. Specifically, we implemented a generalized version of GradCAM-like explainability, which involves conducting a backward pass relative to the predicted class to calculate the gradients; Subsequently, we combined these gradients with activations to create a Class Activation Map (CAM) for each attention head. We then interpolated the most dominants outputs to the dimension of the input sequences to have a residue-level activation map. Those active input residues are highlighted by drawing connections between target residues and the mutated residue. For visualization, we rendered the proteins using the NGLView package^49^, adjusting the connection lines’ width to reflect each residue’s relative contribution. Although experimental, this suggested approach not only provides an interpretable justification for our predictions but also ensures that the model captures meaningful biological characteristics of the proteins and the mutations. Figures 10-17 present visualizations that illustrate the impact of missense mutations on four proteins associated with cancer. Analyses and interpretation of these mutations—namely BRAF p.V600E, JAK2 p.V617F, KRAS p.G12D, and VHL p.R161Q—are provided in the supplementary material.

**Figure 10:**
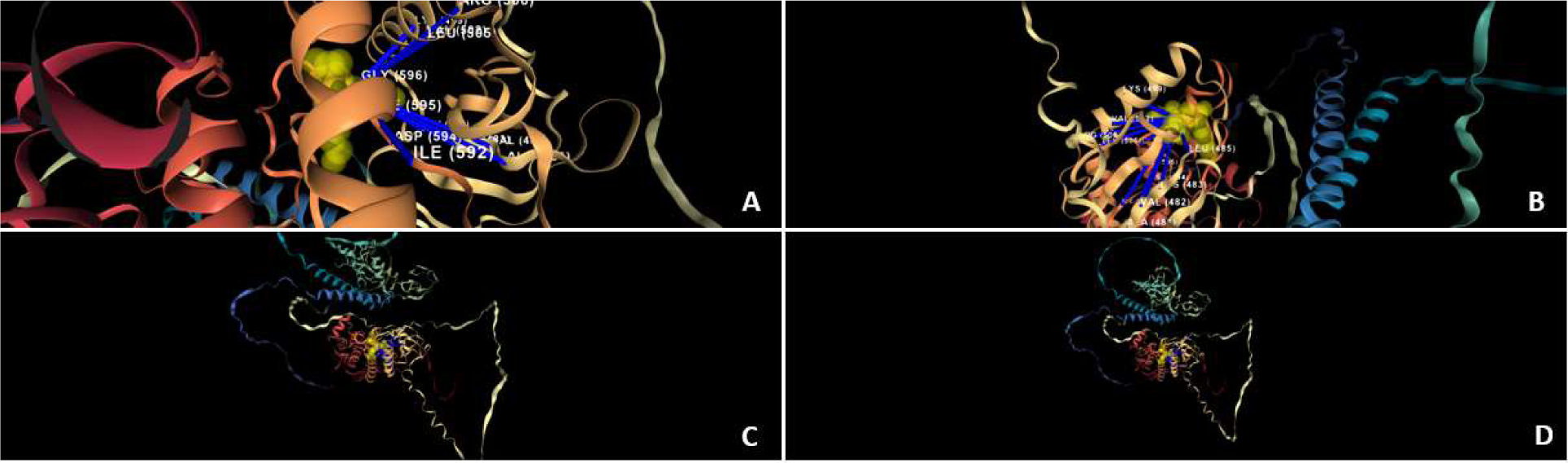
Sequence model explanation of BRAF V600E mutation. Blue lines refer to the residues that contributed to the predicted outcome according to the protein language model, with connection width reflecting the relative contribution. The sequence model uses mainly residues from the DFG motif (594 to 596) and from the ATP binding site, such as 483, 501 and nearby residues. 10A: Focus on the DGF motif area. 10B: Focus on the ATP binding site area. 10C: Focus on the mutation area. 10D: Overall view of the BRAF protein.

**Figure 11:**
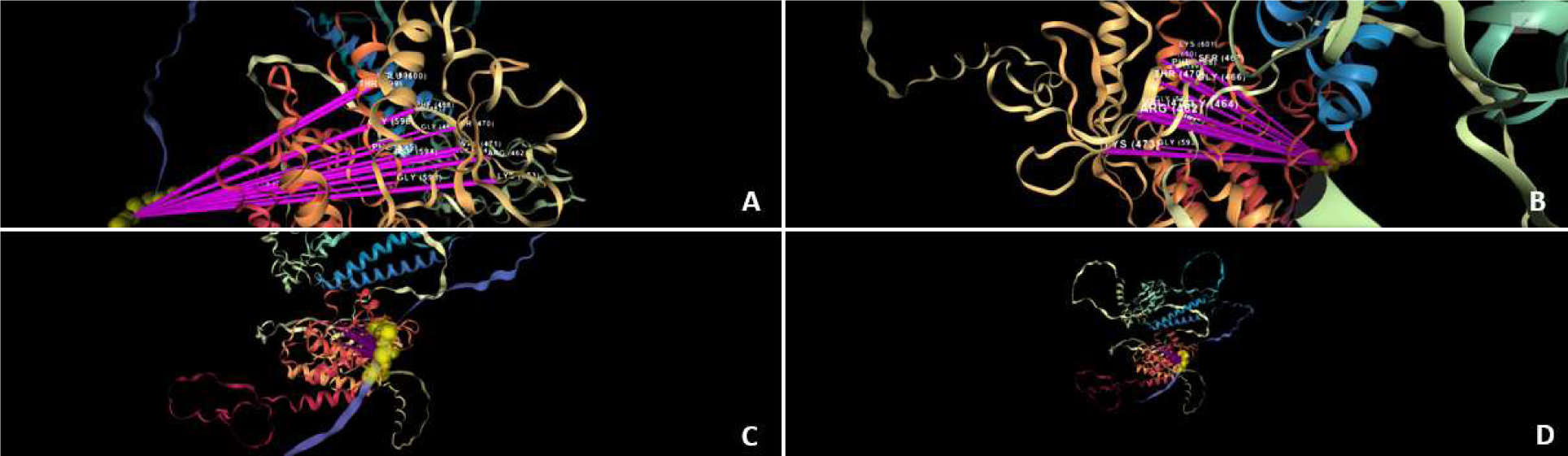
Structure model explanation of BRAF V600E mutation. Pink lines refer to the residues that contributed to the predicted outcome, with connection width reflecting the relative contribution. The structure model finds mainly the residues from the activation loop (A-loop): residues 594 to 600, and the Glycine-rich loop (P-loop) – residues 464 to 471 - as contributing the most to the classifier decision. 11A: Focus on the A-loop area. 11B: focus on the P-loop area. 11C: Focus on the mutation area. 11D: Overall view of the BRAF protein.

**Figure 12:**
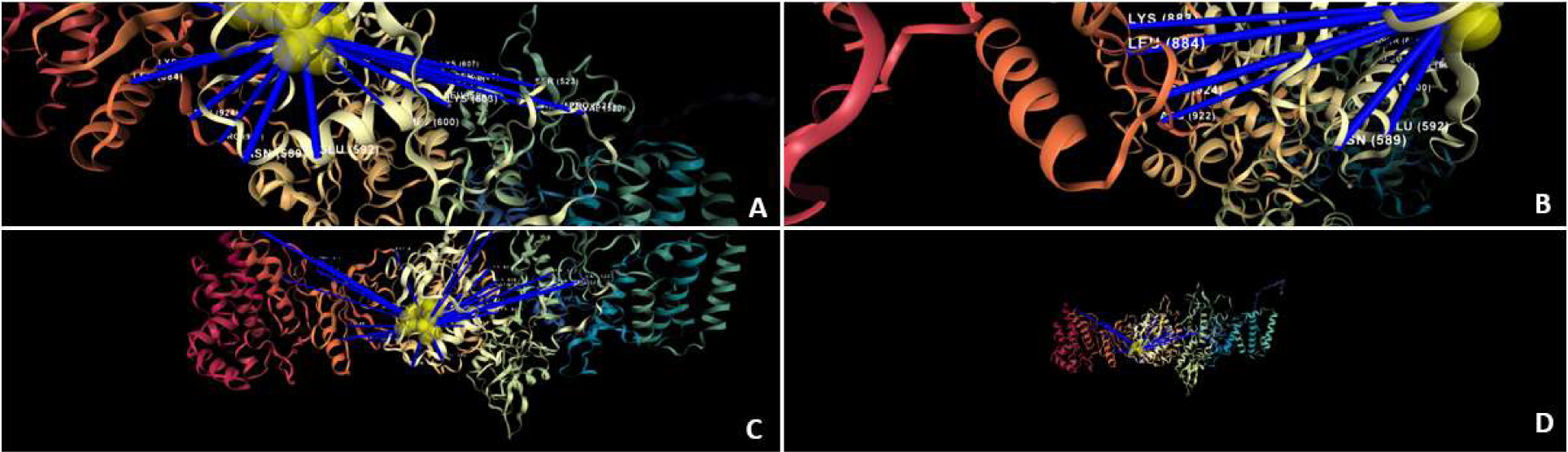
JAK2 V617F Sequence model explanation. Blue lines refer to the residues that contributed to the predicted outcome, with connection width reflecting the relative contribution. The sequence model focuses on residues around 588-607, 683, 592, in JH1 domain, and residues around 883-885, 922-925, 935-947 in JH2 pseudokinase domain, which contribute to the stabilization of the JH2-JH1 interaction through salt bridging or van der Waals forces. 12A: Focus on JH1 (the kinase domain) area. 12B: Focus on JH2 (the pseudokinase domain) area. 12C: Focus on the mutation area. 12D: Overall view of the JAK2 protein.

**Figure 13:**
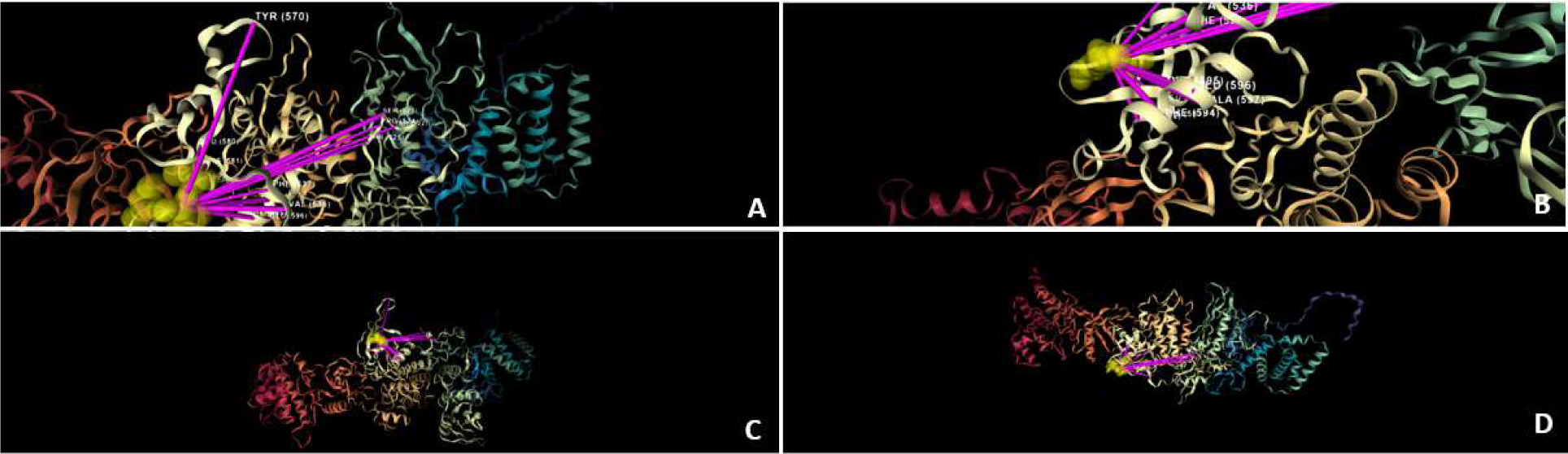
JAK2 V617F Structure model explanation. Pink lines refer to the residues that contributed to the predicted outcome, with connection width reflecting the relative contribution. The structure model finds, besides residues 523 and 570 which are crucial to the auto-inhibition process of the wild type form, those in the Phenyl ring of residues around 594 and the residues involved in the phosphorylation of JH2 domain and thus the SH2-JH1 interaction. 13A: Focus on the auto-inhibition area in JH1. 13B: Focus on the Phenyl ring area. 13C: Focus on the mutation area. 13D: Overall view of the JAK2 protein.

**Figure 14:**
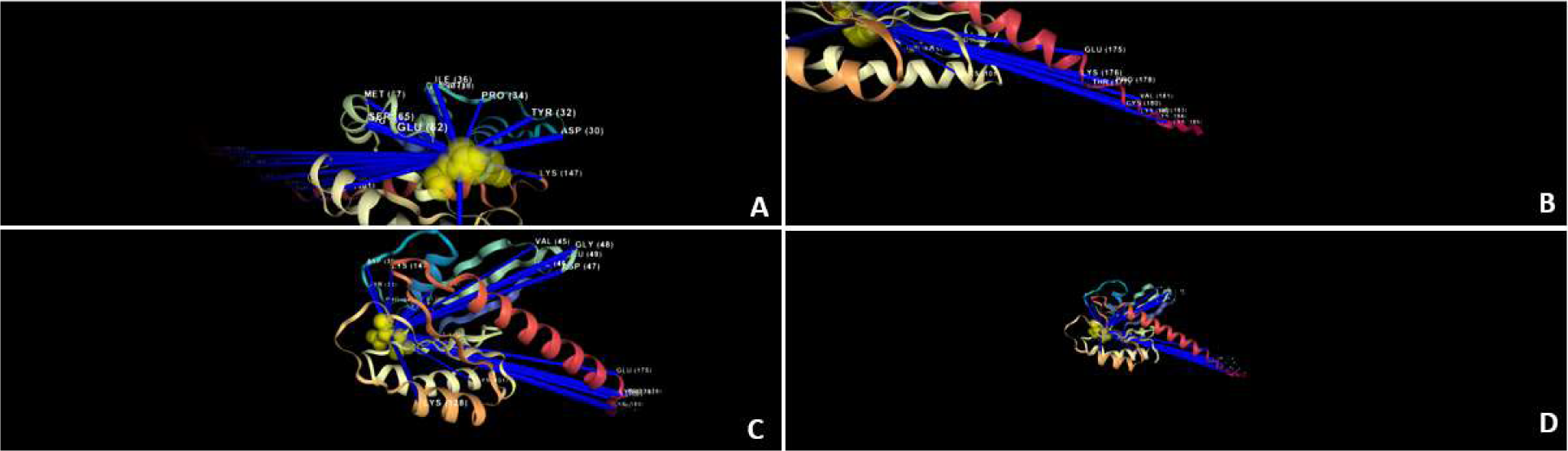
KRAS G12D Sequence model explanation. Blue lines refer to the residues that contributed to the predicted outcome, with connection width reflecting the relative contribution. The sequence model utilizes residues mainly from the allosteric sites in switch I and II: residues 30-40 and 58-70, respectively; and the membrane association domain - C-terminal structural element, named the hypervariable region (HVR), which plays a crucial role in anchoring RAS to the membrane in residues 180-188. In addition, it utilizes lysine residues such as K101, K104, K128 and K147 which were observed to function as acetylation sites. 14A: Focus on switches I and II area. 14B: Focus on the C terminal area. 14C: Focus on the mutation area. 14D: Overall view of the KRAS protein.

**Figure 15:**
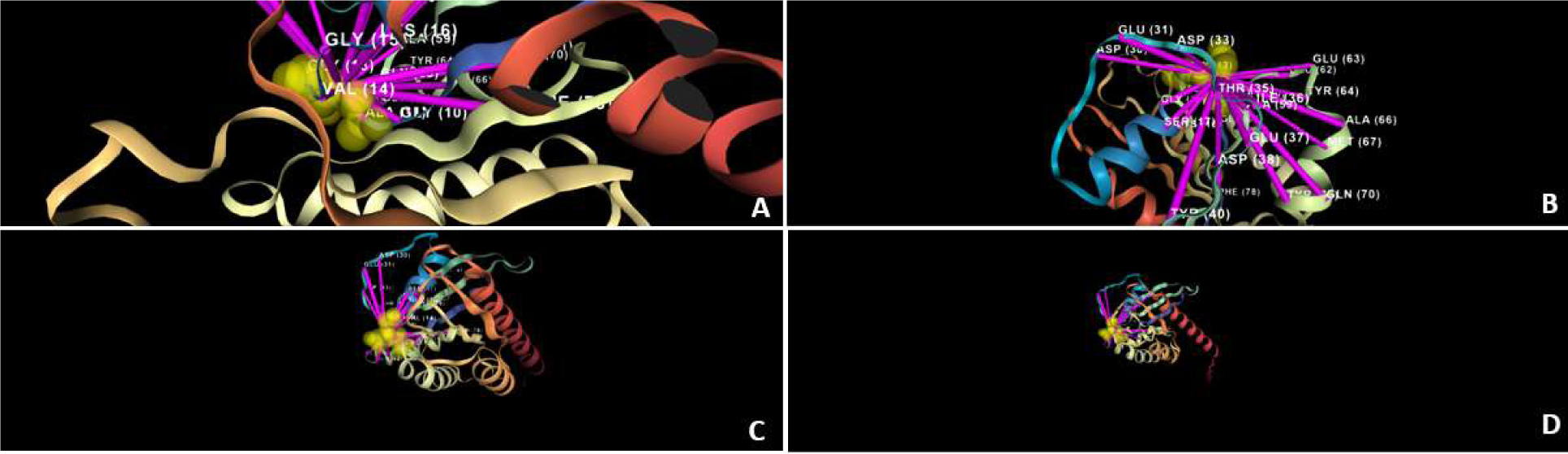
KRAS G12D Structure model explanation. Pink lines refer to the residues that contributed to the predicted outcome, with connection width reflecting the relative contribution. The structure model focuses on the P-loop, in residues 10-17, and Switch I and II regions, which undergo conformational changes upon GTP or GDP binding. The G12D mutation alters the dynamics of these regions, affecting how KRAS interacts with GTPase-activating proteins (GAPs), Guanine nucleotide exchange factors (GEFs), and downstream effectors. 15A: Focus on the P-loop area. 15B: Focus on switches I and II area. 15C: Focus on the mutation area. 15D: Overall view of the KRAS protein.

**Figure 16:**
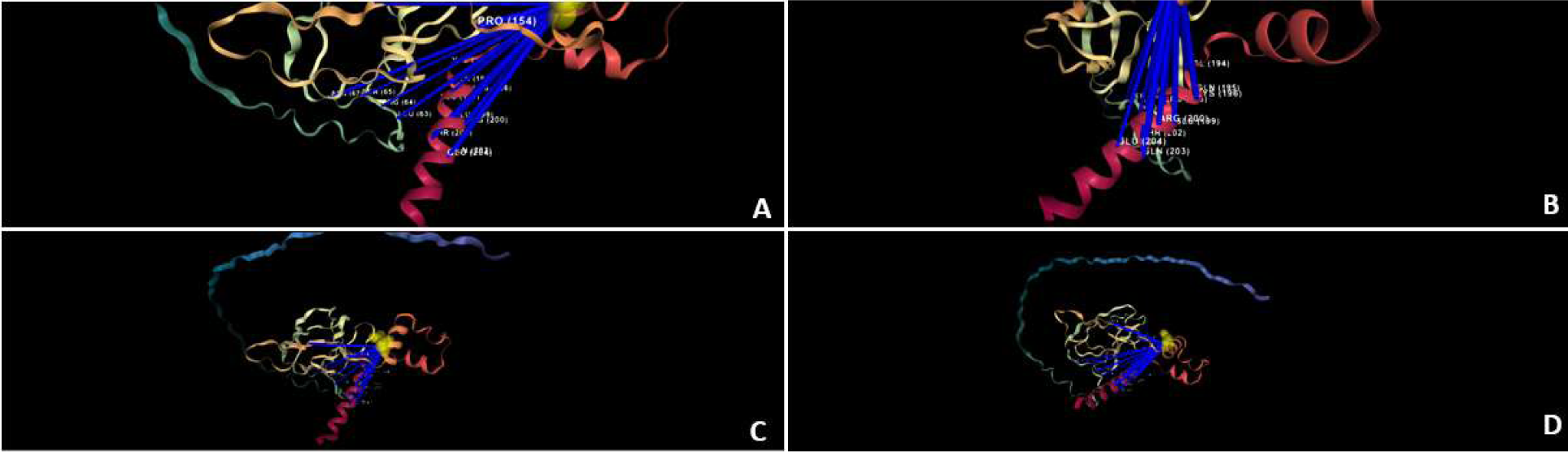
VHL R161Q Sequence model explanation. Blue lines refer to the residues that contributed to the predicted outcome, with connection width reflecting the relative contribution. The sequence model predicts the pathogenicity using residues from the β-domain of pVHL (residues 63–154 and 193–204), which are involved in the binding sites of both elongin-C and HIF. 16A: Focus on the β-domain. 16B: Different view of the β-domain. 16C: Focus on the mutation area. 16D: Overall view of the VHL protein.

**Figure 17:**
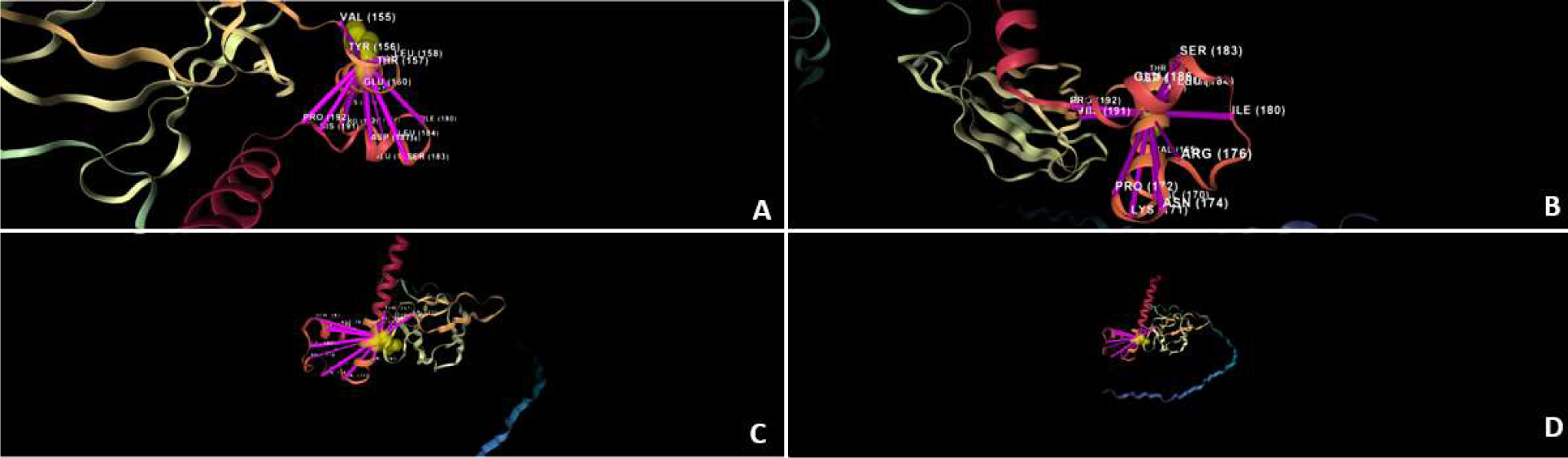
VHL R161Q Structure model explanation. Pink lines refer to the residues that contributed to the predicted outcome, with connection width reflecting the relative contribution. The structure model focuses on residues from α-helical domain (residues 155–192) which directly contacts elongin-C. 17A: Focus on the α-helical domain. 17B: A different view of the α-helical domain. 17C: Focus on the mutation area. 17D: Overall view of the VHL protein.

## Discussion

Since the impressive decrease in sequencing prices in the last years, genomic testing such as whole exome or whole genome sequencing is more rapid and affordable than ever. Rare disease diagnosis, prenatal genetic tests, and oncogenetics among many other medical fields are utilizing the next generation sequencing (NGS) technologies in the daily clinical routine. In this genomic arena, the prevalent missense mutations are unique compared to other types of mutations. While nonsense and frameshift mutations frequently result in significantly dysfunctional proteins with altered function and/or stability, the effect of missense mutations is sometimes more nuanced and its biological and/or clinical implications are not self-evident. The wide range of possible effects on protein structure and function caused by missense mutations motivated the development of a multitude of variant effect prediction methods over the last two decades, aiming to determine the pathogenicity of the mutations whose effect is not clear and are therefore termed VUS. Notable tools such as CADD^1^, DOUGEN2^50^, REVEL^2^ and lately AlphaMissense^51^ were built upon various structural and evolutionary data to try and classify different variants.

In this article, we have used a novel approach by integrating state-of-the-art deep learning algorithms such as the protein language model (PLM) and graph neural network (GNN) with calculation of more traditional features such as ΔΔG, Root Mean Square Deviation (RMSD), BLOSUM substitution values and molecular weight, amongst others. This ensemble algorithm, called TriVIAl, shows cutting edge classification accuracy in the benign versus pathogenic discrimination challenge.

The TriVIAl algorithm achieved state-of-the-art performance with an AUC-ROC of 0.887, marginally better than AlphaMissense score of 0.886 and superior to, metaRNN and ClinPred, whose score was 0.82. Furthermore, TriVIAl demonstrated superior performance in additional metrics; it achieved a precision-recall curve (PRC) score of 0.67, compared to 0.63 by AlphaMissense. Similarly, TriVIAl’s Brier score was 0.16, better than AlphaMissense’s 0.32.

In the era of affordable and accessible genomic sequencing the interpretation of VUS is a major challenge in the clinical practice of genetics and oncology. Critical decisions such as the recommendation for termination of pregnancy or the use of a targeted drug for cancer patients are often faced with the uncertain implications of VUS reported in such cases. There are several criteria that are currently taken into consideration when the meaning and pathogenicity of a point mutation is explored such as the information regarding previous identification of similar mutations in the particular gene or the identification of a mutation in the vicinity of a known mutation, the type of amino acid substitution, evolutionary conservation and more. Yet the number of VUS reported is significant, resulting in a practical and emotional burden for the patients, families and caregivers. The AlphaMissense approach illustrates in a very convincing way the contribution of PLM to the exploration of VUS. The AlphaMissense and similar approaches increase our understanding of the predicted damage attributed to the protein structure and function by the specific alteration identified. There can be however some context dependent disagreement between the effect of a mutation on the structure and the function of the protein and its relevance to the disease question. The TriVIAl approach we developed is aimed to take such context understanding into consideration when determining the clinical relevance of a missense mutation. There are numerous examples where the biological and medical effect of a mutated protein is dependent not only on the demonstration that the protein product is crippled but also on whether the mutation results in a loss of function or gain of function This is best exemplified when dealing with mutated genes in cancer, as depicted in table 2 and supplementary case studies 1-4. Some mutations indiscriminately distributed along the KRAS oncogene, including truncating mutations, can inactivate this protein but usually will not contribute to cancer development and progression. On the other hand, activating mutations that change specific amino acids in the hotspot positions 12, 13, 59, 61 of the protein results in the GTP-bound conformation and constitutive signaling which will contribute to malignant transformation. Similarly, activating mutations in the catalytic domain of protein tyrosine kinase growth factor receptors result in gain of function, activation and enhanced signaling thus supporting malignant transformation. An inactivating or a neutral mutation in the same receptors usually will not enhance cancer development. A special form of gain of function is seen in some unique proteins, such as IDH1 and IDH2. The pathogenic mutations which are localized to a very precise hotspot, mainly codons 132 and 172 respectively provide the protein with “neooncogene” biochemical activity. This novel biochemical function prevalent in AML, brain tumors and other cancers results in the generation of 2-hydroxyglutarate instead of α-ketoglutarate and contributes to the malignant phenotype by affecting demethylation of DNA, RNA and histones.

**Table 2:**
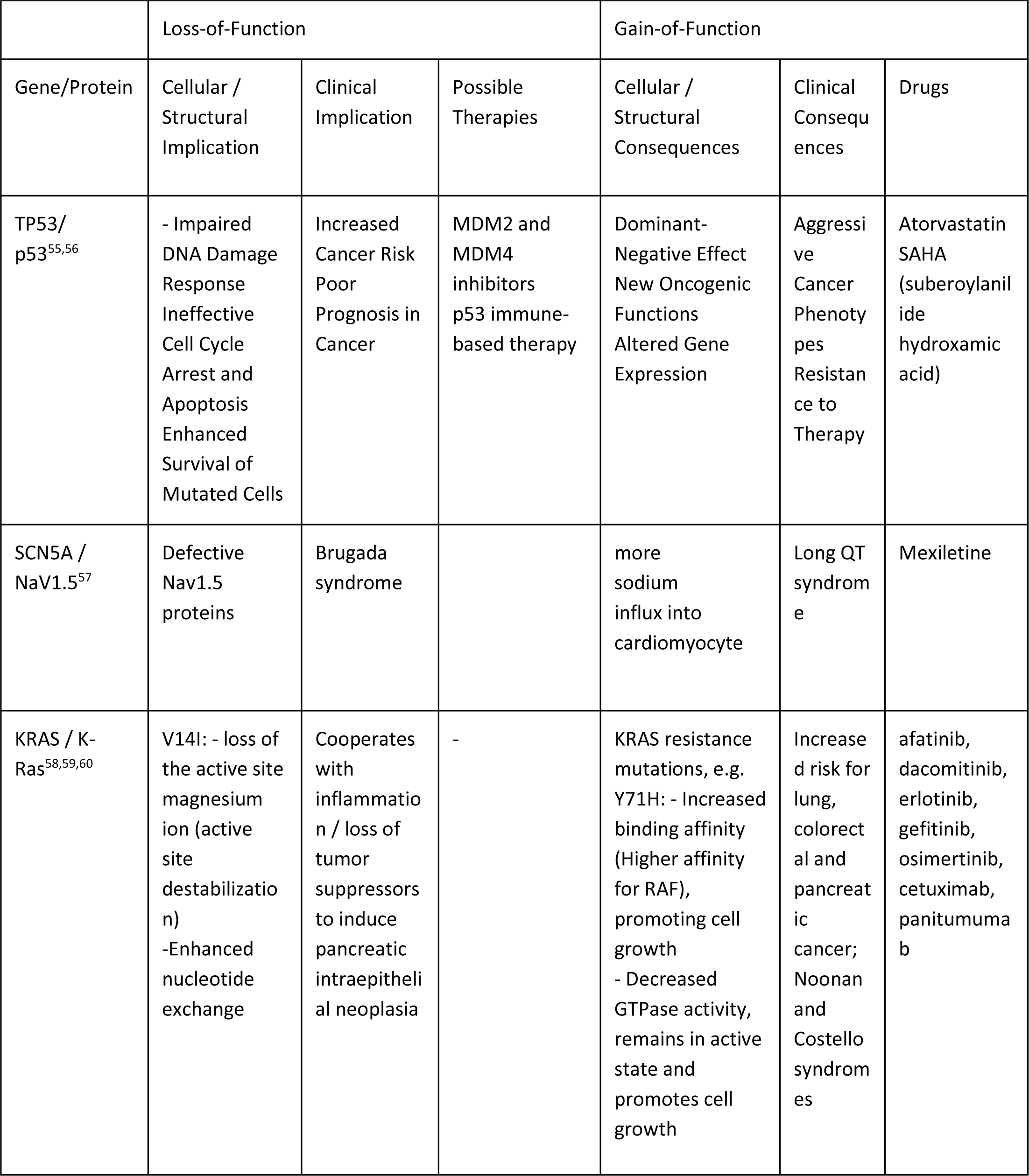
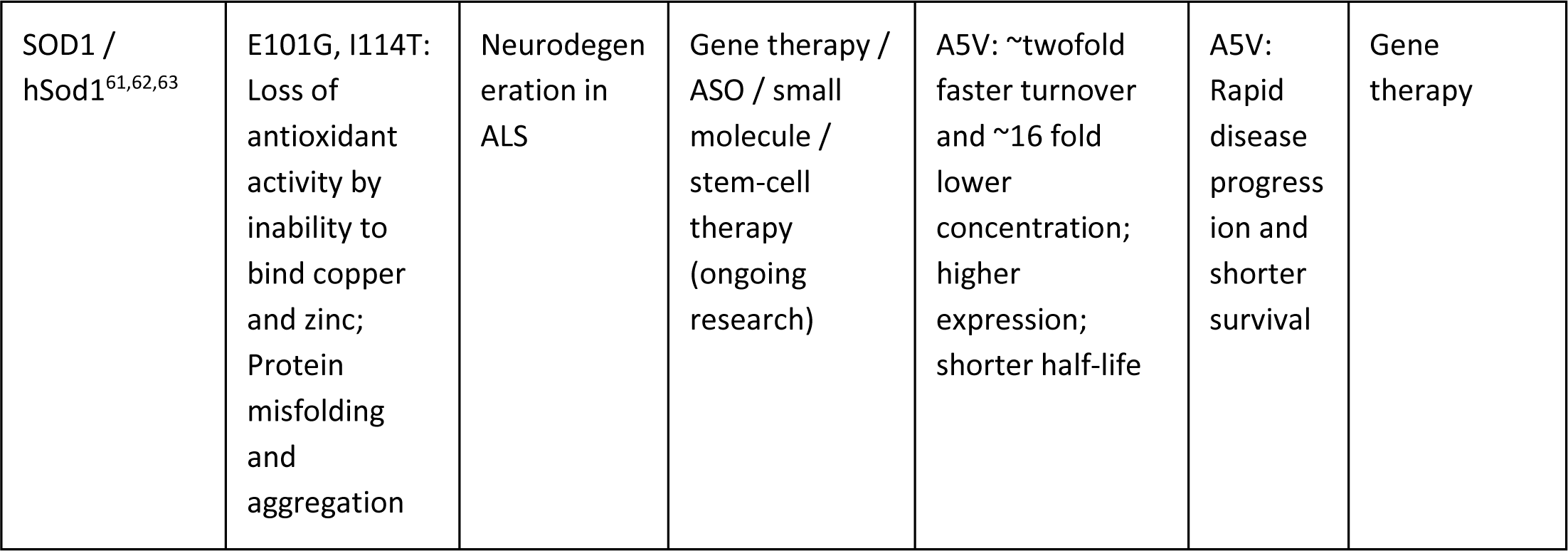
Examples for genes with both gain and loss of function and their putative direct and indirect implications in molecular and clinical aspects.

When dealing with tumor suppressor genes, the contribution of mutations to cancer depends on elimination or reduction of the activity. Multiple sites spread along the protein are expected to inactivate tumor suppression, including truncation of the protein due to nonsense mutations. This principle for differentiating oncogenes from tumor suppressor genes was termed the 20/20 rule as discussed in Vogelstein 2013.^52^ The above distinction is not without exceptions, where some cancer related proteins can function as either oncogenes or tumor suppressor genes, depending on the tumor type. For example, activating mutations turn the NOTCH protein into an active oncogene in T cell acute lymphoblastic leukemia and chronic lymphocytic leukemia while in head and neck cancer mutated NOTCH functions as a tumor suppressor. The FMS gene, encoding for macrophage colony stimulating receptor, can contribute to cancerogenesis when both activated or inactivated by mutations, depending on the specific malignant state. p53 the prototypic tumor suppressor gene may have in addition to loss of function effects also gain of function features ^53^. In some genes mutations in different regions may affect differently the biological activity of the mutated protein. For example, the APC tumor suppressor gene predisposes to familial polyposis syndrome and the development of colon cancer when mutations are located in the 5’ region encoding for the first N terminal 1600 amino acids, only when the mutations result in truncation. Mutations in the APC 3’ part, encoding for the terminal 1200 amino acids, are not associated with increased cancer predisposition. It is clear that an algorithm that takes into consideration the gain or loss of function attributed by the mutation contributes to improved characterization of the effect of the mutation.

The nature of the biochemical outcome of a mutation GOF vs LOF is relevant also to genetic disorders. The study of the globin genes and proteins paved the way for basic molecular research and its translation to clinical applications. Mutations in these genes result in thalassemia and hemoglobinopathies, very prevalent genetic diseases associated with major morbidity and mortality. Mutations that cripple the α-globin genes usually result in alpha thalassemia, and are definitely pathogenic. Yet in patients with beta thalassemia major, who usually are dependent on frequent blood transfusions, the co-inheritance of an α-globin mutation decreases the globin chain imbalance and attenuates the severity of anemia. The α-globin mutation in this case is therefore beneficial. Homozygotes for the single base-pair point mutation at position 6 in the β-globin gene resulting in the substitution of the amino acid valine for glutamic acid at position 6 in the β-globin chain (sickle gene mutation) are clearly affected by severe disease. On the other hand, the hematologic effect of this mutation in the heterozygous state is minimal while carriers are relatively protected against deadly malaria. Therefore, sickle gene mutation is actually beneficial for those living in areas where malaria is endemic. This further illustrates the importance of taking into consideration the specific context when interpreting whether a mutation in pathogenic, neutral or beneficial. This principle of context dependent interpretation was surprisingly demonstrated when mutations in the erythropoietin receptor resulting in erythrocytosis, known to be a risk factor for thrombotic events, proved to be beneficial for E. Mäntyranta, the Finnish cross-country skier who won four Olympic gold medals, thanks to the improved tissue oxygen delivery.

Our TriVIAl approach takes into consideration the relevance of the effect of specific mutations, be it gain or loss of function, to the specific cellular, tissue, organismal and environmental setting. Yet the current ensemble presented here does not take into consideration all possible interacting and milieu associated factors.

Future tools for prediction of mutation pathogenicity will benefit by adding to the interpretation, the information not only of the biochemical effects but also the incorporation of medical and contextual data directly extracted from the Electronic Health Record (EHR) and comprehensive databases.

The algorithm we developed has an additional advantage when aiming at providing the clinician or geneticist with a practical tool for improving medical decisions. The use of AI based medicine is expected to improve diagnosis and care yet like all new tools it is expected not to be error free. The question of who bears the blame when AI Medicine goes wrong is actively discussed^54^ The TriVIAI ensemble, when interpreting the mutation meaning and significance, is based on an explainability feature instead of a black box situation, thus enabling the clinician to follow and visualize the processes that lead to the AI derived conclusion and recommendation. This feature allows the analysts to delve into the detailed structure of the protein itself and to explore different active sites or functions of the protein, giving more biological reasoning and insights into the decision. We expect that this feature will be useful when handling VUS variants in well-defined and studied proteins and also for structure-driven novel drug design. It will ease the burden and responsibility of the caregiver when adopting the AI recommendation.

In conclusion, the new ensemble which fuses classical features such as energy and molecular properties, along with a strong PLM model that has learnt the distribution of protein sequences and their underlying derivatives, and the 3D GNN that enabled fine-grained amino acid interactions provides a useful AI tool that increases the ability to decipher the interpretation of missense mutations while integrating biological insights with visualization and explainability.

## Acknowledgements

This work was supported by the infrastructure grant of the Israel Innovation Authority, Israel Ministry of Health and the National Headquarter “Digital Israel”. The authors thank the Kahn Family Foundation for the continuous support of their research. G.R. is supported by the Flight Attendant Medical Research Institute (FAMRI) and by a grant from the Varda and Boaz Dotan Research Center in Hemato-Oncology, Tel Aviv University. G.R. holds the Djerassi Chair in Oncology at the Tel Aviv University.

## Data Availability

Public datasets including ClinVar, COSMIC, and dbNSFP4.4a, as well as the AlphaFold Protein Structure Database, are freely accessible for download and use. The test datasets were sourced from Zhang et. al.^38^ and are available in Supplementary Table 2. The LOF and GOF annotations taken from the GOF/LOF database^,48^ are also freely available. HGMD Pro 2022.2 is available only under a paid license.

## Code Availability

The code will be made public soon in GitHub.

## Case Studies

### Case Study 1: BRAF p.V600E

The BRAF protein takes role in intracellular signaling pathways that regulate cell growth. It is known to be mutated in several human cancers, such as melanoma, non-small-cells lung carcinoma, adenocarcinoma of the lung, and glioblastoma among others.

The V600E mutation, a substitution of Valine with Glutamic acid, causes the protein to be phosphomimetic because of the negative charge of the acidic Glutamic acid. It mimics the phosphorylation of other residues in the activation segment – specifically Threonine (T) at position 599 and Serine (S) at position 602 which activate the wild-type protein. The V600E mutation disrupts the normal interaction between the protein’s Glycine-rich loop (P-loop) and the activation loop (A-loop), which in their usual state help regulate the protein’s activity. The loss of BRAF inhibition control is oncogenic, leading to poor suppression of tumor growth. Figures 10 and 11 present visualizations showing the classifier explanations for the “Pathogenic” outcome prediction. The sequence model uses mainly residues from the DFG motif (594 to 596) and from the ATP binding, such as 483, 501 and nearby residues (Figure 10). The structure model finds mainly the residues from the activation loop (A-loop) – residues 594 to 600, and the Glycine-rich loop (P-loop) – residues 464 to 471 - as contributing the most to the classifier decision (Figure 11).

### Case Study 2: JAK2 p.V617F

The Janus Kinase 2 (JAK2) gene encodes a key protein in the signaling pathways of several growth factors and cytokines, particularly those involved in hematopoiesis (blood cell production).

The JAK2 V617F mutation is a missense mutation where Valine (V) is substituted by Phenylalanine (F) at the 617th amino acid position in the JAK2 protein.

This mutation is strongly associated with myeloproliferative neoplasms (MPNs), a group of disorders characterized by the excessive production of one or more types of blood cells. The most common MPNs associated with the JAK2 V617F mutation include polycythemia vera, essential thrombocythemia, and primary myelofibrosis.

In the wild type form, the JAK2 protein is activated by the binding of cytokines to their receptors, leading to the controlled production of blood cells. However, the V617F mutation results in a structural change that causes the JAK2 protein to be constitutively active, meaning it signals for blood cell production even in the absence of cytokines. This leads to the overproduction of erythrocytes. The sequence model focuses on residues around 588-607, 683, 592, in JH1; residues around 883-885, 922-925, 935-947 in JH2 pseudokinase domain, which contribute to the stabilization of the JH2-JH1 interaction through salt bridging or van der Waals forces (Figure 12). The structure model finds, besides residues 523 and 570 which are crucial to the auto-inhibition process of the wild type form, those in the Phenyl ring of residues around 594 and the residues involved in the phosphorylation of JH2 domain and thus the SH2-JH1 interaction (Figure 13).

These explanations are in-line with experimental structural biology findings, such as Chen, E. et al.^70^ and Gou, P. et al.^71^.

### Case study 3: KRAS p.G12D

GTPase KRas (KRAS) is a signal transducer protein, which plays an essential role in various cellular signaling events such as in regulation of cell proliferation, differentiation and survival. KRAS G12D mutation results in an amino acid substitution at codon 12 from Glycine (G) to Aspartic acid (D), contributing to uncontrolled cell growth and cancer development. The sequence model utilizes residues mainly from the allosteric sites in switch I and II, residues 30-40 and 58-70, respectively, and the membrane association domain - C-terminal structural element, named the hypervariable region (HVR), which plays a crucial role in anchoring RAS to the membrane in residues 180-188 ^72^. In addition, it utilizes lysine residues such as K101, K104, K128 and K147 which were observed to be acetylation sites by Knyphausen P., et al^73^ (Figure 14). The structure model focuses on the P-loop, in residues 10-17, and Switch I and II regions, which undergo conformational changes upon GTP or GDP binding. The G12D mutation alters the dynamics of these regions, affecting how KRAS interacts with GTPase-activating proteins (GAPs), Guanine nucleotide exchange factors (GEFs), and downstream effectors^74^ (Figure 15)

### Case Study 4: VHL p.R161Q

The von-Hippel – Lindau gene (VHL) is a tumor suppression gene which plays a critical role in in regulating cell growth and division. The VHL protein which is encoded by this gene, pVHL, is involved in the degradation of hypoxia-inducible factors (HIFs) transcription factors that respond to change in the oxygen levels within the cellular environment. Under normal oxygen conditions, VHL binds to HIFs and targets them for degradation, preventing the activation of genes that promote cell survival and angiogenesis. pVHL forms a ternary complex with the elongin C and elongin B proteins (also known as VCB complex), which is essential for pVHL stability and function.

The R161Q mutation in pVHL isoform 30 results in the substitution of Arginine (R) with Glutamine (Q) at position 161 position of the VHL amino acid sequence.^75^

The sequence model predicts the pathogenicity using residues from the β-domain of pVHL (residues 63–154 and 193–204), which are involved in the binding sites of both elongin-C and HIF (Figure 16). The structure model focuses on residues from α-helical domain (residues 155–192) which directly contacts elongin-C (Figure 17).

The presented case studies demonstrate the model capabilities in modelling the molecular properties involved in different pathogenicity mechanisms. These modeling also provides the end user with valuable insights into the etiology of the mutations and their implications. Understanding the mechanisms of mutations also paves the way for the development of more effective and targeted treatment strategies.

