## Supplemental Table 1 + Appendix for "Mutation Pathogenicity Prediction by a Biology Based Explainable AI Multi-Modal Algorithm"

### Supplementary

| Feature name | Feature type | Explanation |
|---|---|---|
| Total energy | float | ddG between the wild type and the mutant protein |
| Backbone Hbond | float | |
| Sidechain Hbond | float | |
| Van der Waals | float | |
| Electrostatics | float | |
| Solvation Polar | float | |
| Solvation Hydrophobic | float | |
| Van der Waals clashes | float | |
| Entropy sidechain | float | |
| Entropy mainchain | float | |
| CIS bond | float | |
| Torsional clash | float | |
| Backbone clash | float | |
| Helix dipole | float | |
| Disulfide | float | |
| Energy ionization | float | |
| RMSD | float | Root mean square deviation between the superimposed wild type and mutant proteins |
| BLOSUM45 | integer | Blocks Substitution Matrix generated for sequences with minimum percentage identity of 45 |
| BLOSUM62 | integer | Blocks Substitution Matrix generated for sequences with minimum percentage identity of 62 |
| BLOSUM80 | integer | Blocks Substitution Matrix generated for sequences with minimum percentage identity of 80 |
| Molecular weight * | float | Molecular weight |
| Aromaticity* | float | Aromaticity calculated according to Lobry, 1994 |
| Instability index* | float | Instability index calculated according to Guruprasad et al 1990 |
| Isoelectric point* | float | Isoelectric point |
| Flexibility* | float | Flexibility calculated according to Vihinen, 1994 |
| Gravy* | float | Gravy calculated according to Kyte and Doolittle |

**Supplementary Table 1**: Protein tabular features. Features marked with a start (*) refer to features that were computed for both the wild-type and the mutant protein

### Appendix

#### A. Software and Frameworks

TriVIAl was developed using Python3.9 with the following libraries and frameworks: BioPython[64] for sequence analysis algorithms; BioPandas for efficient protein structure parsing[65]; Scikit-Learn[66], PyTorch[67], Deep Graph Library (DGL)[68], and Hugging Face Transformers[69] for the models' development and training. For full details please refer to the public repository at GitHub.

#### A.1 Tabular Model Description

The tabular features as described in Supplementary Table 1 were first transformed by scaling column-wise to a normalized range. The tabular model consists of seven linear layers with increased number of output features, and a final output layer with a size unified across all modalities, of 256 channels. All linear layers are followed by Leaky ReLU activations, and dropout with ratio = 0.25 was applied after the 1$^{st}$, 2$^{nd}$, 3$^{rd}$, and 6$^{th}$ layers.

#### A.2 Structural Model Description

TriVIAl GNN module contains several blocks for updating the nodes features and a readout function that obtains the final graph-level representation. The readout function consists of fused edges and nodes readout followed by linear layers. In addition, the global readout is enhanced with an attention layer running on the concatenated pooling of the mutation across several intermediate layers.

##### A.2.1 E(3) Equivariant Graph Convolution Network (EGCN):

In the EGCN backbone, the model is built upon nine graph convolutional layers, chosen due to their E(3) equivariance properties: These layers can handle translation, rotation, and reflections, making them particularly suitable for molecular data analysis, such as protein structures. The reflection equivariance is important for correctly interpreting chiral molecules, which have non-superimposable mirror images, a common characteristic in molecular biology. The network uses residual connections and attention operations for aggregated embedding as part of the equivariant message passing.

##### A.2.1 Graph Attention Network (GAT):

The GAT backbone uses attention mechanism to dynamically determine the importance of each neighbor's features. In addition to the original GAT proposal which uses nodes features, we augment the model with edge attention as well. The attention layer computes the coefficients between nodes and edges; these coefficients indicate the importance of one node's and edge's features to another. For each pair of nodes, and similarly for each pair of edges, an attention score is computed, using a shared learnable linear transformation applied to their features, followed by Leaky ReLU and softmax normalization. The GAT backbone is built with six GAT blocks, each with five heads for multi-head attention.
