## Supplemental Table 2 for "Mutation Pathogenicity Prediction by a Biology Based Explainable AI Multi-Modal Algorithm"

**Variants with functional readout data from deep mutational scan experiments of BRCA1**

| chrom | pos | ref | alt | ref_aa | alt_aa | genename | transcript_id | CADD_raw_rankscore | MPC_rankscore | PrimateAI_rankscore | M-CAP_rankscore | REVEL_rankscore | gMVP_rankscore | MVP_rankscore | ClinPred | BayesDel | EVmutation | target |
|---|---|---|---|---|---|---|---|---|---|---|---|---|---|---|---|---|---|---|
| 17 | 43045705 | T | C | I | M | BRCA1 | ENST00000471181 | 0.47797 | 0.43105 | 0.22602 | 0.90532 | 0.84159 | 0.2783061203 | 0.8581 | 0.864531219 | 0.177813 | 5.92524034 | 0 |
| 17 | 43045706 | A | C | I | R | BRCA1 | ENST00000471181 | 0.71746 | 0.54049 | 0.29109 | 0.98001 | 0.88307 | 0.5842704821 | 0.97297 | 0.9831917882 | 0.297077 | 5.92524034 | 0 |
| 17 | 43045706 | A | G | I | T | BRCA1 | ENST00000471181 | 0.68699 | 0.4426 | 0.35451 | 0.95567 | 0.87099 | 0.3682824377 | 0.97061 | 0.9608803988 | 0.260788 | 5.92524034 | 0 |
| 17 | 43045707 | T | A | I | L | BRCA1 | ENST00000471181 | 0.46314 | 0.21961 | 0.251 | 0.92776 | 0.78535 | 0.2632659336 | 0.70751 | 0.7369521856 | 0.101854 | 4.500494054 | 0 |
| 17 | 43045707 | T | C | I | V | BRCA1 | ENST00000471181 | 0.17781 | 0.0826 | 0.18219 | 0.64684 | 0.74306 | 0.1605788825 | 0.49429 | 0.0641624406 | 0.0513818 | 1.440830029 | 0 |
| 17 | 43045707 | T | G | I | L | BRCA1 | ENST00000471181 | 0.46955 | 0.21961 | 0.251 | 0.92776 | 0.78535 | 0.2632659336 | 0.70751 | 0.7369521856 | 0.101855 | 4.500494054 | 0 |
| 17 | 43045709 | A | C | L | R | BRCA1 | ENST00000471181 | 0.88591 | 0.51675 | 0.3709 | 0.98287 | 0.89963 | 0.5319424791 | 0.87142 | 0.9828765988 | 0.442171 | 6.411878665 | 0 |
| 17 | 43045709 | A | G | L | P | BRCA1 | ENST00000471181 | 0.88904 | 0.53493 | 0.41387 | 0.98326 | 0.90711 | 0.6909943191 | 0.88874 | 0.9940621257 | 0.448673 | 6.411878665 | 1 |
| 17 | 43045709 | A | T | L | Q | BRCA1 | ENST00000471181 | 0.87821 | 0.45914 | 0.3092 | 0.98203 | 0.89739 | 0.496012473 | 0.90834 | 0.9828765988 | 0.409497 | 6.411878665 | 0 |
| 17 | 43045710 | G | C | L | V | BRCA1 | ENST00000471181 | 0.77537 | 0.45046 | 0.23016 | 0.94638 | 0.86098 | 0.2570332047 | 0.95426 | 0.9807201028 | 0.263265 | 6.411878665 | 0 |
| 17 | 43045710 | G | T | L | M | BRCA1 | ENST00000471181 | 0.80424 | 0.447 | 0.25889 | 0.93374 | 0.8233 | 0.2666808609 | 0.96712 | 0.8409406738 | 0.272501 | 5.608416024 | 0 |
| 17 | 43045712 | T | A | Y | F | BRCA1 | ENST00000471181 | 0.75798 | 0.4381 | 0.40157 | 0.98258 | 0.93162 | 0.4984845252 | 0.96327 | 0.9910620451 | 0.32623 | 6.530175343 | 0 |
| 17 | 43045712 | T | C | Y | C | BRCA1 | ENST00000471181 | 0.86018 | 0.48925 | 0.48549 | 0.99236 | 0.94913 | 0.7473910293 | 0.97984 | 0.9884004593 | 0.489707 | 6.530175343 | 1 |
| 17 | 43045712 | T | G | Y | S | BRCA1 | ENST00000471181 | 0.82895 | 0.51182 | 0.42422 | 0.99216 | 0.95097 | 0.7953782095 | 0.96907 | 0.9911102057 | 0.50832 | 6.530175343 | 1 |
| 17 | 43045713 | A | C | Y | D | BRCA1 | ENST00000471181 | 0.87496 | 0.55271 | 0.48709 | 0.99227 | 0.94949 | 0.8360414302 | 0.97553 | 0.9911102057 | 0.521449 | 6.530175343 | 1 |
| 17 | 43045713 | A | G | Y | H | BRCA1 | ENST00000471181 | 0.86333 | 0.49506 | 0.49911 | 0.98879 | 0.92844 | 0.734437292 | 0.97155 | 0.9875670671 | 0.372996 | 6.530175343 | 1 |
| 17 | 43045713 | A | T | Y | N | BRCA1 | ENST00000471181 | 0.87038 | 0.53743 | 0.49445 | 0.99248 | 0.94876 | 0.8011276577 | 0.97975 | 0.9821776152 | 0.503117 | 6.530175343 | 1 |
| 17 | 43045715 | G | A | T | I | BRCA1 | ENST00000471181 | 0.51468 | 0.43034 | 0.20571 | 0.9306 | 0.85482 | 0.2453957579 | 0.89987 | 0.9269515872 | 0.248321 | 5.831847079 | 0 |
| 17 | 43045715 | G | C | T | S | BRCA1 | ENST00000471181 | 0.44973 | 0.18543 | 0.11005 | 0.83339 | 0.79998 | 0.1690646628 | 0.87387 | 0.7792036533 | 0.200729 | 3.916008707 | 0 |
| 17 | 43045715 | G | T | T | N | BRCA1 | ENST00000471181 | 0.48704 | 0.42292 | 0.15923 | 0.9161 | 0.82046 | 0.2475052601 | 0.85444 | 0.8949037194 | 0.149408 | 5.831847079 | 0 |
| 17 | 43045716 | T | A | T | S | BRCA1 | ENST00000471181 | 0.35797 | 0.18543 | 0.11005 | 0.78446 | 0.77972 | 0.1690646628 | 0.81827 | 0.5173198581 | 0.165393 | 3.916008707 | 0 |
| 17 | 43045716 | T | C | T | A | BRCA1 | ENST00000471181 | 0.23562 | 0.14724 | 0.0931 | 0.55972 | 0.76289 | 0.1938308768 | 0.74043 | 0.1093363017 | 0.158324 | 2.232428582 | 0 |
| 17 | 43045716 | T | G | T | P | BRCA1 | ENST00000471181 | 0.3075 | 0.12534 | 0.11995 | 0.83127 | 0.77654 | 0.5501090487 | 0.78415 | 0.2493893504 | 0.223937 | 3.042921075 | 0 |
| 17 | 43045717 | G | C | D | E | BRCA1 | ENST00000471181 | 0.33153 | 0.08989 | 0.19504 | 0.81127 | 0.81124 | 0.2179803825 | 0.84939 | 0.7843420506 | 0.178 | 3.645657332 | 0 |
| 17 | 43045717 | G | T | D | E | BRCA1 | ENST00000471181 | 0.32717 | 0.08989 | 0.19504 | 0.81163 | 0.80772 | 0.2179803825 | 0.84939 | 0.8636924624 | 0.177999 | 3.645657332 | 0 |
| 17 | 43045718 | T | A | D | V | BRCA1 | ENST00000471181 | 0.77537 | 0.47964 | 0.17135 | 0.94973 | 0.84321 | 0.5659941297 | 0.83191 | 0.9882785082 | 0.328375 | 5.943015398 | 0 |
| 17 | 43045718 | T | C | D | G | BRCA1 | ENST00000471181 | 0.60164 | 0.46284 | 0.18734 | 0.94292 | 0.83116 | 0.4877248166 | 0.87607 | 0.8591641188 | 0.261679 | 3.602689319 | 0 |
| 17 | 43045718 | T | G | D | A | BRCA1 | ENST00000471181 | 0.63489 | 0.39867 | 0.1808 | 0.94356 | 0.83394 | 0.4623180292 | 0.86266 | 0.7630452895 | 0.253742 | 4.428118983 | 0 |
| 17 | 43045719 | C | A | D | Y | BRCA1 | ENST00000471181 | 0.71634 | 0.47551 | 0.22468 | 0.9807 | 0.90229 | 0.572338404 | 0.88021 | 0.9783862233 | 0.359675 | 5.943015398 | 0 |
| 17 | 43045719 | C | G | D | H | BRCA1 | ENST00000471181 | 0.7088 | 0.4623 | 0.23501 | 0.97351 | 0.8933 | 0.4776192322 | 0.92209 | 0.980559051 | 0.252975 | 5.943015398 | 0 |
| 17 | 43045719 | C | T | D | N | BRCA1 | ENST00000471181 | 0.70696 | 0.36118 | 0.21667 | 0.95593 | 0.82556 | 0.3339691099 | 0.88228 | 0.9610207677 | 0.136659 | 3.302954526 | 0 |
| 17 | 43045721 | A | C | L | R | BRCA1 | ENST00000471181 | 0.86254 | 0.51636 | 0.4047 | 0.98293 | 0.88022 | 0.5419780791 | 0.97463 | 0.9869763255 | 0.313198 | 6.684540167 | 0 |
| 17 | 43045721 | A | G | L | P | BRCA1 | ENST00000471181 | 0.8665 | 0.53457 | 0.44036 | 0.98557 | 0.91357 | 0.6916553976 | 0.97749 | 0.9867870808 | 0.370817 | 6.684540167 | 0 |
| 17 | 43045721 | A | T | L | Q | BRCA1 | ENST00000471181 | 0.85404 | 0.45873 | 0.34778 | 0.98249 | 0.88069 | 0.3698061752 | 0.97861 | 0.9869763255 | 0.298759 | 6.684540167 | 0 |
| 17 | 43045722 | G | C | L | V | BRCA1 | ENST00000471181 | 0.60882 | 0.446 | 0.2833 | 0.9282 | 0.862 | 0.2787234929 | 0.93406 | 0.9742180705 | 0.229526 | 6.684540167 | 0 |
| 17 | 43045722 | G | T | L | M | BRCA1 | ENST00000471181 | 0.64151 | 0.44665 | 0.30936 | 0.93061 | 0.82724 | 0.2733913621 | 0.92467 | 0.9785038829 | 0.197971 | 6.684540167 | 0 |
| 17 | 43045723 | C | A | E | D | BRCA1 | ENST00000471181 | 0.27831 | 0.42993 | 0.24527 | 0.84576 | 0.81356 | 0.2493533053 | 0.83715 | 0.8583576083 | 0.165692 | 3.422035565 | 0 |
| 17 | 43045723 | C | G | E | D | BRCA1 | ENST00000471181 | 0.28344 | 0.42993 | 0.24527 | 0.83139 | 0.81876 | 0.2493533053 | 0.83715 | 0.8583576083 | 0.1657 | 3.422035565 | 0 |
| 17 | 43045724 | T | C | E | G | BRCA1 | ENST00000471181 | 0.9077 | 0.47791 | 0.22853 | 0.95549 | 0.89009 | 0.4331577536 | 0.90376 | 0.8876031637 | 0.221357 | 6.263951092 | 0 |
| 17 | 43045725 | C | G | E | Q | BRCA1 | ENST00000471181 | 0.70439 | 0.44993 | 0.24333 | 0.93073 | 0.8345 | 0.3716605964 | 0.86631 | 0.8895577788 | 0.154198 | 6.263951092 | 0 |
| 17 | 43045725 | C | T | E | K | BRCA1 | ENST00000471181 | 0.76942 | 0.47024 | 0.31623 | 0.96143 | 0.83834 | 0.4087811359 | 0.83326 | 0.9270783067 | 0.237376 | 4.39731932 | 0 |
| 17 | 43045726 | C | G | Q | H | BRCA1 | ENST00000471181 | 0.68875 | 0.4312 | 0.09657 | 0.96559 | 0.88496 | 0.6965603509 | 0.94344 | 0.9699732661 | 0.253928 | 6.265976383 | 0 |
| 17 | 43045727 | T | A | Q | L | BRCA1 | ENST00000471181 | 0.8336 | 0.41409 | 0.06593 | 0.96794 | 0.89422 | 0.5784669412 | 0.89217 | 0.9718090296 | 0.247994 | 6.265976383 | 0 |
| 17 | 43045727 | T | C | Q | R | BRCA1 | ENST00000471181 | 0.46531 | 0.44725 | 0.09533 | 0.90301 | 0.85012 | 0.6502420273 | 0.88862 | 0.8691961169 | 0.142387 | 3.221344429 | 0 |
| 17 | 43045727 | T | G | Q | P | BRCA1 | ENST00000471181 | 0.81418 | 0.15974 | 0.08522 | 0.97066 | 0.91736 | 0.7792248274 | 0.93408 | 0.9098798037 | 0.361284 | 6.265976383 | 1 |
| 17 | 43045728 | G | C | Q | E | BRCA1 | ENST00000471181 | 0.80866 | 0.364 | 0.07168 | 0.94211 | 0.87781 | 0.6225113641 | 0.95113 | 0.9399325848 | 0.23168 | 6.265976383 | 0 |
| 17 | 43045728 | G | T | Q | K | BRCA1 | ENST00000471181 | 0.83821 | 0.39674 | 0.10122 | 0.92843 | 0.86951 | 0.6202189688 | 0.95001 | 0.9565668106 | 0.268528 | 6.265976383 | 0 |
| 17 | 43045729 | G | C | C | W | BRCA1 | ENST00000471181 | 0.74213 | 0.49712 | 0.17008 | 0.91091 | 0.89829 | 0.5732378316 | 0.95741 | 0.8719896078 | 0.148267 | 5.962524673 | 0 |
| 17 | 43045730 | C | A | C | F | BRCA1 | ENST00000471181 | 0.61171 | 0.50397 | 0.13649 | 0.94954 | 0.91314 | 0.4567240147 | 0.95341 | 0.8984569907 | 0.225327 | 5.962524673 | 0 |
| 17 | 43045730 | C | G | C | S | BRCA1 | ENST00000471181 | 0.56269 | 0.41486 | 0.12171 | 0.93116 | 0.87491 | 0.5134191708 | 0.92844 | 0.9016986489 | 0.219715 | 5.962524673 | 0 |
| 17 | 43045730 | C | T | C | Y | BRCA1 | ENST00000471181 | 0.59089 | 0.5327 | 0.21036 | 0.93077 | 0.91778 | 0.4847494882 | 0.97353 | 0.9613015056 | 0.219015 | 5.962524673 | 0 |
| 17 | 43045731 | A | C | C | G | BRCA1 | ENST00000471181 | 0.5234 | 0.45269 | 0.11237 | 0.91022 | 0.87733 | 0.5585846246 | 0.9078 | 0.88608253 | 0.210167 | 5.962524673 | 0 |
| 17 | 43045731 | A | G | C | R | BRCA1 | ENST00000471181 | 0.38112 | 0.54168 | 0.19561 | 0.8256 | 0.69678 | 0.6059728612 | 0.75058 | 0.7334851623 | 0.086372 | 1.574436189 | 0 |
| 17 | 43045731 | A | T | C | S | BRCA1 | ENST00000471181 | 0.53401 | 0.41486 | 0.12171 | 0.94274 | 0.848 | 0.5134191708 | 0.92066 | 0.8147726059 | 0.198794 | 5.962524673 | 0 |
| 17 | 43045732 | C | A | Q | H | BRCA1 | ENST00000471181 | 0.62113 | 0.44086 | 0.08375 | 0.93258 | 0.86703 | 0.4111467242 | 0.92038 | 0.9117088914 | 0.231785 | 5.093086601 | 0 |
| 17 | 43045732 | C | G | Q | H | BRCA1 | ENST00000471181 | 0.62916 | 0.44086 | 0.08375 | 0.93045 | 0.86703 | 0.4111467242 | 0.92038 | 0.7060866952 | 0.10541 | 5.093086601 | 0 |
| 17 | 43045733 | T | A | Q | L | BRCA1 | ENST00000471181 | 0.74531 | 0.44332 | 0.05634 | 0.97303 | 0.88022 | 0.3990634815 | 0.8925 | 0.9592486024 | 0.241615 | 6.260781246 | 0 |
| 17 | 43045733 | T | C | Q | R | BRCA1 | ENST00000471181 | 0.41034 | 0.45747 | 0.07848 | 0.90655 | 0.82836 | 0.2447520309 | 0.80675 | 0.7667226195 | 0.180426 | 3.292420797 | 0 |
| 17 | 43045733 | T | G | Q | P | BRCA1 | ENST00000471181 | 0.74459 | 0.48318 | 0.07403 | 0.97756 | 0.89648 | 0.6248846779 | 0.92285 | 0.8987398148 | 0.288107 | 6.260781246 | 1 |
| 17 | 43045734 | G | C | Q | E | BRCA1 | ENST00000471181 | 0.35819 | 0.44459 | 0.06611 | 0.90475 | 0.83228 | 0.2384864581 | 0.88057 | 0.7634592652 | 0.228173 | 3.478010567 | 0 |
| 17 | 43045734 | G | T | Q | K | BRCA1 | ENST00000471181 | 0.49711 | 0.47796 | 0.08665 | 0.90818 | 0.83172 | 0.2870937755 | 0.88698 | 0.8781657219 | 0.230739 | 5.386970229 | 0 |
| 17 | 43045736 | T | A | Y | F | BRCA1 | ENST00000471181 | 0.74994 | 0.42452 | 0.21557 | 0.93584 | 0.86302 | 0.4909174657 | 0.846 | 0.6780180335 | 0.16697 | 6.286835563 | 0 |
| 17 | 43045736 | T | C | Y | C | BRCA1 | ENST00000471181 | 0.83166 | 0.48724 | 0.27844 | 0.97417 | 0.8873 | 0.6787483716 | 0.91247 | 0.9509183764 | 0.237772 | 6.286835563 | 0 |
| 17 | 43045736 | T | G | Y | S | BRCA1 | ENST00000471181 | 0.82209 | 0.51102 | 0.22858 | 0.97978 | 0.8887 | 0.6698082987 | 0.90933 | 0.9434607625 | 0.278212 | 6.286835563 | 0 |
| 17 | 43045737 | A | G | Y | H | BRCA1 | ENST00000471181 | 0.58996 | 0.49225 | 0.29054 | 0.95091 | 0.90972 | 0.6710803016 | 0.94497 | 0.9380555749 | 0.221516 | 4.421818064 | 0 |
| 17 | 43045737 | A | T | Y | N | BRCA1 | ENST00000471181 | 0.73397 | 0.53701 | 0.28923 | 0.97806 | 0.9045 | 0.6806083151 | 0.95979 | 0.9921137094 | 0.258275 | 6.286835563 | 0 |
| 17 | 43045739 | A | C | L | R | BRCA1 | ENST00000471181 | 0.80361 | 0.51153 | 0.17467 | 0.96859 | 0.88963 | 0.44288082 | 0.85077 | 0.3856360025 | 0.194735 | 6.24638303 | 0 |
| 17 | 43045739 | A | G | L | P | BRCA1 | ENST00000471181 | 0.80929 | 0.53217 | 0.19466 | 0.95423 | 0.89739 | 0.6847149477 | 0.84761 | 0.7964447873 | 0.386983 | 6.24638303 | 0 |
| 17 | 43045739 | A | T | L | H | BRCA1 | ENST00000471181 | 0.78653 | 0.48817 | 0.16002 | 0.96925 | 0.86352 | 0.4194760243 | 0.86443 | 0.8036332471 | 0.256909 | 6.24638303 | 0 |
| 17 | 43045740 | G | A | L | F | BRCA1 | ENST00000471181 | 0.6666 | 0.44968 | 0.10241 | 0.91588 | 0.84267 | 0.258222843 | 0.95942 | 0.7738852807 | 0.208182 | 6.24638303 | 0 |
| 17 | 43045740 | G | C | L | V | BRCA1 | ENST00000471181 | 0.3327 | 0.19917 | 0.07481 | 0.86124 | 0.81298 | 0.2936266854 | 0.89462 | 0.4387552139 | 0.161869 | 3.249021019 | 0 |
| 17 | 43045740 | G | T | L | I | BRCA1 | ENST00000471181 | 0.60332 | 0.20173 | 0.11705 | 0.8543 | 0.78473 | 0.2145050829 | 0.88616 | 0.5877593269 | 0.120642 | 6.24638303 | 0 |
| 17 | 43045742 | G | A | A | V | BRCA1 | ENST00000471181 | 0.78099 | 0.3503 | 0.19839 | 0.96404 | 0.88354 | 0.5375680843 | 0.95483 | 0.9010221362 | 0.185145 | 6.153867861 | 0 |
| 17 | 43045742 | G | C | A | G | BRCA1 | ENST00000471181 | 0.76824 | 0.40229 | 0.18007 | 0.96665 | 0.89784 | 0.4765098789 | 0.96608 | 0.9435557723 | 0.242137 | 6.153867861 | 0 |
| 17 | 43045742 | G | T | A | E | BRCA1 | ENST00000471181 | 0.77283 | 0.4714 | 0.23803 | 0.96778 | 0.8873 | 0.7420767596 | 0.96776 | 0.9725813866 | 0.360123 | 6.153867861 | 1 |
| 17 | 43045743 | C | A | A | S | BRCA1 | ENST00000471181 | 0.46849 | 0.41862 | 0.16545 | 0.9 | 0.84106 | 0.4320735876 | 0.91523 | 0.9149045348 | 0.13052 | 3.102142122 | 0 |
| 17 | 43045743 | C | G | A | P | BRCA1 | ENST00000471181 | 0.68059 | 0.48668 | 0.24918 | 0.97065 | 0.89513 | 0.8314781501 | 0.97541 | 0.9679126143 | 0.364479 | 6.153867861 | 1 |
| 17 | 43045743 | C | T | A | T | BRCA1 | ENST00000471181 | 0.66723 | 0.42365 | 0.19243 | 0.96831 | 0.89648 | 0.4460004964 | 0.96905 | 0.9719617367 | 0.233553 | 6.153867861 | 0 |
| 17 | 43045745 | A | C | V | G | BRCA1 | ENST00000471181 | 0.83197 | 0.47522 | 0.10523 | 0.95838 | 0.90494 | 0.8317857812 | 0.9444 | 0.9876223207 | 0.435617 | 6.042451599 | 1 |
| 17 | 43045745 | A | G | V | A | BRCA1 | ENST00000471181 | 0.79081 | 0.37662 | 0.17952 | 0.93297 | 0.85378 | 0.6918128418 | 0.92918 | 0.9865043163 | 0.29917 | 6.042451599 | 1 |
| 17 | 43045745 | A | T | V | E | BRCA1 | ENST00000471181 | 0.83676 | 0.49213 | 0.16154 | 0.97376 | 0.8933 | 0.8778763043 | 0.92088 | 0.992426753 | 0.43119 | 6.042451599 | 1 |
| 17 | 43045746 | C | A | V | L | BRCA1 | ENST00000471181 | 0.23511 | 0.11632 | 0.15787 | 0.70664 | 0.67661 | 0.6225352844 | 0.76516 | 0.2203506976 | 0.203111 | 5.007432197 | 0 |
| 17 | 43045746 | C | G | V | L | BRCA1 | ENST00000471181 | 0.23965 | 0.11632 | 0.15787 | 0.70384 | 0.67843 | 0.6225352844 | 0.76516 | 0.2203506976 | 0.203111 | 5.007432197 | 0 |
| 17 | 43045746 | C | T | V | I | BRCA1 | ENST00000471181 | 0.14933 | 0.07725 | 0.07242 | 0.61417 | 0.61184 | 0.3724726606 | 0.60621 | 0.06992497385 | 0.0831237 | 2.336496447 | 0 |
| 17 | 43045747 | A | C | S | R | BRCA1 | ENST00000471181 | 0.45942 | 0.46381 | 0.42873 | 0.98951 | 0.90929 | 0.8065436678 | 0.84705 | 0.9738906622 | 0.425935 | 6.530175608 | 1 |
| 17 | 43045747 | A | T | S | R | BRCA1 | ENST00000471181 | 0.45425 | 0.46381 | 0.42873 | 0.98951 | 0.90929 | 0.8065436678 | 0.84705 | 0.9738906622 | 0.425929 | 6.530175608 | 1 |
| 17 | 43045748 | C | A | S | I | BRCA1 | ENST00000471181 | 0.72492 | 0.46677 | 0.40035 | 0.98725 | 0.91986 | 0.6955771816 | 0.97861 | 0.9939095974 | 0.475534 | 6.530175608 | 1 |
| 17 | 43045748 | C | G | S | T | BRCA1 | ENST00000471181 | 0.65445 | 0.09929 | 0.36675 | 0.96889 | 0.8933 | 0.5025590989 | 0.98372 | 0.974770844 | 0.424854 | 6.530175608 | 1 |
| 17 | 43045748 | C | T | S | N | BRCA1 | ENST00000471181 | 0.66288 | 0.35887 | 0.41294 | 0.97784 | 0.85741 | 0.6631355715 | 0.98791 | 0.9848355651 | 0.265914 | 6.530175608 | 1 |
| 17 | 43045749 | T | A | S | C | BRCA1 | ENST00000471181 | 0.85902 | 0.43977 | 0.37559 | 0.9818 | 0.92884 | 0.5383855054 | 0.97194 | 0.988276422 | 0.38003 | 6.530175608 | 0 |
| 17 | 43045749 | T | G | S | R | BRCA1 | ENST00000471181 | 0.85077 | 0.46381 | 0.42873 | 0.98971 | 0.94543 | 0.8065436678 | 0.98174 | 0.9944150448 | 0.483671 | 6.530175608 | 1 |
| 17 | 43045750 | G | C | D | E | BRCA1 | ENST00000471181 | 0.75368 | 0.43837 | 0.42385 | 0.97468 | 0.92642 | 0.5472754793 | 0.97917 | 0.9848355651 | 0.424869 | 6.530176209 | 1 |
| 17 | 43045751 | T | A | D | V | BRCA1 | ENST00000471181 | 0.81292 | 0.51286 | 0.37754 | 0.98992 | 0.95023 | 0.783753914 | 0.96653 | 0.9957713485 | 0.47486 | 6.530176209 | 1 |
| 17 | 43045751 | T | C | D | G | BRCA1 | ENST00000471181 | 0.82981 | 0.49709 | 0.39751 | 0.98816 | 0.94356 | 0.7467861642 | 0.97967 | 0.9952281713 | 0.47138 | 6.530176209 | 1 |
| 17 | 43045751 | T | G | D | A | BRCA1 | ENST00000471181 | 0.82287 | 0.48774 | 0.38959 | 0.98787 | 0.94876 | 0.7427366676 | 0.97513 | 0.9957713485 | 0.458895 | 6.530176209 | 0 |
| 17 | 43045752 | C | G | D | H | BRCA1 | ENST00000471181 | 0.78725 | 0.47983 | 0.4819 | 0.98411 | 0.93475 | 0.7444547005 | 0.98452 | 0.9939545989 | 0.471148 | 6.530176209 | 0 |
| 17 | 43045752 | C | T | D | N | BRCA1 | ENST00000471181 | 0.79469 | 0.44853 | 0.45189 | 0.98461 | 0.89055 | 0.6634907953 | 0.98425 | 0.9842811823 | 0.194588 | 6.530176209 | 0 |
| 17 | 43045753 | C | A | L | F | BRCA1 | ENST00000471181 | 0.57491 | 0.46518 | 0.3068 | 0.97871 | 0.83943 | 0.4629200162 | 0.94947 | 0.9892064333 | 0.337786 | 6.53017639 | 0 |
| 17 | 43045753 | C | G | L | F | BRCA1 | ENST00000471181 | 0.5824 | 0.46518 | 0.3068 | 0.97888 | 0.83943 | 0.4629200162 | 0.94947 | 0.9867265224 | 0.33779 | 6.53017639 | 0 |

| | | | | | | | | | | | | | | | | | |
|---|---|---|---|---|---|---|---|---|---|---|---|---|---|---|---|---|---|
| 17 | 43045754 | A | G | L | S | BRCA1 | ENST00000471181 | 0.8437 | 0.50737 | 0.33545 | 0.98377 | 0.90755 | 0.6721993358 | 0.94558 | 0.9917297959 | 0.447566 | 6.53017639 | 1 |
| 17 | 43045755 | A | C | L | V | BRCA1 | ENST00000471181 | 0.41878 | 0.45205 | 0.26271 | 0.95461 | 0.84588 | 0.4463231242 | 0.86844 | 0.9573149085 | 0.32001 | 6.53017639 | 1 |
| 17 | 43045757 | A | C | V | G | BRCA1 | ENST00000471181 | 0.86403 | 0.48504 | 0.38303 | 0.98367 | 0.93358 | 0.7466409899 | 0.94114 | 0.9909467697 | 0.460779 | 6.52815715 | 1 |
| 17 | 43045757 | A | G | V | A | BRCA1 | ENST00000471181 | 0.83406 | 0.43665 | 0.49701 | 0.97691 | 0.91736 | 0.5569963424 | 0.93979 | 0.984226644 | 0.375653 | 6.52815715 | 0 |
| 17 | 43045757 | A | T | V | E | BRCA1 | ENST00000471181 | 0.86955 | 0.23763 | 0.47967 | 0.98085 | 0.92924 | 0.8105425284 | 0.91142 | 0.9482374191 | 0.460108 | 6.52815715 | 1 |
| 17 | 43045758 | C | A | V | L | BRCA1 | ENST00000471181 | 0.71281 | 0.29206 | 0.47864 | 0.93491 | 0.84052 | 0.397943263 | 0.92847 | 0.9880070686 | 0.267193 | 6.52815715 | 0 |
| 17 | 43045758 | C | G | V | L | BRCA1 | ENST00000471181 | 0.72132 | 0.29206 | 0.47864 | 0.93108 | 0.84267 | 0.397943263 | 0.92847 | 0.9880070686 | 0.267195 | 6.52815715 | 0 |
| 17 | 43045758 | C | T | V | M | BRCA1 | ENST00000471181 | 0.74978 | 0.44471 | 0.5059 | 0.96835 | 0.85792 | 0.4134853626 | 0.97274 | 0.9942343831 | 0.327263 | 6.52815715 | 0 |
| 17 | 43045759 | C | A | W | C | BRCA1 | ENST00000471181 | 0.83221 | 0.52734 | 0.52142 | 0.99511 | 0.95863 | 0.8506528145 | 0.98036 | 0.9987760186 | 0.533264 | 6.530175352 | 1 |
| 17 | 43045759 | C | G | W | C | BRCA1 | ENST00000471181 | 0.84035 | 0.52734 | 0.52142 | 0.99511 | 0.95863 | 0.8506528145 | 0.98036 | 0.9956784844 | 0.533265 | 6.530175352 | 1 |
| 17 | 43045760 | C | A | W | L | BRCA1 | ENST00000471181 | 0.78399 | 0.5373 | 0.5339 | 0.99597 | 0.95753 | 0.7400357408 | 0.98615 | 0.9962674975 | 0.517972 | 6.530175352 | 1 |
| 17 | 43045760 | C | G | W | S | BRCA1 | ENST00000471181 | 0.82163 | 0.52151 | 0.50575 | 0.99543 | 0.95572 | 0.8414423473 | 0.98977 | 0.994707942 | 0.529147 | 6.530175352 | 1 |
| 17 | 43045761 | A | C | W | G | BRCA1 | ENST00000471181 | 0.90095 | 0.5373 | 0.47573 | 0.99778 | 0.9681 | 0.8001986637 | 0.9775 | 0.989980638 | 0.536607 | 6.530175352 | 1 |
| 17 | 43045761 | A | G | W | R | BRCA1 | ENST00000471181 | 0.87362 | 0.53294 | 0.58764 | 0.99547 | 0.97252 | 0.8665895507 | 0.99518 | 0.9966059923 | 0.536605 | 6.530175352 | 1 |
| 17 | 43045761 | A | T | W | R | BRCA1 | ENST00000471181 | 0.86204 | 0.53294 | 0.58764 | 0.99564 | 0.97252 | 0.8665895507 | 0.99518 | 0.9966059923 | 0.536605 | 6.530175352 | 1 |
| 17 | 43045762 | C | A | E | D | BRCA1 | ENST00000471181 | 0.26851 | 0.29702 | 0.26425 | 0.75571 | 0.74936 | 0.343608834 | 0.83632 | 0.8204373121 | 0.145378 | 3.150678316 | 0 |
| 17 | 43045762 | C | G | E | D | BRCA1 | ENST00000471181 | 0.2735 | 0.29702 | 0.26425 | 0.72323 | 0.74936 | 0.343608834 | 0.83632 | 0.8134340048 | 0.145374 | 3.150678316 | 0 |
| 17 | 43045763 | T | A | E | V | BRCA1 | ENST00000471181 | 0.60794 | 0.39738 | 0.24175 | 0.97212 | 0.8887 | 0.4933782256 | 0.92388 | 0.9803704023 | 0.368583 | 6.256227711 | 1 |
| 17 | 43045763 | T | C | E | G | BRCA1 | ENST00000471181 | 0.83097 | 0.45545 | 0.23596 | 0.95664 | 0.89918 | 0.4191693846 | 0.92348 | 0.9658320546 | 0.38133 | 6.256227711 | 0 |
| 17 | 43045763 | T | G | E | A | BRCA1 | ENST00000471181 | 0.77077 | 0.35663 | 0.26309 | 0.96213 | 0.88165 | 0.4292566949 | 0.93545 | 0.9409117103 | 0.314655 | 6.256227711 | 0 |
| 17 | 43045764 | C | G | E | Q | BRCA1 | ENST00000471181 | 0.70262 | 0.4182 | 0.25524 | 0.92573 | 0.80476 | 0.368562513 | 0.94492 | 0.9411853552 | 0.246596 | 6.256227711 | 0 |
| 17 | 43045764 | C | T | E | K | BRCA1 | ENST00000471181 | 0.77251 | 0.43605 | 0.3548 | 0.96284 | 0.83116 | 0.4505624132 | 0.93501 | 0.8495799303 | 0.267513 | 6.256227711 | 0 |
| 17 | 43045767 | G | C | R | G | BRCA1 | ENST00000471181 | 0.48202 | 0.48932 | 0.15835 | 0.94892 | 0.78035 | 0.5831315762 | 0.91784 | 0.9585673809 | 0.343351 | 6.256228434 | 1 |
| 17 | 43045769 | G | A | T | I | BRCA1 | ENST00000471181 | 0.62597 | 0.44238 | 0.28081 | 0.94257 | 0.84213 | 0.6220639569 | 0.927 | 0.7215113833 | 0.119374 | 5.423925471 | 0 |
| 17 | 43045769 | G | C | T | S | BRCA1 | ENST00000471181 | 0.41469 | 0.10288 | 0.1642 | 0.76878 | 0.7562 | 0.339696125 | 0.89212 | 0.5306283236 | 0.161627 | 3.932252053 | 0 |
| 17 | 43045769 | G | T | T | N | BRCA1 | ENST00000471181 | 0.58275 | 0.41139 | 0.22509 | 0.9356 | 0.81298 | 0.5614226971 | 0.93237 | 0.8001120687 | 0.160531 | 5.979522417 | 0 |
| 17 | 43045770 | T | A | T | S | BRCA1 | ENST00000471181 | 0.44704 | 0.10288 | 0.1642 | 0.84141 | 0.78411 | 0.339696125 | 0.9141 | 0.7497035265 | 0.173143 | 3.932252053 | 0 |
| 17 | 43045770 | T | C | T | A | BRCA1 | ENST00000471181 | 0.49847 | 0.17468 | 0.14503 | 0.87142 | 0.80713 | 0.5203952463 | 0.94179 | 0.8908872008 | 0.234092 | 5.979522417 | 0 |
| 17 | 43045770 | T | G | T | P | BRCA1 | ENST00000471181 | 0.70992 | 0.47123 | 0.16516 | 0.95698 | 0.87539 | 0.6986086408 | 0.96639 | 0.9446070194 | 0.315588 | 5.979522417 | 0 |
| 17 | 43045772 | A | G | V | A | BRCA1 | ENST00000471181 | 0.72284 | 0.44799 | 0.42354 | 0.98912 | 0.93044 | 0.6321492566 | 0.99276 | 0.984226644 | 0.455658 | 6.530175437 | 0 |
| 17 | 43045772 | A | T | V | E | BRCA1 | ENST00000471181 | 0.76658 | 0.52478 | 0.40869 | 0.99224 | 0.94913 | 0.8946380494 | 0.99575 | 0.9788354635 | 0.50902 | 6.530175437 | 1 |
| 17 | 43045773 | C | T | V | M | BRCA1 | ENST00000471181 | 0.6934 | 0.43583 | 0.43556 | 0.98392 | 0.91526 | 0.6808606142 | 0.99493 | 0.9627158046 | 0.500987 | 6.530175437 | 1 |
| 17 | 43045775 | A | C | V | G | BRCA1 | ENST00000471181 | 0.76658 | 0.45073 | 0.13037 | 0.97818 | 0.90274 | 0.826329298 | 0.97567 | 0.9724556804 | 0.345508 | 6.17177218 | 1 |
| 17 | 43045776 | C | G | V | L | BRCA1 | ENST00000471181 | 0.19888 | 0.13585 | 0.19021 | 0.82844 | 0.77399 | 0.4812556666 | 0.8879 | 0.6808077693 | 0.170472 | 3.018976197 | 0 |
| 17 | 43045778 | G | A | P | L | BRCA1 | ENST00000471181 | 0.34533 | 0.25318 | 0.13468 | 0.91178 | 0.82613 | 0.3420317476 | 0.84548 | 0.677098453 | 0.194758 | 5.274704149 | 0 |
| 17 | 43045778 | G | C | P | R | BRCA1 | ENST00000471181 | 0.16419 | 0.11325 | 0.13075 | 0.79308 | 0.79637 | 0.3111113272 | 0.82078 | 0.6048014164 | 0.195697 | 4.175599526 | 0 |
| 17 | 43045778 | G | T | P | H | BRCA1 | ENST00000471181 | 0.15842 | 0.42607 | 0.11664 | 0.89999 | 0.8517 | 0.3436739985 | 0.85005 | 0.753020227 | 0.201193 | 3.178677714 | 0 |
| 17 | 43045779 | G | A | P | S | BRCA1 | ENST00000471181 | 0.15297 | 0.10581 | 0.08157 | 0.74607 | 0.79637 | 0.2892902968 | 0.71439 | 0.1895111054 | 0.144762 | 3.198172087 | 0 |
| 17 | 43045779 | G | C | P | A | BRCA1 | ENST00000471181 | 0.19937 | 0.10875 | 0.08584 | 0.71543 | 0.77782 | 0.2305320194 | 0.7353 | 0.1522688191 | 0.142892 | 3.49643701 | 0 |
| 17 | 43045779 | G | T | P | T | BRCA1 | ENST00000471181 | 0.15416 | 0.18659 | 0.09697 | 0.803 | 0.80476 | 0.320389479 | 0.79583 | 0.3320328593 | 0.212012 | 3.597039313 | 0 |
| 17 | 43045781 | G | A | A | V | BRCA1 | ENST00000471181 | 0.20264 | 0.17138 | 0.12225 | 0.75577 | 0.77718 | 0.3766252747 | 0.6754 | 0.359401226 | 0.157696 | 1.969151936 | 0 |
| 17 | 43045782 | C | A | A | S | BRCA1 | ENST00000471181 | 0.36598 | 0.25632 | 0.09649 | 0.91396 | 0.78411 | 0.4870678281 | 0.79724 | 0.904476285 | 0.18463 | 6.055465026 | 0 |
| 17 | 43045782 | C | G | A | P | BRCA1 | ENST00000471181 | 0.41985 | 0.48627 | 0.16231 | 0.9357 | 0.79028 | 0.7273499052 | 0.88278 | 0.904476285 | 0.271313 | 6.055465026 | 0 |
| 17 | 43045782 | C | T | A | T | BRCA1 | ENST00000471181 | 0.40027 | 0.38103 | 0.11413 | 0.91682 | 0.77972 | 0.4462947971 | 0.85174 | 0.7612768412 | 0.203926 | 6.055465026 | 0 |
| 17 | 43045783 | C | A | E | D | BRCA1 | ENST00000471181 | 0.10092 | 0.08253 | 0.1282 | 0.7677 | 0.78473 | 0.1873754289 | 0.45498 | 0.5786334872 | 0.115094 | 2.871292325 | 0 |
| 17 | 43045783 | C | G | E | D | BRCA1 | ENST00000471181 | 0.10412 | 0.08253 | 0.1282 | 0.76464 | 0.78473 | 0.1873754289 | 0.45498 | 0.5786334872 | 0.115105 | 2.871292325 | 0 |
| 17 | 43045784 | T | A | E | V | BRCA1 | ENST00000471181 | 0.39284 | 0.14714 | 0.10661 | | 0.78658 | 0.3868396228 | 0.85096 | 0.920634985 | 0.232733 | 5.4190138 | 0 |
| 17 | 43045784 | T | C | E | G | BRCA1 | ENST00000471181 | 0.30417 | 0.16976 | 0.10113 | | 0.68646 | 0.2377707224 | 0.84371 | 0.8346920013 | 0.147198 | 2.909316818 | 0 |
| 17 | 43045784 | T | G | E | A | BRCA1 | ENST00000471181 | 0.26758 | 0.10002 | 0.12808 | | 0.74093 | 0.2685236594 | 0.85805 | 0.4860637188 | 0.143264 | 2.640050166 | 0 |
| 17 | 43045785 | C | G | E | Q | BRCA1 | ENST00000471181 | 0.22881 | 0.14139 | 0.11371 | | 0.7528 | 0.1744673013 | 0.85131 | 0.5000983477 | 0.143154 | 3.055429156 | 0 |
| 17 | 43045786 | A | C | C | W | BRCA1 | ENST00000471181 | 0.79223 | 0.49712 | 0.47239 | | 0.91569 | 0.5382533035 | 0.94585 | 0.9781780243 | 0.346891 | 5.942181497 | 0 |
| 17 | 43045787 | C | A | C | F | BRCA1 | ENST00000471181 | 0.68859 | 0.50678 | 0.42012 | 0.96389 | 0.81933 | 0.4373616235 | 0.98502 | 0.9844745398 | 0.389047 | 5.942181497 | 0 |
| 17 | 43045787 | C | G | C | S | BRCA1 | ENST00000471181 | 0.64314 | 0.46119 | 0.40184 | 0.93009 | 0.79998 | 0.3551678823 | 0.96987 | 0.9780826569 | 0.371827 | 3.463075379 | 0 |
| 17 | 43045787 | C | T | C | Y | BRCA1 | ENST00000471181 | 0.67072 | 0.53867 | 0.5307 | 0.95402 | 0.82159 | 0.4671813802 | 0.97031 | 0.9786148071 | 0.362324 | 4.107619445 | 0 |
| 17 | 43045788 | A | C | C | G | BRCA1 | ENST00000471181 | 0.53529 | 0.48047 | 0.37232 | 0.94027 | 0.89009 | 0.3831907806 | 0.96862 | 0.9767137766 | 0.385293 | 3.319022138 | 0 |
| 17 | 43045788 | A | T | C | S | BRCA1 | ENST00000471181 | 0.65791 | 0.46119 | 0.40184 | 0.96241 | 0.83997 | 0.3551678823 | 0.97109 | 0.9808604717 | 0.358242 | 3.463075379 | 0 |
| 17 | 43045789 | C | A | M | I | BRCA1 | ENST00000471181 | 0.45998 | 0.12028 | 0.17825 | 0.93746 | 0.76157 | 0.2455296399 | 0.92022 | 0.9208710194 | 0.184546 | 3.602287935 | 0 |
| 17 | 43045789 | C | G | M | I | BRCA1 | ENST00000471181 | 0.46655 | 0.12028 | 0.17825 | 0.93388 | 0.76157 | 0.2455296399 | 0.92022 | 0.9208710194 | 0.184554 | 3.602287935 | 0 |
| 17 | 43045789 | C | T | M | I | BRCA1 | ENST00000471181 | 0.47087 | 0.12028 | 0.17825 | 0.94433 | 0.7562 | 0.2455296399 | 0.93696 | 0.6405775547 | 0.0197625 | 3.602287935 | 0 |
| 17 | 43045790 | A | C | M | R | BRCA1 | ENST00000471181 | 0.59212 | | | 0.94047 | 0.65077 | 0.4344600941 | 0.58208 | 0.9198993444 | 0.0377334 | 5.371487045 | 0 |
| 17 | 43045790 | A | G | M | T | BRCA1 | ENST00000471181 | 0.53791 | 0.26437 | 0.26943 | 0.91287 | 0.80238 | 0.2953467014 | 0.96601 | 0.9670513868 | 0.251249 | 3.566771984 | 0 |
| 17 | 43045790 | A | T | M | K | BRCA1 | ENST00000471181 | 0.43424 | | | 0.92958 | 0.65277 | 0.4118397102 | 0.53657 | 0.9321523905 | 0.0376506 | 2.871213417 | 0 |
| 17 | 43045791 | T | A | M | L | BRCA1 | ENST00000471181 | 0.23044 | | | 0.71225 | 0.60952 | 0.2536716814 | 0.15292 | 0.02074185781 | -0.0336574 | 1.256559666 | 0 |
| 17 | 43045791 | T | C | M | V | BRCA1 | ENST00000471181 | 0.29829 | | | 0.79453 | 0.60717 | 0.2905370161 | 0.17906 | 0.03442526961 | -0.165098 | 2.290548772 | 0 |
| 17 | 43045791 | T | G | M | L | BRCA1 | ENST00000471181 | 0.23478 | | | 0.6404 | 0.60952 | 0.2536716814 | 0.152 | 0.01930561898 | -0.0575019 | 1.256559666 | 0 |
| 17 | 43045792 | C | A | Q | H | BRCA1 | ENST00000471181 | 0.62704 | | | 0.93218 | 0.59021 | 0.2955336982 | 0.64539 | 0.9585067034 | 0.0243252 | 6.260135595 | 0 |
| 17 | 43045792 | C | G | Q | H | BRCA1 | ENST00000471181 | 0.63512 | | | 0.94226 | 0.56561 | 0.2955336982 | 0.67553 | 0.9724463224 | 0.0231661 | 6.260135595 | 0 |
| 17 | 43045793 | T | A | Q | L | BRCA1 | ENST00000471181 | 0.68139 | 0.22521 | 0.20341 | 0.83698 | 0.7816 | 0.3931017666 | 0.84604 | 0.65452981 | 0.177022 | 3.929073725 | 0 |
| 17 | 43045793 | T | C | Q | R | BRCA1 | ENST00000471181 | 0.76784 | 0.39976 | 0.24416 | 0.88015 | 0.80118 | 0.2357979941 | 0.87407 | 0.9478456974 | 0.226987 | 4.061930865 | 0 |
| 17 | 43045793 | T | G | Q | P | BRCA1 | ENST00000471181 | 0.80258 | 0.45175 | 0.23044 | 0.93072 | 0.8543 | 0.6726567821 | 0.90645 | 0.9531556964 | 0.349867 | 6.260135595 | 0 |
| 17 | 43045794 | G | C | Q | E | BRCA1 | ENST00000471181 | 0.62165 | | | 0.93985 | 0.66639 | 0.2374785974 | 0.81337 | 0.9513159394 | 0.0589854 | 6.260135595 | 0 |
| 17 | 43045794 | G | T | Q | K | BRCA1 | ENST00000471181 | 0.50505 | | | 0.93968 | 0.58896 | 0.3102496185 | 0.71707 | 0.5927984853 | 0.00959493 | 3.573184485 | 0 |
| 17 | 43045796 | C | A | G | V | BRCA1 | ENST00000471181 | 0.37566 | 0.48461 | 0.14965 | 0.94209 | 0.80536 | 0.4288645923 | 0.79356 | 0.961902678 | 0.279003 | 5.854049685 | 0 |
| 17 | 43045796 | C | G | G | A | BRCA1 | ENST00000471181 | 0.32712 | 0.38073 | 0.14388 | 0.92697 | 0.72701 | 0.3089986301 | 0.8406 | 0.9162338376 | 0.161549 | 5.854049685 | 0 |
| 17 | 43045796 | C | T | G | E | BRCA1 | ENST00000471181 | 0.18104 | 0.25976 | 0.19268 | | 0.81933 | 0.5696340896 | 0.81806 | 0.9081710577 | 0.229591 | 3.029218296 | 0 |
| 17 | 43045797 | C | A | G | W | BRCA1 | ENST00000471181 | 0.53636 | 0.47845 | 0.1925 | 0.9521 | 0.81298 | 0.6152809788 | 0.46967 | 0.9904873967 | 0.263009 | 4.104621483 | 0 |
| 17 | 43045797 | C | G | G | R | BRCA1 | ENST00000471181 | 0.48927 | 0.47231 | 0.20413 | 0.90967 | 0.79998 | 0.6024563036 | 0.54898 | 0.9782237411 | 0.233628 | 4.429892157 | 0 |
| 17 | 43045797 | C | T | G | R | BRCA1 | ENST00000471181 | 0.4937 | 0.47231 | 0.20413 | | 0.81472 | 0.6024563036 | 0.65613 | 0.9613280296 | 0.236956 | 4.429892157 | 0 |
| 17 | 43045798 | A | C | I | M | BRCA1 | ENST00000471181 | 0.45487 | 0.44433 | 0.22574 | 0.90445 | 0.81006 | 0.3352009427 | 0.71755 | 0.9813063741 | 0.122237 | 6.06381981 | 1 |
| 17 | 43045799 | A | C | I | S | BRCA1 | ENST00000471181 | 0.66335 | 0.51153 | 0.2703 | 0.9566 | 0.82556 | 0.5986559112 | 0.82995 | 0.9953398705 | 0.343527 | 6.06381981 | 0 |
| 17 | 43045799 | A | G | I | T | BRCA1 | ENST00000471181 | 0.59791 | 0.48662 | 0.34928 | 0.94786 | 0.82046 | 0.3419797757 | 0.92466 | 0.9947854877 | 0.272687 | 5.613288335 | 0 |
| 17 | 43045799 | A | T | I | N | BRCA1 | ENST00000471181 | 0.65586 | 0.55114 | 0.3246 | 0.95499 | 0.81876 | 0.5753215957 | 0.77439 | 0.9957330823 | 0.253014 | 6.06381981 | 0 |
| 17 | 43045800 | T | A | I | F | BRCA1 | ENST00000471181 | 0.605 | 0.51644 | 0.26316 | 0.92948 | 0.83283 | 0.4482676631 | 0.91958 | 0.9400733113 | 0.24798 | 6.06381981 | 0 |
| 17 | 43045800 | T | C | I | V | BRCA1 | ENST00000471181 | 0.50393 | 0.16629 | 0.1857 | 0.92068 | 0.80238 | 0.1881692878 | 0.72309 | 0.9653446078 | 0.203002 | 4.773654329 | 0 |
| 17 | 43045800 | T | G | I | L | BRCA1 | ENST00000471181 | 0.44423 | 0.31353 | 0.25609 | 0.91605 | 0.74936 | 0.3541835563 | 0.75954 | 0.9647496343 | 0.139446 | 3.063336067 | 0 |
| 17 | 43045802 | G | A | A | V | BRCA1 | ENST00000471181 | 0.21521 | 0.09155 | 0.07575 | 0.55373 | 0.75417 | 0.2860013055 | 0.48522 | 0.9620807171 | 0.173484 | 1.866242609 | 0 |
| 17 | 43045802 | G | T | A | E | BRCA1 | ENST00000471181 | 0.21669 | 0.24623 | 0.09465 | 0.83481 | 0.76816 | 0.4345280817 | 0.7762 | 0.9924736023 | 0.157981 | 2.673614249 | 0 |
| 17 | 43047643 | C | A | A | S | BRCA1 | ENST00000471181 | 0.39231 | 0.11996 | 0.06116 | 0.95379 | 0.71627 | 0.2349657967 | 0.87835 | 0.9874442816 | 0.0935976 | 5.464316923 | 1 |
| 17 | 43047643 | C | G | A | P | BRCA1 | ENST00000471181 | 0.69444 | 0.30079 | 0.11154 | 0.98089 | 0.78411 | 0.6747215146 | 0.88486 | 0.8092108965 | 0.186482 | 5.464316923 | 1 |
| 17 | 43047643 | C | T | A | T | BRCA1 | ENST00000471181 | 0.41463 | 0.09039 | 0.07342 | 0.96741 | 0.77654 | 0.2286914656 | 0.8814 | 0.6804724336 | 0.247613 | 4.598345435 | 1 |
| 17 | 43047644 | A | C | H | Q | BRCA1 | ENST00000471181 | 0.1202 | | | 0.88715 | 0.59881 | 0.1902808556 | 0.09354 | 0.2655356228 | -0.0493435 | 2.558458538 | 0 |
| 17 | 43047644 | A | T | H | Q | BRCA1 | ENST00000471181 | 0.12263 | | | 0.891 | 0.61416 | 0.1902808556 | 0.09552 | 0.2655356228 | -0.0462797 | 2.558458538 | 0 |
| 17 | 43047645 | T | A | H | L | BRCA1 | ENST00000471181 | 0.14906 | 0.15268 | 0.09507 | 0.88652 | 0.70994 | 0.4413074518 | 0.53079 | 0.1831178167 | 0.0566156 | 2.738256308 | 0 |
| 17 | 43047645 | T | C | H | R | BRCA1 | ENST00000471181 | 0.08614 | 0.18538 | 0.11591 | 0.83468 | 0.81182 | 0.3149960947 | 0.75739 | 0.1025550073 | 0.138616 | 2.394526317 | 0 |
| 17 | 43047645 | T | G | H | P | BRCA1 | ENST00000471181 | 0.14045 | 0.15943 | 0.09338 | 0.83228 | 0.78535 | 0.6914424699 | 0.52585 | 0.09003805629 | 0.173812 | 2.518964477 | 0 |
| 17 | 43047646 | G | A | H | Y | BRCA1 | ENST00000471181 | 0.33548 | 0.42117 | 0.12446 | 0.96993 | 0.6993 | 0.3109826919 | 0.82699 | 0.8822077513 | 0.176283 | 5.612270674 | 0 |
| 17 | 43047646 | G | C | H | D | BRCA1 | ENST00000471181 | 0.4836 | 0.49752 | 0.11768 | 0.9622 | 0.75755 | 0.5114274247 | 0.88959 | 0.8536531329 | 0.183351 | 5.612270674 | 0 |
| 17 | 43047646 | G | T | H | N | BRCA1 | ENST00000471181 | 0.24585 | 0.37297 | 0.118 | 0.97327 | 0.6757 | 0.3026714044 | 0.92142 | 0.8607518077 | 0.087417 | 5.612270674 | 0 |
| 17 | 43047647 | G | C | F | L | BRCA1 | ENST00000471181 | 0.18654 | 0.10736 | 0.29852 | 0.92293 | 0.76685 | 0.3815198722 | 0.80893 | 0.7518070936 | 0.0949008 | 2.928558179 | 0 |
| 17 | 43047647 | G | T | F | L | BRCA1 | ENST00000471181 | 0.18357 | 0.10736 | 0.29852 | 0.91416 | 0.7675 | 0.3815198722 | 0.80907 | 0.7518070936 | 0.0948987 | 2.928558179 | 0 |
| 17 | 43047648 | A | C | F | C | BRCA1 | ENST00000471181 | 0.40816 | 0.45319 | 0.29342 | 0.92454 | 0.81006 | 0.5410389221 | 0.78159 | 0.9319531322 | 0.187809 | 2.810141592 | 0 |

| | | | | | | | | | | | | | | | | | |
|---|---|---|---|---|---|---|---|---|---|---|---|---|---|---|---|---|---|
| 17 | 43047648 | A | G | F | S | BRCA1 | ENST00000471181 | 0.73005 | 0.41463 | 0.28103 | 0.96634 | 0.83779 | 0.6039610508 | 0.89177 | 0.9589788318 | 0.199729 | 3.710028014 | 0 |
| 17 | 43047648 | A | T | F | Y | BRCA1 | ENST00000471181 | 0.34347 | 0.21646 | 0.30119 | 0.85286 | 0.74235 | 0.3904019415 | 0.84007 | 0.8601202369 | 0.10519 | 3.060050192 | 0 |
| 17 | 43047649 | A | C | F | V | BRCA1 | ENST00000471181 | 0.70671 | 0.1247 | 0.24349 | 0.97595 | 0.83779 | 0.4527430289 | 0.94148 | 0.9505161047 | 0.225745 | 5.774065688 | 0 |
| 17 | 43047649 | A | G | F | L | BRCA1 | ENST00000471181 | 0.43956 | 0.10736 | 0.29852 | 0.97648 | 0.76881 | 0.3815198722 | 0.96603 | 0.9115201831 | 0.141599 | 2.928558179 | 0 |
| 17 | 43047649 | A | T | F | I | BRCA1 | ENST00000471181 | 0.56179 | 0.14379 | 0.28667 | 0.98029 | 0.81646 | 0.4594251067 | 0.976 | 0.9596922994 | 0.147704 | 5.774065688 | 0 |
| 17 | 43047651 | C | A | G | V | BRCA1 | ENST00000471181 | 0.32552 | 0.46743 | 0.1209 | 0.97558 | 0.84213 | 0.3869750884 | 0.86443 | 0.9813201427 | 0.228154 | 6.218370177 | 0 |
| 17 | 43047651 | C | T | G | D | BRCA1 | ENST00000471181 | 0.21454 | 0.14323 | 0.18646 | | 0.72626 | 0.4159055806 | 0.86524 | 0.8512828946 | 0.149873 | 3.332139725 | 0 |
| 17 | 43047652 | C | A | G | C | BRCA1 | ENST00000471181 | 0.40506 | 0.44449 | 0.14692 | 0.95211 | 0.80831 | 0.4255268791 | 0.66797 | 0.9156537652 | 0.222728 | 6.218370177 | 0 |
| 17 | 43047652 | C | G | G | R | BRCA1 | ENST00000471181 | 0.38484 | 0.42776 | 0.17543 | 0.94997 | 0.79334 | 0.4459056139 | 0.66736 | 0.8477653265 | 0.188771 | 6.218370177 | 0 |
| 17 | 43047652 | C | T | G | S | BRCA1 | ENST00000471181 | 0.25539 | 0.36897 | 0.11296 | | 0.74376 | 0.3152916073 | 0.82035 | 0.8188928962 | 0.0947574 | 3.182597257 | 0 |
| 17 | 43047653 | A | C | N | K | BRCA1 | ENST00000471181 | 0.27545 | 0.23511 | 0.05461 | 0.94978 | 0.75074 | 0.1847551729 | 0.56169 | 0.5247277021 | 0.0635634 | 4.13471359 | 0 |
| 17 | 43047653 | A | T | N | K | BRCA1 | ENST00000471181 | 0.27147 | 0.23511 | 0.05461 | 0.91977 | 0.74797 | 0.1847551729 | 0.51342 | 0.5247277021 | 0.0635565 | 4.13471359 | 0 |
| 17 | 43047654 | T | A | N | I | BRCA1 | ENST00000471181 | 0.28524 | 0.33351 | 0.05268 | 0.96656 | 0.80178 | 0.3175012937 | 0.67293 | 0.6707013845 | 0.083806 | 4.13471359 | 0 |
| 17 | 43047654 | T | C | N | S | BRCA1 | ENST00000471181 | 0.03715 | 0.07277 | 0.02653 | 0.63685 | 0.73296 | 0.131326699 | 0.35965 | 0.004140317906 | -0.20054 | -0.5504123676 | 0 |
| 17 | 43047654 | T | G | N | T | BRCA1 | ENST00000471181 | 0.09035 | 0.10002 | 0.0353 | 0.83754 | 0.76946 | 0.2054968571 | 0.7217 | 0.03430571251 | 0.0833103 | 1.909200083 | 0 |
| 17 | 43047655 | T | A | N | Y | BRCA1 | ENST00000471181 | 0.31421 | 0.29946 | 0.05645 | 0.95044 | 0.80889 | 0.3055516576 | 0.54854 | 0.6662432551 | 0.150698 | 4.13471359 | 0 |
| 17 | 43047655 | T | C | N | D | BRCA1 | ENST00000471181 | 0.10833 | 0.10371 | 0.05729 | 0.85049 | 0.59393 | 0.20416641 | 0.61737 | 0.1505633891 | 0.0313823 | 1.539248011 | 0 |
| 17 | 43047655 | T | G | N | H | BRCA1 | ENST00000471181 | 0.1996 | 0.09429 | 0.03726 | 0.93952 | 0.75349 | 0.2350807436 | 0.79237 | 0.5443611145 | 0.0644963 | 2.398285801 | 0 |
| 17 | 43047656 | G | C | D | E | BRCA1 | ENST00000471181 | 0.25448 | 0.0852 | 0.22794 | 0.9404 | 0.83228 | 0.1902758567 | 0.83365 | 0.9633061886 | 0.167533 | 3.503049034 | 0 |
| 17 | 43047656 | G | T | D | E | BRCA1 | ENST00000471181 | 0.2508 | 0.0852 | 0.22794 | 0.96433 | 0.83283 | 0.1902758567 | 0.83698 | 0.9633061886 | 0.167507 | 3.503049034 | 0 |
| 17 | 43047657 | T | A | D | V | BRCA1 | ENST00000471181 | 0.72404 | 0.17899 | 0.20775 | 0.9783 | 0.91862 | 0.4773759531 | 0.85616 | 0.9852287769 | 0.249558 | 6.187671839 | 0 |
| 17 | 43047657 | T | C | D | G | BRCA1 | ENST00000471181 | 0.54984 | 0.12824 | 0.22151 | 0.97409 | 0.85586 | 0.4242137283 | 0.85909 | 0.9801591635 | 0.229686 | 4.282395436 | 1 |
| 17 | 43047657 | T | G | D | A | BRCA1 | ENST00000471181 | 0.56964 | 0.14408 | 0.21639 | 0.97807 | 0.87442 | 0.3822625991 | 0.87374 | 0.9808026552 | 0.201804 | 6.187671839 | 0 |
| 17 | 43047658 | C | A | D | Y | BRCA1 | ENST00000471181 | 0.65688 | 0.40026 | 0.2534 | 0.96933 | 0.86603 | 0.4938828789 | 0.85457 | 0.9917325974 | 0.244972 | 5.793412474 | 0 |
| 17 | 43047658 | C | G | D | H | BRCA1 | ENST00000471181 | 0.63527 | 0.2927 | 0.2625 | 0.96597 | 0.85326 | 0.4290158808 | 0.89689 | 0.9851549268 | 0.222241 | 6.187671839 | 1 |
| 17 | 43047658 | C | T | D | N | BRCA1 | ENST00000471181 | 0.36013 | 0.08989 | 0.24673 | 0.9548 | 0.672 | 0.2555578725 | 0.89663 | 0.9818469882 | 0.0724514 | 3.265306691 | 0 |
| 17 | 43047659 | C | A | E | D | BRCA1 | ENST00000471181 | 0.32336 | 0.08227 | 0.17608 | | 0.79818 | 0.3373843264 | 0.91168 | 0.9293009043 | 0.172716 | 2.674217419 | 0 |
| 17 | 43047659 | C | G | E | D | BRCA1 | ENST00000471181 | 0.32897 | 0.08227 | 0.17608 | 0.93077 | 0.80178 | 0.3373843264 | 0.8396 | 0.8831019998 | 0.166781 | 2.674217419 | 0 |
| 17 | 43047660 | T | A | E | V | BRCA1 | ENST00000471181 | 0.84831 | 0.12354 | 0.16053 | 0.9828 | 0.911 | 0.5052659885 | 0.92896 | 0.9814782143 | 0.253171 | 5.97254384 | 0 |
| 17 | 43047660 | T | C | E | G | BRCA1 | ENST00000471181 | 0.86768 | 0.15806 | 0.1523 | 0.97948 | 0.91058 | 0.4121945898 | 0.94068 | 0.9656748772 | 0.223715 | 4.601954388 | 0 |
| 17 | 43047660 | T | G | E | A | BRCA1 | ENST00000471181 | 0.81771 | 0.11348 | 0.17842 | 0.98253 | 0.91611 | 0.4165726357 | 0.93731 | 0.9680300951 | 0.207736 | 5.97254384 | 0 |
| 17 | 43047661 | C | G | E | Q | BRCA1 | ENST00000471181 | 0.67995 | 0.15322 | 0.16639 | 0.98801 | 0.88354 | 0.3851981292 | 0.94403 | 0.9581565857 | 0.206285 | 5.97254384 | 0 |
| 17 | 43047661 | C | T | E | K | BRCA1 | ENST00000471181 | 0.73285 | 0.12824 | 0.25057 | 0.97868 | 0.88354 | 0.3883727383 | 0.89809 | 0.967092216 | 0.235485 | 5.97254384 | 0 |
| 17 | 43047663 | G | A | T | I | BRCA1 | ENST00000471181 | 0.47388 | 0.12694 | 0.31844 | 0.94505 | 0.81065 | 0.2270670343 | 0.89179 | 0.9307233691 | 0.226417 | 5.120775573 | 0 |
| 17 | 43047663 | G | C | T | R | BRCA1 | ENST00000471181 | 0.61365 | | | 0.97606 | 0.77972 | 0.3285862328 | 0.60073 | 0.939232111 | 0.0858019 | 5.742008117 | 0 |
| 17 | 43047663 | G | T | T | K | BRCA1 | ENST00000471181 | 0.47797 | | | 0.96311 | 0.77909 | 0.2963079196 | 0.51975 | 0.917639792 | 0.0856383 | 4.399606886 | 0 |
| 17 | 43047664 | T | A | T | S | BRCA1 | ENST00000471181 | 0.30622 | | | 0.93629 | 0.56426 | 0.166279774 | 0.35192 | 0.8465998173 | 0.0242799 | 3.732993192 | 0 |
| 17 | 43047664 | T | C | T | A | BRCA1 | ENST00000471181 | 0.3251 | | | 0.91765 | 0.51233 | 0.195160842 | 0.3094 | 0.4246710241 | -0.00717998 | 2.480855509 | 0 |
| 17 | 43047664 | T | G | T | P | BRCA1 | ENST00000471181 | 0.35719 | | | 0.96559 | 0.66922 | 0.4869405424 | 0.55119 | 0.8027839065 | 0.0393994 | 3.544986809 | 0 |
| 17 | 43047665 | C | A | W | C | BRCA1 | ENST00000471181 | 0.8402 | | | 0.98577 | 0.81006 | 0.5563840271 | 0.88609 | 0.9887433052 | 0.109242 | 6.602799641 | 1 |
| 17 | 43047665 | C | G | W | C | BRCA1 | ENST00000471181 | 0.84824 | | | 0.98581 | 0.77654 | 0.5563840271 | 0.71672 | 0.9844692349 | 0.098815 | 6.602799641 | 1 |
| 17 | 43047666 | C | A | W | L | BRCA1 | ENST00000471181 | 0.804 | 0.5053 | 0.55695 | 0.98265 | 0.86653 | 0.4039380972 | 0.91631 | 0.9848945737 | 0.205271 | 6.602799641 | 0 |
| 17 | 43047666 | C | G | W | S | BRCA1 | ENST00000471181 | 0.84127 | 0.49369 | 0.53698 | 0.98304 | 0.9248 | 0.4532235141 | 0.90331 | 0.9756383896 | 0.27144 | 5.272673354 | 1 |
| 17 | 43047667 | A | C | W | G | BRCA1 | ENST00000471181 | 0.92151 | | | 0.97506 | 0.81414 | 0.3932777191 | 0.79697 | 0.9901124835 | 0.108598 | 6.602799641 | 0 |
| 17 | 43047667 | A | G | W | R | BRCA1 | ENST00000471181 | 0.90058 | | | 0.97461 | 0.8153 | 0.4980888558 | 0.84314 | 0.9952451587 | 0.11356 | 6.602799641 | 0 |
| 17 | 43047667 | A | T | W | R | BRCA1 | ENST00000471181 | 0.89113 | | | 0.97678 | 0.79456 | 0.4980888558 | 0.77513 | 0.9952451587 | 0.105668 | 6.602799641 | 0 |
| 17 | 43047669 | G | A | A | V | BRCA1 | ENST00000471181 | 0.86545 | 0.43706 | 0.32085 | 0.97859 | 0.91015 | 0.4504765095 | 0.82732 | 0.9881035686 | 0.266088 | 6.347099224 | 0 |
| 17 | 43047669 | G | C | A | G | BRCA1 | ENST00000471181 | 0.86502 | 0.11188 | 0.28998 | 0.95609 | 0.89694 | 0.3572595372 | 0.85257 | 0.9836771488 | 0.258039 | 6.347099224 | 1 |
| 17 | 43047670 | C | A | A | S | BRCA1 | ENST00000471181 | 0.58133 | 0.38779 | 0.27126 | 0.95119 | 0.83943 | 0.322243859 | 0.79465 | 0.9881035686 | 0.2356 | 4.588701747 | 0 |
| 17 | 43047670 | C | T | A | T | BRCA1 | ENST00000471181 | 0.62476 | 0.43001 | 0.31541 | 0.9663 | 0.85326 | 0.3689644069 | 0.83207 | 0.9881035686 | 0.248887 | 6.347099224 | 0 |
| 17 | 43047671 | A | C | D | E | BRCA1 | ENST00000471181 | 0.27189 | | | 0.9488 | 0.71233 | 0.26071193 | 0.37211 | 0.8400523067 | -0.0255944 | 6.267925912 | 0 |
| 17 | 43047671 | A | T | D | E | BRCA1 | ENST00000471181 | 0.2679 | | | 0.95322 | 0.67933 | 0.26071193 | 0.30959 | 0.8400523067 | -0.0402383 | 6.267925912 | 0 |
| 17 | 43047672 | T | A | D | V | BRCA1 | ENST00000471181 | 0.67207 | 0.28869 | 0.34377 | 0.96836 | 0.91652 | 0.5906610132 | 0.84129 | 0.9840788841 | 0.385665 | 6.267925912 | 0 |
| 17 | 43047672 | T | C | D | G | BRCA1 | ENST00000471181 | 0.68771 | 0.46653 | 0.35912 | 0.94861 | 0.9045 | 0.5003379136 | 0.89512 | 0.958663106 | 0.315809 | 6.267925912 | 0 |
| 17 | 43047672 | T | G | D | A | BRCA1 | ENST00000471181 | 0.67684 | 0.3366 | 0.34907 | 0.95204 | 0.88354 | 0.479453892 | 0.87388 | 0.9646285772 | 0.244971 | 6.267925912 | 0 |
| 17 | 43047673 | C | T | D | N | BRCA1 | ENST00000471181 | 0.54977 | 0.3872 | 0.40483 | 0.974 | 0.77909 | 0.3663798041 | 0.86511 | 0.9574999213 | 0.101802 | 6.267925912 | 0 |
| 17 | 43047675 | G | C | P | R | BRCA1 | ENST00000471181 | 0.86516 | 0.45143 | 0.51453 | 0.97435 | 0.91569 | 0.6163551608 | 0.95587 | 0.9960331321 | 0.448198 | 6.053421395 | 0 |
| 17 | 43047676 | G | A | P | S | BRCA1 | ENST00000471181 | 0.73197 | | | 0.9745 | 0.75417 | 0.5163284808 | 0.85774 | 0.9960331321 | 0.0994207 | 4.136432359 | 0 |
| 17 | 43047676 | G | C | P | A | BRCA1 | ENST00000471181 | 0.68243 | | | 0.97436 | 0.73949 | 0.4289662776 | 0.859 | 0.986117065 | 0.0979892 | 6.053421395 | 1 |
| 17 | 43047676 | G | T | P | T | BRCA1 | ENST00000471181 | 0.70567 | | | 0.96855 | 0.77011 | 0.5165545599 | 0.83245 | 0.9960331321 | 0.10229 | 6.053421395 | 0 |
| 17 | 43047677 | C | A | Q | H | BRCA1 | ENST00000471181 | 0.59849 | | | 0.97464 | 0.71861 | 0.7285617149 | 0.75476 | 0.9747804999 | 0.0731622 | 6.279103722 | 0 |
| 17 | 43047677 | C | G | Q | H | BRCA1 | ENST00000471181 | 0.60625 | | | 0.9782 | 0.78286 | 0.7285617149 | 0.7516 | 0.9754245281 | 0.0849212 | 6.279103722 | 0 |
| 17 | 43047678 | T | A | Q | L | BRCA1 | ENST00000471181 | 0.84226 | 0.40857 | 0.28846 | 0.98418 | 0.9248 | 0.6638824986 | 0.91739 | 0.9875673056 | 0.363663 | 6.279103722 | 1 |
| 17 | 43047678 | T | C | Q | R | BRCA1 | ENST00000471181 | 0.8115 | 0.42041 | 0.33917 | 0.97013 | 0.89739 | 0.6577701561 | 0.9593 | 0.9866105914 | 0.29922 | 6.279103722 | 1 |
| 17 | 43047678 | T | G | Q | P | BRCA1 | ENST00000471181 | 0.83337 | 0.47059 | 0.3182 | 0.97683 | 0.89963 | 0.7966903274 | 0.95052 | 0.9884384274 | 0.388123 | 6.279103722 | 1 |
| 17 | 43047679 | G | C | Q | E | BRCA1 | ENST00000471181 | 0.5232 | | | 0.9755 | 0.50304 | 0.6471018183 | 0.77865 | 0.9867417216 | 0.0140358 | 4.276617096 | 0 |
| 17 | 43047679 | G | T | Q | K | BRCA1 | ENST00000471181 | 0.68403 | | | 0.97793 | 0.75349 | 0.6575618141 | 0.78178 | 0.9889861345 | 0.0778767 | 6.279103722 | 1 |
| 17 | 43047681 | A | C | V | G | BRCA1 | ENST00000471181 | 0.76387 | | | 0.98531 | 0.66828 | 0.8273101537 | 0.78202 | 0.98373276 | 0.0146394 | 5.542157915 | 1 |
| 17 | 43047681 | A | G | V | A | BRCA1 | ENST00000471181 | 0.70848 | 0.2518 | 0.23862 | 0.97857 | 0.9045 | 0.5421284588 | 0.96169 | 0.8693732023 | 0.161333 | 5.159013473 | 0 |
| 17 | 43047681 | A | T | V | E | BRCA1 | ENST00000471181 | 0.76966 | | | | | 0.8406078639 | | 0.9840629101 | 0.376545 | 5.542157915 | 1 |
| 17 | 43047682 | C | A | V | L | BRCA1 | ENST00000471181 | 0.22386 | | | 0.95079 | 0.48868 | 0.5702387757 | 0.46822 | 0.9379248023 | -0.0260928 | 3.5666164 | 0 |
| 17 | 43047682 | C | G | V | L | BRCA1 | ENST00000471181 | 0.22822 | | | 0.96224 | 0.53625 | 0.5702387757 | 0.48732 | 0.9647747278 | -0.0308616 | 3.5666164 | 0 |
| 17 | 43047682 | C | T | V | M | BRCA1 | ENST00000471181 | 0.28189 | | | 0.94612 | 0.51843 | 0.5888935836 | 0.67332 | 0.8474328518 | -0.0544868 | 3.029917603 | 0 |
| 17 | 43047684 | A | G | V | A | BRCA1 | ENST00000471181 | 0.83744 | 0.36029 | 0.40167 | 0.98585 | 0.9211 | 0.5820812869 | 0.94569 | 0.9842317104 | 0.469562 | 6.249824603 | 1 |
| 17 | 43047684 | A | T | V | D | BRCA1 | ENST00000471181 | 0.89796 | 0.5018 | 0.40758 | 0.98761 | 0.92561 | 0.9035998393 | 0.91102 | 0.9781660438 | 0.477181 | 6.249824603 | 1 |
| 17 | 43047685 | C | T | V | I | BRCA1 | ENST00000471181 | 0.30596 | | | 0.98208 | 0.44999 | 0.1869111385 | 0.70005 | 0.9543676376 | 0.0522729 | 3.104453977 | 0 |
| 17 | 43047687 | A | C | V | G | BRCA1 | ENST00000471181 | 0.87476 | | | 0.99165 | 0.80298 | 0.8637555544 | 0.8116 | 0.9951920509 | 0.0867389 | 6.497601963 | 1 |
| 17 | 43047688 | C | G | V | L | BRCA1 | ENST00000471181 | 0.48614 | | | 0.97615 | 0.73223 | 0.5032317451 | 0.77529 | 0.9867016673 | 0.0660829 | 5.297401563 | 0 |
| 17 | 43047689 | A | C | I | M | BRCA1 | ENST00000471181 | 0.5429 | | | 0.95699 | 0.51539 | 0.4288140801 | 0.62129 | 0.8278336525 | -0.00660912 | 5.499631881 | 0 |
| 17 | 43047690 | A | C | I | S | BRCA1 | ENST00000471181 | 0.74221 | 0.44202 | 0.0805 | 0.97166 | 0.84694 | 0.7809510679 | 0.90525 | 0.9848060012 | 0.261361 | 5.499631881 | 1 |
| 17 | 43047690 | A | G | I | T | BRCA1 | ENST00000471181 | 0.64423 | 0.39366 | 0.12265 | 0.97732 | 0.86098 | 0.546592615 | 0.98577 | 0.9646333456 | 0.235793 | 5.499631881 | 0 |
| 17 | 43047690 | A | T | I | N | BRCA1 | ENST00000471181 | 0.74714 | 0.50448 | 0.10234 | 0.97076 | 0.8367 | 0.7345685162 | 0.86864 | 0.9836657643 | 0.236539 | 5.499631881 | 1 |
| 17 | 43047691 | T | C | I | V | BRCA1 | ENST00000471181 | 0.03657 | 0.07644 | 0.04455 | 0.83861 | 0.78286 | 0.2112561103 | 0.28531 | 0.01529788544 | -0.110307 | 0.4070365927 | 0 |
| 17 | 43047691 | T | G | I | L | BRCA1 | ENST00000471181 | 0.16398 | 0.09137 | 0.07149 | 0.90508 | 0.77011 | 0.4271099284 | 0.33225 | 0.3216858506 | 0.0460708 | 3.854286331 | 0 |
| 17 | 43047693 | G | A | P | L | BRCA1 | ENST00000471181 | 0.29642 | 0.33589 | 0.14054 | 0.91229 | 0.78348 | 0.4074932826 | 0.67137 | 0.9294394255 | 0.180618 | 5.143312194 | 0 |
| 17 | 43047693 | G | C | P | R | BRCA1 | ENST00000471181 | 0.43859 | 0.40894 | 0.13865 | 0.9317 | 0.82556 | 0.3904154647 | 0.86847 | 0.9617601037 | 0.204731 | 3.559455979 | 0 |
| 17 | 43047693 | G | T | P | Q | BRCA1 | ENST00000471181 | 0.42359 | 0.31557 | 0.09907 | 0.91774 | 0.79577 | 0.3557532752 | 0.84718 | 0.9576046467 | 0.182631 | 5.543833297 | 0 |
| 17 | 43047694 | G | A | P | S | BRCA1 | ENST00000471181 | 0.08616 | | | 0.88517 | 0.37446 | 0.2863240297 | 0.16869 | 0.3094584048 | -0.0910949 | 3.70083636 | 0 |
| 17 | 43047694 | G | C | P | A | BRCA1 | ENST00000471181 | 0.06631 | 0.08898 | 0.10152 | 0.85474 | 0.76881 | 0.2240527064 | 0.62061 | 0.03419035301 | -0.00385282 | 1.634643081 | 0 |
| 17 | 43047694 | G | T | P | T | BRCA1 | ENST00000471181 | 0.1392 | | | 0.95024 | 0.43577 | 0.3375218302 | 0.26916 | 0.7495135665 | -0.0288427 | 5.146475138 | 0 |
| 17 | 43047695 | G | C | H | Q | BRCA1 | ENST00000471181 | 0.07226 | 0.09232 | 0.11786 | 0.91821 | 0.82556 | 0.2723782409 | 0.62199 | 0.1000139166 | 0.132507 | 2.797386362 | 0 |
| 17 | 43047695 | G | T | H | Q | BRCA1 | ENST00000471181 | 0.07011 | 0.09232 | 0.11786 | 0.92256 | 0.82556 | 0.2723782409 | 0.63647 | 0.1021193392 | 0.132503 | 2.797386362 | 0 |
| 17 | 43047696 | T | A | H | L | BRCA1 | ENST00000471181 | 0.15155 | 0.11856 | 0.11614 | 0.91532 | 0.79212 | 0.4383195642 | 0.39908 | 0.7541745305 | 0.159826 | 5.553426128 | 0 |
| 17 | 43047696 | T | C | H | R | BRCA1 | ENST00000471181 | 0.06364 | 0.11157 | 0.12942 | 0.64853 | 0.77972 | 0.3090253322 | 0.65892 | 0.2173109055 | 0.163684 | 2.868383559 | 0 |
| 17 | 43047696 | T | G | H | P | BRCA1 | ENST00000471181 | 0.14736 | 0.24967 | 0.10718 | 0.92042 | 0.80713 | 0.6216890681 | 0.54491 | 0.8566746712 | 0.189053 | 4.435807423 | 0 |
| 17 | 43047697 | G | A | H | Y | BRCA1 | ENST00000471181 | 0.35819 | 0.16864 | 0.13374 | 0.96692 | 0.7589 | 0.2868647219 | 0.46318 | 0.8070001006 | 0.161097 | 3.536214693 | 0 |
| 17 | 43047697 | G | C | H | D | BRCA1 | ENST00000471181 | 0.29099 | 0.15346 | 0.12875 | 0.96633 | 0.76553 | 0.5086285033 | 0.62162 | 0.8517965078 | 0.205316 | 5.553426128 | 0 |
| 17 | 43047697 | G | T | H | N | BRCA1 | ENST00000471181 | 0.26842 | 0.13261 | 0.13226 | 0.96392 | 0.68292 | 0.2435508249 | 0.62413 | 0.6737209558 | 0.0995666 | 5.039656557 | 0 |
| 17 | 43047699 | A | C | V | G | BRCA1 | ENST00000471181 | 0.22139 | | | 0.95896 | 0.34701 | 0.298929897 | 0.25527 | 0.2435555905 | -0.0696216 | 2.598532752 | 0 |
| 17 | 43047699 | A | G | V | A | BRCA1 | ENST00000471181 | 0.0866 | 0.1097 | 0.09191 | 0.70412 | 0.76223 | 0.1732577913 | 0.47177 | 0.02963586699 | 0.0476963 | -1.097260531 | 0 |
| 17 | 43047699 | A | T | V | D | BRCA1 | ENST00000471181 | 0.23061 | | | | | 0.3244336325 | | 0.05925958411 | -0.4536 | 2.598532752 | 0 |

| | | | | | | | | | | | | | | | | | | |
|---|---|---|---|---|---|---|---|---|---|---|---|---|---|---|---|---|---|---|
| 17 | 43047700 | C | A | V | F | BRCA1 | ENST00000471181 | 0.05501 | | | 0.90098 | 0.44293 | 0.3052279557 | 0.05136 | 0.02823626592 | -0.137402 | 2.002419677 | 0 |
| 17 | 43047700 | C | G | V | L | BRCA1 | ENST00000471181 | 0.04649 | | | 0.78212 | 0.47225 | 0.2103403365 | 0.04254 | 0.04092926417 | -0.148171 | 0.899892611 | 0 |
| 17 | 43047700 | C | T | V | I | BRCA1 | ENST00000471181 | 0.03539 | | | 0.8257 | 0.50304 | 0.1454043731 | 0.04254 | 0.03567127018 | -0.140909 | 0.01087628823 | 0 |
| 17 | 43047702 | C | A | G | V | BRCA1 | ENST00000471181 | 0.19998 | 0.40942 | 0.12439 | 0.97313 | 0.82046 | 0.5198381253 | 0.65725 | 0.7972319722 | 0.159144 | 3.583887355 | 0 |
| 17 | 43047702 | C | G | G | A | BRCA1 | ENST00000471181 | 0.10415 | 0.10942 | 0.12947 | 0.95943 | 0.79758 | 0.3610935859 | 0.62034 | 0.3792591095 | 0.0584828 | 3.412287713 | 0 |
| 17 | 43047703 | C | A | G | C | BRCA1 | ENST00000471181 | 0.81284 | 0.43397 | 0.1482 | 0.98573 | 0.85586 | 0.5158118191 | 0.90823 | 0.9876246452 | 0.227495 | 5.538616889 | 0 |
| 17 | 43047703 | C | G | G | R | BRCA1 | ENST00000471181 | 0.58403 | | | 0.98671 | 0.65178 | 0.4513390444 | 0.74191 | 0.9854255915 | 0.0772453 | 5.538616889 | 0 |
| 17 | 43047703 | C | T | G | S | BRCA1 | ENST00000471181 | 0.53408 | | | 0.97638 | 0.66449 | 0.3591894788 | 0.62503 | 0.9438294768 | 0.0800731 | 3.481177726 | 0 |
| 17 | 43049122 | G | A | T | I | BRCA1 | ENST00000471181 | 0.29642 | 0.10761 | 0.10847 | 0.78768 | 0.77654 | 0.1586723103 | 0.73568 | 0.2356425822 | 0.177876 | 3.817594421 | 0 |
| 17 | 43049122 | G | C | T | R | BRCA1 | ENST00000471181 | 0.38759 | 0.13114 | 0.08943 | 0.70457 | 0.7675 | 0.2212880473 | 0.70576 | 0.1396568865 | 0.128128 | 5.038349233 | 0 |
| 17 | 43049122 | G | T | T | K | BRCA1 | ENST00000471181 | 0.2004 | 0.11029 | 0.09568 | 0.47973 | 0.74658 | 0.2152272911 | 0.61513 | 0.08649008461 | 0.101626 | 1.529680273 | 0 |
| 17 | 43049123 | T | A | T | S | BRCA1 | ENST00000471181 | 0.24146 | 0.08366 | 0.0708 | 0.76202 | 0.75687 | 0.1437237836 | 0.62385 | 0.1275887286 | 0.0740164 | 5.038349233 | 0 |
| 17 | 43049123 | T | C | T | A | BRCA1 | ENST00000471181 | 0.16644 | 0.09137 | 0.06848 | 0.73228 | 0.77399 | 0.1566513973 | 0.61781 | 0.05076632347 | 0.0840587 | 2.927562066 | 0 |
| 17 | 43049123 | T | G | T | P | BRCA1 | ENST00000471181 | 0.40354 | 0.16135 | 0.07613 | 0.82177 | 0.80058 | 0.3579016529 | 0.63229 | 0.1951369508 | 0.132418 | 3.297380128 | 0 |
| 17 | 43049125 | C | A | G | V | BRCA1 | ENST00000471181 | 0.29692 | 0.1202 | 0.05905 | 0.77096 | 0.80118 | 0.4118559325 | 0.66566 | 0.2690842656 | 0.156328 | 3.790841557 | 0 |
| 17 | 43049125 | C | G | G | A | BRCA1 | ENST00000471181 | 0.24371 | 0.10581 | 0.05882 | 0.69574 | 0.77399 | 0.3081271853 | 0.64668 | 0.05147963168 | 0.105348 | 5.345944639 | 0 |
| 17 | 43049125 | C | T | G | D | BRCA1 | ENST00000471181 | 0.15926 | 0.11996 | 0.08612 | 0.45795 | 0.74867 | 0.3581833532 | 0.47304 | 0.02343201673 | -0.139374 | 2.112416173 | 0 |
| 17 | 43049126 | C | A | G | C | BRCA1 | ENST00000471181 | 0.39847 | 0.32233 | 0.0646 | 0.83975 | 0.78286 | 0.4510906287 | 0.60445 | 0.586087036 | 0.13443 | 5.345944639 | 0 |
| 17 | 43049126 | C | G | G | R | BRCA1 | ENST00000471181 | 0.16574 | 0.11101 | 0.08272 | 0.69385 | 0.77782 | 0.3251390262 | 0.38032 | 0.1248819605 | 0.107905 | 2.247622086 | 0 |
| 17 | 43049126 | C | T | G | S | BRCA1 | ENST00000471181 | 0.13727 | 0.09137 | 0.05027 | 0.54861 | 0.71073 | 0.2615987157 | 0.46053 | 0.1147575155 | 0.0930511 | 1.70869722 | 0 |
| 17 | 43049128 | A | C | L | R | BRCA1 | ENST00000471181 | 0.22956 | 0.13549 | 0.09777 | 0.59724 | 0.79273 | 0.3650154388 | 0.31575 | 0.05765585229 | 0.0632738 | 2.692979958 | 0 |
| 17 | 43049128 | A | G | L | P | BRCA1 | ENST00000471181 | 0.24137 | 0.15057 | 0.10374 | 0.70584 | 0.79028 | 0.5679628368 | 0.29149 | 0.1206412762 | 0.108296 | 2.511215374 | 0 |
| 17 | 43049128 | A | T | L | H | BRCA1 | ENST00000471181 | 0.21363 | 0.23543 | 0.08986 | 0.75411 | 0.80653 | 0.3415062036 | 0.3102 | 0.430475086 | 0.0703553 | 2.523865187 | 0 |
| 17 | 43049129 | G | A | L | F | BRCA1 | ENST00000471181 | 0.18658 | 0.09913 | 0.0705 | 0.59932 | 0.78535 | 0.2023713556 | 0.63916 | 0.07557454705 | 0.0959438 | 2.333629515 | 0 |
| 17 | 43049129 | G | C | L | V | BRCA1 | ENST00000471181 | 0.16874 | 0.09179 | 0.05953 | 0.57756 | 0.78098 | 0.2123937217 | 0.64072 | 0.03628909588 | 0.114987 | 2.503330998 | 0 |
| 17 | 43049129 | G | T | L | I | BRCA1 | ENST00000471181 | 0.17892 | 0.09878 | 0.07796 | 0.61452 | 0.74658 | 0.1374765262 | 0.51735 | 0.04445133731 | 0.0667561 | 5.295892233 | 0 |
| 17 | 43049131 | G | A | T | I | BRCA1 | ENST00000471181 | 0.75869 | 0.16042 | 0.27735 | 0.89062 | 0.84106 | 0.3609902343 | 0.93842 | 0.5703939143 | 0.0907932 | 5.883500223 | 0 |
| 17 | 43049131 | G | C | T | S | BRCA1 | ENST00000471181 | 0.40553 | 0.0855 | 0.19716 | 0.80623 | 0.8233 | 0.2130361129 | 0.93566 | 0.7133171558 | 0.145218 | 2.713592555 | 0 |
| 17 | 43049131 | G | T | T | N | BRCA1 | ENST00000471181 | 0.71161 | 0.27023 | 0.24061 | 0.87712 | 0.82273 | 0.287648638 | 0.93309 | 0.8769288063 | 0.120496 | 5.883500223 | 0 |
| 17 | 43049132 | T | A | T | S | BRCA1 | ENST00000471181 | 0.36002 | 0.0855 | 0.19716 | 0.81028 | 0.8233 | 0.2130361129 | 0.92241 | 0.7136557102 | 0.141107 | 2.713592555 | 0 |
| 17 | 43049132 | T | C | T | A | BRCA1 | ENST00000471181 | 0.64151 | 0.10076 | 0.18895 | 0.80259 | 0.82556 | 0.3011671392 | 0.92471 | 0.7136557102 | 0.181108 | 5.883500223 | 0 |
| 17 | 43049132 | T | G | T | P | BRCA1 | ENST00000471181 | 0.56929 | 0.39101 | 0.19892 | 0.852 | 0.81298 | 0.5229485614 | 0.88276 | 0.8532059193 | 0.177699 | 3.310630934 | 0 |
| 17 | 43049133 | G | C | F | L | BRCA1 | ENST00000471181 | 0.40693 | 0.10982 | 0.27441 | 0.65778 | 0.72926 | 0.4779347541 | 0.89226 | 0.5584899187 | 0.0915536 | 1.880314925 | 0 |
| 17 | 43049133 | G | T | F | L | BRCA1 | ENST00000471181 | 0.4022 | 0.10982 | 0.27441 | 0.6759 | 0.72626 | 0.4779347541 | 0.89226 | 0.5584899187 | 0.0915606 | 1.880314925 | 0 |
| 17 | 43049134 | A | C | F | C | BRCA1 | ENST00000471181 | 0.9059 | 0.49279 | 0.27551 | 0.9252 | 0.79273 | 0.6615798301 | 0.91081 | 0.9518437982 | 0.227994 | 5.793312176 | 0 |
| 17 | 43049134 | A | G | F | S | BRCA1 | ENST00000471181 | 0.90058 | 0.47806 | 0.25728 | 0.92189 | 0.76421 | 0.6619798649 | 0.89999 | 0.946547389 | 0.204717 | 3.744724596 | 0 |
| 17 | 43049134 | A | T | F | Y | BRCA1 | ENST00000471181 | 0.82116 | 0.35735 | 0.27274 | 0.81829 | 0.74235 | 0.4839791779 | 0.91655 | 0.9063339233 | 0.131504 | 5.793312176 | 0 |
| 17 | 43049135 | A | C | F | V | BRCA1 | ENST00000471181 | 0.73221 | 0.1439 | 0.2269 | 0.88098 | 0.79938 | 0.5914492948 | 0.91838 | 0.5163696247 | 0.204825 | 5.793312176 | 0 |
| 17 | 43049135 | A | G | F | L | BRCA1 | ENST00000471181 | 0.37278 | 0.10982 | 0.27441 | 0.6334 | 0.71233 | 0.4779347541 | 0.90063 | 0.7140513062 | 0.0791952 | 1.880314925 | 0 |
| 17 | 43049135 | A | T | F | I | BRCA1 | ENST00000471181 | 0.71105 | 0.1975 | 0.25356 | 0.81835 | 0.78473 | 0.5615750599 | 0.96028 | 0.6489474773 | 0.130343 | 5.793312176 | 0 |
| 17 | 43049137 | G | A | S | L | BRCA1 | ENST00000471181 | 0.24787 | 0.08157 | 0.07936 | 0.74908 | 0.78098 | 0.2707048949 | 0.39223 | 0.0829372853 | 0.0647567 | 1.805842323 | 0 |
| 17 | 43049138 | A | C | S | A | BRCA1 | ENST00000471181 | 0.19491 | 0.08601 | 0.07329 | 0.75569 | 0.73877 | 0.1968113418 | 0.86762 | 0.5191012025 | 0.1166 | 3.68938104 | 0 |
| 17 | 43049138 | A | G | S | P | BRCA1 | ENST00000471181 | 0.41777 | 0.11614 | 0.09661 | 0.8799 | 0.78967 | 0.6681171881 | 0.90696 | 0.7268800139 | 0.214606 | 4.99712119 | 0 |
| 17 | 43049138 | A | T | S | T | BRCA1 | ENST00000471181 | 0.23861 | 0.09339 | 0.08727 | 0.83972 | 0.74936 | 0.2535616922 | 0.88062 | 0.4649033248 | 0.187356 | 5.61104387 | 0 |
| 17 | 43049141 | A | C | S | A | BRCA1 | ENST00000471181 | 0.24384 | 0.0765 | 0.16972 | 0.80359 | 0.74021 | 0.1955349046 | 0.7896 | 0.06796205363 | 0.0794605 | 5.896868364 | 0 |
| 17 | 43049141 | A | G | S | P | BRCA1 | ENST00000471181 | 0.27199 | 0.25897 | 0.19231 | 0.85459 | 0.81933 | 0.6095866698 | 0.77576 | 0.3180524111 | 0.128721 | 3.594851877 | 0 |
| 17 | 43049141 | A | T | S | T | BRCA1 | ENST00000471181 | 0.21577 | 0.08323 | 0.18655 | 0.77481 | 0.78906 | 0.2458083519 | 0.80208 | 0.1218478978 | 0.104332 | 4.084314083 | 0 |
| 17 | 43049143 | A | C | L | R | BRCA1 | ENST00000471181 | 0.55415 | 0.37861 | 0.12852 | 0.91345 | 0.83505 | 0.6070451979 | 0.89644 | 0.6598553163 | 0.180152 | 4.687291845 | 0 |
| 17 | 43049143 | A | G | L | P | BRCA1 | ENST00000471181 | 0.29954 | 0.15289 | 0.13417 | 0.80571 | 0.80298 | 0.6225819131 | 0.49253 | 0.1222885177 | 0.165081 | 0.05436919229 | 0 |
| 17 | 43049143 | A | T | L | H | BRCA1 | ENST00000471181 | 0.54814 | 0.44778 | 0.12255 | 0.93025 | 0.82046 | 0.5186968509 | 0.89509 | 0.6830230081 | 0.164152 | 4.687291845 | 0 |
| 17 | 43049144 | G | A | L | F | BRCA1 | ENST00000471181 | 0.32068 | 0.32958 | 0.07759 | 0.85781 | 0.76356 | 0.3651256071 | 0.96204 | 0.4800254728 | 0.110082 | 4.139406512 | 0 |
| 17 | 43049144 | G | C | L | V | BRCA1 | ENST00000471181 | 0.29288 | 0.10624 | 0.06448 | 0.78683 | 0.78906 | 0.3739060738 | 0.8722 | 0.4942937493 | 0.185966 | 3.554377893 | 0 |
| 17 | 43049144 | G | T | L | I | BRCA1 | ENST00000471181 | 0.28583 | 0.10753 | 0.10633 | 0.81489 | 0.77782 | 0.2535176248 | 0.80177 | 0.6666688919 | 0.102746 | 2.58648705 | 0 |
| 17 | 43049145 | C | A | E | D | BRCA1 | ENST00000471181 | 0.32552 | 0.08273 | 0.19991 | 0.77119 | 0.81065 | 0.2738512182 | 0.70625 | 0.7171704769 | 0.327958 | 3.601760271 | 0 |
| 17 | 43049145 | C | G | E | D | BRCA1 | ENST00000471181 | 0.33115 | 0.08273 | 0.19991 | 0.76484 | 0.79878 | 0.2738512182 | 0.70625 | 0.2316276133 | 0.283651 | 3.601760271 | 0 |
| 17 | 43049146 | T | A | E | V | BRCA1 | ENST00000471181 | 0.77742 | 0.19695 | 0.17847 | 0.95659 | 0.88637 | 0.5571874843 | 0.83866 | 0.940534234 | 0.246457 | 5.750801491 | 0 |
| 17 | 43049146 | T | C | E | G | BRCA1 | ENST00000471181 | 0.79587 | 0.29747 | 0.1686 | 0.95175 | 0.87684 | 0.4724188489 | 0.88382 | 0.9115646482 | 0.22374 | 5.750801491 | 0 |
| 17 | 43049146 | T | G | E | A | BRCA1 | ENST00000471181 | 0.73405 | 0.18105 | 0.19866 | 0.94172 | 0.86047 | 0.4460787158 | 0.86538 | 0.7616618276 | 0.209625 | 4.027981973 | 0 |
| 17 | 43049147 | C | G | E | Q | BRCA1 | ENST00000471181 | 0.48506 | 0.22604 | 0.18212 | 0.87777 | 0.8306 | 0.3869657379 | 0.89075 | 0.7149195671 | 0.229917 | 3.143240096 | 0 |
| 17 | 43049147 | C | T | E | K | BRCA1 | ENST00000471181 | 0.5728 | 0.21061 | 0.2706 | 0.80484 | 0.80417 | 0.384306799 | 0.84949 | 0.7130561471 | 0.067731 | 2.790598725 | 0 |
| 17 | 43049148 | C | G | K | N | BRCA1 | ENST00000471181 | 0.64867 | 0.37065 | 0.33111 | 0.9543 | 0.88259 | 0.6389852119 | 0.87008 | 0.9822084308 | 0.146055 | 6.098933495 | 0 |
| 17 | 43049149 | T | A | K | M | BRCA1 | ENST00000471181 | 0.7434 | 0.16271 | 0.23617 | 0.97233 | 0.91314 | 0.6863085456 | 0.8724 | 0.9772709012 | 0.293112 | 6.098933495 | 0 |
| 17 | 43049149 | T | C | K | R | BRCA1 | ENST00000471181 | 0.41357 | 0.08036 | 0.24275 | 0.90648 | 0.83505 | 0.4198685263 | 0.84697 | 0.878059566 | 0.210827 | 2.927250997 | 0 |
| 17 | 43049149 | T | G | K | T | BRCA1 | ENST00000471181 | 0.57645 | 0.29852 | 0.31983 | 0.9743 | 0.91526 | 0.6596442699 | 0.85528 | 0.9702286124 | 0.250019 | 4.691139473 | 0 |
| 17 | 43049150 | T | C | K | E | BRCA1 | ENST00000471181 | 0.75408 | 0.25583 | 0.29935 | 0.93119 | 0.88684 | 0.6719363917 | 0.83922 | 0.9514852762 | 0.275416 | 6.098933495 | 0 |
| 17 | 43049150 | T | G | K | Q | BRCA1 | ENST00000471181 | 0.72427 | 0.21991 | 0.25909 | 0.94003 | 0.86553 | 0.5505345599 | 0.85536 | 0.9248048663 | 0.238234 | 6.098933495 | 0 |
| 17 | 43049152 | A | C | V | G | BRCA1 | ENST00000471181 | 0.81653 | 0.48666 | 0.31794 | 0.98648 | 0.93554 | 0.850686829 | 0.99202 | 0.9930023551 | 0.451874 | 6.375466993 | 0 |
| 17 | 43049152 | A | G | V | A | BRCA1 | ENST00000471181 | 0.78495 | 0.44312 | 0.42858 | 0.98316 | 0.92763 | 0.6349415542 | 0.98123 | 0.9812527299 | 0.417136 | 6.375466993 | 0 |
| 17 | 43049153 | C | A | V | L | BRCA1 | ENST00000471181 | 0.71041 | 0.21536 | 0.41186 | 0.96695 | 0.85586 | 0.6452662221 | 0.98335 | 0.9742401242 | 0.337669 | 6.375466993 | 0 |
| 17 | 43049153 | C | G | V | L | BRCA1 | ENST00000471181 | 0.71891 | 0.21536 | 0.41186 | 0.96563 | 0.85586 | 0.6452662221 | 0.98335 | 0.9672945738 | 0.33767 | 6.375466993 | 0 |
| 17 | 43049153 | C | T | V | M | BRCA1 | ENST00000471181 | 0.74722 | 0.44272 | 0.44161 | 0.97598 | 0.8887 | 0.6375573622 | 0.98808 | 0.991306603 | 0.354172 | 6.375466993 | 0 |
| 17 | 43049155 | A | C | V | G | BRCA1 | ENST00000471181 | 0.81528 | 0.14329 | 0.40699 | 0.98022 | 0.94431 | 0.8772505485 | 0.98698 | 0.9676528573 | 0.45796 | 6.410703152 | 0 |
| 17 | 43049155 | A | G | V | A | BRCA1 | ENST00000471181 | 0.78043 | 0.29656 | 0.53319 | 0.97914 | 0.90929 | 0.6358664857 | 0.98792 | 0.8678325415 | 0.274638 | 5.178155399 | 0 |
| 17 | 43049156 | C | A | V | L | BRCA1 | ENST00000471181 | 0.64539 | 0.12879 | 0.51304 | 0.96248 | 0.86403 | 0.5918844182 | 0.97978 | 0.9446972013 | 0.291897 | 6.410703152 | 0 |
| 17 | 43049156 | C | G | V | L | BRCA1 | ENST00000471181 | 0.65366 | 0.12879 | 0.51304 | 0.96201 | 0.86403 | 0.5918844182 | 0.97978 | 0.9397184253 | 0.291898 | 6.410703152 | 0 |
| 17 | 43049156 | C | T | V | M | BRCA1 | ENST00000471181 | 0.72804 | 0.42664 | 0.54951 | 0.97001 | 0.87148 | 0.6382755079 | 0.98711 | 0.9806551933 | 0.307633 | 6.410703152 | 0 |
| 17 | 43049158 | G | A | S | F | BRCA1 | ENST00000471181 | 0.82248 | 0.11741 | 0.34041 | 0.95272 | 0.82668 | 0.4563134313 | 0.95512 | 0.9620023966 | 0.319395 | 6.038437767 | 0 |
| 17 | 43049158 | G | C | S | C | BRCA1 | ENST00000471181 | 0.80606 | 0.39763 | 0.2998 | 0.95386 | 0.83943 | 0.4882332707 | 0.97382 | 0.9654983878 | 0.253733 | 4.905324986 | 0 |
| 17 | 43049158 | G | T | S | Y | BRCA1 | ENST00000471181 | 0.77766 | 0.39859 | 0.36714 | 0.95873 | 0.86453 | 0.4848214143 | 0.95627 | 0.9790061116 | 0.305197 | 6.038437767 | 0 |
| 17 | 43049159 | A | C | S | A | BRCA1 | ENST00000471181 | 0.47753 | 0.16604 | 0.27401 | 0.945 | 0.77654 | 0.2450395701 | 0.85008 | 0.8311126828 | 0.184963 | 4.795059315 | 0 |
| 17 | 43049159 | A | G | S | P | BRCA1 | ENST00000471181 | 0.71706 | 0.42406 | 0.30905 | 0.97588 | 0.84906 | 0.8005578674 | 0.91701 | 0.8756486177 | 0.297424 | 6.038437767 | 0 |
| 17 | 43049159 | A | T | S | T | BRCA1 | ENST00000471181 | 0.46593 | 0.15383 | 0.30108 | 0.91669 | 0.76223 | 0.2812332914 | 0.80184 | 0.3540291952 | 0.156324 | 3.232496084 | 0 |
| 17 | 43049161 | G | A | A | V | BRCA1 | ENST00000471181 | 0.80874 | 0.13296 | 0.2983 | 0.97891 | 0.91652 | 0.8682206471 | 0.97538 | 0.9672945738 | 0.470221 | 6.541286629 | 1 |
| 17 | 43049161 | G | C | A | G | BRCA1 | ENST00000471181 | 0.8066 | 0.24205 | 0.26284 | 0.96286 | 0.87781 | 0.7642663166 | 0.9509 | 0.9778698683 | 0.287836 | 6.541286629 | 0 |
| 17 | 43049161 | G | T | A | D | BRCA1 | ENST00000471181 | 0.82006 | 0.49743 | 0.38521 | 0.98622 | 0.93747 | 0.9484033491 | 0.97764 | 0.992405951 | 0.482561 | 6.541286629 | 1 |
| 17 | 43049162 | C | A | A | S | BRCA1 | ENST00000471181 | 0.70038 | 0.19022 | 0.24405 | 0.97861 | 0.90842 | 0.7755930888 | 0.95689 | 0.9705228209 | 0.438509 | 6.541286629 | 0 |
| 17 | 43049162 | C | G | A | P | BRCA1 | ENST00000471181 | 0.71386 | 0.4773 | 0.34402 | 0.98553 | 0.93358 | 0.9382548403 | 0.97847 | 0.9929260015 | 0.47533 | 6.541286629 | 1 |
| 17 | 43049162 | C | T | A | T | BRCA1 | ENST00000471181 | 0.7426 | 0.33317 | 0.27968 | 0.98401 | 0.9328 | 0.8339468559 | 0.97396 | 0.9287589192 | 0.473837 | 6.541286629 | 1 |
| 17 | 43049164 | C | A | G | V | BRCA1 | ENST00000471181 | 0.70487 | 0.47559 | 0.4138 | 0.99254 | 0.95645 | 0.9165863792 | 0.97308 | 0.9946420193 | 0.53403 | 6.410634448 | 1 |
| 17 | 43049164 | C | G | G | A | BRCA1 | ENST00000471181 | 0.67517 | 0.2903 | 0.39578 | 0.98975 | 0.96008 | 0.8392247563 | 0.98775 | 0.9796954393 | 0.533374 | 6.410634448 | 1 |
| 17 | 43049164 | C | T | G | D | BRCA1 | ENST00000471181 | 0.70744 | 0.47395 | 0.53797 | 0.99057 | 0.95863 | 0.8705809198 | 0.98957 | 0.9697861075 | 0.466255 | 6.410634448 | 0 |
| 17 | 43049165 | C | A | G | C | BRCA1 | ENST00000471181 | 0.82553 | 0.45422 | 0.45303 | 0.99245 | 0.9608 | 0.8819313644 | 0.97011 | 0.9946420193 | 0.53403 | 6.410634448 | 1 |
| 17 | 43049165 | C | G | G | R | BRCA1 | ENST00000471181 | 0.79832 | 0.46743 | 0.52166 | 0.99218 | 0.9528 | 0.8829322157 | 0.96977 | 0.9946420193 | 0.533874 | 6.410634448 | 1 |
| 17 | 43049165 | C | T | G | S | BRCA1 | ENST00000471181 | 0.76848 | 0.25737 | 0.38511 | 0.98936 | 0.95935 | 0.8253359382 | 0.98963 | 0.9936221242 | 0.533308 | 6.410634448 | 1 |
| 17 | 43049166 | A | C | C | W | BRCA1 | ENST00000471181 | 0.78811 | 0.48572 | 0.56817 | 0.98623 | 0.93123 | 0.8749118137 | 0.97102 | 0.9950957298 | 0.430692 | 6.482816679 | 1 |
| 17 | 43049167 | C | A | C | F | BRCA1 | ENST00000471181 | 0.813 | 0.16946 | 0.51346 | 0.97537 | 0.92275 | 0.8481751988 | 0.98907 | 0.9787903428 | 0.442562 | 6.482816679 | 0 |
| 17 | 43049167 | C | G | C | S | BRCA1 | ENST00000471181 | 0.77124 | 0.36277 | 0.50243 | 0.97608 | 0.91399 | 0.7707658455 | 0.98869 | 0.9849267006 | 0.444662 | 6.482816679 | 0 |
| 17 | 43049167 | C | T | C | Y | BRCA1 | ENST00000471181 | 0.79524 | 0.46887 | 0.62517 | 0.97796 | 0.93123 | 0.8447876758 | 0.99426 | 0.9876686931 | 0.444238 | 6.482816679 | 1 |
| 17 | 43049168 | A | C | C | G | BRCA1 | ENST00000471181 | 0.80684 | 0.4369 | 0.47036 | 0.97407 | 0.93358 | 0.7714216084 | 0.98907 | 0.9540185928 | 0.43835 | 4.066964389 | 0 |
| 17 | 43049168 | A | G | C | R | BRCA1 | ENST00000471181 | 0.8217 | 0.53095 | 0.60971 | 0.98177 | 0.94129 | 0.9089639051 | 0.98741 | 0.9833028913 | 0.441747 | 6.482816679 | 1 |
| 17 | 43049168 | A | T | C | S | BRCA1 | ENST00000471181 | 0.75805 | 0.36277 | 0.50243 | 0.97603 | 0.94015 | 0.7707658455 | 0.98469 | 0.9848070145 | 0.441745 | 6.482816679 | 0 |

| | | | | | | | | | | | | | | | | | | |
|---|---|---|---|---|---|---|---|---|---|---|---|---|---|---|---|---|---|---|
| 17 | 43049170 | A | C | L | R | BRCA1 | ENST00000471181 | 0.81418 | 0.48394 | 0.52029 | 0.98349 | 0.89239 | 0.7885883734 | 0.98514 | 0.9876952171 | 0.419233 | 6.541287373 | 0 |
| 17 | 43049170 | A | G | L | P | BRCA1 | ENST00000471181 | 0.81904 | 0.52378 | 0.5563 | 0.98446 | 0.90711 | 0.8887133235 | 0.98551 | 0.9634432793 | 0.448922 | 6.541287373 | 1 |
| 17 | 43049170 | A | T | L | Q | BRCA1 | ENST00000471181 | 0.80463 | 0.40063 | 0.44628 | 0.98364 | 0.86803 | 0.6119397954 | 0.9886 | 0.9888792038 | 0.436012 | 6.541287373 | 0 |
| 17 | 43049171 | G | C | L | V | BRCA1 | ENST00000471181 | 0.75551 | 0.3052 | 0.3616 | 0.94992 | 0.81818 | 0.4907609166 | 0.98593 | 0.9649550319 | 0.3028 | 6.541287373 | 0 |
| 17 | 43049171 | G | T | L | M | BRCA1 | ENST00000471181 | 0.7965 | 0.43492 | 0.39861 | 0.94963 | 0.78286 | 0.4115677874 | 0.97564 | 0.9829454422 | 0.261036 | 6.541287373 | 0 |
| 17 | 43049172 | C | A | Q | H | BRCA1 | ENST00000471181 | 0.21138 | 0.08593 | 0.01485 | 0.75729 | 0.77141 | 0.3467166122 | 0.65868 | 0.03151377533 | 0.225916 | 3.700521135 | 0 |
| 17 | 43049172 | C | G | Q | H | BRCA1 | ENST00000471181 | 0.21558 | 0.08593 | 0.01485 | 0.75215 | 0.77141 | 0.3467166122 | 0.65868 | 0.03599596586 | 0.225913 | 3.700521135 | 0 |
| 17 | 43049173 | T | A | Q | L | BRCA1 | ENST00000471181 | 0.27222 | 0.08922 | 0.00942 | 0.91524 | 0.81006 | 0.4705229355 | 0.59868 | 0.09103874352 | 0.103063 | 4.206269831 | 0 |
| 17 | 43049173 | T | C | Q | R | BRCA1 | ENST00000471181 | 0.22652 | 0.11829 | 0.01709 | 0.76846 | 0.7589 | 0.3241969084 | 0.59858 | 0.02826620574 | 0.144134 | 1.805588261 | 0 |
| 17 | 43049173 | T | G | Q | P | BRCA1 | ENST00000471181 | 0.36811 | 0.25884 | 0.01347 | 0.95906 | 0.84853 | 0.7524430573 | 0.77534 | 0.4754921496 | 0.222267 | 5.51402118 | 1 |
| 17 | 43049174 | G | C | Q | E | BRCA1 | ENST00000471181 | 0.25903 | 0.10718 | 0.01232 | 0.75871 | 0.69594 | 0.2742215214 | 0.87052 | 0.1730521172 | 0.182149 | 2.990208176 | 0 |
| 17 | 43049174 | G | T | Q | K | BRCA1 | ENST00000471181 | 0.38799 | 0.11694 | 0.01923 | 0.80928 | 0.72474 | 0.4240918271 | 0.89937 | 0.5502777696 | 0.193505 | 4.411067666 | 0 |
| 17 | 43049176 | A | C | V | G | BRCA1 | ENST00000471181 | 0.76371 | 0.41941 | 0.08412 | 0.96967 | 0.87442 | 0.8398109755 | 0.98438 | 0.6403096383 | 0.438798 | 6.077571437 | 0 |
| 17 | 43049176 | A | G | V | A | BRCA1 | ENST00000471181 | 0.60471 | 0.43186 | 0.15239 | 0.96736 | 0.87296 | 0.6803038513 | 0.9842 | 0.9022991061 | 0.353497 | 5.079385229 | 0 |
| 17 | 43049176 | A | T | V | E | BRCA1 | ENST00000471181 | 0.80337 | 0.53551 | 0.13732 | 0.97224 | 0.85945 | 0.8989918997 | 0.98024 | 0.9654320478 | 0.438473 | 6.077571437 | 1 |
| 17 | 43049177 | C | A | V | L | BRCA1 | ENST00000471181 | 0.24568 | 0.37553 | 0.13056 | 0.8775 | 0.75143 | 0.5889241966 | 0.8968 | 0.7870392799 | 0.164549 | 2.21460635 | 0 |
| 17 | 43049177 | C | G | V | L | BRCA1 | ENST00000471181 | 0.25036 | 0.37553 | 0.13056 | 0.8751 | 0.75143 | 0.5889241966 | 0.8968 | 0.7442888021 | 0.164529 | 2.21460635 | 0 |
| 17 | 43049177 | C | T | V | I | BRCA1 | ENST00000471181 | 0.38903 | 0.3693 | 0.06763 | 0.91923 | 0.65673 | 0.3219014009 | 0.82587 | 0.910831809 | 0.112948 | 6.077571437 | 0 |
| 17 | 43049178 | C | A | M | I | BRCA1 | ENST00000471181 | 0.40904 | 0.15665 | 0.35603 | 0.94196 | 0.76487 | 0.7695472742 | 0.76942 | 0.6233174205 | 0.172545 | 4.387135917 | 0 |
| 17 | 43049178 | C | G | M | I | BRCA1 | ENST00000471181 | 0.41528 | 0.15665 | 0.35603 | 0.93464 | 0.76487 | 0.7695472742 | 0.76942 | 0.6233174205 | 0.172547 | 4.387135917 | 0 |
| 17 | 43049178 | C | T | M | I | BRCA1 | ENST00000471181 | 0.41932 | 0.15665 | 0.35603 | 0.93599 | 0.76487 | 0.7695472742 | 0.76942 | 0.6233174205 | 0.172525 | 4.387135917 | 0 |
| 17 | 43049179 | A | C | M | R | BRCA1 | ENST00000471181 | 0.78344 | 0.41303 | 0.40908 | 0.97097 | 0.8859 | 0.9642800711 | 0.95582 | 0.9729756713 | 0.373917 | 6.246425503 | 1 |
| 17 | 43049179 | A | G | M | T | BRCA1 | ENST00000471181 | 0.70832 | 0.36357 | 0.46764 | 0.96882 | 0.85689 | 0.8473644705 | 0.94417 | 0.1209916355 | -0.0044913 | 6.246425503 | 0 |
| 17 | 43049179 | A | T | M | K | BRCA1 | ENST00000471181 | 0.7536 | 0.27978 | 0.43148 | 0.95955 | 0.86653 | 0.9557007442 | 0.95406 | 0.9728270769 | 0.34099 | 6.246425503 | 1 |
| 17 | 43049180 | T | A | M | L | BRCA1 | ENST00000471181 | 0.57914 | 0.13733 | 0.34714 | 0.88529 | 0.75822 | 0.6979287923 | 0.80067 | 0.9228143096 | 0.223444 | 5.831418744 | 0 |
| 17 | 43049180 | T | C | M | V | BRCA1 | ENST00000471181 | 0.52008 | 0.09959 | 0.28652 | 0.956 | 0.82668 | 0.7489110899 | 0.88426 | 0.7836143374 | 0.196752 | 6.246425503 | 0 |
| 17 | 43049180 | T | G | M | L | BRCA1 | ENST00000471181 | 0.58645 | 0.13733 | 0.34714 | 0.88529 | 0.75822 | 0.6979287923 | 0.80067 | 0.2783330624 | -0.0292879 | 5.831418744 | 0 |
| 17 | 43049181 | C | A | W | C | BRCA1 | ENST00000471181 | 0.73005 | 0.52572 | 0.36497 | 0.93292 | 0.80713 | 0.7562311659 | 0.80343 | 0.9905490279 | 0.279111 | 4.349867921 | 0 |
| 17 | 43049181 | C | G | W | C | BRCA1 | ENST00000471181 | 0.73853 | 0.52572 | 0.36497 | 0.92551 | 0.80595 | 0.7562311659 | 0.80343 | 0.9905490279 | 0.279109 | 4.349867921 | 0 |
| 17 | 43049182 | C | A | W | L | BRCA1 | ENST00000471181 | 0.7023 | 0.5053 | 0.36887 | 0.92817 | 0.79273 | 0.6563761418 | 0.84647 | 0.9784641266 | 0.238842 | 5.985989333 | 0 |
| 17 | 43049182 | C | G | W | S | BRCA1 | ENST00000471181 | 0.74053 | 0.32536 | 0.35392 | 0.92946 | 0.77909 | 0.6920895983 | 0.84468 | 0.9584062696 | 0.24855 | 5.985989333 | 0 |
| 17 | 43049183 | A | C | W | G | BRCA1 | ENST00000471181 | 0.53616 | 0.48825 | 0.32407 | 0.95175 | 0.83725 | 0.6372961395 | 0.75462 | 0.963567853 | 0.246385 | 5.985989333 | 0 |
| 17 | 43049183 | A | G | W | R | BRCA1 | ENST00000471181 | 0.37816 | 0.53771 | 0.41464 | 0.94336 | 0.79938 | 0.7169085553 | 0.89232 | 0.9627318382 | 0.223644 | 3.004405794 | 0 |
| 17 | 43049183 | A | T | W | R | BRCA1 | ENST00000471181 | 0.36873 | 0.53771 | 0.41464 | 0.94171 | 0.77718 | 0.7169085553 | 0.89232 | 0.9627318382 | 0.223626 | 3.004405794 | 0 |
| 17 | 43049184 | T | A | E | D | BRCA1 | ENST00000471181 | 0.30822 | 0.17281 | 0.27895 | 0.92309 | 0.80238 | 0.6410131758 | 0.77756 | 0.7951770425 | 0.170275 | 6.52861499 | 0 |
| 17 | 43049184 | T | G | E | D | BRCA1 | ENST00000471181 | 0.31353 | 0.17281 | 0.27895 | 0.92181 | 0.80238 | 0.6410131758 | 0.77756 | 0.8468011618 | 0.170277 | 6.52861499 | 0 |
| 17 | 43049185 | T | A | E | V | BRCA1 | ENST00000471181 | 0.76539 | 0.31797 | 0.25155 | 0.97493 | 0.85586 | 0.6914001239 | 0.87098 | 0.8796597719 | 0.301591 | 6.52861499 | 0 |
| 17 | 43049185 | T | G | E | A | BRCA1 | ENST00000471181 | 0.73957 | 0.12733 | 0.27705 | 0.96706 | 0.88401 | 0.6677788613 | 0.89249 | 0.8914160132 | 0.241491 | 6.52861499 | 0 |
| 17 | 43049186 | C | G | E | Q | BRCA1 | ENST00000471181 | 0.46239 | 0.32404 | 0.26828 | 0.89879 | 0.74867 | 0.5883801581 | 0.9632 | 0.8245936632 | 0.237293 | 6.52861499 | 0 |
| 17 | 43049186 | C | T | E | K | BRCA1 | ENST00000471181 | 0.51362 | 0.1243 | 0.3608 | 0.91856 | 0.79151 | 0.7330509069 | 0.95377 | 0.7980544661 | 0.252714 | 6.52861499 | 0 |
| 17 | 43049188 | A | C | L | R | BRCA1 | ENST00000471181 | 0.62567 | 0.49051 | 0.39565 | 0.98495 | 0.88117 | 0.90662904 | 0.96581 | 0.9648061991 | 0.447114 | 6.528614956 | 1 |
| 17 | 43049188 | A | G | L | P | BRCA1 | ENST00000471181 | 0.6365 | 0.52297 | 0.44492 | 0.98574 | 0.87733 | 0.9245295532 | 0.95948 | 0.9631689191 | 0.331444 | 6.528614956 | 1 |
| 17 | 43049189 | G | C | L | V | BRCA1 | ENST00000471181 | 0.35168 | 0.20516 | 0.25208 | 0.95566 | 0.83394 | 0.5634402639 | 0.95372 | 0.7460424304 | 0.267804 | 6.528614956 | 0 |
| 17 | 43049190 | T | A | Q | H | BRCA1 | ENST00000471181 | 0.32462 | 0.42685 | 0.15783 | 0.94062 | 0.80058 | 0.4085054811 | 0.66638 | 0.6677427888 | 0.186748 | 2.890214557 | 0 |
| 17 | 43049190 | T | G | Q | H | BRCA1 | ENST00000471181 | 0.33003 | 0.42685 | 0.15783 | 0.94068 | 0.80653 | 0.4085054811 | 0.66638 | 0.6677427888 | 0.186749 | 2.890214557 | 0 |
| 17 | 43049191 | T | A | Q | L | BRCA1 | ENST00000471181 | 0.71442 | 0.27514 | 0.12525 | 0.96845 | 0.81588 | 0.5388051639 | 0.88927 | 0.92724967 | 0.213874 | 6.099747959 | 0 |
| 17 | 43049191 | T | C | Q | R | BRCA1 | ENST00000471181 | 0.655 | 0.43233 | 0.15868 | 0.95071 | 0.8306 | 0.4562801329 | 0.90019 | 0.8123737844 | 0.247246 | 6.099747959 | 0 |
| 17 | 43049191 | T | G | Q | P | BRCA1 | ENST00000471181 | 0.71506 | 0.46932 | 0.13463 | 0.9644 | 0.84482 | 0.8025122388 | 0.90012 | 0.8492360262 | 0.261543 | 6.099747959 | 0 |
| 17 | 43049192 | G | C | Q | E | BRCA1 | ENST00000471181 | 0.27015 | 0.2494 | 0.13327 | 0.8518 | 0.62547 | 0.3819232533 | 0.82804 | 0.3886963469 | 0.166213 | 3.13969027 | 0 |
| 17 | 43049192 | G | T | Q | K | BRCA1 | ENST00000471181 | 0.40366 | 0.37403 | 0.1701 | 0.94894 | 0.81876 | 0.525844665 | 0.87167 | 0.7846821094 | 0.218548 | 5.469545787 | 0 |
| 17 | 43049193 | A | C | D | E | BRCA1 | ENST00000471181 | 0.31655 | 0.106 | 0.24676 | | 0.80238 | 0.223530192 | 0.73885 | 0.798484683 | 0.177911 | 3.746272049 | 0 |
| 17 | 43049193 | A | T | D | E | BRCA1 | ENST00000471181 | 0.32157 | 0.106 | 0.24676 | | 0.80238 | 0.223530192 | 0.73885 | 0.798484683 | 0.177908 | 3.746272049 | 0 |
| 17 | 43049194 | T | A | D | V | BRCA1 | ENST00000471181 | 0.6628 | 0.31276 | 0.21647 | 0.98525 | 0.91015 | 0.5542150203 | 0.77678 | 0.9284366369 | 0.311007 | 6.494923171 | 0 |
| 17 | 43049194 | T | C | D | G | BRCA1 | ENST00000471181 | 0.3662 | 0.12917 | 0.22946 | 0.96404 | 0.82159 | 0.5359268938 | 0.68923 | 0.4480015039 | 0.0712186 | 3.652314046 | 0 |
| 17 | 43049194 | T | G | D | A | BRCA1 | ENST00000471181 | 0.52241 | 0.1832 | 0.22979 | 0.98454 | 0.84906 | 0.4582082568 | 0.71386 | 0.8968839645 | 0.21293 | 6.494923171 | 0 |
| 17 | 43051065 | G | A | T | I | BRCA1 | ENST00000471181 | 0.6132 | 0.43875 | 0.27028 | 0.95662 | 0.87099 | 0.2968297729 | 0.83783 | 0.9386041164 | 0.302574 | 5.000303961 | 0 |
| 17 | 43051065 | G | C | T | R | BRCA1 | ENST00000471181 | 0.72828 | 0.13767 | 0.2081 | 0.93967 | 0.80118 | 0.5051874523 | 0.79003 | 0.8966028094 | 0.255904 | 6.28384365 | 0 |
| 17 | 43051065 | G | T | T | K | BRCA1 | ENST00000471181 | 0.44698 | 0.23798 | 0.22443 | 0.90435 | 0.79273 | 0.4868432638 | 0.75664 | 0.7279514074 | 0.227024 | 3.193773255 | 0 |
| 17 | 43051066 | T | A | T | S | BRCA1 | ENST00000471181 | 0.53421 | 0.15679 | 0.13694 | 0.89837 | 0.84267 | 0.193076527 | 0.85227 | 0.8943631649 | 0.208295 | 6.28384365 | 0 |
| 17 | 43051066 | T | C | T | A | BRCA1 | ENST00000471181 | 0.57202 | 0.14344 | 0.12504 | 0.901 | 0.86603 | 0.2338883923 | 0.85931 | 0.8542965055 | 0.249322 | 6.28384365 | 0 |
| 17 | 43051066 | T | G | T | P | BRCA1 | ENST00000471181 | 0.78788 | 0.13383 | 0.13849 | 0.96089 | 0.89193 | 0.6613901755 | 0.84505 | 0.8704865575 | 0.313962 | 5.090133346 | 0 |
| 17 | 43051068 | G | A | P | L | BRCA1 | ENST00000471181 | 0.81418 | 0.34819 | 0.38 | 0.95493 | 0.85996 | 0.4010410985 | 0.85046 | 0.778183639 | 0.223603 | 6.36194143 | 0 |
| 17 | 43051068 | G | C | P | R | BRCA1 | ENST00000471181 | 0.65226 | 0.45342 | 0.38184 | 0.94619 | 0.84321 | 0.3632392508 | 0.90293 | 0.984652698 | 0.39522 | 4.931577222 | 0 |
| 17 | 43051068 | G | T | P | H | BRCA1 | ENST00000471181 | 0.79603 | 0.46289 | 0.361 | 0.92343 | 0.82387 | 0.3616712946 | 0.91 | 0.984652698 | 0.354122 | 5.009707894 | 0 |
| 17 | 43051069 | G | A | P | S | BRCA1 | ENST00000471181 | 0.56352 | 0.40106 | 0.28694 | 0.90706 | 0.78782 | 0.2828660266 | 0.81372 | 0.87147367 | 0.182799 | 3.286649534 | 0 |
| 17 | 43051069 | G | C | P | A | BRCA1 | ENST00000471181 | 0.51783 | 0.1705 | 0.28708 | 0.91588 | 0.81703 | 0.2771792917 | 0.81658 | 0.462915201 | 0.239101 | 5.709255429 | 0 |
| 17 | 43051069 | G | T | P | T | BRCA1 | ENST00000471181 | 0.54406 | 0.4 | 0.32861 | 0.94985 | 0.77399 | 0.3278969926 | 0.8096 | 0.6680822968 | 0.327314 | 3.551367753 | 0 |
| 17 | 43051070 | C | T | M | I | BRCA1 | ENST00000471181 | 0.66011 | 0.25683 | 0.39511 | 0.94884 | 0.84694 | 0.6686151074 | 0.95408 | 0.9819744229 | 0.261262 | 6.541287201 | 0 |
| 17 | 43051071 | A | C | M | R | BRCA1 | ENST00000471181 | 0.81803 | 0.52177 | 0.45041 | 0.98402 | 0.90581 | 0.8749656723 | 0.96346 | 0.9386549592 | 0.241697 | 6.541287201 | 1 |
| 17 | 43051071 | A | G | M | T | BRCA1 | ENST00000471181 | 0.75224 | 0.47024 | 0.5228 | 0.98605 | 0.89513 | 0.7010316681 | 0.95803 | 0.9930410385 | 0.29835 | 6.541287201 | 0 |
| 17 | 43051071 | A | T | M | K | BRCA1 | ENST00000471181 | 0.80092 | 0.52568 | 0.47688 | 0.98475 | 0.90842 | 0.8696247969 | 0.96308 | 0.9909239411 | 0.436191 | 6.541287201 | 1 |
| 17 | 43051072 | T | A | M | L | BRCA1 | ENST00000471181 | 0.7282 | 0.11035 | 0.3927 | 0.93926 | 0.85326 | 0.5968304091 | 0.90487 | 0.973870635 | 0.242632 | 6.541287201 | 0 |
| 17 | 43051072 | T | C | M | V | BRCA1 | ENST00000471181 | 0.70061 | 0.11943 | 0.32277 | 0.96577 | 0.87877 | 0.6682912677 | 0.92048 | 0.9611440897 | 0.257496 | 6.541287201 | 1 |
| 17 | 43051072 | T | G | M | L | BRCA1 | ENST00000471181 | 0.73645 | 0.11035 | 0.3927 | 0.93869 | 0.85326 | 0.5968304091 | 0.90487 | 0.973870635 | 0.242633 | 6.541287201 | 0 |
| 17 | 43051073 | G | C | N | K | BRCA1 | ENST00000471181 | 0.5961 | 0.41975 | 0.3888 | 0.96652 | 0.83997 | 0.2333508262 | 0.72902 | 0.8920674324 | 0.101758 | 6.615269461 | 0 |
| 17 | 43051073 | G | T | N | K | BRCA1 | ENST00000471181 | 0.59017 | 0.41975 | 0.3888 | 0.96683 | 0.83997 | 0.2333508262 | 0.72902 | 0.9061606526 | 0.101755 | 6.615269461 | 0 |
| 17 | 43051074 | T | A | N | I | BRCA1 | ENST00000471181 | 0.851 | 0.28552 | 0.31026 | 0.97353 | 0.90668 | 0.364588523 | 0.75761 | 0.9771137834 | 0.289162 | 6.615269461 | 0 |
| 17 | 43051074 | T | C | N | S | BRCA1 | ENST00000471181 | 0.73013 | 0.12603 | 0.25586 | 0.93806 | 0.80358 | 0.1634484907 | 0.75673 | 0.7483670712 | 0.108082 | 6.615269461 | 0 |
| 17 | 43051074 | T | G | N | T | BRCA1 | ENST00000471181 | 0.80811 | 0.23204 | 0.30181 | 0.95725 | 0.86251 | 0.2549205077 | 0.76804 | 0.9093100429 | 0.143701 | 6.615269461 | 0 |
| 17 | 43051075 | T | A | N | Y | BRCA1 | ENST00000471181 | 0.81174 | 0.15447 | 0.3419 | 0.97067 | 0.88354 | 0.3517541805 | 0.73662 | 0.9205858111 | 0.286655 | 6.615269461 | 0 |
| 17 | 43051075 | T | C | N | D | BRCA1 | ENST00000471181 | 0.45579 | 0.317 | 0.35381 | 0.90073 | 0.65967 | 0.1793992871 | 0.69555 | 0.6337531805 | 0.0826287 | 3.318164578 | 0 |
| 17 | 43051075 | T | G | N | H | BRCA1 | ENST00000471181 | 0.76515 | 0.39459 | 0.32054 | 0.94595 | 0.82387 | 0.2748418376 | 0.85535 | 0.8630669713 | 0.146371 | 6.615269461 | 0 |
| 17 | 43051077 | G | A | T | I | BRCA1 | ENST00000471181 | 0.72885 | 0.37744 | 0.50551 | 0.97546 | 0.88212 | 0.4928549952 | 0.86781 | 0.9860658646 | 0.285232 | 3.758414384 | 0 |
| 17 | 43051077 | G | T | T | N | BRCA1 | ENST00000471181 | 0.67955 | 0.44134 | 0.45197 | 0.97501 | 0.87345 | 0.5270681525 | 0.88671 | 0.9838222861 | 0.150666 | 6.371745323 | 0 |
| 17 | 43051078 | T | A | T | S | BRCA1 | ENST00000471181 | 0.58297 | 0.26202 | 0.36426 | 0.97004 | 0.88496 | 0.2764434227 | 0.91897 | 0.896451056 | 0.200734 | 5.18221096 | 0 |
| 17 | 43051078 | T | G | T | P | BRCA1 | ENST00000471181 | 0.83306 | 0.4716 | 0.3649 | 0.98221 | 0.89239 | 0.6962133897 | 0.91875 | 0.9860658646 | 0.391832 | 6.371745323 | 0 |
| 17 | 43051079 | G | T | F | L | BRCA1 | ENST00000471181 | 0.61484 | 0.12275 | 0.55638 | 0.98796 | 0.87733 | 0.631877802 | 0.92457 | 0.9586142302 | 0.337719 | 6.541286461 | 0 |
| 17 | 43051080 | A | C | F | C | BRCA1 | ENST00000471181 | 0.89667 | 0.50617 | 0.54853 | 0.99207 | 0.95207 | 0.7714354758 | 0.88617 | 0.9952366352 | 0.510159 | 6.541286461 | 0 |
| 17 | 43051080 | A | G | F | S | BRCA1 | ENST00000471181 | 0.89679 | 0.50573 | 0.51823 | 0.99287 | 0.94913 | 0.769397349 | 0.90559 | 0.9957713485 | 0.509639 | 6.541286461 | 0 |
| 17 | 43051080 | A | T | F | Y | BRCA1 | ENST00000471181 | 0.82139 | 0.42198 | 0.53935 | 0.98654 | 0.88117 | 0.5658699839 | 0.96026 | 0.9890440702 | 0.331502 | 6.541286461 | 0 |
| 17 | 43051081 | A | G | F | L | BRCA1 | ENST00000471181 | 0.85705 | 0.12275 | 0.55638 | 0.98525 | 0.92723 | 0.631877802 | 0.96579 | 0.9808641076 | 0.373552 | 6.541286461 | 0 |
| 17 | 43051081 | A | T | F | I | BRCA1 | ENST00000471181 | 0.83966 | 0.36164 | 0.5116 | 0.98784 | 0.92233 | 0.7312468325 | 0.95398 | 0.9901183248 | 0.370564 | 6.541286461 | 0 |
| 17 | 43051083 | G | A | P | L | BRCA1 | ENST00000471181 | 0.7965 | 0.16913 | 0.37849 | 0.97039 | 0.90842 | 0.5749030801 | 0.84861 | 0.9918612242 | 0.350135 | 6.538263732 | 0 |
| 17 | 43051083 | G | C | P | R | BRCA1 | ENST00000471181 | 0.78019 | 0.42207 | 0.36747 | 0.97189 | 0.91862 | 0.5156635189 | 0.93673 | 0.9934025407 | 0.425275 | 6.538263732 | 0 |
| 17 | 43051083 | G | T | P | H | BRCA1 | ENST00000471181 | 0.77814 | 0.45675 | 0.35664 | 0.97416 | 0.911 | 0.4979310122 | 0.93769 | 0.9946420193 | 0.355715 | 6.538263732 | 0 |
| 17 | 43051084 | G | A | P | S | BRCA1 | ENST00000471181 | 0.69212 | 0.3651 | 0.28301 | 0.95973 | 0.8873 | 0.4890116096 | 0.94679 | 0.9790488482 | 0.266941 | 5.290440941 | 0 |
| 17 | 43051084 | G | C | P | A | BRCA1 | ENST00000471181 | 0.64298 | 0.19744 | 0.27897 | 0.95615 | 0.89603 | 0.385464888 | 0.94989 | 0.8710693717 | 0.230368 | 6.538263732 | 0 |
| 17 | 43051084 | G | T | P | T | BRCA1 | ENST00000471181 | 0.66597 | 0.4 | 0.32682 | 0.9694 | 0.89918 | 0.5039465719 | 0.93121 | 0.9857566953 | 0.336182 | 6.538263732 | 0 |
| 17 | 43051086 | C | A | G | V | BRCA1 | ENST00000471181 | 0.65893 | 0.40942 | 0.36809 | 0.98322 | 0.94876 | 0.833521207 | 0.93833 | 0.9770092368 | 0.420504 | 6.267926013 | 1 |
| 17 | 43051086 | C | G | G | A | BRCA1 | ENST00000471181 | 0.61865 | 0.17759 | 0.35659 | 0.95628 | 0.911 | 0.738062985 | 0.94034 | 0.8822761774 | 0.270759 | 6.267926013 | 0 |
| 17 | 43051086 | C | T | G | E | BRCA1 | ENST00000471181 | 0.50524 | 0.2253 | 0.44877 | 0.94063 | 0.88259 | 0.8321840533 | 0.91497 | 0.881978631 | 0.224685 | 3.174916089 | 0 |

| | | | | | | | | | | | | | | | | |
|---|---|---|---|---|---|---|---|---|---|---|---|---|---|---|---|---|
| 17 | 43051087 | C | A | G | W | BRCA1 | ENST00000471181 | 0.80858 | 0.47556 | 0.43937 | 0.96412 | 0.91058 | 0.7973742383 | 0.93306 | 0.9967247844 | 0.365969 | 6.267926013 | 1 |
| 17 | 43051087 | C | G | G | R | BRCA1 | ENST00000471181 | 0.74666 | 0.447 | 0.46 | 0.96166 | 0.91058 | 0.7969343501 | 0.94697 | 0.9726752043 | 0.403638 | 6.267926013 | 1 |
| 17 | 43051089 | T | A | Y | F | BRCA1 | ENST00000471181 | 0.41463 | 0.26863 | 0.12438 | 0.84857 | 0.85326 | 0.546825676 | 0.87855 | 0.7399432659 | 0.147868 | 3.92798934 | 0 |
| 17 | 43051089 | T | G | Y | S | BRCA1 | ENST00000471181 | 0.63812 | 0.25258 | 0.14234 | 0.92881 | 0.87974 | 0.71352442 | 0.90099 | 0.8457730412 | 0.299667 | 4.528420452 | 0 |
| 17 | 43051090 | A | C | Y | D | BRCA1 | ENST00000471181 | 0.83506 | 0.43136 | 0.20205 | 0.9182 | 0.94318 | 0.8322175858 | 0.94155 | 0.9479261637 | 0.391294 | 6.121695804 | 1 |
| 17 | 43051090 | A | T | Y | N | BRCA1 | ENST00000471181 | 0.82988 | 0.41592 | 0.21092 | 0.9362 | 0.92682 | 0.7405347755 | 0.95505 | 0.9466201663 | 0.318552 | 6.121695804 | 0 |
| 17 | 43051091 | G | C | C | W | BRCA1 | ENST00000471181 | 0.63222 | 0.50156 | 0.49155 | 0.99552 | 0.8543 | 0.7881842073 | 0.91623 | 0.9901560545 | 0.406461 | 6.541286629 | 1 |
| 17 | 43051092 | C | A | C | F | BRCA1 | ENST00000471181 | 0.73117 | 0.52322 | 0.4337 | 0.97606 | 0.89147 | 0.6735709171 | 0.97233 | 0.9926800728 | 0.425403 | 6.541286629 | 1 |
| 17 | 43051092 | C | G | C | S | BRCA1 | ENST00000471181 | 0.69364 | 0.45753 | 0.40818 | 0.98077 | 0.90798 | 0.7185255089 | 0.99503 | 0.9857566953 | 0.428985 | 6.541286629 | 0 |
| 17 | 43051092 | C | T | C | Y | BRCA1 | ENST00000471181 | 0.71321 | 0.54188 | 0.56125 | 0.98297 | 0.91862 | 0.7193743279 | 0.99537 | 0.995549202 | 0.432845 | 6.541286629 | 0 |
| 17 | 43051095 | C | A | C | F | BRCA1 | ENST00000471181 | 0.69444 | 0.51483 | 0.51047 | 0.97691 | 0.87588 | 0.7755815212 | 0.90146 | 0.9806365371 | 0.26005 | 6.251343891 | 0 |
| 17 | 43051095 | C | T | C | Y | BRCA1 | ENST00000471181 | 0.52632 | 0.53867 | 0.62887 | 0.94417 | 0.83228 | 0.821136778 | 0.9165 | 0.9581320286 | 0.207967 | 3.526155797 | 0 |
| 17 | 43051096 | A | T | C | S | BRCA1 | ENST00000471181 | 0.79934 | 0.38064 | 0.49414 | 0.97579 | 0.94765 | 0.7559528533 | 0.97226 | 0.9552016854 | 0.345787 | 6.251343891 | 0 |
| 17 | 43051097 | G | C | I | M | BRCA1 | ENST00000471181 | 0.54399 | 0.41237 | 0.26682 | 0.96032 | 0.78035 | 0.6367690944 | 0.97898 | 0.7862426043 | 0.234316 | 5.830593513 | 0 |
| 17 | 43051098 | A | C | I | S | BRCA1 | ENST00000471181 | 0.90422 | 0.50872 | 0.31921 | 0.97608 | 0.92397 | 0.9206834548 | 0.9978 | 0.9796691537 | 0.388074 | 5.830593513 | 1 |
| 17 | 43051098 | A | G | I | T | BRCA1 | ENST00000471181 | 0.85152 | 0.48171 | 0.40683 | 0.97103 | 0.90842 | 0.7686198777 | 0.9974 | 0.9608945847 | 0.339874 | 5.830593513 | 1 |
| 17 | 43051098 | A | T | I | N | BRCA1 | ENST00000471181 | 0.89911 | 0.44913 | 0.37039 | 0.98072 | 0.9014 | 0.8949965095 | 0.99785 | 0.9824072123 | 0.277107 | 5.830593513 | 1 |
| 17 | 43051099 | T | A | I | F | BRCA1 | ENST00000471181 | 0.68275 | 0.42644 | 0.31325 | 0.94966 | 0.7562 | 0.7544445394 | 0.99274 | 0.8578502536 | 0.218765 | 5.830593513 | 0 |
| 17 | 43051099 | T | C | I | V | BRCA1 | ENST00000471181 | 0.29352 | 0.08109 | 0.22418 | 0.76351 | 0.72775 | 0.3265482574 | 0.77595 | 0.178227514 | 0.07635 | 2.366339514 | 0 |
| 17 | 43051099 | T | G | I | L | BRCA1 | ENST00000471181 | 0.63819 | 0.09929 | 0.29335 | 0.91234 | 0.80772 | 0.6020540517 | 0.90351 | 0.7663351297 | 0.177281 | 3.758912542 | 0 |
| 17 | 43051100 | T | A | E | D | BRCA1 | ENST00000471181 | 0.51619 | 0.16971 | 0.16572 | 0.87051 | 0.78098 | 0.5186055764 | 0.87505 | 0.661581099 | 0.167388 | 3.63812632 | 0 |
| 17 | 43051100 | T | G | E | D | BRCA1 | ENST00000471181 | 0.523 | 0.16971 | 0.16572 | 0.86663 | 0.78098 | 0.5186055764 | 0.87505 | 0.661581099 | 0.167391 | 3.63812632 | 0 |
| 17 | 43051101 | T | A | E | V | BRCA1 | ENST00000471181 | 0.76499 | 0.26575 | 0.146 | 0.97549 | 0.85996 | 0.5529637015 | 0.90946 | 0.9491487741 | 0.31606 | 5.73481829 | 0 |
| 17 | 43051101 | T | G | E | A | BRCA1 | ENST00000471181 | 0.72877 | 0.29633 | 0.16399 | 0.9636 | 0.88543 | 0.5216079098 | 0.92704 | 0.8947607279 | 0.232525 | 4.19726238 | 0 |
| 17 | 43051102 | C | G | E | Q | BRCA1 | ENST00000471181 | 0.35619 | 0.31978 | 0.16554 | 0.88415 | 0.78658 | 0.4674975218 | 0.87494 | 0.733351449 | 0.161447 | 4.093915144 | 0 |
| 17 | 43051102 | C | T | E | K | BRCA1 | ENST00000471181 | 0.38386 | 0.12343 | 0.2352 | 0.93493 | 0.83283 | 0.569739617 | 0.88219 | 0.797532022 | 0.182384 | 2.687363737 | 0 |
| 17 | 43051104 | A | G | L | P | BRCA1 | ENST00000471181 | 0.88861 | 0.53095 | 0.46523 | 0.98862 | 0.88212 | 0.8263812286 | 0.98791 | 0.9942643046 | 0.436743 | 6.597026022 | 1 |
| 17 | 43051104 | A | T | L | Q | BRCA1 | ENST00000471181 | 0.87768 | 0.45017 | 0.34347 | 0.9848 | 0.90537 | 0.6984605333 | 0.98582 | 0.9948493838 | 0.381783 | 6.597026022 | 1 |
| 17 | 43051105 | G | C | L | V | BRCA1 | ENST00000471181 | 0.61761 | 0.27649 | 0.27255 | 0.95662 | 0.82948 | 0.4116530025 | 0.91027 | 0.927449584 | 0.241364 | 6.597026022 | 0 |
| 17 | 43051105 | G | T | L | I | BRCA1 | ENST00000471181 | 0.62521 | 0.34604 | 0.34798 | 0.95275 | 0.79758 | 0.354352761 | 0.87871 | 0.9471259117 | 0.150408 | 6.180825885 | 0 |
| 17 | 43051107 | C | A | G | V | BRCA1 | ENST00000471181 | 0.70133 | 0.27215 | 0.27927 | 0.98568 | 0.91778 | 0.8007763311 | 0.93724 | 0.993391037 | 0.343104 | 6.638585512 | 1 |
| 17 | 43051107 | C | G | G | A | BRCA1 | ENST00000471181 | 0.67136 | 0.22374 | 0.2864 | 0.95959 | 0.90318 | 0.6564252769 | 0.94062 | 0.9638263583 | 0.267312 | 6.638585512 | 0 |
| 17 | 43051108 | C | G | G | R | BRCA1 | ENST00000471181 | 0.76403 | 0.46621 | 0.37164 | 0.97431 | 0.8705 | 0.6875546865 | 0.94511 | 0.9914570451 | 0.281027 | 6.638585512 | 0 |
| 17 | 43051109 | C | A | R | S | BRCA1 | ENST00000471181 | 0.11534 | 0.10659 | 0.04128 | 0.90676 | 0.79637 | 0.4512042811 | 0.34646 | 0.4103640914 | 0.184467 | 3.701040408 | 0 |
| 17 | 43051109 | C | G | R | S | BRCA1 | ENST00000471181 | 0.11866 | 0.10659 | 0.04128 | 0.90545 | 0.79637 | 0.4512042811 | 0.34646 | 0.2229092866 | 0.184471 | 3.701040408 | 0 |
| 17 | 43051110 | C | A | R | M | BRCA1 | ENST00000471181 | 0.39487 | 0.30458 | 0.03923 | 0.94629 | 0.83283 | 0.540468278 | 0.62306 | 0.8174147606 | 0.16654 | 4.209074542 | 0 |
| 17 | 43051110 | C | G | R | T | BRCA1 | ENST00000471181 | 0.34565 | 0.16836 | 0.05887 | 0.88795 | 0.81761 | 0.4839225927 | 0.40795 | 0.1410006234 | 0.123311 | 4.208194808 | 0 |
| 17 | 43051110 | C | T | R | K | BRCA1 | ENST00000471181 | 0.11051 | 0.09225 | 0.0415 | 0.63776 | 0.79089 | 0.4180865407 | 0.29733 | 0.02499362408 | 0.0798587 | 0.9651976022 | 0 |
| 17 | 43051111 | T | A | R | W | BRCA1 | ENST00000471181 | 0.60683 | 0.41201 | 0.0608 | 0.93346 | 0.82836 | 0.6071548291 | 0.85967 | 0.9229151011 | 0.234878 | 5.33572743 | 0 |
| 17 | 43051111 | T | C | R | G | BRCA1 | ENST00000471181 | 0.27279 | 0.11892 | 0.04145 | 0.87917 | 0.80713 | 0.5639802537 | 0.57971 | 0.1764542942 | 0.2535 | 4.868971534 | 0 |
| 17 | 43051112 | G | C | F | L | BRCA1 | ENST00000471181 | 0.66699 | 0.1365 | 0.45208 | 0.94028 | 0.90318 | 0.7594064497 | 0.94851 | 0.9195697904 | 0.419631 | 5.290440915 | 0 |
| 17 | 43051112 | G | T | F | L | BRCA1 | ENST00000471181 | 0.66059 | 0.1365 | 0.45208 | 0.94077 | 0.90318 | 0.7594064497 | 0.94851 | 0.9195697904 | 0.419629 | 5.290440915 | 0 |
| 17 | 43051113 | A | C | F | C | BRCA1 | ENST00000471181 | 0.88628 | 0.14172 | 0.44677 | 0.986 | 0.9528 | 0.8983705093 | 0.95177 | 0.9926835895 | 0.490468 | 6.538263705 | 1 |
| 17 | 43051113 | A | G | F | S | BRCA1 | ENST00000471181 | 0.88635 | 0.2394 | 0.41574 | 0.98515 | 0.96226 | 0.9236490746 | 0.95518 | 0.9827687144 | 0.491052 | 6.538263705 | 1 |
| 17 | 43051114 | A | C | F | V | BRCA1 | ENST00000471181 | 0.87623 | 0.18736 | 0.389 | 0.98585 | 0.96517 | 0.8481082165 | 0.97967 | 0.9594798088 | 0.488522 | 6.538263705 | 1 |
| 17 | 43051114 | A | G | F | L | BRCA1 | ENST00000471181 | 0.8869 | 0.1365 | 0.45208 | 0.97158 | 0.93515 | 0.7594064497 | 0.98544 | 0.9656034112 | 0.449737 | 5.290440915 | 0 |
| 17 | 43051114 | A | T | F | I | BRCA1 | ENST00000471181 | 0.87187 | 0.19898 | 0.41245 | 0.97943 | 0.9528 | 0.8477690222 | 0.98785 | 0.9819489121 | 0.462518 | 6.538263705 | 1 |
| 17 | 43051115 | G | C | I | M | BRCA1 | ENST00000471181 | 0.68131 | 0.4231 | 0.39946 | 0.94129 | 0.86902 | 0.5405893804 | 0.94737 | 0.8435865045 | 0.206659 | 3.890714928 | 0 |
| 17 | 43051116 | A | C | I | S | BRCA1 | ENST00000471181 | 0.9351 | 0.48603 | 0.45022 | 0.97794 | 0.90624 | 0.8539206948 | 0.98166 | 0.9935699701 | 0.296774 | 6.078703977 | 1 |
| 17 | 43051116 | A | G | I | T | BRCA1 | ENST00000471181 | 0.87569 | 0.4426 | 0.53539 | 0.9731 | 0.91399 | 0.7209668241 | 0.9797 | 0.9771898389 | 0.288735 | 6.078703977 | 1 |
| 17 | 43051116 | A | T | I | N | BRCA1 | ENST00000471181 | 0.93357 | 0.53254 | 0.50734 | 0.98072 | 0.91271 | 0.8619998026 | 0.98519 | 0.9939052463 | 0.271308 | 6.078703977 | 1 |
| 17 | 43051117 | T | C | I | V | BRCA1 | ENST00000471181 | 0.49139 | 0.13654 | 0.3621 | 0.91015 | 0.81298 | 0.4264963186 | 0.74152 | 0.9037949443 | 0.171429 | 6.078703977 | 0 |
| 17 | 43051117 | T | G | I | L | BRCA1 | ENST00000471181 | 0.39452 | 0.10555 | 0.43131 | 0.80353 | 0.71627 | 0.5278119674 | 0.64847 | 0.7662667036 | 0.110618 | 2.755744482 | 0 |
| 17 | 43057052 | C | A | K | N | BRCA1 | ENST00000471181 | 0.85152 | 0.16857 | 0.32142 | 0.96729 | 0.825 | 0.6654171014 | 0.89556 | 0.9616745114 | 0.170069 | 6.379106282 | 0 |
| 17 | 43057052 | C | G | K | N | BRCA1 | ENST00000471181 | 0.85974 | 0.16857 | 0.32142 | 0.96675 | 0.82556 | 0.6654171014 | 0.89556 | 0.9594130516 | 0.170084 | 6.379106282 | 0 |
| 17 | 43057053 | T | A | K | M | BRCA1 | ENST00000471181 | 0.92257 | 0.25525 | 0.24529 | 0.98351 | 0.92275 | 0.7128489921 | 0.90305 | 0.9773367047 | 0.221018 | 5.151595955 | 0 |
| 17 | 43057053 | T | C | K | R | BRCA1 | ENST00000471181 | 0.90047 | 0.08843 | 0.24401 | 0.96379 | 0.88916 | 0.48612633 | 0.85238 | 0.9208989739 | 0.182986 | 6.379106282 | 0 |
| 17 | 43057053 | T | G | K | T | BRCA1 | ENST00000471181 | 0.90344 | 0.19324 | 0.30628 | 0.97806 | 0.92397 | 0.7078025552 | 0.88542 | 0.9520997405 | 0.23625 | 4.339383334 | 0 |
| 17 | 43057054 | T | C | K | E | BRCA1 | ENST00000471181 | 0.7015 | 0.13719 | 0.29626 | 0.96983 | 0.8933 | 0.6919596686 | 0.8378 | 0.9483684301 | 0.262956 | 6.379106282 | 0 |
| 17 | 43057054 | T | G | K | Q | BRCA1 | ENST00000471181 | 0.54136 | 0.09821 | 0.265 | 0.96605 | 0.86251 | 0.6146728362 | 0.83433 | 0.7659621835 | 0.219109 | 4.823681583 | 0 |
| 17 | 43057055 | T | A | R | S | BRCA1 | ENST00000471181 | 0.43412 | 0.28511 | 0.08003 | 0.91353 | 0.80536 | 0.4347867843 | 0.76726 | 0.5764744996 | 0.195039 | 5.594738152 | 0 |
| 17 | 43057055 | T | G | R | S | BRCA1 | ENST00000471181 | 0.44035 | 0.28511 | 0.08003 | 0.91384 | 0.80536 | 0.4347867843 | 0.76726 | 0.5719499209 | 0.195044 | 5.594738152 | 0 |
| 17 | 43057056 | C | A | R | I | BRCA1 | ENST00000471181 | 0.3958 | 0.1748 | 0.10514 | 0.92974 | 0.82668 | 0.5038784466 | 0.84173 | 0.6883405447 | 0.170554 | 5.594738152 | 0 |
| 17 | 43057056 | C | G | R | T | BRCA1 | ENST00000471181 | 0.35808 | 0.35931 | 0.1021 | 0.91829 | 0.81472 | 0.4508733081 | 0.86338 | 0.6207020879 | 0.163506 | 5.594738152 | 0 |
| 17 | 43057056 | C | T | R | K | BRCA1 | ENST00000471181 | 0.17091 | 0.09707 | 0.10751 | 0.64095 | 0.71861 | 0.4225402998 | 0.61965 | 0.1066146269 | 0.104715 | 3.175662077 | 0 |
| 17 | 43057057 | T | C | R | G | BRCA1 | ENST00000471181 | 0.28271 | 0.33584 | 0.0833 | 0.90307 | 0.8199 | 0.5081748028 | 0.88049 | 0.6806866527 | 0.199629 | 2.355170766 | 0 |
| 17 | 43057058 | G | C | D | E | BRCA1 | ENST00000471181 | 0.24003 | 0.08473 | 0.14223 | 0.7489 | 0.73734 | 0.268735155 | 0.66278 | 0.1936408877 | 0.118092 | 2.447502443 | 0 |
| 17 | 43057058 | G | T | D | E | BRCA1 | ENST00000471181 | 0.23652 | 0.08473 | 0.14223 | 0.7489 | 0.72626 | 0.268735155 | 0.66278 | 0.1936408877 | 0.118093 | 2.447502443 | 0 |
| 17 | 43057059 | T | A | D | V | BRCA1 | ENST00000471181 | 0.44576 | 0.13311 | 0.11853 | 0.95626 | 0.79577 | 0.6162862643 | 0.68047 | 0.6520062685 | 0.187921 | 5.349204731 | 0 |
| 17 | 43057059 | T | C | D | G | BRCA1 | ENST00000471181 | 0.24068 | 0.12074 | 0.13477 | 0.77213 | 0.72626 | 0.5252012409 | 0.45291 | 0.05228244513 | 0.107157 | 1.576018953 | 0 |
| 17 | 43057059 | T | G | D | A | BRCA1 | ENST00000471181 | 0.23797 | 0.11836 | 0.12526 | 0.86871 | 0.74446 | 0.5071162371 | 0.49342 | 0.258038789 | 0.124333 | 3.603841054 | 0 |
| 17 | 43057060 | C | A | D | Y | BRCA1 | ENST00000471181 | 0.50877 | 0.2794 | 0.16615 | 0.95071 | 0.8306 | 0.6097770957 | 0.86767 | 0.8423552513 | 0.19384 | 5.349204731 | 0 |
| 17 | 43057060 | C | G | D | H | BRCA1 | ENST00000471181 | 0.49113 | 0.21979 | 0.18158 | 0.93292 | 0.82387 | 0.5293828431 | 0.86012 | 0.5467602611 | 0.172976 | 5.349204731 | 0 |
| 17 | 43057060 | C | T | D | N | BRCA1 | ENST00000471181 | 0.25985 | 0.08892 | 0.1593 | 0.89025 | 0.73949 | 0.3411292491 | 0.78787 | 0.5985450149 | 0.0750286 | 2.976330942 | 0 |
| 17 | 43057061 | C | A | Q | H | BRCA1 | ENST00000471181 | 0.30198 | 0.12399 | 0.02302 | 0.94815 | 0.83228 | 0.4617684548 | 0.67564 | 0.42752707 | 0.200998 | 3.85991306 | 0 |
| 17 | 43057061 | C | G | Q | H | BRCA1 | ENST00000471181 | 0.3074 | 0.12399 | 0.02302 | 0.94825 | 0.83228 | 0.4617684548 | 0.67564 | 0.4623464048 | 0.200996 | 3.85991306 | 0 |
| 17 | 43057062 | T | A | Q | L | BRCA1 | ENST00000471181 | 0.29854 | 0.08898 | 0.02278 | 0.96387 | 0.80831 | 0.4947714687 | 0.32431 | 0.09125710279 | 0.128264 | 3.270167923 | 0 |
| 17 | 43057062 | T | C | Q | R | BRCA1 | ENST00000471181 | 0.1894 | 0.09792 | 0.02482 | 0.81065 | 0.80536 | 0.3451329708 | 0.27821 | 0.02399974689 | 0.133758 | 2.565982882 | 0 |
| 17 | 43057062 | T | G | Q | P | BRCA1 | ENST00000471181 | 0.20276 | 0.11127 | 0.02233 | 0.89374 | 0.79998 | 0.7164981234 | 0.31517 | 0.04256455228 | 0.171613 | 3.237780109 | 0 |
| 17 | 43057063 | G | C | Q | E | BRCA1 | ENST00000471181 | 0.42282 | 0.10264 | 0.01986 | 0.91137 | 0.79637 | 0.4041471691 | 0.86181 | 0.2240182073 | 0.187196 | 3.886996392 | 0 |
| 17 | 43057063 | G | T | Q | K | BRCA1 | ENST00000471181 | 0.35625 | 0.11956 | 0.02642 | 0.90192 | 0.80536 | 0.4312151839 | 0.85441 | 0.1984506689 | 0.21118 | 3.864108996 | 0 |
| 17 | 43057065 | G | A | S | F | BRCA1 | ENST00000471181 | 0.62916 | 0.45205 | 0.19093 | 0.94261 | 0.83172 | 0.5687619424 | 0.85385 | 0.7845430374 | 0.223505 | 3.097034263 | 0 |
| 17 | 43057065 | G | C | S | C | BRCA1 | ENST00000471181 | 0.79911 | 0.42141 | 0.17255 | 0.95948 | 0.88259 | 0.5262578511 | 0.91135 | 0.8420654535 | 0.245056 | 5.86899923 | 0 |
| 17 | 43057065 | G | T | S | Y | BRCA1 | ENST00000471181 | 0.76626 | 0.48227 | 0.20969 | 0.96616 | 0.88165 | 0.5846305258 | 0.89031 | 0.8222616985 | 0.252422 | 5.392433169 | 0 |
| 17 | 43057066 | A | C | S | A | BRCA1 | ENST00000471181 | 0.71089 | 0.08366 | 0.15796 | 0.93298 | 0.82892 | 0.3316078732 | 0.85935 | 0.7068371773 | 0.204663 | 4.093420507 | 0 |
| 17 | 43057066 | A | G | S | P | BRCA1 | ENST00000471181 | 0.7783 | 0.44012 | 0.18373 | 0.96544 | 0.87 | 0.77767823 | 0.90155 | 0.9287512898 | 0.288803 | 5.86899923 | 0 |
| 17 | 43057066 | A | T | S | T | BRCA1 | ENST00000471181 | 0.67136 | 0.09187 | 0.17256 | 0.93343 | 0.80476 | 0.4191841885 | 0.86796 | 0.8610393405 | 0.222709 | 5.86899923 | 0 |
| 17 | 43057067 | T | A | E | D | BRCA1 | ENST00000471181 | 0.68915 | 0.15305 | 0.24273 | 0.9341 | 0.85792 | 0.5011476367 | 0.79442 | 0.9055006504 | 0.252355 | 6.569213729 | 0 |
| 17 | 43057067 | T | G | E | D | BRCA1 | ENST00000471181 | 0.6974 | 0.15305 | 0.24273 | 0.93195 | 0.85792 | 0.5011476367 | 0.79442 | 0.8730254173 | 0.252331 | 6.569213729 | 0 |
| 17 | 43057068 | T | A | E | V | BRCA1 | ENST00000471181 | 0.91078 | 0.12844 | 0.22431 | 0.97644 | 0.92028 | 0.5553418077 | 0.85387 | 0.9576286674 | 0.399273 | 6.569213729 | 0 |
| 17 | 43057068 | T | C | E | G | BRCA1 | ENST00000471181 | 0.92194 | 0.11719 | 0.21342 | 0.97226 | 0.91526 | 0.5281465485 | 0.86992 | 0.9525215626 | 0.327574 | 6.569213729 | 0 |
| 17 | 43057068 | T | G | E | A | BRCA1 | ENST00000471181 | 0.89243 | 0.242 | 0.24237 | 0.97106 | 0.90052 | 0.520024957 | 0.84748 | 0.9527734518 | 0.288456 | 6.569213729 | 0 |
| 17 | 43057069 | C | G | E | Q | BRCA1 | ENST00000471181 | 0.64267 | 0.29688 | 0.23479 | 0.95752 | 0.80772 | 0.4646924172 | 0.82624 | 0.2608781212 | 0.0627644 | 3.730234159 | 0 |
| 17 | 43057069 | C | T | E | K | BRCA1 | ENST00000471181 | 0.84249 | 0.21697 | 0.32217 | 0.9561 | 0.84267 | 0.5274900006 | 0.83754 | 0.9526699781 | 0.361523 | 5.902643601 | 0 |
| 17 | 43057070 | T | A | R | S | BRCA1 | ENST00000471181 | 0.80487 | 0.42399 | 0.24995 | 0.98558 | 0.89739 | 0.8801594868 | 0.79165 | 0.9641472101 | 0.40148 | 6.541287772 | 1 |
| 17 | 43057070 | T | G | R | S | BRCA1 | ENST00000471181 | 0.81292 | 0.42399 | 0.24995 | 0.986 | 0.89739 | 0.8801594868 | 0.79165 | 0.9561644793 | 0.401482 | 6.541287772 | 1 |
| 17 | 43057071 | C | A | R | I | BRCA1 | ENST00000471181 | 0.89451 | 0.44831 | 0.28223 | 0.97457 | 0.92275 | 0.8086616529 | 0.87304 | 0.952780664 | 0.417875 | 6.541287772 | 1 |
| 17 | 43057071 | C | G | R | T | BRCA1 | ENST00000471181 | 0.85981 | 0.48279 | 0.28389 | 0.97613 | 0.91652 | 0.8733795107 | 0.87502 | 0.9243597388 | 0.41308 | 6.541287772 | 1 |
| 17 | 43057071 | C | T | R | K | BRCA1 | ENST00000471181 | 0.85836 | 0.38714 | 0.30218 | 0.96433 | 0.89963 | 0.8059516097 | 0.84831 | 0.9727433324 | 0.416105 | 6.541287772 | 1 |
| 17 | 43057072 | T | C | R | G | BRCA1 | ENST00000471181 | 0.85397 | 0.4579 | 0.24462 | 0.98116 | 0.94728 | 0.8742499503 | 0.83858 | 0.9772220254 | 0.415236 | 6.541287772 | 1 |
| 17 | 43057074 | G | A | A | V | BRCA1 | ENST00000471181 | 0.88727 | 0.09965 | 0.21499 | 0.92289 | 0.87247 | 0.6992790836 | 0.93762 | 0.9348021746 | 0.250104 | 6.118537494 | 1 |

| | | | | | | | | | | | | | | | | | | |
|---|---|---|---|---|---|---|---|---|---|---|---|---|---|---|---|---|---|---|
| 17 | 43057074 | G | C | A | G | BRCA1 | ENST00000471181 | 0.8665 | 0.09416 | 0.18671 | 0.9122 | 0.86302 | 0.5677910432 | 0.91328 | 0.8950039744 | 0.218846 | 6.118537494 | 0 |
| 17 | 43057074 | G | T | A | E | BRCA1 | ENST00000471181 | 0.88145 | 0.10632 | 0.2508 | 0.95305 | 0.88212 | 0.8776424584 | 0.94224 | 0.9361892939 | 0.287205 | 6.118537494 | 1 |
| 17 | 43057075 | C | A | A | S | BRCA1 | ENST00000471181 | 0.49997 | 0.08394 | 0.17305 | 0.8958 | 0.81182 | 0.4912644269 | 0.8183 | 0.7152122855 | 0.142155 | 2.584136916 | 0 |
| 17 | 43057075 | C | G | A | P | BRCA1 | ENST00000471181 | 0.72676 | 0.14551 | 0.2546 | 0.95551 | 0.8873 | 0.9064089238 | 0.91563 | 0.8732268214 | 0.394069 | 6.118537494 | 1 |
| 17 | 43057075 | C | T | A | T | BRCA1 | ENST00000471181 | 0.70519 | 0.09363 | 0.19971 | 0.94425 | 0.88776 | 0.5763821581 | 0.89557 | 0.8878396153 | 0.236051 | 6.118537494 | 1 |
| 17 | 43057077 | C | A | R | L | BRCA1 | ENST00000471181 | 0.82685 | 0.11407 | 0.32824 | 0.95533 | 0.90537 | 0.8110614072 | 0.86731 | 0.9588401914 | 0.419473 | 6.559579587 | 0 |
| 17 | 43057077 | C | G | R | P | BRCA1 | ENST00000471181 | 0.84696 | 0.22139 | 0.34134 | 0.95944 | 0.90185 | 0.931025722 | 0.8574 | 0.9588401914 | 0.420063 | 6.559579587 | 1 |
| 17 | 43057077 | C | T | R | Q | BRCA1 | ENST00000471181 | 0.83498 | 0.11092 | 0.25 | 0.94406 | 0.88543 | 0.6880949793 | 0.8398 | 0.2221175779 | 0.282686 | 6.559579587 | 0 |
| 17 | 43057078 | G | C | R | G | BRCA1 | ENST00000471181 | 0.78685 | 0.1181 | 0.30121 | 0.96438 | 0.89963 | 0.8115317293 | 0.81509 | 0.8901241422 | 0.413828 | 6.559579587 | 1 |
| 17 | 43057079 | C | A | K | N | BRCA1 | ENST00000471181 | 0.5702 | 0.10668 | 0.14253 | 0.94477 | 0.8306 | 0.6385820649 | 0.74601 | 0.9194481373 | 0.10077 | 5.895130245 | 0 |
| 17 | 43057080 | T | A | K | M | BRCA1 | ENST00000471181 | 0.65972 | 0.16234 | 0.09369 | 0.94898 | 0.86503 | 0.6348648909 | 0.70101 | 0.6408944093 | 0.194439 | 5.895130245 | 0 |
| 17 | 43057080 | T | C | K | R | BRCA1 | ENST00000471181 | 0.32094 | 0.07473 | 0.08339 | 0.66755 | 0.76487 | 0.4480487312 | 0.37235 | 0.1404971778 | 0.108196 | 1.894576534 | 0 |
| 17 | 43057080 | T | G | K | T | BRCA1 | ENST00000471181 | 0.62681 | 0.11741 | 0.12915 | 0.94587 | 0.86403 | 0.6325471018 | 0.70033 | 0.7281394601 | 0.182823 | 5.895130245 | 0 |
| 17 | 43057081 | T | C | K | E | BRCA1 | ENST00000471181 | 0.65736 | 0.13156 | 0.12412 | 0.82403 | 0.78286 | 0.5878850343 | 0.70898 | 0.4486021583 | 0.207272 | 3.511845666 | 0 |
| 17 | 43057081 | T | G | K | Q | BRCA1 | ENST00000471181 | 0.76171 | 0.09744 | 0.09077 | 0.89749 | 0.82443 | 0.5456121725 | 0.74771 | 0.7819780111 | 0.198696 | 5.895130245 | 0 |
| 17 | 43057083 | G | A | P | L | BRCA1 | ENST00000471181 | 0.84816 | 0.40026 | 0.35454 | 0.97789 | 0.9014 | 0.8992464848 | 0.9075 | 0.9950510859 | 0.284213 | 6.541287067 | 1 |
| 17 | 43057083 | G | C | P | R | BRCA1 | ENST00000471181 | 0.83244 | 0.12188 | 0.34511 | 0.97106 | 0.94167 | 0.9182466152 | 0.91112 | 0.9865800738 | 0.426096 | 6.541287067 | 1 |
| 17 | 43057083 | G | T | P | Q | BRCA1 | ENST00000471181 | 0.81912 | 0.23183 | 0.28752 | 0.97473 | 0.92561 | 0.8438767495 | 0.92813 | 0.993062079 | 0.374433 | 6.541287067 | 1 |
| 17 | 43057084 | G | A | P | S | BRCA1 | ENST00000471181 | 0.82342 | 0.15049 | 0.27831 | 0.97116 | 0.90052 | 0.859479001 | 0.93154 | 0.9757707119 | 0.27327 | 6.541287067 | 1 |
| 17 | 43057084 | G | C | P | A | BRCA1 | ENST00000471181 | 0.77458 | 0.08873 | 0.26711 | 0.96359 | 0.92316 | 0.7786595677 | 0.93069 | 0.8986198306 | 0.280894 | 6.541287067 | 0 |
| 17 | 43057084 | G | T | P | T | BRCA1 | ENST00000471181 | 0.79761 | 0.20992 | 0.30651 | 0.97384 | 0.9182 | 0.8408291369 | 0.93348 | 0.9757707119 | 0.336639 | 6.541287067 | 0 |
| 17 | 43057086 | C | G | G | A | BRCA1 | ENST00000471181 | 0.70286 | 0.1307 | 0.36129 | 0.96473 | 0.88963 | 0.7612716812 | 0.89849 | 0.9635987282 | 0.287109 | 6.541286482 | 0 |
| 17 | 43057086 | C | T | G | D | BRCA1 | ENST00000471181 | 0.73509 | 0.30423 | 0.48458 | 0.97617 | 0.90755 | 0.9132658661 | 0.90584 | 0.9908012748 | 0.419193 | 6.541286482 | 1 |
| 17 | 43057087 | C | A | G | C | BRCA1 | ENST00000471181 | 0.8356 | 0.40301 | 0.4074 | 0.97654 | 0.91015 | 0.8338924052 | 0.91998 | 0.9941008687 | 0.371518 | 6.541286482 | 1 |
| 17 | 43057087 | C | G | G | R | BRCA1 | ENST00000471181 | 0.80858 | 0.13319 | 0.47825 | 0.97338 | 0.89467 | 0.8804340123 | 0.91879 | 0.9850801826 | 0.26802 | 6.541286482 | 1 |
| 17 | 43057087 | C | T | G | S | BRCA1 | ENST00000471181 | 0.77877 | 0.09851 | 0.3611 | 0.96886 | 0.90929 | 0.7370936006 | 0.90009 | 0.9832825065 | 0.408152 | 6.541286482 | 0 |
| 17 | 43057088 | T | A | Q | H | BRCA1 | ENST00000471181 | 0.45107 | 0.09619 | 0.09071 | 0.91878 | 0.86352 | 0.6856787824 | 0.72626 | 0.8232312799 | 0.329975 | 4.411429706 | 0 |
| 17 | 43057088 | T | G | Q | H | BRCA1 | ENST00000471181 | 0.4574 | 0.09619 | 0.09071 | 0.91532 | 0.86352 | 0.6856787824 | 0.72626 | 0.8232312799 | 0.329975 | 4.411429706 | 0 |
| 17 | 43057089 | T | A | Q | L | BRCA1 | ENST00000471181 | 0.75193 | 0.08873 | 0.06239 | 0.94356 | 0.86951 | 0.6864703002 | 0.80181 | 0.905845046 | 0.248185 | 6.270540908 | 0 |
| 17 | 43057089 | T | C | Q | R | BRCA1 | ENST00000471181 | 0.41831 | 0.0997 | 0.09089 | 0.77277 | 0.77654 | 0.6230218701 | 0.71134 | 0.447504878 | 0.206294 | 3.385902462 | 0 |
| 17 | 43057089 | T | G | Q | P | BRCA1 | ENST00000471181 | 0.75041 | 0.11127 | 0.0727 | 0.92857 | 0.87296 | 0.8753022776 | 0.78939 | 0.8700632453 | 0.311917 | 6.270540908 | 1 |
| 17 | 43057090 | G | C | Q | E | BRCA1 | ENST00000471181 | 0.59255 | 0.09904 | 0.06673 | 0.91508 | 0.82724 | 0.6612386528 | 0.76103 | 0.8568902016 | 0.231311 | 4.945809968 | 0 |
| 17 | 43057090 | G | T | Q | K | BRCA1 | ENST00000471181 | 0.76164 | 0.1161 | 0.10194 | 0.85758 | 0.81933 | 0.6673560768 | 0.75688 | 0.8528017998 | 0.242959 | 6.270540908 | 0 |
| 17 | 43057092 | T | A | H | L | BRCA1 | ENST00000471181 | 0.79034 | 0.11683 | 0.37543 | 0.97335 | 0.95207 | 0.8161464167 | 0.87691 | 0.9504985809 | 0.357702 | 6.541286976 | 0 |
| 17 | 43057092 | T | C | H | R | BRCA1 | ENST00000471181 | 0.74308 | 0.17289 | 0.42025 | 0.9574 | 0.93786 | 0.7486943476 | 0.87368 | 0.950999856 | 0.360662 | 6.541286976 | 0 |
| 17 | 43057092 | T | G | H | P | BRCA1 | ENST00000471181 | 0.78867 | 0.14537 | 0.34836 | 0.97376 | 0.97141 | 0.9091481753 | 0.87006 | 0.8818798065 | 0.418409 | 6.541286976 | 1 |
| 17 | 43057093 | G | A | H | Y | BRCA1 | ENST00000471181 | 0.83591 | 0.22433 | 0.42087 | 0.94217 | 0.88916 | 0.7420785498 | 0.89205 | 0.968919754 | 0.395163 | 6.541286976 | 1 |
| 17 | 43057093 | G | C | H | D | BRCA1 | ENST00000471181 | 0.87788 | 0.25013 | 0.41411 | 0.95609 | 0.94015 | 0.8599119072 | 0.8887 | 0.9752618074 | 0.425301 | 6.541286976 | 1 |
| 17 | 43057094 | G | C | N | K | BRCA1 | ENST00000471181 | 0.71658 | 0.18368 | 0.3911 | 0.92675 | 0.848 | 0.7466990762 | 0.75002 | 0.8003519177 | 0.112057 | 6.3643411 | 0 |
| 17 | 43057094 | G | T | N | K | BRCA1 | ENST00000471181 | 0.71008 | 0.18368 | 0.3911 | 0.92651 | 0.825 | 0.7466990762 | 0.75002 | 0.8130992055 | 0.112055 | 6.3643411 | 0 |
| 17 | 43057095 | T | A | N | I | BRCA1 | ENST00000471181 | 0.78067 | 0.11719 | 0.31704 | 0.96397 | 0.91652 | 0.805378597 | 0.837 | 0.953902185 | 0.222422 | 6.3643411 | 0 |
| 17 | 43057095 | T | C | N | S | BRCA1 | ENST00000471181 | 0.54439 | 0.07595 | 0.2579 | 0.90205 | 0.73734 | 0.5481654462 | 0.74941 | 0.6960099339 | 0.065755 | 3.553440616 | 0 |
| 17 | 43057095 | T | G | N | T | BRCA1 | ENST00000471181 | 0.73934 | 0.10371 | 0.30957 | 0.94225 | 0.87296 | 0.6725951567 | 0.79067 | 0.854855597 | 0.13922 | 6.3643411 | 0 |
| 17 | 43057096 | T | A | N | Y | BRCA1 | ENST00000471181 | 0.78447 | 0.29946 | 0.34559 | 0.96217 | 0.8705 | 0.7825876727 | 0.86129 | 0.9790586829 | 0.224571 | 6.3643411 | 0 |
| 17 | 43057096 | T | C | N | D | BRCA1 | ENST00000471181 | 0.76578 | 0.1375 | 0.36149 | 0.90775 | 0.7727 | 0.7091903037 | 0.78749 | 0.8650875688 | 0.113561 | 5.906584744 | 0 |
| 17 | 43057096 | T | G | N | H | BRCA1 | ENST00000471181 | 0.73813 | 0.2665 | 0.32393 | 0.9442 | 0.80595 | 0.6777160398 | 0.84902 | 0.940058589 | 0.132805 | 6.3643411 | 0 |
| 17 | 43057097 | T | A | R | S | BRCA1 | ENST00000471181 | 0.69965 | 0.1044 | 0.30845 | 0.89621 | 0.82948 | 0.6038769648 | 0.6887 | 0.6362742782 | 0.202535 | 6.057057906 | 0 |
| 17 | 43057097 | T | G | R | S | BRCA1 | ENST00000471181 | 0.70784 | 0.1044 | 0.30845 | 0.89601 | 0.81356 | 0.6038769648 | 0.6887 | 0.6728895307 | 0.202528 | 6.057057906 | 0 |
| 17 | 43057098 | C | A | R | I | BRCA1 | ENST00000471181 | 0.81857 | 0.16885 | 0.32962 | 0.93327 | 0.86149 | 0.5972561544 | 0.85397 | 0.9448749423 | 0.267024 | 6.482030025 | 0 |
| 17 | 43057098 | C | G | R | T | BRCA1 | ENST00000471181 | 0.77434 | 0.1365 | 0.35554 | 0.92752 | 0.86753 | 0.6234270965 | 0.81852 | 0.9134963751 | 0.289624 | 6.482030025 | 0 |
| 17 | 43057098 | C | T | R | K | BRCA1 | ENST00000471181 | 0.76594 | 0.09179 | 0.3815 | 0.9099 | 0.81065 | 0.4984873758 | 0.79207 | 0.8581369519 | 0.203054 | 5.243354857 | 0 |
| 17 | 43057099 | T | C | R | G | BRCA1 | ENST00000471181 | 0.84673 | 0.11951 | 0.29941 | 0.96021 | 0.8873 | 0.5915351159 | 0.82228 | 0.7968190908 | 0.313197 | 6.482030025 | 0 |
| 17 | 43057101 | C | A | G | V | BRCA1 | ENST00000471181 | 0.74666 | 0.31166 | 0.44199 | 0.97036 | 0.9182 | 0.8556450073 | 0.96253 | 0.9790488482 | 0.416723 | 6.541286482 | 1 |
| 17 | 43057101 | C | T | G | E | BRCA1 | ENST00000471181 | 0.75567 | 0.13848 | 0.53072 | 0.97173 | 0.9248 | 0.8430255894 | 0.96824 | 0.9620783925 | 0.416313 | 6.541286482 | 1 |
| 17 | 43057102 | C | G | G | R | BRCA1 | ENST00000471181 | 0.83683 | 0.24092 | 0.55264 | 0.97537 | 0.90096 | 0.8536698278 | 0.96508 | 0.982719779 | 0.385516 | 6.541286482 | 1 |
| 17 | 43057102 | C | T | G | R | BRCA1 | ENST00000471181 | 0.84211 | 0.24092 | 0.55264 | 0.97644 | 0.89784 | 0.8536698278 | 0.96508 | 0.9864353538 | 0.385507 | 6.541286482 | 1 |
| 17 | 43057103 | A | C | N | K | BRCA1 | ENST00000471181 | 0.70286 | 0.12857 | 0.42503 | 0.95092 | 0.8345 | 0.7749739018 | 0.77061 | 0.6752702594 | 0.12052 | 6.26792448 | 0 |
| 17 | 43057103 | A | T | N | K | BRCA1 | ENST00000471181 | 0.69604 | 0.12857 | 0.42503 | 0.95244 | 0.8345 | 0.7749739018 | 0.77061 | 0.597058571 | 0.120517 | 6.26792448 | 0 |
| 17 | 43057104 | T | A | N | I | BRCA1 | ENST00000471181 | 0.83244 | 0.24467 | 0.35996 | 0.96923 | 0.91399 | 0.8453764977 | 0.82972 | 0.9234006405 | 0.228934 | 6.26792448 | 0 |
| 17 | 43057104 | T | C | N | S | BRCA1 | ENST00000471181 | 0.70744 | 0.07296 | 0.27631 | 0.90432 | 0.8356 | 0.5719466043 | 0.77478 | 0.6667488813 | 0.109437 | 6.26792448 | 0 |
| 17 | 43057104 | T | G | N | T | BRCA1 | ENST00000471181 | 0.48031 | 0.09369 | 0.33063 | 0.90537 | 0.81933 | 0.7135917742 | 0.74681 | 0.7845112681 | 0.143499 | 3.17491437 | 0 |
| 17 | 43057105 | T | G | N | H | BRCA1 | ENST00000471181 | 0.82241 | 0.23874 | 0.35097 | 0.92276 | 0.85012 | 0.7174558713 | 0.83772 | 0.8792163134 | 0.142985 | 6.26792448 | 0 |
| 17 | 43057107 | A | G | V | A | BRCA1 | ENST00000471181 | 0.81347 | 0.14079 | 0.20616 | 0.9438 | 0.86047 | 0.557769944 | 0.91815 | 0.8900480866 | 0.245988 | 6.071002443 | 0 |
| 17 | 43057107 | A | T | V | D | BRCA1 | ENST00000471181 | 0.87268 | 0.31261 | 0.20954 | 0.96199 | 0.8933 | 0.8284999986 | 0.92159 | 0.9172283411 | 0.392664 | 6.071002443 | 1 |
| 17 | 43057108 | C | T | V | I | BRCA1 | ENST00000471181 | 0.09088 | 0.07746 | 0.10704 | 0.52294 | 0.79273 | 0.3030795228 | 0.25859 | 0.03136929125 | -0.0222259 | 1.695568002 | 0 |
| 17 | 43057110 | A | C | V | G | BRCA1 | ENST00000471181 | 0.8933 | 0.11968 | 0.32472 | 0.95911 | 0.93747 | 0.7885483272 | 0.9831 | 0.9475073218 | 0.420298 | 6.131085886 | 0 |
| 17 | 43057110 | A | G | V | A | BRCA1 | ENST00000471181 | 0.8609 | 0.11002 | 0.47296 | 0.94366 | 0.86852 | 0.6424781522 | 0.98123 | 0.9525767565 | 0.267712 | 6.131085886 | 0 |
| 17 | 43057111 | C | G | V | L | BRCA1 | ENST00000471181 | 0.48101 | 0.08942 | 0.44634 | 0.93896 | 0.8517 | 0.6704674492 | 0.91447 | 0.9387472868 | 0.236541 | 6.131085886 | 0 |
| 17 | 43057112 | A | C | D | E | BRCA1 | ENST00000471181 | 0.67883 | 0.0997 | 0.37918 | 0.94319 | 0.89558 | 0.6265101144 | 0.86682 | 0.8974894881 | 0.361757 | 6.541286319 | 1 |
| 17 | 43057112 | A | T | D | E | BRCA1 | ENST00000471181 | 0.67215 | 0.0997 | 0.37918 | 0.94319 | 0.89558 | 0.6265101144 | 0.86682 | 0.8974894881 | 0.361751 | 6.541286319 | 1 |
| 17 | 43057113 | T | A | D | V | BRCA1 | ENST00000471181 | 0.84711 | 0.19527 | 0.32435 | 0.98297 | 0.9528 | 0.8650375138 | 0.83074 | 0.9763120413 | 0.423613 | 6.541286319 | 1 |
| 17 | 43057113 | T | C | D | G | BRCA1 | ENST00000471181 | 0.8629 | 0.17896 | 0.34313 | 0.97905 | 0.92682 | 0.8231733762 | 0.87189 | 0.9717485905 | 0.411721 | 6.541286319 | 1 |
| 17 | 43057113 | T | G | D | A | BRCA1 | ENST00000471181 | 0.85647 | 0.15111 | 0.33431 | 0.9798 | 0.94129 | 0.823696951 | 0.87546 | 0.9748970866 | 0.404297 | 6.541286319 | 1 |
| 17 | 43057114 | C | A | D | Y | BRCA1 | ENST00000471181 | 0.82342 | 0.22979 | 0.42686 | 0.97398 | 0.92397 | 0.8671898714 | 0.81142 | 0.9741367102 | 0.423179 | 6.541286319 | 1 |
| 17 | 43057114 | C | G | D | H | BRCA1 | ENST00000471181 | 0.81724 | 0.35378 | 0.45182 | 0.97529 | 0.90362 | 0.8069876047 | 0.9139 | 0.9796130657 | 0.421771 | 6.541286319 | 1 |
| 17 | 43057114 | C | T | D | N | BRCA1 | ENST00000471181 | 0.82459 | 0.09032 | 0.41397 | 0.97017 | 0.825 | 0.721531409 | 0.91557 | 0.9620016217 | 0.132302 | 6.541286319 | 0 |
| 17 | 43057116 | C | A | G | V | BRCA1 | ENST00000471181 | 0.807 | 0.1159 | 0.50067 | 0.97375 | 0.91862 | 0.8406988629 | 0.90295 | 0.9837046266 | 0.419373 | 6.541286482 | 1 |
| 17 | 43057116 | C | G | G | A | BRCA1 | ENST00000471181 | 0.77766 | 0.09878 | 0.48402 | 0.95924 | 0.87974 | 0.8022979064 | 0.90221 | 0.9579486859 | 0.334826 | 6.541286482 | 1 |
| 17 | 43057116 | C | T | G | E | BRCA1 | ENST00000471181 | 0.81583 | 0.14451 | 0.58926 | 0.97332 | 0.91399 | 0.8666234688 | 0.90836 | 0.9837046266 | 0.419262 | 6.541286482 | 1 |
| 17 | 43057117 | C | G | G | R | BRCA1 | ENST00000471181 | 0.9071 | 0.11022 | 0.6156 | 0.97233 | 0.92028 | 0.8491739431 | 0.87048 | 0.9837046266 | 0.419927 | 6.541286482 | 1 |
| 17 | 43057117 | C | T | G | R | BRCA1 | ENST00000471181 | 0.9104 | 0.11022 | 0.6156 | 0.97328 | 0.92028 | 0.8491739431 | 0.87048 | 0.9864353538 | 0.419918 | 6.541286482 | 1 |
| 17 | 43057118 | T | A | R | S | BRCA1 | ENST00000471181 | 0.87689 | 0.12392 | 0.24905 | 0.95315 | 0.86352 | 0.7299491879 | 0.81223 | 0.8868048191 | 0.309859 | 5.905959776 | 0 |
| 17 | 43057118 | T | G | R | S | BRCA1 | ENST00000471181 | 0.88354 | 0.12392 | 0.24905 | 0.95291 | 0.88776 | 0.7299491879 | 0.81223 | 0.8868048191 | 0.309861 | 5.905959776 | 0 |
| 17 | 43057119 | C | A | R | I | BRCA1 | ENST00000471181 | 0.73918 | 0.16401 | 0.28412 | 0.94628 | 0.84853 | 0.739136327 | 0.73441 | 0.7691034382 | 0.237603 | 5.579494545 | 0 |
| 17 | 43057119 | C | G | R | T | BRCA1 | ENST00000471181 | 0.69596 | 0.15408 | 0.29168 | 0.92227 | 0.82836 | 0.7315122279 | 0.74252 | 0.5989818171 | 0.221556 | 5.905959776 | 0 |
| 17 | 43057120 | T | C | R | G | BRCA1 | ENST00000471181 | 0.88093 | 0.14885 | 0.24948 | 0.95208 | 0.89376 | 0.788095453 | 0.70069 | 0.8864928484 | 0.333462 | 5.905959776 | 0 |
| 17 | 43057122 | A | C | V | G | BRCA1 | ENST00000471181 | 0.88398 | 0.3804 | 0.38006 | 0.98805 | 0.95717 | 0.933612239 | 0.87529 | 0.9957568049 | 0.458973 | 6.541288293 | 1 |
| 17 | 43057122 | A | G | V | A | BRCA1 | ENST00000471181 | 0.85661 | 0.13763 | 0.50112 | 0.98041 | 0.9458 | 0.7921065422 | 0.89261 | 0.9823827147 | 0.456703 | 6.541288293 | 1 |
| 17 | 43057122 | A | T | V | D | BRCA1 | ENST00000471181 | 0.89634 | 0.15535 | 0.50607 | 0.98762 | 0.94949 | 0.9640847428 | 0.84897 | 0.9891754985 | 0.459038 | 6.541288293 | 1 |
| 17 | 43057123 | C | A | V | F | BRCA1 | ENST00000471181 | 0.84127 | 0.1879 | 0.45168 | 0.98068 | 0.93319 | 0.9138417157 | 0.89868 | 0.9805431962 | 0.45086 | 6.541288293 | 1 |
| 17 | 43057123 | C | G | V | L | BRCA1 | ENST00000471181 | 0.81716 | 0.08942 | 0.47817 | 0.95048 | 0.82387 | 0.750143322 | 0.81825 | 0.9368805289 | 0.290019 | 6.541288293 | 1 |
| 17 | 43057123 | C | T | V | I | BRCA1 | ENST00000471181 | 0.78519 | 0.07604 | 0.32182 | 0.92219 | 0.848 | 0.4689780595 | 0.87824 | 0.9368805289 | 0.234266 | 6.541288293 | 0 |
| 17 | 43057124 | T | A | E | D | BRCA1 | ENST00000471181 | 0.77703 | 0.08227 | 0.44107 | 0.98469 | 0.93083 | 0.7139880632 | 0.92858 | 0.992028892 | 0.436665 | 6.541286163 | 1 |
| 17 | 43057125 | T | A | E | V | BRCA1 | ENST00000471181 | 0.89944 | 0.10704 | 0.4196 | 0.98739 | 0.96189 | 0.8163630213 | 0.86071 | 0.9877266288 | 0.448969 | 6.541286163 | 0 |
| 17 | 43057125 | T | C | E | G | BRCA1 | ENST00000471181 | 0.91223 | 0.10736 | 0.39024 | 0.98529 | 0.96153 | 0.8544848252 | 0.91907 | 0.9934003353 | 0.457206 | 6.541286163 | 1 |
| 17 | 43057125 | T | G | E | A | BRCA1 | ENST00000471181 | 0.87853 | 0.10259 | 0.44533 | 0.986 | 0.96517 | 0.793005116 | 0.92428 | 0.9930375218 | 0.455622 | 6.541286163 | 0 |
| 17 | 43057126 | C | G | E | Q | BRCA1 | ENST00000471181 | 0.84635 | 0.09628 | 0.42447 | 0.97475 | 0.92723 | 0.7462027956 | 0.96608 | 0.980740726 | 0.437948 | 6.541286163 | 0 |
| 17 | 43057126 | C | T | E | K | BRCA1 | ENST00000471181 | 0.90196 | 0.10264 | 0.54571 | 0.9824 | 0.94469 | 0.8667216012 | 0.94387 | 0.9923228025 | 0.454957 | 6.541286163 | 1 |
| 17 | 43057127 | A | C | F | L | BRCA1 | ENST00000471181 | 0.88379 | 0.10659 | 0.51634 | 0.97804 | 0.94506 | 0.8757095698 | 0.94891 | 0.9909716845 | 0.354828 | 6.541286461 | 1 |
| 17 | 43057127 | A | T | F | L | BRCA1 | ENST00000471181 | 0.8784 | 0.10659 | 0.51634 | 0.97903 | 0.93939 | 0.8757095698 | 0.94891 | 0.9896653295 | 0.354826 | 6.541286461 | 1 |
| 17 | 43057128 | A | C | F | C | BRCA1 | ENST00000471181 | 0.91617 | 0.33256 | 0.50606 | 0.98542 | 0.9703 | 0.9382378606 | 0.91972 | 0.9939714074 | 0.465651 | 6.541286461 | 1 |

| | | | | | | | | | | | | | | | | | |
|---|---|---|---|---|---|---|---|---|---|---|---|---|---|---|---|---|---|
| 17 | 43057128 | A | G | F | S | BRCA1 | ENST00000471181 | 0.91622 | 0.19097 | 0.4801 | 0.9849 | 0.96993 | 0.9418082354 | 0.91751 | 0.990474999 | 0.465567 | 6.541286461 | 1 |
| 17 | 43057128 | A | T | F | Y | BRCA1 | ENST00000471181 | 0.85137 | 0.10386 | 0.50246 | 0.94232 | 0.7528 | 0.8284392959 | 0.87639 | 0.9574756622 | 0.175492 | 6.541286461 | 0 |
| 17 | 43057129 | A | C | F | V | BRCA1 | ENST00000471181 | 0.89048 | 0.1217 | 0.44684 | 0.98556 | 0.9681 | 0.9278382095 | 0.94792 | 0.9910199046 | 0.465334 | 6.541286461 | 1 |
| 17 | 43057129 | A | G | F | L | BRCA1 | ENST00000471181 | 0.89998 | 0.10659 | 0.51634 | 0.9753 | 0.96153 | 0.8757095698 | 0.98373 | 0.9899529815 | 0.363176 | 6.541286461 | 1 |
| 17 | 43057129 | A | T | F | I | BRCA1 | ENST00000471181 | 0.88653 | 0.13371 | 0.4826 | 0.97921 | 0.96189 | 0.9298794624 | 0.97511 | 0.9905275106 | 0.334258 | 6.541286461 | 1 |
| 17 | 43057130 | A | C | D | E | BRCA1 | ENST00000471181 | 0.29437 | 0.08852 | 0.23764 | 0.77507 | 0.80536 | 0.3681707409 | 0.70289 | 0.4627877772 | 0.150663 | 2.212427633 | 0 |
| 17 | 43057130 | A | T | D | E | BRCA1 | ENST00000471181 | 0.2902 | 0.08852 | 0.23764 | 0.77974 | 0.80536 | 0.3681707409 | 0.70289 | 0.4627877772 | 0.150661 | 2.212427633 | 0 |
| 17 | 43057131 | T | A | D | V | BRCA1 | ENST00000471181 | 0.66216 | 0.19825 | 0.2077 | 0.97816 | 0.90274 | 0.6463444114 | 0.77316 | 0.4995364753 | 0.33861 | 5.575203538 | 0 |
| 17 | 43057131 | T | C | D | G | BRCA1 | ENST00000471181 | 0.49789 | 0.12158 | 0.22193 | 0.95056 | 0.91904 | 0.6169183824 | 0.86052 | 0.4577309893 | 0.115462 | 3.835929922 | 0 |
| 17 | 43057131 | T | G | D | A | BRCA1 | ENST00000471181 | 0.39289 | 0.12158 | 0.2211 | 0.95221 | 0.86703 | 0.5461125155 | 0.81126 | 0.9359226227 | 0.202246 | 3.582012198 | 0 |
| 17 | 43057132 | C | A | D | Y | BRCA1 | ENST00000471181 | 0.61948 | 0.32934 | 0.27652 | 0.97108 | 0.8859 | 0.6617877452 | 0.78286 | 0.9910219312 | 0.334717 | 5.575203538 | 0 |
| 17 | 43057132 | C | G | D | H | BRCA1 | ENST00000471181 | 0.61223 | 0.27649 | 0.28861 | 0.94496 | 0.87636 | 0.5644357306 | 0.89163 | 0.9607174397 | 0.255717 | 5.575203538 | 0 |
| 17 | 43057132 | C | T | D | N | BRCA1 | ENST00000471181 | 0.60295 | 0.1044 | 0.26437 | 0.9429 | 0.76157 | 0.4616434689 | 0.87722 | 0.9396597743 | 0.0955378 | 2.727272762 | 0 |
| 17 | 43057133 | A | C | H | Q | BRCA1 | ENST00000471181 | 0.5185 | 0.12733 | 0.234 | 0.92088 | 0.81356 | 0.2486931357 | 0.7976 | 0.2219481699 | 0.170808 | 5.606035494 | 0 |
| 17 | 43057133 | A | T | H | Q | BRCA1 | ENST00000471181 | 0.5129 | 0.12733 | 0.234 | 0.92088 | 0.81356 | 0.2486931357 | 0.7976 | 0.2222087063 | 0.170807 | 5.606035494 | 0 |
| 17 | 43057134 | T | A | H | L | BRCA1 | ENST00000471181 | 0.51441 | 0.11884 | 0.22057 | 0.83854 | 0.80772 | 0.5219129933 | 0.59885 | 0.4357538223 | 0.18153 | 3.848606408 | 0 |
| 17 | 43057134 | T | C | H | R | BRCA1 | ENST00000471181 | 0.38748 | 0.11157 | 0.25524 | 0.81431 | 0.79697 | 0.3718387523 | 0.65259 | 0.06628328819 | 0.186563 | 2.77742867 | 0 |
| 17 | 43057134 | T | G | H | P | BRCA1 | ENST00000471181 | 0.55752 | 0.2975 | 0.22726 | 0.93503 | 0.84906 | 0.7563128832 | 0.80826 | 0.4183955015 | 0.259557 | 5.606035494 | 0 |
| 17 | 43057135 | G | A | H | Y | BRCA1 | ENST00000471181 | 0.36968 | 0.18349 | 0.26165 | 0.88898 | 0.85064 | 0.3867218667 | 0.81753 | 0.5926917791 | 0.183405 | 3.243367421 | 0 |
| 17 | 43057135 | G | C | H | D | BRCA1 | ENST00000471181 | 0.58846 | 0.12824 | 0.25808 | 0.90152 | 0.85689 | 0.4927394286 | 0.82865 | 0.2623936653 | 0.184593 | 4.352009894 | 0 |
| 17 | 43057135 | G | T | H | N | BRCA1 | ENST00000471181 | 0.54243 | 0.13697 | 0.25697 | 0.92011 | 0.78597 | 0.3605819232 | 0.82108 | 0.2445503526 | 0.104098 | 5.606035494 | 0 |
| 17 | 43063333 | C | A | E | D | BRCA1 | ENST00000471181 | 0.89202 | 0.12723 | 0.27429 | 0.94873 | 0.84052 | 0.7905235756 | 0.95433 | 0.9787413478 | 0.247648 | 6.293421327 | 0 |
| 17 | 43063333 | C | G | E | D | BRCA1 | ENST00000471181 | 0.90276 | 0.12723 | 0.27429 | 0.94815 | 0.84321 | 0.7905235756 | 0.95433 | 0.9787413478 | 0.24765 | 6.293421327 | 0 |
| 17 | 43063334 | T | A | E | V | BRCA1 | ENST00000471181 | 0.59849 | 0.47742 | 0.24899 | 0.96622 | 0.85274 | 0.8327176809 | 0.85061 | 0.9858539104 | 0.189614 | 3.215549035 | 0 |
| 17 | 43063334 | T | C | E | G | BRCA1 | ENST00000471181 | 0.93406 | 0.47749 | 0.23857 | 0.97723 | 0.92316 | 0.8545024934 | 0.91633 | 0.9835872054 | 0.24479 | 6.293421327 | 0 |
| 17 | 43063334 | T | G | E | A | BRCA1 | ENST00000471181 | 0.8948 | 0.46163 | 0.27412 | 0.96917 | 0.91986 | 0.8312038586 | 0.88904 | 0.9743973017 | 0.227881 | 6.293421327 | 0 |
| 17 | 43063335 | C | T | E | K | BRCA1 | ENST00000471181 | 0.94138 | 0.47059 | 0.36107 | 0.96016 | 0.83615 | 0.8948589506 | 0.95396 | 0.9915617108 | 0.259369 | 6.293421327 | 1 |
| 17 | 43063336 | A | C | N | K | BRCA1 | ENST00000471181 | 0.68867 | 0.36244 | 0.09297 | 0.951 | 0.79516 | 0.4201305202 | 0.85825 | 0.9312272668 | 0.0976429 | 3.920520555 | 0 |
| 17 | 43063336 | A | T | N | K | BRCA1 | ENST00000471181 | 0.68187 | 0.36244 | 0.09297 | 0.95106 | 0.79516 | 0.4201305202 | 0.85825 | 0.9312272668 | 0.0976343 | 3.920520555 | 0 |
| 17 | 43063337 | T | A | N | I | BRCA1 | ENST00000471181 | 0.7149 | 0.4689 | 0.08543 | 0.98103 | 0.80713 | 0.5581617162 | 0.82898 | 0.9136493802 | 0.215355 | 3.920520555 | 0 |
| 17 | 43063337 | T | C | N | S | BRCA1 | ENST00000471181 | 0.33035 | 0.08188 | 0.06537 | 0.82245 | 0.66922 | 0.3020293437 | 0.74498 | 0.03914898454 | -0.110817 | 1.661158344 | 0 |
| 17 | 43063337 | T | G | N | T | BRCA1 | ENST00000471181 | 0.50302 | 0.14859 | 0.09288 | 0.93861 | 0.79273 | 0.3806672937 | 0.80948 | 0.416194737 | 0.105465 | 3.920520555 | 0 |
| 17 | 43063338 | T | A | N | Y | BRCA1 | ENST00000471181 | 0.44613 | 0.47591 | 0.0878 | 0.98241 | 0.86653 | 0.5133668821 | 0.80824 | 0.7099617414 | 0.177527 | 2.611397825 | 0 |
| 17 | 43063338 | T | C | N | D | BRCA1 | ENST00000471181 | 0.20036 | 0.10371 | 0.07566 | 0.68169 | 0.71861 | 0.3069999986 | 0.52084 | 0.02436743918 | 0.0149582 | -1.32863339 | 0 |
| 17 | 43063338 | T | G | N | H | BRCA1 | ENST00000471181 | 0.4138 | 0.35224 | 0.0847 | 0.93707 | 0.7816 | 0.4258319901 | 0.82411 | 0.6735623479 | 0.0775056 | 1.754496339 | 0 |
| 17 | 43063340 | A | C | L | R | BRCA1 | ENST00000471181 | 0.91125 | 0.50988 | 0.51544 | 0.98674 | 0.94318 | 0.9284527006 | 0.99087 | 0.9921656847 | 0.461688 | 6.560063351 | 0 |
| 17 | 43063340 | A | T | L | Q | BRCA1 | ENST00000471181 | 0.90534 | 0.45213 | 0.43682 | 0.98699 | 0.94506 | 0.8882774014 | 0.99313 | 0.9933299422 | 0.44407 | 6.560063351 | 0 |
| 17 | 43063341 | G | C | L | V | BRCA1 | ENST00000471181 | 0.6966 | 0.37815 | 0.3551 | 0.93607 | 0.84535 | 0.5939078238 | 0.98951 | 0.9539468288 | 0.219246 | 6.560063351 | 0 |
| 17 | 43063341 | G | T | L | M | BRCA1 | ENST00000471181 | 0.72588 | 0.44258 | 0.3888 | 0.96122 | 0.79212 | 0.5134868692 | 0.99172 | 0.9153336461 | 0.236422 | 6.560063351 | 0 |
| 17 | 43063342 | C | A | M | I | BRCA1 | ENST00000471181 | 0.1115 | 0.10092 | 0.24036 | 0.56766 | 0.74728 | 0.2878244253 | 0.43419 | 0.0374273964 | 0.0284159 | 0.8517420902 | 0 |
| 17 | 43063342 | C | G | M | I | BRCA1 | ENST00000471181 | 0.11481 | 0.10092 | 0.24036 | 0.56436 | 0.74235 | 0.2878244253 | 0.43419 | 0.03446265818 | 0.0283031 | 0.8517420902 | 0 |
| 17 | 43063342 | C | T | M | I | BRCA1 | ENST00000471181 | 0.11694 | 0.10092 | 0.24036 | 0.56766 | 0.74728 | 0.2878244253 | 0.43419 | 0.03402659705 | 0.0282787 | 0.8517420902 | 0 |
| 17 | 43063343 | A | C | M | R | BRCA1 | ENST00000471181 | 0.53758 | 0.17909 | 0.26436 | 0.88045 | 0.75005 | 0.7535751327 | 0.90856 | 0.9116148949 | 0.118961 | 3.464737447 | 0 |
| 17 | 43063343 | A | G | M | T | BRCA1 | ENST00000471181 | 0.46724 | 0.10881 | 0.338 | 0.88436 | 0.79395 | 0.547740775 | 0.88565 | 0.7966253161 | 0.171975 | 5.140935888 | 0 |
| 17 | 43063343 | A | T | M | K | BRCA1 | ENST00000471181 | 0.62802 | 0.15712 | 0.2828 | 0.87886 | 0.76881 | 0.7361500093 | 0.89352 | 0.8870053887 | 0.123505 | 5.140935888 | 0 |
| 17 | 43063344 | T | A | M | L | BRCA1 | ENST00000471181 | 0.24297 | 0.08613 | 0.21919 | 0.5581 | 0.73 | 0.3150461248 | 0.58009 | 0.06448108703 | 0.0291803 | 1.960715976 | 0 |
| 17 | 43063344 | T | C | M | V | BRCA1 | ENST00000471181 | 0.21869 | 0.09269 | 0.17378 | 0.54014 | 0.77335 | 0.3525021818 | 0.77187 | 0.257863313 | 0.0755877 | 2.017306302 | 0 |
| 17 | 43063344 | T | G | M | L | BRCA1 | ENST00000471181 | 0.24747 | 0.08613 | 0.21919 | 0.55101 | 0.73 | 0.3150461248 | 0.58009 | 0.06448108703 | 0.0291559 | 1.960715976 | 0 |
| 17 | 43063345 | T | A | K | N | BRCA1 | ENST00000471181 | 0.54855 | 0.46591 | 0.29045 | 0.94733 | 0.78223 | 0.5957874874 | 0.8808 | 0.9535613656 | 0.114287 | 6.406715041 | 0 |
| 17 | 43063345 | T | G | K | N | BRCA1 | ENST00000471181 | 0.55559 | 0.46591 | 0.29045 | 0.94131 | 0.78098 | 0.5957874874 | 0.8808 | 0.9413758516 | 0.114287 | 6.406715041 | 0 |
| 17 | 43063346 | T | A | K | I | BRCA1 | ENST00000471181 | 0.68643 | 0.52896 | 0.24877 | 0.96713 | 0.84159 | 0.7137023969 | 0.82856 | 0.9338057637 | 0.214255 | 6.406715041 | 0 |
| 17 | 43063346 | T | C | K | R | BRCA1 | ENST00000471181 | 0.44716 | 0.11002 | 0.21372 | 0.82958 | 0.80595 | 0.3568473288 | 0.79046 | 0.04901405796 | 0.144299 | 2.734715931 | 0 |
| 17 | 43063346 | T | G | K | T | BRCA1 | ENST00000471181 | 0.64198 | 0.46876 | 0.29032 | 0.96011 | 0.85843 | 0.6524194895 | 0.85084 | 0.8981749415 | 0.19714 | 5.955294523 | 0 |
| 17 | 43063347 | T | C | K | E | BRCA1 | ENST00000471181 | 0.80764 | 0.4555 | 0.26367 | 0.95594 | 0.84321 | 0.6305675707 | 0.8846 | 0.9199564457 | 0.243777 | 6.406715041 | 0 |
| 17 | 43063347 | T | G | K | Q | BRCA1 | ENST00000471181 | 0.78075 | 0.37987 | 0.22787 | 0.93431 | 0.85117 | 0.5474695821 | 0.90481 | 0.8043550849 | 0.220052 | 6.406715041 | 0 |
| 17 | 43063348 | T | A | R | S | BRCA1 | ENST00000471181 | 0.49551 | 0.10471 | 0.18494 | 0.8699 | 0.8278 | 0.5371835971 | 0.75671 | 0.1532021165 | 0.173009 | 5.009905637 | 0 |
| 17 | 43063348 | T | G | R | S | BRCA1 | ENST00000471181 | 0.50218 | 0.10471 | 0.18494 | 0.8699 | 0.8278 | 0.5371835971 | 0.75671 | 0.154324457 | 0.173008 | 5.009905637 | 0 |
| 17 | 43063349 | C | A | R | I | BRCA1 | ENST00000471181 | 0.60193 | 0.20959 | 0.20692 | 0.92797 | 0.79878 | 0.6430580642 | 0.82086 | 0.4533387721 | 0.190283 | 5.732780829 | 0 |
| 17 | 43063349 | C | G | R | T | BRCA1 | ENST00000471181 | 0.56428 | 0.16423 | 0.21908 | 0.90187 | 0.80536 | 0.5979566043 | 0.82937 | 0.197838366 | 0.18837 | 5.732780829 | 0 |
| 17 | 43063349 | C | T | R | K | BRCA1 | ENST00000471181 | 0.31587 | 0.09862 | 0.22546 | 0.84417 | 0.78348 | 0.5087275721 | 0.85221 | 0.1587455361 | 0.142879 | 2.865697182 | 0 |
| 17 | 43063350 | T | C | R | G | BRCA1 | ENST00000471181 | 0.34544 | 0.11752 | 0.18031 | 0.73834 | 0.77591 | 0.5254020639 | 0.35686 | 0.0115276781 | -0.0460034 | 2.40089749 | 0 |
| 17 | 43063351 | T | A | E | D | BRCA1 | ENST00000471181 | 0.76179 | 0.14277 | 0.36657 | 0.89305 | 0.82836 | 0.5223055504 | 0.84038 | 0.788441956 | 0.201617 | 6.114754902 | 0 |
| 17 | 43063351 | T | G | E | D | BRCA1 | ENST00000471181 | 0.76998 | 0.14277 | 0.36657 | 0.89785 | 0.82948 | 0.5223055504 | 0.84038 | 0.788441956 | 0.20162 | 6.114754902 | 0 |
| 17 | 43063352 | T | A | E | V | BRCA1 | ENST00000471181 | 0.85242 | 0.33537 | 0.33652 | 0.9519 | 0.86149 | 0.6114536916 | 0.70467 | 0.8542157412 | 0.215248 | 6.114754902 | 0 |
| 17 | 43063352 | T | C | E | G | BRCA1 | ENST00000471181 | 0.86361 | 0.33425 | 0.32375 | 0.9419 | 0.80476 | 0.5478703467 | 0.75723 | 0.802066803 | 0.194905 | 4.316041379 | 0 |
| 17 | 43063352 | T | G | E | A | BRCA1 | ENST00000471181 | 0.68675 | 0.22521 | 0.36606 | 0.92266 | 0.79637 | 0.5247057867 | 0.67422 | 0.2601154813 | 0.137121 | 4.568273159 | 0 |
| 17 | 43063353 | C | G | E | Q | BRCA1 | ENST00000471181 | 0.73069 | 0.28029 | 0.35251 | 0.90824 | 0.84213 | 0.452716079 | 0.80953 | 0.8393669128 | 0.215192 | 4.956178463 | 0 |
| 17 | 43063353 | C | T | E | K | BRCA1 | ENST00000471181 | 0.798 | 0.20194 | 0.46342 | 0.95168 | 0.85012 | 0.5430573013 | 0.76114 | 0.8608608842 | 0.216206 | 5.360479554 | 0 |
| 17 | 43063354 | T | A | K | N | BRCA1 | ENST00000471181 | 0.768 | 0.46955 | 0.28661 | 0.93994 | 0.85792 | 0.6874067718 | 0.84925 | 0.9761528969 | 0.135661 | 6.010096911 | 0 |
| 17 | 43063354 | T | G | K | N | BRCA1 | ENST00000471181 | 0.77616 | 0.46955 | 0.28661 | 0.93982 | 0.85792 | 0.6874067718 | 0.84925 | 0.9722495675 | 0.135665 | 6.010096911 | 0 |
| 17 | 43063355 | T | A | K | I | BRCA1 | ENST00000471181 | 0.82201 | 0.53929 | 0.2439 | 0.98205 | 0.86553 | 0.7545610837 | 0.82227 | 0.9910514355 | 0.252682 | 6.010096911 | 0 |
| 17 | 43063355 | T | C | K | R | BRCA1 | ENST00000471181 | 0.61895 | 0.38243 | 0.20749 | 0.89114 | 0.82892 | 0.4334405555 | 0.86475 | 0.9059051871 | 0.180432 | 6.010096911 | 0 |
| 17 | 43063355 | T | G | K | T | BRCA1 | ENST00000471181 | 0.62559 | 0.46833 | 0.27793 | 0.97912 | 0.89376 | 0.6771205114 | 0.86529 | 0.9520347714 | 0.231725 | 4.503202849 | 0 |
| 17 | 43063356 | T | C | K | E | BRCA1 | ENST00000471181 | 0.5814 | 0.45181 | 0.2592 | 0.94782 | 0.86098 | 0.616027121 | 0.80445 | 0.7743372321 | 0.17407 | 3.876194228 | 0 |
| 17 | 43063356 | T | G | K | Q | BRCA1 | ENST00000471181 | 0.41005 | 0.41728 | 0.22854 | 0.87773 | 0.83172 | 0.5589835577 | 0.78371 | 0.8693973422 | 0.138076 | 3.045304472 | 0 |
| 17 | 43063357 | A | C | I | M | BRCA1 | ENST00000471181 | 0.34565 | 0.10117 | 0.25289 | 0.88224 | 0.78658 | 0.5141373578 | 0.82145 | 0.09768181266 | 0.138168 | 2.161168316 | 0 |
| 17 | 43063358 | A | C | I | S | BRCA1 | ENST00000471181 | 0.90014 | 0.2785 | 0.31411 | 0.96992 | 0.87247 | 0.7985367064 | 0.89793 | 0.9711208344 | 0.379476 | 5.567046617 | 0 |
| 17 | 43063358 | A | G | I | T | BRCA1 | ENST00000471181 | 0.83767 | 0.254 | 0.40585 | 0.95877 | 0.86902 | 0.5541342394 | 0.92532 | 0.9523636699 | 0.285905 | 5.567046617 | 0 |
| 17 | 43063358 | A | T | I | N | BRCA1 | ENST00000471181 | 0.90799 | 0.42299 | 0.37333 | 0.97688 | 0.85378 | 0.791002298 | 0.90071 | 0.987781167 | 0.27073 | 5.567046617 | 0 |
| 17 | 43063359 | T | A | I | F | BRCA1 | ENST00000471181 | 0.33121 | 0.23928 | 0.29264 | 0.92756 | 0.76487 | 0.6118342679 | 0.82408 | 0.1617644578 | 0.185824 | 2.800635851 | 0 |
| 17 | 43063359 | T | C | I | V | BRCA1 | ENST00000471181 | 0.38164 | 0.0809 | 0.21135 | 0.87103 | 0.73223 | 0.3504538506 | 0.63723 | 0.436286509 | 0.102849 | 2.60153989 | 0 |
| 17 | 43063359 | T | G | I | L | BRCA1 | ENST00000471181 | 0.34413 | 0.09117 | 0.28933 | 0.85935 | 0.68734 | 0.5487011119 | 0.60112 | 0.3033594787 | 0.124068 | 3.466531453 | 0 |
| 17 | 43063361 | G | C | S | C | BRCA1 | ENST00000471181 | 0.84089 | 0.44665 | 0.44165 | 0.9612 | 0.86703 | 0.6915695627 | 0.86881 | 0.9407616854 | 0.271115 | 4.025755867 | 0 |
| 17 | 43063362 | A | C | S | A | BRCA1 | ENST00000471181 | 0.84226 | 0.25778 | 0.41912 | 0.97611 | 0.92642 | 0.6260154451 | 0.94549 | 0.9905011654 | 0.324872 | 6.495886828 | 0 |
| 17 | 43063362 | A | G | S | P | BRCA1 | ENST00000471181 | 0.90828 | 0.47633 | 0.46064 | 0.98491 | 0.96226 | 0.9393824402 | 0.95886 | 0.996650517 | 0.47191 | 6.495886828 | 1 |
| 17 | 43063362 | A | T | S | T | BRCA1 | ENST00000471181 | 0.81763 | 0.30031 | 0.45264 | 0.97702 | 0.93824 | 0.686398016 | 0.94789 | 0.9905011654 | 0.408274 | 6.495886828 | 0 |
| 17 | 43063363 | C | A | Q | H | BRCA1 | ENST00000471181 | 0.70799 | 0.4336 | 0.24879 | 0.96353 | 0.84375 | 0.5137684455 | 0.93987 | 0.9655738473 | 0.229464 | 6.301108771 | 0 |
| 17 | 43063363 | C | G | Q | H | BRCA1 | ENST00000471181 | 0.7165 | 0.4336 | 0.24879 | 0.96028 | 0.84375 | 0.5137684455 | 0.93987 | 0.9655738473 | 0.229466 | 6.301108771 | 0 |
| 17 | 43063364 | T | A | Q | L | BRCA1 | ENST00000471181 | 0.57822 | 0.25083 | 0.20772 | 0.97467 | 0.88823 | 0.637235781 | 0.81658 | 0.9713051319 | 0.22154 | 6.301108771 | 0 |
| 17 | 43063364 | T | C | Q | R | BRCA1 | ENST00000471181 | 0.41445 | 0.43593 | 0.24955 | 0.91675 | 0.83615 | 0.5252844318 | 0.82109 | 0.9341394901 | 0.196608 | 3.418025216 | 0 |
| 17 | 43063364 | T | G | Q | P | BRCA1 | ENST00000471181 | 0.73749 | 0.47135 | 0.23191 | 0.97939 | 0.93201 | 0.8551204824 | 0.87901 | 0.9629846215 | 0.354847 | 6.301108771 | 1 |
| 17 | 43063365 | G | C | Q | E | BRCA1 | ENST00000471181 | 0.47338 | 0.35168 | 0.21678 | 0.90727 | 0.81006 | 0.4110665768 | 0.83325 | 0.8909872174 | 0.168169 | 5.117653265 | 0 |
| 17 | 43063365 | G | T | Q | K | BRCA1 | ENST00000471181 | 0.48806 | 0.40777 | 0.25764 | 0.87642 | 0.8153 | 0.6129145918 | 0.80468 | 0.8145667315 | 0.179463 | 5.896183827 | 0 |
| 17 | 43063368 | T | A | T | S | BRCA1 | ENST00000471181 | 0.36259 | 0.25083 | 0.2872 | 0.94969 | 0.74658 | 0.2308195999 | 0.77331 | 0.8412092924 | 0.135817 | 6.179657447 | 0 |
| 17 | 43063368 | T | C | T | A | BRCA1 | ENST00000471181 | 0.38473 | 0.22366 | 0.26674 | 0.91295 | 0.75005 | 0.265251221 | 0.78709 | 0.03117927279 | -0.108625 | 6.179657447 | 0 |
| 17 | 43063370 | A | C | V | G | BRCA1 | ENST00000471181 | 0.88055 | 0.45726 | 0.44219 | 0.99039 | 0.96044 | 0.8922156418 | 0.94747 | 0.9937282801 | 0.454409 | 6.679299815 | 1 |
| 17 | 43063370 | A | G | V | A | BRCA1 | ENST00000471181 | 0.82895 | 0.38553 | 0.55291 | 0.98664 | 0.94167 | 0.7301847415 | 0.89929 | 0.9872301221 | 0.409913 | 6.679299815 | 0 |
| 17 | 43063370 | A | T | V | E | BRCA1 | ENST00000471181 | 0.88202 | 0.49821 | 0.53378 | 0.98978 | 0.95681 | 0.9255366565 | 0.90907 | 0.9716984034 | 0.454028 | 6.679299815 | 1 |
| 17 | 43063371 | C | A | V | L | BRCA1 | ENST00000471181 | 0.491 | 0.27817 | 0.53078 | 0.95647 | 0.76356 | 0.5651798072 | 0.88898 | 0.9204894304 | 0.290956 | 6.679299815 | 0 |
| 17 | 43063371 | C | G | V | L | BRCA1 | ENST00000471181 | 0.49782 | 0.27817 | 0.53078 | 0.95438 | 0.76356 | 0.5651798072 | 0.88898 | 0.9837778807 | 0.348646 | 6.679299815 | 0 |
| 17 | 43063371 | C | T | V | M | BRCA1 | ENST00000471181 | 0.71593 | 0.44312 | 0.55344 | 0.97355 | 0.88259 | 0.5966926712 | 0.94271 | 0.9878394604 | 0.40973 | 6.679299815 | 0 |

| | | | | | | | | | | | | | | | | | | |
|---|---|---|---|---|---|---|---|---|---|---|---|---|---|---|---|---|---|---|
| 17 | 43063372 | C | A | W | C | BRCA1 | ENST00000471181 | 0.94983 | 0.52692 | 0.60104 | 0.99694 | 0.98621 | 0.9775807834 | 0.99273 | 0.9987737536 | 0.539788 | 6.560063911 | 1 |
| 17 | 43063372 | C | G | W | C | BRCA1 | ENST00000471181 | 0.95069 | 0.52692 | 0.60104 | 0.99694 | 0.98621 | 0.9775807834 | 0.99273 | 0.9956703186 | 0.539789 | 6.560063911 | 1 |
| 17 | 43063373 | C | A | W | L | BRCA1 | ENST00000471181 | 0.94661 | 0.52418 | 0.60143 | 0.99563 | 0.98477 | 0.9331403057 | 0.99414 | 0.9984385371 | 0.551306 | 6.560063911 | 1 |
| 17 | 43063874 | A | C | W | G | BRCA1 | ENST00000471181 | 0.95089 | 0.51462 | 0.54739 | 0.99403 | 0.97438 | 0.962232277 | 0.99595 | 0.9508325458 | 0.531854 | 6.560063911 | 1 |
| 17 | 43063874 | A | G | W | R | BRCA1 | ENST00000471181 | 0.95518 | 0.26095 | 0.65155 | 0.9889 | 0.9781 | 0.9795897019 | 0.9989 | 0.9890664816 | 0.555085 | 6.560063911 | 1 |
| 17 | 43063874 | A | T | W | R | BRCA1 | ENST00000471181 | 0.95341 | 0.26095 | 0.65155 | 0.98861 | 0.9781 | 0.9795897019 | 0.9989 | 0.9890664816 | 0.555087 | 6.560063911 | 1 |
| 17 | 43063875 | G | C | F | L | BRCA1 | ENST00000471181 | 0.43406 | 0.12667 | 0.28403 | 0.69069 | 0.72093 | 0.4526117634 | 0.75424 | 0.09424667588 | 0.105598 | 3.148215415 | 0 |
| 17 | 43063875 | G | T | F | L | BRCA1 | ENST00000471181 | 0.4352 | 0.12667 | 0.28403 | 0.69069 | 0.72093 | 0.4526117634 | 0.75424 | 0.09200580062 | 0.105607 | 3.148215415 | 0 |
| 17 | 43063876 | A | C | F | C | BRCA1 | ENST00000471181 | 0.84605 | 0.46915 | 0.27801 | 0.95786 | 0.79151 | 0.5388782055 | 0.51911 | 0.6909816265 | 0.19007 | 6.21205732 | 0 |
| 17 | 43063876 | A | G | F | S | BRCA1 | ENST00000471181 | 0.48114 | 0.41658 | 0.24736 | 0.83708 | 0.70671 | 0.4646590913 | 0.38305 | 0.3930902779 | 0.166192 | 3.059127373 | 0 |
| 17 | 43063876 | A | T | F | Y | BRCA1 | ENST00000471181 | 0.61208 | 0.33194 | 0.28314 | 0.8543 | 0.69168 | 0.4209035159 | 0.69944 | 0.7184662223 | 0.0642505 | 6.21205732 | 0 |
| 17 | 43063877 | A | C | F | V | BRCA1 | ENST00000471181 | 0.60691 | 0.18771 | 0.22134 | 0.90265 | 0.7727 | 0.4863767976 | 0.67905 | 0.4195759786 | 0.184588 | 6.21205732 | 0 |
| 17 | 43063877 | A | G | F | L | BRCA1 | ENST00000471181 | 0.52167 | 0.12667 | 0.28403 | 0.68392 | 0.73223 | 0.4526117634 | 0.72467 | 0.1946502696 | 0.139779 | 3.148215415 | 0 |
| 17 | 43063877 | A | T | F | I | BRCA1 | ENST00000471181 | 0.59516 | 0.13563 | 0.2603 | 0.86449 | 0.73296 | 0.5061093817 | 0.74757 | 0.4168102495 | 0.14693 | 6.21205732 | 0 |
| 17 | 43063879 | T | A | Y | F | BRCA1 | ENST00000471181 | 0.62241 | 0.42251 | 0.53048 | 0.88114 | 0.70427 | 0.5445890397 | 0.84985 | 0.718942523 | 0.142308 | 4.028610414 | 0 |
| 17 | 43063879 | T | C | Y | C | BRCA1 | ENST00000471181 | 0.86845 | 0.30214 | 0.63526 | 0.96644 | 0.90668 | 0.7633605268 | 0.94367 | 0.9695740342 | 0.386284 | 6.465505821 | 0 |
| 17 | 43063879 | T | G | Y | S | BRCA1 | ENST00000471181 | 0.83483 | 0.49362 | 0.55711 | 0.97268 | 0.8933 | 0.7846793551 | 0.92039 | 0.9682629704 | 0.311278 | 6.465505821 | 0 |
| 17 | 43063880 | A | C | Y | D | BRCA1 | ENST00000471181 | 0.89757 | 0.54049 | 0.62471 | 0.96966 | 0.9045 | 0.8583070309 | 0.94831 | 0.9742265344 | 0.393408 | 6.465505821 | 0 |
| 17 | 43063880 | A | G | Y | H | BRCA1 | ENST00000471181 | 0.8877 | 0.49023 | 0.6455 | 0.94367 | 0.77718 | 0.718294307 | 0.93208 | 0.9289300442 | 0.231194 | 6.465505821 | 0 |
| 17 | 43063880 | A | T | Y | N | BRCA1 | ENST00000471181 | 0.89376 | 0.35456 | 0.63358 | 0.9739 | 0.80948 | 0.7947680012 | 0.96412 | 0.9519457817 | 0.337941 | 6.465505821 | 0 |
| 17 | 43063881 | G | C | S | R | BRCA1 | ENST00000471181 | 0.81497 | 0.45969 | 0.64084 | 0.94887 | 0.88776 | 0.9270436621 | 0.91578 | 0.980918467 | 0.397653 | 6.560063803 | 1 |
| 17 | 43063881 | G | T | S | R | BRCA1 | ENST00000471181 | 0.8085 | 0.45969 | 0.64084 | 0.95101 | 0.88307 | 0.9270436621 | 0.91578 | 0.980918467 | 0.397651 | 6.560063803 | 1 |
| 17 | 43063882 | C | A | S | I | BRCA1 | ENST00000471181 | 0.85924 | 0.35417 | 0.60745 | 0.96426 | 0.93901 | 0.9031845738 | 0.95515 | 0.9569734335 | 0.412918 | 6.560063803 | 1 |
| 17 | 43063882 | C | G | S | T | BRCA1 | ENST00000471181 | 0.79104 | 0.2087 | 0.58142 | 0.8756 | 0.82159 | 0.7352719819 | 0.94275 | 0.8145945072 | 0.319414 | 6.560063803 | 0 |
| 17 | 43063882 | C | T | S | N | BRCA1 | ENST00000471181 | 0.7995 | 0.34889 | 0.63364 | 0.87454 | 0.62325 | 0.8328081841 | 0.93093 | 0.8893971443 | 0.133582 | 6.560063803 | 1 |
| 17 | 43063883 | T | A | S | C | BRCA1 | ENST00000471181 | 0.87496 | 0.42859 | 0.57835 | 0.96421 | 0.9248 | 0.8449632428 | 0.95721 | 0.9685192704 | 0.407107 | 6.560063803 | 1 |
| 17 | 43063883 | T | G | S | R | BRCA1 | ENST00000471181 | 0.86741 | 0.45969 | 0.64084 | 0.9664 | 0.91314 | 0.9270436621 | 0.934 | 0.9557685852 | 0.403311 | 6.560063803 | 1 |
| 17 | 43063885 | A | C | V | G | BRCA1 | ENST00000471181 | 0.869 | 0.49191 | 0.30647 | 0.98461 | 0.95645 | 0.9646431444 | 0.99918 | 0.9799349904 | 0.509109 | 6.560064113 | 1 |
| 17 | 43063885 | A | T | V | D | BRCA1 | ENST00000471181 | 0.88272 | 0.52911 | 0.44937 | 0.98539 | 0.9579 | 0.9840826672 | 0.99914 | 0.9571502209 | 0.509057 | 6.560064113 | 1 |
| 17 | 43063886 | C | A | V | F | BRCA1 | ENST00000471181 | 0.80187 | 0.49665 | 0.38521 | 0.98316 | 0.94015 | 0.9341618823 | 0.99883 | 0.9582858086 | 0.501283 | 6.560064113 | 1 |
| 17 | 43063886 | C | G | V | L | BRCA1 | ENST00000471181 | 0.77703 | 0.3681 | 0.40996 | 0.92919 | 0.87198 | 0.7658742364 | 0.99291 | 0.8342872417 | 0.412047 | 6.560064113 | 0 |
| 17 | 43063886 | C | T | V | I | BRCA1 | ENST00000471181 | 0.74483 | 0.38138 | 0.25611 | 0.94328 | 0.83834 | 0.4680779159 | 0.99069 | 0.844199703 | 0.293863 | 6.560064113 | 0 |
| 17 | 43063888 | A | C | V | G | BRCA1 | ENST00000471181 | 0.91383 | 0.47806 | 0.25594 | 0.98438 | 0.91778 | 0.9726832803 | 0.99845 | 0.9535521269 | 0.446714 | 6.212216053 | 1 |
| 17 | 43063888 | A | G | V | A | BRCA1 | ENST00000471181 | 0.89113 | 0.44272 | 0.36034 | 0.97788 | 0.90229 | 0.8357715317 | 0.99631 | 0.9425508976 | 0.446814 | 6.212216053 | 1 |
| 17 | 43063888 | A | T | V | E | BRCA1 | ENST00000471181 | 0.91695 | 0.46806 | 0.33429 | 0.98761 | 0.91945 | 0.9733398281 | 0.99791 | 0.9564117193 | 0.452997 | 6.212216053 | 1 |
| 17 | 43063889 | C | A | V | L | BRCA1 | ENST00000471181 | 0.60852 | 0.15209 | 0.32885 | 0.90205 | 0.82387 | 0.7744005586 | 0.94292 | 0.6455096217 | 0.24027 | 6.212216053 | 1 |
| 17 | 43063889 | C | G | V | L | BRCA1 | ENST00000471181 | 0.61641 | 0.15209 | 0.32885 | 0.89961 | 0.82387 | 0.7744005586 | 0.94292 | 0.56520297 | 0.240272 | 6.212216053 | 0 |
| 17 | 43063889 | C | T | V | I | BRCA1 | ENST00000471181 | 0.34098 | 0.08171 | 0.20783 | 0.68571 | 0.7609 | 0.3984513177 | 0.72542 | 0.347958535 | 0.0741041 | 2.604653884 | 0 |
| 17 | 43063890 | C | A | W | C | BRCA1 | ENST00000471181 | 0.90297 | 0.52734 | 0.74896 | 0.96808 | 0.9328 | 0.9062367721 | 0.98543 | 0.9374461739 | 0.42982 | 6.560063911 | 0 |
| 17 | 43063890 | C | G | W | C | BRCA1 | ENST00000471181 | 0.90833 | 0.52734 | 0.74896 | 0.97057 | 0.9328 | 0.9062367721 | 0.98543 | 0.9372795072 | 0.429821 | 6.560063911 | 0 |
| 17 | 43063891 | C | A | W | L | BRCA1 | ENST00000471181 | 0.8665 | 0.48825 | 0.75658 | 0.97561 | 0.91904 | 0.7940087076 | 0.99316 | 0.9882537127 | 0.413216 | 6.560063911 | 0 |
| 17 | 43063891 | C | G | W | S | BRCA1 | ENST00000471181 | 0.89673 | 0.51914 | 0.73959 | 0.97022 | 0.93083 | 0.8695710072 | 0.99363 | 0.9596031308 | 0.437488 | 6.560063911 | 0 |
| 17 | 43063892 | A | C | W | G | BRCA1 | ENST00000471181 | 0.94621 | 0.2394 | 0.71003 | 0.98172 | 0.9681 | 0.8556716405 | 0.98367 | 0.9859831929 | 0.445525 | 6.560063911 | 1 |
| 17 | 43063892 | A | G | W | R | BRCA1 | ENST00000471181 | 0.93696 | 0.54721 | 0.79849 | 0.97632 | 0.9648 | 0.8721669023 | 0.99252 | 0.9927089214 | 0.445298 | 6.560063911 | 0 |
| 17 | 43063892 | A | T | W | R | BRCA1 | ENST00000471181 | 0.93226 | 0.54721 | 0.79849 | 0.97644 | 0.96553 | 0.8721669023 | 0.99252 | 0.9927089214 | 0.445298 | 6.560063911 | 0 |
| 17 | 43063893 | T | A | K | N | BRCA1 | ENST00000471181 | 0.76942 | 0.47119 | 0.65036 | 0.95192 | 0.76946 | 0.8629738968 | 0.72732 | 0.9458722472 | 0.105174 | 6.253562274 | 0 |
| 17 | 43063893 | T | G | K | N | BRCA1 | ENST00000471181 | 0.77758 | 0.47119 | 0.65036 | 0.95269 | 0.74936 | 0.8629738968 | 0.72732 | 0.9458722472 | 0.105174 | 6.253562274 | 0 |
| 17 | 43063894 | T | A | K | I | BRCA1 | ENST00000471181 | 0.78693 | 0.54606 | 0.59331 | 0.9722 | 0.8278 | 0.8414006211 | 0.71789 | 0.9458578229 | 0.193658 | 6.253562274 | 0 |
| 17 | 43063894 | T | C | K | R | BRCA1 | ENST00000471181 | 0.56428 | 0.13549 | 0.5391 | 0.87114 | 0.75822 | 0.6919918515 | 0.67918 | 0.3620545684 | 0.143766 | 4.139306713 | 0 |
| 17 | 43063894 | T | G | K | T | BRCA1 | ENST00000471181 | 0.74229 | 0.47406 | 0.6331 | 0.96136 | 0.82216 | 0.8450446709 | 0.77173 | 0.7849877481 | 0.195486 | 6.253562274 | 0 |
| 17 | 43063895 | T | C | K | E | BRCA1 | ENST00000471181 | 0.77585 | 0.47668 | 0.61773 | 0.96041 | 0.86803 | 0.8960990736 | 0.8064 | 0.9289503098 | 0.250901 | 6.253562274 | 1 |
| 17 | 43063895 | T | G | K | Q | BRCA1 | ENST00000471181 | 0.74858 | 0.41053 | 0.5647 | 0.95677 | 0.87148 | 0.8206728594 | 0.80598 | 0.9289503098 | 0.199522 | 6.253562274 | 0 |
| 17 | 43063897 | C | A | G | V | BRCA1 | ENST00000471181 | 0.77014 | 0.38153 | 0.5328 | 0.97864 | 0.95097 | 0.8345154894 | 0.96589 | 0.9849244952 | 0.456138 | 6.550696838 | 0 |
| 17 | 43063897 | C | G | G | A | BRCA1 | ENST00000471181 | 0.73557 | 0.38189 | 0.51123 | 0.97333 | 0.93631 | 0.7056946505 | 0.96943 | 0.9771686792 | 0.420018 | 6.550696838 | 0 |
| 17 | 43063897 | C | T | G | E | BRCA1 | ENST00000471181 | 0.77093 | 0.4278 | 0.61934 | 0.9799 | 0.94765 | 0.7614125589 | 0.9643 | 0.9849244952 | 0.442996 | 6.550696838 | 0 |
| 17 | 43063898 | C | G | G | R | BRCA1 | ENST00000471181 | 0.59183 | 0.47886 | 0.62899 | 0.97528 | 0.87829 | 0.6906749688 | 0.91039 | 0.8191790233 | 0.404728 | 3.163768657 | 0 |
| 17 | 43063898 | C | T | G | R | BRCA1 | ENST00000471181 | 0.59675 | 0.47886 | 0.62899 | 0.9762 | 0.87829 | 0.6906749688 | 0.91039 | 0.501897815 | 0.223924 | 3.163768657 | 0 |
| 17 | 43063900 | C | A | G | V | BRCA1 | ENST00000471181 | 0.77006 | 0.4805 | 0.60355 | 0.96073 | 0.94728 | 0.806869463 | 0.9261 | 0.9889349341 | 0.423774 | 6.385763915 | 1 |
| 17 | 43063900 | C | G | G | A | BRCA1 | ENST00000471181 | 0.74061 | 0.40189 | 0.58935 | 0.94132 | 0.89467 | 0.6931855661 | 0.87209 | 0.9473128915 | 0.256859 | 6.385763915 | 0 |
| 17 | 43063900 | C | T | G | E | BRCA1 | ENST00000471181 | 0.779 | 0.38701 | 0.70747 | 0.96147 | 0.92924 | 0.8865032515 | 0.89628 | 0.9596950412 | 0.40507 | 6.385763915 | 0 |
| 17 | 43063901 | C | G | G | R | BRCA1 | ENST00000471181 | 0.72861 | 0.47806 | 0.72518 | 0.97514 | 0.92804 | 0.8746639213 | 0.85995 | 0.9849244952 | 0.40395 | 5.151119149 | 0 |
| 17 | 43063901 | C | T | G | R | BRCA1 | ENST00000471181 | 0.73405 | 0.47806 | 0.72518 | 0.97834 | 0.92804 | 0.8746639213 | 0.85995 | 0.9849244952 | 0.403938 | 5.151119149 | 0 |
| 17 | 43063903 | G | A | A | V | BRCA1 | ENST00000471181 | 0.78796 | 0.395 | 0.68136 | 0.98355 | 0.93123 | 0.8112857511 | 0.96604 | 0.5933313443 | 0.243063 | 6.560063124 | 0 |
| 17 | 43063903 | G | C | A | G | BRCA1 | ENST00000471181 | 0.78756 | 0.41217 | 0.63638 | 0.97296 | 0.93123 | 0.7096079516 | 0.95765 | 0.9386454821 | 0.304463 | 6.560063124 | 1 |
| 17 | 43063903 | G | T | A | E | BRCA1 | ENST00000471181 | 0.77774 | 0.45995 | 0.7288 | 0.98219 | 0.94728 | 0.9448097839 | 0.96811 | 0.8381022215 | 0.337714 | 6.560063124 | 1 |
| 17 | 43063904 | C | A | A | S | BRCA1 | ENST00000471181 | 0.68595 | 0.40038 | 0.61363 | 0.97576 | 0.86149 | 0.6727332664 | 0.94115 | 0.8958111095 | 0.393348 | 6.560063124 | 0 |
| 17 | 43063904 | C | G | A | P | BRCA1 | ENST00000471181 | 0.73837 | 0.48668 | 0.72875 | 0.98111 | 0.94839 | 0.9595682704 | 0.96345 | 0.9696070552 | 0.455445 | 6.560063124 | 1 |
| 17 | 43063904 | C | T | A | T | BRCA1 | ENST00000471181 | 0.72869 | 0.42406 | 0.66287 | 0.98034 | 0.90842 | 0.7469530278 | 0.95019 | 0.9258556962 | 0.331245 | 6.560063124 | 1 |
| 17 | 43063905 | A | C | I | M | BRCA1 | ENST00000471181 | 0.61439 | 0.44516 | 0.71459 | 0.97501 | 0.90842 | 0.7600767135 | 0.85111 | 0.9546717405 | 0.382039 | 6.560063403 | 0 |
| 17 | 43063906 | A | C | I | S | BRCA1 | ENST00000471181 | 0.87782 | 0.49826 | 0.7749 | 0.98223 | 0.95023 | 0.937587799 | 0.98093 | 0.9856491089 | 0.447059 | 6.560063403 | 1 |
| 17 | 43063906 | A | G | I | T | BRCA1 | ENST00000471181 | 0.81716 | 0.47723 | 0.86056 | 0.97807 | 0.94876 | 0.8037101122 | 0.97289 | 0.9580516219 | 0.446101 | 6.560063403 | 0 |
| 17 | 43063906 | A | T | I | N | BRCA1 | ENST00000471181 | 0.8714 | 0.53013 | 0.84066 | 0.98397 | 0.9506 | 0.9453875338 | 0.98763 | 0.9868988991 | 0.366246 | 6.560063403 | 0 |
| 17 | 43063907 | T | A | I | F | BRCA1 | ENST00000471181 | 0.7438 | 0.51764 | 0.76346 | 0.97485 | 0.93554 | 0.8727070849 | 0.95582 | 0.9818675518 | 0.394019 | 6.560063403 | 1 |
| 17 | 43063907 | T | C | I | V | BRCA1 | ENST00000471181 | 0.66161 | 0.29753 | 0.66169 | 0.91179 | 0.8306 | 0.5103966353 | 0.85139 | 0.9321967959 | 0.1495 | 6.560063403 | 0 |
| 17 | 43063907 | T | G | I | L | BRCA1 | ENST00000471181 | 0.72748 | 0.36824 | 0.75351 | 0.92809 | 0.82387 | 0.7461057924 | 0.83731 | 0.8527542354 | 0.206853 | 6.560063403 | 0 |
| 17 | 43063909 | C | A | G | V | BRCA1 | ENST00000471181 | 0.6938 | 0.48461 | 0.8091 | 0.97741 | 0.96371 | 0.9313839617 | 0.97519 | 0.9873235822 | 0.435932 | 6.560064042 | 1 |
| 17 | 43063909 | C | G | G | A | BRCA1 | ENST00000471181 | 0.6643 | 0.41298 | 0.80261 | 0.92879 | 0.79697 | 0.8632494276 | 0.85311 | 0.1742708225 | 0.220015 | 6.560064042 | 0 |
| 17 | 43063909 | C | T | G | E | BRCA1 | ENST00000471181 | 0.70286 | 0.49031 | 0.87053 | 0.97482 | 0.95899 | 0.9788375418 | 0.97609 | 0.9873235822 | 0.43643 | 6.560064042 | 1 |
| 17 | 43063910 | C | G | G | R | BRCA1 | ENST00000471181 | 0.7995 | 0.47638 | 0.89067 | 0.97943 | 0.96664 | 0.9768317218 | 0.97919 | 0.9873235822 | 0.436582 | 6.560064042 | 1 |
| 17 | 43063910 | C | T | G | R | BRCA1 | ENST00000471181 | 0.80495 | 0.47638 | 0.89067 | 0.98168 | 0.96664 | 0.9768317218 | 0.97919 | 0.9873235822 | 0.436572 | 6.560064042 | 1 |
| 17 | 43063912 | A | G | L | P | BRCA1 | ENST00000471181 | 0.80155 | 0.47239 | 0.81844 | 0.98258 | 0.93044 | 0.9519501479 | 0.97158 | 0.9701038003 | 0.401446 | 6.385765452 | 1 |
| 17 | 43063912 | A | T | L | Q | BRCA1 | ENST00000471181 | 0.78804 | 0.4571 | 0.70312 | 0.9663 | 0.82046 | 0.7493831747 | 0.93392 | 0.945707798 | 0.289929 | 5.151120697 | 0 |
| 17 | 43063913 | G | C | L | V | BRCA1 | ENST00000471181 | 0.65508 | 0.37464 | 0.63097 | 0.94889 | 0.8199 | 0.6346292822 | 0.82993 | 0.9107264876 | 0.212535 | 6.385765452 | 0 |
| 17 | 43063913 | G | T | L | I | BRCA1 | ENST00000471181 | 0.65531 | 0.44793 | 0.71176 | 0.95784 | 0.79878 | 0.5157669669 | 0.81588 | 0.855127372 | 0.169439 | 6.385765452 | 0 |
| 17 | 43063914 | A | C | F | L | BRCA1 | ENST00000471181 | 0.63168 | 0.31306 | 0.85628 | 0.95169 | 0.84747 | 0.781555754 | 0.93555 | 0.9313581586 | 0.288807 | 6.560064 | 1 |
| 17 | 43063914 | A | T | F | L | BRCA1 | ENST00000471181 | 0.62529 | 0.31306 | 0.85628 | 0.95102 | 0.84747 | 0.781555754 | 0.93555 | 0.9313581586 | 0.288802 | 6.560064 | 1 |
| 17 | 43063915 | A | C | F | C | BRCA1 | ENST00000471181 | 0.83652 | 0.50935 | 0.84645 | 0.9869 | 0.95935 | 0.8674407246 | 0.99637 | 0.9868988991 | 0.440052 | 6.560064 | 1 |
| 17 | 43063915 | A | G | F | S | BRCA1 | ENST00000471181 | 0.84279 | 0.53493 | 0.81791 | 0.98786 | 0.9579 | 0.9113930467 | 0.99548 | 0.9905520678 | 0.439419 | 6.560064 | 1 |
| 17 | 43063915 | A | T | F | Y | BRCA1 | ENST00000471181 | 0.75647 | 0.4416 | 0.84014 | 0.96732 | 0.90624 | 0.7613621707 | 0.97439 | 0.8987220231 | 0.301289 | 6.560064 | 1 |
| 17 | 43063916 | A | C | F | V | BRCA1 | ENST00000471181 | 0.82272 | 0.44381 | 0.80106 | 0.98692 | 0.93786 | 0.8315295023 | 0.99226 | 0.9711878896 | 0.429081 | 6.560064 | 1 |
| 17 | 43063916 | A | G | F | L | BRCA1 | ENST00000471181 | 0.83567 | 0.31306 | 0.85628 | 0.94292 | 0.85996 | 0.781555754 | 0.96517 | 0.937466085 | 0.322452 | 6.560064 | 1 |
| 17 | 43063916 | A | T | F | I | BRCA1 | ENST00000471181 | 0.81771 | 0.43926 | 0.81511 | 0.97081 | 0.92069 | 0.8618078206 | 0.98602 | 0.9864664674 | 0.430396 | 6.560064 | 1 |
| 17 | 43063918 | T | A | Y | F | BRCA1 | ENST00000471181 | 0.72556 | 0.38073 | 0.88656 | 0.96353 | 0.87198 | 0.7670800006 | 0.91874 | 0.848907418 | 0.230973 | 6.560064193 | 0 |
| 17 | 43063918 | T | C | Y | C | BRCA1 | ENST00000471181 | 0.83298 | 0.4848 | 0.94279 | 0.98499 | 0.95389 | 0.9290081828 | 0.9805 | 0.9905520678 | 0.454723 | 6.560064193 | 1 |
| 17 | 43063918 | T | G | Y | S | BRCA1 | ENST00000471181 | 0.7999 | 0.49362 | 0.90086 | 0.98837 | 0.96773 | 0.9422145498 | 0.97793 | 0.9910926223 | 0.45518 | 6.560064193 | 1 |
| 17 | 43063919 | A | C | Y | D | BRCA1 | ENST00000471181 | 0.83506 | 0.53653 | 0.93695 | 0.9866 | 0.9659 | 0.9767928599 | 0.979 | 0.9910926223 | 0.462083 | 6.560064193 | 1 |
| 17 | 43063919 | A | G | Y | H | BRCA1 | ENST00000471181 | 0.78622 | 0.48901 | 0.9441 | 0.97707 | 0.93123 | 0.9041501161 | 0.97324 | 0.9808411002 | 0.42126 | 6.560064193 | 1 |
| 17 | 43063919 | A | T | Y | N | BRCA1 | ENST00000471181 | 0.82872 | 0.3503 | 0.93846 | 0.98567 | 0.96298 | 0.9546703541 | 0.97632 | 0.9910926223 | 0.450649 | 6.560064193 | 1 |
| 17 | 43063920 | T | A | K | N | BRCA1 | ENST00000471181 | 0.66067 | 0.47119 | 0.84565 | 0.98399 | 0.88022 | 0.9641312751 | 0.90163 | 0.9613983035 | 0.30306 | 6.560063397 | 1 |
| 17 | 43063920 | T | G | K | N | BRCA1 | ENST00000471181 | 0.66874 | 0.47119 | 0.84565 | 0.9838 | 0.87198 | 0.9641312751 | 0.90163 | 0.9613983035 | 0.303063 | 6.560063397 | 1 |
| 17 | 43063921 | T | A | K | I | BRCA1 | ENST00000471181 | 0.82225 | 0.52657 | 0.78352 | 0.99228 | 0.96919 | 0.9625713887 | 0.96841 | 0.9905520678 | 0.489853 | 6.560063397 | 1 |

| | | | | | | | | | | | | | | | | | | |
|---|---|---|---|---|---|---|---|---|---|---|---|---|---|---|---|---|---|---|
| 17 | 43063921 | T | C | K | R | BRCA1 | ENST00000471181 | 0.77116 | 0.39933 | 0.75192 | 0.98267 | 0.95316 | 0.8949649325 | 0.97021 | 0.9445506556 | 0.504957 | 6.560063397 | 1 |
| 17 | 43063921 | T | G | K | T | BRCA1 | ENST00000471181 | 0.7779 | 0.35567 | 0.82683 | 0.99026 | 0.97364 | 0.9534052504 | 0.97684 | 0.9856491089 | 0.514526 | 6.560063397 | 1 |
| 17 | 43063922 | T | C | K | E | BRCA1 | ENST00000471181 | 0.85234 | 0.48493 | 0.81664 | 0.98931 | 0.94802 | 0.9694814321 | 0.96421 | 0.9808411002 | 0.51096 | 6.560063397 | 1 |
| 17 | 43063922 | T | G | K | Q | BRCA1 | ENST00000471181 | 0.77402 | 0.42365 | 0.77865 | 0.98576 | 0.9528 | 0.9458077157 | 0.97191 | 0.9808411002 | 0.492122 | 6.560063397 | 1 |
| 17 | 43063924 | A | C | L | R | BRCA1 | ENST00000471181 | 0.87683 | 0.48843 | 0.77064 | 0.97876 | 0.93162 | 0.874969776 | 0.98632 | 0.9821840525 | 0.415955 | 6.366443769 | 0 |
| 17 | 43063924 | A | T | L | Q | BRCA1 | ENST00000471181 | 0.86872 | 0.44771 | 0.69577 | 0.97781 | 0.94015 | 0.7498788356 | 0.98693 | 0.9821840525 | 0.416625 | 6.366443769 | 0 |
| 17 | 43063925 | G | C | L | V | BRCA1 | ENST00000471181 | 0.59718 | 0.34611 | 0.61555 | 0.94335 | 0.83505 | 0.6083611166 | 0.91059 | 0.8503775894 | 0.215148 | 6.366443769 | 0 |
| 17 | 43063925 | G | T | L | M | BRCA1 | ENST00000471181 | 0.6197 | 0.44173 | 0.66231 | 0.94967 | 0.8233 | 0.5085210204 | 0.9261 | 0.8973980796 | 0.21159 | 4.620061101 | 0 |
| 17 | 43063927 | G | A | T | I | BRCA1 | ENST00000471181 | 0.79294 | 0.45825 | 0.83522 | 0.97747 | 0.94802 | 0.9133025521 | 0.96313 | 0.9500310421 | 0.440764 | 6.560064253 | 1 |
| 17 | 43063927 | G | C | T | R | BRCA1 | ENST00000471181 | 0.80274 | 0.42114 | 0.80118 | 0.97681 | 0.94394 | 0.9396798394 | 0.96037 | 0.9818655849 | 0.455021 | 6.560064253 | 1 |
| 17 | 43063927 | G | T | T | K | BRCA1 | ENST00000471181 | 0.81732 | 0.46225 | 0.81844 | 0.97705 | 0.94205 | 0.9463266495 | 0.96055 | 0.9818655849 | 0.453655 | 6.560064253 | 1 |
| 17 | 43063928 | T | C | T | A | BRCA1 | ENST00000471181 | 0.78177 | 0.27744 | 0.70951 | 0.97059 | 0.94949 | 0.8581725292 | 0.9614 | 0.9577012658 | 0.451455 | 6.560064253 | 1 |
| 17 | 43063928 | T | G | T | P | BRCA1 | ENST00000471181 | 0.81826 | 0.17311 | 0.70794 | 0.98017 | 0.9506 | 0.9152661502 | 0.9696 | 0.9840706587 | 0.459871 | 6.560064253 | 1 |
| 17 | 43063930 | C | A | R | L | BRCA1 | ENST00000471181 | 0.85537 | 0.45918 | 0.71483 | 0.98349 | 0.9517 | 0.9287022455 | 0.96721 | 0.9955496788 | 0.459576 | 6.560063882 | 1 |
| 17 | 43063930 | C | G | R | P | BRCA1 | ENST00000471181 | 0.87376 | 0.50835 | 0.72181 | 0.98582 | 0.9517 | 0.9596233407 | 0.97298 | 0.9936571717 | 0.467529 | 6.560063882 | 1 |
| 17 | 43063931 | G | C | R | G | BRCA1 | ENST00000471181 | 0.8619 | 0.17892 | 0.67644 | 0.97646 | 0.94281 | 0.9439637604 | 0.95284 | 0.9855345488 | 0.463265 | 6.560063882 | 1 |
| 17 | 43063932 | T | A | E | D | BRCA1 | ENST00000471181 | 0.72196 | 0.21571 | 0.71368 | 0.88358 | 0.78348 | 0.6324653019 | 0.72471 | 0.907804966 | 0.176936 | 6.410233347 | 0 |
| 17 | 43063932 | T | G | E | D | BRCA1 | ENST00000471181 | 0.73021 | 0.21571 | 0.71368 | 0.88358 | 0.7872 | 0.6324653019 | 0.72471 | 0.907804966 | 0.176938 | 6.410233347 | 0 |
| 17 | 43063933 | T | A | E | V | BRCA1 | ENST00000471181 | 0.9368 | 0.42392 | 0.68599 | 0.96861 | 0.91652 | 0.7697314618 | 0.88054 | 0.9720848203 | 0.388848 | 6.410233347 | 0 |
| 17 | 43063933 | T | C | E | G | BRCA1 | ENST00000471181 | 0.94266 | 0.44285 | 0.64623 | 0.96223 | 0.89513 | 0.7915542686 | 0.89917 | 0.932423532 | 0.331625 | 6.410233347 | 0 |
| 17 | 43063933 | T | G | E | A | BRCA1 | ENST00000471181 | 0.92606 | 0.37089 | 0.71199 | 0.94775 | 0.83283 | 0.7084224996 | 0.87559 | 0.9346379022 | 0.284539 | 6.410233347 | 0 |
| 17 | 43063934 | C | G | E | Q | BRCA1 | ENST00000471181 | 0.76634 | 0.38153 | 0.68492 | 0.9078 | 0.76223 | 0.6648142881 | 0.84193 | 0.8475667133 | 0.244626 | 6.410233347 | 0 |
| 17 | 43063934 | C | T | E | K | BRCA1 | ENST00000471181 | 0.6974 | 0.12399 | 0.81184 | 0.91628 | 0.81472 | 0.7716092801 | 0.74927 | 0.6498169314 | 0.196301 | 4.788118698 | 0 |
| 17 | 43063935 | A | C | C | W | BRCA1 | ENST00000471181 | 0.8755 | 0.50075 | 0.8426 | 0.98243 | 0.95462 | 0.9306437961 | 0.94279 | 0.9889427423 | 0.406943 | 6.560063124 | 1 |
| 17 | 43063936 | C | A | C | F | BRCA1 | ENST00000471181 | 0.82201 | 0.51685 | 0.79389 | 0.98491 | 0.97735 | 0.9116363947 | 0.99011 | 0.9860268831 | 0.434314 | 6.560063124 | 1 |
| 17 | 43063936 | C | G | C | S | BRCA1 | ENST00000471181 | 0.78511 | 0.47914 | 0.77778 | 0.97989 | 0.96919 | 0.8795885672 | 0.99284 | 0.9481102228 | 0.377318 | 6.560063124 | 0 |
| 17 | 43063936 | C | T | C | Y | BRCA1 | ENST00000471181 | 0.8044 | 0.53986 | 0.88734 | 0.98224 | 0.98549 | 0.9201147386 | 0.99469 | 0.9849244952 | 0.434537 | 6.560063124 | 1 |
| 17 | 43063937 | A | C | C | G | BRCA1 | ENST00000471181 | 0.90774 | 0.48698 | 0.74978 | 0.98511 | 0.96846 | 0.9068210082 | 0.9903 | 0.9786413312 | 0.442702 | 6.560063124 | 0 |
| 17 | 43063937 | A | G | C | R | BRCA1 | ENST00000471181 | 0.91595 | 0.55154 | 0.87648 | 0.98708 | 0.97773 | 0.9534277799 | 0.99137 | 0.9905520678 | 0.4427 | 6.560063124 | 1 |
| 17 | 43063937 | A | T | C | S | BRCA1 | ENST00000471181 | 0.87436 | 0.47914 | 0.77778 | 0.98352 | 0.97104 | 0.8795885672 | 0.99254 | 0.9910156131 | 0.441566 | 6.560063124 | 0 |
| 17 | 43063939 | A | C | V | G | BRCA1 | ENST00000471181 | 0.90218 | 0.49274 | 0.57382 | 0.98792 | 0.93241 | 0.8436004337 | 0.98805 | 0.9861073494 | 0.437671 | 6.560064113 | 1 |
| 17 | 43063939 | A | G | V | A | BRCA1 | ENST00000471181 | 0.87847 | 0.40245 | 0.70975 | 0.97868 | 0.86098 | 0.6670611839 | 0.97308 | 0.9150724411 | 0.272096 | 6.560064113 | 0 |
| 17 | 43063939 | A | T | V | E | BRCA1 | ENST00000471181 | 0.90625 | 0.53073 | 0.69173 | 0.98536 | 0.90755 | 0.8420236778 | 0.97483 | 0.9864664674 | 0.456927 | 6.560064113 | 1 |
| 17 | 43063940 | C | G | V | L | BRCA1 | ENST00000471181 | 0.79176 | 0.27076 | 0.68723 | 0.97183 | 0.85945 | 0.6808024178 | 0.98398 | 0.847240451 | 0.27045 | 6.560064113 | 0 |
| 17 | 43063940 | C | T | V | M | BRCA1 | ENST00000471181 | 0.81967 | 0.44553 | 0.72358 | 0.98082 | 0.80831 | 0.6064405531 | 0.98663 | 0.8535016029 | 0.261928 | 6.560064113 | 0 |
| 17 | 43063941 | A | C | F | L | BRCA1 | ENST00000471181 | 0.43896 | 0.28284 | 0.58219 | 0.98148 | 0.79818 | 0.4676384821 | 0.83055 | 0.5108150244 | 0.169897 | 2.296938979 | 0 |
| 17 | 43063941 | A | T | F | L | BRCA1 | ENST00000471181 | 0.43382 | 0.28284 | 0.58219 | 0.98167 | 0.77527 | 0.4676384821 | 0.83055 | 0.3246143469 | 0.169896 | 2.296938979 | 0 |
| 17 | 43063942 | A | C | F | C | BRCA1 | ENST00000471181 | 0.93223 | 0.50776 | 0.58132 | 0.97862 | 0.85945 | 0.719339377 | 0.95837 | 0.9488047957 | 0.270659 | 6.312530206 | 0 |
| 17 | 43063942 | A | G | F | S | BRCA1 | ENST00000471181 | 0.93201 | 0.52058 | 0.54619 | 0.97802 | 0.88963 | 0.6824533996 | 0.94536 | 0.940335691 | 0.265685 | 6.312530206 | 0 |
| 17 | 43063942 | A | T | F | Y | BRCA1 | ENST00000471181 | 0.87709 | 0.43115 | 0.58127 | 0.96183 | 0.83339 | 0.5080220407 | 0.96409 | 0.8262205638 | 0.192585 | 6.312530206 | 0 |
| 17 | 43063943 | A | C | F | V | BRCA1 | ENST00000471181 | 0.67811 | 0.37495 | 0.49995 | 0.97831 | 0.87491 | 0.6286945035 | 0.91869 | 0.8880725503 | 0.21301 | 6.312530206 | 1 |
| 17 | 43063943 | A | G | F | L | BRCA1 | ENST00000471181 | 0.36297 | 0.28284 | 0.58219 | 0.95167 | 0.79273 | 0.4676384821 | 0.8749 | 0.5456359386 | 0.154202 | 2.296938979 | 0 |
| 17 | 43063944 | C | A | E | D | BRCA1 | ENST00000471181 | 0.26968 | 0.21571 | 0.5476 | 0.8992 | 0.81006 | 0.4006104782 | 0.75455 | 0.6169785857 | 0.145464 | 2.892659933 | 0 |
| 17 | 43063944 | C | G | E | D | BRCA1 | ENST00000471181 | 0.27469 | 0.21571 | 0.5476 | 0.89706 | 0.80713 | 0.4006104782 | 0.75455 | 0.6098405719 | 0.145463 | 2.892659933 | 0 |
| 17 | 43063945 | T | A | E | V | BRCA1 | ENST00000471181 | 0.90715 | 0.36189 | 0.50888 | 0.97167 | 0.83779 | 0.6276291762 | 0.93069 | 0.9685317874 | 0.261627 | 6.189177457 | 0 |
| 17 | 43063945 | T | C | E | G | BRCA1 | ENST00000471181 | 0.91893 | 0.40736 | 0.47339 | 0.96313 | 0.84106 | 0.5039217014 | 0.93793 | 0.9298161864 | 0.225089 | 4.969002816 | 0 |
| 17 | 43063945 | T | G | E | A | BRCA1 | ENST00000471181 | 0.88807 | 0.34506 | 0.54792 | 0.96434 | 0.8278 | 0.5365716123 | 0.94378 | 0.9080473781 | 0.178242 | 6.189177457 | 0 |
| 17 | 43063946 | C | G | E | Q | BRCA1 | ENST00000471181 | 0.66723 | 0.39804 | 0.51381 | 0.93033 | 0.7609 | 0.4654901181 | 0.93725 | 0.7892357593 | 0.181192 | 4.390608542 | 0 |
| 17 | 43063946 | C | T | E | K | BRCA1 | ENST00000471181 | 0.73405 | 0.37561 | 0.67255 | 0.97015 | 0.8153 | 0.4935083481 | 0.92885 | 0.9055244327 | 0.233328 | 6.189177457 | 1 |
| 17 | 43063948 | G | A | A | V | BRCA1 | ENST00000471181 | 0.57575 | 0.094 | 0.32191 | 0.92564 | 0.85482 | 0.3668657426 | 0.85636 | 0.7163401246 | 0.224449 | 2.709604691 | 1 |
| 17 | 43063948 | G | T | A | D | BRCA1 | ENST00000471181 | 0.61462 | 0.34964 | 0.40203 | 0.95868 | 0.86653 | 0.4252404829 | 0.88495 | 0.7867617607 | 0.232471 | 5.646480554 | 1 |
| 17 | 43063949 | C | A | A | S | BRCA1 | ENST00000471181 | 0.41807 | 0.16194 | 0.25664 | 0.93579 | 0.8199 | 0.2880397905 | 0.76716 | 0.6548841 | 0.131354 | 5.646480554 | 1 |
| 17 | 43063949 | C | G | A | P | BRCA1 | ENST00000471181 | 0.23724 | 0.12261 | 0.36449 | 0.88214 | 0.81472 | 0.5776377873 | 0.6689 | 0.152168259 | 0.173188 | 3.827562038 | 0 |
| 17 | 43063949 | C | T | A | T | BRCA1 | ENST00000471181 | 0.24853 | 0.17223 | 0.28038 | 0.87435 | 0.82613 | 0.3219002166 | 0.7571 | 0.4088478982 | 0.150897 | 2.302548063 | 0 |
| 17 | 43063950 | A | C | D | E | BRCA1 | ENST00000471181 | 0.43774 | 0.08968 | 0.55868 | 0.9611 | 0.85274 | 0.6273476963 | 0.93789 | 0.8906274438 | 0.360203 | 6.560063735 | 1 |
| 17 | 43063950 | A | T | D | E | BRCA1 | ENST00000471181 | 0.43315 | 0.08968 | 0.55868 | 0.96136 | 0.85117 | 0.6273476963 | 0.93789 | 0.8840181231 | 0.360199 | 6.560063735 | 1 |
| 17 | 43063951 | T | A | D | V | BRCA1 | ENST00000471181 | 0.92322 | 0.21697 | 0.51608 | 0.985 | 0.94691 | 0.8006901106 | 0.95752 | 0.9904845953 | 0.538123 | 6.560063735 | 1 |
| 17 | 43063951 | T | G | D | A | BRCA1 | ENST00000471181 | 0.91909 | 0.13156 | 0.52443 | 0.97949 | 0.92924 | 0.7516246998 | 0.97234 | 0.9740822315 | 0.521223 | 6.560063735 | 1 |
| 17 | 43067608 | C | G | D | H | BRCA1 | ENST00000471181 | 0.91905 | 0.27304 | 0.62993 | 0.98386 | 0.94129 | 0.7365246228 | 0.93336 | 0.9432914853 | 0.330058 | 6.560063735 | 1 |
| 17 | 43067608 | C | T | D | N | BRCA1 | ENST00000471181 | 0.74117 | 0.08961 | 0.59634 | 0.96994 | 0.76223 | 0.6064171836 | 0.89658 | 0.8991907239 | 0.0957369 | 6.560063735 | 1 |
| 17 | 43067610 | G | A | T | I | BRCA1 | ENST00000471181 | 0.89617 | 0.20107 | 0.8026 | 0.97904 | 0.90581 | 0.7593582511 | 0.95595 | 0.983474791 | 0.375627 | 6.709413834 | 1 |
| 17 | 43067610 | G | C | T | R | BRCA1 | ENST00000471181 | 0.87416 | 0.15735 | 0.76699 | 0.98318 | 0.94986 | 0.8006351366 | 0.95002 | 0.983474791 | 0.409641 | 6.709413834 | 1 |
| 17 | 43067610 | G | T | T | K | BRCA1 | ENST00000471181 | 0.88316 | 0.15865 | 0.78423 | 0.97217 | 0.94167 | 0.8246692411 | 0.95051 | 0.9844139814 | 0.406783 | 6.709413834 | 1 |
| 17 | 43067611 | T | A | T | S | BRCA1 | ENST00000471181 | 0.50569 | 0.08297 | 0.69204 | 0.97461 | 0.90007 | 0.5271110218 | 0.93675 | 0.9175677299 | 0.220385 | 6.709413834 | 0 |
| 17 | 43067611 | T | G | T | P | BRCA1 | ENST00000471181 | 0.77561 | 0.33566 | 0.67712 | 0.99046 | 0.95753 | 0.8370556531 | 0.95175 | 0.9863265753 | 0.370649 | 6.709413834 | 1 |
| 17 | 43067612 | T | A | K | N | BRCA1 | ENST00000471181 | 0.73717 | 0.17838 | 0.86873 | 0.98218 | 0.89648 | 0.6745811877 | 0.89941 | 0.9217121005 | 0.180942 | 6.709413646 | 0 |
| 17 | 43067612 | T | G | K | N | BRCA1 | ENST00000471181 | 0.74531 | 0.17838 | 0.86873 | 0.98209 | 0.89648 | 0.6745811877 | 0.89941 | 0.9217121005 | 0.180943 | 6.709413646 | 0 |
| 17 | 43067613 | T | C | K | R | BRCA1 | ENST00000471181 | 0.68787 | 0.09786 | 0.79988 | 0.97275 | 0.89963 | 0.4649521664 | 0.94892 | 0.8858281374 | 0.207854 | 6.709413646 | 0 |
| 17 | 43067614 | T | C | K | E | BRCA1 | ENST00000471181 | 0.64492 | 0.146 | 0.84708 | 0.98385 | 0.87684 | 0.7104839546 | 0.85923 | 0.8528624177 | 0.241796 | 6.709413646 | 0 |
| 17 | 43067614 | T | G | K | Q | BRCA1 | ENST00000471181 | 0.75368 | 0.12043 | 0.81671 | 0.98323 | 0.90229 | 0.6214349925 | 0.91431 | 0.386101199 | 0.00261458 | 6.709413646 | 0 |
| 17 | 43067615 | C | A | M | I | BRCA1 | ENST00000471181 | 0.30714 | 0.1065 | 0.67788 | 0.88224 | 0.78035 | 0.6555722439 | 0.91689 | 0.616486311 | 0.108253 | 2.715933919 | 0 |
| 17 | 43067615 | C | G | M | I | BRCA1 | ENST00000471181 | 0.3126 | 0.1065 | 0.67788 | 0.87837 | 0.78035 | 0.6555722439 | 0.91689 | 0.627497077 | 0.10826 | 2.715933919 | 0 |
| 17 | 43067615 | C | T | M | I | BRCA1 | ENST00000471181 | 0.31613 | 0.1065 | 0.67788 | 0.88224 | 0.78035 | 0.6555722439 | 0.91689 | 0.616486311 | 0.108236 | 2.715933919 | 0 |
| 17 | 43067616 | A | C | M | R | BRCA1 | ENST00000471181 | 0.74946 | 0.45036 | 0.73624 | 0.98098 | 0.91357 | 0.9342833153 | 0.9822 | 0.967697084 | 0.39965 | 6.269774189 | 1 |
| 17 | 43067616 | A | G | M | T | BRCA1 | ENST00000471181 | 0.68851 | 0.19976 | 0.78826 | 0.97506 | 0.84535 | 0.7362676002 | 0.95905 | 0.9487380385 | 0.245471 | 6.269774189 | 0 |
| 17 | 43067616 | A | T | M | K | BRCA1 | ENST00000471181 | 0.73381 | 0.392 | 0.75973 | 0.9774 | 0.91778 | 0.9158138517 | 0.97959 | 0.9762248993 | 0.383884 | 6.269774189 | 1 |
| 17 | 43067617 | T | A | M | L | BRCA1 | ENST00000471181 | 0.50426 | 0.08633 | 0.67639 | 0.9168 | 0.79938 | 0.653806274 | 0.92552 | 0.9253697395 | 0.165018 | 6.269774189 | 0 |
| 17 | 43067617 | T | C | M | V | BRCA1 | ENST00000471181 | 0.49145 | 0.13049 | 0.61068 | 0.9303 | 0.78597 | 0.6388211576 | 0.93473 | 0.8085352182 | 0.174293 | 6.269774189 | 0 |
| 17 | 43067617 | T | G | M | L | BRCA1 | ENST00000471181 | 0.511 | 0.08633 | 0.67639 | 0.91441 | 0.79938 | 0.653806274 | 0.92552 | 0.9253697395 | 0.165019 | 6.269774189 | 0 |
| 17 | 43067619 | A | C | V | G | BRCA1 | ENST00000471181 | 0.77243 | 0.17697 | 0.26655 | 0.9895 | 0.93123 | 0.9435711619 | 0.97415 | 0.9332769513 | 0.404774 | 4.796127105 | 1 |
| 17 | 43067619 | A | G | V | A | BRCA1 | ENST00000471181 | 0.58183 | 0.12958 | 0.32778 | 0.97607 | 0.8859 | 0.7661297717 | 0.9783 | 0.8190752864 | 0.288524 | 4.796127105 | 0 |
| 17 | 43067619 | A | T | V | D | BRCA1 | ENST00000471181 | 0.65594 | 0.20821 | 0.34091 | 0.99001 | 0.91229 | 0.9687737249 | 0.9614 | 0.9433001876 | 0.424014 | 4.796127105 | 1 |
| 17 | 43067620 | C | A | V | F | BRCA1 | ENST00000471181 | 0.38426 | 0.12219 | 0.31447 | 0.98071 | 0.94205 | 0.8725440083 | 0.89248 | 0.7167183161 | 0.307364 | 4.796127105 | 1 |
| 17 | 43067620 | C | G | V | L | BRCA1 | ENST00000471181 | 0.36643 | 0.08942 | 0.30097 | 0.94062 | 0.80831 | 0.5947058002 | 0.86444 | 0.2729465663 | 0.219236 | 4.796127105 | 0 |
| 17 | 43067620 | C | T | V | I | BRCA1 | ENST00000471181 | 0.18259 | 0.07418 | 0.25166 | 0.84408 | 0.77141 | 0.2838259641 | 0.35926 | 0.03545985371 | 0.0828905 | -0.6809242374 | 0 |
| 17 | 43067622 | A | C | V | G | BRCA1 | ENST00000471181 | 0.82693 | 0.37101 | 0.49113 | 0.99167 | 0.96117 | 0.9380739578 | 0.98685 | 0.9926355481 | 0.448011 | 6.53017713 | 1 |
| 17 | 43067622 | A | G | V | A | BRCA1 | ENST00000471181 | 0.76171 | 0.16931 | 0.63464 | 0.98669 | 0.95645 | 0.7361740535 | 0.98331 | 0.9554655552 | 0.440143 | 6.53017713 | 0 |
| 17 | 43067622 | A | T | V | D | BRCA1 | ENST00000471181 | 0.83982 | 0.37937 | 0.63905 | 0.99234 | 0.96153 | 0.9644176162 | 0.98419 | 0.9888609052 | 0.447939 | 6.53017713 | 1 |
| 17 | 43067623 | C | A | V | F | BRCA1 | ENST00000471181 | 0.58867 | 0.28668 | 0.57748 | 0.98843 | 0.93939 | 0.8822034388 | 0.98471 | 0.9830822945 | 0.416447 | 6.53017713 | 1 |
| 17 | 43067623 | C | G | V | L | BRCA1 | ENST00000471181 | 0.40325 | 0.08961 | 0.61725 | 0.94558 | 0.81356 | 0.6367027869 | 0.93523 | 0.9423413873 | 0.312769 | 6.53017713 | 0 |
| 17 | 43067623 | C | T | V | I | BRCA1 | ENST00000471181 | 0.37992 | 0.07473 | 0.4188 | 0.94864 | 0.84213 | 0.3363161623 | 0.94815 | 0.9198652506 | 0.200898 | 5.691927761 | 0 |
| 17 | 43067624 | A | C | H | Q | BRCA1 | ENST00000471181 | 0.58974 | 0.38103 | 0.60461 | 0.98757 | 0.95935 | 0.9460106456 | 0.94235 | 0.981033206 | 0.399458 | 6.709413538 | 1 |
| 17 | 43067624 | A | T | H | Q | BRCA1 | ENST00000471181 | 0.58368 | 0.38103 | 0.60461 | 0.98758 | 0.95935 | 0.9460106456 | 0.94235 | 0.981033206 | 0.399456 | 6.709413538 | 1 |
| 17 | 43067625 | T | A | H | L | BRCA1 | ENST00000471181 | 0.73277 | 0.33721 | 0.59659 | 0.99366 | 0.97438 | 0.9549984627 | 0.92558 | 0.9904273748 | 0.426519 | 6.709413538 | 1 |
| 17 | 43067625 | T | C | H | R | BRCA1 | ENST00000471181 | 0.68107 | 0.37561 | 0.64861 | 0.98491 | 0.97438 | 0.9640302921 | 0.97402 | 0.9906214476 | 0.428791 | 6.709413538 | 1 |
| 17 | 43067625 | T | G | H | P | BRCA1 | ENST00000471181 | 0.72877 | 0.41087 | 0.5654 | 0.98979 | 0.9781 | 0.9827087173 | 0.96504 | 0.988447845 | 0.429378 | 6.709413538 | 1 |
| 17 | 43067626 | G | A | H | Y | BRCA1 | ENST00000471181 | 0.72869 | 0.21984 | 0.64833 | 0.98109 | 0.9528 | 0.9306907967 | 0.95439 | 0.983474791 | 0.415897 | 6.709413538 | 1 |
| 17 | 43067626 | G | C | H | D | BRCA1 | ENST00000471181 | 0.7438 | 0.16256 | 0.63189 | 0.98299 | 0.97141 | 0.9797805685 | 0.96964 | 0.989394486 | 0.436628 | 6.709413538 | 1 |
| 17 | 43067626 | G | T | H | N | BRCA1 | ENST00000471181 | 0.73245 | 0.21479 | 0.633 | 0.97969 | 0.93747 | 0.9368111952 | 0.96985 | 0.989394486 | 0.265598 | 6.709413538 | 1 |
| 17 | 43067628 | G | A | T | I | BRCA1 | ENST00000471181 | 0.66019 | 0.19591 | 0.74958 | 0.97781 | 0.96517 | 0.9105552858 | 0.96177 | 0.983474791 | 0.438305 | 6.709413834 | 1 |
| 17 | 43067628 | G | C | T | S | BRCA1 | ENST00000471181 | 0.44722 | 0.08234 | 0.64476 | 0.96767 | 0.94091 | 0.7405431346 | 0.95025 | 0.973551929 | 0.332058 | 6.709413834 | 1 |

| | | | | | | | | | | | | | | | | | | |
|---|---|---|---|---|---|---|---|---|---|---|---|---|---|---|---|---|---|---|
| 17 | 43067628 | G | T | T | N | BRCA1 | ENST00000471181 | 0.61447 | 0.17371 | 0.71644 | 0.97779 | 0.93397 | 0.921152207 | 0.95339 | 0.9827303886 | 0.267044 | 6.709413834 | 1 |
| 17 | 43067629 | T | A | T | S | BRCA1 | ENST00000471181 | 0.46134 | 0.08234 | 0.64476 | 0.97958 | 0.94543 | 0.7405431346 | 0.93899 | 0.9683006406 | 0.324992 | 6.709413834 | 0 |
| 17 | 43067629 | T | C | T | A | BRCA1 | ENST00000471181 | 0.50004 | 0.0931 | 0.62922 | 0.98047 | 0.9659 | 0.8659960742 | 0.95242 | 0.9705802798 | 0.418536 | 6.709413834 | 1 |
| 17 | 43067629 | T | G | T | P | BRCA1 | ENST00000471181 | 0.71265 | 0.24371 | 0.63619 | 0.98954 | 0.96298 | 0.9378197307 | 0.95704 | 0.9826409221 | 0.430895 | 6.709413834 | 1 |
| 17 | 43067631 | G | A | T | I | BRCA1 | ENST00000471181 | 0.67851 | 0.44606 | 0.64202 | 0.98686 | 0.91986 | 0.6504030107 | 0.9461 | 0.9814019799 | 0.240471 | 6.709413834 | 1 |
| 17 | 43067632 | T | A | T | S | BRCA1 | ENST00000471181 | 0.46308 | 0.08211 | 0.53661 | 0.97697 | 0.92356 | 0.5309860119 | 0.93882 | 0.9683006406 | 0.233477 | 6.709413834 | 0 |
| 17 | 43067632 | T | C | T | A | BRCA1 | ENST00000471181 | 0.5129 | 0.0992 | 0.518 | 0.98049 | 0.92069 | 0.6099061716 | 0.94065 | 0.9705802798 | 0.222536 | 6.709413834 | 0 |
| 17 | 43067632 | T | G | T | P | BRCA1 | ENST00000471181 | 0.72276 | 0.32792 | 0.53176 | 0.99347 | 0.93824 | 0.7981633737 | 0.94918 | 0.9863265753 | 0.254864 | 6.709413834 | 1 |
| 17 | 43067633 | C | A | E | D | BRCA1 | ENST00000471181 | 0.26345 | 0.08684 | 0.4774 | 0.94079 | 0.84535 | 0.5493347447 | 0.75345 | 0.740231514 | 0.172545 | 4.878214947 | 0 |
| 17 | 43067633 | C | G | E | D | BRCA1 | ENST00000471181 | 0.26837 | 0.08684 | 0.4774 | 0.94033 | 0.84535 | 0.5493347447 | 0.75345 | 0.7650667429 | 0.172542 | 4.878214947 | 0 |
| 17 | 43067634 | T | A | E | V | BRCA1 | ENST00000471181 | 0.59682 | 0.12093 | 0.43942 | 0.99328 | 0.88069 | 0.7348500512 | 0.77265 | 0.820915401 | 0.15071 | 6.627189414 | 1 |
| 17 | 43067634 | T | G | E | A | BRCA1 | ENST00000471181 | 0.54089 | 0.09771 | 0.46958 | 0.96892 | 0.82892 | 0.6543653642 | 0.75826 | 0.6292396489 | 0.104576 | 6.627189414 | 0 |
| 17 | 43067635 | C | G | E | Q | BRCA1 | ENST00000471181 | 0.48246 | 0.1111 | 0.46284 | 0.96348 | 0.80118 | 0.6037045653 | 0.83251 | 0.8048781157 | 0.171495 | 6.627189414 | 1 |
| 17 | 43067635 | C | T | E | K | BRCA1 | ENST00000471181 | 0.50955 | 0.10314 | 0.57045 | 0.96482 | 0.85012 | 0.6388643436 | 0.75698 | 0.8327643275 | 0.173673 | 6.627189414 | 1 |
| 17 | 43067636 | T | G | E | D | BRCA1 | ENST00000471181 | 0.376 | 0.08358 | 0.37768 | 0.97896 | 0.86803 | 0.4203844994 | 0.85952 | 0.5704740286 | 0.170448 | 4.96788141 | 0 |
| 17 | 43067637 | T | A | E | V | BRCA1 | ENST00000471181 | 0.65672 | 0.11227 | 0.35692 | 0.99229 | 0.92192 | 0.5804634934 | 0.80167 | 0.8141777961 | 0.227892 | 6.349044872 | 1 |
| 17 | 43067637 | T | C | E | G | BRCA1 | ENST00000471181 | 0.67891 | 0.11632 | 0.34337 | 0.98813 | 0.8887 | 0.472408135 | 0.85805 | 0.8824033737 | 0.175833 | 4.804398592 | 0 |
| 17 | 43067637 | T | G | E | A | BRCA1 | ENST00000471181 | 0.4015 | 0.09744 | 0.37784 | 0.98027 | 0.88496 | 0.4870892421 | 0.83972 | 0.8342170715 | 0.106675 | 6.349044872 | 0 |
| 17 | 43067638 | C | T | E | K | BRCA1 | ENST00000471181 | 0.42144 | 0.10851 | 0.48085 | 0.96608 | 0.85378 | 0.5035579533 | 0.84595 | 0.814396441 | 0.209411 | 4.172786516 | 1 |
| 17 | 43067640 | G | C | T | S | BRCA1 | ENST00000471181 | 0.27455 | 0.08152 | 0.32006 | 0.70662 | 0.71153 | 0.4946781562 | 0.55893 | 0.3214621544 | 0.0867104 | 2.748065343 | 0 |
| 17 | 43067640 | G | T | T | N | BRCA1 | ENST00000471181 | 0.3944 | 0.11074 | 0.38745 | 0.92121 | 0.78782 | 0.6987391351 | 0.6409 | 0.5325635672 | 0.0917924 | 6.016927684 | 0 |
| 17 | 43067641 | T | A | T | S | BRCA1 | ENST00000471181 | 0.29357 | 0.08152 | 0.32006 | 0.76635 | 0.71312 | 0.4946781562 | 0.69658 | 0.2827510834 | 0.0826858 | 2.748065343 | 0 |
| 17 | 43067641 | T | C | T | A | BRCA1 | ENST00000471181 | 0.41878 | 0.08917 | 0.31442 | 0.95031 | 0.82613 | 0.6953639372 | 0.79093 | 0.8804332018 | 0.177082 | 6.016927684 | 0 |
| 17 | 43067641 | T | G | T | P | BRCA1 | ENST00000471181 | 0.59682 | 0.14172 | 0.33136 | 0.9708 | 0.81472 | 0.8178376925 | 0.78396 | 0.9373443127 | 0.21194 | 6.016927684 | 1 |
| 17 | 43067642 | A | C | I | M | BRCA1 | ENST00000471181 | 0.54351 | 0.25718 | 0.42671 | 0.9697 | 0.81124 | 0.432870958 | 0.92047 | 0.7512642145 | 0.122117 | 5.990485038 | 0 |
| 17 | 43067643 | A | C | I | S | BRCA1 | ENST00000471181 | 0.71032 | 0.21991 | 0.47827 | 0.98605 | 0.88117 | 0.8086825436 | 0.97453 | 0.9138073325 | 0.209904 | 5.990485038 | 0 |
| 17 | 43067643 | A | G | I | T | BRCA1 | ENST00000471181 | 0.64508 | 0.1961 | 0.56746 | 0.98073 | 0.88637 | 0.5950645356 | 0.97515 | 0.809979856 | 0.218511 | 5.990485038 | 0 |
| 17 | 43067643 | A | T | I | N | BRCA1 | ENST00000471181 | 0.7145 | 0.3113 | 0.53655 | 0.98837 | 0.87636 | 0.7317484975 | 0.97192 | 0.9455958009 | 0.198277 | 5.990485038 | 0 |
| 17 | 43067644 | T | A | I | F | BRCA1 | ENST00000471181 | 0.32589 | 0.29058 | 0.47222 | 0.93772 | 0.84853 | 0.6297179943 | 0.86231 | 0.6614652276 | 0.0925199 | 5.990485038 | 0 |
| 17 | 43067644 | T | C | I | V | BRCA1 | ENST00000471181 | 0.10662 | 0.07664 | 0.38233 | 0.7468 | 0.74936 | 0.2617643813 | 0.32538 | 0.07155850312 | 0.0359631 | 1.703235513 | 0 |
| 17 | 43067644 | T | G | I | L | BRCA1 | ENST00000471181 | 0.20962 | 0.10371 | 0.46017 | 0.94818 | 0.80595 | 0.4260571879 | 0.63271 | 0.5113272071 | 0.0837865 | 5.990485038 | 0 |
| 17 | 43067646 | A | C | L | R | BRCA1 | ENST00000471181 | 0.17127 | 0.17672 | 0.15606 | 0.89048 | 0.77591 | 0.3102483928 | 0.31862 | 0.1235827037 | 0.0492383 | 3.669417739 | 0 |
| 17 | 43067646 | A | G | L | P | BRCA1 | ENST00000471181 | 0.18585 | 0.29149 | 0.17971 | 0.91284 | 0.84159 | 0.6392181765 | 0.37855 | 0.1208760142 | 0.0208358 | 4.845245152 | 0 |
| 17 | 43067646 | A | T | L | Q | BRCA1 | ENST00000471181 | 0.09247 | 0.09619 | 0.13804 | 0.70259 | 0.78782 | 0.2747399044 | 0.2529 | 0.06000712514 | -0.0169591 | 0.8526355802 | 0 |
| 17 | 43067647 | G | C | L | V | BRCA1 | ENST00000471181 | 0.06329 | 0.09179 | 0.09603 | 0.80025 | 0.77845 | 0.2727874196 | 0.42699 | 0.06936883926 | 0.0653449 | 1.693779102 | 0 |
| 17 | 43067647 | G | T | L | I | BRCA1 | ENST00000471181 | 0.09465 | 0.10456 | 0.13377 | 0.85249 | 0.74936 | 0.1960174967 | 0.34151 | 0.0755568498 | 0.00797007 | 3.815692651 | 0 |
| 17 | 43067648 | A | C | N | K | BRCA1 | ENST00000471181 | 0.49254 | 0.16535 | 0.34865 | 0.96085 | 0.76223 | 0.2286725028 | 0.85708 | 0.4265828518 | 0.0792321 | 5.914045025 | 0 |
| 17 | 43067648 | A | T | N | K | BRCA1 | ENST00000471181 | 0.4871 | 0.16535 | 0.34865 | 0.96085 | 0.76289 | 0.2286725028 | 0.85708 | 0.428069569 | 0.0792273 | 5.914045025 | 0 |
| 17 | 43067649 | T | A | N | I | BRCA1 | ENST00000471181 | 0.5154 | 0.13463 | 0.31259 | 0.97398 | 0.85792 | 0.4041311258 | 0.85441 | 0.7865610719 | 0.205155 | 5.914045025 | 0 |
| 17 | 43067649 | T | C | N | S | BRCA1 | ENST00000471181 | 0.30766 | 0.07746 | 0.25746 | 0.88997 | 0.57487 | 0.1318392707 | 0.75894 | 0.152473554 | 0.0367358 | 4.017371328 | 0 |
| 17 | 43067649 | T | G | N | T | BRCA1 | ENST00000471181 | 0.47043 | 0.10471 | 0.29629 | 0.95088 | 0.77399 | 0.2428017081 | 0.7976 | 0.3820723593 | 0.097705 | 5.914045025 | 0 |
| 17 | 43067650 | T | A | N | Y | BRCA1 | ENST00000471181 | 0.49357 | 0.18394 | 0.32323 | 0.97387 | 0.84588 | 0.358061576 | 0.86432 | 0.6149440408 | 0.191356 | 5.914045025 | 0 |
| 17 | 43067650 | T | C | N | D | BRCA1 | ENST00000471181 | 0.21985 | 0.10371 | 0.33171 | 0.77104 | 0.71548 | 0.2129021483 | 0.59906 | 0.0727004847 | 0.0441403 | 2.030357117 | 0 |
| 17 | 43067650 | T | G | N | H | BRCA1 | ENST00000471181 | 0.35669 | 0.26878 | 0.30233 | 0.93081 | 0.76223 | 0.25663894 | 0.75006 | 0.1653719842 | 0.0786244 | 4.316185611 | 0 |
| 17 | 43067652 | G | C | T | S | BRCA1 | ENST00000471181 | 0.20002 | 0.08171 | 0.38278 | 0.77672 | 0.75417 | 0.255315695 | 0.57001 | 0.190699473 | 0.0913037 | 2.612201818 | 0 |
| 17 | 43067652 | G | T | T | N | BRCA1 | ENST00000471181 | 0.29984 | 0.10479 | 0.43798 | 0.91685 | 0.75687 | 0.4646671611 | 0.74794 | 0.5416994691 | 0.106917 | 4.311305838 | 0 |
| 17 | 43067653 | T | A | T | S | BRCA1 | ENST00000471181 | 0.22488 | 0.08171 | 0.38278 | 0.78123 | 0.77076 | 0.255315695 | 0.60405 | 0.321598053 | 0.0944929 | 2.612201818 | 0 |
| 17 | 43067653 | T | C | T | A | BRCA1 | ENST00000471181 | 0.35752 | 0.08917 | 0.3695 | 0.91021 | 0.77909 | 0.3144852444 | 0.72678 | 0.7279246449 | 0.162444 | 4.520772272 | 0 |
| 17 | 43067653 | T | G | T | P | BRCA1 | ENST00000471181 | 0.41416 | 0.16401 | 0.39128 | 0.96834 | 0.8199 | 0.703989645 | 0.83329 | 0.8775971532 | 0.222975 | 6.158277786 | 0 |
| 17 | 43067654 | T | A | L | F | BRCA1 | ENST00000471181 | 0.29059 | 0.31486 | 0.33042 | 0.95946 | 0.7562 | 0.4288164763 | 0.92707 | 0.5241733193 | 0.142612 | 3.155145399 | 0 |
| 17 | 43067654 | T | G | L | F | BRCA1 | ENST00000471181 | 0.29572 | 0.31486 | 0.33042 | 0.96126 | 0.7562 | 0.4288164763 | 0.92707 | 0.5172675848 | 0.142612 | 3.155145399 | 0 |
| 17 | 43067656 | A | C | L | V | BRCA1 | ENST00000471181 | 0.13678 | 0.12085 | 0.28635 | 0.63206 | 0.69168 | 0.2837392617 | 0.46097 | 0.07643921636 | 0.0687053 | 3.136903183 | 0 |
| 17 | 43067656 | A | T | L | I | BRCA1 | ENST00000471181 | 0.20795 | 0.22834 | 0.34221 | 0.83677 | 0.77972 | 0.2174990848 | 0.43999 | 0.2808268368 | 0.0596983 | 3.874407416 | 0 |
| 17 | 43067658 | G | A | T | I | BRCA1 | ENST00000471181 | 0.19435 | 0.0992 | 0.24295 | 0.8943 | 0.77011 | 0.341278169 | 0.50877 | 0.1127164774 | 0.169439 | 4.388096721 | 0 |
| 17 | 43067658 | G | T | T | N | BRCA1 | ENST00000471181 | 0.17863 | 0.12343 | 0.19684 | 0.92106 | 0.77011 | 0.3807672025 | 0.56915 | 0.3057265282 | 0.100494 | 3.16740243 | 0 |
| 17 | 43067659 | T | A | T | S | BRCA1 | ENST00000471181 | 0.06547 | 0.08171 | 0.14718 | 0.81487 | 0.78658 | 0.236817574 | 0.19356 | 0.04137590604 | 0.0687708 | 2.877796372 | 0 |
| 17 | 43067659 | T | C | T | A | BRCA1 | ENST00000471181 | 0.0687 | 0.08898 | 0.13928 | 0.83309 | 0.80948 | 0.3036059619 | 0.20273 | 0.04549454676 | 0.0236901 | 1.826486857 | 0 |
| 17 | 43067659 | T | G | T | P | BRCA1 | ENST00000471181 | 0.15978 | 0.16401 | 0.163 | 0.94197 | 0.8153 | 0.7080764198 | 0.20884 | 0.2959975004 | 0.149876 | 5.158971718 | 0 |
| 17 | 43067661 | A | G | I | T | BRCA1 | ENST00000471181 | 0.52685 | 0.12108 | 0.24221 | 0.97076 | 0.79998 | 0.4976690872 | 0.77778 | 0.3336541454 | 0.151934 | 5.570803476 | 0 |
| 17 | 43067662 | T | C | I | V | BRCA1 | ENST00000471181 | 0.18925 | 0.07664 | 0.11329 | 0.75904 | 0.77399 | 0.4152658334 | 0.17975 | 0.0416812225 | 0.0418394 | 3.59348084 | 0 |
| 17 | 43067662 | T | G | I | L | BRCA1 | ENST00000471181 | 0.20032 | 0.09089 | 0.16235 | 0.80115 | 0.74587 | 0.5067570197 | 0.23763 | 0.05047253892 | 0.0151518 | 2.819422206 | 0 |
| 17 | 43067663 | G | C | H | Q | BRCA1 | ENST00000471181 | 0.1923 | 0.09821 | 0.29143 | 0.85178 | 0.7528 | 0.3327774196 | 0.68722 | 0.1197413005 | 0.106353 | 3.153057017 | 0 |
| 17 | 43067663 | G | T | H | Q | BRCA1 | ENST00000471181 | 0.18932 | 0.09821 | 0.29143 | 0.85189 | 0.7528 | 0.3327774196 | 0.68722 | 0.1203111753 | 0.106356 | 3.153057017 | 0 |
| 17 | 43067664 | T | A | H | L | BRCA1 | ENST00000471181 | 0.34817 | 0.1154 | 0.28056 | 0.94813 | 0.76553 | 0.5431759663 | 0.68532 | 0.380877167 | 0.143945 | 5.936800001 | 0 |
| 17 | 43067664 | T | C | H | R | BRCA1 | ENST00000471181 | 0.21997 | 0.11157 | 0.31765 | 0.81378 | 0.75485 | 0.4538796764 | 0.63697 | 0.1219973788 | 0.11768 | 2.719521992 | 0 |
| 17 | 43067664 | T | G | H | P | BRCA1 | ENST00000471181 | 0.47759 | 0.22181 | 0.27705 | 0.96447 | 0.81356 | 0.7539890211 | 0.79955 | 0.3759800196 | 0.160417 | 5.936800001 | 0 |
| 17 | 43067665 | G | A | H | Y | BRCA1 | ENST00000471181 | 0.24524 | 0.10573 | 0.32328 | 0.85366 | 0.80536 | 0.4418262617 | 0.70413 | 0.2058498717 | 0.0595815 | 4.619388471 | 0 |
| 17 | 43067665 | G | C | H | D | BRCA1 | ENST00000471181 | 0.36962 | 0.13936 | 0.31403 | 0.93848 | 0.78535 | 0.5634301835 | 0.6191 | 0.3408206701 | 0.143246 | 5.936800001 | 0 |
| 17 | 43067665 | G | T | H | N | BRCA1 | ENST00000471181 | 0.2641 | 0.1162 | 0.31687 | 0.90171 | 0.71548 | 0.4018150995 | 0.59599 | 0.1649151078 | 0.0576919 | 4.236549285 | 0 |
| 17 | 43067666 | G | C | H | Q | BRCA1 | ENST00000471181 | 0.33371 | 0.10026 | 0.33886 | 0.91476 | 0.79818 | 0.4003819066 | 0.7266 | 0.2825654745 | 0.17901 | 5.666732974 | 0 |
| 17 | 43067666 | G | T | H | Q | BRCA1 | ENST00000471181 | 0.3294 | 0.10026 | 0.33886 | 0.93175 | 0.79818 | 0.4003819066 | 0.7266 | 0.2825654745 | 0.179008 | 5.666732974 | 0 |
| 17 | 43067667 | T | A | H | L | BRCA1 | ENST00000471181 | 0.27884 | 0.11798 | 0.32172 | 0.93108 | 0.77463 | 0.4268379367 | 0.58555 | 0.3892680407 | 0.154358 | 5.666732974 | 0 |
| 17 | 43067667 | T | C | H | R | BRCA1 | ENST00000471181 | 0.32998 | 0.11557 | 0.35009 | 0.86666 | 0.79456 | 0.5095557897 | 0.71596 | 0.149214214 | 0.177181 | 5.666732974 | 0 |
| 17 | 43067667 | T | G | H | P | BRCA1 | ENST00000471181 | 0.48614 | 0.19188 | 0.32307 | 0.93312 | 0.81646 | 0.7218396598 | 0.79746 | 0.2384453351 | 0.21856 | 5.666732974 | 1 |
| 17 | 43067668 | G | A | H | Y | BRCA1 | ENST00000471181 | 0.12196 | 0.10573 | 0.35555 | 0.72413 | 0.78535 | 0.392577283 | 0.25393 | 0.0249574855 | 0.0847534 | 1.629762887 | 0 |
| 17 | 43067668 | G | T | H | N | BRCA1 | ENST00000471181 | 0.15824 | 0.10997 | 0.36125 | 0.89663 | 0.76157 | 0.4111497125 | 0.31001 | 0.2243418396 | 0.0414412 | 3.626096382 | 0 |
| 17 | 43067669 | T | A | K | N | BRCA1 | ENST00000471181 | 0.58767 | 0.11481 | 0.2837 | 0.95266 | 0.73223 | 0.4722610328 | 0.69741 | 0.879771769 | 0.102258 | 6.079973395 | 0 |
| 17 | 43067669 | T | G | K | N | BRCA1 | ENST00000471181 | 0.59508 | 0.11481 | 0.2837 | 0.95464 | 0.7337 | 0.4722610328 | 0.69741 | 0.8817432523 | 0.10226 | 6.079973395 | 0 |
| 17 | 43067670 | T | A | K | I | BRCA1 | ENST00000471181 | 0.51067 | 0.26711 | 0.24737 | 0.97375 | 0.80417 | 0.5746611782 | 0.38632 | 0.9113871455 | 0.198972 | 6.079973395 | 0 |
| 17 | 43067670 | T | C | K | R | BRCA1 | ENST00000471181 | 0.32083 | 0.07553 | 0.2228 | 0.77033 | 0.73223 | 0.2531682675 | 0.25729 | 0.1750264317 | 0.127545 | 3.053697868 | 0 |
| 17 | 43067670 | T | G | K | T | BRCA1 | ENST00000471181 | 0.47118 | 0.17205 | 0.28597 | 0.9539 | 0.79334 | 0.5009249728 | 0.36252 | 0.4332506061 | 0.163403 | 6.079973395 | 0 |
| 17 | 43067671 | T | C | K | E | BRCA1 | ENST00000471181 | 0.35991 | 0.13904 | 0.26362 | 0.9492 | 0.81006 | 0.40580903 | 0.56258 | 0.8558703661 | 0.184127 | 6.079973395 | 0 |
| 17 | 43067671 | T | G | K | Q | BRCA1 | ENST00000471181 | 0.32812 | 0.09797 | 0.23187 | 0.90011 | 0.7675 | 0.382526645 | 0.5143 | 0.2402563006 | 0.169262 | 4.401643193 | 0 |
| 17 | 43067672 | T | G | R | S | BRCA1 | ENST00000471181 | 0.35875 | 0.1132 | 0.14172 | 0.95679 | 0.85117 | 0.4433944245 | 0.81472 | 0.8089458346 | 0.199476 | 5.957562343 | 0 |
| 17 | 43067673 | C | A | R | I | BRCA1 | ENST00000471181 | 0.35069 | 0.16919 | 0.15408 | 0.97557 | 0.88307 | 0.4418452795 | 0.85373 | 0.8571855426 | 0.21426 | 5.957562343 | 0 |
| 17 | 43067673 | C | T | R | K | BRCA1 | ENST00000471181 | 0.21157 | 0.09202 | 0.17458 | 0.76131 | 0.69762 | 0.3214456485 | 0.6707 | 0.161007598 | 0.0460443 | 2.647004363 | 1 |
| 17 | 43067674 | T | C | R | G | BRCA1 | ENST00000471181 | 0.42401 | 0.134 | 0.13678 | 0.9785 | 0.8873 | 0.4588882429 | 0.84781 | 0.8149478436 | 0.287651 | 5.957562343 | 0 |
| 17 | 43067676 | G | C | A | G | BRCA1 | ENST00000471181 | 0.61074 | 0.11784 | 0.35989 | 0.9597 | 0.86753 | 0.6314040233 | 0.88013 | 0.8748479486 | 0.246232 | 6.68241714 | 0 |
| 17 | 43067677 | C | A | A | S | BRCA1 | ENST00000471181 | 0.61626 | 0.14802 | 0.34101 | 0.96777 | 0.89513 | 0.6043268508 | 0.87941 | 0.2764928344 | 0.0385132 | 6.68241714 | 0 |
| 17 | 43067677 | C | G | A | P | BRCA1 | ENST00000471181 | 0.69477 | 0.47204 | 0.44045 | 0.98092 | 0.92561 | 0.8975910412 | 0.90908 | 0.9830822945 | 0.399381 | 6.68241714 | 1 |
| 17 | 43067677 | C | T | A | T | BRCA1 | ENST00000471181 | 0.66691 | 0.1818 | 0.38088 | 0.96951 | 0.86803 | 0.5640569859 | 0.88296 | 0.9221466184 | 0.247966 | 5.378161705 | 0 |
| 17 | 43067678 | A | C | F | L | BRCA1 | ENST00000471181 | 0.50498 | 0.10819 | 0.61696 | 0.96945 | 0.81124 | 0.6457981696 | 0.92654 | 0.7992803148 | 0.15732 | 6.709413449 | 0 |
| 17 | 43067678 | A | T | F | L | BRCA1 | ENST00000471181 | 0.49945 | 0.10819 | 0.61696 | 0.96945 | 0.81124 | 0.6457981696 | 0.92654 | 0.9156208634 | 0.157319 | 6.709413449 | 1 |
| 17 | 43067679 | A | C | F | C | BRCA1 | ENST00000471181 | 0.75081 | 0.4557 | 0.60231 | 0.99075 | 0.93631 | 0.7377724437 | 0.906 | 0.9846643806 | 0.27315 | 6.709413449 | 0 |
| 17 | 43067679 | A | G | F | S | BRCA1 | ENST00000471181 | 0.75145 | 0.44097 | 0.57938 | 0.99178 | 0.93939 | 0.8093198808 | 0.92845 | 0.9926355481 | 0.335217 | 6.709413449 | 1 |
| 17 | 43067679 | A | T | F | Y | BRCA1 | ENST00000471181 | 0.655 | 0.26615 | 0.6017 | 0.97437 | 0.89055 | 0.5960216363 | 0.90829 | 0.9510608912 | 0.207252 | 6.709413449 | 0 |
| 17 | 43067680 | A | C | F | V | BRCA1 | ENST00000471181 | 0.73437 | 0.20209 | 0.55594 | 0.98409 | 0.89739 | 0.6891962902 | 0.92166 | 0.9821764231 | 0.250338 | 6.709413449 | 0 |
| 17 | 43067680 | A | G | F | L | BRCA1 | ENST00000471181 | 0.55209 | 0.10819 | 0.61696 | 0.96945 | 0.84428 | 0.6457981696 | 0.9308 | 0.7800738215 | 0.0981775 | 6.709413449 | 0 |
| 17 | 43067680 | A | T | F | I | BRCA1 | ENST00000471181 | 0.70623 | 0.16218 | 0.58127 | 0.97587 | 0.86803 | 0.739226665 | 0.94171 | 0.9815266728 | 0.219139 | 6.709413449 | 0 |
| 17 | 43067681 | C | G | K | N | BRCA1 | ENST00000471181 | 0.49299 | 0.2896 | 0.42507 | 0.96409 | 0.89101 | 0.6705550881 | 0.90055 | 0.8989596963 | 0.0813673 | 6.546049954 | 0 |
| 17 | 43067682 | T | A | K | M | BRCA1 | ENST00000471181 | 0.73086 | 0.3391 | 0.33525 | 0.98721 | 0.93162 | 0.701919707 | 0.89756 | 0.9643391371 | 0.230069 | 6.546049954 | 0 |

| | | | | | | | | | | | | | | | | | | |
|---|---|---|---|---|---|---|---|---|---|---|---|---|---|---|---|---|---|---|
| 17 | 43067682 | T | C | K | R | BRCA1 | ENST00000471181 | 0.50185 | 0.13393 | 0.33988 | 0.9556 | 0.83615 | 0.4070696029 | 0.86881 | 0.7449114323 | 0.152686 | 4.326556187 | 0 |
| 17 | 43067682 | T | G | K | T | BRCA1 | ENST00000471181 | 0.69228 | 0.19251 | 0.41365 | 0.98604 | 0.92561 | 0.6510975115 | 0.89208 | 0.8973681927 | 0.229182 | 6.546049954 | 0 |
| 17 | 43067683 | T | C | K | E | BRCA1 | ENST00000471181 | 0.64563 | 0.1901 | 0.39474 | 0.95852 | 0.87148 | 0.6252325618 | 0.88222 | 0.7291494012 | 0.2274 | 6.546049954 | 0 |
| 17 | 43067683 | T | G | K | Q | BRCA1 | ENST00000471181 | 0.61261 | 0.18538 | 0.36024 | 0.96924 | 0.87877 | 0.5654233617 | 0.91197 | 0.7993603945 | 0.201299 | 6.546049954 | 0 |
| 17 | 43067685 | T | A | Y | F | BRCA1 | ENST00000471181 | 0.42825 | 0.08735 | 0.14745 | 0.91463 | 0.75211 | 0.2852496687 | 0.64722 | 0.3897509266 | 0.0950004 | 2.788047822 | 0 |
| 17 | 43067685 | T | C | Y | C | BRCA1 | ENST00000471181 | 0.66288 | 0.1922 | 0.12926 | 0.9437 | 0.81703 | 0.4193393192 | 0.82916 | 0.3093556166 | 0.137078 | 2.788047822 | 0 |
| 17 | 43067685 | T | G | Y | S | BRCA1 | ENST00000471181 | 0.48018 | 0.13114 | 0.1254 | 0.89927 | 0.74517 | 0.3956418201 | 0.55722 | 0.4821357131 | 0.140099 | 2.788047822 | 0 |
| 17 | 43067686 | A | C | Y | D | BRCA1 | ENST00000471181 | 0.48088 | 0.17061 | 0.18241 | 0.90608 | 0.73734 | 0.545414985 | 0.48855 | 0.2146231979 | 0.161669 | 2.788047822 | 0 |
| 17 | 43067686 | A | G | Y | H | BRCA1 | ENST00000471181 | 0.33559 | 0.1187 | 0.16807 | 0.78929 | 0.69846 | 0.3471075168 | 0.58844 | 0.1314341666 | 0.154414 | -1.915045918 | 0 |
| 17 | 43067686 | A | T | Y | N | BRCA1 | ENST00000471181 | 0.4742 | 0.15565 | 0.17556 | 0.90965 | 0.74235 | 0.4335744788 | 0.58134 | 0.2393526882 | 0.124426 | 2.788047822 | 0 |
| 17 | 43067688 | A | C | V | G | BRCA1 | ENST00000471181 | 0.61514 | 0.14423 | 0.30081 | 0.9819 | 0.92316 | 0.8233531709 | 0.93179 | 0.9893029332 | 0.366172 | 6.028271739 | 0 |
| 17 | 43067688 | A | G | V | A | BRCA1 | ENST00000471181 | 0.42849 | 0.11481 | 0.38986 | 0.96898 | 0.89918 | 0.6576279013 | 0.91865 | 0.9679824114 | 0.285972 | 3.53744798 | 0 |
| 17 | 43067688 | A | T | V | E | BRCA1 | ENST00000471181 | 0.62612 | 0.15111 | 0.37745 | 0.98103 | 0.92561 | 0.8843682867 | 0.91758 | 0.9862077236 | 0.38717 | 6.028271739 | 1 |
| 17 | 43067689 | C | G | V | L | BRCA1 | ENST00000471181 | 0.25031 | 0.08917 | 0.36822 | 0.95641 | 0.84321 | 0.5934893358 | 0.7926 | 0.8856687546 | 0.195181 | 6.028271739 | 0 |
| 17 | 43067689 | C | T | V | M | BRCA1 | ENST00000471181 | 0.27355 | 0.09219 | 0.39043 | 0.94952 | 0.83339 | 0.5888976874 | 0.87465 | 0.7279686332 | 0.103023 | 3.878208614 | 0 |
| 17 | 43067691 | A | C | L | R | BRCA1 | ENST00000471181 | 0.33393 | 0.13491 | 0.28011 | 0.96467 | 0.82159 | 0.5257804782 | 0.62914 | 0.4824355245 | 0.185182 | 4.313047413 | 0 |
| 17 | 43067691 | A | G | L | P | BRCA1 | ENST00000471181 | 0.34011 | 0.14931 | 0.31673 | 0.96523 | 0.82668 | 0.7721413102 | 0.61619 | 0.07468677331 | 0.191231 | 5.201788016 | 0 |
| 17 | 43067691 | A | T | L | H | BRCA1 | ENST00000471181 | 0.4523 | 0.14605 | 0.27682 | 0.96935 | 0.85117 | 0.3892861572 | 0.80871 | 0.666002214 | 0.200347 | 5.129940968 | 0 |
| 17 | 43067692 | G | A | L | F | BRCA1 | ENST00000471181 | 0.36264 | 0.09892 | 0.21358 | 0.94523 | 0.85222 | 0.2415110868 | 0.95777 | 0.5295164585 | 0.00212808 | 5.751893646 | 0 |
| 17 | 43067692 | G | C | L | V | BRCA1 | ENST00000471181 | 0.31441 | 0.09179 | 0.17154 | 0.88876 | 0.80536 | 0.2517427726 | 0.83591 | 0.233262226 | 0.166197 | 3.479228939 | 0 |
| 17 | 43067692 | G | T | L | I | BRCA1 | ENST00000471181 | 0.23148 | 0.10086 | 0.2356 | 0.91034 | 0.79516 | 0.1629640394 | 0.71594 | 0.1218275726 | 0.107474 | 2.23425245 | 0 |
| 17 | 43067693 | C | A | M | I | BRCA1 | ENST00000471181 | 0.31618 | 0.10092 | 0.18088 | 0.95129 | 0.82103 | 0.1946614492 | 0.81322 | 0.5119693279 | 0.176152 | 5.337811906 | 0 |
| 17 | 43067693 | C | G | M | I | BRCA1 | ENST00000471181 | 0.32173 | 0.10092 | 0.18088 | 0.95022 | 0.83228 | 0.1946614492 | 0.81322 | 0.8250141144 | 0.176155 | 5.337811906 | 0 |
| 17 | 43067694 | A | C | M | R | BRCA1 | ENST00000471181 | 0.37725 | 0.15425 | 0.22526 | 0.98448 | 0.88401 | 0.4869750526 | 0.9259 | 0.7879825234 | 0.223421 | 5.337811906 | 1 |
| 17 | 43067694 | A | T | M | K | BRCA1 | ENST00000471181 | 0.35797 | 0.15377 | 0.23639 | 0.98074 | 0.85894 | 0.4636050976 | 0.86538 | 0.8340230584 | 0.21106 | 5.337811906 | 1 |
| 17 | 43067695 | T | A | M | L | BRCA1 | ENST00000471181 | 0.17774 | 0.08473 | 0.18116 | 0.9534 | 0.79456 | 0.2368652493 | 0.62969 | 0.2491087317 | 0.115785 | 2.857515451 | 1 |
| 17 | 43067695 | T | C | M | V | BRCA1 | ENST00000471181 | 0.1595 | 0.09219 | 0.14017 | 0.83815 | 0.78535 | 0.2591711578 | 0.75603 | 0.05810140818 | 0.131309 | 1.902674786 | 0 |
| 17 | 43070928 | A | C | F | L | BRCA1 | ENST00000471181 | 0.10687 | 0.10632 | 0.14289 | 0.63732 | 0.70833 | 0.2860780928 | 0.54795 | 0.04060572386 | 0.054752 | 0.497100437 | 0 |
| 17 | 43070928 | A | T | F | L | BRCA1 | ENST00000471181 | 0.10379 | 0.10632 | 0.14289 | 0.63732 | 0.70833 | 0.2860780928 | 0.54795 | 0.04060572386 | 0.0547931 | 0.497100437 | 0 |
| 17 | 43070929 | A | C | F | C | BRCA1 | ENST00000471181 | 0.29462 | 0.13296 | 0.15061 | 0.73507 | 0.79998 | 0.3840029962 | 0.35094 | 0.02726667188 | 0.120711 | 4.650551067 | 0 |
| 17 | 43070929 | A | G | F | S | BRCA1 | ENST00000471181 | 0.149 | 0.18189 | 0.14463 | 0.76516 | 0.79878 | 0.3843288052 | 0.34258 | 0.01019230265 | -0.0538666 | 2.389127956 | 0 |
| 17 | 43070929 | A | T | F | Y | BRCA1 | ENST00000471181 | 0.12136 | 0.11232 | 0.15538 | 0.65065 | 0.77845 | 0.2799334435 | 0.26553 | 0.1475963145 | 0.00631667 | 3.18706005 | 0 |
| 17 | 43070930 | A | C | F | V | BRCA1 | ENST00000471181 | 0.17468 | 0.12022 | 0.10528 | 0.70485 | 0.78597 | 0.2961041358 | 0.39776 | 0.01316688662 | 0.0795176 | 2.569056739 | 0 |
| 17 | 43070930 | A | G | F | L | BRCA1 | ENST00000471181 | 0.16111 | 0.10632 | 0.14289 | 0.53338 | 0.68734 | 0.2860780928 | 0.46806 | 0.03271147981 | 0.0454031 | 0.497100437 | 0 |
| 17 | 43070930 | A | T | F | I | BRCA1 | ENST00000471181 | 0.20127 | 0.12974 | 0.14087 | 0.68847 | 0.73589 | 0.2993996545 | 0.49812 | 0.03891295195 | 0.0374717 | 4.671422081 | 0 |
| 17 | 43070931 | T | A | E | D | BRCA1 | ENST00000471181 | 0.60485 | 0.09938 | 0.34669 | 0.9404 | 0.85996 | 0.6509533563 | 0.87623 | 0.9461736083 | 0.172734 | 6.79421325 | 0 |
| 17 | 43070931 | T | G | E | D | BRCA1 | ENST00000471181 | 0.61246 | 0.09938 | 0.34669 | 0.94136 | 0.85996 | 0.6509533563 | 0.87623 | 0.9494435191 | 0.172734 | 6.79421325 | 0 |
| 17 | 43070932 | T | A | E | V | BRCA1 | ENST00000471181 | 0.80274 | 0.23489 | 0.32629 | 0.97714 | 0.91611 | 0.7291831328 | 0.88605 | 0.9950450659 | 0.279746 | 6.79421325 | 0 |
| 17 | 43070932 | T | G | E | A | BRCA1 | ENST00000471181 | 0.77521 | 0.15377 | 0.35086 | 0.9738 | 0.90711 | 0.7224270633 | 0.92536 | 0.994453609 | 0.255168 | 6.79421325 | 0 |
| 17 | 43070933 | C | G | E | Q | BRCA1 | ENST00000471181 | 0.72973 | 0.14296 | 0.34024 | 0.92914 | 0.84052 | 0.6577924376 | 0.92448 | 0.9870196581 | 0.203025 | 6.79421325 | 0 |
| 17 | 43070933 | C | T | E | K | BRCA1 | ENST00000471181 | 0.79721 | 0.10393 | 0.43171 | 0.94924 | 0.88684 | 0.8054441196 | 0.90908 | 0.9926409125 | 0.275432 | 6.79421325 | 0 |
| 17 | 43070934 | T | A | E | D | BRCA1 | ENST00000471181 | 0.45616 | 0.08253 | 0.19688 | 0.86398 | 0.80476 | 0.2555154852 | 0.78314 | 0.4511530794 | 0.20539 | 2.897130805 | 0 |
| 17 | 43070934 | T | G | E | D | BRCA1 | ENST00000471181 | 0.46252 | 0.08253 | 0.19688 | 0.86351 | 0.80476 | 0.2555154852 | 0.78314 | 0.4439899294 | 0.205405 | 2.897130805 | 0 |
| 17 | 43070935 | T | A | E | V | BRCA1 | ENST00000471181 | 0.55546 | 0.11512 | 0.13241 | 0.95432 | 0.87442 | 0.3360281136 | 0.78581 | 0.8174248338 | 0.157089 | 2.897130805 | 0 |
| 17 | 43070935 | T | C | E | G | BRCA1 | ENST00000471181 | 0.57456 | 0.12607 | 0.14319 | 0.94174 | 0.84853 | 0.2405557488 | 0.81814 | 0.6075444221 | 0.130594 | 2.897130805 | 0 |
| 17 | 43070935 | T | G | E | A | BRCA1 | ENST00000471181 | 0.51955 | 0.1077 | 0.14664 | 0.93457 | 0.83615 | 0.2476563972 | 0.76582 | 0.6075444221 | 0.152499 | 2.897130805 | 0 |
| 17 | 43070936 | C | G | E | Q | BRCA1 | ENST00000471181 | 0.41281 | 0.10162 | 0.15902 | 0.88392 | 0.82159 | 0.2088176732 | 0.77692 | 0.7081924081 | 0.189792 | 2.257597816 | 0 |
| 17 | 43070936 | C | T | E | K | BRCA1 | ENST00000471181 | 0.25958 | 0.10264 | 0.16821 | 0.72853 | 0.7337 | 0.2138857169 | 0.46616 | 0.3389938176 | 0.13681 | -2.665316335 | 0 |
| 17 | 43070938 | G | A | P | L | BRCA1 | ENST00000471181 | 0.28461 | 0.09744 | 0.10061 | 0.94225 | 0.86753 | 0.3528207885 | 0.85579 | 0.9360610247 | 0.215742 | 2.780443676 | 0 |
| 17 | 43070938 | G | C | P | R | BRCA1 | ENST00000471181 | 0.51395 | 0.20833 | 0.10505 | 0.93071 | 0.86653 | 0.3041796494 | 0.87283 | 0.7938757539 | 0.21396 | 5.479073101 | 0 |
| 17 | 43070938 | G | T | P | Q | BRCA1 | ENST00000471181 | 0.2387 | 0.25137 | 0.06783 | 0.85355 | 0.82668 | 0.1868170273 | 0.7569 | 0.477342993 | 0.172823 | 1.849059928 | 0 |
| 17 | 43070939 | G | A | P | S | BRCA1 | ENST00000471181 | 0.23098 | 0.11585 | 0.05646 | 0.92261 | 0.80238 | 0.2342347201 | 0.6963 | 0.5781190395 | 0.165467 | 2.460120968 | 0 |
| 17 | 43070939 | G | C | P | A | BRCA1 | ENST00000471181 | 0.2071 | 0.09666 | 0.06277 | 0.88844 | 0.81933 | 0.1928833194 | 0.70066 | 0.08649852127 | 0.162276 | 3.53308735 | 0 |
| 17 | 43070939 | G | T | P | T | BRCA1 | ENST00000471181 | 0.23178 | 0.10245 | 0.06624 | 0.94662 | 0.84747 | 0.2222798372 | 0.76628 | 0.3009990156 | 0.181621 | 4.897365219 | 0 |
| 17 | 43070942 | T | A | T | S | BRCA1 | ENST00000471181 | 0.58724 | 0.09187 | 0.27429 | 0.93771 | 0.862 | 0.1936491817 | 0.75652 | 0.7261965871 | 0.149149 | 4.983036749 | 0 |
| 17 | 43070942 | T | C | T | A | BRCA1 | ENST00000471181 | 0.75264 | 0.09718 | 0.25982 | 0.95762 | 0.92356 | 0.3278251904 | 0.81721 | 0.9415749311 | 0.285873 | 5.268147249 | 0 |
| 17 | 43070944 | A | C | L | R | BRCA1 | ENST00000471181 | 0.87214 | 0.21761 | 0.4674 | 0.98245 | 0.95608 | 0.8660711401 | 0.9197 | 0.9930995703 | 0.45777 | 6.794212507 | 1 |
| 17 | 43070944 | A | G | L | P | BRCA1 | ENST00000471181 | 0.87556 | 0.46179 | 0.50998 | 0.98832 | 0.95207 | 0.8763733612 | 0.94193 | 0.9964382648 | 0.460105 | 6.794212507 | 1 |
| 17 | 43070944 | A | T | L | Q | BRCA1 | ENST00000471181 | 0.86375 | 0.23312 | 0.39444 | 0.98322 | 0.95243 | 0.8150615759 | 0.93865 | 0.994125843 | 0.459321 | 6.794212507 | 1 |
| 17 | 43070945 | G | C | L | V | BRCA1 | ENST00000471181 | 0.51197 | 0.10977 | 0.31643 | 0.97659 | 0.90185 | 0.488574696 | 0.82995 | 0.9747264385 | 0.293734 | 6.794212507 | 0 |
| 17 | 43070945 | G | T | L | M | BRCA1 | ENST00000471181 | 0.53623 | 0.35014 | 0.35407 | 0.96954 | 0.82443 | 0.4229448928 | 0.79082 | 0.980718255 | 0.253172 | 6.794212507 | 0 |
| 17 | 43070947 | C | A | G | V | BRCA1 | ENST00000471181 | 0.71201 | 0.20688 | 0.43382 | 0.97448 | 0.94765 | 0.8376658614 | 0.95452 | 0.9939077497 | 0.454762 | 6.690187835 | 0 |
| 17 | 43070947 | C | G | G | A | BRCA1 | ENST00000471181 | 0.68227 | 0.11672 | 0.43326 | 0.95619 | 0.91569 | 0.7082673001 | 0.93937 | 0.9861046672 | 0.395955 | 6.690187835 | 0 |
| 17 | 43070947 | C | T | G | D | BRCA1 | ENST00000471181 | 0.71458 | 0.22834 | 0.55327 | 0.97428 | 0.94618 | 0.7740874879 | 0.95753 | 0.9856675267 | 0.431691 | 6.690187835 | 0 |
| 17 | 43070948 | C | A | G | C | BRCA1 | ENST00000471181 | 0.71962 | 0.38767 | 0.47818 | 0.97883 | 0.93786 | 0.7845389732 | 0.93902 | 0.9953488708 | 0.440441 | 6.690187835 | 0 |
| 17 | 43070948 | C | G | G | R | BRCA1 | ENST00000471181 | 0.69188 | 0.13319 | 0.54337 | 0.97354 | 0.94091 | 0.7928525054 | 0.93206 | 0.991466403 | 0.439932 | 6.690187835 | 0 |
| 17 | 43070948 | C | T | G | S | BRCA1 | ENST00000471181 | 0.66185 | 0.09137 | 0.41668 | 0.95921 | 0.90842 | 0.7114966627 | 0.93311 | 0.9801039696 | 0.410498 | 5.135726345 | 0 |
| 17 | 43070950 | G | A | S | F | BRCA1 | ENST00000471181 | 0.77593 | 0.34653 | 0.39021 | 0.98504 | 0.94243 | 0.8348538988 | 0.9726 | 0.9962857962 | 0.521509 | 6.7942122 | 1 |
| 17 | 43070950 | G | C | S | C | BRCA1 | ENST00000471181 | 0.76044 | 0.36522 | 0.35202 | 0.9818 | 0.94431 | 0.7621995184 | 0.97818 | 0.9952161312 | 0.519888 | 6.7942122 | 1 |
| 17 | 43070950 | G | T | S | Y | BRCA1 | ENST00000471181 | 0.73021 | 0.42198 | 0.41966 | 0.98503 | 0.94281 | 0.8260157866 | 0.97271 | 0.9962857962 | 0.522078 | 6.7942122 | 1 |
| 17 | 43070951 | A | G | S | P | BRCA1 | ENST00000471181 | 0.73589 | 0.26705 | 0.36473 | 0.98368 | 0.95316 | 0.9055920398 | 0.97796 | 0.9918777943 | 0.515251 | 6.7942122 | 1 |
| 17 | 43070951 | A | T | S | T | BRCA1 | ENST00000471181 | 0.64384 | 0.09724 | 0.35259 | 0.95466 | 0.92438 | 0.6636798854 | 0.95501 | 0.8307671547 | 0.359633 | 6.7942122 | 0 |
| 17 | 43070953 | A | C | V | G | BRCA1 | ENST00000471181 | 0.74778 | 0.41385 | 0.16016 | 0.92113 | 0.84694 | 0.5215579485 | 0.87974 | 0.7626087666 | 0.362611 | 5.44395931 | 0 |
| 17 | 43070953 | A | G | V | A | BRCA1 | ENST00000471181 | 0.33147 | 0.13205 | 0.20471 | 0.77672 | 0.79697 | 0.2294566669 | 0.78139 | 0.09096242276 | 0.145852 | 0.3798635454 | 0 |
| 17 | 43070954 | C | A | V | L | BRCA1 | ENST00000471181 | 0.56595 | 0.09433 | 0.18997 | 0.7794 | 0.80178 | 0.3290694033 | 0.88498 | 0.8557356596 | 0.255819 | 5.44395931 | 0 |
| 17 | 43070954 | C | G | V | L | BRCA1 | ENST00000471181 | 0.57336 | 0.09433 | 0.18997 | 0.76893 | 0.80178 | 0.3290694033 | 0.88498 | 0.8557356596 | 0.255832 | 5.44395931 | 0 |
| 17 | 43070954 | C | T | V | M | BRCA1 | ENST00000471181 | 0.65304 | 0.41031 | 0.20764 | 0.89588 | 0.81414 | 0.2919978337 | 0.93347 | 0.9070158005 | 0.201501 | 5.44395931 | 0 |
| 17 | 43070956 | A | C | V | G | BRCA1 | ENST00000471181 | 0.73717 | 0.36634 | 0.36715 | 0.98077 | 0.97178 | 0.8569547842 | 0.99492 | 0.9964752793 | 0.446579 | 6.794212638 | 0 |
| 17 | 43070956 | A | G | V | A | BRCA1 | ENST00000471181 | 0.70495 | 0.28347 | 0.47347 | 0.97756 | 0.93824 | 0.6030578637 | 0.98548 | 0.9790180922 | 0.29853 | 6.794212638 | 0 |
| 17 | 43070956 | A | T | V | E | BRCA1 | ENST00000471181 | 0.74348 | 0.40573 | 0.45918 | 0.97939 | 0.95645 | 0.8622146032 | 0.99073 | 0.99665988994 | 0.44626 | 6.794212638 | 1 |
| 17 | 43070957 | C | A | V | L | BRCA1 | ENST00000471181 | 0.64906 | 0.09816 | 0.45508 | 0.94524 | 0.90096 | 0.6471905313 | 0.98129 | 0.9659355283 | 0.2834 | 6.794212638 | 1 |
| 17 | 43070957 | C | G | V | L | BRCA1 | ENST00000471181 | 0.65736 | 0.09816 | 0.45508 | 0.94323 | 0.90096 | 0.6471905313 | 0.98129 | 0.9659355283 | 0.283401 | 6.794212638 | 1 |
| 17 | 43070958 | C | A | M | I | BRCA1 | ENST00000471181 | 0.28714 | 0.10252 | 0.19449 | 0.68882 | 0.74446 | 0.3029600591 | 0.86287 | 0.3192876577 | 0.147325 | 3.279601041 | 0 |
| 17 | 43070958 | C | G | M | I | BRCA1 | ENST00000471181 | 0.29238 | 0.10252 | 0.19449 | 0.68629 | 0.73734 | 0.3029600591 | 0.86287 | 0.4033564031 | 0.14736 | 3.279601041 | 0 |
| 17 | 43070958 | C | T | M | I | BRCA1 | ENST00000471181 | 0.29577 | 0.10252 | 0.19449 | | 0.73661 | 0.3029600591 | | 0.007210041803 | -0.195766 | 3.279601041 | 0 |
| 17 | 43070959 | A | C | M | R | BRCA1 | ENST00000471181 | 0.72556 | 0.15858 | 0.23151 | 0.93952 | 0.90362 | 0.8668831216 | 0.9502 | 0.9349498153 | 0.413836 | 6.270277205 | 1 |
| 17 | 43070959 | A | G | M | T | BRCA1 | ENST00000471181 | 0.64937 | 0.10581 | 0.29373 | 0.94149 | 0.89558 | 0.6371524249 | 0.9344 | 0.3823396266 | 0.322922 | 6.270277205 | 0 |
| 17 | 43070959 | A | T | M | K | BRCA1 | ENST00000471181 | 0.69163 | 0.15245 | 0.25699 | 0.94827 | 0.90185 | 0.8268712295 | 0.94685 | 0.9609320164 | 0.414296 | 6.270277205 | 1 |
| 17 | 43070960 | T | A | M | L | BRCA1 | ENST00000471181 | 0.21264 | 0.08473 | 0.18233 | 0.61926 | 0.80298 | 0.3857975273 | 0.47488 | 0.09452105314 | 0.0977433 | 3.24682372 | 0 |
| 17 | 43070960 | T | C | M | V | BRCA1 | ENST00000471181 | 0.28773 | 0.09247 | 0.12864 | 0.92302 | 0.79637 | 0.37747408 | 0.70284 | 0.2650720775 | 0.186399 | 3.513545644 | 0 |
| 17 | 43070960 | T | G | M | L | BRCA1 | ENST00000471181 | 0.21673 | 0.08473 | 0.18233 | 0.63278 | 0.80298 | 0.3857975273 | 0.47488 | 0.09452105314 | 0.0977408 | 3.24682372 | 0 |
| 17 | 43070962 | G | A | S | F | BRCA1 | ENST00000471181 | 0.74722 | 0.33346 | 0.35745 | 0.98039 | 0.93319 | 0.5316627893 | 0.97208 | 0.9899010062 | 0.4266 | 4.158007608 | 0 |
| 17 | 43070962 | G | C | S | C | BRCA1 | ENST00000471181 | 0.73157 | 0.3996 | 0.32749 | 0.97064 | 0.88022 | 0.471212644 | 0.98234 | 0.9627897739 | 0.282246 | 6.279741005 | 0 |
| 17 | 43070962 | G | T | S | Y | BRCA1 | ENST00000471181 | 0.70118 | 0.4529 | 0.38137 | 0.97924 | 0.90842 | 0.5135226463 | 0.96839 | 0.9838535786 | 0.279513 | 6.279741005 | 0 |
| 17 | 43070963 | A | C | S | A | BRCA1 | ENST00000471181 | 0.66874 | 0.12164 | 0.30863 | 0.93489 | 0.89055 | 0.2416230591 | 0.97275 | 0.971840024 | 0.252199 | 5.549621169 | 0 |
| 17 | 43070963 | A | G | S | P | BRCA1 | ENST00000471181 | 0.71995 | 0.40229 | 0.3481 | 0.97603 | 0.92964 | 0.8106621849 | 0.98522 | 0.9930143356 | 0.416285 | 6.279741005 | 1 |
| 17 | 43070963 | A | T | S | T | BRCA1 | ENST00000471181 | 0.62863 | 0.12016 | 0.3345 | 0.9394 | 0.87877 | 0.305284954 | 0.97079 | 0.9742385745 | 0.249343 | 6.279741005 | 0 |
| 17 | 43070964 | C | A | M | I | BRCA1 | ENST00000471181 | 0.05441 | 0.10066 | 0.23882 | 0.5942 | 0.78967 | 0.2279927233 | 0.37788 | 0.03400951162 | 0.0816137 | -1.148449709 | 0 |
| 17 | 43070964 | C | G | M | I | BRCA1 | ENST00000471181 | 0.05687 | 0.10066 | 0.23882 | 0.5952 | 0.78967 | 0.2279927233 | 0.37788 | 0.03807189352 | 0.0816186 | -1.148449709 | 0 |
| 17 | 43070964 | C | T | M | I | BRCA1 | ENST00000471181 | 0.05849 | 0.10066 | 0.23882 | 0.5942 | 0.79028 | 0.2279927233 | 0.37788 | 0.03346100502 | 0.081558 | -1.148449709 | 0 |
| 17 | 43070965 | A | C | M | R | BRCA1 | ENST00000471181 | 0.26581 | 0.14072 | 0.25471 | 0.92332 | 0.83004 | 0.7320023803 | 0.8701 | 0.2215554267 | 0.18502 | 4.105919849 | 0 |

| | | | | | | | | | | | | | | | | | |
|---|---|---|---|---|---|---|---|---|---|---|---|---|---|---|---|---|---|
| 17 | 43070965 | A | G | M | T | BRCA1 | ENST00000471181 | 0.31904 | 0.10638 | 0.309 | 0.94662 | 0.82216 | 0.486201754 | 0.835 | 0.1100118393 | 0.0592197 | 4.105919849 | 0 |
| 17 | 43070965 | A | T | M | K | BRCA1 | ENST00000471181 | 0.23073 | 0.14414 | 0.26915 | 0.82729 | 0.80118 | 0.7011915498 | 0.78175 | 0.07553444678 | 0.174976 | 3.567618328 | 0 |
| 17 | 43070966 | T | A | M | L | BRCA1 | ENST00000471181 | 0.12901 | 0.08499 | 0.22545 | 0.57671 | 0.78906 | 0.2714703579 | 0.4665 | 0.107176654 | 0.0580379 | 1.394243917 | 0 |
| 17 | 43070966 | T | C | M | V | BRCA1 | ENST00000471181 | 0.10577 | 0.09269 | 0.18203 | 0.67751 | 0.79089 | 0.284017409 | 0.79117 | 0.03905834258 | 0.129236 | 1.04654723 | 0 |
| 17 | 43070966 | T | G | M | L | BRCA1 | ENST00000471181 | 0.13233 | 0.08499 | 0.22545 | 0.57627 | 0.78967 | 0.2714703579 | 0.4665 | 0.107176654 | 0.0580289 | 1.394243917 | 0 |
| 17 | 43070967 | T | A | R | S | BRCA1 | ENST00000471181 | 0.27459 | 0.17961 | 0.08199 | 0.87281 | 0.85378 | 0.3526649279 | 0.66706 | 0.2234081551 | 0.150355 | 6.110473082 | 0 |
| 17 | 43070967 | T | G | R | S | BRCA1 | ENST00000471181 | 0.27952 | 0.17961 | 0.08199 | 0.86821 | 0.85378 | 0.3526649279 | 0.66706 | 0.1672692768 | 0.150358 | 6.110473082 | 0 |
| 17 | 43070968 | C | A | R | I | BRCA1 | ENST00000471181 | 0.34353 | 0.1678 | 0.09744 | 0.87587 | 0.83888 | 0.3999204805 | 0.6926 | 0.4092948051 | 0.166808 | 6.110473082 | 0 |
| 17 | 43070968 | C | G | R | T | BRCA1 | ENST00000471181 | 0.23503 | 0.24889 | 0.10764 | 0.86913 | 0.8543 | 0.3717576134 | 0.68063 | 0.218770207 | 0.134725 | 5.520232321 | 0 |
| 17 | 43070968 | C | T | R | K | BRCA1 | ENST00000471181 | 0.14041 | 0.09304 | 0.10788 | 0.69844 | 0.82159 | 0.2593346613 | 0.49678 | 0.02993948199 | 0.096935 | 3.380322069 | 0 |
| 17 | 43070969 | T | C | R | G | BRCA1 | ENST00000471181 | 0.19953 | 0.19932 | 0.08553 | 0.90219 | 0.84159 | 0.3742012697 | 0.59015 | 0.05769517108 | 0.152628 | 2.903862667 | 0 |
| 17 | 43070970 | T | A | K | N | BRCA1 | ENST00000471181 | 0.23499 | 0.10668 | 0.11603 | 0.92469 | 0.7609 | 0.3377249943 | 0.72707 | 0.4705256224 | 0.0808639 | 4.603129186 | 0 |
| 17 | 43070970 | T | G | K | N | BRCA1 | ENST00000471181 | 0.23939 | 0.10668 | 0.11603 | 0.9243 | 0.7609 | 0.3377249943 | 0.72707 | 0.4705256224 | 0.0808612 | 4.603129186 | 0 |
| 17 | 43070971 | T | A | K | I | BRCA1 | ENST00000471181 | 0.3327 | 0.16431 | 0.10111 | 0.95309 | 0.82159 | 0.399851226 | 0.63409 | 0.7462979555 | 0.130724 | 5.663802227 | 0 |
| 17 | 43070971 | T | C | K | R | BRCA1 | ENST00000471181 | 0.08705 | 0.07473 | 0.0771 | 0.62485 | 0.825 | 0.2162307588 | 0.32801 | 0.02585477568 | 0.0825577 | 1.704201906 | 0 |
| 17 | 43070971 | T | G | K | T | BRCA1 | ENST00000471181 | 0.16391 | 0.11169 | 0.12171 | 0.89394 | 0.83283 | 0.3796607412 | 0.63593 | 0.2029763013 | 0.126533 | 4.637356165 | 0 |
| 17 | 43070972 | T | C | K | E | BRCA1 | ENST00000471181 | 0.21577 | 0.12824 | 0.10483 | 0.8788 | 0.82159 | 0.3425476105 | 0.57135 | 0.06813939457 | 0.103563 | 5.081852268 | 0 |
| 17 | 43070972 | T | G | K | Q | BRCA1 | ENST00000471181 | 0.14315 | 0.09797 | 0.07876 | 0.81634 | 0.8233 | 0.2804686272 | 0.57219 | 0.08415193111 | 0.157095 | 3.31101373 | 0 |
| 17 | 43070973 | G | C | N | K | BRCA1 | ENST00000471181 | 0.09371 | 0.11101 | 0.24019 | 0.69207 | 0.72926 | 0.2368888115 | 0.54598 | 0.02406061429 | 0.0149554 | 3.149234535 | 0 |
| 17 | 43070973 | G | T | N | K | BRCA1 | ENST00000471181 | 0.09132 | 0.11101 | 0.24019 | 0.68859 | 0.73661 | 0.2368888115 | 0.54598 | 0.02901271738 | 0.0149715 | 3.149234535 | 0 |
| 17 | 43070974 | T | A | N | I | BRCA1 | ENST00000471181 | 0.15712 | 0.11752 | 0.21187 | 0.72563 | 0.84428 | 0.3297830321 | 0.44934 | 0.05163758234 | 0.0644893 | 4.058271298 | 0 |
| 17 | 43070974 | T | C | N | S | BRCA1 | ENST00000471181 | 0.10562 | 0.07333 | 0.17834 | 0.68793 | 0.74658 | 0.1852510265 | 0.59492 | 0.02466535538 | 0.0127173 | 2.782984287 | 0 |
| 17 | 43070974 | T | G | N | T | BRCA1 | ENST00000471181 | 0.13646 | 0.09369 | 0.21194 | 0.85017 | 0.84535 | 0.2498080524 | 0.68695 | 0.07912257314 | 0.0517244 | 4.194525973 | 0 |
| 17 | 43070975 | T | A | N | Y | BRCA1 | ENST00000471181 | 0.35019 | 0.26975 | 0.21885 | 0.93685 | 0.85064 | 0.2871085931 | 0.74837 | 0.4539555849 | 0.116708 | 5.614210821 | 0 |
| 17 | 43070975 | T | C | N | D | BRCA1 | ENST00000471181 | 0.13894 | 0.12795 | 0.23217 | 0.73599 | 0.71627 | 0.2517956121 | 0.60365 | 0.04425867647 | 0.0188902 | 3.815067803 | 0 |
| 17 | 43070975 | T | G | N | H | BRCA1 | ENST00000471181 | 0.23511 | 0.09583 | 0.20916 | 0.8906 | 0.80713 | 0.2633816516 | 0.79201 | 0.3994719088 | 0.0501336 | 3.448016218 | 0 |
| 17 | 43070977 | A | C | V | G | BRCA1 | ENST00000471181 | 0.09194 | 0.12055 | 0.14396 | 0.78697 | 0.82387 | 0.3077111761 | 0.28455 | 0.09373340756 | 0.0561715 | 2.631498369 | 0 |
| 17 | 43070977 | A | G | V | A | BRCA1 | ENST00000471181 | 0.0545 | 0.12507 | 0.18018 | 0.77631 | 0.82613 | 0.2394337813 | 0.37686 | 0.04193964058 | 0.0746562 | 2.048449721 | 0 |
| 17 | 43070977 | A | T | V | D | BRCA1 | ENST00000471181 | 0.12783 | 0.24844 | 0.19159 | 0.88482 | 0.83339 | 0.4890649448 | 0.31499 | 0.05862213703 | 0.115123 | 5.437057859 | 0 |
| 17 | 43070978 | C | A | V | F | BRCA1 | ENST00000471181 | 0.18411 | 0.22898 | 0.16765 | 0.8775 | 0.82948 | 0.4065380823 | 0.82728 | 0.1288475301 | 0.106439 | 5.437057859 | 0 |
| 17 | 43070978 | C | G | V | L | BRCA1 | ENST00000471181 | 0.10531 | 0.102 | 0.16235 | 0.81038 | 0.82103 | 0.2545434017 | 0.73809 | 0.0661713928 | 0.112849 | 5.437057859 | 0 |
| 17 | 43070978 | C | T | V | I | BRCA1 | ENST00000471181 | 0.0905 | 0.07497 | 0.12766 | 0.72094 | 0.81761 | 0.173917837 | 0.46685 | 0.1089373156 | 0.0559682 | 3.258591502 | 0 |
| 17 | 43070979 | C | A | R | S | BRCA1 | ENST00000471181 | 0.17158 | 0.13491 | 0.05256 | 0.92249 | 0.84482 | 0.3342201558 | 0.57519 | 0.2367749214 | 0.108533 | 2.970310998 | 0 |
| 17 | 43070979 | C | G | R | S | BRCA1 | ENST00000471181 | 0.17532 | 0.13491 | 0.05256 | 0.92292 | 0.84482 | 0.3342201558 | 0.57519 | 0.1250923723 | 0.108556 | 2.970310998 | 0 |
| 17 | 43070980 | C | G | R | T | BRCA1 | ENST00000471181 | 0.11919 | 0.13713 | 0.0662 | 0.74397 | 0.83116 | 0.3592290154 | 0.54683 | 0.008449902017 | -0.132548 | 2.057739096 | 0 |
| 17 | 43070980 | C | T | R | K | BRCA1 | ENST00000471181 | 0.10285 | 0.09327 | 0.06579 | 0.7161 | 0.85326 | 0.253830186 | 0.62494 | 0.03934157635 | 0.0950261 | 2.906982823 | 0 |
| 17 | 43070981 | T | A | R | W | BRCA1 | ENST00000471181 | 0.43829 | 0.41278 | 0.07841 | 0.94921 | 0.84052 | 0.4215077201 | 0.7901 | 0.6010514239 | 0.11434 | 5.334917839 | 0 |
| 17 | 43070981 | T | C | R | G | BRCA1 | ENST00000471181 | 0.17511 | 0.11924 | 0.05359 | 0.81542 | 0.86951 | 0.3086737577 | 0.57616 | 0.03183675232 | 0.0949008 | 3.352858554 | 0 |
| 17 | 43070982 | T | A | E | D | BRCA1 | ENST00000471181 | 0.22366 | 0.12179 | 0.03439 | 0.73091 | 0.83834 | 0.2631119321 | 0.53231 | 0.0609038706 | 0.136126 | 3.299582186 | 0 |
| 17 | 43070982 | T | G | E | D | BRCA1 | ENST00000471181 | 0.22789 | 0.12179 | 0.03439 | 0.73251 | 0.83725 | 0.2631119321 | 0.53231 | 0.05953806973 | 0.136126 | 3.299582186 | 0 |
| 17 | 43070983 | T | A | E | V | BRCA1 | ENST00000471181 | 0.24401 | 0.11146 | 0.02002 | 0.90469 | 0.84106 | 0.4015639158 | 0.66301 | 0.148554042 | 0.149535 | 2.821601642 | 0 |
| 17 | 43070983 | T | C | E | G | BRCA1 | ENST00000471181 | 0.15651 | 0.13995 | 0.02562 | 0.79479 | 0.79577 | 0.2932611333 | 0.59282 | 0.1261317879 | 0.079537 | 2.337566174 | 0 |
| 17 | 43070983 | T | G | E | A | BRCA1 | ENST00000471181 | 0.16353 | 0.12788 | 0.0282 | 0.86181 | 0.82892 | 0.3365901922 | 0.63539 | 0.08019827589 | 0.118838 | 3.620550919 | 0 |
| 17 | 43070984 | C | G | E | Q | BRCA1 | ENST00000471181 | 0.1253 | 0.10218 | 0.02827 | 0.76367 | 0.85945 | 0.3173774921 | 0.63481 | 0.0479525913 | 0.142367 | 3.732091117 | 0 |
| 17 | 43070984 | C | T | E | K | BRCA1 | ENST00000471181 | 0.11803 | 0.10393 | 0.04049 | 0.61492 | 0.84159 | 0.309564179 | 0.50341 | 0.03177763149 | 0.10668 | 1.407890632 | 0 |
| 17 | 43070986 | G | A | T | I | BRCA1 | ENST00000471181 | 0.08536 | 0.12634 | 0.17459 | 0.79098 | 0.84428 | 0.1907867896 | 0.50166 | 0.05263416568 | 0.102502 | 4.213271808 | 0 |
| 17 | 43070986 | G | C | T | R | BRCA1 | ENST00000471181 | 0.15852 | 0.152 | 0.13358 | 0.83022 | 0.85482 | 0.3004814106 | 0.52289 | 0.07073947787 | 0.120582 | 4.404189877 | 0 |
| 17 | 43070986 | G | T | T | K | BRCA1 | ENST00000471181 | 0.07392 | 0.11279 | 0.14471 | 0.75971 | 0.83505 | 0.2916215401 | 0.40277 | 0.01738540618 | 0.0936199 | 2.987022268 | 0 |
| 17 | 43070987 | T | A | T | S | BRCA1 | ENST00000471181 | 0.0778 | 0.0855 | 0.10745 | 0.83924 | 0.82273 | 0.1767334627 | 0.36201 | 0.06196664936 | 0.0506091 | 5.329163356 | 0 |
| 17 | 43070987 | T | C | T | A | BRCA1 | ENST00000471181 | 0.0718 | 0.09334 | 0.09737 | 0.81153 | 0.82724 | 0.1865155105 | 0.34923 | 0.04045096738 | 0.111776 | 2.456263972 | 0 |
| 17 | 43070987 | T | G | T | P | BRCA1 | ENST00000471181 | 0.09604 | 0.26684 | 0.11434 | 0.88078 | 0.84853 | 0.2863017207 | 0.3663 | 0.05338700487 | 0.103304 | 3.385527118 | 0 |
| 17 | 43070989 | G | A | S | L | BRCA1 | ENST00000471181 | 0.4824 | 0.10534 | 0.15092 | 0.87146 | 0.7872 | 0.2791240785 | 0.83465 | 0.9404530525 | 0.191269 | 3.991298529 | 0 |
| 17 | 43070990 | A | C | S | A | BRCA1 | ENST00000471181 | 0.27928 | 0.11533 | 0.1382 | 0.76024 | 0.8278 | 0.1681666812 | 0.80932 | 0.1623328151 | 0.110123 | 4.807565893 | 0 |
| 17 | 43070990 | A | G | S | P | BRCA1 | ENST00000471181 | 0.29183 | 0.11931 | 0.15909 | 0.83525 | 0.83888 | 0.2368023432 | 0.70401 | 0.06366592191 | 0.148138 | 2.890425377 | 0 |
| 17 | 43070990 | A | T | S | T | BRCA1 | ENST00000471181 | 0.35619 | 0.08898 | 0.15544 | 0.82512 | 0.84106 | 0.2295909895 | 0.83162 | 0.2345938055 | 0.184741 | 3.896409343 | 0 |
| 17 | 43070992 | G | A | A | V | BRCA1 | ENST00000471181 | 0.42424 | 0.09269 | 0.03277 | 0.83927 | 0.80476 | 0.205817915 | 0.8232 | 0.6539843082 | 0.171863 | 1.831722245 | 0 |
| 17 | 43070992 | G | C | A | G | BRCA1 | ENST00000471181 | 0.31369 | 0.12137 | 0.03232 | 0.8021 | 0.79456 | 0.2104533416 | 0.77607 | 0.4828009307 | 0.12438 | 2.803380693 | 0 |
| 17 | 43070992 | G | T | A | D | BRCA1 | ENST00000471181 | 0.43883 | 0.2131 | 0.05569 | 0.85462 | 0.80595 | 0.2706461064 | 0.80785 | 0.5997284651 | 0.185023 | 2.803380693 | 0 |
| 17 | 43070993 | C | G | A | P | BRCA1 | ENST00000471181 | 0.18771 | 0.12382 | 0.04883 | 0.84933 | 0.81182 | 0.2213974168 | 0.39173 | 0.05972462902 | 0.124268 | -0.1407175741 | 0 |
| 17 | 43070993 | C | T | A | T | BRCA1 | ENST00000471181 | 0.18726 | 0.09208 | 0.03934 | 0.83632 | 0.81414 | 0.1757675623 | 0.39128 | 0.08629737226 | 0.140028 | 0.9802357453 | 0 |
| 17 | 43070995 | G | A | T | I | BRCA1 | ENST00000471181 | 0.03742 | 0.09892 | 0.11135 | 0.66628 | 0.8367 | 0.156426048 | 0.35317 | 0.01993471757 | 0.0691126 | 0.3907725818 | 0 |
| 17 | 43070995 | G | C | T | R | BRCA1 | ENST00000471181 | 0.14474 | 0.16211 | 0.09689 | 0.87874 | 0.85012 | 0.2151412359 | 0.59812 | 0.08896785975 | -0.0264031 | 4.707391241 | 0 |
| 17 | 43070995 | G | T | T | K | BRCA1 | ENST00000471181 | 0.14239 | 0.14207 | 0.10242 | 0.86172 | 0.86653 | 0.2251006683 | 0.54014 | 0.1104049605 | 0.100043 | 2.08643806 | 0 |
| 17 | 43070996 | T | A | T | S | BRCA1 | ENST00000471181 | 0.07986 | 0.09897 | 0.07918 | 0.78704 | 0.79938 | 0.1267493035 | 0.39917 | 0.0484022091 | 0.0688996 | 4.707391241 | 0 |
| 17 | 43070996 | T | C | T | A | BRCA1 | ENST00000471181 | 0.04601 | 0.08989 | 0.07556 | 0.64151 | 0.80653 | 0.1598157056 | 0.34979 | 0.03000065699 | 0.0614707 | 1.817947427 | 0 |
| 17 | 43070996 | T | G | T | P | BRCA1 | ENST00000471181 | 0.12171 | 0.23951 | 0.08653 | 0.85369 | 0.83004 | 0.1714941487 | 0.55376 | 0.1112570252 | 0.095723 | 4.053464432 | 0 |
| 17 | 43070997 | C | A | L | F | BRCA1 | ENST00000471181 | 0.07645 | 0.1002 | 0.07578 | 0.84139 | 0.83725 | 0.1188539013 | 0.77339 | 0.04091932281 | 0.0868691 | 2.354726146 | 0 |
| 17 | 43070997 | C | G | L | F | BRCA1 | ENST00000471181 | 0.07938 | 0.1002 | 0.07578 | 0.83904 | 0.83779 | 0.1188539013 | 0.77339 | 0.03093664476 | 0.0868914 | 2.354726146 | 0 |
| 17 | 43070998 | A | C | L | W | BRCA1 | ENST00000471181 | 0.30218 | 0.44877 | 0.09356 | 0.91376 | 0.7675 | 0.2065688358 | 0.83203 | 0.3894628849 | 0.0895849 | 4.99186041 | 0 |
| 17 | 43070998 | A | G | L | S | BRCA1 | ENST00000471181 | 0.12199 | 0.1932 | 0.08672 | 0.86564 | 0.77591 | 0.2093239929 | 0.65597 | 0.07689037867 | 0.0539208 | 2.386157056 | 0 |
| 17 | 43070999 | A | C | L | V | BRCA1 | ENST00000471181 | 0.01773 | 0.09247 | 0.05774 | 0.79515 | 0.82613 | 0.1470661789 | 0.21205 | 0.02184124105 | 0.0585782 | 0.4524539675 | 0 |
| 17 | 43070999 | A | T | L | M | BRCA1 | ENST00000471181 | 0.06219 | 0.27716 | 0.07192 | 0.85642 | 0.8199 | 0.1572086422 | 0.19296 | 0.0580947484 | 0.0223209 | 4.345380371 | 0 |
| 17 | 43071000 | T | A | E | D | BRCA1 | ENST00000471181 | 0.22922 | 0.10659 | 0.0915 | 0.84079 | 0.83615 | 0.1795363915 | 0.61193 | 0.1200427339 | 0.13687 | 4.319628738 | 0 |
| 17 | 43071000 | T | G | E | D | BRCA1 | ENST00000471181 | 0.23355 | 0.10659 | 0.0915 | 0.83108 | 0.83615 | 0.1795363915 | 0.61193 | 0.1190301234 | 0.136872 | 4.319628738 | 0 |
| 17 | 43071001 | T | A | E | V | BRCA1 | ENST00000471181 | 0.40121 | 0.17814 | 0.07221 | 0.91916 | 0.80713 | 0.3495728487 | 0.67389 | 0.2120187653 | 0.141727 | 5.788382628 | 0 |
| 17 | 43071001 | T | C | E | G | BRCA1 | ENST00000471181 | 0.20986 | 0.13557 | 0.07384 | 0.78997 | 0.76289 | 0.2590492841 | 0.60303 | 0.1129573985 | 0.0665103 | 2.52876466 | 0 |
| 17 | 43071001 | T | G | E | A | BRCA1 | ENST00000471181 | 0.26465 | 0.1147 | 0.08734 | 0.88528 | 0.80772 | 0.2620186634 | 0.65214 | 0.186812113 | 0.101557 | 4.177958297 | 0 |
| 17 | 43071002 | C | G | E | Q | BRCA1 | ENST00000471181 | 0.27971 | 0.12561 | 0.0818 | 0.85929 | 0.78035 | 0.2589269147 | 0.57538 | 0.1839993149 | 0.134976 | 5.788382628 | 0 |
| 17 | 43071002 | C | T | E | K | BRCA1 | ENST00000471181 | 0.15324 | 0.10314 | 0.12852 | 0.74884 | 0.75552 | 0.2577335857 | 0.4452 | 0.09748895174 | 0.110558 | 2.885999119 | 0 |
| 17 | 43071004 | G | A | P | L | BRCA1 | ENST00000471181 | 0.1865 | 0.09821 | 0.09886 | 0.9447 | 0.8278 | 0.2742403739 | 0.90452 | 0.2312545776 | 0.206619 | 5.87618231 | 0 |
| 17 | 43071004 | G | C | P | R | BRCA1 | ENST00000471181 | 0.37493 | 0.32929 | 0.09687 | 0.94364 | 0.78967 | 0.2497520181 | 0.88843 | 0.5334623881 | 0.152104 | 6.476248728 | 0 |
| 17 | 43071004 | G | T | P | Q | BRCA1 | ENST00000471181 | 0.36314 | 0.24871 | 0.07009 | 0.94583 | 0.77972 | 0.2568224115 | 0.88494 | 0.555571504 | 0.125753 | 6.476248728 | 0 |
| 17 | 43071005 | G | A | P | S | BRCA1 | ENST00000471181 | 0.41422 | 0.21549 | 0.0666 | 0.91254 | 0.80358 | 0.2377194666 | 0.83873 | 0.3302540183 | 0.137204 | 3.770420626 | 0 |
| 17 | 43071005 | G | C | P | A | BRCA1 | ENST00000471181 | 0.36292 | 0.17539 | 0.06664 | 0.88643 | 0.80889 | 0.2024193477 | 0.87295 | 0.2999010569 | 0.116018 | 6.476248728 | 0 |
| 17 | 43071005 | G | T | P | T | BRCA1 | ENST00000471181 | 0.39428 | 0.29149 | 0.08326 | 0.90706 | 0.82613 | 0.2681472833 | 0.87451 | 0.6324163079 | 0.161536 | 5.184792487 | 0 |
| 17 | 43071006 | C | A | K | N | BRCA1 | ENST00000471181 | 0.08516 | 0.10753 | 0.19408 | 0.77854 | 0.71938 | 0.2658642937 | 0.37955 | 0.03659600096 | 0.024722 | 3.054172841 | 0 |
| 17 | 43071006 | C | G | K | N | BRCA1 | ENST00000471181 | 0.08821 | 0.10753 | 0.19408 | 0.76702 | 0.7285 | 0.2658642937 | 0.37955 | 0.0377162787 | 0.0247345 | 3.054172841 | 0 |
| 17 | 43071007 | T | A | K | M | BRCA1 | ENST00000471181 | 0.34006 | 0.27147 | 0.14246 | 0.9338 | 0.8124 | 0.3697235627 | 0.66553 | 0.4339178801 | 0.112316 | 6.027488709 | 0 |
| 17 | 43071007 | T | C | K | R | BRCA1 | ENST00000471181 | 0.19245 | 0.0753 | 0.15185 | 0.86549 | 0.79878 | 0.1982896761 | 0.56794 | 0.08786828578 | 0.101897 | 4.991615031 | 0 |
| 17 | 43071007 | T | G | K | T | BRCA1 | ENST00000471181 | 0.19693 | 0.13763 | 0.19509 | 0.86411 | 0.79818 | 0.2831093057 | 0.55819 | 0.03373385023 | 0.0683816 | 3.669309928 | 0 |
| 17 | 43071008 | T | C | K | E | BRCA1 | ENST00000471181 | 0.20849 | 0.1595 | 0.17936 | 0.85353 | 0.79089 | 0.2752116725 | 0.595 | 0.09742783215 | 0.123922 | 6.027488709 | 0 |
| 17 | 43071008 | T | G | K | Q | BRCA1 | ENST00000471181 | 0.29173 | 0.20155 | 0.16272 | 0.90204 | 0.79697 | 0.2704048792 | 0.61494 | 0.2032162696 | 0.146302 | 5.283190695 | 0 |
| 17 | 43071009 | C | A | E | D | BRCA1 | ENST00000471181 | 0.05696 | 0.09995 | 0.08829 | 0.72181 | 0.83228 | 0.1752020273 | 0.52574 | 0.01401276048 | 0.0695957 | 2.404952313 | 0 |
| 17 | 43071009 | C | G | E | D | BRCA1 | ENST00000471181 | 0.05952 | 0.09995 | 0.08829 | 0.72413 | 0.83228 | 0.1752020273 | 0.52574 | 0.01401276048 | 0.069609 | 2.404952313 | 0 |
| 17 | 43071010 | T | A | E | V | BRCA1 | ENST00000471181 | 0.20218 | 0.18416 | 0.07245 | 0.83768 | 0.84641 | 0.3423004618 | 0.58988 | 0.08688140724 | 0.128503 | 3.796753743 | 0 |
| 17 | 43071010 | T | C | E | G | BRCA1 | ENST00000471181 | 0.15037 | 0.12788 | 0.08147 | 0.77881 | 0.77845 | 0.2604130435 | 0.58065 | 0.05648664385 | 0.0419244 | 2.03229065 | 0 |
| 17 | 43071010 | T | G | E | A | BRCA1 | ENST00000471181 | 0.13101 | 0.11638 | 0.08754 | 0.75145 | 0.82668 | 0.2679397125 | 0.61912 | 0.03181308639 | 0.0631221 | 4.40365509 | 0 |
| 17 | 43071011 | C | G | E | Q | BRCA1 | ENST00000471181 | 0.06732 | 0.09219 | 0.08886 | 0.62047 | 0.81182 | 0.2725377232 | 0.53385 | 0.2254919261 | 0.0933957 | 4.526216509 | 0 |
| 17 | 43071011 | C | T | E | K | BRCA1 | ENST00000471181 | 0.07287 | 0.10393 | 0.12127 | 0.66824 | 0.79395 | 0.2552744095 | 0.4953 | 0.2260139734 | 0.0643285 | 1.176491587 | 0 |
| 17 | 43071012 | C | A | R | S | BRCA1 | ENST00000471181 | 0.07239 | 0.12275 | 0.01238 | 0.85011 | 0.84482 | 0.2401315183 | 0.35551 | 0.4267455041 | -0.0497124 | 3.783264683 | 0 |
| 17 | 43071012 | C | G | R | S | BRCA1 | ENST00000471181 | 0.07525 | 0.12275 | 0.01238 | 0.8338 | 0.84482 | 0.2401315183 | 0.35551 | 0.1806913167 | 0.0766619 | 3.783264683 | 0 |

| | | | | | | | | | | | | | | | | | |
|---|---|---|---|---|---|---|---|---|---|---|---|---|---|---|---|---|---|
| 17 | 43071013 | C | G | R | T | BRCA1 | ENST00000471181 | 0.05524 | 0.13741 | 0.01953 | 0.85338 | 0.83004 | 0.2264655293 | 0.26553 | 0.1149087101 | 0.049404 | 3.149847465 | 0 |
| 17 | 43071013 | C | T | R | K | BRCA1 | ENST00000471181 | 0.03806 | 0.09327 | 0.01578 | 0.63732 | 0.8124 | 0.2089567331 | 0.32085 | 0.02388435873 | 0.0219401 | 1.477072739 | 0 |
| 17 | 43071014 | T | C | R | G | BRCA1 | ENST00000471181 | 0.25702 | 0.13936 | 0.0128 | 0.89835 | 0.84588 | 0.2595729415 | 0.45765 | 0.2268367708 | 0.11329 | 3.723156332 | 0 |
| 17 | 43071015 | G | T | S | R | BRCA1 | ENST00000471181 | 0.36811 | 0.41067 | 0.14926 | 0.87783 | 0.77845 | 0.2523227948 | 0.75991 | 0.5891799331 | 0.160767 | 3.86462505 | 0 |
| 17 | 43071016 | C | A | S | I | BRCA1 | ENST00000471181 | 0.46432 | 0.3164 | 0.14414 | 0.93644 | 0.81933 | 0.315408606 | 0.65199 | 0.4458212256 | 0.170679 | 5.947457164 | 0 |
| 17 | 43071016 | C | G | S | T | BRCA1 | ENST00000471181 | 0.30213 | 0.29141 | 0.13059 | 0.91068 | 0.78348 | 0.19351384 | 0.66526 | 0.2906372547 | 0.0932293 | 4.695151697 | 0 |
| 17 | 43071016 | C | T | S | N | BRCA1 | ENST00000471181 | 0.27081 | 0.33693 | 0.14967 | 0.86387 | 0.67933 | 0.1601895616 | 0.64517 | 0.08260722895 | 0.04429 | 3.154087392 | 0 |
| 17 | 43071017 | T | A | S | C | BRCA1 | ENST00000471181 | 0.52453 | 0.41578 | 0.1267 | 0.90833 | 0.75143 | 0.252024707 | 0.80743 | 0.3749583903 | 0.124807 | 5.947457164 | 0 |
| 17 | 43071017 | T | C | S | G | BRCA1 | ENST00000471181 | 0.24743 | 0.08735 | 0.11017 | 0.74611 | 0.73296 | 0.1847413881 | 0.71175 | 0.0458585023 | 0.0612542 | 3.21904541 | 0 |
| 17 | 43071017 | T | G | S | R | BRCA1 | ENST00000471181 | 0.35476 | 0.41067 | 0.14926 | 0.90494 | 0.7727 | 0.2523227948 | 0.72849 | 0.5363667011 | 0.149946 | 3.86462505 | 0 |
| 17 | 43071019 | A | C | V | G | BRCA1 | ENST00000471181 | 0.12017 | 0.12022 | 0.06617 | 0.92172 | 0.87345 | 0.2823292592 | 0.62543 | 0.2312387079 | 0.143799 | 2.945450974 | 0 |
| 17 | 43071019 | A | G | V | A | BRCA1 | ENST00000471181 | 0.10669 | 0.11856 | 0.09994 | 0.83674 | 0.8543 | 0.2032992617 | 0.86103 | 0.08056306094 | 0.121247 | 2.160829379 | 0 |
| 17 | 43071019 | A | T | V | E | BRCA1 | ENST00000471181 | 0.12051 | 0.15111 | 0.09232 | 0.91062 | 0.87099 | 0.2479949305 | 0.65224 | 0.1271461099 | 0.120891 | 2.94236424 | 0 |
| 17 | 43071020 | C | G | V | L | BRCA1 | ENST00000471181 | 0.04812 | 0.08961 | 0.08284 | 0.79898 | 0.82387 | 0.1992762743 | 0.45568 | 0.074830231 | 0.0586004 | 2.135639158 | 0 |
| 17 | 43071020 | C | T | V | M | BRCA1 | ENST00000471181 | 0.06079 | 0.09377 | 0.09232 | 0.76871 | 0.83997 | 0.1897797139 | 0.44353 | 0.05848999462 | 0.0364991 | 1.758710413 | 0 |
| 17 | 43071021 | A | C | S | R | BRCA1 | ENST00000471181 | 0.18487 | 0.22381 | 0.09547 | 0.89252 | 0.84106 | 0.318127201 | 0.73883 | 0.1340848827 | 0.149874 | 3.005625231 | 0 |
| 17 | 43071021 | A | T | S | R | BRCA1 | ENST00000471181 | 0.18183 | 0.22381 | 0.09547 | 0.89252 | 0.84106 | 0.318127201 | 0.73883 | 0.1349818877 | 0.149873 | 3.005625231 | 0 |
| 17 | 43071022 | C | T | S | N | BRCA1 | ENST00000471181 | 0.04776 | 0.08873 | 0.09054 | 0.61161 | 0.78658 | 0.2074684011 | 0.45975 | 0.02831266337 | -0.107577 | 3.922669312 | 0 |
| 17 | 43071023 | T | A | S | C | BRCA1 | ENST00000471181 | 0.38438 | 0.37803 | 0.07708 | 0.92996 | 0.77463 | 0.300700673 | 0.88837 | 0.4717423973 | 0.133518 | 4.225614787 | 0 |
| 17 | 43071023 | T | C | S | G | BRCA1 | ENST00000471181 | 0.16832 | 0.13844 | 0.06855 | 0.88226 | 0.78535 | 0.2334506938 | 0.85339 | 0.1657795651 | 0.113424 | 3.564655155 | 0 |
| 17 | 43071023 | T | G | S | R | BRCA1 | ENST00000471181 | 0.24612 | 0.22381 | 0.09547 | 0.90406 | 0.79334 | 0.318127201 | 0.82706 | 0.3004317582 | 0.158339 | 3.005625231 | 0 |
| 17 | 43104868 | A | C | Y | D | BRCA1 | ENST00000471181 | 0.32467 | 0.17657 | 0.41534 | 0.9389 | 0.82159 | 0.681465741 | 0.69996 | 0.1256449201 | 0.233436 | 1.112405748 | 0 |
| 17 | 43104868 | A | T | Y | N | BRCA1 | ENST00000471181 | 0.32057 | 0.16892 | 0.4341 | 0.95234 | 0.82836 | 0.5795530214 | 0.64546 | 0.1474502846 | 0.220184 | 1.112405748 | 0 |
| 17 | 43104869 | C | A | E | D | BRCA1 | ENST00000471181 | 0.21929 | 0.08543 | 0.15673 | 0.79261 | 0.76487 | 0.3949957796 | 0.71696 | 0.4148139358 | 0.141737 | 0.5546928722 | 0 |
| 17 | 43104869 | C | G | E | D | BRCA1 | ENST00000471181 | 0.22345 | 0.08543 | 0.15673 | 0.7905 | 0.76487 | 0.3949957796 | 0.71696 | 0.3110077381 | 0.141736 | 0.5546928722 | 0 |
| 17 | 43104874 | A | C | L | V | BRCA1 | ENST00000471181 | 0.25076 | 0.36765 | 0.23331 | 0.92877 | 0.85064 | 0.3025291496 | 0.71312 | 0.1850685304 | 0.171375 | 3.923124662 | 0 |
| 17 | 43104876 | C | A | G | V | BRCA1 | ENST00000471181 | 0.64555 | 0.45518 | 0.46598 | 0.92063 | 0.9014 | 0.6363694039 | 0.95931 | 0.9810838103 | 0.495717 | 6.377996923 | 1 |
| 17 | 43104876 | C | G | G | A | BRCA1 | ENST00000471181 | 0.61551 | 0.4373 | 0.46105 | 0.88693 | 0.84321 | 0.4990739806 | 0.97418 | 0.9500960708 | 0.359131 | 6.377996923 | 0 |
| 17 | 43104876 | C | T | G | D | BRCA1 | ENST00000471181 | 0.64843 | 0.48662 | 0.58515 | 0.88515 | 0.88448 | 0.601221152 | 0.97282 | 0.9783811569 | 0.493647 | 6.377996923 | 0 |
| 17 | 43104877 | C | G | G | R | BRCA1 | ENST00000471181 | 0.56693 | 0.47111 | 0.57261 | 0.95035 | 0.93358 | 0.6399561662 | 0.94447 | 0.984716177 | 0.52428 | 6.377996923 | 0 |
| 17 | 43104877 | C | T | G | S | BRCA1 | ENST00000471181 | 0.43075 | 0.43845 | 0.45026 | 0.97426 | 0.86703 | 0.5465525413 | 0.98533 | 0.894462347 | 0.340364 | 4.270677769 | 0 |
| 17 | 43104880 | T | A | T | S | BRCA1 | ENST00000471181 | 0.40693 | 0.35125 | 0.39999 | 0.93842 | 0.85945 | 0.4334958462 | 0.88926 | 0.8806759119 | 0.223193 | 4.069108922 | 0 |
| 17 | 43104880 | T | C | T | A | BRCA1 | ENST00000471181 | 0.44948 | 0.31114 | 0.37444 | 0.95741 | 0.89648 | 0.5432003961 | 0.91344 | 0.9387990832 | 0.294278 | 4.555946561 | 0 |
| 17 | 43104880 | T | G | T | P | BRCA1 | ENST00000471181 | 0.59458 | 0.48019 | 0.39204 | 0.97407 | 0.90537 | 0.7754981789 | 0.93114 | 0.9641208053 | 0.353856 | 6.291935476 | 1 |
| 17 | 43104881 | G | C | D | E | BRCA1 | ENST00000471181 | 0.58967 | 0.40021 | 0.60194 | 0.91731 | 0.9014 | 0.4394487339 | 0.93605 | 0.9834271669 | 0.375416 | 6.680664366 | 1 |
| 17 | 43104881 | G | T | D | E | BRCA1 | ENST00000471181 | 0.58382 | 0.40021 | 0.60194 | 0.92211 | 0.9014 | 0.4394487339 | 0.93605 | 0.9834271669 | 0.375415 | 6.680664366 | 1 |
| 17 | 43104882 | T | A | D | V | BRCA1 | ENST00000471181 | 0.73749 | 0.4788 | 0.5563 | 0.95899 | 0.95353 | 0.7645069792 | 0.95592 | 0.993458569 | 0.510059 | 6.680664366 | 1 |
| 17 | 43104882 | T | C | D | G | BRCA1 | ENST00000471181 | 0.75424 | 0.47019 | 0.57708 | 0.95173 | 0.93901 | 0.6767982003 | 0.95417 | 0.9837375879 | 0.509016 | 6.680664366 | 1 |
| 17 | 43104882 | T | G | D | A | BRCA1 | ENST00000471181 | 0.74555 | 0.43841 | 0.56452 | 0.95525 | 0.94876 | 0.6970473084 | 0.95561 | 0.984577477 | 0.509047 | 6.680664366 | 1 |
| 17 | 43104883 | C | A | D | Y | BRCA1 | ENST00000471181 | 0.71875 | 0.47961 | 0.63703 | 0.97567 | 0.9367 | 0.7512419752 | 0.97126 | 0.9951253533 | 0.509982 | 6.680664366 | 1 |
| 17 | 43104883 | C | G | D | H | BRCA1 | ENST00000471181 | 0.71209 | 0.47286 | 0.66079 | 0.97543 | 0.92356 | 0.6939213112 | 0.97056 | 0.9936667085 | 0.509412 | 6.680664366 | 1 |
| 17 | 43104883 | C | T | D | N | BRCA1 | ENST00000471181 | 0.71826 | 0.42198 | 0.63146 | 0.97595 | 0.84052 | 0.5820309814 | 0.98613 | 0.9868932962 | 0.219158 | 6.680664366 | 1 |
| 17 | 43104885 | A | C | L | R | BRCA1 | ENST00000471181 | 0.81284 | 0.49409 | 0.69145 | 0.98903 | 0.90096 | 0.4840636356 | 0.97396 | 0.9811190367 | 0.504915 | 5.814224536 | 0 |
| 17 | 43104885 | A | G | L | P | BRCA1 | ENST00000471181 | 0.83483 | 0.51939 | 0.72953 | 0.98889 | 0.91945 | 0.7462758785 | 0.99278 | 0.989606142 | 0.506373 | 6.503435781 | 1 |
| 17 | 43104885 | A | T | L | H | BRCA1 | ENST00000471181 | 0.81261 | 0.47796 | 0.69062 | 0.98867 | 0.89784 | 0.4420569265 | 0.97405 | 0.9821922183 | 0.500462 | 6.503435781 | 0 |
| 17 | 43104886 | G | A | L | F | BRCA1 | ENST00000471181 | 0.66153 | 0.44443 | 0.60068 | 0.9677 | 0.89558 | 0.3076320065 | 0.98898 | 0.9775394201 | 0.329948 | 6.080440739 | 0 |
| 17 | 43104886 | G | T | L | I | BRCA1 | ENST00000471181 | 0.62909 | 0.24293 | 0.60531 | 0.88336 | 0.85274 | 0.2347922955 | 0.94107 | 0.9638406634 | 0.237804 | 5.077741615 | 0 |
| 17 | 43104887 | C | A | Q | H | BRCA1 | ENST00000471181 | 0.37498 | 0.13433 | 0.31662 | 0.8605 | 0.84159 | 0.4732948106 | 0.77226 | 0.4261256754 | 0.214957 | 1.063145639 | 0 |
| 17 | 43104887 | C | G | Q | H | BRCA1 | ENST00000471181 | 0.38101 | 0.13433 | 0.31662 | 0.85958 | 0.84159 | 0.4732948106 | 0.77226 | 0.6391490698 | 0.214959 | 1.063145639 | 0 |
| 17 | 43104888 | T | A | Q | L | BRCA1 | ENST00000471181 | 0.5429 | 0.08873 | 0.30563 | 0.83532 | 0.8367 | 0.5379168082 | 0.95196 | 0.6207103003 | 0.236897 | 1.063145639 | 1 |
| 17 | 43104888 | T | C | Q | R | BRCA1 | ENST00000471181 | 0.50094 | 0.10363 | 0.34525 | 0.81272 | 0.82724 | 0.4132729857 | 0.94677 | 0.5461423575 | 0.246922 | 1.063145639 | 0 |
| 17 | 43104889 | G | C | Q | E | BRCA1 | ENST00000471181 | 0.15365 | 0.09826 | 0.27786 | 0.86866 | 0.75417 | 0.3194783461 | 0.53946 | 0.08790089935 | 0.158498 | -5.131568666 | 0 |
| 17 | 43104889 | G | T | Q | K | BRCA1 | ENST00000471181 | 0.32717 | 0.1187 | 0.38156 | 0.93167 | 0.79273 | 0.5415045759 | 0.83868 | 0.3272785983 | 0.22659 | 1.063145639 | 0 |
| 17 | 43104890 | A | C | F | L | BRCA1 | ENST00000471181 | 0.67716 | 0.40968 | 0.63741 | 0.97506 | 0.91143 | 0.5312796378 | 0.97075 | 0.9440840483 | 0.303237 | 6.680663937 | 1 |
| 17 | 43104891 | A | G | F | S | BRCA1 | ENST00000471181 | 0.71361 | 0.51294 | 0.60094 | 0.99026 | 0.89558 | 0.6710917453 | 0.98916 | 0.9848339558 | 0.497142 | 6.680663937 | 1 |
| 17 | 43104891 | A | T | F | Y | BRCA1 | ENST00000471181 | 0.6643 | 0.45012 | 0.62229 | 0.9689 | 0.87198 | 0.4396969155 | 0.96619 | 0.9484209418 | 0.342588 | 6.680663937 | 0 |
| 17 | 43104892 | A | C | F | V | BRCA1 | ENST00000471181 | 0.73421 | 0.46423 | 0.57391 | 0.98793 | 0.91945 | 0.6077021451 | 0.99057 | 0.949044168 | 0.474217 | 6.680663937 | 0 |
| 17 | 43104892 | A | G | F | L | BRCA1 | ENST00000471181 | 0.74515 | 0.40968 | 0.63741 | 0.97012 | 0.90581 | 0.5312796378 | 0.98539 | 0.9574085474 | 0.315036 | 6.680663937 | 1 |
| 17 | 43104892 | A | T | F | I | BRCA1 | ENST00000471181 | 0.72949 | 0.48493 | 0.60325 | 0.98239 | 0.91357 | 0.6415545289 | 0.98528 | 0.9517472386 | 0.348649 | 6.680663937 | 0 |
| 17 | 43104894 | G | A | A | V | BRCA1 | ENST00000471181 | 0.68619 | 0.41631 | 0.44537 | 0.97362 | 0.91986 | 0.5380311354 | 0.99407 | 0.9834023714 | 0.486309 | 6.680142566 | 1 |
| 17 | 43104894 | G | T | A | D | BRCA1 | ENST00000471181 | 0.69612 | 0.51234 | 0.53045 | 0.98007 | 0.90581 | 0.768881307 | 0.99708 | 0.9925268888 | 0.493061 | 6.680142566 | 1 |
| 17 | 43104895 | C | A | A | S | BRCA1 | ENST00000471181 | 0.57659 | 0.41941 | 0.38516 | 0.97127 | 0.86453 | 0.4080257671 | 0.9906 | 0.9774441123 | 0.314568 | 6.653258247 | 0 |
| 17 | 43104895 | C | G | A | P | BRCA1 | ENST00000471181 | 0.6243 | 0.48587 | 0.49522 | 0.97818 | 0.88637 | 0.8219290256 | 0.99634 | 0.9898102283 | 0.48476 | 6.680142566 | 1 |
| 17 | 43104895 | C | T | A | T | BRCA1 | ENST00000471181 | 0.61529 | 0.42806 | 0.42467 | 0.97559 | 0.88212 | 0.4280633659 | 0.99425 | 0.9774441123 | 0.469571 | 6.680142566 | 1 |
| 17 | 43104897 | C | A | C | F | BRCA1 | ENST00000471181 | 0.29074 | 0.14952 | 0.15952 | 0.92879 | 0.80178 | 0.3783476731 | 0.59063 | 0.2979259789 | 0.181573 | 0.9683349512 | 0 |
| 17 | 43104897 | C | G | C | S | BRCA1 | ENST00000471181 | 0.1976 | 0.11295 | 0.14399 | 0.8563 | 0.80358 | 0.310181383 | 0.52084 | 0.01647592336 | 0.140694 | -0.2169266493 | 0 |
| 17 | 43104897 | C | T | C | Y | BRCA1 | ENST00000471181 | 0.27923 | 0.15908 | 0.19725 | 0.90373 | 0.80772 | 0.4264895019 | 0.55419 | 0.04996513611 | 0.172938 | -0.4502952262 | 0 |
| 17 | 43104898 | A | C | C | G | BRCA1 | ENST00000471181 | 0.31988 | 0.11446 | 0.13111 | 0.88307 | 0.80653 | 0.3429321391 | 0.60167 | 0.02337835788 | 0.185589 | 0.9683349512 | 0 |
| 17 | 43104898 | A | G | C | R | BRCA1 | ENST00000471181 | 0.30606 | 0.16675 | 0.19094 | 0.87338 | 0.79516 | 0.4537848903 | 0.69978 | 0.05141867325 | 0.176027 | -2.14440343 | 0 |
| 17 | 43104898 | A | T | C | S | BRCA1 | ENST00000471181 | 0.29173 | 0.11295 | 0.14399 | 0.8574 | 0.78597 | 0.310181383 | 0.70798 | 0.02005246971 | 0.15913 | -0.2169266493 | 0 |
| 17 | 43104899 | A | C | I | M | BRCA1 | ENST00000471181 | 0.40845 | 0.43951 | 0.38193 | 0.90715 | 0.82836 | 0.4667602069 | 0.95496 | 0.6788136363 | 0.174103 | 4.616959688 | 0 |
| 17 | 43104900 | A | C | I | S | BRCA1 | ENST00000471181 | 0.52281 | 0.4758 | 0.44863 | 0.97282 | 0.89285 | 0.7401761778 | 0.99403 | 0.9543861151 | 0.401532 | 6.177134313 | 0 |
| 17 | 43104900 | A | G | I | T | BRCA1 | ENST00000471181 | 0.36131 | 0.44665 | 0.54616 | 0.96963 | 0.89918 | 0.4729432499 | 0.99567 | 0.8811936975 | 0.371246 | 3.365749167 | 0 |
| 17 | 43104901 | T | A | I | F | BRCA1 | ENST00000471181 | 0.67231 | 0.5007 | 0.42969 | 0.95903 | 0.86852 | 0.6058789152 | 0.99669 | 0.9062334299 | 0.291416 | 6.177134313 | 0 |
| 17 | 43104901 | T | C | I | V | BRCA1 | ENST00000471181 | 0.42193 | 0.31003 | 0.33219 | 0.92318 | 0.85117 | 0.2372020475 | 0.93888 | 0.8788395524 | 0.224022 | 5.646894704 | 0 |
| 17 | 43104901 | T | G | I | L | BRCA1 | ENST00000471181 | 0.4935 | 0.30657 | 0.42388 | 0.90992 | 0.83725 | 0.4627263541 | 0.96044 | 0.9367668033 | 0.189976 | 5.867456475 | 0 |
| 17 | 43104902 | G | C | I | M | BRCA1 | ENST00000471181 | 0.34981 | 0.20665 | 0.31515 | 0.95063 | 0.86352 | 0.3040566465 | 0.93992 | 0.1379853793 | 0.187828 | 4.39187974 | 0 |
| 17 | 43104903 | A | G | I | T | BRCA1 | ENST00000471181 | 0.28295 | 0.26636 | 0.46534 | 0.94807 | 0.85064 | 0.3029444565 | 0.9598 | 0.2829928696 | 0.132062 | 3.186586388 | 0 |
| 17 | 43104903 | A | T | I | N | BRCA1 | ENST00000471181 | 0.64329 | 0.53373 | 0.43868 | 0.98504 | 0.87198 | 0.553388235 | 0.96524 | 0.9208096266 | 0.267496 | 6.127706635 | 1 |
| 17 | 43104904 | T | A | I | F | BRCA1 | ENST00000471181 | 0.405 | 0.49339 | 0.35506 | 0.97202 | 0.86453 | 0.4028338393 | 0.98943 | 0.906057179 | 0.255758 | 6.127706635 | 0 |
| 17 | 43104904 | T | G | I | L | BRCA1 | ENST00000471181 | 0.40447 | 0.20346 | 0.3515 | 0.93505 | 0.825 | 0.3367433674 | 0.91115 | 0.8101133108 | 0.217036 | 5.104751313 | 0 |
| 17 | 43104905 | T | A | K | N | BRCA1 | ENST00000471181 | 0.42615 | 0.21256 | 0.35363 | 0.96277 | 0.82103 | 0.3805649611 | 0.80483 | 0.5792331696 | 0.11331 | 6.100757444 | 0 |
| 17 | 43104905 | T | G | K | N | BRCA1 | ENST00000471181 | 0.43232 | 0.21256 | 0.35363 | 0.96157 | 0.82103 | 0.3805649611 | 0.80483 | 0.6785624623 | 0.1133 | 6.100757444 | 0 |
| 17 | 43104906 | T | C | K | R | BRCA1 | ENST00000471181 | 0.34877 | 0.08722 | 0.25883 | 0.77966 | 0.78348 | 0.2513444868 | 0.77545 | 0.09865392715 | 0.164115 | 4.193979626 | 0 |
| 17 | 43104906 | T | G | K | T | BRCA1 | ENST00000471181 | 0.62075 | 0.39535 | 0.3394 | 0.96045 | 0.88448 | 0.4140192656 | 0.88965 | 0.8092424273 | 0.209318 | 6.48859099 | 0 |
| 17 | 43104907 | T | C | K | E | BRCA1 | ENST00000471181 | 0.52473 | 0.32581 | 0.32111 | 0.82005 | 0.84428 | 0.2888089716 | 0.85173 | 0.7362760305 | 0.23244 | 3.884027437 | 0 |
| 17 | 43104907 | T | G | K | Q | BRCA1 | ENST00000471181 | 0.67334 | 0.37834 | 0.28148 | 0.94705 | 0.86098 | 0.3184501731 | 0.89101 | 0.8127345443 | 0.218056 | 6.48859099 | 0 |
| 17 | 43104908 | C | A | L | F | BRCA1 | ENST00000471181 | 0.61186 | 0.45622 | 0.48577 | 0.94104 | 0.87099 | 0.3893844136 | 0.99376 | 0.9307463169 | 0.288724 | 5.762411073 | 1 |
| 17 | 43104908 | C | G | L | F | BRCA1 | ENST00000471181 | 0.61978 | 0.45622 | 0.48577 | 0.94095 | 0.87099 | 0.3893844136 | 0.99376 | 0.9307463169 | 0.288726 | 5.762411073 | 0 |
| 17 | 43104909 | A | C | L | W | BRCA1 | ENST00000471181 | 0.62498 | 0.5366 | 0.57978 | 0.98057 | 0.89239 | 0.6176600352 | 0.98837 | 0.8358715773 | 0.297722 | 6.140783793 | 0 |
| 17 | 43104909 | A | G | L | S | BRCA1 | ENST00000471181 | 0.44472 | 0.48259 | 0.51345 | 0.97812 | 0.87684 | 0.5316606411 | 0.98846 | 0.9588330388 | 0.388281 | 4.39972383 | 0 |
| 17 | 43104910 | A | C | L | V | BRCA1 | ENST00000471181 | 0.32849 | 0.4223 | 0.43403 | 0.93483 | 0.85586 | 0.3811904002 | 0.93804 | 0.7241452932 | 0.256313 | 6.017177822 | 0 |
| 17 | 43104910 | A | T | L | M | BRCA1 | ENST00000471181 | 0.35897 | 0.44052 | 0.4709 | 0.92975 | 0.80476 | 0.3440675196 | 0.91628 | 0.8400174379 | 0.182579 | 5.381010967 | 0 |
| 17 | 43104912 | A | C | L | R | BRCA1 | ENST00000471181 | 0.73981 | 0.51675 | 0.71688 | 0.98826 | 0.91986 | 0.9089098676 | 0.98995 | 0.9638209343 | 0.492225 | 6.674973442 | 1 |
| 17 | 43104912 | A | G | L | P | BRCA1 | ENST00000471181 | 0.74388 | 0.53457 | 0.74312 | 0.99001 | 0.92233 | 0.9328755162 | 0.9917 | 0.9650481939 | 0.493102 | 6.674973442 | 1 |
| 17 | 43104912 | A | T | L | Q | BRCA1 | ENST00000471181 | 0.73013 | 0.45914 | 0.64764 | 0.98674 | 0.91058 | 0.7981753131 | 0.98772 | 0.9638209343 | 0.492158 | 6.674973442 | 0 |
| 17 | 43104913 | G | C | L | V | BRCA1 | ENST00000471181 | 0.49718 | 0.45285 | 0.58609 | 0.9718 | 0.85792 | 0.5936382007 | 0.96519 | 0.9581444263 | 0.300536 | 6.499647507 | 0 |
| 17 | 43104913 | G | T | L | I | BRCA1 | ENST00000471181 | 0.50328 | 0.46381 | 0.63643 | 0.9705 | 0.83172 | 0.445238848 | 0.94267 | 0.9665762782 | 0.259984 | 6.674973442 | 0 |
| 17 | 43104914 | C | A | E | D | BRCA1 | ENST00000471181 | 0.43804 | 0.34616 | 0.47154 | 0.95711 | 0.81703 | 0.5365653052 | 0.89882 | 0.4987432361 | 0.267524 | 6.865889774 | 0 |
| 17 | 43104915 | T | A | E | V | BRCA1 | ENST00000471181 | 0.74722 | 0.34145 | 0.44196 | 0.97775 | 0.91442 | 0.5703174909 | 0.92121 | 0.8535923362 | 0.324244 | 5.963096437 | 0 |
| 17 | 43104915 | T | C | E | G | BRCA1 | ENST00000471181 | 0.43093 | 0.16284 | 0.42193 | 0.96316 | 0.85378 | 0.5009559577 | 0.90756 | 0.378878057 | 0.235792 | 3.935425531 | 1 |

| | | | | | | | | | | | | | | | | | | |
|---|---|---|---|---|---|---|---|---|---|---|---|---|---|---|---|---|---|---|
| 17 | 43104915 | T | G | E | A | BRCA1 | ENST00000471181 | 0.70559 | 0.31153 | 0.46759 | 0.94683 | 0.87636 | 0.5027047413 | 0.93131 | 0.8349848986 | 0.260401 | 5.030214662 | 0 |
| 17 | 43104916 | C | G | E | Q | BRCA1 | ENST00000471181 | 0.65964 | 0.40435 | 0.45406 | 0.94755 | 0.86951 | 0.4694286891 | 0.95177 | 0.9408112168 | 0.29787 | 6.865889774 | 0 |
| 17 | 43104916 | C | T | E | K | BRCA1 | ENST00000471181 | 0.71361 | 0.37407 | 0.57476 | 0.95392 | 0.88401 | 0.5603087582 | 0.95215 | 0.9408112168 | 0.313007 | 6.865889774 | 1 |
| 17 | 43104917 | T | A | E | D | BRCA1 | ENST00000471181 | 0.68027 | 0.4363 | 0.45488 | 0.94321 | 0.85996 | 0.4381439009 | 0.84868 | 0.9703956842 | 0.23903 | 4.533202455 | 0 |
| 17 | 43104917 | T | G | E | D | BRCA1 | ENST00000471181 | 0.68851 | 0.4363 | 0.45488 | 0.94356 | 0.85996 | 0.4381439009 | 0.84868 | 0.9628904462 | 0.239031 | 4.533202455 | 0 |
| 17 | 43104918 | T | A | E | V | BRCA1 | ENST00000471181 | 0.798 | 0.47742 | 0.43305 | 0.98067 | 0.93554 | 0.5732821605 | 0.8514 | 0.9812770486 | 0.357381 | 6.326590181 | 1 |
| 17 | 43104918 | T | C | E | G | BRCA1 | ENST00000471181 | 0.81755 | 0.47791 | 0.39989 | 0.97889 | 0.91229 | 0.5091531109 | 0.8868 | 0.9812770486 | 0.276559 | 6.326590181 | 0 |
| 17 | 43104918 | T | G | E | A | BRCA1 | ENST00000471181 | 0.76824 | 0.46163 | 0.45182 | 0.97868 | 0.92028 | 0.4945155065 | 0.87283 | 0.9788236618 | 0.28269 | 6.326590181 | 0 |
| 17 | 43104919 | C | G | E | Q | BRCA1 | ENST00000471181 | 0.58652 | 0.45036 | 0.43198 | 0.94283 | 0.8306 | 0.42597839 | 0.90495 | 0.9821062088 | 0.231823 | 4.42875983 | 0 |
| 17 | 43104919 | C | T | E | K | BRCA1 | ENST00000471181 | 0.64734 | 0.47059 | 0.56791 | 0.94887 | 0.85843 | 0.5254493124 | 0.8756 | 0.9806548357 | 0.301755 | 6.326590181 | 1 |
| 17 | 43104921 | A | C | V | G | BRCA1 | ENST00000471181 | 0.69324 | 0.46586 | 0.29501 | 0.98425 | 0.94394 | 0.7773044979 | 0.98794 | 0.8738025052 | 0.484379 | 6.997362731 | 0 |
| 17 | 43104921 | A | T | V | D | BRCA1 | ENST00000471181 | 0.71442 | 0.48067 | 0.40928 | 0.98518 | 0.92763 | 0.9207792048 | 0.98776 | 0.9023621082 | 0.486316 | 6.997362731 | 1 |
| 17 | 43104922 | C | A | V | F | BRCA1 | ENST00000471181 | 0.59276 | 0.48771 | 0.36581 | 0.97399 | 0.90007 | 0.822404925 | 0.99127 | 0.9647145867 | 0.49048 | 6.997362731 | 1 |
| 17 | 43104922 | C | G | V | L | BRCA1 | ENST00000471181 | 0.53791 | 0.36306 | 0.38903 | 0.96028 | 0.88496 | 0.5757822781 | 0.98601 | 0.9100224376 | 0.328455 | 6.997362731 | 0 |
| 17 | 43104922 | C | T | V | I | BRCA1 | ENST00000471181 | 0.28417 | 0.37478 | 0.25473 | 0.89352 | 0.83004 | 0.3171273275 | 0.95696 | 0.8457580209 | 0.236127 | 4.141939115 | 0 |
| 17 | 43104924 | A | C | L | R | BRCA1 | ENST00000471181 | 0.723 | 0.51028 | 0.62341 | 0.98513 | 0.93709 | 0.8930656314 | 0.96544 | 0.969014883 | 0.427 | 6.671660367 | 1 |
| 17 | 43104924 | A | G | L | P | BRCA1 | ENST00000471181 | 0.727 | 0.52818 | 0.66366 | 0.9882 | 0.93397 | 0.9281889027 | 0.97192 | 0.9719974399 | 0.42729 | 6.671660367 | 1 |
| 17 | 43104924 | A | T | L | H | BRCA1 | ENST00000471181 | 0.70455 | 0.48693 | 0.61933 | 0.98774 | 0.92561 | 0.786818823 | 0.9744 | 0.8871542441 | 0.427484 | 6.671660367 | 1 |
| 17 | 43104925 | G | A | L | F | BRCA1 | ENST00000471181 | 0.697 | 0.44722 | 0.52749 | 0.95873 | 0.89009 | 0.6268195771 | 0.97855 | 0.9802761078 | 0.273927 | 6.671660367 | 1 |
| 17 | 43104925 | G | T | L | I | BRCA1 | ENST00000471181 | 0.66122 | 0.32673 | 0.55039 | 0.94053 | 0.82387 | 0.4105679965 | 0.95848 | 0.9843622446 | 0.213324 | 6.057400082 | 0 |
| 17 | 43104926 | T | A | Q | H | BRCA1 | ENST00000471181 | 0.49493 | 0.40813 | 0.43074 | 0.97307 | 0.82159 | 0.3365497466 | 0.91147 | 0.9451591372 | 0.2707 | 5.367167946 | 1 |
| 17 | 43104927 | T | A | Q | L | BRCA1 | ENST00000471181 | 0.72116 | 0.09262 | 0.36438 | 0.97285 | 0.90668 | 0.4995468366 | 0.91914 | 0.936578095 | 0.357645 | 4.781234228 | 0 |
| 17 | 43104927 | T | C | Q | R | BRCA1 | ENST00000471181 | 0.68731 | 0.37353 | 0.43188 | 0.97171 | 0.85117 | 0.2896678297 | 0.94041 | 0.9423229694 | 0.342298 | 6.537855628 | 0 |
| 17 | 43104927 | T | G | Q | P | BRCA1 | ENST00000471181 | 0.70888 | 0.43757 | 0.40493 | 0.98086 | 0.90185 | 0.7750138241 | 0.95443 | 0.9571671486 | 0.489592 | 6.537855628 | 0 |
| 17 | 43104928 | G | T | Q | K | BRCA1 | ENST00000471181 | 0.63451 | 0.33204 | 0.44175 | 0.96826 | 0.87247 | 0.3816538092 | 0.91495 | 0.9444692731 | 0.331289 | 4.541040855 | 0 |
| 17 | 43104929 | A | C | S | R | BRCA1 | ENST00000471181 | 0.63015 | 0.33486 | 0.36966 | 0.98257 | 0.86403 | 0.4216789491 | 0.95006 | 0.9512912631 | 0.297981 | 6.55627346 | 0 |
| 17 | 43104930 | C | A | S | I | BRCA1 | ENST00000471181 | 0.49447 | 0.43306 | 0.35319 | 0.98144 | 0.88354 | 0.4165997509 | 0.95124 | 0.9683761597 | 0.305241 | 6.55627346 | 1 |
| 17 | 43104930 | C | G | S | T | BRCA1 | ENST00000471181 | 0.58474 | 0.28907 | 0.31545 | 0.96335 | 0.84588 | 0.2521098808 | 0.9624 | 0.9006797671 | 0.260522 | 6.55627346 | 0 |
| 17 | 43104930 | C | T | S | N | BRCA1 | ENST00000471181 | 0.4831 | 0.32113 | 0.36103 | 0.93601 | 0.78597 | 0.2977815993 | 0.9367 | 0.7487822771 | 0.163001 | 4.509337993 | 0 |
| 17 | 43104931 | T | A | S | C | BRCA1 | ENST00000471181 | 0.68739 | 0.4238 | 0.30999 | 0.98482 | 0.88823 | 0.3290355128 | 0.96166 | 0.9493345022 | 0.278371 | 6.55627346 | 0 |
| 17 | 43104931 | T | C | S | G | BRCA1 | ENST00000471181 | 0.36047 | 0.2898 | 0.26254 | 0.95825 | 0.85378 | 0.2640097622 | 0.93476 | 0.8878896236 | 0.244553 | 4.417336564 | 0 |
| 17 | 43104932 | A | C | F | L | BRCA1 | ENST00000471181 | 0.66786 | 0.38919 | 0.72468 | 0.97585 | 0.91526 | 0.5536016309 | 0.9717 | 0.9731118083 | 0.308141 | 6.680209121 | 0 |
| 17 | 43104932 | A | T | F | L | BRCA1 | ENST00000471181 | 0.66114 | 0.38919 | 0.72468 | 0.97648 | 0.91526 | 0.5536016309 | 0.9717 | 0.9731118083 | 0.30814 | 6.680209121 | 0 |
| 17 | 43104933 | A | C | F | C | BRCA1 | ENST00000471181 | 0.76299 | 0.50212 | 0.71963 | 0.98946 | 0.93593 | 0.6971236413 | 0.98088 | 0.9808686972 | 0.479535 | 6.680209121 | 0 |
| 17 | 43104933 | A | G | F | S | BRCA1 | ENST00000471181 | 0.76203 | 0.32054 | 0.69213 | 0.98613 | 0.95426 | 0.7685830126 | 0.9768 | 0.9716718197 | 0.477185 | 6.680209121 | 0 |
| 17 | 43104933 | A | T | F | Y | BRCA1 | ENST00000471181 | 0.62688 | 0.45012 | 0.7084 | 0.96932 | 0.90581 | 0.5524659888 | 0.97953 | 0.9753752947 | 0.344169 | 6.662904041 | 0 |
| 17 | 43104934 | A | C | F | V | BRCA1 | ENST00000471181 | 0.74061 | 0.45275 | 0.66662 | 0.98751 | 0.91945 | 0.6193065414 | 0.98243 | 0.9719974399 | 0.438619 | 6.680209121 | 0 |
| 17 | 43104934 | A | G | F | L | BRCA1 | ENST00000471181 | 0.74858 | 0.38919 | 0.72468 | 0.97381 | 0.90581 | 0.5536016309 | 0.97953 | 0.9753752947 | 0.319515 | 6.680209121 | 0 |
| 17 | 43104935 | T | A | R | S | BRCA1 | ENST00000471181 | 0.60485 | 0.42799 | 0.55269 | 0.98934 | 0.88022 | 0.6028637885 | 0.93019 | 0.9711030722 | 0.432155 | 6.657980371 | 0 |
| 17 | 43104935 | T | G | R | S | BRCA1 | ENST00000471181 | 0.61246 | 0.42799 | 0.55269 | 0.98942 | 0.88069 | 0.6028637885 | 0.93019 | 0.9711030722 | 0.432156 | 6.657980371 | 0 |
| 17 | 43104936 | C | A | R | I | BRCA1 | ENST00000471181 | 0.68187 | 0.48129 | 0.5771 | 0.98056 | 0.92397 | 0.5755324577 | 0.97331 | 0.978042841 | 0.447363 | 5.680621303 | 0 |
| 17 | 43104936 | C | G | R | T | BRCA1 | ENST00000471181 | 0.63989 | 0.47462 | 0.59442 | 0.98062 | 0.91143 | 0.600295587 | 0.97741 | 0.943408668 | 0.454684 | 5.707974101 | 0 |
| 17 | 43104936 | C | T | R | K | BRCA1 | ENST00000471181 | 0.6385 | 0.39108 | 0.62089 | 0.9665 | 0.8705 | 0.5317230238 | 0.9781 | 0.9787411094 | 0.335769 | 6.427817373 | 0 |
| 17 | 43104937 | T | C | R | G | BRCA1 | ENST00000471181 | 0.76164 | 0.44977 | 0.54245 | 0.9882 | 0.93241 | 0.5649702946 | 0.93025 | 0.9796789885 | 0.458136 | 6.657980371 | 0 |
| 17 | 43104939 | G | A | T | M | BRCA1 | ENST00000471181 | 0.79895 | 0.435 | 0.53388 | 0.96868 | 0.85689 | 0.3003683917 | 0.98009 | 0.8716692328 | 0.178359 | 6.88027466 | 0 |
| 17 | 43104939 | G | C | T | R | BRCA1 | ENST00000471181 | 0.78535 | 0.15073 | 0.56991 | 0.95125 | 0.84213 | 0.444916234 | 0.97949 | 0.9577011466 | 0.384519 | 6.88027466 | 0 |
| 17 | 43104939 | G | T | T | K | BRCA1 | ENST00000471181 | 0.79919 | 0.39459 | 0.59107 | 0.9645 | 0.87 | 0.4245926934 | 0.98154 | 0.975625217 | 0.365442 | 6.88027466 | 0 |
| 17 | 43104940 | T | A | T | S | BRCA1 | ENST00000471181 | 0.43004 | 0.28675 | 0.48193 | 0.94684 | 0.80713 | 0.2156043282 | 0.92297 | 0.7692920566 | 0.233096 | 4.373193875 | 0 |
| 17 | 43104940 | T | C | T | A | BRCA1 | ENST00000471181 | 0.45224 | 0.29946 | 0.46499 | 0.93711 | 0.83116 | 0.2272505334 | 0.9257 | 0.7945339084 | 0.25147 | 5.414654639 | 0 |
| 17 | 43104940 | T | G | T | P | BRCA1 | ENST00000471181 | 0.61015 | 0.20246 | 0.48126 | 0.97023 | 0.88069 | 0.4659858203 | 0.94615 | 0.8710036278 | 0.315505 | 4.262757053 | 0 |
| 17 | 43104941 | A | T | S | R | BRCA1 | ENST00000471181 | 0.61903 | 0.44788 | 0.50914 | 0.98404 | 0.89147 | 0.6630985825 | 0.94831 | 0.9703956842 | 0.401815 | 6.833717258 | 0 |
| 17 | 43104942 | C | G | S | T | BRCA1 | ENST00000471181 | 0.58981 | 0.13438 | 0.45214 | 0.96016 | 0.88307 | 0.494981959 | 0.96309 | 0.9519395232 | 0.303752 | 6.833717258 | 0 |
| 17 | 43104942 | C | T | S | N | BRCA1 | ENST00000471181 | 0.32653 | 0.39862 | 0.48884 | 0.93317 | 0.78782 | 0.3767757646 | 0.91599 | 0.9573819041 | 0.16879 | 3.66682371 | 0 |
| 17 | 43104943 | T | A | S | C | BRCA1 | ENST00000471181 | 0.81009 | 0.43657 | 0.44854 | 0.98246 | 0.91862 | 0.5819267897 | 0.98669 | 0.9217536449 | 0.32575 | 6.833717258 | 0 |
| 17 | 43104943 | T | C | S | G | BRCA1 | ENST00000471181 | 0.75416 | 0.37328 | 0.4005 | 0.97196 | 0.90842 | 0.5213934673 | 0.98583 | 0.9658761024 | 0.314766 | 6.833717258 | 0 |
| 17 | 43104943 | T | G | S | R | BRCA1 | ENST00000471181 | 0.79737 | 0.44788 | 0.50914 | 0.9804 | 0.90711 | 0.6630985825 | 0.98524 | 0.9753752947 | 0.429007 | 6.833717258 | 0 |
| 17 | 43104944 | T | A | E | D | BRCA1 | ENST00000471181 | 0.624 | 0.32879 | 0.4607 | 0.8923 | 0.79938 | 0.4118690701 | 0.82549 | 0.8956930637 | 0.224834 | 6.400723947 | 0 |
| 17 | 43104944 | T | G | E | D | BRCA1 | ENST00000471181 | 0.63183 | 0.32879 | 0.4607 | 0.89165 | 0.79938 | 0.4118690701 | 0.82549 | 0.8956930637 | 0.224835 | 6.400723947 | 0 |
| 17 | 43104945 | T | C | E | G | BRCA1 | ENST00000471181 | 0.46357 | 0.40106 | 0.4099 | 0.93039 | 0.81182 | 0.6032644981 | 0.83057 | 0.8885934353 | 0.111609 | 3.620678311 | 0 |
| 17 | 43104945 | T | G | E | A | BRCA1 | ENST00000471181 | 0.73061 | 0.35121 | 0.45722 | 0.94472 | 0.86352 | 0.5014069315 | 0.85385 | 0.9132928848 | 0.176005 | 5.427038446 | 0 |
| 17 | 43104946 | C | T | E | K | BRCA1 | ENST00000471181 | 0.74316 | 0.38458 | 0.57215 | 0.92112 | 0.85274 | 0.5507311001 | 0.91239 | 0.9510563612 | 0.226722 | 6.098446189 | 0 |
| 17 | 43104947 | T | A | Q | H | BRCA1 | ENST00000471181 | 0.49383 | 0.42086 | 0.46438 | 0.97069 | 0.8233 | 0.6002869801 | 0.84442 | 0.94037956 | 0.198166 | 6.956321337 | 0 |
| 17 | 43104947 | T | G | Q | H | BRCA1 | ENST00000471181 | 0.50049 | 0.42086 | 0.46438 | 0.97069 | 0.82103 | 0.6002869801 | 0.84442 | 0.94037956 | 0.198168 | 6.956321337 | 0 |
| 17 | 43104948 | T | C | Q | R | BRCA1 | ENST00000471181 | 0.55999 | 0.38912 | 0.46248 | 0.90145 | 0.80713 | 0.4758869875 | 0.77416 | 0.7067289352 | 0.164504 | 4.403447628 | 0 |
| 17 | 43104948 | T | G | Q | P | BRCA1 | ENST00000471181 | 0.71233 | 0.45419 | 0.42831 | 0.95176 | 0.88637 | 0.7612578 | 0.86303 | 0.893279016 | 0.335725 | 6.956321337 | 0 |
| 17 | 43104949 | G | C | Q | E | BRCA1 | ENST00000471181 | 0.61522 | 0.12707 | 0.41922 | 0.87591 | 0.84588 | 0.5187493736 | 0.82991 | 0.8839221001 | 0.188917 | 6.956321337 | 0 |
| 17 | 43104951 | A | C | L | R | BRCA1 | ENST00000471181 | 0.8174 | 0.4448 | 0.7048 | 0.98349 | 0.91652 | 0.8654238189 | 0.94409 | 0.9822522402 | 0.421781 | 6.608568926 | 1 |
| 17 | 43104951 | A | G | L | P | BRCA1 | ENST00000471181 | 0.82131 | 0.50493 | 0.73738 | 0.98696 | 0.93004 | 0.8848533852 | 0.95681 | 0.9839816689 | 0.448239 | 6.608568926 | 0 |
| 17 | 43104951 | A | T | L | Q | BRCA1 | ENST00000471181 | 0.80787 | 0.20341 | 0.62276 | 0.98038 | 0.91229 | 0.8155888413 | 0.95704 | 0.9737026691 | 0.446007 | 6.608568926 | 1 |
| 17 | 43104952 | G | C | L | V | BRCA1 | ENST00000471181 | 0.59624 | 0.19695 | 0.54201 | 0.95543 | 0.84694 | 0.4971227765 | 0.92376 | 0.9355646968 | 0.233788 | 5.691237734 | 0 |
| 17 | 43104953 | G | C | S | R | BRCA1 | ENST00000471181 | 0.6194 | 0.42418 | 0.61565 | 0.97689 | 0.82724 | 0.6390077138 | 0.91906 | 0.781711638 | 0.177461 | 6.57570597 | 0 |
| 17 | 43104953 | G | T | S | R | BRCA1 | ENST00000471181 | 0.61328 | 0.42418 | 0.61565 | 0.97903 | 0.82724 | 0.6390077138 | 0.91906 | 0.96100986 | 0.334585 | 6.57570597 | 0 |
| 17 | 43104954 | C | G | S | T | BRCA1 | ENST00000471181 | 0.62635 | 0.31736 | 0.55587 | 0.96337 | 0.81298 | 0.3774627464 | 0.9536 | 0.9787411094 | 0.284574 | 6.57570597 | 0 |
| 17 | 43104954 | C | T | S | N | BRCA1 | ENST00000471181 | 0.63451 | 0.15575 | 0.6024 | 0.96637 | 0.73877 | 0.4580111657 | 0.96142 | 0.9630215168 | 0.200507 | 5.110941674 | 1 |
| 17 | 43104955 | T | A | S | C | BRCA1 | ENST00000471181 | 0.76784 | 0.4238 | 0.55158 | 0.98262 | 0.85843 | 0.451169165 | 0.94614 | 0.679955037 | 0.303801 | 6.57570597 | 0 |
| 17 | 43104955 | T | C | S | G | BRCA1 | ENST00000471181 | 0.73661 | 0.34783 | 0.4883 | 0.97258 | 0.8367 | 0.3780493511 | 0.91535 | 0.6714552376 | 0.299192 | 5.950198218 | 0 |
| 17 | 43104955 | T | G | S | R | BRCA1 | ENST00000471181 | 0.75813 | 0.42418 | 0.61565 | 0.9827 | 0.84747 | 0.6390077138 | 0.94292 | 0.6892348921 | 0.346933 | 6.57570597 | 0 |
| 17 | 43104956 | C | A | R | S | BRCA1 | ENST00000471181 | 0.61425 | | | 0.98892 | 0.57877 | 0.6143526183 | 0.71813 | 0.8906769156 | 0.0815023 | 5.696735852 | 1 |
| 17 | 43104956 | C | G | R | S | BRCA1 | ENST00000471181 | 0.56359 | | | | 0.57877 | 0.6143526183 | 0.71813 | 0.8969510198 | 0.0814947 | 5.696735852 | 1 |
| 17 | 43106456 | C | A | R | M | BRCA1 | ENST00000471181 | 0.86824 | 0.36858 | 0.50471 | 0.97463 | 0.92804 | 0.5760633311 | 0.97364 | 0.9037867785 | 0.42502 | 6.638217599 | 1 |
| 17 | 43106456 | C | G | R | T | BRCA1 | ENST00000471181 | 0.83506 | 0.27433 | 0.51457 | 0.97065 | 0.92397 | 0.6177915348 | 0.95964 | 0.9219440818 | 0.429761 | 6.638217599 | 1 |
| 17 | 43106456 | C | T | R | K | BRCA1 | ENST00000471181 | 0.83236 | 0.18061 | 0.54359 | 0.95359 | 0.90972 | 0.4770912094 | 0.96842 | 0.9607485533 | 0.411382 | 6.638217599 | 1 |
| 17 | 43106457 | T | A | R | W | BRCA1 | ENST00000471181 | 0.95466 | 0.41475 | 0.57325 | 0.98615 | 0.9528 | 0.5858245295 | 0.97092 | 0.9748387337 | 0.429476 | 6.638217599 | 1 |
| 17 | 43106457 | T | C | R | G | BRCA1 | ENST00000471181 | 0.94793 | 0.25078 | 0.45949 | 0.9844 | 0.96008 | 0.5739693076 | 0.93509 | 0.8674880862 | 0.464929 | 6.638217599 | 1 |
| 17 | 43106458 | T | A | K | N | BRCA1 | ENST00000471181 | 0.86297 | 0.39253 | 0.62953 | 0.96995 | 0.84747 | 0.6453209344 | 0.83285 | 0.9315660596 | 0.117314 | 6.119268702 | 1 |
| 17 | 43106458 | T | G | K | N | BRCA1 | ENST00000471181 | 0.75097 | 0.39253 | 0.62953 | 0.97053 | 0.84747 | 0.6453209344 | 0.83285 | 0.9315660596 | 0.117316 | 6.119268702 | 0 |
| 17 | 43106459 | T | A | K | I | BRCA1 | ENST00000471181 | 0.70431 | 0.49151 | 0.55078 | 0.97966 | 0.91484 | 0.6751098715 | 0.82586 | 0.962872982 | 0.290261 | 6.119268702 | 0 |
| 17 | 43106459 | T | C | K | R | BRCA1 | ENST00000471181 | 0.47879 | 0.08722 | 0.50791 | 0.93515 | 0.83725 | 0.3789100959 | 0.87694 | 0.8794846535 | 0.222917 | 2.34468578 | 0 |
| 17 | 43106459 | T | G | K | T | BRCA1 | ENST00000471181 | 0.6594 | 0.41468 | 0.59773 | 0.97735 | 0.88684 | 0.6130699428 | 0.87126 | 0.9589595795 | 0.26939 | 6.119268702 | 0 |
| 17 | 43106460 | T | C | K | E | BRCA1 | ENST00000471181 | 0.77085 | 0.35705 | 0.59338 | 0.97034 | 0.88354 | 0.6173444582 | 0.88311 | 0.9423453808 | 0.393128 | 6.119268702 | 0 |
| 17 | 43106460 | T | G | K | Q | BRCA1 | ENST00000471181 | 0.74874 | 0.35625 | 0.53448 | 0.9671 | 0.88117 | 0.5100302156 | 0.91476 | 0.9175157547 | 0.260896 | 6.030484745 | 0 |
| 17 | 43106462 | G | A | T | I | BRCA1 | ENST00000471181 | 0.74157 | 0.32879 | 0.7652 | 0.97807 | 0.86553 | 0.6744454742 | 0.98051 | 0.9685572982 | 0.339868 | 6.552315626 | 0 |
| 17 | 43106462 | G | C | T | S | BRCA1 | ENST00000471181 | 0.6978 | 0.25737 | 0.64419 | 0.96948 | 0.86753 | 0.422098043 | 0.98158 | 0.9361602664 | 0.28049 | 6.456512597 | 0 |
| 17 | 43106462 | G | T | T | N | BRCA1 | ENST00000471181 | 0.70423 | 0.38853 | 0.71514 | 0.97337 | 0.84588 | 0.5882916379 | 0.97499 | 0.9565452337 | 0.206832 | 6.02199239 | 0 |
| 17 | 43106463 | T | A | T | S | BRCA1 | ENST00000471181 | 0.63175 | 0.25737 | 0.64419 | 0.97794 | 0.87974 | 0.422098043 | 0.97488 | 0.8980134726 | 0.270172 | 6.456512597 | 0 |
| 17 | 43106463 | T | C | T | A | BRCA1 | ENST00000471181 | 0.66628 | 0.23658 | 0.63552 | 0.97768 | 0.87877 | 0.5959906377 | 0.97992 | 0.8980134726 | 0.296728 | 5.964324544 | 0 |
| 17 | 43106463 | T | G | T | P | BRCA1 | ENST00000471181 | 0.70799 | 0.44887 | 0.64378 | 0.98608 | 0.89648 | 0.7148054016 | 0.9739 | 0.9589595795 | 0.363033 | 6.552315626 | 0 |
| 17 | 43106464 | T | C | I | M | BRCA1 | ENST00000471181 | 0.58782 | 0.43065 | 0.57584 | 0.97894 | 0.86753 | 0.4549135368 | 0.96441 | 0.9500963688 | 0.269935 | 6.634437766 | 0 |
| 17 | 43106465 | A | C | I | R | BRCA1 | ENST00000471181 | 0.77593 | 0.50693 | 0.67583 | 0.98632 | 0.9506 | 0.7861419078 | 0.98167 | 0.968298018 | 0.419369 | 6.634437766 | 1 |
| 17 | 43106465 | A | G | I | T | BRCA1 | ENST00000471181 | 0.74866 | 0.13538 | 0.73507 | 0.97677 | 0.92601 | 0.5948376854 | 0.98833 | 0.9500447512 | 0.350568 | 6.634437766 | 0 |
| 17 | 43106465 | A | T | I | K | BRCA1 | ENST00000471181 | 0.76379 | 0.49151 | 0.69282 | 0.98569 | 0.9506 | 0.814989457 | 0.97799 | 0.9605305195 | 0.418643 | 6.634437766 | 1 |

| | | | | | | | | | | | | | | | | | | |
|---|---|---|---|---|---|---|---|---|---|---|---|---|---|---|---|---|---|---|
| 17 | 43106466 | T | A | I | L | BRCA1 | ENST00000471181 | 0.64229 | 0.20047 | 0.61231 | 0.96683 | 0.84159 | 0.4350578671 | 0.922 | 0.8290872574 | 0.219673 | 6.634437766 | 0 |
| 17 | 43106466 | T | C | I | V | BRCA1 | ENST00000471181 | 0.4286 | 0.10393 | 0.50956 | 0.91232 | 0.84588 | 0.2534169999 | 0.81575 | 0.3960707784 | 0.197833 | 3.065311656 | 0 |
| 17 | 43106466 | T | G | I | L | BRCA1 | ENST00000471181 | 0.65023 | 0.20047 | 0.61231 | 0.96683 | 0.84213 | 0.4350578671 | 0.922 | 0.8164978027 | 0.219674 | 6.634437766 | 0 |
| 17 | 43106467 | A | C | D | E | BRCA1 | ENST00000471181 | 0.22394 | 0.09232 | 0.22923 | 0.74301 | 0.82273 | 0.2148008572 | 0.82463 | 0.03310659682 | 0.189215 | 2.817297236 | 0 |
| 17 | 43106467 | A | T | D | E | BRCA1 | ENST00000471181 | 0.22054 | 0.09232 | 0.22923 | 0.74113 | 0.82273 | 0.2148008572 | 0.82463 | 0.03316472737 | 0.189164 | 2.817297236 | 0 |
| 17 | 43106468 | T | A | D | V | BRCA1 | ENST00000471181 | 0.47036 | 0.16742 | 0.20368 | 0.93213 | 0.8705 | 0.422934413 | 0.7553 | 0.3485430777 | 0.275775 | 5.543504085 | 0 |
| 17 | 43106468 | T | C | D | G | BRCA1 | ENST00000471181 | 0.3509 | 0.15812 | 0.22623 | 0.91637 | 0.83779 | 0.4869463125 | 0.81531 | 0.1506749243 | 0.251718 | 5.110334886 | 0 |
| 17 | 43106468 | T | G | D | A | BRCA1 | ENST00000471181 | 0.34669 | 0.11667 | 0.23062 | 0.89463 | 0.83283 | 0.3506731543 | 0.7897 | 0.2295843512 | 0.211003 | 2.928776072 | 0 |
| 17 | 43106469 | C | A | D | Y | BRCA1 | ENST00000471181 | 0.28261 | 0.11672 | 0.29127 | 0.90279 | 0.87345 | 0.4385609153 | 0.86345 | 0.03945846483 | 0.108459 | 5.543504085 | 0 |
| 17 | 43106469 | C | G | D | H | BRCA1 | ENST00000471181 | 0.27526 | 0.112 | 0.28499 | 0.87874 | 0.83943 | 0.352124401 | 0.84538 | 0.07631146163 | 0.214801 | 4.474160276 | 0 |
| 17 | 43106469 | C | T | D | N | BRCA1 | ENST00000471181 | 0.24198 | 0.09083 | 0.26267 | 0.78176 | 0.66922 | 0.269101437 | 0.77743 | 0.08278644085 | 0.107845 | 2.775445704 | 0 |
| 17 | 43106470 | A | C | N | K | BRCA1 | ENST00000471181 | 0.53859 | 0.35247 | 0.76649 | 0.93937 | 0.81761 | 0.2877859077 | 0.92451 | 0.918514967 | 0.142078 | 6.545542956 | 0 |
| 17 | 43106470 | A | T | N | K | BRCA1 | ENST00000471181 | 0.53293 | 0.35247 | 0.76649 | 0.93928 | 0.81818 | 0.2877859077 | 0.92451 | 0.918514967 | 0.142101 | 6.545542956 | 0 |
| 17 | 43106471 | T | A | N | I | BRCA1 | ENST00000471181 | 0.63912 | 0.45545 | 0.71361 | 0.98608 | 0.87442 | 0.4322160214 | 0.91484 | 0.9371104836 | 0.280225 | 6.545542956 | 0 |
| 17 | 43106471 | T | C | N | S | BRCA1 | ENST00000471181 | 0.30714 | 0.13456 | 0.63971 | 0.93358 | 0.77654 | 0.146127077 | 0.9119 | 0.7063105702 | 0.121359 | 3.572735956 | 0 |
| 17 | 43106471 | T | G | N | T | BRCA1 | ENST00000471181 | 0.32621 | 0.33401 | 0.68786 | 0.92223 | 0.8543 | 0.1973118775 | 0.89951 | 0.8236886263 | 0.182126 | 3.806157548 | 0 |
| 17 | 43106472 | T | A | N | Y | BRCA1 | ENST00000471181 | 0.76371 | 0.46 | 0.73211 | 0.98448 | 0.92356 | 0.3591254985 | 0.94966 | 0.9786425829 | 0.313839 | 6.545542956 | 1 |
| 17 | 43106472 | T | C | N | D | BRCA1 | ENST00000471181 | 0.71562 | 0.11413 | 0.74313 | 0.96274 | 0.85482 | 0.2698078635 | 0.95506 | 0.8944306374 | 0.177011 | 4.735693714 | 0 |
| 17 | 43106472 | T | G | N | H | BRCA1 | ENST00000471181 | 0.71962 | 0.43306 | 0.70548 | 0.97055 | 0.84906 | 0.2899433192 | 0.96363 | 0.9434948564 | 0.192268 | 6.545542956 | 0 |
| 17 | 43106473 | C | A | K | N | BRCA1 | ENST00000471181 | 0.70615 | 0.12323 | 0.74083 | 0.96681 | 0.8543 | 0.6774169467 | 0.9049 | 0.9509145617 | 0.193364 | 6.419077952 | 0 |
| 17 | 43106473 | C | G | K | N | BRCA1 | ENST00000471181 | 0.71466 | 0.12323 | 0.74083 | 0.96673 | 0.8543 | 0.6774169467 | 0.9049 | 0.9509145617 | 0.193366 | 6.419077952 | 0 |
| 17 | 43106474 | T | A | K | M | BRCA1 | ENST00000471181 | 0.75615 | 0.47607 | 0.63665 | 0.98443 | 0.91484 | 0.7032885754 | 0.92177 | 0.9719340205 | 0.3239 | 4.819184091 | 0 |
| 17 | 43106474 | T | C | K | R | BRCA1 | ENST00000471181 | 0.66161 | 0.08722 | 0.63832 | 0.95498 | 0.87925 | 0.4222882623 | 0.88694 | 0.876729846 | 0.240934 | 6.419077952 | 0 |
| 17 | 43106474 | T | G | K | T | BRCA1 | ENST00000471181 | 0.72252 | 0.37306 | 0.7191 | 0.98395 | 0.90755 | 0.6922029615 | 0.91715 | 0.9719340205 | 0.35742 | 6.419077952 | 0 |
| 17 | 43106475 | T | C | K | E | BRCA1 | ENST00000471181 | 0.78733 | 0.32581 | 0.70856 | 0.97764 | 0.87925 | 0.6734287862 | 0.90774 | 0.9229376316 | 0.235505 | 6.419077952 | 0 |
| 17 | 43106475 | T | G | K | Q | BRCA1 | ENST00000471181 | 0.76379 | 0.35625 | 0.66174 | 0.97571 | 0.87296 | 0.5495577666 | 0.90981 | 0.9719340205 | 0.276086 | 6.419077952 | 0 |
| 17 | 43106476 | A | C | C | W | BRCA1 | ENST00000471181 | 0.76211 | 0.49956 | 0.76582 | 0.99745 | 0.98621 | 0.8940653396 | 0.99926 | 0.9634407163 | 0.54574 | 6.675636481 | 1 |
| 17 | 43106477 | C | A | C | F | BRCA1 | ENST00000471181 | 0.62962 | 0.50959 | 0.71921 | 0.99535 | 0.98972 | 0.8875793477 | 0.99948 | 0.998311162 | 0.553375 | 6.675636481 | 1 |
| 17 | 43106477 | C | G | C | S | BRCA1 | ENST00000471181 | 0.59487 | 0.41048 | 0.69614 | 0.9973 | 0.98071 | 0.8626820059 | 0.99944 | 0.9973466396 | 0.553376 | 6.675636481 | 1 |
| 17 | 43106477 | C | T | C | Y | BRCA1 | ENST00000471181 | 0.61276 | 0.52834 | 0.82222 | 0.99628 | 0.99202 | 0.8871226589 | 0.99954 | 0.998311162 | 0.553373 | 6.675636481 | 1 |
| 17 | 43106478 | A | C | C | G | BRCA1 | ENST00000471181 | 0.75057 | 0.44742 | 0.66181 | 0.99708 | 0.98972 | 0.8553342639 | 0.9996 | 0.7738491297 | 0.431805 | 6.675636481 | 1 |
| 17 | 43106478 | A | G | C | R | BRCA1 | ENST00000471181 | 0.76467 | 0.48129 | 0.8061 | 0.99701 | 0.98657 | 0.9046337823 | 0.99962 | 0.9338114858 | 0.558556 | 6.675636481 | 1 |
| 17 | 43106478 | A | T | C | S | BRCA1 | ENST00000471181 | 0.70495 | 0.41048 | 0.69614 | 0.99767 | 0.9822 | 0.8626820059 | 0.99945 | 0.9188253284 | 0.558558 | 6.675636481 | 1 |
| 17 | 43106479 | T | A | L | F | BRCA1 | ENST00000471181 | 0.516 | 0.44443 | 0.52944 | 0.97605 | 0.85064 | 0.5290546243 | 0.90893 | 0.8668829799 | 0.239101 | 6.664488347 | 0 |
| 17 | 43106479 | T | G | L | F | BRCA1 | ENST00000471181 | 0.52281 | 0.44443 | 0.52944 | 0.97519 | 0.85064 | 0.5290546243 | 0.90893 | 0.9330402017 | 0.239103 | 6.664488347 | 0 |
| 17 | 43106480 | A | G | L | S | BRCA1 | ENST00000471181 | 0.74762 | 0.46671 | 0.57195 | 0.98193 | 0.91526 | 0.6385192552 | 0.94538 | 0.9483236074 | 0.408694 | 6.664488347 | 0 |
| 17 | 43106481 | A | C | L | V | BRCA1 | ENST00000471181 | 0.53771 | 0.28657 | 0.46454 | 0.95949 | 0.82103 | 0.4542876708 | 0.8006 | 0.5203748832 | 0.204441 | 6.508176334 | 0 |
| 17 | 43106481 | A | T | L | I | BRCA1 | ENST00000471181 | 0.54426 | 0.37848 | 0.54304 | 0.95662 | 0.82724 | 0.3223034462 | 0.79439 | 0.5688520427 | 0.125371 | 6.664488347 | 0 |
| 17 | 43106483 | G | A | P | L | BRCA1 | ENST00000471181 | 0.69147 | 0.11672 | 0.6801 | 0.97974 | 0.9796 | 0.8048285268 | 0.97221 | 0.9912266135 | 0.504376 | 6.67563828 | 0 |
| 17 | 43106483 | G | C | P | R | BRCA1 | ENST00000471181 | 0.67772 | 0.43845 | 0.67908 | 0.98084 | 0.97215 | 0.8662829799 | 0.98075 | 0.9990285635 | 0.50671 | 6.67563828 | 0 |
| 17 | 43106484 | G | A | P | S | BRCA1 | ENST00000471181 | 0.68011 | 0.39674 | 0.58327 | 0.98083 | 0.9681 | 0.8178648903 | 0.97404 | 0.9848254323 | 0.491001 | 6.67563828 | 0 |
| 17 | 43106484 | G | C | P | A | BRCA1 | ENST00000471181 | 0.62848 | 0.2809 | 0.57874 | 0.97874 | 0.95753 | 0.7249255834 | 0.97407 | 0.9774869084 | 0.406454 | 6.67563828 | 0 |
| 17 | 43106485 | A | C | C | W | BRCA1 | ENST00000471181 | 0.70968 | 0.49956 | 0.70781 | 0.99828 | 0.97512 | 0.9185455292 | 0.99804 | 0.997744441 | 0.544369 | 6.675636481 | 1 |
| 17 | 43106486 | C | A | C | F | BRCA1 | ENST00000471181 | 0.62143 | 0.50959 | 0.64865 | 0.99627 | 0.99265 | 0.8984901245 | 0.99865 | 0.9980607629 | 0.553754 | 6.675636481 | 1 |
| 17 | 43106486 | C | G | C | S | BRCA1 | ENST00000471181 | 0.58702 | 0.42762 | 0.63595 | 0.9976 | 0.98071 | 0.887695286 | 0.99641 | 0.9980456829 | 0.553707 | 6.675636481 | 1 |
| 17 | 43106486 | C | T | C | Y | BRCA1 | ENST00000471181 | 0.60471 | 0.52834 | 0.76753 | 0.99713 | 0.99386 | 0.9078823969 | 0.99778 | 0.9983478785 | 0.553751 | 6.675636481 | 1 |
| 17 | 43106487 | A | C | C | G | BRCA1 | ENST00000471181 | 0.70583 | 0.44742 | 0.59828 | 0.99762 | 0.99006 | 0.9059908076 | 0.99848 | 0.9838856459 | 0.433365 | 6.675636481 | 1 |
| 17 | 43106487 | A | T | C | S | BRCA1 | ENST00000471181 | 0.66027 | 0.42762 | 0.63595 | 0.99768 | 0.97885 | 0.887695286 | 0.99479 | 0.9969351292 | 0.558833 | 6.675636481 | 1 |
| 17 | 43106488 | C | A | Q | H | BRCA1 | ENST00000471181 | 0.32955 | 0.40813 | 0.37662 | 0.95161 | 0.82216 | 0.3605633185 | 0.7939 | 0.843886137 | 0.22044 | 6.574187885 | 0 |
| 17 | 43106488 | C | G | Q | H | BRCA1 | ENST00000471181 | 0.33521 | 0.40813 | 0.37662 | 0.95055 | 0.82216 | 0.3605633185 | 0.7939 | 0.843886137 | 0.220437 | 6.574187885 | 0 |
| 17 | 43106489 | T | A | Q | L | BRCA1 | ENST00000471181 | 0.73885 | 0.22979 | 0.31484 | 0.96967 | 0.89285 | 0.4571799461 | 0.91223 | 0.7796255866 | 0.296107 | 6.456700431 | 0 |
| 17 | 43106489 | T | C | Q | R | BRCA1 | ENST00000471181 | 0.69885 | 0.39782 | 0.38009 | 0.95853 | 0.88117 | 0.3291316622 | 0.88347 | 0.4606664788 | 0.17455 | 6.501641979 | 0 |
| 17 | 43106489 | T | G | Q | P | BRCA1 | ENST00000471181 | 0.72043 | 0.43757 | 0.34237 | 0.96873 | 0.89009 | 0.7600994495 | 0.89385 | 0.4380369144 | 0.393134 | 6.523296632 | 0 |
| 17 | 43106490 | G | C | Q | E | BRCA1 | ENST00000471181 | 0.5156 | 0.34087 | 0.33142 | 0.902 | 0.82387 | 0.3438595219 | 0.85299 | 0.5232063419 | 0.218615 | 4.158124375 | 0 |
| 17 | 43106490 | G | T | Q | K | BRCA1 | ENST00000471181 | 0.55121 | 0.36622 | 0.39052 | 0.94102 | 0.85741 | 0.3601275066 | 0.8607 | 0.7129054378 | 0.276635 | 4.490325864 | 0 |
| 17 | 43106492 | G | A | S | L | BRCA1 | ENST00000471181 | 0.50831 | 0.18665 | 0.43592 | 0.9114 | 0.87442 | 0.4534956297 | 0.86363 | 0.4259993407 | 0.277201 | 7.079527351 | 0 |
| 17 | 43106493 | A | C | S | A | BRCA1 | ENST00000471181 | 0.27388 | 0.10186 | 0.39533 | 0.89452 | 0.81933 | 0.2312936402 | 0.82178 | 0.3380980598 | 0.203388 | 4.489031076 | 0 |
| 17 | 43106493 | A | G | S | P | BRCA1 | ENST00000471181 | 0.4507 | 0.4245 | 0.4392 | 0.95115 | 0.87 | 0.6605944162 | 0.86073 | 0.7752785087 | 0.364337 | 5.542427484 | 0 |
| 17 | 43106493 | A | T | S | T | BRCA1 | ENST00000471181 | 0.46494 | 0.25087 | 0.43118 | 0.92488 | 0.81876 | 0.3908559588 | 0.83477 | 0.4894862161 | 0.221116 | 4.858049346 | 0 |
| 17 | 43106495 | G | A | P | L | BRCA1 | ENST00000471181 | 0.32115 | 0.2744 | 0.40681 | 0.95433 | 0.90711 | 0.3420486722 | 0.8992 | 0.2995494306 | 0.195688 | 3.634772003 | 0 |
| 17 | 43106495 | G | C | P | R | BRCA1 | ENST00000471181 | 0.55827 | 0.40026 | 0.4013 | 0.97315 | 0.86902 | 0.2742430317 | 0.90702 | 0.6767026782 | 0.240463 | 6.580980513 | 0 |
| 17 | 43106495 | G | T | P | H | BRCA1 | ENST00000471181 | 0.55704 | 0.45104 | 0.38647 | 0.9789 | 0.86951 | 0.3088791526 | 0.92083 | 0.8382867575 | 0.236087 | 6.580980513 | 1 |
| 17 | 43106496 | G | C | P | A | BRCA1 | ENST00000471181 | 0.46799 | 0.13875 | 0.32134 | 0.93953 | 0.89147 | 0.2411366386 | 0.90812 | 0.4925810397 | 0.301645 | 5.45883997 | 0 |
| 17 | 43106496 | G | T | P | T | BRCA1 | ENST00000471181 | 0.39058 | 0.288 | 0.35984 | 0.95808 | 0.86503 | 0.3301168145 | 0.89915 | 0.6120112538 | 0.280039 | 5.004476782 | 0 |
| 17 | 43106498 | C | A | G | V | BRCA1 | ENST00000471181 | 0.58332 | 0.47599 | 0.46044 | 0.967 | 0.87099 | 0.5124847785 | 0.90057 | 0.9541471601 | 0.252638 | 6.687487227 | 0 |
| 17 | 43106498 | C | G | G | A | BRCA1 | ENST00000471181 | 0.55731 | 0.40861 | 0.44986 | 0.93525 | 0.848 | 0.4006251857 | 0.87197 | 0.8174296021 | 0.215444 | 5.74531953 | 0 |
| 17 | 43106499 | C | G | G | R | BRCA1 | ENST00000471181 | 0.61746 | 0.47111 | 0.57489 | 0.94548 | 0.84588 | 0.4306747121 | 0.92346 | 0.9184286594 | 0.254212 | 6.687487227 | 0 |
| 17 | 43106500 | T | A | K | N | BRCA1 | ENST00000471181 | 0.6507 | 0.36295 | 0.56451 | 0.96161 | 0.80476 | 0.3963627613 | 0.71792 | 0.7734529458 | 0.0805298 | 4.160259029 | 0 |
| 17 | 43106500 | T | G | K | N | BRCA1 | ENST00000471181 | 0.65877 | 0.36295 | 0.56451 | 0.96183 | 0.80476 | 0.3963627613 | 0.71792 | 0.7746307236 | 0.0805312 | 4.160259029 | 0 |
| 17 | 43106501 | T | A | K | I | BRCA1 | ENST00000471181 | 0.70984 | 0.41463 | 0.50817 | 0.97523 | 0.91143 | 0.508647948 | 0.8466 | 0.8537909623 | 0.154685 | 6.025166925 | 0 |
| 17 | 43106501 | T | C | K | R | BRCA1 | ENST00000471181 | 0.47936 | 0.08936 | 0.46223 | 0.86833 | 0.85945 | 0.2401094571 | 0.72016 | 0.6866875291 | 0.177351 | 3.020440972 | 0 |
| 17 | 43106501 | T | G | K | T | BRCA1 | ENST00000471181 | 0.66826 | 0.36563 | 0.54787 | 0.95847 | 0.88776 | 0.4465951847 | 0.8006 | 0.712116333 | 0.121134 | 6.025166925 | 0 |
| 17 | 43106502 | T | C | K | E | BRCA1 | ENST00000471181 | 0.73981 | 0.34859 | 0.53135 | 0.92616 | 0.86852 | 0.3854381721 | 0.79706 | 0.8474942446 | 0.18637 | 6.025166925 | 0 |
| 17 | 43106502 | T | G | K | Q | BRCA1 | ENST00000471181 | 0.7173 | 0.34895 | 0.48383 | 0.93969 | 0.86302 | 0.3075503718 | 0.78901 | 0.6844492825 | 0.165786 | 6.025166925 | 0 |
| 17 | 43106503 | C | A | K | N | BRCA1 | ENST00000471181 | 0.56964 | 0.34172 | 0.50665 | 0.96487 | 0.8306 | 0.5280274979 | 0.85159 | 0.7028209716 | 0.156134 | 5.516514392 | 0 |
| 17 | 43106503 | C | G | K | N | BRCA1 | ENST00000471181 | 0.57709 | 0.34172 | 0.50665 | 0.96417 | 0.8306 | 0.5280274979 | 0.85159 | 0.6972354372 | 0.156135 | 5.516514392 | 0 |
| 17 | 43106504 | T | A | K | M | BRCA1 | ENST00000471181 | 0.67684 | 0.47607 | 0.38682 | 0.98053 | 0.91778 | 0.6156612659 | 0.88281 | 0.9269780517 | 0.197016 | 6.119682897 | 0 |
| 17 | 43106504 | T | C | K | R | BRCA1 | ENST00000471181 | 0.36805 | 0.08289 | 0.38792 | 0.82547 | 0.79577 | 0.3259634705 | 0.7669 | 0.1662745772 | 0.140641 | 2.636027944 | 0 |
| 17 | 43106504 | T | G | K | T | BRCA1 | ENST00000471181 | 0.64027 | 0.34449 | 0.49849 | 0.97215 | 0.87539 | 0.5819017264 | 0.85517 | 0.7010811929 | 0.180914 | 6.119682897 | 0 |
| 17 | 43106505 | T | C | K | E | BRCA1 | ENST00000471181 | 0.75439 | 0.29212 | 0.47076 | 0.95864 | 0.87829 | 0.5388997296 | 0.8501 | 0.6293765792 | 0.236278 | 5.453488475 | 0 |
| 17 | 43106505 | T | G | K | Q | BRCA1 | ENST00000471181 | 0.73221 | 0.35625 | 0.41109 | 0.95337 | 0.88165 | 0.4477243131 | 0.84959 | 0.6533076086 | 0.212173 | 6.119682897 | 0 |
| 17 | 43106506 | C | A | Q | H | BRCA1 | ENST00000471181 | 0.4074 | 0.0931 | 0.36747 | 0.93987 | 0.83997 | 0.4892234081 | 0.8831 | 0.6119066914 | 0.198691 | 6.893102872 | 0 |
| 17 | 43106506 | C | G | Q | H | BRCA1 | ENST00000471181 | 0.41363 | 0.0931 | 0.36747 | 0.9398 | 0.83997 | 0.4892234081 | 0.8831 | 0.5028559021 | 0.128637 | 6.893102872 | 0 |
| 17 | 43106507 | T | A | Q | L | BRCA1 | ENST00000471181 | 0.74682 | 0.27129 | 0.30417 | 0.96442 | 0.89147 | 0.5177994889 | 0.86613 | 0.6982655372 | 0.212125 | 6.893102872 | 0 |
| 17 | 43106507 | T | C | Q | R | BRCA1 | ENST00000471181 | 0.68267 | 0.19441 | 0.35864 | 0.91688 | 0.85222 | 0.3512501744 | 0.83181 | 0.5101328113 | 0.229668 | 4.856234841 | 0 |
| 17 | 43106507 | T | G | Q | P | BRCA1 | ENST00000471181 | 0.7278 | 0.3771 | 0.34042 | 0.97374 | 0.8933 | 0.6684117367 | 0.89646 | 0.9154081345 | 0.271696 | 6.893102872 | 0 |
| 17 | 43106508 | G | C | Q | E | BRCA1 | ENST00000471181 | 0.59443 | 0.16638 | 0.32929 | 0.91403 | 0.83116 | 0.4171156001 | 0.84891 | 0.5438986176 | 0.176488 | 6.233750423 | 0 |
| 17 | 43106508 | G | T | Q | K | BRCA1 | ENST00000471181 | 0.35311 | 0.12248 | 0.37348 | 0.84352 | 0.83004 | 0.4276910522 | 0.82526 | 0.3367268741 | 0.204101 | 3.896491859 | 0 |
| 17 | 43106509 | G | C | N | K | BRCA1 | ENST00000471181 | 0.68915 | 0.4617 | 0.55787 | 0.95627 | 0.85378 | 0.2051728798 | 0.69197 | 0.7224243239 | 0.0958031 | 6.232970205 | 0 |
| 17 | 43106510 | T | A | N | I | BRCA1 | ENST00000471181 | 0.67557 | 0.45461 | 0.48048 | 0.981 | 0.87 | 0.3994475694 | 0.79496 | 0.882457912 | 0.191548 | 6.232970205 | 0 |
| 17 | 43106510 | T | C | N | S | BRCA1 | ENST00000471181 | 0.4295 | 0.39859 | 0.39095 | 0.91509 | 0.73443 | 0.1557477557 | 0.65964 | 0.1390588433 | -0.0207936 | 3.692238902 | 0 |
| 17 | 43106511 | T | A | N | Y | BRCA1 | ENST00000471181 | 0.63099 | 0.47796 | 0.51126 | 0.98159 | 0.85843 | 0.3613085105 | 0.7799 | 0.883795619 | 0.205298 | 6.232970205 | 0 |
| 17 | 43106511 | T | C | N | D | BRCA1 | ENST00000471181 | 0.47382 | 0.45962 | 0.529 | 0.9281 | 0.78535 | 0.2470882869 | 0.73723 | 0.2695226145 | 0.0681435 | 3.469648946 | 0 |
| 17 | 43106511 | T | G | N | H | BRCA1 | ENST00000471181 | 0.58645 | 0.45084 | 0.47453 | 0.96459 | 0.78348 | 0.2474115482 | 0.73369 | 0.6743603725 | 0.10297 | 6.232970205 | 0 |
| 17 | 43106514 | G | A | L | F | BRCA1 | ENST00000471181 | 0.62316 | 0.46518 | 0.49467 | 0.96716 | 0.88543 | 0.4388326592 | 0.99051 | 0.2231394432 | -0.0451815 | 4.370063734 | 0 |
| 17 | 43106514 | G | C | L | V | BRCA1 | ENST00000471181 | 0.58674 | 0.45285 | 0.43465 | 0.96675 | 0.88684 | 0.4341795919 | 0.98441 | 0.9248082042 | 0.267517 | 6.720419347 | 0 |
| 17 | 43106514 | G | T | L | I | BRCA1 | ENST00000471181 | 0.59269 | 0.46381 | 0.51306 | 0.95684 | 0.85894 | 0.3108130879 | 0.96924 | 0.9327133894 | 0.176526 | 4.011656237 | 0 |
| 17 | 43106516 | A | C | L | R | BRCA1 | ENST00000471181 | 0.84195 | 0.4448 | 0.70667 | 0.98638 | 0.93083 | 0.8467047966 | 0.99198 | 0.9348634481 | 0.466027 | 6.488528524 | 0 |
| 17 | 43106516 | A | G | L | P | BRCA1 | ENST00000471181 | 0.84605 | 0.50493 | 0.74682 | 0.98828 | 0.94356 | 0.9141458765 | 0.99495 | 0.9355834723 | 0.500687 | 6.488528524 | 1 |
| 17 | 43106516 | A | T | L | H | BRCA1 | ENST00000471181 | 0.8249 | 0.47796 | 0.70544 | 0.98103 | 0.89513 | 0.7334126719 | 0.98783 | 0.8270638088 | 0.410809 | 6.392790909 | 0 |
| 17 | 43106517 | G | A | L | F | BRCA1 | ENST00000471181 | 0.57829 | 0.44443 | 0.59355 | 0.96421 | 0.88776 | 0.5363852076 | 0.98138 | 0.8236959577 | 0.267931 | 5.129273075 | 0 |

| | | | | | | | | | | | | | | | | |
|---|---|---|---|---|---|---|---|---|---|---|---|---|---|---|---|---|
| 17 | 43106517 | G | C | L | V | BRCA1 | ENST00000471181 | 0.66446 | 0.28657 | 0.53469 | 0.96651 | 0.88401 | 0.5357470991 | 0.97209 | 0.7787536383 | 0.300096 | 6.488528524 | 0 |
| 17 | 43106517 | G | T | L | I | BRCA1 | ENST00000471181 | 0.67525 | 0.37848 | 0.6102 | 0.9669 | 0.87394 | 0.4035422626 | 0.96628 | 0.9045881629 | 0.220177 | 6.488528524 | 0 |
| 17 | 43106518 | T | A | K | N | BRCA1 | ENST00000471181 | 0.69548 | 0.39253 | 0.66158 | 0.96218 | 0.84588 | 0.7073771129 | 0.89093 | 0.7882775847 | 0.101998 | 5.736494633 | 0 |
| 17 | 43106518 | T | G | K | N | BRCA1 | ENST00000471181 | 0.70374 | 0.39253 | 0.66158 | 0.96 | 0.84535 | 0.7073771129 | 0.89093 | 0.7915380401 | 0.102001 | 5.736494633 | 0 |
| 17 | 43106519 | T | A | K | I | BRCA1 | ENST00000471181 | 0.84302 | 0.44869 | 0.59077 | 0.97284 | 0.91484 | 0.7582235867 | 0.94066 | 0.9564186931 | 0.257746 | 6.687517143 | 0 |
| 17 | 43106519 | T | C | K | R | BRCA1 | ENST00000471181 | 0.73789 | 0.08722 | 0.54769 | 0.9283 | 0.83172 | 0.4515577147 | 0.9048 | 0.6917782709 | 0.217862 | 5.667628446 | 0 |
| 17 | 43106519 | T | G | K | T | BRCA1 | ENST00000471181 | 0.7984 | 0.39535 | 0.64047 | 0.96116 | 0.87588 | 0.6821606274 | 0.91759 | 0.8452923339 | 0.256173 | 6.687517143 | 0 |
| 17 | 43106520 | T | C | K | E | BRCA1 | ENST00000471181 | 0.77505 | 0.35705 | 0.62533 | 0.95114 | 0.84641 | 0.664416498 | 0.89814 | 0.6756175908 | 0.268943 | 5.426993225 | 0 |
| 17 | 43106520 | T | G | K | Q | BRCA1 | ENST00000471181 | 0.75153 | 0.35625 | 0.57789 | 0.95632 | 0.84213 | 0.5892115981 | 0.90956 | 0.7636390062 | 0.218142 | 4.976017325 | 0 |
| 17 | 43106522 | A | C | L | R | BRCA1 | ENST00000471181 | 0.77283 | 0.49944 | 0.59647 | 0.95207 | 0.86902 | 0.6793106843 | 0.95659 | 0.7568658321 | 0.237325 | 6.766065965 | 0 |
| 17 | 43106522 | A | G | L | P | BRCA1 | ENST00000471181 | 0.77703 | 0.52579 | 0.64213 | 0.98179 | 0.91143 | 0.8588869154 | 0.98417 | 0.9580077529 | 0.396789 | 6.766065965 | 1 |
| 17 | 43106522 | A | T | L | Q | BRCA1 | ENST00000471181 | 0.75543 | 0.4416 | 0.50358 | 0.96759 | 0.848 | 0.4880057457 | 0.96214 | 0.9220617414 | 0.347274 | 6.766065965 | 0 |
| 17 | 43106523 | G | T | L | M | BRCA1 | ENST00000471181 | 0.63842 | 0.24327 | 0.47508 | 0.92754 | 0.82613 | 0.3817768672 | 0.88548 | 0.7367253356 | 0.171532 | 5.772919066 | 0 |
| 17 | 43106524 | C | A | M | I | BRCA1 | ENST00000471181 | 0.56117 | 0.10566 | 0.54441 | 0.87926 | 0.80536 | 0.6078354349 | 0.80718 | 0.8777412176 | 0.207859 | 4.198614314 | 0 |
| 17 | 43106524 | C | G | M | I | BRCA1 | ENST00000471181 | 0.56852 | 0.10566 | 0.54441 | 0.87516 | 0.80536 | 0.6078354349 | 0.80718 | 0.8296602964 | 0.20786 | 4.198614314 | 0 |
| 17 | 43106524 | C | T | M | I | BRCA1 | ENST00000471181 | 0.57329 | 0.10566 | 0.54441 | 0.87568 | 0.80536 | 0.6078354349 | 0.80718 | 0.8777412176 | 0.207849 | 4.198614314 | 0 |
| 17 | 43106525 | A | C | M | R | BRCA1 | ENST00000471181 | 0.69997 | 0.38584 | 0.61997 | 0.9646 | 0.91569 | 0.9381597513 | 0.95756 | 0.9580696821 | 0.410289 | 6.368404467 | 1 |
| 17 | 43106525 | A | G | M | T | BRCA1 | ENST00000471181 | 0.62985 | 0.17688 | 0.68428 | 0.95155 | 0.90229 | 0.7824993728 | 0.95309 | 0.9333351254 | 0.363476 | 6.368404467 | 0 |
| 17 | 43106525 | A | T | M | K | BRCA1 | ENST00000471181 | 0.68428 | 0.35356 | 0.64373 | 0.96473 | 0.90755 | 0.9236867797 | 0.95817 | 0.9428873062 | 0.410656 | 6.368404467 | 1 |
| 17 | 43106526 | T | A | M | L | BRCA1 | ENST00000471181 | 0.61499 | 0.09759 | 0.54884 | 0.92094 | 0.84428 | 0.5813152456 | 0.8387 | 0.7349604964 | 0.216538 | 4.865637954 | 0 |
| 17 | 43106526 | T | C | M | V | BRCA1 | ENST00000471181 | 0.35025 | 0.09501 | 0.4709 | 0.94212 | 0.86753 | 0.6030539527 | 0.88083 | 0.7733387351 | 0.255864 | 6.368404467 | 0 |
| 17 | 43106526 | T | G | M | L | BRCA1 | ENST00000471181 | 0.62263 | 0.09759 | 0.54884 | 0.92094 | 0.84428 | 0.5813152456 | 0.8387 | 0.689422965 | 0.21654 | 4.865637954 | 0 |
| 17 | 43106527 | G | C | C | W | BRCA1 | ENST00000471181 | 0.59762 | 0.46613 | 0.63718 | 0.99841 | 0.96553 | 0.9064099566 | 0.99783 | 0.9972888231 | 0.534793 | 6.530946705 | 1 |
| 17 | 43106528 | C | A | C | F | BRCA1 | ENST00000471181 | 0.64423 | 0.15377 | 0.59127 | 0.99515 | 0.97885 | 0.8854062096 | 0.99954 | 0.9923970103 | 0.562575 | 6.530946705 | 1 |
| 17 | 43106528 | C | G | C | S | BRCA1 | ENST00000471181 | 0.5695 | 0.12551 | 0.56703 | 0.99609 | 0.97438 | 0.8504917898 | 0.99875 | 0.9980688691 | 0.552782 | 6.530946705 | 1 |
| 17 | 43106528 | C | T | C | Y | BRCA1 | ENST00000471181 | 0.62704 | 0.4057 | 0.70586 | 0.99676 | 0.9796 | 0.8989645642 | 0.99905 | 0.9977073669 | 0.562575 | 6.530946705 | 1 |
| 17 | 43106529 | A | C | C | G | BRCA1 | ENST00000471181 | 0.73685 | 0.2663 | 0.53434 | 0.99618 | 0.98728 | 0.8672005302 | 0.99967 | 0.9977038503 | 0.568059 | 6.530946705 | 1 |
| 17 | 43106529 | A | G | C | R | BRCA1 | ENST00000471181 | 0.75296 | 0.46746 | 0.68102 | 0.99648 | 0.98657 | 0.9258664453 | 0.99966 | 0.9966281056 | 0.568053 | 6.530946705 | 1 |
| 17 | 43106530 | A | C | F | L | BRCA1 | ENST00000471181 | 0.48602 | 0.42123 | 0.73575 | 0.94702 | 0.85638 | 0.5302113774 | 0.93254 | 0.7485562707 | 0.250125 | 3.852068698 | 0 |
| 17 | 43106530 | A | T | F | L | BRCA1 | ENST00000471181 | 0.48063 | 0.42123 | 0.73575 | 0.94701 | 0.85638 | 0.5302113774 | 0.93254 | 0.7579231392 | 0.25012 | 3.852068698 | 0 |
| 17 | 43106531 | A | C | F | C | BRCA1 | ENST00000471181 | 0.74316 | 0.50693 | 0.7378 | 0.97239 | 0.93436 | 0.5952843764 | 0.99077 | 0.9713304639 | 0.410166 | 6.0991475 | 0 |
| 17 | 43106531 | A | G | F | S | BRCA1 | ENST00000471181 | 0.74316 | 0.52896 | 0.69633 | 0.95918 | 0.87491 | 0.5867724174 | 0.98307 | 0.9549073577 | 0.395624 | 6.0991475 | 0 |
| 17 | 43106531 | A | T | F | Y | BRCA1 | ENST00000471181 | 0.65555 | 0.45989 | 0.72294 | 0.94476 | 0.86803 | 0.4201475688 | 0.94448 | 0.7855051648 | 0.241541 | 6.0991475 | 0 |
| 17 | 43106532 | A | C | F | V | BRCA1 | ENST00000471181 | 0.7954 | 0.47766 | 0.67882 | 0.95703 | 0.88354 | 0.5893033269 | 0.98617 | 0.8860567808 | 0.359832 | 6.0991475 | 0 |
| 17 | 43106532 | A | G | F | L | BRCA1 | ENST00000471181 | 0.65062 | 0.42123 | 0.73575 | 0.93974 | 0.87394 | 0.5302113774 | 0.96972 | 0.8436216712 | 0.272433 | 3.852068698 | 0 |
| 17 | 43106532 | A | T | F | I | BRCA1 | ENST00000471181 | 0.78249 | 0.48858 | 0.69749 | 0.95504 | 0.88823 | 0.6412513596 | 0.96238 | 0.90370363 | 0.265329 | 6.0991475 | 0 |
| 17 | 43106533 | T | A | K | N | BRCA1 | ENST00000471181 | 0.46793 | 0.10942 | 0.39774 | 0.94201 | 0.81298 | 0.6521103436 | 0.89369 | 0.8868661523 | 0.17412 | 6.239416086 | 0 |
| 17 | 43106533 | T | G | K | N | BRCA1 | ENST00000471181 | 0.45783 | 0.10942 | 0.39774 | 0.94076 | 0.81298 | 0.6521103436 | 0.89369 | 0.8868661523 | 0.174121 | 6.239416086 | 0 |
| 17 | 43115726 | T | A | K | I | BRCA1 | ENST00000471181 | 0.56373 | 0.34443 | 0.33681 | 0.99131 | 0.88212 | 0.7109868453 | 0.91576 | 0.8929001689 | 0.248727 | 6.239416086 | 1 |
| 17 | 43115726 | T | C | K | R | BRCA1 | ENST00000471181 | 0.24284 | 0.07553 | 0.29951 | 0.9314 | 0.76881 | 0.3941595886 | 0.51857 | 0.07732956111 | 0.129126 | 2.371866056 | 0 |
| 17 | 43115726 | T | G | K | T | BRCA1 | ENST00000471181 | 0.50283 | 0.21039 | 0.39446 | 0.98141 | 0.85996 | 0.6560061141 | 0.76066 | 0.2686519295 | 0.225128 | 6.239416086 | 0 |
| 17 | 43115727 | T | C | K | E | BRCA1 | ENST00000471181 | 0.82334 | 0.16702 | 0.36165 | 0.9284 | 0.82273 | 0.6316293037 | 0.90833 | 0.7304461598 | 0.299557 | 6.239416086 | 0 |
| 17 | 43115727 | T | G | K | Q | BRCA1 | ENST00000471181 | 0.80668 | 0.16946 | 0.30579 | 0.89637 | 0.82892 | 0.5531403288 | 0.88529 | 0.2982670367 | 0.186907 | 6.239416086 | 0 |
| 17 | 43115728 | G | C | C | W | BRCA1 | ENST00000471181 | 0.91831 | 0.46653 | 0.65436 | 0.97148 | 0.98477 | 0.9226276907 | 0.99919 | 0.9979792237 | 0.56591 | 6.553294284 | 1 |
| 17 | 43115729 | C | A | C | F | BRCA1 | ENST00000471181 | 0.68267 | 0.17153 | 0.60339 | 0.98174 | 0.98477 | 0.8982672403 | 0.9995 | 0.9906427264 | 0.562895 | 6.553294284 | 1 |
| 17 | 43115729 | C | G | C | S | BRCA1 | ENST00000471181 | 0.64804 | 0.41964 | 0.57313 | 0.97916 | 0.98183 | 0.8869805142 | 0.99842 | 0.9976931214 | 0.562848 | 6.553294284 | 1 |
| 17 | 43115729 | C | T | C | Y | BRCA1 | ENST00000471181 | 0.61551 | 0.18824 | 0.71483 | 0.97762 | 0.95535 | 0.9204042333 | 0.99892 | 0.9872590899 | 0.562886 | 6.03413286 | 1 |
| 17 | 43115730 | A | C | C | G | BRCA1 | ENST00000471181 | 0.74635 | 0.44134 | 0.55114 | 0.98603 | 0.99234 | 0.9119822817 | 0.99962 | 0.9585968256 | 0.561248 | 6.553294284 | 1 |
| 17 | 43115730 | A | G | C | R | BRCA1 | ENST00000471181 | 0.76076 | 0.51777 | 0.68461 | 0.98651 | 0.99105 | 0.9265915866 | 0.99962 | 0.9969582558 | 0.564566 | 6.454406029 | 1 |
| 17 | 43115730 | A | T | C | S | BRCA1 | ENST00000471181 | 0.70021 | 0.41964 | 0.57313 | 0.98296 | 0.99138 | 0.8869805142 | 0.9987 | 0.9979967475 | 0.564518 | 6.553294284 | 1 |
| 17 | 43115731 | A | T | F | L | BRCA1 | ENST00000471181 | 0.64734 | 0.21991 | 0.80239 | 0.95078 | 0.93515 | 0.7788799594 | 0.96484 | 0.8893490434 | 0.421299 | 6.414530199 | 1 |
| 17 | 43115732 | A | T | F | Y | BRCA1 | ENST00000471181 | 0.67279 | 0.44238 | 0.79092 | 0.88403 | 0.87442 | 0.6727044161 | 0.91381 | 0.7772912618 | 0.316178 | 6.414530199 | 0 |
| 17 | 43115734 | T | C | I | M | BRCA1 | ENST00000471181 | 0.65791 | 0.43385 | 0.58745 | 0.94646 | 0.84959 | 0.3116320926 | 0.97302 | 0.866387248 | 0.352634 | 5.918107353 | 0 |
| 17 | 43115735 | A | C | I | R | BRCA1 | ENST00000471181 | 0.73869 | 0.26827 | 0.68627 | 0.93588 | 0.91945 | 0.5424839443 | 0.99035 | 0.7027803098 | 0.450447 | 5.933239561 | 0 |
| 17 | 43115735 | A | G | I | T | BRCA1 | ENST00000471181 | 0.71217 | 0.43254 | 0.75042 | 0.92987 | 0.84321 | 0.2899207761 | 0.98108 | 0.9713988304 | 0.407966 | 4.707294625 | 0 |
| 17 | 43115735 | A | T | I | K | BRCA1 | ENST00000471181 | 0.72668 | 0.47723 | 0.70766 | 0.94794 | 0.92151 | 0.5895003905 | 0.99076 | 0.7849817177 | 0.456006 | 6.880301456 | 0 |
| 17 | 43115736 | T | A | I | L | BRCA1 | ENST00000471181 | 0.61933 | 0.30694 | 0.628 | 0.85543 | 0.82159 | 0.3304077551 | 0.93511 | 0.666590049 | 0.299156 | 5.505059289 | 0 |
| 17 | 43115736 | T | C | I | V | BRCA1 | ENST00000471181 | 0.56609 | 0.31578 | 0.52932 | 0.77427 | 0.77399 | 0.1887360485 | 0.90849 | 0.7323348522 | 0.198342 | 4.692964812 | 0 |
| 17 | 43115736 | T | G | I | L | BRCA1 | ENST00000471181 | 0.62711 | 0.30694 | 0.628 | 0.84994 | 0.82159 | 0.3304077551 | 0.93511 | 0.6649164533 | 0.299158 | 5.505059289 | 0 |
| 17 | 43115737 | G | C | H | Q | BRCA1 | ENST00000471181 | 0.56554 | 0.43557 | 0.64109 | 0.99353 | 0.96153 | 0.9140455545 | 0.99306 | 0.9978321195 | 0.553458 | 6.56441227 | 1 |
| 17 | 43115737 | G | T | H | Q | BRCA1 | ENST00000471181 | 0.55985 | 0.43557 | 0.64109 | 0.99355 | 0.96153 | 0.9140455545 | 0.99306 | 0.9978321195 | 0.553452 | 6.56441227 | 1 |
| 17 | 43115738 | T | A | H | L | BRCA1 | ENST00000471181 | 0.66818 | 0.46964 | 0.62974 | 0.99138 | 0.99296 | 0.9393494172 | 0.99554 | 0.9925912619 | 0.562833 | 6.56441227 | 1 |
| 17 | 43115738 | T | C | H | R | BRCA1 | ENST00000471181 | 0.62195 | 0.46692 | 0.66788 | 0.99129 | 0.99498 | 0.9435604894 | 0.99633 | 0.998141408 | 0.562828 | 6.56441227 | 1 |
| 17 | 43115738 | T | G | H | P | BRCA1 | ENST00000471181 | 0.66652 | 0.5168 | 0.59864 | 0.99014 | 0.99138 | 0.9546586625 | 0.99684 | 0.9908818007 | 0.562883 | 6.56441227 | 1 |
| 17 | 43115739 | G | A | H | Y | BRCA1 | ENST00000471181 | 0.67406 | 0.45783 | 0.6706 | 0.99012 | 0.97327 | 0.9297748439 | 0.99384 | 0.9952718616 | 0.563492 | 6.391273675 | 1 |
| 17 | 43115739 | G | C | H | D | BRCA1 | ENST00000471181 | 0.72059 | 0.17441 | 0.66322 | 0.99222 | 0.98257 | 0.9677321252 | 0.99644 | 0.996338129 | 0.56591 | 6.56441227 | 1 |
| 17 | 43115739 | G | T | H | N | BRCA1 | ENST00000471181 | 0.68611 | 0.40477 | 0.66606 | 0.99168 | 0.95972 | 0.8774377382 | 0.99406 | 0.9978985786 | 0.386214 | 6.56441227 | 1 |
| 17 | 43115740 | G | C | D | E | BRCA1 | ENST00000471181 | 0.53267 | 0.42957 | 0.69564 | 0.92772 | 0.8517 | 0.5432249223 | 0.80459 | 0.9295815229 | 0.229233 | 6.543660463 | 0 |
| 17 | 43115740 | G | T | D | E | BRCA1 | ENST00000471181 | 0.52719 | 0.42957 | 0.69564 | 0.92698 | 0.85222 | 0.5432249223 | 0.80459 | 0.9295815229 | 0.229229 | 6.543660463 | 0 |
| 17 | 43115741 | T | A | D | V | BRCA1 | ENST00000471181 | 0.72925 | 0.49835 | 0.64689 | 0.93069 | 0.9252 | 0.8215515202 | 0.91837 | 0.8493895905 | 0.400881 | 6.543660463 | 0 |
| 17 | 43115741 | T | C | D | G | BRCA1 | ENST00000471181 | 0.74643 | 0.48572 | 0.67212 | 0.88578 | 0.87829 | 0.7170289417 | 0.82825 | 0.9001214504 | 0.383747 | 5.878360764 | 0 |
| 17 | 43115741 | T | G | D | A | BRCA1 | ENST00000471181 | 0.73934 | 0.45833 | 0.66419 | 0.90623 | 0.90052 | 0.7565693549 | 0.89928 | 0.8690462708 | 0.391688 | 6.088247352 | 0 |
| 17 | 43115742 | C | A | D | Y | BRCA1 | ENST00000471181 | 0.70519 | 0.48241 | 0.7341 | 0.89293 | 0.91442 | 0.7986324839 | 0.9417 | 0.8489458308 | 0.400989 | 6.543660463 | 0 |
| 17 | 43115742 | C | G | D | H | BRCA1 | ENST00000471181 | 0.69885 | 0.47571 | 0.75425 | 0.88729 | 0.89648 | 0.7549973087 | 0.93129 | 0.8592190742 | 0.392112 | 6.543660463 | 0 |
| 17 | 43115742 | C | T | D | N | BRCA1 | ENST00000471181 | 0.70639 | 0.44363 | 0.72483 | 0.89337 | 0.79151 | 0.5927581214 | 0.87806 | 0.7225487461 | 0.102433 | 6.514276351 | 0 |
| 17 | 43115743 | A | C | C | W | BRCA1 | ENST00000471181 | 0.73781 | 0.50075 | 0.73629 | 0.99385 | 0.9822 | 0.9225236093 | 0.99596 | 0.9991029501 | 0.553429 | 6.565801107 | 1 |
| 17 | 43115744 | C | A | C | F | BRCA1 | ENST00000471181 | 0.67525 | 0.51605 | 0.68539 | 0.99 | 0.98764 | 0.9232333959 | 0.99902 | 0.9958381653 | 0.562895 | 6.565801107 | 1 |
| 17 | 43115744 | C | G | C | S | BRCA1 | ENST00000471181 | 0.6385 | 0.43882 | 0.67205 | 0.99087 | 0.97587 | 0.9055423539 | 0.99836 | 0.9980086684 | 0.562848 | 6.565801107 | 1 |
| 17 | 43115744 | C | T | C | Y | BRCA1 | ENST00000471181 | 0.65751 | 0.53471 | 0.79708 | 0.98682 | 0.9844 | 0.9194013715 | 0.99916 | 0.9992095232 | 0.562886 | 6.565801107 | 1 |
| 17 | 43115745 | A | C | C | G | BRCA1 | ENST00000471181 | 0.75009 | 0.46372 | 0.63158 | 0.99445 | 0.98728 | 0.9023610108 | 0.99935 | 0.9959942102 | 0.564889 | 6.565801107 | 1 |
| 17 | 43115745 | A | G | C | R | BRCA1 | ENST00000471181 | 0.76418 | 0.50888 | 0.78017 | 0.99314 | 0.98513 | 0.940454708 | 0.99933 | 0.9988405108 | 0.564887 | 6.565801107 | 1 |
| 17 | 43115745 | A | T | C | S | BRCA1 | ENST00000471181 | 0.70447 | 0.43882 | 0.67205 | 0.9918 | 0.98146 | 0.9055423539 | 0.99839 | 0.9974339604 | 0.564839 | 6.565801107 | 1 |
| 17 | 43115746 | C | A | K | N | BRCA1 | ENST00000471181 | 0.48685 | 0.4238 | 0.6248 | 0.97346 | 0.78597 | 0.5732879168 | 0.5288 | 0.824539125 | 0.0625387 | 6.173507059 | 0 |
| 17 | 43115746 | C | G | K | N | BRCA1 | ENST00000471181 | 0.49363 | 0.4238 | 0.6248 | 0.97009 | 0.78597 | 0.5732879168 | 0.5288 | 0.824539125 | 0.0625345 | 6.173507059 | 0 |
| 17 | 43115747 | T | A | K | M | BRCA1 | ENST00000471181 | 0.61432 | 0.4863 | 0.47875 | 0.88399 | 0.86302 | 0.6128682798 | 0.86714 | 0.8905242085 | 0.183012 | 6.173507059 | 0 |
| 17 | 43115747 | T | C | K | R | BRCA1 | ENST00000471181 | 0.43993 | 0.11309 | 0.48484 | 0.67455 | 0.81588 | 0.3353464474 | 0.82696 | 0.298636833 | 0.113675 | 3.128632371 | 0 |
| 17 | 43115747 | T | G | K | T | BRCA1 | ENST00000471181 | 0.46531 | 0.1914 | 0.59984 | 0.77667 | 0.83116 | 0.5809227023 | 0.77793 | 0.3669247329 | 0.167497 | 4.000690433 | 0 |
| 17 | 43115748 | T | C | K | E | BRCA1 | ENST00000471181 | 0.67358 | 0.40224 | 0.58651 | 0.84084 | 0.82556 | 0.5960777257 | 0.82851 | 0.8790517449 | 0.216629 | 6.173507059 | 0 |
| 17 | 43115748 | T | G | K | Q | BRCA1 | ENST00000471181 | 0.62856 | 0.40936 | 0.52819 | 0.78336 | 0.82216 | 0.4587173031 | 0.88403 | 0.8132905364 | 0.172948 | 6.173507059 | 0 |
| 17 | 43115750 | G | C | T | R | BRCA1 | ENST00000471181 | 0.62985 | 0.46317 | 0.71691 | 0.96446 | 0.93631 | 0.8887893534 | 0.97117 | 0.9909331799 | 0.479082 | 6.382029316 | 1 |
| 17 | 43115750 | G | T | T | K | BRCA1 | ENST00000471181 | 0.64074 | 0.46837 | 0.73843 | 0.96445 | 0.94431 | 0.8804758212 | 0.98003 | 0.9926330447 | 0.478281 | 6.382029316 | 1 |
| 17 | 43115751 | T | A | T | S | BRCA1 | ENST00000471181 | 0.56373 | 0.26741 | 0.63324 | 0.94601 | 0.89739 | 0.5797923069 | 0.97536 | 0.8955072761 | 0.319744 | 6.382029316 | 0 |
| 17 | 43115751 | T | C | T | A | BRCA1 | ENST00000471181 | 0.58916 | 0.38845 | 0.60641 | 0.92956 | 0.91015 | 0.7020170682 | 0.97408 | 0.8140078187 | 0.40855 | 6.382029316 | 0 |
| 17 | 43115751 | T | G | T | P | BRCA1 | ENST00000471181 | 0.4708 | 0.48794 | 0.62916 | 0.96989 | 0.94356 | 0.8476921247 | 0.9656 | 0.9742149115 | 0.465013 | 5.887449591 | 0 |
| 17 | 43115754 | A | C | S | A | BRCA1 | ENST00000471181 | 0.6489 | 0.30031 | 0.65343 | 0.94451 | 0.87198 | 0.3231128526 | 0.97601 | 0.8827190399 | 0.318948 | 6.565743261 | 0 |
| 17 | 43115754 | A | T | S | T | BRCA1 | ENST00000471181 | 0.60978 | 0.39751 | 0.68104 | 0.93087 | 0.84641 | 0.4013353579 | 0.97428 | 0.8843592405 | 0.34934 | 6.565743261 | 0 |
| 17 | 43115756 | A | C | V | G | BRCA1 | ENST00000471181 | 0.71225 | 0.46828 | 0.48226 | 0.97315 | 0.94205 | 0.858408909 | 0.98777 | 0.9733402133 | 0.441945 | 6.335062213 | 1 |
| 17 | 43115756 | A | G | V | A | BRCA1 | ENST00000471181 | 0.67827 | 0.35258 | 0.63541 | 0.94335 | 0.89376 | 0.5907637038 | 0.98043 | 0.9687743187 | 0.456911 | 6.335062213 | 1 |
| 17 | 43115756 | A | T | V | D | BRCA1 | ENST00000471181 | 0.72909 | 0.50788 | 0.64719 | 0.96975 | 0.94506 | 0.9038512846 | 0.98814 | 0.9859365225 | 0.526191 | 6.335062213 | 1 |
| 17 | 43115757 | C | T | V | I | BRCA1 | ENST00000471181 | 0.35636 | 0.17535 | 0.4109 | 0.86428 | 0.79334 | 0.3375084447 | 0.95323 | 0.7645992637 | 0.156804 | 3.337477946 | 0 |
| 17 | 43115759 | G | A | P | L | BRCA1 | ENST00000471181 | 0.67708 | 0.44821 | 0.69464 | 0.96317 | 0.94949 | 0.8095381793 | 0.97629 | 0.9888013601 | 0.478941 | 6.566327159 | 1 |

| | | | | | | | | | | | | | | | | | |
|---|---|---|---|---|---|---|---|---|---|---|---|---|---|---|---|---|---|
| 17 | 43115759 | G | C | P | R | BRCA1 | ENST00000471181 | 0.66074 | 0.45342 | 0.69295 | 0.96417 | 0.95426 | 0.8249037894 | 0.99046 | 0.9921758771 | 0.496977 | 6.566327159 | 1 |
| 17 | 43115759 | G | T | P | H | BRCA1 | ENST00000471181 | 0.65869 | 0.47184 | 0.67541 | 0.95047 | 0.9458 | 0.788518389 | 0.99389 | 0.9973959923 | 0.512307 | 6.566327159 | 1 |
| 17 | 43115760 | G | A | P | S | BRCA1 | ENST00000471181 | 0.71962 | 0.43965 | 0.601 | 0.96449 | 0.95316 | 0.7668592371 | 0.99267 | 0.9936443567 | 0.495494 | 6.566327159 | 1 |
| 17 | 43115760 | G | C | P | A | BRCA1 | ENST00000471181 | 0.67016 | 0.42685 | 0.5911 | 0.95846 | 0.93554 | 0.660368709 | 0.98924 | 0.9710563421 | 0.43682 | 6.566327159 | 0 |
| 17 | 43115761 | T | A | E | D | BRCA1 | ENST00000471181 | 0.58902 | 0.4037 | 0.65135 | 0.91175 | 0.83725 | 0.2753448109 | 0.84217 | 0.7453271855 | 0.228895 | 6.494519319 | 0 |
| 17 | 43115761 | T | G | E | D | BRCA1 | ENST00000471181 | 0.59639 | 0.4037 | 0.65135 | 0.9119 | 0.83725 | 0.2753448109 | 0.84217 | 0.8408778906 | 0.228897 | 6.494519319 | 0 |
| 17 | 43115762 | T | A | E | V | BRCA1 | ENST00000471181 | 0.75655 | 0.47006 | 0.62861 | 0.91854 | 0.92601 | 0.597622161 | 0.90116 | 0.9771971107 | 0.409293 | 6.560046108 | 0 |
| 17 | 43115762 | T | G | E | A | BRCA1 | ENST00000471181 | 0.72877 | 0.44083 | 0.65536 | 0.90128 | 0.89694 | 0.4653543357 | 0.90312 | 0.9750452638 | 0.407066 | 6.560046108 | 0 |
| 17 | 43115763 | C | G | E | Q | BRCA1 | ENST00000471181 | 0.66067 | 0.42978 | 0.63684 | 0.83175 | 0.81876 | 0.404233059 | 0.90272 | 0.7717859153 | 0.260469 | 6.560046108 | 0 |
| 17 | 43115763 | C | T | E | K | BRCA1 | ENST00000471181 | 0.72836 | 0.24205 | 0.75789 | 0.86108 | 0.86703 | 0.4334182878 | 0.89574 | 0.9175257683 | 0.39522 | 6.560046108 | 0 |
| 17 | 43115764 | C | A | K | N | BRCA1 | ENST00000471181 | 0.53859 | 0.44955 | 0.62443 | 0.96381 | 0.85117 | 0.4686748902 | 0.91349 | 0.7841031605 | 0.158911 | 6.509408713 | 0 |
| 17 | 43115764 | C | G | K | N | BRCA1 | ENST00000471181 | 0.54576 | 0.44955 | 0.62443 | 0.96637 | 0.85117 | 0.4686748902 | 0.91349 | 0.8866645098 | 0.158912 | 6.509408713 | 0 |
| 17 | 43115765 | T | A | K | M | BRCA1 | ENST00000471181 | 0.70038 | 0.47979 | 0.50242 | 0.96481 | 0.90494 | 0.5211073327 | 0.93986 | 0.9609534144 | 0.279757 | 6.509408713 | 0 |
| 17 | 43115765 | T | C | K | R | BRCA1 | ENST00000471181 | 0.65806 | 0.18055 | 0.49321 | 0.76353 | 0.86047 | 0.2901775508 | 0.89088 | 0.5168938262 | 0.190678 | 4.094879867 | 0 |
| 17 | 43115765 | T | G | K | T | BRCA1 | ENST00000471181 | 0.66755 | 0.42664 | 0.60169 | 0.93818 | 0.88212 | 0.4904118071 | 0.91647 | 0.801881423 | 0.272431 | 6.509408713 | 0 |
| 17 | 43115766 | T | C | K | E | BRCA1 | ENST00000471181 | 0.7497 | 0.46329 | 0.58558 | 0.92098 | 0.85638 | 0.5001174807 | 0.92135 | 0.9691259861 | 0.291601 | 4.154158126 | 0 |
| 17 | 43115766 | T | G | K | Q | BRCA1 | ENST00000471181 | 0.71634 | 0.43482 | 0.53141 | 0.91475 | 0.82948 | 0.3794828194 | 0.93914 | 0.8982334733 | 0.225741 | 3.640953638 | 0 |
| 17 | 43115767 | G | C | I | M | BRCA1 | ENST00000471181 | 0.57294 | 0.4179 | 0.6921 | 0.87963 | 0.82387 | 0.3124065343 | 0.92358 | 0.7511663437 | 0.247529 | 4.607673315 | 0 |
| 17 | 43115768 | A | C | I | S | BRCA1 | ENST00000471181 | 0.73237 | 0.48806 | 0.74703 | 0.92985 | 0.89101 | 0.7497670423 | 0.98058 | 0.8165606837 | 0.397946 | 6.820846946 | 0 |
| 17 | 43115768 | A | G | I | T | BRCA1 | ENST00000471181 | 0.66652 | 0.4499 | 0.82928 | 0.92667 | 0.8859 | 0.5056798081 | 0.98645 | 0.917252779 | 0.402284 | 6.820846946 | 0 |
| 17 | 43115769 | T | A | I | F | BRCA1 | ENST00000471181 | 0.70615 | 0.4604 | 0.73785 | 0.88899 | 0.86098 | 0.4693027117 | 0.97147 | 0.9588987231 | 0.305059 | 6.820846946 | 0 |
| 17 | 43115769 | T | C | I | V | BRCA1 | ENST00000471181 | 0.62271 | 0.22786 | 0.64685 | 0.82486 | 0.8517 | 0.2132191162 | 0.90992 | 0.6727738203 | 0.170225 | 4.136163339 | 0 |
| 17 | 43115769 | T | G | I | L | BRCA1 | ENST00000471181 | 0.69083 | 0.10057 | 0.73706 | 0.81488 | 0.78782 | 0.3370510949 | 0.81758 | 0.6035323145 | 0.173862 | 4.91037982 | 0 |
| 17 | 43115770 | C | G | L | F | BRCA1 | ENST00000471181 | 0.55428 | 0.46025 | 0.71262 | 0.94415 | 0.83725 | 0.5124326552 | 0.96987 | 0.8739020228 | 0.241299 | 6.968068048 | 0 |
| 17 | 43115771 | A | C | L | W | BRCA1 | ENST00000471181 | 0.72404 | 0.53697 | 0.7876 | 0.96141 | 0.89873 | 0.7131489115 | 0.98716 | 0.8993965983 | 0.397025 | 6.968068048 | 0 |
| 17 | 43115771 | A | G | L | S | BRCA1 | ENST00000471181 | 0.72204 | 0.49279 | 0.73387 | 0.93729 | 0.85792 | 0.7644941033 | 0.97939 | 0.9664606452 | 0.400602 | 6.968068048 | 0 |
| 17 | 43115772 | A | C | L | V | BRCA1 | ENST00000471181 | 0.3001 | 0.43548 | 0.67065 | 0.83921 | 0.79089 | 0.4030458168 | 0.89885 | 0.500477612 | 0.146832 | 4.201009451 | 0 |
| 17 | 43115772 | A | T | L | M | BRCA1 | ENST00000471181 | 0.42962 | 0.44339 | 0.69962 | 0.91342 | 0.78658 | 0.3823199004 | 0.90371 | 0.8931191564 | 0.17919 | 4.75038762 | 0 |
| 17 | 43115773 | C | A | E | D | BRCA1 | ENST00000471181 | 0.22689 | 0.41678 | 0.69506 | 0.84295 | 0.80358 | 0.3190424515 | 0.85722 | 0.8476831317 | 0.194353 | 3.796560679 | 0 |
| 17 | 43115773 | C | G | E | D | BRCA1 | ENST00000471181 | 0.23132 | 0.41678 | 0.69506 | 0.81039 | 0.80417 | 0.3190424515 | 0.85722 | 0.8476831317 | 0.194356 | 3.796560679 | 0 |
| 17 | 43115774 | T | A | E | V | BRCA1 | ENST00000471181 | 0.69412 | 0.42952 | 0.66774 | 0.9512 | 0.93123 | 0.6477514118 | 0.95421 | 0.9908639789 | 0.423235 | 6.839725181 | 0 |
| 17 | 43115774 | T | C | E | G | BRCA1 | ENST00000471181 | 0.71281 | 0.46935 | 0.64344 | 0.90413 | 0.90711 | 0.56710811 | 0.94651 | 0.878349483 | 0.372045 | 6.839725181 | 0 |
| 17 | 43115774 | T | G | E | A | BRCA1 | ENST00000471181 | 0.6647 | 0.42045 | 0.68608 | 0.92958 | 0.92356 | 0.5300152365 | 0.95146 | 0.9761231542 | 0.394289 | 6.839725181 | 0 |
| 17 | 43115775 | C | G | E | Q | BRCA1 | ENST00000471181 | 0.61082 | 0.43776 | 0.68209 | 0.85755 | 0.84052 | 0.4923782282 | 0.95191 | 0.8076155873 | 0.261016 | 6.839725181 | 0 |
| 17 | 43115775 | C | T | E | K | BRCA1 | ENST00000471181 | 0.67557 | 0.458 | 0.77525 | 0.91494 | 0.89918 | 0.5788801411 | 0.95847 | 0.9837852716 | 0.405755 | 6.839725181 | 0 |
| 17 | 43115777 | A | C | L | R | BRCA1 | ENST00000471181 | 0.70792 | 0.51312 | 0.72763 | 0.96402 | 0.91271 | 0.7459675175 | 0.98522 | 0.9855260849 | 0.448034 | 6.568056894 | 0 |
| 17 | 43115777 | A | G | L | P | BRCA1 | ENST00000471181 | 0.71193 | 0.53095 | 0.75574 | 0.97229 | 0.94765 | 0.8492252265 | 0.9937 | 0.9972961545 | 0.479893 | 6.568056894 | 1 |
| 17 | 43115777 | A | T | L | Q | BRCA1 | ENST00000471181 | 0.6982 | 0.45545 | 0.64406 | 0.95886 | 0.90581 | 0.6069820852 | 0.9854 | 0.9811867476 | 0.444554 | 6.568056894 | 0 |
| 17 | 43115778 | G | C | L | V | BRCA1 | ENST00000471181 | 0.50004 | 0.4203 | 0.57692 | 0.954 | 0.89376 | 0.5196740297 | 0.96779 | 0.9762944579 | 0.300208 | 6.568056894 | 0 |
| 17 | 43115778 | G | T | L | M | BRCA1 | ENST00000471181 | 0.52672 | 0.44339 | 0.61988 | 0.93672 | 0.83834 | 0.3405058619 | 0.92688 | 0.9630194306 | 0.270194 | 6.118842278 | 0 |
| 17 | 43115779 | A | C | C | W | BRCA1 | ENST00000471181 | 0.70423 | 0.50075 | 0.72164 | | 0.9833 | 0.9464819455 | 0.99794 | 0.9981954694 | 0.514496 | 6.581416725 | 1 |
| 17 | 43124017 | C | A | C | F | BRCA1 | ENST00000471181 | 0.86699 | 0.50717 | 0.66234 | 0.99438 | 0.99296 | 0.9419153465 | 0.99918 | 0.9794338346 | 0.539304 | 6.581416725 | 1 |
| 17 | 43124017 | C | G | C | S | BRCA1 | ENST00000471181 | 0.82996 | 0.46773 | 0.64644 | 0.99584 | 0.99072 | 0.9244110122 | 0.99943 | 0.9959779978 | 0.500108 | 6.581416725 | 1 |
| 17 | 43124017 | C | T | C | Y | BRCA1 | ENST00000471181 | 0.84726 | 0.53471 | 0.78553 | 0.99465 | 0.9917 | 0.9410546706 | 0.99946 | 0.9915230274 | 0.538525 | 6.581416725 | 1 |
| 17 | 43124018 | A | C | C | G | BRCA1 | ENST00000471181 | 0.88868 | 0.47595 | 0.60837 | 0.99421 | 0.9917 | 0.9241559175 | 0.99928 | 0.9963377714 | 0.539545 | 6.581416725 | 1 |
| 17 | 43124018 | A | G | C | R | BRCA1 | ENST00000471181 | 0.85793 | 0.54367 | 0.7604 | 0.99327 | 0.98938 | 0.9534401463 | 0.99922 | 0.9960098267 | 0.542692 | 6.581416725 | 1 |
| 17 | 43124018 | A | T | C | S | BRCA1 | ENST00000471181 | 0.82412 | 0.46773 | 0.64644 | 0.9953 | 0.99039 | 0.9244110122 | 0.99924 | 0.9959779978 | 0.504263 | 6.581416725 | 1 |
| 17 | 43124019 | G | C | I | M | BRCA1 | ENST00000471181 | 0.54793 | 0.44069 | 0.58836 | 0.9771 | 0.85326 | 0.6817038284 | 0.91323 | 0.9718171358 | 0.267181 | 6.564776998 | 0 |
| 17 | 43124020 | A | G | I | T | BRCA1 | ENST00000471181 | 0.68171 | 0.4499 | 0.75681 | 0.98519 | 0.93824 | 0.7839156411 | 0.97882 | 0.9748427868 | 0.440272 | 6.564776998 | 1 |
| 17 | 43124020 | A | T | I | N | BRCA1 | ENST00000471181 | 0.73981 | 0.53373 | 0.73323 | 0.98813 | 0.93824 | 0.8913645781 | 0.98719 | 0.9875292778 | 0.369667 | 6.564776998 | 1 |
| 17 | 43124021 | T | C | I | V | BRCA1 | ENST00000471181 | 0.6086 | 0.1422 | 0.52762 | 0.8278 | 0.84535 | 0.4876467073 | 0.85145 | 0.7157424092 | 0.184308 | 5.035622419 | 0 |
| 17 | 43124023 | G | A | P | L | BRCA1 | ENST00000471181 | 0.73797 | 0.43568 | 0.72989 | 0.98627 | 0.95207 | 0.7512045455 | 0.9541 | 0.9701777697 | 0.410837 | 6.580438659 | 1 |
| 17 | 43124023 | G | C | P | R | BRCA1 | ENST00000471181 | 0.72148 | 0.45753 | 0.7234 | 0.98589 | 0.95972 | 0.7915342731 | 0.96594 | 0.9710247517 | 0.449876 | 6.580438659 | 0 |
| 17 | 43124024 | G | A | P | S | BRCA1 | ENST00000471181 | 0.72869 | 0.44373 | 0.63055 | 0.97601 | 0.91442 | 0.7086079679 | 0.95759 | 0.9461869001 | 0.272743 | 6.580438659 | 0 |
| 17 | 43124024 | G | C | P | A | BRCA1 | ENST00000471181 | 0.67915 | 0.41888 | 0.63476 | 0.98189 | 0.92884 | 0.6026484371 | 0.96663 | 0.932682097 | 0.288595 | 5.257121995 | 0 |
| 17 | 43124024 | G | T | P | T | BRCA1 | ENST00000471181 | 0.70238 | 0.46258 | 0.67314 | 0.98356 | 0.94243 | 0.7211569195 | 0.97372 | 0.9576455355 | 0.426941 | 6.580438659 | 0 |
| 17 | 43124025 | A | C | C | W | BRCA1 | ENST00000471181 | 0.83736 | 0.49831 | 0.7755 | 0.99695 | 0.96737 | 0.9139679961 | 0.99594 | 0.9928746223 | 0.466371 | 6.581416725 | 1 |
| 17 | 43124026 | C | G | C | S | BRCA1 | ENST00000471181 | 0.65218 | 0.43882 | 0.72794 | 0.99561 | 0.99105 | 0.8828225569 | 0.999 | 0.9959779978 | 0.551764 | 6.581416725 | 1 |
| 17 | 43124026 | C | T | C | Y | BRCA1 | ENST00000471181 | 0.67144 | 0.53471 | 0.82963 | 0.99461 | 0.98938 | 0.9054356559 | 0.99938 | 0.9978685379 | 0.551752 | 6.581416725 | 1 |
| 17 | 43124027 | A | C | C | G | BRCA1 | ENST00000471181 | 0.78638 | 0.47028 | 0.69452 | 0.99533 | 0.98972 | 0.9036868309 | 0.99931 | 0.9963377714 | 0.556289 | 6.581416725 | 1 |
| 17 | 43124027 | A | G | C | R | BRCA1 | ENST00000471181 | 0.80037 | 0.53493 | 0.80996 | 0.99364 | 0.98728 | 0.9187767449 | 0.99934 | 0.9969949722 | 0.556285 | 6.581416725 | 1 |
| 17 | 43124027 | A | T | C | S | BRCA1 | ENST00000471181 | 0.74101 | 0.43882 | 0.72794 | 0.99532 | 0.98869 | 0.8828225569 | 0.99923 | 0.9959779978 | 0.556286 | 6.581416725 | 1 |
| 17 | 43124029 | T | A | E | V | BRCA1 | ENST00000471181 | 0.77275 | 0.42952 | 0.55265 | 0.97378 | 0.93162 | 0.5054219317 | 0.88155 | 0.8529191017 | 0.25281 | 6.581416391 | 0 |
| 17 | 43124029 | T | G | E | A | BRCA1 | ENST00000471181 | 0.74507 | 0.42045 | 0.58472 | 0.97333 | 0.9328 | 0.4553266127 | 0.88322 | 0.8440711498 | 0.206526 | 6.581416391 | 0 |
| 17 | 43124030 | C | G | E | Q | BRCA1 | ENST00000471181 | 0.67128 | 0.43776 | 0.57107 | 0.93514 | 0.88448 | 0.3607249767 | 0.85087 | 0.7581776155 | 0.205576 | 6.581416391 | 0 |
| 17 | 43124030 | C | T | E | K | BRCA1 | ENST00000471181 | 0.73909 | 0.458 | 0.70483 | 0.96468 | 0.90842 | 0.5047664855 | 0.85958 | 0.8732894659 | 0.218747 | 6.581416391 | 0 |
| 17 | 43124031 | T | A | L | F | BRCA1 | ENST00000471181 | 0.58197 | 0.45497 | 0.62356 | 0.96977 | 0.86902 | 0.5877128689 | 0.92726 | 0.7228525246 | 0.28334 | 6.581416081 | 0 |
| 17 | 43124031 | T | G | L | F | BRCA1 | ENST00000471181 | 0.58931 | 0.45497 | 0.62356 | 0.96995 | 0.86852 | 0.5877128689 | 0.92726 | 0.7166642811 | 0.283341 | 6.581416081 | 0 |
| 17 | 43124032 | A | G | L | S | BRCA1 | ENST00000471181 | 0.80913 | 0.50212 | 0.65409 | 0.98808 | 0.9328 | 0.8396595354 | 0.96777 | 0.9466410875 | 0.414856 | 6.581416081 | 1 |
| 17 | 43124033 | A | C | L | V | BRCA1 | ENST00000471181 | 0.61865 | 0.3607 | 0.55913 | 0.95309 | 0.89376 | 0.5182106094 | 0.94692 | 0.9172233343 | 0.243362 | 6.581416081 | 0 |
| 17 | 43124033 | A | T | L | I | BRCA1 | ENST00000471181 | 0.62468 | 0.28302 | 0.64257 | 0.95474 | 0.89101 | 0.3945090837 | 0.93837 | 0.9172233343 | 0.157082 | 6.581416081 | 0 |
| 17 | 43124034 | G | C | I | M | BRCA1 | ENST00000471181 | 0.69428 | 0.43385 | 0.68982 | 0.93341 | 0.86098 | 0.4041477474 | 0.96033 | 0.7708864202 | 0.242336 | 6.284957288 | 0 |
| 17 | 43124035 | A | C | I | S | BRCA1 | ENST00000471181 | 0.73581 | 0.37861 | 0.7498 | 0.9635 | 0.87829 | 0.559455078 | 0.99158 | 0.8031124784 | 0.288625 | 5.67132036 | 0 |
| 17 | 43124035 | A | G | I | T | BRCA1 | ENST00000471181 | 0.51705 | 0.37996 | 0.83405 | 0.95898 | 0.87781 | 0.3905816535 | 0.98134 | 0.7710319161 | 0.280075 | 4.732929608 | 0 |
| 17 | 43124035 | A | T | I | N | BRCA1 | ENST00000471181 | 0.44832 | 0.26827 | 0.81828 | 0.9119 | 0.86503 | 0.5218187719 | 0.97353 | 0.863640368 | 0.256756 | 3.736381597 | 0 |
| 17 | 43124036 | T | A | I | F | BRCA1 | ENST00000471181 | 0.61566 | 0.50275 | 0.74072 | 0.91955 | 0.86403 | 0.4797526684 | 0.9617 | 0.7693007588 | 0.238217 | 6.284957288 | 0 |
| 17 | 43124036 | T | C | I | V | BRCA1 | ENST00000471181 | 0.29874 | 0.2825 | 0.64547 | 0.84605 | 0.84959 | 0.3356229009 | 0.88477 | 0.2277928785 | 0.0989214 | 4.922335202 | 0 |
| 17 | 43124036 | T | G | I | L | BRCA1 | ENST00000471181 | 0.32918 | 0.27677 | 0.73588 | 0.8826 | 0.8367 | 0.4496371511 | 0.87968 | 0.8741663694 | 0.154752 | 4.619396013 | 0 |
| 17 | 43124037 | T | A | K | N | BRCA1 | ENST00000471181 | 0.68147 | 0.46012 | 0.78278 | 0.9718 | 0.88212 | 0.5623898644 | 0.88116 | 0.8732894659 | 0.157429 | 6.573338777 | 0 |
| 17 | 43124037 | T | G | K | N | BRCA1 | ENST00000471181 | 0.68971 | 0.46012 | 0.78278 | 0.97387 | 0.88212 | 0.5623898644 | 0.88116 | 0.7694233284 | 0.157431 | 6.573338777 | 0 |
| 17 | 43124038 | T | A | K | I | BRCA1 | ENST00000471181 | 0.73045 | 0.54328 | 0.72298 | 0.98723 | 0.94949 | 0.6488513456 | 0.95384 | 0.9634467959 | 0.270806 | 6.573338777 | 0 |
| 17 | 43124038 | T | C | K | R | BRCA1 | ENST00000471181 | 0.68115 | 0.37662 | 0.68975 | 0.96492 | 0.89422 | 0.3618869215 | 0.9157 | 0.7842309861 | 0.231816 | 6.573338777 | 0 |
| 17 | 43124038 | T | G | K | T | BRCA1 | ENST00000471181 | 0.68531 | 0.46306 | 0.7665 | 0.98244 | 0.94129 | 0.5837594252 | 0.91754 | 0.8440711498 | 0.245565 | 6.573338777 | 0 |
| 17 | 43124039 | T | C | K | E | BRCA1 | ENST00000471181 | 0.75049 | 0.43401 | 0.75207 | 0.96067 | 0.88963 | 0.4686687483 | 0.88784 | 0.8560277224 | 0.247673 | 5.488695778 | 0 |
| 17 | 43124040 | C | A | Q | H | BRCA1 | ENST00000471181 | 0.56825 | 0.16736 | 0.60598 | 0.96798 | 0.90624 | 0.4760934979 | 0.94662 | 0.7586685795 | 0.276544 | 5.441578437 | 0 |
| 17 | 43124040 | C | G | Q | H | BRCA1 | ENST00000471181 | 0.57568 | 0.16736 | 0.60598 | 0.96856 | 0.90624 | 0.4760934979 | 0.94662 | 0.6915295538 | 0.276546 | 5.441578437 | 0 |
| 17 | 43124041 | T | A | Q | L | BRCA1 | ENST00000471181 | 0.69724 | 0.20274 | 0.53766 | 0.98236 | 0.94205 | 0.5962928017 | 0.94281 | 0.744521016 | 0.26668 | 5.269120373 | 0 |
| 17 | 43124041 | T | C | Q | R | BRCA1 | ENST00000471181 | 0.64976 | 0.37786 | 0.60424 | 0.9656 | 0.90929 | 0.4676634766 | 0.89575 | 0.7341243373 | 0.257292 | 5.466622434 | 0 |
| 17 | 43124041 | T | G | Q | P | BRCA1 | ENST00000471181 | 0.67247 | 0.45622 | 0.57285 | 0.98119 | 0.94469 | 0.8432424418 | 0.93893 | 0.7831301122 | 0.395029 | 6.373229155 | 1 |
| 17 | 43124042 | G | C | Q | E | BRCA1 | ENST00000471181 | 0.61179 | 0.31937 | 0.55912 | 0.94141 | 0.88684 | 0.4647921883 | 0.89929 | 0.7112174219 | 0.195434 | 5.399587511 | 0 |
| 17 | 43124042 | G | T | Q | K | BRCA1 | ENST00000471181 | 0.64074 | 0.38027 | 0.61516 | 0.9656 | 0.91862 | 0.5712810089 | 0.89701 | 0.7310216622 | 0.25799 | 4.434242405 | 0 |
| 17 | 43124043 | C | A | M | I | BRCA1 | ENST00000471181 | 0.57062 | 0.36413 | 0.67558 | 0.93225 | 0.88307 | 0.4828103613 | 0.84566 | 0.7250443697 | 0.20091 | 6.581331489 | 0 |
| 17 | 43124043 | C | G | M | I | BRCA1 | ENST00000471181 | 0.57808 | 0.36413 | 0.67558 | 0.9288 | 0.88307 | 0.4828103613 | 0.84566 | 0.7006139354 | 0.200913 | 6.581331489 | 0 |
| 17 | 43124043 | C | T | M | I | BRCA1 | ENST00000471181 | 0.5829 | 0.36413 | 0.67558 | 0.93043 | 0.88307 | 0.4828103613 | 0.84566 | 0.7250443697 | 0.200901 | 6.581331489 | 0 |
| 17 | 43124044 | A | C | M | R | BRCA1 | ENST00000471181 | 0.69188 | 0.4964 | 0.73912 | 0.96819 | 0.93747 | 0.878192446 | 0.94172 | 0.7668827997 | 0.385513 | 6.581331489 | 1 |
| 17 | 43124044 | A | G | M | T | BRCA1 | ENST00000471181 | 0.62681 | 0.42131 | 0.79385 | 0.97049 | 0.92356 | 0.6801701345 | 0.95244 | 0.7578753529 | 0.380399 | 6.581331489 | 1 |
| 17 | 43124044 | A | T | M | K | BRCA1 | ENST00000471181 | 0.67621 | 0.47389 | 0.76011 | 0.96624 | 0.9252 | 0.8553564628 | 0.9324 | 0.7607134022 | 0.382709 | 6.581331489 | 1 |
| 17 | 43124045 | T | A | M | L | BRCA1 | ENST00000471181 | 0.64579 | 0.27614 | 0.67825 | 0.91702 | 0.84482 | 0.4525504686 | 0.83081 | 0.6576683341 | 0.185389 | 6.581331489 | 0 |
| 17 | 43124045 | T | G | M | L | BRCA1 | ENST00000471181 | 0.65382 | 0.27614 | 0.67825 | 0.91796 | 0.84482 | 0.4525504686 | 0.83081 | 0.6595312228 | 0.185392 | 6.581331489 | 0 |
| 17 | 43124047 | G | A | A | V | BRCA1 | ENST00000471181 | 0.71434 | 0.13767 | 0.60758 | 0.93146 | 0.84106 | 0.4524135156 | 0.91685 | 0.7152889528 | 0.217245 | 4.725822301 | 0 |
| 17 | 43124047 | G | C | A | G | BRCA1 | ENST00000471181 | 0.57808 | 0.13875 | 0.56336 | 0.89183 | 0.8124 | 0.4590836263 | 0.88357 | 0.8006814122 | 0.161222 | 4.123595105 | 0 |

| | | | | | | | | | | | | | | | | |
|---|---|---|---|---|---|---|---|---|---|---|---|---|---|---|---|---|
| 17 | 43124047 | G | T | A | D | BRCA1 | ENST00000471181 | 0.72427 | 0.28933 | 0.7039 | 0.94929 | 0.84906 | 0.7516457419 | 0.90874 | 0.7249199723 | 0.217611 | 6.288051153 | 0 |
| 17 | 43124048 | C | A | A | S | BRCA1 | ENST00000471181 | 0.6215 | 0.12771 | 0.5475 | 0.93851 | 0.83394 | 0.4130944305 | 0.90827 | 0.923887074 | 0.16683 | 5.273853797 | 0 |
| 17 | 43124048 | C | G | A | P | BRCA1 | ENST00000471181 | 0.67231 | 0.389 | 0.66353 | 0.94982 | 0.86403 | 0.8250985944 | 0.93821 | 0.8883513212 | 0.314886 | 6.288051153 | 1 |
| 17 | 43124048 | C | T | A | T | BRCA1 | ENST00000471181 | 0.66272 | 0.19785 | 0.59102 | 0.9343 | 0.8278 | 0.4036324353 | 0.94022 | 0.7711977533 | 0.232022 | 4.528840436 | 0 |
| 17 | 43124049 | A | C | N | K | BRCA1 | ENST00000471181 | 0.56062 | 0.17371 | 0.48366 | 0.90384 | 0.82216 | 0.3082298759 | 0.81168 | 0.867667973 | 0.0699466 | 6.078709012 | 0 |
| 17 | 43124049 | A | T | N | K | BRCA1 | ENST00000471181 | 0.55477 | 0.17371 | 0.48366 | 0.90403 | 0.82216 | 0.3082298759 | 0.81168 | 0.867667973 | 0.069948 | 6.078709012 | 0 |
| 17 | 43124050 | T | A | N | I | BRCA1 | ENST00000471181 | 0.46799 | 0.25766 | 0.43521 | 0.93541 | 0.85843 | 0.4371987121 | 0.88563 | 0.5915572643 | 0.157185 | 4.166438621 | 0 |
| 17 | 43124050 | T | C | N | S | BRCA1 | ENST00000471181 | 0.26456 | 0.07396 | 0.35864 | 0.73075 | 0.71705 | 0.1782922061 | 0.7638 | 0.4543565214 | 0.0526607 | 3.308108319 | 0 |
| 17 | 43124050 | T | G | N | T | BRCA1 | ENST00000471181 | 0.55532 | 0.14036 | 0.42318 | 0.89762 | 0.81761 | 0.2551053287 | 0.83795 | 0.5870239735 | 0.0933428 | 4.694615661 | 0 |
| 17 | 43124051 | T | A | N | Y | BRCA1 | ENST00000471181 | 0.60449 | 0.37169 | 0.44734 | 0.95005 | 0.89422 | 0.4286504527 | 0.90642 | 0.8092691512 | 0.158741 | 6.078709012 | 0 |
| 17 | 43124051 | T | C | N | D | BRCA1 | ENST00000471181 | 0.46562 | 0.17194 | 0.46249 | 0.89056 | 0.76946 | 0.3419101356 | 0.81251 | 0.7759510279 | 0.0517306 | 6.078709012 | 0 |
| 17 | 43124051 | T | G | N | H | BRCA1 | ENST00000471181 | 0.28872 | 0.34611 | 0.42156 | 0.88875 | 0.74164 | 0.2942503342 | 0.82312 | 0.6759373546 | 0.0535163 | 3.036245645 | 0 |
| 17 | 43124052 | A | C | I | M | BRCA1 | ENST00000471181 | 0.38685 | 0.0935 | 0.37033 | 0.86175 | 0.81006 | 0.5084765537 | 0.8026 | 0.3752331734 | 0.128338 | 2.908933041 | 0 |
| 17 | 43124053 | A | C | I | S | BRCA1 | ENST00000471181 | 0.58517 | 0.15408 | 0.44954 | 0.95085 | 0.85274 | 0.8149671755 | 0.96775 | 0.615202393 | 0.376377 | 2.908933041 | 1 |
| 17 | 43124053 | A | G | I | T | BRCA1 | ENST00000471181 | 0.51711 | 0.12439 | 0.5538 | 0.94223 | 0.84052 | 0.5933033442 | 0.95732 | 0.544923633 | 0.291582 | 2.908933041 | 1 |
| 17 | 43124053 | A | T | I | N | BRCA1 | ENST00000471181 | 0.72612 | 0.2272 | 0.53177 | 0.96743 | 0.89603 | 0.820083404 | 0.95872 | 0.6202192629 | 0.220747 | 2.908933041 | 1 |
| 17 | 43124054 | T | C | I | V | BRCA1 | ENST00000471181 | 0.33425 | 0.07644 | 0.33298 | 0.86234 | 0.74376 | 0.4107763247 | 0.6392 | 0.6398671865 | 0.065787 | 2.908933041 | 0 |
| 17 | 43124054 | T | G | I | L | BRCA1 | ENST00000471181 | 0.23844 | 0.09021 | 0.40102 | 0.73478 | 0.75074 | 0.5543708258 | 0.44866 | 0.04177673534 | 0.043442 | -2.914575802 | 0 |
| 17 | 43124056 | A | C | V | G | BRCA1 | ENST00000471181 | 0.75551 | 0.40353 | 0.49503 | 0.95723 | 0.88212 | 0.7494375565 | 0.98043 | 0.8223862755 | 0.354317 | 6.220129361 | 1 |
| 17 | 43124056 | A | G | V | A | BRCA1 | ENST00000471181 | 0.72164 | 0.37734 | 0.61695 | 0.93998 | 0.86603 | 0.6490787054 | 0.97784 | 0.7657382622 | 0.207886 | 6.220129361 | 0 |
| 17 | 43124057 | C | A | V | F | BRCA1 | ENST00000471181 | 0.56172 | 0.46158 | 0.57475 | 0.96314 | 0.84747 | 0.8165219666 | 0.98783 | 0.7945968509 | 0.256363 | 6.220129361 | 1 |
| 17 | 43124057 | C | G | V | L | BRCA1 | ENST00000471181 | 0.5316 | 0.20186 | 0.59909 | 0.9223 | 0.84375 | 0.675917887 | 0.90598 | 0.6013406868 | 0.194025 | 5.589943712 | 0 |
| 17 | 43124057 | C | T | V | I | BRCA1 | ENST00000471181 | 0.48685 | 0.0997 | 0.43217 | 0.85432 | 0.83339 | 0.5078013736 | 0.84327 | 0.5887858222 | 0.140283 | 4.491955493 | 0 |
| 17 | 43124058 | A | C | N | K | BRCA1 | ENST00000471181 | 0.60375 | 0.20459 | 0.60639 | 0.92832 | 0.81646 | 0.3003543177 | 0.70575 | 0.5696079731 | 0.0976419 | 5.226761756 | 0 |
| 17 | 43124058 | A | T | N | K | BRCA1 | ENST00000471181 | 0.59762 | 0.20459 | 0.60639 | 0.92782 | 0.81646 | 0.3003543177 | 0.70575 | 0.5696079731 | 0.097642 | 5.226761756 | 0 |
| 17 | 43124059 | T | A | N | I | BRCA1 | ENST00000471181 | 0.60025 | 0.4093 | 0.5484 | 0.97209 | 0.88069 | 0.4421023572 | 0.86612 | 0.6227391461 | 0.154891 | 6.218643279 | 0 |
| 17 | 43124059 | T | C | N | S | BRCA1 | ENST00000471181 | 0.36142 | 0.09661 | 0.47603 | 0.86966 | 0.74021 | 0.2076338463 | 0.62528 | 0.6098393202 | 0.0687394 | 3.394977267 | 0 |
| 17 | 43124059 | T | G | N | T | BRCA1 | ENST00000471181 | 0.55121 | 0.24515 | 0.52946 | 0.94733 | 0.83615 | 0.2741182524 | 0.75532 | 0.5402295169 | 0.1154 | 5.643203997 | 0 |
| 17 | 43124060 | T | A | N | Y | BRCA1 | ENST00000471181 | 0.6001 | 0.44044 | 0.57232 | 0.97138 | 0.90842 | 0.4171607691 | 0.87551 | 0.9570525885 | 0.191794 | 6.218643279 | 0 |
| 17 | 43124060 | T | C | N | D | BRCA1 | ENST00000471181 | 0.41588 | 0.10907 | 0.58615 | 0.88835 | 0.80772 | 0.3450038122 | 0.68478 | 0.7381125093 | 0.0921872 | 4.470953295 | 0 |
| 17 | 43124060 | T | G | N | H | BRCA1 | ENST00000471181 | 0.40115 | 0.39658 | 0.53966 | 0.94085 | 0.81761 | 0.2913079737 | 0.81182 | 0.658666356 | 0.104814 | 4.345098884 | 0 |
| 17 | 43124061 | T | A | Q | H | BRCA1 | ENST00000471181 | 0.54351 | 0.43438 | 0.44769 | 0.94011 | 0.84535 | 0.418314465 | 0.80789 | 0.7579387426 | 0.175226 | 4.597216407 | 0 |
| 17 | 43124061 | T | G | Q | H | BRCA1 | ENST00000471181 | 0.55052 | 0.43438 | 0.44769 | 0.94052 | 0.84535 | 0.418314465 | 0.80789 | 0.7579387426 | 0.175228 | 4.597216407 | 0 |
| 17 | 43124062 | T | A | Q | L | BRCA1 | ENST00000471181 | 0.74746 | 0.35705 | 0.38889 | 0.94819 | 0.87829 | 0.5765865478 | 0.82583 | 0.795156002 | 0.186298 | 6.32869647 | 0 |
| 17 | 43124062 | T | C | Q | R | BRCA1 | ENST00000471181 | 0.7015 | 0.44561 | 0.44485 | 0.92738 | 0.83116 | 0.3534799253 | 0.72263 | 0.782828927 | 0.188831 | 4.307842087 | 0 |
| 17 | 43124062 | T | G | Q | P | BRCA1 | ENST00000471181 | 0.72244 | 0.47259 | 0.41766 | 0.95295 | 0.88117 | 0.8238495478 | 0.8093 | 0.7699840677 | 0.319494 | 6.32869647 | 1 |
| 17 | 43124063 | G | C | Q | E | BRCA1 | ENST00000471181 | 0.62294 | 0.42572 | 0.40807 | 0.90923 | 0.82556 | 0.4164218429 | 0.77185 | 0.641978534 | 0.165505 | 5.380025657 | 0 |
| 17 | 43124063 | G | T | Q | K | BRCA1 | ENST00000471181 | 0.533 | 0.45024 | 0.45525 | 0.8383 | 0.82387 | 0.4761934894 | 0.70985 | 0.7341580987 | 0.1511 | 3.729214074 | 0 |
| 17 | 43124065 | A | C | V | G | BRCA1 | ENST00000471181 | 0.74571 | 0.48292 | 0.55857 | 0.9725 | 0.91652 | 0.8000511897 | 0.98501 | 0.8244869379 | 0.391935 | 6.608442631 | 1 |
| 17 | 43124065 | A | G | V | A | BRCA1 | ENST00000471181 | 0.71353 | 0.40956 | 0.64449 | 0.95627 | 0.90537 | 0.6742784453 | 0.96767 | 0.8418642282 | 0.333327 | 6.608442631 | 0 |
| 17 | 43124065 | A | T | V | E | BRCA1 | ENST00000471181 | 0.75201 | 0.2998 | 0.63675 | 0.97522 | 0.91569 | 0.8746429756 | 0.98525 | 0.8173181985 | 0.382154 | 6.608442631 | 1 |
| 17 | 43124066 | C | T | V | I | BRCA1 | ENST00000471181 | 0.57638 | 0.39352 | 0.5196 | 0.93094 | 0.85064 | 0.4701417808 | 0.91031 | 0.7428113571 | 0.170649 | 4.904544265 | 0 |
| 17 | 43124067 | T | A | E | D | BRCA1 | ENST00000471181 | 0.43629 | 0.4363 | 0.46978 | 0.87259 | 0.84482 | 0.4012382308 | 0.82243 | 0.7555491627 | 0.19143 | 4.093584373 | 0 |
| 17 | 43124067 | T | G | E | D | BRCA1 | ENST00000471181 | 0.44247 | 0.4363 | 0.46978 | 0.86952 | 0.84428 | 0.4012382308 | 0.82243 | 0.7563239447 | 0.191435 | 4.093584373 | 0 |
| 17 | 43124068 | T | C | E | G | BRCA1 | ENST00000471181 | 0.78122 | 0.47791 | 0.43448 | 0.97284 | 0.93044 | 0.4945862345 | 0.88846 | 0.8334181425 | 0.213646 | 6.988443224 | 0 |
| 17 | 43124068 | T | G | E | A | BRCA1 | ENST00000471181 | 0.73365 | 0.46163 | 0.4679 | 0.9672 | 0.90274 | 0.489951042 | 0.86603 | 0.818449627 | 0.127017 | 6.988443224 | 0 |
| 17 | 43124069 | C | G | E | Q | BRCA1 | ENST00000471181 | 0.6218 | 0.45036 | 0.45753 | 0.9361 | 0.88543 | 0.4343039856 | 0.89632 | 0.7822559011 | 0.191484 | 5.884648043 | 0 |
| 17 | 43124070 | T | A | E | D | BRCA1 | ENST00000471181 | 0.59443 | 0.41678 | 0.35426 | 0.89575 | 0.8306 | 0.4148737445 | 0.85595 | 0.7989346981 | 0.184601 | 4.114721994 | 0 |
| 17 | 43124070 | T | G | E | D | BRCA1 | ENST00000471181 | 0.60193 | 0.41678 | 0.35426 | 0.89348 | 0.8306 | 0.4148737445 | 0.85595 | 0.789776206 | 0.184601 | 4.114721994 | 0 |
| 17 | 43124071 | T | A | E | V | BRCA1 | ENST00000471181 | 0.70109 | 0.24377 | 0.34143 | 0.97328 | 0.90972 | 0.5364102984 | 0.87431 | 0.8989931941 | 0.179304 | 6.068026769 | 0 |
| 17 | 43124071 | T | C | E | G | BRCA1 | ENST00000471181 | 0.41576 | 0.46935 | 0.32086 | 0.93102 | 0.84694 | 0.4611472159 | 0.86867 | 0.7447452146 | 0.148706 | 3.554725007 | 0 |
| 17 | 43124071 | T | G | E | A | BRCA1 | ENST00000471181 | 0.66652 | 0.38919 | 0.35555 | 0.94309 | 0.86352 | 0.4559927039 | 0.85324 | 0.7701702118 | 0.13523 | 5.026551978 | 0 |
| 17 | 43124072 | C | G | E | Q | BRCA1 | ENST00000471181 | 0.38885 | 0.43776 | 0.34005 | 0.88469 | 0.83615 | 0.3959555932 | 0.81774 | 0.5166123509 | 0.144164 | 3.799193339 | 0 |
| 17 | 43124072 | C | T | E | K | BRCA1 | ENST00000471181 | 0.56748 | 0.45186 | 0.42766 | 0.92362 | 0.8345 | 0.4532646482 | 0.82631 | 0.8326400518 | 0.158627 | 6.068026769 | 0 |
| 17 | 43124074 | A | C | V | G | BRCA1 | ENST00000471181 | 0.53589 | 0.44955 | 0.1472 | 0.97118 | 0.87198 | 0.547495871 | 0.89408 | 0.616193327 | 0.304497 | 5.764978116 | 0 |
| 17 | 43124074 | A | G | V | A | BRCA1 | ENST00000471181 | 0.36816 | 0.30124 | 0.19693 | 0.95654 | 0.83888 | 0.4263607565 | 0.91422 | 0.5388216027 | 0.178859 | 3.486661082 | 0 |
| 17 | 43124075 | C | A | V | F | BRCA1 | ENST00000471181 | 0.17091 | 0.46365 | 0.17501 | 0.9463 | 0.83283 | 0.6059072285 | 0.87187 | 0.4623709878 | 0.150734 | 5.284373031 | 0 |
| 17 | 43124075 | C | G | V | L | BRCA1 | ENST00000471181 | 0.03351 | 0.20715 | 0.17029 | 0.79954 | 0.80772 | 0.3931364007 | 0.71461 | 0.1484317944 | 0.104827 | 4.264658944 | 0 |
| 17 | 43124075 | C | T | V | I | BRCA1 | ENST00000471181 | 0.01559 | 0.08323 | 0.12193 | 0.60755 | 0.78535 | 0.2488737153 | 0.36704 | 0.02126318694 | 0.014402 | 0.9916267495 | 0 |
| 17 | 43124077 | C | A | R | L | BRCA1 | ENST00000471181 | 0.05589 | 0.14065 | 0.02172 | 0.87458 | 0.84052 | 0.4447589688 | 0.42616 | 0.4348624945 | 0.0955768 | 5.391428874 | 0 |
| 17 | 43124077 | C | G | R | P | BRCA1 | ENST00000471181 | 0.11778 | 0.23274 | 0.02317 | 0.87407 | 0.83779 | 0.6548694667 | 0.44222 | 0.4003046453 | 0.141656 | 4.98548147 | 0 |
| 17 | 43124077 | C | T | R | H | BRCA1 | ENST00000471181 | 0.00761 | 0.11355 | 0.01744 | 0.56824 | 0.80536 | 0.3189401465 | 0.37093 | 0.03728073386 | -0.108434 | 1.095640953 | 0 |
| 17 | 43124078 | G | A | R | C | BRCA1 | ENST00000471181 | 0.46637 | 0.31669 | 0.02789 | 0.91713 | 0.82046 | 0.5069287445 | 0.87173 | 0.2830934525 | 0.0713739 | 5.391428874 | 0 |
| 17 | 43124078 | G | C | R | G | BRCA1 | ENST00000471181 | 0.28383 | 0.13713 | 0.01913 | 0.85459 | 0.80595 | 0.475257844 | 0.77602 | 0.2318910608 | 0.155148 | 4.83064527 | 0 |
| 17 | 43124080 | A | C | L | R | BRCA1 | ENST00000471181 | 0.29477 | 0.14724 | 0.20962 | 0.85583 | 0.78473 | 0.6005190496 | 0.67757 | 0.1043171734 | 0.124862 | 0.9733234191 | 0 |
| 17 | 43124080 | A | G | L | P | BRCA1 | ENST00000471181 | 0.3003 | 0.1668 | 0.20833 | 0.86543 | 0.79395 | 0.6203929934 | 0.63444 | 0.07324178016 | 0.132662 | 1.903174421 | 0 |
| 17 | 43124080 | A | T | L | H | BRCA1 | ENST00000471181 | 0.29783 | 0.16946 | 0.20652 | 0.88598 | 0.79998 | 0.513042051 | 0.65523 | 0.1119397136 | 0.125789 | 1.903174421 | 0 |
| 17 | 43124081 | G | A | L | F | BRCA1 | ENST00000471181 | 0.16022 | 0.10624 | 0.19349 | 0.83001 | 0.75074 | 0.4005016595 | 0.82289 | 0.0953318684 | 0.127044 | 1.903174421 | 0 |
| 17 | 43124081 | G | C | L | V | BRCA1 | ENST00000471181 | 0.0567 | 0.09155 | 0.17921 | 0.60656 | 0.75143 | 0.3766482586 | 0.48842 | 0.02684499957 | 0.0965654 | -3.249257289 | 0 |
| 17 | 43124081 | G | T | L | I | BRCA1 | ENST00000471181 | 0.08902 | 0.09904 | 0.16333 | 0.66026 | 0.71312 | 0.2776919873 | 0.46594 | 0.04023227617 | 0.0389058 | -0.3095554663 | 0 |
| 17 | 43124083 | G | A | A | V | BRCA1 | ENST00000471181 | 0.05212 | 0.09297 | 0.09308 | 0.74137 | 0.80713 | 0.3791519841 | 0.74074 | 0.0179609958 | 0.107418 | 1.315992152 | 0 |
| 17 | 43124083 | G | C | A | G | BRCA1 | ENST00000471181 | 0.06342 | 0.09262 | 0.07731 | 0.84964 | 0.82216 | 0.3769899455 | 0.78356 | 0.08267901348 | 0.117647 | 2.83712531 | 0 |
| 17 | 43124083 | G | T | A | D | BRCA1 | ENST00000471181 | 0.2097 | 0.34736 | 0.13189 | 0.94908 | 0.84482 | 0.5466729277 | 0.80919 | 0.4554655326 | 0.126412 | 3.677171374 | 0 |
| 17 | 43124084 | C | A | A | S | BRCA1 | ENST00000471181 | 0.08155 | 0.14643 | 0.06807 | 0.91958 | 0.79758 | 0.3437063742 | 0.7872 | 0.258529272 | 0.084498 | 3.057060581 | 0 |
| 17 | 43124084 | C | T | A | T | BRCA1 | ENST00000471181 | 0.1564 | 0.15346 | 0.081 | 0.88315 | 0.80118 | 0.3425090103 | 0.79861 | 0.1232540488 | 0.107082 | 2.165060408 | 0 |
| 17 | 43124086 | G | A | S | F | BRCA1 | ENST00000471181 | 0.44478 | 0.38833 | 0.2359 | 0.96551 | 0.85945 | 0.4594807279 | 0.8764 | 0.8231654167 | 0.202498 | 5.769938979 | 0 |
| 17 | 43124086 | G | C | S | C | BRCA1 | ENST00000471181 | 0.42933 | 0.4238 | 0.20969 | 0.95856 | 0.80536 | 0.4254792589 | 0.77989 | 0.5484864116 | 0.129049 | 2.118511227 | 0 |
| 17 | 43124086 | G | T | S | Y | BRCA1 | ENST00000471181 | 0.4104 | 0.46885 | 0.25756 | 0.9638 | 0.86098 | 0.4416963871 | 0.87645 | 0.7865451574 | 0.189758 | 5.769938979 | 0 |
| 17 | 43124087 | A | C | S | A | BRCA1 | ENST00000471181 | 0.28685 | 0.17688 | 0.20271 | 0.93436 | 0.77909 | 0.2435718119 | 0.78902 | 0.1440020344 | 0.058386 | 4.271899964 | 0 |
| 17 | 43124087 | A | G | S | P | BRCA1 | ENST00000471181 | 0.16153 | 0.13252 | 0.22276 | 0.86701 | 0.80178 | 0.4869787708 | 0.72235 | 0.06361240894 | 0.158519 | 2.612763813 | 0 |
| 17 | 43124087 | A | T | S | T | BRCA1 | ENST00000471181 | 0.2897 | 0.28907 | 0.21124 | 0.95875 | 0.80653 | 0.3589461308 | 0.80096 | 0.5529299974 | 0.114134 | 4.80885971 | 0 |
| 17 | 43124088 | T | A | L | F | BRCA1 | ENST00000471181 | 0.15199 | 0.16552 | 0.17749 | 0.90134 | 0.79212 | 0.3927161501 | 0.8631 | 0.07084684819 | 0.0948034 | 2.678374302 | 0 |
| 17 | 43124088 | T | G | L | F | BRCA1 | ENST00000471181 | 0.15549 | 0.16552 | 0.17749 | 0.90134 | 0.79212 | 0.3927161501 | 0.8631 | 0.07084684819 | 0.0948061 | 2.678374302 | 0 |
| 17 | 43124089 | A | G | L | S | BRCA1 | ENST00000471181 | 0.50289 | 0.48956 | 0.19912 | 0.97075 | 0.81472 | 0.5193058749 | 0.93763 | 0.8561949134 | 0.128733 | 3.584039064 | 0 |
| 17 | 43124090 | A | C | L | V | BRCA1 | ENST00000471181 | 0.26354 | 0.1389 | 0.14905 | 0.76959 | 0.78597 | 0.4434125472 | 0.815 | 0.1052278429 | 0.13476 | 4.302541343 | 0 |
| 17 | 43124090 | A | T | L | I | BRCA1 | ENST00000471181 | 0.48672 | 0.30004 | 0.19553 | 0.88841 | 0.77205 | 0.3352999702 | 0.76976 | 0.4455613809 | 0.0855617 | 6.146839612 | 0 |
| 17 | 43124091 | A | C | D | E | BRCA1 | ENST00000471181 | 0.55114 | 0.43397 | 0.43984 | 0.92904 | 0.80948 | 0.5025076227 | 0.84891 | 0.7796400648 | 0.176142 | 4.582855261 | 0 |
| 17 | 43124091 | A | T | D | E | BRCA1 | ENST00000471181 | 0.54535 | 0.43397 | 0.43984 | 0.92904 | 0.80948 | 0.5025076227 | 0.84891 | 0.7820523788 | 0.176137 | 4.582855261 | 0 |
| 17 | 43124092 | T | G | D | A | BRCA1 | ENST00000471181 | 0.677 | 0.43605 | 0.41518 | 0.96066 | 0.85378 | 0.7206029108 | 0.87521 | 0.8964475989 | 0.209208 | 4.46974985 | 0 |
| 17 | 43124093 | C | A | D | Y | BRCA1 | ENST00000471181 | 0.60801 | 0.48572 | 0.47949 | 0.95844 | 0.89558 | 0.7825119733 | 0.89659 | 0.9657840133 | 0.23938 | 6.482502851 | 0 |
| 17 | 43124093 | C | G | D | H | BRCA1 | ENST00000471181 | 0.60229 | 0.47901 | 0.49145 | 0.95217 | 0.8517 | 0.6557136036 | 0.88575 | 0.9614516497 | 0.217801 | 6.482502851 | 1 |
| 17 | 43124093 | C | T | D | N | BRCA1 | ENST00000471181 | 0.60911 | 0.44726 | 0.46809 | 0.93698 | 0.80653 | 0.5585264007 | 0.87804 | 0.7423120141 | 0.023897 | 5.401408085 | 0 |

| Variants with functional readout data from deep mutational scan experiments of TP53 | | | | | | | | | | | | | | | | | | |
|---|---|---|---|---|---|---|---|---|---|---|---|---|---|---|---|---|---|---|
| chrom | pos | ref | alt | ref_aa | alt_aa | genename | transcript_id | CADD_raw_rankscore | MPC_rankscore | PrimateAI_rankscore | M-CAP_rankscore | REVEL_rankscore | gMVP_rankscore | MVP_rankscore | ClinPred | BayesDel | EVmutation | target |
| 17 | 7673744 | T | A | K | N | TP53 | ENST00000269305 | 0.22822 | 0.10127 | 0.27026 | 0.95095 | 0.75349 | 0.4974426638 | 0.93397 | 0.8700563908 | 0.126405 | 4.407020432 | 0 |
| 17 | 7673745 | T | A | K | I | TP53 | ENST00000269305 | 0.56117 | 0.33184 | 0.24599 | 0.97957 | 0.94765 | 0.5321627467 | 0.99158 | 0.97812289 | 0.384261 | 5.947037608 | 0 |
| 17 | 7673745 | T | C | K | R | TP53 | ENST00000269305 | 0.40664 | 0.06219 | 0.21045 | 0.96604 | 0.76881 | 0.3405155154 | 0.90262 | 0.5357489586 | 0.172479 | 3.510391614 | 0 |
| 17 | 7673745 | T | G | K | T | TP53 | ENST00000269305 | 0.40887 | 0.2794 | 0.25581 | 0.96065 | 0.83283 | 0.5045632251 | 0.9199 | 0.8576375842 | 0.23434 | 5.080210572 | 0 |
| 17 | 7673746 | T | C | K | E | TP53 | ENST00000269305 | 0.41576 | 0.16648 | 0.25128 | 0.97311 | 0.79212 | 0.4849134324 | 0.91859 | 0.8941676617 | 0.221091 | 5.327015969 | 0 |
| 17 | 7673746 | T | G | K | Q | TP53 | ENST00000269305 | 0.38158 | 0.12641 | 0.22012 | 0.90679 | 0.78098 | 0.3919950023 | 0.8875 | 0.2872547209 | 0.160599 | 4.378140134 | 0 |
| 17 | 7673747 | C | A | K | N | TP53 | ENST00000269305 | 0.3379 | 0.40106 | 0.30342 | 0.96448 | 0.82613 | 0.6761153636 | 0.95576 | 0.978140831 | 0.19388 | 3.7950656 | 0 |
| 17 | 7673747 | C | G | K | N | TP53 | ENST00000269305 | 0.34364 | 0.40106 | 0.30342 | 0.96335 | 0.82613 | 0.6761153636 | 0.95615 | 0.9786029458 | 0.193873 | 3.7950656 | 0 |
| 17 | 7673748 | T | A | K | M | TP53 | ENST00000269305 | 0.81763 | 0.42384 | 0.22741 | 0.97756 | 0.93397 | 0.7025081158 | 0.95587 | 0.9970832467 | 0.306282 | 7.000524199 | 0 |
| 17 | 7673748 | T | G | K | T | TP53 | ENST00000269305 | 0.78265 | 0.40346 | 0.299 | 0.97684 | 0.93201 | 0.6779373403 | 0.94784 | 0.9908474684 | 0.293972 | 6.281862679 | 0 |
| 17 | 7673749 | T | C | K | E | TP53 | ENST00000269305 | 0.54116 | 0.30966 | 0.27707 | 0.93335 | 0.8199 | 0.6901077123 | 0.84652 | 0.7758240104 | 0.22313 | 5.286875547 | 0 |
| 17 | 7673749 | T | G | K | Q | TP53 | ENST00000269305 | 0.29308 | 0.10555 | 0.25028 | 0.94187 | 0.84906 | 0.5878242628 | 0.93204 | 0.9765283465 | 0.250922 | 4.867577472 | 0 |
| 17 | 7673751 | C | G | R | P | TP53 | ENST00000269305 | 0.2441 | 0.39216 | 0.00881 | 0.94697 | 0.85064 | 0.5830560558 | 0.938 | 0.9427092671 | 0.230939 | 4.285262547 | 0 |
| 17 | 7673751 | C | T | R | H | TP53 | ENST00000269305 | 0.11443 | 0.08455 | 0.00592 | 0.69735 | 0.83505 | 0.2301681611 | 0.97725 | 0.01741066346 | 0.0347643 | 1.106219382 | 0 |
| 17 | 7673752 | G | A | R | C | TP53 | ENST00000269305 | 0.8418 | 0.38958 | 0.01149 | 0.94495 | 0.89739 | 0.3737970898 | 0.94702 | 0.8905134966 | 0.150931 | 5.214138143 | 0 |
| 17 | 7673752 | G | C | R | G | TP53 | ENST00000269305 | 0.45992 | 0.288 | 0.00657 | 0.94091 | 0.82556 | 0.3003742306 | 0.93734 | 0.9541941285 | 0.241864 | 4.157947532 | 0 |
| 17 | 7673754 | A | C | L | R | TP53 | ENST00000269305 | 0.29069 | 0.14065 | 0.30988 | 0.95216 | 0.73661 | 0.5337280865 | 0.84329 | 0.06614343745 | 0.111817 | 2.673451657 | 0 |
| 17 | 7673754 | A | G | L | P | TP53 | ENST00000269305 | 0.51021 | 0.26711 | 0.29185 | 0.95205 | 0.79697 | 0.7023459343 | 0.89582 | 0.556392312 | 0.155922 | 3.431823096 | 0 |
| 17 | 7673754 | A | T | L | H | TP53 | ENST00000269305 | 0.29188 | 0.22627 | 0.31075 | 0.95328 | 0.71627 | 0.4258572875 | 0.84121 | 0.2454970926 | 0.120422 | 1.440616914 | 0 |
| 17 | 7673755 | G | A | L | F | TP53 | ENST00000269305 | 0.11735 | 0.07378 | 0.24497 | 0.53276 | 0.66922 | 0.2674843732 | 0.66111 | 0.02327830156 | -0.00242514 | -2.640309762 | 0 |
| 17 | 7673755 | G | C | L | V | TP53 | ENST00000269305 | 0.16156 | 0.07301 | 0.24847 | 0.83807 | 0.71627 | 0.3195617159 | 0.84692 | 0.03456749945 | 0.093757 | 2.70187261 | 0 |
| 17 | 7673755 | G | T | L | I | TP53 | ENST00000269305 | 0.16033 | 0.08464 | 0.27136 | 0.85375 | 0.67386 | 0.2429299853 | 0.71983 | 0.0740763247 | 0.0471554 | 2.893915558 | 0 |
| 17 | 7673756 | A | T | N | K | TP53 | ENST00000269305 | 0.50302 | 0.40936 | 0.40701 | 0.97765 | 0.88354 | 0.6533560713 | 0.97331 | 0.9904262424 | 0.217171 | 4.608662806 | 0 |
| 17 | 7673757 | T | A | N | I | TP53 | ENST00000269305 | 0.63359 | 0.42762 | 0.34636 | 0.97926 | 0.9014 | 0.7251122222 | 0.96628 | 0.990213573 | 0.237229 | 7.201840719 | 0 |
| 17 | 7673757 | T | C | N | S | TP53 | ENST00000269305 | 0.28876 | 0.11875 | 0.28956 | 0.95122 | 0.77335 | 0.4536839486 | 0.84983 | 0.5171658397 | 0.0928534 | 4.388860528 | 0 |
| 17 | 7673757 | T | G | N | T | TP53 | ENST00000269305 | 0.58027 | 0.37051 | 0.3332 | 0.96187 | 0.86098 | 0.5720126915 | 0.92424 | 0.9755263925 | 0.187065 | 5.504409008 | 0 |
| 17 | 7673760 | T | A | E | V | TP53 | ENST00000269305 | 0.89101 | 0.3981 | 0.20726 | 0.97535 | 0.86653 | 0.5509981067 | 0.95951 | 0.9718739986 | 0.244149 | 7.02946755 | 0 |
| 17 | 7673760 | T | C | E | G | TP53 | ENST00000269305 | 0.75973 | 0.40007 | 0.21562 | 0.95806 | 0.85741 | 0.5179557489 | 0.90338 | 0.9155527353 | 0.223868 | 5.142811071 | 0 |
| 17 | 7673760 | T | G | E | A | TP53 | ENST00000269305 | 0.70911 | 0.36221 | 0.22922 | 0.96321 | 0.82273 | 0.5071649728 | 0.89981 | 0.8596503139 | 0.211845 | 4.747650876 | 0 |
| 17 | 7673761 | C | G | E | Q | TP53 | ENST00000269305 | 0.65617 | 0.33948 | 0.22597 | 0.95298 | 0.85586 | 0.4249006826 | 0.92494 | 0.7873991728 | 0.204575 | 5.085051184 | 0 |
| 17 | 7673761 | C | T | E | K | TP53 | ENST00000269305 | 0.55985 | 0.12932 | 0.29323 | 0.95765 | 0.8345 | 0.4563433282 | 0.87938 | 0.4434299773 | 0.198359 | 4.491590255 | 0 |
| 17 | 7673763 | T | C | E | G | TP53 | ENST00000269305 | 0.94774 | 0.41701 | 0.27927 | 0.97781 | 0.97735 | 0.892192658 | 0.9726 | 0.997256577 | 0.427816 | 6.121041571 | 1 |
| 17 | 7673763 | T | G | E | A | TP53 | ENST00000269305 | 0.93573 | 0.40365 | 0.31863 | 0.97722 | 0.97735 | 0.8912348412 | 0.98938 | 0.9974761605 | 0.53692 | 6.102505088 | 1 |
| 17 | 7673764 | C | G | E | Q | TP53 | ENST00000269305 | 0.71914 | 0.39117 | 0.30929 | 0.95541 | 0.97438 | 0.8572991151 | 0.95886 | 0.9969265764 | 0.351212 | 6.179523889 | 1 |
| 17 | 7673764 | C | T | E | K | TP53 | ENST00000269305 | 0.78598 | 0.40891 | 0.41541 | 0.96675 | 0.99039 | 0.9371994971 | 0.98741 | 0.9952715039 | 0.566471 | 5.110920898 | 1 |
| 17 | 7673765 | C | A | E | D | TP53 | ENST00000269305 | 0.43762 | 0.35186 | 0.24381 | 0.92778 | 0.80058 | 0.7065233776 | 0.88304 | 0.620823741 | 0.111635 | 3.89213725 | 0 |
| 17 | 7673765 | C | G | E | D | TP53 | ENST00000269305 | 0.44405 | 0.35186 | 0.24381 | 0.92753 | 0.80058 | 0.7065233776 | 0.88298 | 0.527033329 | 0.111634 | 3.89213725 | 0 |
| 17 | 7673766 | T | A | E | V | TP53 | ENST00000269305 | 0.87403 | 0.41274 | 0.22107 | 0.9853 | 0.98585 | 0.8230809726 | 0.97079 | 0.998267889 | 0.481554 | 7.789208427 | 1 |
| 17 | 7673766 | T | G | E | A | TP53 | ENST00000269305 | 0.84965 | 0.40251 | 0.23852 | 0.97821 | 0.97624 | 0.8300324806 | 0.96179 | 0.9979998308 | 0.345004 | 6.817510539 | 0 |
| 17 | 7673767 | C | G | E | Q | TP53 | ENST00000269305 | 0.61015 | 0.39001 | 0.22922 | 0.94838 | 0.96627 | 0.7864248337 | 0.95342 | 0.9753563404 | 0.414826 | 6.521399541 | 0 |
| 17 | 7673767 | C | T | E | K | TP53 | ENST00000269305 | 0.67987 | 0.41008 | 0.3381 | 0.96318 | 0.97624 | 0.8809610849 | 0.96722 | 0.9959677823 | 0.474993 | 6.279950097 | 1 |
| 17 | 7673769 | G | A | T | I | TP53 | ENST00000269305 | 0.82717 | 0.25262 | 0.4063 | 0.9675 | 0.89285 | 0.4470010997 | 0.91946 | 0.9775036573 | 0.29903 | 4.760669965 | 0 |
| 17 | 7673769 | G | C | T | R | TP53 | ENST00000269305 | 0.84876 | 0.40471 | 0.34106 | 0.96943 | 0.90581 | 0.634386072 | 0.95531 | 0.9905267613 | 0.297801 | 5.596454046 | 0 |
| 17 | 7673769 | G | T | T | K | TP53 | ENST00000269305 | 0.86169 | 0.36107 | 0.35693 | 0.96743 | 0.89963 | 0.6022413928 | 0.9483 | 0.9818831682 | 0.283178 | 5.196703232 | 0 |
| 17 | 7673770 | T | A | T | S | TP53 | ENST00000269305 | 0.45425 | 0.21512 | 0.24899 | 0.94711 | 0.78286 | 0.2498591843 | 0.9092 | 0.6973876953 | 0.178939 | 3.779918844 | 0 |
| 17 | 7673770 | T | C | T | A | TP53 | ENST00000269305 | 0.61447 | 0.2269 | 0.23529 | 0.92833 | 0.76024 | 0.3615558383 | 0.89638 | 0.8183570504 | 0.120101 | 5.425784312 | 0 |
| 17 | 7673770 | T | G | T | P | TP53 | ENST00000269305 | 0.65838 | 0.42477 | 0.2556 | 0.96867 | 0.82668 | 0.733242641 | 0.97559 | 0.9898770452 | 0.293877 | 8.201038801 | 1 |
| 17 | 7673772 | C | A | R | L | TP53 | ENST00000269305 | 0.51514 | 0.10693 | 0.31241 | 0.95677 | 0.88637 | 0.4608860482 | 0.95729 | 0.9849463701 | 0.281861 | 7.214202298 | 0 |
| 17 | 7673772 | C | G | R | P | TP53 | ENST00000269305 | 0.86389 | 0.44402 | 0.33572 | 0.975 | 0.93709 | 0.7721982672 | 0.96051 | 0.9922806025 | 0.35506 | 8.031268711 | 1 |
| 17 | 7673772 | C | T | R | H | TP53 | ENST00000269305 | 0.83391 | 0.41306 | 0.30405 | 0.97553 | 0.94913 | 0.2880269696 | 0.97105 | 0.6829593182 | 0.234006 | 6.998628364 | 0 |
| 17 | 7673773 | G | A | R | C | TP53 | ENST00000269305 | 0.42389 | 0.43557 | 0.39057 | 0.97848 | 0.911 | 0.4523532398 | 0.98529 | 0.309286728 | 0.201378 | 5.357912795 | 0 |
| 17 | 7673773 | G | C | R | G | TP53 | ENST00000269305 | 0.46457 | 0.11793 | 0.28948 | 0.95907 | 0.89963 | 0.4396685196 | 0.95594 | 0.9872627854 | 0.339958 | 7.110242224 | 0 |
| 17 | 7673773 | G | T | R | S | TP53 | ENST00000269305 | 0.36426 | 0.20173 | 0.29376 | 0.95791 | 0.8873 | 0.4239926481 | 0.94559 | 0.7482590079 | 0.284729 | 4.889703596 | 0 |
| 17 | 7673775 | C | A | R | L | TP53 | ENST00000269305 | 0.89167 | 0.41566 | 0.50449 | 0.96683 | 0.97178 | 0.8202670132 | 0.94266 | 0.9989080429 | 0.416469 | 5.325227392 | 0 |
| 17 | 7673775 | C | G | R | P | TP53 | ENST00000269305 | 0.90645 | 0.45036 | 0.5228 | 0.96334 | 0.97661 | 0.9163334742 | 0.96982 | 0.9979202151 | 0.55318 | 8.505666317 | 1 |
| 17 | 7673775 | C | T | R | Q | TP53 | ENST00000269305 | 0.89718 | 0.39651 | 0.39442 | 0.95487 | 0.96773 | 0.7779293173 | 0.97018 | 0.9949803948 | 0.49809 | 7.499249124 | 0 |
| 17 | 7673776 | G | A | R | W | TP53 | ENST00000269305 | 0.92599 | 0.40396 | 0.58617 | 0.97303 | 0.97104 | 0.8391295158 | 0.98002 | 0.9974831939 | 0.542691 | 6.608720965 | 1 |
| 17 | 7673776 | G | C | R | G | TP53 | ENST00000269305 | 0.84567 | 0.16573 | 0.46789 | 0.97549 | 0.95899 | 0.848248144 | 0.98853 | 0.9972184896 | 0.501805 | 6.431325375 | 1 |
| 17 | 7673777 | G | C | D | E | TP53 | ENST00000269305 | 0.71369 | 0.38046 | 0.54204 | 0.98728 | 0.97587 | 0.8587520143 | 0.99711 | 0.996170342 | 0.545911 | 4.581932314 | 1 |
| 17 | 7673777 | G | T | D | E | TP53 | ENST00000269305 | 0.7072 | 0.38046 | 0.54204 | 0.98765 | 0.97587 | 0.8587520143 | 0.99712 | 0.996170342 | 0.54591 | 4.581932314 | 1 |
| 17 | 7673778 | T | C | D | G | TP53 | ENST00000269305 | 0.87543 | 0.43684 | 0.5175 | 0.9793 | 0.99744 | 0.9198621503 | 0.99639 | 0.9977051616 | 0.583801 | 6.60525829 | 1 |
| 17 | 7673779 | C | A | D | Y | TP53 | ENST00000269305 | 0.84935 | 0.42828 | 0.5759 | 0.95948 | 0.99296 | 0.9435691789 | 0.99309 | 0.9994951487 | 0.583114 | 6.60525829 | 1 |
| 17 | 7673779 | C | G | D | H | TP53 | ENST00000269305 | 0.67772 | 0.42219 | 0.59749 | 0.95553 | 0.99414 | 0.9184710279 | 0.99556 | 0.9976904392 | 0.565173 | 3.584086431 | 1 |
| 17 | 7673779 | C | T | D | N | TP53 | ENST00000269305 | 0.84786 | 0.3954 | 0.57064 | 0.95263 | 0.98834 | 0.8911851829 | 0.96246 | 0.9982839227 | 0.162963 | 6.60525829 | 1 |
| 17 | 7673780 | T | A | R | S | TP53 | ENST00000269305 | 0.66858 | 0.28235 | 0.61261 | 0.99387 | 0.96444 | 0.9131487297 | 0.99716 | 0.9948951602 | 0.493751 | 5.773911464 | 1 |
| 17 | 7673780 | T | G | R | S | TP53 | ENST00000269305 | 0.67676 | 0.28235 | 0.61261 | 0.99386 | 0.96444 | 0.9131487297 | 0.99717 | 0.9948951602 | 0.493753 | 5.773911464 | 1 |
| 17 | 7673781 | C | A | R | I | TP53 | ENST00000269305 | 0.89249 | 0.4777 | 0.63086 | 0.96269 | 0.98799 | 0.8548397736 | 0.98408 | 0.9993172884 | 0.581044 | 7.950445767 | 1 |
| 17 | 7673781 | C | G | R | T | TP53 | ENST00000269305 | 0.85654 | 0.43536 | 0.65454 | 0.96574 | 0.98728 | 0.9134043614 | 0.98971 | 0.9993113279 | 0.580771 | 7.950445767 | 1 |
| 17 | 7673781 | C | T | R | K | TP53 | ENST00000269305 | 0.84385 | 0.3374 | 0.66808 | 0.95916 | 0.98549 | 0.8650137588 | 0.99455 | 0.9944867492 | 0.57776 | 6.756093346 | 1 |
| 17 | 7673784 | C | G | G | A | TP53 | ENST00000269305 | 0.80321 | 0.40777 | 0.56085 | 0.96464 | 0.97512 | 0.6160006118 | 0.9593 | 0.9982738495 | 0.377886 | 6.835976193 | 0 |
| 17 | 7673785 | C | A | G | W | TP53 | ENST00000269305 | 0.82366 | 0.40207 | 0.65218 | 0.95141 | 0.98367 | 0.7750144851 | 0.96188 | 0.9997983575 | 0.465304 | 6.262337564 | 0 |
| 17 | 7673785 | C | G | G | R | TP53 | ENST00000269305 | 0.76776 | 0.42384 | 0.67802 | 0.9734 | 0.97104 | 0.7644145756 | 0.93341 | 0.9996900558 | 0.400384 | 6.760508882 | 0 |
| 17 | 7673787 | G | A | P | L | TP53 | ENST00000269305 | 0.89729 | 0.40282 | 0.75325 | 0.98077 | 0.98728 | 0.833514831 | 0.97315 | 0.9999285936 | 0.571886 | 6.717694022 | 1 |
| 17 | 7673787 | G | C | P | R | TP53 | ENST00000269305 | 0.88517 | 0.40761 | 0.744 | 0.97904 | 0.99105 | 0.8864413369 | 0.98644 | 0.9997503161 | 0.58868 | 7.749665806 | 1 |
| 17 | 7673787 | G | T | P | H | TP53 | ENST00000269305 | 0.88348 | 0.41969 | 0.7303 | 0.9781 | 0.99072 | 0.8233362463 | 0.98099 | 0.999797523 | 0.58868 | 7.749665806 | 1 |
| 17 | 7673788 | G | A | P | S | TP53 | ENST00000269305 | 0.89656 | 0.40145 | 0.67447 | 0.95782 | 0.99471 | 0.8260852202 | 0.9876 | 0.9991906285 | 0.583515 | 5.663284145 | 1 |
| 17 | 7673788 | G | C | P | A | TP53 | ENST00000269305 | 0.85938 | 0.3963 | 0.65567 | 0.95376 | 0.99326 | 0.7517920178 | 0.98319 | 0.9985657334 | 0.560575 | 7.749665806 | 1 |
| 17 | 7673788 | G | T | P | T | TP53 | ENST00000269305 | 0.88145 | 0.4149 | 0.71041 | 0.96064 | 0.99202 | 0.8150315551 | 0.99065 | 0.999353826 | 0.579476 | 7.749665806 | 1 |
| 17 | 7673789 | A | C | C | W | TP53 | ENST00000269305 | 0.83982 | 0.21337 | 0.70092 | 0.98661 | 0.96334 | 0.918294621 | 0.98875 | 0.9807364941 | 0.482617 | 5.883476351 | 0 |
| 17 | 7673790 | C | G | C | S | TP53 | ENST00000269305 | 0.8712 | 0.39159 | 0.62583 | 0.93574 | 0.98585 | 0.878003893 | 0.98948 | 0.9995818734 | 0.563797 | 4.896403105 | 0 |
| 17 | 7673790 | C | T | C | Y | TP53 | ENST00000269305 | 0.88898 | 0.45861 | 0.75368 | 0.95737 | 0.98728 | 0.899671211 | 0.99154 | 0.9995818734 | 0.566646 | 7.963171592 | 0 |
| 17 | 7673791 | A | G | C | R | TP53 | ENST00000269305 | 0.93576 | 0.48283 | 0.73747 | 0.98652 | 0.96846 | 0.9266041183 | 0.99505 | 0.9998114705 | 0.585806 | 7.963171592 | 0 |
| 17 | 7673791 | A | T | C | S | TP53 | ENST00000269305 | 0.90504 | 0.39159 | 0.62583 | 0.9845 | 0.9703 | 0.878003893 | 0.99539 | 0.9997389913 | 0.583563 | 4.896403105 | 0 |
| 17 | 7673793 | G | A | A | V | TP53 | ENST00000269305 | 0.87636 | 0.39231 | 0.61813 | 0.96494 | 0.98513 | 0.8324980604 | 0.97474 | 0.9983562827 | 0.53291 | 8.331162455 | 0 |
| 17 | 7673793 | G | C | A | G | TP53 | ENST00000269305 | 0.87536 | 0.38208 | 0.566 | 0.9563 | 0.98367 | 0.7012847797 | 0.96607 | 0.99784863 | 0.449773 | 8.331162455 | 1 |
| 17 | 7673794 | C | G | A | P | TP53 | ENST00000269305 | 0.83345 | 0.29596 | 0.66473 | 0.95329 | 0.99039 | 0.9268610858 | 0.99288 | 0.9969621301 | 0.560163 | 8.331162455 | 1 |
| 17 | 7673795 | A | C | C | W | TP53 | ENST00000269305 | 0.9325 | 0.43982 | 0.76029 | 0.98341 | 0.98071 | 0.9446544466 | 0.99706 | 0.9999047518 | 0.551479 | 7.890788795 | 1 |
| 17 | 7673796 | C | A | C | F | TP53 | ENST00000269305 | 0.79761 | 0.45555 | 0.71112 | 0.97022 | 0.99876 | 0.9350205339 | 0.99498 | 0.9997780919 | 0.583481 | 7.890788795 | 1 |
| 17 | 7673796 | C | T | C | Y | TP53 | ENST00000269305 | 0.78059 | 0.47645 | 0.81092 | 0.9728 | 0.997 | 0.9447487644 | 0.99423 | 0.999764502 | 0.583481 | 7.890788795 | 1 |
| 17 | 7673797 | A | C | C | G | TP53 | ENST00000269305 | 0.88454 | 0.42658 | 0.65849 | 0.98135 | 0.99296 | 0.9338312673 | 0.99548 | 0.9996167421 | 0.586616 | 6.73224249 | 1 |
| 17 | 7673797 | A | G | C | R | TP53 | ENST00000269305 | 0.89515 | 0.48324 | 0.78778 | 0.97715 | 0.99443 | 0.9588389013 | 0.99581 | 0.9998316765 | 0.586616 | 7.890788795 | 1 |
| 17 | 7673799 | A | G | V | A | TP53 | ENST00000269305 | 0.82833 | 0.3563 | 0.2933 | 0.9665 | 0.95316 | 0.7421934829 | 0.95231 | 0.995747149 | 0.381747 | 7.552728224 | 1 |
| 17 | 7673799 | A | T | V | D | TP53 | ENST00000269305 | 0.88029 | 0.46905 | 0.28708 | 0.97925 | 0.96846 | 0.9524099076 | 0.97627 | 0.9955891371 | 0.54013 | 7.552728224 | 1 |
| 17 | 7673800 | C | A | V | F | TP53 | ENST00000269305 | 0.26991 | 0.16892 | 0.25098 | 0.95472 | 0.9248 | 0.8946949376 | 0.98548 | 0.9693212509 | 0.385401 | 5.536307609 | 1 |
| 17 | 7673800 | C | G | V | L | TP53 | ENST00000269305 | 0.18165 | 0.09377 | 0.26385 | 0.95859 | 0.85945 | 0.6645524456 | 0.92814 | 0.822691381 | 0.211738 | 5.297312573 | 1 |
| 17 | 7673800 | C | T | V | I | TP53 | ENST00000269305 | 0.09524 | 0.06084 | 0.13107 | 0.88615 | 0.68995 | 0.2999136859 | 0.70666 | 0.1405421048 | 0.0357549 | 2.088511997 | 0 |
| 17 | 7673802 | C | A | R | L | TP53 | ENST00000269305 | 0.9068 | 0.42733 | 0.65316 | 0.971 | 0.98034 | 0.8869682993 | 0.9969 | 0.9964241022 | 0.5772 | 8.064547545 | 1 |
| 17 | 7673802 | C | G | R | P | TP53 | ENST00000269305 | 0.91897 | 0.45228 | 0.66258 | 0.97086 | 0.9844 | 0.9665304234 | 0.99726 | 0.9976357222 | 0.57871 | 8.064547545 | 1 |
| 17 | 7673802 | C | T | R | H | TP53 | ENST00000269305 | 0.74077 | 0.28988 | 0.64306 | 0.95386 | 0.9608 | 0.7930177715 | 0.99676 | 0.9090914726 | 0.487327 | 4.189200897 | 1 |

| | | | | | | | | | | | | | | | | |
|---|---|---|---|---|---|---|---|---|---|---|---|---|---|---|---|---|
| 17 | 7673803 | G | A | R | C | TP53 | ENST00000269305 | 0.76068 | 0.44258 | 0.71999 | 0.97479 | 0.9729 | 0.8501711313 | 0.99664 | 0.9945346117 | 0.433271 | 6.897058132 | 1 |
| 17 | 7673803 | G | C | R | G | TP53 | ENST00000269305 | 0.68251 | 0.43365 | 0.61098 | 0.9754 | 0.97327 | 0.9192115653 | 0.99825 | 0.9971434474 | 0.554478 | 8.064547545 | 1 |
| 17 | 7673803 | G | T | R | S | TP53 | ENST00000269305 | 0.6514 | 0.41457 | 0.62507 | 0.97266 | 0.97475 | 0.8868314979 | 0.9985 | 0.9982738495 | 0.559385 | 8.064547545 | 1 |
| 17 | 7673805 | A | C | V | G | TP53 | ENST00000269305 | 0.7536 | 0.43706 | 0.33991 | 0.96429 | 0.89648 | 0.9247582075 | 0.9687 | 0.9659199119 | 0.347787 | 5.858937741 | 1 |
| 17 | 7673805 | A | T | V | E | TP53 | ENST00000269305 | 0.75973 | 0.47176 | 0.43895 | 0.98201 | 0.93786 | 0.9403770531 | 0.98838 | 0.9901693463 | 0.486724 | 8.260032787 | 1 |
| 17 | 7673806 | C | A | V | L | TP53 | ENST00000269305 | 0.78574 | 0.36863 | 0.43506 | 0.94824 | 0.96444 | 0.7234170217 | 0.95879 | 0.9835625291 | 0.470042 | 5.262642875 | 1 |
| 17 | 7673806 | C | G | V | L | TP53 | ENST00000269305 | 0.79422 | 0.36863 | 0.43506 | 0.9444 | 0.96444 | 0.7234170217 | 0.95878 | 0.9835625291 | 0.470042 | 5.262642875 | 1 |
| 17 | 7673806 | C | T | V | M | TP53 | ENST00000269305 | 0.82599 | 0.39947 | 0.4732 | 0.94727 | 0.97512 | 0.773994134 | 0.96489 | 0.9396519496 | 0.51421 | 7.145877698 | 1 |
| 17 | 7673807 | C | A | E | D | TP53 | ENST00000269305 | 0.68171 | 0.36761 | 0.55926 | 0.9786 | 0.9517 | 0.6090492965 | 0.97068 | 0.9849085808 | 0.392358 | 5.391983801 | 0 |
| 17 | 7673807 | C | G | E | D | TP53 | ENST00000269305 | 0.69019 | 0.36761 | 0.55926 | 0.97578 | 0.95207 | 0.6090492965 | 0.97065 | 0.9849085808 | 0.392361 | 5.391983801 | 0 |
| 17 | 7673808 | T | G | E | A | TP53 | ENST00000269305 | 0.8755 | 0.40747 | 0.56791 | 0.98155 | 0.99072 | 0.7119952981 | 0.9894 | 0.9990352392 | 0.566356 | 6.800326488 | 0 |
| 17 | 7673809 | C | G | E | Q | TP53 | ENST00000269305 | 0.81536 | 0.39654 | 0.54322 | 0.94427 | 0.98146 | 0.6360052427 | 0.97647 | 0.9965957913 | 0.527745 | 6.842434127 | 0 |
| 17 | 7673809 | C | T | E | K | TP53 | ENST00000269305 | 0.87821 | 0.41549 | 0.67082 | 0.96659 | 0.98549 | 0.8017278682 | 0.99412 | 0.9996361732 | 0.573901 | 6.485433601 | 1 |
| 17 | 7673810 | A | C | F | L | TP53 | ENST00000269305 | 0.75312 | 0.41053 | 0.6957 | 0.98327 | 0.9458 | 0.584984221 | 0.96538 | 0.9942641854 | 0.415089 | 5.357068672 | 1 |
| 17 | 7673810 | A | T | F | L | TP53 | ENST00000269305 | 0.74643 | 0.41053 | 0.6957 | 0.98353 | 0.9458 | 0.584984221 | 0.96536 | 0.9953424931 | 0.415085 | 5.357068672 | 1 |
| 17 | 7673811 | A | C | F | C | TP53 | ENST00000269305 | 0.95063 | 0.4514 | 0.68754 | 0.97554 | 0.9833 | 0.7895189924 | 0.99536 | 0.9994244576 | 0.556308 | 6.577148041 | 1 |
| 17 | 7673811 | A | G | F | S | TP53 | ENST00000269305 | 0.95064 | 0.48645 | 0.65321 | 0.98277 | 0.98904 | 0.8308097729 | 0.9906 | 0.9990692735 | 0.569316 | 9.002588097 | 1 |
| 17 | 7673811 | A | T | F | Y | TP53 | ENST00000269305 | 0.92806 | 0.4144 | 0.67883 | 0.98019 | 0.98585 | 0.5673828697 | 0.96301 | 0.9909986258 | 0.499907 | 9.002588097 | 0 |
| 17 | 7673812 | A | C | F | V | TP53 | ENST00000269305 | 0.90794 | 0.43579 | 0.63804 | 0.97634 | 0.9822 | 0.6383753066 | 0.97074 | 0.9873221085 | 0.525671 | 5.409349919 | 1 |
| 17 | 7673812 | A | G | F | L | TP53 | ENST00000269305 | 0.91534 | 0.41053 | 0.6957 | 0.96969 | 0.98549 | 0.584984221 | 0.95804 | 0.9955747128 | 0.485455 | 5.357068672 | 1 |
| 17 | 7673812 | A | T | F | I | TP53 | ENST00000269305 | 0.90463 | 0.44238 | 0.66069 | 0.97477 | 0.98657 | 0.6235625898 | 0.97856 | 0.9963092208 | 0.518909 | 5.607328123 | 1 |
| 17 | 7673814 | C | A | S | I | TP53 | ENST00000269305 | 0.62143 | 0.4144 | 0.39469 | 0.97097 | 0.93939 | 0.5612354386 | 0.9012 | 0.9794399142 | 0.308912 | 5.82535975 | 0 |
| 17 | 7673814 | C | G | S | T | TP53 | ENST00000269305 | 0.30198 | 0.0852 | 0.36181 | 0.93482 | 0.84535 | 0.3967961358 | 0.85746 | 0.712559104 | 0.173 | 5.499288379 | 0 |
| 17 | 7673814 | C | T | S | N | TP53 | ENST00000269305 | 0.53879 | 0.33463 | 0.40695 | 0.96459 | 0.90624 | 0.5648274064 | 0.88791 | 0.9459285736 | 0.149235 | 6.53075107 | 0 |
| 17 | 7673815 | T | A | S | C | TP53 | ENST00000269305 | 0.52227 | 0.39259 | 0.35858 | 0.94268 | 0.76024 | 0.5237989778 | 0.84529 | 0.934962213 | 0.137425 | 3.889274887 | 0 |
| 17 | 7673815 | T | G | S | R | TP53 | ENST00000269305 | 0.76363 | 0.41103 | 0.42878 | 0.98415 | 0.9328 | 0.7442311553 | 0.9627 | 0.9778914452 | 0.382358 | 6.152639 | 0 |
| 17 | 7673817 | T | C | N | S | TP53 | ENST00000269305 | 0.22026 | 0.06076 | 0.12999 | 0.93685 | 0.65277 | 0.2869323514 | 0.9154 | 0.1052036285 | 0.0557266 | 3.549459685 | 0 |
| 17 | 7673817 | T | G | N | T | TP53 | ENST00000269305 | 0.4622 | 0.24653 | 0.16821 | 0.95103 | 0.70427 | 0.3634384901 | 0.93991 | 0.6044644117 | 0.105795 | 5.746624087 | 0 |
| 17 | 7673818 | T | A | N | Y | TP53 | ENST00000269305 | 0.45579 | 0.42609 | 0.18436 | 0.94033 | 0.82216 | 0.469014236 | 0.94453 | 0.8373394608 | 0.152211 | 5.402022509 | 0 |
| 17 | 7673818 | T | C | N | D | TP53 | ENST00000269305 | 0.19138 | 0.08543 | 0.18358 | 0.94 | 0.52145 | 0.4314085843 | 0.85187 | 0.049855683 | -0.0136439 | 2.153012137 | 0 |
| 17 | 7673818 | T | G | N | H | TP53 | ENST00000269305 | 0.41263 | 0.38636 | 0.1586 | 0.8994 | 0.72016 | 0.3796510189 | 0.92998 | 0.6461204886 | 0.0656171 | 5.777335463 | 0 |
| 17 | 7673820 | C | G | R | P | TP53 | ENST00000269305 | 0.94514 | 0.4515 | 0.57748 | 0.983 | 0.98257 | 0.9653487585 | 0.99063 | 0.9971268773 | 0.576331 | 7.081391087 | 1 |
| 17 | 7673821 | G | C | R | G | TP53 | ENST00000269305 | 0.79856 | 0.4325 | 0.52163 | 0.97747 | 0.98938 | 0.9060900141 | 0.99703 | 0.9968980551 | 0.536921 | 7.081391087 | 1 |
| 17 | 7673823 | C | A | G | V | TP53 | ENST00000269305 | 0.81339 | 0.43077 | 0.62946 | 0.97102 | 0.99744 | 0.8789899266 | 0.99296 | 0.9996961355 | 0.582542 | 7.630309665 | 1 |
| 17 | 7673823 | C | G | G | A | TP53 | ENST00000269305 | 0.7813 | 0.4062 | 0.60737 | 0.9403 | 0.99386 | 0.8274233654 | 0.99512 | 0.9984715581 | 0.570268 | 4.589556259 | 0 |
| 17 | 7673823 | C | T | G | E | TP53 | ENST00000269305 | 0.82225 | 0.43557 | 0.69809 | 0.97265 | 0.99722 | 0.9301020574 | 0.99049 | 0.9994071722 | 0.582542 | 7.630309665 | 1 |
| 17 | 7673824 | C | G | G | R | TP53 | ENST00000269305 | 0.90863 | 0.42384 | 0.71113 | 0.97547 | 0.99471 | 0.9188782925 | 0.99153 | 0.999221921 | 0.582236 | 7.261087845 | 1 |
| 17 | 7673824 | C | T | G | R | TP53 | ENST00000269305 | 0.9119 | 0.42384 | 0.71113 | 0.97547 | 0.99471 | 0.9188782925 | 0.99153 | 0.9994383454 | 0.582236 | 7.261087845 | 1 |
| 17 | 7673826 | A | C | L | R | TP53 | ENST00000269305 | 0.94739 | 0.45515 | 0.53624 | 0.96657 | 0.99525 | 0.9420652442 | 0.99785 | 0.9994831085 | 0.584143 | 7.683747695 | 1 |
| 17 | 7673826 | A | G | L | P | TP53 | ENST00000269305 | 0.94809 | 0.47093 | 0.57878 | 0.97267 | 0.99006 | 0.9510199283 | 0.99823 | 0.9994968176 | 0.574854 | 7.413398225 | 1 |
| 17 | 7673827 | G | C | L | V | TP53 | ENST00000269305 | 0.62886 | 0.40095 | 0.38093 | 0.96041 | 0.96846 | 0.6271518171 | 0.98334 | 0.9812684655 | 0.485215 | 5.383101428 | 0 |
| 17 | 7673827 | G | T | L | M | TP53 | ENST00000269305 | 0.66003 | 0.39573 | 0.41897 | 0.95407 | 0.96153 | 0.5419986806 | 0.98041 | 0.9468710536 | 0.527209 | 6.731011496 | 0 |
| 17 | 7673829 | A | C | L | R | TP53 | ENST00000269305 | 0.63835 | 0.16355 | 0.32411 | 0.9493 | 0.94394 | 0.8362594395 | 0.9752 | 0.9895590544 | 0.394044 | 8.189706984 | 1 |
| 17 | 7673829 | A | G | L | P | TP53 | ENST00000269305 | 0.64345 | 0.46692 | 0.36407 | 0.96036 | 0.94728 | 0.8847001273 | 0.97944 | 0.9923098087 | 0.382644 | 5.354389331 | 0 |
| 17 | 7673829 | A | T | L | Q | TP53 | ENST00000269305 | 0.92904 | 0.3886 | 0.25767 | 0.96771 | 0.94053 | 0.7359551905 | 0.95546 | 0.9922836423 | 0.347698 | 8.189706984 | 0 |
| 17 | 7673830 | G | C | L | V | TP53 | ENST00000269305 | 0.1776 | 0.07671 | 0.20216 | 0.72763 | 0.70179 | 0.3356851185 | 0.71612 | 0.02773053944 | 0.0580477 | 3.816851414 | 0 |
| 17 | 7673830 | G | T | L | I | TP53 | ENST00000269305 | 0.29627 | 0.25167 | 0.23942 | 0.93655 | 0.6993 | 0.2717053055 | 0.79594 | 0.2706365585 | 0.116639 | 4.419581631 | 0 |
| 17 | 7673831 | A | C | N | K | TP53 | ENST00000269305 | 0.12447 | 0.08101 | 0.35863 | 0.77198 | 0.50926 | 0.2508341184 | 0.71267 | 0.02143384915 | -0.0945855 | 3.855714055 | 0 |
| 17 | 7673831 | A | T | N | K | TP53 | ENST00000269305 | 0.12177 | 0.08101 | 0.35863 | 0.77198 | 0.50926 | 0.2508341184 | 0.71616 | 0.02143384915 | -0.0945737 | 3.855714055 | 0 |
| 17 | 7673832 | T | A | N | I | TP53 | ENST00000269305 | 0.37879 | 0.2927 | 0.30167 | 0.96378 | 0.64364 | 0.3079110902 | 0.85621 | 0.1032965445 | 0.0850298 | 5.792520113 | 0 |
| 17 | 7673832 | T | C | N | S | TP53 | ENST00000269305 | 0.27236 | 0.05865 | 0.24323 | 0.95635 | 0.5533 | 0.1470846458 | 0.72508 | 0.05801216514 | -0.0254468 | 4.047154896 | 0 |
| 17 | 7673832 | T | G | N | T | TP53 | ENST00000269305 | 0.32299 | 0.07282 | 0.29267 | 0.96818 | 0.66354 | 0.1802173141 | 0.82513 | 0.06592519171 | 0.0685118 | 5.396183903 | 0 |
| 17 | 7673833 | T | A | N | Y | TP53 | ENST00000269305 | 0.37431 | 0.36037 | 0.32458 | 0.96518 | 0.75074 | 0.3127957175 | 0.88778 | 0.6740375161 | 0.159412 | 6.194209101 | 0 |
| 17 | 7673833 | T | C | N | D | TP53 | ENST00000269305 | 0.30786 | 0.11789 | 0.34013 | 0.95977 | 0.64053 | 0.2323019003 | 0.9086 | 0.09246008333 | -0.212129 | 5.699357467 | 0 |
| 17 | 7673833 | T | G | N | H | TP53 | ENST00000269305 | 0.3165 | 0.32734 | 0.30179 | 0.95976 | 0.64053 | 0.2593211657 | 0.90521 | 0.5210543871 | 0.0809893 | 5.065656118 | 0 |
| 17 | 7673835 | C | A | G | V | TP53 | ENST00000269305 | 0.73845 | 0.42882 | 0.47869 | 0.96794 | 0.9833 | 0.8664125103 | 0.99658 | 0.9993005991 | 0.576512 | 8.528225146 | 1 |
| 17 | 7673835 | C | G | G | A | TP53 | ENST00000269305 | 0.70551 | 0.392 | 0.47018 | 0.94381 | 0.98183 | 0.7702095921 | 0.99684 | 0.9965540171 | 0.568711 | 6.353030993 | 0 |
| 17 | 7673835 | C | T | G | D | TP53 | ENST00000269305 | 0.73869 | 0.43474 | 0.59574 | 0.96178 | 0.98404 | 0.804278856 | 0.99555 | 0.996573329 | 0.569688 | 6.399764934 | 0 |
| 17 | 7673836 | C | A | G | C | TP53 | ENST00000269305 | 0.90867 | 0.40786 | 0.51898 | 0.97212 | 0.95972 | 0.8418871793 | 0.99712 | 0.9996961355 | 0.5775 | 6.75446916 | 0 |
| 17 | 7673837 | A | C | S | R | TP53 | ENST00000269305 | 0.35774 | 0.08995 | 0.2552 | | 0.70994 | 0.2534710512 | 0.86055 | 0.09172847867 | 0.127014 | 5.660020631 | 0 |
| 17 | 7673837 | A | T | S | R | TP53 | ENST00000269305 | 0.35306 | 0.08995 | 0.2552 | 0.95561 | 0.70914 | 0.2534710512 | 0.85804 | 0.09172847867 | 0.126994 | 5.660020631 | 0 |
| 17 | 7674181 | C | G | S | T | TP53 | ENST00000269305 | 0.35306 | | | 0.94401 | 0.78844 | 0.1833591067 | 0.73986 | 0.3413962424 | 0.052415 | 5.089716846 | 0 |
| 17 | 7674181 | C | T | S | N | TP53 | ENST00000269305 | 0.32961 | | | 0.95197 | 0.75074 | 0.1386231852 | 0.69774 | 0.1031617075 | 0.0299224 | 2.355930028 | 0 |
| 17 | 7674182 | T | A | S | C | TP53 | ENST00000269305 | 0.59003 | | | 0.98813 | 0.81414 | 0.2925216838 | 0.72988 | 0.6404613853 | 0.0978505 | 5.273704918 | 0 |
| 17 | 7674182 | T | C | S | G | TP53 | ENST00000269305 | 0.41647 | | | 0.98516 | 0.80358 | 0.163659036 | 0.78852 | 0.177859515 | 0.0751428 | 3.97562965 | 0 |
| 17 | 7674182 | T | G | S | R | TP53 | ENST00000269305 | 0.41724 | 0.08995 | 0.2552 | 0.97037 | 0.75755 | 0.2534710512 | 0.87709 | 0.2017507285 | 0.166403 | 5.660020631 | 0 |
| 17 | 7674184 | G | A | S | F | TP53 | ENST00000269305 | 0.52234 | 0.1205 | 0.30749 | 0.95347 | 0.86251 | 0.3140546241 | 0.97325 | 0.8117013574 | 0.325249 | 5.83262457 | 0 |
| 17 | 7674184 | G | C | S | C | TP53 | ENST00000269305 | 0.68771 | 0.36074 | 0.28492 | 0.94704 | 0.87442 | 0.3403140451 | 0.87043 | 0.9432165623 | 0.280464 | 6.655503447 | 0 |
| 17 | 7674184 | G | T | S | Y | TP53 | ENST00000269305 | 0.65062 | 0.38465 | 0.33373 | 0.92803 | 0.87829 | 0.3270314968 | 0.91673 | 0.8952660561 | 0.325511 | 6.028867145 | 0 |
| 17 | 7674185 | A | C | S | A | TP53 | ENST00000269305 | 0.26968 | 0.06474 | 0.2531 | 0.94656 | 0.84321 | 0.1489351837 | 0.96518 | 0.1140545536 | 0.191458 | 4.094848306 | 0 |
| 17 | 7674185 | A | G | S | P | TP53 | ENST00000269305 | 0.28242 | 0.09363 | 0.2904 | 0.94131 | 0.87394 | 0.4116439412 | 0.97836 | 0.3184995651 | 0.30768 | 3.253530793 | 0 |
| 17 | 7674186 | G | C | D | E | TP53 | ENST00000269305 | 0.30858 | 0.34392 | 0.54015 | 0.98252 | 0.82443 | 0.4193565743 | 0.93076 | 0.8622378111 | 0.211563 | 6.819033431 | 0 |
| 17 | 7674186 | G | T | D | E | TP53 | ENST00000269305 | 0.30437 | 0.34392 | 0.54015 | 0.98277 | 0.82387 | 0.4193565743 | 0.9308 | 0.8622378111 | 0.211561 | 6.819033431 | 0 |
| 17 | 7674187 | T | A | D | V | TP53 | ENST00000269305 | 0.71979 | 0.44782 | 0.49058 | 0.99351 | 0.91186 | 0.6190419447 | 0.97455 | 0.958301723 | 0.398164 | 8.75953363 | 1 |
| 17 | 7674187 | T | C | D | G | TP53 | ENST00000269305 | 0.43297 | 0.23928 | 0.51121 | 0.97963 | 0.87198 | 0.5487095949 | 0.98601 | 0.6645794511 | 0.332605 | 5.258192703 | 0 |
| 17 | 7674188 | C | A | D | Y | TP53 | ENST00000269305 | 0.49106 | 0.42711 | 0.57579 | 0.96506 | 0.90972 | 0.6542522214 | 0.95488 | 0.9581631422 | 0.355197 | 6.534075333 | 1 |
| 17 | 7674188 | C | G | D | H | TP53 | ENST00000269305 | 0.65696 | 0.4206 | 0.60011 | 0.95561 | 0.89963 | 0.554575064 | 0.9164 | 0.9266043901 | 0.331516 | 5.743071394 | 1 |
| 17 | 7674189 | T | A | E | D | TP53 | ENST00000269305 | 0.64586 | 0.29769 | 0.63094 | 0.94699 | 0.97141 | 0.8531540337 | 0.97116 | 0.9686909914 | 0.42527 | 8.490461999 | 1 |
| 17 | 7674189 | T | G | E | D | TP53 | ENST00000269305 | 0.65382 | 0.29769 | 0.63094 | 0.94624 | 0.97141 | 0.8531540337 | 0.97121 | 0.9686909914 | 0.425272 | 8.490461999 | 1 |
| 17 | 7674190 | T | A | E | V | TP53 | ENST00000269305 | 0.91934 | 0.42048 | 0.60827 | 0.97062 | 0.99471 | 0.9053367936 | 0.99273 | 0.9969170094 | 0.567546 | 8.490461999 | 1 |
| 17 | 7674190 | T | G | E | A | TP53 | ENST00000269305 | 0.90318 | 0.40747 | 0.62665 | 0.97656 | 0.99234 | 0.9065148506 | 0.98361 | 0.9986800551 | 0.543458 | 8.490461999 | 1 |
| 17 | 7674191 | C | G | E | Q | TP53 | ENST00000269305 | 0.81826 | 0.38352 | 0.61883 | 0.95865 | 0.98585 | 0.8662519813 | 0.96539 | 0.9827663898 | 0.396954 | 5.857053314 | 1 |
| 17 | 7674191 | C | T | E | K | TP53 | ENST00000269305 | 0.88042 | 0.41392 | 0.74356 | 0.96827 | 0.99414 | 0.9564128168 | 0.99119 | 0.9872169455 | 0.58127 | 7.361719082 | 1 |
| 17 | 7674193 | A | C | L | R | TP53 | ENST00000269305 | 0.91264 | 0.45435 | 0.74874 | 0.98344 | 0.98071 | 0.9814981745 | 0.99876 | 0.9977442026 | 0.574778 | 4.838506184 | 1 |
| 17 | 7674193 | A | G | L | P | TP53 | ENST00000269305 | 0.91503 | 0.47093 | 0.78504 | 0.98883 | 0.98477 | 0.9808664008 | 0.9986 | 0.9991853833 | 0.586651 | 6.907071349 | 1 |
| 17 | 7674193 | A | T | L | Q | TP53 | ENST00000269305 | 0.90685 | 0.40548 | 0.67762 | 0.98293 | 0.97848 | 0.950647105 | 0.99857 | 0.9969896063 | 0.584126 | 6.907071349 | 1 |
| 17 | 7674194 | G | C | L | V | TP53 | ENST00000269305 | 0.47457 | 0.39897 | 0.60624 | 0.96805 | 0.97104 | 0.7209817795 | 0.99172 | 0.9809491038 | 0.532324 | 6.907071349 | 1 |
| 17 | 7674194 | G | T | L | M | TP53 | ENST00000269305 | 0.49757 | 0.39573 | 0.6376 | 0.96609 | 0.9517 | 0.6588032591 | 0.97532 | 0.9693465829 | 0.473525 | 3.872350313 | 0 |
| 17 | 7674196 | G | T | T | K | TP53 | ENST00000269305 | 0.91708 | 0.42533 | 0.73811 | 0.98266 | 0.98585 | 0.9057323391 | 0.99256 | 0.9994267225 | 0.583456 | 7.778298516 | 1 |
| 17 | 7674197 | T | A | T | S | TP53 | ENST00000269305 | 0.68779 | 0.3075 | 0.63179 | 0.98418 | 0.9528 | 0.6559553541 | 0.9452 | 0.9821579456 | 0.266989 | 4.225062349 | 0 |
| 17 | 7674197 | T | C | T | A | TP53 | ENST00000269305 | 0.73757 | 0.37518 | 0.6192 | 0.9869 | 0.97587 | 0.7914067395 | 0.99593 | 0.9953930378 | 0.545239 | 7.778298516 | 0 |
| 17 | 7674197 | T | G | T | P | TP53 | ENST00000269305 | 0.77561 | 0.41468 | 0.61167 | 0.9891 | 0.98904 | 0.9005055843 | 0.99503 | 0.9987612963 | 0.563464 | 6.832626205 | 1 |
| 17 | 7674198 | G | C | I | M | TP53 | ENST00000269305 | 0.66248 | 0.40193 | 0.53508 | 0.97997 | 0.93201 | 0.4873357987 | 0.96011 | 0.9637226462 | 0.337638 | 5.66975482 | 0 |
| 17 | 7674199 | A | C | I | S | TP53 | ENST00000269305 | 0.88016 | 0.45406 | 0.60945 | 0.98886 | 0.98834 | 0.8709784207 | 0.99306 | 0.9981081486 | 0.562206 | 8.735628957 | 1 |
| 17 | 7674199 | A | T | I | N | TP53 | ENST00000269305 | 0.87403 | 0.48645 | 0.70137 | 0.98658 | 0.98294 | 0.8299893911 | 0.96657 | 0.9976697564 | 0.407497 | 8.77807309 | 1 |
| 17 | 7674200 | T | A | I | F | TP53 | ENST00000269305 | 0.62552 | 0.46452 | 0.58855 | 0.98605 | 0.92964 | 0.5992655962 | 0.95555 | 0.9540543556 | 0.261726 | 5.55676853 | 1 |
| 17 | 7674200 | T | C | I | V | TP53 | ENST00000269305 | 0.52234 | 0.36664 | 0.4705 | 0.9738 | 0.85843 | 0.2744640017 | 0.89583 | 0.7773415446 | 0.148776 | 4.610719486 | 0 |
| 17 | 7674200 | T | G | I | L | TP53 | ENST00000269305 | 0.32262 | 0.39521 | 0.58208 | 0.97442 | 0.86302 | 0.4544420854 | 0.85782 | 0.7305125594 | 0.153214 | 4.537339198 | 0 |
| 17 | 7674201 | G | C | I | M | TP53 | ENST00000269305 | 0.43069 | 0.3981 | 0.68399 | 0.97736 | 0.96407 | 0.7389670121 | 0.98856 | 0.9863378406 | 0.500309 | 9.187015788 | 0 |
| 17 | 7674202 | A | C | I | S | TP53 | ENST00000269305 | 0.74293 | 0.45448 | 0.75386 | 0.99144 | 0.9917 | 0.9544261248 | 0.99737 | 0.9994267225 | 0.582326 | 9.187015788 | 1 |
| 17 | 7674202 | A | G | I | T | TP53 | ENST00000269305 | 0.67716 | 0.43178 | 0.84495 | 0.98931 | 0.99202 | 0.8666886195 | 0.99422 | 0.9989325404 | 0.581981 | 9.187015788 | 1 |

| | | | | | | | | | | | | | | | | | | |
|---|---|---|---|---|---|---|---|---|---|---|---|---|---|---|---|---|---|---|
| 17 | 7674202 | A | T | I | N | TP53 | ENST00000269305 | 0.73525 | 0.48683 | 0.82552 | 0.99046 | 0.99443 | 0.9398855098 | 0.98245 | 0.9993522763 | 0.54488 | 9.187015788 | 1 |
| 17 | 7674203 | T | C | I | V | TP53 | ENST00000269305 | 0.65767 | 0.2881 | 0.62338 | 0.96443 | 0.92763 | 0.4331694865 | 0.91311 | 0.8211653233 | 0.240123 | 4.961127174 | 0 |
| 17 | 7674203 | T | G | I | L | TP53 | ENST00000269305 | 0.69756 | 0.13782 | 0.72606 | 0.97274 | 0.96189 | 0.6742168062 | 0.97368 | 0.9491357803 | 0.396977 | 5.822447414 | 0 |
| 17 | 7674206 | T | C | T | A | TP53 | ENST00000269305 | 0.54304 | 0.3973 | 0.38651 | 0.97602 | 0.95572 | 0.5838858158 | 0.92612 | 0.9736407399 | 0.313137 | 4.453281401 | 1 |
| 17 | 7674206 | T | G | T | P | TP53 | ENST00000269305 | 0.71995 | 0.43641 | 0.41407 | 0.9843 | 0.98799 | 0.8444506711 | 0.96891 | 0.9881706238 | 0.486879 | 8.978805049 | 1 |
| 17 | 7674208 | A | C | L | R | TP53 | ENST00000269305 | 0.87834 | 0.45476 | 0.7096 | 0.98048 | 0.98257 | 0.8653067513 | 0.99487 | 0.9989155531 | 0.556 | 6.340663161 | 0 |
| 17 | 7674208 | A | G | L | P | TP53 | ENST00000269305 | 0.88157 | 0.47093 | 0.75325 | 0.98712 | 0.99138 | 0.9270449704 | 0.99373 | 0.9992169142 | 0.569275 | 8.190708757 | 1 |
| 17 | 7674208 | A | T | L | H | TP53 | ENST00000269305 | 0.86226 | 0.43447 | 0.71274 | 0.97826 | 0.97773 | 0.755403692 | 0.96546 | 0.9985752106 | 0.411351 | 7.206206011 | 0 |
| 17 | 7674209 | G | A | L | F | TP53 | ENST00000269305 | 0.65132 | 0.23117 | 0.60314 | 0.96514 | 0.90052 | 0.5458472027 | 0.95537 | 0.9879187942 | 0.309538 | 6.984382322 | 0 |
| 17 | 7674209 | G | C | L | V | TP53 | ENST00000269305 | 0.62589 | 0.09938 | 0.53815 | 0.93367 | 0.84747 | 0.4739327676 | 0.94215 | 0.9807886481 | 0.175956 | 4.511362784 | 0 |
| 17 | 7674209 | G | T | L | I | TP53 | ENST00000269305 | 0.61417 | 0.18646 | 0.61211 | 0.95134 | 0.7727 | 0.3405239708 | 0.88572 | 0.8055405617 | 0.14409 | 4.573154625 | 0 |
| 17 | 7674211 | A | C | I | S | TP53 | ENST00000269305 | 0.82918 | 0.44814 | 0.73771 | 0.99081 | 0.99356 | 0.9414049645 | 0.99735 | 0.9994267225 | 0.582634 | 9.567559616 | 1 |
| 17 | 7674211 | A | G | I | T | TP53 | ENST00000269305 | 0.76475 | 0.42907 | 0.8325 | 0.98831 | 0.98071 | 0.8411205595 | 0.98951 | 0.9972928166 | 0.536891 | 5.968417179 | 1 |
| 17 | 7674211 | A | T | I | N | TP53 | ENST00000269305 | 0.81135 | 0.42795 | 0.81749 | 0.9909 | 0.98257 | 0.926513615 | 0.97734 | 0.998061955 | 0.461402 | 9.567559616 | 1 |
| 17 | 7674212 | T | A | I | F | TP53 | ENST00000269305 | 0.85588 | 0.46487 | 0.72525 | 0.98712 | 0.99443 | 0.8431185852 | 0.99545 | 0.9927582384 | 0.579794 | 8.52787211 | 1 |
| 17 | 7674212 | T | C | I | V | TP53 | ENST00000269305 | 0.77021 | 0.3352 | 0.60494 | 0.97206 | 0.95645 | 0.4261106608 | 0.95765 | 0.9434313774 | 0.413862 | 5.871719187 | 0 |
| 17 | 7674212 | T | G | I | L | TP53 | ENST00000269305 | 0.83859 | 0.38224 | 0.7184 | 0.97165 | 0.9659 | 0.6506125094 | 0.98854 | 0.9830006361 | 0.523267 | 6.088427915 | 1 |
| 17 | 7674214 | G | A | P | L | TP53 | ENST00000269305 | 0.85033 | 0.40282 | 0.90319 | 0.97834 | 0.99443 | 0.7911113922 | 0.994 | 0.9992566705 | 0.585222 | 6.418205667 | 1 |
| 17 | 7674215 | G | C | P | A | TP53 | ENST00000269305 | 0.72588 | 0.3963 | 0.84202 | 0.95644 | 0.96956 | 0.6585172623 | 0.96802 | 0.9971824884 | 0.520185 | 4.568124891 | 0 |
| 17 | 7674215 | G | T | P | T | TP53 | ENST00000269305 | 0.75687 | 0.4149 | 0.87888 | 0.97694 | 0.99039 | 0.7320367115 | 0.97974 | 0.9987440109 | 0.584586 | 6.98721634 | 0 |
| 17 | 7674216 | C | A | R | S | TP53 | ENST00000269305 | 0.47564 | 0.41385 | 0.70958 | 0.98736 | 0.92316 | 0.9009793216 | 0.99478 | 0.9880681634 | 0.465778 | 4.250290998 | 1 |
| 17 | 7674216 | C | G | R | S | TP53 | ENST00000269305 | 0.48234 | 0.41385 | 0.70958 | 0.98724 | 0.92316 | 0.9009793216 | 0.99477 | 0.9860873818 | 0.465776 | 4.250290998 | 1 |
| 17 | 7674217 | C | A | R | M | TP53 | ENST00000269305 | 0.75861 | 0.41217 | 0.72929 | 0.98165 | 0.97661 | 0.8740845602 | 0.98535 | 0.9977337122 | 0.563746 | 4.805464207 | 1 |
| 17 | 7674217 | C | G | R | T | TP53 | ENST00000269305 | 0.73701 | 0.45383 | 0.74742 | 0.98253 | 0.98034 | 0.894018284 | 0.99252 | 0.9990002513 | 0.560359 | 6.779615058 | 1 |
| 17 | 7674218 | T | A | R | W | TP53 | ENST00000269305 | 0.43309 | 0.29109 | 0.78687 | 0.98308 | 0.9528 | 0.8955091775 | 0.9779 | 0.9879944324 | 0.51236 | 6.779615058 | 1 |
| 17 | 7674218 | T | C | R | G | TP53 | ENST00000269305 | 0.39952 | 0.43131 | 0.69067 | 0.98234 | 0.9681 | 0.9168133534 | 0.99639 | 0.9967085123 | 0.50533 | 6.779615058 | 1 |
| 17 | 7674220 | C | A | R | L | TP53 | ENST00000269305 | 0.84347 | 0.43001 | 0.6903 | 0.98457 | 0.99386 | 0.9146786228 | 0.99332 | 0.9980717897 | 0.562571 | 7.112700364 | 1 |
| 17 | 7674220 | C | G | R | P | TP53 | ENST00000269305 | 0.86304 | 0.45269 | 0.69501 | 0.9845 | 0.99525 | 0.959622542 | 0.98667 | 0.9978740215 | 0.562621 | 6.045533254 | 1 |
| 17 | 7674220 | C | T | R | Q | TP53 | ENST00000269305 | 0.85122 | 0.4192 | 0.56979 | 0.97876 | 0.98513 | 0.8749344809 | 0.9747 | 0.9815758467 | 0.377622 | 4.687437015 | 1 |
| 17 | 7674221 | G | A | R | W | TP53 | ENST00000269305 | 0.87879 | 0.40509 | 0.75558 | 0.9859 | 0.98257 | 0.9142616771 | 0.98397 | 0.9982162118 | 0.537007 | 7.112700364 | 1 |
| 17 | 7674221 | G | C | R | G | TP53 | ENST00000269305 | 0.77719 | 0.43482 | 0.64059 | 0.99119 | 0.9844 | 0.9398272033 | 0.99693 | 0.9982859492 | 0.536999 | 5.202921328 | 1 |
| 17 | 7674222 | G | T | N | K | TP53 | ENST00000269305 | 0.75973 | 0.41904 | 0.75528 | 0.98911 | 0.96846 | 0.8931579249 | 0.97034 | 0.9987046719 | 0.256613 | 8.699778479 | 0 |
| 17 | 7674223 | T | A | N | I | TP53 | ENST00000269305 | 0.82389 | 0.43034 | 0.67049 | 0.98939 | 0.9917 | 0.8937757348 | 0.97382 | 0.9997751117 | 0.500739 | 8.699778479 | 1 |
| 17 | 7674223 | T | C | N | S | TP53 | ENST00000269305 | 0.72836 | 0.37101 | 0.60636 | 0.98111 | 0.98549 | 0.7609181099 | 0.9674 | 0.9986293316 | 0.264055 | 5.965675259 | 0 |
| 17 | 7674224 | T | G | N | H | TP53 | ENST00000269305 | 0.76808 | 0.40439 | 0.68504 | 0.98475 | 0.98657 | 0.8455158744 | 0.9797 | 0.9993114471 | 0.36014 | 6.620378683 | 0 |
| 17 | 7674225 | C | G | M | I | TP53 | ENST00000269305 | 0.69204 | 0.34191 | 0.72662 | 0.94191 | 0.94986 | 0.570141263 | 0.9298 | 0.9921453595 | 0.354602 | 5.153550425 | 0 |
| 17 | 7674226 | A | G | M | T | TP53 | ENST00000269305 | 0.7657 | 0.41935 | 0.8476 | 0.9908 | 0.99265 | 0.7652920108 | 0.94551 | 0.9988079071 | 0.426885 | 8.819318962 | 1 |
| 17 | 7674226 | A | T | M | K | TP53 | ENST00000269305 | 0.81567 | 0.46443 | 0.80576 | 0.98903 | 0.99471 | 0.85785636 | 0.97028 | 0.9995203018 | 0.524814 | 8.819318962 | 1 |
| 17 | 7674229 | C | A | G | V | TP53 | ENST00000269305 | 0.73429 | 0.43112 | 0.66999 | 0.97819 | 0.997 | 0.8650895408 | 0.99406 | 0.998462677 | 0.464504 | 7.536227782 | 1 |
| 17 | 7674229 | C | A | G | V | TP53 | ENST00000269305 | 0.73429 | 0.43112 | 0.66999 | 0.97819 | 0.997 | 0.8650895408 | 0.99406 | 0.998462677 | 0.464504 | 7.536227782 | 1 |
| 17 | 7674229 | C | A | G | V | TP53 | ENST00000269305 | 0.73429 | 0.43112 | 0.66999 | 0.97819 | 0.997 | 0.8650895408 | 0.99406 | 0.998462677 | 0.464504 | 7.536227782 | 1 |
| 17 | 7674229 | C | G | G | A | TP53 | ENST00000269305 | 0.70423 | 0.41004 | 0.65861 | 0.96676 | 0.98938 | 0.8088383766 | 0.98376 | 0.9972850084 | 0.576197 | 6.515365948 | 1 |
| 17 | 7674229 | C | G | G | A | TP53 | ENST00000269305 | 0.70423 | 0.41004 | 0.65861 | 0.96676 | 0.98938 | 0.8088383766 | 0.98376 | 0.9972850084 | 0.576197 | 6.515365948 | 1 |
| 17 | 7674229 | C | G | G | A | TP53 | ENST00000269305 | 0.70423 | 0.41004 | 0.65861 | 0.96676 | 0.98938 | 0.8088383766 | 0.98376 | 0.9972850084 | 0.576197 | 6.515365948 | 1 |
| 17 | 7674229 | C | T | G | D | TP53 | ENST00000269305 | 0.73685 | 0.43672 | 0.79225 | 0.97589 | 0.99744 | 0.9005840103 | 0.9952 | 0.9963680506 | 0.514803 | 7.536227782 | 1 |
| 17 | 7674229 | C | T | G | D | TP53 | ENST00000269305 | 0.73685 | 0.43672 | 0.79225 | 0.97589 | 0.99744 | 0.9005840103 | 0.9952 | 0.9963680506 | 0.514803 | 7.536227782 | 1 |
| 17 | 7674229 | C | T | G | D | TP53 | ENST00000269305 | 0.73685 | 0.43672 | 0.79225 | 0.97589 | 0.99744 | 0.9005840103 | 0.9952 | 0.9963680506 | 0.514803 | 7.536227782 | 1 |
| 17 | 7674230 | C | A | G | C | TP53 | ENST00000269305 | 0.73389 | 0.4083 | 0.72017 | 0.97531 | 0.99652 | 0.8480159919 | 0.99468 | 0.9998431206 | 0.581969 | 7.536227782 | 1 |
| 17 | 7674230 | C | A | G | C | TP53 | ENST00000269305 | 0.73389 | 0.4083 | 0.72017 | 0.97531 | 0.99652 | 0.8480159919 | 0.99468 | 0.9998431206 | 0.581969 | 7.536227782 | 1 |
| 17 | 7674230 | C | A | G | C | TP53 | ENST00000269305 | 0.73389 | 0.4083 | 0.72017 | 0.97531 | 0.99652 | 0.8480159919 | 0.99468 | 0.9998431206 | 0.581969 | 7.536227782 | 1 |
| 17 | 7674230 | C | G | G | R | TP53 | ENST00000269305 | 0.70527 | 0.42422 | 0.77688 | 0.97812 | 0.99414 | 0.8708231385 | 0.9952 | 0.9994951487 | 0.581969 | 5.792374112 | 1 |
| 17 | 7674230 | C | T | G | S | TP53 | ENST00000269305 | 0.67581 | 0.39939 | 0.64362 | 0.96673 | 0.98972 | 0.803881961 | 0.99348 | 0.9986068606 | 0.550935 | 4.694036397 | 1 |
| 17 | 7674230 | C | T | G | S | TP53 | ENST00000269305 | 0.67581 | 0.39939 | 0.64362 | 0.96673 | 0.98972 | 0.803881961 | 0.99348 | 0.9986068606 | 0.550935 | 4.694036397 | 1 |
| 17 | 7674230 | C | T | G | S | TP53 | ENST00000269305 | 0.67581 | 0.39939 | 0.64362 | 0.96673 | 0.98972 | 0.803881961 | 0.99348 | 0.9986068606 | 0.550935 | 4.694036397 | 1 |
| 17 | 7674232 | C | A | G | V | TP53 | ENST00000269305 | 0.72412 | 0.43112 | 0.67902 | 0.97949 | 0.99039 | 0.8311846342 | 0.98683 | 0.9994433522 | 0.581131 | 8.420993708 | 1 |
| 17 | 7674232 | C | A | G | V | TP53 | ENST00000269305 | 0.72412 | 0.43112 | 0.67902 | 0.97949 | 0.99039 | 0.8311846342 | 0.98683 | 0.9994433522 | 0.581131 | 8.420993708 | 1 |
| 17 | 7674232 | C | A | G | V | TP53 | ENST00000269305 | 0.72412 | 0.43112 | 0.67902 | 0.97949 | 0.99039 | 0.8311846342 | 0.98683 | 0.9994433522 | 0.581131 | 8.420993708 | 1 |
| 17 | 7674232 | C | G | G | A | TP53 | ENST00000269305 | 0.69444 | 0.41004 | 0.67254 | 0.96784 | 0.98071 | 0.7644240087 | 0.96854 | 0.9987832904 | 0.470752 | 8.420993708 | 1 |
| 17 | 7674232 | C | G | G | A | TP53 | ENST00000269305 | 0.69444 | 0.41004 | 0.67254 | 0.96784 | 0.98071 | 0.7644240087 | 0.96854 | 0.9987832904 | 0.470752 | 8.420993708 | 1 |
| 17 | 7674232 | C | G | G | A | TP53 | ENST00000269305 | 0.69444 | 0.41004 | 0.67254 | 0.96784 | 0.98071 | 0.7644240087 | 0.96854 | 0.9987832904 | 0.470752 | 8.420993708 | 1 |
| 17 | 7674232 | C | T | G | D | TP53 | ENST00000269305 | 0.7266 | 0.43672 | 0.78776 | 0.97693 | 0.9917 | 0.8320746011 | 0.98489 | 0.9989257455 | 0.581109 | 7.434508176 | 1 |
| 17 | 7674232 | C | T | G | D | TP53 | ENST00000269305 | 0.7266 | 0.43672 | 0.78776 | 0.97693 | 0.9917 | 0.8320746011 | 0.98489 | 0.9989257455 | 0.581109 | 7.434508176 | 1 |
| 17 | 7674232 | C | T | G | D | TP53 | ENST00000269305 | 0.7266 | 0.43672 | 0.78776 | 0.97693 | 0.9917 | 0.8320746011 | 0.98489 | 0.9989257455 | 0.581109 | 7.434508176 | 1 |
| 17 | 7674233 | C | A | G | C | TP53 | ENST00000269305 | 0.90863 | 0.4083 | 0.73032 | 0.97959 | 0.98728 | 0.8393413557 | 0.9728 | 0.9995982051 | 0.570103 | 8.373027178 | 1 |
| 17 | 7674233 | C | A | G | C | TP53 | ENST00000269305 | 0.90863 | 0.4083 | 0.73032 | 0.97959 | 0.98728 | 0.8393413557 | 0.9728 | 0.9995982051 | 0.570103 | 8.373027178 | 1 |
| 17 | 7674233 | C | A | G | C | TP53 | ENST00000269305 | 0.90863 | 0.4083 | 0.73032 | 0.97959 | 0.98728 | 0.8393413557 | 0.9728 | 0.9995982051 | 0.570103 | 8.373027178 | 1 |
| 17 | 7674233 | C | G | G | R | TP53 | ENST00000269305 | 0.88988 | 0.42422 | 0.77668 | 0.97884 | 0.98799 | 0.8444693446 | 0.97856 | 0.9994433522 | 0.577391 | 8.420993708 | 1 |
| 17 | 7674233 | C | G | G | R | TP53 | ENST00000269305 | 0.88988 | 0.42422 | 0.77668 | 0.97884 | 0.98799 | 0.8444693446 | 0.97856 | 0.9994433522 | 0.577391 | 8.420993708 | 1 |
| 17 | 7674233 | C | G | G | R | TP53 | ENST00000269305 | 0.88988 | 0.42422 | 0.77668 | 0.97884 | 0.98799 | 0.8444693446 | 0.97856 | 0.9994433522 | 0.577391 | 8.420993708 | 1 |
| 17 | 7674233 | C | T | G | S | TP53 | ENST00000269305 | 0.86524 | 0.39939 | 0.67714 | 0.9709 | 0.98585 | 0.7669178742 | 0.96951 | 0.9988603592 | 0.487533 | 5.393392412 | 1 |
| 17 | 7674233 | C | T | G | S | TP53 | ENST00000269305 | 0.86524 | 0.39939 | 0.67714 | 0.9709 | 0.98585 | 0.7669178742 | 0.96951 | 0.9988603592 | 0.487533 | 5.393392412 | 1 |
| 17 | 7674233 | C | T | G | S | TP53 | ENST00000269305 | 0.86524 | 0.39939 | 0.67714 | 0.9709 | 0.98585 | 0.7669178742 | 0.96951 | 0.9988603592 | 0.487533 | 5.393392412 | 1 |
| 17 | 7674234 | C | A | M | I | TP53 | ENST00000269305 | 0.73333 | 0.41245 | 0.42958 | 0.98278 | 0.94318 | 0.5115582358 | 0.87197 | 0.9836936593 | 0.237795 | 6.299480579 | 0 |
| 17 | 7674234 | C | A | M | I | TP53 | ENST00000269305 | 0.73333 | 0.41245 | 0.42958 | 0.98278 | 0.94318 | 0.5115582358 | 0.87197 | 0.9836936593 | 0.237795 | 6.299480579 | 0 |
| 17 | 7674234 | C | A | M | I | TP53 | ENST00000269305 | 0.73333 | 0.41245 | 0.42958 | 0.98278 | 0.94318 | 0.5115582358 | 0.87197 | 0.9836936593 | 0.237795 | 6.299480579 | 0 |
| 17 | 7674234 | C | G | M | I | TP53 | ENST00000269305 | 0.74181 | 0.41245 | 0.42958 | 0.98183 | 0.94318 | 0.5115582358 | 0.87257 | 0.9836936593 | 0.237802 | 6.299480579 | 0 |
| 17 | 7674234 | C | G | M | I | TP53 | ENST00000269305 | 0.74181 | 0.41245 | 0.42958 | 0.98183 | 0.94318 | 0.5115582358 | 0.87257 | 0.9836936593 | 0.237802 | 6.299480579 | 0 |
| 17 | 7674234 | C | G | M | I | TP53 | ENST00000269305 | 0.74181 | 0.41245 | 0.42958 | 0.98183 | 0.94318 | 0.5115582358 | 0.87257 | 0.9836936593 | 0.237802 | 6.299480579 | 0 |
| 17 | 7674234 | C | T | M | I | TP53 | ENST00000269305 | 0.74722 | 0.41245 | 0.42958 | 0.98159 | 0.94318 | 0.5115582358 | 0.87191 | 0.9836936593 | 0.237795 | 6.299480579 | 0 |
| 17 | 7674234 | C | T | M | I | TP53 | ENST00000269305 | 0.74722 | 0.41245 | 0.42958 | 0.98159 | 0.94318 | 0.5115582358 | 0.87191 | 0.9836936593 | 0.237795 | 6.299480579 | 0 |
| 17 | 7674235 | A | C | M | R | TP53 | ENST00000269305 | 0.90488 | 0.46101 | 0.50066 | 0.98617 | 0.96883 | 0.8232339275 | 0.94108 | 0.9992597699 | 0.397906 | 7.064572929 | 0 |
| 17 | 7674235 | A | C | M | R | TP53 | ENST00000269305 | 0.90488 | 0.46101 | 0.50066 | 0.98617 | 0.96883 | 0.8232339275 | 0.94108 | 0.9992597699 | 0.397906 | 7.064572929 | 0 |
| 17 | 7674235 | A | C | M | R | TP53 | ENST00000269305 | 0.90488 | 0.46101 | 0.50066 | 0.98617 | 0.96883 | 0.8232339275 | 0.94108 | 0.9992597699 | 0.397906 | 7.064572929 | 0 |
| 17 | 7674235 | A | G | M | T | TP53 | ENST00000269305 | 0.85212 | 0.41895 | 0.58861 | 0.98821 | 0.9648 | 0.664552845 | 0.93393 | 0.9978519678 | 0.373759 | 5.444527323 | 0 |
| 17 | 7674235 | A | G | M | T | TP53 | ENST00000269305 | 0.85212 | 0.41895 | 0.58861 | 0.98821 | 0.9648 | 0.664552845 | 0.93393 | 0.9978519678 | 0.373759 | 5.444527323 | 0 |
| 17 | 7674235 | A | G | M | T | TP53 | ENST00000269305 | 0.85212 | 0.41895 | 0.58861 | 0.98821 | 0.9648 | 0.664552845 | 0.93393 | 0.9978519678 | 0.373759 | 5.444527323 | 0 |
| 17 | 7674235 | A | T | M | K | TP53 | ENST00000269305 | 0.89417 | 0.46402 | 0.52936 | 0.98862 | 0.98477 | 0.7970705732 | 0.93273 | 0.9995650649 | 0.405479 | 7.618893393 | 0 |
| 17 | 7674235 | A | T | M | K | TP53 | ENST00000269305 | 0.89417 | 0.46402 | 0.52936 | 0.98862 | 0.98477 | 0.7970705732 | 0.93273 | 0.9995650649 | 0.405479 | 7.618893393 | 0 |
| 17 | 7674235 | A | T | M | K | TP53 | ENST00000269305 | 0.89417 | 0.46402 | 0.52936 | 0.98862 | 0.98477 | 0.7970705732 | 0.93273 | 0.9995650649 | 0.405479 | 7.618893393 | 0 |
| 17 | 7674236 | T | A | M | L | TP53 | ENST00000269305 | 0.70077 | 0.38952 | 0.42144 | 0.98489 | 0.92069 | 0.4245412586 | 0.89552 | 0.8278625011 | 0.226227 | 5.805369334 | 0 |
| 17 | 7674236 | T | A | M | L | TP53 | ENST00000269305 | 0.70077 | 0.38952 | 0.42144 | 0.98489 | 0.92069 | 0.4245412586 | 0.89552 | 0.8278625011 | 0.226227 | 5.805369334 | 0 |
| 17 | 7674236 | T | A | M | L | TP53 | ENST00000269305 | 0.70077 | 0.38952 | 0.42144 | 0.98489 | 0.92069 | 0.4245412586 | 0.89552 | 0.8278625011 | 0.226227 | 5.805369334 | 0 |
| 17 | 7674236 | T | C | M | V | TP53 | ENST00000269305 | 0.67223 | 0.40038 | 0.34025 | 0.93283 | 0.89558 | 0.5425333272 | 0.86472 | 0.7435705662 | 0.171079 | 5.796715666 | 0 |
| 17 | 7674236 | T | C | M | V | TP53 | ENST00000269305 | 0.67223 | 0.40038 | 0.34025 | 0.93283 | 0.89558 | 0.5425333272 | 0.86472 | 0.7435705662 | 0.171079 | 5.796715666 | 0 |
| 17 | 7674236 | T | C | M | V | TP53 | ENST00000269305 | 0.67223 | 0.40038 | 0.34025 | 0.93283 | 0.89558 | 0.5425333272 | 0.86472 | 0.7435705662 | 0.171079 | 5.796715666 | 0 |
| 17 | 7674236 | T | G | M | L | TP53 | ENST00000269305 | 0.70903 | 0.38952 | 0.42144 | 0.98371 | 0.92069 | 0.4245412586 | 0.89573 | 0.8359814868 | 0.226229 | 5.805369334 | 0 |
| 17 | 7674236 | T | G | M | L | TP53 | ENST00000269305 | 0.70903 | 0.38952 | 0.42144 | 0.98371 | 0.92069 | 0.4245412586 | 0.89573 | 0.8359814868 | 0.226229 | 5.805369334 | 0 |
| 17 | 7674236 | T | G | M | L | TP53 | ENST00000269305 | 0.70903 | 0.38952 | 0.42144 | 0.98371 | 0.92069 | 0.4245412586 | 0.89573 | 0.8359814868 | 0.226229 | 5.805369334 | 0 |
| 17 | 7674237 | G | C | C | W | TP53 | ENST00000269305 | 0.92511 | 0.44218 | 0.83564 | 0.97918 | 0.98728 | 0.910769811 | 0.99497 | 0.9998545647 | 0.541846 | 8.535452659 | 0 |
| 17 | 7674238 | C | A | C | F | TP53 | ENST00000269305 | 0.78946 | 0.4623 | 0.79124 | 0.97953 | 0.99806 | 0.9064284511 | 0.98934 | 0.9998233914 | 0.583204 | 8.535452659 | 1 |
| 17 | 7674238 | C | A | C | F | TP53 | ENST00000269305 | 0.78946 | 0.4623 | 0.79124 | 0.97953 | 0.99806 | 0.9064284511 | 0.98934 | 0.9998233914 | 0.583204 | 8.535452659 | 1 |

| | | | | | | | | | | | | | | | | | | |
|---|---|---|---|---|---|---|---|---|---|---|---|---|---|---|---|---|---|---|
| 17 | 7674238 | C | G | C | S | TP53 | ENST00000269305 | 0.75232 | 0.42828 | 0.767 | 0.97896 | 0.98972 | 0.8894311524 | 0.95493 | 0.9991539717 | 0.512496 | 7.332145152 | 1 |
| 17 | 7674238 | C | G | C | S | TP53 | ENST00000269305 | 0.75232 | 0.42828 | 0.767 | 0.97896 | 0.98972 | 0.8894311524 | 0.95493 | 0.9991539717 | 0.512496 | 7.332145152 | 1 |
| 17 | 7674238 | C | G | C | S | TP53 | ENST00000269305 | 0.75232 | 0.42828 | 0.767 | 0.97896 | 0.98972 | 0.8894311524 | 0.95493 | 0.9991539717 | 0.512496 | 7.332145152 | 1 |
| 17 | 7674238 | C | T | C | Y | TP53 | ENST00000269305 | 0.77172 | 0.47886 | 0.88864 | 0.97925 | 0.99722 | 0.9066372751 | 0.98049 | 0.9998125434 | 0.583201 | 8.535452659 | 1 |
| 17 | 7674238 | C | T | C | Y | TP53 | ENST00000269305 | 0.77172 | 0.47886 | 0.88864 | 0.97925 | 0.99722 | 0.9066372751 | 0.98049 | 0.9998125434 | 0.583201 | 8.535452659 | 1 |
| 17 | 7674238 | C | T | C | Y | TP53 | ENST00000269305 | 0.77172 | 0.47886 | 0.88864 | 0.97925 | 0.99722 | 0.9066372751 | 0.98049 | 0.9998125434 | 0.583201 | 8.535452659 | 1 |
| 17 | 7674239 | A | G | C | R | TP53 | ENST00000269305 | 0.93265 | 0.48683 | 0.8722 | 0.98863 | 0.99552 | 0.9218795793 | 0.99179 | 0.9999185801 | 0.588171 | 7.346504617 | 1 |
| 17 | 7674239 | A | G | C | R | TP53 | ENST00000269305 | 0.93265 | 0.48683 | 0.8722 | 0.98863 | 0.99552 | 0.9218795793 | 0.99179 | 0.9999185801 | 0.588171 | 7.346504617 | 1 |
| 17 | 7674239 | A | G | C | R | TP53 | ENST00000269305 | 0.93265 | 0.48683 | 0.8722 | 0.98863 | 0.99552 | 0.9218795793 | 0.99179 | 0.9999185801 | 0.588171 | 7.346504617 | 1 |
| 17 | 7674239 | A | T | C | S | TP53 | ENST00000269305 | 0.9018 | 0.42828 | 0.767 | 0.99156 | 0.99386 | 0.8894311524 | 0.96429 | 0.9998753071 | 0.575151 | 7.332145152 | 1 |
| 17 | 7674239 | A | T | C | S | TP53 | ENST00000269305 | 0.9018 | 0.42828 | 0.767 | 0.99156 | 0.99386 | 0.8894311524 | 0.96429 | 0.9998753071 | 0.575151 | 7.332145152 | 1 |
| 17 | 7674239 | A | T | C | S | TP53 | ENST00000269305 | 0.9018 | 0.42828 | 0.767 | 0.99156 | 0.99386 | 0.8894311524 | 0.96429 | 0.9998753071 | 0.575151 | 7.332145152 | 1 |
| 17 | 7674241 | G | A | S | F | TP53 | ENST00000269305 | 0.8305 | 0.41226 | 0.6957 | 0.98952 | 0.99326 | 0.9049090378 | 0.99196 | 0.9978739023 | 0.544964 | 7.94203853 | 1 |
| 17 | 7674241 | G | A | S | F | TP53 | ENST00000269305 | 0.8305 | 0.41226 | 0.6957 | 0.98952 | 0.99326 | 0.9049090378 | 0.99196 | 0.9978739023 | 0.544964 | 7.94203853 | 1 |
| 17 | 7674241 | G | A | S | F | TP53 | ENST00000269305 | 0.8305 | 0.41226 | 0.6957 | 0.98952 | 0.99326 | 0.9049090378 | 0.99196 | 0.9978739023 | 0.544964 | 7.94203853 | 1 |
| 17 | 7674241 | G | C | S | C | TP53 | ENST00000269305 | 0.81528 | 0.39643 | 0.65808 | 0.98687 | 0.99138 | 0.8499942974 | 0.97137 | 0.9982457161 | 0.520653 | 7.94203853 | 1 |
| 17 | 7674241 | G | C | S | C | TP53 | ENST00000269305 | 0.81528 | 0.39643 | 0.65808 | 0.98687 | 0.99138 | 0.8499942974 | 0.97137 | 0.9982457161 | 0.520653 | 7.94203853 | 1 |
| 17 | 7674241 | G | C | S | C | TP53 | ENST00000269305 | 0.81528 | 0.39643 | 0.65808 | 0.98687 | 0.99138 | 0.8499942974 | 0.97137 | 0.9982457161 | 0.520653 | 7.94203853 | 1 |
| 17 | 7674241 | G | T | S | Y | TP53 | ENST00000269305 | 0.78535 | 0.43474 | 0.72259 | 0.98941 | 0.99386 | 0.891875814 | 0.99159 | 0.9986697435 | 0.544724 | 7.94203853 | 1 |
| 17 | 7674241 | G | T | S | Y | TP53 | ENST00000269305 | 0.78535 | 0.43474 | 0.72259 | 0.98941 | 0.99386 | 0.891875814 | 0.99159 | 0.9986697435 | 0.544724 | 7.94203853 | 1 |
| 17 | 7674242 | A | C | S | A | TP53 | ENST00000269305 | 0.74938 | 0.37662 | 0.63033 | 0.98881 | 0.95717 | 0.7536346786 | 0.96055 | 0.9967104197 | 0.405599 | 6.803310191 | 1 |
| 17 | 7674242 | A | C | S | A | TP53 | ENST00000269305 | 0.74938 | 0.37662 | 0.63033 | 0.98881 | 0.95717 | 0.7536346786 | 0.96055 | 0.9967104197 | 0.405599 | 6.803310191 | 1 |
| 17 | 7674242 | A | G | S | P | TP53 | ENST00000269305 | 0.80013 | 0.43212 | 0.6646 | 0.99261 | 0.99202 | 0.9086019472 | 0.98865 | 0.9984089732 | 0.542659 | 7.821626889 | 1 |
| 17 | 7674242 | A | G | S | P | TP53 | ENST00000269305 | 0.80013 | 0.43212 | 0.6646 | 0.99261 | 0.99202 | 0.9086019472 | 0.98865 | 0.9984089732 | 0.542659 | 7.821626889 | 1 |
| 17 | 7674242 | A | G | S | P | TP53 | ENST00000269305 | 0.80013 | 0.43212 | 0.6646 | 0.99261 | 0.99202 | 0.9086019472 | 0.98865 | 0.9984089732 | 0.542659 | 7.821626889 | 1 |
| 17 | 7674242 | A | T | S | T | TP53 | ENST00000269305 | 0.70823 | 0.38736 | 0.65342 | 0.99246 | 0.98071 | 0.8207573584 | 0.98757 | 0.9966920614 | 0.530973 | 4.554669141 | 1 |
| 17 | 7674242 | A | T | S | T | TP53 | ENST00000269305 | 0.70823 | 0.38736 | 0.65342 | 0.99246 | 0.98071 | 0.8207573584 | 0.98757 | 0.9966920614 | 0.530973 | 4.554669141 | 1 |
| 17 | 7674242 | A | T | S | T | TP53 | ENST00000269305 | 0.70823 | 0.38736 | 0.65342 | 0.99246 | 0.98071 | 0.8207573584 | 0.98757 | 0.9966920614 | 0.530973 | 4.554669141 | 1 |
| 17 | 7674243 | A | C | S | R | TP53 | ENST00000269305 | 0.71546 | 0.41524 | 0.6046 | 0.99283 | 0.9755 | 0.786339412 | 0.97992 | 0.9991233945 | 0.501083 | 9.51794321 | 1 |
| 17 | 7674243 | A | T | S | R | TP53 | ENST00000269305 | 0.70872 | 0.41524 | 0.6046 | 0.99328 | 0.9755 | 0.786339412 | 0.97964 | 0.9992496371 | 0.501047 | 9.51794321 | 1 |
| 17 | 7674243 | A | T | S | R | TP53 | ENST00000269305 | 0.70872 | 0.41524 | 0.6046 | 0.99328 | 0.9755 | 0.786339412 | 0.97964 | 0.9992496371 | 0.501047 | 9.51794321 | 1 |
| 17 | 7674244 | C | A | S | I | TP53 | ENST00000269305 | 0.77386 | 0.42299 | 0.56095 | 0.98116 | 0.96627 | 0.7740949518 | 0.97429 | 0.9986257553 | 0.513756 | 6.908200992 | 0 |
| 17 | 7674244 | C | A | S | I | TP53 | ENST00000269305 | 0.77386 | 0.42299 | 0.56095 | 0.98116 | 0.96627 | 0.7740949518 | 0.97429 | 0.9986257553 | 0.513756 | 6.908200992 | 0 |
| 17 | 7674244 | C | G | S | T | TP53 | ENST00000269305 | 0.70366 | 0.38813 | 0.52816 | 0.96732 | 0.93554 | 0.5873397289 | 0.92536 | 0.9961435795 | 0.326603 | 5.837412677 | 0 |
| 17 | 7674244 | C | G | S | T | TP53 | ENST00000269305 | 0.70366 | 0.38813 | 0.52816 | 0.96732 | 0.93554 | 0.5873397289 | 0.92536 | 0.9961435795 | 0.326603 | 5.837412677 | 0 |
| 17 | 7674244 | C | G | S | T | TP53 | ENST00000269305 | 0.70366 | 0.38813 | 0.52816 | 0.96732 | 0.93554 | 0.5873397289 | 0.92536 | 0.9961435795 | 0.326603 | 5.837412677 | 0 |
| 17 | 7674244 | C | T | S | N | TP53 | ENST00000269305 | 0.71209 | 0.39071 | 0.59234 | 0.97268 | 0.91526 | 0.6084769586 | 0.91135 | 0.9968777895 | 0.236908 | 5.810677889 | 0 |
| 17 | 7674244 | C | T | S | N | TP53 | ENST00000269305 | 0.71209 | 0.39071 | 0.59234 | 0.97268 | 0.91526 | 0.6084769586 | 0.91135 | 0.9968777895 | 0.236908 | 5.810677889 | 0 |
| 17 | 7674245 | T | A | S | C | TP53 | ENST00000269305 | 0.85916 | 0.39643 | 0.53525 | 0.98809 | 0.98109 | 0.6586525489 | 0.96102 | 0.9989868999 | 0.455538 | 7.749082447 | 0 |
| 17 | 7674245 | T | A | S | C | TP53 | ENST00000269305 | 0.85916 | 0.39643 | 0.53525 | 0.98809 | 0.98109 | 0.6586525489 | 0.96102 | 0.9989868999 | 0.455538 | 7.749082447 | 1 |
| 17 | 7674245 | T | C | S | G | TP53 | ENST00000269305 | 0.8263 | 0.38919 | 0.47018 | 0.98565 | 0.96226 | 0.613556501 | 0.93638 | 0.997825563 | 0.339231 | 6.59390832 | 1 |
| 17 | 7674245 | T | G | S | R | TP53 | ENST00000269305 | 0.851 | 0.41524 | 0.6046 | 0.98784 | 0.98693 | 0.786339412 | 0.97273 | 0.9993194342 | 0.521431 | 9.51794321 | 1 |
| 17 | 7674245 | T | G | S | R | TP53 | ENST00000269305 | 0.851 | 0.41524 | 0.6046 | 0.98784 | 0.98693 | 0.786339412 | 0.97273 | 0.9993194342 | 0.521431 | 9.51794321 | 1 |
| 17 | 7674246 | G | C | N | K | TP53 | ENST00000269305 | 0.79302 | 0.41904 | 0.68135 | 0.98846 | 0.93977 | 0.7480218942 | 0.95505 | 0.9960423708 | 0.241124 | 5.74714985 | 1 |
| 17 | 7674246 | G | T | N | K | TP53 | ENST00000269305 | 0.78653 | 0.41904 | 0.68135 | 0.98843 | 0.93977 | 0.7480218942 | 0.95495 | 0.9967874289 | 0.241091 | 5.74714985 | 1 |
| 17 | 7674247 | T | A | N | I | TP53 | ENST00000269305 | 0.83637 | 0.43034 | 0.59668 | 0.99339 | 0.98834 | 0.7586555427 | 0.96436 | 0.9997751117 | 0.44401 | 8.526922906 | 1 |
| 17 | 7674247 | T | C | N | S | TP53 | ENST00000269305 | 0.71674 | 0.37178 | 0.52963 | 0.9888 | 0.97141 | 0.5963004584 | 0.97206 | 0.9935675263 | 0.238888 | 6.535703573 | 1 |
| 17 | 7674247 | T | C | N | S | TP53 | ENST00000269305 | 0.71674 | 0.37178 | 0.52963 | 0.9888 | 0.97141 | 0.5963004584 | 0.97206 | 0.9935675263 | 0.238888 | 6.535703573 | 1 |
| 17 | 7674247 | T | C | N | S | TP53 | ENST00000269305 | 0.71674 | 0.37178 | 0.52963 | 0.9888 | 0.97141 | 0.5963004584 | 0.97206 | 0.9935675263 | 0.238888 | 6.535703573 | 1 |
| 17 | 7674247 | T | G | N | T | TP53 | ENST00000269305 | 0.79453 | 0.40251 | 0.57733 | 0.99028 | 0.98549 | 0.6798587714 | 0.96352 | 0.9981423616 | 0.310525 | 6.837283226 | 1 |
| 17 | 7674247 | T | G | N | T | TP53 | ENST00000269305 | 0.79453 | 0.40251 | 0.57733 | 0.99028 | 0.98549 | 0.6798587714 | 0.96352 | 0.9981423616 | 0.310525 | 6.837283226 | 1 |
| 17 | 7674247 | T | G | N | T | TP53 | ENST00000269305 | 0.79453 | 0.40251 | 0.57733 | 0.99028 | 0.98549 | 0.6798587714 | 0.96352 | 0.9981423616 | 0.310525 | 6.837283226 | 1 |
| 17 | 7674248 | T | A | N | Y | TP53 | ENST00000269305 | 0.89515 | 0.42727 | 0.63796 | 0.98916 | 0.97438 | 0.7495569514 | 0.97345 | 0.9994698167 | 0.531089 | 6.009155813 | 0 |
| 17 | 7674248 | T | A | N | Y | TP53 | ENST00000269305 | 0.89515 | 0.42727 | 0.63796 | 0.98916 | 0.97438 | 0.7495569514 | 0.97345 | 0.9994698167 | 0.531089 | 6.009155813 | 0 |
| 17 | 7674248 | T | A | N | Y | TP53 | ENST00000269305 | 0.89515 | 0.42727 | 0.63796 | 0.98916 | 0.97438 | 0.7495569514 | 0.97345 | 0.9994698167 | 0.531089 | 6.009155813 | 0 |
| 17 | 7674248 | T | C | N | D | TP53 | ENST00000269305 | 0.88022 | 0.41725 | 0.64976 | 0.98631 | 0.99006 | 0.7234787159 | 0.96949 | 0.9973301888 | 0.308347 | 8.494736683 | 1 |
| 17 | 7674248 | T | C | N | D | TP53 | ENST00000269305 | 0.88022 | 0.41725 | 0.64976 | 0.98631 | 0.99006 | 0.7234787159 | 0.96949 | 0.9973301888 | 0.308347 | 8.494736683 | 1 |
| 17 | 7674248 | T | C | N | D | TP53 | ENST00000269305 | 0.88022 | 0.41725 | 0.64976 | 0.98631 | 0.99006 | 0.7234787159 | 0.96949 | 0.9973301888 | 0.308347 | 8.494736683 | 1 |
| 17 | 7674248 | T | G | N | H | TP53 | ENST00000269305 | 0.85691 | 0.40439 | 0.60393 | 0.98735 | 0.98257 | 0.6971962973 | 0.96965 | 0.9990971088 | 0.304082 | 6.690484971 | 0 |
| 17 | 7674248 | T | G | N | H | TP53 | ENST00000269305 | 0.85691 | 0.40439 | 0.60393 | 0.98735 | 0.98257 | 0.6971962973 | 0.96965 | 0.9990971088 | 0.304082 | 6.690484971 | 0 |
| 17 | 7674249 | A | C | C | W | TP53 | ENST00000269305 | 0.82576 | 0.44218 | 0.93339 | 0.99621 | 0.97104 | 0.9469452857 | 0.99768 | 0.9998728037 | 0.538724 | 7.774754717 | 1 |
| 17 | 7674249 | A | C | C | W | TP53 | ENST00000269305 | 0.82576 | 0.44218 | 0.93339 | 0.99621 | 0.97104 | 0.9469452857 | 0.99768 | 0.9998728037 | 0.538724 | 7.774754717 | 1 |
| 17 | 7674249 | A | C | C | W | TP53 | ENST00000269305 | 0.82576 | 0.44218 | 0.93339 | 0.99621 | 0.97104 | 0.9469452857 | 0.99768 | 0.9998728037 | 0.538724 | 7.774754717 | 1 |
| 17 | 7674250 | C | A | C | F | TP53 | ENST00000269305 | 0.7878 | 0.4623 | 0.9013 | 0.98648 | 0.99326 | 0.9360855168 | 0.99387 | 0.9997820258 | 0.583032 | 7.774754717 | 1 |
| 17 | 7674250 | C | A | C | F | TP53 | ENST00000269305 | 0.7878 | 0.4623 | 0.9013 | 0.98648 | 0.99326 | 0.9360855168 | 0.99387 | 0.9997820258 | 0.583032 | 7.774754717 | 1 |
| 17 | 7674250 | C | A | C | F | TP53 | ENST00000269305 | 0.7878 | 0.4623 | 0.9013 | 0.98648 | 0.99326 | 0.9360855168 | 0.99387 | 0.9997820258 | 0.583032 | 7.774754717 | 1 |
| 17 | 7674250 | C | G | C | S | TP53 | ENST00000269305 | 0.75065 | 0.42828 | 0.88946 | 0.98577 | 0.98938 | 0.9257870139 | 0.99137 | 0.9997686744 | 0.583033 | 7.774754717 | 1 |
| 17 | 7674250 | C | G | C | S | TP53 | ENST00000269305 | 0.75065 | 0.42828 | 0.88946 | 0.98577 | 0.98938 | 0.9257870139 | 0.99137 | 0.9997686744 | 0.583033 | 7.774754717 | 1 |
| 17 | 7674250 | C | G | C | S | TP53 | ENST00000269305 | 0.75065 | 0.42828 | 0.88946 | 0.98577 | 0.98938 | 0.9257870139 | 0.99137 | 0.9997686744 | 0.583033 | 7.774754717 | 1 |
| 17 | 7674250 | C | T | C | Y | TP53 | ENST00000269305 | 0.77006 | 0.47886 | 0.95995 | 0.98645 | 0.99296 | 0.9480238331 | 0.99269 | 0.9994176626 | 0.583029 | 7.774754717 | 1 |
| 17 | 7674250 | C | T | C | Y | TP53 | ENST00000269305 | 0.77006 | 0.47886 | 0.95995 | 0.98645 | 0.99296 | 0.9480238331 | 0.99269 | 0.9994176626 | 0.583029 | 7.774754717 | 1 |
| 17 | 7674250 | C | T | C | Y | TP53 | ENST00000269305 | 0.77006 | 0.47886 | 0.95995 | 0.98645 | 0.99296 | 0.9480238331 | 0.99269 | 0.9994176626 | 0.583029 | 7.774754717 | 1 |
| 17 | 7674251 | A | C | C | G | TP53 | ENST00000269305 | 0.84134 | 0.43007 | 0.86449 | 0.99522 | 0.98657 | 0.9398723723 | 0.995 | 0.9998657703 | 0.579568 | 7.740015724 | 1 |
| 17 | 7674251 | A | C | C | G | TP53 | ENST00000269305 | 0.84134 | 0.43007 | 0.86449 | 0.99522 | 0.98657 | 0.9398723723 | 0.995 | 0.9998657703 | 0.579568 | 7.740015724 | 1 |
| 17 | 7674251 | A | C | C | G | TP53 | ENST00000269305 | 0.84134 | 0.43007 | 0.86449 | 0.99522 | 0.98657 | 0.9398723723 | 0.995 | 0.9998657703 | 0.579568 | 7.740015724 | 1 |
| 17 | 7674251 | A | G | C | R | TP53 | ENST00000269305 | 0.85456 | 0.48683 | 0.95384 | 0.99476 | 0.99202 | 0.9589191038 | 0.99457 | 0.9999172688 | 0.588435 | 7.774754717 | 1 |
| 17 | 7674251 | A | G | C | R | TP53 | ENST00000269305 | 0.85456 | 0.48683 | 0.95384 | 0.99476 | 0.99202 | 0.9589191038 | 0.99457 | 0.9999172688 | 0.588435 | 7.774754717 | 1 |
| 17 | 7674251 | A | G | C | R | TP53 | ENST00000269305 | 0.85456 | 0.48683 | 0.95384 | 0.99476 | 0.99202 | 0.9589191038 | 0.99457 | 0.9999172688 | 0.588435 | 7.774754717 | 1 |
| 17 | 7674251 | A | T | C | S | TP53 | ENST00000269305 | 0.79674 | 0.42828 | 0.88946 | 0.99461 | 0.99265 | 0.9257870139 | 0.99323 | 0.9999150038 | 0.588435 | 7.774754717 | 1 |
| 17 | 7674251 | A | T | C | S | TP53 | ENST00000269305 | 0.79674 | 0.42828 | 0.88946 | 0.99461 | 0.99265 | 0.9257870139 | 0.99323 | 0.9999150038 | 0.588435 | 7.774754717 | 1 |
| 17 | 7674251 | A | T | C | S | TP53 | ENST00000269305 | 0.79674 | 0.42828 | 0.88946 | 0.99461 | 0.99265 | 0.9257870139 | 0.99323 | 0.9999150038 | 0.588435 | 7.774754717 | 1 |
| 17 | 7674252 | C | A | M | I | TP53 | ENST00000269305 | 0.69525 | 0.41245 | 0.70604 | 0.98699 | 0.98109 | 0.67765396 | 0.9645 | 0.9977282882 | 0.473184 | 7.021959488 | 1 |
| 17 | 7674252 | C | A | M | I | TP53 | ENST00000269305 | 0.69525 | 0.41245 | 0.70604 | 0.98699 | 0.98109 | 0.67765396 | 0.9645 | 0.9977282882 | 0.473184 | 7.021959488 | 1 |
| 17 | 7674252 | C | G | M | I | TP53 | ENST00000269305 | 0.70374 | 0.41245 | 0.70604 | 0.98596 | 0.98109 | 0.67765396 | 0.96461 | 0.9977282882 | 0.473186 | 7.021959488 | 1 |
| 17 | 7674252 | C | G | M | I | TP53 | ENST00000269305 | 0.70374 | 0.41245 | 0.70604 | 0.98596 | 0.98109 | 0.67765396 | 0.96461 | 0.9977282882 | 0.473186 | 7.021959488 | 1 |
| 17 | 7674252 | C | G | M | I | TP53 | ENST00000269305 | 0.70374 | 0.41245 | 0.70604 | 0.98596 | 0.98109 | 0.67765396 | 0.96461 | 0.9977282882 | 0.473186 | 7.021959488 | 1 |
| 17 | 7674252 | C | T | M | I | TP53 | ENST00000269305 | 0.70919 | 0.41245 | 0.70604 | 0.98691 | 0.98109 | 0.67765396 | 0.96382 | 0.9900884032 | 0.355437 | 7.021959488 | 1 |
| 17 | 7674252 | C | T | M | I | TP53 | ENST00000269305 | 0.70919 | 0.41245 | 0.70604 | 0.98691 | 0.98109 | 0.67765396 | 0.96382 | 0.9900884032 | 0.355437 | 7.021959488 | 1 |
| 17 | 7674252 | C | T | M | I | TP53 | ENST00000269305 | 0.70919 | 0.41245 | 0.70604 | 0.98691 | 0.98109 | 0.67765396 | 0.96382 | 0.9900884032 | 0.355437 | 7.021959488 | 1 |
| 17 | 7674253 | A | C | M | R | TP53 | ENST00000269305 | 0.83197 | 0.46101 | 0.76079 | 0.99238 | 0.99006 | 0.9195180121 | 0.99035 | 0.999260962 | 0.586906 | 8.319859716 | 0 |
| 17 | 7674253 | A | C | M | R | TP53 | ENST00000269305 | 0.83197 | 0.46101 | 0.76079 | 0.99238 | 0.99006 | 0.9195180121 | 0.99035 | 0.999260962 | 0.586906 | 8.319859716 | 0 |
| 17 | 7674253 | A | G | M | T | TP53 | ENST00000269305 | 0.76674 | 0.41895 | 0.81769 | 0.99161 | 0.99006 | 0.7697370804 | 0.97167 | 0.9992324114 | 0.525286 | 5.281244871 | 0 |
| 17 | 7674253 | A | G | M | T | TP53 | ENST00000269305 | 0.76674 | 0.41895 | 0.81769 | 0.99161 | 0.99006 | 0.7697370804 | 0.97167 | 0.9992324114 | 0.525286 | 5.281244871 | 0 |
| 17 | 7674253 | A | T | M | K | TP53 | ENST00000269305 | 0.81661 | 0.46402 | 0.78262 | 0.99143 | 0.99296 | 0.9029880749 | 0.97782 | 0.9994004965 | 0.552028 | 5.605781683 | 1 |
| 17 | 7674253 | A | T | M | K | TP53 | ENST00000269305 | 0.81661 | 0.46402 | 0.78262 | 0.99143 | 0.99296 | 0.9029880749 | 0.97782 | 0.9994004965 | 0.552028 | 5.605781683 | 1 |
| 17 | 7674253 | A | T | M | K | TP53 | ENST00000269305 | 0.81661 | 0.46402 | 0.78262 | 0.99143 | 0.99296 | 0.9029880749 | 0.97782 | 0.9994004965 | 0.552028 | 5.605781683 | 1 |
| 17 | 7674254 | T | A | M | L | TP53 | ENST00000269305 | 0.84876 | 0.38952 | 0.71026 | 0.98016 | 0.98764 | 0.5883932406 | 0.95425 | 0.9774301052 | 0.424856 | 6.090996763 | 1 |
| 17 | 7674254 | T | A | M | L | TP53 | ENST00000269305 | 0.84876 | 0.38952 | 0.71026 | 0.98016 | 0.98764 | 0.5883932406 | 0.95425 | 0.9774301052 | 0.424856 | 6.090996763 | 0 |
| 17 | 7674254 | T | A | M | L | TP53 | ENST00000269305 | 0.84876 | 0.38952 | 0.71026 | 0.98016 | 0.98764 | 0.5883932406 | 0.95425 | 0.9774301052 | 0.424856 | 6.090996763 | 0 |
| 17 | 7674254 | T | C | M | V | TP53 | ENST00000269305 | 0.81489 | 0.40038 | 0.6394 | 0.98542 | 0.98621 | 0.6738641162 | 0.97392 | 0.9946507817 | 0.485997 | 5.166198375 | 1 |
| 17 | 7674254 | T | G | M | L | TP53 | ENST00000269305 | 0.85632 | 0.38952 | 0.71026 | 0.97644 | 0.98764 | 0.5883932406 | 0.95436 | 0.9774301052 | 0.424856 | 6.090996763 | 1 |

| | | | | | | | | | | | | | | | | | | |
|---|---|---|---|---|---|---|---|---|---|---|---|---|---|---|---|---|---|---|
| 17 | 7674254 | T | G | M | L | TP53 | ENST00000269305 | 0.85632 | 0.38952 | 0.71026 | 0.97644 | 0.98764 | 0.5883932406 | 0.95436 | 0.9774301052 | 0.424856 | 6.090996763 | 0 |
| 17 | 7674254 | T | G | M | L | TP53 | ENST00000269305 | 0.85632 | 0.38952 | 0.71026 | 0.97644 | 0.98764 | 0.5883932406 | 0.95436 | 0.9774301052 | 0.424856 | 6.090996763 | 0 |
| 17 | 7674256 | T | A | Y | F | TP53 | ENST00000269305 | 0.29512 | 0.09187 | 0.31399 | 0.91202 | 0.76553 | 0.4010931667 | 0.81478 | 0.09490258992 | 0.141824 | 0.5171350184 | 0 |
| 17 | 7674256 | T | A | Y | F | TP53 | ENST00000269305 | 0.29512 | 0.09187 | 0.31399 | 0.91202 | 0.76553 | 0.4010931667 | 0.81478 | 0.09490258992 | 0.141824 | 0.5171350184 | 0 |
| 17 | 7674256 | T | C | Y | C | TP53 | ENST00000269305 | 0.81087 | 0.42944 | 0.41403 | 0.9911 | 0.93977 | 0.7336701627 | 0.97891 | 0.9957493544 | 0.447987 | 5.357038984 | 1 |
| 17 | 7674256 | T | C | Y | C | TP53 | ENST00000269305 | 0.81087 | 0.42944 | 0.41403 | 0.9911 | 0.93977 | 0.7336701627 | 0.97891 | 0.9957493544 | 0.447987 | 5.357038984 | 1 |
| 17 | 7674256 | T | C | Y | C | TP53 | ENST00000269305 | 0.81087 | 0.42944 | 0.41403 | 0.9911 | 0.93977 | 0.7336701627 | 0.97891 | 0.9957493544 | 0.447987 | 5.357038984 | 1 |
| 17 | 7674256 | T | G | Y | S | TP53 | ENST00000269305 | 0.77869 | 0.45012 | 0.34684 | 0.99311 | 0.92763 | 0.8147265129 | 0.97279 | 0.9969935417 | 0.371772 | 6.50451077 | 1 |
| 17 | 7674256 | T | G | Y | S | TP53 | ENST00000269305 | 0.77869 | 0.45012 | 0.34684 | 0.99311 | 0.92763 | 0.8147265129 | 0.97279 | 0.9969935417 | 0.371772 | 6.50451077 | 1 |
| 17 | 7674256 | T | G | Y | S | TP53 | ENST00000269305 | 0.77869 | 0.45012 | 0.34684 | 0.99311 | 0.92763 | 0.8147265129 | 0.97279 | 0.9969935417 | 0.371772 | 6.50451077 | 1 |
| 17 | 7674257 | A | C | Y | D | TP53 | ENST00000269305 | 0.90843 | 0.48645 | 0.39933 | 0.99597 | 0.95243 | 0.8567960592 | 0.9768 | 0.9981405735 | 0.504604 | 5.416082886 | 1 |
| 17 | 7674257 | A | C | Y | D | TP53 | ENST00000269305 | 0.90843 | 0.48645 | 0.39933 | 0.99597 | 0.95243 | 0.8567960592 | 0.9768 | 0.9981405735 | 0.504604 | 5.416082886 | 1 |
| 17 | 7674257 | A | C | Y | D | TP53 | ENST00000269305 | 0.90843 | 0.48645 | 0.39933 | 0.99597 | 0.95243 | 0.8567960592 | 0.9768 | 0.9981405735 | 0.504604 | 5.416082886 | 1 |
| 17 | 7674257 | A | G | Y | H | TP53 | ENST00000269305 | 0.89911 | 0.4336 | 0.40798 | 0.99309 | 0.92438 | 0.691526184 | 0.94367 | 0.9968084693 | 0.287845 | 6.50451077 | 1 |
| 17 | 7674257 | A | G | Y | H | TP53 | ENST00000269305 | 0.89911 | 0.4336 | 0.40798 | 0.99309 | 0.92438 | 0.691526184 | 0.94367 | 0.9968084693 | 0.287845 | 6.50451077 | 1 |
| 17 | 7674257 | A | G | Y | H | TP53 | ENST00000269305 | 0.89911 | 0.4336 | 0.40798 | 0.99309 | 0.92438 | 0.691526184 | 0.94367 | 0.9968084693 | 0.287845 | 6.50451077 | 1 |
| 17 | 7674257 | A | T | Y | N | TP53 | ENST00000269305 | 0.90509 | 0.47271 | 0.41077 | 0.99651 | 0.94949 | 0.8116629122 | 0.96098 | 0.9979302883 | 0.424015 | 6.50451077 | 1 |
| 17 | 7674257 | A | T | Y | N | TP53 | ENST00000269305 | 0.90509 | 0.47271 | 0.41077 | 0.99651 | 0.94949 | 0.8116629122 | 0.96098 | 0.9979302883 | 0.424015 | 6.50451077 | 1 |
| 17 | 7674257 | A | T | Y | N | TP53 | ENST00000269305 | 0.90509 | 0.47271 | 0.41077 | 0.99651 | 0.94949 | 0.8116629122 | 0.96098 | 0.9979302883 | 0.424015 | 6.50451077 | 1 |
| 17 | 7674258 | G | C | N | K | TP53 | ENST00000269305 | 0.49357 | 0.24159 | 0.57203 | 0.96596 | 0.82443 | 0.4429900105 | 0.90286 | 0.9860575795 | 0.146344 | 4.002209452 | 0 |
| 17 | 7674258 | G | C | N | K | TP53 | ENST00000269305 | 0.49357 | 0.24159 | 0.57203 | 0.96596 | 0.82443 | 0.4429900105 | 0.90286 | 0.9860575795 | 0.146344 | 4.002209452 | 0 |
| 17 | 7674258 | G | C | N | K | TP53 | ENST00000269305 | 0.49357 | 0.24159 | 0.57203 | 0.96596 | 0.82443 | 0.4429900105 | 0.90286 | 0.9860575795 | 0.146344 | 4.002209452 | 0 |
| 17 | 7674258 | G | T | N | K | TP53 | ENST00000269305 | 0.48838 | 0.24159 | 0.57203 | 0.96718 | 0.825 | 0.4429900105 | 0.90292 | 0.9940109849 | 0.146327 | 4.002209452 | 0 |
| 17 | 7674258 | G | T | N | K | TP53 | ENST00000269305 | 0.48838 | 0.24159 | 0.57203 | 0.96718 | 0.825 | 0.4429900105 | 0.90292 | 0.9940109849 | 0.146327 | 4.002209452 | 0 |
| 17 | 7674258 | G | T | N | K | TP53 | ENST00000269305 | 0.48838 | 0.24159 | 0.57203 | 0.96718 | 0.825 | 0.4429900105 | 0.90292 | 0.9940109849 | 0.146327 | 4.002209452 | 0 |
| 17 | 7674259 | T | A | N | I | TP53 | ENST00000269305 | 0.90468 | 0.42838 | 0.48892 | 0.98602 | 0.96737 | 0.5374742485 | 0.93453 | 0.9922704697 | 0.253559 | 7.298304009 | 0 |
| 17 | 7674259 | T | A | N | I | TP53 | ENST00000269305 | 0.90468 | 0.42838 | 0.48892 | 0.98602 | 0.96737 | 0.5374742485 | 0.93453 | 0.9922704697 | 0.253559 | 7.298304009 | 0 |
| 17 | 7674259 | T | C | N | S | TP53 | ENST00000269305 | 0.47772 | 0.12131 | 0.42355 | 0.96026 | 0.83997 | 0.240764559 | 0.87869 | 0.121167317 | -0.071271 | 3.776453917 | 0 |
| 17 | 7674259 | T | C | N | S | TP53 | ENST00000269305 | 0.47772 | 0.12131 | 0.42355 | 0.96026 | 0.83997 | 0.240764559 | 0.87869 | 0.121167317 | -0.071271 | 3.776453917 | 0 |
| 17 | 7674259 | T | G | N | T | TP53 | ENST00000269305 | 0.869 | 0.38492 | 0.47191 | 0.98042 | 0.96226 | 0.3412916096 | 0.92652 | 0.9925650358 | 0.254796 | 5.640882178 | 0 |
| 17 | 7674259 | T | G | N | T | TP53 | ENST00000269305 | 0.869 | 0.38492 | 0.47191 | 0.98042 | 0.96226 | 0.3412916096 | 0.92652 | 0.9925650358 | 0.254796 | 5.640882178 | 0 |
| 17 | 7674259 | T | G | N | T | TP53 | ENST00000269305 | 0.869 | 0.38492 | 0.47191 | 0.98042 | 0.96226 | 0.3412916096 | 0.92652 | 0.9925650358 | 0.254796 | 5.640882178 | 0 |
| 17 | 7674260 | T | A | N | Y | TP53 | ENST00000269305 | 0.91101 | 0.42649 | 0.5315 | 0.98455 | 0.9844 | 0.5302834412 | 0.93905 | 0.9971495271 | 0.399646 | 6.721180161 | 0 |
| 17 | 7674260 | T | A | N | Y | TP53 | ENST00000269305 | 0.91101 | 0.42649 | 0.5315 | 0.98455 | 0.9844 | 0.5302834412 | 0.93905 | 0.9971495271 | 0.399646 | 6.721180161 | 0 |
| 17 | 7674260 | T | A | N | Y | TP53 | ENST00000269305 | 0.91101 | 0.42649 | 0.5315 | 0.98455 | 0.9844 | 0.5302834412 | 0.93905 | 0.9971495271 | 0.399646 | 6.721180161 | 0 |
| 17 | 7674260 | T | C | N | D | TP53 | ENST00000269305 | 0.89382 | 0.38919 | 0.54346 | 0.97503 | 0.97698 | 0.4066479201 | 0.93226 | 0.9918147922 | 0.265924 | 8.424139611 | 1 |
| 17 | 7674260 | T | C | N | D | TP53 | ENST00000269305 | 0.89382 | 0.38919 | 0.54346 | 0.97503 | 0.97698 | 0.4066479201 | 0.93226 | 0.9918147922 | 0.265924 | 8.424139611 | 1 |
| 17 | 7674260 | T | C | N | D | TP53 | ENST00000269305 | 0.89382 | 0.38919 | 0.54346 | 0.97503 | 0.97698 | 0.4066479201 | 0.93226 | 0.9918147922 | 0.265924 | 8.424139611 | 1 |
| 17 | 7674260 | T | G | N | H | TP53 | ENST00000269305 | 0.87775 | 0.40208 | 0.49669 | 0.98063 | 0.97327 | 0.400724282 | 0.94297 | 0.9975247337 | 0.27756 | 6.532665429 | 0 |
| 17 | 7674260 | T | G | N | H | TP53 | ENST00000269305 | 0.87775 | 0.40208 | 0.49669 | 0.98063 | 0.97327 | 0.400724282 | 0.94297 | 0.9975247337 | 0.27756 | 6.532665429 | 0 |
| 17 | 7674260 | T | G | N | H | TP53 | ENST00000269305 | 0.87775 | 0.40208 | 0.49669 | 0.98063 | 0.97327 | 0.400724282 | 0.94297 | 0.9975247337 | 0.27756 | 6.532665429 | 0 |
| 17 | 7674262 | T | A | Y | F | TP53 | ENST00000269305 | 0.40969 | 0.63316 | 0.67839 | 0.97829 | 0.88684 | 0.3631931316 | 0.95277 | 0.4129514396 | 0.192759 | 2.615626043 | 0 |
| 17 | 7674262 | T | A | Y | F | TP53 | ENST00000269305 | 0.40969 | 0.63316 | 0.67839 | 0.97829 | 0.88684 | 0.3631931316 | 0.95277 | 0.4129514396 | 0.192759 | 2.615626043 | 0 |
| 17 | 7674262 | T | A | Y | F | TP53 | ENST00000269305 | 0.40969 | 0.63316 | 0.67839 | 0.97829 | 0.88684 | 0.3631931316 | 0.95277 | 0.4129514396 | 0.192759 | 2.615626043 | 0 |
| 17 | 7674262 | T | C | Y | C | TP53 | ENST00000269305 | 0.84165 | 0.92236 | 0.7728 | 0.99138 | 0.98904 | 0.6887327159 | 0.99416 | 0.9955471754 | 0.533688 | 7.566311762 | 1 |
| 17 | 7674262 | T | C | Y | C | TP53 | ENST00000269305 | 0.84165 | 0.92236 | 0.7728 | 0.99138 | 0.98904 | 0.6887327159 | 0.99416 | 0.9955471754 | 0.533688 | 7.566311762 | 1 |
| 17 | 7674262 | T | C | Y | C | TP53 | ENST00000269305 | 0.84165 | 0.92236 | 0.7728 | 0.99138 | 0.98904 | 0.6887327159 | 0.99416 | 0.9955471754 | 0.533688 | 7.566311762 | 1 |
| 17 | 7674262 | T | G | Y | S | TP53 | ENST00000269305 | 0.79414 | 0.92971 | 0.7015 | 0.99397 | 0.98585 | 0.7827264296 | 0.98198 | 0.9979867935 | 0.456489 | 7.566311762 | 1 |
| 17 | 7674262 | T | G | Y | S | TP53 | ENST00000269305 | 0.79414 | 0.92971 | 0.7015 | 0.99397 | 0.98585 | 0.7827264296 | 0.98198 | 0.9979867935 | 0.456489 | 7.566311762 | 1 |
| 17 | 7674262 | T | G | Y | S | TP53 | ENST00000269305 | 0.79414 | 0.92971 | 0.7015 | 0.99397 | 0.98585 | 0.7827264296 | 0.98198 | 0.9979867935 | 0.456489 | 7.566311762 | 1 |
| 17 | 7674263 | A | C | Y | D | TP53 | ENST00000269305 | 0.65194 | 0.88634 | 0.76174 | 0.97771 | 0.96371 | 0.8309765264 | 0.99229 | 0.9956741929 | 0.520495 | 7.566311762 | 1 |
| 17 | 7674263 | A | C | Y | D | TP53 | ENST00000269305 | 0.65194 | 0.88634 | 0.76174 | 0.97771 | 0.96371 | 0.8309765264 | 0.99229 | 0.9956741929 | 0.520495 | 7.566311762 | 1 |
| 17 | 7674263 | A | C | Y | D | TP53 | ENST00000269305 | 0.65194 | 0.88634 | 0.76174 | 0.97771 | 0.96371 | 0.8309765264 | 0.99229 | 0.9956741929 | 0.520495 | 7.566311762 | 1 |
| 17 | 7674263 | A | G | Y | H | TP53 | ENST00000269305 | 0.84188 | 0.92559 | 0.78108 | 0.98298 | 0.96883 | 0.6664266559 | 0.96231 | 0.9951966405 | 0.365702 | 7.566311762 | 1 |
| 17 | 7674263 | A | G | Y | H | TP53 | ENST00000269305 | 0.84188 | 0.92559 | 0.78108 | 0.98298 | 0.96883 | 0.6664266559 | 0.96231 | 0.9951966405 | 0.365702 | 7.566311762 | 1 |
| 17 | 7674263 | A | G | Y | H | TP53 | ENST00000269305 | 0.84188 | 0.92559 | 0.78108 | 0.98298 | 0.96883 | 0.6664266559 | 0.96231 | 0.9951966405 | 0.365702 | 7.566311762 | 1 |
| 17 | 7674263 | A | T | Y | N | TP53 | ENST00000269305 | 0.84317 | 0.94597 | 0.77155 | 0.98905 | 0.98764 | 0.7729218524 | 0.97927 | 0.9968078732 | 0.506211 | 7.566311762 | 1 |
| 17 | 7674263 | A | T | Y | N | TP53 | ENST00000269305 | 0.84317 | 0.94597 | 0.77155 | 0.98905 | 0.98764 | 0.7729218524 | 0.97927 | 0.9968078732 | 0.506211 | 7.566311762 | 1 |
| 17 | 7674263 | A | T | Y | N | TP53 | ENST00000269305 | 0.84317 | 0.94597 | 0.77155 | 0.98905 | 0.98764 | 0.7729218524 | 0.97927 | 0.9968078732 | 0.506211 | 7.566311762 | 1 |
| 17 | 7674264 | G | C | H | Q | TP53 | ENST00000269305 | 0.6946 | 0.89279 | 0.377 | 0.98891 | 0.87877 | 0.4281840553 | 0.91606 | 0.9192498326 | 0.223429 | 8.061454106 | 0 |
| 17 | 7674264 | G | C | H | Q | TP53 | ENST00000269305 | 0.6946 | 0.89279 | 0.377 | 0.98891 | 0.87877 | 0.4281840553 | 0.91606 | 0.9192498326 | 0.223429 | 8.061454106 | 0 |
| 17 | 7674264 | G | T | H | Q | TP53 | ENST00000269305 | 0.68811 | 0.89279 | 0.377 | 0.98925 | 0.87877 | 0.4281840553 | 0.9179 | 0.9320356846 | 0.223456 | 8.061454106 | 0 |
| 17 | 7674264 | G | T | H | Q | TP53 | ENST00000269305 | 0.68811 | 0.89279 | 0.377 | 0.98925 | 0.87877 | 0.4281840553 | 0.9179 | 0.9320356846 | 0.223456 | 8.061454106 | 0 |
| 17 | 7674265 | T | A | H | L | TP53 | ENST00000269305 | 0.39841 | 0.90673 | 0.35638 | 0.94599 | 0.83228 | 0.3764230884 | 0.89461 | 0.9115791917 | 0.202091 | 3.447744669 | 0 |
| 17 | 7674265 | T | A | H | L | TP53 | ENST00000269305 | 0.39841 | 0.90673 | 0.35638 | 0.94599 | 0.83228 | 0.3764230884 | 0.89461 | 0.9115791917 | 0.202091 | 3.447744669 | 0 |
| 17 | 7674265 | T | A | H | L | TP53 | ENST00000269305 | 0.39841 | 0.90673 | 0.35638 | 0.94599 | 0.83228 | 0.3764230884 | 0.89461 | 0.9115791917 | 0.202091 | 3.447744669 | 0 |
| 17 | 7674265 | T | C | H | R | TP53 | ENST00000269305 | 0.77869 | 0.91449 | 0.39847 | 0.98932 | 0.90096 | 0.5389250132 | 0.91324 | 0.9193108678 | 0.318885 | 5.537659827 | 0 |
| 17 | 7674265 | T | C | H | R | TP53 | ENST00000269305 | 0.77869 | 0.91449 | 0.39847 | 0.98932 | 0.90096 | 0.5389250132 | 0.91324 | 0.9193108678 | 0.318885 | 5.537659827 | 0 |
| 17 | 7674265 | T | G | H | P | TP53 | ENST00000269305 | 0.84598 | 0.94484 | 0.35826 | 0.9936 | 0.95207 | 0.7683339221 | 0.96224 | 0.9222623706 | 0.410445 | 6.649072412 | 0 |
| 17 | 7674265 | T | G | H | P | TP53 | ENST00000269305 | 0.84598 | 0.94484 | 0.35826 | 0.9936 | 0.95207 | 0.7683339221 | 0.96224 | 0.9222623706 | 0.410445 | 6.649072412 | 0 |
| 17 | 7674266 | G | A | H | Y | TP53 | ENST00000269305 | 0.71867 | 0.89831 | 0.4101 | 0.9789 | 0.8887 | 0.3955975187 | 0.88098 | 0.8954969645 | 0.249996 | 5.720227897 | 0 |
| 17 | 7674266 | G | A | H | Y | TP53 | ENST00000269305 | 0.71867 | 0.89831 | 0.4101 | 0.9789 | 0.8887 | 0.3955975187 | 0.88098 | 0.8954969645 | 0.249996 | 5.720227897 | 0 |
| 17 | 7674266 | G | A | H | Y | TP53 | ENST00000269305 | 0.71867 | 0.89831 | 0.4101 | 0.9789 | 0.8887 | 0.3955975187 | 0.88098 | 0.8954969645 | 0.249996 | 5.720227897 | 0 |
| 17 | 7674266 | G | C | H | D | TP53 | ENST00000269305 | 0.80187 | 0.9318 | 0.39193 | 0.98664 | 0.96371 | 0.6256099983 | 0.98087 | 0.9700000286 | 0.464211 | 6.074319381 | 0 |
| 17 | 7674266 | G | C | H | D | TP53 | ENST00000269305 | 0.80187 | 0.9318 | 0.39193 | 0.98664 | 0.96371 | 0.6256099983 | 0.98087 | 0.9700000286 | 0.464211 | 6.074319381 | 1 |
| 17 | 7674266 | G | T | H | N | TP53 | ENST00000269305 | 0.76179 | 0.90529 | 0.39892 | 0.98092 | 0.89829 | 0.4300419744 | 0.88871 | 0.9484375715 | 0.224544 | 6.100922105 | 0 |
| 17 | 7674266 | G | T | H | N | TP53 | ENST00000269305 | 0.76179 | 0.90529 | 0.39892 | 0.98092 | 0.89829 | 0.4300419744 | 0.88871 | 0.9484375715 | 0.224544 | 6.100922105 | 0 |
| 17 | 7674266 | G | T | H | N | TP53 | ENST00000269305 | 0.76179 | 0.90529 | 0.39892 | 0.98092 | 0.89829 | 0.4300419744 | 0.88871 | 0.9484375715 | 0.224544 | 6.100922105 | 0 |
| 17 | 7674267 | G | C | I | M | TP53 | ENST00000269305 | 0.65234 | 0.89319 | 0.33838 | 0.98882 | 0.92601 | 0.5009551727 | 0.95296 | 0.9793543816 | 0.372412 | 5.558917264 | 0 |
| 17 | 7674267 | G | C | I | M | TP53 | ENST00000269305 | 0.65234 | 0.89319 | 0.33838 | 0.98882 | 0.92601 | 0.5009551727 | 0.95296 | 0.9793543816 | 0.372412 | 5.558917264 | 0 |
| 17 | 7674268 | A | C | I | S | TP53 | ENST00000269305 | 0.9289 | 0.93568 | 0.40294 | 0.99299 | 0.98549 | 0.8572855369 | 0.98754 | 0.9985045195 | 0.567063 | 6.920059412 | 1 |
| 17 | 7674268 | A | C | I | S | TP53 | ENST00000269305 | 0.9289 | 0.93568 | 0.40294 | 0.99299 | 0.98549 | 0.8572855369 | 0.98754 | 0.9985045195 | 0.567063 | 6.920059412 | 1 |
| 17 | 7674268 | A | C | I | S | TP53 | ENST00000269305 | 0.9289 | 0.93568 | 0.40294 | 0.99299 | 0.98549 | 0.8572855369 | 0.98754 | 0.9985045195 | 0.567063 | 6.920059412 | 1 |
| 17 | 7674268 | A | G | I | T | TP53 | ENST00000269305 | 0.86418 | 0.86069 | 0.51567 | 0.98799 | 0.97997 | 0.6788680006 | 0.99079 | 0.9940000772 | 0.54468 | 6.369258118 | 1 |
| 17 | 7674268 | A | G | I | T | TP53 | ENST00000269305 | 0.86418 | 0.86069 | 0.51567 | 0.98799 | 0.97997 | 0.6788680006 | 0.99079 | 0.9940000772 | 0.54468 | 6.369258118 | 1 |
| 17 | 7674268 | A | G | I | T | TP53 | ENST00000269305 | 0.86418 | 0.86069 | 0.51567 | 0.98799 | 0.97997 | 0.6788680006 | 0.99079 | 0.9940000772 | 0.54468 | 6.369258118 | 1 |
| 17 | 7674268 | A | T | I | N | TP53 | ENST00000269305 | 0.92624 | 0.95547 | 0.47784 | 0.99073 | 0.97848 | 0.8140009584 | 0.94531 | 0.9972500205 | 0.421117 | 5.373980883 | 1 |
| 17 | 7674268 | A | T | I | N | TP53 | ENST00000269305 | 0.92624 | 0.95547 | 0.47784 | 0.99073 | 0.97848 | 0.8140009584 | 0.94531 | 0.9972500205 | 0.421117 | 5.373980883 | 1 |
| 17 | 7674268 | A | T | I | N | TP53 | ENST00000269305 | 0.92624 | 0.95547 | 0.47784 | 0.99073 | 0.97848 | 0.8140009584 | 0.94531 | 0.9972500205 | 0.421117 | 5.373980883 | 1 |
| 17 | 7674269 | T | A | I | F | TP53 | ENST00000269305 | 0.52699 | 0.94186 | 0.38309 | 0.99066 | 0.9506 | 0.6641465031 | 0.98584 | 0.9951661825 | 0.457394 | 6.203803862 | 1 |
| 17 | 7674269 | T | A | I | F | TP53 | ENST00000269305 | 0.52699 | 0.94186 | 0.38309 | 0.99066 | 0.9506 | 0.6641465031 | 0.98584 | 0.9951661825 | 0.457394 | 6.203803862 | 1 |
| 17 | 7674269 | T | A | I | F | TP53 | ENST00000269305 | 0.52699 | 0.94186 | 0.38309 | 0.99066 | 0.9506 | 0.6641465031 | 0.98584 | 0.9951661825 | 0.457394 | 6.203803862 | 1 |
| 17 | 7674269 | T | C | I | V | TP53 | ENST00000269305 | 0.19797 | 0.53909 | 0.28765 | 0.9007 | 0.73148 | 0.2089848398 | 0.84612 | 0.1679874957 | 0.140064 | 2.331767541 | 0 |
| 17 | 7674269 | T | C | I | V | TP53 | ENST00000269305 | 0.19797 | 0.53909 | 0.28765 | 0.9007 | 0.73148 | 0.2089848398 | 0.84612 | 0.1679874957 | 0.140064 | 2.331767541 | 0 |
| 17 | 7674269 | T | C | I | V | TP53 | ENST00000269305 | 0.19797 | 0.53909 | 0.28765 | 0.9007 | 0.73148 | 0.2089848398 | 0.84612 | 0.1679874957 | 0.140064 | 2.331767541 | 0 |
| 17 | 7674269 | T | G | I | L | TP53 | ENST00000269305 | 0.27416 | 0.67927 | 0.37145 | 0.97779 | 0.88776 | 0.4464471874 | 0.95992 | 0.8247543573 | 0.262086 | 3.965010971 | 0 |
| 17 | 7674269 | T | G | I | L | TP53 | ENST00000269305 | 0.27416 | 0.67927 | 0.37145 | 0.97779 | 0.88776 | 0.4464471874 | 0.95992 | 0.8247543573 | 0.262086 | 3.965010971 | 0 |
| 17 | 7674269 | T | G | I | L | TP53 | ENST00000269305 | 0.27416 | 0.67927 | 0.37145 | 0.97779 | 0.88776 | 0.4464471874 | 0.95992 | 0.8247543573 | 0.262086 | 3.965010971 | 0 |
| 17 | 7674271 | G | A | T | I | TP53 | ENST00000269305 | 0.89143 | 0.90247 | 0.37316 | 0.96517 | 0.98799 | 0.6457986102 | 0.95682 | 0.9986492991 | 0.527987 | 5.149082331 | 0 |
| 17 | 7674271 | G | A | T | I | TP53 | ENST00000269305 | 0.89143 | 0.90247 | 0.37316 | 0.96517 | 0.98799 | 0.6457986102 | 0.95682 | 0.9986492991 | 0.527987 | 5.149082331 | 0 |
| 17 | 7674271 | G | A | T | I | TP53 | ENST00000269305 | 0.89143 | 0.90247 | 0.37316 | 0.96517 | 0.98799 | 0.6457986102 | 0.95682 | 0.9986492991 | 0.527987 | 5.149082331 | 0 |

| | | | | | | | | | | | | | | | | |
|---|---|---|---|---|---|---|---|---|---|---|---|---|---|---|---|---|
| 17 | 7674271 | G | C | T | S | TP53 | ENST00000269305 | 0.83583 | 0.82653 | 0.24873 | 0.95761 | 0.97848 | 0.3859515425 | 0.93213 | 0.9966282248 | 0.388367 | 5.826339637 | 0 |
| 17 | 7674271 | G | C | T | S | TP53 | ENST00000269305 | 0.83583 | 0.82653 | 0.24873 | 0.95761 | 0.97848 | 0.3859515425 | 0.93213 | 0.9966282248 | 0.388367 | 5.826339637 | 0 |
| 17 | 7674271 | G | C | T | S | TP53 | ENST00000269305 | 0.83583 | 0.82653 | 0.24873 | 0.95761 | 0.97848 | 0.3859515425 | 0.93213 | 0.9966282248 | 0.388367 | 5.826339637 | 0 |
| 17 | 7674271 | G | T | T | N | TP53 | ENST00000269305 | 0.86269 | 0.91035 | 0.31372 | 0.95949 | 0.967 | 0.6535109955 | 0.93196 | 0.9987271428 | 0.296283 | 6.298835199 | 0 |
| 17 | 7674271 | G | T | T | N | TP53 | ENST00000269305 | 0.86269 | 0.91035 | 0.31372 | 0.95949 | 0.967 | 0.6535109955 | 0.93196 | 0.9987271428 | 0.296283 | 6.298835199 | 0 |
| 17 | 7674272 | T | A | T | S | TP53 | ENST00000269305 | 0.88709 | 0.82653 | 0.24873 | 0.98134 | 0.98071 | 0.3859515425 | 0.94007 | 0.9960716963 | 0.385542 | 5.826339637 | 0 |
| 17 | 7674272 | T | A | T | S | TP53 | ENST00000269305 | 0.88709 | 0.82653 | 0.24873 | 0.98134 | 0.98071 | 0.3859515425 | 0.94007 | 0.9960716963 | 0.385542 | 5.826339637 | 0 |
| 17 | 7674272 | T | C | T | A | TP53 | ENST00000269305 | 0.74611 | 0.63432 | 0.2378 | 0.97414 | 0.97438 | 0.5591280434 | 0.97792 | 0.989788413 | 0.445824 | 4.554559588 | 0 |
| 17 | 7674272 | T | C | T | A | TP53 | ENST00000269305 | 0.74611 | 0.63432 | 0.2378 | 0.97414 | 0.97438 | 0.5591280434 | 0.97792 | 0.989788413 | 0.445824 | 4.554559588 | 0 |
| 17 | 7674272 | T | G | T | P | TP53 | ENST00000269305 | 0.93924 | 0.92508 | 0.24001 | 0.98596 | 0.99356 | 0.7517084552 | 0.97809 | 0.9985830784 | 0.574992 | 5.395500112 | 0 |
| 17 | 7674272 | T | G | T | P | TP53 | ENST00000269305 | 0.93924 | 0.92508 | 0.24001 | 0.98596 | 0.99356 | 0.7517084552 | 0.97809 | 0.9985830784 | 0.574992 | 5.395500112 | 0 |
| 17 | 7674272 | T | G | T | P | TP53 | ENST00000269305 | 0.93924 | 0.92508 | 0.24001 | 0.98596 | 0.99356 | 0.7517084552 | 0.97809 | 0.9985830784 | 0.574992 | 5.395500112 | 1 |
| 17 | 7674274 | G | A | T | I | TP53 | ENST00000269305 | 0.92768 | 0.90234 | 0.57034 | 0.95071 | 0.98585 | 0.5859221799 | 0.98835 | 0.996214807 | 0.542867 | 4.824652985 | 0 |
| 17 | 7674274 | G | A | T | I | TP53 | ENST00000269305 | 0.92768 | 0.90234 | 0.57034 | 0.95071 | 0.98585 | 0.5859221799 | 0.98835 | 0.996214807 | 0.542867 | 4.824652985 | 0 |
| 17 | 7674274 | G | A | T | I | TP53 | ENST00000269305 | 0.92768 | 0.90234 | 0.57034 | 0.95071 | 0.98585 | 0.5859221799 | 0.98835 | 0.996214807 | 0.542867 | 4.824652985 | 0 |
| 17 | 7674274 | G | C | T | S | TP53 | ENST00000269305 | 0.76387 | 0.80419 | 0.44019 | 0.93546 | 0.96044 | 0.3642332303 | 0.93572 | 0.9728785157 | 0.339614 | 4.311039472 | 0 |
| 17 | 7674274 | G | C | T | S | TP53 | ENST00000269305 | 0.76387 | 0.80419 | 0.44019 | 0.93546 | 0.96044 | 0.3642332303 | 0.93572 | 0.9728785157 | 0.339614 | 4.311039472 | 0 |
| 17 | 7674274 | G | C | T | S | TP53 | ENST00000269305 | 0.76387 | 0.80419 | 0.44019 | 0.93546 | 0.96044 | 0.3642332303 | 0.93572 | 0.9728785157 | 0.339614 | 4.311039472 | 0 |
| 17 | 7674274 | G | T | T | N | TP53 | ENST00000269305 | 0.90848 | 0.90904 | 0.51576 | 0.94769 | 0.96334 | 0.6315761337 | 0.94198 | 0.9920193553 | 0.270118 | 7.177781895 | 0 |
| 17 | 7674275 | T | A | T | S | TP53 | ENST00000269305 | 0.65633 | 0.80419 | 0.44019 | 0.9717 | 0.95899 | 0.3642332303 | 0.94859 | 0.9698975682 | 0.330558 | 4.311039472 | 0 |
| 17 | 7674275 | T | A | T | S | TP53 | ENST00000269305 | 0.65633 | 0.80419 | 0.44019 | 0.9717 | 0.95899 | 0.3642332303 | 0.94859 | 0.9698975682 | 0.330558 | 4.311039472 | 0 |
| 17 | 7674275 | T | A | T | S | TP53 | ENST00000269305 | 0.65633 | 0.80419 | 0.44019 | 0.9717 | 0.95899 | 0.3642332303 | 0.94859 | 0.9698975682 | 0.330558 | 4.311039472 | 0 |
| 17 | 7674275 | T | C | T | A | TP53 | ENST00000269305 | 0.66011 | 0.61281 | 0.42247 | 0.96437 | 0.95645 | 0.4953834948 | 0.98772 | 0.978585124 | 0.460141 | 4.644041245 | 0 |
| 17 | 7674275 | T | C | T | A | TP53 | ENST00000269305 | 0.66011 | 0.61281 | 0.42247 | 0.96437 | 0.95645 | 0.4953834948 | 0.98772 | 0.978585124 | 0.460141 | 4.644041245 | 0 |
| 17 | 7674275 | T | C | T | A | TP53 | ENST00000269305 | 0.66011 | 0.61281 | 0.42247 | 0.96437 | 0.95645 | 0.4953834948 | 0.98772 | 0.978585124 | 0.460141 | 4.644041245 | 0 |
| 17 | 7674275 | T | G | T | P | TP53 | ENST00000269305 | 0.88259 | 0.92482 | 0.41818 | 0.98063 | 0.97997 | 0.7616122389 | 0.99448 | 0.9982738495 | 0.547084 | 8.260959427 | 0 |
| 17 | 7674275 | T | G | T | P | TP53 | ENST00000269305 | 0.88259 | 0.92482 | 0.41818 | 0.98063 | 0.97997 | 0.7616122389 | 0.99448 | 0.9982738495 | 0.547084 | 8.260959427 | 1 |
| 17 | 7674276 | A | C | C | W | TP53 | ENST00000269305 | 0.42538 | 0.92971 | 0.34272 | 0.97686 | 0.84052 | 0.6249934829 | 0.95234 | 0.5036587119 | 0.180481 | 5.11331367 | 0 |
| 17 | 7674276 | A | C | C | W | TP53 | ENST00000269305 | 0.42538 | 0.92971 | 0.34272 | 0.97686 | 0.84052 | 0.6249934829 | 0.95234 | 0.5036587119 | 0.180481 | 5.11331367 | 0 |
| 17 | 7674276 | A | C | C | W | TP53 | ENST00000269305 | 0.42538 | 0.92971 | 0.34272 | 0.97686 | 0.84052 | 0.6249934829 | 0.95234 | 0.5036587119 | 0.180481 | 5.11331367 | 0 |
| 17 | 7674277 | C | A | C | F | TP53 | ENST00000269305 | 0.34424 | 0.93087 | 0.29285 | 0.95748 | 0.71783 | 0.5130767953 | 0.87522 | 0.5481108427 | 0.118635 | 4.427881457 | 0 |
| 17 | 7674277 | C | A | C | F | TP53 | ENST00000269305 | 0.34424 | 0.93087 | 0.29285 | 0.95748 | 0.71783 | 0.5130767953 | 0.87522 | 0.5481108427 | 0.118635 | 4.427881457 | 0 |
| 17 | 7674277 | C | A | C | F | TP53 | ENST00000269305 | 0.34424 | 0.93087 | 0.29285 | 0.95748 | 0.71783 | 0.5130767953 | 0.87522 | 0.5481108427 | 0.118635 | 4.427881457 | 0 |
| 17 | 7674277 | C | G | C | S | TP53 | ENST00000269305 | 0.33019 | 0.72229 | 0.28513 | 0.9137 | 0.80713 | 0.4983319284 | 0.8907 | 0.1659323573 | 0.161334 | 4.247997671 | 0 |
| 17 | 7674277 | C | G | C | S | TP53 | ENST00000269305 | 0.33019 | 0.72229 | 0.28513 | 0.9137 | 0.80713 | 0.4983319284 | 0.8907 | 0.1659323573 | 0.161334 | 4.247997671 | 0 |
| 17 | 7674277 | C | G | C | S | TP53 | ENST00000269305 | 0.33019 | 0.72229 | 0.28513 | 0.9137 | 0.80713 | 0.4983319284 | 0.8907 | 0.1659323573 | 0.161334 | 4.247997671 | 0 |
| 17 | 7674277 | C | T | C | Y | TP53 | ENST00000269305 | 0.3005 | 0.80901 | 0.38808 | 0.89347 | 0.70833 | 0.5523440876 | 0.7862 | 0.168099612 | 0.0918238 | 2.358011961 | 0 |
| 17 | 7674277 | C | T | C | Y | TP53 | ENST00000269305 | 0.3005 | 0.80901 | 0.38808 | 0.89347 | 0.70833 | 0.5523440876 | 0.7862 | 0.168099612 | 0.0918238 | 2.358011961 | 0 |
| 17 | 7674277 | C | T | C | Y | TP53 | ENST00000269305 | 0.3005 | 0.80901 | 0.38808 | 0.89347 | 0.70833 | 0.5523440876 | 0.7862 | 0.168099612 | 0.0918238 | 2.358011961 | 0 |
| 17 | 7674278 | A | C | C | G | TP53 | ENST00000269305 | 0.61888 | 0.89749 | 0.26745 | 0.98345 | 0.91015 | 0.5458069638 | 0.94072 | 0.9346579909 | 0.240501 | 4.97321897 | 1 |
| 17 | 7674278 | A | C | C | G | TP53 | ENST00000269305 | 0.61888 | 0.89749 | 0.26745 | 0.98345 | 0.91015 | 0.5458069638 | 0.94072 | 0.9346579909 | 0.240501 | 4.97321897 | 0 |
| 17 | 7674278 | A | C | C | G | TP53 | ENST00000269305 | 0.61888 | 0.89749 | 0.26745 | 0.98345 | 0.91015 | 0.5458069638 | 0.94072 | 0.9346579909 | 0.240501 | 4.97321897 | 0 |
| 17 | 7674278 | A | G | C | R | TP53 | ENST00000269305 | 0.66304 | 0.95374 | 0.37147 | 0.98637 | 0.91694 | 0.6666794921 | 0.94361 | 0.9144310355 | 0.216782 | 6.201318125 | 1 |
| 17 | 7674278 | A | G | C | R | TP53 | ENST00000269305 | 0.66304 | 0.95374 | 0.37147 | 0.98637 | 0.91694 | 0.6666794921 | 0.94361 | 0.9144310355 | 0.216782 | 6.201318125 | 0 |
| 17 | 7674278 | A | T | C | S | TP53 | ENST00000269305 | 0.41943 | 0.72229 | 0.28513 | 0.95291 | 0.84375 | 0.4983319284 | 0.91623 | 0.432444036 | 0.202173 | 4.247997671 | 0 |
| 17 | 7674278 | A | T | C | S | TP53 | ENST00000269305 | 0.41943 | 0.72229 | 0.28513 | 0.95291 | 0.84375 | 0.4983319284 | 0.91623 | 0.432444036 | 0.202173 | 4.247997671 | 0 |
| 17 | 7674279 | G | C | D | E | TP53 | ENST00000269305 | 0.17317 | 0.55563 | 0.22888 | 0.95329 | 0.69254 | 0.1411909874 | 0.73241 | 0.03791374713 | 0.0725677 | 3.302453057 | 0 |
| 17 | 7674279 | G | C | D | E | TP53 | ENST00000269305 | 0.17317 | 0.55563 | 0.22888 | 0.95329 | 0.69254 | 0.1411909874 | 0.73241 | 0.03791374713 | 0.0725677 | 3.302453057 | 0 |
| 17 | 7674279 | G | C | D | E | TP53 | ENST00000269305 | 0.17317 | 0.55563 | 0.22888 | 0.95329 | 0.69254 | 0.1411909874 | 0.73241 | 0.03791374713 | 0.0725677 | 3.302453057 | 0 |
| 17 | 7674279 | G | T | D | E | TP53 | ENST00000269305 | 0.17028 | 0.55563 | 0.22888 | 0.95287 | 0.69339 | 0.1411909874 | 0.73847 | 0.03216172755 | 0.0725642 | 3.302453057 | 0 |
| 17 | 7674279 | G | T | D | E | TP53 | ENST00000269305 | 0.17028 | 0.55563 | 0.22888 | 0.95287 | 0.69339 | 0.1411909874 | 0.73847 | 0.03216172755 | 0.0725642 | 3.302453057 | 0 |
| 17 | 7674279 | G | T | D | E | TP53 | ENST00000269305 | 0.17028 | 0.55563 | 0.22888 | 0.95287 | 0.69339 | 0.1411909874 | 0.73847 | 0.03216172755 | 0.0725642 | 3.302453057 | 0 |
| 17 | 7674280 | T | A | D | V | TP53 | ENST00000269305 | 0.6842 | 0.92569 | 0.20719 | 0.98999 | 0.94655 | 0.4778284142 | 0.96766 | 0.9970265031 | 0.363681 | 5.634075282 | 0 |
| 17 | 7674280 | T | A | D | V | TP53 | ENST00000269305 | 0.6842 | 0.92569 | 0.20719 | 0.98999 | 0.94655 | 0.4778284142 | 0.96766 | 0.9970265031 | 0.363681 | 5.634075282 | 0 |
| 17 | 7674280 | T | A | D | V | TP53 | ENST00000269305 | 0.6842 | 0.92569 | 0.20719 | 0.98999 | 0.94655 | 0.4778284142 | 0.96766 | 0.9970265031 | 0.363681 | 5.634075282 | 0 |
| 17 | 7674280 | T | C | D | G | TP53 | ENST00000269305 | 0.55066 | 0.71774 | 0.22167 | 0.97762 | 0.89513 | 0.4601788505 | 0.97981 | 0.9724898338 | 0.288914 | 4.392534296 | 0 |
| 17 | 7674280 | T | C | D | G | TP53 | ENST00000269305 | 0.55066 | 0.71774 | 0.22167 | 0.97762 | 0.89513 | 0.4601788505 | 0.97981 | 0.9724898338 | 0.288914 | 4.392534296 | 0 |
| 17 | 7674280 | T | C | D | G | TP53 | ENST00000269305 | 0.55066 | 0.71774 | 0.22167 | 0.97762 | 0.89513 | 0.4601788505 | 0.97981 | 0.9724898338 | 0.288914 | 4.392534296 | 0 |
| 17 | 7674280 | T | G | D | A | TP53 | ENST00000269305 | 0.54998 | 0.84334 | 0.23181 | 0.98355 | 0.91862 | 0.3957390023 | 0.93738 | 0.9816460013 | 0.284203 | 4.633831302 | 0 |
| 17 | 7674280 | T | G | D | A | TP53 | ENST00000269305 | 0.54998 | 0.84334 | 0.23181 | 0.98355 | 0.91862 | 0.3957390023 | 0.93738 | 0.9816460013 | 0.284203 | 4.633831302 | 0 |
| 17 | 7674280 | T | G | D | A | TP53 | ENST00000269305 | 0.54998 | 0.84334 | 0.23181 | 0.98355 | 0.91862 | 0.3957390023 | 0.93738 | 0.9816460013 | 0.284203 | 4.633831302 | 0 |
| 17 | 7674281 | C | A | D | Y | TP53 | ENST00000269305 | 0.77585 | 0.91844 | 0.27402 | 0.97748 | 0.95389 | 0.5163056898 | 0.95548 | 0.997901082 | 0.374087 | 7.911457809 | 0 |
| 17 | 7674281 | C | A | D | Y | TP53 | ENST00000269305 | 0.77585 | 0.91844 | 0.27402 | 0.97748 | 0.95389 | 0.5163056898 | 0.95548 | 0.997901082 | 0.374087 | 7.911457809 | 0 |
| 17 | 7674281 | C | G | D | H | TP53 | ENST00000269305 | 0.75877 | 0.90913 | 0.28192 | 0.97174 | 0.92642 | 0.4122957243 | 0.90432 | 0.9884771109 | 0.325564 | 7.015000141 | 0 |
| 17 | 7674281 | C | G | D | H | TP53 | ENST00000269305 | 0.75877 | 0.90913 | 0.28192 | 0.97174 | 0.92642 | 0.4122957243 | 0.90432 | 0.9884771109 | 0.325564 | 7.015000141 | 0 |
| 17 | 7674281 | C | G | D | H | TP53 | ENST00000269305 | 0.75877 | 0.90913 | 0.28192 | 0.97174 | 0.92642 | 0.4122957243 | 0.90432 | 0.9884771109 | 0.325564 | 7.015000141 | 0 |
| 17 | 7674281 | C | T | D | N | TP53 | ENST00000269305 | 0.46059 | 0.61917 | 0.26345 | 0.96114 | 0.81356 | 0.2498365585 | 0.90788 | 0.9510923624 | 0.177758 | 5.382878558 | 0 |
| 17 | 7674281 | C | T | D | N | TP53 | ENST00000269305 | 0.46059 | 0.61917 | 0.26345 | 0.96114 | 0.81356 | 0.2498365585 | 0.90788 | 0.9510923624 | 0.177758 | 5.382878558 | 0 |
| 17 | 7674283 | G | A | S | F | TP53 | ENST00000269305 | 0.72877 | 0.68008 | 0.32188 | 0.96099 | 0.90537 | 0.6542864011 | 0.95766 | 0.9933579564 | 0.276472 | 6.413821236 | 0 |
| 17 | 7674283 | G | A | S | F | TP53 | ENST00000269305 | 0.72877 | 0.68008 | 0.32188 | 0.96099 | 0.90537 | 0.6542864011 | 0.95766 | 0.9933579564 | 0.276472 | 6.413821236 | 0 |
| 17 | 7674283 | G | A | S | F | TP53 | ENST00000269305 | 0.72877 | 0.68008 | 0.32188 | 0.96099 | 0.90537 | 0.6542864011 | 0.95766 | 0.9933579564 | 0.276472 | 6.413821236 | 0 |
| 17 | 7674283 | G | C | S | C | TP53 | ENST00000269305 | 0.69732 | 0.59825 | 0.30344 | 0.95905 | 0.85326 | 0.5912813433 | 0.91633 | 0.9344560504 | 0.221942 | 3.648697107 | 0 |
| 17 | 7674283 | G | C | S | C | TP53 | ENST00000269305 | 0.69732 | 0.59825 | 0.30344 | 0.95905 | 0.85326 | 0.5912813433 | 0.91633 | 0.9344560504 | 0.221942 | 3.648697107 | 0 |
| 17 | 7674283 | G | C | S | C | TP53 | ENST00000269305 | 0.69732 | 0.59825 | 0.30344 | 0.95905 | 0.85326 | 0.5912813433 | 0.91633 | 0.9344560504 | 0.221942 | 3.648697107 | 0 |
| 17 | 7674283 | G | T | S | Y | TP53 | ENST00000269305 | 0.87187 | 0.90195 | 0.33914 | 0.96809 | 0.92438 | 0.6390764726 | 0.95984 | 0.9968292117 | 0.293973 | 7.742240035 | 0 |
| 17 | 7674283 | G | T | S | Y | TP53 | ENST00000269305 | 0.87187 | 0.90195 | 0.33914 | 0.96809 | 0.92438 | 0.6390764726 | 0.95984 | 0.9968292117 | 0.293973 | 7.742240035 | 0 |
| 17 | 7674284 | A | C | S | A | TP53 | ENST00000269305 | 0.21713 | 0.51528 | 0.27618 | 0.95807 | 0.69425 | 0.3665026968 | 0.94005 | 0.1700840145 | 0.124413 | 3.485657645 | 0 |
| 17 | 7674284 | A | C | S | A | TP53 | ENST00000269305 | 0.21713 | 0.51528 | 0.27618 | 0.95807 | 0.69425 | 0.3665026968 | 0.94005 | 0.1700840145 | 0.124413 | 3.485657645 | 0 |
| 17 | 7674284 | A | C | S | A | TP53 | ENST00000269305 | 0.21713 | 0.51528 | 0.27618 | 0.95807 | 0.69425 | 0.3665026968 | 0.94005 | 0.1700840145 | 0.124413 | 3.485657645 | 0 |
| 17 | 7674284 | A | G | S | P | TP53 | ENST00000269305 | 0.24167 | 0.67388 | 0.3002 | 0.96847 | 0.88448 | 0.6989216977 | 0.98902 | 0.6262049079 | 0.314175 | 6.203435342 | 0 |
| 17 | 7674284 | A | G | S | P | TP53 | ENST00000269305 | 0.24167 | 0.67388 | 0.3002 | 0.96847 | 0.88448 | 0.6989216977 | 0.98902 | 0.6262049079 | 0.314175 | 6.203435342 | 0 |
| 17 | 7674284 | A | G | S | P | TP53 | ENST00000269305 | 0.24167 | 0.67388 | 0.3002 | 0.96847 | 0.88448 | 0.6989216977 | 0.98902 | 0.6262049079 | 0.314175 | 6.203435342 | 1 |
| 17 | 7674284 | A | T | S | T | TP53 | ENST00000269305 | 0.18668 | 0.62293 | 0.28859 | 0.90941 | 0.84641 | 0.489779634 | 0.98343 | 0.05895629898 | 0.227452 | 4.168613058 | 0 |
| 17 | 7674284 | A | T | S | T | TP53 | ENST00000269305 | 0.18668 | 0.62293 | 0.28859 | 0.90941 | 0.84641 | 0.489779634 | 0.98343 | 0.05895629898 | 0.227452 | 4.168613058 | 0 |
| 17 | 7674284 | A | T | S | T | TP53 | ENST00000269305 | 0.18668 | 0.62293 | 0.28859 | 0.90941 | 0.84641 | 0.489779634 | 0.98343 | 0.05895629898 | 0.227452 | 4.168613058 | 0 |
| 17 | 7674286 | C | A | G | V | TP53 | ENST00000269305 | 0.69581 | 0.91904 | 0.28487 | 0.97964 | 0.94986 | 0.7995238279 | 0.98142 | 0.9987810254 | 0.420671 | 7.340224108 | 0 |
| 17 | 7674286 | C | A | G | V | TP53 | ENST00000269305 | 0.69581 | 0.91904 | 0.28487 | 0.97964 | 0.94986 | 0.7995238279 | 0.98142 | 0.9987810254 | 0.420671 | 7.340224108 | 0 |
| 17 | 7674286 | C | A | G | V | TP53 | ENST00000269305 | 0.69581 | 0.91904 | 0.28487 | 0.97964 | 0.94986 | 0.7995238279 | 0.98142 | 0.9987810254 | 0.420671 | 7.340224108 | 0 |
| 17 | 7674286 | C | G | G | A | TP53 | ENST00000269305 | 0.52966 | 0.77124 | 0.2832 | 0.97252 | 0.92233 | 0.6929371504 | 0.97569 | 0.9915546775 | 0.340717 | 4.921241263 | 0 |
| 17 | 7674286 | C | G | G | A | TP53 | ENST00000269305 | 0.52966 | 0.77124 | 0.2832 | 0.97252 | 0.92233 | 0.6929371504 | 0.97569 | 0.9915546775 | 0.340717 | 4.921241263 | 0 |
| 17 | 7674286 | C | G | G | A | TP53 | ENST00000269305 | 0.52966 | 0.77124 | 0.2832 | 0.97252 | 0.92233 | 0.6929371504 | 0.97569 | 0.9915546775 | 0.340717 | 4.921241263 | 0 |
| 17 | 7674286 | C | T | G | D | TP53 | ENST00000269305 | 0.55525 | 0.78817 | 0.38634 | 0.97631 | 0.89239 | 0.7172620853 | 0.94594 | 0.9812096953 | 0.26543 | 5.358890818 | 0 |
| 17 | 7674286 | C | T | G | D | TP53 | ENST00000269305 | 0.55525 | 0.78817 | 0.38634 | 0.97631 | 0.89239 | 0.7172620853 | 0.94594 | 0.9812096953 | 0.26543 | 5.358890818 | 0 |
| 17 | 7674286 | C | T | G | D | TP53 | ENST00000269305 | 0.55525 | 0.78817 | 0.38634 | 0.97631 | 0.89239 | 0.7172620853 | 0.94594 | 0.9812096953 | 0.26543 | 5.358890818 | 0 |
| 17 | 7674287 | C | A | G | C | TP53 | ENST00000269305 | 0.57956 | 0.90241 | 0.32149 | 0.95065 | 0.96407 | 0.7689813811 | 0.9736 | 0.9992780089 | 0.452762 | 6.455887522 | 0 |
| 17 | 7674287 | C | A | G | C | TP53 | ENST00000269305 | 0.57956 | 0.90241 | 0.32149 | 0.95065 | 0.96407 | 0.7689813811 | 0.9736 | 0.9992780089 | 0.452762 | 6.455887522 | 0 |
| 17 | 7674287 | C | A | G | C | TP53 | ENST00000269305 | 0.57956 | 0.90241 | 0.32149 | 0.95065 | 0.96407 | 0.7689813811 | 0.9736 | 0.9992780089 | 0.452762 | 6.455887522 | 0 |
| 17 | 7674287 | C | G | G | R | TP53 | ENST00000269305 | 0.55291 | 0.91327 | 0.37781 | 0.94875 | 0.9648 | 0.7545295342 | 0.96023 | 0.9990720749 | 0.406523 | 6.3594482 | 0 |
| 17 | 7674287 | C | G | G | R | TP53 | ENST00000269305 | 0.55291 | 0.91327 | 0.37781 | 0.94875 | 0.9648 | 0.7545295342 | 0.96023 | 0.9990720749 | 0.406523 | 6.3594482 | 0 |
| 17 | 7674287 | C | T | G | S | TP53 | ENST00000269305 | 0.40027 | 0.87751 | 0.27786 | 0.92875 | 0.96008 | 0.6671023042 | 0.95424 | 0.9966452718 | 0.364074 | 4.248370256 | 0 |
| 17 | 7674287 | C | T | G | S | TP53 | ENST00000269305 | 0.40027 | 0.87751 | 0.27786 | 0.92875 | 0.96008 | 0.6671023042 | 0.95424 | 0.9966452718 | 0.364074 | 4.248370256 | 0 |

| | | | | | | | | | | | | | | | | | | |
|---|---|---|---|---|---|---|---|---|---|---|---|---|---|---|---|---|---|---|
| 17 | 7674287 | C | T | G | S | TP53 | ENST00000269305 | 0.40027 | 0.87751 | 0.27786 | 0.92875 | 0.96008 | 0.6671023042 | 0.95424 | 0.9966452718 | 0.364074 | 4.248370256 | 0 |
| 17 | 7674289 | A | C | V | G | TP53 | ENST00000269305 | 0.76387 | 0.8993 | 0.11625 | 0.98138 | 0.86653 | 0.5452310453 | 0.95882 | 0.9992477894 | 0.285468 | 6.559061438 | 0 |
| 17 | 7674289 | A | C | V | G | TP53 | ENST00000269305 | 0.76387 | 0.8993 | 0.11625 | 0.98138 | 0.86653 | 0.5452310453 | 0.95882 | 0.9992477894 | 0.285468 | 6.559061438 | 0 |
| 17 | 7674289 | A | C | V | G | TP53 | ENST00000269305 | 0.76387 | 0.8993 | 0.11625 | 0.98138 | 0.86653 | 0.5452310453 | 0.95882 | 0.9992477894 | 0.285468 | 6.559061438 | 0 |
| 17 | 7674289 | A | G | V | A | TP53 | ENST00000269305 | 0.43786 | 0.68423 | 0.20143 | 0.98 | 0.7528 | 0.343751557 | 0.86852 | 0.8164128065 | 0.169825 | 5.02456599 | 0 |
| 17 | 7674289 | A | G | V | A | TP53 | ENST00000269305 | 0.43786 | 0.68423 | 0.20143 | 0.98 | 0.7528 | 0.343751557 | 0.86852 | 0.8164128065 | 0.169825 | 5.02456599 | 0 |
| 17 | 7674289 | A | G | V | A | TP53 | ENST00000269305 | 0.43786 | 0.68423 | 0.20143 | 0.98 | 0.7528 | 0.343751557 | 0.86852 | 0.8164128065 | 0.169825 | 5.02456599 | 0 |
| 17 | 7674289 | A | T | V | D | TP53 | ENST00000269305 | 0.79096 | 0.91936 | 0.20603 | 0.98426 | 0.86703 | 0.6281460858 | 0.9622 | 0.9994139671 | 0.317358 | 6.880114319 | 0 |
| 17 | 7674289 | A | T | V | D | TP53 | ENST00000269305 | 0.79096 | 0.91936 | 0.20603 | 0.98426 | 0.86703 | 0.6281460858 | 0.9622 | 0.9994139671 | 0.317358 | 6.880114319 | 0 |
| 17 | 7674290 | C | A | V | F | TP53 | ENST00000269305 | 0.8465 | 0.91011 | 0.16666 | 0.98298 | 0.89513 | 0.5171926683 | 0.94083 | 0.9788967967 | 0.250919 | 8.465179731 | 0 |
| 17 | 7674290 | C | A | V | F | TP53 | ENST00000269305 | 0.8465 | 0.91011 | 0.16666 | 0.98298 | 0.89513 | 0.5171926683 | 0.94083 | 0.9788967967 | 0.250919 | 8.465179731 | 0 |
| 17 | 7674290 | C | G | V | L | TP53 | ENST00000269305 | 0.4186 | 0.57491 | 0.1794 | 0.98253 | 0.71705 | 0.286637789 | 0.84131 | 0.08223528415 | 0.112563 | 4.179528126 | 0 |
| 17 | 7674290 | C | G | V | L | TP53 | ENST00000269305 | 0.4186 | 0.57491 | 0.1794 | 0.98253 | 0.71705 | 0.286637789 | 0.84131 | 0.08223528415 | 0.112563 | 4.179528126 | 0 |
| 17 | 7674290 | C | G | V | L | TP53 | ENST00000269305 | 0.4186 | 0.57491 | 0.1794 | 0.98253 | 0.71705 | 0.286637789 | 0.84131 | 0.08223528415 | 0.112563 | 4.179528126 | 0 |
| 17 | 7674290 | C | T | V | I | TP53 | ENST00000269305 | 0.35674 | 0.50209 | 0.08976 | 0.97593 | 0.70752 | 0.1349480817 | 0.87649 | 0.1684433778 | 0.140677 | 6.053133735 | 0 |
| 17 | 7674290 | C | T | V | I | TP53 | ENST00000269305 | 0.35674 | 0.50209 | 0.08976 | 0.97593 | 0.70752 | 0.1349480817 | 0.87649 | 0.1684433778 | 0.140677 | 6.053133735 | 0 |
| 17 | 7674290 | C | T | V | I | TP53 | ENST00000269305 | 0.35674 | 0.50209 | 0.08976 | 0.97593 | 0.70752 | 0.1349480817 | 0.87649 | 0.1684433778 | 0.140677 | 6.053133735 | 0 |
| 17 | 7674859 | C | A | E | D | TP53 | ENST00000269305 | 0.65578 | 0.5836 | 0.25482 | 0.97608 | 0.89193 | 0.2665891596 | 0.94518 | 0.7854799032 | 0.124928 | 6.367667059 | 0 |
| 17 | 7674859 | C | A | E | D | TP53 | ENST00000269305 | 0.65578 | 0.5836 | 0.25482 | 0.97608 | 0.89193 | 0.2665891596 | 0.94518 | 0.7854799032 | 0.124928 | 6.367667059 | 0 |
| 17 | 7674859 | C | A | E | D | TP53 | ENST00000269305 | 0.65578 | 0.5836 | 0.25482 | 0.97608 | 0.89193 | 0.2665891596 | 0.94518 | 0.7854799032 | 0.124928 | 6.367667059 | 0 |
| 17 | 7674859 | C | G | E | D | TP53 | ENST00000269305 | 0.66414 | 0.5836 | 0.25482 | 0.97648 | 0.89193 | 0.2665891596 | 0.94514 | 0.9553804398 | 0.251332 | 6.367667059 | 0 |
| 17 | 7674859 | C | G | E | D | TP53 | ENST00000269305 | 0.66414 | 0.5836 | 0.25482 | 0.97648 | 0.89193 | 0.2665891596 | 0.94514 | 0.9553804398 | 0.251332 | 6.367667059 | 0 |
| 17 | 7674860 | T | A | E | V | TP53 | ENST00000269305 | 0.95882 | 0.91151 | 0.22993 | 0.98751 | 0.9608 | 0.430084279 | 0.94947 | 0.9979683757 | 0.352418 | 6.595043383 | 0 |
| 17 | 7674860 | T | A | E | V | TP53 | ENST00000269305 | 0.95882 | 0.91151 | 0.22993 | 0.98751 | 0.9608 | 0.430084279 | 0.94947 | 0.9979683757 | 0.352418 | 6.595043383 | 0 |
| 17 | 7674860 | T | A | E | V | TP53 | ENST00000269305 | 0.95882 | 0.91151 | 0.22993 | 0.98751 | 0.9608 | 0.430084279 | 0.94947 | 0.9979683757 | 0.352418 | 6.595043383 | 0 |
| 17 | 7674860 | T | C | E | G | TP53 | ENST00000269305 | 0.95936 | 0.91072 | 0.2156 | 0.98401 | 0.96883 | 0.3632117363 | 0.91688 | 0.9973008037 | 0.288856 | 8.523080987 | 0 |
| 17 | 7674860 | T | C | E | G | TP53 | ENST00000269305 | 0.95936 | 0.91072 | 0.2156 | 0.98401 | 0.96883 | 0.3632117363 | 0.91688 | 0.9973008037 | 0.288856 | 8.523080987 | 0 |
| 17 | 7674860 | T | C | E | G | TP53 | ENST00000269305 | 0.95936 | 0.91072 | 0.2156 | 0.98401 | 0.96883 | 0.3632117363 | 0.91688 | 0.9973008037 | 0.288856 | 8.523080987 | 0 |
| 17 | 7674860 | T | G | E | A | TP53 | ENST00000269305 | 0.95788 | 0.89749 | 0.24557 | 0.97917 | 0.94506 | 0.3186753846 | 0.91733 | 0.9935583472 | 0.280673 | 5.831375629 | 0 |
| 17 | 7674860 | T | G | E | A | TP53 | ENST00000269305 | 0.95788 | 0.89749 | 0.24557 | 0.97917 | 0.94506 | 0.3186753846 | 0.91733 | 0.9935583472 | 0.280673 | 5.831375629 | 0 |
| 17 | 7674860 | T | G | E | A | TP53 | ENST00000269305 | 0.95788 | 0.89749 | 0.24557 | 0.97917 | 0.94506 | 0.3186753846 | 0.91733 | 0.9935583472 | 0.280673 | 5.831375629 | 0 |
| 17 | 7674861 | C | G | E | Q | TP53 | ENST00000269305 | 0.23457 | 0.8023 | 0.22105 | 0.9315 | 0.80595 | 0.2577328559 | 0.86128 | 0.8447921872 | 0.161851 | 4.193225748 | 0 |
| 17 | 7674861 | C | G | E | Q | TP53 | ENST00000269305 | 0.23457 | 0.8023 | 0.22105 | 0.9315 | 0.80595 | 0.2577328559 | 0.86128 | 0.8447921872 | 0.161851 | 4.193225748 | 0 |
| 17 | 7674861 | C | G | E | Q | TP53 | ENST00000269305 | 0.23457 | 0.8023 | 0.22105 | 0.9315 | 0.80595 | 0.2577328559 | 0.86128 | 0.8447921872 | 0.161851 | 4.193225748 | 0 |
| 17 | 7674861 | C | T | E | K | TP53 | ENST00000269305 | 0.68915 | 0.89268 | 0.32627 | 0.97274 | 0.89376 | 0.3758080464 | 0.89331 | 0.9794366956 | 0.30296 | 7.547560689 | 0 |
| 17 | 7674861 | C | T | E | K | TP53 | ENST00000269305 | 0.68915 | 0.89268 | 0.32627 | 0.97274 | 0.89376 | 0.3758080464 | 0.89331 | 0.9794366956 | 0.30296 | 7.547560689 | 0 |
| 17 | 7674863 | G | A | P | L | TP53 | ENST00000269305 | 0.81771 | 0.86582 | 0.41406 | 0.92056 | 0.86149 | 0.7460017385 | 0.97351 | 0.9990677238 | 0.270664 | 4.516582644 | 1 |
| 17 | 7674863 | G | A | P | L | TP53 | ENST00000269305 | 0.81771 | 0.86582 | 0.41406 | 0.92056 | 0.86149 | 0.7460017385 | 0.97351 | 0.9990677238 | 0.270664 | 4.516582644 | 0 |
| 17 | 7674863 | G | C | P | R | TP53 | ENST00000269305 | 0.92782 | 0.9038 | 0.41262 | 0.95342 | 0.911 | 0.7887191707 | 0.96623 | 0.9988800883 | 0.417603 | 7.792396548 | 1 |
| 17 | 7674863 | G | T | P | H | TP53 | ENST00000269305 | 0.92559 | 0.91425 | 0.39475 | 0.95121 | 0.90581 | 0.7332878375 | 0.9644 | 0.9989446998 | 0.384756 | 6.886588494 | 1 |
| 17 | 7674863 | G | T | P | H | TP53 | ENST00000269305 | 0.92559 | 0.91425 | 0.39475 | 0.95121 | 0.90581 | 0.7332878375 | 0.9644 | 0.9989446998 | 0.384756 | 6.886588494 | 0 |
| 17 | 7674864 | G | A | P | S | TP53 | ENST00000269305 | 0.73365 | 0.89766 | 0.3391 | 0.9571 | 0.92151 | 0.7190282481 | 0.96331 | 0.9993984699 | 0.383259 | 5.457276833 | 0 |
| 17 | 7674864 | G | A | P | S | TP53 | ENST00000269305 | 0.73365 | 0.89766 | 0.3391 | 0.9571 | 0.92151 | 0.7190282481 | 0.96331 | 0.9993984699 | 0.383259 | 5.457276833 | 0 |
| 17 | 7674864 | G | C | P | A | TP53 | ENST00000269305 | 0.67676 | 0.88625 | 0.32354 | 0.95406 | 0.91904 | 0.6247192052 | 0.95569 | 0.999120295 | 0.277334 | 5.716825086 | 0 |
| 17 | 7674864 | G | C | P | A | TP53 | ENST00000269305 | 0.67676 | 0.88625 | 0.32354 | 0.95406 | 0.91904 | 0.6247192052 | 0.95569 | 0.999120295 | 0.277334 | 5.716825086 | 0 |
| 17 | 7674864 | G | C | P | A | TP53 | ENST00000269305 | 0.67676 | 0.88625 | 0.32354 | 0.95406 | 0.91904 | 0.6247192052 | 0.95569 | 0.999120295 | 0.277334 | 5.716825086 | 0 |
| 17 | 7674864 | G | T | P | T | TP53 | ENST00000269305 | 0.70776 | 0.90935 | 0.37599 | 0.92903 | 0.93004 | 0.7093038872 | 0.95196 | 0.999052465 | 0.37817 | 6.235924718 | 1 |
| 17 | 7674864 | G | T | P | T | TP53 | ENST00000269305 | 0.70776 | 0.90935 | 0.37599 | 0.92903 | 0.93004 | 0.7093038872 | 0.95196 | 0.999052465 | 0.37817 | 6.235924718 | 0 |
| 17 | 7674864 | G | T | P | T | TP53 | ENST00000269305 | 0.70776 | 0.90935 | 0.37599 | 0.92903 | 0.93004 | 0.7093038872 | 0.95196 | 0.999052465 | 0.37817 | 6.235924718 | 0 |
| 17 | 7674866 | G | A | P | L | TP53 | ENST00000269305 | 0.33131 | 0.61393 | 0.19238 | 0.84933 | 0.8356 | 0.3432725866 | 0.97693 | 0.8196352124 | 0.25443 | 4.114365011 | 0 |
| 17 | 7674866 | G | A | P | L | TP53 | ENST00000269305 | 0.33131 | 0.61393 | 0.19238 | 0.84933 | 0.8356 | 0.3432725866 | 0.97693 | 0.8196352124 | 0.25443 | 4.114365011 | 0 |
| 17 | 7674866 | G | A | P | L | TP53 | ENST00000269305 | 0.33131 | 0.61393 | 0.19238 | 0.84933 | 0.8356 | 0.3432725866 | 0.97693 | 0.8196352124 | 0.25443 | 4.114365011 | 0 |
| 17 | 7674866 | G | C | P | R | TP53 | ENST00000269305 | 0.35482 | 0.89572 | 0.19818 | 0.93802 | 0.84482 | 0.3292903183 | 0.94165 | 0.9734208584 | 0.292202 | 4.616719205 | 0 |
| 17 | 7674866 | G | C | P | R | TP53 | ENST00000269305 | 0.35482 | 0.89572 | 0.19818 | 0.93802 | 0.84482 | 0.3292903183 | 0.94165 | 0.9734208584 | 0.292202 | 4.616719205 | 0 |
| 17 | 7674866 | G | C | P | R | TP53 | ENST00000269305 | 0.35482 | 0.89572 | 0.19818 | 0.93802 | 0.84482 | 0.3292903183 | 0.94165 | 0.9734208584 | 0.292202 | 4.616719205 | 0 |
| 17 | 7674866 | G | T | P | Q | TP53 | ENST00000269305 | 0.47564 | 0.87637 | 0.14571 | 0.96419 | 0.85741 | 0.3428510415 | 0.95493 | 0.9837927818 | 0.273639 | 4.878355239 | 0 |
| 17 | 7674866 | G | T | P | Q | TP53 | ENST00000269305 | 0.47564 | 0.87637 | 0.14571 | 0.96419 | 0.85741 | 0.3428510415 | 0.95493 | 0.9837927818 | 0.273639 | 4.878355239 | 0 |
| 17 | 7674866 | G | T | P | Q | TP53 | ENST00000269305 | 0.47564 | 0.87637 | 0.14571 | 0.96419 | 0.85741 | 0.3428510415 | 0.95493 | 0.9837927818 | 0.273639 | 4.878355239 | 0 |
| 17 | 7674867 | G | A | P | S | TP53 | ENST00000269305 | 0.27517 | 0.60047 | 0.13484 | 0.8597 | 0.66922 | 0.3030578472 | 0.89804 | 0.2020139694 | 0.154628 | 3.306534499 | 0 |
| 17 | 7674867 | G | A | P | S | TP53 | ENST00000269305 | 0.27517 | 0.60047 | 0.13484 | 0.8597 | 0.66922 | 0.3030578472 | 0.89804 | 0.2020139694 | 0.154628 | 3.306534499 | 0 |
| 17 | 7674867 | G | A | P | S | TP53 | ENST00000269305 | 0.27517 | 0.60047 | 0.13484 | 0.8597 | 0.66922 | 0.3030578472 | 0.89804 | 0.2020139694 | 0.154628 | 3.306534499 | 0 |
| 17 | 7674867 | G | C | P | A | TP53 | ENST00000269305 | 0.25151 | 0.6433 | 0.13291 | 0.8797 | 0.71312 | 0.2234715687 | 0.92927 | 0.1980649233 | 0.191222 | 4.135490597 | 0 |
| 17 | 7674867 | G | C | P | A | TP53 | ENST00000269305 | 0.25151 | 0.6433 | 0.13291 | 0.8797 | 0.71312 | 0.2234715687 | 0.92927 | 0.1980649233 | 0.191222 | 4.135490597 | 0 |
| 17 | 7674867 | G | T | P | T | TP53 | ENST00000269305 | 0.26091 | 0.8133 | 0.17092 | 0.76512 | 0.80595 | 0.3371916834 | 0.95019 | 0.8622416258 | 0.234075 | 3.669781329 | 0 |
| 17 | 7674867 | G | T | P | T | TP53 | ENST00000269305 | 0.26091 | 0.8133 | 0.17092 | 0.76512 | 0.80595 | 0.3371916834 | 0.95019 | 0.8622416258 | 0.234075 | 3.669781329 | 0 |
| 17 | 7674867 | G | T | P | T | TP53 | ENST00000269305 | 0.26091 | 0.8133 | 0.17092 | 0.76512 | 0.80595 | 0.3371916834 | 0.95019 | 0.8622416258 | 0.234075 | 3.669781329 | 0 |
| 17 | 7674868 | C | A | E | D | TP53 | ENST00000269305 | 0.31862 | 0.57025 | 0.42012 | 0.95009 | 0.87877 | 0.4675029752 | 0.96853 | 0.9725052118 | 0.295891 | 5.260048105 | 0 |
| 17 | 7674868 | C | A | E | D | TP53 | ENST00000269305 | 0.31862 | 0.57025 | 0.42012 | 0.95009 | 0.87877 | 0.4675029752 | 0.96853 | 0.9725052118 | 0.295891 | 5.260048105 | 0 |
| 17 | 7674868 | C | A | E | D | TP53 | ENST00000269305 | 0.31862 | 0.57025 | 0.42012 | 0.95009 | 0.87877 | 0.4675029752 | 0.96853 | 0.9725052118 | 0.295891 | 5.260048105 | 0 |
| 17 | 7674868 | C | G | E | D | TP53 | ENST00000269305 | 0.3242 | 0.57025 | 0.42012 | 0.94929 | 0.87877 | 0.4675029752 | 0.96858 | 0.9740513563 | 0.295907 | 5.260048105 | 0 |
| 17 | 7674868 | C | G | E | D | TP53 | ENST00000269305 | 0.3242 | 0.57025 | 0.42012 | 0.94929 | 0.87877 | 0.4675029752 | 0.96858 | 0.9740513563 | 0.295907 | 5.260048105 | 0 |
| 17 | 7674868 | C | G | E | D | TP53 | ENST00000269305 | 0.3242 | 0.57025 | 0.42012 | 0.94929 | 0.87877 | 0.4675029752 | 0.96858 | 0.9740513563 | 0.295907 | 5.260048105 | 0 |
| 17 | 7674869 | T | A | E | V | TP53 | ENST00000269305 | 0.92229 | 0.90943 | 0.40001 | 0.97885 | 0.9729 | 0.7080863625 | 0.99355 | 0.9979250431 | 0.531919 | 8.382888006 | 0 |
| 17 | 7674869 | T | A | E | V | TP53 | ENST00000269305 | 0.92229 | 0.90943 | 0.40001 | 0.97885 | 0.9729 | 0.7080863625 | 0.99355 | 0.9979250431 | 0.531919 | 8.382888006 | 0 |
| 17 | 7674869 | T | A | E | V | TP53 | ENST00000269305 | 0.92229 | 0.90943 | 0.40001 | 0.97885 | 0.9729 | 0.7080863625 | 0.99355 | 0.9979250431 | 0.531919 | 8.382888006 | 0 |
| 17 | 7674869 | T | C | E | G | TP53 | ENST00000269305 | 0.90452 | 0.7526 | 0.37671 | 0.96297 | 0.94543 | 0.7025807855 | 0.98731 | 0.9919867516 | 0.495452 | 5.545651475 | 0 |
| 17 | 7674869 | T | C | E | G | TP53 | ENST00000269305 | 0.90452 | 0.7526 | 0.37671 | 0.96297 | 0.94543 | 0.7025807855 | 0.98731 | 0.9919867516 | 0.495452 | 5.545651475 | 0 |
| 17 | 7674869 | T | G | E | A | TP53 | ENST00000269305 | 0.90625 | 0.88459 | 0.4207 | 0.96964 | 0.96517 | 0.6443436316 | 0.98729 | 0.9961245656 | 0.490505 | 6.471536621 | 0 |
| 17 | 7674869 | T | G | E | A | TP53 | ENST00000269305 | 0.90625 | 0.88459 | 0.4207 | 0.96964 | 0.96517 | 0.6443436316 | 0.98729 | 0.9961245656 | 0.490505 | 6.471536621 | 0 |
| 17 | 7674869 | T | G | E | A | TP53 | ENST00000269305 | 0.90625 | 0.88459 | 0.4207 | 0.96964 | 0.96517 | 0.6443436316 | 0.98729 | 0.9961245656 | 0.490505 | 6.471536621 | 0 |
| 17 | 7674870 | C | G | E | Q | TP53 | ENST00000269305 | 0.72188 | 0.88057 | 0.41519 | 0.96039 | 0.9506 | 0.5891078884 | 0.96132 | 0.9905416369 | 0.372954 | 5.024474667 | 0 |
| 17 | 7674870 | C | G | E | Q | TP53 | ENST00000269305 | 0.72188 | 0.88057 | 0.41519 | 0.96039 | 0.9506 | 0.5891078884 | 0.96132 | 0.9905416369 | 0.372954 | 5.024474667 | 0 |
| 17 | 7674870 | C | G | E | Q | TP53 | ENST00000269305 | 0.72188 | 0.88057 | 0.41519 | 0.96039 | 0.9506 | 0.5891078884 | 0.96132 | 0.9905416369 | 0.372954 | 5.024474667 | 0 |
| 17 | 7674870 | C | T | E | K | TP53 | ENST00000269305 | 0.79104 | 0.89442 | 0.52548 | 0.96854 | 0.97141 | 0.7075444585 | 0.98715 | 0.9919374585 | 0.514723 | 5.547411151 | 0 |
| 17 | 7674870 | C | T | E | K | TP53 | ENST00000269305 | 0.79104 | 0.89442 | 0.52548 | 0.96854 | 0.97141 | 0.7075444585 | 0.98715 | 0.9919374585 | 0.514723 | 5.547411151 | 0 |
| 17 | 7674870 | C | T | E | K | TP53 | ENST00000269305 | 0.79104 | 0.89442 | 0.52548 | 0.96854 | 0.97141 | 0.7075444585 | 0.98715 | 0.9919374585 | 0.514723 | 5.547411151 | 0 |
| 17 | 7674872 | T | A | Y | F | TP53 | ENST00000269305 | 0.92545 | 0.886 | 0.50434 | 0.97549 | 0.94986 | 0.5317621336 | 0.96469 | 0.996691227 | 0.331693 | 4.707860577 | 0 |
| 17 | 7674872 | T | A | Y | F | TP53 | ENST00000269305 | 0.92545 | 0.886 | 0.50434 | 0.97549 | 0.94986 | 0.5317621336 | 0.96469 | 0.996691227 | 0.331693 | 4.707860577 | 0 |
| 17 | 7674872 | T | A | Y | F | TP53 | ENST00000269305 | 0.92545 | 0.886 | 0.50434 | 0.97549 | 0.94986 | 0.5317621336 | 0.96469 | 0.996691227 | 0.331693 | 4.707860577 | 0 |
| 17 | 7674872 | T | C | Y | C | TP53 | ENST00000269305 | 0.95219 | 0.9234 | 0.61177 | 0.97993 | 0.98585 | 0.8003334684 | 0.98217 | 0.9971929193 | 0.546373 | 8.545484831 | 1 |
| 17 | 7674872 | T | C | Y | C | TP53 | ENST00000269305 | 0.95219 | 0.9234 | 0.61177 | 0.97993 | 0.98585 | 0.8003334684 | 0.98217 | 0.9971929193 | 0.546373 | 8.545484831 | 1 |
| 17 | 7674872 | T | C | Y | C | TP53 | ENST00000269305 | 0.95219 | 0.9234 | 0.61177 | 0.97993 | 0.98585 | 0.8003334684 | 0.98217 | 0.9971929193 | 0.546373 | 8.545484831 | 1 |
| 17 | 7674872 | T | G | Y | S | TP53 | ENST00000269305 | 0.94707 | 0.93697 | 0.52333 | 0.98468 | 0.97923 | 0.8464889908 | 0.99562 | 0.9997316003 | 0.566832 | 8.14179178 | 1 |
| 17 | 7674872 | T | G | Y | S | TP53 | ENST00000269305 | 0.94707 | 0.93697 | 0.52333 | 0.98468 | 0.97923 | 0.8464889908 | 0.99562 | 0.9997316003 | 0.566832 | 8.14179178 | 1 |
| 17 | 7674872 | T | G | Y | S | TP53 | ENST00000269305 | 0.94707 | 0.93697 | 0.52333 | 0.98468 | 0.97923 | 0.8464889908 | 0.99562 | 0.9997316003 | 0.566832 | 8.14179178 | 1 |
| 17 | 7674873 | A | C | Y | D | TP53 | ENST00000269305 | 0.91805 | 0.9568 | 0.60066 | 0.98177 | 0.98513 | 0.8924600639 | 0.99668 | 0.9999278784 | 0.585502 | 8.545484831 | 1 |
| 17 | 7674873 | A | C | Y | D | TP53 | ENST00000269305 | 0.91805 | 0.9568 | 0.60066 | 0.98177 | 0.98513 | 0.8924600639 | 0.99668 | 0.9999278784 | 0.585502 | 8.545484831 | 1 |
| 17 | 7674873 | A | C | Y | D | TP53 | ENST00000269305 | 0.91805 | 0.9568 | 0.60066 | 0.98177 | 0.98513 | 0.8924600639 | 0.99668 | 0.9999278784 | 0.585502 | 8.545484831 | 1 |
| 17 | 7674873 | A | G | Y | H | TP53 | ENST00000269305 | 0.91059 | 0.92711 | 0.61929 | 0.97361 | 0.97587 | 0.7534409753 | 0.97624 | 0.9993275404 | 0.494776 | 5.878046872 | 1 |
| 17 | 7674873 | A | G | Y | H | TP53 | ENST00000269305 | 0.91059 | 0.92711 | 0.61929 | 0.97361 | 0.97587 | 0.7534409753 | 0.97624 | 0.9993275404 | 0.494776 | 5.878046872 | 1 |
| 17 | 7674873 | A | T | Y | N | TP53 | ENST00000269305 | 0.91508 | 0.94997 | 0.60861 | 0.98307 | 0.97624 | 0.8355466094 | 0.9806 | 0.9998053908 | 0.511416 | 8.545484831 | 1 |

| | | | | | | | | | | | | | | | | | | |
|---|---|---|---|---|---|---|---|---|---|---|---|---|---|---|---|---|---|---|
| 17 | 7674873 | A | T | Y | N | TP53 | ENST00000269305 | 0.91508 | 0.94997 | 0.60861 | 0.98307 | 0.97624 | 0.8355466094 | 0.9806 | 0.9998053908 | 0.511416 | 8.545484831 | 1 |
| 17 | 7674873 | A | T | Y | N | TP53 | ENST00000269305 | 0.91508 | 0.94997 | 0.60861 | 0.98307 | 0.97624 | 0.8355466094 | 0.9806 | 0.9998053908 | 0.511416 | 8.545484831 | 1 |
| 17 | 7674875 | G | A | P | L | TP53 | ENST00000269305 | 0.91966 | 0.89976 | 0.71256 | 0.96362 | 0.98972 | 0.7740781373 | 0.99633 | 0.9995616078 | 0.573276 | 4.961597315 | 0 |
| 17 | 7674875 | G | C | P | R | TP53 | ENST00000269305 | 0.91157 | 0.90412 | 0.70615 | 0.95105 | 0.99552 | 0.8346273929 | 0.99253 | 0.9997420907 | 0.588101 | 7.8190432 | 1 |
| 17 | 7674875 | G | C | P | R | TP53 | ENST00000269305 | 0.91157 | 0.90412 | 0.70615 | 0.95105 | 0.99552 | 0.8346273929 | 0.99253 | 0.9997420907 | 0.588101 | 7.8190432 | 0 |
| 17 | 7674875 | G | C | P | R | TP53 | ENST00000269305 | 0.91157 | 0.90412 | 0.70615 | 0.95105 | 0.99552 | 0.8346273929 | 0.99253 | 0.9997420907 | 0.588101 | 7.8190432 | 0 |
| 17 | 7674875 | G | T | P | H | TP53 | ENST00000269305 | 0.91031 | 0.91454 | 0.69386 | 0.95141 | 0.99296 | 0.7638773813 | 0.98889 | 0.9997420907 | 0.588101 | 8.001643256 | 0 |
| 17 | 7674875 | G | T | P | H | TP53 | ENST00000269305 | 0.91031 | 0.91454 | 0.69386 | 0.95141 | 0.99296 | 0.7638773813 | 0.98889 | 0.9997420907 | 0.588101 | 8.001643256 | 0 |
| 17 | 7674875 | G | T | P | H | TP53 | ENST00000269305 | 0.91031 | 0.91454 | 0.69386 | 0.95141 | 0.99296 | 0.7638773813 | 0.98889 | 0.9997420907 | 0.588101 | 8.001643256 | 0 |
| 17 | 7674876 | G | A | P | S | TP53 | ENST00000269305 | 0.89307 | 0.89849 | 0.64636 | 0.9466 | 0.99386 | 0.7347056344 | 0.99534 | 0.9975780845 | 0.578601 | 5.619502275 | 0 |
| 17 | 7674876 | G | C | P | A | TP53 | ENST00000269305 | 0.8507 | 0.89354 | 0.63214 | 0.93051 | 0.98621 | 0.6689425964 | 0.98117 | 0.9990837574 | 0.578016 | 5.920761291 | 0 |
| 17 | 7674876 | G | C | P | A | TP53 | ENST00000269305 | 0.8507 | 0.89354 | 0.63214 | 0.93051 | 0.98621 | 0.6689425964 | 0.98117 | 0.9990837574 | 0.578016 | 5.920761291 | 0 |
| 17 | 7674876 | G | C | P | A | TP53 | ENST00000269305 | 0.8507 | 0.89354 | 0.63214 | 0.93051 | 0.98621 | 0.6689425964 | 0.98117 | 0.9990837574 | 0.578016 | 5.920761291 | 0 |
| 17 | 7674876 | G | T | P | T | TP53 | ENST00000269305 | 0.87268 | 0.91059 | 0.68325 | 0.94851 | 0.99443 | 0.7342602516 | 0.99196 | 0.9994065762 | 0.588196 | 8.001643256 | 0 |
| 17 | 7674878 | A | C | V | G | TP53 | ENST00000269305 | 0.95016 | 0.92792 | 0.37002 | 0.98002 | 0.997 | 0.8811889267 | 0.99409 | 0.9994161129 | 0.584902 | 7.211852798 | 1 |
| 17 | 7674878 | A | C | V | G | TP53 | ENST00000269305 | 0.95016 | 0.92792 | 0.37002 | 0.98002 | 0.997 | 0.8811889267 | 0.99409 | 0.9994161129 | 0.584902 | 7.211852798 | 1 |
| 17 | 7674878 | A | G | V | A | TP53 | ENST00000269305 | 0.94452 | 0.91917 | 0.51347 | 0.97597 | 0.99744 | 0.6203837117 | 0.98304 | 0.9976678491 | 0.536694 | 6.238974161 | 0 |
| 17 | 7674878 | A | G | V | A | TP53 | ENST00000269305 | 0.94452 | 0.91917 | 0.51347 | 0.97597 | 0.99744 | 0.6203837117 | 0.98304 | 0.9976678491 | 0.536694 | 6.238974161 | 0 |
| 17 | 7674878 | A | T | V | E | TP53 | ENST00000269305 | 0.95092 | 0.94947 | 0.4962 | 0.9835 | 0.99806 | 0.8855698645 | 0.99574 | 0.9994977713 | 0.584627 | 8.258755699 | 1 |
| 17 | 7674878 | A | T | V | E | TP53 | ENST00000269305 | 0.95092 | 0.94947 | 0.4962 | 0.9835 | 0.99806 | 0.8855698645 | 0.99574 | 0.9994977713 | 0.584627 | 8.258755699 | 1 |
| 17 | 7674878 | A | T | V | E | TP53 | ENST00000269305 | 0.95092 | 0.94947 | 0.4962 | 0.9835 | 0.99806 | 0.8855698645 | 0.99574 | 0.9994977713 | 0.584627 | 8.258755699 | 1 |
| 17 | 7674879 | C | A | V | L | TP53 | ENST00000269305 | 0.65296 | 0.89498 | 0.48818 | 0.9416 | 0.95935 | 0.5564954623 | 0.97027 | 0.9670380354 | 0.396209 | 4.241382891 | 0 |
| 17 | 7674879 | C | A | V | L | TP53 | ENST00000269305 | 0.65296 | 0.89498 | 0.48818 | 0.9416 | 0.95935 | 0.5564954623 | 0.97027 | 0.9670380354 | 0.396209 | 4.241382891 | 0 |
| 17 | 7674879 | C | G | V | L | TP53 | ENST00000269305 | 0.6613 | 0.89498 | 0.48818 | 0.93855 | 0.95935 | 0.5564954623 | 0.96986 | 0.9617013335 | 0.396219 | 4.241382891 | 0 |
| 17 | 7674879 | C | G | V | L | TP53 | ENST00000269305 | 0.6613 | 0.89498 | 0.48818 | 0.93855 | 0.95935 | 0.5564954623 | 0.96986 | 0.9617013335 | 0.396219 | 4.241382891 | 0 |
| 17 | 7674879 | C | G | V | L | TP53 | ENST00000269305 | 0.6613 | 0.89498 | 0.48818 | 0.93855 | 0.95935 | 0.5564954623 | 0.96986 | 0.9617013335 | 0.396219 | 4.241382891 | 0 |
| 17 | 7674879 | C | T | V | M | TP53 | ENST00000269305 | 0.72308 | 0.89766 | 0.52729 | 0.95407 | 0.96262 | 0.6391558901 | 0.9729 | 0.9942829609 | 0.41 | 5.54274953 | 1 |
| 17 | 7674879 | C | T | V | M | TP53 | ENST00000269305 | 0.72308 | 0.89766 | 0.52729 | 0.95407 | 0.96262 | 0.6391558901 | 0.9729 | 0.9942829609 | 0.41 | 5.54274953 | 0 |
| 17 | 7674879 | C | T | V | M | TP53 | ENST00000269305 | 0.72308 | 0.89766 | 0.52729 | 0.95407 | 0.96262 | 0.6391558901 | 0.9729 | 0.9942829609 | 0.41 | 5.54274953 | 0 |
| 17 | 7674881 | A | C | V | G | TP53 | ENST00000269305 | 0.36153 | 0.92355 | 0.13834 | 0.99075 | 0.90711 | 0.6911910659 | 0.97842 | 0.9911351204 | 0.329911 | 4.915232773 | 1 |
| 17 | 7674881 | A | G | V | A | TP53 | ENST00000269305 | 0.58531 | 0.90852 | 0.23173 | 0.98797 | 0.86803 | 0.4011806679 | 0.95961 | 0.823267762 | 0.208085 | 4.748784018 | 0 |
| 17 | 7674881 | A | G | V | A | TP53 | ENST00000269305 | 0.58531 | 0.90852 | 0.23173 | 0.98797 | 0.86803 | 0.4011806679 | 0.95961 | 0.823267762 | 0.208085 | 4.748784018 | 0 |
| 17 | 7674882 | C | A | V | L | TP53 | ENST00000269305 | 0.23389 | 0.61413 | 0.21706 | 0.92978 | 0.8124 | 0.3478113681 | 0.91839 | 0.2768159509 | 0.146338 | 3.279992371 | 0 |
| 17 | 7674882 | C | A | V | L | TP53 | ENST00000269305 | 0.23389 | 0.61413 | 0.21706 | 0.92978 | 0.8124 | 0.3478113681 | 0.91839 | 0.2768159509 | 0.146338 | 3.279992371 | 0 |
| 17 | 7674882 | C | G | V | L | TP53 | ENST00000269305 | 0.2384 | 0.61413 | 0.21706 | 0.9272 | 0.8124 | 0.3478113681 | 0.91837 | 0.2257343531 | 0.146336 | 3.279992371 | 0 |
| 17 | 7674882 | C | T | V | M | TP53 | ENST00000269305 | 0.49151 | 0.89143 | 0.23997 | 0.9618 | 0.75417 | 0.3607688787 | 0.93748 | 0.8717166781 | 0.0350284 | 5.041502843 | 0 |
| 17 | 7674884 | A | C | V | G | TP53 | ENST00000269305 | 0.84605 | 0.92792 | 0.48646 | 0.97773 | 0.9986 | 0.8957952157 | 0.99655 | 0.9990986586 | 0.576068 | 8.030851319 | 1 |
| 17 | 7674884 | A | C | V | G | TP53 | ENST00000269305 | 0.84605 | 0.92792 | 0.48646 | 0.97773 | 0.9986 | 0.8957952157 | 0.99655 | 0.9990986586 | 0.576068 | 8.030851319 | 1 |
| 17 | 7674884 | A | C | V | G | TP53 | ENST00000269305 | 0.84605 | 0.92792 | 0.48646 | 0.97773 | 0.9986 | 0.8957952157 | 0.99655 | 0.9990986586 | 0.576068 | 8.030851319 | 1 |
| 17 | 7674884 | A | G | V | A | TP53 | ENST00000269305 | 0.81347 | 0.91917 | 0.62679 | 0.97309 | 0.99414 | 0.646837442 | 0.98987 | 0.9984121323 | 0.566936 | 5.830284797 | 0 |
| 17 | 7674884 | A | G | V | A | TP53 | ENST00000269305 | 0.81347 | 0.91917 | 0.62679 | 0.97309 | 0.99414 | 0.646837442 | 0.98987 | 0.9984121323 | 0.566936 | 5.830284797 | 0 |
| 17 | 7674884 | A | T | V | E | TP53 | ENST00000269305 | 0.85197 | 0.94947 | 0.61089 | 0.97863 | 0.99892 | 0.9093197899 | 0.99804 | 0.9994630218 | 0.585406 | 8.030851319 | 1 |
| 17 | 7674884 | A | T | V | E | TP53 | ENST00000269305 | 0.85197 | 0.94947 | 0.61089 | 0.97863 | 0.99892 | 0.9093197899 | 0.99804 | 0.9994630218 | 0.585406 | 8.030851319 | 1 |
| 17 | 7674884 | A | T | V | E | TP53 | ENST00000269305 | 0.85197 | 0.94947 | 0.61089 | 0.97863 | 0.99892 | 0.9093197899 | 0.99804 | 0.9994630218 | 0.585406 | 8.030851319 | 1 |
| 17 | 7674885 | C | A | V | L | TP53 | ENST00000269305 | 0.6514 | 0.89481 | 0.61967 | 0.9461 | 0.99006 | 0.6275933991 | 0.99111 | 0.9888322353 | 0.551947 | 4.70237639 | 1 |
| 17 | 7674885 | C | A | V | L | TP53 | ENST00000269305 | 0.6514 | 0.89481 | 0.61967 | 0.9461 | 0.99006 | 0.6275933991 | 0.99111 | 0.9888322353 | 0.551947 | 4.70237639 | 1 |
| 17 | 7674885 | C | A | V | L | TP53 | ENST00000269305 | 0.6514 | 0.89481 | 0.61967 | 0.9461 | 0.99006 | 0.6275933991 | 0.99111 | 0.9888322353 | 0.551947 | 4.70237639 | 1 |
| 17 | 7674885 | C | G | V | L | TP53 | ENST00000269305 | 0.65972 | 0.89481 | 0.61967 | 0.94501 | 0.99006 | 0.6275933991 | 0.9911 | 0.9888322353 | 0.551947 | 4.70237639 | 1 |
| 17 | 7674885 | C | G | V | L | TP53 | ENST00000269305 | 0.65972 | 0.89481 | 0.61967 | 0.94501 | 0.99006 | 0.6275933991 | 0.9911 | 0.9888322353 | 0.551947 | 4.70237639 | 1 |
| 17 | 7674885 | C | G | V | L | TP53 | ENST00000269305 | 0.65972 | 0.89481 | 0.61967 | 0.94501 | 0.99006 | 0.6275933991 | 0.9911 | 0.9888322353 | 0.551947 | 4.70237639 | 1 |
| 17 | 7674885 | C | T | V | M | TP53 | ENST00000269305 | 0.68763 | 0.89766 | 0.65403 | 0.95334 | 0.99265 | 0.6947806512 | 0.9827 | 0.9970505238 | 0.54851 | 8.030851319 | 1 |
| 17 | 7674885 | C | T | V | M | TP53 | ENST00000269305 | 0.68763 | 0.89766 | 0.65403 | 0.95334 | 0.99265 | 0.6947806512 | 0.9827 | 0.9970505238 | 0.54851 | 8.030851319 | 1 |
| 17 | 7674885 | C | T | V | M | TP53 | ENST00000269305 | 0.68763 | 0.89766 | 0.65403 | 0.95334 | 0.99265 | 0.6947806512 | 0.9827 | 0.9970505238 | 0.54851 | 8.030851319 | 1 |
| 17 | 7674886 | A | C | S | R | TP53 | ENST00000269305 | 0.47551 | 0.91086 | 0.56941 | 0.98544 | 0.94015 | 0.8950592916 | 0.97872 | 0.9984602928 | 0.417681 | 7.830788214 | 1 |
| 17 | 7674886 | A | C | S | R | TP53 | ENST00000269305 | 0.47551 | 0.91086 | 0.56941 | 0.98544 | 0.94015 | 0.8950592916 | 0.97872 | 0.9984602928 | 0.417681 | 7.830788214 | 1 |
| 17 | 7674886 | A | C | S | R | TP53 | ENST00000269305 | 0.47551 | 0.91086 | 0.56941 | 0.98544 | 0.94015 | 0.8950592916 | 0.97872 | 0.9984602928 | 0.417681 | 7.830788214 | 1 |
| 17 | 7674886 | A | T | S | R | TP53 | ENST00000269305 | 0.47024 | 0.91086 | 0.56941 | 0.98544 | 0.94015 | 0.8950592916 | 0.97856 | 0.9984602928 | 0.41768 | 7.830788214 | 1 |
| 17 | 7674886 | A | T | S | R | TP53 | ENST00000269305 | 0.47024 | 0.91086 | 0.56941 | 0.98544 | 0.94015 | 0.8950592916 | 0.97856 | 0.9984602928 | 0.41768 | 7.830788214 | 1 |
| 17 | 7674886 | A | T | S | R | TP53 | ENST00000269305 | 0.47024 | 0.91086 | 0.56941 | 0.98544 | 0.94015 | 0.8950592916 | 0.97856 | 0.9984602928 | 0.41768 | 7.830788214 | 1 |
| 17 | 7674887 | C | A | S | I | TP53 | ENST00000269305 | 0.80874 | 0.91723 | 0.52304 | 0.96573 | 0.97735 | 0.7964408376 | 0.99103 | 0.9989297986 | 0.563937 | 7.830788214 | 1 |
| 17 | 7674887 | C | A | S | I | TP53 | ENST00000269305 | 0.80874 | 0.91723 | 0.52304 | 0.96573 | 0.97735 | 0.7964408376 | 0.99103 | 0.9989297986 | 0.563937 | 7.830788214 | 1 |
| 17 | 7674887 | C | A | S | I | TP53 | ENST00000269305 | 0.80874 | 0.91723 | 0.52304 | 0.96573 | 0.97735 | 0.7964408376 | 0.99103 | 0.9989297986 | 0.563937 | 7.830788214 | 1 |
| 17 | 7674887 | C | G | S | T | TP53 | ENST00000269305 | 0.73901 | 0.88544 | 0.49698 | 0.95458 | 0.95499 | 0.6163586862 | 0.94624 | 0.9938447536 | 0.395759 | 7.830788214 | 1 |
| 17 | 7674887 | C | G | S | T | TP53 | ENST00000269305 | 0.73901 | 0.88544 | 0.49698 | 0.95458 | 0.95499 | 0.6163586862 | 0.94624 | 0.9938447536 | 0.395759 | 7.830788214 | 0 |
| 17 | 7674887 | C | G | S | T | TP53 | ENST00000269305 | 0.73901 | 0.88544 | 0.49698 | 0.95458 | 0.95499 | 0.6163586862 | 0.94624 | 0.9938447536 | 0.395759 | 7.830788214 | 0 |
| 17 | 7674887 | C | T | S | N | TP53 | ENST00000269305 | 0.74746 | 0.88829 | 0.55663 | 0.95199 | 0.92964 | 0.770738813 | 0.94014 | 0.9949259758 | 0.2567 | 5.139359696 | 1 |
| 17 | 7674888 | T | A | S | C | TP53 | ENST00000269305 | 0.76156 | 0.89365 | 0.50707 | 0.98453 | 0.97327 | 0.7218276653 | 0.98212 | 0.9989381433 | 0.492254 | 4.400884385 | 0 |
| 17 | 7674888 | T | A | S | C | TP53 | ENST00000269305 | 0.76156 | 0.89365 | 0.50707 | 0.98453 | 0.97327 | 0.7218276653 | 0.98212 | 0.9989381433 | 0.492254 | 4.400884385 | 0 |
| 17 | 7674888 | T | C | S | G | TP53 | ENST00000269305 | 0.72853 | 0.88625 | 0.4359 | 0.97647 | 0.97104 | 0.6967994711 | 0.97677 | 0.9948195815 | 0.389015 | 7.830788214 | 1 |
| 17 | 7674888 | T | C | S | G | TP53 | ENST00000269305 | 0.72853 | 0.88625 | 0.4359 | 0.97647 | 0.97104 | 0.6967994711 | 0.97677 | 0.9948195815 | 0.389015 | 7.830788214 | 1 |
| 17 | 7674888 | T | C | S | G | TP53 | ENST00000269305 | 0.72853 | 0.88625 | 0.4359 | 0.97647 | 0.97104 | 0.6967994711 | 0.97677 | 0.9948195815 | 0.389015 | 7.830788214 | 1 |
| 17 | 7674888 | T | G | S | R | TP53 | ENST00000269305 | 0.75591 | 0.91086 | 0.56941 | 0.98024 | 0.97885 | 0.8950592916 | 0.9798 | 0.9993194342 | 0.500989 | 7.830788214 | 1 |
| 17 | 7674888 | T | G | S | R | TP53 | ENST00000269305 | 0.75591 | 0.91086 | 0.56941 | 0.98024 | 0.97885 | 0.8950592916 | 0.9798 | 0.9993194342 | 0.500989 | 7.830788214 | 1 |
| 17 | 7674888 | T | G | S | R | TP53 | ENST00000269305 | 0.75591 | 0.91086 | 0.56941 | 0.98024 | 0.97885 | 0.8950592916 | 0.9798 | 0.9993194342 | 0.500989 | 7.830788214 | 1 |
| 17 | 7674889 | A | C | H | Q | TP53 | ENST00000269305 | 0.15681 | 0.71867 | 0.30358 | 0.91138 | 0.81588 | 0.4346936921 | 0.93032 | 0.1247367486 | -0.0482403 | 3.70688694 | 0 |
| 17 | 7674889 | A | C | H | Q | TP53 | ENST00000269305 | 0.15681 | 0.71867 | 0.30358 | 0.91138 | 0.81588 | 0.4346936921 | 0.93032 | 0.1247367486 | -0.0482403 | 3.70688694 | 0 |
| 17 | 7674889 | A | C | H | Q | TP53 | ENST00000269305 | 0.15681 | 0.71867 | 0.30358 | 0.91138 | 0.81588 | 0.4346936921 | 0.93032 | 0.1247367486 | -0.0482403 | 3.70688694 | 0 |
| 17 | 7674889 | A | T | H | Q | TP53 | ENST00000269305 | 0.15392 | 0.71867 | 0.30358 | 0.91138 | 0.81588 | 0.4346936921 | 0.93182 | 0.2053401619 | 0.129342 | 3.70688694 | 0 |
| 17 | 7674889 | A | T | H | Q | TP53 | ENST00000269305 | 0.15392 | 0.71867 | 0.30358 | 0.91138 | 0.81588 | 0.4346936921 | 0.93182 | 0.2053401619 | 0.129342 | 3.70688694 | 0 |
| 17 | 7674889 | A | T | H | Q | TP53 | ENST00000269305 | 0.15392 | 0.71867 | 0.30358 | 0.91138 | 0.81588 | 0.4346936921 | 0.93182 | 0.2053401619 | 0.129342 | 3.70688694 | 0 |
| 17 | 7674890 | T | A | H | L | TP53 | ENST00000269305 | 0.78622 | 0.91469 | 0.29361 | 0.98602 | 0.96117 | 0.5660199091 | 0.9733 | 0.9909523726 | 0.390539 | 5.731562913 | 1 |
| 17 | 7674890 | T | C | H | R | TP53 | ENST00000269305 | 0.73141 | 0.90959 | 0.33395 | 0.97919 | 0.967 | 0.6484827226 | 0.98591 | 0.9442492127 | 0.451423 | 6.750824022 | 1 |
| 17 | 7674890 | T | C | H | R | TP53 | ENST00000269305 | 0.73141 | 0.90959 | 0.33395 | 0.97919 | 0.967 | 0.6484827226 | 0.98591 | 0.9442492127 | 0.451423 | 6.750824022 | 1 |
| 17 | 7674890 | T | G | H | P | TP53 | ENST00000269305 | 0.78622 | 0.94498 | 0.27811 | 0.98258 | 0.9844 | 0.8104140446 | 0.97857 | 0.9969773293 | 0.466454 | 8.90840985 | 1 |
| 17 | 7674890 | T | G | H | P | TP53 | ENST00000269305 | 0.78622 | 0.94498 | 0.27811 | 0.98258 | 0.9844 | 0.8104140446 | 0.97857 | 0.9969773293 | 0.466454 | 8.90840985 | 1 |
| 17 | 7674891 | G | A | H | Y | TP53 | ENST00000269305 | 0.73581 | 0.91275 | 0.33112 | 0.974 | 0.90929 | 0.4867559417 | 0.95818 | 0.9910234213 | 0.296918 | 6.500355744 | 0 |
| 17 | 7674891 | G | A | H | Y | TP53 | ENST00000269305 | 0.73581 | 0.91275 | 0.33112 | 0.974 | 0.90929 | 0.4867559417 | 0.95818 | 0.9910234213 | 0.296918 | 6.500355744 | 0 |
| 17 | 7674891 | G | C | H | D | TP53 | ENST00000269305 | 0.77956 | 0.92909 | 0.32776 | 0.97367 | 0.95207 | 0.7162808165 | 0.97235 | 0.9891036749 | 0.413615 | 8.90840985 | 1 |
| 17 | 7674891 | G | T | H | N | TP53 | ENST00000269305 | 0.74547 | 0.90455 | 0.31869 | 0.97136 | 0.90885 | 0.5097666104 | 0.95453 | 0.9800041318 | 0.269751 | 7.583318163 | 1 |
| 17 | 7674891 | G | T | H | N | TP53 | ENST00000269305 | 0.74547 | 0.90455 | 0.31869 | 0.97136 | 0.90885 | 0.5097666104 | 0.95453 | 0.9800041318 | 0.269751 | 7.583318163 | 0 |
| 17 | 7674893 | C | A | R | L | TP53 | ENST00000269305 | 0.87126 | 0.92269 | 0.38516 | 0.96501 | 0.9833 | 0.8385167323 | 0.9951 | 0.9983338714 | 0.57366 | 8.463795278 | 1 |
| 17 | 7674893 | C | A | R | L | TP53 | ENST00000269305 | 0.87126 | 0.92269 | 0.38516 | 0.96501 | 0.9833 | 0.8385167323 | 0.9951 | 0.9983338714 | 0.57366 | 8.463795278 | 1 |
| 17 | 7674893 | C | G | R | P | TP53 | ENST00000269305 | 0.88916 | 0.9383 | 0.40823 | 0.96943 | 0.99105 | 0.9242619958 | 0.9975 | 0.9976927042 | 0.573851 | 8.463795278 | 1 |
| 17 | 7674893 | C | G | R | P | TP53 | ENST00000269305 | 0.88916 | 0.9383 | 0.40823 | 0.96943 | 0.99105 | 0.9242619958 | 0.9975 | 0.9976927042 | 0.573851 | 8.463795278 | 1 |
| 17 | 7674893 | C | T | R | Q | TP53 | ENST00000269305 | 0.87892 | 0.91415 | 0.30228 | 0.96289 | 0.97997 | 0.7874391668 | 0.98913 | 0.9905337095 | 0.515915 | 6.622224816 | 1 |
| 17 | 7674894 | G | C | R | G | TP53 | ENST00000269305 | 0.52672 | 0.9263 | 0.36002 | 0.96946 | 0.9781 | 0.8867237395 | 0.99801 | 0.9978598952 | 0.528671 | 8.463795278 | 1 |
| 17 | 7674894 | G | C | R | G | TP53 | ENST00000269305 | 0.52672 | 0.9263 | 0.36002 | 0.96946 | 0.9781 | 0.8867237395 | 0.99801 | 0.9978598952 | 0.528671 | 8.463795278 | 1 |
| 17 | 7674894 | G | C | R | G | TP53 | ENST00000269305 | 0.52672 | 0.9263 | 0.36002 | 0.96946 | 0.9781 | 0.8867237395 | 0.99801 | 0.9978598952 | 0.528671 | 8.463795278 | 1 |
| 17 | 7674895 | A | C | F | L | TP53 | ENST00000269305 | 0.13477 | 0.65049 | 0.2843 | 0.78303 | 0.77782 | 0.3080230625 | 0.98549 | 0.1074723676 | 0.130533 | 2.480066794 | 0 |
| 17 | 7674895 | A | C | F | L | TP53 | ENST00000269305 | 0.13477 | 0.65049 | 0.2843 | 0.78303 | 0.77782 | 0.3080230625 | 0.98549 | 0.1074723676 | 0.130533 | 2.480066794 | 0 |
| 17 | 7674895 | A | C | F | L | TP53 | ENST00000269305 | 0.13477 | 0.65049 | 0.2843 | 0.78303 | 0.77782 | 0.3080230625 | 0.98549 | 0.1074723676 | 0.130533 | 2.480066794 | 0 |

| | | | | | | | | | | | | | | | | | | |
|---|---|---|---|---|---|---|---|---|---|---|---|---|---|---|---|---|---|---|
| 17 | 7674895 | A | T | F | L | TP53 | ENST00000269305 | 0.13204 | 0.65049 | 0.2843 | 0.78203 | 0.77909 | 0.3080230625 | 0.98555 | 0.1074723676 | 0.130539 | 2.480066794 | 0 |
| 17 | 7674895 | A | T | F | L | TP53 | ENST00000269305 | 0.13204 | 0.65049 | 0.2843 | 0.78203 | 0.77909 | 0.3080230625 | 0.98555 | 0.1074723676 | 0.130539 | 2.480066794 | 0 |
| 17 | 7674896 | A | C | F | C | TP53 | ENST00000269305 | 0.37143 | 0.93532 | 0.281 | 0.97628 | 0.86803 | 0.5185970384 | 0.97178 | 0.5550843477 | 0.22661 | 7.636715839 | 0 |
| 17 | 7674896 | A | C | F | C | TP53 | ENST00000269305 | 0.37143 | 0.93532 | 0.281 | 0.97628 | 0.86803 | 0.5185970384 | 0.97178 | 0.5550843477 | 0.22661 | 7.636715839 | 0 |
| 17 | 7674896 | A | C | F | C | TP53 | ENST00000269305 | 0.37143 | 0.93532 | 0.281 | 0.97628 | 0.86803 | 0.5185970384 | 0.97178 | 0.5550843477 | 0.22661 | 7.636715839 | 0 |
| 17 | 7674896 | A | G | F | S | TP53 | ENST00000269305 | 0.34734 | 0.94555 | 0.25182 | 0.93081 | 0.81818 | 0.3846062777 | 0.96159 | 0.4372469783 | 0.175066 | 5.293866356 | 0 |
| 17 | 7674896 | A | G | F | S | TP53 | ENST00000269305 | 0.34734 | 0.94555 | 0.25182 | 0.93081 | 0.81818 | 0.3846062777 | 0.96159 | 0.4372469783 | 0.175066 | 5.293866356 | 0 |
| 17 | 7674896 | A | G | F | S | TP53 | ENST00000269305 | 0.34734 | 0.94555 | 0.25182 | 0.93081 | 0.81818 | 0.3846062777 | 0.96159 | 0.4372469783 | 0.175066 | 5.293866356 | 0 |
| 17 | 7674896 | A | T | F | Y | TP53 | ENST00000269305 | 0.24458 | 0.87683 | 0.27475 | 0.95662 | 0.71312 | 0.2717927929 | 0.93791 | 0.3169496655 | 0.147591 | 5.918757534 | 0 |
| 17 | 7674896 | A | T | F | Y | TP53 | ENST00000269305 | 0.24458 | 0.87683 | 0.27475 | 0.95662 | 0.71312 | 0.2717927929 | 0.93791 | 0.3169496655 | 0.147591 | 5.918757534 | 0 |
| 17 | 7674896 | A | T | F | Y | TP53 | ENST00000269305 | 0.24458 | 0.87683 | 0.27475 | 0.95662 | 0.71312 | 0.2717927929 | 0.93791 | 0.3169496655 | 0.147591 | 5.918757534 | 0 |
| 17 | 7674897 | A | C | F | V | TP53 | ENST00000269305 | 0.21193 | 0.87994 | 0.23348 | 0.94276 | 0.7675 | 0.4539688851 | 0.95013 | 0.2255321834 | 0.165315 | 7.636715839 | 1 |
| 17 | 7674897 | A | C | F | V | TP53 | ENST00000269305 | 0.21193 | 0.87994 | 0.23348 | 0.94276 | 0.7675 | 0.4539688851 | 0.95013 | 0.2255321834 | 0.165315 | 7.636715839 | 0 |
| 17 | 7674897 | A | C | F | V | TP53 | ENST00000269305 | 0.21193 | 0.87994 | 0.23348 | 0.94276 | 0.7675 | 0.4539688851 | 0.95013 | 0.2255321834 | 0.165315 | 7.636715839 | 0 |
| 17 | 7674897 | A | G | F | L | TP53 | ENST00000269305 | 0.19877 | 0.65049 | 0.2843 | 0.79782 | 0.77399 | 0.3080230625 | 0.97828 | 0.09025730938 | 0.148143 | 2.480066794 | 0 |
| 17 | 7674897 | A | G | F | L | TP53 | ENST00000269305 | 0.19877 | 0.65049 | 0.2843 | 0.79782 | 0.77399 | 0.3080230625 | 0.97828 | 0.09025730938 | 0.148143 | 2.480066794 | 0 |
| 17 | 7674897 | A | G | F | L | TP53 | ENST00000269305 | 0.19877 | 0.65049 | 0.2843 | 0.79782 | 0.77399 | 0.3080230625 | 0.97828 | 0.09025730938 | 0.148143 | 2.480066794 | 0 |
| 17 | 7674897 | A | T | F | I | TP53 | ENST00000269305 | 0.21287 | 0.84102 | 0.24863 | 0.95837 | 0.77205 | 0.4689659272 | 0.95536 | 0.2640517056 | 0.16084 | 7.636715839 | 0 |
| 17 | 7674897 | A | T | F | I | TP53 | ENST00000269305 | 0.21287 | 0.84102 | 0.24863 | 0.95837 | 0.77205 | 0.4689659272 | 0.95536 | 0.2640517056 | 0.16084 | 7.636715839 | 1 |
| 17 | 7674899 | G | A | T | I | TP53 | ENST00000269305 | 0.89348 | 0.89993 | 0.44659 | 0.96947 | 0.98869 | 0.7165783948 | 0.99522 | 0.9984715581 | 0.577827 | 7.512905167 | 1 |
| 17 | 7674899 | G | A | T | I | TP53 | ENST00000269305 | 0.89348 | 0.89993 | 0.44659 | 0.96947 | 0.98869 | 0.7165783948 | 0.99522 | 0.9984715581 | 0.577827 | 7.512905167 | 1 |
| 17 | 7674899 | G | C | T | S | TP53 | ENST00000269305 | 0.54677 | 0.58701 | 0.31823 | 0.9241 | 0.91652 | 0.4279526744 | 0.96342 | 0.9412460923 | 0.38477 | 5.155573214 | 0 |
| 17 | 7674899 | G | C | T | S | TP53 | ENST00000269305 | 0.54677 | 0.58701 | 0.31823 | 0.9241 | 0.91652 | 0.4279526744 | 0.96342 | 0.9412460923 | 0.38477 | 5.155573214 | 0 |
| 17 | 7674899 | G | C | T | S | TP53 | ENST00000269305 | 0.54677 | 0.58701 | 0.31823 | 0.9241 | 0.91652 | 0.4279526744 | 0.96342 | 0.9412460923 | 0.38477 | 5.155573214 | 0 |
| 17 | 7674899 | G | T | T | N | TP53 | ENST00000269305 | 0.83236 | 0.88818 | 0.38575 | 0.96612 | 0.91778 | 0.7210080684 | 0.93148 | 0.9948590994 | 0.30673 | 5.886151178 | 0 |
| 17 | 7674899 | G | T | T | N | TP53 | ENST00000269305 | 0.83236 | 0.88818 | 0.38575 | 0.96612 | 0.91778 | 0.7210080684 | 0.93148 | 0.9948590994 | 0.30673 | 5.886151178 | 0 |
| 17 | 7674899 | G | T | T | N | TP53 | ENST00000269305 | 0.83236 | 0.88818 | 0.38575 | 0.96612 | 0.91778 | 0.7210080684 | 0.93148 | 0.9948590994 | 0.30673 | 5.886151178 | 0 |
| 17 | 7674900 | T | A | T | S | TP53 | ENST00000269305 | 0.48666 | 0.58701 | 0.31823 | 0.93248 | 0.93475 | 0.4279526744 | 0.97164 | 0.9392846227 | 0.381315 | 5.155573214 | 0 |
| 17 | 7674900 | T | A | T | S | TP53 | ENST00000269305 | 0.48666 | 0.58701 | 0.31823 | 0.93248 | 0.93475 | 0.4279526744 | 0.97164 | 0.9392846227 | 0.381315 | 5.155573214 | 0 |
| 17 | 7674900 | T | C | T | A | TP53 | ENST00000269305 | 0.80124 | 0.76605 | 0.30192 | 0.96904 | 0.96553 | 0.6402646925 | 0.99348 | 0.9953173399 | 0.528719 | 6.659372151 | 1 |
| 17 | 7674900 | T | C | T | A | TP53 | ENST00000269305 | 0.80124 | 0.76605 | 0.30192 | 0.96904 | 0.96553 | 0.6402646925 | 0.99348 | 0.9953173399 | 0.528719 | 6.659372151 | 0 |
| 17 | 7674900 | T | G | T | P | TP53 | ENST00000269305 | 0.86133 | 0.91961 | 0.29744 | 0.97993 | 0.97661 | 0.8027183086 | 0.99599 | 0.9981716871 | 0.575931 | 8.967333419 | 1 |
| 17 | 7674900 | T | G | T | P | TP53 | ENST00000269305 | 0.86133 | 0.91961 | 0.29744 | 0.97993 | 0.97661 | 0.8027183086 | 0.99599 | 0.9981716871 | 0.575931 | 8.967333419 | 0 |
| 17 | 7674900 | T | G | T | P | TP53 | ENST00000269305 | 0.86133 | 0.91961 | 0.29744 | 0.97993 | 0.97661 | 0.8027183086 | 0.99599 | 0.9981716871 | 0.575931 | 8.967333419 | 1 |
| 17 | 7674901 | G | C | N | K | TP53 | ENST00000269305 | 0.20655 | 0.77969 | 0.07168 | 0.90915 | 0.69168 | 0.3022842318 | 0.88027 | 0.09500435609 | 0.10919 | 3.398805293 | 0 |
| 17 | 7674901 | G | C | N | K | TP53 | ENST00000269305 | 0.20655 | 0.77969 | 0.07168 | 0.90915 | 0.69168 | 0.3022842318 | 0.88027 | 0.09500435609 | 0.10919 | 3.398805293 | 0 |
| 17 | 7674901 | G | C | N | K | TP53 | ENST00000269305 | 0.20655 | 0.77969 | 0.07168 | 0.90915 | 0.69168 | 0.3022842318 | 0.88027 | 0.09500435609 | 0.10919 | 3.398805293 | 0 |
| 17 | 7674901 | G | T | N | K | TP53 | ENST00000269305 | 0.20344 | 0.77969 | 0.07168 | 0.91191 | 0.69254 | 0.3022842318 | 0.88083 | 0.1152562032 | 0.109193 | 3.398805293 | 0 |
| 17 | 7674901 | G | T | N | K | TP53 | ENST00000269305 | 0.20344 | 0.77969 | 0.07168 | 0.91191 | 0.69254 | 0.3022842318 | 0.88083 | 0.1152562032 | 0.109193 | 3.398805293 | 0 |
| 17 | 7674901 | G | T | N | K | TP53 | ENST00000269305 | 0.20344 | 0.77969 | 0.07168 | 0.91191 | 0.69254 | 0.3022842318 | 0.88083 | 0.1152562032 | 0.109193 | 3.398805293 | 0 |
| 17 | 7674902 | T | A | N | I | TP53 | ENST00000269305 | 0.20226 | 0.67498 | 0.05531 | 0.96336 | 0.7872 | 0.3789207547 | 0.91602 | 0.1270339181 | 0.12187 | 2.598685239 | 0 |
| 17 | 7674902 | T | A | N | I | TP53 | ENST00000269305 | 0.20226 | 0.67498 | 0.05531 | 0.96336 | 0.7872 | 0.3789207547 | 0.91602 | 0.1270339181 | 0.12187 | 2.598685239 | 0 |
| 17 | 7674902 | T | A | N | I | TP53 | ENST00000269305 | 0.20226 | 0.67498 | 0.05531 | 0.96336 | 0.7872 | 0.3789207547 | 0.91602 | 0.1270339181 | 0.12187 | 2.598685239 | 0 |
| 17 | 7674902 | T | C | N | S | TP53 | ENST00000269305 | 0.16125 | 0.51968 | 0.04263 | 0.93263 | 0.57877 | 0.1756455785 | 0.8164 | 0.07781996578 | 0.0382646 | 3.41600959 | 0 |
| 17 | 7674902 | T | C | N | S | TP53 | ENST00000269305 | 0.16125 | 0.51968 | 0.04263 | 0.93263 | 0.57877 | 0.1756455785 | 0.8164 | 0.07781996578 | 0.0382646 | 3.41600959 | 0 |
| 17 | 7674902 | T | C | N | S | TP53 | ENST00000269305 | 0.16125 | 0.51968 | 0.04263 | 0.93263 | 0.57877 | 0.1756455785 | 0.8164 | 0.07781996578 | 0.0382646 | 3.41600959 | 0 |
| 17 | 7674902 | T | G | N | T | TP53 | ENST00000269305 | 0.17398 | 0.58797 | 0.05563 | 0.90785 | 0.70097 | 0.2557365516 | 0.85179 | 0.01564835571 | 0.0954717 | 1.65515428 | 0 |
| 17 | 7674902 | T | G | N | T | TP53 | ENST00000269305 | 0.17398 | 0.58797 | 0.05563 | 0.90785 | 0.70097 | 0.2557365516 | 0.85179 | 0.01564835571 | 0.0954717 | 1.65515428 | 0 |
| 17 | 7674902 | T | G | N | T | TP53 | ENST00000269305 | 0.17398 | 0.58797 | 0.05563 | 0.90785 | 0.70097 | 0.2557365516 | 0.85179 | 0.01564835571 | 0.0954717 | 1.65515428 | 0 |
| 17 | 7674903 | T | A | N | Y | TP53 | ENST00000269305 | 0.21857 | 0.71392 | 0.06655 | 0.97093 | 0.77335 | 0.3798511671 | 0.91501 | 0.07965318024 | 0.121662 | 4.251200653 | 0 |
| 17 | 7674903 | T | A | N | Y | TP53 | ENST00000269305 | 0.21857 | 0.71392 | 0.06655 | 0.97093 | 0.77335 | 0.3798511671 | 0.91501 | 0.07965318024 | 0.121662 | 4.251200653 | 0 |
| 17 | 7674903 | T | C | N | D | TP53 | ENST00000269305 | 0.15321 | 0.71252 | 0.07106 | 0.97381 | 0.62436 | 0.3334543348 | 0.88572 | 0.09309458843 | 0.0681818 | 4.512474191 | 0 |
| 17 | 7674903 | T | C | N | D | TP53 | ENST00000269305 | 0.15321 | 0.71252 | 0.07106 | 0.97381 | 0.62436 | 0.3334543348 | 0.88572 | 0.09309458843 | 0.0681818 | 4.512474191 | 0 |
| 17 | 7674903 | T | C | N | D | TP53 | ENST00000269305 | 0.15321 | 0.71252 | 0.07106 | 0.97381 | 0.62436 | 0.3334543348 | 0.88572 | 0.09309458843 | 0.0681818 | 4.512474191 | 0 |
| 17 | 7674903 | T | G | N | H | TP53 | ENST00000269305 | 0.11462 | 0.59546 | 0.05732 | 0.96021 | 0.66544 | 0.2951185979 | 0.9126 | 0.02451925538 | 0.0743972 | 2.966028942 | 0 |
| 17 | 7674903 | T | G | N | H | TP53 | ENST00000269305 | 0.11462 | 0.59546 | 0.05732 | 0.96021 | 0.66544 | 0.2951185979 | 0.9126 | 0.02451925538 | 0.0743972 | 2.966028942 | 0 |
| 17 | 7674903 | T | G | N | H | TP53 | ENST00000269305 | 0.11462 | 0.59546 | 0.05732 | 0.96021 | 0.66544 | 0.2951185979 | 0.9126 | 0.02451925538 | 0.0743972 | 2.966028942 | 0 |
| 17 | 7674904 | T | A | R | S | TP53 | ENST00000269305 | 0.11859 | 0.62887 | 0.01488 | 0.97609 | 0.83725 | 0.3122093744 | 0.92818 | 0.05501764215 | 0.121439 | 1.959614861 | 0 |
| 17 | 7674904 | T | A | R | S | TP53 | ENST00000269305 | 0.11859 | 0.62887 | 0.01488 | 0.97609 | 0.83725 | 0.3122093744 | 0.92818 | 0.05501764215 | 0.121439 | 1.959614861 | 0 |
| 17 | 7674904 | T | A | R | S | TP53 | ENST00000269305 | 0.11859 | 0.62887 | 0.01488 | 0.97609 | 0.83725 | 0.3122093744 | 0.92818 | 0.05501764215 | 0.121439 | 1.959614861 | 0 |
| 17 | 7674904 | T | G | R | S | TP53 | ENST00000269305 | 0.12184 | 0.62887 | 0.01488 | 0.97619 | 0.83725 | 0.3122093744 | 0.92811 | 0.05558907072 | 0.121438 | 1.959614861 | 0 |
| 17 | 7674904 | T | G | R | S | TP53 | ENST00000269305 | 0.12184 | 0.62887 | 0.01488 | 0.97619 | 0.83725 | 0.3122093744 | 0.92811 | 0.05558907072 | 0.121438 | 1.959614861 | 0 |
| 17 | 7674904 | T | G | R | S | TP53 | ENST00000269305 | 0.12184 | 0.62887 | 0.01488 | 0.97619 | 0.83725 | 0.3122093744 | 0.92811 | 0.05558907072 | 0.121438 | 1.959614861 | 0 |
| 17 | 7674905 | C | A | R | I | TP53 | ENST00000269305 | 0.1126 | 0.78321 | 0.02387 | 0.9229 | 0.80536 | 0.3058902047 | 0.94075 | 0.08549435611 | 0.135559 | 2.667508366 | 0 |
| 17 | 7674905 | C | A | R | I | TP53 | ENST00000269305 | 0.1126 | 0.78321 | 0.02387 | 0.9229 | 0.80536 | 0.3058902047 | 0.94075 | 0.08549435611 | 0.135559 | 2.667508366 | 0 |
| 17 | 7674905 | C | A | R | I | TP53 | ENST00000269305 | 0.1126 | 0.78321 | 0.02387 | 0.9229 | 0.80536 | 0.3058902047 | 0.94075 | 0.08549435611 | 0.135559 | 2.667508366 | 0 |
| 17 | 7674905 | C | G | R | T | TP53 | ENST00000269305 | 0.07328 | 0.72555 | 0.01927 | 0.91547 | 0.78782 | 0.3379448901 | 0.89219 | 0.08859390765 | 0.106462 | 1.989977687 | 0 |
| 17 | 7674905 | C | G | R | T | TP53 | ENST00000269305 | 0.07328 | 0.72555 | 0.01927 | 0.91547 | 0.78782 | 0.3379448901 | 0.89219 | 0.08859390765 | 0.106462 | 1.989977687 | 0 |
| 17 | 7674905 | C | G | R | T | TP53 | ENST00000269305 | 0.07328 | 0.72555 | 0.01927 | 0.91547 | 0.78782 | 0.3379448901 | 0.89219 | 0.08859390765 | 0.106462 | 1.989977687 | 0 |
| 17 | 7674905 | C | T | R | K | TP53 | ENST00000269305 | 0.05501 | 0.58346 | 0.02254 | 0.83771 | 0.75552 | 0.2734669238 | 0.82711 | 0.04003203401 | 0.0795301 | 0.8667595539 | 0 |
| 17 | 7674905 | C | T | R | K | TP53 | ENST00000269305 | 0.05501 | 0.58346 | 0.02254 | 0.83771 | 0.75552 | 0.2734669238 | 0.82711 | 0.04003203401 | 0.0795301 | 0.8667595539 | 0 |
| 17 | 7674905 | C | T | R | K | TP53 | ENST00000269305 | 0.05501 | 0.58346 | 0.02254 | 0.83771 | 0.75552 | 0.2734669238 | 0.82711 | 0.04003203401 | 0.0795301 | 0.8667595539 | 0 |
| 17 | 7674906 | T | C | R | G | TP53 | ENST00000269305 | 0.2754 | 0.72188 | 0.01559 | 0.96367 | 0.862 | 0.2656221163 | 0.98439 | 0.09679087996 | 0.198211 | 2.954291832 | 0 |
| 17 | 7674906 | T | C | R | G | TP53 | ENST00000269305 | 0.2754 | 0.72188 | 0.01559 | 0.96367 | 0.862 | 0.2656221163 | 0.98439 | 0.09679087996 | 0.198211 | 2.954291832 | 0 |
| 17 | 7674906 | T | C | R | G | TP53 | ENST00000269305 | 0.2754 | 0.72188 | 0.01559 | 0.96367 | 0.862 | 0.2656221163 | 0.98439 | 0.09679087996 | 0.198211 | 2.954291832 | 0 |
| 17 | 7674907 | G | C | D | E | TP53 | ENST00000269305 | 0.56034 | 0.88846 | 0.34254 | 0.98396 | 0.92233 | 0.6538705847 | 0.99223 | 0.9913034439 | 0.482585 | 5.700756032 | 0 |
| 17 | 7674907 | G | C | D | E | TP53 | ENST00000269305 | 0.56034 | 0.88846 | 0.34254 | 0.98396 | 0.92233 | 0.6538705847 | 0.99223 | 0.9913034439 | 0.482585 | 5.700756032 | 0 |
| 17 | 7674907 | G | T | D | E | TP53 | ENST00000269305 | 0.55477 | 0.88846 | 0.34254 | 0.9845 | 0.92233 | 0.6538705847 | 0.9922 | 0.9913034439 | 0.482569 | 5.700756032 | 0 |
| 17 | 7674907 | G | T | D | E | TP53 | ENST00000269305 | 0.55477 | 0.88846 | 0.34254 | 0.9845 | 0.92233 | 0.6538705847 | 0.9922 | 0.9913034439 | 0.482569 | 5.700756032 | 0 |
| 17 | 7674908 | T | A | D | V | TP53 | ENST00000269305 | 0.85175 | 0.93746 | 0.31198 | 0.97226 | 0.99072 | 0.8305786261 | 0.99762 | 0.9996232986 | 0.566266 | 6.471831391 | 1 |
| 17 | 7674908 | T | A | D | V | TP53 | ENST00000269305 | 0.85175 | 0.93746 | 0.31198 | 0.97226 | 0.99072 | 0.8305786261 | 0.99762 | 0.9996232986 | 0.566266 | 6.471831391 | 1 |
| 17 | 7674908 | T | A | D | V | TP53 | ENST00000269305 | 0.85175 | 0.93746 | 0.31198 | 0.97226 | 0.99072 | 0.8305786261 | 0.99762 | 0.9996232986 | 0.566266 | 6.471831391 | 1 |
| 17 | 7674908 | T | C | D | G | TP53 | ENST00000269305 | 0.86097 | 0.92856 | 0.32952 | 0.97315 | 0.98869 | 0.8338576471 | 0.99457 | 0.9985243678 | 0.566065 | 5.684208944 | 0 |
| 17 | 7674908 | T | C | D | G | TP53 | ENST00000269305 | 0.86097 | 0.92856 | 0.32952 | 0.97315 | 0.98869 | 0.8338576471 | 0.99457 | 0.9985243678 | 0.566065 | 5.684208944 | 0 |
| 17 | 7674908 | T | G | D | A | TP53 | ENST00000269305 | 0.86018 | 0.92269 | 0.31633 | 0.97376 | 0.98404 | 0.8065210145 | 0.98976 | 0.9993115664 | 0.580697 | 6.565131863 | 1 |
| 17 | 7674908 | T | G | D | A | TP53 | ENST00000269305 | 0.86018 | 0.92269 | 0.31633 | 0.97376 | 0.98404 | 0.8065210145 | 0.98976 | 0.9993115664 | 0.580697 | 6.565131863 | 0 |
| 17 | 7674909 | C | A | D | Y | TP53 | ENST00000269305 | 0.6514 | 0.92161 | 0.37141 | 0.95204 | 0.98834 | 0.8612693181 | 0.9963 | 0.9994754195 | 0.578399 | 6.9047381 | 1 |
| 17 | 7674909 | C | A | D | Y | TP53 | ENST00000269305 | 0.6514 | 0.92161 | 0.37141 | 0.95204 | 0.98834 | 0.8612693181 | 0.9963 | 0.9994754195 | 0.578399 | 6.9047381 | 0 |
| 17 | 7674909 | C | A | D | Y | TP53 | ENST00000269305 | 0.6514 | 0.92161 | 0.37141 | 0.95204 | 0.98834 | 0.8612693181 | 0.9963 | 0.9994754195 | 0.578399 | 6.9047381 | 0 |
| 17 | 7674909 | C | G | D | H | TP53 | ENST00000269305 | 0.63566 | 0.91723 | 0.39221 | 0.95863 | 0.98367 | 0.7652643861 | 0.98647 | 0.9962415695 | 0.550033 | 5.233458992 | 0 |
| 17 | 7674909 | C | G | D | H | TP53 | ENST00000269305 | 0.63566 | 0.91723 | 0.39221 | 0.95863 | 0.98367 | 0.7652643861 | 0.98647 | 0.9962415695 | 0.550033 | 5.233458992 | 0 |
| 17 | 7674909 | C | T | D | N | TP53 | ENST00000269305 | 0.61633 | 0.89413 | 0.36756 | 0.94701 | 0.91945 | 0.7270538692 | 0.95596 | 0.9973323345 | 0.195515 | 4.047645949 | 0 |
| 17 | 7674909 | C | T | D | N | TP53 | ENST00000269305 | 0.61633 | 0.89413 | 0.36756 | 0.94701 | 0.91945 | 0.7270538692 | 0.95596 | 0.9973323345 | 0.195515 | 4.047645949 | 0 |
| 17 | 7674909 | C | T | D | N | TP53 | ENST00000269305 | 0.61633 | 0.89413 | 0.36756 | 0.94701 | 0.91945 | 0.7270538692 | 0.95596 | 0.9973323345 | 0.195515 | 4.047645949 | 0 |
| 17 | 7674910 | A | C | D | E | TP53 | ENST00000269305 | 0.11437 | 0.55511 | 0.16916 | 0.94262 | 0.74376 | 0.2240072206 | 0.79294 | 0.05142727867 | 0.0679375 | 2.172750222 | 0 |
| 17 | 7674910 | A | C | D | E | TP53 | ENST00000269305 | 0.11437 | 0.55511 | 0.16916 | 0.94262 | 0.74376 | 0.2240072206 | 0.79294 | 0.05142727867 | 0.0679375 | 2.172750222 | 0 |
| 17 | 7674910 | A | C | D | E | TP53 | ENST00000269305 | 0.11437 | 0.55511 | 0.16916 | 0.94262 | 0.74376 | 0.2240072206 | 0.79294 | 0.05142727867 | 0.0679375 | 2.172750222 | 0 |
| 17 | 7674910 | A | T | D | E | TP53 | ENST00000269305 | 0.11174 | 0.55511 | 0.16916 | 0.94262 | 0.74306 | 0.2240072206 | 0.79244 | 0.05142727867 | 0.067934 | 2.172750222 | 0 |
| 17 | 7674910 | A | T | D | E | TP53 | ENST00000269305 | 0.11174 | 0.55511 | 0.16916 | 0.94262 | 0.74306 | 0.2240072206 | 0.79244 | 0.05142727867 | 0.067934 | 2.172750222 | 0 |
| 17 | 7674910 | A | T | D | E | TP53 | ENST00000269305 | 0.11174 | 0.55511 | 0.16916 | 0.94262 | 0.74306 | 0.2240072206 | 0.79244 | 0.05142727867 | 0.067934 | 2.172750222 | 0 |
| 17 | 7674911 | T | A | D | V | TP53 | ENST00000269305 | 0.6402 | 0.90175 | 0.15656 | 0.98534 | 0.85222 | 0.5237886909 | 0.97738 | 0.9869203568 | 0.324915 | 4.556911955 | 0 |

| | | | | | | | | | | | | | | | | | |
|---|---|---|---|---|---|---|---|---|---|---|---|---|---|---|---|---|---|
| 17 | 7674911 | T | A | D | V | TP53 | ENST00000269305 | 0.6402 | 0.90175 | 0.15656 | 0.98534 | 0.85222 | 0.5237886909 | 0.97738 | 0.9869203568 | 0.324915 | 4.556911955 | 0 |
| 17 | 7674911 | T | A | D | V | TP53 | ENST00000269305 | 0.6402 | 0.90175 | 0.15656 | 0.98534 | 0.85222 | 0.5237886909 | 0.97738 | 0.9869203568 | 0.324915 | 4.556911955 | 0 |
| 17 | 7674911 | T | C | D | G | TP53 | ENST00000269305 | 0.68491 | 0.87067 | 0.16529 | 0.976 | 0.85222 | 0.5567395401 | 0.96669 | 0.9921653271 | 0.301609 | 4.770921864 | 0 |
| 17 | 7674911 | T | C | D | G | TP53 | ENST00000269305 | 0.68491 | 0.87067 | 0.16529 | 0.976 | 0.85222 | 0.5567395401 | 0.96669 | 0.9921653271 | 0.301609 | 4.770921864 | 0 |
| 17 | 7674911 | T | C | D | G | TP53 | ENST00000269305 | 0.68491 | 0.87067 | 0.16529 | 0.976 | 0.85222 | 0.5567395401 | 0.96669 | 0.9921653271 | 0.301609 | 4.770921864 | 0 |
| 17 | 7674911 | T | G | D | A | TP53 | ENST00000269305 | 0.5304 | 0.8144 | 0.15971 | 0.97882 | 0.79697 | 0.4416008161 | 0.93484 | 0.9795885682 | 0.24441 | 5.858072918 | 0 |
| 17 | 7674911 | T | G | D | A | TP53 | ENST00000269305 | 0.5304 | 0.8144 | 0.15971 | 0.97882 | 0.79697 | 0.4416008161 | 0.93484 | 0.9795885682 | 0.24441 | 5.858072918 | 0 |
| 17 | 7674911 | T | G | D | A | TP53 | ENST00000269305 | 0.5304 | 0.8144 | 0.15971 | 0.97882 | 0.79697 | 0.4416008161 | 0.93484 | 0.9795885682 | 0.24441 | 5.858072918 | 0 |
| 17 | 7674912 | C | A | D | Y | TP53 | ENST00000269305 | 0.60978 | 0.91805 | 0.20038 | 0.98397 | 0.84694 | 0.5292298882 | 0.98184 | 0.998221457 | 0.320107 | 4.931804634 | 0 |
| 17 | 7674912 | C | A | D | Y | TP53 | ENST00000269305 | 0.60978 | 0.91805 | 0.20038 | 0.98397 | 0.84694 | 0.5292298882 | 0.98184 | 0.998221457 | 0.320107 | 4.931804634 | 0 |
| 17 | 7674912 | C | A | D | Y | TP53 | ENST00000269305 | 0.60978 | 0.91805 | 0.20038 | 0.98397 | 0.84694 | 0.5292298882 | 0.98184 | 0.998221457 | 0.320107 | 4.931804634 | 0 |
| 17 | 7674912 | C | G | D | H | TP53 | ENST00000269305 | 0.61111 | 0.90472 | 0.20513 | 0.97838 | 0.80889 | 0.4055629003 | 0.9624 | 0.9910855889 | 0.239515 | 5.273793359 | 0 |
| 17 | 7674912 | C | G | D | H | TP53 | ENST00000269305 | 0.61111 | 0.90472 | 0.20513 | 0.97838 | 0.80889 | 0.4055629003 | 0.9624 | 0.9910855889 | 0.239515 | 5.273793359 | 0 |
| 17 | 7674912 | C | G | D | H | TP53 | ENST00000269305 | 0.61111 | 0.90472 | 0.20513 | 0.97838 | 0.80889 | 0.4055629003 | 0.9624 | 0.9910855889 | 0.239515 | 5.273793359 | 0 |
| 17 | 7674912 | C | T | D | N | TP53 | ENST00000269305 | 0.35674 | 0.59521 | 0.19032 | 0.96467 | 0.7217 | 0.319440035 | 0.93379 | 0.867898941 | 0.128017 | 3.487467438 | 0 |
| 17 | 7674912 | C | T | D | N | TP53 | ENST00000269305 | 0.35674 | 0.59521 | 0.19032 | 0.96467 | 0.7217 | 0.319440035 | 0.93379 | 0.867898941 | 0.128017 | 3.487467438 | 0 |
| 17 | 7674913 | C | A | L | F | TP53 | ENST00000269305 | 0.18187 | 0.63826 | 0.15474 | 0.9324 | 0.73296 | 0.250028568 | 0.87484 | 0.08513224125 | 0.134101 | 4.318904086 | 0 |
| 17 | 7674913 | C | A | L | F | TP53 | ENST00000269305 | 0.18187 | 0.63826 | 0.15474 | 0.9324 | 0.73296 | 0.250028568 | 0.87484 | 0.08513224125 | 0.134101 | 4.318904086 | 0 |
| 17 | 7674913 | C | A | L | F | TP53 | ENST00000269305 | 0.18187 | 0.63826 | 0.15474 | 0.9324 | 0.73296 | 0.250028568 | 0.87484 | 0.08513224125 | 0.134101 | 4.318904086 | 0 |
| 17 | 7674913 | C | G | L | F | TP53 | ENST00000269305 | 0.1857 | 0.63826 | 0.15474 | 0.93416 | 0.73296 | 0.250028568 | 0.87554 | 0.08513224125 | 0.134114 | 4.318904086 | 0 |
| 17 | 7674913 | C | G | L | F | TP53 | ENST00000269305 | 0.1857 | 0.63826 | 0.15474 | 0.93416 | 0.73296 | 0.250028568 | 0.87554 | 0.08513224125 | 0.134114 | 4.318904086 | 0 |
| 17 | 7674913 | C | G | L | F | TP53 | ENST00000269305 | 0.1857 | 0.63826 | 0.15474 | 0.93416 | 0.73296 | 0.250028568 | 0.87554 | 0.08513224125 | 0.134114 | 4.318904086 | 0 |
| 17 | 7674914 | A | C | L | W | TP53 | ENST00000269305 | 0.29627 | 0.94631 | 0.20571 | 0.97512 | 0.80178 | 0.5199260672 | 0.94003 | 0.44288674 | 0.207371 | 3.893576299 | 0 |
| 17 | 7674914 | A | C | L | W | TP53 | ENST00000269305 | 0.29627 | 0.94631 | 0.20571 | 0.97512 | 0.80178 | 0.5199260672 | 0.94003 | 0.44288674 | 0.207371 | 3.893576299 | 0 |
| 17 | 7674914 | A | C | L | W | TP53 | ENST00000269305 | 0.29627 | 0.94631 | 0.20571 | 0.97512 | 0.80178 | 0.5199260672 | 0.94003 | 0.44288674 | 0.207371 | 3.893576299 | 0 |
| 17 | 7674914 | A | G | L | S | TP53 | ENST00000269305 | 0.18266 | 0.74842 | 0.1602 | 0.96138 | 0.84641 | 0.5793013007 | 0.91146 | 0.6936503649 | 0.243186 | 3.729779351 | 0 |
| 17 | 7674914 | A | G | L | S | TP53 | ENST00000269305 | 0.18266 | 0.74842 | 0.1602 | 0.96138 | 0.84641 | 0.5793013007 | 0.91146 | 0.6936503649 | 0.243186 | 3.729779351 | 0 |
| 17 | 7674914 | A | G | L | S | TP53 | ENST00000269305 | 0.18266 | 0.74842 | 0.1602 | 0.96138 | 0.84641 | 0.5793013007 | 0.91146 | 0.6936503649 | 0.243186 | 3.729779351 | 0 |
| 17 | 7674915 | A | C | L | V | TP53 | ENST00000269305 | 0.14084 | 0.58542 | 0.1215 | 0.87019 | 0.80358 | 0.2361947927 | 0.86965 | 0.04567645118 | 0.103284 | 3.306920655 | 0 |
| 17 | 7674915 | A | C | L | V | TP53 | ENST00000269305 | 0.14084 | 0.58542 | 0.1215 | 0.87019 | 0.80358 | 0.2361947927 | 0.86965 | 0.04567645118 | 0.103284 | 3.306920655 | 0 |
| 17 | 7674915 | A | T | L | M | TP53 | ENST00000269305 | 0.1562 | 0.85993 | 0.14758 | 0.97407 | 0.74376 | 0.2169863755 | 0.8289 | 0.3265433609 | 0.122317 | 4.268487734 | 0 |
| 17 | 7674915 | A | T | L | M | TP53 | ENST00000269305 | 0.1562 | 0.85993 | 0.14758 | 0.97407 | 0.74376 | 0.2169863755 | 0.8289 | 0.3265433609 | 0.122317 | 4.268487734 | 0 |
| 17 | 7674915 | A | T | L | M | TP53 | ENST00000269305 | 0.1562 | 0.85993 | 0.14758 | 0.97407 | 0.74376 | 0.2169863755 | 0.8289 | 0.3265433609 | 0.122317 | 4.268487734 | 0 |
| 17 | 7674917 | T | A | Y | F | TP53 | ENST00000269305 | 0.88566 | 0.87402 | 0.45008 | 0.97239 | 0.94431 | 0.5614879443 | 0.96379 | 0.995644331 | 0.313765 | 5.064885481 | 1 |
| 17 | 7674917 | T | A | Y | F | TP53 | ENST00000269305 | 0.88566 | 0.87402 | 0.45008 | 0.97239 | 0.94431 | 0.5614879443 | 0.96379 | 0.995644331 | 0.313765 | 5.064885481 | 1 |
| 17 | 7674917 | T | A | Y | F | TP53 | ENST00000269305 | 0.88566 | 0.87402 | 0.45008 | 0.97239 | 0.94431 | 0.5614879443 | 0.96379 | 0.995644331 | 0.313765 | 5.064885481 | 1 |
| 17 | 7674917 | T | C | Y | C | TP53 | ENST00000269305 | 0.94114 | 0.92209 | 0.52737 | 0.97536 | 0.96262 | 0.860365415 | 0.99604 | 0.9980540276 | 0.571744 | 5.748515634 | 1 |
| 17 | 7674917 | T | C | Y | C | TP53 | ENST00000269305 | 0.94114 | 0.92209 | 0.52737 | 0.97536 | 0.96262 | 0.860365415 | 0.99604 | 0.9980540276 | 0.571744 | 5.748515634 | 1 |
| 17 | 7674917 | T | C | Y | C | TP53 | ENST00000269305 | 0.94114 | 0.92209 | 0.52737 | 0.97536 | 0.96262 | 0.860365415 | 0.99604 | 0.9980540276 | 0.571744 | 5.748515634 | 1 |
| 17 | 7674917 | T | G | Y | S | TP53 | ENST00000269305 | 0.91725 | 0.86282 | 0.46993 | 0.97868 | 0.95499 | 0.853146983 | 0.9985 | 0.9983412027 | 0.558673 | 7.220104814 | 1 |
| 17 | 7674917 | T | G | Y | S | TP53 | ENST00000269305 | 0.91725 | 0.86282 | 0.46993 | 0.97868 | 0.95499 | 0.853146983 | 0.9985 | 0.9983412027 | 0.558673 | 7.220104814 | 1 |
| 17 | 7674917 | T | G | Y | S | TP53 | ENST00000269305 | 0.91725 | 0.86282 | 0.46993 | 0.97868 | 0.95499 | 0.853146983 | 0.9985 | 0.9983412027 | 0.558673 | 7.220104814 | 1 |
| 17 | 7674918 | A | C | Y | D | TP53 | ENST00000269305 | 0.87099 | 0.95231 | 0.52196 | 0.97148 | 0.98183 | 0.8964550135 | 0.998 | 0.9996414185 | 0.568646 | 7.220104814 | 1 |
| 17 | 7674918 | A | C | Y | D | TP53 | ENST00000269305 | 0.87099 | 0.95231 | 0.52196 | 0.97148 | 0.98183 | 0.8964550135 | 0.998 | 0.9996414185 | 0.568646 | 7.220104814 | 1 |
| 17 | 7674918 | A | C | Y | D | TP53 | ENST00000269305 | 0.87099 | 0.95231 | 0.52196 | 0.97148 | 0.98183 | 0.8964550135 | 0.998 | 0.9996414185 | 0.568646 | 7.220104814 | 1 |
| 17 | 7674918 | A | G | Y | H | TP53 | ENST00000269305 | 0.82186 | 0.8107 | 0.53941 | 0.95021 | 0.97252 | 0.7778036153 | 0.99065 | 0.9909805655 | 0.5587 | 5.507312301 | 1 |
| 17 | 7674918 | A | G | Y | H | TP53 | ENST00000269305 | 0.82186 | 0.8107 | 0.53941 | 0.95021 | 0.97252 | 0.7778036153 | 0.99065 | 0.9909805655 | 0.5587 | 5.507312301 | 1 |
| 17 | 7674918 | A | G | Y | H | TP53 | ENST00000269305 | 0.82186 | 0.8107 | 0.53941 | 0.95021 | 0.97252 | 0.7778036153 | 0.99065 | 0.9909805655 | 0.5587 | 5.507312301 | 1 |
| 17 | 7674918 | A | T | Y | N | TP53 | ENST00000269305 | 0.86629 | 0.94498 | 0.52948 | 0.97421 | 0.97141 | 0.8596400119 | 0.99094 | 0.998678267 | 0.566016 | 7.220104814 | 1 |
| 17 | 7674918 | A | T | Y | N | TP53 | ENST00000269305 | 0.86629 | 0.94498 | 0.52948 | 0.97421 | 0.97141 | 0.8596400119 | 0.99094 | 0.998678267 | 0.566016 | 7.220104814 | 1 |
| 17 | 7674918 | A | T | Y | N | TP53 | ENST00000269305 | 0.86629 | 0.94498 | 0.52948 | 0.97421 | 0.97141 | 0.8596400119 | 0.99094 | 0.998678267 | 0.566016 | 7.220104814 | 1 |
| 17 | 7674919 | C | A | E | D | TP53 | ENST00000269305 | 0.23537 | 0.7464 | 0.07412 | 0.96514 | 0.78223 | 0.331122637 | 0.90647 | 0.667263329 | 0.19411 | 3.32833841 | 0 |
| 17 | 7674919 | C | A | E | D | TP53 | ENST00000269305 | 0.23537 | 0.7464 | 0.07412 | 0.96514 | 0.78223 | 0.331122637 | 0.90647 | 0.667263329 | 0.19411 | 3.32833841 | 0 |
| 17 | 7674919 | C | A | E | D | TP53 | ENST00000269305 | 0.23537 | 0.7464 | 0.07412 | 0.96514 | 0.78223 | 0.331122637 | 0.90647 | 0.667263329 | 0.19411 | 3.32833841 | 0 |
| 17 | 7674919 | C | G | E | D | TP53 | ENST00000269305 | 0.2399 | 0.7464 | 0.07412 | 0.96394 | 0.78223 | 0.331122637 | 0.90668 | 0.6775787473 | 0.194115 | 3.32833841 | 0 |
| 17 | 7674919 | C | G | E | D | TP53 | ENST00000269305 | 0.2399 | 0.7464 | 0.07412 | 0.96394 | 0.78223 | 0.331122637 | 0.90668 | 0.6775787473 | 0.194115 | 3.32833841 | 0 |
| 17 | 7674919 | C | G | E | D | TP53 | ENST00000269305 | 0.2399 | 0.7464 | 0.07412 | 0.96394 | 0.78223 | 0.331122637 | 0.90668 | 0.6775787473 | 0.194115 | 3.32833841 | 0 |
| 17 | 7674920 | T | A | E | V | TP53 | ENST00000269305 | 0.50844 | 0.90707 | 0.05883 | 0.97022 | 0.83725 | 0.3810682236 | 0.94136 | 0.9657635689 | 0.236114 | 3.954052364 | 0 |
| 17 | 7674920 | T | A | E | V | TP53 | ENST00000269305 | 0.50844 | 0.90707 | 0.05883 | 0.97022 | 0.83725 | 0.3810682236 | 0.94136 | 0.9657635689 | 0.236114 | 3.954052364 | 0 |
| 17 | 7674920 | T | A | E | V | TP53 | ENST00000269305 | 0.50844 | 0.90707 | 0.05883 | 0.97022 | 0.83725 | 0.3810682236 | 0.94136 | 0.9657635689 | 0.236114 | 3.954052364 | 0 |
| 17 | 7674920 | T | C | E | G | TP53 | ENST00000269305 | 0.66739 | 0.90807 | 0.0578 | 0.97431 | 0.84588 | 0.4753399056 | 0.92999 | 0.9762138724 | 0.251373 | 6.7347697 | 0 |
| 17 | 7674920 | T | C | E | G | TP53 | ENST00000269305 | 0.66739 | 0.90807 | 0.0578 | 0.97431 | 0.84588 | 0.4753399056 | 0.92999 | 0.9762138724 | 0.251373 | 6.7347697 | 0 |
| 17 | 7674920 | T | G | E | A | TP53 | ENST00000269305 | 0.50062 | 0.88459 | 0.07348 | 0.9476 | 0.84375 | 0.3503323212 | 0.92894 | 0.9424406886 | 0.232265 | 4.824962215 | 0 |
| 17 | 7674920 | T | G | E | A | TP53 | ENST00000269305 | 0.50062 | 0.88459 | 0.07348 | 0.9476 | 0.84375 | 0.3503323212 | 0.92894 | 0.9424406886 | 0.232265 | 4.824962215 | 0 |
| 17 | 7674920 | T | G | E | A | TP53 | ENST00000269305 | 0.50062 | 0.88459 | 0.07348 | 0.9476 | 0.84375 | 0.3503323212 | 0.92894 | 0.9424406886 | 0.232265 | 4.824962215 | 0 |
| 17 | 7674921 | C | G | E | Q | TP53 | ENST00000269305 | 0.12467 | 0.72274 | 0.06614 | 0.96476 | 0.7589 | 0.2674342467 | 0.88823 | 0.1649941802 | 0.144273 | 3.253237031 | 0 |
| 17 | 7674921 | C | G | E | Q | TP53 | ENST00000269305 | 0.12467 | 0.72274 | 0.06614 | 0.96476 | 0.7589 | 0.2674342467 | 0.88823 | 0.1649941802 | 0.144273 | 3.253237031 | 0 |
| 17 | 7674921 | C | G | E | Q | TP53 | ENST00000269305 | 0.12467 | 0.72274 | 0.06614 | 0.96476 | 0.7589 | 0.2674342467 | 0.88823 | 0.1649941802 | 0.144273 | 3.253237031 | 0 |
| 17 | 7674921 | C | T | E | K | TP53 | ENST00000269305 | 0.19094 | 0.69194 | 0.11049 | 0.86793 | 0.85638 | 0.399493909 | 0.92839 | 0.2163199633 | 0.202426 | 3.501408799 | 0 |
| 17 | 7674921 | C | T | E | K | TP53 | ENST00000269305 | 0.19094 | 0.69194 | 0.11049 | 0.86793 | 0.85638 | 0.399493909 | 0.92839 | 0.2163199633 | 0.202426 | 3.501408799 | 0 |
| 17 | 7674923 | A | C | V | G | TP53 | ENST00000269305 | 0.50478 | 0.70704 | 0.12562 | 0.97427 | 0.83615 | 0.5786431967 | 0.96737 | 0.486097157 | 0.298136 | 4.373258635 | 0 |
| 17 | 7674923 | A | C | V | G | TP53 | ENST00000269305 | 0.50478 | 0.70704 | 0.12562 | 0.97427 | 0.83615 | 0.5786431967 | 0.96737 | 0.486097157 | 0.298136 | 4.373258635 | 0 |
| 17 | 7674923 | A | C | V | G | TP53 | ENST00000269305 | 0.50478 | 0.70704 | 0.12562 | 0.97427 | 0.83615 | 0.5786431967 | 0.96737 | 0.486097157 | 0.298136 | 4.373258635 | 0 |
| 17 | 7674923 | A | G | V | A | TP53 | ENST00000269305 | 0.2744 | 0.64964 | 0.14716 | 0.8271 | 0.80178 | 0.3895876189 | 0.87287 | 0.137090534 | 0.162598 | -1.338339257 | 0 |
| 17 | 7674923 | A | G | V | A | TP53 | ENST00000269305 | 0.2744 | 0.64964 | 0.14716 | 0.8271 | 0.80178 | 0.3895876189 | 0.87287 | 0.137090534 | 0.162598 | -1.338339257 | 0 |
| 17 | 7674923 | A | G | V | A | TP53 | ENST00000269305 | 0.2744 | 0.64964 | 0.14716 | 0.8271 | 0.80178 | 0.3895876189 | 0.87287 | 0.137090534 | 0.162598 | -1.338339257 | 0 |
| 17 | 7674923 | A | T | V | E | TP53 | ENST00000269305 | 0.51152 | 0.78709 | 0.15905 | 0.98098 | 0.85945 | 0.8081491777 | 0.97911 | 0.6967709064 | 0.312334 | 5.914842335 | 1 |
| 17 | 7674923 | A | T | V | E | TP53 | ENST00000269305 | 0.51152 | 0.78709 | 0.15905 | 0.98098 | 0.85945 | 0.8081491777 | 0.97911 | 0.6967709064 | 0.312334 | 5.914842335 | 1 |
| 17 | 7674923 | A | T | V | E | TP53 | ENST00000269305 | 0.51152 | 0.78709 | 0.15905 | 0.98098 | 0.85945 | 0.8081491777 | 0.97911 | 0.6967709064 | 0.312334 | 5.914842335 | 1 |
| 17 | 7674924 | C | A | V | L | TP53 | ENST00000269305 | 0.16633 | 0.57644 | 0.15026 | 0.95491 | 0.78967 | 0.5489581758 | 0.89928 | 0.2468054305 | 0.177028 | 4.655042657 | 1 |
| 17 | 7674924 | C | G | V | L | TP53 | ENST00000269305 | 0.17004 | 0.57644 | 0.15026 | 0.9531 | 0.78967 | 0.5489581758 | 0.9006 | 0.2177571739 | 0.177044 | 4.655042657 | 1 |
| 17 | 7674924 | C | T | V | M | TP53 | ENST00000269305 | 0.20036 | 0.76013 | 0.15832 | 0.95817 | 0.79758 | 0.5297766121 | 0.88947 | 0.477420032 | 0.183943 | 5.914842335 | 1 |
| 17 | 7674926 | C | A | R | L | TP53 | ENST00000269305 | 0.14192 | 0.74076 | 0.01082 | 0.92043 | 0.80417 | 0.3753303017 | 0.91065 | 0.07642903924 | 0.12418 | 3.258037213 | 0 |
| 17 | 7674926 | C | A | R | L | TP53 | ENST00000269305 | 0.14192 | 0.74076 | 0.01082 | 0.92043 | 0.80417 | 0.3753303017 | 0.91065 | 0.07642903924 | 0.12418 | 3.258037213 | 0 |
| 17 | 7674926 | C | A | R | L | TP53 | ENST00000269305 | 0.14192 | 0.74076 | 0.01082 | 0.92043 | 0.80417 | 0.3753303017 | 0.91065 | 0.07642903924 | 0.12418 | 3.258037213 | 0 |
| 17 | 7674926 | C | G | R | P | TP53 | ENST00000269305 | 0.33941 | 0.90884 | 0.01419 | 0.97624 | 0.80298 | 0.6991054584 | 0.94498 | 0.568992734 | 0.278986 | 5.982576952 | 0 |
| 17 | 7674926 | C | G | R | P | TP53 | ENST00000269305 | 0.33941 | 0.90884 | 0.01419 | 0.97624 | 0.80298 | 0.6991054584 | 0.94498 | 0.568992734 | 0.278986 | 5.982576952 | 0 |
| 17 | 7674926 | C | T | R | H | TP53 | ENST00000269305 | 0.13249 | 0.66246 | 0.00996 | 0.89396 | 0.80298 | 0.2986061675 | 0.91929 | 0.104153119 | 0.107211 | 0.9543017468 | 0 |
| 17 | 7674926 | C | T | R | H | TP53 | ENST00000269305 | 0.13249 | 0.66246 | 0.00996 | 0.89396 | 0.80298 | 0.2986061675 | 0.91929 | 0.104153119 | 0.107211 | 0.9543017468 | 0 |
| 17 | 7674926 | C | T | R | H | TP53 | ENST00000269305 | 0.13249 | 0.66246 | 0.00996 | 0.89396 | 0.80298 | 0.2986061675 | 0.91929 | 0.104153119 | 0.107211 | 0.9543017468 | 0 |
| 17 | 7674927 | G | A | R | C | TP53 | ENST00000269305 | 0.41075 | 0.70936 | 0.01902 | 0.97946 | 0.78473 | 0.4533878576 | 0.97622 | 0.2080634534 | 0.12263 | 4.634644593 | 0 |
| 17 | 7674927 | G | A | R | C | TP53 | ENST00000269305 | 0.41075 | 0.70936 | 0.01902 | 0.97946 | 0.78473 | 0.4533878576 | 0.97622 | 0.2080634534 | 0.12263 | 4.634644593 | 0 |
| 17 | 7674927 | G | A | R | C | TP53 | ENST00000269305 | 0.41075 | 0.70936 | 0.01902 | 0.97946 | 0.78473 | 0.4533878576 | 0.97622 | 0.2080634534 | 0.12263 | 4.634644593 | 0 |
| 17 | 7674927 | G | C | R | G | TP53 | ENST00000269305 | 0.46943 | 0.79994 | 0.01088 | 0.96598 | 0.77527 | 0.3615166597 | 0.95219 | 0.4692607224 | 0.256783 | 3.970829105 | 0 |
| 17 | 7674927 | G | C | R | G | TP53 | ENST00000269305 | 0.46943 | 0.79994 | 0.01088 | 0.96598 | 0.77527 | 0.3615166597 | 0.95219 | 0.4692607224 | 0.256783 | 3.970829105 | 0 |
| 17 | 7674927 | G | C | R | G | TP53 | ENST00000269305 | 0.46943 | 0.79994 | 0.01088 | 0.96598 | 0.77527 | 0.3615166597 | 0.95219 | 0.4692607224 | 0.256783 | 3.970829105 | 0 |
| 17 | 7674927 | G | T | R | S | TP53 | ENST00000269305 | 0.353 | 0.76723 | 0.00972 | 0.94991 | 0.77718 | 0.4002706779 | 0.92342 | 0.2669836581 | 0.213588 | 4.322451767 | 0 |
| 17 | 7674927 | G | T | R | S | TP53 | ENST00000269305 | 0.353 | 0.76723 | 0.00972 | 0.94991 | 0.77718 | 0.4002706779 | 0.92342 | 0.2669836581 | 0.213588 | 4.322451767 | 0 |
| 17 | 7674927 | G | T | R | S | TP53 | ENST00000269305 | 0.353 | 0.76723 | 0.00972 | 0.94991 | 0.77718 | 0.4002706779 | 0.92342 | 0.2669836581 | 0.213588 | 4.322451767 | 0 |
| 17 | 7674928 | C | A | L | F | TP53 | ENST00000269305 | 0.09759 | 0.89442 | 0.18232 | 0.98064 | 0.75074 | 0.1989053239 | 0.89636 | 0.4269631505 | 0.135624 | 5.572389032 | 0 |
| 17 | 7674928 | C | A | L | F | TP53 | ENST00000269305 | 0.09759 | 0.89442 | 0.18232 | 0.98064 | 0.75074 | 0.1989053239 | 0.89636 | 0.4269631505 | 0.135624 | 5.572389032 | 0 |

| | | | | | | | | | | | | | | | | |
|---|---|---|---|---|---|---|---|---|---|---|---|---|---|---|---|---|
| 17 | 7674928 | C | A | L | F | TP53 | ENST00000269305 | 0.09759 | 0.89442 | 0.18232 | 0.98064 | 0.75074 | 0.1989053239 | 0.89636 | 0.4269631505 | 0.135624 | 5.572389032 | 0 |
| 17 | 7674928 | C | G | L | F | TP53 | ENST00000269305 | 0.10077 | 0.89442 | 0.18232 | 0.9795 | 0.75074 | 0.1989053239 | 0.89378 | 0.6047585011 | 0.135617 | 5.572389032 | 0 |
| 17 | 7674928 | C | G | L | F | TP53 | ENST00000269305 | 0.10077 | 0.89442 | 0.18232 | 0.9795 | 0.75074 | 0.1989053239 | 0.89378 | 0.6047585011 | 0.135617 | 5.572389032 | 0 |
| 17 | 7674928 | C | G | L | F | TP53 | ENST00000269305 | 0.10077 | 0.89442 | 0.18232 | 0.9795 | 0.75074 | 0.1989053239 | 0.89378 | 0.6047585011 | 0.135617 | 5.572389032 | 0 |
| 17 | 7674929 | A | C | L | W | TP53 | ENST00000269305 | 0.31488 | 0.94935 | 0.22438 | 0.95078 | 0.76881 | 0.3634405971 | 0.98015 | 0.4140223861 | 0.184161 | 7.335593755 | 0 |
| 17 | 7674929 | A | C | L | W | TP53 | ENST00000269305 | 0.31488 | 0.94935 | 0.22438 | 0.95078 | 0.76881 | 0.3634405971 | 0.98015 | 0.4140223861 | 0.184161 | 7.335593755 | 0 |
| 17 | 7674929 | A | C | L | W | TP53 | ENST00000269305 | 0.31488 | 0.94935 | 0.22438 | 0.95078 | 0.76881 | 0.3634405971 | 0.98015 | 0.4140223861 | 0.184161 | 7.335593755 | 0 |
| 17 | 7674929 | A | G | L | S | TP53 | ENST00000269305 | 0.17809 | 0.92762 | 0.17812 | 0.89023 | 0.78906 | 0.2556965192 | 0.95027 | 0.2963080704 | 0.155974 | 3.364757365 | 0 |
| 17 | 7674929 | A | G | L | S | TP53 | ENST00000269305 | 0.17809 | 0.92762 | 0.17812 | 0.89023 | 0.78906 | 0.2556965192 | 0.95027 | 0.2963080704 | 0.155974 | 3.364757365 | 0 |
| 17 | 7674929 | A | G | L | S | TP53 | ENST00000269305 | 0.17809 | 0.92762 | 0.17812 | 0.89023 | 0.78906 | 0.2556965192 | 0.95027 | 0.2963080704 | 0.155974 | 3.364757365 | 0 |
| 17 | 7674930 | A | C | L | V | TP53 | ENST00000269305 | 0.1524 | 0.7728 | 0.14123 | 0.99207 | 0.75143 | 0.213902187 | 0.94535 | 0.4302001297 | 0.139149 | 5.305274442 | 0 |
| 17 | 7674930 | A | C | L | V | TP53 | ENST00000269305 | 0.1524 | 0.7728 | 0.14123 | 0.99207 | 0.75143 | 0.213902187 | 0.94535 | 0.4302001297 | 0.139149 | 5.305274442 | 0 |
| 17 | 7674930 | A | C | L | V | TP53 | ENST00000269305 | 0.1524 | 0.7728 | 0.14123 | 0.99207 | 0.75143 | 0.213902187 | 0.94535 | 0.4302001297 | 0.139149 | 5.305274442 | 0 |
| 17 | 7674930 | A | T | L | M | TP53 | ENST00000269305 | 0.18422 | 0.76751 | 0.16292 | 0.99385 | 0.77654 | 0.1819453035 | 0.95259 | 0.2937461436 | 0.151785 | 4.866328178 | 0 |
| 17 | 7674930 | A | T | L | M | TP53 | ENST00000269305 | 0.18422 | 0.76751 | 0.16292 | 0.99385 | 0.77654 | 0.1819453035 | 0.95259 | 0.2937461436 | 0.151785 | 4.866328178 | 0 |
| 17 | 7674930 | A | T | L | M | TP53 | ENST00000269305 | 0.18422 | 0.76751 | 0.16292 | 0.99385 | 0.77654 | 0.1819453035 | 0.95259 | 0.2937461436 | 0.151785 | 4.866328178 | 0 |
| 17 | 7674931 | A | C | N | K | TP53 | ENST00000269305 | 0.49421 | 0.914 | 0.5604 | 0.9644 | 0.83943 | 0.5906963635 | 0.96512 | 0.9724074602 | 0.208551 | 5.372907599 | 0 |
| 17 | 7674931 | A | C | N | K | TP53 | ENST00000269305 | 0.49421 | 0.914 | 0.5604 | 0.9644 | 0.83943 | 0.5906963635 | 0.96512 | 0.9724074602 | 0.208551 | 5.372907599 | 0 |
| 17 | 7674931 | A | C | N | K | TP53 | ENST00000269305 | 0.49421 | 0.914 | 0.5604 | 0.9644 | 0.83943 | 0.5906963635 | 0.96512 | 0.9724074602 | 0.208551 | 5.372907599 | 0 |
| 17 | 7674931 | A | T | N | K | TP53 | ENST00000269305 | 0.48876 | 0.914 | 0.5604 | 0.96432 | 0.83943 | 0.5906963635 | 0.96487 | 0.9724074602 | 0.208569 | 5.372907599 | 0 |
| 17 | 7674931 | A | T | N | K | TP53 | ENST00000269305 | 0.48876 | 0.914 | 0.5604 | 0.96432 | 0.83943 | 0.5906963635 | 0.96487 | 0.9724074602 | 0.208569 | 5.372907599 | 0 |
| 17 | 7674931 | A | T | N | K | TP53 | ENST00000269305 | 0.48876 | 0.914 | 0.5604 | 0.96432 | 0.83943 | 0.5906963635 | 0.96487 | 0.9724074602 | 0.208569 | 5.372907599 | 0 |
| 17 | 7674932 | T | A | N | I | TP53 | ENST00000269305 | 0.83882 | 0.92292 | 0.49078 | 0.98398 | 0.93241 | 0.7010323428 | 0.96566 | 0.9872969389 | 0.370317 | 4.602370057 | 0 |
| 17 | 7674932 | T | A | N | I | TP53 | ENST00000269305 | 0.83882 | 0.92292 | 0.49078 | 0.98398 | 0.93241 | 0.7010323428 | 0.96566 | 0.9872969389 | 0.370317 | 4.602370057 | 0 |
| 17 | 7674932 | T | A | N | I | TP53 | ENST00000269305 | 0.83882 | 0.92292 | 0.49078 | 0.98398 | 0.93241 | 0.7010323428 | 0.96566 | 0.9872969389 | 0.370317 | 4.602370057 | 0 |
| 17 | 7674932 | T | C | N | S | TP53 | ENST00000269305 | 0.45573 | 0.86678 | 0.43309 | 0.96203 | 0.88307 | 0.327046342 | 0.90474 | 0.8938180804 | 0.164596 | 3.947373653 | 0 |
| 17 | 7674932 | T | C | N | S | TP53 | ENST00000269305 | 0.45573 | 0.86678 | 0.43309 | 0.96203 | 0.88307 | 0.327046342 | 0.90474 | 0.8938180804 | 0.164596 | 3.947373653 | 0 |
| 17 | 7674932 | T | C | N | S | TP53 | ENST00000269305 | 0.45573 | 0.86678 | 0.43309 | 0.96203 | 0.88307 | 0.327046342 | 0.90474 | 0.8938180804 | 0.164596 | 3.947373653 | 0 |
| 17 | 7674932 | T | G | N | T | TP53 | ENST00000269305 | 0.532 | 0.8993 | 0.47258 | 0.97012 | 0.92275 | 0.4649432704 | 0.96023 | 0.9914741516 | 0.280949 | 4.993435094 | 0 |
| 17 | 7674932 | T | G | N | T | TP53 | ENST00000269305 | 0.532 | 0.8993 | 0.47258 | 0.97012 | 0.92275 | 0.4649432704 | 0.96023 | 0.9914741516 | 0.280949 | 4.993435094 | 0 |
| 17 | 7674933 | T | A | N | Y | TP53 | ENST00000269305 | 0.76323 | 0.92061 | 0.5244 | 0.9855 | 0.97252 | 0.656187396 | 0.97486 | 0.9985966086 | 0.422038 | 7.983205784 | 1 |
| 17 | 7674933 | T | A | N | Y | TP53 | ENST00000269305 | 0.76323 | 0.92061 | 0.5244 | 0.9855 | 0.97252 | 0.656187396 | 0.97486 | 0.9985966086 | 0.422038 | 7.983205784 | 0 |
| 17 | 7674933 | T | A | N | Y | TP53 | ENST00000269305 | 0.76323 | 0.92061 | 0.5244 | 0.9855 | 0.97252 | 0.656187396 | 0.97486 | 0.9985966086 | 0.422038 | 7.983205784 | 0 |
| 17 | 7674933 | T | C | N | D | TP53 | ENST00000269305 | 0.73973 | 0.91241 | 0.53372 | 0.97158 | 0.9506 | 0.4951484232 | 0.9705 | 0.9905883074 | 0.276718 | 5.434267555 | 0 |
| 17 | 7674933 | T | C | N | D | TP53 | ENST00000269305 | 0.73973 | 0.91241 | 0.53372 | 0.97158 | 0.9506 | 0.4951484232 | 0.9705 | 0.9905883074 | 0.276718 | 5.434267555 | 0 |
| 17 | 7674933 | T | C | N | D | TP53 | ENST00000269305 | 0.73973 | 0.91241 | 0.53372 | 0.97158 | 0.9506 | 0.4951484232 | 0.9705 | 0.9905883074 | 0.276718 | 5.434267555 | 0 |
| 17 | 7674933 | T | G | N | H | TP53 | ENST00000269305 | 0.71618 | 0.9012 | 0.49458 | 0.97225 | 0.95426 | 0.5483905062 | 0.97284 | 0.995994091 | 0.301717 | 5.181601275 | 0 |
| 17 | 7674933 | T | G | N | H | TP53 | ENST00000269305 | 0.71618 | 0.9012 | 0.49458 | 0.97225 | 0.95426 | 0.5483905062 | 0.97284 | 0.995994091 | 0.301717 | 5.181601275 | 0 |
| 17 | 7674933 | T | G | N | H | TP53 | ENST00000269305 | 0.71618 | 0.9012 | 0.49458 | 0.97225 | 0.95426 | 0.5483905062 | 0.97284 | 0.995994091 | 0.301717 | 5.181601275 | 0 |
| 17 | 7674935 | C | A | G | V | TP53 | ENST00000269305 | 0.73845 | 0.92355 | 0.61398 | 0.97043 | 0.97215 | 0.7142070089 | 0.96509 | 0.9987642765 | 0.550106 | 8.225197885 | 1 |
| 17 | 7674935 | C | A | G | V | TP53 | ENST00000269305 | 0.73845 | 0.92355 | 0.61398 | 0.97043 | 0.97215 | 0.7142070089 | 0.96509 | 0.9987642765 | 0.550106 | 8.225197885 | 1 |
| 17 | 7674935 | C | A | G | V | TP53 | ENST00000269305 | 0.73845 | 0.92355 | 0.61398 | 0.97043 | 0.97215 | 0.7142070089 | 0.96509 | 0.9987642765 | 0.550106 | 8.225197885 | 1 |
| 17 | 7674935 | C | G | G | A | TP53 | ENST00000269305 | 0.57801 | 0.90636 | 0.60378 | 0.96073 | 0.9681 | 0.5569776551 | 0.94059 | 0.9893766642 | 0.383107 | 5.154966497 | 0 |
| 17 | 7674935 | C | G | G | A | TP53 | ENST00000269305 | 0.57801 | 0.90636 | 0.60378 | 0.96073 | 0.9681 | 0.5569776551 | 0.94059 | 0.9893766642 | 0.383107 | 5.154966497 | 0 |
| 17 | 7674935 | C | G | G | A | TP53 | ENST00000269305 | 0.57801 | 0.90636 | 0.60378 | 0.96073 | 0.9681 | 0.5569776551 | 0.94059 | 0.9893766642 | 0.383107 | 5.154966497 | 0 |
| 17 | 7674935 | C | T | G | E | TP53 | ENST00000269305 | 0.73765 | 0.92717 | 0.68679 | 0.96279 | 0.96517 | 0.6469917189 | 0.95255 | 0.9988040924 | 0.465056 | 4.928340483 | 1 |
| 17 | 7674935 | C | T | G | E | TP53 | ENST00000269305 | 0.73765 | 0.92717 | 0.68679 | 0.96279 | 0.96517 | 0.6469917189 | 0.95255 | 0.9988040924 | 0.465056 | 4.928340483 | 0 |
| 17 | 7674935 | C | T | G | E | TP53 | ENST00000269305 | 0.73765 | 0.92717 | 0.68679 | 0.96279 | 0.96517 | 0.6469917189 | 0.95255 | 0.9988040924 | 0.465056 | 4.928340483 | 0 |
| 17 | 7674936 | C | G | G | R | TP53 | ENST00000269305 | 0.88473 | 0.91823 | 0.70343 | 0.9871 | 0.96627 | 0.6808809953 | 0.97292 | 0.9948759675 | 0.394092 | 7.989272575 | 0 |
| 17 | 7674936 | C | G | G | R | TP53 | ENST00000269305 | 0.88473 | 0.91823 | 0.70343 | 0.9871 | 0.96627 | 0.6808809953 | 0.97292 | 0.9948759675 | 0.394092 | 7.989272575 | 0 |
| 17 | 7674936 | C | G | G | R | TP53 | ENST00000269305 | 0.88473 | 0.91823 | 0.70343 | 0.9871 | 0.96627 | 0.6808809953 | 0.97292 | 0.9948759675 | 0.394092 | 7.989272575 | 0 |
| 17 | 7674936 | C | T | G | R | TP53 | ENST00000269305 | 0.88892 | 0.91823 | 0.70343 | 0.9874 | 0.96627 | 0.6808809953 | 0.97265 | 0.9957427382 | 0.39409 | 7.989272575 | 0 |
| 17 | 7674936 | C | T | G | R | TP53 | ENST00000269305 | 0.88892 | 0.91823 | 0.70343 | 0.9874 | 0.96627 | 0.6808809953 | 0.97265 | 0.9957427382 | 0.39409 | 7.989272575 | 0 |
| 17 | 7674937 | T | A | E | D | TP53 | ENST00000269305 | 0.72483 | 0.88401 | 0.49761 | 0.98598 | 0.94469 | 0.5347098097 | 0.99492 | 0.9763873816 | 0.499399 | 4.992187778 | 0 |
| 17 | 7674937 | T | A | E | D | TP53 | ENST00000269305 | 0.72483 | 0.88401 | 0.49761 | 0.98598 | 0.94469 | 0.5347098097 | 0.99492 | 0.9763873816 | 0.499399 | 4.992187778 | 0 |
| 17 | 7674937 | T | A | E | D | TP53 | ENST00000269305 | 0.72483 | 0.88401 | 0.49761 | 0.98598 | 0.94469 | 0.5347098097 | 0.99492 | 0.9763873816 | 0.499399 | 4.992187778 | 0 |
| 17 | 7674937 | T | G | E | D | TP53 | ENST00000269305 | 0.73301 | 0.88401 | 0.49761 | 0.98581 | 0.94469 | 0.5347098097 | 0.99494 | 0.9763873816 | 0.499458 | 4.992187778 | 0 |
| 17 | 7674938 | T | A | E | V | TP53 | ENST00000269305 | 0.87503 | 0.91578 | 0.48034 | 0.98366 | 0.9822 | 0.6854144199 | 0.99335 | 0.9978922009 | 0.570268 | 6.449780903 | 0 |
| 17 | 7674938 | T | A | E | V | TP53 | ENST00000269305 | 0.87503 | 0.91578 | 0.48034 | 0.98366 | 0.9822 | 0.6854144199 | 0.99335 | 0.9978922009 | 0.570268 | 6.449780903 | 0 |
| 17 | 7674938 | T | A | E | V | TP53 | ENST00000269305 | 0.87503 | 0.91578 | 0.48034 | 0.98366 | 0.9822 | 0.6854144199 | 0.99335 | 0.9978922009 | 0.570268 | 6.449780903 | 0 |
| 17 | 7674938 | T | C | E | G | TP53 | ENST00000269305 | 0.72588 | 0.91619 | 0.44931 | 0.97421 | 0.9781 | 0.743168596 | 0.98851 | 0.9940906167 | 0.545096 | 4.485185148 | 0 |
| 17 | 7674938 | T | C | E | G | TP53 | ENST00000269305 | 0.72588 | 0.91619 | 0.44931 | 0.97421 | 0.9781 | 0.743168596 | 0.98851 | 0.9940906167 | 0.545096 | 4.485185148 | 0 |
| 17 | 7674938 | T | G | E | A | TP53 | ENST00000269305 | 0.84801 | 0.90472 | 0.50355 | 0.9786 | 0.98621 | 0.659922734 | 0.98861 | 0.9972991329 | 0.576373 | 6.498500385 | 0 |
| 17 | 7674938 | T | G | E | A | TP53 | ENST00000269305 | 0.84801 | 0.90472 | 0.50355 | 0.9786 | 0.98621 | 0.659922734 | 0.98861 | 0.9972991329 | 0.576373 | 6.498500385 | 0 |
| 17 | 7674938 | T | G | E | A | TP53 | ENST00000269305 | 0.84801 | 0.90472 | 0.50355 | 0.9786 | 0.98621 | 0.659922734 | 0.98861 | 0.9972991329 | 0.576373 | 6.498500385 | 0 |
| 17 | 7674939 | C | G | E | Q | TP53 | ENST00000269305 | 0.86183 | 0.89582 | 0.48024 | 0.96685 | 0.96846 | 0.6091460105 | 0.98643 | 0.9872708321 | 0.54325 | 6.123175157 | 0 |
| 17 | 7674939 | C | G | E | Q | TP53 | ENST00000269305 | 0.86183 | 0.89582 | 0.48024 | 0.96685 | 0.96846 | 0.6091460105 | 0.98643 | 0.9872708321 | 0.54325 | 6.123175157 | 0 |
| 17 | 7674939 | C | G | E | Q | TP53 | ENST00000269305 | 0.86183 | 0.89582 | 0.48024 | 0.96685 | 0.96846 | 0.6091460105 | 0.98643 | 0.9872708321 | 0.54325 | 6.123175157 | 0 |
| 17 | 7674939 | C | T | E | K | TP53 | ENST00000269305 | 0.91209 | 0.91136 | 0.61811 | 0.97405 | 0.97624 | 0.7561271946 | 0.99373 | 0.9969528913 | 0.578534 | 5.802649073 | 0 |
| 17 | 7674939 | C | T | E | K | TP53 | ENST00000269305 | 0.91209 | 0.91136 | 0.61811 | 0.97405 | 0.97624 | 0.7561271946 | 0.99373 | 0.9969528913 | 0.578534 | 5.802649073 | 0 |
| 17 | 7674941 | A | G | V | A | TP53 | ENST00000269305 | 0.92007 | 0.91917 | 0.58038 | 0.97689 | 0.94431 | 0.556147124 | 0.98338 | 0.9806013528 | 0.417873 | 5.690116663 | 0 |
| 17 | 7674941 | A | T | V | E | TP53 | ENST00000269305 | 0.93619 | 0.94947 | 0.56436 | 0.98506 | 0.97141 | 0.8310347778 | 0.99723 | 0.9985249639 | 0.563636 | 8.760438011 | 1 |
| 17 | 7674942 | C | G | V | L | TP53 | ENST00000269305 | 0.51507 | 0.89498 | 0.5576 | 0.95847 | 0.90668 | 0.5432432791 | 0.96963 | 0.9532197118 | 0.345106 | 6.323163372 | 0 |
| 17 | 7674944 | C | A | R | L | TP53 | ENST00000269305 | 0.90079 | 0.92201 | 0.57632 | 0.98302 | 0.98477 | 0.8402241616 | 0.9973 | 0.99817276 | 0.57174 | 6.031151296 | 0 |
| 17 | 7674944 | C | G | R | P | TP53 | ENST00000269305 | 0.91396 | 0.93818 | 0.58656 | 0.98016 | 0.98764 | 0.9420639773 | 0.99386 | 0.9985722303 | 0.576167 | 8.283702361 | 1 |
| 17 | 7674945 | G | C | R | G | TP53 | ENST00000269305 | 0.89543 | 0.92614 | 0.5291 | 0.98397 | 0.97141 | 0.8988550156 | 0.99716 | 0.9976861477 | 0.541566 | 8.283702361 | 0 |
| 17 | 7674946 | G | C | I | M | TP53 | ENST00000269305 | 0.81292 | 0.89928 | 0.58483 | 0.95943 | 0.91736 | 0.5958261427 | 0.97246 | 0.9864000082 | 0.358423 | 5.41842787 | 0 |
| 17 | 7674947 | A | C | I | S | TP53 | ENST00000269305 | 0.85353 | 0.93938 | 0.64847 | 0.9891 | 0.95972 | 0.923171261 | 0.99836 | 0.9988412261 | 0.570381 | 8.837713275 | 1 |
| 17 | 7674947 | A | G | I | T | TP53 | ENST00000269305 | 0.78962 | 0.92429 | 0.75045 | 0.98355 | 0.95462 | 0.79778865 | 0.98683 | 0.99941504 | 0.525761 | 8.837713275 | 1 |
| 17 | 7674947 | A | T | I | N | TP53 | ENST00000269305 | 0.84658 | 0.9568 | 0.73171 | 0.98918 | 0.9506 | 0.8994668902 | 0.9853 | 0.9990060925 | 0.437222 | 8.837713275 | 1 |
| 17 | 7674948 | T | A | I | F | TP53 | ENST00000269305 | 0.79034 | 0.94684 | 0.64064 | 0.97594 | 0.97401 | 0.781161985 | 0.98837 | 0.9961848855 | 0.533913 | 6.947676296 | 1 |
| 17 | 7674948 | T | C | I | V | TP53 | ENST00000269305 | 0.64641 | 0.87344 | 0.53048 | 0.95811 | 0.9045 | 0.2789768799 | 0.92808 | 0.8504542112 | 0.277769 | 4.965741166 | 0 |
| 17 | 7674948 | T | G | I | L | TP53 | ENST00000269305 | 0.59704 | 0.8964 | 0.62654 | 0.9694 | 0.90406 | 0.5218755637 | 0.95649 | 0.923396945 | 0.319913 | 4.796714177 | 0 |
| 17 | 7674950 | A | C | L | R | TP53 | ENST00000269305 | 0.94193 | 0.9398 | 0.75312 | 0.95767 | 0.98477 | 0.909369283 | 0.99642 | 0.9922944307 | 0.571373 | 9.43677829 | 1 |
| 17 | 7674950 | A | G | L | P | TP53 | ENST00000269305 | 0.94294 | 0.9489 | 0.79317 | 0.96037 | 0.98621 | 0.9227325572 | 0.99829 | 0.9986838698 | 0.584663 | 9.43677829 | 1 |
| 17 | 7674950 | A | T | L | H | TP53 | ENST00000269305 | 0.93638 | 0.92635 | 0.75803 | 0.95929 | 0.97923 | 0.7939757673 | 0.9932 | 0.998496294 | 0.575042 | 9.43677829 | 1 |
| 17 | 7674951 | G | A | L | F | TP53 | ENST00000269305 | 0.8761 | 0.90712 | 0.66109 | 0.97484 | 0.95462 | 0.5524234914 | 0.99508 | 0.9927470088 | 0.554285 | 5.527336725 | 1 |
| 17 | 7674951 | G | C | L | V | TP53 | ENST00000269305 | 0.84043 | 0.89818 | 0.61626 | 0.9773 | 0.94802 | 0.4056207661 | 0.98889 | 0.9816762209 | 0.478331 | 5.996455853 | 0 |
| 17 | 7674951 | G | T | L | I | TP53 | ENST00000269305 | 0.84665 | 0.90673 | 0.67856 | 0.97787 | 0.93201 | 0.2752461689 | 0.9707 | 0.9605309963 | 0.41203 | 5.825958521 | 0 |
| 17 | 7674952 | A | C | H | Q | TP53 | ENST00000269305 | 0.65972 | 0.89783 | 0.74659 | 0.96442 | 0.93824 | 0.8002416293 | 0.99604 | 0.9951312542 | 0.520113 | 6.556410006 | 0 |
| 17 | 7674953 | T | A | H | L | TP53 | ENST00000269305 | 0.85771 | 0.9223 | 0.73773 | 0.97681 | 0.95935 | 0.8478920664 | 0.9976 | 0.999766767 | 0.582967 | 8.475558754 | 1 |
| 17 | 7674953 | T | C | H | R | TP53 | ENST00000269305 | 0.81214 | 0.92061 | 0.77585 | 0.97719 | 0.96664 | 0.8994405324 | 0.99763 | 0.9990561604 | 0.554369 | 7.675627177 | 1 |
| 17 | 7674953 | T | G | H | P | TP53 | ENST00000269305 | 0.85624 | 0.94597 | 0.70736 | 0.97614 | 0.96153 | 0.9184214522 | 0.99654 | 0.999725163 | 0.577785 | 8.475558754 | 1 |
| 17 | 7674954 | G | A | H | Y | TP53 | ENST00000269305 | 0.90458 | 0.91476 | 0.77238 | 0.96592 | 0.96773 | 0.8429484027 | 0.98946 | 0.9989527464 | 0.5603 | 5.865844991 | 1 |
| 17 | 7674954 | G | C | H | D | TP53 | ENST00000269305 | 0.92904 | 0.93335 | 0.76042 | 0.96803 | 0.98071 | 0.9065533131 | 0.9943 | 0.9994570613 | 0.582403 | 8.475558754 | 1 |
| 17 | 7674954 | G | T | H | N | TP53 | ENST00000269305 | 0.91199 | 0.90843 | 0.76791 | 0.94864 | 0.96334 | 0.8194750823 | 0.9754 | 0.9988184571 | 0.386962 | 8.475558754 | 1 |
| 17 | 7674955 | C | A | Q | H | TP53 | ENST00000269305 | 0.3179 | 0.69443 | 0.27703 | 0.97795 | 0.80653 | 0.4164719144 | 0.88447 | 0.531229496 | 0.189051 | 4.878923982 | 0 |
| 17 | 7674955 | C | G | Q | H | TP53 | ENST00000269305 | 0.32346 | 0.69443 | 0.27703 | 0.97697 | 0.80831 | 0.4164719144 | 0.88403 | 0.4207128286 | 0.189049 | 4.878923982 | 0 |
| 17 | 7674956 | T | A | Q | L | TP53 | ENST00000269305 | 0.85822 | 0.88393 | 0.23164 | 0.99406 | 0.9014 | 0.5020094556 | 0.9063 | 0.9436573386 | 0.242767 | 6.927611917 | 0 |
| 17 | 7674956 | T | C | Q | R | TP53 | ENST00000269305 | 0.68915 | 0.89749 | 0.2717 | 0.99416 | 0.88259 | 0.4153825154 | 0.87566 | 0.8623945088 | 0.23366 | 5.547710739 | 0 |
| 17 | 7674956 | T | G | Q | P | TP53 | ENST00000269305 | 0.74906 | 0.91936 | 0.23719 | 0.99254 | 0.92316 | 0.7622894295 | 0.90944 | 0.8729514608 | 0.314519 | 6.922176514 | 0 |

| | | | | | | | | | | | | | | | | |
|---|---|---|---|---|---|---|---|---|---|---|---|---|---|---|---|---|
| 17 | 7674957 | G | C | Q | E | TP53 | ENST00000269305 | 0.45193 | 0.86723 | 0.24729 | 0.95776 | 0.8199 | 0.399645955 | 0.86118 | 0.7614748478 | 0.202527 | 4.997568177 | 0 |
| 17 | 7674957 | G | T | Q | K | TP53 | ENST00000269305 | 0.50661 | 0.90718 | 0.28473 | 0.97622 | 0.86453 | 0.4461203044 | 0.88413 | 0.7731629014 | 0.231761 | 5.777552239 | 0 |
| 17 | 7674959 | G | A | P | L | TP53 | ENST00000269305 | 0.91729 | 0.89976 | 0.34237 | 0.9843 | 0.91442 | 0.528584247 | 0.92766 | 0.9613342881 | 0.342845 | 6.282076449 | 0 |
| 17 | 7674959 | G | C | P | R | TP53 | ENST00000269305 | 0.56117 | 0.90412 | 0.34116 | 0.85253 | 0.85274 | 0.4995068455 | 0.88227 | 0.5849393606 | 0.108709 | 5.129428433 | 0 |
| 17 | 7674959 | G | T | P | H | TP53 | ENST00000269305 | 0.91007 | 0.91454 | 0.30936 | 0.97986 | 0.93436 | 0.4128418697 | 0.9269 | 0.9811261296 | 0.361416 | 7.033438991 | 0 |
| 17 | 7674960 | G | A | P | S | TP53 | ENST00000269305 | 0.36008 | 0.89849 | 0.23833 | 0.97953 | 0.90318 | 0.4500106767 | 0.9419 | 0.8951999545 | 0.202427 | 5.611667505 | 0 |
| 17 | 7674960 | G | C | P | A | TP53 | ENST00000269305 | 0.19532 | 0.89354 | 0.24461 | 0.97622 | 0.84267 | 0.2985044546 | 0.90284 | 0.4077565074 | 0.173265 | 4.267526704 | 0 |
| 17 | 7674960 | G | T | P | T | TP53 | ENST00000269305 | 0.43297 | 0.91059 | 0.27227 | 0.98515 | 0.90537 | 0.4621169032 | 0.95611 | 0.9593343735 | 0.300041 | 6.58310455 | 0 |
| 17 | 7674962 | G | A | P | L | TP53 | ENST00000269305 | 0.92555 | 0.86582 | 0.66058 | 0.97696 | 0.94469 | 0.5169395292 | 0.95635 | 0.9954833984 | 0.37133 | 6.515109201 | 1 |
| 17 | 7674962 | G | T | P | H | TP53 | ENST00000269305 | 0.78859 | 0.91425 | 0.64149 | 0.95882 | 0.90711 | 0.47064704 | 0.91334 | 0.9804931283 | 0.253747 | 5.265516218 | 0 |
| 17 | 7674963 | G | A | P | S | TP53 | ENST00000269305 | 0.86432 | 0.89766 | 0.58364 | 0.97422 | 0.93083 | 0.4695845772 | 0.95526 | 0.9828373194 | 0.306569 | 5.810256102 | 0 |
| 17 | 7674963 | G | C | P | A | TP53 | ENST00000269305 | 0.50792 | 0.88625 | 0.5728 | 0.96487 | 0.9182 | 0.3702240985 | 0.93748 | 0.9692134261 | 0.273556 | 5.336406937 | 0 |
| 17 | 7674963 | G | T | P | T | TP53 | ENST00000269305 | 0.82116 | 0.90935 | 0.62579 | 0.97926 | 0.94394 | 0.4842908852 | 0.96467 | 0.9950730205 | 0.351808 | 8.218052907 | 1 |
| 17 | 7674965 | G | A | A | V | TP53 | ENST00000269305 | 0.84028 | 0.8967 | 0.39721 | 0.97286 | 0.93162 | 0.3732544422 | 0.95741 | 0.5499962879 | 0.246169 | 5.567302411 | 0 |
| 17 | 7674965 | G | C | A | G | TP53 | ENST00000269305 | 0.94006 | 0.89783 | 0.35275 | 0.9766 | 0.91694 | 0.3273122607 | 0.91736 | 0.974219203 | 0.362731 | 7.383210453 | 0 |
| 17 | 7674965 | G | T | A | D | TP53 | ENST00000269305 | 0.80692 | 0.94218 | 0.50066 | 0.97507 | 0.94205 | 0.5780859518 | 0.97105 | 0.9765934944 | 0.50627 | 5.078244504 | 0 |
| 17 | 7674966 | C | A | A | S | TP53 | ENST00000269305 | 0.69917 | 0.88566 | 0.33441 | 0.96399 | 0.8933 | 0.2709971025 | 0.87252 | 0.9516767263 | 0.325239 | 6.060725369 | 0 |
| 17 | 7674966 | C | G | A | P | TP53 | ENST00000269305 | 0.75392 | 0.92717 | 0.44372 | 0.95889 | 0.90274 | 0.6172050127 | 0.94688 | 0.9831594229 | 0.393976 | 6.136305671 | 0 |
| 17 | 7674966 | C | T | A | T | TP53 | ENST00000269305 | 0.61477 | 0.89319 | 0.38102 | 0.97629 | 0.90581 | 0.320196657 | 0.92212 | 0.9716159105 | 0.364085 | 4.902223837 | 0 |
| 17 | 7674968 | A | C | L | R | TP53 | ENST00000269305 | 0.50648 | 0.86227 | 0.40731 | 0.95489 | 0.86302 | 0.4078213361 | 0.95275 | 0.1469866037 | 0.207848 | 5.90413054 | 0 |
| 17 | 7674968 | A | G | L | P | TP53 | ENST00000269305 | 0.40156 | 0.94855 | 0.43968 | 0.97082 | 0.85274 | 0.5115343293 | 0.97375 | 0.3233830631 | 0.299311 | 4.283569644 | 0 |
| 17 | 7674968 | A | T | L | Q | TP53 | ENST00000269305 | 0.67461 | 0.89969 | 0.33126 | 0.95862 | 0.83339 | 0.3653487667 | 0.94886 | 0.3925437033 | 0.232423 | 6.786536993 | 0 |
| 17 | 7674969 | G | C | L | V | TP53 | ENST00000269305 | 0.22001 | 0.87883 | 0.26288 | 0.93826 | 0.82836 | 0.1690896985 | 0.93329 | 0.3125796914 | 0.197119 | 4.852974152 | 0 |
| 17 | 7674969 | G | T | L | M | TP53 | ENST00000269305 | 0.50811 | 0.89172 | 0.2931 | 0.90361 | 0.76881 | 0.1557713042 | 0.92848 | 0.5116526484 | 0.197604 | 6.256765836 | 0 |
| 17 | 7674971 | C | A | G | V | TP53 | ENST00000269305 | 0.64961 | 0.91936 | 0.40064 | 0.98223 | 0.87733 | 0.4988948469 | 0.89971 | 0.9941698313 | 0.314281 | 5.707582178 | 0 |
| 17 | 7674971 | C | G | G | A | TP53 | ENST00000269305 | 0.44089 | 0.71722 | 0.39516 | 0.92082 | 0.84906 | 0.2899008218 | 0.92476 | 0.6865307689 | 0.226758 | 3.457567969 | 0 |
| 17 | 7674971 | C | T | G | D | TP53 | ENST00000269305 | 0.42412 | 0.81321 | 0.52861 | 0.9514 | 0.85689 | 0.3521097349 | 0.91488 | 0.6161835194 | 0.243721 | 4.473017846 | 0 |
| 17 | 7675053 | C | A | G | C | TP53 | ENST00000269305 | 0.4074 | 0.90322 | 0.44452 | 0.9651 | 0.89558 | 0.4977534623 | 0.91324 | 0.9886997342 | 0.293484 | 5.382552297 | 0 |
| 17 | 7675053 | C | G | G | R | TP53 | ENST00000269305 | 0.6962 | 0.91644 | 0.51151 | 0.97175 | 0.8933 | 0.4308480481 | 0.89508 | 0.975055933 | 0.296313 | 6.835122041 | 0 |
| 17 | 7675053 | C | T | G | S | TP53 | ENST00000269305 | 0.36153 | 0.79054 | 0.38085 | 0.9316 | 0.86902 | 0.2811754255 | 0.84417 | 0.8525909185 | 0.216163 | 4.656824284 | 0 |
| 17 | 7675054 | A | C | D | E | TP53 | ENST00000269305 | 0.26322 | 0.83308 | 0.49194 | 0.96369 | 0.86603 | 0.1611862127 | 0.9275 | 0.8450309038 | 0.1682 | 3.612783673 | 0 |
| 17 | 7675054 | A | T | D | E | TP53 | ENST00000269305 | 0.26063 | 0.83308 | 0.49194 | 0.96565 | 0.86603 | 0.1611862127 | 0.92754 | 0.7202209234 | 0.1682 | 3.612783673 | 0 |
| 17 | 7675055 | T | A | D | V | TP53 | ENST00000269305 | 0.80369 | 0.93612 | 0.44856 | 0.98562 | 0.93554 | 0.5253373814 | 0.97766 | 0.9973305464 | 0.395549 | 7.66191602 | 0 |
| 17 | 7675055 | T | C | D | G | TP53 | ENST00000269305 | 0.7794 | 0.92707 | 0.46503 | 0.97826 | 0.88401 | 0.3940530283 | 0.94673 | 0.9642323256 | 0.234519 | 6.652554055 | 0 |
| 17 | 7675055 | T | G | D | A | TP53 | ENST00000269305 | 0.75989 | 0.91773 | 0.45716 | 0.97797 | 0.92356 | 0.4266505542 | 0.971 | 0.9952943921 | 0.335936 | 6.102115504 | 0 |
| 17 | 7675056 | C | A | D | Y | TP53 | ENST00000269305 | 0.47823 | 0.92151 | 0.54067 | 0.98068 | 0.94129 | 0.5124360428 | 0.95264 | 0.9943408966 | 0.367802 | 7.666669351 | 0 |
| 17 | 7675056 | C | G | D | H | TP53 | ENST00000269305 | 0.47288 | 0.91679 | 0.55566 | 0.98124 | 0.88963 | 0.462398273 | 0.95171 | 0.9981645942 | 0.325572 | 5.848457822 | 0 |
| 17 | 7675056 | C | T | D | N | TP53 | ENST00000269305 | 0.33489 | 0.8899 | 0.52886 | 0.96325 | 0.80536 | 0.2437277138 | 0.92506 | 0.791811049 | 0.00709879 | 4.168816643 | 0 |
| 17 | 7675057 | G | C | S | R | TP53 | ENST00000269305 | 0.12196 | 0.87701 | 0.25143 | 0.98447 | 0.79818 | 0.1824082444 | 0.87964 | 0.5649839044 | 0.122461 | 5.523815496 | 0 |
| 17 | 7675057 | G | T | S | R | TP53 | ENST00000269305 | 0.11941 | 0.87701 | 0.25143 | 0.98249 | 0.79818 | 0.1824082444 | 0.8806 | 0.6414471865 | 0.122456 | 5.523815496 | 0 |
| 17 | 7675058 | C | A | S | I | TP53 | ENST00000269305 | 0.27251 | 0.71244 | 0.23183 | 0.98024 | 0.75485 | 0.2125974642 | 0.92635 | 0.6856076717 | 0.156195 | 4.479725945 | 0 |
| 17 | 7675058 | C | G | S | T | TP53 | ENST00000269305 | 0.08916 | 0.59857 | 0.20173 | 0.97765 | 0.65673 | 0.138610929 | 0.87676 | 0.02965255387 | 0.114732 | 4.217974531 | 0 |
| 17 | 7675058 | C | T | S | N | TP53 | ENST00000269305 | 0.06787 | 0.55202 | 0.234 | 0.90585 | 0.65077 | 0.09442896106 | 0.74873 | 0.02032452974 | -0.0718235 | 2.317565805 | 0 |
| 17 | 7675059 | T | A | S | C | TP53 | ENST00000269305 | 0.30025 | 0.87013 | 0.22526 | 0.97633 | 0.73589 | 0.2286950873 | 0.93859 | 0.8529621363 | 0.175391 | 6.547948388 | 0 |
| 17 | 7675059 | T | C | S | G | TP53 | ENST00000269305 | 0.10011 | 0.5496 | 0.18648 | 0.92003 | 0.78411 | 0.105877125 | 0.81391 | 0.02831061661 | 0.108347 | 2.610433779 | 0 |
| 17 | 7675059 | T | G | S | R | TP53 | ENST00000269305 | 0.20375 | 0.87701 | 0.25143 | 0.98315 | 0.77141 | 0.1824082444 | 0.90366 | 0.2937707007 | 0.160291 | 5.523815496 | 0 |
| 17 | 7675060 | A | C | D | E | TP53 | ENST00000269305 | 0.03345 | 0.55827 | 0.48586 | 0.95804 | 0.82046 | 0.167570478 | 0.94396 | 0.1325591356 | 0.111481 | 2.299509279 | 0 |
| 17 | 7675060 | A | T | D | E | TP53 | ENST00000269305 | 0.03195 | 0.55827 | 0.48586 | 0.9581 | 0.82046 | 0.167570478 | 0.94447 | 0.178870514 | 0.111465 | 2.299509279 | 0 |
| 17 | 7675061 | T | A | D | V | TP53 | ENST00000269305 | 0.68267 | 0.93017 | 0.44637 | 0.97369 | 0.95133 | 0.4861392197 | 0.97144 | 0.9901211262 | 0.387739 | 5.784386818 | 0 |
| 17 | 7675061 | T | C | D | G | TP53 | ENST00000269305 | 0.41493 | 0.91495 | 0.46243 | 0.96842 | 0.87733 | 0.4149266253 | 0.98498 | 0.9473131895 | 0.319556 | 4.679638143 | 0 |
| 17 | 7675061 | T | G | D | A | TP53 | ENST00000269305 | 0.66067 | 0.86874 | 0.45806 | 0.96817 | 0.91271 | 0.4205226642 | 0.94665 | 0.9772455096 | 0.31486 | 6.42693251 | 0 |
| 17 | 7675062 | C | A | D | Y | TP53 | ENST00000269305 | 0.22648 | 0.91961 | 0.52823 | 0.92314 | 0.90007 | 0.5098198079 | 0.95083 | 0.9700231552 | 0.366219 | 6.375901524 | 0 |
| 17 | 7675062 | C | G | D | H | TP53 | ENST00000269305 | 0.3406 | 0.91229 | 0.54864 | 0.92678 | 0.8933 | 0.3922021738 | 0.92843 | 0.9760554433 | 0.334544 | 5.222895242 | 0 |
| 17 | 7675062 | C | T | D | N | TP53 | ENST00000269305 | 0.14127 | 0.86141 | 0.52133 | 0.91751 | 0.74164 | 0.2029519287 | 0.85668 | 0.8837015629 | 0.139097 | 5.098348548 | 0 |
| 17 | 7675064 | G | A | S | L | TP53 | ENST00000269305 | 0.30827 | 0.86128 | 0.26365 | 0.98234 | 0.81933 | 0.2388586203 | 0.88877 | 0.4867640436 | 0.187426 | 5.693716054 | 0 |
| 17 | 7675065 | A | C | S | A | TP53 | ENST00000269305 | 0.22998 | 0.73465 | 0.24166 | 0.96776 | 0.78286 | 0.1206759055 | 0.94491 | 0.1285410821 | 0.15864 | 4.300389077 | 0 |
| 17 | 7675065 | A | G | S | P | TP53 | ENST00000269305 | 0.26026 | 0.72766 | 0.25324 | 0.96429 | 0.75822 | 0.4220433031 | 0.94129 | 0.06829273701 | 0.149823 | 3.805158428 | 0 |
| 17 | 7675066 | G | C | C | W | TP53 | ENST00000269305 | 0.60054 | 0.93062 | 0.26082 | 0.97882 | 0.87394 | 0.4718377112 | 0.9736 | 0.9788378477 | 0.28869 | 8.373960609 | 0 |
| 17 | 7675067 | C | A | C | F | TP53 | ENST00000269305 | 0.4189 | 0.94179 | 0.23405 | 0.97218 | 0.89285 | 0.371315453 | 0.91599 | 0.987601459 | 0.205236 | 8.373960609 | 0 |
| 17 | 7675067 | C | G | C | S | TP53 | ENST00000269305 | 0.15474 | 0.70438 | 0.19841 | 0.88387 | 0.74517 | 0.2481436026 | 0.81833 | 0.1291141063 | 0.138367 | 3.490216292 | 0 |
| 17 | 7675067 | C | T | C | Y | TP53 | ENST00000269305 | 0.28174 | 0.95159 | 0.30757 | 0.96826 | 0.86149 | 0.4003484154 | 0.9164 | 0.968490541 | 0.199388 | 7.299725457 | 0 |
| 17 | 7675068 | A | C | C | G | TP53 | ENST00000269305 | 0.35344 | 0.89599 | 0.18498 | 0.95101 | 0.86653 | 0.3557951942 | 0.90505 | 0.8858264089 | 0.155635 | 5.787214441 | 0 |
| 17 | 7675068 | A | G | C | R | TP53 | ENST00000269305 | 0.24224 | 0.95226 | 0.28928 | 0.96995 | 0.86653 | 0.4172668336 | 0.90535 | 0.8363568187 | 0.167945 | 4.570923563 | 0 |
| 17 | 7675068 | A | T | C | S | TP53 | ENST00000269305 | 0.181 | 0.70438 | 0.19841 | 0.87486 | 0.75687 | 0.2481436026 | 0.86804 | 0.1487656236 | 0.115008 | 3.490216292 | 0 |
| 17 | 7675070 | C | A | R | L | TP53 | ENST00000269305 | 0.66327 | 0.90877 | 0.33893 | 0.97501 | 0.88401 | 0.4173687117 | 0.91588 | 0.9233490825 | 0.28437 | 6.199477197 | 0 |
| 17 | 7675070 | C | G | R | P | TP53 | ENST00000269305 | 0.72244 | 0.93746 | 0.36456 | 0.98472 | 0.9703 | 0.6602543956 | 0.98142 | 0.9797382355 | 0.469839 | 9.209716555 | 1 |
| 17 | 7675070 | C | T | R | H | TP53 | ENST00000269305 | 0.69836 | 0.91945 | 0.33537 | 0.98263 | 0.93436 | 0.2904626525 | 0.96128 | 0.803822944 | 0.301127 | 7.15544788 | 0 |
| 17 | 7675071 | G | A | R | C | TP53 | ENST00000269305 | 0.655 | 0.82037 | 0.40732 | 0.97259 | 0.96919 | 0.4205410485 | 0.98312 | 0.9550928473 | 0.492215 | 5.309173718 | 0 |
| 17 | 7675071 | G | C | R | G | TP53 | ENST00000269305 | 0.89867 | 0.92397 | 0.31328 | 0.97551 | 0.96737 | 0.4114262074 | 0.937 | 0.96473068 | 0.342749 | 6.594763875 | 0 |
| 17 | 7675071 | G | T | R | S | TP53 | ENST00000269305 | 0.57315 | 0.90791 | 0.32152 | 0.9443 | 0.92682 | 0.3852856855 | 0.873 | 0.9534432292 | 0.307554 | 5.209390999 | 0 |
| 17 | 7675072 | C | A | E | D | TP53 | ENST00000269305 | 0.52201 | 0.8844 | 0.53691 | 0.97358 | 0.93475 | 0.3165018747 | 0.93926 | 0.9074921271 | 0.257958 | 8.744466842 | 0 |
| 17 | 7675072 | C | G | E | D | TP53 | ENST00000269305 | 0.52905 | 0.8844 | 0.53691 | 0.97637 | 0.93475 | 0.3165018747 | 0.93948 | 0.9893823862 | 0.257976 | 8.744466842 | 0 |
| 17 | 7675073 | T | A | E | V | TP53 | ENST00000269305 | 0.92588 | 0.91578 | 0.51382 | 0.98567 | 0.98477 | 0.401890069 | 0.95986 | 0.9981815815 | 0.409833 | 7.199703465 | 0 |
| 17 | 7675073 | T | C | E | G | TP53 | ENST00000269305 | 0.93417 | 0.91619 | 0.47959 | 0.98137 | 0.9781 | 0.481109253 | 0.95587 | 0.9987111092 | 0.403359 | 8.656847377 | 0 |
| 17 | 7675073 | T | G | E | A | TP53 | ENST00000269305 | 0.91153 | 0.90472 | 0.53866 | 0.97551 | 0.96627 | 0.3584883266 | 0.95667 | 0.9949948788 | 0.35912 | 6.49841845 | 0 |
| 17 | 7675074 | C | G | E | Q | TP53 | ENST00000269305 | 0.47545 | 0.89599 | 0.51328 | 0.9149 | 0.87394 | 0.2871488872 | 0.87678 | 0.8826995492 | 0.219941 | 5.481039314 | 0 |
| 17 | 7675075 | A | C | H | Q | TP53 | ENST00000269305 | 0.34915 | 0.89783 | 0.54275 | 0.98105 | 0.93162 | 0.779163863 | 0.98034 | 0.9988791347 | 0.474835 | 7.595815899 | 1 |
| 17 | 7675075 | A | T | H | Q | TP53 | ENST00000269305 | 0.34456 | 0.89783 | 0.54275 | 0.98105 | 0.93162 | 0.779163863 | 0.98017 | 0.9988791347 | 0.474833 | 7.595815899 | 1 |
| 17 | 7675076 | T | A | H | L | TP53 | ENST00000269305 | 0.45949 | 0.9223 | 0.53759 | 0.97775 | 0.9659 | 0.8116328638 | 0.95448 | 0.9992964268 | 0.481659 | 4.145977502 | 1 |
| 17 | 7675076 | T | C | H | R | TP53 | ENST00000269305 | 0.71113 | 0.92061 | 0.59069 | 0.97278 | 0.98549 | 0.8845544298 | 0.9765 | 0.9985765219 | 0.539639 | 7.595815899 | 1 |
| 17 | 7675076 | T | G | H | P | TP53 | ENST00000269305 | 0.71345 | 0.94597 | 0.49646 | 0.979 | 0.97661 | 0.8937794805 | 0.96654 | 0.9996702671 | 0.53845 | 7.595815899 | 1 |
| 17 | 7675077 | G | A | H | Y | TP53 | ENST00000269305 | 0.88653 | 0.91476 | 0.58709 | 0.97421 | 0.99006 | 0.8275034715 | 0.99261 | 0.9980379939 | 0.5848 | 7.595815899 | 1 |
| 17 | 7675077 | G | C | H | D | TP53 | ENST00000269305 | 0.91674 | 0.93335 | 0.57127 | 0.96878 | 0.97848 | 0.8909666366 | 0.96576 | 0.9989199638 | 0.567077 | 7.595815899 | 1 |
| 17 | 7675077 | G | T | H | N | TP53 | ENST00000269305 | 0.89555 | 0.90843 | 0.5784 | 0.96956 | 0.97327 | 0.7979842951 | 0.95649 | 0.992550175 | 0.40446 | 6.425810889 | 1 |
| 17 | 7675079 | T | A | H | L | TP53 | ENST00000269305 | 0.86755 | 0.9223 | 0.35434 | 0.99226 | 0.9517 | 0.511078329 | 0.91733 | 0.9994543195 | 0.351088 | 6.161536658 | 0 |
| 17 | 7675079 | T | C | H | R | TP53 | ENST00000269305 | 0.82287 | 0.92061 | 0.39829 | 0.98786 | 0.9659 | 0.5181535009 | 0.93189 | 0.9970743656 | 0.341689 | 7.085767183 | 0 |
| 17 | 7675080 | G | A | H | Y | TP53 | ENST00000269305 | 0.65688 | 0.91476 | 0.40979 | 0.98289 | 0.96153 | 0.5052800487 | 0.93157 | 0.9963124394 | 0.339316 | 5.762290643 | 0 |
| 17 | 7675080 | G | C | H | D | TP53 | ENST00000269305 | 0.70318 | 0.93335 | 0.38754 | 0.98528 | 0.96956 | 0.6063396665 | 0.93879 | 0.9957249165 | 0.461365 | 8.791912812 | 1 |
| 17 | 7675080 | G | T | H | N | TP53 | ENST00000269305 | 0.6689 | 0.90843 | 0.38372 | 0.92427 | 0.90318 | 0.3467291438 | 0.87436 | 0.965094924 | 0.171088 | 5.706645175 | 0 |
| 17 | 7675082 | G | C | P | R | TP53 | ENST00000269305 | 0.82443 | 0.90243 | 0.67656 | 0.97637 | 0.96993 | 0.5716557738 | 0.91384 | 0.9990082383 | 0.477786 | 6.900793173 | 1 |
| 17 | 7675082 | G | T | P | H | TP53 | ENST00000269305 | 0.82576 | 0.81743 | 0.66372 | 0.97359 | 0.95207 | 0.4630958723 | 0.88077 | 0.9977843165 | 0.400682 | 5.903078065 | 0 |
| 17 | 7675083 | G | A | P | S | TP53 | ENST00000269305 | 0.87801 | 0.89247 | 0.60433 | 0.95559 | 0.96407 | 0.4911926247 | 0.89953 | 0.99913764 | 0.408658 | 5.186151817 | 0 |
| 17 | 7675083 | G | C | P | A | TP53 | ENST00000269305 | 0.82467 | 0.86902 | 0.59423 | 0.96063 | 0.96008 | 0.3578030661 | 0.87875 | 0.9987811446 | 0.400416 | 5.589212801 | 0 |
| 17 | 7675083 | G | T | P | T | TP53 | ENST00000269305 | 0.85713 | 0.90775 | 0.64604 | 0.95906 | 0.96444 | 0.4673261 | 0.90902 | 0.99913764 | 0.42346 | 6.458329394 | 0 |
| 17 | 7675084 | G | C | C | W | TP53 | ENST00000269305 | 0.67851 | 0.93139 | 0.79918 | 0.99627 | 0.95899 | 0.9031624299 | 0.97413 | 0.9997493625 | 0.472628 | 8.245307739 | 1 |
| 17 | 7675085 | C | A | C | F | TP53 | ENST00000269305 | 0.76832 | 0.94333 | 0.74394 | 0.98805 | 0.99105 | 0.8906092507 | 0.98539 | 0.9996084571 | 0.569042 | 8.245307739 | 1 |
| 17 | 7675085 | C | G | C | S | TP53 | ENST00000269305 | 0.72756 | 0.79536 | 0.73188 | 0.97822 | 0.98904 | 0.8776730852 | 0.98503 | 0.9993796349 | 0.552205 | 8.245307739 | 1 |
| 17 | 7675086 | A | C | C | G | TP53 | ENST00000269305 | 0.90223 | 0.91737 | 0.69342 | 0.98774 | 0.98938 | 0.8951998526 | 0.98521 | 0.9995525479 | 0.579978 | 7.511338597 | 1 |
| 17 | 7675086 | A | G | C | R | TP53 | ENST00000269305 | 0.91111 | 0.95605 | 0.83255 | 0.98645 | 0.98728 | 0.9127916743 | 0.98002 | 0.9999172688 | 0.586306 | 7.062580419 | 1 |
| 17 | 7675086 | A | T | C | S | TP53 | ENST00000269305 | 0.86382 | 0.79536 | 0.73188 | 0.96893 | 0.98938 | 0.8776730852 | 0.98797 | 0.9993632436 | 0.564236 | 8.245307739 | 1 |
| 17 | 7675088 | C | A | R | L | TP53 | ENST00000269305 | 0.86176 | 0.91816 | 0.61279 | 0.97533 | 0.99386 | 0.8479766894 | 0.99675 | 0.9997362494 | 0.564459 | 7.12957347 | 0 |

| | | | | | | | | | | | | | | | | |
|---|---|---|---|---|---|---|---|---|---|---|---|---|---|---|---|---|
| 17 | 7675088 | C | G | R | P | TP53 | ENST00000269305 | 0.88825 | 0.93777 | 0.62424 | 0.97351 | 0.99552 | 0.9196423233 | 0.99437 | 0.9991239905 | 0.56446 | 6.192298646 | 0 |
| 17 | 7675088 | C | T | R | H | TP53 | ENST00000269305 | 0.5495 | 0.77718 | 0.60661 | 0.94427 | 0.98071 | 0.784526593 | 0.99718 | 0.9928691983 | 0.545774 | 3.950892957 | 1 |
| 17 | 7675089 | G | A | R | C | TP53 | ENST00000269305 | 0.88348 | 0.93139 | 0.68246 | 0.99055 | 0.94802 | 0.8257595077 | 0.99899 | 0.9970322847 | 0.418053 | 3.738069619 | 0 |
| 17 | 7675089 | G | C | R | G | TP53 | ENST00000269305 | 0.81119 | 0.92438 | 0.57335 | 0.99221 | 0.94655 | 0.8884294199 | 0.99905 | 0.998558104 | 0.551568 | 7.12957347 | 1 |
| 17 | 7675089 | G | T | R | S | TP53 | ENST00000269305 | 0.78011 | 0.90938 | 0.58988 | 0.99235 | 0.94728 | 0.8758885794 | 0.99831 | 0.9989534616 | 0.551292 | 6.125535337 | 0 |
| 17 | 7675090 | C | A | R | S | TP53 | ENST00000269305 | 0.39336 | 0.8931 | 0.29162 | 0.95709 | 0.86403 | 0.5111192014 | 0.94713 | 0.9908866882 | 0.239247 | 5.502671421 | 0 |
| 17 | 7675090 | C | G | R | S | TP53 | ENST00000269305 | 0.39952 | 0.8931 | 0.29162 | 0.95567 | 0.86403 | 0.5111192014 | 0.94693 | 0.9893687367 | 0.239247 | 5.502671421 | 0 |
| 17 | 7675091 | C | A | R | M | TP53 | ENST00000269305 | 0.4747 | 0.90501 | 0.29907 | 0.97943 | 0.85843 | 0.5092159481 | 0.97246 | 0.9941357374 | 0.246387 | 6.64751593 | 0 |
| 17 | 7675091 | C | G | R | T | TP53 | ENST00000269305 | 0.40928 | 0.93062 | 0.32575 | 0.96143 | 0.86098 | 0.5034614735 | 0.96138 | 0.9857090712 | 0.227219 | 5.199974028 | 0 |
| 17 | 7675091 | C | T | R | K | TP53 | ENST00000269305 | 0.19457 | 0.60259 | 0.31453 | 0.88605 | 0.70179 | 0.3826353948 | 0.87544 | 0.09054736048 | 0.120412 | 2.653987861 | 0 |
| 17 | 7675092 | T | A | R | W | TP53 | ENST00000269305 | 0.5716 | 0.90136 | 0.35396 | 0.97649 | 0.86149 | 0.5657347248 | 0.95089 | 0.9967926145 | 0.269475 | 8.25567194 | 1 |
| 17 | 7675092 | T | C | R | G | TP53 | ENST00000269305 | 0.50511 | 0.91462 | 0.28066 | 0.98189 | 0.90406 | 0.5787137733 | 0.98034 | 0.9920166731 | 0.356477 | 8.25567194 | 0 |
| 17 | 7675094 | A | C | V | G | TP53 | ENST00000269305 | 0.72388 | 0.92792 | 0.57785 | 0.98759 | 0.92316 | 0.880900754 | 0.99779 | 0.9985664487 | 0.574698 | 8.338882545 | 1 |
| 17 | 7675094 | A | G | V | A | TP53 | ENST00000269305 | 0.69139 | 0.91917 | 0.71726 | 0.98578 | 0.91862 | 0.6983041907 | 0.99147 | 0.9982470274 | 0.560238 | 6.474659973 | 1 |
| 17 | 7675094 | A | T | V | E | TP53 | ENST00000269305 | 0.72997 | 0.94947 | 0.70545 | 0.98657 | 0.91484 | 0.8969254733 | 0.9922 | 0.9987396598 | 0.54141 | 8.338882545 | 1 |
| 17 | 7675095 | C | A | V | L | TP53 | ENST00000269305 | 0.533 | 0.89481 | 0.69582 | 0.96174 | 0.96517 | 0.6477653343 | 0.97557 | 0.992857337 | 0.508586 | 7.323505413 | 1 |
| 17 | 7675095 | C | G | V | L | TP53 | ENST00000269305 | 0.54015 | 0.89481 | 0.69582 | 0.96443 | 0.96517 | 0.6477653343 | 0.97557 | 0.9912086725 | 0.508586 | 7.323505413 | 1 |
| 17 | 7675095 | C | T | V | M | TP53 | ENST00000269305 | 0.57385 | 0.89766 | 0.73254 | 0.96487 | 0.96956 | 0.698973642 | 0.99224 | 0.9953399897 | 0.578176 | 7.921841595 | 1 |
| 17 | 7675097 | A | C | V | G | TP53 | ENST00000269305 | 0.75495 | 0.92742 | 0.5368 | 0.98103 | 0.9917 | 0.7116659638 | 0.99527 | 0.9984624386 | 0.565532 | 7.644088674 | 0 |
| 17 | 7675097 | A | G | V | A | TP53 | ENST00000269305 | 0.71434 | 0.91384 | 0.67842 | 0.97256 | 0.95426 | 0.4845607975 | 0.95986 | 0.9900872111 | 0.408278 | 5.372597322 | 0 |
| 17 | 7675097 | A | T | V | D | TP53 | ENST00000269305 | 0.7714 | 0.94855 | 0.68548 | 0.98272 | 0.99138 | 0.7897323195 | 0.99579 | 0.9987414479 | 0.575188 | 8.672178094 | 1 |
| 17 | 7675098 | C | A | V | F | TP53 | ENST00000269305 | 0.4298 | 0.92781 | 0.62685 | 0.98793 | 0.97885 | 0.7190038596 | 0.99012 | 0.9985021353 | 0.545893 | 7.706066797 | 1 |
| 17 | 7675098 | C | G | V | L | TP53 | ENST00000269305 | 0.40769 | 0.78383 | 0.66094 | 0.97744 | 0.94129 | 0.4809952013 | 0.96907 | 0.9741591215 | 0.376417 | 5.535072423 | 0 |
| 17 | 7675099 | C | A | E | D | TP53 | ENST00000269305 | 0.07919 | 0.86751 | 0.54175 | 0.94965 | 0.84213 | 0.2882249283 | 0.88248 | 0.6062341332 | 0.172876 | 4.588092685 | 0 |
| 17 | 7675099 | C | G | E | D | TP53 | ENST00000269305 | 0.08217 | 0.86751 | 0.54175 | 0.94997 | 0.84213 | 0.2882249283 | 0.88274 | 0.5116734505 | 0.172877 | 4.588092685 | 0 |
| 17 | 7675100 | T | A | E | V | TP53 | ENST00000269305 | 0.6681 | 0.91476 | 0.51523 | 0.98758 | 0.98071 | 0.5750317567 | 0.97981 | 0.9985951781 | 0.422908 | 7.051442178 | 0 |
| 17 | 7675100 | T | C | E | G | TP53 | ENST00000269305 | 0.68875 | 0.91524 | 0.49261 | 0.98351 | 0.97885 | 0.5680135555 | 0.94982 | 0.9955216646 | 0.377208 | 8.49496588 | 1 |
| 17 | 7675100 | T | G | E | A | TP53 | ENST00000269305 | 0.64058 | 0.90322 | 0.54372 | 0.98197 | 0.97104 | 0.4881526551 | 0.94835 | 0.9971518517 | 0.288561 | 7.801614007 | 0 |
| 17 | 7675101 | C | G | E | Q | TP53 | ENST00000269305 | 0.43896 | 0.89328 | 0.52529 | 0.96453 | 0.95389 | 0.4367404121 | 0.95954 | 0.9889292121 | 0.352981 | 6.811150178 | 0 |
| 17 | 7675101 | C | T | E | K | TP53 | ENST00000269305 | 0.46961 | 0.86621 | 0.65933 | 0.9747 | 0.96262 | 0.5242642873 | 0.94579 | 0.9923363328 | 0.380519 | 6.569994603 | 0 |
| 17 | 7675103 | G | A | T | M | TP53 | ENST00000269305 | 0.40436 | 0.90068 | 0.37745 | 0.97063 | 0.9608 | 0.2708205027 | 0.97234 | 0.7052611113 | 0.251623 | 4.677002164 | 0 |
| 17 | 7675103 | G | C | T | R | TP53 | ENST00000269305 | 0.68027 | 0.9111 | 0.41533 | 0.98244 | 0.96044 | 0.4242902815 | 0.97663 | 0.9874067903 | 0.392087 | 5.57994786 | 0 |
| 17 | 7675103 | G | T | T | K | TP53 | ENST00000269305 | 0.50492 | 0.83185 | 0.43564 | 0.96946 | 0.9458 | 0.3696870282 | 0.97906 | 0.9604715705 | 0.357777 | 5.433616262 | 0 |
| 17 | 7675104 | T | A | T | S | TP53 | ENST00000269305 | 0.26252 | 0.8357 | 0.33193 | 0.94825 | 0.85894 | 0.1407703511 | 0.95056 | 0.9085736871 | 0.229822 | 4.819481862 | 0 |
| 17 | 7675104 | T | C | T | A | TP53 | ENST00000269305 | 0.21056 | 0.81691 | 0.3159 | 0.93152 | 0.79697 | 0.1711382776 | 0.93717 | 0.2609120011 | 0.159343 | 3.653371719 | 0 |
| 17 | 7675104 | T | G | T | P | TP53 | ENST00000269305 | 0.56769 | 0.92466 | 0.32092 | 0.97987 | 0.93863 | 0.5457450079 | 0.98022 | 0.9681432843 | 0.355803 | 7.204518506 | 0 |
| 17 | 7675105 | C | A | M | I | TP53 | ENST00000269305 | 0.28817 | 0.64371 | 0.31144 | 0.97441 | 0.84213 | 0.2407233561 | 0.94788 | 0.5471804142 | 0.200077 | 4.5978830917 | 0 |
| 17 | 7675105 | C | G | M | I | TP53 | ENST00000269305 | 0.29343 | 0.64371 | 0.31144 | 0.96719 | 0.84213 | 0.2407233561 | 0.94772 | 0.4269660115 | 0.200075 | 4.597830917 | 0 |
| 17 | 7675105 | C | T | M | I | TP53 | ENST00000269305 | 0.29682 | 0.64371 | 0.31144 | 0.97362 | 0.84213 | 0.2407233561 | 0.94729 | 0.5376009345 | 0.200084 | 4.597830917 | 0 |
| 17 | 7675106 | A | C | M | R | TP53 | ENST00000269305 | 0.56276 | 0.9087 | 0.38739 | 0.96972 | 0.911 | 0.7014863739 | 0.96245 | 0.9808149338 | 0.272745 | 4.516484856 | 0 |
| 17 | 7675106 | A | G | M | T | TP53 | ENST00000269305 | 0.40278 | 0.67392 | 0.44573 | 0.95145 | 0.90494 | 0.4434420998 | 0.96621 | 0.8314960003 | 0.296923 | 7.16738854 | 0 |
| 17 | 7675106 | A | T | M | K | TP53 | ENST00000269305 | 0.56069 | 0.88449 | 0.40458 | 0.98171 | 0.94839 | 0.6516117082 | 0.97361 | 0.984983623 | 0.367318 | 7.998072637 | 0 |
| 17 | 7675107 | T | A | M | L | TP53 | ENST00000269305 | 0.32786 | 0.5831 | 0.32473 | 0.94182 | 0.82273 | 0.2880665338 | 0.92989 | 0.5356974006 | 0.186027 | 5.361754233 | 0 |
| 17 | 7675107 | T | C | M | V | TP53 | ENST00000269305 | 0.1577 | 0.58785 | 0.26364 | 0.84764 | 0.78782 | 0.2438364774 | 0.86558 | 0.1684140712 | 0.15319 | 3.538333669 | 0 |
| 17 | 7675107 | T | G | M | L | TP53 | ENST00000269305 | 0.33334 | 0.5831 | 0.32473 | 0.94203 | 0.82159 | 0.2880665338 | 0.92985 | 0.3837579489 | 0.186043 | 5.361754233 | 0 |
| 17 | 7675108 | G | T | H | Q | TP53 | ENST00000269305 | 0.4673 | 0.87117 | 0.5247 | 0.93654 | 0.82613 | 0.5171910296 | 0.96521 | 0.9984282255 | 0.255559 | 5.996903845 | 0 |
| 17 | 7675109 | T | A | H | L | TP53 | ENST00000269305 | 0.57723 | 0.76337 | 0.51306 | 0.95723 | 0.81588 | 0.6700099066 | 0.97284 | 0.9892958999 | 0.326349 | 6.450395764 | 1 |
| 17 | 7675109 | T | C | H | R | TP53 | ENST00000269305 | 0.55834 | 0.87797 | 0.56188 | 0.94807 | 0.82273 | 0.7019504715 | 0.96917 | 0.96935004 | 0.277113 | 7.347475708 | 1 |
| 17 | 7675109 | T | G | H | P | TP53 | ENST00000269305 | 0.6075 | 0.94073 | 0.49836 | 0.95715 | 0.80178 | 0.8200407413 | 0.96132 | 0.9990879297 | 0.293435 | 7.814917847 | 1 |
| 17 | 7675110 | G | A | H | Y | TP53 | ENST00000269305 | 0.22127 | 0.6471 | 0.5571 | 0.89799 | 0.78658 | 0.6202445692 | 0.87481 | 0.8315386772 | 0.182472 | 2.902928305 | 0 |
| 17 | 7675110 | G | C | H | D | TP53 | ENST00000269305 | 0.34445 | 0.92004 | 0.56332 | 0.97691 | 0.81646 | 0.7452008289 | 0.92141 | 0.9282943606 | 0.213924 | 4.183470603 | 0 |
| 17 | 7675110 | G | T | H | N | TP53 | ENST00000269305 | 0.45052 | 0.89156 | 0.56038 | 0.9737 | 0.825 | 0.5678938853 | 0.95054 | 0.9656882286 | 0.202852 | 4.909789803 | 0 |
| 17 | 7675111 | C | A | Q | H | TP53 | ENST00000269305 | 0.38027 | 0.8588 | 0.2158 | 0.98536 | 0.83779 | 0.3424163175 | 0.91813 | 0.9681936502 | 0.189673 | 4.700798763 | 0 |
| 17 | 7675111 | C | G | Q | H | TP53 | ENST00000269305 | 0.38633 | 0.8588 | 0.2158 | 0.98478 | 0.83943 | 0.3424163175 | 0.91824 | 0.9701681137 | 0.189687 | 4.700798763 | 0 |
| 17 | 7675112 | T | C | Q | R | TP53 | ENST00000269305 | 0.57273 | 0.78886 | 0.22483 | 0.98292 | 0.825 | 0.3265358635 | 0.94063 | 0.9809383154 | 0.261707 | 5.317845213 | 0 |
| 17 | 7675112 | T | G | Q | P | TP53 | ENST00000269305 | 0.76148 | 0.90391 | 0.1924 | 0.98392 | 0.86098 | 0.6842063696 | 0.93695 | 0.9865800738 | 0.265735 | 7.056986784 | 0 |
| 17 | 7675113 | G | C | Q | E | TP53 | ENST00000269305 | 0.1062 | 0.61099 | 0.16939 | 0.95283 | 0.76487 | 0.1805068501 | 0.75572 | 0.2094604969 | 0.147637 | 0.8228290561 | 0 |
| 17 | 7675113 | G | T | Q | K | TP53 | ENST00000269305 | 0.26275 | 0.77074 | 0.24807 | 0.98821 | 0.86803 | 0.3982199643 | 0.9546 | 0.920755744 | 0.313808 | 6.074210867 | 0 |
| 17 | 7675115 | G | A | S | L | TP53 | ENST00000269305 | 0.26986 | 0.64516 | 0.30806 | 0.96334 | 0.85064 | 0.3486453833 | 0.93644 | 0.8088536263 | 0.20665 | 4.053944412 | 0 |
| 17 | 7675116 | A | C | S | A | TP53 | ENST00000269305 | 0.21114 | 0.79831 | 0.2992 | 0.96833 | 0.77782 | 0.1553661328 | 0.87576 | 0.4369901419 | 0.12454 | 4.389752387 | 0 |
| 17 | 7675116 | A | G | S | P | TP53 | ENST00000269305 | 0.34538 | 0.92033 | 0.33755 | 0.98574 | 0.89648 | 0.5789039512 | 0.96517 | 0.9656996727 | 0.266329 | 4.411193657 | 0 |
| 17 | 7675116 | A | T | S | T | TP53 | ENST00000269305 | 0.11214 | 0.81571 | 0.31849 | 0.98635 | 0.79334 | 0.2195376937 | 0.89331 | 0.270740211 | 0.1436 | 4.337812521 | 0 |
| 17 | 7675117 | C | A | Q | H | TP53 | ENST00000269305 | 0.14582 | 0.76018 | 0.3662 | 0.98379 | 0.84375 | 0.453552931 | 0.93194 | 0.4887601435 | 0.165572 | 4.603218823 | 0 |
| 17 | 7675117 | C | G | Q | H | TP53 | ENST00000269305 | 0.14937 | 0.76018 | 0.3662 | 0.9837 | 0.84375 | 0.453552931 | 0.93202 | 0.4335253835 | 0.165573 | 4.603218823 | 0 |
| 17 | 7675118 | T | A | Q | L | TP53 | ENST00000269305 | 0.59161 | 0.64661 | 0.32009 | 0.97902 | 0.82948 | 0.4624744819 | 0.97122 | 0.9519104362 | 0.249226 | 3.089634452 | 0 |
| 17 | 7675118 | T | C | Q | R | TP53 | ENST00000269305 | 0.43707 | 0.63835 | 0.31467 | 0.94048 | 0.77011 | 0.2545839436 | 0.95371 | 0.7674856186 | 0.213071 | 1.625802549 | 0 |
| 17 | 7675118 | T | G | Q | P | TP53 | ENST00000269305 | 0.59429 | 0.81783 | 0.34265 | 0.98415 | 0.87099 | 0.7586293364 | 0.96589 | 0.9076171517 | 0.291155 | 5.79585443 | 0 |
| 17 | 7675119 | G | C | Q | E | TP53 | ENST00000269305 | 0.40746 | 0.63259 | 0.3701 | 0.94878 | 0.82836 | 0.4031437564 | 0.8983 | 0.6103248 | 0.195203 | 3.058446803 | 0 |
| 17 | 7675119 | G | T | Q | K | TP53 | ENST00000269305 | 0.24642 | 0.66982 | 0.33777 | 0.85133 | 0.77399 | 0.3170719129 | 0.75475 | 0.1122064739 | 0.123022 | -0.8609252896 | 0 |
| 17 | 7675120 | C | A | K | N | TP53 | ENST00000269305 | 0.53968 | 0.9161 | 0.64496 | 0.97519 | 0.93319 | 0.6962585174 | 0.95183 | 0.9971358776 | 0.259974 | 8.608064491 | 0 |
| 17 | 7675120 | C | G | K | N | TP53 | ENST00000269305 | 0.54684 | 0.9161 | 0.64496 | 0.97672 | 0.93319 | 0.6962585174 | 0.95204 | 0.996471405 | 0.259975 | 8.608064491 | 0 |
| 17 | 7675121 | T | A | K | M | TP53 | ENST00000269305 | 0.69251 | 0.93062 | 0.55409 | 0.97797 | 0.98183 | 0.7162324802 | 0.9896 | 0.9963114858 | 0.52263 | 6.109371344 | 0 |
| 17 | 7675121 | T | C | K | R | TP53 | ENST00000269305 | 0.49699 | 0.87103 | 0.54934 | 0.96521 | 0.97475 | 0.508090345 | 0.97564 | 0.9420812726 | 0.413775 | 4.553362884 | 0 |
| 17 | 7675121 | T | G | K | T | TP53 | ENST00000269305 | 0.65995 | 0.91805 | 0.61997 | 0.97259 | 0.98257 | 0.6899285099 | 0.98671 | 0.9947426319 | 0.524092 | 5.621359676 | 0 |
| 17 | 7675122 | T | C | K | E | TP53 | ENST00000269305 | 0.754 | 0.93489 | 0.6172 | 0.9822 | 0.96334 | 0.7404223075 | 0.99773 | 0.9944609404 | 0.553451 | 6.432579913 | 1 |
| 17 | 7675122 | T | G | K | Q | TP53 | ENST00000269305 | 0.69868 | 0.90521 | 0.58066 | 0.98495 | 0.97215 | 0.6117356122 | 0.98542 | 0.9944995642 | 0.481519 | 7.03772341 | 0 |
| 17 | 7675124 | T | A | Y | F | TP53 | ENST00000269305 | 0.55449 | 0.78618 | 0.60273 | 0.96721 | 0.90362 | 0.477377702 | 0.9281 | 0.9759922028 | 0.227187 | 4.817096392 | 0 |
| 17 | 7675124 | T | C | Y | C | TP53 | ENST00000269305 | 0.69155 | 0.92329 | 0.69925 | 0.98166 | 0.96226 | 0.8151402085 | 0.9929 | 0.9987146854 | 0.539375 | 8.134909234 | 1 |
| 17 | 7675124 | T | G | Y | S | TP53 | ENST00000269305 | 0.65869 | 0.93697 | 0.63196 | 0.98057 | 0.9729 | 0.8402855253 | 0.98932 | 0.9990340471 | 0.538766 | 9.194323714 | 1 |
| 17 | 7675125 | A | C | Y | D | TP53 | ENST00000269305 | 0.80163 | 0.9568 | 0.68983 | 0.97677 | 0.97475 | 0.8814821946 | 0.99492 | 0.9998813868 | 0.579668 | 9.194323714 | 1 |
| 17 | 7675125 | A | G | Y | H | TP53 | ENST00000269305 | 0.78819 | 0.92696 | 0.70746 | 0.9697 | 0.97401 | 0.7535257084 | 0.97956 | 0.9955434203 | 0.476435 | 8.0903455051 | 1 |
| 17 | 7675125 | A | T | Y | N | TP53 | ENST00000269305 | 0.79634 | 0.94997 | 0.69855 | 0.97927 | 0.97512 | 0.843082808 | 0.98771 | 0.9995812774 | 0.572081 | 9.194323714 | 1 |
| 17 | 7675126 | G | C | I | M | TP53 | ENST00000269305 | 0.54453 | 0.89182 | 0.423 | 0.97727 | 0.85894 | 0.545251812 | 0.97029 | 0.9879155159 | 0.245187 | 5.133962237 | 0 |
| 17 | 7675127 | A | G | I | T | TP53 | ENST00000269305 | 0.77972 | 0.91384 | 0.6025 | 0.98445 | 0.93475 | 0.6331678174 | 0.95814 | 0.9937624335 | 0.367486 | 8.13985482 | 0 |
| 17 | 7675127 | A | T | I | N | TP53 | ENST00000269305 | 0.83928 | 0.95598 | 0.5733 | 0.98712 | 0.93554 | 0.7985862408 | 0.96905 | 0.9942282438 | 0.309717 | 8.13985482 | 1 |
| 17 | 7675128 | T | C | I | V | TP53 | ENST00000269305 | 0.19017 | 0.52157 | 0.35755 | 0.91943 | 0.78473 | 0.1755599915 | 0.87308 | 0.09228493809 | 0.140725 | 3.190061714 | 0 |
| 17 | 7675128 | T | G | I | L | TP53 | ENST00000269305 | 0.52234 | 0.77444 | 0.46417 | 0.98771 | 0.87684 | 0.4569120306 | 0.94331 | 0.8562820554 | 0.211693 | 5.401945784 | 0 |
| 17 | 7675130 | G | A | A | V | TP53 | ENST00000269305 | 0.84104 | 0.8967 | 0.50279 | 0.98878 | 0.9367 | 0.5644868487 | 0.9479 | 0.9751605392 | 0.397165 | 6.26285097 | 0 |
| 17 | 7675130 | G | C | A | G | TP53 | ENST00000269305 | 0.8402 | 0.89783 | 0.45822 | 0.98599 | 0.93901 | 0.5501265792 | 0.94184 | 0.9758018851 | 0.384656 | 8.022458196 | 0 |
| 17 | 7675130 | G | T | A | D | TP53 | ENST00000269305 | 0.85055 | 0.94218 | 0.60449 | 0.98836 | 0.9755 | 0.8384932114 | 0.98765 | 0.9959677458 | 0.574477 | 9.148536085 | 1 |
| 17 | 7675131 | C | A | A | S | TP53 | ENST00000269305 | 0.5679 | 0.88566 | 0.43794 | 0.97976 | 0.91611 | 0.5463432629 | 0.92394 | 0.962379396 | 0.327408 | 7.570976219 | 0 |
| 17 | 7675131 | C | G | A | P | TP53 | ENST00000269305 | 0.61626 | 0.92717 | 0.55939 | 0.93296 | 0.86503 | 0.7892002067 | 0.90006 | 0.797522366 | 0.275838 | 5.025890289 | 0 |
| 17 | 7675131 | C | T | A | T | TP53 | ENST00000269305 | 0.6072 | 0.89319 | 0.48309 | 0.98702 | 0.93162 | 0.5912807649 | 0.93766 | 0.9794812799 | 0.358081 | 8.069766862 | 1 |
| 17 | 7675132 | C | A | M | I | TP53 | ENST00000269305 | 0.3004 | 0.84503 | 0.45984 | 0.95095 | 0.87296 | 0.4409378785 | 0.92882 | 0.7929543257 | 0.304324 | 5.243089673 | 0 |
| 17 | 7675132 | C | G | M | I | TP53 | ENST00000269305 | 0.30586 | 0.84503 | 0.45984 | 0.9506 | 0.87296 | 0.4409378785 | 0.92979 | 0.7331977487 | 0.30433 | 5.243089673 | 0 |
| 17 | 7675132 | C | T | M | I | TP53 | ENST00000269305 | 0.30935 | 0.84503 | 0.45984 | 0.95053 | 0.87247 | 0.4409378785 | 0.9285 | 0.5962894063 | 0.293554 | 5.243089673 | 0 |
| 17 | 7675133 | A | C | M | R | TP53 | ENST00000269305 | 0.67724 | 0.93996 | 0.52522 | 0.97943 | 0.95972 | 0.7943565227 | 0.99159 | 0.9804691672 | 0.50514 | 8.537458877 | 0 |
| 17 | 7675133 | A | G | M | T | TP53 | ENST00000269305 | 0.27741 | 0.90976 | 0.5832 | 0.96545 | 0.87877 | 0.4980993631 | 0.90883 | 0.5476892591 | 0.261709 | 3.971706849 | 0 |
| 17 | 7675133 | A | T | M | K | TP53 | ENST00000269305 | 0.6475 | 0.9404 | 0.54843 | 0.98102 | 0.9579 | 0.7615086119 | 0.99055 | 0.9804691672 | 0.493382 | 7.068489394 | 0 |
| 17 | 7675134 | T | A | M | L | TP53 | ENST00000269305 | 0.30056 | 0.59513 | 0.4478 | 0.95232 | 0.82668 | 0.3850624295 | 0.88072 | 0.4643385708 | 0.190198 | 4.982891111 | 0 |
| 17 | 7675134 | T | C | M | V | TP53 | ENST00000269305 | 0.30218 | 0.83491 | 0.37748 | 0.96055 | 0.88823 | 0.422251411 | 0.96583 | 0.3318133627 | 0.241894 | 4.262327146 | 0 |

| | | | | | | | | | | | | | | | | | | |
|---|---|---|---|---|---|---|---|---|---|---|---|---|---|---|---|---|---|---|
| 17 | 7675134 | T | G | M | L | TP53 | ENST00000269305 | 0.30581 | 0.59513 | 0.4478 | 0.94048 | 0.82668 | 0.3850624295 | 0.88073 | 0.4643385708 | 0.190184 | 4.982891111 | 0 |
| 17 | 7675136 | G | A | A | V | TP53 | ENST00000269305 | 0.75957 | 0.87918 | 0.42819 | 0.9818 | 0.9252 | 0.5053663656 | 0.95124 | 0.9937378168 | 0.355457 | 3.791728235 | 1 |
| 17 | 7675136 | G | C | A | G | TP53 | ENST00000269305 | 0.58254 | 0.6517 | 0.37233 | 0.9804 | 0.91015 | 0.5556282727 | 0.96203 | 0.984739542 | 0.369139 | 5.060327999 | 0 |
| 17 | 7675136 | G | T | A | D | TP53 | ENST00000269305 | 0.79271 | 0.93962 | 0.51489 | 0.98063 | 0.94913 | 0.8624930811 | 0.97556 | 0.9977391958 | 0.458875 | 7.21991828 | 1 |
| 17 | 7675137 | C | A | A | S | TP53 | ENST00000269305 | 0.38955 | 0.8754 | 0.35245 | 0.97832 | 0.90537 | 0.5221304242 | 0.95435 | 0.9740346074 | 0.330198 | 4.731872926 | 0 |
| 17 | 7675137 | C | G | A | P | TP53 | ENST00000269305 | 0.43087 | 0.92603 | 0.45787 | 0.97746 | 0.94129 | 0.8675463071 | 0.98622 | 0.9948969483 | 0.515812 | 7.21991828 | 1 |
| 17 | 7675137 | C | T | A | T | TP53 | ENST00000269305 | 0.28058 | 0.73623 | 0.38526 | 0.95847 | 0.90007 | 0.5500990096 | 0.9589 | 0.9337339944 | 0.26813 | 2.017745157 | 0 |
| 17 | 7675139 | C | A | R | L | TP53 | ENST00000269305 | 0.74293 | 0.92085 | 0.45679 | 0.98335 | 0.96919 | 0.873361691 | 0.99512 | 0.9984660149 | 0.566223 | 6.634582344 | 1 |
| 17 | 7675139 | C | G | R | P | TP53 | ENST00000269305 | 0.76475 | 0.93796 | 0.47965 | 0.98275 | 0.97848 | 0.9557548093 | 0.99728 | 0.9975389242 | 0.566223 | 7.683583566 | 1 |
| 17 | 7675140 | G | C | R | G | TP53 | ENST00000269305 | 0.70895 | 0.92554 | 0.42956 | 0.97953 | 0.95863 | 0.9181133942 | 0.9963 | 0.9954811335 | 0.530867 | 7.683583566 | 1 |
| 17 | 7675140 | G | T | R | S | TP53 | ENST00000269305 | 0.6774 | 0.91029 | 0.44067 | 0.97775 | 0.95935 | 0.8909447268 | 0.99693 | 0.9969124794 | 0.530587 | 6.275619281 | 0 |
| 17 | 7675142 | A | C | V | G | TP53 | ENST00000269305 | 0.74388 | 0.92792 | 0.28479 | 0.98705 | 0.96226 | 0.8654858436 | 0.98694 | 0.997757256 | 0.565523 | 8.696709808 | 1 |
| 17 | 7675142 | A | T | V | D | TP53 | ENST00000269305 | 0.76036 | 0.94913 | 0.36321 | 0.98917 | 0.95899 | 0.8939817357 | 0.98796 | 0.9979228377 | 0.566697 | 8.696709808 | 1 |
| 17 | 7675143 | C | A | V | F | TP53 | ENST00000269305 | 0.26804 | 0.9294 | 0.32475 | 0.99087 | 0.89694 | 0.733504993 | 0.97712 | 0.9772703052 | 0.384181 | 6.686888504 | 1 |
| 17 | 7675143 | C | G | V | L | TP53 | ENST00000269305 | 0.08144 | 0.86526 | 0.34695 | 0.98478 | 0.81356 | 0.3340288762 | 0.88336 | 0.2696185112 | 0.153998 | 3.702351861 | 0 |
| 17 | 7675143 | C | T | V | I | TP53 | ENST00000269305 | 0.13763 | 0.67834 | 0.23352 | 0.99427 | 0.73589 | 0.1111248118 | 0.82353 | 0.1082348078 | -0.031324 | 3.764646449 | 0 |
| 17 | 7675145 | C | A | R | L | TP53 | ENST00000269305 | 0.18007 | 0.90999 | 0.14367 | 0.95585 | 0.79334 | 0.4724805707 | 0.90697 | 0.7024038434 | 0.18677 | 3.834538419 | 0 |
| 17 | 7675145 | C | T | R | H | TP53 | ENST00000269305 | 0.32278 | 0.91961 | 0.13722 | 0.96236 | 0.85792 | 0.3668095705 | 0.96172 | 0.6953080893 | 0.171905 | 4.382644354 | 0 |
| 17 | 7675146 | G | A | R | C | TP53 | ENST00000269305 | 0.27199 | 0.82575 | 0.19027 | 0.94459 | 0.7816 | 0.5387843421 | 0.88336 | 0.1869009882 | 0.0791594 | 3.530548905 | 0 |
| 17 | 7675146 | G | T | R | S | TP53 | ENST00000269305 | 0.20062 | 0.90698 | 0.13751 | 0.93811 | 0.79998 | 0.5884391119 | 0.89068 | 0.4730595946 | 0.161482 | 5.702045819 | 0 |
| 17 | 7675149 | T | A | T | S | TP53 | ENST00000269305 | 0.04083 | 0.54702 | 0.14752 | 0.7946 | 0.68292 | 0.2287753036 | 0.90937 | 0.07113493351 | 0.0909877 | 1.182904279 | 0 |
| 17 | 7675149 | T | C | T | A | TP53 | ENST00000269305 | 0.0438 | 0.57664 | 0.15446 | 0.83311 | 0.66258 | 0.2973906809 | 0.91896 | 0.05820205251 | -0.0273894 | 1.862675614 | 0 |
| 17 | 7675149 | T | G | T | P | TP53 | ENST00000269305 | 0.25939 | 0.91229 | 0.16066 | 0.95819 | 0.83339 | 0.6152269964 | 0.96347 | 0.6946588159 | 0.200663 | 7.441369703 | 0 |
| 17 | 7675151 | C | A | G | V | TP53 | ENST00000269305 | 0.34114 | 0.91834 | 0.34337 | 0.96919 | 0.91694 | 0.6914485567 | 0.9909 | 0.9990044236 | 0.502691 | 6.178683284 | 1 |
| 17 | 7675151 | C | G | G | A | TP53 | ENST00000269305 | 0.31291 | 0.80585 | 0.34491 | 0.93515 | 0.90581 | 0.5182705823 | 0.96916 | 0.9930839539 | 0.326592 | 5.181799519 | 0 |
| 17 | 7675151 | C | T | G | D | TP53 | ENST00000269305 | 0.23086 | 0.7348 | 0.44079 | 0.94571 | 0.90274 | 0.5741458385 | 0.98376 | 0.47398148 | 0.345031 | 3.890389049 | 0 |
| 17 | 7675152 | C | G | G | R | TP53 | ENST00000269305 | 0.29392 | 0.91415 | 0.42829 | 0.96181 | 0.89829 | 0.6284194959 | 0.98186 | 0.9987829328 | 0.395828 | 5.055360035 | 0 |
| 17 | 7675152 | C | T | G | S | TP53 | ENST00000269305 | 0.17735 | 0.6865 | 0.33882 | 0.93338 | 0.86352 | 0.5120271808 | 0.95173 | 0.9007704854 | -0.0118271 | 2.935844251 | 0 |
| 17 | 7675154 | G | A | P | L | TP53 | ENST00000269305 | 0.10029 | 0.59345 | 0.07403 | 0.94385 | 0.69254 | 0.2372535925 | 0.90073 | 0.1252915263 | 0.149099 | 2.710434418 | 0 |
| 17 | 7675154 | G | C | P | R | TP53 | ENST00000269305 | 0.08462 | 0.61928 | 0.08023 | 0.93949 | 0.71627 | 0.2345506276 | 0.89727 | 0.07246731501 | 0.143592 | 4.339506756 | 0 |
| 17 | 7675154 | G | T | P | H | TP53 | ENST00000269305 | 0.10959 | 0.87402 | 0.06707 | 0.97577 | 0.70994 | 0.2331078225 | 0.90342 | 0.5858921409 | 0.159419 | 4.389111412 | 0 |
| 17 | 7675155 | G | A | P | S | TP53 | ENST00000269305 | 0.04759 | 0.66869 | 0.05009 | 0.96943 | 0.68292 | 0.213168604 | 0.88625 | 0.08242703629 | 0.149279 | 4.452531284 | 0 |
| 17 | 7675155 | G | C | P | A | TP53 | ENST00000269305 | 0.02528 | 0.56687 | 0.05461 | 0.97411 | 0.69168 | 0.1694280529 | 0.8282 | 0.04255959476 | 0.101659 | 3.286249503 | 0 |
| 17 | 7675155 | G | T | P | T | TP53 | ENST00000269305 | 0.03685 | 0.62587 | 0.06017 | 0.97659 | 0.70262 | 0.2546013089 | 0.89314 | 0.2586918175 | 0.149612 | 5.01336849 | 0 |
| 17 | 7675157 | G | C | P | R | TP53 | ENST00000269305 | 0.72027 | 0.90395 | 0.36124 | 0.98351 | 0.95535 | 0.7547282503 | 0.96196 | 0.9989445806 | 0.535677 | 7.778109899 | 1 |
| 17 | 7675158 | G | C | P | A | TP53 | ENST00000269305 | 0.61731 | 0.89027 | 0.29689 | 0.96891 | 0.9367 | 0.5878590346 | 0.93721 | 0.9979732633 | 0.373056 | 6.345859955 | 0 |
| 17 | 7675158 | G | T | P | T | TP53 | ENST00000269305 | 0.64291 | 0.90976 | 0.33848 | 0.9775 | 0.96008 | 0.6791688977 | 0.94445 | 0.9985483289 | 0.416222 | 6.296084012 | 0 |
| 17 | 7675160 | G | A | P | L | TP53 | ENST00000269305 | 0.6942 | 0.89943 | 0.44049 | 0.97191 | 0.92397 | 0.6876621832 | 0.93202 | 0.9946616292 | 0.322493 | 4.452068638 | 0 |
| 17 | 7675160 | G | C | P | R | TP53 | ENST00000269305 | 0.70752 | 0.90368 | 0.43918 | 0.96297 | 0.97698 | 0.743787907 | 0.97182 | 0.9983375072 | 0.506834 | 7.670916454 | 1 |
| 17 | 7675161 | G | A | P | S | TP53 | ENST00000269305 | 0.53314 | 0.87675 | 0.3694 | 0.96141 | 0.97178 | 0.6606268057 | 0.96852 | 0.9876742959 | 0.506784 | 5.783947356 | 1 |
| 17 | 7675163 | G | A | T | I | TP53 | ENST00000269305 | 0.30478 | 0.6737 | 0.18141 | 0.93545 | 0.70179 | 0.2103653447 | 0.85822 | 0.6443403959 | 0.141523 | 6.675902365 | 0 |
| 17 | 7675163 | G | C | T | R | TP53 | ENST00000269305 | 0.23292 | 0.83761 | 0.16455 | 0.95371 | 0.74728 | 0.3198061104 | 0.86223 | 0.6649679542 | 0.147403 | 4.171342275 | 0 |
| 17 | 7675163 | G | T | T | K | TP53 | ENST00000269305 | 0.22447 | 0.69427 | 0.17729 | 0.89188 | 0.71233 | 0.3425052233 | 0.82055 | 0.4545400739 | 0.137554 | 3.786262437 | 0 |
| 17 | 7675164 | T | A | T | S | TP53 | ENST00000269305 | 0.09447 | 0.55178 | 0.11181 | 0.89911 | 0.60599 | 0.1661421463 | 0.77709 | 0.087959548 | 0.0746743 | 3.251293459 | 0 |
| 17 | 7675164 | T | C | T | A | TP53 | ENST00000269305 | 0.09232 | 0.57042 | 0.10992 | 0.84641 | 0.65869 | 0.2000795525 | 0.76402 | 0.05842984596 | 0.0641099 | 1.58078309 | 0 |
| 17 | 7675164 | T | G | T | P | TP53 | ENST00000269305 | 0.07454 | 0.67851 | 0.11273 | 0.78074 | 0.7589 | 0.2764277926 | 0.74305 | 0.04345820472 | 0.095313 | -0.1538498207 | 0 |
| 17 | 7675166 | G | A | S | F | TP53 | ENST00000269305 | 0.24659 | 0.77248 | 0.39997 | 0.89047 | 0.75822 | 0.4459058067 | 0.91919 | 0.222212651 | 0.193616 | 5.980561541 | 0 |
| 17 | 7675166 | G | C | S | C | TP53 | ENST00000269305 | 0.47036 | 0.88742 | 0.37375 | 0.91028 | 0.77463 | 0.4677497796 | 0.93657 | 0.8280644417 | 0.206518 | 7.034324782 | 0 |
| 17 | 7675166 | G | T | S | Y | TP53 | ENST00000269305 | 0.44253 | 0.91961 | 0.42396 | 0.91557 | 0.75211 | 0.4385158978 | 0.92259 | 0.8036612272 | 0.210111 | 6.531306259 | 0 |
| 17 | 7675167 | A | C | S | A | TP53 | ENST00000269305 | 0.17021 | 0.55769 | 0.34595 | 0.94206 | 0.63736 | 0.2119535444 | 0.84178 | 0.03900651634 | 0.0870981 | 4.540305622 | 0 |
| 17 | 7675167 | A | T | S | T | TP53 | ENST00000269305 | 0.1592 | 0.76255 | 0.3611 | 0.89875 | 0.65277 | 0.3253340791 | 0.84296 | 0.08450940968 | 0.113005 | 4.503696115 | 0 |
| 17 | 7675168 | A | C | D | E | TP53 | ENST00000269305 | 0.06974 | 0.55195 | 0.09293 | 0.96519 | 0.61184 | 0.1647578269 | 0.71588 | 0.08941229433 | 0.0150376 | 2.810238213 | 0 |
| 17 | 7675168 | A | T | D | E | TP53 | ENST00000269305 | 0.06753 | 0.55195 | 0.09293 | 0.97101 | 0.61184 | 0.1647578269 | 0.71486 | 0.08941229433 | 0.0150348 | 2.810238213 | 0 |
| 17 | 7675169 | T | A | D | V | TP53 | ENST00000269305 | 0.10549 | 0.80316 | 0.07517 | 0.97411 | 0.81646 | 0.4230046453 | 0.90062 | 0.07249975266 | 0.133945 | 3.828478877 | 0 |
| 17 | 7675169 | T | C | D | G | TP53 | ENST00000269305 | 0.10953 | 0.77765 | 0.0744 | 0.88192 | 0.76487 | 0.2809140651 | 0.86082 | 0.07391748029 | 0.0928826 | 2.634478269 | 0 |
| 17 | 7675169 | T | G | D | A | TP53 | ENST00000269305 | 0.09158 | 0.66446 | 0.07757 | 0.97206 | 0.72016 | 0.2887093657 | 0.79484 | 0.4371226728 | 0.0779143 | 2.520415464 | 0 |
| 17 | 7675170 | C | G | D | H | TP53 | ENST00000269305 | 0.28039 | 0.86978 | 0.10654 | 0.97945 | 0.77463 | 0.3558990416 | 0.84331 | 0.5413062572 | 0.0958469 | 4.296208185 | 0 |
| 17 | 7675170 | C | T | D | N | TP53 | ENST00000269305 | 0.10892 | 0.56926 | 0.10349 | 0.97004 | 0.5976 | 0.183784315 | 0.64179 | 0.07480045026 | -0.0401861 | 1.185994321 | 1 |
| 17 | 7675172 | A | G | V | A | TP53 | ENST00000269305 | 0.70655 | 0.90611 | 0.50032 | 0.96639 | 0.94015 | 0.4871672963 | 0.97065 | 0.9866468906 | 0.409304 | 5.914355111 | 0 |
| 17 | 7675172 | A | T | V | D | TP53 | ENST00000269305 | 0.75989 | 0.94702 | 0.50399 | 0.97491 | 0.97104 | 0.8569798337 | 0.99629 | 0.9977234006 | 0.545724 | 8.822317061 | 1 |
| 17 | 7675173 | C | A | V | F | TP53 | ENST00000269305 | 0.48985 | 0.79328 | 0.44895 | 0.9552 | 0.91399 | 0.7263751225 | 0.97144 | 0.994615376 | 0.484809 | 6.814143116 | 0 |
| 17 | 7675173 | C | G | V | L | TP53 | ENST00000269305 | 0.24423 | 0.68286 | 0.47217 | 0.96259 | 0.86653 | 0.4769773505 | 0.94889 | 0.5841627717 | 0.366146 | 4.561494757 | 0 |
| 17 | 7675173 | C | T | V | I | TP53 | ENST00000269305 | 0.31245 | 0.5813 | 0.30305 | 0.96492 | 0.8153 | 0.215936637 | 0.84818 | 0.8868983984 | 0.211549 | 6.207626558 | 0 |
| 17 | 7675174 | C | A | W | C | TP53 | ENST00000269305 | 0.50185 | 0.94555 | 0.35779 | 0.98157 | 0.85894 | 0.5774124931 | 0.95694 | 0.8819436431 | 0.217388 | 8.319908657 | 0 |
| 17 | 7675174 | C | G | W | C | TP53 | ENST00000269305 | 0.50883 | 0.94555 | 0.35779 | 0.97972 | 0.85894 | 0.5774124931 | 0.95724 | 0.8290438652 | 0.217394 | 8.319908657 | 0 |
| 17 | 7675175 | C | A | W | L | TP53 | ENST00000269305 | 0.16843 | 0.83478 | 0.3688 | 0.95486 | 0.71783 | 0.3450637025 | 0.91624 | 0.04432035915 | 0.149213 | 5.061780983 | 0 |
| 17 | 7675175 | C | G | W | S | TP53 | ENST00000269305 | 0.20675 | 0.94091 | 0.34149 | 0.92346 | 0.79878 | 0.4875265275 | 0.93268 | 0.2998377958 | 0.186153 | 5.3301148 | 0 |
| 17 | 7675176 | A | C | W | G | TP53 | ENST00000269305 | 0.24893 | 0.94913 | 0.32357 | 0.95731 | 0.80772 | 0.5321228796 | 0.96899 | 0.5886206031 | 0.189961 | 6.907673645 | 0 |
| 17 | 7675176 | A | G | W | R | TP53 | ENST00000269305 | 0.1923 | 0.87981 | 0.39763 | 0.84328 | 0.76289 | 0.4941986488 | 0.94355 | 0.09313575333 | 0.151548 | 4.628534004 | 0 |
| 17 | 7675176 | A | T | W | R | TP53 | ENST00000269305 | 0.18618 | 0.87981 | 0.39763 | 0.84328 | 0.76289 | 0.4941986488 | 0.94174 | 0.09210904392 | 0.15152 | 4.628534004 | 0 |
| 17 | 7675178 | A | C | L | R | TP53 | ENST00000269305 | 0.82615 | 0.93962 | 0.51823 | 0.9774 | 0.95243 | 0.8515221524 | 0.99066 | 0.9942399263 | 0.549395 | 7.404138754 | 1 |
| 17 | 7675178 | A | G | L | P | TP53 | ENST00000269305 | 0.51889 | 0.9489 | 0.55042 | 0.96976 | 0.94243 | 0.8948755447 | 0.98976 | 0.9968139529 | 0.54572 | 6.930104932 | 1 |
| 17 | 7675178 | A | T | L | Q | TP53 | ENST00000269305 | 0.82084 | 0.90241 | 0.44545 | 0.97304 | 0.9528 | 0.756610489 | 0.97843 | 0.9922946692 | 0.499337 | 8.367095783 | 1 |
| 17 | 7675179 | G | C | L | V | TP53 | ENST00000269305 | 0.21733 | 0.69733 | 0.35873 | 0.87682 | 0.76157 | 0.2636238841 | 0.8629 | 0.07227434764 | 0.132585 | 4.077029804 | 0 |
| 17 | 7675179 | G | T | L | M | TP53 | ENST00000269305 | 0.29717 | 0.89143 | 0.4008 | 0.96733 | 0.78906 | 0.3544069638 | 0.89143 | 0.3395527627 | 0.154937 | 4.385651328 | 0 |
| 17 | 7675180 | C | A | Q | H | TP53 | ENST00000269305 | 0.55786 | 0.88922 | 0.53361 | 0.97039 | 0.89694 | 0.5578745075 | 0.97054 | 0.983299315 | 0.310555 | 5.457137201 | 0 |
| 17 | 7675180 | C | G | Q | H | TP53 | ENST00000269305 | 0.56519 | 0.88922 | 0.53361 | 0.97 | 0.89694 | 0.5578745075 | 0.97056 | 0.983299315 | 0.310554 | 5.457137201 | 0 |
| 17 | 7675181 | T | A | Q | L | TP53 | ENST00000269305 | 0.60669 | 0.89304 | 0.4772 | 0.95618 | 0.9211 | 0.5526323016 | 0.96602 | 0.9924185276 | 0.388025 | 5.272519283 | 0 |
| 17 | 7675181 | T | C | Q | R | TP53 | ENST00000269305 | 0.75376 | 0.9054 | 0.53988 | 0.95707 | 0.91569 | 0.5690797917 | 0.95875 | 0.9528914094 | 0.315858 | 6.037094615 | 0 |
| 17 | 7675181 | T | G | Q | P | TP53 | ENST00000269305 | 0.77378 | 0.92039 | 0.49395 | 0.96689 | 0.93593 | 0.8697231358 | 0.97573 | 0.9966308475 | 0.434997 | 9.083753427 | 1 |
| 17 | 7675182 | G | C | Q | E | TP53 | ENST00000269305 | 0.39585 | 0.90501 | 0.48608 | 0.9055 | 0.86653 | 0.4968176654 | 0.90049 | 0.8337612748 | 0.209848 | 5.591221936 | 0 |
| 17 | 7675182 | G | T | Q | K | TP53 | ENST00000269305 | 0.73173 | 0.92418 | 0.54542 | 0.94679 | 0.92069 | 0.5387353172 | 0.94924 | 0.9310956001 | 0.353424 | 5.748283979 | 0 |
| 17 | 7675184 | A | G | V | A | TP53 | ENST00000269305 | 0.81661 | 0.91917 | 0.41168 | 0.97961 | 0.82948 | 0.5936516275 | 0.9705 | 0.8911115583 | 0.291245 | 7.184708407 | 1 |
| 17 | 7675184 | A | T | V | E | TP53 | ENST00000269305 | 0.85353 | 0.94947 | 0.38909 | 0.98059 | 0.88543 | 0.8767571461 | 0.98619 | 0.9540998732 | 0.529103 | 9.083585176 | 1 |
| 17 | 7675185 | C | A | V | L | TP53 | ENST00000269305 | 0.26331 | 0.89481 | 0.38463 | 0.96659 | 0.84747 | 0.4645927562 | 0.92533 | 0.8925095201 | 0.192742 | 5.442187346 | 0 |
| 17 | 7675185 | C | G | V | L | TP53 | ENST00000269305 | 0.26823 | 0.89481 | 0.38463 | 0.96597 | 0.84747 | 0.4645927562 | 0.92585 | 0.8630171418 | 0.192749 | 5.442187346 | 0 |
| 17 | 7675187 | G | A | P | L | TP53 | ENST00000269305 | 0.9274 | 0.89976 | 0.72439 | 0.9689 | 0.9648 | 0.7182201087 | 0.99152 | 0.9926712513 | 0.546788 | 5.151727737 | 0 |
| 17 | 7675187 | G | C | P | R | TP53 | ENST00000269305 | 0.92174 | 0.90412 | 0.72019 | 0.97392 | 0.97512 | 0.7807594301 | 0.99232 | 0.9994040728 | 0.578234 | 8.218610126 | 0 |
| 17 | 7675187 | G | T | P | H | TP53 | ENST00000269305 | 0.92151 | 0.91454 | 0.69975 | 0.97396 | 0.97773 | 0.691345205 | 0.9901 | 0.9995946288 | 0.583417 | 6.451693624 | 0 |
| 17 | 7675188 | G | A | P | S | TP53 | ENST00000269305 | 0.88138 | 0.89849 | 0.64476 | 0.96861 | 0.96773 | 0.6553614094 | 0.98977 | 0.9982047081 | 0.571773 | 6.43767039 | 0 |
| 17 | 7675188 | G | C | P | A | TP53 | ENST00000269305 | 0.83536 | 0.89354 | 0.63974 | 0.96571 | 0.9729 | 0.5809510844 | 0.97738 | 0.9988020658 | 0.57154 | 6.674629752 | 0 |
| 17 | 7675188 | G | T | P | T | TP53 | ENST00000269305 | 0.85894 | 0.91059 | 0.68507 | 0.96277 | 0.96919 | 0.6622369839 | 0.98819 | 0.9989848733 | 0.586315 | 7.21988815 | 0 |
| 17 | 7675189 | G | C | C | W | TP53 | ENST00000269305 | 0.8044 | 0.93153 | 0.80362 | 0.9872 | 0.93863 | 0.7963135244 | 0.98264 | 0.9991814494 | 0.511969 | 7.728099117 | 1 |
| 17 | 7675190 | C | A | C | F | TP53 | ENST00000269305 | 0.86671 | 0.94409 | 0.75705 | 0.98227 | 0.96517 | 0.7379999688 | 0.99503 | 0.9993928671 | 0.537949 | 5.828699236 | 0 |
| 17 | 7675190 | C | G | C | S | TP53 | ENST00000269305 | 0.67549 | 0.92136 | 0.74026 | 0.97314 | 0.94053 | 0.6763243528 | 0.91033 | 0.9687892795 | 0.353601 | 5.519874464 | 0 |
| 17 | 7675190 | C | T | C | Y | TP53 | ENST00000269305 | 0.85048 | 0.95294 | 0.84669 | 0.97841 | 0.97141 | 0.7685044902 | 0.99128 | 0.9993620515 | 0.561428 | 8.757699733 | 1 |
| 17 | 7675191 | A | C | C | G | TP53 | ENST00000269305 | 0.85279 | 0.92269 | 0.71031 | 0.98577 | 0.95753 | 0.7478258773 | 0.99148 | 0.9985363483 | 0.550155 | 8.757699733 | 1 |
| 17 | 7675191 | A | G | C | R | TP53 | ENST00000269305 | 0.86699 | 0.9568 | 0.83126 | 0.98331 | 0.95826 | 0.8573517067 | 0.99477 | 0.9993907213 | 0.557561 | 8.757699733 | 1 |
| 17 | 7675191 | A | T | C | S | TP53 | ENST00000269305 | 0.65359 | 0.92136 | 0.74026 | 0.97585 | 0.93162 | 0.6763243528 | 0.92697 | 0.9555379152 | 0.346874 | 5.519874464 | 0 |
| 17 | 7675193 | G | A | T | I | TP53 | ENST00000269305 | 0.92468 | 0.90482 | 0.63152 | 0.98049 | 0.95133 | 0.5796418584 | 0.97739 | 0.9948955178 | 0.494837 | 5.548206942 | 0 |
| 17 | 7675193 | G | C | T | S | TP53 | ENST00000269305 | 0.88641 | 0.67099 | 0.49628 | 0.9728 | 0.91945 | 0.3163375725 | 0.94319 | 0.9866922498 | 0.289397 | 4.376276064 | 0 |

| | | | | | | | | | | | | | | | | | | |
|---|---|---|---|---|---|---|---|---|---|---|---|---|---|---|---|---|---|---|
| 17 | 7675193 | G | T | T | N | TP53 | ENST00000269305 | 0.90244 | 0.90807 | 0.57605 | 0.98105 | 0.92884 | 0.561322141 | 0.94813 | 0.9956983328 | 0.27412 | 5.778014823 | 0 |
| 17 | 7675194 | T | A | T | S | TP53 | ENST00000269305 | 0.68699 | 0.67099 | 0.49628 | 0.95611 | 0.94876 | 0.3163375725 | 0.94415 | 0.9885896444 | 0.268173 | 4.376276064 | 0 |
| 17 | 7675194 | T | C | T | A | TP53 | ENST00000269305 | 0.44832 | 0.84503 | 0.48152 | 0.95967 | 0.93786 | 0.495833367 | 0.96131 | 0.9868406057 | 0.355891 | 4.132576785 | 0 |
| 17 | 7675194 | T | G | T | P | TP53 | ENST00000269305 | 0.73117 | 0.92579 | 0.47984 | 0.96714 | 0.95097 | 0.6699924725 | 0.9948 | 0.9888476133 | 0.518855 | 4.215953872 | 0 |
| 17 | 7675195 | C | A | K | N | TP53 | ENST00000269305 | 0.59276 | 0.88981 | 0.75097 | 0.98493 | 0.90668 | 0.6809282437 | 0.92399 | 0.9860040545 | 0.200393 | 6.737532682 | 0 |
| 17 | 7675195 | C | G | K | N | TP53 | ENST00000269305 | 0.60047 | 0.88981 | 0.75097 | 0.98475 | 0.90668 | 0.6809282437 | 0.92394 | 0.9860040545 | 0.200395 | 6.737532682 | 0 |
| 17 | 7675196 | T | A | K | M | TP53 | ENST00000269305 | 0.80076 | 0.9303 | 0.64605 | 0.9884 | 0.94243 | 0.7210195121 | 0.96229 | 0.9970416427 | 0.419105 | 7.418292812 | 0 |
| 17 | 7675196 | T | C | K | R | TP53 | ENST00000269305 | 0.7567 | 0.86889 | 0.64498 | 0.9756 | 0.91945 | 0.4701380764 | 0.93013 | 0.969689548 | 0.24806 | 4.43970648 | 0 |
| 17 | 7675196 | T | G | K | T | TP53 | ENST00000269305 | 0.76832 | 0.91712 | 0.73065 | 0.98196 | 0.93554 | 0.6670858891 | 0.96416 | 0.9938115478 | 0.389933 | 5.484834459 | 0 |
| 17 | 7675197 | T | G | K | Q | TP53 | ENST00000269305 | 0.66771 | 0.90195 | 0.67899 | 0.96304 | 0.92275 | 0.6223021132 | 0.93426 | 0.9902443886 | 0.316757 | 6.30587221 | 0 |
| 17 | 7675199 | G | A | A | V | TP53 | ENST00000269305 | 0.84043 | 0.82293 | 0.66877 | 0.96068 | 0.96153 | 0.4564792345 | 0.9809 | 0.9870674014 | 0.525704 | 6.953757079 | 1 |
| 17 | 7675199 | G | C | A | G | TP53 | ENST00000269305 | 0.86112 | 0.89442 | 0.63183 | 0.94901 | 0.95133 | 0.2674713183 | 0.94034 | 0.9882232547 | 0.396359 | 4.145809552 | 0 |
| 17 | 7675199 | G | T | A | D | TP53 | ENST00000269305 | 0.87643 | 0.94168 | 0.75614 | 0.96719 | 0.9729 | 0.5887365387 | 0.98353 | 0.9772150061 | 0.541811 | 5.181060922 | 0 |
| 17 | 7675200 | C | A | A | S | TP53 | ENST00000269305 | 0.72116 | 0.88367 | 0.60848 | 0.95287 | 0.97661 | 0.2919004862 | 0.96016 | 0.9927960756 | 0.504434 | 5.009781909 | 0 |
| 17 | 7675200 | C | G | A | P | TP53 | ENST00000269305 | 0.7737 | 0.92655 | 0.71508 | 0.96046 | 0.99039 | 0.6934556712 | 0.98819 | 0.9975588322 | 0.572539 | 8.446920871 | 1 |
| 17 | 7675200 | C | T | A | T | TP53 | ENST00000269305 | 0.76355 | 0.88956 | 0.6555 | 0.96503 | 0.97624 | 0.4262258555 | 0.95615 | 0.9968959093 | 0.48215 | 7.044763477 | 0 |
| 17 | 7675202 | A | C | L | R | TP53 | ENST00000269305 | 0.90715 | 0.93828 | 0.60504 | 0.98528 | 0.99525 | 0.7399178332 | 0.98946 | 0.9982945323 | 0.559536 | 5.603940848 | 0 |
| 17 | 7675202 | A | G | L | P | TP53 | ENST00000269305 | 0.90993 | 0.94855 | 0.65524 | 0.98829 | 0.99072 | 0.7892921146 | 0.98998 | 0.9982945323 | 0.561973 | 6.000784042 | 0 |
| 17 | 7675202 | A | T | L | Q | TP53 | ENST00000269305 | 0.90106 | 0.90096 | 0.54195 | 0.98325 | 0.99105 | 0.6268169744 | 0.97084 | 0.9982945323 | 0.511957 | 6.835333343 | 0 |
| 17 | 7675203 | G | C | L | V | TP53 | ENST00000269305 | 0.66422 | 0.84991 | 0.44638 | 0.98473 | 0.90711 | 0.3906479197 | 0.94873 | 0.9306654334 | 0.301793 | 5.650301174 | 0 |
| 17 | 7675203 | G | T | L | M | TP53 | ENST00000269305 | 0.56256 | 0.8452 | 0.49151 | 0.97151 | 0.86302 | 0.3015509793 | 0.9442 | 0.532854259 | 0.153619 | 3.676295352 | 0 |
| 17 | 7675204 | T | G | Q | H | TP53 | ENST00000269305 | 0.65163 | 0.88956 | 0.53177 | 0.99007 | 0.98938 | 0.68785484 | 0.99496 | 0.9970133305 | 0.530134 | 6.574853745 | 0 |
| 17 | 7675205 | T | A | Q | L | TP53 | ENST00000269305 | 0.92084 | 0.89442 | 0.467 | 0.98198 | 0.99356 | 0.6340915647 | 0.98638 | 0.9965008497 | 0.517457 | 6.766247413 | 0 |
| 17 | 7675205 | T | C | Q | R | TP53 | ENST00000269305 | 0.86054 | 0.90581 | 0.5334 | 0.95625 | 0.96407 | 0.6276278955 | 0.99634 | 0.9681409001 | 0.501727 | 4.79112548 | 0 |
| 17 | 7675205 | T | G | Q | P | TP53 | ENST00000269305 | 0.9076 | 0.92061 | 0.48988 | 0.97523 | 0.99498 | 0.8566981747 | 0.99648 | 0.9965923429 | 0.552179 | 8.849687384 | 1 |
| 17 | 7675206 | G | C | Q | E | TP53 | ENST00000269305 | 0.8495 | 0.90619 | 0.47529 | 0.96622 | 0.95863 | 0.6168381248 | 0.97369 | 0.9873487949 | 0.436822 | 6.168223629 | 1 |
| 17 | 7675206 | G | T | Q | K | TP53 | ENST00000269305 | 0.88454 | 0.92482 | 0.54331 | 0.92791 | 0.96117 | 0.5863092149 | 0.98841 | 0.9892341496 | 0.457256 | 5.361591905 | 0 |
| 17 | 7675207 | G | C | C | W | TP53 | ENST00000269305 | 0.84241 | 0.93153 | 0.74962 | 0.98671 | 0.96664 | 0.8448867447 | 0.99644 | 0.999732554 | 0.515775 | 9.127507215 | 1 |
| 17 | 7675208 | C | A | C | F | TP53 | ENST00000269305 | 0.77687 | 0.94409 | 0.69489 | 0.97271 | 0.99386 | 0.8087124405 | 0.98775 | 0.9995927215 | 0.560231 | 9.127507215 | 1 |
| 17 | 7675208 | C | T | C | Y | TP53 | ENST00000269305 | 0.75901 | 0.95294 | 0.80904 | 0.97493 | 0.99234 | 0.8294633789 | 0.98305 | 0.9998121858 | 0.56611 | 9.127507215 | 1 |
| 17 | 7675209 | A | C | C | G | TP53 | ENST00000269305 | 0.92694 | 0.92269 | 0.63851 | 0.98504 | 0.99525 | 0.8101690441 | 0.99676 | 0.9998053908 | 0.582637 | 9.127507215 | 1 |
| 17 | 7675209 | A | G | C | R | TP53 | ENST00000269305 | 0.93271 | 0.9568 | 0.79086 | 0.98289 | 0.99498 | 0.9124874308 | 0.99704 | 0.999820888 | 0.582637 | 9.127507215 | 1 |
| 17 | 7675210 | A | C | F | L | TP53 | ENST00000269305 | 0.75248 | 0.91566 | 0.57838 | 0.98854 | 0.96226 | 0.6954221749 | 0.97501 | 0.9935411215 | 0.38596 | 7.873858486 | 1 |
| 17 | 7675210 | A | T | F | L | TP53 | ENST00000269305 | 0.74571 | 0.91566 | 0.57838 | 0.98935 | 0.96226 | 0.6954221749 | 0.97477 | 0.9935411215 | 0.385848 | 7.873858486 | 1 |
| 17 | 7675211 | A | C | F | C | TP53 | ENST00000269305 | 0.91909 | 0.93796 | 0.55852 | 0.98729 | 0.99039 | 0.825341185 | 0.98657 | 0.9931615591 | 0.510619 | 9.177383383 | 1 |
| 17 | 7675211 | A | G | F | S | TP53 | ENST00000269305 | 0.91913 | 0.9568 | 0.52437 | 0.98834 | 0.98799 | 0.832829901 | 0.99395 | 0.9982738495 | 0.54329 | 9.177383383 | 1 |
| 17 | 7675211 | A | T | F | Y | TP53 | ENST00000269305 | 0.66953 | 0.91275 | 0.56737 | 0.97195 | 0.86302 | 0.5803925174 | 0.94256 | 0.8549967408 | 0.313943 | 4.64498979 | 0 |
| 17 | 7675212 | A | C | F | V | TP53 | ENST00000269305 | 0.85018 | 0.92838 | 0.49153 | 0.98464 | 0.99356 | 0.7485199512 | 0.9717 | 0.9984527826 | 0.439927 | 9.177383383 | 1 |
| 17 | 7675212 | A | G | F | L | TP53 | ENST00000269305 | 0.86233 | 0.91566 | 0.57838 | 0.98008 | 0.98799 | 0.6954221749 | 0.97251 | 0.9966059923 | 0.434243 | 7.873858486 | 1 |
| 17 | 7675212 | A | T | F | I | TP53 | ENST00000269305 | 0.84537 | 0.93482 | 0.53795 | 0.97814 | 0.99234 | 0.7723263791 | 0.97197 | 0.9986557961 | 0.423426 | 9.177383383 | 1 |
| 17 | 7675213 | C | A | M | I | TP53 | ENST00000269305 | 0.33699 | 0.62515 | 0.42212 | 0.94283 | 0.7727 | 0.201590896 | 0.91795 | 0.8926575184 | 0.133895 | 1.950340023 | 0 |
| 17 | 7675213 | C | G | M | I | TP53 | ENST00000269305 | 0.34271 | 0.62515 | 0.42212 | 0.93907 | 0.77141 | 0.201590896 | 0.91801 | 0.8845660686 | 0.13391 | 1.950340023 | 0 |
| 17 | 7675213 | C | T | M | I | TP53 | ENST00000269305 | 0.34642 | 0.62515 | 0.42212 | 0.94049 | 0.7727 | 0.201590896 | 0.91799 | 0.3730205894 | -0.0308637 | 1.950340023 | 0 |
| 17 | 7675214 | A | C | M | R | TP53 | ENST00000269305 | 0.72596 | 0.80604 | 0.47682 | 0.96617 | 0.95389 | 0.8934246423 | 0.9942 | 0.9930433035 | 0.511277 | 4.750112935 | 1 |
| 17 | 7675214 | A | G | M | T | TP53 | ENST00000269305 | 0.65562 | 0.73286 | 0.5148 | 0.97697 | 0.92763 | 0.6195741401 | 0.99235 | 0.9768328667 | 0.428613 | 4.750112935 | 1 |
| 17 | 7675214 | A | T | M | K | TP53 | ENST00000269305 | 0.7031 | 0.7813 | 0.4999 | 0.97136 | 0.95243 | 0.8342492816 | 0.99102 | 0.9929726124 | 0.463849 | 4.750112935 | 1 |
| 17 | 7675215 | T | A | M | L | TP53 | ENST00000269305 | 0.28111 | 0.55132 | 0.39411 | 0.89461 | 0.73661 | 0.2434065871 | 0.76047 | 0.1377478838 | 0.131052 | -2.003677141 | 0 |
| 17 | 7675215 | T | C | M | V | TP53 | ENST00000269305 | 0.45285 | 0.59235 | 0.3528 | 0.89788 | 0.78782 | 0.3010324723 | 0.91177 | 0.1924657404 | 0.162035 | 1.49443111 | 0 |
| 17 | 7675215 | T | G | M | L | TP53 | ENST00000269305 | 0.28612 | 0.55132 | 0.39411 | 0.89609 | 0.73661 | 0.2434065871 | 0.72768 | 0.1377478838 | 0.130983 | -2.003677141 | 0 |
| 17 | 7675216 | C | A | K | N | TP53 | ENST00000269305 | 0.69917 | 0.9161 | 0.56308 | 0.969 | 0.97067 | 0.8768943469 | 0.97518 | 0.9984770417 | 0.497111 | 7.3599996912 | 1 |
| 17 | 7675216 | C | G | K | N | TP53 | ENST00000269305 | 0.70768 | 0.9161 | 0.56308 | 0.96798 | 0.97067 | 0.8768943469 | 0.97516 | 0.9984770417 | 0.461628 | 7.3599996912 | 1 |
| 17 | 7675217 | T | A | K | M | TP53 | ENST00000269305 | 0.75177 | 0.93062 | 0.45632 | 0.96872 | 0.98146 | 0.8653063933 | 0.99177 | 0.9979128242 | 0.3714 | 4.701278602 | 1 |
| 17 | 7675217 | T | C | K | R | TP53 | ENST00000269305 | 0.85522 | 0.87129 | 0.45069 | 0.98329 | 0.9755 | 0.7893867629 | 0.99673 | 0.9849087 | 0.42107 | 4.899982349 | 1 |
| 17 | 7675218 | T | C | K | E | TP53 | ENST00000269305 | 0.91045 | 0.93503 | 0.53183 | 0.97825 | 0.99603 | 0.899964782 | 0.99591 | 0.9822501367 | 0.582874 | 7.3599996912 | 1 |
| 17 | 7675218 | T | G | K | Q | TP53 | ENST00000269305 | 0.89255 | 0.90521 | 0.48569 | 0.97835 | 0.98869 | 0.8367125065 | 0.99691 | 0.9968397617 | 0.582874 | 6.225552758 | 1 |
| 17 | 7675219 | G | C | N | K | TP53 | ENST00000269305 | 0.69107 | 0.91327 | 0.47327 | 0.95487 | 0.87491 | 0.6233156751 | 0.95776 | 0.8632742167 | 0.169115 | 5.400769168 | 0 |
| 17 | 7675219 | G | T | N | K | TP53 | ENST00000269305 | 0.68459 | 0.91327 | 0.47327 | 0.95879 | 0.87539 | 0.6233156751 | 0.95853 | 0.7746642232 | 0.0513884 | 5.400769168 | 0 |
| 17 | 7675220 | T | A | N | I | TP53 | ENST00000269305 | 0.83544 | 0.92279 | 0.40659 | 0.98862 | 0.98693 | 0.7346039353 | 0.97259 | 0.9991142154 | 0.388401 | 6.764884019 | 1 |
| 17 | 7675220 | T | C | N | S | TP53 | ENST00000269305 | 0.74157 | 0.85639 | 0.34031 | 0.97774 | 0.95753 | 0.2998532999 | 0.96839 | 0.9579774141 | 0.263987 | 6.45655376 | 0 |
| 17 | 7675220 | T | G | N | T | TP53 | ENST00000269305 | 0.79089 | 0.898 | 0.39975 | 0.98218 | 0.97512 | 0.5042471798 | 0.95127 | 0.9841371775 | 0.276647 | 6.658692748 | 0 |
| 17 | 7675221 | T | A | N | Y | TP53 | ENST00000269305 | 0.86587 | 0.92045 | 0.43994 | 0.99265 | 0.9648 | 0.6092331398 | 0.9782 | 0.9939680099 | 0.388611 | 8.905851801 | 1 |
| 17 | 7675221 | T | C | N | D | TP53 | ENST00000269305 | 0.80519 | 0.83116 | 0.46032 | 0.98075 | 0.90406 | 0.4674462798 | 0.97605 | 0.9727401137 | 0.24639 | 6.104149053 | 0 |
| 17 | 7675221 | T | G | N | H | TP53 | ENST00000269305 | 0.82193 | 0.90085 | 0.40777 | 0.98776 | 0.94318 | 0.556177861 | 0.97959 | 0.9650812745 | 0.279717 | 6.024707542 | 0 |
| 17 | 7675223 | A | C | L | R | TP53 | ENST00000269305 | 0.93031 | 0.9398 | 0.67093 | 0.99534 | 0.97923 | 0.856694911 | 0.99877 | 0.9984565973 | 0.585345 | 5.73550118 | 1 |
| 17 | 7675223 | A | G | L | P | TP53 | ENST00000269305 | 0.93189 | 0.9489 | 0.70738 | 0.99509 | 0.98257 | 0.8785073895 | 0.99861 | 0.9986373782 | 0.585606 | 8.534588601 | 1 |
| 17 | 7675223 | A | T | L | H | TP53 | ENST00000269305 | 0.92194 | 0.92635 | 0.66512 | 0.99347 | 0.98146 | 0.7572569565 | 0.99869 | 0.9964367151 | 0.583389 | 7.736656557 | 1 |
| 17 | 7675224 | G | C | L | V | TP53 | ENST00000269305 | 0.67597 | 0.89783 | 0.53165 | 0.97578 | 0.96773 | 0.5065423155 | 0.99332 | 0.9857584238 | 0.549576 | 6.023575162 | 1 |
| 17 | 7675224 | G | T | L | I | TP53 | ENST00000269305 | 0.68259 | 0.90658 | 0.58917 | 0.97787 | 0.93358 | 0.2886664414 | 0.94429 | 0.9830259681 | 0.406426 | 6.048726259 | 0 |
| 17 | 7675226 | G | A | A | V | TP53 | ENST00000269305 | 0.02124 | 0.57572 | 0.06402 | 0.82301 | 0.76553 | 0.1141322542 | 0.83434 | 0.128028959 | 0.0812225 | 0.305188872 | 0 |
| 17 | 7675226 | G | C | A | G | TP53 | ENST00000269305 | 0.01921 | 0.57989 | 0.06145 | 0.90669 | 0.72246 | 0.1258775695 | 0.79038 | 0.0904841274 | 0.0645485 | 1.224987035 | 0 |
| 17 | 7675226 | G | T | A | D | TP53 | ENST00000269305 | 0.01262 | 0.72994 | 0.07465 | 0.88254 | 0.81761 | 0.1836362488 | 0.89455 | 0.08664891044 | 0.107381 | -1.457605098 | 0 |
| 17 | 7675227 | C | A | A | S | TP53 | ENST00000269305 | 0.00288 | 0.5425 | 0.05821 | 0.64721 | 0.77011 | 0.1103446551 | 0.75185 | 0.04683424905 | 0.0653491 | -0.7140157162 | 0 |
| 17 | 7675227 | C | G | A | P | TP53 | ENST00000269305 | 0.01643 | 0.67759 | 0.07836 | 0.83445 | 0.80298 | 0.3616874067 | 0.90499 | 0.2910496294 | 0.097827 | 0.5744775121 | 0 |
| 17 | 7675227 | C | T | A | T | TP53 | ENST00000269305 | 0.00809 | 0.56523 | 0.05913 | 0.80735 | 0.74517 | 0.1197413893 | 0.7728 | 0.05353296368 | 0.0616065 | -0.7653766682 | 0 |
| 17 | 7675229 | G | A | P | L | TP53 | ENST00000269305 | 0.56845 | 0.89404 | 0.50341 | 0.97029 | 0.78658 | 0.3868310159 | 0.97888 | 0.9662901163 | 0.224004 | 5.992522385 | 0 |
| 17 | 7675229 | G | T | P | H | TP53 | ENST00000269305 | 0.56943 | 0.91395 | 0.47754 | 0.96076 | 0.80238 | 0.353529198 | 0.95423 | 0.9913056493 | 0.231729 | 5.685372002 | 0 |
| 17 | 7675230 | G | A | P | S | TP53 | ENST00000269305 | 0.30956 | 0.8964 | 0.40329 | 0.94816 | 0.79577 | 0.3432561165 | 0.94559 | 0.9635483027 | 0.185421 | 3.757085962 | 0 |
| 17 | 7675230 | G | C | P | A | TP53 | ENST00000269305 | 0.2787 | 0.87659 | 0.3942 | 0.94278 | 0.77463 | 0.2645704224 | 0.94599 | 0.9084156156 | 0.184309 | 4.735117001 | 0 |
| 17 | 7675230 | G | T | P | T | TP53 | ENST00000269305 | 0.21641 | 0.90791 | 0.44557 | 0.91592 | 0.75822 | 0.3563171577 | 0.89274 | 0.9214270115 | 0.161191 | 4.732607098 | 0 |
| 17 | 7675232 | G | A | S | F | TP53 | ENST00000269305 | 0.92937 | 0.90827 | 0.51551 | 0.98533 | 0.98183 | 0.8758460683 | 0.98766 | 0.9995063543 | 0.558936 | 8.259737906 | 1 |
| 17 | 7675232 | G | C | S | C | TP53 | ENST00000269305 | 0.92237 | 0.89365 | 0.48053 | 0.98373 | 0.97848 | 0.8156360897 | 0.97952 | 0.9991958737 | 0.553125 | 8.259737906 | 0 |
| 17 | 7675232 | G | T | S | Y | TP53 | ENST00000269305 | 0.90544 | 0.92626 | 0.54662 | 0.98491 | 0.97923 | 0.8691244263 | 0.99471 | 0.9995349646 | 0.583331 | 8.259737906 | 1 |
| 17 | 7675233 | A | C | S | A | TP53 | ENST00000269305 | 0.76084 | 0.87322 | 0.44952 | 0.97796 | 0.97661 | 0.6689558165 | 0.9544 | 0.9949840307 | 0.413239 | 6.346657324 | 0 |
| 17 | 7675233 | A | G | S | P | TP53 | ENST00000269305 | 0.81135 | 0.92429 | 0.48153 | 0.98136 | 0.99296 | 0.930378084 | 0.9883 | 0.9993859529 | 0.573683 | 8.259737906 | 1 |
| 17 | 7675233 | A | T | S | T | TP53 | ENST00000269305 | 0.71979 | 0.88544 | 0.47154 | 0.97791 | 0.98146 | 0.7347877373 | 0.96781 | 0.9955838323 | 0.464114 | 8.259737906 | 1 |
| 17 | 7675235 | T | A | Y | F | TP53 | ENST00000269305 | 0.74157 | 0.886 | 0.67069 | 0.98257 | 0.96189 | 0.4889026394 | 0.95983 | 0.950124681 | 0.360164 | 3.962871947 | 0 |
| 17 | 7675235 | T | C | Y | C | TP53 | ENST00000269305 | 0.9322 | 0.9234 | 0.75835 | 0.9864 | 0.99876 | 0.8451041755 | 0.99438 | 0.9996919632 | 0.580142 | 7.512555089 | 1 |
| 17 | 7675235 | T | G | Y | S | TP53 | ENST00000269305 | 0.92225 | 0.93697 | 0.68734 | 0.98908 | 0.997 | 0.84575118 | 0.99537 | 0.9994581342 | 0.580142 | 8.598265368 | 1 |
| 17 | 7675236 | A | C | Y | D | TP53 | ENST00000269305 | 0.95332 | 0.9568 | 0.75341 | 0.9856 | 0.99843 | 0.8944064344 | 0.99779 | 0.9997919202 | 0.582939 | 8.598265368 | 1 |
| 17 | 7675236 | A | G | Y | H | TP53 | ENST00000269305 | 0.95333 | 0.92711 | 0.76856 | 0.98813 | 0.99414 | 0.754573932 | 0.98791 | 0.9993798733 | 0.582212 | 6.527826707 | 1 |
| 17 | 7675236 | A | T | Y | N | TP53 | ENST00000269305 | 0.95244 | 0.94997 | 0.75966 | 0.98779 | 0.99525 | 0.8537901454 | 0.98616 | 0.9998053908 | 0.564512 | 8.598265368 | 1 |
| 17 | 7675995 | G | A | T | M | TP53 | ENST00000269305 | 0.90779 | 0.90115 | 0.52665 | 0.97636 | 0.98183 | 0.5756093827 | 0.97663 | 0.999397397 | 0.513779 | 5.178351102 | 0 |
| 17 | 7675995 | G | C | T | R | TP53 | ENST00000269305 | 0.89684 | 0.91287 | 0.56191 | 0.97391 | 0.99471 | 0.8032064366 | 0.99416 | 0.9994321465 | 0.582935 | 8.077827595 | 1 |
| 17 | 7675995 | G | T | T | K | TP53 | ENST00000269305 | 0.89634 | 0.91883 | 0.58464 | 0.97549 | 0.99471 | 0.7500744944 | 0.99013 | 0.9995650649 | 0.582889 | 6.558098756 | 1 |
| 17 | 7675996 | T | A | T | S | TP53 | ENST00000269305 | 0.86545 | 0.87806 | 0.46454 | 0.97409 | 0.97364 | 0.3970907808 | 0.95975 | 0.9910257459 | 0.394418 | 6.632046042 | 0 |
| 17 | 7675996 | T | C | T | A | TP53 | ENST00000269305 | 0.85989 | 0.88099 | 0.44544 | 0.97062 | 0.98834 | 0.5484931968 | 0.98243 | 0.996876359 | 0.493338 | 5.899890165 | 0 |
| 17 | 7675996 | T | G | T | P | TP53 | ENST00000269305 | 0.91468 | 0.92696 | 0.44735 | 0.96792 | 0.98904 | 0.7955108796 | 0.99073 | 0.9967903495 | 0.566581 | 7.758974511 | 1 |
| 17 | 7675997 | G | C | C | W | TP53 | ENST00000269305 | 0.56082 | 0.87498 | 0.5897 | 0.95906 | 0.80831 | 0.740726909 | 0.90183 | 0.8475254178 | 0.165741 | 4.892144189 | 0 |
| 17 | 7675998 | C | A | C | F | TP53 | ENST00000269305 | 0.70927 | 0.94196 | 0.51943 | 0.95557 | 0.93241 | 0.6428054759 | 0.91815 | 0.863723617 | 0.271296 | 5.642494854 | 0 |
| 17 | 7675998 | C | G | C | S | TP53 | ENST00000269305 | 0.34489 | 0.91763 | 0.48108 | 0.89527 | 0.87829 | 0.5856315147 | 0.82986 | 0.8218544722 | 0.202197 | 4.906955945 | 0 |
| 17 | 7675998 | C | T | C | Y | TP53 | ENST00000269305 | 0.3915 | 0.95105 | 0.66584 | 0.94673 | 0.89918 | 0.674771049 | 0.89045 | 0.8068944024 | 0.243661 | 4.169492503 | 0 |

| | | | | | | | | | | | | | | | | | | |
|---|---|---|---|---|---|---|---|---|---|---|---|---|---|---|---|---|---|---|
| 17 | 7675999 | A | G | C | R | TP53 | ENST00000269305 | 0.61761 | 0.95633 | 0.64582 | 0.97854 | 0.96627 | 0.7981489967 | 0.95796 | 0.9764578938 | 0.34284 | 7.377320566 | 1 |
| 17 | 7675999 | A | T | C | S | TP53 | ENST00000269305 | 0.26888 | 0.91763 | 0.48108 | 0.95507 | 0.90052 | 0.5856315147 | 0.85976 | 0.3985470235 | 0.0421429 | 4.906955945 | 0 |
| 17 | 7676001 | G | A | T | I | TP53 | ENST00000269305 | 0.70736 | 0.85803 | 0.55549 | 0.95225 | 0.95753 | 0.6033412441 | 0.95639 | 0.9972906709 | 0.391925 | 6.825904302 | 0 |
| 17 | 7676001 | G | C | T | S | TP53 | ENST00000269305 | 0.61477 | 0.64573 | 0.38796 | 0.93099 | 0.90624 | 0.2848983008 | 0.94035 | 0.974604249 | 0.276022 | 5.44760376 | 0 |
| 17 | 7676001 | G | T | T | N | TP53 | ENST00000269305 | 0.67294 | 0.67055 | 0.47927 | 0.94255 | 0.93162 | 0.5565735165 | 0.94286 | 0.9934393764 | 0.252137 | 5.247879718 | 0 |
| 17 | 7676002 | T | A | T | S | TP53 | ENST00000269305 | 0.62241 | 0.64573 | 0.38796 | 0.95015 | 0.91569 | 0.2848983008 | 0.95713 | 0.9696699381 | 0.292779 | 5.44760376 | 0 |
| 17 | 7676002 | T | C | T | A | TP53 | ENST00000269305 | 0.67963 | 0.57713 | 0.36356 | 0.95556 | 0.92561 | 0.5087155225 | 0.95307 | 0.9853899479 | 0.261558 | 5.792301612 | 0 |
| 17 | 7676002 | T | G | T | P | TP53 | ENST00000269305 | 0.7548 | 0.90482 | 0.3699 | 0.96696 | 0.95826 | 0.7031773605 | 0.98287 | 0.9952068925 | 0.427822 | 5.48753155 | 0 |
| 17 | 7676004 | A | C | V | G | TP53 | ENST00000269305 | 0.76379 | 0.92614 | 0.4676 | 0.98329 | 0.97067 | 0.7209103078 | 0.98501 | 0.9948195815 | 0.515203 | 7.390846812 | 0 |
| 17 | 7676004 | A | G | V | A | TP53 | ENST00000269305 | 0.5824 | 0.87327 | 0.61669 | 0.93974 | 0.86951 | 0.4588011687 | 0.92989 | 0.8486627936 | 0.227529 | 4.705831753 | 0 |
| 17 | 7676004 | A | T | V | E | TP53 | ENST00000269305 | 0.77021 | 0.94843 | 0.59826 | 0.97961 | 0.9703 | 0.7632228165 | 0.97043 | 0.9949980974 | 0.402994 | 8.938762385 | 0 |
| 17 | 7676005 | C | A | V | L | TP53 | ENST00000269305 | 0.43987 | 0.87051 | 0.59162 | 0.96239 | 0.90668 | 0.5423494701 | 0.9099 | 0.8737661242 | 0.277801 | 6.189931895 | 0 |
| 17 | 7676005 | C | G | V | L | TP53 | ENST00000269305 | 0.44631 | 0.87051 | 0.59162 | 0.9554 | 0.90668 | 0.5423494701 | 0.91017 | 0.8737661242 | 0.277802 | 6.189931895 | 0 |
| 17 | 7676005 | C | T | V | M | TP53 | ENST00000269305 | 0.5001 | 0.89702 | 0.63712 | 0.96402 | 0.92356 | 0.5250524311 | 0.93672 | 0.9724715948 | 0.258902 | 6.638872102 | 0 |
| 17 | 7676007 | G | A | S | F | TP53 | ENST00000269305 | 0.79919 | 0.90769 | 0.62201 | 0.97431 | 0.98294 | 0.5956724579 | 0.97822 | 0.9984715581 | 0.49995 | 7.856066825 | 0 |
| 17 | 7676007 | G | C | S | C | TP53 | ENST00000269305 | 0.80479 | 0.89211 | 0.5905 | 0.95902 | 0.9796 | 0.5612701278 | 0.95236 | 0.9983863831 | 0.359989 | 6.725902561 | 0 |
| 17 | 7676007 | G | T | S | Y | TP53 | ENST00000269305 | 0.7745 | 0.92573 | 0.6481 | 0.9689 | 0.98034 | 0.5954800903 | 0.97555 | 0.9984715581 | 0.453921 | 7.619961463 | 0 |
| 17 | 7676008 | A | C | S | A | TP53 | ENST00000269305 | 0.81865 | 0.82884 | 0.55805 | 0.96054 | 0.95207 | 0.2957675579 | 0.9462 | 0.9902668595 | 0.336906 | 5.795588243 | 0 |
| 17 | 7676008 | A | G | S | P | TP53 | ENST00000269305 | 0.86851 | 0.92269 | 0.59374 | 0.96716 | 0.98869 | 0.738176541 | 0.97423 | 0.9986108541 | 0.518338 | 8.64061016 | 0 |
| 17 | 7676008 | A | T | S | T | TP53 | ENST00000269305 | 0.77758 | 0.85564 | 0.58267 | 0.96227 | 0.96371 | 0.4328909122 | 0.94987 | 0.9843044877 | 0.367708 | 6.274463599 | 0 |
| 17 | 7676009 | C | A | K | N | TP53 | ENST00000269305 | 0.65226 | 0.9161 | 0.68786 | 0.98353 | 0.95863 | 0.781915357 | 0.98142 | 0.9991707802 | 0.242277 | 6.074127797 | 0 |
| 17 | 7676010 | T | A | K | M | TP53 | ENST00000269305 | 0.81606 | 0.93062 | 0.57775 | 0.98236 | 0.97997 | 0.8126442361 | 0.97637 | 0.9982738495 | 0.407981 | 8.387480822 | 0 |
| 17 | 7676010 | T | C | K | R | TP53 | ENST00000269305 | 0.77972 | 0.87103 | 0.5693 | 0.97769 | 0.95826 | 0.6194543459 | 0.97861 | 0.9899033308 | 0.310679 | 5.885090456 | 0 |
| 17 | 7676010 | T | G | K | T | TP53 | ENST00000269305 | 0.78384 | 0.91805 | 0.66158 | 0.98004 | 0.97885 | 0.7860149113 | 0.97474 | 0.9993194342 | 0.383211 | 7.22528668 | 0 |
| 17 | 7676011 | T | C | K | E | TP53 | ENST00000269305 | 0.88342 | 0.93489 | 0.65104 | 0.97902 | 0.98183 | 0.8120749415 | 0.97842 | 0.9958615899 | 0.432393 | 7.116373675 | 1 |
| 17 | 7676011 | T | G | K | Q | TP53 | ENST00000269305 | 0.86112 | 0.90521 | 0.60385 | 0.97758 | 0.97215 | 0.7423039954 | 0.97995 | 0.99090904 | 0.343527 | 6.349124215 | 0 |
| 17 | 7676013 | G | A | A | V | TP53 | ENST00000269305 | 0.82116 | 0.89656 | 0.55695 | 0.97841 | 0.97475 | 0.4808104215 | 0.96304 | 0.9953991771 | 0.406181 | 6.561620885 | 0 |
| 17 | 7676013 | G | C | A | G | TP53 | ENST00000269305 | 0.81881 | 0.89749 | 0.50867 | 0.97128 | 0.95645 | 0.3351035264 | 0.96861 | 0.9932703376 | 0.402426 | 6.041162394 | 0 |
| 17 | 7676013 | G | T | A | D | TP53 | ENST00000269305 | 0.82918 | 0.94218 | 0.65031 | 0.9772 | 0.98869 | 0.6564272599 | 0.9687 | 0.9984715581 | 0.545649 | 8.936785939 | 0 |
| 17 | 7676014 | C | A | A | S | TP53 | ENST00000269305 | 0.61649 | 0.88544 | 0.48499 | 0.97526 | 0.95097 | 0.3007087704 | 0.95375 | 0.9743137956 | 0.370458 | 6.044335308 | 0 |
| 17 | 7676014 | C | G | A | P | TP53 | ENST00000269305 | 0.6704 | 0.92717 | 0.60029 | 0.97036 | 0.97215 | 0.6570488293 | 0.96962 | 0.9960676432 | 0.464459 | 5.736566107 | 0 |
| 17 | 7676014 | C | T | A | T | TP53 | ENST00000269305 | 0.64337 | 0.893 | 0.52767 | 0.96993 | 0.95426 | 0.406059291 | 0.96065 | 0.9831724167 | 0.371133 | 4.933495443 | 0 |
| 17 | 7676016 | G | A | T | I | TP53 | ENST00000269305 | 0.79247 | 0.90622 | 0.55363 | 0.94174 | 0.97141 | 0.5346833005 | 0.95507 | 0.9981380701 | 0.406846 | 6.263466054 | 0 |
| 17 | 7676016 | G | C | T | R | TP53 | ENST00000269305 | 0.80266 | 0.91302 | 0.50306 | 0.95332 | 0.97512 | 0.6286553799 | 0.96961 | 0.9981380701 | 0.429014 | 6.717059097 | 0 |
| 17 | 7676016 | G | T | T | K | TP53 | ENST00000269305 | 0.81685 | 0.91936 | 0.52287 | 0.95655 | 0.96737 | 0.6262059674 | 0.96107 | 0.9972272515 | 0.413992 | 6.414159877 | 0 |
| 17 | 7676017 | T | A | T | S | TP53 | ENST00000269305 | 0.66509 | 0.88325 | 0.40555 | 0.96749 | 0.95462 | 0.2772197372 | 0.96471 | 0.9873425987 | 0.362281 | 6.29785801 | 0 |
| 17 | 7676017 | T | C | T | A | TP53 | ENST00000269305 | 0.68683 | 0.8947 | 0.386 | 0.95414 | 0.95133 | 0.4572335016 | 0.96845 | 0.9875621796 | 0.367834 | 5.793953685 | 0 |
| 17 | 7676017 | T | G | T | P | TP53 | ENST00000269305 | 0.74404 | 0.92744 | 0.38337 | 0.96878 | 0.97327 | 0.5919757477 | 0.96672 | 0.9970652461 | 0.415653 | 6.746665575 | 0 |
| 17 | 7676019 | C | A | G | V | TP53 | ENST00000269305 | 0.71281 | 0.9234 | 0.51577 | 0.97735 | 0.94765 | 0.6089981234 | 0.96565 | 0.9982738495 | 0.347093 | 6.790810649 | 0 |
| 17 | 7676019 | C | G | G | A | TP53 | ENST00000269305 | 0.68091 | 0.90449 | 0.50515 | 0.96428 | 0.92682 | 0.4262275631 | 0.97322 | 0.9203212318 | 0.277867 | 7.145091546 | 0 |
| 17 | 7676019 | C | T | G | E | TP53 | ENST00000269305 | 0.71562 | 0.92036 | 0.59607 | 0.96119 | 0.92924 | 0.4807511785 | 0.94129 | 0.9757712483 | 0.183227 | 6.177683645 | 0 |
| 17 | 7676020 | C | A | G | W | TP53 | ENST00000269305 | 0.60647 | 0.91663 | 0.59898 | 0.97246 | 0.92151 | 0.5329090404 | 0.97507 | 0.9988600016 | 0.331286 | 8.995488465 | 0 |
| 17 | 7676020 | C | G | G | R | TP53 | ENST00000269305 | 0.55175 | 0.91805 | 0.6239 | 0.96246 | 0.92356 | 0.522557161 | 0.95727 | 0.9970909357 | 0.293522 | 6.96182898 | 0 |
| 17 | 7676022 | G | A | S | F | TP53 | ENST00000269305 | 0.83437 | 0.90827 | 0.56211 | 0.95371 | 0.90581 | 0.6276417905 | 0.96727 | 0.9962499738 | 0.361356 | 6.103246245 | 0 |
| 17 | 7676022 | G | C | S | C | TP53 | ENST00000269305 | 0.82217 | 0.89365 | 0.52365 | 0.94028 | 0.89603 | 0.5776069401 | 0.95376 | 0.9955242872 | 0.286371 | 6.840204836 | 0 |
| 17 | 7676022 | G | T | S | Y | TP53 | ENST00000269305 | 0.79247 | 0.92626 | 0.59462 | 0.95232 | 0.90885 | 0.6204545499 | 0.97086 | 0.9976168871 | 0.362027 | 8.65938987 | 0 |
| 17 | 7676023 | A | C | S | A | TP53 | ENST00000269305 | 0.6061 | | | 0.80283 | | 0.3212130006 | | 0.77784127 | 0.23155 | 5.48763436 | 0 |
| 17 | 7676023 | A | G | S | P | TP53 | ENST00000269305 | 0.67533 | | | 0.82207 | | 0.7117716978 | | 0.9955627918 | 0.395276 | 6.108283332 | 0 |
| 17 | 7676023 | A | T | S | T | TP53 | ENST00000269305 | 0.58211 | | | 0.77268 | | 0.4715668625 | | 0.9146715675 | 0.353538 | 4.992832998 | 0 |
| 17 | 7676024 | A | C | H | Q | TP53 | ENST00000269305 | 0.07419 | | | 0.78113 | | 0.1689933839 | | 0.06652393192 | -0.423557 | 0.4089931295 | 0 |
| 17 | 7676024 | A | T | H | Q | TP53 | ENST00000269305 | 0.07194 | | | 0.78113 | | 0.1689933839 | | 0.06652393192 | -0.422118 | 0.4089931295 | 0 |
| 17 | 7676025 | T | A | H | L | TP53 | ENST00000269305 | 0.47747 | 0.71745 | 0.27585 | 0.65356 | 0.81818 | 0.3840301803 | 0.95552 | 0.8620537519 | 0.227912 | 5.283394129 | 0 |
| 17 | 7676025 | T | C | H | R | TP53 | ENST00000269305 | 0.33914 | 0.65732 | 0.29165 | 0.5145 | 0.77591 | 0.2348306754 | 0.91988 | 0.538482964 | 0.197857 | 3.392801741 | 0 |
| 17 | 7676025 | T | G | H | P | TP53 | ENST00000269305 | 0.40109 | 0.90412 | 0.26077 | 0.79825 | 0.85117 | 0.6542624808 | 0.93644 | 0.6693623662 | 0.224253 | 2.968613908 | 0 |
| 17 | 7676026 | G | A | H | Y | TP53 | ENST00000269305 | 0.33441 | 0.83714 | 0.30076 | 0.91288 | 0.79637 | 0.2444802595 | 0.8476 | 0.8071393371 | 0.18496 | 4.780805456 | 0 |
| 17 | 7676026 | G | C | H | D | TP53 | ENST00000269305 | 0.33823 | | | 0.61906 | | 0.4226621046 | 0.93391 | 0.2341152249 | -0.0621203 | 3.684123433 | 0 |
| 17 | 7676026 | G | T | H | N | TP53 | ENST00000269305 | 0.23174 | | | 0.41638 | | 0.2011545195 | | 0.1363924593 | -0.111071 | 1.052253007 | 0 |
| 17 | 7676027 | C | A | L | F | TP53 | ENST00000269305 | 0.4346 | | | 0.80012 | | 0.2852790974 | 0.95036 | 0.8787848949 | 0.222698 | 8.031861964 | 0 |
| 17 | 7676027 | C | G | L | F | TP53 | ENST00000269305 | 0.44101 | | | 0.796 | | 0.2852790974 | 0.95167 | 0.740955472 | 0.0670093 | 8.031861964 | 0 |
| 17 | 7676028 | A | C | L | W | TP53 | ENST00000269305 | 0.84233 | | | 0.86723 | | 0.4663044132 | 0.94638 | 0.9067656994 | 0.416912 | 8.960990323 | 0 |
| 17 | 7676028 | A | G | L | S | TP53 | ENST00000269305 | 0.68315 | | | 0.79755 | | 0.4454029849 | 0.95037 | 0.8295521736 | 0.361268 | 6.19515721 | 0 |
| 17 | 7676029 | A | C | L | V | TP53 | ENST00000269305 | 0.2903 | 0.709 | 0.39988 | 0.94845 | 0.82892 | 0.2631562197 | 0.89422 | 0.4201987684 | 0.207638 | 6.065209781 | 0 |
| 17 | 7676029 | A | T | L | M | TP53 | ENST00000269305 | 0.47658 | 0.88525 | 0.44246 | 0.9661 | 0.81006 | 0.2035708403 | 0.91916 | 0.8001813293 | 0.202686 | 8.960990323 | 0 |
| 17 | 7676030 | G | C | F | L | TP53 | ENST00000269305 | 0.70447 | 0.91566 | 0.68777 | 0.84909 | 0.92884 | 0.6405131908 | 0.97468 | 0.9988388419 | 0.384141 | 5.751722757 | 0 |
| 17 | 7676030 | G | T | F | L | TP53 | ENST00000269305 | 0.69788 | 0.91566 | 0.68777 | 0.85737 | 0.92884 | 0.6405131908 | 0.97488 | 0.9988388419 | 0.384142 | 5.751722757 | 0 |
| 17 | 7676031 | A | C | F | C | TP53 | ENST00000269305 | 0.89555 | 0.93796 | 0.68149 | 0.83629 | 0.99138 | 0.8240918629 | 0.99112 | 0.9991069436 | 0.559033 | 5.911399543 | 1 |
| 17 | 7676031 | A | G | F | S | TP53 | ENST00000269305 | 0.8956 | 0.9568 | 0.65604 | 0.88392 | 0.99138 | 0.8460898236 | 0.99339 | 0.9993373752 | 0.558909 | 8.558386291 | 1 |
| 17 | 7676031 | A | T | F | Y | TP53 | ENST00000269305 | 0.8199 | 0.91275 | 0.67759 | 0.89368 | 0.96846 | 0.5702422873 | 0.96444 | 0.9930666685 | 0.416975 | 7.198928346 | 0 |
| 17 | 7676032 | A | C | F | V | TP53 | ENST00000269305 | 0.81638 | 0.92838 | 0.624 | 0.88271 | 0.98477 | 0.7040596569 | 0.99183 | 0.9993410707 | 0.461628 | 5.95912318 | 1 |
| 17 | 7676032 | A | G | F | L | TP53 | ENST00000269305 | 0.82958 | | | 0.88011 | | 0.6405131908 | | 0.999166131 | 0.501539 | 5.751722757 | 0 |
| 17 | 7676032 | A | T | F | I | TP53 | ENST00000269305 | 0.81143 | | | 0.86221 | | 0.7079912459 | | 0.9994008541 | 0.473239 | 5.638611849 | 1 |
| 17 | 7676034 | C | A | G | V | TP53 | ENST00000269305 | 0.30807 | | | 0.86777 | | 0.5499505441 | | 0.7472158426 | 0.228609 | 4.954022487 | 0 |
| 17 | 7676034 | C | G | G | A | TP53 | ENST00000269305 | 0.24323 | 0.88793 | 0.33676 | 0.74467 | 0.75822 | 0.4351626096 | 0.94774 | 0.3262233436 | -0.142587 | 4.625248872 | 0 |
| 17 | 7676034 | C | T | G | D | TP53 | ENST00000269305 | 0.20306 | | | 0.81805 | | 0.6435967182 | | 0.5416831374 | 0.039297 | 5.28023147 | 0 |
| 17 | 7676035 | C | A | G | C | TP53 | ENST00000269305 | 0.4094 | | | 0.85425 | | 0.5959718815 | | 0.9018861651 | 0.322078 | 6.307533678 | 0 |
| 17 | 7676035 | C | G | G | R | TP53 | ENST00000269305 | 0.19767 | | | 0.80436 | | 0.6154287558 | | 0.697508812 | 0.135536 | 4.618367156 | 0 |
| 17 | 7676035 | C | T | G | S | TP53 | ENST00000269305 | 0.28461 | 0.89044 | 0.29879 | 0.76116 | 0.70671 | 0.4458231668 | 0.91804 | 0.6316589713 | 0.155812 | 5.514005248 | 0 |
| 17 | 7676037 | A | C | L | R | TP53 | ENST00000269305 | 0.70519 | 0.93946 | 0.56336 | 0.97403 | 0.95863 | 0.9374488768 | 0.98934 | 0.9986277819 | 0.501177 | 8.918287811 | 1 |
| 17 | 7676037 | A | G | L | P | TP53 | ENST00000269305 | 0.70943 | 0.94878 | 0.60218 | 0.98195 | 0.95935 | 0.9474194362 | 0.98779 | 0.9989359975 | 0.487441 | 8.918287811 | 1 |
| 17 | 7676037 | A | T | L | Q | TP53 | ENST00000269305 | 0.69564 | 0.90222 | 0.48164 | 0.97451 | 0.96117 | 0.8772264768 | 0.98352 | 0.9986277819 | 0.421605 | 8.918287811 | 1 |
| 17 | 7676038 | G | C | L | V | TP53 | ENST00000269305 | 0.19065 | 0.7725 | 0.41988 | 0.95341 | 0.75755 | 0.4162154013 | 0.88498 | 0.4380608201 | 0.152722 | 4.112956342 | 0 |
| 17 | 7676040 | C | A | R | L | TP53 | ENST00000269305 | 0.29512 | 0.90835 | 0.18459 | 0.94766 | 0.86047 | 0.374165823 | 0.98137 | 0.9724482298 | 0.313988 | 5.075063609 | 1 |
| 17 | 7676040 | C | G | R | P | TP53 | ENST00000269305 | 0.4349 | 0.93628 | 0.18743 | 0.95672 | 0.83997 | 0.7965287794 | 0.99007 | 0.9032248855 | 0.431694 | 7.609550191 | 1 |
| 17 | 7676040 | C | T | R | H | TP53 | ENST00000269305 | 0.19528 | 0.70413 | 0.16718 | 0.92506 | 0.83943 | 0.1932140997 | 0.94199 | 0.04055010875 | 0.0320992 | 2.677024773 | 0 |
| 17 | 7676041 | G | A | R | C | TP53 | ENST00000269305 | 0.38788 | 0.93051 | 0.22987 | 0.89448 | 0.7675 | 0.4456848505 | 0.97207 | 0.7571222186 | 0.191065 | 4.046618159 | 0 |
| 17 | 7676041 | G | C | R | G | TP53 | ENST00000269305 | 0.59096 | 0.92161 | 0.1614 | 0.94084 | 0.83116 | 0.4538489394 | 0.96309 | 0.7138000131 | 0.204773 | 6.013708302 | 0 |
| 17 | 7676041 | G | T | R | S | TP53 | ENST00000269305 | 0.35003 | 0.9015 | 0.1621 | 0.89564 | 0.80118 | 0.295801297 | 0.93841 | 0.6278755665 | 0.17205 | 4.882028213 | 0 |
| 17 | 7676042 | G | C | F | L | TP53 | ENST00000269305 | 0.70728 | 0.9155 | 0.70868 | 0.87815 | 0.90007 | 0.8447340652 | 0.93825 | 0.9884454012 | 0.316817 | 4.56466865 | 0 |
| 17 | 7676042 | G | T | F | L | TP53 | ENST00000269305 | 0.70069 | 0.9155 | 0.70868 | 0.87956 | 0.90007 | 0.8447340652 | 0.93855 | 0.9884454012 | 0.316818 | 4.56466865 | 0 |
| 17 | 7676043 | A | C | F | C | TP53 | ENST00000269305 | 0.91503 | 0.93796 | 0.71092 | 0.96675 | 0.99525 | 0.9302398642 | 0.99574 | 0.9994921684 | 0.572235 | 8.413780125 | 1 |
| 17 | 7676043 | A | G | F | S | TP53 | ENST00000269305 | 0.91512 | 0.9568 | 0.68295 | 0.97556 | 0.99138 | 0.9375601605 | 0.99524 | 0.9991144538 | 0.57212 | 8.413780125 | 1 |
| 17 | 7676044 | A | G | F | L | TP53 | ENST00000269305 | 0.68867 | 0.9155 | 0.70868 | 0.95832 | 0.92884 | 0.8447340652 | 0.93729 | 0.9888008833 | 0.338239 | 4.56466865 | 0 |
| 17 | 7676044 | A | T | F | I | TP53 | ENST00000269305 | 0.69348 | 0.93482 | 0.68557 | 0.97736 | 0.94949 | 0.8950653784 | 0.97675 | 0.9989908338 | 0.459363 | 8.413780125 | 1 |
| 17 | 7676046 | C | A | G | V | TP53 | ENST00000269305 | 0.38874 | 0.92209 | 0.41717 | 0.97173 | 0.87345 | 0.6993249687 | 0.96465 | 0.9940646291 | 0.239783 | 6.703525343 | 0 |
| 17 | 7676046 | C | G | G | A | TP53 | ENST00000269305 | 0.33952 | 0.87042 | 0.41096 | 0.94292 | 0.87829 | 0.5045861951 | 0.92598 | 0.8041670918 | 0.170664 | 5.367434071 | 0 |
| 17 | 7676046 | C | T | G | D | TP53 | ENST00000269305 | 0.17742 | 0.8072 | 0.51597 | 0.90716 | 0.87 | 0.5627221733 | 0.91778 | 0.5888659358 | 0.245964 | 2.80398252 | 0 |
| 17 | 7676047 | C | A | G | C | TP53 | ENST00000269305 | 0.47174 | 0.90412 | 0.44868 | 0.98127 | 0.87148 | 0.7051994303 | 0.97099 | 0.9700192213 | 0.248264 | 6.425527303 | 0 |
| 17 | 7676047 | C | G | G | R | TP53 | ENST00000269305 | 0.31998 | 0.91712 | 0.49712 | 0.96343 | 0.90406 | 0.6347443943 | 0.97246 | 0.9237528443 | 0.321078 | 5.549883728 | 0 |
| 17 | 7676047 | C | T | G | S | TP53 | ENST00000269305 | 0.28965 | 0.78847 | 0.38014 | 0.95012 | 0.85012 | 0.4840537618 | 0.92121 | 0.5570960641 | 0.189721 | 4.596615425 | 0 |
| 17 | 7676049 | T | A | Y | F | TP53 | ENST00000269305 | 0.33228 | 0.71232 | 0.52441 | 0.96158 | 0.86149 | 0.4910205694 | 0.90965 | 0.8307790756 | 0.185125 | 4.391799974 | 0 |
| 17 | 7676049 | T | C | Y | C | TP53 | ENST00000269305 | 0.42615 | 0.923 | 0.59719 | 0.97239 | 0.96627 | 0.7419704885 | 0.96969 | 0.99616611 | 0.371733 | 4.613729413 | 0 |

| | | | | | | | | | | | | | | | | | |
|---|---|---|---|---|---|---|---|---|---|---|---|---|---|---|---|---|---|
| 17 | 7676049 | T | G | Y | S | TP53 | ENST00000269305 | 0.61 | 0.90835 | 0.53539 | 0.97559 | 0.95572 | 0.7831031913 | 0.97655 | 0.9840509295 | 0.321652 | 7.09186249 | 0 |
| 17 | 7676050 | A | C | Y | D | TP53 | ENST00000269305 | 0.67024 | 0.95573 | 0.59656 | 0.97375 | 0.96993 | 0.8422497017 | 0.98629 | 0.993645668 | 0.464647 | 7.128658715 | 1 |
| 17 | 7676050 | A | T | Y | N | TP53 | ENST00000269305 | 0.66304 | 0.94843 | 0.60352 | 0.97775 | 0.95535 | 0.7994742109 | 0.96764 | 0.9914435744 | 0.343621 | 6.524305941 | 0 |
| 17 | 7676051 | G | C | S | R | TP53 | ENST00000269305 | 0.14058 | 0.73642 | 0.31547 | 0.92974 | 0.862 | 0.1981145085 | 0.94853 | 0.1326787598 | 0.256664 | 4.272483396 | 0 |
| 17 | 7676051 | G | T | S | R | TP53 | ENST00000269305 | 0.13792 | 0.73642 | 0.31547 | 0.92271 | 0.862 | 0.1981145085 | 0.94904 | 0.1370231972 | 0.256665 | 4.272483396 | 0 |
| 17 | 7676052 | C | A | S | I | TP53 | ENST00000269305 | 0.07761 | 0.71902 | 0.31674 | 0.95275 | 0.75349 | 0.2057235284 | 0.87611 | 0.1071050819 | 0.118861 | 4.123882714 | 0 |
| 17 | 7676052 | C | G | S | T | TP53 | ENST00000269305 | 0.03639 | 0.5423 | 0.28308 | 0.84568 | 0.68821 | 0.1514085992 | 0.76786 | 0.1538217068 | 0.0535852 | 0.9777300949 | 0 |
| 17 | 7676052 | C | T | S | N | TP53 | ENST00000269305 | 0.03992 | 0.55043 | 0.29902 | 0.87386 | 0.66544 | 0.1100090137 | 0.63779 | 0.02056215098 | -0.00769932 | 0.8308934499 | 0 |
| 17 | 7676053 | T | A | S | C | TP53 | ENST00000269305 | 0.11625 | 0.57186 | 0.29438 | 0.97099 | 0.72398 | 0.2459542835 | 0.8759 | 0.07963645942 | 0.126926 | 3.694105496 | 0 |
| 17 | 7676053 | T | C | S | G | TP53 | ENST00000269305 | 0.0907 | 0.59201 | 0.27419 | 0.96987 | 0.68381 | 0.1381673226 | 0.82961 | 0.05751505735 | 0.084042 | 4.107730597 | 0 |
| 17 | 7676053 | T | G | S | R | TP53 | ENST00000269305 | 0.0983 | 0.73642 | 0.31547 | 0.97738 | 0.85117 | 0.1981145085 | 0.93813 | 0.09794850712 | 0.232289 | 4.272483396 | 0 |
| 17 | 7676055 | C | A | G | V | TP53 | ENST00000269305 | 0.55738 | 0.92355 | 0.67534 | 0.97597 | 0.9796 | 0.9060291875 | 0.99381 | 0.9991476536 | 0.561254 | 7.453117131 | 1 |
| 17 | 7676055 | C | T | G | D | TP53 | ENST00000269305 | 0.55937 | 0.92762 | 0.77135 | 0.97375 | 0.96664 | 0.9271601513 | 0.98968 | 0.9985570312 | 0.56114 | 6.238992566 | 0 |
| 17 | 7676056 | C | A | G | C | TP53 | ENST00000269305 | 0.53616 | 0.90472 | 0.71369 | 0.97098 | 0.98799 | 0.8748764773 | 0.99646 | 0.998955369 | 0.561254 | 7.453117131 | 1 |
| 17 | 7676056 | C | G | G | R | TP53 | ENST00000269305 | 0.5131 | 0.91823 | 0.76207 | 0.97332 | 0.97923 | 0.9096168036 | 0.9937 | 0.9987259507 | 0.561254 | 7.453117131 | 1 |
| 17 | 7676056 | C | T | G | S | TP53 | ENST00000269305 | 0.38101 | 0.89652 | 0.66144 | 0.96437 | 0.96627 | 0.8433186645 | 0.98087 | 0.9965245128 | 0.541976 | 5.185835261 | 0 |
| 17 | 7676057 | C | A | Q | H | TP53 | ENST00000269305 | 0.02152 | 0.55638 | 0.2617 | 0.85433 | 0.66161 | 0.3579379947 | 0.86129 | 0.1884273291 | 0.0634792 | 0.5673812396 | 0 |
| 17 | 7676057 | C | G | Q | H | TP53 | ENST00000269305 | 0.02327 | 0.55638 | 0.2617 | 0.85463 | 0.66161 | 0.3579379947 | 0.86536 | 0.1884273291 | 0.0635133 | 0.5673812396 | 0 |
| 17 | 7676058 | T | A | Q | L | TP53 | ENST00000269305 | 0.222 | 0.61344 | 0.25294 | 0.95612 | 0.79818 | 0.4363517384 | 0.91095 | 0.3322257996 | 0.165861 | 2.909036223 | 0 |
| 17 | 7676058 | T | C | Q | R | TP53 | ENST00000269305 | 0.26372 | 0.60858 | 0.26571 | 0.86649 | 0.71073 | 0.3230232307 | 0.83183 | 0.1099302173 | 0.0918008 | 2.666462006 | 0 |
| 17 | 7676058 | T | G | Q | P | TP53 | ENST00000269305 | 0.13966 | 0.65587 | 0.2267 | 0.73353 | 0.8199 | 0.4307184489 | 0.89846 | 0.0393085219 | 0.143643 | -0.9434672959 | 0 |
| 17 | 7676059 | G | T | Q | K | TP53 | ENST00000269305 | 0.46612 | 0.69256 | 0.27585 | 0.87081 | 0.79697 | 0.3782282093 | 0.88509 | 0.5372508764 | 0.176546 | 3.312888901 | 0 |
| 17 | 7676061 | T | A | Y | F | TP53 | ENST00000269305 | 0.49151 | 0.67022 | 0.74203 | 0.94685 | 0.92561 | 0.4582339397 | 0.97414 | 0.9358246922 | 0.308196 | 3.707745513 | 0 |
| 17 | 7676061 | T | C | Y | C | TP53 | ENST00000269305 | 0.77211 | 0.92269 | 0.81196 | 0.96345 | 0.96153 | 0.8194156878 | 0.96759 | 0.9957531691 | 0.388355 | 8.182751448 | 0 |
| 17 | 7676061 | T | G | Y | S | TP53 | ENST00000269305 | 0.73797 | 0.93579 | 0.75998 | 0.96969 | 0.97401 | 0.8392521744 | 0.98741 | 0.9973684549 | 0.477918 | 6.614558485 | 0 |
| 17 | 7676062 | A | C | Y | D | TP53 | ENST00000269305 | 0.77837 | 0.95633 | 0.80669 | 0.95443 | 0.9781 | 0.8847764877 | 0.99254 | 0.9949041307 | 0.537331 | 7.153920007 | 0 |
| 17 | 7676062 | A | G | Y | H | TP53 | ENST00000269305 | 0.73301 | 0.92644 | 0.82037 | 0.93406 | 0.96153 | 0.7249739335 | 0.95498 | 0.9792761803 | 0.397856 | 4.723602933 | 0 |
| 17 | 7676062 | A | T | Y | N | TP53 | ENST00000269305 | 0.77306 | 0.94955 | 0.81381 | 0.95644 | 0.97475 | 0.8235701335 | 0.9795 | 0.9967706203 | 0.446032 | 6.62052479 | 0 |
| 17 | 7676064 | G | A | T | I | TP53 | ENST00000269305 | 0.51028 | 0.64535 | 0.60865 | 0.90509 | 0.81006 | 0.3319697347 | 0.94441 | 0.7006106377 | 0.207177 | 5.708347634 | 0 |
| 17 | 7676064 | G | C | T | S | TP53 | ENST00000269305 | 0.23784 | 0.64573 | 0.51194 | 0.90261 | 0.79637 | 0.1721261841 | 0.93164 | 0.1551420391 | 0.200558 | 4.90335249 | 0 |
| 17 | 7676064 | G | T | T | N | TP53 | ENST00000269305 | 0.24506 | 0.84789 | 0.55975 | 0.88951 | 0.73589 | 0.1998871161 | 0.915 | 0.3289909065 | 0.158736 | 4.174235203 | 0 |
| 17 | 7676065 | T | A | T | S | TP53 | ENST00000269305 | 0.25372 | 0.64573 | 0.51194 | 0.8901 | 0.78967 | 0.1721261841 | 0.94128 | 0.1864254773 | 0.203074 | 4.90335249 | 0 |
| 17 | 7676065 | T | C | T | A | TP53 | ENST00000269305 | 0.28408 | 0.64698 | 0.49436 | 0.93326 | 0.81006 | 0.3304537641 | 0.95388 | 0.412592411 | 0.206711 | 5.329111601 | 0 |
| 17 | 7676065 | T | G | T | P | TP53 | ENST00000269305 | 0.29974 | 0.68707 | 0.51286 | 0.92513 | 0.81124 | 0.4536673959 | 0.9308 | 0.1012281254 | 0.187483 | 4.670457615 | 0 |

Variants with functional readout data from deep mutational scan experiments of *MSH2*

| chrom | pos | ref | alt | ref_aa | alt_aa | genename | transcript_id | CADD_raw_rankscore | MPC_rankscore | PrimateAI_rankscore | M-CAP_rankscore | REVEL_rankscore | gMVP_rankscore | MVP_rankscore | ClinPred | BayesDel | EVmutation | target |
|---|---|---|---|---|---|---|---|---|---|---|---|---|---|---|---|---|---|---|
| 2 | 47403195 | G | A | A | T | MSH2 | ENST00000233146 | 0.90838 | 0.03299 | 0.75513 | 0.95594 | 0.95133 | 0.6311779305 | 0.96544 | 0.4857534766 | 0.256866 | 3.13276487 | 0 |
| 2 | 47403195 | G | C | A | P | MSH2 | ENST00000233146 | 0.9145 | 0.03137 | 0.8102 | 0.95216 | 0.89101 | 0.6801755052 | 0.90074 | 0.9863920212 | 0.396182 | 7.503905917 | 0 |
| 2 | 47403195 | G | T | A | S | MSH2 | ENST00000233146 | 0.87794 | 0.02667 | 0.71884 | 0.96417 | 0.82046 | 0.5744117159 | 0.89608 | 0.9816287756 | 0.300643 | 4.218996865 | 0 |
| 2 | 47403196 | C | A | A | E | MSH2 | ENST00000233146 | 0.67645 | 0.01401 | 0.81397 | 0.95161 | 0.81646 | 0.6873533264 | 0.92159 | 0.9853185415 | 0.405006 | 7.503905917 | 0 |
| 2 | 47403196 | C | G | A | G | MSH2 | ENST00000233146 | 0.82381 | 0.03299 | 0.73668 | 0.95263 | 0.80772 | 0.5592785471 | 0.83887 | 0.9856464267 | 0.275855 | 7.503905917 | 0 |
| 2 | 47403196 | C | T | A | V | MSH2 | ENST00000233146 | 0.82934 | 0.03419 | 0.77612 | 0.94244 | 0.88448 | 0.6168499954 | 0.87138 | 0.988702774 | 0.273624 | 7.503905917 | 0 |
| 2 | 47403198 | G | A | V | M | MSH2 | ENST00000233146 | 0.76531 | 0.0218 | 0.50517 | 0.91043 | 0.77205 | 0.4128792168 | 0.83808 | 0.9618543983 | 0.231576 | 7.84558159 | 0 |
| 2 | 47403198 | G | C | V | L | MSH2 | ENST00000233146 | 0.56062 | 0.00592 | 0.45502 | 0.88561 | 0.69168 | 0.4115967754 | 0.93118 | 0.8073754311 | 0.195029 | 1.725407967 | 0 |
| 2 | 47403198 | G | T | V | L | MSH2 | ENST00000233146 | 0.55497 | 0.00592 | 0.45502 | 0.88711 | 0.69168 | 0.4115967754 | 0.92457 | 0.5099654794 | 0.195044 | 1.725407967 | 0 |
| 2 | 47403199 | T | C | V | A | MSH2 | ENST00000233146 | 0.56242 | 0.00898 | 0.48807 | 0.94027 | 0.79089 | 0.39700843 | 0.85418 | 0.6823380589 | 0.221073 | 7.84558159 | 0 |
| 2 | 47403199 | T | G | V | G | MSH2 | ENST00000233146 | 0.59726 | 0.01699 | 0.34541 | 0.9515 | 0.84106 | 0.5008347863 | 0.80239 | 0.9536565542 | 0.259281 | 7.84558159 | 0 |
| 2 | 47403201 | C | A | Q | K | MSH2 | ENST00000233146 | 0.46158 | 0.00664 | 0.58173 | 0.93847 | 0.78906 | 0.5083993808 | 0.90047 | 0.9467238784 | 0.262369 | 6.318875862 | 0 |
| 2 | 47403202 | A | C | Q | P | MSH2 | ENST00000233146 | 0.58225 | 0.00958 | 0.5361 | 0.96934 | 0.88496 | 0.5325370848 | 0.94099 | 0.9625825882 | 0.316007 | 8.080462938 | 0 |
| 2 | 47403202 | A | G | Q | R | MSH2 | ENST00000233146 | 0.45236 | 0.00996 | 0.57451 | 0.94307 | 0.85792 | 0.5050236183 | 0.96303 | 0.7128264904 | 0.336993 | 8.080462938 | 0 |
| 2 | 47403202 | A | T | Q | L | MSH2 | ENST00000233146 | 0.62068 | 0.00574 | 0.50971 | 0.97085 | 0.90274 | 0.4863100081 | 0.90723 | 0.9347090721 | 0.246138 | 6.102756817 | 0 |
| 2 | 47403203 | G | C | Q | H | MSH2 | ENST00000233146 | 0.48354 | 0.01649 | 0.56533 | 0.97707 | 0.85482 | 0.5489221508 | 0.91839 | 0.9112414122 | 0.215477 | 7.361606867 | 0 |
| 2 | 47403203 | G | T | Q | H | MSH2 | ENST00000233146 | 0.47842 | 0.01649 | 0.56533 | 0.97757 | 0.85482 | 0.5489221508 | 0.92115 | 0.9112414122 | 0.215453 | 7.361606867 | 0 |
| 2 | 47403204 | C | A | P | T | MSH2 | ENST00000233146 | 0.78019 | 0.01396 | 0.69796 | 0.9489 | 0.90929 | 0.4586275572 | 0.88578 | 0.9793134332 | 0.270277 | 7.625669846 | 0 |
| 2 | 47403204 | C | G | P | A | MSH2 | ENST00000233146 | 0.61686 | 0.00783 | 0.65772 | 0.91251 | 0.79516 | 0.3986568366 | 0.79796 | 0.9776819348 | 0.163309 | 3.36745798 | 0 |
| 2 | 47403204 | C | T | P | S | MSH2 | ENST00000233146 | 0.52261 | 0.00638 | 0.66273 | 0.91298 | 0.83725 | 0.4425350431 | 0.82149 | 0.9341100454 | 0.222721 | 2.198642748 | 0 |
| 2 | 47403205 | C | A | P | Q | MSH2 | ENST00000233146 | 0.54875 | 0.03287 | 0.67635 | 0.96818 | 0.9458 | 0.4786200421 | 0.95651 | 0.7730855346 | 0.210997 | 6.238880716 | 0 |
| 2 | 47403205 | C | G | P | R | MSH2 | ENST00000233146 | 0.53515 | 0.03124 | 0.75319 | 0.96975 | 0.93044 | 0.4693377039 | 0.89036 | 0.9952286482 | 0.318416 | 5.583744386 | 0 |
| 2 | 47403205 | C | T | P | L | MSH2 | ENST00000233146 | 0.73093 | 0.02319 | 0.75629 | 0.97258 | 0.89918 | 0.4553307716 | 0.89424 | 0.9966928959 | 0.257841 | 7.625669846 | 0 |
| 2 | 47403207 | A | C | K | Q | MSH2 | ENST00000233146 | 0.59523 | 0.00608 | 0.44276 | 0.94638 | 0.80358 | 0.4134596383 | 0.91972 | 0.8740203381 | 0.206395 | 4.190422762 | 0 |
| 2 | 47403207 | A | G | K | E | MSH2 | ENST00000233146 | 0.61298 | 0.00527 | 0.50523 | 0.9505 | 0.79637 | 0.4162148367 | 0.92023 | 0.6844915152 | 0.223929 | 3.82920829 | 0 |
| 2 | 47403208 | A | C | K | T | MSH2 | ENST00000233146 | 0.56304 | 0.0054 | 0.51588 | 0.95536 | 0.85534 | 0.4600005982 | 0.92863 | 0.835580647 | 0.233706 | 3.31391277 | 0 |
| 2 | 47403208 | A | G | K | R | MSH2 | ENST00000233146 | 0.54358 | 0.00603 | 0.42095 | 0.92701 | 0.79697 | 0.3444736825 | 0.94539 | 0.8718270063 | 0.172063 | 2.931850393 | 0 |
| 2 | 47403208 | A | T | K | M | MSH2 | ENST00000233146 | 0.75678 | 0.00754 | 0.4275 | 0.95266 | 0.84588 | 0.5314745393 | 0.91589 | 0.8560296893 | 0.19573 | 7.2908998 | 0 |
| 2 | 47403209 | G | C | K | N | MSH2 | ENST00000233146 | 0.49196 | 0.00543 | 0.54427 | 0.92252 | 0.7675 | 0.4150779965 | 0.89006 | 0.5483329296 | 0.109426 | 2.04829938 | 0 |
| 2 | 47403209 | G | T | K | N | MSH2 | ENST00000233146 | 0.48678 | 0.00543 | 0.54427 | 0.92241 | 0.76881 | 0.4150779965 | 0.89037 | 0.5645893216 | 0.109425 | 2.04829938 | 0 |
| 2 | 47403210 | G | A | E | K | MSH2 | ENST00000233146 | 0.70262 | 0.00566 | 0.58273 | 0.91627 | 0.73805 | 0.3866472414 | 0.8796 | 0.9390713573 | 0.160846 | 1.915526275 | 0 |
| 2 | 47403210 | G | C | E | Q | MSH2 | ENST00000233146 | 0.50374 | 0.00574 | 0.45413 | 0.91467 | 0.6993 | 0.3869954558 | 0.87017 | 0.6036504507 | 0.12959 | 0.05422241217 | 0 |
| 2 | 47403211 | A | C | E | A | MSH2 | ENST00000233146 | 0.48253 | 0.00562 | 0.48848 | 0.92764 | 0.70097 | 0.3835972465 | 0.93807 | 0.8414189816 | 0.168893 | 2.829006537 | 0 |
| 2 | 47403211 | A | G | E | G | MSH2 | ENST00000233146 | 0.63788 | 0.00556 | 0.43896 | 0.93829 | 0.72398 | 0.3761041099 | 0.85749 | 0.9459339976 | 0.187633 | 6.25860915 | 0 |
| 2 | 47403211 | A | T | E | V | MSH2 | ENST00000233146 | 0.62772 | 0.0058 | 0.46056 | 0.95637 | 0.81646 | 0.4590157351 | 0.9329 | 0.9406650066 | 0.234572 | 6.25860915 | 0 |
| 2 | 47403212 | G | C | E | D | MSH2 | ENST00000233146 | 0.3351 | 0.00586 | 0.47869 | 0.90219 | 0.7727 | 0.2884774615 | 0.93082 | 0.2759201755 | 0.0837454 | 3.582487193 | 0 |
| 2 | 47403212 | G | T | E | D | MSH2 | ENST00000233146 | 0.33078 | 0.00586 | 0.47869 | 0.90488 | 0.7727 | 0.2884774615 | 0.93056 | 0.2854825196 | 0.0837558 | 3.582487193 | 0 |
| 2 | 47403213 | A | C | T | P | MSH2 | ENST00000233146 | 0.56311 | 0.00562 | 0.49143 | 0.93662 | 0.81646 | 0.3403530722 | 0.93283 | 0.4541276693 | 0.198995 | 5.208999721 | 0 |
| 2 | 47403213 | A | G | T | A | MSH2 | ENST00000233146 | 0.44893 | 0.0058 | 0.48435 | 0.87159 | 0.63631 | 0.3452732012 | 0.90754 | 0.3025704317 | 0.0969959 | 1.883028011 | 0 |
| 2 | 47403213 | A | T | T | S | MSH2 | ENST00000233146 | 0.43196 | 0.00592 | 0.47416 | 0.84031 | 0.66828 | 0.2774029195 | 0.88849 | 0.3225203172 | 0.0863045 | 0.6553484321 | 0 |
| 2 | 47403214 | C | A | T | K | MSH2 | ENST00000233146 | 0.2745 | 0.00535 | 0.57363 | 0.84024 | 0.72246 | 0.4092863814 | 0.89741 | 0.3937238157 | 0.120244 | 0.3733864196 | 0 |
| 2 | 47403214 | C | G | T | R | MSH2 | ENST00000233146 | 0.3075 | 0.00574 | 0.56105 | 0.89968 | 0.79577 | 0.4146536834 | 0.92936 | 0.638328135 | 0.18764 | 2.199079244 | 0 |
| 2 | 47403214 | C | T | T | M | MSH2 | ENST00000233146 | 0.36297 | 0.00566 | 0.52503 | | 0.82836 | 0.367996496 | 0.98347 | 0.06071456036 | 0.0178849 | 5.208999721 | 0 |
| 2 | 47403216 | C | A | L | M | MSH2 | ENST00000233146 | 0.64182 | 0.00804 | 0.53664 | 0.93786 | 0.80238 | 0.3309000145 | 0.80825 | 0.8796981573 | 0.14012 | 6.049681419 | 0 |
| 2 | 47403216 | C | G | L | V | MSH2 | ENST00000233146 | 0.47658 | 0.00696 | 0.49567 | 0.95854 | 0.85945 | 0.3531889295 | 0.86881 | 0.8025496006 | 0.205985 | 4.524800162 | 0 |
| 2 | 47403217 | T | A | L | Q | MSH2 | ENST00000233146 | 0.93999 | 0.0245 | 0.56082 | 0.98687 | 0.94281 | 0.5315019574 | 0.89815 | 0.980483532 | 0.327148 | 4.161934888 | 0 |
| 2 | 47403217 | T | C | L | P | MSH2 | ENST00000233146 | 0.94208 | 0.0227 | 0.68461 | 0.99122 | 0.95753 | 0.6095097586 | 0.92934 | 0.987677753 | 0.401191 | 7.795468358 | 0 |
| 2 | 47403217 | T | G | L | R | MSH2 | ENST00000233146 | 0.94003 | 0.01924 | 0.63667 | 0.9868 | 0.94091 | 0.5595575345 | 0.91918 | 0.980483532 | 0.382098 | 4.682859896 | 0 |
| 2 | 47403219 | C | A | Q | K | MSH2 | ENST00000233146 | 0.31821 | 0.00532 | 0.47229 | 0.86848 | 0.7562 | 0.4130826012 | 0.96727 | 0.2815915383 | 0.174801 | 2.332014104 | 0 |
| 2 | 47403219 | C | G | Q | E | MSH2 | ENST00000233146 | 0.27952 | 0.00569 | 0.42256 | 0.82847 | 0.72398 | 0.3309659778 | 0.92749 | 0.182268256 | 0.10461 | 2.704145609 | 0 |
| 2 | 47403220 | A | C | Q | P | MSH2 | ENST00000233146 | 0.46649 | 0.00608 | 0.42554 | 0.91317 | 0.79395 | 0.40621416 | 0.93959 | 0.5328235626 | 0.192406 | 5.613522057 | 0 |
| 2 | 47403220 | A | G | Q | R | MSH2 | ENST00000233146 | 0.42723 | 0.0058 | 0.45692 | 0.90605 | 0.77527 | 0.3743200586 | 0.96293 | 0.4827729166 | 0.181203 | 2.924980471 | 0 |
| 2 | 47403220 | A | T | Q | L | MSH2 | ENST00000233146 | 0.48678 | 0.00599 | 0.4028 | 0.91556 | 0.76553 | 0.4239180918 | 0.93868 | 0.5205268264 | 0.134336 | 3.344822406 | 0 |
| 2 | 47403221 | G | C | Q | H | MSH2 | ENST00000233146 | 0.35328 | 0.01096 | 0.45435 | 0.93237 | 0.77909 | 0.4483550541 | 0.92857 | 0.7646977901 | 0.134449 | 5.361417298 | 0 |
| 2 | 47403221 | G | T | Q | H | MSH2 | ENST00000233146 | 0.34882 | 0.01096 | 0.45435 | 0.93237 | 0.77909 | 0.4483550541 | 0.92901 | 0.5475869179 | 0.134476 | 5.361417298 | 0 |
| 2 | 47403222 | T | A | L | M | MSH2 | ENST00000233146 | 0.37776 | 0.00615 | 0.52084 | 0.85093 | 0.6331 | 0.3138305556 | 0.75515 | 0.2640322149 | 0.0766633 | 1.451047444 | 0 |
| 2 | 47403222 | T | G | L | V | MSH2 | ENST00000233146 | 0.50838 | 0.00633 | 0.48066 | 0.94499 | 0.81414 | 0.3166539069 | 0.91401 | 0.3350179493 | 0.155975 | 3.402346678 | 0 |
| 2 | 47403223 | T | C | L | S | MSH2 | ENST00000233146 | 0.87523 | 0.01285 | 0.57351 | 0.99301 | 0.90537 | 0.4891032558 | 0.78989 | 0.9844741821 | 0.212217 | 4.048684774 | 0 |
| 2 | 47403223 | T | G | L | W | MSH2 | ENST00000233146 | 0.95028 | 0.02023 | 0.64325 | 0.99409 | 0.89694 | 0.4583315213 | 0.78941 | 0.9651252627 | 0.214485 | 5.133390082 | 0 |
| 2 | 47403224 | G | C | L | F | MSH2 | ENST00000233146 | 0.7395 | 0.0058 | 0.54397 | 0.9508 | 0.81876 | 0.3095182526 | 0.76886 | 0.972674787 | 0.189505 | 7.426261263 | 0 |
| 2 | 47403224 | G | T | L | F | MSH2 | ENST00000233146 | 0.73301 | 0.0058 | 0.54397 | 0.9511 | 0.81876 | 0.3095182526 | 0.76645 | 0.972674787 | 0.189516 | 7.426261263 | 0 |
| 2 | 47403225 | G | A | E | K | MSH2 | ENST00000233146 | 0.68691 | 0.00608 | 0.68158 | 0.92614 | 0.77463 | 0.4867117229 | 0.89116 | 0.7990281582 | 0.204157 | 1.677454098 | 0 |
| 2 | 47403225 | G | C | E | Q | MSH2 | ENST00000233146 | 0.63367 | 0.0058 | 0.57235 | 0.92102 | 0.83505 | 0.5041213126 | 0.93317 | 0.9460274577 | 0.22351 | 2.118276022 | 0 |
| 2 | 47403226 | A | C | E | A | MSH2 | ENST00000233146 | 0.64617 | 0.00622 | 0.58413 | 0.95349 | 0.81124 | 0.5207026709 | 0.90642 | 0.9770415425 | 0.167556 | 3.164146314 | 0 |
| 2 | 47403226 | A | G | E | G | MSH2 | ENST00000233146 | 0.69163 | 0.00603 | 0.54666 | 0.94834 | 0.7562 | 0.480375009 | 0.88121 | 0.9757660627 | 0.132763 | 4.591826621 | 0 |
| 2 | 47403226 | A | T | E | V | MSH2 | ENST00000233146 | 0.67533 | 0.00804 | 0.56878 | 0.97935 | 0.88259 | 0.5654685444 | 0.96047 | 0.9820929766 | 0.250917 | 6.305428003 | 0 |
| 2 | 47403227 | G | C | E | D | MSH2 | ENST00000233146 | 0.26008 | 0.00586 | 0.58596 | 0.79129 | 0.69254 | 0.3323431087 | 0.84629 | 0.1720790714 | 0.0510846 | -1.133147578 | 0 |
| 2 | 47403227 | G | T | E | D | MSH2 | ENST00000233146 | 0.25638 | 0.00586 | 0.58596 | 0.79838 | 0.69254 | 0.3323431087 | 0.84589 | 0.1720790714 | 0.0511187 | -1.133147578 | 0 |
| 2 | 47403228 | A | C | S | R | MSH2 | ENST00000233146 | 0.40226 | 0.00551 | 0.5424 | 0.88966 | 0.84375 | 0.4892771978 | 0.97544 | 0.1712723225 | 0.213682 | 3.669056113 | 0 |
| 2 | 47403228 | A | G | S | G | MSH2 | ENST00000233146 | 0.37981 | 0.00586 | 0.44429 | 0.82725 | 0.7285 | 0.3667564832 | 0.90296 | 0.1338950396 | 0.0505576 | 1.055403878 | 0 |
| 2 | 47403228 | A | T | S | C | MSH2 | ENST00000233146 | 0.46886 | 0.00935 | 0.49346 | 0.90886 | 0.88916 | 0.4250458155 | 0.95536 | 0.6844548583 | 0.217601 | 4.863981947 | 0 |
| 2 | 47403229 | G | A | S | N | MSH2 | ENST00000233146 | 0.48653 | 0.0058 | 0.52757 | 0.86462 | 0.62547 | 0.3784733338 | 0.87874 | 0.5180840492 | -0.0194579 | 2.643167183 | 0 |
| 2 | 47403229 | G | C | S | T | MSH2 | ENST00000233146 | 0.47394 | 0.00586 | 0.47715 | 0.84719 | 0.65869 | 0.3908996542 | 0.92828 | 0.5851323009 | 0.118473 | 3.705663071 | 0 |
| 2 | 47403229 | G | T | S | I | MSH2 | ENST00000233146 | 0.56006 | 0.00543 | 0.52528 | 0.84186 | 0.81761 | 0.5022461246 | 0.98291 | 0.5953429937 | 0.361673 | 3.262728253 | 0 |
| 2 | 47403230 | C | A | S | R | MSH2 | ENST00000233146 | 0.40716 | 0.00551 | 0.5424 | 0.90514 | 0.84535 | 0.4892771978 | 0.96597 | 0.5887029171 | 0.235543 | 3.669056113 | 0 |
| 2 | 47403230 | C | G | S | R | MSH2 | ENST00000233146 | 0.41345 | 0.00551 | 0.5424 | 0.90445 | 0.84535 | 0.4892771978 | 0.96847 | 0.5443150997 | 0.235524 | 3.669056113 | 0 |
| 2 | 47403231 | G | A | A | T | MSH2 | ENST00000233146 | 0.50955 | 0.0058 | 0.37407 | 0.83743 | 0.61068 | 0.367114475 | 0.84488 | 0.2584283508 | 0.103911 | 2.048672196 | 0 |
| 2 | 47403231 | G | C | A | P | MSH2 | ENST00000233146 | 0.52699 | 0.00527 | 0.4381 | 0.86648 | 0.75755 | 0.4648686727 | 0.94308 | 0.4493049804 | 0.180682 | 2.038479578 | 0 |
| 2 | 47403231 | G | T | A | S | MSH2 | ENST00000233146 | 0.47766 | 0.00622 | 0.34487 | 0.85145 | 0.64157 | 0.3119981267 | 0.85193 | 0.5077838302 | 0.103827 | 1.366580933 | 0 |
| 2 | 47403232 | C | A | A | E | MSH2 | ENST00000233146 | 0.34038 | 0.00596 | 0.43743 | 0.81856 | 0.80948 | 0.4554247176 | 0.92112 | 0.2301711238 | 0.127327 | 2.675921277 | 0 |
| 2 | 47403232 | C | G | A | G | MSH2 | ENST00000233146 | 0.36036 | 0.00569 | 0.35891 | 0.87607 | 0.66161 | 0.2947081798 | 0.8788 | 0.2848976851 | 0.0897157 | 1.99659677 | 0 |
| 2 | 47403232 | C | T | A | V | MSH2 | ENST00000233146 | 0.35256 | 0.00569 | 0.40624 | 0.83693 | 0.65869 | 0.3505992727 | 0.87863 | 0.3463382125 | 0.0812698 | 1.919166245 | 0 |
| 2 | 47403234 | G | A | A | T | MSH2 | ENST00000233146 | 0.55511 | 0.00586 | 0.60963 | 0.90437 | 0.75005 | 0.4635465707 | 0.90915 | 0.7788310647 | 0.21773 | 3.677007615 | 0 |
| 2 | 47403234 | G | C | A | P | MSH2 | ENST00000233146 | 0.63084 | 0.00799 | 0.673 | 0.90923 | 0.88637 | 0.5789764557 | 0.91026 | 0.962495029 | 0.333093 | 5.691370807 | 0 |
| 2 | 47403234 | G | T | A | S | MSH2 | ENST00000233146 | 0.50661 | 0.00586 | 0.5718 | 0.88393 | 0.672 | 0.3954719682 | 0.84662 | 0.7165377736 | 0.122175 | 2.212704701 | 0 |
| 2 | 47403235 | C | A | A | D | MSH2 | ENST00000233146 | 0.36682 | 0.0054 | 0.71255 | 0.905 | 0.81472 | 0.5564800112 | 0.82499 | 0.8414033651 | 0.208952 | 3.266069873 | 0 |
| 2 | 47403235 | C | G | A | G | MSH2 | ENST00000233146 | 0.37998 | 0.00569 | 0.59279 | 0.90578 | 0.79697 | 0.3854862193 | 0.85444 | 0.7379422784 | 0.215305 | 5.194197047 | 0 |
| 2 | 47403235 | C | T | A | V | MSH2 | ENST00000233146 | 0.38329 | 0.0058 | 0.63586 | 0.9003 | 0.81124 | 0.4590380442 | 0.89785 | 0.9445663691 | 0.24681 | 6.048657574 | 0 |
| 2 | 47403237 | G | A | E | K | MSH2 | ENST00000233146 | 0.94001 | 0.02589 | 0.76278 | 0.96048 | 0.96627 | 0.5547959514 | 0.97962 | 0.9983715415 | 0.481131 | 6.10318734 | 0 |
| 2 | 47403237 | G | C | E | Q | MSH2 | ENST00000233146 | 0.6138 | 0.0176 | 0.64472 | 0.94634 | 0.91058 | 0.511437395 | 0.94448 | 0.9853634834 | 0.379246 | 4.731505634 | 0 |
| 2 | 47403238 | A | C | E | A | MSH2 | ENST00000233146 | 0.91574 | 0.03231 | 0.66948 | 0.98066 | 0.94281 | 0.5644965711 | 0.93192 | 0.9978625178 | 0.340544 | 5.971252978 | 0 |
| 2 | 47403238 | A | G | E | G | MSH2 | ENST00000233146 | 0.94075 | 0.03099 | 0.61413 | 0.98587 | 0.96883 | 0.5331769559 | 0.95329 | 0.998536706 | 0.49328 | 6.586528125 | 0 |
| 2 | 47403238 | A | T | E | V | MSH2 | ENST00000233146 | 0.93633 | 0.03287 | 0.64618 | 0.98957 | 0.97141 | 0.5999840449 | 0.96997 | 0.9988955259 | 0.483475 | 4.414650581 | 0 |
| 2 | 47403240 | G | A | V | I | MSH2 | ENST00000233146 | 0.18183 | 0.00608 | 0.30484 | 0.79295 | 0.58771 | 0.1763838575 | 0.76314 | 0.06518859858 | -0.000706293 | 0.9477816311 | 0 |
| 2 | 47403240 | G | C | V | L | MSH2 | ENST00000233146 | 0.17714 | 0.00574 | 0.34502 | 0.79918 | 0.61416 | 0.2431943479 | 0.78057 | 0.0464336036 | -0.00606193 | -1.814225345 | 0 |
| 2 | 47403240 | G | T | V | F | MSH2 | ENST00000233146 | 0.18745 | 0.00527 | 0.35334 | 0.82101 | 0.65967 | 0.3136010338 | 0.74506 | 0.07444553762 | 0.0222708 | 0.8212463932 | 0 |
| 2 | 47403241 | T | A | V | D | MSH2 | ENST00000233146 | 0.24638 | 0.00535 | 0.38118 | 0.88967 | 0.75822 | 0.3438590399 | 0.86988 | 0.2066265047 | 0.0308796 | 0.09083793486 | 0 |
| 2 | 47403241 | T | C | V | A | MSH2 | ENST00000233146 | 0.19581 | 0.00535 | 0.36304 | 0.85476 | 0.69846 | 0.2269321195 | 0.58875 | 0.06419224292 | -0.0613615 | -1.395337 | 0 |
| 2 | 47403241 | T | G | V | G | MSH2 | ENST00000233146 | 0.22698 | 0.00527 | 0.3068 | 0.8875 | 0.73949 | 0.3100169155 | 0.81297 | 0.04707598086 | 0.0201732 | -0.4236786095 | 0 |

| | | | | | | | | | | | | | | | | | | |
|---|---|---|---|---|---|---|---|---|---|---|---|---|---|---|---|---|---|---|
| 2 | 47403243 | G | A | G | S | MSH2 | ENST00000233146 | 0.67517 | 0.01133 | 0.52485 | 0.8918 | 0.85012 | 0.5352095881 | 0.80688 | 0.9858420491 | 0.146212 | 5.11048577 | 0 |
| 2 | 47403243 | G | C | G | R | MSH2 | ENST00000233146 | 0.94974 | 0.03243 | 0.64851 | 0.95705 | 0.9648 | 0.6476323337 | 0.92941 | 0.9982151985 | 0.378669 | 6.952079475 | 0 |
| 2 | 47403243 | G | T | G | C | MSH2 | ENST00000233146 | 0.95359 | 0.03576 | 0.60857 | 0.96359 | 0.96371 | 0.5936291669 | 0.91336 | 0.9978321195 | 0.393553 | 6.952079475 | 0 |
| 2 | 47403244 | G | A | G | D | MSH2 | ENST00000233146 | 0.69196 | 0.02708 | 0.67566 | 0.94884 | 0.93241 | 0.5998252235 | 0.86804 | 0.9971061349 | 0.345829 | 6.952079475 | 0 |
| 2 | 47403244 | G | C | G | A | MSH2 | ENST00000233146 | 0.38645 | 0.00754 | 0.54712 | 0.90608 | 0.88354 | 0.4807800288 | 0.80014 | 0.9925487041 | 0.213718 | 5.602981772 | 0 |
| 2 | 47403244 | G | T | G | V | MSH2 | ENST00000233146 | 0.69196 | 0.02755 | 0.55564 | 0.96915 | 0.94765 | 0.5737925288 | 0.8601 | 0.9973828197 | 0.378437 | 6.952079475 | 0 |
| 2 | 47403246 | T | A | F | I | MSH2 | ENST00000233146 | 0.9333 | 0.03224 | 0.83192 | 0.98235 | 0.98657 | 0.6674584369 | 0.93182 | 0.9996098876 | 0.452745 | 6.780751947 | 0 |
| 2 | 47403246 | T | C | F | L | MSH2 | ENST00000233146 | 0.93908 | 0.03491 | 0.85842 | 0.9781 | 0.98294 | 0.5918764172 | 0.92405 | 0.2967583119 | 0.10786 | 3.000025341 | 0 |
| 2 | 47403246 | T | G | F | V | MSH2 | ENST00000233146 | 0.93562 | 0.03313 | 0.81298 | 0.98537 | 0.98938 | 0.6247905529 | 0.95819 | 0.999506712 | 0.486256 | 5.794164769 | 0 |
| 2 | 47403247 | T | A | F | Y | MSH2 | ENST00000233146 | 0.91785 | 0.03527 | 0.84715 | 0.97592 | 0.95353 | 0.5035445541 | 0.88146 | 0.9977353811 | 0.308037 | 7.415397436 | 0 |
| 2 | 47403247 | T | C | F | S | MSH2 | ENST00000233146 | 0.94775 | 0.02864 | 0.83042 | 0.99092 | 0.99234 | 0.7363748765 | 0.96584 | 0.9995535016 | 0.488299 | 7.415397436 | 0 |
| 2 | 47403247 | T | G | F | C | MSH2 | ENST00000233146 | 0.94764 | 0.03175 | 0.86087 | 0.99018 | 0.99202 | 0.6955253062 | 0.95691 | 0.9992172718 | 0.491385 | 7.415397436 | 0 |
| 2 | 47403248 | C | A | F | L | MSH2 | ENST00000233146 | 0.72372 | 0.03491 | 0.85842 | 0.978 | 0.9328 | 0.5918764172 | 0.93498 | 0.9985200763 | 0.375816 | 3.000025341 | 0 |
| 2 | 47403248 | C | G | F | L | MSH2 | ENST00000233146 | 0.73221 | 0.03491 | 0.85842 | 0.97845 | 0.93319 | 0.5918764172 | 0.93481 | 0.9985200763 | 0.375817 | 3.000025341 | 0 |
| 2 | 47403249 | G | C | V | L | MSH2 | ENST00000233146 | 0.4857 | 0.00574 | 0.71211 | 0.84979 | 0.61988 | 0.2746968424 | 0.86371 | 0.5492821336 | 0.0866972 | 0.3318608271 | 0 |
| 2 | 47403249 | G | T | V | L | MSH2 | ENST00000233146 | 0.48056 | 0.00574 | 0.71211 | 0.84763 | 0.61988 | 0.2746968424 | 0.86336 | 0.584883213 | 0.0866992 | 0.3318608271 | 0 |
| 2 | 47403250 | T | A | V | E | MSH2 | ENST00000233146 | 0.68379 | 0.00705 | 0.72092 | 0.98291 | 0.95863 | 0.6061664131 | 0.92109 | 0.9903214574 | 0.301894 | 5.895093337 | 0 |
| 2 | 47403250 | T | C | V | A | MSH2 | ENST00000233146 | 0.50498 | 0.00615 | 0.74284 | 0.94596 | 0.90972 | 0.3864406621 | 0.87812 | 0.9620699286 | 0.241988 | 4.80670498 | 0 |
| 2 | 47403250 | T | G | V | G | MSH2 | ENST00000233146 | 0.81967 | 0.00812 | 0.6089 | 0.98114 | 0.9648 | 0.543347939 | 0.89958 | 0.9905564785 | 0.343758 | 5.895093337 | 0 |
| 2 | 47403252 | C | A | R | S | MSH2 | ENST00000233146 | 0.53408 | 0.00556 | 0.58758 | 0.91184 | 0.68203 | 0.4104656226 | 0.85251 | 0.7888861299 | 0.102627 | 0.7128464864 | 0 |
| 2 | 47403252 | C | G | R | G | MSH2 | ENST00000233146 | 0.63912 | 0.00747 | 0.59319 | 0.95105 | 0.90362 | 0.399562833 | 0.95278 | 0.9735046625 | 0.263868 | 6.114026091 | 0 |
| 2 | 47403252 | C | T | R | C | MSH2 | ENST00000233146 | 0.94916 | 0.02143 | 0.70573 | 0.97837 | 0.90494 | 0.4629243679 | 0.93595 | 0.9988891482 | 0.29478 | 5.773201038 | 0 |
| 2 | 47403253 | G | A | R | H | MSH2 | ENST00000233146 | 0.78939 | 0.01538 | 0.61511 | 0.98348 | 0.9045 | 0.3750049472 | 0.88047 | 0.9840445518 | 0.162882 | 4.645499669 | 0 |
| 2 | 47403253 | G | C | R | P | MSH2 | ENST00000233146 | 0.62886 | 0.00522 | 0.65583 | 0.97569 | 0.94394 | 0.6921393255 | 0.98056 | 0.9849165082 | 0.398549 | 6.114026091 | 0 |
| 2 | 47403253 | G | T | R | L | MSH2 | ENST00000233146 | 0.64579 | 0.01002 | 0.64368 | 0.98348 | 0.93475 | 0.474336369 | 0.93215 | 0.977811873 | 0.247943 | 3.613844335 | 0 |
| 2 | 47403255 | T | A | F | I | MSH2 | ENST00000233146 | 0.77972 | 0.0058 | 0.82052 | 0.96651 | 0.94091 | 0.6169813022 | 0.90557 | 0.9942128062 | 0.316726 | 6.414295166 | 0 |
| 2 | 47403255 | T | C | F | L | MSH2 | ENST00000233146 | 0.92847 | 0.00777 | 0.85006 | 0.97062 | 0.95645 | 0.5288306384 | 0.92882 | 0.9942128062 | 0.40289 | 4.112324015 | 0 |
| 2 | 47403255 | T | G | F | V | MSH2 | ENST00000233146 | 0.92386 | 0.00705 | 0.8001 | 0.97504 | 0.9659 | 0.5622185803 | 0.95802 | 0.9964147806 | 0.432864 | 5.533513012 | 0 |
| 2 | 47403256 | T | A | F | Y | MSH2 | ENST00000233146 | 0.54684 | 0.00562 | 0.84351 | 0.93085 | 0.90007 | 0.4261184827 | 0.77353 | 0.9759514332 | 0.194331 | 2.966221077 | 0 |
| 2 | 47403256 | T | C | F | S | MSH2 | ENST00000233146 | 0.91499 | 0.01369 | 0.82404 | 0.98584 | 0.9681 | 0.6531325124 | 0.96492 | 0.9921414852 | 0.43687 | 6.40007346 | 0 |
| 2 | 47403256 | T | G | F | C | MSH2 | ENST00000233146 | 0.91991 | 0.00574 | 0.84982 | 0.98012 | 0.97178 | 0.616557719 | 0.95863 | 0.9955926538 | 0.453463 | 5.351371836 | 0 |
| 2 | 47403257 | C | A | F | L | MSH2 | ENST00000233146 | 0.66399 | 0.00777 | 0.85006 | 0.93157 | 0.92233 | 0.5288306384 | 0.88338 | 0.9955854416 | 0.381912 | 4.112324015 | 0 |
| 2 | 47403257 | C | G | F | L | MSH2 | ENST00000233146 | 0.67239 | 0.00777 | 0.85006 | 0.92021 | 0.92233 | 0.5288306384 | 0.88353 | 0.9955854416 | 0.38191 | 4.112324015 | 0 |
| 2 | 47403258 | T | A | F | I | MSH2 | ENST00000233146 | 0.42717 | 0.00527 | 0.73616 | 0.96655 | 0.825 | 0.4283401224 | 0.84012 | 0.8926301599 | 0.117765 | 2.989114232 | 0 |
| 2 | 47403258 | T | C | F | L | MSH2 | ENST00000233146 | 0.42933 | 0.00543 | 0.77696 | 0.96942 | 0.77141 | 0.3726105637 | 0.85071 | 0.05335165013 | -0.215385 | 1.028388617 | 0 |
| 2 | 47403258 | T | G | F | V | MSH2 | ENST00000233146 | 0.42532 | 0.00527 | 0.71853 | 0.97326 | 0.75687 | 0.4255809029 | 0.77152 | 0.7869489193 | 0.0882921 | 1.632783293 | 0 |
| 2 | 47403259 | T | A | F | Y | MSH2 | ENST00000233146 | 0.40523 | 0.00551 | 0.76857 | 0.82904 | 0.52745 | 0.3651202226 | 0.74347 | 0.2313827425 | 0.0333121 | -0.0422761736 | 0 |
| 2 | 47403259 | T | C | F | S | MSH2 | ENST00000233146 | 0.81229 | 0.00773 | 0.7325 | 0.98635 | 0.91986 | 0.608892472 | 0.91059 | 0.9738422632 | 0.230945 | 4.577255464 | 0 |
| 2 | 47403259 | T | G | F | C | MSH2 | ENST00000233146 | 0.64035 | 0.00527 | 0.76852 | 0.97484 | 0.90668 | 0.5447052259 | 0.8992 | 0.9676846862 | 0.212594 | 4.577255464 | 0 |
| 2 | 47403260 | T | A | F | L | MSH2 | ENST00000233146 | 0.36325 | 0.00543 | 0.77696 | 0.94528 | 0.74164 | 0.3726105637 | 0.85586 | 0.5547229648 | 0.0698443 | 1.028388617 | 0 |
| 2 | 47403260 | T | G | F | L | MSH2 | ENST00000233146 | 0.36901 | 0.00543 | 0.77696 | 0.94493 | 0.74164 | 0.3726105637 | 0.85639 | 0.3135632277 | 0.0698401 | 1.028388617 | 0 |
| 2 | 47403261 | C | A | Q | K | MSH2 | ENST00000233146 | 0.24616 | 0.00527 | 0.59475 | 0.87592 | 0.67386 | 0.3048991309 | 0.82408 | 0.1053873331 | 0.0488666 | 0.5476251584 | 0 |
| 2 | 47403261 | C | G | Q | E | MSH2 | ENST00000233146 | 0.23516 | 0.00556 | 0.53915 | 0.89854 | 0.56291 | 0.2813392458 | 0.70289 | 0.1900562942 | 0.0412728 | 2.910787437 | 0 |
| 2 | 47403262 | A | C | Q | P | MSH2 | ENST00000233146 | 0.48921 | 0.00622 | 0.55791 | 0.9641 | 0.8873 | 0.6194524868 | 0.93375 | 0.5271365593 | 0.187072 | 4.663996526 | 0 |
| 2 | 47403262 | A | G | Q | R | MSH2 | ENST00000233146 | 0.41652 | 0.00556 | 0.57918 | 0.9241 | 0.65673 | 0.2611822521 | 0.86259 | 0.3006833221 | 0.0654013 | -0.4349325388 | 0 |
| 2 | 47403262 | A | T | Q | L | MSH2 | ENST00000233146 | 0.48525 | 0.00574 | 0.53235 | 0.92465 | 0.62547 | 0.3460422171 | 0.77288 | 0.4639486969 | 0.0486306 | -0.8692614151 | 0 |
| 2 | 47403263 | G | C | Q | H | MSH2 | ENST00000233146 | 0.37645 | 0.00916 | 0.57805 | 0.95688 | 0.72775 | 0.3771923659 | 0.82737 | 0.4032074213 | 0.0715408 | 1.121654223 | 0 |
| 2 | 47403263 | G | T | Q | H | MSH2 | ENST00000233146 | 0.37188 | 0.00916 | 0.57805 | 0.95627 | 0.72775 | 0.3771923659 | 0.82851 | 0.4032074213 | 0.0715527 | 1.121654223 | 0 |
| 2 | 47403264 | G | A | G | S | MSH2 | ENST00000233146 | 0.20857 | 0.00569 | 0.4553 | 0.83339 | 0.66161 | 0.2560976144 | 0.7328 | 0.1109719803 | 0.029032 | -1.736680693 | 0 |
| 2 | 47403264 | G | C | G | R | MSH2 | ENST00000233146 | 0.30591 | 0.00543 | 0.57346 | 0.90836 | 0.83228 | 0.3765318382 | 0.89832 | 0.1774146024 | 0.104671 | 1.266561796 | 0 |
| 2 | 47403264 | G | T | G | C | MSH2 | ENST00000233146 | 0.57801 | 0.01346 | 0.54275 | 0.96656 | 0.84535 | 0.3699561417 | 0.8835 | 0.9147737622 | 0.122597 | 5.365707251 | 0 |
| 2 | 47403265 | G | A | G | D | MSH2 | ENST00000233146 | 0.42764 | 0.00527 | 0.59527 | 0.89248 | 0.75349 | 0.3396952712 | 0.7887 | 0.3227879405 | 0.0840865 | 0.6627994211 | 0 |
| 2 | 47403265 | G | C | G | A | MSH2 | ENST00000233146 | 0.39278 | 0.00556 | 0.50497 | 0.84067 | 0.54628 | 0.2335447912 | 0.80286 | 0.2755623162 | 0.0413347 | -0.2617567341 | 0 |
| 2 | 47403265 | G | T | G | V | MSH2 | ENST00000233146 | 0.54183 | 0.00574 | 0.51931 | 0.92534 | 0.79334 | 0.3633818086 | 0.8807 | 0.7763541341 | 0.111063 | 4.682435073 | 0 |
| 2 | 47403267 | A | C | M | L | MSH2 | ENST00000233146 | 0.46258 | 0.0058 | 0.72911 | 0.84596 | 0.65673 | 0.4235351193 | 0.66846 | 0.2796547718 | 0.0640695 | 0.6995204463 | 0 |
| 2 | 47403267 | A | G | M | V | MSH2 | ENST00000233146 | 0.67891 | 0.00769 | 0.66277 | 0.97312 | 0.89376 | 0.5073803931 | 0.89701 | 0.9560918212 | 0.224131 | 3.357992527 | 0 |
| 2 | 47403267 | A | T | M | L | MSH2 | ENST00000233146 | 0.4574 | 0.0058 | 0.72911 | 0.84656 | 0.65673 | 0.4235351193 | 0.66901 | 0.3622895982 | 0.0640737 | 0.6995204463 | 0 |
| 2 | 47403268 | T | A | M | K | MSH2 | ENST00000233146 | 0.84928 | 0.01281 | 0.80533 | 0.98371 | 0.98183 | 0.8183082763 | 0.95135 | 0.9986300468 | 0.445919 | 7.522289562 | 0 |
| 2 | 47403268 | T | C | M | T | MSH2 | ENST00000233146 | 0.78162 | 0.00945 | 0.83892 | 0.97514 | 0.96008 | 0.6690474628 | 0.86863 | 0.9938620329 | 0.288172 | 3.70858333 | 0 |
| 2 | 47403268 | T | G | M | R | MSH2 | ENST00000233146 | 0.87221 | 0.01591 | 0.78609 | 0.98133 | 0.98183 | 0.8406634163 | 0.9374 | 0.998463273 | 0.430371 | 7.522289562 | 0 |
| 2 | 47403270 | C | A | P | T | MSH2 | ENST00000233146 | 0.76076 | 0.02414 | 0.73717 | 0.97258 | 0.91271 | 0.478515217 | 0.84192 | 0.9978524446 | 0.253923 | 6.645768384 | 0 |
| 2 | 47403270 | C | G | P | A | MSH2 | ENST00000233146 | 0.71482 | 0.00812 | 0.70365 | 0.96076 | 0.86603 | 0.3860970611 | 0.76603 | 0.9927234054 | 0.145716 | 6.569247847 | 0 |
| 2 | 47403270 | C | T | P | S | MSH2 | ENST00000233146 | 0.49538 | 0.00816 | 0.69917 | 0.92713 | 0.84428 | 0.47610877 | 0.76659 | 0.9619117379 | 0.133224 | 3.475847639 | 0 |
| 2 | 47403271 | C | A | P | Q | MSH2 | ENST00000233146 | 0.68979 | 0.03398 | 0.71441 | 0.96508 | 0.8873 | 0.520512245 | 0.84199 | 0.9976086617 | 0.268977 | 4.841089978 | 0 |
| 2 | 47403271 | C | G | P | R | MSH2 | ENST00000233146 | 0.52267 | 0.00978 | 0.77209 | 0.93538 | 0.91736 | 0.4796765008 | 0.81658 | 0.9475505333 | 0.293036 | 4.464943154 | 0 |
| 2 | 47403271 | C | T | P | L | MSH2 | ENST00000233146 | 0.70615 | 0.02835 | 0.77795 | 0.96315 | 0.88543 | 0.5215129586 | 0.86409 | 0.9529110193 | 0.166663 | 6.487764913 | 0 |
| 2 | 47403273 | G | C | E | Q | MSH2 | ENST00000233146 | 0.63735 | 0.03205 | 0.64669 | 0.88765 | 0.79878 | 0.4309865847 | 0.88508 | 0.9707192183 | 0.198179 | 3.372202857 | 0 |
| 2 | 47403274 | A | C | E | A | MSH2 | ENST00000233146 | 0.44168 | 0.00696 | 0.6632 | 0.94414 | 0.73734 | 0.4736466468 | 0.8451 | 0.7064627409 | 0.112537 | 1.924248331 | 0 |
| 2 | 47403274 | A | G | E | G | MSH2 | ENST00000233146 | 0.60222 | 0.00763 | 0.61894 | 0.95717 | 0.86302 | 0.4528469452 | 0.78229 | 0.9938440323 | 0.181612 | 4.777486738 | 0 |
| 2 | 47403274 | A | T | E | V | MSH2 | ENST00000233146 | 0.76291 | 0.02646 | 0.65034 | 0.97911 | 0.90972 | 0.5421014951 | 0.88092 | 0.995906353 | 0.228432 | 6.305267152 | 0 |
| 2 | 47403275 | G | C | E | D | MSH2 | ENST00000233146 | 0.48417 | 0.00681 | 0.66953 | 0.92709 | 0.79818 | 0.3399984957 | 0.92643 | 0.2790341675 | 0.102316 | 2.615843253 | 0 |
| 2 | 47403275 | G | T | E | D | MSH2 | ENST00000233146 | 0.47905 | 0.00681 | 0.66953 | 0.92854 | 0.79818 | 0.3399984957 | 0.92678 | 0.2790341675 | 0.102321 | 2.615843253 | 0 |
| 2 | 47403276 | A | C | K | Q | MSH2 | ENST00000233146 | 0.90969 | 0.03189 | 0.78002 | 0.9608 | 0.93631 | 0.6879136561 | 0.93685 | 0.9981355667 | 0.282301 | 4.516102189 | 0 |
| 2 | 47403276 | A | G | K | E | MSH2 | ENST00000233146 | 0.91979 | 0.02679 | 0.81432 | 0.97651 | 0.92233 | 0.730485253 | 0.89478 | 0.9963368177 | 0.291255 | 4.437742793 | 0 |
| 2 | 47403277 | A | C | K | T | MSH2 | ENST00000233146 | 0.88616 | 0.03231 | 0.82047 | 0.97471 | 0.9579 | 0.7093897358 | 0.87795 | 0.9969190359 | 0.300874 | 7.670110308 | 0 |
| 2 | 47403277 | A | G | K | R | MSH2 | ENST00000233146 | 0.73061 | 0.01158 | 0.75759 | 0.94482 | 0.90096 | 0.5838129394 | 0.92753 | 0.9889118671 | 0.260433 | 5.590433848 | 0 |
| 2 | 47403277 | A | T | K | M | MSH2 | ENST00000233146 | 0.93354 | 0.03205 | 0.75684 | 0.98467 | 0.96407 | 0.7578840067 | 0.91309 | 0.9991612434 | 0.381827 | 8.576547329 | 0 |
| 2 | 47403278 | G | C | K | N | MSH2 | ENST00000233146 | 0.76148 | 0.0318 | 0.83886 | 0.98205 | 0.88448 | 0.6974725855 | 0.93649 | 0.9966926575 | 0.132917 | 5.320812541 | 0 |
| 2 | 47403278 | G | T | K | N | MSH2 | ENST00000233146 | 0.75495 | 0.0318 | 0.83886 | 0.98221 | 0.88448 | 0.6974725855 | 0.93643 | 0.9966926575 | 0.132913 | 5.320812541 | 0 |
| 2 | 47403279 | C | A | P | T | MSH2 | ENST00000233146 | 0.7426 | 0.0315 | 0.7539 | 0.91856 | 0.90274 | 0.519291429 | 0.83602 | 0.9955093861 | 0.238342 | 7.316608073 | 0 |
| 2 | 47403279 | C | G | P | A | MSH2 | ENST00000233146 | 0.66834 | 0.01305 | 0.70634 | 0.88389 | 0.84267 | 0.4318402374 | 0.76597 | 0.8149137497 | 0.144439 | 3.474273287 | 0 |
| 2 | 47403279 | C | T | P | S | MSH2 | ENST00000233146 | 0.70623 | 0.01855 | 0.71954 | 0.87669 | 0.83172 | 0.5075434009 | 0.79367 | 0.9689295292 | 0.14718 | 4.322566184 | 0 |
| 2 | 47403280 | C | G | P | R | MSH2 | ENST00000233146 | 0.60934 | 0.03547 | 0.78611 | 0.91644 | 0.93004 | 0.5569840999 | 0.89454 | 0.9949421287 | 0.381763 | 7.316608073 | 0 |
| 2 | 47403280 | C | T | P | L | MSH2 | ENST00000233146 | 0.43847 | 0.00832 | 0.79636 | 0.89771 | 0.92151 | 0.5919211593 | 0.91192 | 0.974999845 | 0.260939 | 3.838360138 | 0 |
| 2 | 47403282 | A | C | T | P | MSH2 | ENST00000233146 | 0.42193 | 0.00608 | 0.5853 | 0.92217 | 0.78348 | 0.4800117153 | 0.87578 | 0.5925393701 | 0.0944782 | -0.8356582788 | 0 |
| 2 | 47403282 | A | G | T | A | MSH2 | ENST00000233146 | 0.30771 | 0.00574 | 0.56222 | 0.85766 | 0.71548 | 0.2753931059 | 0.80706 | 0.5911263824 | 0.0330273 | -0.2405022087 | 0 |
| 2 | 47403282 | A | T | T | S | MSH2 | ENST00000233146 | 0.28636 | 0.00586 | 0.56835 | 0.8529 | 0.70179 | 0.2276474559 | 0.82977 | 0.2032096893 | 0.0531988 | -1.385013249 | 0 |
| 2 | 47403283 | C | A | T | N | MSH2 | ENST00000233146 | 0.36459 | 0.00547 | 0.61719 | 0.86654 | 0.60835 | 0.307261483 | 0.79356 | 0.10476771 | 0.0526433 | -0.3062249202 | 0 |
| 2 | 47403283 | C | G | T | S | MSH2 | ENST00000233146 | 0.30889 | 0.00586 | 0.56835 | 0.81869 | 0.65575 | 0.2276474559 | 0.74841 | 0.07746808738 | 0.0387331 | -1.385013249 | 0 |
| 2 | 47403283 | C | T | T | I | MSH2 | ENST00000233146 | 0.43926 | 0.00556 | 0.66315 | 0.89854 | 0.79697 | 0.312697709 | 0.89212 | 0.154634852 | 0.10844 | 3.56572025 | 0 |
| 2 | 47403285 | A | C | T | P | MSH2 | ENST00000233146 | 0.94492 | 0.02979 | 0.77811 | 0.975 | 0.9248 | 0.713780878 | 0.9103 | 0.9525856972 | 0.30994 | 7.346359886 | 0 |
| 2 | 47403285 | A | G | T | A | MSH2 | ENST00000233146 | 0.78891 | 0.00877 | 0.78232 | 0.96108 | 0.86149 | 0.548483984 | 0.8763 | 0.9764458537 | 0.157239 | 4.141731247 | 0 |
| 2 | 47403285 | A | T | T | S | MSH2 | ENST00000233146 | 0.58389 | 0.01053 | 0.78932 | 0.95659 | 0.83997 | 0.3830445185 | 0.88886 | 0.9147455096 | 0.162066 | 6.361703705 | 0 |
| 2 | 47403286 | C | A | T | N | MSH2 | ENST00000233146 | 0.62627 | 0.02869 | 0.86271 | 0.95103 | 0.80772 | 0.5835338832 | 0.87747 | 0.9853371978 | 0.135656 | 4.6057577 | 0 |
| 2 | 47403286 | C | G | T | S | MSH2 | ENST00000233146 | 0.47501 | 0.01053 | 0.78932 | 0.92832 | 0.81182 | 0.3830445185 | 0.85477 | 0.6897229552 | 0.104132 | 6.361703705 | 0 |
| 2 | 47403286 | C | T | T | I | MSH2 | ENST00000233146 | 0.85742 | 0.03224 | 0.906 | 0.95366 | 0.94167 | 0.6096759475 | 0.9165 | 0.9967054725 | 0.416319 | 7.346359886 | 0 |
| 2 | 47403288 | A | C | T | P | MSH2 | ENST00000233146 | 0.93619 | 0.03322 | 0.83525 | 0.98612 | 0.94281 | 0.8390329258 | 0.99253 | 0.9095207453 | 0.315799 | 9.220584995 | 0 |
| 2 | 47403288 | A | G | T | A | MSH2 | ENST00000233146 | 0.92044 | 0.03697 | 0.84281 | 0.95837 | 0.87 | 0.7244864664 | 0.93406 | 0.8984457254 | 0.193392 | 4.268712058 | 0 |
| 2 | 47403288 | A | T | T | S | MSH2 | ENST00000233146 | 0.89617 | 0.03848 | 0.84536 | 0.97977 | 0.87733 | 0.5990664809 | 0.96058 | 0.9859636426 | 0.214158 | 4.977069467 | 0 |
| 2 | 47403289 | C | A | T | K | MSH2 | ENST00000233146 | 0.71249 | 0.03426 | 0.9311 | 0.96531 | 0.96444 | 0.8465144121 | 0.94635 | 0.9987798333 | 0.459696 | 6.636425882 | 0 |
| 2 | 47403289 | C | G | T | R | MSH2 | ENST00000233146 | 0.69973 | 0.03518 | 0.92099 | 0.97294 | 0.96664 | 0.8751694836 | 0.95642 | 0.9987572432 | 0.472863 | 9.220584995 | 0 |

| | | | | | | | | | | | | | | | | | | |
|---|---|---|---|---|---|---|---|---|---|---|---|---|---|---|---|---|---|---|
| 2 | 47403289 | C | T | T | I | MSH2 | ENST00000233146 | 0.69123 | 0.03582 | 0.94527 | 0.95887 | 0.93863 | 0.7725186365 | 0.92089 | 0.9541234481 | 0.388086 | 9.220584995 | 0 |
| 2 | 47403291 | G | A | V | M | MSH2 | ENST00000233146 | 0.92809 | 0.01871 | 0.8757 | 0.97474 | 0.88259 | 0.5123986958 | 0.85747 | 0.938442409 | 0.199384 | 5.435615742 | 0 |
| 2 | 47403291 | G | C | V | L | MSH2 | ENST00000233146 | 0.58624 | 0.0058 | 0.84714 | 0.93674 | 0.76157 | 0.447577927 | 0.85322 | 0.7010916471 | 0.139483 | 1.616031477 | 0 |
| 2 | 47403291 | G | T | V | L | MSH2 | ENST00000233146 | 0.58041 | 0.0058 | 0.84714 | 0.94182 | 0.76157 | 0.447577927 | 0.85405 | 0.7010916471 | 0.13947 | 1.616031477 | 0 |
| 2 | 47403292 | T | A | V | E | MSH2 | ENST00000233146 | 0.90848 | 0.01019 | 0.84638 | 0.99386 | 0.95499 | 0.8160079077 | 0.98647 | 0.9972205162 | 0.4828 | 6.992468118 | 0 |
| 2 | 47403292 | T | C | V | A | MSH2 | ENST00000233146 | 0.74467 | 0.00722 | 0.85843 | 0.99035 | 0.93319 | 0.5463694003 | 0.94172 | 0.9837166071 | 0.305065 | 6.992468118 | 0 |
| 2 | 47403292 | T | G | V | G | MSH2 | ENST00000233146 | 0.91097 | 0.0129 | 0.75861 | 0.99361 | 0.95681 | 0.8179431924 | 0.97675 | 0.998141408 | 0.482225 | 6.992468118 | 0 |
| 2 | 47403294 | C | A | R | S | MSH2 | ENST00000233146 | 0.93092 | 0.03155 | 0.90643 | 0.97342 | 0.95608 | 0.8606867758 | 0.97224 | 0.9986373782 | 0.463137 | 8.130029835 | 0 |
| 2 | 47403294 | C | G | R | G | MSH2 | ENST00000233146 | 0.79041 | 0.00615 | 0.89793 | 0.94526 | 0.92844 | 0.8886280119 | 0.97057 | 0.9908910394 | 0.322603 | 7.986442734 | 0 |
| 2 | 47403294 | C | T | R | C | MSH2 | ENST00000233146 | 0.95397 | 0.03212 | 0.95915 | 0.96262 | 0.93083 | 0.8100191464 | 0.93896 | 0.9985167384 | 0.448421 | 8.130029835 | 0 |
| 2 | 47403295 | G | C | R | P | MSH2 | ENST00000233146 | 0.95472 | 0.0307 | 0.93146 | 0.98308 | 0.98071 | 0.9314738728 | 0.97337 | 0.9977261424 | 0.491251 | 8.130029835 | 0 |
| 2 | 47403295 | G | T | R | L | MSH2 | ENST00000233146 | 0.95344 | 0.03197 | 0.92303 | 0.97508 | 0.97327 | 0.8479753674 | 0.96448 | 0.9965738058 | 0.471139 | 3.360211823 | 0 |
| 2 | 47403297 | C | A | L | I | MSH2 | ENST00000233146 | 0.43647 | 0.00566 | 0.83401 | 0.84048 | 0.67108 | 0.2517791144 | 0.76994 | 0.2322661575 | 0.0721054 | 1.834616979 | 0 |
| 2 | 47403297 | C | G | L | V | MSH2 | ENST00000233146 | 0.3853 | 0.00569 | 0.77944 | 0.79252 | 0.71312 | 0.3210796557 | 0.82265 | 0.1144010563 | 0.0762825 | 0.3332931479 | 0 |
| 2 | 47403297 | C | T | L | F | MSH2 | ENST00000233146 | 0.40946 | 0.00556 | 0.82894 | 0.84011 | 0.69339 | 0.4786582567 | 0.85314 | 0.3462142948 | 0.076657 | 0.3440640436 | 0 |
| 2 | 47403298 | T | A | L | H | MSH2 | ENST00000233146 | 0.83073 | 0.02143 | 0.90013 | 0.99025 | 0.95608 | 0.6982573279 | 0.86137 | 0.984872818 | 0.312258 | 5.27382039 | 0 |
| 2 | 47403298 | T | C | L | P | MSH2 | ENST00000233146 | 0.83135 | 0.01315 | 0.93016 | 0.98366 | 0.96371 | 0.8594081077 | 0.97324 | 0.989865005 | 0.455132 | 5.27382039 | 0 |
| 2 | 47403298 | T | G | L | R | MSH2 | ENST00000233146 | 0.82685 | 0.01369 | 0.91067 | 0.98526 | 0.96117 | 0.8109394371 | 0.96701 | 0.9869693518 | 0.445543 | 5.27382039 | 1 |
| 2 | 47403300 | T | A | F | I | MSH2 | ENST00000233146 | 0.90202 | 0.02835 | 0.92079 | 0.97554 | 0.96664 | 0.8044548222 | 0.92546 | 0.9965961576 | 0.404573 | 9.724496782 | 0 |
| 2 | 47403300 | T | C | F | L | MSH2 | ENST00000233146 | 0.89411 | 0.0178 | 0.95103 | 0.97653 | 0.9608 | 0.7818006305 | 0.89646 | 0.9977232814 | 0.330944 | 9.487393832 | 0 |
| 2 | 47403300 | T | G | F | V | MSH2 | ENST00000233146 | 0.90625 | 0.0292 | 0.91564 | 0.97504 | 0.96883 | 0.7743459013 | 0.95498 | 0.9986019731 | 0.450486 | 9.724496782 | 0 |
| 2 | 47403301 | T | A | F | Y | MSH2 | ENST00000233146 | 0.94001 | 0.03274 | 0.94091 | 0.96358 | 0.92723 | 0.7047751723 | 0.91192 | 0.9878175855 | 0.294131 | 4.663295089 | 0 |
| 2 | 47403301 | T | C | F | S | MSH2 | ENST00000233146 | 0.9557 | 0.02683 | 0.92189 | 0.98991 | 0.9755 | 0.8690994457 | 0.96917 | 0.9990202188 | 0.499372 | 9.724496782 | 1 |
| 2 | 47403301 | T | G | F | C | MSH2 | ENST00000233146 | 0.95576 | 0.03099 | 0.94851 | 0.98945 | 0.96298 | 0.8393588586 | 0.92279 | 0.999022007 | 0.374118 | 6.972328515 | 0 |
| 2 | 47403302 | C | A | F | L | MSH2 | ENST00000233146 | 0.70511 | 0.0178 | 0.95103 | 0.99006 | 0.95863 | 0.7818006305 | 0.89287 | 0.9941549897 | 0.290241 | 9.487393832 | 0 |
| 2 | 47403302 | C | G | F | L | MSH2 | ENST00000233146 | 0.71361 | 0.0178 | 0.95103 | 0.98899 | 0.95863 | 0.7818006305 | 0.89284 | 0.9887868762 | 0.290241 | 9.487393832 | 0 |
| 2 | 47403303 | G | A | D | N | MSH2 | ENST00000233146 | 0.655 | 0.03447 | 0.8917 | 0.95313 | 0.8543 | 0.5457735828 | 0.92041 | 0.9892790914 | 0.147007 | 6.057209392 | 0 |
| 2 | 47403303 | G | C | D | H | MSH2 | ENST00000233146 | 0.93915 | 0.03398 | 0.9106 | 0.98333 | 0.91484 | 0.6213478218 | 0.90943 | 0.9974996448 | 0.264193 | 6.28202528 | 0 |
| 2 | 47403303 | G | T | D | Y | MSH2 | ENST00000233146 | 0.94149 | 0.0334 | 0.90198 | 0.98713 | 0.95681 | 0.7110606856 | 0.9731 | 0.9976100922 | 0.466661 | 6.28202528 | 0 |
| 2 | 47403304 | A | C | D | A | MSH2 | ENST00000233146 | 0.95045 | 0.02819 | 0.83978 | 0.98302 | 0.9528 | 0.6441736419 | 0.87266 | 0.9960075617 | 0.319208 | 6.28202528 | 0 |
| 2 | 47403304 | A | G | D | G | MSH2 | ENST00000233146 | 0.95206 | 0.03099 | 0.83804 | 0.97525 | 0.95717 | 0.7045467935 | 0.95367 | 0.9970356226 | 0.443814 | 6.28202528 | 0 |
| 2 | 47403304 | A | T | D | V | MSH2 | ENST00000233146 | 0.95024 | 0.02589 | 0.83306 | 0.98541 | 0.96737 | 0.7032189766 | 0.96794 | 0.9978400469 | 0.47215 | 6.28202528 | 0 |
| 2 | 47403305 | C | A | D | E | MSH2 | ENST00000233146 | 0.35405 | 0.00923 | 0.86358 | 0.8976 | 0.82668 | 0.4321112376 | 0.92135 | 0.8870469928 | 0.140487 | 2.423085551 | 0 |
| 2 | 47403305 | C | G | D | E | MSH2 | ENST00000233146 | 0.35991 | 0.00923 | 0.86358 | 0.89742 | 0.82668 | 0.4321112376 | 0.92102 | 0.5033481121 | -0.0629043 | 2.423085551 | 0 |
| 2 | 47403306 | C | G | R | G | MSH2 | ENST00000233146 | 0.88436 | 0.032 | 0.88475 | 0.97871 | 0.95389 | 0.8160194065 | 0.98793 | 0.9989439845 | 0.458011 | 8.317523519 | 0 |
| 2 | 47403307 | G | A | R | Q | MSH2 | ENST00000233146 | 0.85434 | 0.03243 | 0.8314 | 0.95062 | 0.95133 | 0.7043055663 | 0.97009 | 0.9932408333 | 0.478877 | 8.317523519 | 0 |
| 2 | 47403307 | G | C | R | P | MSH2 | ENST00000233146 | 0.901 | 0.03099 | 0.91453 | 0.96762 | 0.98972 | 0.8954353648 | 0.96989 | 0.9809923768 | 0.411757 | 8.317523519 | 0 |
| 2 | 47403307 | G | T | R | L | MSH2 | ENST00000233146 | 0.87396 | 0.02574 | 0.90827 | 0.96752 | 0.98034 | 0.7842843605 | 0.97086 | 0.9988313317 | 0.486032 | 3.414922616 | 0 |
| 2 | 47403309 | G | A | G | S | MSH2 | ENST00000233146 | 0.54156 | 0.00877 | 0.73864 | 0.9251 | 0.81818 | 0.4946691224 | 0.88572 | 0.3428792655 | 0.332423 | 1.6813534 | 0 |
| 2 | 47403309 | G | C | G | R | MSH2 | ENST00000233146 | 0.9188 | 0.02628 | 0.85542 | 0.97134 | 0.90624 | 0.5732785388 | 0.95123 | 0.9851692915 | 0.306878 | 5.43111851 | 0 |
| 2 | 47403309 | G | T | G | C | MSH2 | ENST00000233146 | 0.94033 | 0.03419 | 0.81937 | 0.98124 | 0.92844 | 0.629258868 | 0.92376 | 0.9975517392 | 0.332385 | 5.43111851 | 0 |
| 2 | 47403310 | G | A | G | D | MSH2 | ENST00000233146 | 0.72196 | 0.02277 | 0.86567 | 0.97175 | 0.89193 | 0.6072471639 | 0.91995 | 0.9867972136 | 0.28603 | 5.43111851 | 0 |
| 2 | 47403310 | G | C | G | A | MSH2 | ENST00000233146 | 0.68019 | 0.01236 | 0.76857 | 0.97207 | 0.85843 | 0.5550626 | 0.93836 | 0.9758618474 | 0.247839 | 3.662038338 | 0 |
| 2 | 47403310 | G | T | G | V | MSH2 | ENST00000233146 | 0.85478 | 0.02574 | 0.77908 | 0.97873 | 0.93123 | 0.6389653679 | 0.93182 | 0.996632278 | 0.337283 | 5.43111851 | 0 |
| 2 | 47403312 | G | A | D | N | MSH2 | ENST00000233146 | 0.93571 | 0.03313 | 0.75296 | 0.97737 | 0.94802 | 0.5491278349 | 0.95084 | 0.9974583983 | 0.195189 | 7.49897395 | 0 |
| 2 | 47403312 | G | C | D | H | MSH2 | ENST00000233146 | 0.93414 | 0.03398 | 0.77921 | 0.9679 | 0.96226 | 0.6977484056 | 0.91882 | 0.9981743097 | 0.422058 | 7.49897395 | 0 |
| 2 | 47403312 | G | T | D | Y | MSH2 | ENST00000233146 | 0.93704 | 0.03362 | 0.75981 | 0.972 | 0.97141 | 0.721093738 | 0.95198 | 0.9983541965 | 0.47207 | 7.49897395 | 0 |
| 2 | 47403313 | A | C | D | A | MSH2 | ENST00000233146 | 0.86269 | 0.02835 | 0.6997 | 0.99158 | 0.96189 | 0.6873523074 | 0.91849 | 0.9956021309 | 0.361751 | 7.49897395 | 0 |
| 2 | 47403313 | A | G | D | G | MSH2 | ENST00000233146 | 0.817 | 0.03013 | 0.69956 | 0.99195 | 0.94318 | 0.6427108138 | 0.93146 | 0.9973310232 | 0.311658 | 7.49897395 | 0 |
| 2 | 47403313 | A | T | D | V | MSH2 | ENST00000233146 | 0.85661 | 0.02984 | 0.69052 | 0.99333 | 0.96371 | 0.7195136219 | 0.97775 | 0.9981746674 | 0.487751 | 7.49897395 | 0 |
| 2 | 47403314 | C | A | D | E | MSH2 | ENST00000233146 | 0.53981 | 0.00722 | 0.71867 | 0.93163 | 0.8367 | 0.4280446098 | 0.85194 | 0.7964774966 | 0.113386 | 2.783784577 | 0 |
| 2 | 47403314 | C | G | D | E | MSH2 | ENST00000233146 | 0.54698 | 0.00722 | 0.71867 | 0.92654 | 0.8367 | 0.4280446098 | 0.85173 | 0.2894119099 | 0.11339 | 2.783784577 | 0 |
| 2 | 47403315 | T | A | F | I | MSH2 | ENST00000233146 | 0.79484 | 0.02023 | 0.81621 | 0.98572 | 0.94728 | 0.6009211362 | 0.89225 | 0.9964770675 | 0.277744 | 6.083498597 | 0 |
| 2 | 47403315 | T | C | F | L | MSH2 | ENST00000233146 | 0.66494 | 0.01385 | 0.86406 | 0.97871 | 0.92844 | 0.542379849 | 0.92231 | 0.9951307774 | 0.290317 | 6.083498597 | 0 |
| 2 | 47403315 | T | G | F | V | MSH2 | ENST00000233146 | 0.66185 | 0.01529 | 0.81092 | 0.98665 | 0.93319 | 0.5709494437 | 0.8631 | 0.9955466986 | 0.27177 | 6.083498597 | 0 |
| 2 | 47403316 | T | A | F | Y | MSH2 | ENST00000233146 | 0.46562 | 0.00556 | 0.84495 | 0.9334 | 0.7255 | 0.4293311136 | 0.781 | 0.2763652007 | 0.0982759 | -0.3455193139 | 0 |
| 2 | 47403316 | T | C | F | S | MSH2 | ENST00000233146 | 0.9391 | 0.01827 | 0.82538 | 0.97964 | 0.94129 | 0.6653084892 | 0.95193 | 0.9964277148 | 0.314925 | 6.083498597 | 0 |
| 2 | 47403316 | T | G | F | C | MSH2 | ENST00000233146 | 0.81559 | 0.02864 | 0.84806 | 0.97685 | 0.911 | 0.643491659 | 0.89522 | 0.9962421656 | 0.26247 | 6.083498597 | 0 |
| 2 | 47403317 | C | A | F | L | MSH2 | ENST00000233146 | 0.51139 | 0.01385 | 0.86406 | 0.96063 | 0.89558 | 0.542379849 | 0.93244 | 0.9771581888 | 0.263629 | 6.083498597 | 0 |
| 2 | 47403317 | C | G | F | L | MSH2 | ENST00000233146 | 0.51836 | 0.01385 | 0.86406 | 0.95855 | 0.89558 | 0.542379849 | 0.93233 | 0.9833209515 | 0.26363 | 6.083498597 | 0 |
| 2 | 47403318 | T | A | Y | N | MSH2 | ENST00000233146 | 0.91468 | 0.02965 | 0.88437 | 0.99033 | 0.98764 | 0.8688958822 | 0.95507 | 0.9997271895 | 0.549028 | 9.358510634 | 0 |
| 2 | 47403318 | T | C | Y | H | MSH2 | ENST00000233146 | 0.91115 | 0.03313 | 0.89133 | 0.98914 | 0.98972 | 0.8207810446 | 0.96744 | 0.9995239973 | 0.566972 | 9.358510634 | 0 |
| 2 | 47403318 | T | G | Y | D | MSH2 | ENST00000233146 | 0.91839 | 0.02844 | 0.88256 | 0.99108 | 0.99356 | 0.896412461 | 0.9891 | 0.9998277426 | 0.568663 | 9.358510634 | 0 |
| 2 | 47403319 | A | C | Y | S | MSH2 | ENST00000233146 | 0.92303 | 0.03166 | 0.82242 | 0.99074 | 0.99824 | 0.8672239685 | 0.98836 | 0.999815166 | 0.57731 | 9.358510634 | 0 |
| 2 | 47403319 | A | G | Y | C | MSH2 | ENST00000233146 | 0.93619 | 0.03362 | 0.89266 | 0.98776 | 0.96334 | 0.8389710801 | 0.98208 | 0.997503221 | 0.400182 | 9.358510634 | 0 |
| 2 | 47403319 | A | T | Y | F | MSH2 | ENST00000233146 | 0.72292 | 0.0129 | 0.81068 | 0.97258 | 0.9458 | 0.6045682983 | 0.87631 | 0.9651254416 | 0.316291 | 6.371012237 | 0 |
| 2 | 47403321 | A | C | T | P | MSH2 | ENST00000233146 | 0.89468 | 0.0315 | 0.53962 | 0.99034 | 0.93901 | 0.8105019864 | 0.93092 | 0.9875770807 | 0.311839 | 7.609555298 | 1 |
| 2 | 47403321 | A | G | T | A | MSH2 | ENST00000233146 | 0.84043 | 0.02108 | 0.54949 | 0.9786 | 0.91271 | 0.6444669788 | 0.93321 | 0.9870797992 | 0.275347 | 7.609555298 | 0 |
| 2 | 47403321 | A | T | T | S | MSH2 | ENST00000233146 | 0.48844 | 0.01019 | 0.54755 | 0.94958 | 0.88448 | 0.5024808794 | 0.89956 | 0.8183500171 | 0.212806 | 6.943930205 | 0 |
| 2 | 47403322 | C | A | T | K | MSH2 | ENST00000233146 | 0.69852 | 0.03231 | 0.65463 | 0.9605 | 0.95023 | 0.789407626 | 0.94251 | 0.9981226325 | 0.468331 | 7.609555298 | 0 |
| 2 | 47403322 | C | G | T | R | MSH2 | ENST00000233146 | 0.68651 | 0.0337 | 0.63697 | 0.96159 | 0.95572 | 0.8300184893 | 0.93616 | 0.9994533658 | 0.460272 | 7.609555298 | 0 |
| 2 | 47403324 | G | A | A | T | MSH2 | ENST00000233146 | 0.57491 | 0.00596 | 0.54309 | 0.95034 | 0.80058 | 0.5075818358 | 0.87164 | 0.869207263 | 0.146992 | 1.650765024 | 0 |
| 2 | 47403324 | G | C | A | P | MSH2 | ENST00000233146 | 0.88035 | 0.01133 | 0.59432 | 0.97702 | 0.92642 | 0.8481957452 | 0.95624 | 0.925585866 | 0.361787 | 5.053754839 | 1 |
| 2 | 47403324 | G | T | A | S | MSH2 | ENST00000233146 | 0.54875 | 0.00608 | 0.52101 | 0.93315 | 0.77718 | 0.5902283549 | 0.85909 | 0.6749173403 | 0.125048 | 2.892718194 | 0 |
| 2 | 47403325 | C | G | A | G | MSH2 | ENST00000233146 | 0.30642 | 0.00881 | 0.52738 | 0.97922 | 0.80772 | 0.6424455837 | 0.87989 | 0.876541853 | 0.13524 | 5.053754839 | 0 |
| 2 | 47403325 | C | T | A | V | MSH2 | ENST00000233146 | 0.15071 | 0.00574 | 0.56887 | 0.80288 | 0.7727 | 0.3464390294 | 0.98756 | 0.07561734228 | 0.165144 | -1.400252919 | 0 |
| 2 | 47403327 | C | A | H | N | MSH2 | ENST00000233146 | 0.74165 | 0.03076 | 0.82466 | 0.96777 | 0.94543 | 0.7669371261 | 0.93551 | 0.998423934 | 0.294583 | 7.644573817 | 0 |
| 2 | 47403327 | C | G | H | D | MSH2 | ENST00000233146 | 0.82186 | 0.02896 | 0.81561 | 0.96736 | 0.97885 | 0.8773539277 | 0.97606 | 0.999663949 | 0.489378 | 7.644573817 | 0 |
| 2 | 47403327 | C | T | H | Y | MSH2 | ENST00000233146 | 0.59436 | 0.01048 | 0.83352 | 0.90829 | 0.95023 | 0.7888350264 | 0.95776 | 0.9975332022 | 0.407258 | 6.422786697 | 0 |
| 2 | 47403328 | A | C | H | P | MSH2 | ENST00000233146 | 0.89018 | 0.02869 | 0.76102 | 0.98499 | 0.96444 | 0.8907831513 | 0.97944 | 0.9995580316 | 0.494616 | 7.40918942 | 0 |
| 2 | 47403328 | A | G | H | R | MSH2 | ENST00000233146 | 0.65023 | 0.00827 | 0.83067 | 0.98085 | 0.96846 | 0.8513501384 | 0.98301 | 0.9975209832 | 0.49462 | 7.644573817 | 0 |
| 2 | 47403328 | A | T | H | L | MSH2 | ENST00000233146 | 0.88641 | 0.0276 | 0.794 | 0.98293 | 0.95316 | 0.817722126 | 0.98582 | 0.9998980761 | 0.463382 | 7.644573817 | 0 |
| 2 | 47403329 | C | A | H | Q | MSH2 | ENST00000233146 | 0.7426 | 0.03293 | 0.79504 | 0.9799 | 0.9681 | 0.8011364023 | 0.98273 | 0.9982616305 | 0.458396 | 7.644573817 | 0 |
| 2 | 47403329 | C | G | H | Q | MSH2 | ENST00000233146 | 0.75105 | 0.03293 | 0.79504 | 0.97524 | 0.96773 | 0.8011364023 | 0.98268 | 0.7936590314 | 0.225772 | 7.644573817 | 0 |
| 2 | 47403330 | G | A | G | S | MSH2 | ENST00000233146 | 0.70398 | 0.01037 | 0.33463 | 0.91636 | 0.85945 | 0.6185888226 | 0.92796 | 0.9907941222 | 0.149873 | 6.683849879 | 0 |
| 2 | 47403330 | G | C | G | R | MSH2 | ENST00000233146 | 0.73181 | 0.00904 | 0.42339 | 0.91713 | 0.85064 | 0.7341305009 | 0.9038 | 0.9704287052 | 0.0890686 | 7.298234493 | 0 |
| 2 | 47403330 | G | T | G | C | MSH2 | ENST00000233146 | 0.90695 | 0.03442 | 0.38988 | 0.95717 | 0.87148 | 0.6801295926 | 0.93776 | 0.9885104299 | 0.278402 | 7.857686717 | 0 |
| 2 | 47403331 | G | C | G | A | MSH2 | ENST00000233146 | 0.60105 | 0.01221 | 0.35268 | 0.91427 | 0.84959 | 0.6271294667 | 0.93238 | 0.9916430712 | 0.14631 | 5.044996791 | 0 |
| 2 | 47403331 | G | T | G | V | MSH2 | ENST00000233146 | 0.67247 | 0.02653 | 0.35282 | 0.94843 | 0.88916 | 0.6992807223 | 0.91546 | 0.9963880777 | 0.295933 | 7.635453062 | 0 |
| 2 | 47403333 | G | A | E | K | MSH2 | ENST00000233146 | 0.523 | 0.00547 | 0.62601 | 0.9016 | 0.71861 | 0.4070052784 | 0.7974 | 0.8358274102 | 0.0845752 | 0.9796843351 | 0 |
| 2 | 47403333 | G | C | E | Q | MSH2 | ENST00000233146 | 0.51197 | 0.00596 | 0.51213 | 0.9331 | 0.75485 | 0.3737100982 | 0.87878 | 0.7215909958 | 0.109702 | 1.235062535 | 0 |
| 2 | 47403334 | A | C | E | A | MSH2 | ENST00000233146 | 0.40646 | 0.00608 | 0.55016 | 0.95484 | 0.81876 | 0.4280595789 | 0.89647 | 0.7243471742 | 0.128763 | 2.87304416 | 0 |
| 2 | 47403334 | A | G | E | G | MSH2 | ENST00000233146 | 0.47018 | 0.00543 | 0.5056 | 0.94682 | 0.77718 | 0.4305242085 | 0.89893 | 0.9226583838 | 0.118712 | 3.175949655 | 0 |
| 2 | 47403334 | A | T | E | V | MSH2 | ENST00000233146 | 0.46438 | 0.00619 | 0.53 | 0.96492 | 0.86753 | 0.4749949137 | 0.92535 | 0.9627689719 | 0.204911 | 5.054633879 | 0 |
| 2 | 47403335 | G | C | E | D | MSH2 | ENST00000233146 | 0.48133 | 0.00586 | 0.54371 | 0.87227 | 0.70914 | 0.3603551418 | 0.90372 | 0.3880225718 | 0.0886054 | 1.453012184 | 0 |
| 2 | 47403335 | G | T | E | D | MSH2 | ENST00000233146 | 0.47615 | 0.00586 | 0.54371 | 0.87227 | 0.70914 | 0.3603551418 | 0.9038 | 0.4890603721 | 0.0886088 | 1.453012184 | 0 |
| 2 | 47403336 | G | A | D | N | MSH2 | ENST00000233146 | 0.94142 | 0.03557 | 0.698 | 0.98185 | 0.94129 | 0.7884429237 | 0.95856 | 0.9990912676 | 0.206502 | 9.574422486 | 0 |
| 2 | 47403336 | G | C | D | H | MSH2 | ENST00000233146 | 0.94648 | 0.03426 | 0.72009 | 0.98979 | 0.99138 | 0.8309032369 | 0.97606 | 0.9995372295 | 0.561547 | 9.574422486 | 0 |
| 2 | 47403337 | A | C | D | A | MSH2 | ENST00000233146 | 0.93964 | 0.03219 | 0.65373 | 0.99609 | 0.97997 | 0.8420204553 | 0.95979 | 0.9981430769 | 0.50106 | 9.574422486 | 0 |
| 2 | 47403337 | A | G | D | G | MSH2 | ENST00000233146 | 0.94191 | 0.03189 | 0.65476 | 0.99567 | 0.9822 | 0.8329882404 | 0.97812 | 0.9980247021 | 0.549805 | 9.574422486 | 0 |
| 2 | 47403337 | A | T | D | V | MSH2 | ENST00000233146 | 0.93745 | 0.03059 | 0.64893 | 0.99552 | 0.98693 | 0.8449080209 | 0.98878 | 0.9993226528 | 0.549291 | 9.574422486 | 0 |

| | | | | | | | | | | | | | | | | |
|---|---|---|---|---|---|---|---|---|---|---|---|---|---|---|---|---|
| 2 | 47403338 | C | A | D | E | MSH2 | ENST00000233146 | 0.64648 | 0.03706 | 0.67487 | 0.99185 | 0.92763 | 0.7414763425 | 0.979 | 0.9915002584 | 0.419517 | 9.574422486 | 0 |
| 2 | 47403338 | C | G | D | E | MSH2 | ENST00000233146 | 0.65477 | 0.03706 | 0.67487 | 0.9923 | 0.92763 | 0.7414763425 | 0.97893 | 0.9966552258 | 0.419519 | 9.574422486 | 0 |
| 2 | 47403339 | G | A | A | T | MSH2 | ENST00000233146 | 0.92272 | 0.03534 | 0.69122 | 0.98231 | 0.99356 | 0.7244126124 | 0.98772 | 0.9996342659 | 0.575295 | 9.839260201 | 0 |
| 2 | 47403339 | G | C | A | P | MSH2 | ENST00000233146 | 0.94387 | 0.03265 | 0.74021 | 0.9819 | 0.99722 | 0.8997318725 | 0.98284 | 0.9994922876 | 0.57243 | 9.839260201 | 0 |
| 2 | 47403339 | G | T | A | S | MSH2 | ENST00000233146 | 0.92371 | 0.03475 | 0.65323 | 0.97822 | 0.97885 | 0.6237719783 | 0.95606 | 0.9951375127 | 0.42909 | 9.839260201 | 0 |
| 2 | 47403340 | C | A | A | E | MSH2 | ENST00000233146 | 0.76013 | 0.03257 | 0.74437 | 0.98723 | 0.99578 | 0.8955241053 | 0.98096 | 0.9998879433 | 0.566172 | 9.839260201 | 0 |
| 2 | 47403340 | C | G | A | G | MSH2 | ENST00000233146 | 0.58874 | 0.0143 | 0.6725 | 0.98429 | 0.98693 | 0.6335004568 | 0.94329 | 0.9962282181 | 0.524409 | 9.839260201 | 0 |
| 2 | 47403340 | C | T | A | V | MSH2 | ENST00000233146 | 0.77275 | 0.03497 | 0.71274 | 0.9877 | 0.99786 | 0.757886279 | 0.98159 | 0.9903548956 | 0.565347 | 9.839260201 | 0 |
| 2 | 47403342 | C | A | L | M | MSH2 | ENST00000233146 | 0.78439 | 0.02255 | 0.62783 | 0.92887 | 0.77845 | 0.3022222621 | 0.76657 | 0.9431782961 | 0.0982624 | 3.454164059 | 0 |
| 2 | 47403342 | C | G | L | V | MSH2 | ENST00000233146 | 0.42156 | 0.00592 | 0.57943 | 0.90018 | 0.71627 | 0.2930154856 | 0.8484 | 0.1216070245 | 0.0654424 | 1.78069348 | 0 |
| 2 | 47403343 | T | A | L | Q | MSH2 | ENST00000233146 | 0.65609 | 0.00659 | 0.63726 | 0.96547 | 0.89101 | 0.4167613402 | 0.85845 | 0.9375016093 | 0.216988 | 3.60077576 | 0 |
| 2 | 47403343 | T | C | L | P | MSH2 | ENST00000233146 | 0.70976 | 0.02037 | 0.74421 | 0.98656 | 0.93397 | 0.7516609727 | 0.9369 | 0.9851314425 | 0.346063 | 6.763018279 | 0 |
| 2 | 47403343 | T | G | L | R | MSH2 | ENST00000233146 | 0.69212 | 0.01523 | 0.7184 | 0.97173 | 0.90755 | 0.5929926972 | 0.86992 | 0.9810186625 | 0.233392 | 3.350060388 | 0 |
| 2 | 47403345 | C | A | L | M | MSH2 | ENST00000233146 | 0.61179 | 0.00777 | 0.68683 | 0.91309 | 0.73074 | 0.3693784193 | 0.79654 | 0.908927381 | 0.1303 | 4.871417725 | 0 |
| 2 | 47403345 | C | G | L | V | MSH2 | ENST00000233146 | 0.58581 | 0.00866 | 0.65434 | 0.93202 | 0.81761 | 0.3989997629 | 0.83215 | 0.9502987266 | 0.140891 | 2.595616814 | 0 |
| 2 | 47403346 | T | A | L | Q | MSH2 | ENST00000233146 | 0.81677 | 0.0143 | 0.70255 | 0.98484 | 0.91694 | 0.5579343151 | 0.8982 | 0.9936465025 | 0.266896 | 4.191928443 | 0 |
| 2 | 47403346 | T | C | L | P | MSH2 | ENST00000233146 | 0.94464 | 0.0178 | 0.78687 | 0.99508 | 0.96773 | 0.814376288 | 0.94205 | 0.995790422 | 0.359964 | 7.103808582 | 0 |
| 2 | 47403346 | T | G | L | R | MSH2 | ENST00000233146 | 0.85286 | 0.01263 | 0.76632 | 0.99338 | 0.9328 | 0.7380878831 | 0.87565 | 0.9946377873 | 0.275361 | 3.429782609 | 0 |
| 2 | 47403348 | G | A | A | T | MSH2 | ENST00000233146 | 0.92457 | 0.03618 | 0.79744 | 0.94794 | 0.94506 | 0.5191412146 | 0.80647 | 0.9954720736 | 0.259162 | 4.762990263 | 0 |
| 2 | 47403348 | G | C | A | P | MSH2 | ENST00000233146 | 0.93164 | 0.03287 | 0.844 | 0.94596 | 0.9729 | 0.8160893082 | 0.90666 | 0.995087564 | 0.433367 | 7.742826831 | 0 |
| 2 | 47403348 | G | T | A | S | MSH2 | ENST00000233146 | 0.90565 | 0.03741 | 0.76294 | 0.96119 | 0.94655 | 0.5218888527 | 0.8198 | 0.8640098572 | 0.259487 | 6.202749814 | 0 |
| 2 | 47403349 | C | A | A | D | MSH2 | ENST00000233146 | 0.73645 | 0.03083 | 0.87251 | 0.97694 | 0.96919 | 0.8540551827 | 0.94786 | 0.9938082099 | 0.443655 | 7.742826831 | 1 |
| 2 | 47403349 | C | G | A | G | MSH2 | ENST00000233146 | 0.72764 | 0.03602 | 0.78022 | 0.97448 | 0.92924 | 0.5414405268 | 0.85823 | 0.9942112565 | 0.259838 | 3.645853799 | 0 |
| 2 | 47403349 | C | T | A | V | MSH2 | ENST00000233146 | 0.40068 | 0.01115 | 0.81351 | 0.92093 | 0.8345 | 0.4590147436 | 0.82565 | 0.4721865147 | 0.177418 | 2.098860785 | 0 |
| 2 | 47403351 | G | A | A | T | MSH2 | ENST00000233146 | 0.95321 | 0.03166 | 0.80614 | 0.97573 | 0.96773 | 0.6669653237 | 0.90522 | 0.9971293807 | 0.352977 | 6.635903696 | 0 |
| 2 | 47403351 | G | C | A | P | MSH2 | ENST00000233146 | 0.95434 | 0.03265 | 0.859 | 0.97643 | 0.98071 | 0.8771296526 | 0.94604 | 0.9994546771 | 0.486492 | 8.977526419 | 0 |
| 2 | 47403351 | G | T | A | S | MSH2 | ENST00000233146 | 0.87443 | 0.02349 | 0.7666 | 0.95963 | 0.90668 | 0.5503497388 | 0.83414 | 0.9638078809 | 0.21634 | 2.76038318 | 0 |
| 2 | 47403352 | C | G | A | G | MSH2 | ENST00000233146 | 0.86949 | 0.03243 | 0.78919 | 0.96409 | 0.97178 | 0.5730284568 | 0.88643 | 0.9979064465 | 0.27671 | 8.977526419 | 0 |
| 2 | 47403352 | C | T | A | V | MSH2 | ENST00000233146 | 0.87576 | 0.03484 | 0.83019 | 0.9694 | 0.96956 | 0.7149754188 | 0.91888 | 0.9993498921 | 0.445312 | 7.803893136 | 0 |
| 2 | 47403354 | C | G | R | G | MSH2 | ENST00000233146 | 0.74021 | 0.01492 | 0.6783 | 0.96036 | 0.9045 | 0.4753257903 | 0.93453 | 0.8920362592 | 0.112067 | 6.618491902 | 0 |
| 2 | 47403354 | C | T | R | W | MSH2 | ENST00000233146 | 0.94575 | 0.03431 | 0.78527 | 0.97358 | 0.92316 | 0.5910404879 | 0.89652 | 0.9991073012 | 0.278083 | 6.863307412 | 1 |
| 2 | 47403355 | G | A | R | Q | MSH2 | ENST00000233146 | 0.44869 | 0.01408 | 0.6103 | 0.90171 | 0.78782 | 0.3442430452 | 0.93308 | 0.4827722311 | -0.00118455 | 2.154379662 | 0 |
| 2 | 47403355 | G | C | R | P | MSH2 | ENST00000233146 | 0.78185 | 0.0254 | 0.71999 | 0.96536 | 0.94015 | 0.7705833518 | 0.96852 | 0.988928318 | 0.376513 | 5.823624559 | 1 |
| 2 | 47403355 | G | T | R | L | MSH2 | ENST00000233146 | 0.58247 | 0.01721 | 0.70958 | 0.96135 | 0.8705 | 0.5045363027 | 0.93604 | 0.9952802658 | 0.238597 | 5.169718799 | 0 |
| 2 | 47403357 | G | A | E | K | MSH2 | ENST00000233146 | 0.9556 | 0.02307 | 0.88246 | 0.93371 | 0.91143 | 0.6789875332 | 0.98889 | 0.9889255166 | 0.411512 | 7.731791355 | 0 |
| 2 | 47403357 | G | C | E | Q | MSH2 | ENST00000233146 | 0.95183 | 0.03006 | 0.77064 | 0.90454 | 0.87829 | 0.5504569601 | 0.9318 | 0.9736353159 | 0.228207 | 7.731791355 | 0 |
| 2 | 47403358 | A | C | E | A | MSH2 | ENST00000233146 | 0.84058 | 0.02361 | 0.79204 | 0.949 | 0.91271 | 0.6207466335 | 0.90922 | 0.9873850346 | 0.236266 | 4.601759851 | 0 |
| 2 | 47403358 | A | G | E | G | MSH2 | ENST00000233146 | 0.89325 | 0.0227 | 0.74365 | 0.97161 | 0.91945 | 0.635078879 | 0.89315 | 0.9970707893 | 0.276614 | 7.731791355 | 0 |
| 2 | 47403358 | A | T | E | V | MSH2 | ENST00000233146 | 0.71762 | 0.02857 | 0.76762 | 0.97685 | 0.91611 | 0.6116277299 | 0.95289 | 0.994086504 | 0.316717 | 3.002033214 | 0 |
| 2 | 47403359 | G | C | E | D | MSH2 | ENST00000233146 | 0.42038 | 0.00687 | 0.79083 | 0.89367 | 0.84853 | 0.5313919269 | 0.91726 | 0.2275236547 | 0.140868 | 3.261526501 | 0 |
| 2 | 47403359 | G | T | E | D | MSH2 | ENST00000233146 | 0.41558 | 0.00687 | 0.79083 | 0.90781 | 0.84853 | 0.5313919269 | 0.91711 | 0.2275236547 | 0.14087 | 3.261526501 | 0 |
| 2 | 47403360 | G | A | V | M | MSH2 | ENST00000233146 | 0.95371 | 0.02663 | 0.80398 | 0.95391 | 0.94839 | 0.5169346404 | 0.90588 | 0.9938054085 | 0.385921 | 3.999713125 | 0 |
| 2 | 47403360 | G | C | V | L | MSH2 | ENST00000233146 | 0.73853 | 0.00619 | 0.76767 | 0.91295 | 0.92601 | 0.4961559397 | 0.95156 | 0.9816774726 | 0.409423 | 1.943622275 | 0 |
| 2 | 47403360 | G | T | V | L | MSH2 | ENST00000233146 | 0.73197 | 0.00619 | 0.76767 | 0.9126 | 0.92601 | 0.4961559397 | 0.94865 | 0.9816774726 | 0.40944 | 1.943622275 | 0 |
| 2 | 47403361 | T | A | V | E | MSH2 | ENST00000233146 | 0.94782 | 0.02614 | 0.77564 | 0.99313 | 0.9729 | 0.7800739356 | 0.96871 | 0.9967965484 | 0.486614 | 6.732111825 | 0 |
| 2 | 47403361 | T | C | V | A | MSH2 | ENST00000233146 | 0.93745 | 0.01818 | 0.78719 | 0.98579 | 0.9608 | 0.5588140225 | 0.92004 | 0.9919841886 | 0.33869 | 6.732111825 | 0 |
| 2 | 47403361 | T | G | V | G | MSH2 | ENST00000233146 | 0.94711 | 0.02891 | 0.66857 | 0.99118 | 0.97735 | 0.7127938391 | 0.96906 | 0.9967965484 | 0.487979 | 6.732111825 | 0 |
| 2 | 47403363 | T | A | F | I | MSH2 | ENST00000233146 | 0.93737 | 0.03224 | 0.91937 | 0.99239 | 0.95023 | 0.7295382465 | 0.88083 | 0.998281002 | 0.309595 | 7.474966804 | 1 |
| 2 | 47403363 | T | C | F | L | MSH2 | ENST00000233146 | 0.94301 | 0.03491 | 0.94133 | 0.98543 | 0.95207 | 0.6528612643 | 0.88823 | 0.9959580302 | 0.334241 | 7.474966804 | 0 |
| 2 | 47403363 | T | G | F | V | MSH2 | ENST00000233146 | 0.93936 | 0.03313 | 0.91073 | 0.99388 | 0.9703 | 0.6839994874 | 0.92566 | 0.9987665415 | 0.456948 | 7.474966804 | 0 |
| 2 | 47403364 | T | C | F | S | MSH2 | ENST00000233146 | 0.95185 | 0.02864 | 0.91882 | 0.99199 | 0.98513 | 0.7637433203 | 0.93501 | 0.9995243549 | 0.470658 | 7.474966804 | 0 |
| 2 | 47403365 | C | A | F | L | MSH2 | ENST00000233146 | 0.92044 | 0.03491 | 0.94133 | 0.9884 | 0.94618 | 0.6528612643 | 0.87075 | 0.9971705079 | 0.303539 | 7.474966804 | 0 |
| 2 | 47403365 | C | G | F | L | MSH2 | ENST00000233146 | 0.92453 | 0.03491 | 0.94133 | 0.98822 | 0.94618 | 0.6528612643 | 0.87038 | 0.9917291999 | 0.165006 | 7.474966804 | 0 |
| 2 | 47403366 | A | C | K | Q | MSH2 | ENST00000233146 | 0.95101 | 0.02937 | 0.79633 | 0.98039 | 0.95499 | 0.5697682883 | 0.93113 | 0.9959753156 | 0.310122 | 4.557398796 | 0 |
| 2 | 47403366 | A | G | K | E | MSH2 | ENST00000233146 | 0.95383 | 0.02403 | 0.83907 | 0.98351 | 0.96773 | 0.6360141112 | 0.94539 | 0.9911594987 | 0.400787 | 2.886682376 | 0 |
| 2 | 47403367 | A | C | K | T | MSH2 | ENST00000233146 | 0.93649 | 0.03088 | 0.84854 | 0.98618 | 0.95133 | 0.6318435259 | 0.92646 | 0.9916166663 | 0.317237 | 7.706741973 | 0 |
| 2 | 47403367 | A | G | K | R | MSH2 | ENST00000233146 | 0.81214 | 0.00816 | 0.77441 | 0.93378 | 0.86149 | 0.483768302 | 0.88858 | 0.9335402846 | 0.139447 | 4.789575437 | 0 |
| 2 | 47403367 | A | T | K | M | MSH2 | ENST00000233146 | 0.94562 | 0.0315 | 0.77831 | 0.98992 | 0.95681 | 0.6749520968 | 0.93453 | 0.9935796261 | 0.334002 | 7.706741973 | 1 |
| 2 | 47403368 | G | C | K | N | MSH2 | ENST00000233146 | 0.95087 | 0.02994 | 0.86629 | 0.96062 | 0.88776 | 0.6220363322 | 0.91243 | 0.9929428101 | 0.18083 | 4.729977411 | 0 |
| 2 | 47403368 | G | T | K | N | MSH2 | ENST00000233146 | 0.95006 | 0.02994 | 0.86629 | 0.96062 | 0.88776 | 0.6220363322 | 0.90985 | 0.9929428101 | 0.180846 | 4.729977411 | 0 |
| 2 | 47403369 | A | C | T | P | MSH2 | ENST00000233146 | 0.95311 | 0.03293 | 0.73523 | 0.99507 | 0.99105 | 0.7557054429 | 0.95567 | 0.9986082911 | 0.499247 | 8.181911586 | 1 |
| 2 | 47403369 | A | G | T | A | MSH2 | ENST00000233146 | 0.94664 | 0.03404 | 0.74092 | 0.99404 | 0.98294 | 0.6455647781 | 0.94504 | 0.9976426959 | 0.39122 | 8.181911586 | 0 |
| 2 | 47403369 | A | T | T | S | MSH2 | ENST00000233146 | 0.63627 | 0.01924 | 0.7496 | 0.9801 | 0.91058 | 0.4812032265 | 0.92363 | 0.9788845778 | 0.259302 | 8.181911586 | 0 |
| 2 | 47403370 | C | A | T | N | MSH2 | ENST00000233146 | 0.68571 | 0.03465 | 0.81184 | 0.98702 | 0.94839 | 0.7082755489 | 0.91138 | 0.9964752793 | 0.203037 | 7.948623337 | 0 |
| 2 | 47403370 | C | G | T | S | MSH2 | ENST00000233146 | 0.37634 | 0.01924 | 0.7496 | 0.96434 | 0.90362 | 0.4812032265 | 0.90848 | 0.9836581349 | 0.248553 | 8.181911586 | 0 |
| 2 | 47403370 | C | T | T | I | MSH2 | ENST00000233146 | 0.73086 | 0.0357 | 0.86063 | 0.98657 | 0.96919 | 0.7255921428 | 0.92992 | 0.9988561869 | 0.448499 | 2.812730918 | 0 |
| 2 | 47403372 | C | A | Q | K | MSH2 | ENST00000233146 | 0.46295 | 0.00543 | 0.56282 | 0.94306 | 0.71938 | 0.4236110803 | 0.89516 | 0.6928998828 | 0.118118 | 3.610352993 | 0 |
| 2 | 47403372 | C | G | Q | E | MSH2 | ENST00000233146 | 0.44126 | 0.00747 | 0.50101 | 0.93752 | 0.74235 | 0.3769090544 | 0.85896 | 0.7701351643 | 0.117763 | 3.610352993 | 0 |
| 2 | 47403373 | A | C | Q | P | MSH2 | ENST00000233146 | 0.63336 | 0.01203 | 0.5199 | 0.97637 | 0.90229 | 0.6557071725 | 0.91568 | 0.9025923014 | 0.212871 | 3.610352993 | 0 |
| 2 | 47403373 | A | G | Q | R | MSH2 | ENST00000233146 | 0.54487 | 0.00562 | 0.55007 | 0.96282 | 0.63947 | 0.3747831234 | 0.92887 | 0.6213051081 | 0.100073 | 0.826346671 | 0 |
| 2 | 47403373 | A | T | Q | L | MSH2 | ENST00000233146 | 0.60222 | 0.0058 | 0.48377 | 0.94719 | 0.78906 | 0.4302345485 | 0.90228 | 0.5566065311 | 0.144216 | 0.5900352773 | 0 |
| 2 | 47403374 | G | C | Q | H | MSH2 | ENST00000233146 | 0.57836 | 0.00619 | 0.54248 | 0.94522 | 0.78906 | 0.4566432062 | 0.89109 | 0.1896701673 | 0.0358112 | 3.610352993 | 0 |
| 2 | 47403374 | G | T | Q | H | MSH2 | ENST00000233146 | 0.57266 | 0.00619 | 0.54248 | 0.94486 | 0.78906 | 0.4566432062 | 0.89118 | 0.08775585467 | 0.0982854 | 3.610352993 | 0 |
| 2 | 47403375 | G | A | G | R | MSH2 | ENST00000233146 | 0.95183 | 0.02972 | 0.75415 | 0.97031 | 0.95462 | 0.6627042764 | 0.9597 | 0.9835643768 | 0.372327 | 5.443372801 | 0 |
| 2 | 47403375 | G | C | G | R | MSH2 | ENST00000233146 | 0.95149 | 0.02972 | 0.75415 | 0.96697 | 0.95462 | 0.6627042764 | 0.95786 | 0.9929618239 | 0.372336 | 5.443372801 | 0 |
| 2 | 47403376 | G | A | G | E | MSH2 | ENST00000233146 | 0.83066 | 0.02516 | 0.72724 | 0.93408 | 0.90842 | 0.6110291582 | 0.86133 | 0.9930878282 | 0.266203 | 5.147384761 | 0 |
| 2 | 47403376 | G | C | G | A | MSH2 | ENST00000233146 | 0.4824 | 0.00654 | 0.62725 | 0.85909 | 0.64876 | 0.4734056812 | 0.82935 | 0.8124880791 | 0.10088 | 1.657939237 | 0 |
| 2 | 47403376 | G | T | G | V | MSH2 | ENST00000233146 | 0.82973 | 0.02324 | 0.63694 | 0.94317 | 0.91186 | 0.6309642591 | 0.91399 | 0.9970151186 | 0.333157 | 5.092300661 | 0 |
| 2 | 47403378 | G | A | V | M | MSH2 | ENST00000233146 | 0.93114 | 0.03706 | 0.74675 | 0.98978 | 0.9367 | 0.6221657249 | 0.89674 | 0.9961432815 | 0.285354 | 6.970794323 | 0 |
| 2 | 47403378 | G | C | V | L | MSH2 | ENST00000233146 | 0.91115 | 0.0364 | 0.70402 | 0.98474 | 0.90581 | 0.5806703894 | 0.85384 | 0.989713788 | 0.208536 | 6.743941947 | 0 |
| 2 | 47403378 | G | T | V | L | MSH2 | ENST00000233146 | 0.90725 | 0.0364 | 0.70402 | 0.98439 | 0.90581 | 0.5806703894 | 0.85343 | 0.989713788 | 0.208536 | 6.743941947 | 0 |
| 2 | 47403379 | T | C | V | A | MSH2 | ENST00000233146 | 0.69292 | 0.02516 | 0.72948 | 0.97818 | 0.90318 | 0.591502148 | 0.73257 | 0.9932940006 | 0.24521 | 6.970794323 | 0 |
| 2 | 47403379 | T | G | V | G | MSH2 | ENST00000233146 | 0.95088 | 0.03265 | 0.58986 | 0.99191 | 0.97067 | 0.721383453 | 0.86743 | 0.9987603426 | 0.369616 | 6.970794323 | 0 |
| 2 | 47403381 | A | C | I | L | MSH2 | ENST00000233146 | 0.43841 | 0.00586 | 0.4422 | 0.80509 | 0.69425 | 0.4085560346 | 0.83101 | 0.9940989017 | 0.0573076 | 5.12537617 | 0 |
| 2 | 47403381 | A | G | I | V | MSH2 | ENST00000233146 | 0.35691 | 0.00615 | 0.37234 | 0.7381 | 0.68995 | 0.2833952337 | 0.78776 | 0.08607670665 | 0.0506815 | 1.186928147 | 0 |
| 2 | 47403381 | A | T | I | F | MSH2 | ENST00000233146 | 0.75782 | 0.01492 | 0.46714 | 0.93072 | 0.85894 | 0.6584309454 | 0.92703 | 0.9846877456 | 0.16701 | 6.843772525 | 0 |
| 2 | 47403382 | T | A | I | N | MSH2 | ENST00000233146 | 0.95453 | 0.02324 | 0.5223 | 0.97381 | 0.93397 | 0.7377843419 | 0.80955 | 0.9975110292 | 0.224355 | 5.136677522 | 0 |
| 2 | 47403382 | T | C | I | T | MSH2 | ENST00000233146 | 0.86147 | 0.01365 | 0.55048 | 0.95186 | 0.90624 | 0.6454902492 | 0.85 | 0.9930865169 | 0.262536 | 3.815372046 | 0 |
| 2 | 47403382 | T | G | I | S | MSH2 | ENST00000233146 | 0.91801 | 0.0129 | 0.4717 | 0.97038 | 0.93004 | 0.8054188635 | 0.83707 | 0.9965163469 | 0.279614 | 2.780728011 | 0 |
| 2 | 47403383 | C | G | I | M | MSH2 | ENST00000233146 | 0.76124 | 0.00872 | 0.43551 | 0.89929 | 0.86753 | 0.4921913277 | 0.818 | 0.9028645754 | 0.142392 | 6.843772525 | 0 |
| 2 | 47403384 | A | C | K | Q | MSH2 | ENST00000233146 | 0.95466 | 0.03484 | 0.73287 | 0.9847 | 0.89513 | 0.7004826582 | 0.93866 | 0.9866786599 | 0.222503 | 7.911739371 | 0 |
| 2 | 47403384 | A | G | K | E | MSH2 | ENST00000233146 | 0.9558 | 0.02972 | 0.77588 | 0.98714 | 0.92438 | 0.7413052236 | 0.93008 | 0.9525725245 | 0.313933 | 7.911739371 | 0 |
| 2 | 47403385 | A | C | K | T | MSH2 | ENST00000233146 | 0.94933 | 0.03353 | 0.79088 | 0.98542 | 0.94167 | 0.709631266 | 0.85361 | 0.997512579 | 0.280473 | 5.866420699 | 0 |
| 2 | 47403385 | A | G | K | R | MSH2 | ENST00000233146 | 0.82709 | 0.01686 | 0.71006 | 0.95645 | 0.83615 | 0.6065966478 | 0.8904 | 0.9650547504 | 0.151104 | 1.581173999 | 0 |
| 2 | 47403385 | A | T | K | M | MSH2 | ENST00000233146 | 0.95315 | 0.0325 | 0.70947 | 0.9923 | 0.95608 | 0.7659307802 | 0.89767 | 0.9973827004 | 0.313692 | 7.911739371 | 0 |
| 2 | 47403387 | T | A | Y | N | MSH2 | ENST00000233146 | 0.43623 | 0.00504 | 0.72997 | 0.94066 | 0.80417 | 0.5177524471 | 0.89765 | 0.2844416797 | 0.146162 | 1.498767772 | 0 |
| 2 | 47403387 | T | C | Y | H | MSH2 | ENST00000233146 | 0.41275 | 0.00535 | 0.73824 | 0.92684 | 0.84052 | 0.4487534225 | 0.86828 | 0.3015730679 | 0.118906 | 1.815897181 | 0 |
| 2 | 47403388 | A | C | Y | S | MSH2 | ENST00000233146 | 0.42843 | 0.00664 | 0.65874 | 0.93432 | 0.83943 | 0.5478287719 | 0.88137 | 0.74791646 | 0.159315 | 2.0927012 | 0 |
| 2 | 47403388 | A | G | Y | C | MSH2 | ENST00000233146 | 0.62597 | 0.02094 | 0.74014 | 0.96762 | 0.87974 | 0.5300804286 | 0.87867 | 0.8269864917 | 0.232186 | 5.684101778 | 0 |
| 2 | 47403388 | A | T | Y | F | MSH2 | ENST00000233146 | 0.34282 | 0.00592 | 0.6397 | 0.88709 | 0.71391 | 0.2713351952 | 0.86592 | 0.3682352304 | 0.102999 | 3.733890575 | 0 |
| 2 | 47403390 | A | C | M | L | MSH2 | ENST00000233146 | 0.1891 | 0.0058 | 0.35223 | 0.90579 | 0.69168 | 0.2946262972 | 0.73722 | 0.06619669883 | -0.0165219 | -3.351368104 | 0 |

| | | | | | | | | | | | | | | | | | | |
|---|---|---|---|---|---|---|---|---|---|---|---|---|---|---|---|---|---|---|
| 2 | 47403390 | A | G | M | V | MSH2 | ENST00000233146 | 0.32289 | 0.00569 | 0.30234 | 0.84205 | 0.7285 | 0.3325861262 | 0.94817 | 0.239942044 | 0.0659819 | 1.083316468 | 0 |
| 2 | 47403390 | A | T | M | L | MSH2 | ENST00000233146 | 0.18599 | 0.0058 | 0.35223 | 0.90579 | 0.69168 | 0.2946262972 | 0.73659 | 0.0614983917 | -0.0164919 | -3.351368104 | 0 |
| 2 | 47403391 | T | A | M | K | MSH2 | ENST00000233146 | 0.59545 | 0.00574 | 0.44349 | 0.95458 | 0.90274 | 0.5642567209 | 0.89163 | 0.9757267833 | 0.213496 | -0.4150958545 | 0 |
| 2 | 47403391 | T | C | M | T | MSH2 | ENST00000233146 | 0.548 | 0.00608 | 0.49397 | 0.93395 | 0.8705 | 0.4043544094 | 0.87732 | 0.9248153567 | 0.165267 | 0.9740288473 | 0 |
| 2 | 47403391 | T | G | M | R | MSH2 | ENST00000233146 | 0.6215 | 0.00664 | 0.41848 | 0.966 | 0.89739 | 0.5982250293 | 0.83948 | 0.9757267833 | 0.175364 | 0.04926507288 | 0 |
| 2 | 47403392 | G | A | M | I | MSH2 | ENST00000233146 | 0.38158 | 0.00551 | 0.3996 | 0.69955 | 0.65077 | 0.3162248841 | 0.85148 | 0.1167502999 | 0.0457839 | 0.137947578 | 0 |
| 2 | 47403392 | G | C | M | I | MSH2 | ENST00000233146 | 0.3793 | 0.00551 | 0.3996 | 0.69891 | 0.65077 | 0.3162248841 | 0.85159 | 0.0819268251 | 0.0457839 | 0.137947578 | 0 |
| 2 | 47403392 | G | T | M | I | MSH2 | ENST00000233146 | 0.37465 | 0.00551 | 0.3996 | 0.70008 | 0.65077 | 0.3162248841 | 0.8494 | 0.1167502999 | 0.0457881 | 0.137947578 | 0 |
| 2 | 47403393 | G | A | G | R | MSH2 | ENST00000233146 | 0.83966 | 0.00893 | 0.65928 | 0.97458 | 0.9517 | 0.7363987417 | 0.95363 | 0.9977563024 | 0.463569 | 8.60259372 | 0 |
| 2 | 47403393 | G | C | G | R | MSH2 | ENST00000233146 | 0.83652 | 0.00893 | 0.65928 | 0.9739 | 0.9517 | 0.7363987417 | 0.95386 | 0.9976455569 | 0.463568 | 8.60259372 | 0 |
| 2 | 47403393 | G | T | G | W | MSH2 | ENST00000233146 | 0.95397 | 0.03452 | 0.63906 | 0.98004 | 0.96956 | 0.7249632472 | 0.93408 | 0.9994757771 | 0.466739 | 8.60259372 | 0 |
| 2 | 47403394 | G | A | G | E | MSH2 | ENST00000233146 | 0.91699 | 0.02796 | 0.62677 | 0.97995 | 0.95935 | 0.7543238914 | 0.9479 | 0.9975760579 | 0.489998 | 7.015453127 | 0 |
| 2 | 47403394 | G | C | G | A | MSH2 | ENST00000233146 | 0.75089 | 0.00928 | 0.55075 | 0.95957 | 0.91399 | 0.6878970757 | 0.9269 | 0.9961054921 | 0.337414 | 6.062900991 | 0 |
| 2 | 47403394 | G | T | G | V | MSH2 | ENST00000233146 | 0.92864 | 0.00877 | 0.56065 | 0.96833 | 0.9729 | 0.7504280933 | 0.94819 | 0.9976601601 | 0.498681 | 8.60259372 | 0 |
| 2 | 47403396 | C | A | P | T | MSH2 | ENST00000233146 | 0.41629 | 0.00551 | 0.36618 | 0.76931 | 0.81006 | 0.277959889 | 0.89239 | 0.2664223661 | 0.133995 | 0.7240014584 | 0 |
| 2 | 47403396 | C | G | P | A | MSH2 | ENST00000233146 | 0.36058 | 0.00574 | 0.34749 | 0.6701 | 0.64261 | 0.199111738 | 0.7836 | 0.206889829 | 0.0601035 | 0.6369465971 | 0 |
| 2 | 47403396 | C | T | P | S | MSH2 | ENST00000233146 | 0.40952 | 0.00569 | 0.33867 | 0.674 | 0.76289 | 0.2290087364 | 0.88816 | 0.1958748247 | 0.0826837 | 3.179521641 | 0 |
| 2 | 47403397 | C | A | P | Q | MSH2 | ENST00000233146 | 0.48133 | 0.00722 | 0.3492 | 0.85625 | 0.77076 | 0.2972676918 | 0.74912 | 0.6439916492 | 0.12372 | -1.20055007 | 0 |
| 2 | 47403397 | C | G | P | R | MSH2 | ENST00000233146 | 0.32146 | 0.00566 | 0.42087 | 0.81356 | 0.78658 | 0.2981259302 | 0.81456 | 0.3631125959 | 0.12257 | 0.7552703073 | 0 |
| 2 | 47403397 | C | T | P | L | MSH2 | ENST00000233146 | 0.38066 | 0.00566 | 0.42141 | 0.87904 | 0.7285 | 0.3107265231 | 0.87336 | 0.6927576065 | 0.0963326 | -1.415739358 | 0 |
| 2 | 47403399 | G | A | A | T | MSH2 | ENST00000233146 | 0.43346 | 0.0058 | 0.32135 | 0.82214 | 0.70508 | 0.283248641 | 0.93907 | 0.2470575273 | 0.0975596 | 0.4613234387 | 0 |
| 2 | 47403399 | G | C | A | P | MSH2 | ENST00000233146 | 0.46438 | 0.00527 | 0.37396 | 0.82784 | 0.69846 | 0.4340347068 | 0.90675 | 0.418292731 | 0.111674 | 2.175162079 | 0 |
| 2 | 47403399 | G | T | A | S | MSH2 | ENST00000233146 | 0.33057 | 0.00592 | 0.29275 | 0.65644 | 0.67478 | 0.2683403394 | 0.85565 | 0.147993724 | 0.0731845 | -0.1900805654 | 0 |
| 2 | 47403400 | C | A | A | E | MSH2 | ENST00000233146 | 0.43303 | 0.00603 | 0.37164 | 0.87604 | 0.77527 | 0.3520240377 | 0.85197 | 0.4373786462 | 0.113043 | 3.030729135 | 1 |
| 2 | 47403400 | C | G | A | G | MSH2 | ENST00000233146 | 0.48171 | 0.00569 | 0.30835 | 0.86638 | 0.69846 | 0.2598485274 | 0.89011 | 0.5920259356 | 0.10191 | 2.107918797 | 0 |
| 2 | 47403400 | C | T | A | V | MSH2 | ENST00000233146 | 0.51356 | 0.00633 | 0.33994 | 0.90342 | 0.74658 | 0.320115518 | 0.86316 | 0.8371209502 | 0.125787 | 3.603431757 | 0 |
| 2 | 47403402 | G | A | G | R | MSH2 | ENST00000233146 | 0.7813 | 0.00718 | 0.64988 | 0.97679 | 0.90798 | 0.4509761226 | 0.88738 | 0.9847072959 | 0.3041 | 6.341976404 | 0 |
| 2 | 47403402 | G | C | G | R | MSH2 | ENST00000233146 | 0.77806 | 0.00718 | 0.64988 | | 0.90798 | 0.4509761226 | 0.88824 | 0.9847072959 | 0.304097 | 6.341976404 | 0 |
| 2 | 47408401 | G | A | G | E | MSH2 | ENST00000233146 | 0.58824 | 0.00705 | 0.62576 | 0.96436 | 0.87148 | 0.4050526283 | 0.83594 | 0.7095633149 | 0.237858 | 5.604754304 | 0 |
| 2 | 47408401 | G | C | G | A | MSH2 | ENST00000233146 | 0.45364 | 0.01893 | 0.55148 | 0.97844 | 0.86703 | 0.3380737732 | 0.89184 | 0.9588387609 | 0.240077 | 4.707648261 | 0 |
| 2 | 47408401 | G | T | G | V | MSH2 | ENST00000233146 | 0.60142 | 0.02815 | 0.55905 | 0.97885 | 0.92192 | 0.4817685963 | 0.87968 | 0.9843608737 | 0.350014 | 5.921294104 | 0 |
| 2 | 47408403 | G | A | A | T | MSH2 | ENST00000233146 | 0.15034 | 0.0058 | 0.24395 | 0.42154 | 0.76421 | 0.2137996892 | 0.88103 | 0.03028977906 | 0.0410298 | 1.075670571 | 0 |
| 2 | 47408403 | G | C | A | P | MSH2 | ENST00000233146 | 0.15189 | 0.00527 | 0.29111 | 0.5775 | 0.81703 | 0.412455964 | 0.95914 | 0.04705915696 | 0.0724445 | 2.106058104 | 0 |
| 2 | 47408403 | G | T | A | S | MSH2 | ENST00000233146 | 0.09091 | 0.00586 | 0.23071 | 0.50329 | 0.68646 | 0.2002319566 | 0.65467 | 0.03910071403 | -0.00770141 | 0.09512183757 | 0 |
| 2 | 47408404 | C | A | A | E | MSH2 | ENST00000233146 | 0.21279 | 0.00547 | 0.29647 | 0.74133 | 0.74376 | 0.2676997247 | 0.83645 | 0.04253136739 | 0.0736857 | 1.003592292 | 0 |
| 2 | 47408404 | C | G | A | G | MSH2 | ENST00000233146 | 0.23579 | 0.00574 | 0.24625 | 0.66588 | 0.69509 | 0.2133861864 | 0.8797 | 0.05869097263 | 0.0654354 | 2.262659572 | 0 |
| 2 | 47408404 | C | T | A | V | MSH2 | ENST00000233146 | 0.23737 | 0.00726 | 0.27855 | 0.72585 | 0.72322 | 0.2434892408 | 0.90194 | 0.1252939999 | 0.0829385 | 3.877029486 | 0 |
| 2 | 47408406 | A | C | K | Q | MSH2 | ENST00000233146 | 0.38708 | 0.00615 | 0.48588 | 0.56837 | 0.73589 | 0.2866547274 | 0.9232 | 0.2750944197 | 0.0963788 | 1.344580888 | 0 |
| 2 | 47408406 | A | G | K | E | MSH2 | ENST00000233146 | 0.40634 | 0.00527 | 0.52781 | 0.59196 | 0.68292 | 0.302421887 | 0.85821 | 0.06619045872 | 0.0793943 | 2.200117339 | 0 |
| 2 | 47408407 | A | C | K | T | MSH2 | ENST00000233146 | 0.46326 | 0.01299 | 0.53328 | 0.73921 | 0.8199 | 0.3818534066 | 0.86695 | 0.3629845128 | 0.163213 | 3.769131151 | 0 |
| 2 | 47408407 | A | G | K | R | MSH2 | ENST00000233146 | 0.42746 | 0.00627 | 0.47089 | 0.61784 | 0.672 | 0.2295155656 | 0.86964 | 0.1712753701 | 0.077624 | 0.9252319844 | 0 |
| 2 | 47408407 | A | T | K | M | MSH2 | ENST00000233146 | 0.79737 | 0.02614 | 0.46938 | 0.82191 | 0.87636 | 0.4360801186 | 0.83637 | 0.9688019156 | 0.176746 | 6.176398572 | 0 |
| 2 | 47408408 | G | C | K | N | MSH2 | ENST00000233146 | 0.39562 | 0.00556 | 0.56105 | 0.64134 | 0.60835 | 0.3084877661 | 0.80535 | 0.1654658169 | 0.0571768 | 1.120842707 | 0 |
| 2 | 47408408 | G | T | K | N | MSH2 | ENST00000233146 | 0.39087 | 0.00556 | 0.56105 | 0.65021 | 0.61184 | 0.3084877661 | 0.80361 | 0.2392062396 | 0.0571886 | 1.120842707 | 0 |
| 2 | 47408409 | A | C | N | H | MSH2 | ENST00000233146 | 0.46047 | 0.01002 | 0.35419 | 0.69741 | 0.62547 | 0.2581817916 | 0.82815 | 0.1214233896 | 0.0327468 | 0.2364155279 | 0 |
| 2 | 47408409 | A | G | N | D | MSH2 | ENST00000233146 | 0.3772 | 0.0058 | 0.38727 | 0.65013 | 0.57877 | 0.190333213 | 0.8038 | 0.1916856229 | 0.0280769 | -1.576899501 | 0 |
| 2 | 47408409 | A | T | N | Y | MSH2 | ENST00000233146 | 0.50075 | 0.00832 | 0.38354 | 0.77 | 0.77141 | 0.2707658455 | 0.94199 | 0.3503177839 | 0.12921 | 0.8352526646 | 0 |
| 2 | 47408410 | A | C | N | T | MSH2 | ENST00000233146 | 0.21279 | 0.00566 | 0.35459 | 0.51906 | 0.60362 | 0.2152324965 | 0.66014 | 0.04346035454 | 0.00778977 | -2.105845164 | 0 |
| 2 | 47408410 | A | G | N | S | MSH2 | ENST00000233146 | 0.22325 | 0.00615 | 0.30865 | 0.70078 | 0.53187 | 0.1607826938 | 0.74069 | 0.0485712595 | 0.00559474 | -0.6882646789 | 0 |
| 2 | 47408410 | A | T | N | I | MSH2 | ENST00000233146 | 0.38392 | 0.00608 | 0.39889 | 0.75716 | 0.75687 | 0.3153818351 | 0.94879 | 0.09374661997 | 0.106172 | 1.768642244 | 0 |
| 2 | 47408411 | T | A | N | K | MSH2 | ENST00000233146 | 0.22852 | 0.00543 | 0.38458 | 0.73291 | 0.58391 | 0.178268272 | 0.59687 | 0.04963695231 | -0.0495746 | -5.745481092 | 0 |
| 2 | 47408411 | T | G | N | K | MSH2 | ENST00000233146 | 0.23284 | 0.00543 | 0.38458 | 0.73425 | 0.58391 | 0.178268272 | 0.59149 | 0.05234857173 | -0.0495809 | -5.745481092 | 0 |
| 2 | 47408412 | C | G | L | V | MSH2 | ENST00000233146 | 0.36592 | 0.01376 | 0.57455 | 0.66795 | 0.80417 | 0.4778736245 | 0.85382 | 0.6475411654 | 0.125316 | 2.105838215 | 0 |
| 2 | 47408413 | T | A | L | Q | MSH2 | ENST00000233146 | 0.82693 | 0.03595 | 0.65267 | 0.90688 | 0.96044 | 0.8013584464 | 0.96444 | 0.9838633537 | 0.463514 | 7.358500373 | 0 |
| 2 | 47408413 | T | C | L | P | MSH2 | ENST00000233146 | 0.84226 | 0.0296 | 0.7672 | 0.90261 | 0.96153 | 0.8536156388 | 0.98124 | 0.9690563083 | 0.476841 | 5.74982942 | 0 |
| 2 | 47408413 | T | G | L | R | MSH2 | ENST00000233146 | 0.83244 | 0.03099 | 0.72509 | 0.90927 | 0.95863 | 0.8495150379 | 0.9733 | 0.9872281551 | 0.48113 | 6.407868535 | 0 |
| 2 | 47408415 | C | A | Q | K | MSH2 | ENST00000233146 | 0.47905 | 0.00556 | 0.36181 | 0.73106 | 0.71627 | 0.4680435433 | 0.91478 | 0.7712494731 | 0.0941817 | 1.530561113 | 0 |
| 2 | 47408416 | A | C | Q | P | MSH2 | ENST00000233146 | 0.52074 | 0.00535 | 0.2926 | 0.55255 | 0.77591 | 0.5741779388 | 0.94485 | 0.1636527507 | 0.0755866 | -3.17919821 | 0 |
| 2 | 47408416 | A | G | Q | R | MSH2 | ENST00000233146 | 0.65806 | 0.00638 | 0.33527 | 0.76854 | 0.78844 | 0.4181099376 | 0.90367 | 0.7293316722 | 0.135404 | 1.530561113 | 0 |
| 2 | 47408416 | A | T | Q | L | MSH2 | ENST00000233146 | 0.58552 | 0.0058 | 0.2997 | 0.81367 | 0.83505 | 0.4985783886 | 0.92688 | 0.8141876459 | 0.162264 | 1.530561113 | 0 |
| 2 | 47408417 | G | C | Q | H | MSH2 | ENST00000233146 | 0.48685 | 0.00586 | 0.31952 | 0.83812 | 0.80476 | 0.5206893818 | 0.83886 | 0.2421374008 | 0.0954243 | 0.3910597239 | 0 |
| 2 | 47408417 | G | T | Q | H | MSH2 | ENST00000233146 | 0.48171 | 0.00586 | 0.31952 | 0.83929 | 0.80476 | 0.5206893818 | 0.83739 | 0.1452645804 | 0.0846358 | 0.3910597239 | 0 |
| 2 | 47408418 | A | C | S | R | MSH2 | ENST00000233146 | 0.66232 | 0.02442 | 0.72314 | 0.82484 | 0.93358 | 0.7271613108 | 0.96809 | 0.9743224978 | 0.451678 | 7.112933481 | 0 |
| 2 | 47408418 | A | G | S | G | MSH2 | ENST00000233146 | 0.47608 | 0.00705 | 0.6138 | 0.53753 | 0.85378 | 0.5790160337 | 0.94161 | 0.7266904116 | 0.248197 | 6.610762128 | 0 |
| 2 | 47408418 | A | T | S | C | MSH2 | ENST00000233146 | 0.83767 | 0.03189 | 0.67333 | 0.79544 | 0.92884 | 0.5827252067 | 0.9225 | 0.9716259837 | 0.361647 | 7.112933481 | 0 |
| 2 | 47408419 | G | A | S | N | MSH2 | ENST00000233146 | 0.46525 | 0.01977 | 0.71453 | 0.67028 | 0.84641 | 0.5820787806 | 0.89415 | 0.9759984612 | 0.166688 | 7.112933481 | 0 |
| 2 | 47408419 | G | C | S | T | MSH2 | ENST00000233146 | 0.42562 | 0.00633 | 0.66471 | 0.62409 | 0.78035 | 0.4973755851 | 0.89844 | 0.7044343352 | 0.152732 | 6.969629105 | 0 |
| 2 | 47408419 | G | T | S | I | MSH2 | ENST00000233146 | 0.61268 | 0.02133 | 0.7032 | 0.82015 | 0.91526 | 0.6701436922 | 0.97774 | 0.8976737994 | 0.334947 | 6.891919183 | 0 |
| 2 | 47408420 | T | A | S | R | MSH2 | ENST00000233146 | 0.59581 | 0.02442 | 0.72314 | 0.81366 | 0.93162 | 0.7271613108 | 0.98783 | 0.971922636 | 0.435654 | 7.112933481 | 0 |
| 2 | 47408420 | T | G | S | R | MSH2 | ENST00000233146 | 0.60332 | 0.02442 | 0.72314 | 0.80959 | 0.93162 | 0.7271613108 | 0.98775 | 0.971922636 | 0.435657 | 7.112933481 | 0 |
| 2 | 47408421 | G | A | V | I | MSH2 | ENST00000233146 | 0.72804 | 0.03353 | 0.56722 | 0.74653 | 0.88637 | 0.3405432227 | 0.82909 | 0.4142953492 | -0.0273273 | 8.174235504 | 0 |
| 2 | 47408421 | G | C | V | L | MSH2 | ENST00000233146 | 0.6191 | 0.03475 | 0.7558 | 0.86217 | 0.95207 | 0.6092011222 | 0.94971 | 0.9628771544 | 0.402756 | 8.174235504 | 0 |
| 2 | 47408422 | T | A | V | D | MSH2 | ENST00000233146 | 0.69772 | 0.02989 | 0.78267 | 0.9231 | 0.98657 | 0.8967561172 | 0.97933 | 0.9977839589 | 0.490296 | 8.174235504 | 1 |
| 2 | 47408422 | T | C | V | A | MSH2 | ENST00000233146 | 0.65023 | 0.03353 | 0.77744 | 0.81702 | 0.95243 | 0.6256484195 | 0.92766 | 0.9811228514 | 0.454227 | 3.51205744 | 0 |
| 2 | 47408422 | T | G | V | G | MSH2 | ENST00000233146 | 0.68259 | 0.03304 | 0.63572 | 0.92156 | 0.97885 | 0.8583223167 | 0.97147 | 0.9980739355 | 0.489881 | 8.174235504 | 0 |
| 2 | 47408424 | G | A | V | M | MSH2 | ENST00000233146 | 0.73245 | 0.03205 | 0.80978 | 0.77614 | 0.90842 | 0.524957108 | 0.85998 | 0.9662240148 | 0.256036 | 5.777102534 | 0 |
| 2 | 47408424 | G | C | V | L | MSH2 | ENST00000233146 | 0.42211 | 0.01115 | 0.76907 | 0.73916 | 0.86098 | 0.5029134414 | 0.90957 | 0.9591729045 | 0.257524 | 5.777102534 | 0 |
| 2 | 47408424 | G | T | V | L | MSH2 | ENST00000233146 | 0.4173 | 0.01115 | 0.76907 | 0.73992 | 0.86098 | 0.5029134414 | 0.90959 | 0.9591729045 | 0.257523 | 5.777102534 | 0 |
| 2 | 47408425 | T | A | V | E | MSH2 | ENST00000233146 | 0.62741 | 0.02283 | 0.77654 | 0.88052 | 0.94167 | 0.6916459507 | 0.94908 | 0.9763842225 | 0.344717 | 5.777102534 | 0 |
| 2 | 47408425 | T | C | V | A | MSH2 | ENST00000233146 | 0.31873 | 0.0058 | 0.7952 | 0.65601 | 0.79212 | 0.4731507931 | 0.90902 | 0.6449151039 | 0.170785 | 2.757110947 | 0 |
| 2 | 47408425 | T | G | V | G | MSH2 | ENST00000233146 | 0.48799 | 0.01731 | 0.64054 | 0.86219 | 0.92642 | 0.6861702293 | 0.93611 | 0.9830537438 | 0.354881 | 2.203778311 | 0 |
| 2 | 47408427 | C | A | L | I | MSH2 | ENST00000233146 | 0.60069 | 0.01915 | 0.85823 | 0.79664 | 0.86951 | 0.3933860283 | 0.82273 | 0.9170678854 | 0.222974 | 3.586535579 | 0 |
| 2 | 47408427 | C | G | L | V | MSH2 | ENST00000233146 | 0.47728 | 0.00769 | 0.80832 | 0.77957 | 0.88684 | 0.5160972927 | 0.88253 | 0.9244509935 | 0.243608 | 2.71157396 | 0 |
| 2 | 47408427 | C | T | L | F | MSH2 | ENST00000233146 | 0.61633 | 0.00741 | 0.85506 | 0.82331 | 0.9211 | 0.664623394 | 0.88788 | 0.9724954963 | 0.329619 | 4.646085822 | 0 |
| 2 | 47408428 | T | C | L | P | MSH2 | ENST00000233146 | 0.73909 | 0.0292 | 0.94613 | 0.92835 | 0.98869 | 0.9038857673 | 0.97854 | 0.9969622493 | 0.484275 | 8.211219011 | 1 |
| 2 | 47408428 | T | G | L | R | MSH2 | ENST00000233146 | 0.71914 | 0.03059 | 0.92938 | 0.9271 | 0.98146 | 0.8960140925 | 0.98234 | 0.9981808662 | 0.489173 | 8.211219011 | 0 |
| 2 | 47408430 | A | C | S | R | MSH2 | ENST00000233146 | 0.72108 | 0.03388 | 0.89253 | 0.92244 | 0.94839 | 0.7919945011 | 0.98975 | 0.9776027799 | 0.471736 | 7.949742944 | 0 |
| 2 | 47408430 | A | G | S | G | MSH2 | ENST00000233146 | 0.72692 | 0.03538 | 0.80519 | 0.91002 | 0.93162 | 0.6585218893 | 0.95279 | 0.9880706072 | 0.314557 | 7.80620812 | 0 |
| 2 | 47408430 | A | T | S | C | MSH2 | ENST00000233146 | 0.77545 | 0.03728 | 0.8516 | 0.92889 | 0.95608 | 0.6650285929 | 0.9586 | 0.9757661819 | 0.459675 | 7.949742944 | 0 |
| 2 | 47408431 | G | A | S | N | MSH2 | ENST00000233146 | 0.42323 | 0.015 | 0.88423 | 0.82288 | 0.83394 | 0.6495847496 | 0.82918 | 0.9090102315 | 0.161541 | 7.566035091 | 0 |
| 2 | 47408431 | G | C | S | T | MSH2 | ENST00000233146 | 0.74994 | 0.03626 | 0.847 | 0.82516 | 0.90798 | 0.6222521519 | 0.92602 | 0.8967764974 | 0.313459 | 7.949742944 | 0 |
| 2 | 47408431 | G | T | S | I | MSH2 | ENST00000233146 | 0.82887 | 0.02427 | 0.87256 | 0.88226 | 0.95681 | 0.7739096212 | 0.91409 | 0.9869421124 | 0.475974 | 7.949742944 | 0 |
| 2 | 47408432 | T | A | S | R | MSH2 | ENST00000233146 | 0.54338 | 0.03388 | 0.89253 | 0.94483 | 0.94167 | 0.7919945011 | 0.98935 | 0.9706539512 | 0.418667 | 7.949742944 | 0 |
| 2 | 47408432 | T | G | S | R | MSH2 | ENST00000233146 | 0.55039 | 0.03388 | 0.89253 | 0.94322 | 0.94167 | 0.7919945011 | 0.98931 | 0.9706539512 | 0.418669 | 7.949742944 | 0 |
| 2 | 47408433 | A | C | K | Q | MSH2 | ENST00000233146 | 0.60273 | 0.0143 | 0.88467 | 0.84335 | 0.87491 | 0.5975301154 | 0.95243 | 0.6418251395 | 0.277622 | 8.161368328 | 0 |
| 2 | 47408433 | A | G | K | E | MSH2 | ENST00000233146 | 0.79176 | 0.02206 | 0.91812 | 0.883 | 0.90885 | 0.6782956764 | 0.97585 | 0.6547268629 | 0.404384 | 7.115925289 | 0 |
| 2 | 47408434 | A | C | K | T | MSH2 | ENST00000233146 | 0.77758 | 0.03129 | 0.92291 | 0.90841 | 0.90755 | 0.6700039162 | 0.92019 | 0.9515430927 | 0.297084 | 8.161368328 | 0 |
| 2 | 47408434 | A | G | K | R | MSH2 | ENST00000233146 | 0.62688 | 0.00844 | 0.859 | 0.68981 | 0.8859 | 0.5316684492 | 0.9473 | 0.7874792814 | 0.268942 | 5.771992841 | 0 |
| 2 | 47408434 | A | T | K | I | MSH2 | ENST00000233146 | 0.82568 | 0.02796 | 0.89948 | 0.91849 | 0.92069 | 0.693808127 | 0.93421 | 0.9874404669 | 0.319671 | 8.161368328 | 0 |
| 2 | 47408435 | A | C | K | N | MSH2 | ENST00000233146 | 0.72148 | 0.01058 | 0.93376 | 0.71782 | 0.84747 | 0.6622285974 | 0.86739 | 0.7433065176 | 0.119843 | 8.161368328 | 0 |
| 2 | 47408435 | A | T | K | N | MSH2 | ENST00000233146 | 0.71466 | 0.01058 | 0.93376 | 0.73461 | 0.84747 | 0.6622285974 | 0.86717 | 0.7433065176 | 0.119838 | 8.161368328 | 0 |

| | | | | | | | | | | | | | | | | | | |
|---|---|---|---|---|---|---|---|---|---|---|---|---|---|---|---|---|---|---|
| 2 | 47408436 | A | C | M | L | MSH2 | ENST00000233146 | 0.42217 | 0.00592 | 0.8737 | 0.59403 | 0.7255 | 0.3847405178 | 0.87951 | 0.5137153268 | 0.109803 | 2.658150399 | 0 |
| 2 | 47408436 | A | G | M | V | MSH2 | ENST00000233146 | 0.40301 | 0.00983 | 0.81694 | 0.66444 | 0.85741 | 0.4470290136 | 0.94461 | 0.6897813082 | 0.251039 | 6.869695219 | 0 |
| 2 | 47408436 | A | T | M | L | MSH2 | ENST00000233146 | 0.41712 | 0.00592 | 0.8737 | 0.59403 | 0.7255 | 0.3847405178 | 0.88 | 0.8037725091 | 0.109808 | 2.658150399 | 0 |
| 2 | 47408437 | T | A | M | K | MSH2 | ENST00000233146 | 0.49686 | 0.01002 | 0.92765 | 0.78092 | 0.88069 | 0.6258267819 | 0.96227 | 0.8270785809 | 0.355179 | 4.211227793 | 0 |
| 2 | 47408437 | T | C | M | T | MSH2 | ENST00000233146 | 0.36398 | 0.01115 | 0.94759 | 0.65185 | 0.83228 | 0.4939479195 | 0.88843 | 0.7734621167 | 0.21078 | 6.74045667 | 0 |
| 2 | 47408437 | T | G | M | R | MSH2 | ENST00000233146 | 0.53711 | 0.01834 | 0.91267 | 0.82387 | 0.91484 | 0.6659275936 | 0.96842 | 0.9245921373 | 0.387759 | 4.526863731 | 0 |
| 2 | 47408439 | A | C | N | H | MSH2 | ENST00000233146 | 0.42651 | 0.01295 | 0.86429 | 0.74783 | 0.86753 | 0.6620989706 | 0.92376 | 0.8230873942 | 0.156673 | 8.599953301 | 0 |
| 2 | 47408439 | A | T | N | Y | MSH2 | ENST00000233146 | 0.63804 | 0.03137 | 0.88647 | 0.86523 | 0.94802 | 0.7190241305 | 0.96401 | 0.9535182118 | 0.457052 | 8.599953301 | 0 |
| 2 | 47408440 | A | C | N | T | MSH2 | ENST00000233146 | 0.60142 | 0.02951 | 0.8435 | 0.85009 | 0.87099 | 0.6079151554 | 0.9043 | 0.9702091813 | 0.19459 | 6.910462516 | 0 |
| 2 | 47408440 | A | G | N | S | MSH2 | ENST00000233146 | 0.55428 | 0.01881 | 0.8004 | 0.78727 | 0.85894 | 0.5555848114 | 0.92207 | 0.8982374668 | 0.120796 | 3.292497932 | 0 |
| 2 | 47408440 | A | T | N | I | MSH2 | ENST00000233146 | 0.77188 | 0.02745 | 0.84439 | 0.85908 | 0.93786 | 0.6938695458 | 0.9271 | 0.9859128594 | 0.359519 | 8.599953301 | 0 |
| 2 | 47408441 | T | A | N | K | MSH2 | ENST00000233146 | 0.44181 | 0.01365 | 0.91433 | 0.85057 | 0.80595 | 0.7011661422 | 0.9266 | 0.8565995097 | 0.11382 | 8.599953301 | 0 |
| 2 | 47408441 | T | G | N | K | MSH2 | ENST00000233146 | 0.44807 | 0.01365 | 0.91433 | 0.85379 | 0.80595 | 0.7011661422 | 0.92684 | 0.8512189388 | 0.113822 | 8.599953301 | 0 |
| 2 | 47408442 | T | A | F | I | MSH2 | ENST00000233146 | 0.79603 | 0.03129 | 0.88868 | 0.88898 | 0.9248 | 0.784955726 | 0.92902 | 0.9969123602 | 0.375643 | 5.170127613 | 0 |
| 2 | 47408442 | T | C | F | L | MSH2 | ENST00000233146 | 0.81559 | 0.03189 | 0.90653 | 0.87498 | 0.91736 | 0.7322867797 | 0.92159 | 0.991040647 | 0.369463 | 5.087809897 | 0 |
| 2 | 47408442 | T | G | F | V | MSH2 | ENST00000233146 | 0.80242 | 0.03212 | 0.8648 | 0.908 | 0.93123 | 0.7204455078 | 0.9515 | 0.9960711002 | 0.437887 | 4.124465374 | 0 |
| 2 | 47408443 | T | A | F | Y | MSH2 | ENST00000233146 | 0.76235 | 0.01812 | 0.89355 | 0.81254 | 0.8887 | 0.6242242329 | 0.86045 | 0.8955189586 | 0.240512 | 5.586562708 | 0 |
| 2 | 47408443 | T | C | F | S | MSH2 | ENST00000233146 | 0.86318 | 0.02702 | 0.87713 | 0.93186 | 0.93201 | 0.7920886812 | 0.94482 | 0.998249948 | 0.456697 | 4.547970498 | 0 |
| 2 | 47408443 | T | G | F | C | MSH2 | ENST00000233146 | 0.86133 | 0.03144 | 0.90305 | 0.92718 | 0.9328 | 0.7616045134 | 0.94089 | 0.997107923 | 0.427234 | 5.062925929 | 0 |
| 2 | 47408444 | T | A | F | L | MSH2 | ENST00000233146 | 0.63443 | 0.03189 | 0.90653 | 0.87968 | 0.86703 | 0.7322867797 | 0.92755 | 0.9886955023 | 0.342591 | 5.087809897 | 0 |
| 2 | 47408444 | T | G | F | L | MSH2 | ENST00000233146 | 0.64229 | 0.03189 | 0.90653 | 0.87959 | 0.86703 | 0.7322867797 | 0.92742 | 0.9867823124 | 0.342591 | 5.087809897 | 0 |
| 2 | 47408445 | G | C | E | Q | MSH2 | ENST00000233146 | 0.80282 | 0.03735 | 0.80724 | 0.77974 | 0.8873 | 0.6212326409 | 0.93011 | 0.8921951652 | 0.25596 | 5.633643125 | 0 |
| 2 | 47408446 | A | C | E | A | MSH2 | ENST00000233146 | 0.79856 | 0.03626 | 0.82729 | 0.87759 | 0.93475 | 0.6837203623 | 0.96658 | 0.9953556061 | 0.473168 | 5.896706481 | 0 |
| 2 | 47408446 | A | G | E | G | MSH2 | ENST00000233146 | 0.84491 | 0.03484 | 0.77224 | 0.88674 | 0.9458 | 0.6898491887 | 0.95318 | 0.9959017634 | 0.477628 | 8.243298186 | 0 |
| 2 | 47408446 | A | T | E | V | MSH2 | ENST00000233146 | 0.82732 | 0.03484 | 0.79704 | 0.89688 | 0.95023 | 0.6838898975 | 0.97616 | 0.9976961017 | 0.491257 | 9.10657765 | 0 |
| 2 | 47408447 | A | C | E | D | MSH2 | ENST00000233146 | 0.47501 | 0.03864 | 0.82345 | 0.84986 | 0.86553 | 0.5908038464 | 0.95128 | 0.9930254221 | 0.325328 | 4.054950664 | 0 |
| 2 | 47408447 | A | T | E | D | MSH2 | ENST00000233146 | 0.46974 | 0.03864 | 0.82345 | 0.85138 | 0.86553 | 0.5908038464 | 0.95107 | 0.9928941131 | 0.325324 | 4.054950664 | 0 |
| 2 | 47408448 | T | A | S | T | MSH2 | ENST00000233146 | 0.38381 | 0.00832 | 0.69552 | 0.65077 | 0.79878 | 0.414035722 | 0.93157 | 0.4280016422 | 0.165254 | 4.300693998 | 0 |
| 2 | 47408448 | T | C | S | P | MSH2 | ENST00000233146 | 0.81857 | 0.02745 | 0.72093 | 0.89021 | 0.87588 | 0.7750708912 | 0.93683 | 0.9862182736 | 0.24753 | 5.455899853 | 0 |
| 2 | 47408448 | T | G | S | A | MSH2 | ENST00000233146 | 0.4372 | 0.00654 | 0.67693 | 0.64684 | 0.71312 | 0.31959467 | 0.88718 | 0.3812515736 | 0.0971457 | 2.845681403 | 0 |
| 2 | 47408449 | C | A | S | Y | MSH2 | ENST00000233146 | 0.55656 | 0.029 | 0.76753 | 0.85653 | 0.87781 | 0.5842231097 | 0.8738 | 0.9737019539 | 0.16135 | 7.250201827 | 0 |
| 2 | 47408449 | C | G | S | C | MSH2 | ENST00000233146 | 0.58503 | 0.00696 | 0.70842 | 0.82014 | 0.87345 | 0.4563441407 | 0.83441 | 0.7218225002 | 0.258763 | 8.719844758 | 0 |
| 2 | 47408449 | C | T | S | F | MSH2 | ENST00000233146 | 0.59958 | 0.02914 | 0.74381 | 0.85587 | 0.89422 | 0.5487093332 | 0.8802 | 0.9928357005 | 0.283594 | 7.151322859 | 0 |
| 2 | 47408451 | T | A | F | I | MSH2 | ENST00000233146 | 0.50419 | 0.00608 | 0.87243 | 0.76442 | 0.84959 | 0.6301460944 | 0.96074 | 0.7006846666 | 0.249247 | 5.494298762 | 0 |
| 2 | 47408451 | T | C | F | L | MSH2 | ENST00000233146 | 0.47306 | 0.00547 | 0.90505 | 0.65261 | 0.80476 | 0.5780121528 | 0.97257 | 0.5367742181 | 0.238958 | 1.695235001 | 0 |
| 2 | 47408451 | T | G | F | V | MSH2 | ENST00000233146 | 0.49906 | 0.0054 | 0.86556 | 0.78502 | 0.8278 | 0.5655861215 | 0.88126 | 0.7820414901 | 0.15677 | 2.286543797 | 0 |
| 2 | 47408452 | T | A | F | Y | MSH2 | ENST00000233146 | 0.64345 | 0.01309 | 0.89283 | 0.75269 | 0.86553 | 0.5313086396 | 0.85453 | 0.9731833339 | 0.192298 | 1.992194847 | 0 |
| 2 | 47408452 | T | C | F | S | MSH2 | ENST00000233146 | 0.87958 | 0.01108 | 0.87563 | 0.89992 | 0.95316 | 0.7293040976 | 0.97886 | 0.9937217832 | 0.42287 | 5.494298762 | 0 |
| 2 | 47408452 | T | G | F | C | MSH2 | ENST00000233146 | 0.91073 | 0.02702 | 0.90294 | 0.91184 | 0.97215 | 0.6607751611 | 0.96036 | 0.9955898523 | 0.430469 | 5.494298762 | 0 |
| 2 | 47408453 | T | A | F | L | MSH2 | ENST00000233146 | 0.3124 | 0.00547 | 0.90505 | 0.56849 | 0.81065 | 0.5780121528 | 0.97255 | 0.2449268401 | 0.213873 | 1.695235001 | 0 |
| 2 | 47408453 | T | G | F | L | MSH2 | ENST00000233146 | 0.31774 | 0.00547 | 0.90505 | 0.57866 | 0.81065 | 0.5780121528 | 0.97221 | 0.2449268401 | 0.213877 | 1.695235001 | 0 |
| 2 | 47408454 | G | A | V | I | MSH2 | ENST00000233146 | 0.38868 | 0.00705 | 0.58926 | 0.60158 | 0.79212 | 0.3170314949 | 0.91335 | 0.6234827042 | 0.159375 | 2.441594924 | 0 |
| 2 | 47408454 | G | C | V | L | MSH2 | ENST00000233146 | 0.41005 | 0.00603 | 0.76691 | 0.62993 | 0.78906 | 0.5227695241 | 0.95351 | 0.4198025167 | 0.170364 | 1.843423805 | 0 |
| 2 | 47408454 | G | T | V | L | MSH2 | ENST00000233146 | 0.40529 | 0.00603 | 0.76691 | 0.6292 | 0.78906 | 0.5227695241 | 0.95361 | 0.4198025167 | 0.170359 | 1.843423805 | 0 |
| 2 | 47408455 | T | A | V | E | MSH2 | ENST00000233146 | 0.6218 | 0.0167 | 0.77326 | 0.91231 | 0.96153 | 0.8085493639 | 0.982 | 0.9849246144 | 0.428984 | 6.92913522 | 0 |
| 2 | 47408455 | T | C | V | A | MSH2 | ENST00000233146 | 0.3034 | 0.00586 | 0.78456 | 0.72084 | 0.82216 | 0.5584974265 | 0.9399 | 0.1160983282 | 0.028727 | 3.439448791 | 0 |
| 2 | 47408455 | T | G | V | G | MSH2 | ENST00000233146 | 0.60544 | 0.00592 | 0.64211 | 0.84803 | 0.94765 | 0.7195114048 | 0.98964 | 0.9768890738 | 0.446054 | 6.92913522 | 0 |
| 2 | 47408457 | A | C | K | Q | MSH2 | ENST00000233146 | 0.65956 | 0.02633 | 0.63766 | 0.85243 | 0.85482 | 0.5074308777 | 0.91676 | 0.9465907812 | 0.210687 | 4.166569232 | 0 |
| 2 | 47408458 | A | C | K | T | MSH2 | ENST00000233146 | 0.72628 | 0.02708 | 0.72098 | 0.88759 | 0.90755 | 0.5785116145 | 0.91038 | 0.9869427681 | 0.25983 | 3.29997813 | 0 |
| 2 | 47408458 | A | G | K | R | MSH2 | ENST00000233146 | 0.31234 | 0.00627 | 0.60831 | 0.58716 | 0.67752 | 0.407937784 | 0.84997 | 0.7259396315 | 0.0671752 | -0.5193736961 | 0 |
| 2 | 47408458 | A | T | K | I | MSH2 | ENST00000233146 | 0.7798 | 0.02614 | 0.67332 | 0.9043 | 0.89376 | 0.6095007799 | 0.93401 | 0.9931803942 | 0.271431 | 5.935521405 | 0 |
| 2 | 47408459 | A | C | K | N | MSH2 | ENST00000233146 | 0.76005 | 0.02732 | 0.74333 | 0.8424 | 0.84321 | 0.5648584738 | 0.89901 | 0.9716168046 | 0.141227 | 3.293068267 | 0 |
| 2 | 47408459 | A | T | K | N | MSH2 | ENST00000233146 | 0.75328 | 0.02732 | 0.74333 | 0.84581 | 0.84321 | 0.5648584738 | 0.89831 | 0.9716168046 | 0.141222 | 3.293068267 | 0 |
| 2 | 47408460 | G | A | D | N | MSH2 | ENST00000233146 | 0.66035 | 0.02356 | 0.65937 | 0.79699 | 0.84213 | 0.5984481614 | 0.9325 | 0.9582623839 | 0.129193 | 3.610501389 | 0 |
| 2 | 47408460 | G | C | D | H | MSH2 | ENST00000233146 | 0.66383 | 0.03124 | 0.69207 | 0.86903 | 0.89829 | 0.6758821924 | 0.92685 | 0.992323041 | 0.289668 | 5.523317004 | 0 |
| 2 | 47408460 | G | T | D | Y | MSH2 | ENST00000233146 | 0.67326 | 0.0307 | 0.67228 | 0.89968 | 0.89055 | 0.7267824972 | 0.95727 | 0.9937093258 | 0.310359 | 7.595859837 | 0 |
| 2 | 47408461 | A | C | D | A | MSH2 | ENST00000233146 | 0.80037 | 0.01459 | 0.58285 | 0.83425 | 0.81876 | 0.6793183547 | 0.88786 | 0.9460466504 | 0.234103 | 5.206155515 | 0 |
| 2 | 47408461 | A | G | D | G | MSH2 | ENST00000233146 | 0.61723 | 0.00547 | 0.58858 | 0.81737 | 0.83997 | 0.6745736825 | 0.92925 | 0.9567371011 | 0.243363 | 6.283622333 | 0 |
| 2 | 47408461 | A | T | D | V | MSH2 | ENST00000233146 | 0.80606 | 0.02288 | 0.57369 | 0.90601 | 0.85378 | 0.7105146227 | 0.95471 | 0.9886050224 | 0.324766 | 4.646335931 | 0 |
| 2 | 47408463 | C | A | L | I | MSH2 | ENST00000233146 | 0.55992 | 0.03293 | 0.77297 | 0.778 | 0.85741 | 0.4181380719 | 0.79861 | 0.8845018744 | 0.288039 | 3.625756475 | 0 |
| 2 | 47408463 | C | G | L | V | MSH2 | ENST00000233146 | 0.54984 | 0.02396 | 0.6991 | 0.81538 | 0.91442 | 0.5690204936 | 0.94656 | 0.3463362157 | 0.245836 | 9.047081295 | 0 |
| 2 | 47408463 | C | T | L | F | MSH2 | ENST00000233146 | 0.59312 | 0.03534 | 0.74685 | 0.87764 | 0.94356 | 0.6183785802 | 0.92395 | 0.9936377406 | 0.48242 | 9.047081295 | 0 |
| 2 | 47408464 | T | A | L | H | MSH2 | ENST00000233146 | 0.68715 | 0.03287 | 0.8495 | 0.93281 | 0.9844 | 0.7640570795 | 0.97846 | 0.9838508964 | 0.422838 | 9.047081295 | 0 |
| 2 | 47408464 | T | C | L | P | MSH2 | ENST00000233146 | 0.71193 | 0.02972 | 0.88456 | 0.92638 | 0.98764 | 0.899810326 | 0.97724 | 0.9971725345 | 0.535402 | 9.047081295 | 0 |
| 2 | 47408464 | T | G | L | R | MSH2 | ENST00000233146 | 0.70567 | 0.01597 | 0.84543 | 0.93081 | 0.98938 | 0.8838404154 | 0.97862 | 0.9977455735 | 0.535065 | 9.047081295 | 0 |
| 2 | 47408466 | C | A | L | I | MSH2 | ENST00000233146 | 0.60588 | 0.03518 | 0.8212 | 0.84423 | 0.88117 | 0.5038224536 | 0.81606 | 0.9876240492 | 0.239354 | 8.505354554 | 0 |
| 2 | 47408466 | C | G | L | V | MSH2 | ENST00000233146 | 0.60186 | 0.03582 | 0.7629 | 0.83769 | 0.91399 | 0.6703392684 | 0.86589 | 0.9891343713 | 0.305117 | 9.625784115 | 0 |
| 2 | 47408466 | C | T | L | F | MSH2 | ENST00000233146 | 0.62924 | 0.03557 | 0.80535 | 0.86388 | 0.96153 | 0.7257703675 | 0.94647 | 0.9926452637 | 0.479105 | 7.418358679 | 0 |
| 2 | 47408467 | T | A | L | H | MSH2 | ENST00000233146 | 0.71586 | 0.03327 | 0.8915 | 0.92887 | 0.98799 | 0.8417864166 | 0.95897 | 0.9975875616 | 0.500548 | 9.625784115 | 0 |
| 2 | 47408467 | T | C | L | P | MSH2 | ENST00000233146 | 0.74069 | 0.02972 | 0.91475 | 0.93023 | 0.98728 | 0.9313057422 | 0.9711 | 0.9980739355 | 0.500539 | 9.625784115 | 0 |
| 2 | 47408467 | T | G | L | R | MSH2 | ENST00000233146 | 0.73613 | 0.03099 | 0.89216 | 0.92899 | 0.99039 | 0.9278359648 | 0.95857 | 0.9982530475 | 0.50027 | 9.625784115 | 0 |
| 2 | 47408469 | C | A | L | M | MSH2 | ENST00000233146 | 0.52241 | 0.03767 | 0.74439 | 0.82225 | 0.85843 | 0.4909364973 | 0.88318 | 0.9757474065 | 0.145814 | 8.750603671 | 0 |
| 2 | 47408469 | C | G | L | V | MSH2 | ENST00000233146 | 0.49886 | 0.03706 | 0.70654 | 0.85721 | 0.86453 | 0.5630863207 | 0.88297 | 0.9108248949 | 0.134784 | 4.038759634 | 0 |
| 2 | 47408470 | T | A | L | Q | MSH2 | ENST00000233146 | 0.8376 | 0.03653 | 0.77555 | 0.9348 | 0.95899 | 0.7322870276 | 0.95743 | 0.998603642 | 0.471564 | 9.364555431 | 0 |
| 2 | 47408470 | T | C | L | P | MSH2 | ENST00000233146 | 0.85279 | 0.03006 | 0.87197 | 0.93448 | 0.96153 | 0.8779830161 | 0.96942 | 0.9964900613 | 0.477515 | 9.364555431 | 0 |
| 2 | 47408470 | T | G | L | R | MSH2 | ENST00000233146 | 0.84831 | 0.0315 | 0.8457 | 0.93684 | 0.95863 | 0.8588737916 | 0.96012 | 0.9988698363 | 0.474933 | 9.364555431 | 0 |
| 2 | 47408472 | G | A | V | I | MSH2 | ENST00000233146 | 0.33575 | 0.00827 | 0.51692 | 0.58047 | 0.72016 | 0.3403159593 | 0.8635 | 0.4078218639 | 0.117363 | 3.016713796 | 0 |
| 2 | 47408472 | G | C | V | L | MSH2 | ENST00000233146 | 0.47797 | 0.02049 | 0.71411 | 0.79822 | 0.80118 | 0.4939171275 | 0.87172 | 0.3995691487 | 0.0806362 | 6.460842953 | 0 |
| 2 | 47408472 | G | T | V | F | MSH2 | ENST00000233146 | 0.67557 | 0.02875 | 0.68751 | 0.90013 | 0.92561 | 0.6777792075 | 0.9135 | 0.9886425138 | 0.301757 | 8.265451631 | 0 |
| 2 | 47408473 | T | A | V | D | MSH2 | ENST00000233146 | 0.68587 | 0.02883 | 0.74555 | 0.85885 | 0.92069 | 0.723349998 | 0.89497 | 0.9717434049 | 0.287548 | 3.971967309 | 0 |
| 2 | 47408473 | T | C | V | A | MSH2 | ENST00000233146 | 0.61574 | 0.02741 | 0.73016 | 0.78543 | 0.89147 | 0.4972891719 | 0.85101 | 0.875831604 | 0.252792 | 4.166933058 | 0 |
| 2 | 47408473 | T | G | V | G | MSH2 | ENST00000233146 | 0.68683 | 0.03099 | 0.57175 | 0.88988 | 0.91652 | 0.5997128519 | 0.8825 | 0.9708541632 | 0.323659 | 4.975968149 | 0 |
| 2 | 47408475 | C | A | R | S | MSH2 | ENST00000233146 | 0.80037 | 0.03362 | 0.85225 | 0.9012 | 0.92356 | 0.6685057103 | 0.95237 | 0.9890298247 | 0.314575 | 5.241835716 | 0 |
| 2 | 47408475 | C | G | R | G | MSH2 | ENST00000233146 | 0.83345 | 0.03219 | 0.83851 | 0.90933 | 0.92316 | 0.6311412169 | 0.94709 | 0.9883372188 | 0.29457 | 7.333695901 | 0 |
| 2 | 47408475 | C | T | R | C | MSH2 | ENST00000233146 | 0.90031 | 0.03212 | 0.91375 | 0.93572 | 0.93123 | 0.6155138332 | 0.96907 | 0.9886159301 | 0.287584 | 7.175228666 | 0 |
| 2 | 47408476 | G | A | R | H | MSH2 | ENST00000233146 | 0.75439 | 0.0334 | 0.86421 | 0.78104 | 0.90537 | 0.5866815286 | 0.88647 | 0.4621822273 | 0.212881 | 5.244930736 | 0 |
| 2 | 47408476 | G | C | R | P | MSH2 | ENST00000233146 | 0.79041 | 0.03118 | 0.87573 | 0.83204 | 0.93824 | 0.7910989569 | 0.91124 | 0.9912959337 | 0.466741 | 8.29526319 | 0 |
| 2 | 47408476 | G | T | R | L | MSH2 | ENST00000233146 | 0.62635 | 0.02815 | 0.87124 | 0.77742 | 0.92561 | 0.6283782516 | 0.94248 | 0.9872313142 | 0.350691 | 5.336241197 | 0 |
| 2 | 47408478 | C | A | Q | K | MSH2 | ENST00000233146 | 0.46171 | 0.00638 | 0.70887 | 0.70248 | 0.8517 | 0.6331968881 | 0.92981 | 0.9569167495 | 0.273743 | 5.224360135 | 0 |
| 2 | 47408478 | C | G | Q | E | MSH2 | ENST00000233146 | 0.44985 | 0.01025 | 0.66182 | 0.62639 | 0.84213 | 0.5513370256 | 0.91512 | 0.9485661983 | 0.262672 | 6.304394184 | 0 |
| 2 | 47408479 | A | C | Q | P | MSH2 | ENST00000233146 | 0.78304 | 0.02366 | 0.65944 | 0.88495 | 0.94839 | 0.8143291772 | 0.95642 | 0.9884309769 | 0.435776 | 6.304394184 | 0 |
| 2 | 47408479 | A | G | Q | R | MSH2 | ENST00000233146 | 0.45942 | 0.00664 | 0.70556 | 0.69589 | 0.8543 | 0.6296383426 | 0.96196 | 0.8795618415 | 0.259705 | 5.530900277 | 0 |
| 2 | 47408479 | A | T | Q | L | MSH2 | ENST00000233146 | 0.62233 | 0.01741 | 0.64854 | 0.90694 | 0.92233 | 0.5972118392 | 0.93524 | 0.9930922985 | 0.314164 | 6.304394184 | 0 |
| 2 | 47408481 | T | A | Y | N | MSH2 | ENST00000233146 | 0.7965 | 0.02857 | 0.86967 | 0.9045 | 0.93436 | 0.7827247358 | 0.91711 | 0.990896523 | 0.359309 | 9.74555058 | 0 |
| 2 | 47408481 | T | C | Y | H | MSH2 | ENST00000233146 | 0.80716 | 0.03274 | 0.8757 | 0.85337 | 0.91058 | 0.7665777985 | 0.88684 | 0.9756610394 | 0.308418 | 5.47604755 | 0 |
| 2 | 47408481 | T | G | Y | D | MSH2 | ENST00000233146 | 0.81936 | 0.02741 | 0.86402 | 0.89704 | 0.94167 | 0.8419843201 | 0.9543 | 0.9831449986 | 0.464234 | 9.74555058 | 0 |
| 2 | 47408482 | A | C | Y | S | MSH2 | ENST00000233146 | 0.81489 | 0.01948 | 0.80084 | 0.89939 | 0.96262 | 0.8120400595 | 0.92143 | 0.9856796861 | 0.460612 | 9.74555058 | 0 |
| 2 | 47408482 | A | G | Y | C | MSH2 | ENST00000233146 | 0.85003 | 0.0337 | 0.87531 | 0.86074 | 0.96371 | 0.7688264294 | 0.8583 | 0.976954937 | 0.283669 | 8.748446061 | 0 |
| 2 | 47408484 | A | G | R | G | MSH2 | ENST00000233146 | 0.76084 | 0.03219 | 0.7634 | 0.92747 | 0.90885 | 0.8203787101 | 0.95712 | 0.9981451035 | 0.449279 | 9.218401214 | 0 |
| 2 | 47408485 | G | A | R | K | MSH2 | ENST00000233146 | 0.73621 | 0.0337 | 0.83466 | 0.78842 | 0.88259 | 0.7154527504 | 0.91372 | 0.9798070192 | 0.344709 | 3.920573538 | 0 |
| 2 | 47408485 | G | C | R | T | MSH2 | ENST00000233146 | 0.88266 | 0.0292 | 0.80974 | 0.85046 | 0.93436 | 0.7796240772 | 0.92799 | 0.9974468946 | 0.460174 | 8.092688163 | 0 |

| | | | | | | | | | | | | | | | | |
|---|---|---|---|---|---|---|---|---|---|---|---|---|---|---|---|---|
| 2 | 47408485 | G | T | R | I | MSH2 | ENST00000233146 | 0.91223 | 0.0118 | 0.78128 | 0.88347 | 0.95608 | 0.7380214516 | 0.90227 | 0.9980600476 | 0.474432 | 9.218401214 | 0 |
| 2 | 47408486 | A | C | R | S | MSH2 | ENST00000233146 | 0.85756 | 0.03409 | 0.77694 | 0.93597 | 0.92924 | 0.783202508 | 0.96722 | 0.9983757734 | 0.447509 | 9.218401214 | 0 |
| 2 | 47408486 | A | T | R | S | MSH2 | ENST00000233146 | 0.8513 | 0.03409 | 0.77694 | 0.93765 | 0.92924 | 0.783202508 | 0.96725 | 0.9949944019 | 0.447506 | 9.218401214 | 0 |
| 2 | 47408487 | G | A | V | I | MSH2 | ENST00000233146 | 0.40681 | 0.00696 | 0.50092 | 0.77422 | 0.82103 | 0.3844148328 | 0.8089 | 0.6590860486 | 0.218528 | 3.074380031 | 0 |
| 2 | 47408487 | G | C | V | L | MSH2 | ENST00000233146 | 0.45401 | 0.00627 | 0.6762 | 0.83701 | 0.88684 | 0.6514735985 | 0.95635 | 0.9206491709 | 0.350864 | 5.658731766 | 0 |
| 2 | 47408487 | G | T | V | F | MSH2 | ENST00000233146 | 0.79358 | 0.0264 | 0.65173 | 0.8912 | 0.97848 | 0.8488231536 | 0.94161 | 0.9948068261 | 0.527192 | 7.827890369 | 0 |
| 2 | 47408488 | T | A | V | D | MSH2 | ENST00000233146 | 0.75981 | 0.02547 | 0.70968 | 0.95178 | 0.96553 | 0.9225804148 | 0.98729 | 0.9974447489 | 0.535065 | 7.827890369 | 1 |
| 2 | 47408488 | T | C | V | A | MSH2 | ENST00000233146 | 0.52281 | 0.00659 | 0.69518 | 0.90597 | 0.89193 | 0.6882472181 | 0.88899 | 0.8868834972 | 0.270565 | 4.219985628 | 0 |
| 2 | 47408488 | T | G | V | G | MSH2 | ENST00000233146 | 0.74069 | 0.02193 | 0.54803 | 0.95163 | 0.96334 | 0.8904988758 | 0.96884 | 0.9937933683 | 0.523868 | 7.827890369 | 0 |
| 2 | 47408490 | G | C | E | Q | MSH2 | ENST00000233146 | 0.78622 | 0.02414 | 0.6886 | 0.78878 | 0.91778 | 0.7685965082 | 0.94091 | 0.9781686664 | 0.386013 | 8.161590271 | 0 |
| 2 | 47408491 | A | C | E | A | MSH2 | ENST00000233146 | 0.79239 | 0.015 | 0.70454 | 0.89383 | 0.9608 | 0.8220110459 | 0.91348 | 0.9928043485 | 0.408152 | 3.284103657 | 0 |
| 2 | 47408491 | A | G | E | G | MSH2 | ENST00000233146 | 0.84081 | 0.03274 | 0.64662 | 0.92366 | 0.96883 | 0.8577114612 | 0.92116 | 0.9977766871 | 0.476636 | 5.973443312 | 0 |
| 2 | 47408491 | A | T | E | V | MSH2 | ENST00000233146 | 0.82326 | 0.03281 | 0.68016 | 0.93245 | 0.9681 | 0.8127377552 | 0.95641 | 0.9983267188 | 0.478935 | 5.573532195 | 0 |
| 2 | 47408492 | A | C | E | D | MSH2 | ENST00000233146 | 0.64368 | 0.03686 | 0.71083 | 0.89508 | 0.92964 | 0.7568753611 | 0.96156 | 0.991407454 | 0.345798 | 8.161590271 | 0 |
| 2 | 47408492 | A | T | E | D | MSH2 | ENST00000233146 | 0.6372 | 0.03686 | 0.71083 | 0.89752 | 0.92964 | 0.7568753611 | 0.96187 | 0.991219461 | 0.345791 | 8.161590271 | 0 |
| 2 | 47408493 | G | A | V | I | MSH2 | ENST00000233146 | 0.54882 | 0.00827 | 0.5965 | 0.64074 | 0.89376 | 0.2947369888 | 0.94325 | 0.2247228175 | 0.229058 | 2.623680928 | 0 |
| 2 | 47408493 | G | C | V | L | MSH2 | ENST00000233146 | 0.59147 | 0.00783 | 0.76663 | 0.7427 | 0.90406 | 0.5501317434 | 0.93713 | 0.9266287684 | 0.337231 | 3.649987395 | 0 |
| 2 | 47408493 | G | T | V | F | MSH2 | ENST00000233146 | 0.77029 | 0.02989 | 0.74034 | 0.87581 | 0.92356 | 0.7898658296 | 0.94862 | 0.991592774 | 0.47953 | 7.652661027 | 0 |
| 2 | 47408494 | T | A | V | D | MSH2 | ENST00000233146 | 0.71201 | 0.02708 | 0.79695 | 0.92949 | 0.94356 | 0.8786922106 | 0.99255 | 0.9984167814 | 0.489322 | 7.652661027 | 0 |
| 2 | 47408494 | T | C | V | A | MSH2 | ENST00000233146 | 0.66494 | 0.023 | 0.78671 | 0.88861 | 0.92151 | 0.6244913083 | 0.92293 | 0.9904111624 | 0.315313 | 7.652661027 | 0 |
| 2 | 47408494 | T | G | V | G | MSH2 | ENST00000233146 | 0.69684 | 0.029 | 0.65525 | 0.91521 | 0.94318 | 0.8491731031 | 0.98649 | 0.9979580641 | 0.48896 | 7.652661027 | 0 |
| 2 | 47408496 | T | A | Y | N | MSH2 | ENST00000233146 | 0.81465 | 0.02653 | 0.86055 | 0.90828 | 0.9579 | 0.8575237758 | 0.89479 | 0.9989169836 | 0.343336 | 7.186016495 | 0 |
| 2 | 47408496 | T | C | Y | H | MSH2 | ENST00000233146 | 0.80858 | 0.03129 | 0.86653 | 0.83441 | 0.95645 | 0.808755296 | 0.87526 | 0.9965767264 | 0.335701 | 7.186016495 | 0 |
| 2 | 47408497 | A | C | Y | S | MSH2 | ENST00000233146 | 0.79453 | 0.02574 | 0.80096 | 0.90301 | 0.83615 | 0.8589652313 | 0.91207 | 0.9973878264 | 0.279654 | 7.186016495 | 0 |
| 2 | 47408497 | A | G | Y | C | MSH2 | ENST00000233146 | 0.82662 | 0.03237 | 0.86101 | 0.89098 | 0.87198 | 0.8268068224 | 0.92457 | 0.9963679314 | 0.40932 | 2.70961997 | 0 |
| 2 | 47408497 | A | T | Y | F | MSH2 | ENST00000233146 | 0.57914 | 0.0071 | 0.77843 | 0.44526 | 0.80058 | 0.6005193525 | 0.81939 | 0.7800167203 | 0.140165 | 3.359739491 | 0 |
| 2 | 47408499 | A | C | K | Q | MSH2 | ENST00000233146 | 0.57055 | 0.00935 | 0.73064 | 0.68935 | 0.8306 | 0.4732657537 | 0.91974 | 0.9218927026 | 0.18191 | 3.11942485 | 0 |
| 2 | 47408499 | A | G | K | E | MSH2 | ENST00000233146 | 0.53253 | 0.01263 | 0.76798 | 0.60946 | 0.86603 | 0.5182776881 | 0.95375 | 0.5158883929 | 0.249562 | 2.379625668 | 0 |
| 2 | 47408500 | A | C | K | T | MSH2 | ENST00000233146 | 0.35953 | 0.01549 | 0.78753 | 0.74331 | 0.86302 | 0.5316918738 | 0.9037 | 0.7039122581 | 0.239129 | 5.942849606 | 0 |
| 2 | 47408500 | A | G | K | R | MSH2 | ENST00000233146 | 0.33494 | 0.00638 | 0.69494 | 0.64734 | 0.78286 | 0.3946531287 | 0.91669 | 0.6757801771 | 0.144712 | 1.44210527 | 0 |
| 2 | 47408501 | G | C | K | N | MSH2 | ENST00000233146 | 0.2799 | 0.00664 | 0.80079 | 0.66218 | 0.81933 | 0.5201745242 | 0.89085 | 0.1261517081 | 0.107882 | 5.942849606 | 0 |
| 2 | 47408501 | G | T | K | N | MSH2 | ENST00000233146 | 0.27597 | 0.00664 | 0.80079 | 0.66218 | 0.81933 | 0.5201745242 | 0.89081 | 0.1680568159 | 0.048603 | 5.942849606 | 0 |
| 2 | 47408502 | A | C | N | H | MSH2 | ENST00000233146 | 0.71024 | 0.03442 | 0.74882 | 0.82914 | 0.89918 | 0.6461512864 | 0.93737 | 0.9645889997 | 0.198872 | 7.076300121 | 0 |
| 2 | 47408502 | A | G | N | D | MSH2 | ENST00000233146 | 0.58559 | 0.02442 | 0.78972 | 0.81542 | 0.86403 | 0.6230683336 | 0.90193 | 0.9740079045 | 0.17861 | 7.076300121 | 0 |
| 2 | 47408502 | A | T | N | Y | MSH2 | ENST00000233146 | 0.75917 | 0.03212 | 0.77706 | 0.88051 | 0.94728 | 0.7089258723 | 0.9554 | 0.9926243424 | 0.461938 | 7.076300121 | 0 |
| 2 | 47408503 | A | C | N | T | MSH2 | ENST00000233146 | 0.54169 | 0.00971 | 0.7292 | 0.6969 | 0.86251 | 0.5603038419 | 0.82686 | 0.9244473577 | 0.180735 | 7.076300121 | 0 |
| 2 | 47408503 | A | G | N | S | MSH2 | ENST00000233146 | 0.38089 | 0.00909 | 0.6782 | 0.70022 | 0.78411 | 0.5195610659 | 0.84064 | 0.9630730748 | 0.102436 | 3.879395118 | 0 |
| 2 | 47408503 | A | T | N | I | MSH2 | ENST00000233146 | 0.75726 | 0.02864 | 0.74147 | 0.88867 | 0.92884 | 0.6803086711 | 0.90076 | 0.9934658408 | 0.334678 | 7.076300121 | 0 |
| 2 | 47408504 | T | A | N | K | MSH2 | ENST00000233146 | 0.35741 | 0.0114 | 0.81278 | 0.83374 | 0.81124 | 0.6761892452 | 0.79578 | 0.9470965266 | 0.0794187 | 7.076300121 | 0 |
| 2 | 47408504 | T | G | N | K | MSH2 | ENST00000233146 | 0.36309 | 0.0114 | 0.81278 | 0.83603 | 0.81124 | 0.6761892452 | 0.79582 | 0.9470965266 | 0.0793762 | 7.076300121 | 0 |
| 2 | 47408505 | A | G | R | G | MSH2 | ENST00000233146 | 0.65523 | 0.00551 | 0.30463 | 0.67205 | 0.76223 | 0.4421808108 | 0.94255 | 0.1173715101 | 0.00479914 | 4.666841584 | 0 |
| 2 | 47408506 | G | C | R | T | MSH2 | ENST00000233146 | 0.26437 | 0.00543 | 0.35644 | 0.68281 | 0.80476 | 0.3780900859 | 0.93507 | 0.4093696475 | 0.0778307 | 0.9251566296 | 0 |
| 2 | 47408506 | G | T | R | I | MSH2 | ENST00000233146 | 0.38329 | 0.00586 | 0.33112 | 0.6812 | 0.80238 | 0.4311431476 | 0.94496 | 0.1821259395 | 0.11983 | 1.306952406 | 0 |
| 2 | 47408507 | A | C | R | S | MSH2 | ENST00000233146 | 0.55731 | 0.00551 | 0.31323 | 0.62796 | 0.74728 | 0.3714228258 | 0.94483 | 0.3673717082 | 0.116877 | 2.533553104 | 0 |
| 2 | 47408507 | A | T | R | S | MSH2 | ENST00000233146 | 0.55154 | 0.00551 | 0.31323 | 0.62348 | 0.74728 | 0.3714228258 | 0.94538 | 0.4283432066 | 0.116875 | 2.533553104 | 0 |
| 2 | 47408508 | G | A | A | T | MSH2 | ENST00000233146 | 0.46165 | 0.00622 | 0.53059 | 0.68336 | 0.74306 | 0.3050816935 | 0.93553 | 0.6324093938 | 0.182959 | 2.867645313 | 0 |
| 2 | 47408508 | G | C | A | P | MSH2 | ENST00000233146 | 0.46078 | 0.02005 | 0.60633 | 0.65297 | 0.86603 | 0.4560283571 | 0.98548 | 0.541451688 | 0.360275 | 4.987368913 | 0 |
| 2 | 47408508 | G | T | A | S | MSH2 | ENST00000233146 | 0.4081 | 0.00608 | 0.47948 | 0.49143 | 0.71073 | 0.273493736 | 0.9026 | 0.7485553622 | 0.0891317 | 2.157238167 | 0 |
| 2 | 47408509 | C | A | A | D | MSH2 | ENST00000233146 | 0.36358 | 0.01048 | 0.6569 | 0.68847 | 0.83888 | 0.4247601767 | 0.9547 | 0.9195532799 | 0.344611 | 4.644218729 | 0 |
| 2 | 47408509 | C | G | A | G | MSH2 | ENST00000233146 | 0.24163 | 0.00574 | 0.50436 | 0.66113 | 0.76685 | 0.2879309993 | 0.87646 | 0.3538639843 | 0.120581 | 2.037450439 | 0 |
| 2 | 47408509 | C | T | A | V | MSH2 | ENST00000233146 | 0.43364 | 0.00654 | 0.55492 | 0.68056 | 0.82836 | 0.3456127812 | 0.91055 | 0.893797636 | 0.204651 | 4.11784946 | 1 |
| 2 | 47408511 | G | A | G | R | MSH2 | ENST00000233146 | 0.7532 | 0.0264 | 0.70828 | 0.82247 | 0.9014 | 0.6804570403 | 0.94777 | 0.9829586148 | 0.436147 | 7.010136521 | 0 |
| 2 | 47408511 | G | C | G | R | MSH2 | ENST00000233146 | 0.75002 | 0.0264 | 0.70828 | 0.82416 | 0.9014 | 0.6804570403 | 0.94723 | 0.9829586148 | 0.436152 | 7.010136521 | 0 |
| 2 | 47408512 | G | C | G | A | MSH2 | ENST00000233146 | 0.66786 | 0.01549 | 0.59498 | 0.72034 | 0.862 | 0.6661147557 | 0.88558 | 0.9304255843 | 0.274745 | 6.214666178 | 0 |
| 2 | 47408512 | G | T | G | V | MSH2 | ENST00000233146 | 0.71762 | 0.02324 | 0.60353 | 0.82938 | 0.91314 | 0.7155482663 | 0.94747 | 0.9887586832 | 0.453892 | 7.010136521 | 0 |
| 2 | 47408514 | A | C | N | H | MSH2 | ENST00000233146 | 0.42229 | 0.02119 | 0.4308 | 0.74362 | 0.74093 | 0.2975690571 | 0.90806 | 0.556468308 | 0.0965672 | 4.314844612 | 0 |
| 2 | 47408514 | A | G | N | D | MSH2 | ENST00000233146 | 0.30478 | 0.00735 | 0.48513 | 0.69466 | 0.69339 | 0.2180697565 | 0.89371 | 0.1280613548 | 0.0844966 | 4.046531487 | 0 |
| 2 | 47408514 | A | T | N | Y | MSH2 | ENST00000233146 | 0.46488 | 0.01983 | 0.4656 | 0.82432 | 0.80653 | 0.3440402667 | 0.92191 | 0.7813689709 | 0.145842 | 4.314844612 | 0 |
| 2 | 47408515 | A | C | N | T | MSH2 | ENST00000233146 | 0.23904 | 0.00769 | 0.41646 | 0.67187 | 0.78035 | 0.2733166129 | 0.88427 | 0.1895906627 | 0.0764015 | 2.636863247 | 0 |
| 2 | 47408515 | A | G | N | S | MSH2 | ENST00000233146 | 0.17614 | 0.00615 | 0.36693 | 0.55449 | 0.69082 | 0.1782750198 | 0.73142 | 0.02391634302 | 0.0256389 | -0.154320058 | 0 |
| 2 | 47408515 | A | T | N | I | MSH2 | ENST00000233146 | 0.31322 | 0.01312 | 0.44921 | 0.82257 | 0.84694 | 0.4069560194 | 0.88218 | 0.6729887724 | 0.0992148 | 4.314844612 | 0 |
| 2 | 47408516 | T | A | N | K | MSH2 | ENST00000233146 | 0.24978 | 0.00735 | 0.50381 | 0.6577 | 0.75349 | 0.2683045347 | 0.89949 | 0.1261649877 | 0.0544554 | 2.548670814 | 0 |
| 2 | 47408516 | T | G | N | K | MSH2 | ENST00000233146 | 0.25435 | 0.00735 | 0.50381 | 0.6569 | 0.75349 | 0.2683045347 | 0.89944 | 0.1151455528 | 0.0544728 | 2.548670814 | 0 |
| 2 | 47408517 | A | C | K | Q | MSH2 | ENST00000233146 | 0.74994 | 0.02914 | 0.56858 | 0.77467 | 0.83997 | 0.5807121707 | 0.94176 | 0.6652439473 | 0.253351 | 4.614152045 | 0 |
| 2 | 47408517 | A | G | K | E | MSH2 | ENST00000233146 | 0.77021 | 0.0167 | 0.61684 | 0.79091 | 0.82046 | 0.6479889621 | 0.89714 | 0.8581289649 | 0.221404 | 5.814567587 | 1 |
| 2 | 47408518 | A | C | K | T | MSH2 | ENST00000233146 | 0.7177 | 0.02369 | 0.63575 | 0.8331 | 0.84321 | 0.6355458823 | 0.86915 | 0.9550610781 | 0.166635 | 6.063855586 | 0 |
| 2 | 47408518 | A | G | K | R | MSH2 | ENST00000233146 | 0.66414 | 0.01086 | 0.53596 | 0.65855 | 0.82668 | 0.5196402769 | 0.93743 | 0.6677431464 | 0.154958 | 3.12751625 | 0 |
| 2 | 47408518 | A | T | K | M | MSH2 | ENST00000233146 | 0.75464 | 0.03106 | 0.53901 | 0.8436 | 0.85482 | 0.6859179715 | 0.90286 | 0.965288043 | 0.230854 | 7.939052316 | 0 |
| 2 | 47408519 | G | C | K | N | MSH2 | ENST00000233146 | 0.63804 | 0.02386 | 0.65153 | 0.73255 | 0.82892 | 0.6153384316 | 0.90745 | 0.7279382262 | 0.108411 | 5.220956722 | 0 |
| 2 | 47408519 | G | T | K | N | MSH2 | ENST00000233146 | 0.63175 | 0.02386 | 0.65153 | 0.73255 | 0.82892 | 0.6153384316 | 0.90765 | 0.7355979427 | 0.108408 | 5.220956722 | 0 |
| 2 | 47408520 | G | A | A | T | MSH2 | ENST00000233146 | 0.39405 | 0.00654 | 0.40347 | 0.69118 | 0.77972 | 0.3854772957 | 0.94434 | 0.4232769012 | 0.0835707 | 0.8271453437 | 0 |
| 2 | 47408520 | G | C | A | P | MSH2 | ENST00000233146 | 0.42544 | 0.00547 | 0.48273 | 0.74769 | 0.81646 | 0.5501070657 | 0.95025 | 0.3248135149 | 0.19556 | 5.22817121 | 0 |
| 2 | 47408520 | G | T | A | S | MSH2 | ENST00000233146 | 0.28607 | 0.00599 | 0.36513 | 0.54183 | 0.64775 | 0.3564326967 | 0.81428 | 0.1156118885 | 0.0510338 | -0.4633068956 | 0 |
| 2 | 47408521 | C | A | A | E | MSH2 | ENST00000233146 | 0.29402 | 0.01002 | 0.4866 | 0.76189 | 0.79998 | 0.5064885396 | 0.81147 | 0.5766267776 | 0.117276 | 2.1803672 | 0 |
| 2 | 47408521 | C | G | A | G | MSH2 | ENST00000233146 | 0.26706 | 0.0058 | 0.39238 | 0.65632 | 0.75211 | 0.3941455146 | 0.96084 | 0.2704847455 | 0.139453 | 1.338859641 | 0 |
| 2 | 47408521 | C | T | A | V | MSH2 | ENST00000233146 | 0.30833 | 0.01211 | 0.44076 | 0.73129 | 0.77718 | 0.451184437 | 0.88403 | 0.7114989758 | 0.131704 | 2.761215962 | 0 |
| 2 | 47408523 | T | A | S | T | MSH2 | ENST00000233146 | 0.17603 | 0.00592 | 0.36873 | 0.53467 | 0.77141 | 0.4193474166 | 0.97174 | 0.02979738638 | 0.0828821 | 0.9751045255 | 0 |
| 2 | 47408523 | T | C | S | P | MSH2 | ENST00000233146 | 0.35802 | 0.00928 | 0.38952 | 0.74058 | 0.86603 | 0.5708047239 | 0.95015 | 0.5469272137 | 0.192305 | 5.858038469 | 0 |
| 2 | 47408523 | T | G | S | A | MSH2 | ENST00000233146 | 0.19057 | 0.00608 | 0.35678 | 0.37934 | 0.7147 | 0.3319216325 | 0.93179 | 0.01978945918 | 0.0565996 | 1.162140136 | 0 |
| 2 | 47408524 | C | A | S | Y | MSH2 | ENST00000233146 | 0.36131 | 0.01309 | 0.46954 | 0.77874 | 0.83283 | 0.5107286136 | 0.91928 | 0.9335808754 | 0.181968 | 5.858038469 | 0 |
| 2 | 47408524 | C | G | S | C | MSH2 | ENST00000233146 | 0.39376 | 0.01902 | 0.3847 | 0.66481 | 0.83943 | 0.4774026689 | 0.8475 | 0.683829546 | 0.0301032 | 4.763636973 | 0 |
| 2 | 47408524 | C | T | S | F | MSH2 | ENST00000233146 | 0.3958 | 0.01048 | 0.4342 | 0.72452 | 0.76157 | 0.5015813417 | 0.88261 | 0.873814404 | 0.117294 | 5.858038469 | 0 |
| 2 | 47408526 | A | C | K | Q | MSH2 | ENST00000233146 | 0.72108 | 0.03322 | 0.64075 | 0.7693 | 0.8543 | 0.5637018447 | 0.90433 | 0.902618885 | 0.240203 | 4.787802879 | 0 |
| 2 | 47408527 | A | C | K | T | MSH2 | ENST00000233146 | 0.68259 | 0.01346 | 0.69951 | 0.82631 | 0.86098 | 0.6347350713 | 0.88821 | 0.8469943404 | 0.250607 | 4.323107713 | 0 |
| 2 | 47408527 | A | G | K | R | MSH2 | ENST00000233146 | 0.54698 | 0.0151 | 0.60954 | 0.68641 | 0.80831 | 0.4955601496 | 0.89927 | 0.9341061115 | 0.187848 | 3.458649286 | 0 |
| 2 | 47408527 | A | T | K | M | MSH2 | ENST00000233146 | 0.77545 | 0.0315 | 0.62329 | 0.85222 | 0.86653 | 0.689737244 | 0.89562 | 0.9608645439 | 0.293554 | 7.505576575 | 0 |
| 2 | 47408528 | G | C | K | N | MSH2 | ENST00000233146 | 0.54487 | 0.02943 | 0.71987 | 0.80003 | 0.80889 | 0.6132980876 | 0.8605 | 0.9313650727 | 0.118924 | 3.604573807 | 0 |
| 2 | 47408528 | G | T | K | N | MSH2 | ENST00000233146 | 0.53941 | 0.02943 | 0.71987 | 0.80003 | 0.80889 | 0.6132980876 | 0.8608 | 0.9313650727 | 0.118921 | 3.604573807 | 0 |
| 2 | 47408529 | G | A | E | K | MSH2 | ENST00000233146 | 0.47143 | 0.00615 | 0.76377 | 0.79937 | 0.84375 | 0.4873916815 | 0.94515 | 0.8916897774 | 0.318299 | 4.111918282 | 0 |
| 2 | 47408529 | G | C | E | Q | MSH2 | ENST00000233146 | 0.59096 | 0.01529 | 0.64269 | 0.76168 | 0.8233 | 0.475216228 | 0.87634 | 0.9210603833 | 0.233993 | 4.11816427 | 0 |
| 2 | 47408530 | A | C | E | A | MSH2 | ENST00000233146 | 0.40284 | 0.0058 | 0.65474 | 0.76488 | 0.83834 | 0.5278698884 | 0.89661 | 0.808999002 | 0.240723 | 2.946670122 | 0 |
| 2 | 47408530 | A | G | E | G | MSH2 | ENST00000233146 | 0.44979 | 0.00551 | 0.60376 | 0.77373 | 0.79456 | 0.5140205381 | 0.94058 | 0.8556973934 | 0.259583 | 3.033455698 | 0 |
| 2 | 47408530 | A | T | E | V | MSH2 | ENST00000233146 | 0.45862 | 0.01082 | 0.62803 | 0.82166 | 0.86852 | 0.5823509513 | 0.93118 | 0.9022008181 | 0.28365 | 2.443059826 | 0 |
| 2 | 47408531 | G | C | E | D | MSH2 | ENST00000233146 | 0.27227 | 0.00599 | 0.65102 | 0.61803 | 0.79212 | 0.3637032107 | 0.95172 | 0.1420519203 | 0.154158 | 2.38067749 | 0 |
| 2 | 47408531 | G | T | E | D | MSH2 | ENST00000233146 | 0.26846 | 0.00599 | 0.65102 | 0.61393 | 0.79212 | 0.3637032107 | 0.95122 | 0.1420519203 | 0.154163 | 2.38067749 | 0 |
| 2 | 47408532 | A | C | N | H | MSH2 | ENST00000233146 | 0.39899 | 0.00596 | 0.6139 | 0.60869 | 0.72246 | 0.486706545 | 0.91076 | 0.649189055 | 0.0995718 | 2.647158167 | 0 |
| 2 | 47408532 | A | G | N | D | MSH2 | ENST00000233146 | 0.43762 | 0.0132 | 0.66672 | 0.70589 | 0.8124 | 0.4575107263 | 0.89135 | 0.884404242 | 0.120364 | 5.74626466 | 0 |
| 2 | 47408532 | A | T | N | Y | MSH2 | ENST00000233146 | 0.74635 | 0.01834 | 0.65047 | 0.83564 | 0.86251 | 0.5656398699 | 0.90106 | 0.9748241901 | 0.231976 | 5.74626466 | 0 |
| 2 | 47408533 | A | C | N | T | MSH2 | ENST00000233146 | 0.39145 | 0.01361 | 0.58604 | 0.71769 | 0.81124 | 0.4490005713 | 0.93603 | 0.8889074326 | 0.163252 | 4.652361958 | 0 |
| 2 | 47408533 | A | G | N | S | MSH2 | ENST00000233146 | 0.35112 | 0.00669 | 0.53854 | 0.64996 | 0.71391 | 0.3682545376 | 0.91423 | 0.660328567 | 0.107405 | 3.052510141 | 0 |
| 2 | 47408533 | A | T | N | I | MSH2 | ENST00000233146 | 0.71883 | 0.02524 | 0.60672 | 0.8073 | 0.8859 | 0.5815908316 | 0.91182 | 0.9837201834 | 0.241479 | 3.685826501 | 0 |

| | | | | | | | | | | | | | | | | | | |
|---|---|---|---|---|---|---|---|---|---|---|---|---|---|---|---|---|---|---|
| 2 | 47408534 | T | A | N | K | MSH2 | ENST00000233146 | 0.29188 | 0.00909 | 0.69347 | 0.73015 | 0.75822 | 0.4931798814 | 0.87897 | 0.7296407819 | 0.086537 | 5.690224206 | 0 |
| 2 | 47408534 | T | G | N | K | MSH2 | ENST00000233146 | 0.29702 | 0.00909 | 0.69347 | 0.74866 | 0.75822 | 0.4931798814 | 0.87874 | 0.271128267 | 0.042244 | 5.690224206 | 0 |
| 2 | 47408535 | G | A | D | N | MSH2 | ENST00000233146 | 0.46183 | 0.00654 | 0.67314 | 0.6281 | 0.76289 | 0.4510165819 | 0.83745 | 0.8158563972 | 0.105145 | 4.779035754 | 0 |
| 2 | 47408535 | G | C | D | H | MSH2 | ENST00000233146 | 0.80542 | 0.02533 | 0.70163 | 0.6321 | 0.90096 | 0.6015954487 | 0.95153 | 0.9829168916 | 0.350167 | 6.357716509 | 0 |
| 2 | 47408535 | G | T | D | Y | MSH2 | ENST00000233146 | 0.82373 | 0.02883 | 0.68358 | 0.78451 | 0.91611 | 0.6958857492 | 0.97564 | 0.9932566881 | 0.443531 | 7.042939251 | 0 |
| 2 | 47408536 | A | C | D | A | MSH2 | ENST00000233146 | 0.56568 | 0.01244 | 0.60563 | 0.8276 | 0.82273 | 0.6183126721 | 0.94528 | 0.8505125046 | 0.248585 | 3.764887236 | 0 |
| 2 | 47408536 | A | G | D | G | MSH2 | ENST00000233146 | 0.59929 | 0.01211 | 0.61659 | 0.73337 | 0.88212 | 0.6418342048 | 0.98318 | 0.8286363482 | 0.324219 | 7.042939251 | 0 |
| 2 | 47408536 | A | T | D | V | MSH2 | ENST00000233146 | 0.86495 | 0.02143 | 0.59924 | 0.86747 | 0.89739 | 0.6757967983 | 0.98674 | 0.9869712591 | 0.396763 | 5.974989515 | 0 |
| 2 | 47408537 | T | A | D | E | MSH2 | ENST00000233146 | 0.26832 | 0.00599 | 0.64134 | 0.72735 | 0.80178 | 0.3498269518 | 0.97852 | 0.07937724888 | 0.119793 | 2.457598269 | 0 |
| 2 | 47408537 | T | G | D | E | MSH2 | ENST00000233146 | 0.27317 | 0.00599 | 0.64134 | 0.72735 | 0.80178 | 0.3498269518 | 0.97839 | 0.08146393958 | 0.119781 | 2.457598269 | 0 |
| 2 | 47408538 | T | A | W | R | MSH2 | ENST00000233146 | 0.88745 | 0.02815 | 0.83481 | 0.89123 | 0.87247 | 0.8709411701 | 0.84408 | 0.9998249412 | 0.309643 | 3.021174281 | 0 |
| 2 | 47408538 | T | C | W | R | MSH2 | ENST00000233146 | 0.89862 | 0.02815 | 0.83481 | 0.88829 | 0.87345 | 0.8709411701 | 0.84647 | 0.9998294115 | 0.309639 | 3.021174281 | 0 |
| 2 | 47408538 | T | G | W | G | MSH2 | ENST00000233146 | 0.91979 | 0.02802 | 0.75284 | 0.93336 | 0.91314 | 0.8585947353 | 0.97923 | 0.9998856783 | 0.486241 | 8.144627334 | 0 |
| 2 | 47408539 | G | C | W | S | MSH2 | ENST00000233146 | 0.93164 | 0.02927 | 0.78439 | 0.82326 | 0.92069 | 0.8692410119 | 0.93957 | 0.9999330044 | 0.494336 | 8.144627334 | 0 |
| 2 | 47408539 | G | T | W | L | MSH2 | ENST00000233146 | 0.91463 | 0.02595 | 0.79979 | 0.85283 | 0.90755 | 0.757765025 | 0.9197 | 0.9998518229 | 0.436775 | 8.144627334 | 0 |
| 2 | 47408541 | T | A | Y | N | MSH2 | ENST00000233146 | 0.43424 | 0.00535 | 0.44966 | 0.67659 | 0.82443 | 0.5779869794 | 0.94391 | 0.4776230454 | 0.180688 | 3.61407733 | 0 |
| 2 | 47408541 | T | C | Y | H | MSH2 | ENST00000233146 | 0.42794 | 0.00527 | 0.45108 | 0.50677 | 0.77972 | 0.511497299 | 0.91481 | 0.1868145168 | 0.090226 | 1.963669887 | 0 |
| 2 | 47408541 | T | G | Y | D | MSH2 | ENST00000233146 | 0.43472 | 0.00476 | 0.44051 | 0.60051 | 0.81818 | 0.6420296847 | 0.96111 | 0.3164998889 | 0.187013 | 1.302557633 | 0 |
| 2 | 47408542 | A | C | Y | S | MSH2 | ENST00000233146 | 0.34462 | 0.00517 | 0.34947 | 0.61695 | 0.78658 | 0.5917456061 | 0.97611 | 0.3134746253 | 0.198137 | 1.368378611 | 0 |
| 2 | 47408542 | A | G | Y | C | MSH2 | ENST00000233146 | 0.61813 | 0.00535 | 0.44968 | 0.63628 | 0.85117 | 0.5828015809 | 0.92044 | 0.3322665095 | 0.090995 | 4.321383709 | 0 |
| 2 | 47408542 | A | T | Y | F | MSH2 | ENST00000233146 | 0.32215 | 0.00592 | 0.32904 | 0.58125 | 0.74093 | 0.3694658654 | 0.91912 | 0.1759567559 | 0.0890321 | 2.933129926 | 0 |
| 2 | 47408544 | T | A | L | M | MSH2 | ENST00000233146 | 0.71722 | 0.02108 | 0.27652 | 0.62629 | 0.79151 | 0.4847524077 | 0.88718 | 0.7247098088 | 0.135469 | 7.421129166 | 0 |
| 2 | 47408544 | T | G | L | V | MSH2 | ENST00000233146 | 0.38702 | 0.00696 | 0.22917 | 0.64236 | 0.80476 | 0.4436801321 | 0.96838 | 0.1116253176 | 0.112417 | 2.161022502 | 0 |
| 2 | 47408545 | T | C | L | S | MSH2 | ENST00000233146 | 0.90906 | 0.02614 | 0.31695 | 0.76577 | 0.89467 | 0.8320517136 | 0.9909 | 0.957102716 | 0.470313 | 7.421129166 | 0 |
| 2 | 47408545 | T | G | L | W | MSH2 | ENST00000233146 | 0.72092 | 0.02965 | 0.39609 | 0.75662 | 0.85945 | 0.7703757396 | 0.96362 | 0.8968084455 | 0.334265 | 7.421129166 | 0 |
| 2 | 47408546 | G | C | L | F | MSH2 | ENST00000233146 | 0.7266 | 0.03083 | 0.28971 | 0.7136 | 0.90581 | 0.6265860755 | 0.93029 | 0.958278358 | 0.247084 | 7.421129166 | 0 |
| 2 | 47408546 | G | T | L | F | MSH2 | ENST00000233146 | 0.72003 | 0.03083 | 0.28971 | 0.74097 | 0.90581 | 0.6265860755 | 0.92553 | 0.958278358 | 0.247079 | 7.421129166 | 0 |
| 2 | 47408547 | G | A | A | T | MSH2 | ENST00000233146 | 0.55107 | 0.00735 | 0.53117 | 0.63174 | 0.79334 | 0.4803520389 | 0.96693 | 0.9093466401 | 0.270723 | 2.748067322 | 0 |
| 2 | 47408547 | G | C | A | P | MSH2 | ENST00000233146 | 0.61978 | 0.00599 | 0.61208 | 0.64651 | 0.91862 | 0.8011054037 | 0.9626 | 0.9401412606 | 0.425808 | 6.623874125 | 0 |
| 2 | 47408547 | G | T | A | S | MSH2 | ENST00000233146 | 0.49835 | 0.00696 | 0.48424 | 0.54798 | 0.79577 | 0.4488019104 | 0.93885 | 0.5876100063 | 0.217825 | 4.038030916 | 0 |
| 2 | 47408548 | C | A | A | E | MSH2 | ENST00000233146 | 0.22305 | 0.00574 | 0.60234 | 0.57455 | 0.78597 | 0.6752539303 | 0.95142 | 0.7763765454 | 0.220686 | 2.810262741 | 0 |
| 2 | 47408548 | C | G | A | G | MSH2 | ENST00000233146 | 0.30437 | 0.01172 | 0.50965 | 0.62984 | 0.84535 | 0.4512198974 | 0.8921 | 0.7585769296 | 0.225252 | 5.558963076 | 0 |
| 2 | 47408548 | C | T | A | V | MSH2 | ENST00000233146 | 0.29632 | 0.00674 | 0.55101 | 0.56881 | 0.7816 | 0.4888783611 | 0.96925 | 0.597871542 | 0.237308 | 2.349919788 | 0 |
| 2 | 47408550 | T | A | Y | N | MSH2 | ENST00000233146 | 0.5361 | 0.0058 | 0.59928 | 0.78639 | 0.87491 | 0.689003 | 0.8493 | 0.9258322716 | 0.215014 | 5.497575816 | 0 |
| 2 | 47408550 | T | C | Y | H | MSH2 | ENST00000233146 | 0.39156 | 0.01924 | 0.58541 | 0.70474 | 0.8517 | 0.5992779902 | 0.80574 | 0.8778578043 | 0.158966 | 5.497575816 | 0 |
| 2 | 47408551 | A | C | Y | S | MSH2 | ENST00000233146 | 0.34506 | 0.00763 | 0.5303 | 0.72267 | 0.8367 | 0.7139870717 | 0.91482 | 0.7233681679 | 0.173341 | 4.526202228 | 0 |
| 2 | 47408551 | A | G | Y | C | MSH2 | ENST00000233146 | 0.49686 | 0.02166 | 0.57172 | 0.78617 | 0.88212 | 0.6525361164 | 0.85984 | 0.5704247952 | -0.00232764 | 3.845273749 | 0 |
| 2 | 47408551 | A | T | Y | F | MSH2 | ENST00000233146 | 0.22694 | 0.00592 | 0.48391 | 0.65901 | 0.66258 | 0.3732670565 | 0.85096 | 0.06727331877 | 0.0539396 | 0.2262991713 | 0 |
| 2 | 47408553 | A | C | K | Q | MSH2 | ENST00000233146 | 0.61089 | 0.01692 | 0.67633 | 0.81187 | 0.90229 | 0.7339894304 | 0.94747 | 0.8696975708 | 0.293064 | 8.405002002 | 0 |
| 2 | 47408553 | A | G | K | E | MSH2 | ENST00000233146 | 0.78764 | 0.02646 | 0.71798 | 0.87145 | 0.93475 | 0.8037444985 | 0.97411 | 0.9552889466 | 0.44659 | 8.552151505 | 0 |
| 2 | 47408554 | A | C | K | T | MSH2 | ENST00000233146 | 0.76418 | 0.03219 | 0.72598 | 0.85951 | 0.93786 | 0.736804519 | 0.94911 | 0.9832628965 | 0.422336 | 6.389259491 | 0 |
| 2 | 47408554 | A | G | K | R | MSH2 | ENST00000233146 | 0.72997 | 0.03475 | 0.64386 | 0.78254 | 0.92028 | 0.7063848685 | 0.94678 | 0.9514457583 | 0.291433 | 7.484432469 | 0 |
| 2 | 47408554 | A | T | K | M | MSH2 | ENST00000233146 | 0.8014 | 0.02965 | 0.64954 | 0.87077 | 0.95353 | 0.7964612325 | 0.961 | 0.9888513684 | 0.450858 | 5.400456287 | 0 |
| 2 | 47408555 | G | C | K | N | MSH2 | ENST00000233146 | 0.82318 | 0.03243 | 0.74536 | 0.8797 | 0.90798 | 0.7807606144 | 0.91348 | 0.9797377586 | 0.205709 | 8.552151505 | 0 |
| 2 | 47408555 | G | T | K | N | MSH2 | ENST00000233146 | 0.81677 | 0.03243 | 0.74536 | 0.87936 | 0.90798 | 0.7807606144 | 0.91367 | 0.9797377586 | 0.205709 | 8.552151505 | 0 |
| 2 | 47410094 | G | A | A | T | MSH2 | ENST00000233146 | 0.87194 | 0.01953 | 0.47033 | 0.88129 | 0.90229 | 0.7690354049 | 0.94418 | 0.96194553 | 0.379107 | 6.257250383 | 0 |
| 2 | 47410094 | G | C | A | P | MSH2 | ENST00000233146 | 0.89468 | 0.02506 | 0.54557 | 0.8321 | 0.95316 | 0.9229372223 | 0.95422 | 0.9737392781 | 0.4629 | 7.242720993 | 0 |
| 2 | 47410094 | G | T | A | S | MSH2 | ENST00000233146 | 0.6842 | 0.0199 | 0.41357 | 0.86774 | 0.86149 | 0.715871927 | 0.90651 | 0.8652881328 | 0.283956 | 7.242720993 | 0 |
| 2 | 47410095 | C | G | A | G | MSH2 | ENST00000233146 | 0.21621 | 0.00599 | 0.44387 | 0.47001 | 0.79212 | 0.7184978704 | 0.90127 | 0.3203133047 | 0.111638 | 2.194159026 | 0 |
| 2 | 47410095 | C | T | A | V | MSH2 | ENST00000233146 | 0.4725 | 0.00681 | 0.49167 | 0.81552 | 0.89784 | 0.7962199502 | 0.9308 | 0.8212327957 | 0.258183 | 2.857904818 | 0 |
| 2 | 47410097 | T | A | S | T | MSH2 | ENST00000233146 | 0.67557 | 0.0133 | 0.60463 | 0.78393 | 0.89285 | 0.8104308315 | 0.9768 | 0.8878558897 | 0.368918 | 3.241632813 | 0 |
| 2 | 47410097 | T | C | S | P | MSH2 | ENST00000233146 | 0.82131 | 0.02927 | 0.61771 | 0.91626 | 0.98294 | 0.9228847684 | 0.97985 | 0.9896382093 | 0.486966 | 8.921083615 | 0 |
| 2 | 47410098 | C | G | S | C | MSH2 | ENST00000233146 | 0.61963 | 0.03563 | 0.60855 | 0.93219 | 0.9796 | 0.8364884931 | 0.96711 | 0.9912795332 | 0.491434 | 8.921083615 | 0 |
| 2 | 47410098 | C | T | S | F | MSH2 | ENST00000233146 | 0.63658 | 0.02934 | 0.64519 | 0.93205 | 0.98071 | 0.8966086707 | 0.98891 | 0.9934156984 | 0.49217 | 8.921083615 | 1 |
| 2 | 47410100 | C | A | P | T | MSH2 | ENST00000233146 | 0.64477 | 0.03033 | 0.74553 | 0.93152 | 0.97773 | 0.7921548097 | 0.97998 | 0.9900374011 | 0.492652 | 9.308730905 | 0 |
| 2 | 47410100 | C | G | P | A | MSH2 | ENST00000233146 | 0.6132 | 0.02614 | 0.69224 | 0.8994 | 0.97215 | 0.736273687 | 0.97565 | 0.983457133 | 0.480652 | 6.571654051 | 0 |
| 2 | 47410100 | C | T | P | S | MSH2 | ENST00000233146 | 0.5106 | 0.01172 | 0.71114 | 0.93604 | 0.95207 | 0.799546977 | 0.98056 | 0.8343870044 | 0.351127 | 6.802306829 | 0 |
| 2 | 47410101 | C | A | P | H | MSH2 | ENST00000233146 | 0.60142 | 0.03106 | 0.76123 | 0.93459 | 0.97661 | 0.8215874626 | 0.95948 | 0.9964786469 | 0.493925 | 9.308730905 | 1 |
| 2 | 47410101 | C | G | P | R | MSH2 | ENST00000233146 | 0.5754 | 0.03327 | 0.77008 | 0.92354 | 0.96553 | 0.849876982 | 0.97827 | 0.9903980172 | 0.486541 | 7.763919945 | 0 |
| 2 | 47410101 | C | T | P | L | MSH2 | ENST00000233146 | 0.61776 | 0.03144 | 0.77911 | 0.90068 | 0.96553 | 0.8313950006 | 0.98979 | 0.9990801811 | 0.482121 | 9.308730905 | 0 |
| 2 | 47410103 | G | A | G | S | MSH2 | ENST00000233146 | 0.83982 | 0.03475 | 0.75046 | 0.93398 | 0.99138 | 0.8972601921 | 0.99495 | 0.9944500923 | 0.549915 | 8.660946862 | 0 |
| 2 | 47410103 | G | C | G | R | MSH2 | ENST00000233146 | 0.86858 | 0.03175 | 0.84159 | 0.93858 | 0.99202 | 0.9296369545 | 0.993 | 0.9909552635 | 0.556103 | 9.845154579 | 0 |
| 2 | 47410103 | G | T | G | C | MSH2 | ENST00000233146 | 0.71233 | 0.01108 | 0.78972 | 0.95762 | 0.97067 | 0.8914969178 | 0.99248 | 0.9943134785 | 0.553157 | 9.845154579 | 1 |
| 2 | 47410104 | G | A | G | D | MSH2 | ENST00000233146 | 0.70968 | 0.03161 | 0.84651 | 0.92506 | 0.99471 | 0.940322148 | 0.9937 | 0.9906824872 | 0.556243 | 9.467912217 | 1 |
| 2 | 47410104 | G | C | G | A | MSH2 | ENST00000233146 | 0.6766 | 0.03254 | 0.75033 | 0.93252 | 0.98404 | 0.8871835268 | 0.99417 | 0.9828307776 | 0.547015 | 9.845154579 | 0 |
| 2 | 47410104 | G | T | G | V | MSH2 | ENST00000233146 | 0.71129 | 0.03212 | 0.75748 | 0.93034 | 0.9917 | 0.9226560453 | 0.98851 | 0.9937793612 | 0.555681 | 9.845154579 | 1 |
| 2 | 47410106 | A | C | N | H | MSH2 | ENST00000233146 | 0.75208 | 0.03442 | 0.61133 | 0.88907 | 0.96008 | 0.8308647331 | 0.96295 | 0.9890068558 | 0.227921 | 7.918760001 | 0 |
| 2 | 47410106 | A | G | N | D | MSH2 | ENST00000233146 | 0.76832 | 0.02984 | 0.64976 | 0.87208 | 0.94543 | 0.801499751 | 0.95367 | 0.9821556588 | 0.225159 | 8.146459574 | 0 |
| 2 | 47410106 | A | T | N | Y | MSH2 | ENST00000233146 | 0.79342 | 0.01408 | 0.63843 | 0.92383 | 0.96371 | 0.859177677 | 0.94753 | 0.9802716608 | 0.422913 | 8.146459574 | 0 |
| 2 | 47410107 | A | C | N | T | MSH2 | ENST00000233146 | 0.76642 | 0.03388 | 0.58161 | 0.88159 | 0.91442 | 0.7771150773 | 0.92212 | 0.9890211225 | 0.203323 | 3.39874263 | 0 |
| 2 | 47410107 | A | G | N | S | MSH2 | ENST00000233146 | 0.7035 | 0.03219 | 0.53722 | | 0.91143 | 0.7478477181 | 0.95447 | 0.1027867369 | -0.1723 | 7.020829099 | 0 |
| 2 | 47410107 | A | T | N | I | MSH2 | ENST00000233146 | 0.81135 | 0.03237 | 0.60724 | 0.92199 | 0.94913 | 0.8523414876 | 0.94282 | 0.9981406927 | 0.355608 | 8.146459574 | 0 |
| 2 | 47410108 | T | A | N | K | MSH2 | ENST00000233146 | 0.6778 | 0.03137 | 0.67617 | 0.88927 | 0.82159 | 0.8537618596 | 0.9561 | 0.9949709177 | 0.160458 | 8.146459574 | 0 |
| 2 | 47410108 | T | G | N | K | MSH2 | ENST00000233146 | 0.68603 | 0.03137 | 0.67617 | 0.88395 | 0.82159 | 0.8537618596 | 0.95612 | 0.9946900606 | 0.160457 | 8.146459574 | 0 |
| 2 | 47410109 | C | A | L | I | MSH2 | ENST00000233146 | 0.44734 | 0.00804 | 0.5509 | 0.76213 | 0.74936 | 0.4697902889 | 0.80851 | 0.6967387224 | 0.122997 | 4.267884495 | 0 |
| 2 | 47410109 | C | G | L | V | MSH2 | ENST00000233146 | 0.69612 | 0.01376 | 0.48517 | 0.87575 | 0.85064 | 0.6111561133 | 0.89845 | 0.1039240183 | -0.0677961 | 4.095227584 | 0 |
| 2 | 47410109 | C | T | L | F | MSH2 | ENST00000233146 | 0.7764 | 0.02943 | 0.53537 | 0.86642 | 0.89422 | 0.701097108 | 0.91166 | 0.9728865027 | 0.304536 | 6.100035002 | 0 |
| 2 | 47410110 | T | A | L | H | MSH2 | ENST00000233146 | 0.72869 | 0.0317 | 0.64895 | 0.89175 | 0.95499 | 0.8288913163 | 0.96002 | 0.9887006908 | 0.439268 | 9.502854807 | 0 |
| 2 | 47410110 | T | C | L | P | MSH2 | ENST00000233146 | 0.54358 | 0.00535 | 0.69233 | 0.8792 | 0.9328 | 0.8871793404 | 0.9889 | 0.9807615876 | 0.486162 | 9.502854807 | 0 |
| 2 | 47410110 | T | G | L | R | MSH2 | ENST00000233146 | 0.74563 | 0.02828 | 0.65104 | 0.89039 | 0.96664 | 0.8870089927 | 0.9709 | 0.9885123968 | 0.447263 | 7.202432306 | 0 |
| 2 | 47410112 | T | A | S | T | MSH2 | ENST00000233146 | 0.28111 | 0.00592 | 0.36979 | 0.45752 | 0.79758 | 0.4221288075 | 0.96342 | 0.04006906424 | 0.0908136 | -0.7705367648 | 0 |
| 2 | 47410112 | T | C | S | P | MSH2 | ENST00000233146 | 0.69348 | 0.01414 | 0.39056 | 0.79194 | 0.87 | 0.7422775137 | 0.95549 | 0.4769563973 | 0.0901562 | 4.450339768 | 0 |
| 2 | 47410112 | T | G | S | A | MSH2 | ENST00000233146 | 0.34456 | 0.00608 | 0.34674 | 0.59464 | 0.75005 | 0.3569192962 | 0.95364 | 0.05978730376 | 0.0820314 | 0.5634982715 | 0 |
| 2 | 47410113 | C | A | S | Y | MSH2 | ENST00000233146 | 0.46488 | 0.01249 | 0.44892 | 0.77363 | 0.87198 | 0.5832219417 | 0.89005 | 0.8425139575 | 0.23842 | 4.44121281 | 0 |
| 2 | 47410113 | C | G | S | C | MSH2 | ENST00000233146 | 0.60816 | 0.02307 | 0.38572 | 0.77641 | 0.87099 | 0.4931845911 | 0.81367 | 0.9016498254 | 0.222545 | 3.842973957 | 1 |
| 2 | 47410113 | C | T | S | F | MSH2 | ENST00000233146 | 0.49802 | 0.00586 | 0.42383 | 0.74289 | 0.86703 | 0.5857425506 | 0.88174 | 0.7212879658 | 0.226971 | 2.818470712 | 0 |
| 2 | 47410115 | C | A | Q | K | MSH2 | ENST00000233146 | 0.46152 | 0.00898 | 0.4877 | 0.79719 | 0.84694 | 0.7598841393 | 0.97302 | 0.8607975245 | 0.384991 | 7.558496186 | 0 |
| 2 | 47410115 | C | G | Q | E | MSH2 | ENST00000233146 | 0.3252 | 0.00619 | 0.42335 | 0.63711 | 0.82216 | 0.7200350346 | 0.95988 | 0.4517203271 | 0.245849 | 3.493064529 | 0 |
| 2 | 47410116 | A | C | Q | P | MSH2 | ENST00000233146 | 0.73173 | 0.02366 | 0.43741 | 0.74374 | 0.93554 | 0.8717475054 | 0.98772 | 0.9688944817 | 0.437801 | 7.559982215 | 0 |
| 2 | 47410116 | A | G | Q | R | MSH2 | ENST00000233146 | 0.52566 | 0.00619 | 0.48286 | 0.61115 | 0.8356 | 0.7779993017 | 0.97727 | 0.8387383223 | 0.253092 | 8.422381836 | 0 |
| 2 | 47410116 | A | T | Q | L | MSH2 | ENST00000233146 | 0.5768 | 0.00608 | 0.41288 | 0.81256 | 0.90185 | 0.7440826485 | 0.98109 | 0.9804783463 | 0.422052 | 7.252238269 | 0 |
| 2 | 47410117 | G | C | Q | H | MSH2 | ENST00000233146 | 0.23926 | 0.02835 | 0.47546 | 0.73088 | 0.89918 | 0.8008244195 | 0.94771 | 0.880723536 | 0.291838 | 8.197770828 | 0 |
| 2 | 47410117 | G | T | Q | H | MSH2 | ENST00000233146 | 0.23579 | 0.02835 | 0.47546 | 0.79502 | 0.89918 | 0.8008244195 | 0.94819 | 0.8868304491 | 0.291837 | 8.197770828 | 0 |
| 2 | 47410118 | T | A | F | I | MSH2 | ENST00000233146 | 0.71722 | 0.01133 | 0.69222 | 0.68404 | 0.87781 | 0.7615407672 | 0.90254 | 0.861454308 | 0.273602 | 5.539016725 | 0 |
| 2 | 47410118 | T | C | F | L | MSH2 | ENST00000233146 | 0.57266 | 0.00586 | 0.7145 | 0.67535 | 0.87588 | 0.7122084462 | 0.94797 | 0.6554791331 | 0.374799 | 4.817021849 | 0 |
| 2 | 47410118 | T | G | F | V | MSH2 | ENST00000233146 | 0.42933 | 0.00599 | 0.65573 | 0.45536 | 0.88354 | 0.7048233158 | 0.94418 | 0.1457239687 | 0.154961 | 4.491685793 | 0 |
| 2 | 47410119 | T | A | F | Y | MSH2 | ENST00000233146 | 0.56442 | 0.00654 | 0.70336 | 0.75575 | 0.87539 | 0.634957446 | 0.96844 | 0.8137022853 | 0.305476 | 8.451540076 | 0 |
| 2 | 47410119 | T | C | F | S | MSH2 | ENST00000233146 | 0.84552 | 0.01591 | 0.67673 | 0.85302 | 0.96773 | 0.8187174551 | 0.97297 | 0.9845442176 | 0.459968 | 3.976212056 | 0 |
| 2 | 47410119 | T | G | F | C | MSH2 | ENST00000233146 | 0.85873 | 0.02696 | 0.70136 | 0.82079 | 0.98621 | 0.7702556011 | 0.96629 | 0.9886120809 | 0.469017 | 8.165648751 | 0 |
| 2 | 47410120 | T | A | F | L | MSH2 | ENST00000233146 | 0.4735 | 0.00586 | 0.7145 | 0.55283 | 0.86098 | 0.7122084462 | 0.96826 | 0.5704741478 | 0.35889 | 4.817021849 | 0 |
| 2 | 47410120 | T | G | F | L | MSH2 | ENST00000233146 | 0.47999 | 0.00586 | 0.7145 | 0.69278 | 0.86098 | 0.7122084462 | 0.9667 | 0.5598739982 | 0.358893 | 4.817021849 | 0 |

| | | | | | | | | | | | | | | | | | | |
|---|---|---|---|---|---|---|---|---|---|---|---|---|---|---|---|---|---|---|
| 2 | 47410121 | G | A | E | K | MSH2 | ENST00000233146 | 0.85242 | 0.02056 | 0.6445 | 0.93955 | 0.96044 | 0.8708368407 | 0.99137 | 0.9830626729 | 0.53468 | 5.256023433 | 0 |
| 2 | 47410121 | G | C | E | Q | MSH2 | ENST00000233146 | 0.76642 | 0.01295 | 0.51305 | 0.87008 | 0.91442 | 0.7794461967 | 0.98448 | 0.9050923054 | 0.487142 | 5.561519796 | 0 |
| 2 | 47410122 | A | C | E | A | MSH2 | ENST00000233146 | 0.7897 | 0.02516 | 0.53341 | 0.8676 | 0.9833 | 0.8267377469 | 0.98851 | 0.9897336364 | 0.522425 | 5.845916457 | 0 |
| 2 | 47410122 | A | G | E | G | MSH2 | ENST00000233146 | 0.84408 | 0.02421 | 0.48366 | 0.90613 | 0.97438 | 0.8196193064 | 0.98388 | 0.9946222787 | 0.539518 | 5.480413629 | 0 |
| 2 | 47410122 | A | T | E | V | MSH2 | ENST00000233146 | 0.82825 | 0.03029 | 0.51151 | 0.92275 | 0.99265 | 0.8235285863 | 0.99318 | 0.9951600432 | 0.540263 | 9.045722962 | 0 |
| 2 | 47410123 | A | C | E | D | MSH2 | ENST00000233146 | 0.64027 | 0.02255 | 0.53003 | 0.74976 | 0.94205 | 0.7341870723 | 0.98921 | 0.9251813293 | 0.434549 | 5.222644475 | 0 |
| 2 | 47410123 | A | T | E | D | MSH2 | ENST00000233146 | 0.63382 | 0.02255 | 0.53003 | 0.76743 | 0.94205 | 0.7341870723 | 0.98891 | 0.9251813293 | 0.434548 | 5.222644475 | 0 |
| 2 | 47410124 | G | A | D | N | MSH2 | ENST00000233146 | 0.80945 | 0.01741 | 0.32744 | 0.93286 | 0.88117 | 0.6057607597 | 0.93849 | 0.9738658395 | 0.234171 | 3.587703667 | 0 |
| 2 | 47410124 | G | C | D | H | MSH2 | ENST00000233146 | 0.82022 | 0.0233 | 0.35075 | 0.86498 | 0.93515 | 0.6992849776 | 0.89494 | 0.9868725989 | 0.347208 | 6.941102322 | 0 |
| 2 | 47410124 | G | T | D | Y | MSH2 | ENST00000233146 | 0.83166 | 0.02516 | 0.34216 | 0.86458 | 0.94765 | 0.759201275 | 0.94347 | 0.9865842863 | 0.385981 | 5.301409663 | 0 |
| 2 | 47410125 | A | C | D | A | MSH2 | ENST00000233146 | 0.65335 | 0.01376 | 0.27832 | 0.89829 | 0.93515 | 0.6929332119 | 0.89398 | 0.9825099707 | 0.344574 | 4.009764883 | 0 |
| 2 | 47410125 | A | G | D | G | MSH2 | ENST00000233146 | 0.66834 | 0.01562 | 0.28167 | 0.89059 | 0.94318 | 0.7065128703 | 0.91441 | 0.9865702987 | 0.342321 | 4.134352076 | 0 |
| 2 | 47410125 | A | T | D | V | MSH2 | ENST00000233146 | 0.64648 | 0.01549 | 0.26342 | 0.91336 | 0.95316 | 0.7259349589 | 0.95706 | 0.9893118739 | 0.388153 | 5.444271945 | 0 |
| 2 | 47410127 | A | C | I | L | MSH2 | ENST00000233146 | 0.2083 | 0.0058 | 0.36374 | 0.60452 | 0.77654 | 0.497228717 | 0.89481 | 0.04230891164 | 0.123085 | 3.353301664 | 0 |
| 2 | 47410127 | A | G | I | V | MSH2 | ENST00000233146 | 0.17678 | 0.00603 | 0.27699 | 0.49013 | 0.77335 | 0.343957434 | 0.91988 | 0.02435435727 | 0.135377 | 1.531236999 | 0 |
| 2 | 47410127 | A | T | I | F | MSH2 | ENST00000233146 | 0.23132 | 0.00504 | 0.38249 | 0.69676 | 0.81761 | 0.6816398482 | 0.97915 | 0.08558610827 | 0.239081 | 4.644772503 | 0 |
| 2 | 47410128 | T | A | I | N | MSH2 | ENST00000233146 | 0.65312 | 0.00916 | 0.47905 | 0.87662 | 0.94053 | 0.7723055573 | 0.85916 | 0.972256422 | 0.290965 | 4.665584019 | 0 |
| 2 | 47410128 | T | C | I | T | MSH2 | ENST00000233146 | 0.4735 | 0.00795 | 0.5213 | 0.82113 | 0.91484 | 0.6283759657 | 0.87241 | 0.8325644732 | 0.305097 | 5.028438659 | 0 |
| 2 | 47410128 | T | G | I | S | MSH2 | ENST00000233146 | 0.65609 | 0.00754 | 0.40017 | 0.84695 | 0.95316 | 0.780556569 | 0.94807 | 0.9469535351 | 0.419881 | 3.818674083 | 0 |
| 2 | 47410130 | C | A | L | I | MSH2 | ENST00000233146 | 0.35547 | 0.0058 | 0.53062 | 0.83287 | 0.80831 | 0.4645925359 | 0.91106 | 0.6848363918 | 0.284945 | 5.307951789 | 0 |
| 2 | 47410130 | C | G | L | V | MSH2 | ENST00000233146 | 0.48177 | 0.00735 | 0.45546 | 0.93042 | 0.90494 | 0.63431847 | 0.94258 | 0.2678600857 | 0.113018 | 6.81786681 | 0 |
| 2 | 47410130 | C | T | L | F | MSH2 | ENST00000233146 | 0.49958 | 0.01756 | 0.50626 | 0.91339 | 0.93044 | 0.7056840193 | 0.94426 | 0.6378371119 | 0.257599 | 7.416660299 | 0 |
| 2 | 47410131 | T | A | L | H | MSH2 | ENST00000233146 | 0.67509 | 0.02741 | 0.61319 | 0.91487 | 0.9659 | 0.8200193962 | 0.973 | 0.9896944848 | 0.528632 | 7.149026731 | 0 |
| 2 | 47410131 | T | C | L | P | MSH2 | ENST00000233146 | 0.50036 | 0.00497 | 0.66485 | 0.93037 | 0.90842 | 0.9075371158 | 0.97809 | 0.9566249847 | 0.523197 | 8.129316804 | 0 |
| 2 | 47410131 | T | G | L | R | MSH2 | ENST00000233146 | 0.68747 | 0.022 | 0.61903 | 0.90705 | 0.96371 | 0.8969992723 | 0.98349 | 0.9954726405 | 0.526261 | 7.840018382 | 0 |
| 2 | 47410133 | T | A | F | I | MSH2 | ENST00000233146 | 0.72172 | 0.01436 | 0.54416 | 0.82939 | 0.92397 | 0.7354680815 | 0.93968 | 0.9856268764 | 0.369165 | 6.265156052 | 0 |
| 2 | 47410133 | T | C | F | L | MSH2 | ENST00000233146 | 0.70431 | 0.0103 | 0.58561 | 0.83214 | 0.87733 | 0.6764104906 | 0.9498 | 0.9443204403 | 0.397295 | 4.122737255 | 0 |
| 2 | 47410133 | T | G | F | V | MSH2 | ENST00000233146 | 0.71842 | 0.01221 | 0.51431 | 0.87089 | 0.92561 | 0.7305153289 | 0.96069 | 0.9643537998 | 0.456416 | 6.028655117 | 0 |
| 2 | 47410134 | T | A | F | Y | MSH2 | ENST00000233146 | 0.50739 | 0.00562 | 0.56121 | 0.35983 | 0.80595 | 0.6060505436 | 0.96086 | 0.7477836609 | 0.282784 | 9.046461 | 0 |
| 2 | 47410134 | T | C | F | S | MSH2 | ENST00000233146 | 0.71482 | 0.01309 | 0.53919 | 0.8637 | 0.87539 | 0.7659097243 | 0.92156 | 0.9466807246 | 0.305354 | 5.91553863 | 0 |
| 2 | 47410134 | T | G | F | C | MSH2 | ENST00000233146 | 0.89933 | 0.0245 | 0.58193 | 0.76411 | 0.9248 | 0.750180435 | 0.98 | 0.9709227085 | 0.486836 | 9.202600079 | 1 |
| 2 | 47410135 | T | A | F | L | MSH2 | ENST00000233146 | 0.34981 | 0.0103 | 0.58561 | 0.68578 | 0.86503 | 0.6764104906 | 0.95514 | 0.9376878738 | 0.34822 | 4.122737255 | 0 |
| 2 | 47410135 | T | G | F | L | MSH2 | ENST00000233146 | 0.35542 | 0.0103 | 0.58561 | 0.71831 | 0.86503 | 0.6764104906 | 0.9519 | 0.9376878738 | 0.348222 | 4.122737255 | 0 |
| 2 | 47410136 | G | A | G | S | MSH2 | ENST00000233146 | 0.53019 | 0.00603 | 0.28045 | 0.7721 | 0.80417 | 0.6224923187 | 0.96251 | 0.7947307815 | 0.273392 | 3.186259703 | 0 |
| 2 | 47410136 | G | C | G | R | MSH2 | ENST00000233146 | 0.71795 | 0.00983 | 0.41903 | 0.73159 | 0.88401 | 0.6872951851 | 0.96773 | 0.5696849823 | 0.292713 | 5.681857197 | 0 |
| 2 | 47410136 | G | T | G | C | MSH2 | ENST00000233146 | 0.89302 | 0.02143 | 0.34599 | 0.78428 | 0.94243 | 0.7151603638 | 0.95113 | 0.9776651215 | 0.399364 | 8.13335751 | 0 |
| 2 | 47410137 | G | A | G | D | MSH2 | ENST00000233146 | 0.37476 | 0.01002 | 0.43577 | 0.83148 | 0.89422 | 0.6790128994 | 0.95202 | 0.9774706364 | 0.357125 | 3.582224865 | 0 |
| 2 | 47410137 | G | C | G | A | MSH2 | ENST00000233146 | 0.33823 | 0.00566 | 0.30229 | 0.49802 | 0.78967 | 0.6430177702 | 0.96872 | 0.7306233644 | 0.228831 | 4.176325754 | 0 |
| 2 | 47410137 | G | T | G | V | MSH2 | ENST00000233146 | 0.45838 | 0.00556 | 0.3179 | 0.70141 | 0.88259 | 0.7356602149 | 0.977 | 0.8249972463 | 0.40334 | 6.696563109 | 0 |
| 2 | 47410139 | A | C | N | H | MSH2 | ENST00000233146 | 0.68155 | 0.01492 | 0.23188 | 0.80395 | 0.86603 | 0.5013013352 | 0.94629 | 0.9051929712 | 0.158493 | 7.715262271 | 0 |
| 2 | 47410139 | A | G | N | D | MSH2 | ENST00000233146 | 0.54875 | 0.00715 | 0.2763 | 0.66056 | 0.76816 | 0.4283055433 | 0.93143 | 0.7657676935 | 0.139576 | 3.325023271 | 0 |
| 2 | 47410139 | A | T | N | Y | MSH2 | ENST00000233146 | 0.73893 | 0.01611 | 0.26848 | 0.86365 | 0.9014 | 0.5767897532 | 0.9421 | 0.9798761606 | 0.274791 | 7.715262271 | 0 |
| 2 | 47410140 | A | C | N | T | MSH2 | ENST00000233146 | 0.50172 | 0.00741 | 0.21224 | 0.71757 | 0.84213 | 0.467668503 | 0.95042 | 0.8498336077 | 0.193198 | 4.759443462 | 0 |
| 2 | 47410140 | A | G | N | S | MSH2 | ENST00000233146 | 0.31364 | 0.00615 | 0.17675 | 0.45064 | 0.64571 | 0.3570532195 | 0.90174 | 0.05308916181 | 0.0739642 | 1.657348554 | 0 |
| 2 | 47410140 | A | T | N | I | MSH2 | ENST00000233146 | 0.67613 | 0.01221 | 0.24352 | 0.74141 | 0.88684 | 0.5949575898 | 0.95514 | 0.9809216857 | 0.264203 | 7.715262271 | 0 |
| 2 | 47410141 | C | A | N | K | MSH2 | ENST00000233146 | 0.36833 | 0.0071 | 0.30795 | 0.70995 | 0.84482 | 0.5158987144 | 0.9193 | 0.9452608824 | 0.151161 | 5.26795082 | 0 |
| 2 | 47410141 | C | G | N | K | MSH2 | ENST00000233146 | 0.37431 | 0.0071 | 0.30795 | 0.71533 | 0.84428 | 0.5158987144 | 0.92102 | 0.9335402846 | 0.151209 | 5.26795082 | 0 |
| 2 | 47410142 | A | C | N | H | MSH2 | ENST00000233146 | 0.35058 | 0.00569 | 0.19672 | 0.61542 | 0.74306 | 0.4530701736 | 0.94649 | 0.489244163 | 0.111358 | 2.890988196 | 0 |
| 2 | 47410142 | A | G | N | D | MSH2 | ENST00000233146 | 0.36816 | 0.00547 | 0.2326 | 0.61302 | 0.73949 | 0.4131971762 | 0.9517 | 0.5814990997 | 0.109043 | 2.981371867 | 0 |
| 2 | 47410142 | A | T | N | Y | MSH2 | ENST00000233146 | 0.38771 | 0.00726 | 0.2265 | 0.73362 | 0.83888 | 0.5297853842 | 0.95587 | 0.7777859569 | 0.235431 | 6.807774463 | 0 |
| 2 | 47410143 | A | C | N | T | MSH2 | ENST00000233146 | 0.29168 | 0.00569 | 0.17745 | 0.64555 | 0.80417 | 0.399300605 | 0.94494 | 0.1191793159 | 0.121696 | 2.12021765 | 0 |
| 2 | 47410143 | A | G | N | S | MSH2 | ENST00000233146 | 0.23123 | 0.00608 | 0.14262 | 0.54662 | 0.71861 | 0.2750788922 | 0.87002 | 0.0505444482 | 0.0708405 | 1.171725713 | 0 |
| 2 | 47410143 | A | T | N | I | MSH2 | ENST00000233146 | 0.34255 | 0.00659 | 0.19727 | 0.75041 | 0.81065 | 0.5582783431 | 0.97628 | 0.5706266165 | 0.198431 | 6.807774463 | 0 |
| 2 | 47410144 | T | A | N | K | MSH2 | ENST00000233146 | 0.27355 | 0.0058 | 0.25771 | 0.74825 | 0.70427 | 0.45946003 | 0.91152 | 0.09042663341 | 0.0789557 | 3.668562821 | 0 |
| 2 | 47410144 | T | G | N | K | MSH2 | ENST00000233146 | 0.27841 | 0.0058 | 0.25771 | 0.71584 | 0.70508 | 0.45946003 | 0.91477 | 0.09145660108 | 0.0789704 | 3.668562821 | 0 |
| 2 | 47410145 | G | A | D | N | MSH2 | ENST00000233146 | 0.69909 | 0.00659 | 0.41638 | 0.60169 | 0.79456 | 0.602394513 | 0.88529 | 0.6965873837 | 0.14863 | 3.921847604 | 0 |
| 2 | 47410145 | G | C | D | H | MSH2 | ENST00000233146 | 0.721 | 0.0103 | 0.44262 | 0.67493 | 0.88354 | 0.7119140353 | 0.92668 | 0.7965798378 | 0.301177 | 6.120374958 | 0 |
| 2 | 47410145 | G | T | D | Y | MSH2 | ENST00000233146 | 0.73861 | 0.01164 | 0.42206 | 0.70854 | 0.90096 | 0.7691465647 | 0.97436 | 0.9033248425 | 0.414721 | 7.686752425 | 0 |
| 2 | 47410146 | A | C | D | A | MSH2 | ENST00000233146 | 0.43847 | 0.00586 | 0.35151 | 0.51215 | 0.84321 | 0.7074623418 | 0.98052 | 0.6147221327 | 0.284093 | 4.759335266 | 0 |
| 2 | 47410146 | A | G | D | G | MSH2 | ENST00000233146 | 0.44217 | 0.00527 | 0.36091 | 0.46536 | 0.80713 | 0.6952874942 | 0.98495 | 0.6859399676 | 0.233232 | 3.271548643 | 0 |
| 2 | 47410146 | A | T | D | V | MSH2 | ENST00000233146 | 0.54562 | 0.0063 | 0.34227 | 0.69427 | 0.89147 | 0.7502666141 | 0.98559 | 0.7649704814 | 0.383465 | 7.686752425 | 0 |
| 2 | 47410147 | T | A | D | E | MSH2 | ENST00000233146 | 0.08939 | 0.0058 | 0.38613 | 0.44001 | 0.79334 | 0.5138717283 | 0.97845 | 0.09590527415 | 0.132656 | 2.300897636 | 0 |
| 2 | 47410147 | T | G | D | E | MSH2 | ENST00000233146 | 0.09241 | 0.0058 | 0.38613 | 0.53573 | 0.79334 | 0.5138717283 | 0.97985 | 0.09590527415 | 0.132574 | 2.300897636 | 0 |
| 2 | 47410148 | A | C | M | L | MSH2 | ENST00000233146 | 0.1996 | 0.0058 | 0.20153 | 0.5723 | 0.81006 | 0.4805691255 | 0.97219 | 0.0836448893 | 0.0956896 | 6.096939956 | 0 |
| 2 | 47410148 | A | G | M | V | MSH2 | ENST00000233146 | 0.18298 | 0.00569 | 0.1525 | 0.59303 | 0.8153 | 0.5340978525 | 0.98347 | 0.01359268185 | -0.127756 | 6.407205796 | 0 |
| 2 | 47410148 | A | T | M | L | MSH2 | ENST00000233146 | 0.19644 | 0.0058 | 0.20153 | 0.5723 | 0.81006 | 0.4805691255 | 0.96808 | 0.0836448893 | 0.0957258 | 6.096939956 | 0 |
| 2 | 47410149 | T | A | M | K | MSH2 | ENST00000233146 | 0.21757 | 0.00512 | 0.27326 | 0.70544 | 0.88916 | 0.7451252397 | 0.95312 | 0.1725228578 | 0.243366 | 5.20080046 | 0 |
| 2 | 47410149 | T | C | M | T | MSH2 | ENST00000233146 | 0.20636 | 0.00543 | 0.32504 | 0.64759 | 0.85996 | 0.6045732146 | 0.94229 | 0.06534262518 | 0.0326066 | 4.605870688 | 0 |
| 2 | 47410149 | T | G | M | R | MSH2 | ENST00000233146 | 0.24677 | 0.00547 | 0.25396 | 0.70717 | 0.8873 | 0.7840137046 | 0.95742 | 0.2385817766 | 0.25252 | 5.792682416 | 0 |
| 2 | 47410150 | G | A | M | I | MSH2 | ENST00000233146 | 0.28097 | 0.00551 | 0.20774 | 0.5078 | 0.79334 | 0.4887113047 | 0.9483 | 0.1982885134 | 0.130556 | 4.24591776 | 0 |
| 2 | 47410150 | G | C | M | I | MSH2 | ENST00000233146 | 0.27904 | 0.00551 | 0.20774 | 0.52952 | 0.79334 | 0.4887113047 | 0.94428 | 0.1347402202 | 0.130571 | 4.24591776 | 0 |
| 2 | 47410150 | G | T | M | I | MSH2 | ENST00000233146 | 0.27512 | 0.00551 | 0.20774 | 0.50642 | 0.79334 | 0.4887113047 | 0.94432 | 0.1340807775 | 0.130571 | 4.24591776 | 0 |
| 2 | 47410151 | T | A | S | T | MSH2 | ENST00000233146 | 0.36542 | 0.00599 | 0.39917 | 0.66222 | 0.81703 | 0.4614893021 | 0.96685 | 0.3617576957 | 0.20549 | 3.836226308 | 0 |
| 2 | 47410151 | T | C | S | P | MSH2 | ENST00000233146 | 0.44758 | 0.00535 | 0.41535 | 0.67929 | 0.862 | 0.7505246695 | 0.9694 | 0.7546336651 | 0.278334 | 6.960414901 | 0 |
| 2 | 47410151 | T | G | S | A | MSH2 | ENST00000233146 | 0.35691 | 0.00603 | 0.3777 | 0.49893 | 0.78597 | 0.3663561179 | 0.95897 | 0.09950167032 | 0.119206 | 3.003192914 | 0 |
| 2 | 47410152 | C | T | S | L | MSH2 | ENST00000233146 | 0.28481 | 0.00654 | 0.41721 | 0.64262 | 0.80772 | 0.5863744069 | 0.97459 | 0.31968382 | 0.311327 | 7.928308491 | 0 |
| 2 | 47410154 | G | A | A | T | MSH2 | ENST00000233146 | 0.10479 | 0.00574 | 0.02623 | 0.63785 | 0.83997 | 0.363994234 | 0.99244 | 0.02071684293 | 0.123121 | 0.08238228464 | 0 |
| 2 | 47410154 | G | C | A | P | MSH2 | ENST00000233146 | 0.15838 | 0.00622 | 0.05075 | 0.69676 | 0.83172 | 0.7240478865 | 0.99709 | 0.03184126297 | 0.203036 | 6.068825641 | 0 |
| 2 | 47410154 | G | T | A | S | MSH2 | ENST00000233146 | 0.09161 | 0.00586 | 0.02201 | 0.46432 | 0.81876 | 0.3490083051 | 0.98098 | 0.02035343895 | -0.000787115 | 0.5986013776 | 0 |
| 2 | 47410155 | C | A | A | D | MSH2 | ENST00000233146 | 0.10953 | 0.00504 | 0.06951 | 0.31686 | 0.77972 | 0.6198556889 | 0.99428 | 0.1120921802 | 0.305325 | 2.606773344 | 0 |
| 2 | 47410155 | C | G | A | G | MSH2 | ENST00000233146 | 0.10818 | 0.00569 | 0.02768 | 0.32163 | 0.78844 | 0.4303023158 | 0.97846 | 0.09981750427 | 0.130064 | 1.701919462 | 0 |
| 2 | 47410155 | C | T | A | V | MSH2 | ENST00000233146 | 0.09762 | 0.00574 | 0.03387 | 0.32054 | 0.76553 | 0.4255042395 | 0.98893 | 0.1100713815 | 0.1565 | 1.526342441 | 0 |
| 2 | 47410157 | T | A | S | T | MSH2 | ENST00000233146 | 0.40261 | 0.00808 | 0.30811 | 0.78742 | 0.84694 | 0.4859819958 | 0.93227 | 0.3295729389 | 0.204259 | 5.059806457 | 0 |
| 2 | 47410157 | T | C | S | P | MSH2 | ENST00000233146 | 0.62098 | 0.01533 | 0.32959 | 0.89758 | 0.8859 | 0.8492382401 | 0.97007 | 0.84516114 | 0.348712 | 6.046826852 | 0 |
| 2 | 47410157 | T | G | S | A | MSH2 | ENST00000233146 | 0.37998 | 0.00608 | 0.28993 | 0.66913 | 0.71153 | 0.3963008881 | 0.90851 | 0.2759885192 | 0.0931827 | 1.021402952 | 0 |
| 2 | 47410158 | C | A | S | Y | MSH2 | ENST00000233146 | 0.66003 | 0.01909 | 0.37484 | 0.91891 | 0.88354 | 0.6669256769 | 0.91261 | 0.9139974117 | 0.3091 | 4.278756309 | 0 |
| 2 | 47410158 | C | G | S | C | MSH2 | ENST00000233146 | 0.4001 | 0.00586 | 0.31012 | 0.82382 | 0.87781 | 0.5621065254 | 0.85154 | 0.7361415625 | 0.239189 | 4.183460232 | 0 |
| 2 | 47410158 | C | T | S | F | MSH2 | ENST00000233146 | 0.6954 | 0.01604 | 0.34615 | 0.91528 | 0.89558 | 0.6750385238 | 0.90048 | 0.9112608433 | 0.310959 | 5.909799111 | 0 |
| 2 | 47410160 | A | C | I | L | MSH2 | ENST00000233146 | 0.36514 | 0.00586 | 0.13614 | 0.57836 | 0.76223 | 0.61036505 | 0.95608 | 0.07809857279 | 0.130089 | 3.770563877 | 0 |
| 2 | 47410160 | A | G | I | V | MSH2 | ENST00000233146 | 0.30111 | 0.00599 | 0.07896 | 0.59437 | 0.73949 | 0.4619817819 | 0.93946 | 0.09759677947 | 0.0563462 | 0.5528850099 | 0 |
| 2 | 47410160 | A | T | I | F | MSH2 | ENST00000233146 | 0.3915 | 0.00654 | 0.15077 | 0.74942 | 0.85894 | 0.6672971092 | 0.98207 | 0.3732370138 | 0.210278 | 3.807587867 | 0 |
| 2 | 47410161 | T | A | I | N | MSH2 | ENST00000233146 | 0.40857 | 0.0058 | 0.22815 | 0.62508 | 0.81933 | 0.7295900944 | 0.93678 | 0.4548823833 | 0.191342 | 1.118636814 | 0 |
| 2 | 47410161 | T | C | I | T | MSH2 | ENST00000233146 | 0.3351 | 0.00535 | 0.25681 | 0.58985 | 0.77335 | 0.6271858591 | 0.99144 | 0.08213315546 | 0.247747 | 1.926898402 | 0 |
| 2 | 47410161 | T | G | I | S | MSH2 | ENST00000233146 | 0.35306 | 0.00509 | 0.16096 | 0.43511 | 0.77845 | 0.7391665131 | 0.985 | 0.3933860064 | 0.238911 | 1.953788782 | 0 |
| 2 | 47410163 | G | A | G | S | MSH2 | ENST00000233146 | 0.40716 | 0.01628 | 0.5622 | 0.78737 | 0.88022 | 0.7802160802 | 0.9541 | 0.8999641538 | 0.360569 | 3.638093062 | 0 |
| 2 | 47410163 | G | C | G | R | MSH2 | ENST00000233146 | 0.47728 | 0.03175 | 0.67535 | 0.75673 | 0.89918 | 0.9049397059 | 0.90809 | 0.9158839583 | 0.315851 | 6.744443705 | 0 |
| 2 | 47410163 | G | T | G | C | MSH2 | ENST00000233146 | 0.52872 | 0.01158 | 0.61496 | 0.73794 | 0.86302 | 0.7930941732 | 0.97293 | 0.8848048449 | 0.425107 | 4.096574061 | 0 |
| 2 | 47410164 | G | A | G | D | MSH2 | ENST00000233146 | 0.36665 | 0.03059 | 0.68768 | 0.72809 | 0.92028 | 0.8938553727 | 0.95556 | 0.7870391418 | 0.428591 | 6.744443705 | 0 |
| 2 | 47410164 | G | C | G | A | MSH2 | ENST00000233146 | 0.29198 | 0.01285 | 0.56822 | 0.63962 | 0.8356 | 0.7339993042 | 0.95002 | 0.4918465316 | 0.305663 | 5.816303371 | 0 |
| 2 | 47410164 | G | T | G | V | MSH2 | ENST00000233146 | 0.33936 | 0.01221 | 0.58072 | 0.74347 | 0.86553 | 0.7871910128 | 0.96101 | 0.3236673437 | 0.393553 | 3.67601108 | 0 |
| 2 | 47410166 | G | A | V | I | MSH2 | ENST00000233146 | 0.34762 | 0.00654 | 0.12925 | 0.54819 | 0.79938 | 0.4432917752 | 0.94698 | 0.1371573517 | 0.140149 | 3.011514202 | 0 |

| | | | | | | | | | | | | | | | | | |
|---|---|---|---|---|---|---|---|---|---|---|---|---|---|---|---|---|---|---|
| 2 | 47410166 | G | C | V | L | MSH2 | ENST00000233146 | 0.48335 | 0.00654 | 0.23838 | 0.6773 | 0.86902 | 0.6853043618 | 0.96796 | 0.6361169139 | 0.253651 | 3.106801249 | 0 |
| 2 | 47410166 | G | T | V | F | MSH2 | ENST00000233146 | 0.67414 | 0.0203 | 0.21603 | 0.84209 | 0.90798 | 0.8657140847 | 0.95964 | 0.9723286629 | 0.392733 | 6.756906514 | 0 |
| 2 | 47410167 | T | A | V | D | MSH2 | ENST00000233146 | 0.47325 | 0.02089 | 0.25488 | 0.89977 | 0.92192 | 0.9237022721 | 0.99233 | 0.9863694906 | 0.462935 | 8.363831119 | 0 |
| 2 | 47410167 | T | C | V | A | MSH2 | ENST00000233146 | 0.22902 | 0.00654 | 0.25835 | 0.68388 | 0.86703 | 0.7057529433 | 0.97111 | 0.639070034 | 0.263862 | 8.08208468 | 0 |
| 2 | 47410167 | T | G | V | G | MSH2 | ENST00000233146 | 0.32536 | 0.00627 | 0.16302 | 0.84835 | 0.86703 | 0.8939229334 | 0.97451 | 0.5540670753 | 0.232249 | 7.107453783 | 0 |
| 2 | 47410169 | G | A | V | M | MSH2 | ENST00000233146 | 0.39434 | 0.0058 | 0.35477 | 0.58429 | 0.77527 | 0.6609567184 | 0.93359 | 0.1274304688 | 0.122285 | 2.46888412 | 0 |
| 2 | 47410169 | G | C | V | L | MSH2 | ENST00000233146 | 0.33758 | 0.00574 | 0.32038 | 0.51938 | 0.79212 | 0.6094245985 | 0.96131 | 0.09118381888 | 0.171418 | 2.287902488 | 0 |
| 2 | 47410169 | G | T | V | L | MSH2 | ENST00000233146 | 0.33324 | 0.00574 | 0.32038 | 0.51938 | 0.79212 | 0.6094245985 | 0.96334 | 0.09118381888 | 0.171413 | 2.287902488 | 0 |
| 2 | 47410170 | T | A | V | E | MSH2 | ENST00000233146 | 0.81873 | 0.01221 | 0.335 | 0.83163 | 0.95462 | 0.8579388072 | 0.97588 | 0.9780368209 | 0.385283 | 7.996907335 | 1 |
| 2 | 47410170 | T | C | V | A | MSH2 | ENST00000233146 | 0.50589 | 0.00681 | 0.35248 | 0.56045 | 0.85996 | 0.6561183067 | 0.97271 | 0.860294342 | 0.252765 | 3.28851735 | 0 |
| 2 | 47410170 | T | G | V | G | MSH2 | ENST00000233146 | 0.81402 | 0.01334 | 0.22982 | 0.80108 | 0.95462 | 0.8585757451 | 0.96524 | 0.9850281477 | 0.378939 | 4.598736097 | 0 |
| 2 | 47410172 | G | A | G | S | MSH2 | ENST00000233146 | 0.57195 | 0.00769 | 0.50462 | 0.78863 | 0.7727 | 0.7614275693 | 0.91736 | 0.6630260736 | 0.24444 | 2.788236555 | 0 |
| 2 | 47410172 | G | C | G | R | MSH2 | ENST00000233146 | 0.93235 | 0.03112 | 0.63549 | 0.76878 | 0.91526 | 0.9116942881 | 0.92025 | 0.8344204812 | 0.331477 | 8.280216419 | 1 |
| 2 | 47410172 | G | T | G | C | MSH2 | ENST00000233146 | 0.94385 | 0.03436 | 0.56205 | 0.77126 | 0.90185 | 0.8172823205 | 0.88635 | 0.8405177402 | 0.304859 | 4.286131338 | 0 |
| 2 | 47410173 | G | A | G | D | MSH2 | ENST00000233146 | 0.6246 | 0.02696 | 0.64544 | 0.74322 | 0.94243 | 0.9039036283 | 0.96146 | 0.9515238045 | 0.418312 | 8.280216419 | 1 |
| 2 | 47410173 | G | C | G | A | MSH2 | ENST00000233146 | 0.2372 | 0.00603 | 0.5095 | 0.46619 | 0.81876 | 0.7336755885 | 0.89205 | 0.4244464338 | 0.169292 | 1.450066131 | 0 |
| 2 | 47410173 | G | T | G | V | MSH2 | ENST00000233146 | 0.59262 | 0.02349 | 0.52355 | 0.68806 | 0.89193 | 0.844581854 | 0.86262 | 0.8934476972 | 0.278626 | 5.203479848 | 0 |
| 2 | 47410175 | G | A | V | I | MSH2 | ENST00000233146 | 0.10357 | 0.00615 | 0.06817 | 0.84048 | 0.79456 | 0.3203917925 | 0.90464 | 0.04986201776 | 0.0828926 | 0.6789506529 | 0 |
| 2 | 47410175 | G | C | V | L | MSH2 | ENST00000233146 | 0.13266 | 0.00592 | 0.13127 | 0.82009 | 0.862 | 0.5947836891 | 0.96501 | 0.1609661497 | 0.177847 | 2.76092462 | 0 |
| 2 | 47410175 | G | T | V | F | MSH2 | ENST00000233146 | 0.20845 | 0.00638 | 0.12501 | 0.84115 | 0.84267 | 0.8362130448 | 0.90272 | 0.7983508706 | 0.227513 | 4.849610009 | 0 |
| 2 | 47410176 | T | A | V | D | MSH2 | ENST00000233146 | 0.67637 | 0.02218 | 0.15475 | 0.91823 | 0.9211 | 0.92363215 | 0.93761 | 0.9896514416 | 0.408491 | 8.140526812 | 1 |
| 2 | 47410176 | T | C | V | A | MSH2 | ENST00000233146 | 0.37947 | 0.01082 | 0.15834 | 0.82264 | 0.87 | 0.6906568737 | 0.90382 | 0.9490846395 | 0.337128 | 4.519154823 | 0 |
| 2 | 47410176 | T | G | V | G | MSH2 | ENST00000233146 | 0.66138 | 0.02436 | 0.097 | 0.92112 | 0.91143 | 0.8923450207 | 0.89714 | 0.9846182466 | 0.37693 | 8.140526812 | 1 |
| 2 | 47410178 | A | C | K | Q | MSH2 | ENST00000233146 | 0.78423 | 0.01126 | 0.42559 | 0.67693 | 0.8517 | 0.6715642846 | 0.90491 | 0.9110949636 | 0.244702 | 6.38210181 | 0 |
| 2 | 47410179 | A | C | K | T | MSH2 | ENST00000233146 | 0.48196 | 0.00877 | 0.48959 | 0.78512 | 0.83394 | 0.6831781003 | 0.87049 | 0.8205036521 | 0.24559 | 4.049005016 | 0 |
| 2 | 47410179 | A | G | K | R | MSH2 | ENST00000233146 | 0.33887 | 0.00633 | 0.39143 | 0.63806 | 0.7816 | 0.6112484206 | 0.93763 | 0.440874666 | 0.21188 | 2.838335826 | 0 |
| 2 | 47410179 | A | T | K | I | MSH2 | ENST00000233146 | 0.6689 | 0.02315 | 0.43836 | 0.80805 | 0.89285 | 0.7078815183 | 0.87693 | 0.9850569367 | 0.279049 | 4.379838276 | 0 |
| 2 | 47410180 | A | C | K | N | MSH2 | ENST00000233146 | 0.58481 | 0.02403 | 0.50428 | 0.63632 | 0.81703 | 0.7041544154 | 0.94413 | 0.935634315 | 0.155402 | 4.775094447 | 0 |
| 2 | 47410180 | A | T | K | N | MSH2 | ENST00000233146 | 0.57879 | 0.02403 | 0.50428 | 0.80623 | 0.81588 | 0.7041544154 | 0.94361 | 0.935634315 | 0.155463 | 4.775094447 | 0 |
| 2 | 47410181 | A | C | M | L | MSH2 | ENST00000233146 | 0.27175 | 0.0058 | 0.35604 | 0.70316 | 0.79212 | 0.3587854091 | 0.91704 | 0.05038524743 | 0.0722314 | -1.914732345 | 0 |
| 2 | 47410181 | A | G | M | V | MSH2 | ENST00000233146 | 0.30294 | 0.00569 | 0.29763 | 0.65089 | 0.81818 | 0.3977399336 | 0.98034 | 0.06422669118 | 0.1356 | 0.2370010444 | 0 |
| 2 | 47410181 | A | T | M | L | MSH2 | ENST00000233146 | 0.26781 | 0.0058 | 0.35604 | 0.70316 | 0.79151 | 0.3587854091 | 0.92646 | 0.05038524743 | 0.0722809 | -1.914732345 | 0 |
| 2 | 47410182 | T | A | M | K | MSH2 | ENST00000233146 | 0.44863 | 0.00543 | 0.43488 | 0.78694 | 0.79998 | 0.7403044825 | 0.92195 | 0.4048324444 | 0.179104 | 2.637771609 | 0 |
| 2 | 47410182 | T | C | M | T | MSH2 | ENST00000233146 | 0.40628 | 0.00547 | 0.48786 | 0.66218 | 0.78286 | 0.5635728789 | 0.94879 | 0.5604078174 | 0.159435 | 2.328073997 | 0 |
| 2 | 47410182 | T | G | M | R | MSH2 | ENST00000233146 | 0.46432 | 0.00615 | 0.41619 | 0.70771 | 0.79028 | 0.7784705878 | 0.9126 | 0.3627257645 | 0.177762 | 2.043433879 | 0 |
| 2 | 47410183 | G | A | M | I | MSH2 | ENST00000233146 | 0.13672 | 0.00551 | 0.38451 | 0.52206 | 0.76223 | 0.3285603571 | 0.93561 | 0.0400274992 | 0.0449645 | -0.04593938518 | 0 |
| 2 | 47410183 | G | C | M | I | MSH2 | ENST00000233146 | 0.13542 | 0.00551 | 0.38451 | 0.5333 | 0.76223 | 0.3285603571 | 0.93303 | 0.0291709397 | 0.044975 | -0.04593938518 | 0 |
| 2 | 47410183 | G | T | M | I | MSH2 | ENST00000233146 | 0.13275 | 0.00551 | 0.38451 | 0.52125 | 0.76223 | 0.3285603571 | 0.93265 | 0.0400274992 | 0.0449764 | -0.04593938518 | 0 |
| 2 | 47410184 | T | A | S | T | MSH2 | ENST00000233146 | 0.22257 | 0.00592 | 0.37096 | 0.64581 | 0.73443 | 0.4403088314 | 0.94518 | 0.1375659555 | 0.119164 | 2.359104517 | 0 |
| 2 | 47410184 | T | C | S | P | MSH2 | ENST00000233146 | 0.32304 | 0.01121 | 0.38535 | 0.8012 | 0.86047 | 0.7576071676 | 0.95588 | 0.7160737514 | 0.256766 | 5.173068708 | 0 |
| 2 | 47410184 | T | G | S | A | MSH2 | ENST00000233146 | 0.23065 | 0.00608 | 0.3567 | 0.62281 | 0.69678 | 0.3933407354 | 0.92515 | 0.1498842537 | 0.0805423 | 1.694604294 | 0 |
| 2 | 47410185 | C | A | S | Y | MSH2 | ENST00000233146 | 0.24681 | 0.00543 | 0.43604 | 0.81641 | 0.84106 | 0.5977427539 | 0.95162 | 0.9008414149 | 0.262536 | 4.052842417 | 0 |
| 2 | 47410185 | C | G | S | C | MSH2 | ENST00000233146 | 0.28588 | 0.01871 | 0.37531 | 0.79005 | 0.82668 | 0.5498496299 | 0.86711 | 0.8893690705 | 0.217062 | 4.604411888 | 0 |
| 2 | 47410185 | C | T | S | F | MSH2 | ENST00000233146 | 0.20283 | 0.00763 | 0.41645 | 0.75684 | 0.81065 | 0.5997017524 | 0.95418 | 0.6162106991 | 0.193186 | 2.879311362 | 0 |
| 2 | 47410187 | G | A | A | T | MSH2 | ENST00000233146 | 0.02819 | 0.00574 | 0.03349 | 0.42951 | 0.83394 | 0.2863933807 | 0.9865 | 0.005297055759 | 0.0449744 | -3.113611108 | 0 |
| 2 | 47410187 | G | C | A | P | MSH2 | ENST00000233146 | 0.0749 | 0.00909 | 0.03344 | 0.68086 | 0.86852 | 0.5105380913 | 0.9698 | 0.09152719378 | 0.102978 | 0.9286846218 | 0 |
| 2 | 47410187 | G | T | A | S | MSH2 | ENST00000233146 | 0.02758 | 0.00586 | 0.02942 | 0.58114 | 0.8367 | 0.2873529188 | 0.9577 | 0.02684035152 | 0.0450968 | -1.677885345 | 0 |
| 2 | 47410188 | C | A | A | E | MSH2 | ENST00000233146 | 0.03122 | 0.00619 | 0.038 | 0.45207 | 0.8345 | 0.3692040367 | 0.92683 | 0.05814756752 | 0.109136 | 0.3794568398 | 0 |
| 2 | 47410188 | C | G | A | G | MSH2 | ENST00000233146 | 0.03272 | 0.00569 | 0.03065 | 0.22457 | 0.82892 | 0.2918395769 | 0.96552 | 0.02609111182 | 0.06891 | -1.000739015 | 0 |
| 2 | 47410188 | C | T | A | V | MSH2 | ENST00000233146 | 0.03774 | 0.00574 | 0.03226 | 0.31668 | 0.84747 | 0.3243006318 | 0.9915 | 0.01953214116 | 0.102023 | 0.003318754012 | 0 |
| 2 | 47410190 | G | A | V | I | MSH2 | ENST00000233146 | 0.20184 | 0.00608 | 0.22901 | 0.5467 | 0.75957 | 0.3313103225 | 0.82741 | 0.3695626557 | 0.116993 | 4.690647091 | 0 |
| 2 | 47410190 | G | C | V | L | MSH2 | ENST00000233146 | 0.21083 | 0.00596 | 0.31228 | 0.65748 | 0.80536 | 0.4359975612 | 0.91531 | 0.4687172472 | 0.18407 | 2.690833547 | 0 |
| 2 | 47410190 | G | T | V | F | MSH2 | ENST00000233146 | 0.4471 | 0.00722 | 0.30987 | 0.83815 | 0.84694 | 0.5751377386 | 0.84332 | 0.7637591958 | 0.20283 | 2.902781507 | 0 |
| 2 | 47410191 | T | A | V | D | MSH2 | ENST00000233146 | 0.13295 | 0.00492 | 0.34292 | 0.68784 | 0.79456 | 0.6130735922 | 0.98724 | 0.1272293174 | 0.226894 | 0.7186672462 | 0 |
| 2 | 47410191 | T | C | V | A | MSH2 | ENST00000233146 | 0.10555 | 0.00535 | 0.32895 | 0.47205 | 0.79212 | 0.4038422095 | 0.93024 | 0.04767745701 | 0.099021 | -0.2216290484 | 0 |
| 2 | 47410191 | T | G | V | G | MSH2 | ENST00000233146 | 0.14295 | 0.00527 | 0.25361 | 0.77727 | 0.83172 | 0.4883696178 | 0.95125 | 0.2270772755 | 0.214678 | 2.085145073 | 0 |
| 2 | 47410193 | G | A | D | N | MSH2 | ENST00000233146 | 0.43406 | 0.00654 | 0.48577 | 0.74333 | 0.74446 | 0.5679648336 | 0.83743 | 0.7036201379 | 0.104978 | 2.476462981 | 0 |
| 2 | 47410193 | G | C | D | H | MSH2 | ENST00000233146 | 0.76491 | 0.03112 | 0.50946 | 0.73503 | 0.8873 | 0.6976529998 | 0.87801 | 0.902544525 | 0.338066 | 5.69206831 | 0 |
| 2 | 47410193 | G | T | D | Y | MSH2 | ENST00000233146 | 0.779 | 0.03166 | 0.49204 | 0.77658 | 0.94802 | 0.745813034 | 0.96146 | 0.9881770142 | 0.434772 | 6.736315986 | 0 |
| 2 | 47410194 | A | C | D | A | MSH2 | ENST00000233146 | 0.44174 | 0.00849 | 0.4285 | 0.62787 | 0.86403 | 0.702213994 | 0.88046 | 0.669793129 | 0.255919 | 3.980337889 | 0 |
| 2 | 47410194 | A | G | D | G | MSH2 | ENST00000233146 | 0.43022 | 0.00562 | 0.43942 | 0.53444 | 0.78223 | 0.6623526055 | 0.90089 | 0.6595653296 | 0.207832 | 2.721945125 | 0 |
| 2 | 47410194 | A | T | D | V | MSH2 | ENST00000233146 | 0.50929 | 0.02231 | 0.41874 | 0.78314 | 0.95133 | 0.7377879224 | 0.95475 | 0.9344173074 | 0.412708 | 5.2460185 | 0 |
| 2 | 47410195 | T | A | D | E | MSH2 | ENST00000233146 | 0.24111 | 0.00654 | 0.45814 | 0.44941 | 0.79998 | 0.4571420069 | 0.92957 | 0.08473188657 | 0.133412 | 3.069170021 | 0 |
| 2 | 47410195 | T | G | D | E | MSH2 | ENST00000233146 | 0.24559 | 0.00654 | 0.45814 | 0.45731 | 0.79998 | 0.4571420069 | 0.9303 | 0.08675631092 | 0.133418 | 3.069170021 | 0 |
| 2 | 47410196 | G | A | G | S | MSH2 | ENST00000233146 | 0.74563 | 0.03265 | 0.51917 | 0.59575 | 0.8859 | 0.6523576851 | 0.95303 | 0.8146086369 | 0.406358 | 5.443951479 | 0 |
| 2 | 47410196 | G | C | G | R | MSH2 | ENST00000233146 | 0.92284 | 0.03322 | 0.62319 | 0.78724 | 0.94986 | 0.7337003626 | 0.95689 | 0.9758793208 | 0.445031 | 5.816518064 | 0 |
| 2 | 47410196 | G | T | G | C | MSH2 | ENST00000233146 | 0.93707 | 0.03587 | 0.56624 | 0.74217 | 0.96627 | 0.7247335876 | 0.97354 | 0.9936429896 | 0.466367 | 7.595920561 | 0 |
| 2 | 47410197 | G | C | G | A | MSH2 | ENST00000233146 | 0.57603 | 0.02775 | 0.51416 | 0.67442 | 0.92069 | 0.6383035319 | 0.95274 | 0.7358336449 | 0.42105 | 4.422987627 | 0 |
| 2 | 47410197 | G | T | G | V | MSH2 | ENST00000233146 | 0.63643 | 0.03313 | 0.51931 | 0.8315 | 0.95681 | 0.7593164422 | 0.95376 | 0.9914180159 | 0.454251 | 6.169558398 | 0 |
| 2 | 47410199 | C | A | Q | K | MSH2 | ENST00000233146 | 0.58753 | 0.02143 | 0.55209 | 0.7568 | 0.85792 | 0.7372654769 | 0.91373 | 0.8670170148 | 0.328174 | 4.519232487 | 0 |
| 2 | 47410199 | C | G | Q | E | MSH2 | ENST00000233146 | 0.31857 | 0.00735 | 0.50037 | 0.71389 | 0.78223 | 0.742730567 | 0.90936 | 0.4708183408 | 0.229174 | 3.921205477 | 0 |
| 2 | 47410200 | A | C | Q | P | MSH2 | ENST00000233146 | 0.82545 | 0.03212 | 0.50842 | 0.86952 | 0.94394 | 0.8840737931 | 0.93411 | 0.9831238985 | 0.41769 | 5.419770495 | 0 |
| 2 | 47410200 | A | G | Q | R | MSH2 | ENST00000233146 | 0.60779 | 0.01597 | 0.54515 | 0.61798 | 0.83943 | 0.7909536174 | 0.89388 | 0.9349377155 | 0.188602 | 5.077137755 | 0 |
| 2 | 47410200 | A | T | Q | L | MSH2 | ENST00000233146 | 0.80803 | 0.02984 | 0.48456 | 0.74537 | 0.93824 | 0.7366402581 | 0.92339 | 0.9860723019 | 0.280776 | 6.711091076 | 0 |
| 2 | 47410201 | G | C | Q | H | MSH2 | ENST00000233146 | 0.30172 | 0.01015 | 0.54423 | 0.59082 | 0.83997 | 0.7957763989 | 0.96477 | 0.9286979437 | 0.208123 | 4.856874451 | 0 |
| 2 | 47410201 | G | T | Q | H | MSH2 | ENST00000233146 | 0.29758 | 0.01015 | 0.54423 | 0.60708 | 0.83997 | 0.7957763989 | 0.96335 | 0.9286979437 | 0.208132 | 4.856874451 | 0 |
| 2 | 47410202 | A | G | R | G | MSH2 | ENST00000233146 | 0.79856 | 0.0175 | 0.47267 | 0.88634 | 0.96444 | 0.8343750937 | 0.96962 | 0.9962816834 | 0.426133 | 4.043253512 | 0 |
| 2 | 47410203 | G | A | R | K | MSH2 | ENST00000233146 | 0.35217 | 0.00644 | 0.5508 | 0.70063 | 0.83228 | 0.7368887565 | 0.93203 | 0.8259316683 | 0.228636 | 2.080220048 | 0 |
| 2 | 47410203 | G | C | R | T | MSH2 | ENST00000233146 | 0.55052 | 0.00664 | 0.54019 | 0.83828 | 0.96117 | 0.7764628261 | 0.95718 | 0.9872760177 | 0.421169 | 3.709414328 | 0 |
| 2 | 47410203 | G | T | R | I | MSH2 | ENST00000233146 | 0.69732 | 0.02237 | 0.51484 | 0.85312 | 0.96334 | 0.7370673392 | 0.94856 | 0.998657465 | 0.439809 | 5.518927372 | 0 |
| 2 | 47410204 | A | C | R | S | MSH2 | ENST00000233146 | 0.3269 | 0.00881 | 0.48745 | 0.85067 | 0.87974 | 0.7910028626 | 0.9639 | 0.9553476572 | 0.310971 | 4.47272708 | 0 |
| 2 | 47410204 | A | T | R | S | MSH2 | ENST00000233146 | 0.32241 | 0.00881 | 0.48745 | 0.85067 | 0.87974 | 0.7910028626 | 0.96356 | 0.9553476572 | 0.311003 | 4.47272708 | 0 |
| 2 | 47410205 | C | G | Q | E | MSH2 | ENST00000233146 | 0.4001 | 0.00619 | 0.21124 | 0.65678 | 0.77591 | 0.6259168445 | 0.96519 | 0.1063810959 | 0.119524 | 1.990894616 | 0 |
| 2 | 47410206 | A | C | Q | P | MSH2 | ENST00000233146 | 0.40834 | 0.00783 | 0.20858 | 0.77769 | 0.8199 | 0.8871391703 | 0.98776 | 0.4712482393 | 0.233606 | 3.948225986 | 1 |
| 2 | 47410206 | A | G | Q | R | MSH2 | ENST00000233146 | 0.38409 | 0.00619 | 0.23849 | 0.65794 | 0.77335 | 0.6569232237 | 0.98399 | 0.3355041146 | 0.18311 | 1.433152765 | 0 |
| 2 | 47410206 | A | T | Q | L | MSH2 | ENST00000233146 | 0.29064 | 0.00574 | 0.18944 | 0.6003 | 0.74446 | 0.5994643536 | 0.96124 | 0.4050352871 | 0.105994 | -0.528670528 | 0 |
| 2 | 47410207 | G | C | Q | H | MSH2 | ENST00000233146 | 0.31779 | 0.00586 | 0.23781 | 0.4407 | 0.81006 | 0.6186405879 | 0.98507 | 0.1147821248 | 0.124924 | 2.062004022 | 0 |
| 2 | 47410207 | G | T | Q | H | MSH2 | ENST00000233146 | 0.31353 | 0.00586 | 0.23781 | 0.44919 | 0.81006 | 0.6186405879 | 0.9864 | 0.1147821248 | 0.12491 | 2.062004022 | 0 |
| 2 | 47410208 | G | A | V | I | MSH2 | ENST00000233146 | 0.40687 | 0.00872 | 0.09214 | 0.59308 | 0.77591 | 0.4334357081 | 0.88602 | 0.04480480994 | -0.117984 | 2.934246849 | 0 |
| 2 | 47410208 | G | C | V | L | MSH2 | ENST00000233146 | 0.73389 | 0.01479 | 0.21959 | 0.75507 | 0.93123 | 0.6983248335 | 0.96591 | 0.9639702439 | 0.370809 | 3.90886501 | 0 |
| 2 | 47410208 | G | T | V | F | MSH2 | ENST00000233146 | 0.7497 | 0.03137 | 0.2038 | 0.88109 | 0.95717 | 0.881377039 | 0.96524 | 0.9974648952 | 0.443012 | 4.464316023 | 0 |
| 2 | 47410209 | T | A | V | D | MSH2 | ENST00000233146 | 0.70968 | 0.0296 | 0.24477 | 0.91343 | 0.99443 | 0.9455773676 | 0.9743 | 0.9987647533 | 0.486923 | 8.664788166 | 1 |
| 2 | 47410209 | T | C | V | A | MSH2 | ENST00000233146 | 0.65359 | 0.03059 | 0.24593 | 0.87056 | 0.97848 | 0.7413617537 | 0.94236 | 0.9945241213 | 0.418231 | 4.367476191 | 0 |
| 2 | 47410209 | T | G | V | G | MSH2 | ENST00000233146 | 0.6942 | 0.03265 | 0.13061 | 0.90682 | 0.98869 | 0.920921074 | 0.97685 | 0.9976408482 | 0.486126 | 7.626110405 | 1 |
| 2 | 47410211 | G | A | G | R | MSH2 | ENST00000233146 | 0.87201 | 0.03447 | 0.80937 | 0.91277 | 0.99843 | 0.9440290488 | 0.9845 | 0.9994565845 | 0.549616 | 10.19663767 | 1 |
| 2 | 47410211 | G | C | G | R | MSH2 | ENST00000233146 | 0.86921 | 0.03447 | 0.80937 | 0.90729 | 0.99843 | 0.9440290488 | 0.98426 | 0.9994565845 | 0.549616 | 10.19663767 | 1 |
| 2 | 47410212 | G | A | G | E | MSH2 | ENST00000233146 | 0.73037 | 0.03322 | 0.78962 | 0.90409 | 0.99824 | 0.9434244178 | 0.97341 | 0.9994565845 | 0.549616 | 8.79425331 | 1 |
| 2 | 47410212 | G | C | G | A | MSH2 | ENST00000233146 | 0.69107 | 0.03595 | 0.70842 | 0.88481 | 0.98764 | 0.8629946222 | 0.98302 | 0.9985361099 | 0.52724 | 5.547305278 | 0 |
| 2 | 47410212 | G | T | G | V | MSH2 | ENST00000233146 | 0.72556 | 0.03379 | 0.71653 | 0.90111 | 0.99876 | 0.9037930607 | 0.97472 | 0.9994565845 | 0.549048 | 6.773354887 | 1 |
| 2 | 47410214 | G | A | V | I | MSH2 | ENST00000233146 | 0.43267 | 0.00633 | 0.17883 | 0.64972 | 0.79577 | 0.4334348268 | 0.79164 | 0.710080564 | 0.140622 | 2.425576424 | 0 |
| 2 | 47410214 | G | C | V | L | MSH2 | ENST00000233146 | 0.4358 | 0.00599 | 0.34233 | 0.62979 | 0.79028 | 0.7095397299 | 0.91298 | 0.8161492944 | 0.197079 | 3.73611412 | 0 |

| | | | | | | | | | | | | | | | | | | |
|---|---|---|---|---|---|---|---|---|---|---|---|---|---|---|---|---|---|---|
| 2 | 47410214 | G | T | V | F | MSH2 | ENST00000233146 | 0.82397 | 0.01953 | 0.31688 | 0.88569 | 0.95681 | 0.8834539864 | 0.92362 | 0.9934132695 | 0.389276 | 3.946189985 | 0 |
| 2 | 47410215 | T | A | V | D | MSH2 | ENST00000233146 | 0.78946 | 0.02374 | 0.37795 | 0.90722 | 0.97364 | 0.9439215659 | 0.99174 | 0.9987896085 | 0.484336 | 8.215918788 | 1 |
| 2 | 47410215 | T | C | V | A | MSH2 | ENST00000233146 | 0.43237 | 0.00608 | 0.37555 | 0.79308 | 0.88022 | 0.7264163254 | 0.91638 | 0.9424009919 | 0.24806 | 3.089179048 | 0 |
| 2 | 47410215 | T | G | V | G | MSH2 | ENST00000233146 | 0.76339 | 0.01699 | 0.22839 | 0.8912 | 0.9796 | 0.9157140806 | 0.94702 | 0.9956070781 | 0.32097 | 8.215918788 | 1 |
| 2 | 47410217 | G | A | G | R | MSH2 | ENST00000233146 | 0.87801 | 0.03388 | 0.80641 | 0.83541 | 0.96773 | 0.9326400453 | 0.97543 | 0.9728612304 | 0.43617 | 8.654628028 | 1 |
| 2 | 47410217 | G | C | G | R | MSH2 | ENST00000233146 | 0.87529 | 0.03388 | 0.80641 | 0.83338 | 0.96773 | 0.9326400453 | 0.97515 | 0.9728612304 | 0.43617 | 8.654628028 | 1 |
| 2 | 47410218 | G | A | G | E | MSH2 | ENST00000233146 | 0.71169 | 0.03237 | 0.78625 | 0.82147 | 0.96737 | 0.9300010331 | 0.95762 | 0.9797723787 | 0.43589 | 8.654628028 | 1 |
| 2 | 47410218 | G | C | G | A | MSH2 | ENST00000233146 | 0.29929 | 0.00799 | 0.70631 | 0.67407 | 0.81818 | 0.7852072539 | 0.9454 | 0.4492128085 | 0.356428 | 2.248001716 | 0 |
| 2 | 47410218 | G | T | G | V | MSH2 | ENST00000233146 | 0.68835 | 0.03124 | 0.71625 | 0.76293 | 0.95899 | 0.856531559 | 0.95352 | 0.9637355847 | 0.426805 | 5.023184433 | 0 |
| 2 | 47410220 | T | C | Y | H | MSH2 | ENST00000233146 | 0.86608 | 0.02824 | 0.78221 | 0.7728 | 0.95935 | 0.7911755101 | 0.88405 | 0.9958375692 | 0.327213 | 6.135495696 | 0 |
| 2 | 47410220 | T | G | Y | D | MSH2 | ENST00000233146 | 0.69508 | 0.00562 | 0.7649 | 0.81212 | 0.93977 | 0.8918494563 | 0.96863 | 0.9899191856 | 0.436722 | 8.673892005 | 1 |
| 2 | 47410221 | A | C | Y | S | MSH2 | ENST00000233146 | 0.67684 | 0.01711 | 0.71033 | 0.84605 | 0.85843 | 0.8508023817 | 0.91024 | 0.9755445123 | 0.254185 | 5.720683488 | 0 |
| 2 | 47410221 | A | G | Y | C | MSH2 | ENST00000233146 | 0.55285 | 0.03023 | 0.77843 | 0.80545 | 0.89694 | 0.7861635421 | 0.92939 | 0.9805263877 | 0.363026 | 5.621882842 | 0 |
| 2 | 47410221 | A | T | Y | F | MSH2 | ENST00000233146 | 0.39568 | 0.00599 | 0.68388 | 0.37308 | 0.66733 | 0.563627123 | 0.81446 | 0.798887074 | 0.0806781 | 2.31389089 | 0 |
| 2 | 47410223 | G | A | V | M | MSH2 | ENST00000233146 | 0.82887 | 0.03484 | 0.69194 | 0.8071 | 0.92028 | 0.7559288779 | 0.94549 | 0.8743895888 | 0.417371 | 6.474914154 | 0 |
| 2 | 47410223 | G | C | V | L | MSH2 | ENST00000233146 | 0.64384 | 0.0084 | 0.65813 | 0.70581 | 0.8859 | 0.7165836691 | 0.95385 | 0.8711079359 | 0.394776 | 4.549982105 | 0 |
| 2 | 47410223 | G | T | V | L | MSH2 | ENST00000233146 | 0.63758 | 0.0084 | 0.65813 | 0.70581 | 0.8859 | 0.7165836691 | 0.95397 | 0.7597075183 | 0.394776 | 4.549982105 | 0 |
| 2 | 47410224 | T | C | V | A | MSH2 | ENST00000233146 | 0.30894 | 0.00705 | 0.67638 | 0.64181 | 0.80713 | 0.7001052493 | 0.85506 | 0.5978950262 | 0.235704 | 1.827486899 | 0 |
| 2 | 47410224 | T | G | V | G | MSH2 | ENST00000233146 | 0.4871 | 0.02349 | 0.5442 | 0.91082 | 0.95499 | 0.8939182237 | 0.97512 | 0.9814642065 | 0.449556 | 5.071887312 | 0 |
| 2 | 47410226 | G | A | D | N | MSH2 | ENST00000233146 | 0.86865 | 0.03753 | 0.67935 | 0.8059 | 0.93554 | 0.8392114535 | 0.96154 | 0.9982782602 | 0.243648 | 7.78203442 | 0 |
| 2 | 47410226 | G | C | D | H | MSH2 | ENST00000233146 | 0.86983 | 0.03465 | 0.70328 | 0.88696 | 0.99471 | 0.8630862959 | 0.9878 | 0.7632547119 | 0.355994 | 8.914987945 | 0 |
| 2 | 47410226 | G | T | D | Y | MSH2 | ENST00000233146 | 0.87676 | 0.03409 | 0.68623 | 0.87705 | 0.98972 | 0.9099543041 | 0.99307 | 0.9992120266 | 0.528613 | 8.914987945 | 0 |
| 2 | 47410227 | A | C | D | A | MSH2 | ENST00000233146 | 0.82076 | 0.03394 | 0.60786 | 0.94468 | 0.99326 | 0.8820740461 | 0.97916 | 0.9996658564 | 0.536104 | 8.914987945 | 0 |
| 2 | 47410227 | A | G | D | G | MSH2 | ENST00000233146 | 0.82693 | 0.03313 | 0.6261 | 0.9427 | 0.99676 | 0.8999245155 | 0.98896 | 0.9993661046 | 0.536701 | 8.914987945 | 0 |
| 2 | 47410227 | A | T | D | V | MSH2 | ENST00000233146 | 0.81261 | 0.03189 | 0.60571 | 0.94601 | 0.99628 | 0.8885468455 | 0.99193 | 0.9996905327 | 0.528542 | 8.914987945 | 0 |
| 2 | 47410229 | T | A | S | T | MSH2 | ENST00000233146 | 0.19135 | 0.00586 | 0.33697 | 0.42902 | 0.76024 | 0.5768567217 | 0.94686 | 0.1834745109 | 0.0860824 | 2.058043394 | 0 |
| 2 | 47410229 | T | C | S | P | MSH2 | ENST00000233146 | 0.54149 | 0.00996 | 0.34835 | 0.81826 | 0.89558 | 0.7865582337 | 0.96562 | 0.7145303488 | 0.222132 | 5.143210522 | 0 |
| 2 | 47410229 | T | G | S | A | MSH2 | ENST00000233146 | 0.19918 | 0.00603 | 0.32964 | 0.51659 | 0.7285 | 0.4659627125 | 0.9075 | 0.04958912964 | 0.0611665 | 0.8926639157 | 0 |
| 2 | 47410230 | C | A | S | Y | MSH2 | ENST00000233146 | 0.29597 | 0.00923 | 0.40386 | 0.71843 | 0.8356 | 0.6998777793 | 0.92522 | 0.7749040164 | 0.259511 | 5.051766105 | 1 |
| 2 | 47410230 | C | G | S | C | MSH2 | ENST00000233146 | 0.43762 | 0.02043 | 0.35379 | 0.7168 | 0.83505 | 0.6336395029 | 0.79047 | 0.8052338292 | 0.206414 | 4.133338622 | 0 |
| 2 | 47410230 | C | T | S | F | MSH2 | ENST00000233146 | 0.22443 | 0.00566 | 0.38416 | 0.56409 | 0.81414 | 0.7207193724 | 0.96136 | 0.5651139617 | 0.22365 | 4.465354977 | 0 |
| 2 | 47410232 | A | C | I | L | MSH2 | ENST00000233146 | 0.2039 | 0.00574 | 0.18384 | 0.44345 | 0.73589 | 0.371093161 | 0.96814 | 0.05562121921 | 0.0703692 | 0.8664199512 | 0 |
| 2 | 47410232 | A | G | I | V | MSH2 | ENST00000233146 | 0.1638 | 0.00638 | 0.144 | 0.50875 | 0.7285 | 0.3244448008 | 0.9626 | 0.010076195 | -0.254834 | 4.352542277 | 0 |
| 2 | 47410232 | A | T | I | L | MSH2 | ENST00000233146 | 0.20066 | 0.00574 | 0.18384 | 0.44345 | 0.73589 | 0.371093161 | 0.96771 | 0.04871288912 | 0.0703615 | 0.8664199512 | 0 |
| 2 | 47410233 | T | C | I | T | MSH2 | ENST00000233146 | 0.04917 | 0.00527 | 0.2349 | 0.44583 | 0.77845 | 0.4004972252 | 0.93805 | 0.04141783714 | 0.118656 | -1.832838006 | 0 |
| 2 | 47410233 | T | G | I | R | MSH2 | ENST00000233146 | 0.09208 | 0.0048 | 0.21949 | 0.5547 | 0.80358 | 0.6087763546 | 0.97798 | 0.07243031809 | 0.147072 | 4.488144367 | 0 |
| 2 | 47410234 | A | G | I | M | MSH2 | ENST00000233146 | 0.15277 | 0.0058 | 0.17214 | 0.61557 | 0.80118 | 0.3860119836 | 0.97593 | 0.06740892128 | 0.0456008 | 1.659936595 | 0 |
| 2 | 47410235 | C | A | Q | K | MSH2 | ENST00000233146 | 0.26753 | 0.00627 | 0.40879 | 0.58523 | 0.76553 | 0.6031169277 | 0.90846 | 0.7436513901 | 0.185474 | 3.43913869 | 0 |
| 2 | 47410235 | C | G | Q | E | MSH2 | ENST00000233146 | 0.23643 | 0.00562 | 0.34981 | 0.62629 | 0.75957 | 0.6026122606 | 0.87637 | 0.1429389909 | -0.0545731 | 2.550321632 | 0 |
| 2 | 47410236 | A | C | Q | P | MSH2 | ENST00000233146 | 0.53435 | 0.00935 | 0.3724 | 0.77516 | 0.8859 | 0.8113531603 | 0.97193 | 0.7864656448 | 0.297318 | 3.43913869 | 0 |
| 2 | 47410236 | A | G | Q | R | MSH2 | ENST00000233146 | 0.35322 | 0.00574 | 0.39582 | 0.53022 | 0.7727 | 0.6426241252 | 0.96407 | 0.1805888712 | 0.0244664 | 3.313718302 | 0 |
| 2 | 47410236 | A | T | Q | L | MSH2 | ENST00000233146 | 0.40588 | 0.00574 | 0.32521 | 0.50258 | 0.73589 | 0.5348939285 | 0.9521 | 0.5642337203 | 0.133057 | -1.283728512 | 0 |
| 2 | 47410237 | G | C | Q | H | MSH2 | ENST00000233146 | 0.3699 | 0.01705 | 0.39095 | 0.74196 | 0.9045 | 0.6604025444 | 0.89044 | 0.8881066442 | 0.195516 | 2.575973938 | 0 |
| 2 | 47410237 | G | T | Q | H | MSH2 | ENST00000233146 | 0.36531 | 0.01705 | 0.39095 | 0.74486 | 0.90362 | 0.6604025444 | 0.89036 | 0.8881066442 | 0.195533 | 2.575973938 | 0 |
| 2 | 47410238 | A | G | R | G | MSH2 | ENST00000233146 | 0.67859 | 0.02381 | 0.43805 | 0.85247 | 0.94167 | 0.7962187246 | 0.92517 | 0.9931609631 | 0.364554 | 2.873659042 | 0 |
| 2 | 47410238 | A | T | R | W | MSH2 | ENST00000233146 | 0.89012 | 0.00872 | 0.56218 | 0.83606 | 0.95426 | 0.8367611596 | 0.86364 | 0.9918016791 | 0.38843 | 7.18026599 | 0 |
| 2 | 47410239 | G | A | R | K | MSH2 | ENST00000233146 | 0.4986 | 0.02068 | 0.52078 | 0.59088 | 0.83997 | 0.802235923 | 0.90501 | 0.2657586599 | -0.0610834 | 5.091225196 | 0 |
| 2 | 47410239 | G | C | R | T | MSH2 | ENST00000233146 | 0.49395 | 0.02421 | 0.50337 | 0.81421 | 0.93397 | 0.8218424746 | 0.92686 | 0.9847177863 | 0.337906 | 7.18026599 | 0 |
| 2 | 47410239 | G | T | R | M | MSH2 | ENST00000233146 | 0.85624 | 0.03404 | 0.48546 | 0.84911 | 0.96189 | 0.8069687383 | 0.91256 | 0.9956030846 | 0.319767 | 7.18026599 | 0 |
| 2 | 47410240 | G | C | R | S | MSH2 | ENST00000233146 | 0.41991 | 0.02255 | 0.45099 | 0.75244 | 0.90842 | 0.8043480829 | 0.94607 | 0.990952611 | 0.331044 | 7.18026599 | 0 |
| 2 | 47410240 | G | T | R | S | MSH2 | ENST00000233146 | 0.41505 | 0.02255 | 0.45099 | 0.7509 | 0.90842 | 0.8043480829 | 0.94622 | 0.990952611 | 0.331043 | 7.18026599 | 0 |
| 2 | 47410241 | A | G | K | E | MSH2 | ENST00000233146 | 0.40963 | 0.00592 | 0.44127 | 0.61227 | 0.77845 | 0.7203135676 | 0.94739 | 0.8373345733 | 0.223635 | 2.070695005 | 0 |
| 2 | 47410242 | A | C | K | T | MSH2 | ENST00000233146 | 0.41866 | 0.00674 | 0.45298 | 0.69385 | 0.8199 | 0.6546685749 | 0.95342 | 0.7767741084 | 0.202361 | 3.232779976 | 0 |
| 2 | 47410242 | A | G | K | R | MSH2 | ENST00000233146 | 0.43647 | 0.0103 | 0.35794 | 0.64915 | 0.87148 | 0.6200479188 | 0.93192 | 0.7445693612 | 0.191377 | 3.003539563 | 0 |
| 2 | 47410242 | A | T | K | I | MSH2 | ENST00000233146 | 0.64291 | 0.00958 | 0.4164 | 0.81285 | 0.91652 | 0.7274310028 | 0.92474 | 0.9866135716 | 0.233087 | 6.821506223 | 0 |
| 2 | 47410243 | A | C | K | N | MSH2 | ENST00000233146 | 0.37047 | 0.00747 | 0.474 | 0.75012 | 0.78844 | 0.7022873936 | 0.90204 | 0.6694425344 | 0.111113 | 6.882166353 | 0 |
| 2 | 47410243 | A | T | K | N | MSH2 | ENST00000233146 | 0.3657 | 0.00747 | 0.474 | 0.77505 | 0.78844 | 0.7022873936 | 0.9019 | 0.6694425344 | 0.11111 | 6.882166353 | 0 |
| 2 | 47410244 | C | A | L | I | MSH2 | ENST00000233146 | 0.33419 | 0.01115 | 0.35878 | 0.71491 | 0.78906 | 0.4297322087 | 0.8295 | 0.6460142136 | 0.111436 | 4.858481599 | 0 |
| 2 | 47410244 | C | G | L | V | MSH2 | ENST00000233146 | 0.4655 | 0.01361 | 0.2939 | 0.73608 | 0.86503 | 0.5559792138 | 0.93594 | 0.8000786901 | 0.161317 | 4.67315204 | 0 |
| 2 | 47410245 | T | A | L | Q | MSH2 | ENST00000233146 | 0.79808 | 0.03503 | 0.35619 | 0.91245 | 0.97512 | 0.8677367743 | 0.98376 | 0.9973633885 | 0.474943 | 8.423575243 | 1 |
| 2 | 47410245 | T | C | L | P | MSH2 | ENST00000233146 | 0.81387 | 0.0292 | 0.49331 | 0.9225 | 0.97438 | 0.9362480839 | 0.99121 | 0.9967514277 | 0.478028 | 8.423575243 | 1 |
| 2 | 47410245 | T | G | L | R | MSH2 | ENST00000233146 | 0.80779 | 0.03019 | 0.43976 | 0.9055 | 0.97475 | 0.9211090073 | 0.99288 | 0.9973633885 | 0.474782 | 8.423575243 | 1 |
| 2 | 47410247 | G | A | G | R | MSH2 | ENST00000233146 | 0.55346 | 0.0073 | 0.65652 | 0.76559 | 0.87394 | 0.8527910843 | 0.92993 | 0.9756405354 | 0.253921 | 3.454825096 | 0 |
| 2 | 47410247 | G | C | G | R | MSH2 | ENST00000233146 | 0.55066 | 0.0073 | 0.65652 | 0.76469 | 0.87442 | 0.8527910843 | 0.92521 | 0.96706599 | 0.253934 | 3.454825096 | 0 |
| 2 | 47410248 | G | A | G | E | MSH2 | ENST00000233146 | 0.36665 | 0.00633 | 0.64041 | 0.78597 | 0.8933 | 0.8476040727 | 0.86756 | 0.9872068763 | 0.253014 | 2.816334195 | 0 |
| 2 | 47410248 | G | C | G | A | MSH2 | ENST00000233146 | 0.35168 | 0.01133 | 0.53224 | 0.73637 | 0.88354 | 0.6594068022 | 0.89773 | 0.9404962063 | 0.249945 | 3.117129637 | 0 |
| 2 | 47410248 | G | T | G | V | MSH2 | ENST00000233146 | 0.52879 | 0.00872 | 0.54809 | 0.77091 | 0.94949 | 0.7539367463 | 0.88944 | 0.9910407662 | 0.311538 | 5.006452554 | 0 |
| 2 | 47410250 | C | A | L | M | MSH2 | ENST00000233146 | 0.67406 | 0.02835 | 0.42536 | 0.76923 | 0.8306 | 0.5417263171 | 0.7856 | 0.6575073312 | 0.131507 | 3.900769394 | 0 |
| 2 | 47410250 | C | G | L | V | MSH2 | ENST00000233146 | 0.268 | 0.00574 | 0.36962 | 0.32649 | 0.71312 | 0.4023966365 | 0.88281 | 0.2812829018 | 0.0952816 | -1.512083027 | 0 |
| 2 | 47410251 | T | A | L | Q | MSH2 | ENST00000233146 | 0.73989 | 0.03155 | 0.44576 | 0.95389 | 0.95972 | 0.8062109872 | 0.94766 | 0.9914090037 | 0.322479 | 6.82544804 | 0 |
| 2 | 47410251 | T | C | L | P | MSH2 | ENST00000233146 | 0.75559 | 0.02595 | 0.5789 | 0.96071 | 0.96407 | 0.9176242195 | 0.98043 | 0.9911958178 | 0.438972 | 6.82544804 | 1 |
| 2 | 47410251 | T | G | L | R | MSH2 | ENST00000233146 | 0.74603 | 0.02556 | 0.54643 | 0.94906 | 0.9729 | 0.8732693838 | 0.98253 | 0.9965627789 | 0.438729 | 6.82544804 | 1 |
| 2 | 47410253 | T | A | C | S | MSH2 | ENST00000233146 | 0.50739 | 0.01369 | 0.74498 | 0.75205 | 0.92844 | 0.7776611677 | 0.92671 | 0.8313699961 | 0.377191 | 2.261822505 | 0 |
| 2 | 47410253 | T | C | C | R | MSH2 | ENST00000233146 | 0.91987 | 0.02741 | 0.84649 | 0.87647 | 0.96407 | 0.9191476402 | 0.96005 | 0.9949797988 | 0.431315 | 6.486913814 | 0 |
| 2 | 47410253 | T | G | C | G | MSH2 | ENST00000233146 | 0.71891 | 0.02989 | 0.71209 | 0.87101 | 0.92924 | 0.8230490238 | 0.93764 | 0.9769175053 | 0.408024 | 5.93232585 | 0 |
| 2 | 47410254 | G | A | C | Y | MSH2 | ENST00000233146 | 0.9276 | 0.02883 | 0.86105 | 0.78441 | 0.97438 | 0.8700915661 | 0.94147 | 0.9943318103 | 0.458081 | 5.325219823 | 0 |
| 2 | 47410254 | G | C | C | S | MSH2 | ENST00000233146 | 0.56013 | 0.01369 | 0.74498 | 0.68195 | 0.94053 | 0.7776611677 | 0.91841 | 0.6841517091 | 0.393451 | 2.261822505 | 0 |
| 2 | 47410254 | G | T | C | F | MSH2 | ENST00000233146 | 0.80021 | 0.03029 | 0.7582 | 0.79363 | 0.96664 | 0.8555200902 | 0.95827 | 0.9877607321 | 0.445454 | 7.069182461 | 0 |
| 2 | 47410255 | T | G | C | W | MSH2 | ENST00000233146 | 0.75472 | 0.03265 | 0.80755 | 0.92342 | 0.93475 | 0.8821209916 | 0.98027 | 0.9953672886 | 0.400469 | 8.006291126 | 1 |
| 2 | 47410256 | G | A | E | K | MSH2 | ENST00000233146 | 0.92743 | 0.029 | 0.84617 | 0.88129 | 0.98477 | 0.887779427 | 0.98765 | 0.9730624064 | 0.479454 | 6.415178276 | 0 |
| 2 | 47410256 | G | C | E | Q | MSH2 | ENST00000233146 | 0.70455 | 0.00935 | 0.73776 | 0.63438 | 0.93786 | 0.767648703 | 0.95809 | 0.6790214181 | 0.372746 | 2.665994962 | 0 |
| 2 | 47410257 | A | C | E | A | MSH2 | ENST00000233146 | 0.82248 | 0.02972 | 0.75238 | 0.88127 | 0.98799 | 0.8371265189 | 0.96616 | 0.9959776037 | 0.48459 | 6.234041904 | 0 |
| 2 | 47410257 | A | G | E | G | MSH2 | ENST00000233146 | 0.86755 | 0.03243 | 0.69616 | 0.89006 | 0.98728 | 0.8704461427 | 0.96372 | 0.997931838 | 0.49205 | 7.175080314 | 0 |
| 2 | 47410257 | A | T | E | V | MSH2 | ENST00000233146 | 0.85107 | 0.03243 | 0.72942 | 0.89837 | 0.99006 | 0.827044717 | 0.97059 | 0.9984435439 | 0.497626 | 5.403200305 | 0 |
| 2 | 47410258 | A | C | E | D | MSH2 | ENST00000233146 | 0.53839 | 0.02828 | 0.75263 | 0.80189 | 0.90007 | 0.7707004744 | 0.99018 | 0.9522730112 | 0.395147 | 5.81021223 | 0 |
| 2 | 47410258 | A | T | E | D | MSH2 | ENST00000233146 | 0.53273 | 0.02828 | 0.75263 | 0.80251 | 0.89963 | 0.7707004744 | 0.99025 | 0.9522730112 | 0.395142 | 5.81021223 | 0 |
| 2 | 47410259 | T | A | F | I | MSH2 | ENST00000233146 | 0.82599 | 0.02891 | 0.88009 | 0.89927 | 0.9703 | 0.8726433801 | 0.94706 | 0.9987523556 | 0.443994 | 4.904048723 | 0 |
| 2 | 47410259 | T | C | F | L | MSH2 | ENST00000233146 | 0.84119 | 0.02875 | 0.90202 | 0.91678 | 0.98657 | 0.847626974 | 0.95598 | 0.9985557199 | 0.486323 | 4.641468241 | 0 |
| 2 | 47410259 | T | G | F | V | MSH2 | ENST00000233146 | 0.83236 | 0.0325 | 0.86549 | 0.94906 | 0.98585 | 0.8631996453 | 0.9582 | 0.9985160232 | 0.495311 | 5.104516162 | 0 |
| 2 | 47410260 | T | A | F | Y | MSH2 | ENST00000233146 | 0.80803 | 0.03509 | 0.88682 | 0.88909 | 0.95753 | 0.7983246049 | 0.93253 | 0.9927888513 | 0.393878 | 6.806372094 | 0 |
| 2 | 47410260 | T | C | F | S | MSH2 | ENST00000233146 | 0.8894 | 0.02857 | 0.88012 | 0.94802 | 0.98834 | 0.9173497628 | 0.97291 | 0.9996283054 | 0.495719 | 6.97271074 | 1 |
| 2 | 47410260 | T | G | F | C | MSH2 | ENST00000233146 | 0.88892 | 0.0317 | 0.89964 | 0.94579 | 0.97997 | 0.9031044539 | 0.96554 | 0.9996283054 | 0.50046 | 6.956589572 | 0 |
| 2 | 47410261 | C | A | F | L | MSH2 | ENST00000233146 | 0.59233 | 0.02875 | 0.90202 | 0.89345 | 0.94949 | 0.847626974 | 0.97077 | 0.9979622364 | 0.429157 | 4.641468241 | 0 |
| 2 | 47410261 | C | G | F | L | MSH2 | ENST00000233146 | 0.6001 | 0.02875 | 0.90202 | 0.8934 | 0.94949 | 0.847626974 | 0.97056 | 0.9974888563 | 0.42916 | 4.641468241 | 0 |
| 2 | 47410262 | C | A | P | T | MSH2 | ENST00000233146 | 0.30406 | 0.00562 | 0.80333 | 0.70322 | 0.85945 | 0.596812314 | 0.87735 | 0.5911670327 | 0.197978 | 2.800068226 | 0 |
| 2 | 47410262 | C | G | P | A | MSH2 | ENST00000233146 | 0.25734 | 0.00586 | 0.7579 | 0.47914 | 0.74797 | 0.5061239239 | 0.89266 | 0.5544368029 | 0.17676 | 1.865610503 | 0 |
| 2 | 47410262 | C | T | P | S | MSH2 | ENST00000233146 | 0.28724 | 0.00574 | 0.77321 | 0.61577 | 0.83283 | 0.6007198863 | 0.89991 | 0.6039910316 | 0.195044 | 3.942100052 | 0 |
| 2 | 47410263 | C | A | P | H | MSH2 | ENST00000233146 | 0.41801 | 0.02077 | 0.82584 | 0.71605 | 0.87 | 0.6170314012 | 0.767 | 0.9755136967 | 0.252095 | 4.882595443 | 0 |
| 2 | 47410263 | C | G | P | R | MSH2 | ENST00000233146 | 0.26655 | 0.01492 | 0.8385 | 0.65882 | 0.87247 | 0.6443810062 | 0.90637 | 0.9805516005 | 0.358136 | 4.390699776 | 0 |
| 2 | 47410263 | C | T | P | L | MSH2 | ENST00000233146 | 0.25985 | 0.00705 | 0.84102 | 0.62713 | 0.77845 | 0.6370537417 | 0.87387 | 0.715796113 | 0.214904 | 4.48018607 | 0 |
| 2 | 47410265 | G | T | D | Y | MSH2 | ENST00000233146 | 0.86622 | 0.01774 | 0.84469 | 0.86352 | 0.9729 | 0.9173294505 | 0.96 | 0.9981518388 | 0.516643 | 6.799268259 | 0 |

| | | | | | | | | | | | | | | | | |
|---|---|---|---|---|---|---|---|---|---|---|---|---|---|---|---|---|
| 2 | 47410266 | A | C | D | A | MSH2 | ENST00000233146 | 0.90901 | 0.02896 | 0.77319 | 0.9108 | 0.9528 | 0.8918533672 | 0.90595 | 0.9957597852 | 0.407047 | 5.517408992 | 0 |
| 2 | 47410266 | A | G | D | G | MSH2 | ENST00000233146 | 0.91356 | 0.02955 | 0.78638 | 0.90448 | 0.94506 | 0.8773493419 | 0.89758 | 0.9969710112 | 0.342061 | 3.865160136 | 0 |
| 2 | 47410266 | A | T | D | V | MSH2 | ENST00000233146 | 0.90524 | 0.02984 | 0.76775 | 0.92045 | 0.95535 | 0.8917560749 | 0.9778 | 0.9986886382 | 0.454649 | 7.210076443 | 0 |
| 2 | 47410267 | T | A | D | E | MSH2 | ENST00000233146 | 0.48844 | 0.00849 | 0.81041 | 0.72094 | 0.8345 | 0.7733587523 | 0.91104 | 0.6578501463 | 0.165353 | 3.724447319 | 0 |
| 2 | 47410267 | T | G | D | E | MSH2 | ENST00000233146 | 0.49505 | 0.00849 | 0.81041 | 0.75 | 0.8345 | 0.7733587523 | 0.91069 | 0.6472321749 | 0.165354 | 3.724447319 | 0 |
| 2 | 47410268 | A | C | N | H | MSH2 | ENST00000233146 | 0.68563 | 0.03646 | 0.69646 | 0.85681 | 0.91694 | 0.7791766287 | 0.97474 | 0.9962251186 | 0.247148 | 7.127594454 | 0 |
| 2 | 47410268 | A | G | N | D | MSH2 | ENST00000233146 | 0.51257 | 0.02369 | 0.74542 | 0.72186 | 0.85894 | 0.7615784586 | 0.93626 | 0.9880937934 | 0.173108 | 4.330783666 | 0 |
| 2 | 47410268 | A | T | N | Y | MSH2 | ENST00000233146 | 0.73381 | 0.03398 | 0.73292 | 0.89448 | 0.93786 | 0.8223737749 | 0.96626 | 0.9977303147 | 0.407178 | 7.924892002 | 0 |
| 2 | 47410269 | A | C | N | T | MSH2 | ENST00000233146 | 0.69917 | 0.02904 | 0.66141 | 0.811 | 0.90885 | 0.7360742962 | 0.92704 | 0.9916928411 | 0.258907 | 5.110459649 | 0 |
| 2 | 47410269 | A | G | N | S | MSH2 | ENST00000233146 | 0.66224 | 0.03497 | 0.61573 | 0.8513 | 0.89558 | 0.6700921885 | 0.93469 | 0.9905313849 | 0.197392 | 4.327709112 | 0 |
| 2 | 47410269 | A | T | N | I | MSH2 | ENST00000233146 | 0.76195 | 0.03353 | 0.68386 | 0.91889 | 0.95753 | 0.8207164998 | 0.93593 | 0.9992254972 | 0.375198 | 6.18007002 | 0 |
| 2 | 47410271 | G | T | D | Y | MSH2 | ENST00000233146 | 0.8763 | 0.03362 | 0.74281 | 0.91425 | 0.95133 | 0.7522179697 | 0.96336 | 0.9867255027 | 0.388893 | 7.042050732 | 0 |
| 2 | 47410272 | A | C | D | A | MSH2 | ENST00000233146 | 0.8376 | 0.02094 | 0.65795 | 0.9393 | 0.94205 | 0.7002301525 | 0.90327 | 0.9935763478 | 0.367602 | 5.580024259 | 0 |
| 2 | 47410272 | A | G | D | G | MSH2 | ENST00000233146 | 0.87288 | 0.03083 | 0.67084 | 0.92956 | 0.96627 | 0.6994271773 | 0.94239 | 0.997931838 | 0.451516 | 5.368424293 | 0 |
| 2 | 47410272 | A | T | D | V | MSH2 | ENST00000233146 | 0.8609 | 0.03033 | 0.6559 | 0.94476 | 0.9729 | 0.73929216 | 0.95506 | 0.9981575608 | 0.485717 | 7.428484177 | 0 |
| 2 | 47410273 | T | A | D | E | MSH2 | ENST00000233146 | 0.27831 | 0.01009 | 0.69233 | 0.72759 | 0.86653 | 0.4364689024 | 0.93715 | 0.5129263401 | 0.187421 | 3.186654848 | 0 |
| 2 | 47410273 | T | G | D | E | MSH2 | ENST00000233146 | 0.2833 | 0.01009 | 0.69233 | 0.72719 | 0.86653 | 0.4364689024 | 0.93746 | 0.4469899833 | 0.187416 | 3.186654848 | 0 |
| 2 | 47410274 | C | A | Q | K | MSH2 | ENST00000233146 | 0.51113 | 0.00644 | 0.62951 | 0.68981 | 0.8933 | 0.7636556263 | 0.90288 | 0.6318512559 | 0.275438 | 5.552536635 | 0 |
| 2 | 47410274 | C | G | Q | E | MSH2 | ENST00000233146 | 0.47206 | 0.01133 | 0.57177 | 0.62606 | 0.85945 | 0.7285876595 | 0.86039 | 0.6217615008 | 0.235588 | 5.004057805 | 0 |
| 2 | 47410275 | A | C | Q | P | MSH2 | ENST00000233146 | 0.60383 | 0.02533 | 0.57537 | 0.91568 | 0.91526 | 0.8815009508 | 0.89184 | 0.9577983022 | 0.281779 | 5.41037805 | 0 |
| 2 | 47410275 | A | G | Q | R | MSH2 | ENST00000233146 | 0.53711 | 0.00681 | 0.62101 | 0.82759 | 0.90668 | 0.7748902703 | 0.94737 | 0.5490540266 | 0.147054 | 5.15123968 | 0 |
| 2 | 47410275 | A | T | Q | L | MSH2 | ENST00000233146 | 0.39052 | 0.00622 | 0.55818 | 0.72728 | 0.79212 | 0.7220543778 | 0.97305 | 0.7273999453 | 0.255722 | 4.630898613 | 0 |
| 2 | 47410276 | G | C | Q | H | MSH2 | ENST00000233146 | 0.33377 | 0.00654 | 0.61505 | 0.74069 | 0.87636 | 0.7332185003 | 0.96589 | 0.3403705657 | 0.252514 | 5.439871601 | 0 |
| 2 | 47410276 | G | T | Q | H | MSH2 | ENST00000233146 | 0.32945 | 0.00654 | 0.61505 | 0.74156 | 0.87636 | 0.7332185003 | 0.9659 | 0.3403705657 | 0.252513 | 5.439871601 | 0 |
| 2 | 47410277 | T | A | F | I | MSH2 | ENST00000233146 | 0.81904 | 0.0318 | 0.76577 | 0.9039 | 0.9659 | 0.8169468168 | 0.9263 | 0.997222662 | 0.374905 | 5.351382596 | 0 |
| 2 | 47410277 | T | C | F | L | MSH2 | ENST00000233146 | 0.80566 | 0.03304 | 0.79456 | 0.8736 | 0.92682 | 0.7599998573 | 0.85314 | 0.9906310439 | 0.269328 | 4.485109639 | 0 |
| 2 | 47410277 | T | G | F | V | MSH2 | ENST00000233146 | 0.82545 | 0.03274 | 0.74389 | 0.92421 | 0.96444 | 0.7734596664 | 0.96969 | 0.9969216055 | 0.463399 | 5.554971043 | 0 |
| 2 | 47410278 | T | A | F | Y | MSH2 | ENST00000233146 | 0.65194 | 0.03491 | 0.77358 | 0.73088 | 0.92356 | 0.701658016 | 0.87933 | 0.885730207 | 0.264607 | 5.638353625 | 0 |
| 2 | 47410278 | T | C | F | S | MSH2 | ENST00000233146 | 0.88542 | 0.02851 | 0.76217 | 0.92705 | 0.98477 | 0.8565026398 | 0.94912 | 0.9984782338 | 0.472907 | 6.152200084 | 0 |
| 2 | 47410278 | T | G | F | C | MSH2 | ENST00000233146 | 0.88523 | 0.0317 | 0.78713 | 0.92417 | 0.9833 | 0.8170409694 | 0.95957 | 0.9985900521 | 0.478672 | 7.43565294 | 0 |
| 2 | 47410279 | C | A | F | L | MSH2 | ENST00000233146 | 0.5314 | 0.03304 | 0.79456 | 0.8119 | 0.87 | 0.7599998573 | 0.83713 | 0.9906244874 | 0.23467 | 4.485109639 | 0 |
| 2 | 47410279 | C | G | F | L | MSH2 | ENST00000233146 | 0.53853 | 0.03304 | 0.79456 | 0.81373 | 0.87 | 0.7599998573 | 0.83581 | 0.9884665012 | 0.234672 | 4.485109639 | 0 |
| 2 | 47410280 | T | A | S | T | MSH2 | ENST00000233146 | 0.55587 | 0.01924 | 0.69158 | 0.75261 | 0.86149 | 0.6637381506 | 0.95783 | 0.8715543747 | 0.27817 | 5.781214949 | 0 |
| 2 | 47410280 | T | C | S | P | MSH2 | ENST00000233146 | 0.83135 | 0.03334 | 0.71024 | 0.91976 | 0.95717 | 0.8793786691 | 0.9766 | 0.9954208732 | 0.473821 | 6.897942838 | 0 |
| 2 | 47410280 | T | G | S | A | MSH2 | ENST00000233146 | 0.74277 | 0.01812 | 0.67921 | 0.84684 | 0.86753 | 0.6032279222 | 0.93244 | 0.9148182273 | 0.260339 | 5.76464077 | 0 |
| 2 | 47410281 | C | A | S | Y | MSH2 | ENST00000233146 | 0.69756 | 0.03313 | 0.76518 | 0.92005 | 0.95899 | 0.813482658 | 0.9688 | 0.998559773 | 0.482637 | 7.814445697 | 0 |
| 2 | 47410281 | C | G | S | C | MSH2 | ENST00000233146 | 0.72188 | 0.03728 | 0.70321 | 0.87001 | 0.93397 | 0.6774063292 | 0.92386 | 0.9931894541 | 0.403865 | 8.112473259 | 0 |
| 2 | 47410281 | C | T | S | F | MSH2 | ENST00000233146 | 0.74738 | 0.03547 | 0.73808 | 0.9176 | 0.95899 | 0.8221329471 | 0.97094 | 0.9984715581 | 0.482368 | 9.661528224 | 0 |
| 2 | 47410283 | A | C | N | H | MSH2 | ENST00000233146 | 0.73653 | 0.0366 | 0.67076 | 0.90892 | 0.94318 | 0.7757981533 | 0.96564 | 0.998775661 | 0.218007 | 7.017635926 | 0 |
| 2 | 47410283 | A | G | N | D | MSH2 | ENST00000233146 | 0.72396 | 0.02914 | 0.71945 | 0.86553 | 0.88259 | 0.7871244849 | 0.95169 | 0.9932922125 | 0.177335 | 4.968146819 | 0 |
| 2 | 47410283 | A | T | N | Y | MSH2 | ENST00000233146 | 0.78431 | 0.03419 | 0.71212 | 0.94235 | 0.95426 | 0.8289273827 | 0.95152 | 0.9996371269 | 0.423053 | 8.731141995 | 0 |
| 2 | 47410284 | A | C | N | T | MSH2 | ENST00000233146 | 0.76507 | 0.03686 | 0.63611 | 0.87702 | 0.91058 | 0.722520968 | 0.94282 | 0.9974367023 | 0.212584 | 5.898853585 | 0 |
| 2 | 47410284 | A | G | N | S | MSH2 | ENST00000233146 | 0.71321 | 0.04022 | 0.58614 | 0.87226 | 0.92316 | 0.6931889125 | 0.95703 | 0.8817091584 | -0.0102075 | 5.520897524 | 0 |
| 2 | 47410284 | A | T | N | I | MSH2 | ENST00000233146 | 0.80835 | 0.03379 | 0.64997 | 0.92928 | 0.95023 | 0.8093208861 | 0.93516 | 0.9991555214 | 0.346689 | 6.614714162 | 0 |
| 2 | 47410285 | T | A | N | K | MSH2 | ENST00000233146 | 0.54562 | 0.03491 | 0.75478 | 0.88847 | 0.91442 | 0.812378524 | 0.95322 | 0.995598793 | 0.147964 | 5.001005242 | 0 |
| 2 | 47410285 | T | G | N | K | MSH2 | ENST00000233146 | 0.55264 | 0.03491 | 0.75478 | 0.8863 | 0.91442 | 0.812378524 | 0.95315 | 0.9953821301 | 0.147968 | 5.001005242 | 0 |
| 2 | 47410286 | C | A | L | I | MSH2 | ENST00000233146 | 0.55107 | 0.03006 | 0.69378 | 0.86645 | 0.92642 | 0.5926309459 | 0.83259 | 0.9435276389 | 0.271383 | 5.175779313 | 0 |
| 2 | 47410286 | C | G | L | V | MSH2 | ENST00000233146 | 0.54324 | 0.03093 | 0.65065 | 0.80789 | 0.92964 | 0.7300739535 | 0.89784 | 0.8987826109 | 0.344336 | 4.931999858 | 0 |
| 2 | 47410286 | C | T | L | F | MSH2 | ENST00000233146 | 0.43896 | 0.02225 | 0.69268 | 0.78616 | 0.91569 | 0.7594962781 | 0.87466 | 0.8344211578 | 0.262981 | 5.708150425 | 0 |
| 2 | 47410287 | T | A | L | H | MSH2 | ENST00000233146 | 0.70599 | 0.03327 | 0.77883 | 0.9468 | 0.99265 | 0.8749030967 | 0.96713 | 0.9962176547 | 0.509804 | 9.765664091 | 1 |
| 2 | 47410287 | T | C | L | P | MSH2 | ENST00000233146 | 0.73093 | 0.02994 | 0.81581 | 0.94801 | 0.99265 | 0.9533755188 | 0.97824 | 0.9959011896 | 0.504671 | 8.703907759 | 1 |
| 2 | 47410287 | T | G | L | R | MSH2 | ENST00000233146 | 0.72628 | 0.03137 | 0.78817 | 0.94488 | 0.99414 | 0.9485315299 | 0.98175 | 0.9961735547 | 0.509582 | 9.765664091 | 1 |
| 2 | 47410289 | G | A | E | K | MSH2 | ENST00000233146 | 0.88952 | 0.03518 | 0.72981 | 0.87861 | 0.97885 | 0.8981283732 | 0.97886 | 0.9954702901 | 0.47839 | 5.142437678 | 0 |
| 2 | 47410289 | G | C | E | Q | MSH2 | ENST00000233146 | 0.83197 | 0.03728 | 0.58834 | 0.85831 | 0.94506 | 0.7942304488 | 0.96341 | 0.9850116888 | 0.448845 | 4.796608678 | 0 |
| 2 | 47410290 | A | C | E | A | MSH2 | ENST00000233146 | 0.817 | 0.03602 | 0.60696 | 0.92748 | 0.98549 | 0.8535377498 | 0.93859 | 0.9963507652 | 0.457767 | 5.578584178 | 0 |
| 2 | 47410290 | A | G | E | G | MSH2 | ENST00000233146 | 0.85145 | 0.03458 | 0.547 | 0.92828 | 0.9833 | 0.847067567 | 0.95018 | 0.9967773557 | 0.484023 | 6.962722971 | 0 |
| 2 | 47410290 | A | T | E | V | MSH2 | ENST00000233146 | 0.84612 | 0.03475 | 0.57738 | 0.93681 | 0.98404 | 0.8422001536 | 0.94708 | 0.9962491979 | 0.45708 | 4.584116845 | 0 |
| 2 | 47410291 | G | C | E | D | MSH2 | ENST00000233146 | 0.68163 | 0.03783 | 0.60925 | 0.85996 | 0.93319 | 0.7836839296 | 0.97429 | 0.9809149131 | 0.432294 | 7.135826206 | 0 |
| 2 | 47410291 | G | T | E | D | MSH2 | ENST00000233146 | 0.67517 | 0.03783 | 0.60925 | 0.86899 | 0.93319 | 0.7836839296 | 0.97449 | 0.9821405486 | 0.432288 | 7.135826206 | 0 |
| 2 | 47410292 | G | A | A | T | MSH2 | ENST00000233146 | 0.83951 | 0.00763 | 0.50943 | 0.74799 | 0.91652 | 0.5858444011 | 0.89774 | 0.8321397901 | 0.213829 | 5.411321284 | 0 |
| 2 | 47410292 | G | C | A | P | MSH2 | ENST00000233146 | 0.85412 | 0.02755 | 0.59153 | 0.87372 | 0.94506 | 0.8646091107 | 0.9581 | 0.9810355339 | 0.421295 | 7.393221844 | 0 |
| 2 | 47410292 | G | T | A | S | MSH2 | ENST00000233146 | 0.5369 | 0.00735 | 0.46204 | 0.62338 | 0.82103 | 0.4875629243 | 0.91237 | 0.5831580162 | 0.16177 | 3.475823098 | 0 |
| 2 | 47410293 | C | A | A | D | MSH2 | ENST00000233146 | 0.61171 | 0.02436 | 0.64729 | 0.83239 | 0.95935 | 0.8377193481 | 0.90285 | 0.9868278744 | 0.418142 | 3.851177841 | 0 |
| 2 | 47410293 | C | G | A | G | MSH2 | ENST00000233146 | 0.43744 | 0.01676 | 0.4947 | 0.7182 | 0.88916 | 0.5656222843 | 0.83635 | 0.7029442266 | 0.203632 | 4.574921494 | 0 |
| 2 | 47410293 | C | T | A | V | MSH2 | ENST00000233146 | 0.60618 | 0.02335 | 0.53513 | 0.8009 | 0.92844 | 0.6965743973 | 0.8493 | 0.9786511731 | 0.27674 | 8.381336341 | 0 |
| 2 | 47410295 | C | A | L | I | MSH2 | ENST00000233146 | 0.43956 | 0.03413 | 0.79833 | 0.76763 | 0.85843 | 0.4516367742 | 0.78527 | 0.8988828409 | 0.221631 | 4.64446339 | 0 |
| 2 | 47410295 | C | G | L | V | MSH2 | ENST00000233146 | 0.54136 | 0.03093 | 0.73939 | 0.80748 | 0.88543 | 0.6115702221 | 0.91143 | 0.8755772039 | 0.361964 | 6.419368567 | 0 |
| 2 | 47410295 | C | T | L | F | MSH2 | ENST00000233146 | 0.55758 | 0.02225 | 0.77896 | 0.80699 | 0.88916 | 0.6755410288 | 0.92004 | 0.9514558911 | 0.364798 | 6.195633402 | 0 |
| 2 | 47410296 | T | A | L | H | MSH2 | ENST00000233146 | 0.69452 | 0.03327 | 0.86231 | 0.92215 | 0.96737 | 0.8045349972 | 0.94214 | 0.9944762903 | 0.474713 | 7.452326858 | 0 |
| 2 | 47410296 | T | C | L | P | MSH2 | ENST00000233146 | 0.71946 | 0.02994 | 0.89678 | 0.92126 | 0.97587 | 0.9295213053 | 0.97364 | 0.9928834438 | 0.475198 | 7.452326858 | 1 |
| 2 | 47410296 | T | G | L | R | MSH2 | ENST00000233146 | 0.71418 | 0.03129 | 0.86956 | 0.90097 | 0.97735 | 0.9117096703 | 0.95936 | 0.9944211841 | 0.467571 | 6.432834337 | 0 |
| 2 | 47410298 | C | A | L | I | MSH2 | ENST00000233146 | 0.23174 | 0.00603 | 0.67383 | 0.66555 | 0.78535 | 0.4985156615 | 0.92613 | 0.2382762581 | 0.175149 | 2.684615159 | 0 |
| 2 | 47410298 | C | G | L | V | MSH2 | ENST00000233146 | 0.23626 | 0.00615 | 0.60496 | 0.6995 | 0.80831 | 0.6682943111 | 0.9792 | 0.09931541979 | 0.253616 | 3.337745076 | 0 |
| 2 | 47410298 | C | T | L | F | MSH2 | ENST00000233146 | 0.51028 | 0.02206 | 0.65943 | 0.8683 | 0.93593 | 0.7619394662 | 0.96918 | 0.9459888339 | 0.329803 | 5.394778741 | 0 |
| 2 | 47410299 | T | A | L | H | MSH2 | ENST00000233146 | 0.67787 | 0.02984 | 0.76322 | 0.93329 | 0.99443 | 0.8620462799 | 0.98033 | 0.9948390126 | 0.520941 | 8.662136947 | 1 |
| 2 | 47410299 | T | C | L | P | MSH2 | ENST00000233146 | 0.69812 | 0.02683 | 0.81328 | 0.93537 | 0.99414 | 0.9471202743 | 0.99099 | 0.9882683158 | 0.515344 | 8.662136947 | 1 |
| 2 | 47410299 | T | G | L | R | MSH2 | ENST00000233146 | 0.67883 | 0.02529 | 0.77239 | 0.93237 | 0.99105 | 0.9432079785 | 0.98992 | 0.9932799935 | 0.514297 | 8.662136947 | 1 |
| 2 | 47410301 | A | C | I | L | MSH2 | ENST00000233146 | 0.41552 | 0.00586 | 0.49184 | 0.80836 | 0.77782 | 0.496282041 | 0.90148 | 0.6918572187 | 0.115022 | 2.67330586 | 0 |
| 2 | 47410301 | A | G | I | V | MSH2 | ENST00000233146 | 0.20062 | 0.00608 | 0.38607 | 0.59122 | 0.71783 | 0.2383220457 | 0.8775 | 0.6801710725 | 0.0673708 | -1.749854606 | 0 |
| 2 | 47410301 | A | T | I | F | MSH2 | ENST00000233146 | 0.46121 | 0.01303 | 0.52103 | 0.90064 | 0.90185 | 0.6691952673 | 0.92681 | 0.9566274881 | 0.11598 | 3.15894983 | 0 |
| 2 | 47410302 | T | A | I | N | MSH2 | ENST00000233146 | 0.72893 | 0.02043 | 0.60618 | 0.93298 | 0.92316 | 0.7227522112 | 0.80562 | 0.9920323323 | 0.26258 | 2.84066599 | 0 |
| 2 | 47410302 | T | C | I | T | MSH2 | ENST00000233146 | 0.51132 | 0.00855 | 0.6363 | 0.87037 | 0.88543 | 0.5124101395 | 0.84427 | 0.9426531196 | 0.263421 | 2.757465651 | 0 |
| 2 | 47410302 | T | G | I | S | MSH2 | ENST00000233146 | 0.57111 | 0.01533 | 0.51634 | 0.90173 | 0.8933 | 0.6959361099 | 0.8585 | 0.9895864725 | 0.281467 | 2.004103447 | 0 |
| 2 | 47410303 | C | G | I | M | MSH2 | ENST00000233146 | 0.40074 | 0.02011 | 0.46256 | 0.85888 | 0.8367 | 0.5087796542 | 0.83001 | 0.592293655 | 0.123396 | 2.968085632 | 0 |
| 2 | 47410304 | C | A | Q | K | MSH2 | ENST00000233146 | 0.66122 | 0.02342 | 0.67311 | 0.77904 | 0.93824 | 0.8471798009 | 0.93103 | 0.9893262124 | 0.333668 | 4.448927818 | 0 |
| 2 | 47410304 | C | G | Q | E | MSH2 | ENST00000233146 | 0.62105 | 0.01848 | 0.60776 | 0.77582 | 0.89873 | 0.8075063595 | 0.91504 | 0.9684557403 | 0.28373 | 9.326312372 | 0 |
| 2 | 47410305 | A | C | Q | P | MSH2 | ENST00000233146 | 0.88202 | 0.03083 | 0.62425 | 0.88022 | 0.94318 | 0.9335800699 | 0.96189 | 0.9969602227 | 0.470986 | 8.386593631 | 0 |
| 2 | 47410305 | A | G | Q | R | MSH2 | ENST00000233146 | 0.86361 | 0.02785 | 0.66618 | 0.8057 | 0.93593 | 0.8717124719 | 0.95709 | 0.9875938296 | 0.281588 | 4.226138481 | 0 |
| 2 | 47410305 | A | T | Q | L | MSH2 | ENST00000233146 | 0.85756 | 0.03099 | 0.59603 | 0.88234 | 0.93515 | 0.8259969203 | 0.94746 | 0.9972479939 | 0.362549 | 6.0175248 | 0 |
| 2 | 47410306 | G | C | Q | H | MSH2 | ENST00000233146 | 0.49971 | 0.00836 | 0.66006 | 0.79991 | 0.88401 | 0.8369371947 | 0.9536 | 0.9923101664 | 0.224706 | 6.903435561 | 0 |
| 2 | 47410306 | G | T | Q | H | MSH2 | ENST00000233146 | 0.49447 | 0.00836 | 0.66006 | 0.79991 | 0.88401 | 0.8369371947 | 0.95427 | 0.9923101664 | 0.224701 | 6.903435561 | 0 |
| 2 | 47410307 | A | C | I | L | MSH2 | ENST00000233146 | 0.32131 | 0.00574 | 0.46107 | 0.45252 | 0.71073 | 0.5105092272 | 0.891 | 0.1619814336 | 0.060989 | 1.055767112 | 0 |
| 2 | 47410307 | A | G | I | V | MSH2 | ENST00000233146 | 0.36794 | 0.00638 | 0.35833 | 0.53769 | 0.77718 | 0.3762355269 | 0.95602 | 0.295830071 | 0.159225 | 3.628255283 | 0 |
| 2 | 47410307 | A | T | I | F | MSH2 | ENST00000233146 | 0.59298 | 0.01115 | 0.47804 | 0.81574 | 0.89873 | 0.7025536428 | 0.94346 | 0.912763834 | 0.230727 | 3.37662145 | 0 |
| 2 | 47410308 | T | A | I | N | MSH2 | ENST00000233146 | 0.63023 | 0.01649 | 0.57266 | 0.89012 | 0.93977 | 0.8244606788 | 0.90805 | 0.992422998 | 0.283739 | 5.345511615 | 0 |
| 2 | 47410308 | T | C | I | T | MSH2 | ENST00000233146 | 0.52035 | 0.00574 | 0.60219 | 0.74837 | 0.9014 | 0.659934467 | 0.97333 | 0.5959509015 | 0.269846 | 3.461582287 | 0 |
| 2 | 47410308 | T | G | I | S | MSH2 | ENST00000233146 | 0.61858 | 0.01151 | 0.48397 | 0.88958 | 0.9579 | 0.8256251024 | 0.97254 | 0.9856019616 | 0.419776 | 4.602511455 | 0 |
| 2 | 47410309 | T | G | I | M | MSH2 | ENST00000233146 | 0.37013 | 0.00654 | 0.41406 | 0.68555 | 0.82556 | 0.5374642921 | 0.92592 | 0.163664881 | 0.130978 | 2.89748932 | 0 |
| 2 | 47410310 | G | A | G | R | MSH2 | ENST00000233146 | 0.80148 | 0.02602 | 0.79988 | 0.88814 | 0.91862 | 0.8427130144 | 0.90038 | 0.9818313718 | 0.324615 | 4.847589989 | 0 |
| 2 | 47410310 | G | C | G | R | MSH2 | ENST00000233146 | 0.79832 | 0.02602 | 0.79988 | 0.89081 | 0.91862 | 0.8427130144 | 0.89524 | 0.9833407998 | 0.324638 | 4.847589989 | 0 |
| 2 | 47410311 | G | A | G | E | MSH2 | ENST00000233146 | 0.71386 | 0.03237 | 0.78283 | 0.8645 | 0.92884 | 0.8271991454 | 0.88285 | 0.9954381916 | 0.375769 | 4.969734499 | 0 |

| | | | | | | | | | | | | | | | | | |
|---|---|---|---|---|---|---|---|---|---|---|---|---|---|---|---|---|---|
| 2 | 47410311 | G | C | G | A | MSH2 | ENST00000233146 | 0.51856 | 0.03394 | 0.69929 | 0.78555 | 0.88354 | 0.7599959188 | 0.82854 | 0.9018458128 | 0.245569 | 4.779034282 | 0 |
| 2 | 47410311 | G | T | G | V | MSH2 | ENST00000233146 | 0.70903 | 0.03327 | 0.70713 | 0.87953 | 0.93824 | 0.8627678683 | 0.89396 | 0.9964723035 | 0.409611 | 6.291268172 | 0 |
| 2 | 47410313 | C | A | P | T | MSH2 | ENST00000233146 | 0.6426 | 0.03503 | 0.66701 | 0.86315 | 0.98034 | 0.7991203642 | 0.97189 | 0.9772107139 | 0.493491 | 5.691026178 | 0 |
| 2 | 47410313 | C | G | P | A | MSH2 | ENST00000233146 | 0.48908 | 0.02691 | 0.61663 | 0.79091 | 0.95717 | 0.7306280999 | 0.94956 | 0.8583885431 | 0.430802 | 3.823249936 | 0 |
| 2 | 47410313 | C | T | P | S | MSH2 | ENST00000233146 | 0.67024 | 0.03613 | 0.63389 | 0.8975 | 0.98834 | 0.8078531142 | 0.96208 | 0.9405924678 | 0.424161 | 6.033669497 | 0 |
| 2 | 47410314 | C | A | P | Q | MSH2 | ENST00000233146 | 0.58382 | 0.03802 | 0.64095 | 0.88643 | 0.99234 | 0.8497978123 | 0.97258 | 0.9892156656 | 0.505381 | 9.504533798 | 0 |
| 2 | 47410314 | C | G | P | R | MSH2 | ENST00000233146 | 0.59769 | 0.03608 | 0.69912 | 0.8929 | 0.98693 | 0.8675192332 | 0.98071 | 0.983323703 | 0.505125 | 9.504533798 | 1 |
| 2 | 47410314 | C | T | P | L | MSH2 | ENST00000233146 | 0.61186 | 0.03595 | 0.7071 | 0.87216 | 0.97698 | 0.8413606851 | 0.98725 | 0.9717817903 | 0.376417 | 8.425411601 | 0 |
| 2 | 47410316 | A | G | K | E | MSH2 | ENST00000233146 | 0.81842 | 0.03144 | 0.60574 | 0.91021 | 0.95097 | 0.8304374592 | 0.94412 | 0.9863469886 | 0.353071 | 9.018325832 | 0 |
| 2 | 47410317 | A | C | K | T | MSH2 | ENST00000233146 | 0.78978 | 0.03426 | 0.61356 | 0.92682 | 0.96517 | 0.7952151329 | 0.91634 | 0.9970145763 | 0.394367 | 4.88837322 | 0 |
| 2 | 47410317 | A | G | K | R | MSH2 | ENST00000233146 | 0.63527 | 0.0194 | 0.51268 | 0.80166 | 0.91015 | 0.6876245745 | 0.92574 | 0.8452288332 | 0.239706 | 3.277991025 | 0 |
| 2 | 47410317 | A | T | K | M | MSH2 | ENST00000233146 | 0.82358 | 0.03269 | 0.51469 | 0.9379 | 0.96993 | 0.8195701438 | 0.91893 | 0.9959796965 | 0.355557 | 7.212039447 | 0 |
| 2 | 47410318 | G | C | K | N | MSH2 | ENST00000233146 | 0.60537 | 0.03436 | 0.64264 | 0.89918 | 0.92964 | 0.7893878095 | 0.94536 | 0.9906631708 | 0.166458 | 6.038851182 | 0 |
| 2 | 47410318 | G | T | K | N | MSH2 | ENST00000233146 | 0.59937 | 0.03436 | 0.64264 | 0.90715 | 0.92964 | 0.7893878095 | 0.9457 | 0.9906631708 | 0.166457 | 6.038851182 | 0 |
| 2 | 47410319 | G | A | E | K | MSH2 | ENST00000233146 | 0.88022 | 0.03542 | 0.7216 | 0.94265 | 0.99386 | 0.939427981 | 0.98933 | 0.9950120023 | 0.573146 | 10.19769097 | 1 |
| 2 | 47410320 | A | C | E | A | MSH2 | ENST00000233146 | 0.80882 | 0.03626 | 0.61718 | 0.96614 | 0.99806 | 0.9008769064 | 0.98236 | 0.9963507652 | 0.573779 | 10.19769097 | 1 |
| 2 | 47410320 | A | G | E | G | MSH2 | ENST00000233146 | 0.85463 | 0.03484 | 0.56412 | 0.96695 | 0.99976 | 0.9220702668 | 0.98824 | 0.9967773557 | 0.574359 | 10.19769097 | 1 |
| 2 | 47410320 | A | T | E | V | MSH2 | ENST00000233146 | 0.83744 | 0.03484 | 0.59426 | 0.9654 | 0.99919 | 0.8983643536 | 0.99232 | 0.9964768291 | 0.574691 | 10.19769097 | 1 |
| 2 | 47410321 | A | C | E | D | MSH2 | ENST00000233146 | 0.58006 | 0.03864 | 0.61174 | 0.9646 | 0.98621 | 0.8478233627 | 0.99162 | 0.9852617383 | 0.525636 | 10.19769097 | 0 |
| 2 | 47410321 | A | T | E | D | MSH2 | ENST00000233146 | 0.57406 | 0.03864 | 0.61174 | 0.9646 | 0.98621 | 0.8478233627 | 0.99167 | 0.9852617383 | 0.525632 | 10.19769097 | 0 |
| 2 | 47410322 | T | A | C | S | MSH2 | ENST00000233146 | 0.79469 | 0.03394 | 0.73448 | 0.9323 | 0.98585 | 0.8464009388 | 0.94947 | 0.9961052537 | 0.470366 | 8.596293336 | 0 |
| 2 | 47410322 | T | C | C | R | MSH2 | ENST00000233146 | 0.85485 | 0.02857 | 0.83609 | 0.9335 | 0.98693 | 0.9273721151 | 0.97846 | 0.9990592599 | 0.476697 | 8.922258178 | 1 |
| 2 | 47410322 | T | G | C | G | MSH2 | ENST00000233146 | 0.84096 | 0.03388 | 0.70074 | 0.93305 | 0.98657 | 0.8757960243 | 0.97068 | 0.9981402159 | 0.476376 | 8.922258178 | 0 |
| 2 | 47410323 | G | A | C | Y | MSH2 | ENST00000233146 | 0.85669 | 0.02927 | 0.85382 | 0.88965 | 0.96627 | 0.8876019459 | 0.96154 | 0.9992291927 | 0.479587 | 8.922258178 | 1 |
| 2 | 47410323 | G | C | C | S | MSH2 | ENST00000233146 | 0.84012 | 0.03394 | 0.73448 | 0.91421 | 0.98404 | 0.8464009388 | 0.94738 | 0.9927483257 | 0.473255 | 8.596293336 | 0 |
| 2 | 47410324 | T | G | C | W | MSH2 | ENST00000233146 | 0.75861 | 0.03274 | 0.80355 | 0.9315 | 0.92964 | 0.8899204098 | 0.97117 | 0.9996675253 | 0.424356 | 8.922258178 | 1 |
| 2 | 47410325 | G | A | V | I | MSH2 | ENST00000233146 | 0.31265 | 0.00615 | 0.22526 | 0.47175 | 0.78782 | 0.5685582826 | 0.95498 | 0.204496637 | 0.111912 | 1.48878888 | 0 |
| 2 | 47410325 | G | C | V | L | MSH2 | ENST00000233146 | 0.27875 | 0.0058 | 0.38242 | 0.43239 | 0.7609 | 0.662037056 | 0.96247 | 0.05217646435 | 0.122276 | 0.7984541589 | 0 |
| 2 | 47410325 | G | T | V | F | MSH2 | ENST00000233146 | 0.52626 | 0.01191 | 0.35541 | 0.87355 | 0.90406 | 0.8312327915 | 0.96103 | 0.8922774196 | 0.260955 | 7.709084322 | 0 |
| 2 | 47410326 | T | A | V | D | MSH2 | ENST00000233146 | 0.7031 | 0.01888 | 0.41745 | 0.88988 | 0.96226 | 0.9285526232 | 0.96139 | 0.9885445237 | 0.434462 | 7.709084322 | 1 |
| 2 | 47410326 | T | C | V | A | MSH2 | ENST00000233146 | 0.4988 | 0.00763 | 0.41095 | 0.82091 | 0.93241 | 0.7594233192 | 0.93514 | 0.894195497 | 0.28849 | 7.709084322 | 0 |
| 2 | 47410326 | T | G | V | G | MSH2 | ENST00000233146 | 0.67764 | 0.0178 | 0.27453 | 0.89671 | 0.95608 | 0.9064152171 | 0.92023 | 0.97852844 | 0.381534 | 7.709084322 | 0 |
| 2 | 47410328 | T | A | L | I | MSH2 | ENST00000233146 | 0.31904 | 0.00603 | 0.53261 | 0.46526 | 0.7562 | 0.4178257723 | 0.89317 | 0.1169735715 | 0.0756636 | 3.046537728 | 0 |
| 2 | 47410328 | T | G | L | V | MSH2 | ENST00000233146 | 0.35498 | 0.00648 | 0.44957 | 0.58441 | 0.8367 | 0.5510038079 | 0.97377 | 0.1007079557 | 0.175646 | 3.307795354 | 0 |
| 2 | 47410329 | T | C | L | S | MSH2 | ENST00000233146 | 0.70318 | 0.02237 | 0.54755 | 0.90508 | 0.96189 | 0.8907065568 | 0.95615 | 0.9890198112 | 0.331844 | 5.998121196 | 0 |
| 2 | 47410330 | A | C | L | F | MSH2 | ENST00000233146 | 0.43202 | 0.02206 | 0.51247 | 0.78543 | 0.86352 | 0.6951566005 | 0.98735 | 0.9028235674 | 0.23686 | 4.782265164 | 0 |
| 2 | 47410330 | A | T | L | F | MSH2 | ENST00000233146 | 0.42699 | 0.02206 | 0.51247 | 0.78533 | 0.86352 | 0.6951566005 | 0.98741 | 0.9028235674 | 0.236858 | 4.782265164 | 0 |
| 2 | 47410331 | C | A | P | T | MSH2 | ENST00000233146 | 0.34369 | 0.00777 | 0.48903 | 0.70752 | 0.82103 | 0.6135353625 | 0.90042 | 0.3087636887 | 0.238562 | 4.720613608 | 0 |
| 2 | 47410331 | C | G | P | A | MSH2 | ENST00000233146 | 0.31587 | 0.00619 | 0.44594 | 0.50832 | 0.79395 | 0.4990071635 | 0.83789 | 0.40784958 | 0.181912 | 2.310802009 | 0 |
| 2 | 47410331 | C | T | P | S | MSH2 | ENST00000233146 | 0.48614 | 0.00763 | 0.43907 | 0.69245 | 0.84694 | 0.5900880005 | 0.80664 | 0.7994124293 | 0.211815 | 2.514867912 | 0 |
| 2 | 47410332 | C | A | P | H | MSH2 | ENST00000233146 | 0.41534 | 0.0325 | 0.52435 | 0.81537 | 0.92233 | 0.6524309883 | 0.76457 | 0.9384923577 | 0.264216 | 5.167088615 | 0 |
| 2 | 47410332 | C | G | P | R | MSH2 | ENST00000233146 | 0.43208 | 0.02809 | 0.54957 | 0.77691 | 0.91904 | 0.6974354726 | 0.81115 | 0.9352069497 | 0.302178 | 5.654701507 | 0 |
| 2 | 47410332 | C | T | P | L | MSH2 | ENST00000233146 | 0.43249 | 0.01654 | 0.5602 | 0.74494 | 0.89873 | 0.6812562973 | 0.85343 | 0.9517939091 | 0.25527 | 4.838702225 | 0 |
| 2 | 47410334 | G | A | G | R | MSH2 | ENST00000233146 | 0.27407 | 0.00638 | 0.41639 | 0.62056 | 0.80889 | 0.5321569078 | 0.91879 | 0.2644470334 | 0.130037 | 3.477875318 | 0 |
| 2 | 47410334 | G | C | G | R | MSH2 | ENST00000233146 | 0.27213 | 0.00638 | 0.41639 | 0.648 | 0.8345 | 0.5321569078 | 0.91949 | 0.5635004059 | 0.18796 | 3.477875318 | 0 |
| 2 | 47410335 | G | C | G | A | MSH2 | ENST00000233146 | 0.16312 | 0.00556 | 0.29901 | 0.42302 | 0.76946 | 0.4197702149 | 0.96399 | 0.04346028318 | 0.13105 | 0.7396029196 | 0 |
| 2 | 47410335 | G | T | G | V | MSH2 | ENST00000233146 | 0.21513 | 0.00592 | 0.31398 | 0.72599 | 0.85274 | 0.5099677915 | 0.91448 | 0.2145008855 | 0.176951 | 2.841283623 | 0 |
| 2 | 47410337 | G | A | G | R | MSH2 | ENST00000233146 | 0.85478 | 0.03293 | 0.65899 | 0.84665 | 0.90711 | 0.6745577632 | 0.96483 | 0.9308234081 | 0.470115 | 5.209287378 | 0 |
| 2 | 47410337 | G | C | G | R | MSH2 | ENST00000233146 | 0.85175 | 0.03293 | 0.65899 | 0.84056 | 0.90711 | 0.6745577632 | 0.96481 | 0.95761504 | 0.470115 | 5.209287378 | 0 |
| 2 | 47410338 | G | C | G | A | MSH2 | ENST00000233146 | 0.38765 | 0.03088 | 0.54246 | 0.74552 | 0.89009 | 0.6386506584 | 0.9695 | 0.8789108992 | 0.443424 | 4.154952218 | 0 |
| 2 | 47410338 | G | T | G | V | MSH2 | ENST00000233146 | 0.51949 | 0.0315 | 0.55304 | 0.79883 | 0.94356 | 0.7046834848 | 0.97079 | 0.9841539404 | 0.472923 | 4.648349085 | 0 |
| 2 | 47410340 | G | A | E | K | MSH2 | ENST00000233146 | 0.54521 | 0.00881 | 0.47524 | 0.74767 | 0.80536 | 0.438609472 | 0.98332 | 0.7952474952 | 0.278705 | 5.004349623 | 0 |
| 2 | 47410340 | G | C | E | Q | MSH2 | ENST00000233146 | 0.50049 | 0.01115 | 0.36809 | 0.63201 | 0.8278 | 0.4245582796 | 0.97713 | 0.4388541877 | 0.224154 | 4.040157261 | 0 |
| 2 | 47410341 | A | C | E | A | MSH2 | ENST00000233146 | 0.49679 | 0.01861 | 0.38724 | 0.76556 | 0.79516 | 0.4408524017 | 0.94968 | 0.9512318373 | 0.253364 | 3.953250192 | 0 |
| 2 | 47410341 | A | G | E | G | MSH2 | ENST00000233146 | 0.45033 | 0.00586 | 0.35004 | 0.80047 | 0.85586 | 0.4881775807 | 0.89731 | 0.834415624 | 0.252984 | 3.815884222 | 0 |
| 2 | 47410341 | A | T | E | V | MSH2 | ENST00000233146 | 0.55717 | 0.01058 | 0.36578 | 0.83283 | 0.87 | 0.5173944966 | 0.96098 | 0.9555384517 | 0.302465 | 3.902692645 | 0 |
| 2 | 47410342 | G | C | E | D | MSH2 | ENST00000233146 | 0.29089 | 0.00608 | 0.38087 | 0.54387 | 0.76289 | 0.2705317655 | 0.99082 | 0.3408769667 | 0.171783 | 1.342029197 | 0 |
| 2 | 47410342 | G | T | E | D | MSH2 | ENST00000233146 | 0.28685 | 0.00608 | 0.38087 | 0.56429 | 0.76289 | 0.2705317655 | 0.99063 | 0.4492390752 | 0.171789 | 1.342029197 | 0 |
| 2 | 47410343 | A | C | T | P | MSH2 | ENST00000233146 | 0.45438 | 0.01401 | 0.32791 | 0.79721 | 0.87684 | 0.516551062 | 0.92497 | 0.8693579435 | 0.234924 | 3.530087618 | 0 |
| 2 | 47410343 | A | G | T | A | MSH2 | ENST00000233146 | 0.2448 | 0.0058 | 0.32986 | 0.60002 | 0.76487 | 0.3359967845 | 0.98856 | 0.1023778138 | 0.190414 | 2.137574096 | 0 |
| 2 | 47410343 | A | T | T | S | MSH2 | ENST00000233146 | 0.21056 | 0.00596 | 0.33087 | 0.57658 | 0.71153 | 0.2731060951 | 0.97619 | 0.0952324337 | 0.162025 | 2.030422261 | 0 |
| 2 | 47410344 | C | A | T | N | MSH2 | ENST00000233146 | 0.17703 | 0.00551 | 0.39537 | 0.56663 | 0.67478 | 0.33358454 | 0.94935 | 0.1965308627 | 0.133293 | 2.596680176 | 0 |
| 2 | 47410344 | C | G | T | S | MSH2 | ENST00000233146 | 0.15382 | 0.00596 | 0.33087 | 0.51946 | 0.66258 | 0.2731060951 | 0.96002 | 0.2598313124 | 0.166199 | 2.030422261 | 0 |
| 2 | 47410344 | C | T | T | I | MSH2 | ENST00000233146 | 0.33903 | 0.00816 | 0.45679 | 0.65682 | 0.81298 | 0.380685857 | 0.89421 | 0.7841664553 | 0.231562 | 3.46009877 | 0 |
| 2 | 47410346 | G | A | A | T | MSH2 | ENST00000233146 | 0.21134 | 0.00574 | 0.15456 | 0.47637 | 0.79334 | 0.348639503 | 0.9857 | 0.1348353326 | 0.124599 | 2.562243414 | 0 |
| 2 | 47410346 | G | C | A | P | MSH2 | ENST00000233146 | 0.2562 | 0.00816 | 0.20769 | 0.70316 | 0.84535 | 0.6479792811 | 0.96341 | 0.3030533493 | 0.165833 | 4.115430621 | 0 |
| 2 | 47410346 | G | T | A | S | MSH2 | ENST00000233146 | 0.15267 | 0.00586 | 0.13418 | 0.47236 | 0.75143 | 0.2943611084 | 0.90598 | 0.04227011278 | 0.0385159 | 1.643470025 | 0 |
| 2 | 47410347 | C | A | A | D | MSH2 | ENST00000233146 | 0.15702 | 0.00504 | 0.25419 | 0.52495 | 0.76553 | 0.5487370956 | 0.91063 | 0.6282800436 | 0.152872 | 3.137684447 | 0 |
| 2 | 47410347 | C | G | A | G | MSH2 | ENST00000233146 | 0.16098 | 0.00569 | 0.15092 | 0.44093 | 0.70994 | 0.318766177 | 0.89728 | 0.2281300873 | 0.0642436 | 2.319599806 | 0 |
| 2 | 47410347 | C | T | A | V | MSH2 | ENST00000233146 | 0.16581 | 0.00574 | 0.19158 | 0.44684 | 0.79456 | 0.403583796 | 0.94013 | 0.3313656834 | 0.170407 | 3.334948574 | 0 |
| 2 | 47410349 | G | A | G | R | MSH2 | ENST00000233146 | 0.49673 | 0.00556 | 0.5166 | 0.67989 | 0.77718 | 0.7420429792 | 0.93094 | 0.4193213694 | 0.22411 | 5.105251257 | 0 |
| 2 | 47410349 | G | C | G | R | MSH2 | ENST00000233146 | 0.49408 | 0.00556 | 0.5166 | 0.67926 | 0.77718 | 0.7420429792 | 0.92873 | 0.4193366625 | 0.224125 | 5.105251257 | 0 |
| 2 | 47410350 | G | A | G | E | MSH2 | ENST00000233146 | 0.33828 | 0.01075 | 0.49142 | 0.69764 | 0.80058 | 0.7214786935 | 0.86866 | 0.7126996517 | 0.237178 | 3.63961706 | 0 |
| 2 | 47410350 | G | C | G | A | MSH2 | ENST00000233146 | 0.27493 | 0.00562 | 0.40091 | 0.4422 | 0.70427 | 0.6184403709 | 0.90544 | 0.7422221303 | 0.102391 | 2.462522982 | 0 |
| 2 | 47410350 | G | T | G | V | MSH2 | ENST00000233146 | 0.30198 | 0.0058 | 0.40823 | 0.65663 | 0.76487 | 0.6953424268 | 0.91771 | 0.7737653255 | 0.215749 | 3.836887946 | 0 |
| 2 | 47410352 | G | A | D | N | MSH2 | ENST00000233146 | 0.72869 | 0.00638 | 0.35397 | 0.77669 | 0.87974 | 0.542805374 | 0.89332 | 0.9096022805 | 0.131068 | 4.383045881 | 0 |
| 2 | 47410352 | G | C | D | H | MSH2 | ENST00000233146 | 0.86432 | 0.00599 | 0.38266 | 0.83521 | 0.95243 | 0.6616923946 | 0.92131 | 0.9530545578 | 0.332013 | 4.468083853 | 0 |
| 2 | 47410352 | G | T | D | Y | MSH2 | ENST00000233146 | 0.8765 | 0.02108 | 0.3592 | 0.85009 | 0.97215 | 0.7109900952 | 0.96595 | 0.977622509 | 0.4283 | 4.959369624 | 0 |
| 2 | 47410353 | A | C | D | A | MSH2 | ENST00000233146 | 0.63298 | 0.00644 | 0.30755 | 0.81768 | 0.87877 | 0.6468082612 | 0.89867 | 0.9439392686 | 0.263925 | 5.088704783 | 0 |
| 2 | 47410353 | A | G | D | G | MSH2 | ENST00000233146 | 0.81198 | 0.00556 | 0.31068 | 0.85918 | 0.9506 | 0.6383972163 | 0.9348 | 0.9775031209 | 0.311985 | 5.570016652 | 0 |
| 2 | 47410353 | A | T | D | V | MSH2 | ENST00000233146 | 0.63237 | 0.0134 | 0.29137 | 0.92773 | 0.94839 | 0.693752313 | 0.96887 | 0.9895629287 | 0.407807 | 5.926758352 | 0 |
| 2 | 47410354 | C | A | D | E | MSH2 | ENST00000233146 | 0.22885 | 0.00596 | 0.32675 | 0.50107 | 0.76946 | 0.3848624878 | 0.93557 | 0.2110747695 | 0.106755 | 1.517229445 | 0 |
| 2 | 47410354 | C | G | D | E | MSH2 | ENST00000233146 | 0.2333 | 0.00596 | 0.32675 | 0.46453 | 0.77011 | 0.3848624878 | 0.93665 | 0.1443609446 | 0.10674 | 1.517229445 | 0 |
| 2 | 47410355 | A | C | M | L | MSH2 | ENST00000233146 | 0.30473 | 0.0058 | 0.22262 | 0.45945 | 0.73296 | 0.464562997 | 0.88788 | 0.045639541 | 0.0884195 | 2.902810392 | 0 |
| 2 | 47410355 | A | G | M | V | MSH2 | ENST00000233146 | 0.31364 | 0.00615 | 0.16744 | 0.61237 | 0.80889 | 0.5221603487 | 0.94792 | 0.1487663984 | 0.150758 | 2.616235637 | 0 |
| 2 | 47410355 | A | T | M | L | MSH2 | ENST00000233146 | 0.3004 | 0.0058 | 0.22262 | 0.45945 | 0.73296 | 0.464562997 | 0.88754 | 0.045639541 | 0.0884216 | 2.902810392 | 0 |
| 2 | 47410356 | T | A | M | K | MSH2 | ENST00000233146 | 0.46059 | 0.00586 | 0.3027 | 0.70248 | 0.86753 | 0.7559360251 | 0.91035 | 0.8357647657 | 0.217195 | 3.587607274 | 0 |
| 2 | 47410356 | T | C | M | T | MSH2 | ENST00000233146 | 0.39783 | 0.00547 | 0.33709 | 0.66646 | 0.82273 | 0.587944236 | 0.87893 | 0.5338138938 | 0.184951 | 2.68418795 | 0 |
| 2 | 47410356 | T | G | M | R | MSH2 | ENST00000233146 | 0.46768 | 0.00586 | 0.27852 | 0.69118 | 0.87247 | 0.8006859242 | 0.9181 | 0.8236258626 | 0.218915 | 4.336930173 | 0 |
| 2 | 47410357 | G | A | M | I | MSH2 | ENST00000233146 | 0.4496 | 0.00566 | 0.23046 | 0.53648 | 0.71705 | 0.5092415484 | 0.88981 | 0.2081606714 | -0.00221087 | 3.27105775 | 0 |
| 2 | 47410357 | G | C | M | I | MSH2 | ENST00000233146 | 0.4471 | 0.00566 | 0.23046 | 0.53701 | 0.71705 | 0.5092415484 | 0.89234 | 0.4810345123 | 0.124154 | 3.27105775 | 0 |
| 2 | 47410357 | G | T | M | I | MSH2 | ENST00000233146 | 0.44217 | 0.00566 | 0.23046 | 0.53588 | 0.71705 | 0.5092415484 | 0.8881 | 0.4579983031 | 0.124137 | 3.27105775 | 0 |
| 2 | 47410358 | G | A | G | R | MSH2 | ENST00000233146 | 0.54446 | 0.01433 | 0.51692 | 0.68333 | 0.8367 | 0.6297477259 | 0.90173 | 0.596373634 | 0.1197 | 4.012007818 | 0 |
| 2 | 47410358 | G | C | G | R | MSH2 | ENST00000233146 | 0.54169 | 0.01433 | 0.51692 | 0.6707 | 0.8367 | 0.6297477259 | 0.89711 | 0.7929975112 | 0.242144 | 4.012007818 | 0 |
| 2 | 47410359 | G | A | G | E | MSH2 | ENST00000233146 | 0.30035 | 0.00535 | 0.49434 | 0.70081 | 0.79818 | 0.5937845455 | 0.91342 | 0.4463793933 | 0.122202 | 2.551327904 | 0 |
| 2 | 47410359 | G | C | G | A | MSH2 | ENST00000233146 | 0.2702 | 0.00569 | 0.40725 | 0.49594 | 0.77909 | 0.4859956704 | 0.96732 | 0.1463713497 | 0.150329 | 1.727586532 | 0 |
| 2 | 47410359 | G | T | G | V | MSH2 | ENST00000233146 | 0.33425 | 0.0054 | 0.42207 | 0.74149 | 0.84482 | 0.5801616598 | 0.9449 | 0.5397799015 | 0.185707 | 3.042194175 | 0 |
| 2 | 47410361 | A | C | K | Q | MSH2 | ENST00000233146 | 0.85419 | 0.0278 | 0.45664 | 0.8147 | 0.94356 | 0.6417216542 | 0.93598 | 0.9552939402 | 0.327734 | 5.041397743 | 0 |
| 2 | 47410361 | A | G | K | E | MSH2 | ENST00000233146 | 0.73749 | 0.015 | 0.5022 | 0.78778 | 0.94876 | 0.7202452081 | 0.97087 | 0.9599944353 | 0.429369 | 4.275582247 | 0 |
| 2 | 47410362 | A | C | K | T | MSH2 | ENST00000233146 | 0.473 | 0.00916 | 0.51687 | 0.80449 | 0.94655 | 0.6818463586 | 0.93457 | 0.9403275847 | 0.384677 | 4.955311193 | 0 |
| 2 | 47410362 | A | G | K | R | MSH2 | ENST00000233146 | 0.47734 | 0.00741 | 0.42229 | 0.67052 | 0.8356 | 0.571915165 | 0.96894 | 0.7718638778 | 0.276938 | 3.563631635 | 0 |

| | | | | | | | | | | | | | | | | | | |
|---|---|---|---|---|---|---|---|---|---|---|---|---|---|---|---|---|---|---|
| 2 | 47410362 | A | T | K | I | MSH2 | ENST00000233146 | 0.67748 | 0.00788 | 0.46753 | 0.84396 | 0.94469 | 0.7379506547 | 0.93385 | 0.9777270555 | 0.357055 | 4.693856352 | 0 |
| 2 | 47410364 | C | A | L | M | MSH2 | ENST00000233146 | 0.59385 | 0.03409 | 0.54445 | 0.74376 | 0.89558 | 0.5164472284 | 0.81552 | 0.8922655582 | 0.205718 | 4.255533147 | 0 |
| 2 | 47410364 | C | G | L | V | MSH2 | ENST00000233146 | 0.32838 | 0.02255 | 0.51198 | 0.74731 | 0.88212 | 0.5615496523 | 0.92299 | 0.768255353 | 0.231066 | 3.339687137 | 0 |
| 2 | 47410365 | T | A | L | Q | MSH2 | ENST00000233146 | 0.78273 | 0.03633 | 0.57752 | 0.88792 | 0.98869 | 0.7919732386 | 0.9744 | 0.9967093694 | 0.459866 | 4.789567595 | 0 |
| 2 | 47410365 | T | C | L | P | MSH2 | ENST00000233146 | 0.73525 | 0.01705 | 0.69122 | 0.8961 | 0.98034 | 0.9177579638 | 0.96702 | 0.9919067591 | 0.4336 | 6.436855582 | 0 |
| 2 | 47410365 | T | G | L | R | MSH2 | ENST00000233146 | 0.79334 | 0.03124 | 0.65591 | 0.88408 | 0.98904 | 0.9056562266 | 0.9699 | 0.9967448681 | 0.459607 | 5.240811519 | 1 |
| 2 | 47410367 | A | G | R | G | MSH2 | ENST00000233146 | 0.54793 | 0.01258 | 0.38008 | 0.88026 | 0.94129 | 0.6508409571 | 0.97749 | 0.9750576019 | 0.382262 | 4.874411788 | 0 |
| 2 | 47410368 | G | A | R | K | MSH2 | ENST00000233146 | 0.32637 | 0.00586 | 0.45798 | 0.63509 | 0.71548 | 0.5711620685 | 0.90126 | 0.5909247994 | 0.115614 | 2.094794092 | 0 |
| 2 | 47410368 | G | C | R | T | MSH2 | ENST00000233146 | 0.40728 | 0.00638 | 0.44364 | 0.78587 | 0.89648 | 0.6447300055 | 0.96065 | 0.8964681029 | 0.297509 | 4.540274595 | 0 |
| 2 | 47410368 | G | T | R | I | MSH2 | ENST00000233146 | 0.56186 | 0.01385 | 0.41671 | 0.87072 | 0.93515 | 0.5734858066 | 0.93859 | 0.9886718988 | 0.292218 | 5.081547732 | 0 |
| 2 | 47410369 | A | C | R | S | MSH2 | ENST00000233146 | 0.3034 | 0.00608 | 0.39739 | 0.76182 | 0.82387 | 0.6651367644 | 0.95593 | 0.44565624 | 0.242245 | 4.655486602 | 0 |
| 2 | 47410369 | A | T | R | S | MSH2 | ENST00000233146 | 0.29909 | 0.00608 | 0.39739 | 0.76182 | 0.82387 | 0.6651367644 | 0.95667 | 0.44565624 | 0.242225 | 4.655486602 | 0 |
| 2 | 47410370 | C | A | Q | K | MSH2 | ENST00000233146 | 0.38106 | 0.00615 | 0.36047 | 0.80351 | 0.79577 | 0.5572644507 | 0.93076 | 0.53829593 | 0.211401 | 4.428030307 | 0 |
| 2 | 47410370 | C | G | Q | E | MSH2 | ENST00000233146 | 0.32955 | 0.00608 | 0.3021 | 0.70089 | 0.75005 | 0.4360114562 | 0.89285 | 0.534011662 | 0.132743 | 2.678345915 | 0 |
| 2 | 47410371 | A | C | Q | P | MSH2 | ENST00000233146 | 0.79073 | 0.02533 | 0.31292 | 0.94287 | 0.95462 | 0.775778626 | 0.93578 | 0.9814173579 | 0.396067 | 5.477829501 | 0 |
| 2 | 47410371 | A | G | Q | R | MSH2 | ENST00000233146 | 0.61566 | 0.0158 | 0.34916 | 0.9364 | 0.87247 | 0.5472762092 | 0.9305 | 0.9455161691 | 0.252455 | 4.469445578 | 0 |
| 2 | 47410371 | A | T | Q | L | MSH2 | ENST00000233146 | 0.77227 | 0.01121 | 0.28869 | 0.94052 | 0.90007 | 0.5602895889 | 0.91326 | 0.9813662767 | 0.275276 | 5.117738564 | 0 |
| 2 | 47410372 | G | C | Q | H | MSH2 | ENST00000233146 | 0.6918 | 0.00696 | 0.3392 | 0.84811 | 0.87099 | 0.5071572473 | 0.90027 | 0.9422810674 | 0.225049 | 5.386162398 | 0 |
| 2 | 47410372 | G | T | Q | H | MSH2 | ENST00000233146 | 0.68523 | 0.00696 | 0.3392 | 0.84988 | 0.87099 | 0.5071572473 | 0.90008 | 0.9422810674 | 0.22505 | 5.386162398 | 0 |
| 2 | 47412414 | A | C | I | L | MSH2 | ENST00000233146 | 0.28808 | 0.00574 | 0.0901 | 0.74745 | 0.71938 | 0.5008554842 | 0.83864 | 0.03457491375 | 0.0742218 | 1.799921339 | 0 |
| 2 | 47412414 | A | G | I | V | MSH2 | ENST00000233146 | 0.18137 | 0.00599 | 0.05244 | 0.98027 | 0.66733 | 0.2212377692 | 0.62219 | 0.008513586337 | -0.0928777 | -1.615799275 | 0 |
| 2 | 47412414 | A | T | I | L | MSH2 | ENST00000233146 | 0.28393 | 0.00574 | 0.0901 | 0.74759 | 0.71391 | 0.5008554842 | 0.84591 | 0.1136730984 | 0.0741835 | 1.799921339 | 0 |
| 2 | 47412415 | T | A | I | K | MSH2 | ENST00000233146 | 0.50231 | 0.00681 | 0.12253 | 0.9087 | 0.88543 | 0.7870491849 | 0.92042 | 0.980245173 | 0.23356 | 3.275182401 | 0 |
| 2 | 47412415 | T | C | I | T | MSH2 | ENST00000233146 | 0.39093 | 0.00543 | 0.15711 | 0.79563 | 0.76685 | 0.5550877596 | 0.90488 | 0.7889742851 | 0.155752 | 3.486362453 | 0 |
| 2 | 47412415 | T | G | I | R | MSH2 | ENST00000233146 | 0.52852 | 0.01082 | 0.1139 | 0.91038 | 0.89376 | 0.7893421998 | 0.90045 | 0.9899401069 | 0.224065 | 3.188440585 | 0 |
| 2 | 47412416 | A | G | I | M | MSH2 | ENST00000233146 | 0.29793 | 0.00696 | 0.08743 | 0.66978 | 0.82613 | 0.5146546667 | 0.88459 | 0.4261083603 | 0.136202 | 3.187348984 | 0 |
| 2 | 47412417 | A | C | I | L | MSH2 | ENST00000233146 | 0.28519 | 0.0058 | 0.50033 | 0.62559 | 0.72093 | 0.4187545598 | 0.85857 | 0.405572623 | 0.0656025 | 0.7929949182 | 0 |
| 2 | 47412417 | A | G | I | V | MSH2 | ENST00000233146 | 0.24068 | 0.00615 | 0.39638 | 0.46775 | 0.70345 | 0.2014716526 | 0.86919 | 0.08008762047 | -0.00421082 | -0.06626783374 | 0 |
| 2 | 47412417 | A | T | I | F | MSH2 | ENST00000233146 | 0.61037 | 0.01871 | 0.50834 | 0.85576 | 0.89648 | 0.6528115371 | 0.93345 | 0.9624398947 | 0.224803 | 3.455898391 | 0 |
| 2 | 47412418 | T | A | I | N | MSH2 | ENST00000233146 | 0.7855 | 0.02186 | 0.61084 | 0.93407 | 0.94318 | 0.7534352465 | 0.83302 | 0.9930011034 | 0.249145 | 3.988143948 | 0 |
| 2 | 47412418 | T | C | I | T | MSH2 | ENST00000233146 | 0.56228 | 0.00669 | 0.64348 | 0.8354 | 0.90929 | 0.5439965272 | 0.88159 | 0.9330880642 | 0.269715 | 3.35566543 | 0 |
| 2 | 47412418 | T | G | I | S | MSH2 | ENST00000233146 | 0.4887 | 0.00696 | 0.52572 | 0.78827 | 0.88448 | 0.7400566177 | 0.9115 | 0.9441843629 | 0.254489 | 3.720107428 | 0 |
| 2 | 47412419 | T | G | I | M | MSH2 | ENST00000233146 | 0.29753 | 0.02561 | 0.45861 | 0.81682 | 0.80358 | 0.4602326677 | 0.82948 | 0.6650986075 | 0.0934451 | 4.428503452 | 0 |
| 2 | 47412420 | C | A | Q | K | MSH2 | ENST00000233146 | 0.3371 | 0.00532 | 0.39685 | 0.67017 | 0.74797 | 0.499726163 | 0.98033 | 0.7582424283 | 0.174389 | 3.007864237 | 0 |
| 2 | 47412420 | C | G | Q | E | MSH2 | ENST00000233146 | 0.24362 | 0.00556 | 0.32738 | 0.76314 | 0.74446 | 0.3863775082 | 0.94328 | 0.4312594235 | 0.124045 | 2.142073049 | 0 |
| 2 | 47412421 | A | C | Q | P | MSH2 | ENST00000233146 | 0.59769 | 0.01312 | 0.33609 | 0.86436 | 0.84159 | 0.7506148147 | 0.97703 | 0.9542120695 | 0.250985 | 5.616430355 | 0 |
| 2 | 47412421 | A | G | Q | R | MSH2 | ENST00000233146 | 0.54236 | 0.00596 | 0.3903 | 0.72186 | 0.78223 | 0.4866221837 | 0.9818 | 0.7964445353 | 0.166923 | 3.449080517 | 0 |
| 2 | 47412421 | A | T | Q | L | MSH2 | ENST00000233146 | 0.58211 | 0.0058 | 0.31325 | 0.77317 | 0.76421 | 0.5132333169 | 0.97402 | 0.8208068013 | 0.147893 | 4.59901949 | 0 |
| 2 | 47412422 | A | C | Q | H | MSH2 | ENST00000233146 | 0.5629 | 0.01682 | 0.37746 | 0.80234 | 0.86753 | 0.4631934126 | 0.9489 | 0.8787872791 | 0.205494 | 5.006296363 | 0 |
| 2 | 47412422 | A | T | Q | H | MSH2 | ENST00000233146 | 0.5571 | 0.01682 | 0.37746 | 0.80225 | 0.86753 | 0.4631934126 | 0.94864 | 0.8787872791 | 0.20549 | 5.006296363 | 0 |
| 2 | 47412423 | A | G | R | G | MSH2 | ENST00000233146 | 0.87722 | 0.03322 | 0.46705 | 0.90085 | 0.94839 | 0.6939724981 | 0.92939 | 0.9916046858 | 0.35932 | 6.331258898 | 0 |
| 2 | 47412424 | G | A | R | K | MSH2 | ENST00000233146 | 0.72716 | 0.02741 | 0.55119 | 0.77003 | 0.85945 | 0.6160872453 | 0.93528 | 0.9268354774 | 0.222554 | 4.518948891 | 0 |
| 2 | 47412424 | G | C | R | T | MSH2 | ENST00000233146 | 0.88782 | 0.03129 | 0.53413 | 0.9229 | 0.93083 | 0.7238747846 | 0.94319 | 0.9941382408 | 0.388838 | 5.092503114 | 0 |
| 2 | 47412424 | G | T | R | I | MSH2 | ENST00000233146 | 0.9186 | 0.02857 | 0.52445 | 0.94005 | 0.96334 | 0.6639704404 | 0.93558 | 0.9974164963 | 0.397499 | 5.741245495 | 0 |
| 2 | 47412425 | A | C | R | S | MSH2 | ENST00000233146 | 0.71979 | 0.03518 | 0.47798 | 0.88178 | 0.90406 | 0.7166785791 | 0.97527 | 0.9902963042 | 0.327819 | 5.252113536 | 0 |
| 2 | 47412425 | A | T | R | S | MSH2 | ENST00000233146 | 0.71297 | 0.03518 | 0.47798 | 0.88178 | 0.90406 | 0.7166785791 | 0.97551 | 0.9902963042 | 0.327811 | 5.252113536 | 0 |
| 2 | 47412426 | G | A | G | R | MSH2 | ENST00000233146 | 0.72228 | 0.01505 | 0.55689 | 0.78783 | 0.91526 | 0.7761071891 | 0.89245 | 0.9805712104 | 0.251247 | 4.732518396 | 0 |
| 2 | 47412426 | G | C | G | R | MSH2 | ENST00000233146 | 0.71906 | 0.01505 | 0.55689 | 0.78779 | 0.91526 | 0.7761071891 | 0.88699 | 0.9727396369 | 0.251258 | 4.732518396 | 0 |
| 2 | 47412427 | G | A | G | E | MSH2 | ENST00000233146 | 0.47867 | 0.01009 | 0.53469 | 0.76356 | 0.87296 | 0.7557707451 | 0.86585 | 0.9684038758 | 0.26438 | 5.001110811 | 0 |
| 2 | 47412427 | G | C | G | A | MSH2 | ENST00000233146 | 0.29407 | 0.00566 | 0.41834 | 0.55164 | 0.78223 | 0.5565214069 | 0.90797 | 0.7456593513 | 0.154757 | 4.933682671 | 0 |
| 2 | 47412427 | G | T | G | V | MSH2 | ENST00000233146 | 0.45696 | 0.01019 | 0.43331 | 0.76859 | 0.88354 | 0.664486689 | 0.90798 | 0.9567227364 | 0.26374 | 4.576375972 | 1 |
| 2 | 47412429 | G | A | G | R | MSH2 | ENST00000233146 | 0.79618 | 0.02738 | 0.66676 | 0.87449 | 0.97327 | 0.7453123605 | 0.96387 | 0.9938608408 | 0.457769 | 5.415892761 | 0 |
| 2 | 47412429 | G | C | G | R | MSH2 | ENST00000233146 | 0.79294 | 0.02738 | 0.66676 | 0.87327 | 0.97327 | 0.7453123605 | 0.96361 | 0.9935590625 | 0.457769 | 5.415892761 | 0 |
| 2 | 47412430 | G | A | G | E | MSH2 | ENST00000233146 | 0.76323 | 0.03287 | 0.653 | 0.87647 | 0.95899 | 0.7027278601 | 0.95916 | 0.9968994856 | 0.469992 | 5.224257483 | 0 |
| 2 | 47412430 | G | C | G | A | MSH2 | ENST00000233146 | 0.73981 | 0.03542 | 0.54893 | 0.85619 | 0.94655 | 0.6341872458 | 0.95873 | 0.9942300597 | 0.450988 | 5.765809218 | 0 |
| 2 | 47412430 | G | T | G | V | MSH2 | ENST00000233146 | 0.77734 | 0.03353 | 0.56799 | 0.89407 | 0.97661 | 0.7577367944 | 0.96227 | 0.9986999035 | 0.47601 | 6.667578128 | 0 |
| 2 | 47412432 | A | C | I | L | MSH2 | ENST00000233146 | 0.52619 | 0.00615 | 0.61044 | 0.76754 | 0.74376 | 0.40312153 | 0.91232 | 0.4361606656 | 0.105285 | 2.166360103 | 0 |
| 2 | 47412432 | A | G | I | V | MSH2 | ENST00000233146 | 0.36918 | 0.00638 | 0.51306 | 0.62906 | 0.76356 | 0.2597062864 | 0.91805 | 0.2355584353 | 0.109833 | 2.147425232 | 0 |
| 2 | 47412432 | A | T | I | F | MSH2 | ENST00000233146 | 0.87892 | 0.02133 | 0.62695 | 0.90257 | 0.92438 | 0.6360361862 | 0.93811 | 0.929031074 | 0.265963 | 4.401563832 | 0 |
| 2 | 47412433 | T | A | I | N | MSH2 | ENST00000233146 | 0.82879 | 0.02494 | 0.71462 | 0.89738 | 0.94167 | 0.7082949799 | 0.86768 | 0.9975833893 | 0.277661 | 6.131455139 | 0 |
| 2 | 47412433 | T | C | I | T | MSH2 | ENST00000233146 | 0.74882 | 0.0199 | 0.73624 | 0.81561 | 0.93939 | 0.5942265957 | 0.90451 | 0.9766731858 | 0.297468 | 4.191768589 | 0 |
| 2 | 47412433 | T | G | I | S | MSH2 | ENST00000233146 | 0.813 | 0.01977 | 0.63653 | 0.85616 | 0.93593 | 0.7778685457 | 0.92639 | 0.9770520926 | 0.294717 | 4.543472605 | 0 |
| 2 | 47412434 | T | G | I | M | MSH2 | ENST00000233146 | 0.3074 | 0.01108 | 0.56683 | 0.84236 | 0.85945 | 0.4702789127 | 0.91992 | 0.465857178 | 0.111997 | 4.090034413 | 0 |
| 2 | 47412435 | C | G | L | V | MSH2 | ENST00000233146 | 0.44582 | 0.02172 | 0.38106 | 0.76985 | 0.88212 | 0.4429157433 | 0.91059 | 0.7622386217 | 0.142691 | 4.284976469 | 0 |
| 2 | 47412436 | T | C | L | P | MSH2 | ENST00000233146 | 0.76315 | 0.02844 | 0.61539 | 0.89226 | 0.95899 | 0.8550895664 | 0.97999 | 0.9942125678 | 0.425703 | 6.016313112 | 0 |
| 2 | 47412436 | T | G | L | R | MSH2 | ENST00000233146 | 0.79942 | 0.02828 | 0.56504 | 0.91268 | 0.95935 | 0.7954579851 | 0.96707 | 0.9942295551 | 0.414489 | 6.002378439 | 0 |
| 2 | 47412438 | A | C | I | L | MSH2 | ENST00000233146 | 0.5055 | 0.00669 | 0.57758 | 0.65701 | 0.75822 | 0.4213689631 | 0.89913 | 0.559971869 | 0.104468 | 3.000581802 | 0 |
| 2 | 47412438 | A | G | I | V | MSH2 | ENST00000233146 | 0.36856 | 0.0063 | 0.46244 | 0.51165 | 0.7285 | 0.2415012681 | 0.90189 | 0.4214187264 | 0.099959 | 2.246035197 | 0 |
| 2 | 47412438 | A | T | I | F | MSH2 | ENST00000233146 | 0.65117 | 0.02356 | 0.58778 | 0.79291 | 0.92275 | 0.5773838081 | 0.93385 | 0.9745838642 | 0.289741 | 4.787454396 | 0 |
| 2 | 47412439 | T | A | I | N | MSH2 | ENST00000233146 | 0.75527 | 0.02673 | 0.67944 | 0.92323 | 0.94318 | 0.7041785285 | 0.87819 | 0.9975110292 | 0.278243 | 8.597254654 | 0 |
| 2 | 47412439 | T | C | I | T | MSH2 | ENST00000233146 | 0.42168 | 0.01492 | 0.7084 | 0.83813 | 0.91357 | 0.5728582468 | 0.94287 | 0.9906944036 | 0.261954 | 5.016684064 | 0 |
| 2 | 47412439 | T | G | I | S | MSH2 | ENST00000233146 | 0.75472 | 0.02561 | 0.60646 | 0.90677 | 0.95681 | 0.7684581506 | 0.93775 | 0.9975334406 | 0.346905 | 5.389576646 | 0 |
| 2 | 47412440 | C | G | I | M | MSH2 | ENST00000233146 | 0.35212 | 0.03189 | 0.52918 | 0.73923 | 0.88117 | 0.4663803329 | 0.85673 | 0.8626202941 | 0.0927023 | 5.725856814 | 0 |
| 2 | 47412441 | A | C | T | P | MSH2 | ENST00000233146 | 0.80282 | 0.03293 | 0.44734 | 0.9189 | 0.96008 | 0.736775049 | 0.93136 | 0.9866793752 | 0.396629 | 8.875860209 | 1 |
| 2 | 47412441 | A | G | T | A | MSH2 | ENST00000233146 | 0.75041 | 0.03388 | 0.45104 | 0.77086 | 0.91904 | 0.4629929201 | 0.93644 | 0.9729135036 | 0.236585 | 6.204452654 | 0 |
| 2 | 47412441 | A | T | T | S | MSH2 | ENST00000233146 | 0.56623 | 0.02809 | 0.46074 | 0.67021 | 0.87198 | 0.3353443129 | 0.9103 | 0.6899240017 | 0.158754 | 2.430502442 | 0 |
| 2 | 47412442 | C | G | T | R | MSH2 | ENST00000233146 | 0.60134 | 0.03465 | 0.55413 | 0.87263 | 0.94015 | 0.6876847952 | 0.89693 | 0.9967860225 | 0.381413 | 7.01612803 | 0 |
| 2 | 47412442 | C | T | T | I | MSH2 | ENST00000233146 | 0.5932 | 0.03552 | 0.60212 | 0.89927 | 0.94506 | 0.5717481362 | 0.88255 | 0.9951015115 | 0.3455 | 6.460127777 | 0 |
| 2 | 47412444 | G | A | E | K | MSH2 | ENST00000233146 | 0.63827 | 0.01855 | 0.45233 | 0.77707 | 0.86352 | 0.4479108693 | 0.93564 | 0.9635095596 | 0.246138 | 3.62727061 | 0 |
| 2 | 47412444 | G | C | E | Q | MSH2 | ENST00000233146 | 0.57554 | 0.00935 | 0.33083 | 0.68258 | 0.84106 | 0.3854155463 | 0.95502 | 0.7521740198 | 0.219781 | 4.718798642 | 0 |
| 2 | 47412445 | A | C | E | A | MSH2 | ENST00000233146 | 0.4764 | 0.02172 | 0.34888 | 0.84637 | 0.84641 | 0.4331398374 | 0.84959 | 0.9830073118 | 0.24305 | 4.726835716 | 0 |
| 2 | 47412445 | A | G | E | G | MSH2 | ENST00000233146 | 0.63873 | 0.01827 | 0.31263 | 0.82704 | 0.86653 | 0.4978801007 | 0.8329 | 0.9870826006 | 0.242107 | 5.717037801 | 0 |
| 2 | 47412445 | A | T | E | V | MSH2 | ENST00000233146 | 0.60551 | 0.02809 | 0.32833 | 0.86732 | 0.86553 | 0.4968515008 | 0.91741 | 0.9898045063 | 0.273081 | 4.013587982 | 0 |
| 2 | 47412446 | A | C | E | D | MSH2 | ENST00000233146 | 0.20215 | 0.00615 | 0.3505 | 0.65221 | 0.80058 | 0.3028785345 | 0.96733 | 0.1936282963 | 0.134023 | 0.0808454145 | 0 |
| 2 | 47412446 | A | T | E | D | MSH2 | ENST00000233146 | 0.19896 | 0.00615 | 0.3505 | 0.63129 | 0.79998 | 0.3028785345 | 0.9673 | 0.1936282963 | 0.134003 | 0.0808454145 | 0 |
| 2 | 47412447 | A | G | R | G | MSH2 | ENST00000233146 | 0.84801 | 0.03006 | 0.36117 | 0.92952 | 0.92682 | 0.8296761413 | 0.94436 | 0.9891350269 | 0.222702 | 6.196748131 | 0 |
| 2 | 47412448 | G | A | R | K | MSH2 | ENST00000233146 | 0.40887 | 0.0099 | 0.42217 | 0.68326 | 0.84106 | 0.7304069096 | 0.91638 | 0.9281942248 | 0.172222 | 4.239105335 | 0 |
| 2 | 47412448 | G | C | R | T | MSH2 | ENST00000233146 | 0.71121 | 0.0292 | 0.41496 | 0.8906 | 0.96627 | 0.7866221864 | 0.9565 | 0.9971030354 | 0.401252 | 5.682146794 | 0 |
| 2 | 47412448 | G | T | R | I | MSH2 | ENST00000233146 | 0.74802 | 0.02785 | 0.39454 | 0.92794 | 0.92804 | 0.7293128009 | 0.92243 | 0.9969884753 | 0.291065 | 4.651524475 | 0 |
| 2 | 47412449 | A | C | R | S | MSH2 | ENST00000233146 | 0.7536 | 0.03053 | 0.37013 | 0.8113 | 0.90052 | 0.8058725227 | 0.96706 | 0.9972639084 | 0.357683 | 6.010191598 | 0 |
| 2 | 47412449 | A | T | R | S | MSH2 | ENST00000233146 | 0.74682 | 0.03053 | 0.37013 | 0.81048 | 0.90052 | 0.8058725227 | 0.96733 | 0.9972639084 | 0.357676 | 6.010191598 | 0 |
| 2 | 47412450 | A | C | K | Q | MSH2 | ENST00000233146 | 0.81881 | 0.0357 | 0.42878 | 0.8019 | 0.93939 | 0.6719630662 | 0.91856 | 0.985389173 | 0.301385 | 5.84106487 | 0 |
| 2 | 47412450 | A | G | K | E | MSH2 | ENST00000233146 | 0.84325 | 0.03144 | 0.47058 | 0.83492 | 0.94949 | 0.7312508536 | 0.93704 | 0.9089887142 | 0.2713 | 4.274253044 | 0 |
| 2 | 47412451 | A | C | K | T | MSH2 | ENST00000233146 | 0.82887 | 0.03426 | 0.48008 | 0.86097 | 0.95899 | 0.6998218827 | 0.90638 | 0.9928758144 | 0.416863 | 5.349139759 | 0 |
| 2 | 47412451 | A | G | K | R | MSH2 | ENST00000233146 | 0.66462 | 0.02391 | 0.39512 | 0.71561 | 0.8543 | 0.5587936827 | 0.9484 | 0.946863234 | 0.272778 | 4.870827956 | 0 |
| 2 | 47412451 | A | T | K | I | MSH2 | ENST00000233146 | 0.87201 | 0.02851 | 0.44676 | 0.87801 | 0.96407 | 0.7313202734 | 0.90374 | 0.9962943196 | 0.399791 | 6.674741906 | 0 |
| 2 | 47412452 | A | C | K | N | MSH2 | ENST00000233146 | 0.58041 | 0.03436 | 0.4988 | 0.87094 | 0.89239 | 0.7024759328 | 0.91981 | 0.9931010008 | 0.127922 | 5.217454105 | 0 |
| 2 | 47412452 | A | T | K | N | MSH2 | ENST00000233146 | 0.57449 | 0.03436 | 0.4988 | 0.87094 | 0.89239 | 0.7024759328 | 0.91939 | 0.9931010008 | 0.127918 | 5.217454105 | 0 |
| 2 | 47412453 | A | C | K | Q | MSH2 | ENST00000233146 | 0.77314 | 0.0292 | 0.18025 | 0.77686 | 0.84428 | 0.4895215923 | 0.90519 | 0.9744054675 | 0.222273 | 4.112392408 | 0 |
| 2 | 47412454 | A | C | K | T | MSH2 | ENST00000233146 | 0.60823 | 0.02955 | 0.25508 | 0.77236 | 0.83172 | 0.5697304455 | 0.79547 | 0.9826620817 | 0.131019 | 4.440345974 | 0 |
| 2 | 47412455 | A | C | K | N | MSH2 | ENST00000233146 | 0.67239 | 0.02979 | 0.25171 | 0.74358 | 0.78906 | 0.5296055345 | 0.82586 | 0.9656381607 | 0.103613 | 3.565193427 | 0 |

| | | | | | | | | | | | | | | | | | |
|---|---|---|---|---|---|---|---|---|---|---|---|---|---|---|---|---|---|
| 2 | 47412455 | A | T | K | N | MSH2 | ENST00000233146 | 0.66573 | 0.02979 | 0.25171 | 0.74807 | 0.78906 | 0.5296055345 | 0.82486 | 0.9656381607 | 0.103608 | 3.565193427 | 0 |
| 2 | 47412456 | G | A | A | T | MSH2 | ENST00000233146 | 0.16249 | 0.0058 | 0.08258 | 0.79092 | 0.86047 | 0.2861677974 | 0.96368 | 0.1493084079 | 0.11497 | 1.988898803 | 0 |
| 2 | 47412456 | G | C | A | P | MSH2 | ENST00000233146 | 0.37002 | 0.0134 | 0.11989 | 0.81648 | 0.87442 | 0.6525349734 | 0.9715 | 0.9658949375 | 0.220172 | 4.567957477 | 0 |
| 2 | 47412456 | G | T | A | S | MSH2 | ENST00000233146 | 0.10565 | 0.00592 | 0.06752 | 0.6011 | 0.79456 | 0.24483003 | 0.94691 | 0.03623728082 | 0.039359 | 0.7621639985 | 0 |
| 2 | 47412457 | C | A | A | D | MSH2 | ENST00000233146 | 0.15365 | 0.00889 | 0.15534 | 0.77548 | 0.88963 | 0.5392108999 | 0.95122 | 0.9411801696 | 0.227073 | 3.141739232 | 0 |
| 2 | 47412457 | C | G | A | G | MSH2 | ENST00000233146 | 0.13486 | 0.00569 | 0.08414 | 0.74387 | 0.81818 | 0.2638130843 | 0.93885 | 0.363080889 | 0.0993562 | 1.95780806 | 0 |
| 2 | 47412457 | C | T | A | V | MSH2 | ENST00000233146 | 0.13075 | 0.00574 | 0.10403 | 0.68049 | 0.80238 | 0.3730510991 | 0.9881 | 0.3068090677 | 0.148123 | 1.440228475 | 0 |
| 2 | 47412459 | G | A | D | N | MSH2 | ENST00000233146 | 0.69724 | 0.03106 | 0.24769 | 0.84827 | 0.85064 | 0.4814911651 | 0.88031 | 0.9909079075 | 0.152687 | 4.104391552 | 0 |
| 2 | 47412459 | G | C | D | H | MSH2 | ENST00000233146 | 0.88467 | 0.03118 | 0.27119 | 0.88896 | 0.92316 | 0.5942391824 | 0.85976 | 0.9971123934 | 0.307917 | 4.088428408 | 0 |
| 2 | 47412459 | G | T | D | Y | MSH2 | ENST00000233146 | 0.8926 | 0.03231 | 0.25528 | 0.89897 | 0.9367 | 0.6654874989 | 0.90569 | 0.9960148335 | 0.345266 | 3.538439007 | 0 |
| 2 | 47412460 | A | C | D | A | MSH2 | ENST00000233146 | 0.66929 | 0.02366 | 0.20933 | 0.87936 | 0.9252 | 0.5760646118 | 0.81139 | 0.9912802577 | 0.270249 | 3.520794659 | 0 |
| 2 | 47412460 | A | G | D | G | MSH2 | ENST00000233146 | 0.52307 | 0.02533 | 0.21728 | 0.90123 | 0.92356 | 0.609035195 | 0.90004 | 0.9933633804 | 0.318657 | 4.917567777 | 0 |
| 2 | 47412460 | A | T | D | V | MSH2 | ENST00000233146 | 0.69581 | 0.02765 | 0.19526 | 0.94158 | 0.96226 | 0.6327984507 | 0.93586 | 0.9972863197 | 0.386731 | 3.871313648 | 0 |
| 2 | 47412461 | C | A | D | E | MSH2 | ENST00000233146 | 0.24871 | 0.00644 | 0.22167 | 0.60982 | 0.80118 | 0.3060453216 | 0.92004 | 0.2194713354 | 0.0647504 | -0.07135996367 | 0 |
| 2 | 47412461 | C | G | D | E | MSH2 | ENST00000233146 | 0.25345 | 0.00644 | 0.22167 | 0.59144 | 0.80118 | 0.3060453216 | 0.9202 | 0.2194713354 | 0.0647664 | -0.07135996367 | 0 |
| 2 | 47412462 | T | A | F | I | MSH2 | ENST00000233146 | 0.8329 | 0.0318 | 0.56615 | 0.90153 | 0.9796 | 0.8003078267 | 0.91097 | 0.9991374016 | 0.352474 | 5.354886695 | 0 |
| 2 | 47412462 | T | C | F | L | MSH2 | ENST00000233146 | 0.85018 | 0.03362 | 0.60214 | 0.89159 | 0.98728 | 0.7540722119 | 0.95448 | 0.9969242215 | 0.467239 | 5.655341806 | 0 |
| 2 | 47412462 | T | G | F | V | MSH2 | ENST00000233146 | 0.83859 | 0.03274 | 0.53797 | 0.9256 | 0.99138 | 0.7855905156 | 0.96839 | 0.9986650944 | 0.482425 | 5.66506105 | 0 |
| 2 | 47412463 | T | A | F | Y | MSH2 | ENST00000233146 | 0.6405 | 0.0175 | 0.58564 | 0.8394 | 0.9248 | 0.6780616514 | 0.94178 | 0.9672529697 | 0.302082 | 3.715046622 | 0 |
| 2 | 47412463 | T | C | F | S | MSH2 | ENST00000233146 | 0.89446 | 0.02844 | 0.57281 | 0.9297 | 0.98728 | 0.8256442303 | 0.96171 | 0.9987400174 | 0.479062 | 6.505068636 | 0 |
| 2 | 47412464 | T | A | F | L | MSH2 | ENST00000233146 | 0.54704 | 0.03362 | 0.60214 | 0.91088 | 0.95133 | 0.7540722119 | 0.97668 | 0.9915748239 | 0.424243 | 5.655341806 | 0 |
| 2 | 47412464 | T | G | F | L | MSH2 | ENST00000233146 | 0.55408 | 0.03362 | 0.60214 | 0.9089 | 0.95133 | 0.7540722119 | 0.97667 | 0.9915748239 | 0.424247 | 5.655341806 | 0 |
| 2 | 47412465 | T | A | S | T | MSH2 | ENST00000233146 | 0.3192 | 0.00608 | 0.20242 | 0.49208 | 0.69509 | 0.3300351384 | 0.90953 | 0.09545303881 | 0.0534731 | 0.9533560472 | 0 |
| 2 | 47412465 | T | C | S | P | MSH2 | ENST00000233146 | 0.66193 | 0.01662 | 0.20588 | 0.76719 | 0.78535 | 0.6049145847 | 0.91583 | 0.8337566853 | 0.113848 | 4.460319681 | 0 |
| 2 | 47412465 | T | G | S | A | MSH2 | ENST00000233146 | 0.35697 | 0.00622 | 0.19092 | 0.47307 | 0.63947 | 0.2615679237 | 0.88874 | 0.1207873225 | 0.0391606 | 3.104286126 | 0 |
| 2 | 47412466 | C | A | S | Y | MSH2 | ENST00000233146 | 0.32415 | 0.01505 | 0.23968 | 0.85764 | 0.7727 | 0.4648483466 | 0.92433 | 0.9491206408 | 0.135161 | 4.775872413 | 0 |
| 2 | 47412466 | C | G | S | C | MSH2 | ENST00000233146 | 0.45967 | 0.0264 | 0.20018 | 0.82057 | 0.82273 | 0.4150377162 | 0.86804 | 0.9235178232 | 0.131376 | 5.451186001 | 0 |
| 2 | 47412466 | C | T | S | F | MSH2 | ENST00000233146 | 0.32892 | 0.00603 | 0.22126 | 0.63958 | 0.72322 | 0.4443469532 | 0.93869 | 0.9205506444 | 0.0786473 | 5.076587807 | 0 |
| 2 | 47412468 | A | C | T | P | MSH2 | ENST00000233146 | 0.33516 | 0.00527 | 0.20182 | 0.74562 | 0.81703 | 0.6217551277 | 0.98268 | 0.5797885656 | 0.217371 | 4.121021241 | 0 |
| 2 | 47412468 | A | G | T | A | MSH2 | ENST00000233146 | 0.28315 | 0.0058 | 0.1979 | 0.73456 | 0.79273 | 0.2727243069 | 0.99245 | 0.1311257374 | 0.167872 | 4.244919973 | 0 |
| 2 | 47412468 | A | T | T | S | MSH2 | ENST00000233146 | 0.20986 | 0.00592 | 0.19764 | 0.41408 | 0.78535 | 0.1950911468 | 0.96894 | 0.04991851643 | 0.0999178 | 2.402068894 | 0 |
| 2 | 47412469 | C | A | T | K | MSH2 | ENST00000233146 | 0.20896 | 0.00681 | 0.2729 | 0.73972 | 0.83339 | 0.5388832181 | 0.89387 | 0.9774004817 | 0.211398 | 4.892668635 | 0 |
| 2 | 47412469 | C | G | T | R | MSH2 | ENST00000233146 | 0.2355 | 0.01019 | 0.26194 | 0.77481 | 0.82046 | 0.5926368262 | 0.8775 | 0.7570528388 | 0.21775 | 4.53127141 | 0 |
| 2 | 47412469 | C | T | T | I | MSH2 | ENST00000233146 | 0.22018 | 0.00909 | 0.31942 | 0.7381 | 0.81298 | 0.4289748845 | 0.8853 | 0.436973162 | 0.194301 | 4.537856212 | 0 |
| 2 | 47412471 | A | C | K | Q | MSH2 | ENST00000233146 | 0.80637 | 0.03595 | 0.43182 | 0.87016 | 0.94091 | 0.4803166198 | 0.92531 | 0.9950237274 | 0.313239 | 5.057413663 | 0 |
| 2 | 47412471 | A | G | K | E | MSH2 | ENST00000233146 | 0.6362 | 0.03001 | 0.48145 | 0.86179 | 0.94913 | 0.5594297943 | 0.95088 | 0.9369011521 | 0.422779 | 3.780257987 | 0 |
| 2 | 47412472 | A | C | K | T | MSH2 | ENST00000233146 | 0.78527 | 0.02724 | 0.48599 | 0.89475 | 0.96371 | 0.5502946134 | 0.92399 | 0.9988614321 | 0.438677 | 5.197739175 | 0 |
| 2 | 47412472 | A | G | K | R | MSH2 | ENST00000233146 | 0.67827 | 0.0394 | 0.4001 | 0.83348 | 0.89648 | 0.4103866595 | 0.94237 | 0.9629224539 | 0.28923 | 4.424523415 | 0 |
| 2 | 47412472 | A | T | K | I | MSH2 | ENST00000233146 | 0.86083 | 0.02835 | 0.45262 | 0.9227 | 0.98071 | 0.6368849088 | 0.92189 | 0.9988619089 | 0.440555 | 6.676685983 | 0 |
| 2 | 47412473 | A | C | K | N | MSH2 | ENST00000233146 | 0.54589 | 0.03426 | 0.50839 | 0.90414 | 0.84267 | 0.5821688707 | 0.92593 | 0.9956931472 | 0.140016 | 5.088831901 | 0 |
| 2 | 47412473 | A | T | K | N | MSH2 | ENST00000233146 | 0.54021 | 0.03426 | 0.50839 | 0.90613 | 0.84267 | 0.5821688707 | 0.92607 | 0.9956931472 | 0.14001 | 5.088831901 | 0 |
| 2 | 47412474 | G | A | D | N | MSH2 | ENST00000233146 | 0.5328 | 0.01096 | 0.59589 | 0.76692 | 0.81933 | 0.6041445911 | 0.84358 | 0.9848743081 | 0.0727201 | 4.950587236 | 0 |
| 2 | 47412474 | G | C | D | H | MSH2 | ENST00000233146 | 0.87768 | 0.03413 | 0.62189 | 0.88859 | 0.96993 | 0.6849666134 | 0.96203 | 0.9980134964 | 0.458267 | 6.187175789 | 0 |
| 2 | 47412474 | G | T | D | Y | MSH2 | ENST00000233146 | 0.88579 | 0.0337 | 0.60153 | 0.90158 | 0.96371 | 0.7469901958 | 0.9774 | 0.9993594289 | 0.478979 | 5.210475976 | 0 |
| 2 | 47412475 | A | C | D | A | MSH2 | ENST00000233146 | 0.84218 | 0.03137 | 0.53216 | 0.89685 | 0.97661 | 0.6985825998 | 0.94279 | 0.997541666 | 0.468592 | 5.417563041 | 0 |
| 2 | 47412475 | A | G | D | G | MSH2 | ENST00000233146 | 0.71353 | 0.01459 | 0.5495 | 0.8619 | 0.97252 | 0.6743427698 | 0.97821 | 0.9880902767 | 0.329713 | 5.852336029 | 0 |
| 2 | 47412475 | A | T | D | V | MSH2 | ENST00000233146 | 0.84264 | 0.03124 | 0.53044 | 0.90742 | 0.9729 | 0.7405931096 | 0.97647 | 0.9994176626 | 0.480757 | 6.562895857 | 0 |
| 2 | 47412476 | C | A | D | E | MSH2 | ENST00000233146 | 0.42044 | 0.02547 | 0.56802 | 0.86324 | 0.91357 | 0.4845466271 | 0.97625 | 0.9872861505 | 0.35565 | 4.710906862 | 0 |
| 2 | 47412476 | C | G | D | E | MSH2 | ENST00000233146 | 0.42675 | 0.02547 | 0.56802 | 0.8595 | 0.91357 | 0.4845466271 | 0.97619 | 0.9851775169 | 0.355651 | 4.710906862 | 0 |
| 2 | 47412477 | A | C | I | L | MSH2 | ENST00000233146 | 0.35476 | 0.00599 | 0.1987 | 0.70564 | 0.69425 | 0.372342662 | 0.89538 | 0.7301018834 | 0.0938197 | 3.751326338 | 0 |
| 2 | 47412477 | A | G | I | V | MSH2 | ENST00000233146 | 0.3126 | 0.00633 | 0.12249 | 0.50045 | 0.79273 | 0.1728673825 | 0.92995 | 0.5328033566 | 0.168595 | 1.629013774 | 0 |
| 2 | 47412477 | A | T | I | F | MSH2 | ENST00000233146 | 0.67605 | 0.02143 | 0.21433 | 0.89279 | 0.95826 | 0.5415426803 | 0.92836 | 0.9805186987 | 0.275492 | 4.163555094 | 0 |
| 2 | 47412478 | T | A | I | N | MSH2 | ENST00000233146 | 0.46308 | 0.00527 | 0.29256 | 0.87489 | 0.87148 | 0.6814193326 | 0.92186 | 0.9860851765 | 0.224458 | 2.396501748 | 0 |
| 2 | 47412478 | T | C | I | T | MSH2 | ENST00000233146 | 0.55628 | 0.00722 | 0.33397 | 0.90109 | 0.92804 | 0.4200541736 | 0.92125 | 0.9819062948 | 0.278273 | 3.606006526 | 0 |
| 2 | 47412478 | T | G | I | S | MSH2 | ENST00000233146 | 0.37301 | 0.00615 | 0.22608 | 0.84925 | 0.85326 | 0.6531144998 | 0.86745 | 0.9430510402 | 0.181802 | 3.208915077 | 0 |
| 2 | 47412480 | T | C | Y | H | MSH2 | ENST00000233146 | 0.61119 | 0.00532 | 0.25562 | 0.68847 | 0.82103 | 0.4407003282 | 0.85792 | 0.5480748415 | 0.138919 | 3.085064282 | 0 |
| 2 | 47412480 | T | G | Y | D | MSH2 | ENST00000233146 | 0.49635 | 0.0048 | 0.2605 | 0.7726 | 0.79938 | 0.6269496032 | 0.97457 | 0.7507205009 | 0.241604 | 2.663946194 | 0 |
| 2 | 47412481 | A | C | Y | S | MSH2 | ENST00000233146 | 0.26091 | 0.00543 | 0.19371 | 0.69826 | 0.83004 | 0.4803330624 | 0.97337 | 0.4374337792 | 0.174555 | 2.082807983 | 0 |
| 2 | 47412481 | A | G | Y | C | MSH2 | ENST00000233146 | 0.29163 | 0.00532 | 0.25194 | 0.63011 | 0.79998 | 0.4608781161 | 0.98391 | 0.1789399087 | 0.212495 | 3.273296364 | 0 |
| 2 | 47412481 | A | T | Y | F | MSH2 | ENST00000233146 | 0.21461 | 0.0071 | 0.18944 | 0.59762 | 0.80595 | 0.3264570106 | 0.9401 | 0.2790239155 | 0.102565 | 2.681329577 | 0 |
| 2 | 47412483 | C | A | Q | K | MSH2 | ENST00000233146 | 0.67421 | 0.03001 | 0.45048 | 0.88031 | 0.9367 | 0.6725715394 | 0.92485 | 0.9976325035 | 0.348384 | 5.449938424 | 0 |
| 2 | 47412483 | C | G | Q | E | MSH2 | ENST00000233146 | 0.50432 | 0.03313 | 0.39327 | 0.82249 | 0.87733 | 0.5906526129 | 0.86741 | 0.9947399497 | 0.263892 | 4.367738479 | 0 |
| 2 | 47412484 | A | C | Q | P | MSH2 | ENST00000233146 | 0.79737 | 0.03388 | 0.40227 | 0.94924 | 0.9579 | 0.8406929965 | 0.91366 | 0.9982627034 | 0.353694 | 7.167021099 | 0 |
| 2 | 47412484 | A | G | Q | R | MSH2 | ENST00000233146 | 0.74316 | 0.02037 | 0.44693 | 0.89736 | 0.93475 | 0.6958631647 | 0.95291 | 0.9778223634 | 0.353602 | 5.213924949 | 0 |
| 2 | 47412484 | A | T | Q | L | MSH2 | ENST00000233146 | 0.81747 | 0.02108 | 0.38228 | 0.9387 | 0.95499 | 0.6560620245 | 0.93486 | 0.9985803366 | 0.382125 | 7.237781705 | 0 |
| 2 | 47412485 | G | C | Q | H | MSH2 | ENST00000233146 | 0.58204 | 0.03774 | 0.44657 | 0.91308 | 0.93863 | 0.6396700042 | 0.95045 | 0.9984582663 | 0.309747 | 6.806810719 | 0 |
| 2 | 47412485 | G | T | Q | H | MSH2 | ENST00000233146 | 0.57631 | 0.03774 | 0.44657 | 0.92457 | 0.93863 | 0.6396700042 | 0.95078 | 0.9984582663 | 0.309743 | 6.806810719 | 0 |
| 2 | 47412486 | G | A | D | N | MSH2 | ENST00000233146 | 0.80306 | 0.03741 | 0.63047 | 0.91232 | 0.93083 | 0.7026535241 | 0.95006 | 0.9961330891 | 0.142658 | 5.799513626 | 0 |
| 2 | 47412486 | G | C | D | H | MSH2 | ENST00000233146 | 0.80108 | 0.03458 | 0.65633 | 0.93914 | 0.9844 | 0.7542821514 | 0.94235 | 0.9989491105 | 0.48432 | 6.970406025 | 0 |
| 2 | 47412486 | G | T | D | Y | MSH2 | ENST00000233146 | 0.80929 | 0.03398 | 0.63516 | 0.94327 | 0.97923 | 0.8038603818 | 0.9688 | 0.9997425675 | 0.489184 | 6.647527686 | 0 |
| 2 | 47412487 | A | C | D | A | MSH2 | ENST00000233146 | 0.76602 | 0.0337 | 0.56271 | 0.94947 | 0.96371 | 0.7695320159 | 0.91301 | 0.998971343 | 0.451992 | 5.284537595 | 0 |
| 2 | 47412487 | A | G | D | G | MSH2 | ENST00000233146 | 0.77124 | 0.03304 | 0.58343 | 0.94149 | 0.9648 | 0.7550309789 | 0.94086 | 0.9990392923 | 0.46337 | 6.803867105 | 0 |
| 2 | 47412487 | A | T | D | V | MSH2 | ENST00000233146 | 0.75766 | 0.03144 | 0.55782 | 0.95113 | 0.95826 | 0.7874396901 | 0.97605 | 0.999720037 | 0.482441 | 5.51893765 | 0 |
| 2 | 47412489 | C | A | L | I | MSH2 | ENST00000233146 | 0.68483 | 0.03527 | 0.55184 | 0.90055 | 0.88212 | 0.3611372401 | 0.85505 | 0.9915636778 | 0.162995 | 4.160210123 | 0 |
| 2 | 47412489 | C | G | L | V | MSH2 | ENST00000233146 | 0.68027 | 0.03633 | 0.48405 | 0.90487 | 0.90929 | 0.547493282 | 0.86181 | 0.9826160669 | 0.235627 | 4.871591765 | 0 |
| 2 | 47412489 | C | T | L | F | MSH2 | ENST00000233146 | 0.72212 | 0.03582 | 0.53829 | 0.91924 | 0.93083 | 0.6122313419 | 0.89144 | 0.9979841709 | 0.285559 | 5.352080441 | 0 |
| 2 | 47412490 | T | C | L | P | MSH2 | ENST00000233146 | 0.67708 | 0.03001 | 0.67999 | 0.95176 | 0.97178 | 0.9081763672 | 0.97883 | 0.9991832376 | 0.487088 | 9.043640576 | 1 |
| 2 | 47412490 | T | G | L | R | MSH2 | ENST00000233146 | 0.67255 | 0.03137 | 0.63653 | 0.95337 | 0.97215 | 0.8903567312 | 0.96235 | 0.9990748167 | 0.48682 | 9.369371912 | 1 |
| 2 | 47412492 | A | C | N | H | MSH2 | ENST00000233146 | 0.76792 | 0.03646 | 0.54369 | 0.8562 | 0.93319 | 0.5193028177 | 0.92852 | 0.9895675778 | 0.226051 | 6.734689377 | 0 |
| 2 | 47412492 | A | G | N | D | MSH2 | ENST00000233146 | 0.79002 | 0.0337 | 0.59909 | 0.8226 | 0.92069 | 0.5735051274 | 0.92011 | 0.9847652912 | 0.171673 | 6.267887656 | 0 |
| 2 | 47412492 | A | T | N | Y | MSH2 | ENST00000233146 | 0.81544 | 0.03398 | 0.58833 | 0.90774 | 0.97327 | 0.6197640427 | 0.93905 | 0.9970970154 | 0.451667 | 6.775638391 | 0 |
| 2 | 47412493 | A | C | N | T | MSH2 | ENST00000233146 | 0.63412 | 0.03626 | 0.51404 | 0.8159 | 0.89422 | 0.4307346437 | 0.87466 | 0.9868535995 | 0.151037 | 4.840665327 | 0 |
| 2 | 47412493 | A | G | N | S | MSH2 | ENST00000233146 | 0.72284 | 0.03901 | 0.45068 | 0.82623 | 0.83339 | 0.3746251421 | 0.91143 | 0.9426315427 | -0.0664677 | 5.151874431 | 0 |
| 2 | 47412493 | A | T | N | I | MSH2 | ENST00000233146 | 0.82264 | 0.0337 | 0.54212 | 0.91703 | 0.9506 | 0.6104068176 | 0.86797 | 0.9942665696 | 0.310248 | 5.44227158 | 0 |
| 2 | 47412494 | C | A | N | K | MSH2 | ENST00000233146 | 0.4937 | 0.02765 | 0.62784 | 0.82803 | 0.85482 | 0.5798186922 | 0.89558 | 0.9845501184 | 0.0848203 | 5.043002846 | 0 |
| 2 | 47412494 | C | G | N | K | MSH2 | ENST00000233146 | 0.50056 | 0.02765 | 0.62784 | 0.81111 | 0.85482 | 0.5798186922 | 0.89525 | 0.9584807158 | -0.0375962 | 5.043002846 | 0 |
| 2 | 47412495 | C | G | R | G | MSH2 | ENST00000233146 | 0.72548 | 0.03322 | 0.45934 | 0.94665 | 0.96008 | 0.6635953725 | 0.97222 | 0.9943197966 | 0.435228 | 6.596329262 | 0 |
| 2 | 47412496 | G | A | R | Q | MSH2 | ENST00000233146 | 0.93307 | 0.03491 | 0.38417 | 0.88263 | 0.9506 | 0.5801673197 | 0.95009 | 0.9889027476 | 0.282395 | 6.008412383 | 0 |
| 2 | 47412496 | G | C | R | P | MSH2 | ENST00000233146 | 0.93872 | 0.03166 | 0.52729 | 0.9427 | 0.98109 | 0.8245347119 | 0.96151 | 0.9983849525 | 0.480708 | 7.038899246 | 0 |
| 2 | 47412496 | G | T | R | L | MSH2 | ENST00000233146 | 0.93111 | 0.0337 | 0.50309 | 0.92285 | 0.95753 | 0.6622838605 | 0.97132 | 0.998942554 | 0.460147 | 7.206383645 | 0 |
| 2 | 47412498 | T | A | L | M | MSH2 | ENST00000233146 | 0.52002 | 0.03767 | 0.59624 | 0.88823 | 0.8933 | 0.4721463201 | 0.89776 | 0.9874474406 | 0.243276 | 7.185720979 | 0 |
| 2 | 47412498 | T | G | L | V | MSH2 | ENST00000233146 | 0.49751 | 0.03706 | 0.55791 | 0.89324 | 0.9182 | 0.5613808883 | 0.91574 | 0.9853964448 | 0.257626 | 6.092201085 | 0 |
| 2 | 47412499 | T | C | L | S | MSH2 | ENST00000233146 | 0.91644 | 0.03231 | 0.63517 | 0.9087 | 0.97698 | 0.7895439455 | 0.95267 | 0.999335587 | 0.477592 | 6.34321724 | 1 |
| 2 | 47412499 | T | G | L | W | MSH2 | ENST00000233146 | 0.91708 | 0.02984 | 0.70145 | 0.92284 | 0.97327 | 0.803784889 | 0.90706 | 0.9991675615 | 0.395286 | 9.317641736 | 0 |
| 2 | 47412500 | G | C | L | F | MSH2 | ENST00000233146 | 0.68379 | 0.03595 | 0.60837 | 0.88986 | 0.91736 | 0.5998965162 | 0.90728 | 0.9943615198 | 0.306115 | 6.055516229 | 0 |
| 2 | 47412500 | G | T | L | F | MSH2 | ENST00000233146 | 0.67732 | 0.03595 | 0.60837 | 0.89175 | 0.91736 | 0.5998965162 | 0.90757 | 0.9953149557 | 0.306113 | 6.055516229 | 0 |
| 2 | 47412501 | T | A | L | M | MSH2 | ENST00000233146 | 0.56152 | 0.03753 | 0.3955 | 0.89569 | 0.90362 | 0.4105454671 | 0.8867 | 0.9662991166 | 0.229279 | 6.560626643 | 0 |
| 2 | 47412501 | T | G | L | V | MSH2 | ENST00000233146 | 0.53731 | 0.03313 | 0.36651 | 0.84968 | 0.91186 | 0.4608557932 | 0.89548 | 0.9776253104 | 0.245264 | 5.487137405 | 0 |
| 2 | 47412502 | T | C | L | S | MSH2 | ENST00000233146 | 0.89423 | 0.03224 | 0.44512 | 0.94086 | 0.99072 | 0.7318647113 | 0.96363 | 0.998116374 | 0.494136 | 6.888796378 | 0 |
| 2 | 47412502 | T | G | L | W | MSH2 | ENST00000233146 | 0.89515 | 0.02984 | 0.52153 | 0.93571 | 0.9796 | 0.7480086878 | 0.92616 | 0.9989738464 | 0.42483 | 9.74030503 | 0 |
| 2 | 47412503 | G | C | L | F | MSH2 | ENST00000233146 | 0.63069 | 0.03582 | 0.41351 | 0.88602 | 0.90972 | 0.5365775476 | 0.88608 | 0.9949210286 | 0.276373 | 4.046765581 | 0 |

| | | | | | | | | | | | | | | | | |
|---|---|---|---|---|---|---|---|---|---|---|---|---|---|---|---|---|
| 2 | 47412503 | G | T | L | F | MSH2 | ENST00000233146 | 0.62453 | 0.03582 | 0.41351 | 0.88601 | 0.90972 | 0.5365775476 | 0.88636 | 0.9949210286 | 0.276368 | 4.046765581 | 0 |
| 2 | 47412504 | A | C | K | Q | MSH2 | ENST00000233146 | 0.85448 | 0.03379 | 0.44317 | 0.8575 | 0.96226 | 0.2788639299 | 0.94553 | 0.9806925654 | 0.376767 | 3.897441535 | 0 |
| 2 | 47412504 | A | G | K | E | MSH2 | ENST00000233146 | 0.70358 | 0.02914 | 0.4972 | 0.84823 | 0.91569 | 0.3362681978 | 0.88727 | 0.9860997796 | 0.270439 | 4.646645639 | 0 |
| 2 | 47412505 | A | C | K | T | MSH2 | ENST00000233146 | 0.7214 | 0.03212 | 0.50828 | 0.88869 | 0.91986 | 0.3732025529 | 0.83722 | 0.9920371771 | 0.219368 | 5.832857798 | 0 |
| 2 | 47412505 | A | G | K | R | MSH2 | ENST00000233146 | 0.38341 | 0.00799 | 0.40533 | 0.70202 | 0.79456 | 0.18226055 | 0.8954 | 0.4784414172 | 0.139495 | 2.089365823 | 0 |
| 2 | 47412505 | A | T | K | I | MSH2 | ENST00000233146 | 0.76879 | 0.02614 | 0.468 | 0.93999 | 0.94129 | 0.4120337303 | 0.89129 | 0.9972857237 | 0.251589 | 6.308701847 | 0 |
| 2 | 47412506 | A | C | K | N | MSH2 | ENST00000233146 | 0.86304 | 0.0151 | 0.53559 | 0.83974 | 0.89784 | 0.3322985594 | 0.84813 | 0.9805409312 | 0.156698 | 4.337761409 | 0 |
| 2 | 47412506 | A | T | K | N | MSH2 | ENST00000233146 | 0.85691 | 0.0151 | 0.53559 | 0.8395 | 0.89739 | 0.3322985594 | 0.84779 | 0.9805409312 | 0.156694 | 4.337761409 | 0 |
| 2 | 47412507 | G | A | G | S | MSH2 | ENST00000233146 | 0.35476 | 0.00574 | 0.24135 | 0.55435 | 0.81761 | 0.2178692365 | 0.96044 | 0.2512940764 | 0.110934 | 0.1720672108 | 0 |
| 2 | 47412507 | G | C | G | R | MSH2 | ENST00000233146 | 0.40121 | 0.00633 | 0.37727 | 0.81205 | 0.83888 | 0.3583314469 | 0.97986 | 0.2327372134 | 0.184909 | 2.545869502 | 0 |
| 2 | 47412507 | G | T | G | C | MSH2 | ENST00000233146 | 0.59769 | 0.00562 | 0.31127 | 0.84118 | 0.83779 | 0.3377585818 | 0.94014 | 0.3197589358 | 0.161784 | 2.077423279 | 0 |
| 2 | 47412508 | G | A | G | D | MSH2 | ENST00000233146 | 0.2745 | 0.0058 | 0.40162 | 0.6833 | 0.8153 | 0.2806986999 | 0.94197 | 0.5013739467 | 0.15522 | 1.960778935 | 0 |
| 2 | 47412508 | G | C | G | A | MSH2 | ENST00000233146 | 0.22281 | 0.00556 | 0.27379 | 0.65901 | 0.73661 | 0.2181503446 | 0.89414 | 0.3207725286 | 0.0696466 | -0.2636295406 | 0 |
| 2 | 47412508 | G | T | G | V | MSH2 | ENST00000233146 | 0.2512 | 0.00532 | 0.29414 | 0.74723 | 0.81876 | 0.3620241361 | 0.92324 | 0.3595912457 | 0.139751 | 0.6494994141 | 0 |
| 2 | 47412510 | A | C | K | Q | MSH2 | ENST00000233146 | 0.68755 | 0.01281 | 0.35712 | 0.70164 | 0.81124 | 0.3347371755 | 0.90074 | 0.8819244504 | 0.199319 | 3.605293451 | 0 |
| 2 | 47412510 | A | G | K | E | MSH2 | ENST00000233146 | 0.53846 | 0.0054 | 0.40814 | 0.70138 | 0.74867 | 0.3556168731 | 0.94501 | 0.4886037409 | 0.148794 | 4.253895873 | 0 |
| 2 | 47412511 | A | C | K | T | MSH2 | ENST00000233146 | 0.45438 | 0.00669 | 0.4244 | 0.7563 | 0.81472 | 0.3823936856 | 0.96076 | 0.3769128919 | 0.191896 | 3.924857929 | 0 |
| 2 | 47412511 | A | G | K | R | MSH2 | ENST00000233146 | 0.34163 | 0.00622 | 0.32279 | 0.49585 | 0.76487 | 0.2220831042 | 0.96432 | 0.1026924998 | 0.0760795 | 2.998081301 | 0 |
| 2 | 47412511 | A | T | K | I | MSH2 | ENST00000233146 | 0.49751 | 0.01649 | 0.37953 | 0.83396 | 0.86753 | 0.457006376 | 0.94982 | 0.9834449887 | 0.217631 | 4.46922068 | 0 |
| 2 | 47412512 | A | C | K | N | MSH2 | ENST00000233146 | 0.69163 | 0.01452 | 0.44153 | 0.77405 | 0.79212 | 0.3337038522 | 0.88256 | 0.81266886 | 0.092678 | 4.285283505 | 0 |
| 2 | 47412512 | A | T | K | N | MSH2 | ENST00000233146 | 0.68491 | 0.01452 | 0.44153 | 0.77405 | 0.79212 | 0.3337038522 | 0.8824 | 0.81266886 | 0.092687 | 4.285283505 | 0 |
| 2 | 47412513 | A | C | K | Q | MSH2 | ENST00000233146 | 0.76443 | 0.01396 | 0.48018 | 0.81507 | 0.83888 | 0.3492141959 | 0.9006 | 0.9291075468 | 0.243457 | 5.157134755 | 0 |
| 2 | 47412513 | A | G | K | E | MSH2 | ENST00000233146 | 0.50825 | 0.01831 | 0.52544 | 0.80576 | 0.86503 | 0.3674506122 | 0.96251 | 0.9137182832 | 0.356391 | 4.453469567 | 0 |
| 2 | 47412514 | A | C | K | T | MSH2 | ENST00000233146 | 0.7733 | 0.022 | 0.53318 | 0.87227 | 0.89513 | 0.3919172373 | 0.88037 | 0.9599597454 | 0.275579 | 3.187253819 | 0 |
| 2 | 47412514 | A | G | K | R | MSH2 | ENST00000233146 | 0.47407 | 0.00941 | 0.45448 | 0.74308 | 0.81761 | 0.2344552081 | 0.92777 | 0.8998414278 | 0.205801 | 3.454288554 | 0 |
| 2 | 47412514 | A | T | K | M | MSH2 | ENST00000233146 | 0.82404 | 0.0315 | 0.45655 | 0.92574 | 0.92316 | 0.4881950148 | 0.90106 | 0.9749677777 | 0.3006 | 5.949325291 | 0 |
| 2 | 47412515 | G | C | K | N | MSH2 | ENST00000233146 | 0.62886 | 0.00664 | 0.55724 | 0.71342 | 0.80536 | 0.3452151563 | 0.82679 | 0.8870933056 | 0.0802562 | 3.422492776 | 0 |
| 2 | 47412515 | G | T | K | N | MSH2 | ENST00000233146 | 0.62271 | 0.00664 | 0.55724 | 0.71815 | 0.80536 | 0.3452151563 | 0.82705 | 0.8870933056 | 0.0802499 | 3.422492776 | 0 |
| 2 | 47412516 | G | A | G | R | MSH2 | ENST00000233146 | 0.93592 | 0.02663 | 0.40768 | 0.93033 | 0.89873 | 0.5259209014 | 0.91015 | 0.9912912846 | 0.251032 | 4.462200131 | 0 |
| 2 | 47412516 | G | C | G | R | MSH2 | ENST00000233146 | 0.9348 | 0.02663 | 0.40768 | 0.92683 | 0.89873 | 0.5259209014 | 0.90729 | 0.9890152812 | 0.251041 | 4.462200131 | 0 |
| 2 | 47412517 | G | A | G | E | MSH2 | ENST00000233146 | 0.55909 | 0.00619 | 0.37133 | 0.6891 | 0.81472 | 0.4685133973 | 0.80761 | 0.9636919498 | 0.209133 | 1.742055828 | 0 |
| 2 | 47412517 | G | C | G | A | MSH2 | ENST00000233146 | 0.69364 | 0.01841 | 0.29174 | 0.77912 | 0.8517 | 0.3860678252 | 0.90643 | 0.9791316986 | 0.22756 | 7.595012439 | 0 |
| 2 | 47412517 | G | T | G | V | MSH2 | ENST00000233146 | 0.7266 | 0.00923 | 0.29356 | 0.84178 | 0.89963 | 0.4263687024 | 0.90004 | 0.9726516604 | 0.316139 | 3.693025994 | 0 |
| 2 | 47412519 | G | C | E | Q | MSH2 | ENST00000233146 | 0.51362 | 0.00599 | 0.34659 | 0.7688 | 0.78348 | 0.3540140486 | 0.94243 | 0.5594995618 | 0.22339 | 3.312444306 | 0 |
| 2 | 47412520 | A | C | E | A | MSH2 | ENST00000233146 | 0.73437 | 0.00978 | 0.36251 | 0.82437 | 0.85326 | 0.3867789476 | 0.87739 | 0.9683640003 | 0.257654 | 7.671793614 | 0 |
| 2 | 47412520 | A | G | E | G | MSH2 | ENST00000233146 | 0.75424 | 0.00574 | 0.31443 | 0.78035 | 0.83888 | 0.357477932 | 0.92588 | 0.9579388499 | 0.25871 | 3.845798289 | 0 |
| 2 | 47412520 | A | T | E | V | MSH2 | ENST00000233146 | 0.91712 | 0.02307 | 0.32622 | 0.88247 | 0.94015 | 0.4194639195 | 0.92054 | 0.9834957719 | 0.309229 | 5.031315682 | 0 |
| 2 | 47412521 | G | C | E | D | MSH2 | ENST00000233146 | 0.20579 | 0.00608 | 0.35501 | 0.62961 | 0.7872 | 0.25511764 | 0.96462 | 0.1115943044 | 0.137641 | 2.887015952 | 0 |
| 2 | 47412521 | G | T | E | D | MSH2 | ENST00000233146 | 0.20264 | 0.00608 | 0.35501 | 0.62952 | 0.7872 | 0.25511764 | 0.96483 | 0.151863575 | 0.137638 | 2.887015952 | 0 |
| 2 | 47412522 | C | A | Q | K | MSH2 | ENST00000233146 | 0.25489 | 0.00535 | 0.30515 | 0.63084 | 0.79577 | 0.3512389235 | 0.91972 | 0.6012493372 | 0.202734 | 3.065431859 | 0 |
| 2 | 47412522 | C | G | Q | E | MSH2 | ENST00000233146 | 0.26377 | 0.00562 | 0.2529 | 0.65416 | 0.80238 | 0.3047493433 | 0.89264 | 0.4693272114 | 0.167736 | 3.619740903 | 0 |
| 2 | 47412523 | A | C | Q | P | MSH2 | ENST00000233146 | 0.62248 | 0.01009 | 0.26055 | 0.73501 | 0.84535 | 0.4829237796 | 0.97018 | 0.770898819 | 0.321162 | 7.445956314 | 0 |
| 2 | 47412523 | A | G | Q | R | MSH2 | ENST00000233146 | 0.46599 | 0.00747 | 0.29892 | 0.6958 | 0.85792 | 0.3163635309 | 0.95198 | 0.5331657529 | 0.248623 | 4.886839207 | 0 |
| 2 | 47412523 | A | T | Q | L | MSH2 | ENST00000233146 | 0.50276 | 0.0058 | 0.24285 | 0.79558 | 0.8367 | 0.3613403491 | 0.96075 | 0.8253360391 | 0.263684 | 7.445956314 | 0 |
| 2 | 47412524 | G | C | Q | H | MSH2 | ENST00000233146 | 0.41469 | 0.00603 | 0.29227 | 0.76241 | 0.82273 | 0.4087203781 | 0.98122 | 0.1085446253 | 0.0789597 | 4.88835215 | 0 |
| 2 | 47412524 | G | T | Q | H | MSH2 | ENST00000233146 | 0.40987 | 0.00603 | 0.29227 | 0.8162 | 0.82273 | 0.4087203781 | 0.98123 | 0.3113875985 | 0.205331 | 4.88835215 | 0 |
| 2 | 47412525 | A | C | M | L | MSH2 | ENST00000233146 | 0.29954 | 0.0058 | 0.15745 | 0.51342 | 0.78223 | 0.2239219917 | 0.92786 | 0.04878255725 | 0.0269727 | 0.0179014662 | 0 |
| 2 | 47412525 | A | G | M | V | MSH2 | ENST00000233146 | 0.25498 | 0.00566 | 0.11236 | 0.57117 | 0.76356 | 0.2397831387 | 0.90663 | 0.03195350207 | 0.0348234 | -2.096738342 | 0 |
| 2 | 47412525 | A | T | M | L | MSH2 | ENST00000233146 | 0.29532 | 0.0058 | 0.15745 | 0.51342 | 0.78223 | 0.2239219917 | 0.92745 | 0.04878255725 | 0.026972 | 0.0179014662 | 0 |
| 2 | 47412526 | T | A | M | K | MSH2 | ENST00000233146 | 0.22038 | 0.00556 | 0.22285 | 0.66705 | 0.77654 | 0.4542889102 | 0.88977 | 0.3052513208 | 0.181288 | 1.91433504 | 0 |
| 2 | 47412526 | T | C | M | T | MSH2 | ENST00000233146 | 0.22013 | 0.00543 | 0.27029 | 0.5718 | 0.72926 | 0.3067615944 | 0.93053 | 0.3149370528 | 0.171988 | 1.441115096 | 0 |
| 2 | 47412526 | T | G | M | R | MSH2 | ENST00000233146 | 0.25981 | 0.00627 | 0.20813 | 0.71723 | 0.77463 | 0.467190896 | 0.84335 | 0.3212630656 | 0.156305 | 0.8762912659 | 0 |
| 2 | 47412528 | A | C | N | H | MSH2 | ENST00000233146 | 0.68555 | 0.03409 | 0.48839 | 0.85688 | 0.87296 | 0.2251819725 | 0.89705 | 0.9819511175 | 0.124171 | 7.533991768 | 0 |
| 2 | 47412528 | A | G | N | D | MSH2 | ENST00000233146 | 0.41005 | 0.02077 | 0.53734 | 0.79245 | 0.76685 | 0.2001382447 | 0.88329 | 0.9483432174 | 0.0971015 | 3.197744049 | 0 |
| 2 | 47412528 | A | T | N | Y | MSH2 | ENST00000233146 | 0.73373 | 0.0318 | 0.51559 | 0.90806 | 0.91611 | 0.2816054261 | 0.89265 | 0.9909781218 | 0.234503 | 7.533991768 | 0 |
| 2 | 47412529 | A | C | N | T | MSH2 | ENST00000233146 | 0.28901 | 0.00644 | 0.46618 | 0.66381 | 0.80358 | 0.1745929482 | 0.90024 | 0.7629153728 | 0.127599 | 3.476019226 | 0 |
| 2 | 47412529 | A | G | N | S | MSH2 | ENST00000233146 | 0.22208 | 0.00687 | 0.40376 | 0.65991 | 0.5927 | 0.1295876652 | 0.8332 | 0.9524751902 | 0.0660119 | 1.043901728 | 0 |
| 2 | 47412529 | A | T | N | I | MSH2 | ENST00000233146 | 0.58688 | 0.02414 | 0.4878 | 0.89867 | 0.91314 | 0.2970236691 | 0.88887 | 0.9880937934 | 0.220837 | 6.726143447 | 0 |
| 2 | 47412530 | T | A | N | K | MSH2 | ENST00000233146 | 0.58774 | 0.00872 | 0.56346 | 0.79451 | 0.81124 | 0.2282920092 | 0.93615 | 0.8099267483 | 0.0813916 | 3.330026928 | 0 |
| 2 | 47412530 | T | G | N | K | MSH2 | ENST00000233146 | 0.59508 | 0.00872 | 0.56346 | 0.78857 | 0.81124 | 0.2282920092 | 0.93624 | 0.8099267483 | 0.081361 | 3.330026928 | 0 |
| 2 | 47412531 | A | C | S | R | MSH2 | ENST00000233146 | 0.79997 | 0.03431 | 0.60775 | 0.89722 | 0.9703 | 0.4928592229 | 0.96299 | 0.9841275811 | 0.438443 | 6.855638115 | 0 |
| 2 | 47412531 | A | G | S | G | MSH2 | ENST00000233146 | 0.74834 | 0.02914 | 0.48058 | 0.84263 | 0.93241 | 0.2661491613 | 0.93998 | 0.8962070346 | 0.368297 | 5.771937724 | 0 |
| 2 | 47412532 | G | A | S | N | MSH2 | ENST00000233146 | 0.59733 | 0.03697 | 0.59948 | 0.84342 | 0.87588 | 0.3649445318 | 0.85029 | 0.7643057969 | 0.106314 | 5.663965616 | 0 |
| 2 | 47412532 | G | C | S | T | MSH2 | ENST00000233146 | 0.61008 | 0.03313 | 0.54233 | 0.78564 | 0.87099 | 0.3069602278 | 0.92982 | 0.9600311518 | 0.298613 | 5.421503333 | 0 |
| 2 | 47412532 | G | T | S | I | MSH2 | ENST00000233146 | 0.82623 | 0.03046 | 0.56255 | 0.86221 | 0.94243 | 0.4554544905 | 0.96155 | 0.9872752428 | 0.433764 | 5.036209888 | 0 |
| 2 | 47412533 | T | A | S | R | MSH2 | ENST00000233146 | 0.55724 | 0.03431 | 0.60775 | 0.92767 | 0.93083 | 0.4928592229 | 0.97789 | 0.9769823551 | 0.390044 | 6.855638115 | 0 |
| 2 | 47412533 | T | G | S | R | MSH2 | ENST00000233146 | 0.56435 | 0.03431 | 0.60775 | 0.92104 | 0.93044 | 0.4928592229 | 0.9777 | 0.9707540274 | 0.390045 | 6.855638115 | 0 |
| 2 | 47412534 | G | A | A | T | MSH2 | ENST00000233146 | 0.44807 | 0.00622 | 0.27648 | 0.66913 | 0.86803 | 0.3524254221 | 0.93055 | 0.127814734 | -0.0190095 | 4.874550904 | 0 |
| 2 | 47412534 | G | C | A | P | MSH2 | ENST00000233146 | 0.6329 | 0.02237 | 0.34242 | 0.9181 | 0.94129 | 0.5456525767 | 0.95309 | 0.9769445658 | 0.409644 | 6.514829171 | 0 |
| 2 | 47412534 | G | T | A | S | MSH2 | ENST00000233146 | 0.40232 | 0.00687 | 0.24099 | 0.7524 | 0.79697 | 0.2818669794 | 0.86989 | 0.4757323265 | 0.149361 | 3.170763833 | 0 |
| 2 | 47412535 | C | A | A | D | MSH2 | ENST00000233146 | 0.43418 | 0.01682 | 0.39151 | 0.8599 | 0.90972 | 0.5020877301 | 0.91989 | 0.9514315128 | 0.391278 | 4.270027998 | 0 |
| 2 | 47412535 | C | G | A | G | MSH2 | ENST00000233146 | 0.28179 | 0.00599 | 0.26245 | 0.72556 | 0.84159 | 0.27665036 | 0.93485 | 0.2117834985 | 0.0959876 | 5.104449642 | 0 |
| 2 | 47412535 | C | T | A | V | MSH2 | ENST00000233146 | 0.42992 | 0.00648 | 0.29941 | 0.70637 | 0.90755 | 0.3319442583 | 0.85694 | 0.9603006244 | 0.264674 | 6.434513396 | 0 |
| 2 | 47412537 | G | A | V | I | MSH2 | ENST00000233146 | 0.36548 | 0.00674 | 0.19941 | 0.70135 | 0.76223 | 0.1942754333 | 0.85963 | 0.4335706234 | 0.15123 | 2.156504652 | 0 |
| 2 | 47412537 | G | C | V | L | MSH2 | ENST00000233146 | 0.40523 | 0.00638 | 0.30185 | 0.72183 | 0.7589 | 0.2705294382 | 0.9082 | 0.3984076102 | 0.192042 | 1.660327043 | 0 |
| 2 | 47412537 | G | T | V | L | MSH2 | ENST00000233146 | 0.40045 | 0.00638 | 0.30185 | 0.74024 | 0.75957 | 0.2705294382 | 0.90841 | 0.3746900873 | 0.192025 | 1.660327043 | 0 |
| 2 | 47412538 | T | A | V | E | MSH2 | ENST00000233146 | 0.42115 | 0.00562 | 0.30655 | 0.86083 | 0.85843 | 0.4986061785 | 0.9868 | 0.6161498427 | 0.26003 | 1.769857152 | 0 |
| 2 | 47412538 | T | C | V | A | MSH2 | ENST00000233146 | 0.21371 | 0.00535 | 0.31678 | 0.66404 | 0.77011 | 0.2657146989 | 0.87632 | 0.139439851 | 0.092444 | -1.803645343 | 0 |
| 2 | 47412538 | T | G | V | G | MSH2 | ENST00000233146 | 0.4131 | 0.0058 | 0.21905 | 0.83681 | 0.82046 | 0.3575175099 | 0.97755 | 0.4404756129 | 0.260635 | 0.3328633049 | 0 |
| 2 | 47412540 | T | A | L | M | MSH2 | ENST00000233146 | 0.58168 | 0.03761 | 0.57503 | 0.87813 | 0.90842 | 0.3099131783 | 0.86467 | 0.8990844488 | 0.298001 | 4.827842165 | 0 |
| 2 | 47412540 | T | G | L | V | MSH2 | ENST00000233146 | 0.55793 | 0.03674 | 0.54137 | 0.89645 | 0.91945 | 0.3472104553 | 0.88658 | 0.9723570347 | 0.294891 | 5.563304088 | 0 |
| 2 | 47412541 | T | C | L | S | MSH2 | ENST00000233146 | 0.8145 | 0.03001 | 0.61455 | 0.94101 | 0.97848 | 0.648509039 | 0.96639 | 0.9960826635 | 0.517588 | 6.323051752 | 0 |
| 2 | 47412541 | T | G | L | W | MSH2 | ENST00000233146 | 0.92497 | 0.02979 | 0.67728 | 0.94676 | 0.97885 | 0.6084203596 | 0.93135 | 0.9989957213 | 0.480674 | 8.157061927 | 0 |
| 2 | 47412542 | G | C | L | F | MSH2 | ENST00000233146 | 0.80929 | 0.03161 | 0.58243 | 0.91853 | 0.9608 | 0.4110778277 | 0.92784 | 0.991368711 | 0.424988 | 4.176118265 | 0 |
| 2 | 47412542 | G | T | L | F | MSH2 | ENST00000233146 | 0.8029 | 0.03161 | 0.58243 | 0.92004 | 0.9608 | 0.4110778277 | 0.92805 | 0.9820528626 | 0.424986 | 4.176118265 | 0 |
| 2 | 47412543 | C | A | P | T | MSH2 | ENST00000233146 | 0.67016 | 0.02072 | 0.62584 | 0.93446 | 0.93977 | 0.4217719587 | 0.92147 | 0.9953536987 | 0.433638 | 6.495823919 | 0 |
| 2 | 47412543 | C | G | P | A | MSH2 | ENST00000233146 | 0.48563 | 0.00754 | 0.57117 | 0.86305 | 0.89055 | 0.3337029571 | 0.89239 | 0.9852767587 | 0.294837 | 2.748459802 | 0 |
| 2 | 47412543 | C | T | P | S | MSH2 | ENST00000233146 | 0.54372 | 0.00889 | 0.59251 | 0.86996 | 0.91015 | 0.4283908549 | 0.84361 | 0.8489914536 | 0.266294 | 2.89515039 | 0 |
| 2 | 47412544 | C | A | P | Q | MSH2 | ENST00000233146 | 0.66122 | 0.03313 | 0.60233 | 0.93079 | 0.96117 | 0.4107922991 | 0.90486 | 0.9983002543 | 0.395763 | 4.68942605 | 0 |
| 2 | 47412544 | C | G | P | R | MSH2 | ENST00000233146 | 0.66945 | 0.00978 | 0.66371 | 0.92159 | 0.97698 | 0.5068813722 | 0.94901 | 0.9975968003 | 0.510157 | 5.823616448 | 0 |
| 2 | 47412544 | C | T | P | L | MSH2 | ENST00000233146 | 0.65374 | 0.02547 | 0.67278 | 0.88727 | 0.95097 | 0.4916717328 | 0.92903 | 0.9989769459 | 0.381669 | 6.541730551 | 0 |
| 2 | 47412546 | G | A | E | K | MSH2 | ENST00000233146 | 0.92785 | 0.03388 | 0.64251 | 0.9188 | 0.9659 | 0.5828362288 | 0.9503 | 0.9876487851 | 0.44879 | 5.340806466 | 0 |
| 2 | 47412546 | G | C | E | Q | MSH2 | ENST00000233146 | 0.73197 | 0.02937 | 0.50523 | 0.7773 | 0.86098 | 0.5123263841 | 0.92648 | 0.9424368739 | 0.277236 | 4.701653882 | 0 |
| 2 | 47412547 | A | C | E | A | MSH2 | ENST00000233146 | 0.87847 | 0.0337 | 0.52568 | 0.93134 | 0.97661 | 0.5626012911 | 0.90818 | 0.9961867929 | 0.375635 | 5.446423116 | 0 |
| 2 | 47412547 | A | G | E | G | MSH2 | ENST00000233146 | 0.91036 | 0.01915 | 0.46462 | 0.92391 | 0.96371 | 0.5722840084 | 0.89679 | 0.9972321391 | 0.441126 | 6.10823877 | 0 |
| 2 | 47412547 | A | T | E | V | MSH2 | ENST00000233146 | 0.9017 | 0.03419 | 0.49377 | 0.93273 | 0.98549 | 0.5886011419 | 0.94334 | 0.9943538904 | 0.444235 | 5.296154177 | 0 |
| 2 | 47412548 | A | C | E | D | MSH2 | ENST00000233146 | 0.6978 | 0.03761 | 0.52396 | 0.89274 | 0.88448 | 0.4107740525 | 0.95492 | 0.9845083952 | 0.291581 | 5.279525836 | 0 |
| 2 | 47412548 | A | T | E | D | MSH2 | ENST00000233146 | 0.69107 | 0.03761 | 0.52396 | 0.89471 | 0.88448 | 0.4107740525 | 0.95554 | 0.9853928685 | 0.291577 | 5.279525836 | 0 |
| 2 | 47412549 | A | C | M | L | MSH2 | ENST00000233146 | 0.43093 | 0.00596 | 0.53724 | 0.47367 | 0.68381 | 0.2966256449 | 0.87198 | 0.5323731303 | 0.0830005 | 2.416087316 | 0 |
| 2 | 47412549 | A | G | M | V | MSH2 | ENST00000233146 | 0.37228 | 0.00586 | 0.46714 | 0.57945 | 0.7727 | 0.3852856993 | 0.9211 | 0.8161339164 | 0.158255 | 3.822485896 | 0 |
| 2 | 47412549 | A | T | M | L | MSH2 | ENST00000233146 | 0.42585 | 0.00596 | 0.53724 | 0.47367 | 0.68381 | 0.2966256449 | 0.87071 | 0.5323731303 | 0.0830221 | 2.416087316 | 0 |

| | | | | | | | | | | | | | | | | |
|---|---|---|---|---|---|---|---|---|---|---|---|---|---|---|---|---|
| 2 | 47412550 | T | A | M | K | MSH2 | ENST00000233146 | 0.80511 | 0.01766 | 0.63102 | 0.84522 | 0.93123 | 0.6575319447 | 0.89707 | 0.9900349975 | 0.266671 | 4.397326287 | 0 |
| 2 | 47412550 | T | C | M | T | MSH2 | ENST00000233146 | 0.42579 | 0.0134 | 0.67452 | 0.74639 | 0.82273 | 0.5193161618 | 0.84977 | 0.6174576283 | 0.135492 | 4.111302731 | 0 |
| 2 | 47412550 | T | G | M | R | MSH2 | ENST00000233146 | 0.83852 | 0.023 | 0.60827 | 0.82599 | 0.94205 | 0.7102136981 | 0.88793 | 0.9903309345 | 0.31689 | 5.396072784 | 0 |
| 2 | 47412551 | G | A | M | I | MSH2 | ENST00000233146 | 0.43016 | 0.0058 | 0.54797 | 0.72377 | 0.76289 | 0.3264282153 | 0.84919 | 0.7528046966 | 0.163925 | 3.640018639 | 0 |
| 2 | 47412551 | G | C | M | I | MSH2 | ENST00000233146 | 0.4277 | 0.0058 | 0.54797 | 0.72119 | 0.76289 | 0.3264282153 | 0.8472 | 0.7621446252 | 0.163925 | 3.640018639 | 0 |
| 2 | 47412551 | G | T | M | I | MSH2 | ENST00000233146 | 0.42288 | 0.0058 | 0.54797 | 0.72377 | 0.76289 | 0.3264282153 | 0.84728 | 0.7528046966 | 0.163925 | 3.640018639 | 0 |
| 2 | 47412552 | G | A | E | K | MSH2 | ENST00000233146 | 0.60164 | 0.01002 | 0.76226 | 0.80219 | 0.89739 | 0.3015911632 | 0.9365 | 0.9651043415 | 0.267591 | 5.176370212 | 0 |
| 2 | 47412552 | G | C | E | Q | MSH2 | ENST00000233146 | 0.6766 | 0.03189 | 0.65732 | 0.72416 | 0.8705 | 0.3041665532 | 0.93398 | 0.9491477609 | 0.242306 | 5.353462628 | 0 |
| 2 | 47412553 | A | C | E | A | MSH2 | ENST00000233146 | 0.71802 | 0.01848 | 0.67008 | 0.8856 | 0.92723 | 0.3447551762 | 0.87953 | 0.9929579496 | 0.263329 | 5.823529241 | 0 |
| 2 | 47412553 | A | G | E | G | MSH2 | ENST00000233146 | 0.77124 | 0.02043 | 0.62123 | 0.88217 | 0.93475 | 0.321922746 | 0.85175 | 0.9943025112 | 0.269164 | 4.408966371 | 0 |
| 2 | 47412553 | A | T | E | V | MSH2 | ENST00000233146 | 0.9057 | 0.02984 | 0.64605 | 0.91171 | 0.94949 | 0.3991427338 | 0.94441 | 0.9978570342 | 0.312492 | 6.596224755 | 0 |
| 2 | 47412554 | G | C | E | D | MSH2 | ENST00000233146 | 0.31364 | 0.00615 | 0.66695 | 0.57205 | 0.76816 | 0.1818994597 | 0.95636 | 0.03383370223 | 0.160521 | 1.578338438 | 0 |
| 2 | 47412554 | G | T | E | D | MSH2 | ENST00000233146 | 0.30946 | 0.00615 | 0.66695 | 0.56887 | 0.76816 | 0.1818994597 | 0.95074 | 0.174676761 | 0.160554 | 1.578338438 | 0 |
| 2 | 47412555 | A | C | N | H | MSH2 | ENST00000233146 | 0.43914 | 0.00599 | 0.46729 | 0.82452 | 0.74728 | 0.2415796665 | 0.91154 | 0.7716249228 | 0.174943 | 2.730897028 | 0 |
| 2 | 47412555 | A | G | N | D | MSH2 | ENST00000233146 | 0.48896 | 0.00566 | 0.51395 | 0.82729 | 0.81065 | 0.2152369859 | 0.91416 | 0.7918103933 | 0.171147 | 2.123062844 | 0 |
| 2 | 47412555 | A | T | N | Y | MSH2 | ENST00000233146 | 0.62098 | 0.01376 | 0.49297 | 0.90874 | 0.89873 | 0.2829973885 | 0.89018 | 0.9250774384 | 0.283513 | 3.04742035 | 0 |
| 2 | 47412556 | A | C | N | T | MSH2 | ENST00000233146 | 0.5465 | 0.01048 | 0.46094 | 0.86172 | 0.82724 | 0.1822806144 | 0.92506 | 0.8598690629 | 0.22731 | 3.209480948 | 0 |
| 2 | 47412556 | A | G | N | S | MSH2 | ENST00000233146 | 0.49151 | 0.00889 | 0.40768 | 0.82085 | 0.7816 | 0.1434810002 | 0.9262 | 0.7752548456 | 0.169794 | 2.500895159 | 0 |
| 2 | 47412556 | A | T | N | I | MSH2 | ENST00000233146 | 0.58431 | 0.00773 | 0.46718 | 0.90707 | 0.81006 | 0.3176931242 | 0.91291 | 0.9131017327 | 0.251572 | 4.128394276 | 0 |
| 2 | 47412557 | T | A | N | K | MSH2 | ENST00000233146 | 0.28665 | 0.00547 | 0.52633 | 0.54058 | 0.69594 | 0.1941020973 | 0.86469 | 0.4170852602 | 0.0832971 | -1.091959551 | 0 |
| 2 | 47412557 | T | G | N | K | MSH2 | ENST00000233146 | 0.29173 | 0.00547 | 0.52633 | 0.54424 | 0.69594 | 0.1941020973 | 0.8642 | 0.1648843288 | 0.0832748 | -1.091959551 | 0 |
| 2 | 47412558 | C | A | Q | K | MSH2 | ENST00000233146 | 0.37323 | 0.00535 | 0.43748 | 0.65273 | 0.77972 | 0.5481283195 | 0.83266 | 0.5980745554 | 0.183508 | 2.701407218 | 0 |
| 2 | 47412558 | C | G | Q | E | MSH2 | ENST00000233146 | 0.36704 | 0.00958 | 0.37628 | 0.72323 | 0.77399 | 0.462319186 | 0.85653 | 0.6093080044 | 0.215384 | 3.383002112 | 0 |
| 2 | 47412559 | A | C | Q | P | MSH2 | ENST00000233146 | 0.43871 | 0.00562 | 0.39232 | 0.75417 | 0.86951 | 0.7537872755 | 0.96468 | 0.945433557 | 0.396644 | 3.297539312 | 0 |
| 2 | 47412559 | A | G | Q | R | MSH2 | ENST00000233146 | 0.44442 | 0.0114 | 0.42969 | 0.73836 | 0.8887 | 0.5613064971 | 0.94671 | 0.9352338314 | 0.299516 | 4.875129308 | 0 |
| 2 | 47412559 | A | T | Q | L | MSH2 | ENST00000233146 | 0.46693 | 0.00608 | 0.36503 | 0.84681 | 0.90842 | 0.5440963534 | 0.94798 | 0.9742096066 | 0.365884 | 4.43334385 | 0 |
| 2 | 47412560 | G | C | Q | H | MSH2 | ENST00000233146 | 0.55271 | 0.00715 | 0.42688 | 0.83577 | 0.91229 | 0.5069222446 | 0.87144 | 0.9740983844 | 0.26649 | 4.588978132 | 0 |
| 2 | 47412560 | G | T | Q | H | MSH2 | ENST00000233146 | 0.54711 | 0.00715 | 0.42688 | 0.83586 | 0.91229 | 0.5069222446 | 0.87148 | 0.9740983844 | 0.26649 | 4.588978132 | 0 |
| 2 | 47414269 | G | A | V | I | MSH2 | ENST00000233146 | 0.33806 | 0.00696 | 0.30205 | 0.65644 | 0.68558 | 0.1703781992 | 0.88659 | 0.786718905 | 0.123256 | 2.299145484 | 0 |
| 2 | 47414269 | G | C | V | L | MSH2 | ENST00000233146 | 0.32041 | 0.00622 | 0.48371 | 0.51567 | 0.7147 | 0.3508895524 | 0.96547 | 0.9307223558 | 0.239491 | 2.914464338 | 0 |
| 2 | 47414269 | G | T | V | F | MSH2 | ENST00000233146 | 0.35825 | 0.023 | 0.45409 | 0.79096 | 0.84321 | 0.624081441 | 0.95638 | 0.936938405 | 0.371583 | 4.427054585 | 0 |
| 2 | 47414270 | T | A | V | D | MSH2 | ENST00000233146 | 0.47375 | 0.01654 | 0.51263 | 0.87614 | 0.9796 | 0.7454118976 | 0.97332 | 0.991409719 | 0.431792 | 4.681093204 | 0 |
| 2 | 47414270 | T | C | V | A | MSH2 | ENST00000233146 | 0.38249 | 0.00586 | 0.50671 | 0.63597 | 0.83339 | 0.3802633204 | 0.93098 | 0.8556112647 | 0.244755 | 3.316545627 | 0 |
| 2 | 47414270 | T | G | V | G | MSH2 | ENST00000233146 | 0.44283 | 0.01479 | 0.36926 | 0.78661 | 0.97252 | 0.5497498863 | 0.96484 | 0.9883338213 | 0.430209 | 4.772211623 | 0 |
| 2 | 47414272 | G | A | A | T | MSH2 | ENST00000233146 | 0.85814 | 0.03144 | 0.54732 | 0.91035 | 0.97661 | 0.5643606647 | 0.9585 | 0.9946974516 | 0.490039 | 6.456226985 | 0 |
| 2 | 47414272 | G | C | A | P | MSH2 | ENST00000233146 | 0.86928 | 0.03313 | 0.60925 | 0.89434 | 0.98764 | 0.8386542912 | 0.96763 | 0.9981677532 | 0.488647 | 7.854980465 | 0 |
| 2 | 47414272 | G | T | A | S | MSH2 | ENST00000233146 | 0.67955 | 0.02595 | 0.49536 | 0.81753 | 0.9506 | 0.4566113125 | 0.91337 | 0.9144357246 | 0.351756 | 5.496189247 | 0 |
| 2 | 47414273 | C | A | A | E | MSH2 | ENST00000233146 | 0.61865 | 0.03452 | 0.61543 | 0.91097 | 0.99072 | 0.8265691206 | 0.96895 | 0.9983391762 | 0.480062 | 6.062398461 | 1 |
| 2 | 47414273 | C | G | A | G | MSH2 | ENST00000233146 | 0.60845 | 0.0357 | 0.52378 | 0.82952 | 0.95535 | 0.4763623085 | 0.92757 | 0.990450561 | 0.403194 | 4.39328514 | 0 |
| 2 | 47414273 | C | T | A | V | MSH2 | ENST00000233146 | 0.63252 | 0.0366 | 0.56321 | 0.9046 | 0.97885 | 0.6135064709 | 0.95444 | 0.9974405766 | 0.479397 | 6.106067989 | 0 |
| 2 | 47414275 | G | A | V | I | MSH2 | ENST00000233146 | 0.41688 | 0.00615 | 0.04264 | 0.26632 | 0.70262 | 0.1110362502 | 0.90669 | 0.5115439892 | 0.0708809 | 0.703399931 | 0 |
| 2 | 47414275 | G | C | V | L | MSH2 | ENST00000233146 | 0.45388 | 0.00586 | 0.11407 | 0.36869 | 0.74235 | 0.1943146394 | 0.93815 | 0.8822668195 | 0.0925652 | 2.812524492 | 0 |
| 2 | 47414275 | G | T | V | F | MSH2 | ENST00000233146 | 0.59828 | 0.01037 | 0.11501 | 0.6391 | 0.80831 | 0.479594632 | 0.90869 | 0.8300862908 | 0.193997 | 4.769111979 | 0 |
| 2 | 47414276 | T | A | V | D | MSH2 | ENST00000233146 | 0.54576 | 0.01285 | 0.14899 | 0.80469 | 0.87247 | 0.6756614428 | 0.98155 | 0.9855753183 | 0.295999 | 6.987171803 | 1 |
| 2 | 47414276 | T | C | V | A | MSH2 | ENST00000233146 | 0.38392 | 0.00543 | 0.14488 | 0.31668 | 0.74587 | 0.2824843073 | 0.94409 | 0.3856915086 | 0.103396 | 2.903048989 | 0 |
| 2 | 47414276 | T | G | V | G | MSH2 | ENST00000233146 | 0.52201 | 0.00763 | 0.06741 | 0.71041 | 0.8517 | 0.4611952905 | 0.94643 | 0.977009654 | 0.237248 | 5.704996798 | 0 |
| 2 | 47414278 | T | A | S | T | MSH2 | ENST00000233146 | 0.4227 | 0.00958 | 0.43181 | 0.69604 | 0.8345 | 0.4632362405 | 0.91692 | 0.6452468038 | 0.180047 | 3.516451494 | 0 |
| 2 | 47414278 | T | C | S | P | MSH2 | ENST00000233146 | 0.49641 | 0.01861 | 0.45109 | 0.71708 | 0.86852 | 0.8042771621 | 0.95909 | 0.9680979848 | 0.338644 | 5.347822467 | 0 |
| 2 | 47414278 | T | G | S | A | MSH2 | ENST00000233146 | 0.41416 | 0.00615 | 0.41215 | 0.45902 | 0.69425 | 0.3460586459 | 0.91693 | 0.6730055213 | 0.0899831 | 1.796886856 | 0 |
| 2 | 47414279 | C | T | S | L | MSH2 | ENST00000233146 | 0.38605 | 0.00904 | 0.45487 | 0.7969 | 0.88496 | 0.6601494328 | 0.92415 | 0.9477213025 | 0.153882 | 4.188414008 | 0 |
| 2 | 47414281 | T | A | S | T | MSH2 | ENST00000233146 | 0.52107 | 0.01945 | 0.46902 | 0.70259 | 0.87733 | 0.453918552 | 0.92601 | 0.9355816841 | 0.216995 | 4.580442584 | 0 |
| 2 | 47414281 | T | C | S | P | MSH2 | ENST00000233146 | 0.76649 | 0.03039 | 0.48459 | 0.79009 | 0.93631 | 0.7996222082 | 0.96978 | 0.9375133785 | 0.341901 | 6.131639341 | 0 |
| 2 | 47414281 | T | G | S | A | MSH2 | ENST00000233146 | 0.32717 | 0.00664 | 0.44876 | 0.30386 | 0.57487 | 0.3350926197 | 0.84755 | 0.2806321383 | 0.0208778 | 2.119465592 | 0 |
| 2 | 47414282 | C | T | S | L | MSH2 | ENST00000233146 | 0.45095 | 0.01965 | 0.50221 | 0.83613 | 0.9014 | 0.6546135045 | 0.92853 | 0.9856342673 | 0.271367 | 8.643404452 | 1 |
| 2 | 47414284 | C | A | L | M | MSH2 | ENST00000233146 | 0.41629 | 0.03747 | 0.55028 | 0.76018 | 0.85792 | 0.4231079418 | 0.87398 | 0.9767313004 | 0.223036 | 5.665031363 | 0 |
| 2 | 47414284 | C | G | L | V | MSH2 | ENST00000233146 | 0.18683 | 0.0227 | 0.50187 | 0.73823 | 0.90185 | 0.4939134644 | 0.99357 | 0.3928341866 | 0.206862 | 2.138347071 | 0 |
| 2 | 47414285 | T | A | L | Q | MSH2 | ENST00000233146 | 0.90949 | 0.0364 | 0.59443 | 0.90839 | 0.9579 | 0.712891641 | 0.96635 | 0.9943554401 | 0.442415 | 8.986730749 | 1 |
| 2 | 47414285 | T | C | L | P | MSH2 | ENST00000233146 | 0.91831 | 0.02994 | 0.72249 | 0.8965 | 0.9648 | 0.8828213038 | 0.97098 | 0.9969599247 | 0.440051 | 8.986730749 | 1 |
| 2 | 47414285 | T | G | L | R | MSH2 | ENST00000233146 | 0.91657 | 0.03137 | 0.67824 | 0.89632 | 0.96883 | 0.8599717562 | 0.97291 | 0.9960357547 | 0.443069 | 8.986730749 | 1 |
| 2 | 47414287 | T | A | S | T | MSH2 | ENST00000233146 | 0.50081 | 0.01075 | 0.44553 | 0.54605 | 0.8153 | 0.4104780028 | 0.93128 | 0.8821029067 | 0.143664 | 5.807122118 | 0 |
| 2 | 47414287 | T | C | S | P | MSH2 | ENST00000233146 | 0.89219 | 0.03039 | 0.45823 | 0.76247 | 0.9248 | 0.7699968296 | 0.96831 | 0.9841597676 | 0.348861 | 8.196005862 | 0 |
| 2 | 47414287 | T | G | S | A | MSH2 | ENST00000233146 | 0.43418 | 0.00664 | 0.41416 | 0.25901 | 0.52446 | 0.2671051327 | 0.81978 | 0.7546846867 | 0.0640354 | 1.38071152 | 0 |
| 2 | 47414288 | C | A | S | Y | MSH2 | ENST00000233146 | 0.56138 | 0.03175 | 0.52011 | 0.85109 | 0.90494 | 0.620924872 | 0.92669 | 0.9948949218 | 0.261936 | 6.568111491 | 0 |
| 2 | 47414288 | C | G | S | C | MSH2 | ENST00000233146 | 0.32467 | 0.00827 | 0.44116 | 0.67751 | 0.86453 | 0.4087750904 | 0.94455 | 0.864528656 | 0.21496 | 5.364049564 | 0 |
| 2 | 47414288 | C | T | S | F | MSH2 | ENST00000233146 | 0.60142 | 0.03304 | 0.4947 | 0.85751 | 0.91229 | 0.6073946517 | 0.92394 | 0.9961124063 | 0.249955 | 6.796922952 | 0 |
| 2 | 47414290 | G | A | A | T | MSH2 | ENST00000233146 | 0.88138 | 0.03394 | 0.58497 | 0.83063 | 0.97587 | 0.5484377822 | 0.96511 | 0.989893496 | 0.451009 | 5.894849089 | 0 |
| 2 | 47414290 | G | C | A | P | MSH2 | ENST00000233146 | 0.91934 | 0.03243 | 0.66253 | 0.86099 | 0.98367 | 0.8365858818 | 0.97582 | 0.9946347475 | 0.467398 | 9.018301487 | 0 |
| 2 | 47414290 | G | T | A | S | MSH2 | ENST00000233146 | 0.55738 | 0.01267 | 0.53291 | 0.66322 | 0.89784 | 0.4428277051 | 0.8818 | 0.9628798366 | 0.224608 | 3.787864009 | 0 |
| 2 | 47414291 | C | G | A | G | MSH2 | ENST00000233146 | 0.47848 | 0.03379 | 0.56765 | 0.89883 | 0.9729 | 0.4564332806 | 0.89125 | 0.9923507571 | 0.323623 | 4.679070325 | 0 |
| 2 | 47414291 | C | T | A | V | MSH2 | ENST00000233146 | 0.44692 | 0.03475 | 0.62276 | 0.83446 | 0.96737 | 0.6059215779 | 0.98397 | 0.6442478299 | 0.233001 | 4.690795776 | 0 |
| 2 | 47414293 | G | A | V | I | MSH2 | ENST00000233146 | 0.35897 | 0.00654 | 0.28392 | 0.51995 | 0.79818 | 0.1962946388 | 0.88445 | 0.4583008289 | -0.0378436 | 3.358604927 | 0 |
| 2 | 47414293 | G | C | V | L | MSH2 | ENST00000233146 | 0.31467 | 0.00592 | 0.52331 | 0.47001 | 0.69678 | 0.4180455718 | 0.91812 | 0.9724058509 | 0.159549 | 1.631272267 | 0 |
| 2 | 47414293 | G | T | V | L | MSH2 | ENST00000233146 | 0.31049 | 0.00592 | 0.52331 | 0.47756 | 0.69678 | 0.4180455718 | 0.9166 | 0.9785639048 | 0.159554 | 1.631272267 | 0 |
| 2 | 47414294 | T | C | V | A | MSH2 | ENST00000233146 | 0.31296 | 0.0058 | 0.55294 | 0.61262 | 0.8153 | 0.4919285351 | 0.93883 | 0.167756319 | -0.0240389 | 4.008619199 | 0 |
| 2 | 47414294 | T | G | V | G | MSH2 | ENST00000233146 | 0.60757 | 0.02077 | 0.37302 | 0.888 | 0.95826 | 0.6853863958 | 0.96766 | 0.9968407154 | 0.436516 | 8.071382591 | 0 |
| 2 | 47414296 | A | C | I | L | MSH2 | ENST00000233146 | 0.5919 | 0.01082 | 0.46163 | 0.59252 | 0.83779 | 0.4068097986 | 0.87325 | 0.8838217854 | 0.129556 | 3.260283551 | 0 |
| 2 | 47414296 | A | G | I | V | MSH2 | ENST00000233146 | 0.32373 | 0.00681 | 0.32851 | 0.48769 | 0.73949 | 0.1872695297 | 0.85658 | 0.1579543731 | -0.0795278 | 2.369847737 | 0 |
| 2 | 47414296 | A | T | I | F | MSH2 | ENST00000233146 | 0.66486 | 0.02602 | 0.48881 | 0.87159 | 0.95462 | 0.662866334 | 0.90263 | 0.9931829572 | 0.290024 | 4.283534219 | 0 |
| 2 | 47414297 | T | C | I | T | MSH2 | ENST00000233146 | 0.85471 | 0.0248 | 0.63408 | 0.90036 | 0.97587 | 0.563113491 | 0.87408 | 0.9978558421 | 0.328477 | 6.186619223 | 0 |
| 2 | 47414297 | T | G | I | S | MSH2 | ENST00000233146 | 0.9065 | 0.02809 | 0.493 | 0.93159 | 0.98146 | 0.7491129595 | 0.93707 | 0.9984715581 | 0.465423 | 7.413350063 | 0 |
| 2 | 47414298 | C | G | I | M | MSH2 | ENST00000233146 | 0.59392 | 0.0167 | 0.4164 | 0.72 | 0.90494 | 0.4827348134 | 0.85925 | 0.8599056005 | 0.188902 | 4.053616176 | 0 |
| 2 | 47414299 | A | C | K | Q | MSH2 | ENST00000233146 | 0.88074 | 0.03379 | 0.51215 | 0.82016 | 0.90494 | 0.3154408715 | 0.92204 | 0.9884530902 | 0.251083 | 4.824354449 | 0 |
| 2 | 47414300 | A | C | K | T | MSH2 | ENST00000233146 | 0.81653 | 0.03293 | 0.58733 | 0.84645 | 0.92561 | 0.3729561616 | 0.83779 | 0.9947782755 | 0.251645 | 5.480996745 | 0 |
| 2 | 47414300 | A | G | K | R | MSH2 | ENST00000233146 | 0.46868 | 0.00799 | 0.46562 | 0.58114 | 0.79577 | 0.232487162 | 0.94775 | 0.8982006907 | 0.20429 | 3.921316206 | 0 |
| 2 | 47414300 | A | T | K | M | MSH2 | ENST00000233146 | 0.85115 | 0.03224 | 0.47212 | 0.87213 | 0.96008 | 0.4572558383 | 0.91451 | 0.9979646206 | 0.300723 | 6.425392558 | 0 |
| 2 | 47414301 | G | C | K | N | MSH2 | ENST00000233146 | 0.78891 | 0.03313 | 0.61299 | 0.80987 | 0.85326 | 0.3887080492 | 0.87917 | 0.9787705541 | 0.120282 | 4.809645385 | 0 |
| 2 | 47414301 | G | T | K | N | MSH2 | ENST00000233146 | 0.78241 | 0.03313 | 0.61299 | 0.80616 | 0.85326 | 0.3887080492 | 0.87931 | 0.9827110171 | 0.120273 | 4.809645385 | 0 |
| 2 | 47414302 | T | A | F | I | MSH2 | ENST00000233146 | 0.79492 | 0.0142 | 0.73488 | 0.8728 | 0.94053 | 0.6282040205 | 0.87232 | 0.9835879803 | 0.238544 | 6.374869014 | 0 |
| 2 | 47414302 | T | C | F | L | MSH2 | ENST00000233146 | 0.66502 | 0.00945 | 0.78242 | 0.75316 | 0.94015 | 0.5202562002 | 0.91805 | 0.9881100655 | 0.244142 | 5.739830149 | 0 |
| 2 | 47414302 | T | G | F | V | MSH2 | ENST00000233146 | 0.66193 | 0.01075 | 0.71318 | 0.85017 | 0.95426 | 0.581501788 | 0.90211 | 0.9873319268 | 0.273215 | 5.396288438 | 0 |
| 2 | 47414303 | T | A | F | Y | MSH2 | ENST00000233146 | 0.40576 | 0.00547 | 0.76291 | 0.18233 | 0.59146 | 0.4423452369 | 0.68354 | 0.5893257856 | -0.0178077 | -0.9342113081 | 0 |
| 2 | 47414303 | T | C | F | S | MSH2 | ENST00000233146 | 0.89583 | 0.01475 | 0.73172 | 0.89846 | 0.93201 | 0.6126347093 | 0.94743 | 0.996592164 | 0.288568 | 6.374869014 | 0 |
| 2 | 47414303 | T | G | F | C | MSH2 | ENST00000233146 | 0.921 | 0.02702 | 0.76965 | 0.89716 | 0.92561 | 0.5960883707 | 0.91146 | 0.9967882633 | 0.263572 | 6.374869014 | 1 |
| 2 | 47414304 | T | A | F | L | MSH2 | ENST00000233146 | 0.46612 | 0.00945 | 0.78242 | 0.63828 | 0.85274 | 0.5202562002 | 0.95481 | 0.9878323078 | 0.210112 | 5.739830149 | 0 |
| 2 | 47414304 | T | G | F | L | MSH2 | ENST00000233146 | 0.4725 | 0.00945 | 0.78242 | 0.63075 | 0.85274 | 0.5202562002 | 0.95453 | 0.9879540205 | 0.21013 | 5.739830149 | 0 |
| 2 | 47414305 | T | A | L | I | MSH2 | ENST00000233146 | 0.6039 | 0.03475 | 0.70916 | 0.84237 | 0.90274 | 0.3679513683 | 0.86372 | 0.9897630215 | 0.239839 | 4.677810881 | 0 |
| 2 | 47414305 | T | G | L | V | MSH2 | ENST00000233146 | 0.59944 | 0.0357 | 0.65835 | 0.81097 | 0.92397 | 0.5586214208 | 0.92926 | 0.989358902 | 0.356154 | 5.116608653 | 0 |
| 2 | 47414306 | T | C | L | S | MSH2 | ENST00000233146 | 0.83951 | 0.03212 | 0.72911 | 0.89796 | 0.98693 | 0.786890157 | 0.96789 | 0.9992182255 | 0.480905 | 8.075990636 | 0 |
| 2 | 47414307 | A | C | L | F | MSH2 | ENST00000233146 | 0.71522 | 0.03563 | 0.70498 | 0.89524 | 0.93901 | 0.5998723205 | 0.96298 | 0.9912781119 | 0.426921 | 6.497341168 | 0 |
| 2 | 47414307 | A | T | L | F | MSH2 | ENST00000233146 | 0.70848 | 0.03563 | 0.70498 | 0.89631 | 0.93901 | 0.5998723205 | 0.96308 | 0.991779387 | 0.426913 | 6.497341168 | 0 |
| 2 | 47414308 | G | A | E | K | MSH2 | ENST00000233146 | 0.88994 | 0.02608 | 0.74986 | 0.85759 | 0.94205 | 0.5060228032 | 0.91896 | 0.9977372885 | 0.363786 | 4.63635215 | 0 |

| | | | | | | | | | | | | | | | | |
|---|---|---|---|---|---|---|---|---|---|---|---|---|---|---|---|---|
| 2 | 47414308 | G | C | E | Q | MSH2 | ENST00000233146 | 0.83437 | 0.03322 | 0.6281 | 0.84973 | 0.90274 | 0.3881636527 | 0.90491 | 0.979752183 | 0.265615 | 4.695878737 | 0 |
| 2 | 47414309 | A | C | E | A | MSH2 | ENST00000233146 | 0.72596 | 0.02667 | 0.63532 | 0.8662 | 0.94986 | 0.4893532965 | 0.85741 | 0.9925780892 | 0.264234 | 6.101273126 | 0 |
| 2 | 47414309 | A | G | E | G | MSH2 | ENST00000233146 | 0.63597 | 0.02561 | 0.58899 | 0.81656 | 0.95572 | 0.4984973047 | 0.917 | 0.9907727242 | 0.335653 | 6.700551705 | 0 |
| 2 | 47414309 | A | T | E | V | MSH2 | ENST00000233146 | 0.75893 | 0.03106 | 0.61616 | 0.89376 | 0.97848 | 0.5048295018 | 0.90961 | 0.9973644614 | 0.384865 | 5.768280688 | 0 |
| 2 | 47414310 | A | C | E | D | MSH2 | ENST00000233146 | 0.34506 | 0.00664 | 0.63504 | 0.60551 | 0.8199 | 0.3416896062 | 0.96529 | 0.2111158669 | 0.195002 | 5.348811457 | 0 |
| 2 | 47414310 | A | T | E | D | MSH2 | ENST00000233146 | 0.34049 | 0.00664 | 0.63504 | 0.60398 | 0.8199 | 0.3416896062 | 0.96542 | 0.2111158669 | 0.194991 | 5.348811457 | 0 |
| 2 | 47414311 | C | A | L | I | MSH2 | ENST00000233146 | 0.69692 | 0.01053 | 0.67233 | 0.79342 | 0.87684 | 0.2430762337 | 0.76418 | 0.9560431838 | 0.218336 | 6.156391825 | 0 |
| 2 | 47414311 | C | G | L | V | MSH2 | ENST00000233146 | 0.68747 | 0.01848 | 0.60536 | 0.88432 | 0.85996 | 0.4342153828 | 0.88204 | 0.5621477365 | 0.0991683 | 5.351448469 | 0 |
| 2 | 47414311 | C | T | L | F | MSH2 | ENST00000233146 | 0.73581 | 0.03261 | 0.6562 | 0.88783 | 0.92763 | 0.4560363305 | 0.88216 | 0.9967156649 | 0.367492 | 4.987132889 | 0 |
| 2 | 47414312 | T | A | L | H | MSH2 | ENST00000233146 | 0.73789 | 0.03304 | 0.75007 | 0.93586 | 0.98477 | 0.6420798663 | 0.95438 | 0.9988650084 | 0.477648 | 4.875016195 | 0 |
| 2 | 47414312 | T | C | L | P | MSH2 | ENST00000233146 | 0.76267 | 0.02927 | 0.78209 | 0.93708 | 0.98764 | 0.8417241853 | 0.96371 | 0.9979923964 | 0.45295 | 8.526491066 | 0 |
| 2 | 47414312 | T | G | L | R | MSH2 | ENST00000233146 | 0.75805 | 0.03033 | 0.7494 | 0.91239 | 0.97438 | 0.7808941659 | 0.94727 | 0.999245286 | 0.468581 | 4.19259373 | 0 |
| 2 | 47414314 | T | A | L | I | MSH2 | ENST00000233146 | 0.5232 | 0.03527 | 0.65345 | 0.81921 | 0.89558 | 0.2192661151 | 0.82348 | 0.9739531875 | 0.146868 | 6.605890015 | 0 |
| 2 | 47414314 | T | G | L | V | MSH2 | ENST00000233146 | 0.61372 | 0.03587 | 0.57582 | 0.82179 | 0.90668 | 0.3877887913 | 0.85332 | 0.941241622 | 0.205661 | 5.612082902 | 0 |
| 2 | 47414315 | T | C | L | S | MSH2 | ENST00000233146 | 0.79698 | 0.03212 | 0.6646 | 0.88312 | 0.96773 | 0.6382628248 | 0.89387 | 0.9953824878 | 0.339595 | 4.946012076 | 0 |
| 2 | 47414317 | T | A | S | T | MSH2 | ENST00000233146 | 0.55415 | 0.02714 | 0.62425 | 0.63575 | 0.90885 | 0.3319017058 | 0.908 | 0.9270623326 | 0.20428 | 3.986339375 | 0 |
| 2 | 47414317 | T | C | S | P | MSH2 | ENST00000233146 | 0.80748 | 0.03099 | 0.63492 | 0.82003 | 0.95645 | 0.6402302787 | 0.97807 | 0.9466228485 | 0.288598 | 6.381123639 | 0 |
| 2 | 47414317 | T | G | S | A | MSH2 | ENST00000233146 | 0.43093 | 0.00681 | 0.59428 | 0.56727 | 0.7337 | 0.216682972 | 0.92892 | 0.8219037056 | 0.126508 | 2.863014596 | 0 |
| 2 | 47414318 | C | T | S | L | MSH2 | ENST00000233146 | 0.52586 | 0.00916 | 0.64189 | 0.80096 | 0.93358 | 0.4215342431 | 0.91049 | 0.9764875174 | 0.212363 | 6.248026201 | 0 |
| 2 | 47414320 | G | A | D | N | MSH2 | ENST00000233146 | 0.67279 | 0.01244 | 0.70838 | 0.73539 | 0.8367 | 0.4877577432 | 0.85872 | 0.9829850793 | 0.0817801 | 4.070773553 | 0 |
| 2 | 47414320 | G | C | D | H | MSH2 | ENST00000233146 | 0.85167 | 0.03426 | 0.73301 | 0.89236 | 0.95535 | 0.6119050648 | 0.93747 | 0.9960468239 | 0.47389 | 6.292325024 | 0 |
| 2 | 47414320 | G | T | D | Y | MSH2 | ENST00000233146 | 0.85945 | 0.03388 | 0.7179 | 0.89891 | 0.96153 | 0.6792764908 | 0.9766 | 0.9990122318 | 0.485362 | 6.815234481 | 0 |
| 2 | 47414321 | A | C | D | A | MSH2 | ENST00000233146 | 0.84066 | 0.03287 | 0.63204 | 0.8504 | 0.98693 | 0.6276545287 | 0.94128 | 0.9982670546 | 0.475003 | 6.521567383 | 0 |
| 2 | 47414321 | A | G | D | G | MSH2 | ENST00000233146 | 0.84484 | 0.03124 | 0.64828 | 0.86849 | 0.99443 | 0.5911251246 | 0.9763 | 0.9876127839 | 0.401362 | 6.329662142 | 0 |
| 2 | 47414321 | A | T | D | V | MSH2 | ENST00000233146 | 0.83306 | 0.03137 | 0.62773 | 0.89358 | 0.9833 | 0.6613528009 | 0.97564 | 0.9995065928 | 0.474975 | 7.811604654 | 0 |
| 2 | 47414322 | T | A | D | E | MSH2 | ENST00000233146 | 0.4988 | 0.03563 | 0.67554 | 0.82466 | 0.91526 | 0.3289011902 | 0.97521 | 0.9727830887 | 0.322972 | 7.811604654 | 0 |
| 2 | 47414322 | T | G | D | E | MSH2 | ENST00000233146 | 0.5055 | 0.03563 | 0.67554 | 0.81956 | 0.91526 | 0.3289011902 | 0.97502 | 0.9727830887 | 0.322974 | 7.811604654 | 0 |
| 2 | 47414323 | G | A | D | N | MSH2 | ENST00000233146 | 0.65422 | 0.01848 | 0.44351 | 0.73289 | 0.7609 | 0.2639500786 | 0.85179 | 0.9608843923 | 0.114065 | 2.191330494 | 0 |
| 2 | 47414323 | G | C | D | H | MSH2 | ENST00000233146 | 0.84846 | 0.02951 | 0.47095 | 0.8828 | 0.9248 | 0.4164321161 | 0.85686 | 0.9737901092 | 0.26033 | 3.583885014 | 0 |
| 2 | 47414323 | G | T | D | Y | MSH2 | ENST00000233146 | 0.86226 | 0.02943 | 0.464 | 0.90916 | 0.96444 | 0.4720402143 | 0.97589 | 0.9719162583 | 0.423999 | 5.908513159 | 0 |
| 2 | 47414324 | A | C | D | A | MSH2 | ENST00000233146 | 0.45783 | 0.00674 | 0.38199 | 0.65129 | 0.86951 | 0.3775941496 | 0.89121 | 0.9399976134 | 0.227245 | 2.580010931 | 0 |
| 2 | 47414324 | A | G | D | G | MSH2 | ENST00000233146 | 0.60112 | 0.01485 | 0.3777 | 0.76462 | 0.90406 | 0.4032870991 | 0.88985 | 0.9845585227 | 0.260902 | 2.644251674 | 0 |
| 2 | 47414324 | A | T | D | V | MSH2 | ENST00000233146 | 0.46624 | 0.00615 | 0.3598 | 0.8065 | 0.92561 | 0.4447259871 | 0.96296 | 0.8678542376 | 0.173319 | 2.175225964 | 0 |
| 2 | 47414325 | T | A | D | E | MSH2 | ENST00000233146 | 0.18089 | 0.00596 | 0.40003 | 0.36485 | 0.76356 | 0.1328005439 | 0.92289 | 0.2357464135 | 0.0519346 | -1.682751921 | 0 |
| 2 | 47414325 | T | G | D | E | MSH2 | ENST00000233146 | 0.18461 | 0.00596 | 0.40003 | 0.36344 | 0.76356 | 0.1328005439 | 0.92466 | 0.2357464135 | 0.0518956 | -1.682751921 | 0 |
| 2 | 47414326 | T | A | S | T | MSH2 | ENST00000233146 | 0.41991 | 0.00638 | 0.50995 | 0.6812 | 0.79577 | 0.3081363844 | 0.9547 | 0.7633628249 | 0.181505 | 2.197545613 | 0 |
| 2 | 47414326 | T | C | S | P | MSH2 | ENST00000233146 | 0.83205 | 0.02494 | 0.53524 | 0.83441 | 0.95935 | 0.5919684903 | 0.97315 | 0.9877820611 | 0.384883 | 5.314391713 | 0 |
| 2 | 47414326 | T | G | S | A | MSH2 | ENST00000233146 | 0.4574 | 0.00633 | 0.49711 | 0.49374 | 0.74164 | 0.2058554136 | 0.95712 | 0.7973535061 | 0.127391 | 3.37293607 | 0 |
| 2 | 47414327 | C | A | S | Y | MSH2 | ENST00000233146 | 0.5924 | 0.02529 | 0.60681 | 0.85932 | 0.97885 | 0.4635828436 | 0.93964 | 0.8663218021 | 0.203883 | 7.230319933 | 0 |
| 2 | 47414327 | C | G | S | C | MSH2 | ENST00000233146 | 0.62271 | 0.03362 | 0.52275 | 0.83347 | 0.93977 | 0.3731067479 | 0.84306 | 0.9949914813 | 0.284806 | 6.25487319 | 0 |
| 2 | 47414327 | C | T | S | F | MSH2 | ENST00000233146 | 0.63283 | 0.02896 | 0.57492 | 0.86249 | 0.9781 | 0.4612719952 | 0.95595 | 0.9961887002 | 0.42255 | 5.622398736 | 0 |
| 2 | 47414329 | A | C | N | H | MSH2 | ENST00000233146 | 0.64984 | 0.03265 | 0.53016 | 0.68587 | 0.90494 | 0.5479087128 | 0.92838 | 0.9925985336 | 0.163785 | 7.358081249 | 0 |
| 2 | 47414329 | A | G | N | D | MSH2 | ENST00000233146 | 0.85742 | 0.03112 | 0.58681 | 0.81665 | 0.93162 | 0.5326322839 | 0.93592 | 0.9955766201 | 0.175598 | 7.478312008 | 0 |
| 2 | 47414329 | A | T | N | Y | MSH2 | ENST00000233146 | 0.68803 | 0.00827 | 0.57524 | 0.80638 | 0.9458 | 0.6025253654 | 0.95811 | 0.993078053 | 0.399646 | 4.229864337 | 0 |
| 2 | 47414330 | A | C | N | T | MSH2 | ENST00000233146 | 0.72764 | 0.02951 | 0.49933 | 0.81829 | 0.94543 | 0.492082991 | 0.9012 | 0.9968812466 | 0.205604 | 5.401007242 | 0 |
| 2 | 47414330 | A | G | N | S | MSH2 | ENST00000233146 | 0.64058 | 0.01881 | 0.4352 | 0.75053 | 0.9045 | 0.414138509 | 0.89938 | 0.9922080636 | 0.098889 | 5.848211257 | 0 |
| 2 | 47414330 | A | T | N | I | MSH2 | ENST00000233146 | 0.77489 | 0.02745 | 0.51954 | 0.89003 | 0.97401 | 0.606114992 | 0.90326 | 0.999458611 | 0.347546 | 4.108448897 | 0 |
| 2 | 47414331 | C | A | N | K | MSH2 | ENST00000233146 | 0.44875 | 0.02972 | 0.62311 | 0.86071 | 0.91569 | 0.569632079 | 0.92906 | 0.9962264299 | 0.122111 | 7.478312008 | 0 |
| 2 | 47414331 | C | G | N | K | MSH2 | ENST00000233146 | 0.45524 | 0.02972 | 0.62311 | 0.84924 | 0.91569 | 0.569632079 | 0.92897 | 0.9955939651 | 0.122114 | 7.478312008 | 0 |
| 2 | 47414332 | T | A | F | I | MSH2 | ENST00000233146 | 0.804 | 0.03189 | 0.67092 | 0.81518 | 0.967 | 0.5377334331 | 0.86466 | 0.9956658483 | 0.311903 | 6.677207041 | 0 |
| 2 | 47414332 | T | C | F | L | MSH2 | ENST00000233146 | 0.50153 | 0.03426 | 0.69863 | 0.73209 | 0.93939 | 0.4555869128 | 0.90988 | 0.9922049642 | 0.375101 | 6.302599778 | 0 |
| 2 | 47414332 | T | G | F | V | MSH2 | ENST00000233146 | 0.80637 | 0.03265 | 0.63953 | 0.85169 | 0.9844 | 0.534377556 | 0.93183 | 0.9966897964 | 0.439064 | 6.777780411 | 0 |
| 2 | 47414333 | T | C | F | S | MSH2 | ENST00000233146 | 0.87516 | 0.02802 | 0.65454 | 0.85827 | 0.98367 | 0.6092221506 | 0.96262 | 0.9974967837 | 0.453181 | 4.63630967 | 0 |
| 2 | 47414334 | T | A | F | L | MSH2 | ENST00000233146 | 0.30987 | 0.03426 | 0.69863 | 0.68717 | 0.82892 | 0.4555869128 | 0.94464 | 0.9850494266 | 0.332749 | 6.302599778 | 0 |
| 2 | 47414334 | T | G | F | L | MSH2 | ENST00000233146 | 0.31519 | 0.03426 | 0.69863 | 0.68587 | 0.82892 | 0.4555869128 | 0.9443 | 0.9850494266 | 0.332751 | 6.302599778 | 0 |
| 2 | 47414335 | G | A | G | R | MSH2 | ENST00000233146 | 0.78455 | 0.02984 | 0.61643 | 0.89567 | 0.94543 | 0.4442620685 | 0.93594 | 0.9929407835 | 0.402718 | 3.911802603 | 0 |
| 2 | 47414335 | G | C | G | R | MSH2 | ENST00000233146 | 0.78138 | 0.02984 | 0.61643 | 0.89459 | 0.94543 | 0.4442620685 | 0.93368 | 0.9929407835 | 0.402736 | 3.911802603 | 0 |
| 2 | 47414336 | G | A | G | E | MSH2 | ENST00000233146 | 0.64415 | 0.02691 | 0.59574 | 0.8919 | 0.9458 | 0.4203033605 | 0.85502 | 0.9829197624 | 0.309398 | 7.141542963 | 0 |
| 2 | 47414336 | G | C | G | A | MSH2 | ENST00000233146 | 0.60683 | 0.02342 | 0.49212 | 0.84851 | 0.95353 | 0.3427315778 | 0.93611 | 0.857747972 | 0.152175 | 4.719837039 | 0 |
| 2 | 47414336 | G | T | G | V | MSH2 | ENST00000233146 | 0.64306 | 0.03189 | 0.50784 | 0.89034 | 0.95426 | 0.4815440045 | 0.9313 | 0.9964119792 | 0.430316 | 6.307387738 | 0 |
| 2 | 47414338 | C | A | Q | K | MSH2 | ENST00000233146 | 0.43792 | 0.00654 | 0.48044 | 0.62119 | 0.84375 | 0.2614884098 | 0.95121 | 0.9237300754 | 0.261412 | 7.60518345 | 0 |
| 2 | 47414338 | C | G | Q | E | MSH2 | ENST00000233146 | 0.40869 | 0.00681 | 0.41578 | 0.72111 | 0.84321 | 0.2387722622 | 0.9094 | 0.908901155 | 0.219798 | 6.578623692 | 0 |
| 2 | 47414339 | A | C | Q | P | MSH2 | ENST00000233146 | 0.80329 | 0.02775 | 0.42149 | 0.91582 | 0.96993 | 0.5305677993 | 0.97544 | 0.9932034016 | 0.467989 | 5.276980803 | 1 |
| 2 | 47414339 | A | G | Q | R | MSH2 | ENST00000233146 | 0.47577 | 0.00747 | 0.47023 | 0.68701 | 0.85945 | 0.2697290794 | 0.96974 | 0.6117508411 | 0.220305 | 7.60518345 | 0 |
| 2 | 47414339 | A | T | Q | L | MSH2 | ENST00000233146 | 0.8203 | 0.0246 | 0.40387 | 0.85837 | 0.91526 | 0.3270092015 | 0.88827 | 0.9936336279 | 0.259051 | 3.374771993 | 0 |
| 2 | 47414340 | G | C | Q | H | MSH2 | ENST00000233146 | 0.51021 | 0.03534 | 0.46734 | 0.79733 | 0.87345 | 0.3679650292 | 0.94597 | 0.9604534507 | 0.26616 | 7.60518345 | 0 |
| 2 | 47414340 | G | T | Q | H | MSH2 | ENST00000233146 | 0.50492 | 0.03534 | 0.46734 | 0.79733 | 0.87345 | 0.3679650292 | 0.946 | 0.9604534507 | 0.266162 | 7.60518345 | 0 |
| 2 | 47414341 | T | A | F | I | MSH2 | ENST00000233146 | 0.83081 | 0.03137 | 0.6682 | 0.85739 | 0.95389 | 0.6528003137 | 0.85083 | 0.9971387386 | 0.263892 | 5.369526905 | 1 |
| 2 | 47414341 | T | C | F | L | MSH2 | ENST00000233146 | 0.84446 | 0.03281 | 0.70891 | 0.76425 | 0.9328 | 0.5994999792 | 0.83861 | 0.9951262474 | 0.244665 | 5.29360633 | 0 |
| 2 | 47414341 | T | G | F | V | MSH2 | ENST00000233146 | 0.83714 | 0.03219 | 0.65127 | 0.86807 | 0.98869 | 0.6367396382 | 0.91789 | 0.9971387386 | 0.395845 | 5.187065923 | 1 |
| 2 | 47414342 | T | C | F | S | MSH2 | ENST00000233146 | 0.8856 | 0.02802 | 0.66486 | 0.9063 | 0.96517 | 0.7170970808 | 0.95107 | 0.9706256933 | 0.39708 | 4.009600423 | 1 |
| 2 | 47414342 | T | G | F | C | MSH2 | ENST00000233146 | 0.88922 | 0.0315 | 0.70916 | 0.89142 | 0.91186 | 0.6948926924 | 0.92375 | 0.9863985181 | 0.380548 | 7.471533516 | 0 |
| 2 | 47414343 | T | A | F | L | MSH2 | ENST00000233146 | 0.66731 | 0.03281 | 0.70891 | 0.76122 | 0.90624 | 0.5994999792 | 0.85753 | 0.9938979745 | 0.212279 | 5.29360633 | 0 |
| 2 | 47414343 | T | G | F | L | MSH2 | ENST00000233146 | 0.67549 | 0.03281 | 0.70891 | 0.74771 | 0.90624 | 0.5994999792 | 0.85703 | 0.9938979745 | 0.212282 | 5.29360633 | 0 |
| 2 | 47414344 | G | A | E | K | MSH2 | ENST00000233146 | 0.4946 | 0.00556 | 0.64139 | 0.56396 | 0.76421 | 0.2117747275 | 0.93765 | 0.9428575039 | 0.244364 | 2.130668315 | 0 |
| 2 | 47414344 | G | C | E | Q | MSH2 | ENST00000233146 | 0.48812 | 0.00638 | 0.50024 | 0.57034 | 0.77463 | 0.1834803194 | 0.92793 | 0.4849597216 | 0.228852 | 2.469920901 | 0 |
| 2 | 47414345 | A | C | E | A | MSH2 | ENST00000233146 | 0.44205 | 0.00812 | 0.53293 | 0.64976 | 0.80358 | 0.2174729887 | 0.92536 | 0.8447644114 | 0.245691 | 3.595654704 | 0 |
| 2 | 47414345 | A | G | E | G | MSH2 | ENST00000233146 | 0.53207 | 0.00788 | 0.46887 | 0.75167 | 0.87491 | 0.286852025 | 0.89546 | 0.9593682885 | 0.258008 | 4.290003899 | 0 |
| 2 | 47414345 | A | T | E | V | MSH2 | ENST00000233146 | 0.50426 | 0.00562 | 0.50224 | 0.59839 | 0.8233 | 0.302277966 | 0.96174 | 0.9009338617 | 0.217194 | 4.43405329 | 0 |
| 2 | 47414346 | A | C | E | D | MSH2 | ENST00000233146 | 0.2679 | 0.00599 | 0.52278 | 0.57324 | 0.78223 | 0.1350957898 | 0.97459 | 0.3851017058 | 0.194292 | 3.886002273 | 0 |
| 2 | 47414346 | A | T | E | D | MSH2 | ENST00000233146 | 0.264 | 0.00599 | 0.52278 | 0.57405 | 0.78223 | 0.1350957898 | 0.97468 | 0.4859828949 | 0.194288 | 3.886002273 | 0 |
| 2 | 47414347 | C | A | L | M | MSH2 | ENST00000233146 | 0.37329 | 0.0132 | 0.62405 | 0.61074 | 0.86403 | 0.3436699774 | 0.87982 | 0.8526220322 | 0.192012 | 3.718054014 | 0 |
| 2 | 47414347 | C | G | L | V | MSH2 | ENST00000233146 | 0.56263 | 0.01562 | 0.57806 | 0.82045 | 0.89784 | 0.3418179247 | 0.94529 | 0.9595850706 | 0.27238 | 4.201790281 | 0 |
| 2 | 47414348 | T | A | L | Q | MSH2 | ENST00000233146 | 0.72108 | 0.03287 | 0.66234 | 0.92549 | 0.99498 | 0.7427426855 | 0.96895 | 0.9978141785 | 0.527424 | 8.689014263 | 0 |
| 2 | 47414348 | T | C | L | P | MSH2 | ENST00000233146 | 0.73829 | 0.02851 | 0.78689 | 0.93998 | 0.99471 | 0.8073514629 | 0.96391 | 0.9984715581 | 0.525851 | 5.642715512 | 0 |
| 2 | 47414348 | T | G | L | R | MSH2 | ENST00000233146 | 0.73053 | 0.02835 | 0.7415 | 0.92601 | 0.99824 | 0.7983242056 | 0.98071 | 0.9982543588 | 0.531782 | 8.689014263 | 0 |
| 2 | 47414350 | A | C | T | P | MSH2 | ENST00000233146 | 0.36934 | 0.00681 | 0.54564 | 0.72173 | 0.86753 | 0.5849373444 | 0.96024 | 0.7512625456 | 0.285732 | 7.21556625 | 0 |
| 2 | 47414350 | A | G | T | A | MSH2 | ENST00000233146 | 0.29377 | 0.00574 | 0.54955 | 0.57848 | 0.75822 | 0.2532393673 | 0.96186 | 0.3277072608 | 0.159414 | 3.454062028 | 0 |
| 2 | 47414350 | A | T | T | S | MSH2 | ENST00000233146 | 0.25729 | 0.00586 | 0.55843 | 0.56669 | 0.74867 | 0.1803004085 | 0.93636 | 0.07957062889 | 0.00523113 | 1.337417259 | 0 |
| 2 | 47414351 | C | A | T | N | MSH2 | ENST00000233146 | 0.18588 | 0.00547 | 0.64005 | 0.61803 | 0.74728 | 0.3117285725 | 0.91293 | 0.6380344629 | 0.140725 | 3.427903371 | 0 |
| 2 | 47414351 | C | G | T | S | MSH2 | ENST00000233146 | 0.15875 | 0.00586 | 0.55843 | 0.57399 | 0.74164 | 0.1803004085 | 0.94422 | 0.6943093538 | 0.126756 | 1.337417259 | 0 |
| 2 | 47414351 | C | T | T | I | MSH2 | ENST00000233146 | 0.23259 | 0.00566 | 0.69912 | 0.6972 | 0.80417 | 0.3150973118 | 0.97033 | 0.8169159293 | 0.231663 | 3.261743317 | 0 |
| 2 | 47414353 | A | C | T | P | MSH2 | ENST00000233146 | 0.46394 | 0.01556 | 0.4197 | 0.6026 | 0.86453 | 0.5564209473 | 0.97891 | 0.9511363506 | 0.366527 | 4.778375083 | 0 |
| 2 | 47414353 | A | G | T | A | MSH2 | ENST00000233146 | 0.41712 | 0.00586 | 0.42534 | 0.75155 | 0.77654 | 0.2704261416 | 0.96424 | 0.8459267616 | 0.239991 | 5.090188009 | 0 |
| 2 | 47414353 | A | T | T | S | MSH2 | ENST00000233146 | 0.37504 | 0.00603 | 0.4233 | 0.46651 | 0.69168 | 0.1692292817 | 0.95061 | 0.744799614 | 0.180172 | 2.756683079 | 0 |
| 2 | 47414354 | C | A | T | N | MSH2 | ENST00000233146 | 0.20881 | 0.00556 | 0.51499 | 0.48759 | 0.73589 | 0.3111662736 | 0.94173 | 0.7040702105 | 0.165103 | 6.087791965 | 0 |
| 2 | 47414354 | C | G | T | S | MSH2 | ENST00000233146 | 0.19525 | 0.00603 | 0.4233 | 0.43885 | 0.69846 | 0.1692292817 | 0.94564 | 0.7556863427 | 0.179108 | 2.756683079 | 0 |
| 2 | 47414354 | C | T | T | I | MSH2 | ENST00000233146 | 0.2516 | 0.00648 | 0.57375 | 0.7734 | 0.88776 | 0.3449919141 | 0.93718 | 0.9894324541 | 0.291519 | 4.526841542 | 0 |
| 2 | 47414356 | T | A | F | I | MSH2 | ENST00000233146 | 0.45647 | 0.0227 | 0.52764 | 0.78789 | 0.91526 | 0.414285983 | 0.87808 | 0.9812978506 | 0.257143 | 4.609065535 | 0 |
| 2 | 47414356 | T | C | F | L | MSH2 | ENST00000233146 | 0.42998 | 0.00696 | 0.583 | 0.58796 | 0.84159 | 0.3877181184 | 0.86261 | 0.8782243729 | 0.171594 | 3.539852325 | 0 |
| 2 | 47414356 | T | G | F | V | MSH2 | ENST00000233146 | 0.47533 | 0.0233 | 0.4974 | 0.81887 | 0.94356 | 0.4163088791 | 0.93631 | 0.9846402407 | 0.374174 | 4.892659242 | 0 |

| | | | | | | | | | | | | | | | | | |
|---|---|---|---|---|---|---|---|---|---|---|---|---|---|---|---|---|---|
| 2 | 47414357 | T | A | F | Y | MSH2 | ENST00000233146 | 0.61925 | 0.01249 | 0.55616 | 0.62456 | 0.89918 | 0.3447040306 | 0.88065 | 0.9703350067 | 0.210373 | 3.244391959 | 0 |
| 2 | 47414357 | T | C | F | S | MSH2 | ENST00000233146 | 0.88849 | 0.02595 | 0.52863 | 0.89837 | 0.95717 | 0.6041333126 | 0.93473 | 0.9939029813 | 0.422975 | 8.024359049 | 0 |
| 2 | 47414357 | T | G | F | C | MSH2 | ENST00000233146 | 0.89359 | 0.03019 | 0.57585 | 0.9037 | 0.95389 | 0.5299184124 | 0.9467 | 0.9962707758 | 0.461182 | 8.024359049 | 0 |
| 2 | 47414358 | T | A | F | L | MSH2 | ENST00000233146 | 0.41511 | 0.00696 | 0.583 | 0.61542 | 0.81818 | 0.3877181184 | 0.84894 | 0.8277903199 | 0.15266 | 3.539852325 | 0 |
| 2 | 47414358 | T | G | F | L | MSH2 | ENST00000233146 | 0.42121 | 0.00696 | 0.583 | 0.61417 | 0.81818 | 0.3877181184 | 0.85009 | 0.8277903199 | 0.152652 | 3.539852325 | 0 |
| 2 | 47414359 | G | A | D | N | MSH2 | ENST00000233146 | 0.54677 | 0.00722 | 0.63908 | 0.76209 | 0.82892 | 0.3092163227 | 0.90044 | 0.9894456267 | 0.12501 | 4.072631685 | 0 |
| 2 | 47414359 | G | C | D | H | MSH2 | ENST00000233146 | 0.87669 | 0.0325 | 0.66331 | 0.91914 | 0.96737 | 0.4305092669 | 0.98424 | 0.9978423119 | 0.516537 | 4.983072454 | 0 |
| 2 | 47414359 | G | T | D | Y | MSH2 | ENST00000233146 | 0.88335 | 0.03189 | 0.64407 | 0.92369 | 0.97438 | 0.5462993884 | 0.98818 | 0.9973884225 | 0.531571 | 5.585019779 | 0 |
| 2 | 47414360 | A | C | D | A | MSH2 | ENST00000233146 | 0.92663 | 0.02225 | 0.57321 | 0.90673 | 0.98257 | 0.4729553546 | 0.9802 | 0.997250855 | 0.515615 | 5.633273067 | 0 |
| 2 | 47414360 | A | G | D | G | MSH2 | ENST00000233146 | 0.78177 | 0.00705 | 0.58349 | 0.90203 | 0.9781 | 0.5025536042 | 0.98907 | 0.9945886135 | 0.519139 | 6.377066535 | 0 |
| 2 | 47414360 | A | T | D | V | MSH2 | ENST00000233146 | 0.92464 | 0.02809 | 0.57398 | 0.92704 | 0.98549 | 0.5461094721 | 0.98683 | 0.9996333122 | 0.529172 | 5.731027069 | 0 |
| 2 | 47414361 | C | A | D | E | MSH2 | ENST00000233146 | 0.34871 | 0.00945 | 0.60983 | 0.59525 | 0.87877 | 0.2349547937 | 0.97266 | 0.9699361324 | 0.371823 | 3.334459153 | 0 |
| 2 | 47414361 | C | G | D | E | MSH2 | ENST00000233146 | 0.35454 | 0.00945 | 0.60983 | 0.59542 | 0.87877 | 0.2349547937 | 0.97268 | 0.2839218974 | 0.177288 | 3.334459153 | 0 |
| 2 | 47414362 | T | A | F | I | MSH2 | ENST00000233146 | 0.43738 | 0.00569 | 0.39336 | 0.67798 | 0.75005 | 0.2843953826 | 0.92372 | 0.9317738414 | 0.16556 | 1.542142893 | 0 |
| 2 | 47414362 | T | C | F | L | MSH2 | ENST00000233146 | 0.34201 | 0.00543 | 0.41746 | 0.43931 | 0.70589 | 0.2191100342 | 0.78189 | 0.7285212874 | 0.0938587 | -2.716723751 | 0 |
| 2 | 47414362 | T | G | F | V | MSH2 | ENST00000233146 | 0.44101 | 0.00622 | 0.34837 | 0.76268 | 0.76223 | 0.3217698049 | 0.96791 | 0.8384898305 | 0.261788 | 1.379332679 | 0 |
| 2 | 47414363 | T | A | F | Y | MSH2 | ENST00000233146 | 0.38782 | 0.01102 | 0.42047 | 0.71838 | 0.83228 | 0.3217485699 | 0.88294 | 0.8891139627 | 0.215374 | 3.634654564 | 0 |
| 2 | 47414363 | T | C | F | S | MSH2 | ENST00000233146 | 0.56561 | 0.01164 | 0.38169 | 0.83538 | 0.87491 | 0.518964725 | 0.94161 | 0.9745839834 | 0.292045 | 1.180811608 | 0 |
| 2 | 47414363 | T | G | F | C | MSH2 | ENST00000233146 | 0.73109 | 0.01924 | 0.42779 | 0.89671 | 0.93436 | 0.4511871086 | 0.92959 | 0.9516704082 | 0.323513 | 3.789017237 | 0 |
| 2 | 47414364 | C | A | F | L | MSH2 | ENST00000233146 | 0.17338 | 0.00543 | 0.41746 | 0.53037 | 0.72016 | 0.2191100342 | 0.72086 | 0.1013970971 | 0.051283 | -2.716723751 | 0 |
| 2 | 47414364 | C | G | F | L | MSH2 | ENST00000233146 | 0.17714 | 0.00543 | 0.41746 | 0.53176 | 0.72016 | 0.2191100342 | 0.72025 | 0.1292556524 | 0.0512656 | -2.716723751 | 0 |
| 2 | 47414365 | A | C | S | R | MSH2 | ENST00000233146 | 0.51086 | 0.00556 | 0.25226 | 0.77818 | 0.88069 | 0.3178352275 | 0.9334 | 0.9423385262 | 0.236457 | 4.37135516 | 0 |
| 2 | 47414365 | A | G | S | G | MSH2 | ENST00000233146 | 0.36789 | 0.00586 | 0.16499 | 0.58617 | 0.70427 | 0.1606232115 | 0.93646 | 0.4119708538 | 0.129602 | 2.103488264 | 0 |
| 2 | 47414365 | A | T | S | C | MSH2 | ENST00000233146 | 0.55649 | 0.01628 | 0.20697 | 0.86611 | 0.91652 | 0.2534399838 | 0.91331 | 0.9892086387 | 0.281329 | 5.974112769 | 0 |
| 2 | 47414366 | G | A | S | N | MSH2 | ENST00000233146 | 0.36231 | 0.00586 | 0.23844 | 0.59718 | 0.61416 | 0.1896486549 | 0.8794 | 0.3008764286 | 0.112778 | 1.411144856 | 0 |
| 2 | 47414366 | G | C | S | T | MSH2 | ENST00000233146 | 0.36732 | 0.00592 | 0.19007 | 0.60196 | 0.78967 | 0.1998030577 | 0.93228 | 0.2362959981 | 0.157196 | 2.649380533 | 0 |
| 2 | 47414366 | G | T | S | I | MSH2 | ENST00000233146 | 0.5497 | 0.00996 | 0.22368 | 0.82008 | 0.88307 | 0.3522660361 | 0.94549 | 0.9821042418 | 0.29009 | 5.009713515 | 0 |
| 2 | 47414367 | C | A | S | R | MSH2 | ENST00000233146 | 0.36309 | 0.00556 | 0.25226 | 0.85859 | 0.87198 | 0.3178352275 | 0.90762 | 0.9827511907 | 0.218221 | 4.37135516 | 0 |
| 2 | 47414367 | C | G | S | R | MSH2 | ENST00000233146 | 0.36901 | 0.00556 | 0.25226 | 0.85341 | 0.87198 | 0.3178352275 | 0.90678 | 0.9037322402 | 0.0534631 | 4.37135516 | 0 |
| 2 | 47414368 | C | A | Q | K | MSH2 | ENST00000233146 | 0.48481 | 0.01633 | 0.58338 | 0.89451 | 0.95753 | 0.4948785523 | 0.98561 | 0.9879514575 | 0.476808 | 4.940032622 | 0 |
| 2 | 47414368 | C | G | Q | E | MSH2 | ENST00000233146 | 0.5896 | 0.02212 | 0.5322 | 0.8571 | 0.94618 | 0.4771207207 | 0.96473 | 0.9813541174 | 0.405874 | 4.115426327 | 0 |
| 2 | 47414369 | A | C | Q | P | MSH2 | ENST00000233146 | 0.79374 | 0.03137 | 0.54441 | 0.92509 | 0.99202 | 0.7499969084 | 0.983 | 0.9979154468 | 0.525435 | 9.118050235 | 0 |
| 2 | 47414369 | A | G | Q | R | MSH2 | ENST00000233146 | 0.4249 | 0.00773 | 0.57634 | 0.77061 | 0.91778 | 0.5129198744 | 0.99173 | 0.9516168237 | 0.459479 | 5.383981715 | 0 |
| 2 | 47414369 | A | T | Q | L | MSH2 | ENST00000233146 | 0.7939 | 0.00783 | 0.52554 | 0.92393 | 0.99202 | 0.5055897868 | 0.97485 | 0.9913424253 | 0.497553 | 5.775072575 | 0 |
| 2 | 47414371 | T | A | Y | N | MSH2 | ENST00000233146 | 0.84965 | 0.02937 | 0.84115 | 0.94402 | 0.99722 | 0.6734171084 | 0.95947 | 0.9994416833 | 0.535985 | 7.791898581 | 0 |
| 2 | 47414371 | T | C | Y | H | MSH2 | ENST00000233146 | 0.84385 | 0.03281 | 0.84743 | 0.9116 | 0.99676 | 0.6190058095 | 0.93424 | 0.997384727 | 0.525718 | 5.749193047 | 0 |
| 2 | 47414372 | A | C | Y | S | MSH2 | ENST00000233146 | 0.84522 | 0.03124 | 0.78728 | 0.94589 | 0.99786 | 0.6980200805 | 0.97973 | 0.9995065928 | 0.538937 | 7.01719088 | 0 |
| 2 | 47414372 | A | G | Y | C | MSH2 | ENST00000233146 | 0.8747 | 0.03327 | 0.84192 | 0.93598 | 0.99906 | 0.6313741127 | 0.95187 | 0.9994764924 | 0.542788 | 9.292657023 | 0 |
| 2 | 47414372 | A | T | Y | F | MSH2 | ENST00000233146 | 0.43243 | 0.00799 | 0.76528 | 0.73454 | 0.90318 | 0.3375259753 | 0.87893 | 0.9636397362 | 0.28542 | 3.223149029 | 0 |
| 2 | 47414374 | A | C | M | L | MSH2 | ENST00000233146 | 0.45702 | 0.00592 | 0.57403 | 0.62815 | 0.92561 | 0.6279902941 | 0.90767 | 0.9144801497 | 0.337824 | 3.399059838 | 0 |
| 2 | 47414374 | A | G | M | V | MSH2 | ENST00000233146 | 0.41801 | 0.00596 | 0.50391 | 0.75949 | 0.94431 | 0.6870038313 | 0.91799 | 0.9769332409 | 0.337803 | 3.983394678 | 0 |
| 2 | 47414374 | A | T | M | L | MSH2 | ENST00000233146 | 0.45181 | 0.00592 | 0.57403 | 0.62657 | 0.92561 | 0.6279902941 | 0.9077 | 0.9144801497 | 0.337824 | 3.399059838 | 0 |
| 2 | 47414375 | T | A | M | K | MSH2 | ENST00000233146 | 0.80384 | 0.01372 | 0.65636 | 0.92437 | 0.98657 | 0.8953319856 | 0.97954 | 0.9983955026 | 0.547767 | 8.630432407 | 1 |
| 2 | 47414375 | T | C | M | T | MSH2 | ENST00000233146 | 0.74507 | 0.01523 | 0.69813 | 0.92107 | 0.98034 | 0.8035528883 | 0.97181 | 0.9945592284 | 0.54443 | 8.630432407 | 0 |
| 2 | 47414375 | T | G | M | R | MSH2 | ENST00000233146 | 0.82311 | 0.01803 | 0.63273 | 0.92136 | 0.98728 | 0.9061476872 | 0.98132 | 0.9988402724 | 0.548409 | 8.630432407 | 1 |
| 2 | 47414376 | G | A | M | I | MSH2 | ENST00000233146 | 0.5478 | 0.00718 | 0.57268 | 0.85363 | 0.97364 | 0.6785656851 | 0.95743 | 0.9367388487 | 0.4914 | 7.385197813 | 0 |
| 2 | 47414376 | G | C | M | I | MSH2 | ENST00000233146 | 0.54501 | 0.00718 | 0.57268 | 0.85367 | 0.97364 | 0.6785656851 | 0.95714 | 0.9848697186 | 0.4914 | 7.385197813 | 0 |
| 2 | 47414376 | G | T | M | I | MSH2 | ENST00000233146 | 0.53954 | 0.00718 | 0.57268 | 0.85367 | 0.97364 | 0.6785656851 | 0.95721 | 0.9848697186 | 0.4914 | 7.385197813 | 0 |
| 2 | 47414377 | A | C | K | Q | MSH2 | ENST00000233146 | 0.61037 | 0.00909 | 0.36233 | 0.78734 | 0.87781 | 0.2642127748 | 0.92808 | 0.9410800934 | 0.22803 | 3.563475068 | 0 |
| 2 | 47414378 | A | C | K | T | MSH2 | ENST00000233146 | 0.33796 | 0.00569 | 0.45174 | 0.77636 | 0.81933 | 0.2792194842 | 0.92938 | 0.9875357747 | 0.199475 | 2.058413427 | 0 |
| 2 | 47414378 | A | G | K | R | MSH2 | ENST00000233146 | 0.25363 | 0.00608 | 0.33927 | 0.53253 | 0.5976 | 0.1874878144 | 0.90504 | 0.7362044454 | 0.120111 | 0.4448993679 | 0 |
| 2 | 47414378 | A | T | K | I | MSH2 | ENST00000233146 | 0.38794 | 0.00492 | 0.39134 | 0.81912 | 0.84267 | 0.2849043463 | 0.9345 | 0.994743824 | 0.279633 | 2.502561275 | 0 |
| 2 | 47414379 | A | C | K | N | MSH2 | ENST00000233146 | 0.56076 | 0.01009 | 0.46514 | 0.66418 | 0.8153 | 0.3085349181 | 0.9401 | 0.9733448625 | 0.167062 | 3.854427223 | 0 |
| 2 | 47414379 | A | T | K | N | MSH2 | ENST00000233146 | 0.5549 | 0.01009 | 0.46514 | 0.66418 | 0.81588 | 0.3085349181 | 0.93981 | 0.9733448625 | 0.167046 | 3.854427223 | 0 |
| 2 | 47414380 | T | A | L | M | MSH2 | ENST00000233146 | 0.59212 | 0.03767 | 0.68644 | 0.91288 | 0.95717 | 0.5221425427 | 0.94442 | 0.9879900813 | 0.495454 | 5.669503954 | 0 |
| 2 | 47414380 | T | G | L | V | MSH2 | ENST00000233146 | 0.57666 | 0.03697 | 0.64814 | 0.92584 | 0.97401 | 0.5271339781 | 0.97944 | 0.9902265072 | 0.521497 | 6.917454996 | 0 |
| 2 | 47414381 | T | C | L | S | MSH2 | ENST00000233146 | 0.93111 | 0.03231 | 0.72972 | 0.92111 | 0.9997 | 0.8818758534 | 0.98483 | 0.9992853999 | 0.570242 | 10.53035768 | 0 |
| 2 | 47414381 | T | G | L | W | MSH2 | ENST00000233146 | 0.9318 | 0.02984 | 0.79681 | 0.92163 | 0.9986 | 0.8239039159 | 0.97087 | 0.9993466735 | 0.570636 | 10.53035768 | 0 |
| 2 | 47414382 | G | C | L | F | MSH2 | ENST00000233146 | 0.78748 | 0.03595 | 0.69722 | 0.92111 | 0.96117 | 0.669963333 | 0.97964 | 0.9969115853 | 0.540949 | 8.52785857 | 0 |
| 2 | 47414382 | G | T | L | F | MSH2 | ENST00000233146 | 0.78107 | 0.03595 | 0.69722 | 0.92356 | 0.96117 | 0.669963333 | 0.98039 | 0.9973554611 | 0.540943 | 8.52785857 | 0 |
| 2 | 47414383 | G | A | D | N | MSH2 | ENST00000233146 | 0.87938 | 0.03753 | 0.71345 | 0.95226 | 0.98972 | 0.6790242467 | 0.96429 | 0.9982768297 | 0.277867 | 7.232958475 | 0 |
| 2 | 47414383 | G | C | D | H | MSH2 | ENST00000233146 | 0.87496 | 0.03465 | 0.73879 | 0.92866 | 0.99806 | 0.7503810239 | 0.98754 | 0.997877717 | 0.593168 | 9.538940509 | 0 |
| 2 | 47414384 | A | C | D | A | MSH2 | ENST00000233146 | 0.85152 | 0.03394 | 0.64088 | 0.94122 | 0.99976 | 0.7823466933 | 0.97946 | 0.9998309612 | 0.592852 | 9.538940509 | 0 |
| 2 | 47414384 | A | G | D | G | MSH2 | ENST00000233146 | 0.85756 | 0.03313 | 0.65524 | 0.93741 | 0.99919 | 0.763437259 | 0.98913 | 0.9994817376 | 0.586307 | 6.776141545 | 0 |
| 2 | 47414384 | A | T | D | V | MSH2 | ENST00000233146 | 0.8434 | 0.03189 | 0.63945 | 0.95059 | 0.99962 | 0.7819508174 | 0.9915 | 0.9997856021 | 0.592852 | 9.538940509 | 1 |
| 2 | 47414385 | T | A | D | E | MSH2 | ENST00000233146 | 0.63321 | 0.03815 | 0.6822 | 0.96113 | 0.98869 | 0.630548911 | 0.99526 | 0.9982377291 | 0.526545 | 7.685657201 | 0 |
| 2 | 47414385 | T | G | D | E | MSH2 | ENST00000233146 | 0.64112 | 0.03815 | 0.6822 | 0.96221 | 0.98869 | 0.630548911 | 0.99524 | 0.9978283048 | 0.526546 | 7.685657201 | 0 |
| 2 | 47414386 | A | C | I | L | MSH2 | ENST00000233146 | 0.48563 | 0.00574 | 0.34913 | 0.41266 | 0.69846 | 0.2321913464 | 0.8973 | 0.8971551657 | 0.136185 | 1.089055304 | 0 |
| 2 | 47414386 | A | G | I | V | MSH2 | ENST00000233146 | 0.43255 | 0.00608 | 0.28133 | 0.36823 | 0.71627 | 0.1197694272 | 0.92614 | 0.9311824441 | 0.140203 | 1.656834876 | 0 |
| 2 | 47414386 | A | T | I | F | MSH2 | ENST00000233146 | 0.5027 | 0.00512 | 0.3539 | 0.52093 | 0.76356 | 0.2654093675 | 0.91289 | 0.8949015141 | 0.172578 | 1.645769102 | 0 |
| 2 | 47414387 | T | A | I | N | MSH2 | ENST00000233146 | 0.38799 | 0.00476 | 0.39657 | 0.4694 | 0.65077 | 0.2744919156 | 0.73326 | 0.758802712 | 0.0968097 | -1.467863568 | 0 |
| 2 | 47414387 | T | C | I | T | MSH2 | ENST00000233146 | 0.35875 | 0.00527 | 0.41344 | 0.50098 | 0.72093 | 0.1594779021 | 0.93076 | 0.9195808768 | 0.195794 | 1.459757108 | 0 |
| 2 | 47414387 | T | G | I | S | MSH2 | ENST00000233146 | 0.34707 | 0.00504 | 0.33939 | 0.44082 | 0.74093 | 0.2526413877 | 0.8594 | 0.9561008215 | 0.135705 | -0.6251384115 | 0 |
| 2 | 47414388 | T | G | I | M | MSH2 | ENST00000233146 | 0.29278 | 0.00566 | 0.31617 | 0.31222 | 0.7337 | 0.2102962279 | 0.9083 | 0.1765148491 | 0.0721632 | 0.73991177 | 0 |
| 2 | 47414389 | G | A | A | T | MSH2 | ENST00000233146 | 0.92431 | 0.03753 | 0.67911 | 0.90846 | 0.9833 | 0.4019662366 | 0.93534 | 0.8392497897 | 0.322045 | 5.179185616 | 0 |
| 2 | 47414389 | G | C | A | P | MSH2 | ENST00000233146 | 0.92953 | 0.03322 | 0.73796 | 0.89687 | 0.98294 | 0.7617696143 | 0.90301 | 0.9975492358 | 0.486401 | 7.308007968 | 0 |
| 2 | 47414389 | G | T | A | S | MSH2 | ENST00000233146 | 0.77251 | 0.03821 | 0.63306 | 0.88078 | 0.91778 | 0.2879395924 | 0.83797 | 0.9229159355 | 0.236557 | 3.971650322 | 0 |
| 2 | 47414390 | C | A | A | E | MSH2 | ENST00000233146 | 0.61045 | 0.03484 | 0.74391 | 0.95356 | 0.98621 | 0.7007662038 | 0.92056 | 0.9958030581 | 0.439318 | 7.31004034 | 0 |
| 2 | 47414390 | C | G | A | G | MSH2 | ENST00000233146 | 0.61686 | 0.0199 | 0.65706 | 0.92968 | 0.9648 | 0.3219935566 | 0.87332 | 0.9925909042 | 0.349305 | 6.655543901 | 0 |
| 2 | 47414390 | C | T | A | V | MSH2 | ENST00000233146 | 0.62407 | 0.03692 | 0.69464 | 0.94454 | 0.96444 | 0.501690725 | 0.87806 | 0.9981880784 | 0.371119 | 6.215610162 | 0 |
| 2 | 47414392 | G | A | A | T | MSH2 | ENST00000233146 | 0.85603 | 0.03767 | 0.75703 | 0.91374 | 0.97587 | 0.5687793903 | 0.90417 | 0.9973512888 | 0.413323 | 3.497161316 | 0 |
| 2 | 47414392 | G | C | A | P | MSH2 | ENST00000233146 | 0.87234 | 0.03334 | 0.81427 | 0.91578 | 0.99326 | 0.8465349585 | 0.97023 | 0.9990757704 | 0.539012 | 9.683772211 | 1 |
| 2 | 47414392 | G | T | A | S | MSH2 | ENST00000233146 | 0.81928 | 0.03857 | 0.71559 | 0.92593 | 0.96189 | 0.4406213375 | 0.91067 | 0.9934874773 | 0.364485 | 5.120661484 | 0 |
| 2 | 47414393 | C | A | A | E | MSH2 | ENST00000233146 | 0.62165 | 0.03491 | 0.81858 | 0.92826 | 0.99265 | 0.833672468 | 0.96669 | 0.9994072914 | 0.530155 | 9.683772211 | 1 |
| 2 | 47414393 | C | G | A | G | MSH2 | ENST00000233146 | 0.62098 | 0.03713 | 0.74132 | 0.94925 | 0.97997 | 0.4807831962 | 0.91319 | 0.998159349 | 0.431207 | 9.683772211 | 0 |
| 2 | 47414393 | C | T | A | V | MSH2 | ENST00000233146 | 0.63519 | 0.03728 | 0.7821 | 0.92724 | 0.98257 | 0.6348659788 | 0.93054 | 0.9986709356 | 0.516983 | 6.983643973 | 0 |
| 2 | 47414395 | G | A | V | I | MSH2 | ENST00000233146 | 0.4485 | 0.01108 | 0.48798 | 0.68143 | 0.78658 | 0.2117048395 | 0.79351 | 0.8751331568 | 0.15631 | 5.121504298 | 0 |
| 2 | 47414395 | G | C | V | L | MSH2 | ENST00000233146 | 0.48164 | 0.01285 | 0.67253 | 0.69124 | 0.84747 | 0.4473678775 | 0.93792 | 0.9648604989 | 0.322537 | 4.759801595 | 0 |
| 2 | 47414395 | G | T | V | F | MSH2 | ENST00000233146 | 0.65171 | 0.00898 | 0.64152 | 0.85497 | 0.97067 | 0.7234680433 | 0.954 | 0.9802669287 | 0.452482 | 6.872452002 | 0 |
| 2 | 47414396 | T | C | V | A | MSH2 | ENST00000233146 | 0.47256 | 0.00681 | 0.68849 | 0.76385 | 0.87684 | 0.4879483756 | 0.91012 | 0.9630643129 | 0.243952 | 8.007439194 | 0 |
| 2 | 47414396 | T | G | V | G | MSH2 | ENST00000233146 | 0.67811 | 0.02307 | 0.53463 | 0.92622 | 0.96117 | 0.6674704452 | 0.97868 | 0.9955211878 | 0.458913 | 8.349205601 | 0 |
| 2 | 47414398 | A | G | R | G | MSH2 | ENST00000233146 | 0.4622 | 0.00566 | 0.32897 | 0.52093 | 0.81761 | 0.3329277167 | 0.97499 | 0.6981357336 | 0.179814 | 5.191245187 | 0 |
| 2 | 47414399 | G | A | R | K | MSH2 | ENST00000233146 | 0.41269 | 0.00644 | 0.39342 | 0.43393 | 0.78658 | 0.2642218086 | 0.94954 | 0.7760637999 | 0.187378 | 2.876733352 | 0 |
| 2 | 47414399 | G | C | R | T | MSH2 | ENST00000233146 | 0.58895 | 0.01662 | 0.36311 | 0.76317 | 0.90494 | 0.3332426465 | 0.96867 | 0.9717616439 | 0.327385 | 4.152899621 | 0 |
| 2 | 47414399 | G | T | R | I | MSH2 | ENST00000233146 | 0.77458 | 0.02063 | 0.36869 | 0.8542 | 0.93824 | 0.3457391855 | 0.93873 | 0.9938445091 | 0.292808 | 4.905253897 | 0 |
| 2 | 47414400 | A | C | R | S | MSH2 | ENST00000233146 | 0.58916 | 0.00777 | 0.33083 | 0.59708 | 0.87588 | 0.3377068027 | 0.97443 | 0.9430387616 | 0.26305 | 3.139658384 | 0 |
| 2 | 47414400 | A | T | R | S | MSH2 | ENST00000233146 | 0.58311 | 0.00777 | 0.33083 | 0.60298 | 0.87588 | 0.3377068027 | 0.97438 | 0.9430387616 | 0.263041 | 3.139658384 | 0 |
| 2 | 47414401 | G | A | A | T | MSH2 | ENST00000233146 | 0.90705 | 0.03753 | 0.73307 | 0.94817 | 0.99202 | 0.5007859818 | 0.9673 | 0.9960673451 | 0.530551 | 5.823940752 | 0 |
| 2 | 47414401 | G | C | A | P | MSH2 | ENST00000233146 | 0.91428 | 0.03334 | 0.78346 | 0.92018 | 0.99202 | 0.7975181043 | 0.97318 | 0.9990757704 | 0.539551 | 8.078616609 | 1 |
| 2 | 47414401 | G | T | A | S | MSH2 | ENST00000233146 | 0.87423 | 0.03674 | 0.69032 | 0.9353 | 0.97512 | 0.3708102488 | 0.9211 | 0.9934874773 | 0.410885 | 5.187972794 | 0 |
| 2 | 47414402 | C | A | A | D | MSH2 | ENST00000233146 | 0.87879 | 0.03106 | 0.82918 | 0.92282 | 0.99386 | 0.7750172393 | 0.95194 | 0.9996139407 | 0.526723 | 6.999586971 | 0 |

| | | | | | | | | | | | | | | | | | |
|---|---|---|---|---|---|---|---|---|---|---|---|---|---|---|---|---|---|
| 2 | 47414402 | C | G | A | G | MSH2 | ENST00000233146 | 0.8459 | 0.03697 | 0.70571 | 0.93858 | 0.98728 | 0.3862962728 | 0.92353 | 0.9970957041 | 0.467637 | 7.017237034 | 0 |
| 2 | 47414402 | C | T | A | V | MSH2 | ENST00000233146 | 0.87396 | 0.03718 | 0.74843 | 0.93089 | 0.98764 | 0.5728388021 | 0.9677 | 0.9991185069 | 0.45397 | 8.078616609 | 0 |
| 2 | 47414404 | C | A | L | I | MSH2 | ENST00000233146 | 0.78519 | 0.03602 | 0.75373 | 0.97803 | 0.97624 | 0.4257415695 | 0.90697 | 0.9975848198 | 0.462586 | 9.022981703 | 0 |
| 2 | 47414404 | C | G | L | V | MSH2 | ENST00000233146 | 0.78067 | 0.03706 | 0.70655 | 0.95781 | 0.98799 | 0.5714301217 | 0.96161 | 0.9936946034 | 0.437342 | 6.806127984 | 0 |
| 2 | 47414405 | T | A | L | H | MSH2 | ENST00000233146 | 0.87416 | 0.0334 | 0.83859 | 0.97709 | 0.99876 | 0.8152954907 | 0.98666 | 0.9988543987 | 0.572521 | 9.022981703 | 0 |
| 2 | 47414405 | T | C | L | P | MSH2 | ENST00000233146 | 0.89359 | 0.03006 | 0.87255 | 0.97746 | 0.99744 | 0.880777159 | 0.98774 | 0.9992852807 | 0.572514 | 9.022981703 | 1 |
| 2 | 47414405 | T | G | L | R | MSH2 | ENST00000233146 | 0.89024 | 0.0315 | 0.83985 | 0.97937 | 0.99786 | 0.8755751232 | 0.98702 | 0.9991292357 | 0.572246 | 9.022981703 | 1 |
| 2 | 47414407 | A | C | N | H | MSH2 | ENST00000233146 | 0.78962 | 0.03224 | 0.74145 | 0.88464 | 0.95645 | 0.4690044311 | 0.93551 | 0.9945688248 | 0.287154 | 7.618144533 | 0 |
| 2 | 47414407 | A | G | N | D | MSH2 | ENST00000233146 | 0.83821 | 0.03503 | 0.79022 | 0.89843 | 0.96371 | 0.29114027 | 0.96142 | 0.9976029992 | 0.235418 | 5.700609421 | 0 |
| 2 | 47414407 | A | T | N | Y | MSH2 | ENST00000233146 | 0.85938 | 0.03409 | 0.78038 | 0.94131 | 0.99265 | 0.4830071632 | 0.96833 | 0.9973629117 | 0.529152 | 7.618144533 | 0 |
| 2 | 47414408 | A | C | N | T | MSH2 | ENST00000233146 | 0.82147 | 0.0364 | 0.71284 | 0.91342 | 0.97923 | 0.3621813875 | 0.91729 | 0.9971141815 | 0.30565 | 7.618144533 | 0 |
| 2 | 47414408 | A | G | N | S | MSH2 | ENST00000233146 | 0.62453 | 0.03633 | 0.6622 | 0.88933 | 0.93786 | 0.2159352186 | 0.91877 | 0.9938171506 | 0.163342 | 5.12820798 | 0 |
| 2 | 47414408 | A | T | N | I | MSH2 | ENST00000233146 | 0.86318 | 0.03379 | 0.73633 | 0.94555 | 0.98834 | 0.5906486055 | 0.94021 | 0.9997856021 | 0.447732 | 7.618144533 | 0 |
| 2 | 47414409 | C | A | N | K | MSH2 | ENST00000233146 | 0.59443 | 0.03484 | 0.8131 | 0.92927 | 0.97252 | 0.5020245624 | 0.95484 | 0.9901409745 | 0.0784075 | 7.618144533 | 0 |
| 2 | 47414409 | C | G | N | K | MSH2 | ENST00000233146 | 0.60215 | 0.03484 | 0.8131 | 0.91657 | 0.97252 | 0.5020245624 | 0.95456 | 0.9979718328 | 0.206516 | 7.618144533 | 0 |
| 2 | 47414410 | C | A | L | I | MSH2 | ENST00000233146 | 0.52785 | 0.03413 | 0.71243 | 0.84769 | 0.90972 | 0.2970819619 | 0.8387 | 0.9734823769 | 0.283844 | 3.452827024 | 0 |
| 2 | 47414410 | C | G | L | V | MSH2 | ENST00000233146 | 0.53401 | 0.02506 | 0.65809 | 0.86244 | 0.95097 | 0.461537487 | 0.97611 | 0.6953661442 | 0.469955 | 6.210695107 | 0 |
| 2 | 47414410 | C | T | L | F | MSH2 | ENST00000233146 | 0.75957 | 0.03563 | 0.70672 | 0.89113 | 0.99105 | 0.6223280165 | 0.96572 | 0.9923181534 | 0.535767 | 9.216603289 | 0 |
| 2 | 47414411 | T | A | L | H | MSH2 | ENST00000233146 | 0.75941 | 0.03327 | 0.79091 | 0.9 | 0.99943 | 0.7717152482 | 0.98356 | 0.9993925095 | 0.551593 | 9.216603289 | 0 |
| 2 | 47414411 | T | C | L | P | MSH2 | ENST00000233146 | 0.78407 | 0.02994 | 0.83234 | 0.89579 | 0.997 | 0.8460209133 | 0.9855 | 0.999335587 | 0.551535 | 9.216603289 | 0 |
| 2 | 47414411 | T | G | L | R | MSH2 | ENST00000233146 | 0.77948 | 0.03137 | 0.80478 | 0.89784 | 0.99765 | 0.8432838513 | 0.98492 | 0.999397397 | 0.551314 | 9.216603289 | 0 |
| 2 | 47414413 | T | A | F | I | MSH2 | ENST00000233146 | 0.5106 | 0.02283 | 0.78036 | 0.84487 | 0.93554 | 0.6330343486 | 0.95909 | 0.8946580908 | 0.426349 | 4.100013119 | 0 |
| 2 | 47414413 | T | C | F | L | MSH2 | ENST00000233146 | 0.48063 | 0.00644 | 0.81195 | 0.72221 | 0.87925 | 0.5725601727 | 0.96342 | 0.9374228716 | 0.404763 | 2.084227762 | 0 |
| 2 | 47414413 | T | G | F | V | MSH2 | ENST00000233146 | 0.5123 | 0.02108 | 0.7644 | 0.82345 | 0.9367 | 0.62572405 | 0.96924 | 0.969301343 | 0.425726 | 3.870705753 | 0 |
| 2 | 47414414 | T | A | F | Y | MSH2 | ENST00000233146 | 0.6412 | 0.03313 | 0.79189 | 0.75911 | 0.95207 | 0.4996344617 | 0.92224 | 0.9867401719 | 0.403746 | 5.708799898 | 0 |
| 2 | 47414414 | T | C | F | S | MSH2 | ENST00000233146 | 0.73605 | 0.02307 | 0.78153 | 0.89351 | 0.97401 | 0.7276035125 | 0.97446 | 0.9910494089 | 0.455359 | 4.785381634 | 0 |
| 2 | 47414414 | T | G | F | C | MSH2 | ENST00000233146 | 0.78003 | 0.03083 | 0.80807 | 0.90967 | 0.98257 | 0.6918806917 | 0.95893 | 0.9953448176 | 0.469653 | 4.607465517 | 0 |
| 2 | 47414415 | T | A | F | L | MSH2 | ENST00000233146 | 0.22624 | 0.00644 | 0.81195 | 0.70016 | 0.84641 | 0.5725601727 | 0.98099 | 0.2199557275 | 0.338913 | 2.084227762 | 0 |
| 2 | 47414415 | T | G | F | L | MSH2 | ENST00000233146 | 0.23052 | 0.00644 | 0.81195 | 0.70016 | 0.84641 | 0.5725601727 | 0.98082 | 0.2199557275 | 0.338914 | 2.084227762 | 0 |
| 2 | 47414416 | C | A | Q | K | MSH2 | ENST00000233146 | 0.57744 | 0.01249 | 0.70362 | 0.85347 | 0.92316 | 0.3875683722 | 0.95855 | 0.9625675678 | 0.303887 | 3.168867809 | 0 |
| 2 | 47414416 | C | G | Q | E | MSH2 | ENST00000233146 | 0.5316 | 0.01324 | 0.64805 | 0.72835 | 0.84106 | 0.3476040314 | 0.91326 | 0.927210629 | 0.225253 | 2.914608854 | 0 |
| 2 | 47414417 | A | C | Q | P | MSH2 | ENST00000233146 | 0.40267 | 0.00547 | 0.65549 | 0.54205 | 0.7609 | 0.4758640312 | 0.95329 | 0.6733695865 | 0.198714 | 1.3431594 | 0 |
| 2 | 47414417 | A | G | Q | R | MSH2 | ENST00000233146 | 0.79104 | 0.0203 | 0.69953 | 0.81265 | 0.91058 | 0.3763257134 | 0.96685 | 0.8583104523 | 0.295387 | 3.576755493 | 0 |
| 2 | 47414417 | A | T | Q | L | MSH2 | ENST00000233146 | 0.69589 | 0.01831 | 0.6426 | 0.894 | 0.92964 | 0.4056348677 | 0.96329 | 0.9746379256 | 0.278327 | 5.728200607 | 0 |
| 2 | 47414418 | G | C | Q | H | MSH2 | ENST00000233146 | 0.89371 | 0.03404 | 0.69236 | 0.81507 | 0.92964 | 0.4900533471 | 0.93842 | 0.9572402239 | 0.297364 | 4.787876283 | 0 |
| 2 | 47414418 | G | T | Q | H | MSH2 | ENST00000233146 | 0.88886 | 0.03404 | 0.69236 | 0.81231 | 0.92964 | 0.4900533471 | 0.94283 | 0.9572402239 | 0.297338 | 4.787876283 | 0 |
| 2 | 47416296 | G | A | G | S | MSH2 | ENST00000233146 | 0.49711 | 0.00622 | 0.54012 | 0.63228 | 0.74235 | 0.2710267515 | 0.92964 | 0.8926380277 | 0.178458 | 0.9193834265 | 0 |
| 2 | 47416296 | G | C | G | R | MSH2 | ENST00000233146 | 0.56138 | 0.00654 | 0.67102 | 0.808 | 0.92233 | 0.362623782 | 0.95762 | 0.9780981541 | 0.402406 | 3.58131502 | 0 |
| 2 | 47416297 | G | A | G | D | MSH2 | ENST00000233146 | 0.37879 | 0.01276 | 0.68886 | 0.85359 | 0.89239 | 0.3649649405 | 0.94325 | 0.8586539626 | 0.409463 | 5.134208541 | 0 |
| 2 | 47416297 | G | C | G | A | MSH2 | ENST00000233146 | 0.3438 | 0.00608 | 0.56745 | 0.77688 | 0.84747 | 0.3173152608 | 0.93331 | 0.8972631097 | 0.283241 | 2.427716475 | 0 |
| 2 | 47416297 | G | T | G | V | MSH2 | ENST00000233146 | 0.38238 | 0.00608 | 0.58807 | 0.83619 | 0.93044 | 0.4136366512 | 0.98493 | 0.1537211365 | 0.17945 | 3.70959818 | 0 |
| 2 | 47416299 | T | A | S | T | MSH2 | ENST00000233146 | 0.39702 | 0.00812 | 0.63064 | 0.77584 | 0.82159 | 0.3035722229 | 0.96826 | 0.8370891809 | 0.288474 | 3.579147883 | 0 |
| 2 | 47416299 | T | C | S | P | MSH2 | ENST00000233146 | 0.45322 | 0.00574 | 0.64243 | 0.72676 | 0.85378 | 0.4249107217 | 0.96642 | 0.9054638147 | 0.408303 | 2.259136138 | 0 |
| 2 | 47416299 | T | G | S | A | MSH2 | ENST00000233146 | 0.41629 | 0.01574 | 0.61944 | 0.77622 | 0.80058 | 0.1858354968 | 0.94086 | 0.9339523911 | 0.285725 | 3.180944042 | 0 |
| 2 | 47416300 | C | A | S | Y | MSH2 | ENST00000233146 | 0.42837 | 0.0307 | 0.71087 | 0.90947 | 0.94986 | 0.4067824768 | 0.97853 | 0.9755845666 | 0.450958 | 5.560636667 | 0 |
| 2 | 47416300 | C | G | S | C | MSH2 | ENST00000233146 | 0.52532 | 0.03527 | 0.64376 | 0.85018 | 0.95207 | 0.332060596 | 0.90754 | 0.9748832583 | 0.440094 | 7.013077851 | 0 |
| 2 | 47416300 | C | T | S | F | MSH2 | ENST00000233146 | 0.48666 | 0.03161 | 0.68004 | 0.89103 | 0.95899 | 0.4059741584 | 0.98324 | 0.9305433035 | 0.320603 | 4.981717912 | 0 |
| 2 | 47416302 | G | A | V | I | MSH2 | ENST00000233146 | 0.39585 | 0.00648 | 0.28666 | 0.65077 | 0.77399 | 0.1169291788 | 0.8931 | 0.2007326017 | 0.054546 | 5.855360608 | 0 |
| 2 | 47416302 | G | C | V | L | MSH2 | ENST00000233146 | 0.40232 | 0.00574 | 0.42177 | 0.68774 | 0.7609 | 0.194495522 | 0.95441 | 0.3214052022 | 0.131579 | 0.9514805825 | 0 |
| 2 | 47416302 | G | T | V | F | MSH2 | ENST00000233146 | 0.46936 | 0.00644 | 0.39412 | 0.70887 | 0.81298 | 0.3068942508 | 0.91418 | 0.5890116096 | 0.193332 | 5.855360608 | 0 |
| 2 | 47416303 | T | A | V | D | MSH2 | ENST00000233146 | 0.28073 | 0.00492 | 0.43928 | 0.74401 | 0.82613 | 0.3342094833 | 0.95563 | 0.2898437366 | 0.233811 | 5.855360608 | 0 |
| 2 | 47416303 | T | C | V | A | MSH2 | ENST00000233146 | 0.24107 | 0.00535 | 0.43152 | 0.54234 | 0.79456 | 0.1641374555 | 0.95469 | 0.1861860305 | 0.157222 | 1.531816309 | 0 |
| 2 | 47416303 | T | G | V | G | MSH2 | ENST00000233146 | 0.29407 | 0.00527 | 0.31894 | 0.63919 | 0.79395 | 0.2255111966 | 0.96832 | 0.3511607051 | 0.212706 | 2.299262438 | 0 |
| 2 | 47416305 | G | A | E | K | MSH2 | ENST00000233146 | 0.5716 | 0.02083 | 0.65769 | 0.79996 | 0.81646 | 0.2297896505 | 0.92251 | 0.9068083167 | 0.276807 | 3.265407982 | 0 |
| 2 | 47416305 | G | C | E | Q | MSH2 | ENST00000233146 | 0.64695 | 0.03029 | 0.53191 | 0.73029 | 0.76816 | 0.2427563601 | 0.90139 | 0.8453912735 | 0.237268 | 3.450929905 | 0 |
| 2 | 47416306 | A | C | E | A | MSH2 | ENST00000233146 | 0.4901 | 0.00898 | 0.54693 | 0.72879 | 0.79334 | 0.2656005646 | 0.89642 | 0.8756110668 | 0.235987 | 4.386834736 | 0 |
| 2 | 47416306 | A | G | E | G | MSH2 | ENST00000233146 | 0.53751 | 0.00586 | 0.50574 | 0.65017 | 0.69168 | 0.2330643887 | 0.84703 | 0.8796070218 | 0.133693 | 3.474896792 | 0 |
| 2 | 47416306 | A | T | E | V | MSH2 | ENST00000233146 | 0.54535 | 0.02172 | 0.52538 | 0.82661 | 0.87296 | 0.3227247573 | 0.94931 | 0.9815058112 | 0.28059 | 4.802449688 | 0 |
| 2 | 47416308 | G | A | D | N | MSH2 | ENST00000233146 | 0.48208 | 0.00603 | 0.47833 | 0.52327 | 0.68995 | 0.2373950348 | 0.92196 | 0.8480658531 | 0.14669 | 2.329152511 | 0 |
| 2 | 47416308 | G | C | D | H | MSH2 | ENST00000233146 | 0.52187 | 0.00608 | 0.50899 | 0.67452 | 0.8517 | 0.3712220028 | 0.97521 | 0.962972343 | 0.358709 | 5.611208852 | 0 |
| 2 | 47416308 | G | T | D | Y | MSH2 | ENST00000233146 | 0.70262 | 0.02119 | 0.49876 | 0.9034 | 0.94469 | 0.4556210237 | 0.98356 | 0.9923547506 | 0.452748 | 6.675278464 | 0 |
| 2 | 47416309 | A | C | D | A | MSH2 | ENST00000233146 | 0.51659 | 0.00608 | 0.43793 | 0.78759 | 0.86803 | 0.3913370223 | 0.94202 | 0.9370994568 | 0.319698 | 3.874917601 | 0 |
| 2 | 47416309 | A | G | D | G | MSH2 | ENST00000233146 | 0.50641 | 0.00535 | 0.43919 | 0.79653 | 0.83888 | 0.3744829837 | 0.97709 | 0.4884149227 | 0.319008 | 1.917903366 | 0 |
| 2 | 47416309 | A | T | D | V | MSH2 | ENST00000233146 | 0.54623 | 0.01433 | 0.42596 | 0.92768 | 0.95863 | 0.4354360748 | 0.95774 | 0.9878979325 | 0.429942 | 6.497356176 | 0 |
| 2 | 47416310 | T | A | D | E | MSH2 | ENST00000233146 | 0.30899 | 0.00849 | 0.45691 | 0.74513 | 0.87684 | 0.145424107 | 0.93669 | 0.8980602026 | 0.226026 | 2.938478346 | 0 |
| 2 | 47416310 | T | G | D | E | MSH2 | ENST00000233146 | 0.31426 | 0.00849 | 0.45691 | 0.7394 | 0.87684 | 0.145424107 | 0.93635 | 0.8980602026 | 0.226027 | 2.938478346 | 0 |
| 2 | 47416311 | A | C | T | P | MSH2 | ENST00000233146 | 0.4988 | 0.00532 | 0.36307 | 0.77797 | 0.8124 | 0.3491035181 | 0.94132 | 0.7581439614 | 0.237211 | 4.127513884 | 0 |
| 2 | 47416311 | A | G | T | A | MSH2 | ENST00000233146 | 0.36342 | 0.00574 | 0.35362 | 0.56507 | 0.74517 | 0.2001753301 | 0.94889 | 0.1853335798 | -0.0330205 | 0.7634591072 | 0 |
| 2 | 47416311 | A | T | T | S | MSH2 | ENST00000233146 | 0.33158 | 0.00592 | 0.35945 | 0.60115 | 0.75074 | 0.1570258868 | 0.93639 | 0.4941916168 | 0.1075 | 2.171418239 | 0 |
| 2 | 47416312 | C | A | T | N | MSH2 | ENST00000233146 | 0.31209 | 0.00556 | 0.44332 | 0.59779 | 0.68558 | 0.2630925149 | 0.78914 | 0.7000043988 | 0.114811 | 2.508615897 | 0 |
| 2 | 47416312 | C | G | T | S | MSH2 | ENST00000233146 | 0.26461 | 0.00592 | 0.35945 | 0.5733 | 0.75822 | 0.1570258868 | 0.91719 | 0.572019279 | 0.107783 | 2.171418239 | 0 |
| 2 | 47416312 | C | T | T | I | MSH2 | ENST00000233146 | 0.39568 | 0.00562 | 0.50881 | 0.69186 | 0.8153 | 0.2443402494 | 0.89785 | 0.7759267688 | 0.213411 | 3.724762608 | 0 |
| 2 | 47416314 | A | C | T | P | MSH2 | ENST00000233146 | 0.17507 | 0.00983 | 0.23505 | 0.80216 | 0.83394 | 0.289576872 | 0.95718 | 0.08527062739 | 0.156892 | 5.865635942 | 0 |
| 2 | 47416314 | A | G | T | A | MSH2 | ENST00000233146 | 0.11469 | 0.0058 | 0.23051 | 0.63129 | 0.77718 | 0.1428871932 | 0.94985 | 0.02144748432 | 0.0688313 | 2.539708016 | 0 |
| 2 | 47416314 | A | T | T | S | MSH2 | ENST00000233146 | 0.08974 | 0.00592 | 0.21167 | 0.64968 | 0.74936 | 0.1124521329 | 0.87799 | 0.02414530993 | 0.0424388 | -0.858036736 | 0 |
| 2 | 47416315 | C | A | T | N | MSH2 | ENST00000233146 | 0.08068 | 0.00547 | 0.25901 | 0.55504 | 0.67108 | 0.1532873263 | 0.83939 | 0.06251097471 | 0.0465198 | -0.3910137374 | 0 |
| 2 | 47416315 | C | G | T | S | MSH2 | ENST00000233146 | 0.07835 | 0.00592 | 0.21167 | 0.50978 | 0.6847 | 0.1124521329 | 0.80942 | 0.03022565506 | 0.0387505 | -0.858036736 | 0 |
| 2 | 47416315 | C | T | T | I | MSH2 | ENST00000233146 | 0.15287 | 0.00821 | 0.29906 | 0.77366 | 0.79516 | 0.2117855102 | 0.94735 | 0.1838975218 | 0.163165 | 4.903646166 | 0 |
| 2 | 47416317 | G | A | G | S | MSH2 | ENST00000233146 | 0.47049 | 0.00592 | 0.33775 | 0.67268 | 0.7217 | 0.269766151 | 0.95296 | 0.3195779324 | 0.170977 | 3.845702659 | 0 |
| 2 | 47416317 | G | C | G | R | MSH2 | ENST00000233146 | 0.52041 | 0.00566 | 0.4498 | 0.66437 | 0.74306 | 0.3880872647 | 0.97929 | 0.7544693351 | 0.252428 | 1.525454024 | 0 |
| 2 | 47416317 | G | T | G | C | MSH2 | ENST00000233146 | 0.88739 | 0.02809 | 0.41268 | 0.87363 | 0.91015 | 0.3882343669 | 0.93787 | 0.9896657467 | 0.299595 | 6.365094406 | 0 |
| 2 | 47416318 | G | A | G | D | MSH2 | ENST00000233146 | 0.34948 | 0.00547 | 0.46788 | | 0.82103 | 0.3412169017 | | 0.01590811002 | -0.067059 | 4.992397979 | 0 |
| 2 | 47416318 | G | C | G | A | MSH2 | ENST00000233146 | 0.29567 | 0.00619 | 0.36501 | 0.6052 | 0.79395 | 0.2671549563 | 0.94066 | 0.7863372564 | 0.220376 | 4.611580316 | 0 |
| 2 | 47416318 | G | T | G | V | MSH2 | ENST00000233146 | 0.35953 | 0.01019 | 0.37599 | 0.79339 | 0.86553 | 0.3647530731 | 0.95303 | 0.4921633303 | 0.194427 | 3.785913309 | 0 |
| 2 | 47416320 | T | A | S | T | MSH2 | ENST00000233146 | 0.33489 | 0.00586 | 0.3598 | 0.49811 | 0.74936 | 0.1612734384 | 0.91931 | 0.165725708 | 0.0829949 | -0.07149062552 | 0 |
| 2 | 47416320 | T | C | S | P | MSH2 | ENST00000233146 | 0.43574 | 0.00535 | 0.36727 | 0.80354 | 0.90274 | 0.3477555266 | 0.97024 | 0.8288878798 | 0.237989 | 5.195198665 | 0 |
| 2 | 47416320 | T | G | S | A | MSH2 | ENST00000233146 | 0.38106 | 0.00608 | 0.34996 | 0.6852 | 0.7217 | 0.1045118095 | 0.93655 | 0.3287871778 | 0.0932022 | 1.768488018 | 0 |
| 2 | 47416321 | C | A | S | Y | MSH2 | ENST00000233146 | 0.23482 | 0.01048 | 0.40358 | 0.84019 | 0.85996 | 0.2359821955 | 0.93749 | 0.9691258669 | 0.240741 | 2.645831335 | 0 |
| 2 | 47416321 | C | G | S | C | MSH2 | ENST00000233146 | 0.39649 | 0.00608 | 0.37466 | 0.81076 | 0.92844 | 0.1952887886 | 0.95279 | 0.883681953 | 0.24937 | 6.096020178 | 0 |
| 2 | 47416323 | C | A | Q | K | MSH2 | ENST00000233146 | 0.38633 | 0.00547 | 0.49382 | 0.81814 | 0.87925 | 0.2619309833 | 0.9425 | 0.8214345574 | 0.286909 | 4.991988659 | 0 |
| 2 | 47416323 | C | G | Q | E | MSH2 | ENST00000233146 | 0.34817 | 0.00812 | 0.4472 | 0.76664 | 0.83172 | 0.2667716809 | 0.90318 | 0.5000254443 | 0.233786 | 5.486086876 | 0 |
| 2 | 47416324 | A | C | Q | P | MSH2 | ENST00000233146 | 0.75631 | 0.01676 | 0.45361 | 0.88733 | 0.94839 | 0.4769184655 | 0.95939 | 0.9773534536 | 0.432554 | 6.374975493 | 0 |
| 2 | 47416324 | A | G | Q | R | MSH2 | ENST00000233146 | 0.4864 | 0.0058 | 0.48983 | 0.74043 | 0.83228 | 0.266056978 | 0.97648 | 0.8162915707 | 0.28244 | 5.482000367 | 0 |
| 2 | 47416324 | A | T | Q | L | MSH2 | ENST00000233146 | 0.58789 | 0.0073 | 0.43594 | 0.82604 | 0.8887 | 0.2908431462 | 0.94461 | 0.9071896076 | 0.29013 | 5.419925056 | 0 |
| 2 | 47416325 | G | C | Q | H | MSH2 | ENST00000233146 | 0.39958 | 0.00592 | 0.47875 | 0.69505 | 0.74021 | 0.33913961 | 0.93206 | 0.5741699338 | 0.158702 | 1.984998562 | 0 |
| 2 | 47416325 | G | T | Q | H | MSH2 | ENST00000233146 | 0.39487 | 0.00592 | 0.47875 | 0.69541 | 0.74021 | 0.33913961 | 0.93223 | 0.5741699338 | 0.158641 | 1.984998562 | 0 |
| 2 | 47416326 | T | A | S | T | MSH2 | ENST00000233146 | 0.58631 | 0.01452 | 0.66637 | 0.87451 | 0.9681 | 0.6355429904 | 0.98391 | 0.9171252251 | 0.480796 | 2.797904034 | 0 |
| 2 | 47416326 | T | C | S | P | MSH2 | ENST00000233146 | 0.84249 | 0.03099 | 0.67463 | 0.91201 | 0.99578 | 0.7851597714 | 0.99334 | 0.9969525337 | 0.559739 | 7.940808473 | 0 |
| 2 | 47416326 | T | G | S | A | MSH2 | ENST00000233146 | 0.77664 | 0.02206 | 0.64759 | 0.90664 | 0.98367 | 0.5518519246 | 0.97635 | 0.9866669774 | 0.471984 | 5.495986606 | 0 |
| 2 | 47416327 | C | A | S | Y | MSH2 | ENST00000233146 | 0.60625 | 0.032 | 0.73515 | 0.95268 | 0.99234 | 0.745969652 | 0.99288 | 0.9972909093 | 0.565155 | 7.940808473 | 0 |
| 2 | 47416327 | C | G | S | C | MSH2 | ENST00000233146 | 0.47445 | 0.00885 | 0.66817 | 0.87932 | 0.94318 | 0.6674367337 | 0.98399 | 0.9640501142 | 0.519631 | 3.087448458 | 0 |

| | | | | | | | | | | | | | | | | | | |
|---|---|---|---|---|---|---|---|---|---|---|---|---|---|---|---|---|---|---|
| 2 | 47416327 | C | T | S | F | MSH2 | ENST00000233146 | 0.6539 | 0.03426 | 0.70089 | 0.95238 | 0.99603 | 0.7624830503 | 0.99176 | 0.9947584271 | 0.565164 | 7.940808473 | 0 |
| 2 | 47416329 | C | A | L | M | MSH2 | ENST00000233146 | 0.49679 | 0.03155 | 0.63262 | 0.87139 | 0.93939 | 0.4666753084 | 0.96049 | 0.9872566462 | 0.518861 | 6.44791308 | 0 |
| 2 | 47416329 | C | G | L | V | MSH2 | ENST00000233146 | 0.25062 | 0.01376 | 0.59818 | 0.82407 | 0.90362 | 0.5409072021 | 0.9765 | 0.9452729225 | 0.449313 | 3.36160866 | 0 |
| 2 | 47416330 | T | A | L | Q | MSH2 | ENST00000233146 | 0.92132 | 0.03613 | 0.64831 | 0.95172 | 0.99976 | 0.7599456408 | 0.99148 | 0.9981284738 | 0.586508 | 7.501087444 | 1 |
| 2 | 47416330 | T | C | L | P | MSH2 | ENST00000233146 | 0.9285 | 0.02972 | 0.7475 | 0.9549 | 0.99976 | 0.8935066901 | 0.991 | 0.9981284738 | 0.586731 | 7.501087444 | 1 |
| 2 | 47416330 | T | G | L | R | MSH2 | ENST00000233146 | 0.92653 | 0.03118 | 0.72478 | 0.95615 | 0.99982 | 0.8794452107 | 0.99145 | 0.9981284738 | 0.586537 | 7.501087444 | 1 |
| 2 | 47416332 | G | A | A | T | MSH2 | ENST00000233146 | 0.47357 | 0.00586 | 0.44907 | 0.62801 | 0.79395 | 0.3669122611 | 0.93681 | 0.8851551414 | 0.210101 | 3.119817696 | 0 |
| 2 | 47416332 | G | C | A | P | MSH2 | ENST00000233146 | 0.6905 | 0.01597 | 0.53098 | 0.84463 | 0.96553 | 0.7110186839 | 0.93879 | 0.9706429839 | 0.386627 | 6.480399237 | 0 |
| 2 | 47416332 | G | T | A | S | MSH2 | ENST00000233146 | 0.44223 | 0.00622 | 0.40747 | 0.68189 | 0.82556 | 0.3559644815 | 0.9249 | 0.8182220459 | 0.17179 | 4.07957276 | 0 |
| 2 | 47416333 | C | A | A | D | MSH2 | ENST00000233146 | 0.41428 | 0.01151 | 0.57882 | 0.84077 | 0.93709 | 0.7197050393 | 0.91128 | 0.9815806746 | 0.410957 | 6.480399237 | 0 |
| 2 | 47416333 | C | G | A | G | MSH2 | ENST00000233146 | 0.2901 | 0.00832 | 0.43632 | 0.73724 | 0.87296 | 0.3257054426 | 0.86324 | 0.6881267279 | 0.253054 | 6.480399237 | 0 |
| 2 | 47416333 | C | T | A | V | MSH2 | ENST00000233146 | 0.26303 | 0.00586 | 0.48471 | 0.59464 | 0.77205 | 0.3489343547 | 0.88036 | 0.5437095974 | 0.231494 | 2.73682309 | 0 |
| 2 | 47416335 | G | A | A | T | MSH2 | ENST00000233146 | 0.58895 | 0.00638 | 0.4376 | 0.7933 | 0.80238 | 0.3394360315 | 0.90561 | 0.7518864321 | 0.240169 | 4.218784569 | 0 |
| 2 | 47416335 | G | C | A | P | MSH2 | ENST00000233146 | 0.90437 | 0.01848 | 0.5243 | 0.93295 | 0.94167 | 0.6975160744 | 0.96183 | 0.5000134236 | 0.229105 | 5.356457824 | 0 |
| 2 | 47416335 | G | T | A | S | MSH2 | ENST00000233146 | 0.56103 | 0.00608 | 0.39314 | 0.80847 | 0.77011 | 0.2847927733 | 0.91989 | 0.7327052951 | 0.161246 | 0.6359899126 | 0 |
| 2 | 47416336 | C | A | A | D | MSH2 | ENST00000233146 | 0.40675 | 0.01108 | 0.55315 | 0.91775 | 0.86503 | 0.6187348231 | 0.94798 | 0.8975352049 | 0.273097 | 2.221461928 | 0 |
| 2 | 47416336 | C | G | A | G | MSH2 | ENST00000233146 | 0.21355 | 0.00569 | 0.41021 | 0.71236 | 0.65476 | 0.2791720292 | 0.73412 | 0.373193413 | 0.0824582 | -1.94206021 | 0 |
| 2 | 47416336 | C | T | A | V | MSH2 | ENST00000233146 | 0.41257 | 0.00965 | 0.4578 | 0.904 | 0.89513 | 0.4486813036 | 0.93326 | 0.9384479523 | 0.245594 | 5.356457824 | 0 |
| 2 | 47416338 | T | A | L | M | MSH2 | ENST00000233146 | 0.47244 | 0.03112 | 0.68703 | 0.83651 | 0.93786 | 0.3712492832 | 0.96019 | 0.9698810577 | 0.379762 | 8.027528377 | 0 |
| 2 | 47416338 | T | G | L | V | MSH2 | ENST00000233146 | 0.2203 | 0.00648 | 0.64779 | 0.76147 | 0.87442 | 0.4183365125 | 0.99093 | 0.6764534116 | 0.371104 | 3.867116029 | 0 |
| 2 | 47416339 | T | C | L | S | MSH2 | ENST00000233146 | 0.7874 | 0.02809 | 0.72076 | 0.94174 | 0.99006 | 0.706092055 | 0.97784 | 0.9981284738 | 0.508925 | 8.12483393 | 0 |
| 2 | 47416339 | T | G | L | W | MSH2 | ENST00000233146 | 0.79674 | 0.02857 | 0.79365 | 0.94452 | 0.99414 | 0.7420579208 | 0.95914 | 0.9985507131 | 0.499083 | 9.143652897 | 0 |
| 2 | 47416340 | G | C | L | F | MSH2 | ENST00000233146 | 0.53974 | 0.00735 | 0.69358 | 0.83649 | 0.91694 | 0.5019075775 | 0.98778 | 0.9895415306 | 0.422392 | 6.902308382 | 0 |
| 2 | 47416340 | G | T | L | F | MSH2 | ENST00000233146 | 0.53428 | 0.00735 | 0.69358 | 0.83662 | 0.91694 | 0.5019075775 | 0.98799 | 0.9895415306 | 0.422388 | 6.902308382 | 0 |
| 2 | 47416341 | C | A | L | M | MSH2 | ENST00000233146 | 0.5244 | 0.02633 | 0.78523 | 0.8933 | 0.91143 | 0.4641584866 | 0.90259 | 0.992348671 | 0.419048 | 4.466851593 | 0 |
| 2 | 47416341 | C | G | L | V | MSH2 | ENST00000233146 | 0.50485 | 0.03212 | 0.7573 | 0.9234 | 0.97698 | 0.5452868593 | 0.94876 | 0.9968866706 | 0.493058 | 5.755529727 | 0 |
| 2 | 47416342 | T | A | L | Q | MSH2 | ENST00000233146 | 0.85345 | 0.03557 | 0.81757 | 0.96368 | 0.99824 | 0.7621959793 | 0.98338 | 0.9995238781 | 0.575389 | 9.325882086 | 1 |
| 2 | 47416342 | T | C | L | P | MSH2 | ENST00000233146 | 0.86622 | 0.01244 | 0.89436 | 0.96204 | 0.99876 | 0.8972449889 | 0.98475 | 0.9996061921 | 0.57567 | 9.325882086 | 1 |
| 2 | 47416342 | T | G | L | R | MSH2 | ENST00000233146 | 0.86361 | 0.0307 | 0.87756 | 0.96401 | 0.99806 | 0.8810082369 | 0.98379 | 0.9995238781 | 0.575404 | 9.325882086 | 1 |
| 2 | 47416344 | A | C | N | H | MSH2 | ENST00000233146 | 0.76642 | 0.03509 | 0.78105 | 0.92985 | 0.98477 | 0.6711859392 | 0.96834 | 0.9970846772 | 0.31822 | 7.92379683 | 0 |
| 2 | 47416344 | A | G | N | D | MSH2 | ENST00000233146 | 0.44187 | 0.0084 | 0.81769 | 0.83178 | 0.9182 | 0.6460279944 | 0.93717 | 0.5898199677 | 0.0537102 | 2.967206988 | 0 |
| 2 | 47416344 | A | T | N | Y | MSH2 | ENST00000233146 | 0.81331 | 0.03212 | 0.80974 | 0.95468 | 0.99039 | 0.7348177719 | 0.96194 | 0.9984478951 | 0.506485 | 7.92379683 | 0 |
| 2 | 47416345 | A | C | N | T | MSH2 | ENST00000233146 | 0.7185 | 0.01075 | 0.75325 | 0.91286 | 0.96153 | 0.6053212985 | 0.92176 | 0.9926260114 | 0.29491 | 5.788534491 | 0 |
| 2 | 47416345 | A | G | N | S | MSH2 | ENST00000233146 | 0.71137 | 0.02972 | 0.71184 | 0.90956 | 0.97624 | 0.572597024 | 0.94062 | 0.9766213298 | 0.219896 | 7.92379683 | 0 |
| 2 | 47416345 | A | T | N | I | MSH2 | ENST00000233146 | 0.81528 | 0.02421 | 0.78346 | 0.94489 | 0.98621 | 0.7223268928 | 0.92543 | 0.9993367791 | 0.403317 | 7.92379683 | 0 |
| 2 | 47416346 | T | A | N | K | MSH2 | ENST00000233146 | 0.69163 | 0.03129 | 0.84101 | 0.93161 | 0.97178 | 0.7096930979 | 0.95086 | 0.9961408973 | 0.210864 | 7.92379683 | 0 |
| 2 | 47416346 | T | G | N | K | MSH2 | ENST00000233146 | 0.69989 | 0.03129 | 0.84101 | 0.92995 | 0.97178 | 0.7096930979 | 0.95082 | 0.9961408973 | 0.210868 | 7.92379683 | 0 |
| 2 | 47416347 | A | C | K | Q | MSH2 | ENST00000233146 | 0.83882 | 0.02186 | 0.83112 | 0.79339 | 0.94356 | 0.4861097497 | 0.92077 | 0.9374642372 | 0.249899 | 4.094762731 | 0 |
| 2 | 47416347 | A | G | K | E | MSH2 | ENST00000233146 | 0.74762 | 0.00622 | 0.86257 | 0.78146 | 0.95717 | 0.5884020954 | 0.96237 | 0.9371497035 | 0.376015 | 3.524603313 | 0 |
| 2 | 47416348 | A | C | K | T | MSH2 | ENST00000233146 | 0.85137 | 0.01953 | 0.8688 | 0.82999 | 0.96298 | 0.5641147691 | 0.92408 | 0.9901742935 | 0.366464 | 8.416916284 | 0 |
| 2 | 47416348 | A | G | K | R | MSH2 | ENST00000233146 | 0.49835 | 0.00633 | 0.80347 | 0.56805 | 0.79758 | 0.3978818579 | 0.94617 | 0.8821554184 | 0.180189 | 2.564735242 | 0 |
| 2 | 47416348 | A | T | K | M | MSH2 | ENST00000233146 | 0.88234 | 0.01053 | 0.80938 | 0.83246 | 0.96008 | 0.6190638544 | 0.91313 | 0.9926770926 | 0.342171 | 7.400524025 | 0 |
| 2 | 47416349 | G | C | K | N | MSH2 | ENST00000233146 | 0.49971 | 0.00659 | 0.88252 | 0.75955 | 0.89147 | 0.5602233364 | 0.89874 | 0.9689955711 | 0.104665 | 5.155110573 | 0 |
| 2 | 47416349 | G | T | K | N | MSH2 | ENST00000233146 | 0.49447 | 0.00659 | 0.88252 | 0.76166 | 0.89101 | 0.5602233364 | 0.89915 | 0.9689955711 | 0.10466 | 5.155110573 | 0 |
| 2 | 47416350 | T | A | C | S | MSH2 | ENST00000233146 | 0.82529 | 0.02506 | 0.91027 | 0.91625 | 0.98869 | 0.7304915326 | 0.96438 | 0.9995695949 | 0.514872 | 5.302043584 | 0 |
| 2 | 47416350 | T | C | C | R | MSH2 | ENST00000233146 | 0.8824 | 0.02628 | 0.96246 | 0.93788 | 0.99265 | 0.8277376617 | 0.97877 | 0.9996812344 | 0.536288 | 8.068616885 | 1 |
| 2 | 47416350 | T | G | C | G | MSH2 | ENST00000233146 | 0.86928 | 0.02732 | 0.88183 | 0.93424 | 0.9917 | 0.7699236917 | 0.97711 | 0.9996002316 | 0.51324 | 8.068616885 | 0 |
| 2 | 47416351 | G | A | C | Y | MSH2 | ENST00000233146 | 0.88417 | 0.0275 | 0.97043 | 0.90957 | 0.99105 | 0.7950405575 | 0.95519 | 0.9997765422 | 0.541263 | 8.068616885 | 1 |
| 2 | 47416351 | G | C | C | S | MSH2 | ENST00000233146 | 0.86865 | 0.02506 | 0.91027 | 0.92593 | 0.9755 | 0.7304915326 | 0.95776 | 0.9996979237 | 0.518538 | 5.302043584 | 0 |
| 2 | 47416351 | G | T | C | F | MSH2 | ENST00000233146 | 0.90025 | 0.02614 | 0.91594 | 0.93969 | 0.99326 | 0.7978325659 | 0.96909 | 0.9997894168 | 0.540812 | 8.068616885 | 1 |
| 2 | 47416352 | T | G | C | W | MSH2 | ENST00000233146 | 0.71129 | 0.03212 | 0.94809 | 0.97248 | 0.97364 | 0.8096320289 | 0.99001 | 0.9996526241 | 0.488873 | 8.068616885 | 1 |
| 2 | 47416353 | A | G | K | E | MSH2 | ENST00000233146 | 0.74021 | 0.01692 | 0.65153 | 0.87683 | 0.92844 | 0.3536823457 | 0.89584 | 0.9867222309 | 0.293889 | 4.578370709 | 0 |
| 2 | 47416354 | A | C | K | T | MSH2 | ENST00000233146 | 0.67358 | 0.02623 | 0.6848 | 0.87736 | 0.92723 | 0.387903931 | 0.88598 | 0.9897687435 | 0.254544 | 6.158653002 | 0 |
| 2 | 47416354 | A | G | K | R | MSH2 | ENST00000233146 | 0.29763 | 0.00633 | 0.56897 | 0.58383 | 0.7147 | 0.1874949202 | 0.92616 | 0.7323725224 | 0.130637 | 0.9367214246 | 0 |
| 2 | 47416354 | A | T | K | I | MSH2 | ENST00000233146 | 0.73181 | 0.02582 | 0.61271 | 0.92513 | 0.9328 | 0.3692443445 | 0.89808 | 0.995978117 | 0.260408 | 4.685296634 | 0 |
| 2 | 47416355 | A | C | K | N | MSH2 | ENST00000233146 | 0.61283 | 0.00777 | 0.69069 | 0.74461 | 0.83116 | 0.3360156371 | 0.88684 | 0.9835340381 | 0.151642 | 5.092984984 | 0 |
| 2 | 47416355 | A | T | K | N | MSH2 | ENST00000233146 | 0.60654 | 0.00777 | 0.69069 | 0.74434 | 0.83116 | 0.3360156371 | 0.88683 | 0.9835340381 | 0.151657 | 5.092984984 | 0 |
| 2 | 47416356 | A | C | T | P | MSH2 | ENST00000233146 | 0.81755 | 0.03299 | 0.67767 | 0.93904 | 0.99876 | 0.8383566854 | 0.98177 | 0.9979287386 | 0.585467 | 9.736434532 | 1 |
| 2 | 47416356 | A | G | T | A | MSH2 | ENST00000233146 | 0.77648 | 0.03388 | 0.68786 | 0.93726 | 0.99326 | 0.7906024147 | 0.99552 | 0.9965111613 | 0.565116 | 9.736434532 | 1 |
| 2 | 47416356 | A | T | T | S | MSH2 | ENST00000233146 | 0.723 | 0.02809 | 0.70583 | 0.91003 | 0.99072 | 0.663968182 | 0.98702 | 0.9933186769 | 0.555328 | 7.029909554 | 0 |
| 2 | 47416357 | C | G | T | S | MSH2 | ENST00000233146 | 0.5425 | 0.02809 | 0.70583 | 0.9207 | 0.98549 | 0.663968182 | 0.97829 | 0.9525619149 | 0.491778 | 7.029909554 | 0 |
| 2 | 47416357 | C | T | T | I | MSH2 | ENST00000233146 | 0.60207 | 0.0357 | 0.81575 | 0.96085 | 0.99765 | 0.8360262821 | 0.98539 | 0.9927515984 | 0.584342 | 9.736434532 | 0 |
| 2 | 47416359 | C | A | P | T | MSH2 | ENST00000233146 | 0.54439 | 0.00608 | 0.641 | 0.79785 | 0.91526 | 0.3517770405 | 0.96748 | 0.9827242494 | 0.280278 | 6.499848632 | 0 |
| 2 | 47416359 | C | G | P | A | MSH2 | ENST00000233146 | 0.3075 | 0.0058 | 0.58255 | 0.52359 | 0.77011 | 0.2639470076 | 0.92403 | 0.1032567293 | 0.0687282 | 1.198777385 | 0 |
| 2 | 47416359 | C | T | P | S | MSH2 | ENST00000233146 | 0.40249 | 0.00592 | 0.59901 | 0.7377 | 0.86902 | 0.3666865126 | 0.99058 | 0.7472664118 | 0.372145 | 2.594979395 | 0 |
| 2 | 47416360 | C | A | P | H | MSH2 | ENST00000233146 | 0.51014 | 0.00592 | 0.66132 | 0.84525 | 0.96262 | 0.3853939396 | 0.85395 | 0.9908186793 | 0.34163 | 7.488951783 | 0 |
| 2 | 47416360 | C | G | P | R | MSH2 | ENST00000233146 | 0.6218 | 0.0143 | 0.67689 | 0.8334 | 0.96117 | 0.5219698953 | 0.91092 | 0.9924364686 | 0.401518 | 7.488951783 | 0 |
| 2 | 47416360 | C | T | P | L | MSH2 | ENST00000233146 | 0.36103 | 0.00574 | 0.68579 | 0.677 | 0.88684 | 0.5127945166 | 0.95178 | 0.9938926697 | 0.282596 | 7.488951783 | 0 |
| 2 | 47416362 | C | A | Q | K | MSH2 | ENST00000233146 | 0.56269 | 0.02421 | 0.64185 | 0.79718 | 0.94543 | 0.6945039223 | 0.97698 | 0.9895218611 | 0.448984 | 7.329952079 | 0 |
| 2 | 47416362 | C | G | Q | E | MSH2 | ENST00000233146 | 0.65218 | 0.02335 | 0.57693 | 0.80296 | 0.91862 | 0.6336374235 | 0.94579 | 0.8472200332 | 0.358255 | 7.329952079 | 0 |
| 2 | 47416363 | A | C | Q | P | MSH2 | ENST00000233146 | 0.62476 | 0.00958 | 0.58937 | 0.84083 | 0.94394 | 0.8095192716 | 0.96931 | 0.9782636762 | 0.43889 | 4.983536957 | 0 |
| 2 | 47416363 | A | G | Q | R | MSH2 | ENST00000233146 | 0.48966 | 0.01002 | 0.63067 | 0.82141 | 0.93515 | 0.7206157041 | 0.9875 | 0.946053803 | 0.429218 | 7.329952079 | 0 |
| 2 | 47416363 | A | T | Q | L | MSH2 | ENST00000233146 | 0.83513 | 0.03029 | 0.57087 | 0.92145 | 0.95826 | 0.6334931168 | 0.95549 | 0.9971741438 | 0.422609 | 4.977212649 | 0 |
| 2 | 47416364 | A | C | Q | H | MSH2 | ENST00000233146 | 0.4496 | 0.03646 | 0.63152 | 0.86776 | 0.92438 | 0.6460728879 | 0.9793 | 0.9791849852 | 0.372337 | 7.329952079 | 0 |
| 2 | 47416364 | A | T | Q | H | MSH2 | ENST00000233146 | 0.44442 | 0.03646 | 0.63152 | 0.86761 | 0.92438 | 0.6460728879 | 0.9797 | 0.9886781573 | 0.372332 | 7.329952079 | 0 |
| 2 | 47416365 | G | A | G | R | MSH2 | ENST00000233146 | 0.90318 | 0.03442 | 0.89061 | 0.98481 | 0.99722 | 0.8901504549 | 0.98838 | 0.998980701 | 0.592574 | 10.19769469 | 1 |
| 2 | 47416365 | G | C | G | R | MSH2 | ENST00000233146 | 0.90106 | 0.03442 | 0.89061 | 0.98466 | 0.99765 | 0.8901504549 | 0.98821 | 0.998980701 | 0.592574 | 10.19769469 | 1 |
| 2 | 47416366 | G | A | G | E | MSH2 | ENST00000233146 | 0.80843 | 0.03287 | 0.87819 | 0.98618 | 0.99498 | 0.8919091262 | 0.98678 | 0.998980701 | 0.590941 | 10.19769469 | 1 |
| 2 | 47416366 | G | C | G | A | MSH2 | ENST00000233146 | 0.77267 | 0.03557 | 0.80973 | 0.96955 | 0.99652 | 0.8059994916 | 0.98829 | 0.9977733493 | 0.58525 | 10.19769469 | 1 |
| 2 | 47416366 | G | T | G | V | MSH2 | ENST00000233146 | 0.80408 | 0.03362 | 0.82091 | 0.98143 | 0.99765 | 0.8480628134 | 0.97955 | 0.9989829659 | 0.592991 | 10.19769469 | 1 |
| 2 | 47416368 | C | A | Q | K | MSH2 | ENST00000233146 | 0.65759 | 0.03189 | 0.71678 | 0.76264 | 0.92151 | 0.5529286129 | 0.95683 | 0.9716031551 | 0.399634 | 4.176826279 | 0 |
| 2 | 47416368 | C | G | Q | E | MSH2 | ENST00000233146 | 0.63122 | 0.032 | 0.65224 | 0.78871 | 0.91357 | 0.5052521486 | 0.91122 | 0.8978135586 | 0.289315 | 8.024897494 | 0 |
| 2 | 47416369 | A | C | Q | P | MSH2 | ENST00000233146 | 0.80614 | 0.03362 | 0.671 | 0.89122 | 0.98404 | 0.7775151672 | 0.95706 | 0.9894515872 | 0.456647 | 8.024897494 | 0 |
| 2 | 47416369 | A | G | Q | R | MSH2 | ENST00000233146 | 0.46905 | 0.03484 | 0.70423 | 0.81347 | 0.93863 | 0.582213902 | 0.96057 | 0.9797797799 | 0.387457 | 6.137042247 | 0 |
| 2 | 47416369 | A | T | Q | L | MSH2 | ENST00000233146 | 0.79548 | 0.03166 | 0.644 | 0.9054 | 0.96044 | 0.5700316042 | 0.94453 | 0.988941133 | 0.413105 | 8.024897494 | 0 |
| 2 | 47416370 | A | C | Q | H | MSH2 | ENST00000233146 | 0.19839 | 0.01731 | 0.70368 | 0.80794 | 0.87636 | 0.5107331305 | 0.98844 | 0.9456519485 | 0.337771 | 4.498903705 | 0 |
| 2 | 47416370 | A | T | Q | H | MSH2 | ENST00000233146 | 0.19521 | 0.01731 | 0.70368 | 0.80768 | 0.87636 | 0.5107331305 | 0.9888 | 0.9456519485 | 0.337766 | 4.498903705 | 0 |
| 2 | 47416371 | A | G | R | G | MSH2 | ENST00000233146 | 0.86003 | 0.0325 | 0.73308 | 0.94599 | 0.99105 | 0.7785594385 | 0.99355 | 0.9966049194 | 0.440435 | 10.19772391 | 0 |
| 2 | 47416372 | G | A | R | K | MSH2 | ENST00000233146 | 0.79144 | 0.01459 | 0.79851 | 0.92985 | 0.96444 | 0.7232950929 | 0.99094 | 0.9910061955 | 0.482544 | 10.19772391 | 0 |
| 2 | 47416372 | G | C | R | T | MSH2 | ENST00000233146 | 0.92877 | 0.03076 | 0.78642 | 0.93178 | 0.99265 | 0.7878251551 | 0.98433 | 0.9985845089 | 0.480936 | 10.19772391 | 0 |
| 2 | 47416372 | G | T | R | I | MSH2 | ENST00000233146 | 0.94342 | 0.02835 | 0.76362 | 0.94539 | 0.99202 | 0.7164168743 | 0.97663 | 0.9985389709 | 0.598054 | 10.19772391 | 0 |
| 2 | 47416373 | A | C | R | S | MSH2 | ENST00000233146 | 0.76926 | 0.03398 | 0.74431 | 0.82472 | 0.99105 | 0.7912760662 | 0.997 | 0.9977318645 | 0.53596 | 10.19772391 | 0 |
| 2 | 47416373 | A | T | R | S | MSH2 | ENST00000233146 | 0.76251 | 0.03398 | 0.74431 | 0.82862 | 0.99105 | 0.7912760662 | 0.99722 | 0.9977318645 | 0.535951 | 10.19772391 | 0 |
| 2 | 47416374 | C | A | L | I | MSH2 | ENST00000233146 | 0.52532 | 0.03602 | 0.79977 | 0.87796 | 0.92723 | 0.3511525378 | 0.86997 | 0.9897063971 | 0.330948 | 7.402631613 | 0 |
| 2 | 47416374 | C | G | L | V | MSH2 | ENST00000233146 | 0.52147 | 0.03706 | 0.75217 | 0.8362 | 0.96117 | 0.5462458329 | 0.92354 | 0.9386260509 | 0.250249 | 8.264632306 | 0 |
| 2 | 47416374 | C | T | L | F | MSH2 | ENST00000233146 | 0.55683 | 0.03595 | 0.78701 | 0.84562 | 0.97067 | 0.571033929 | 0.94771 | 0.995567143 | 0.478853 | 8.60000249 | 0 |
| 2 | 47416375 | T | A | L | H | MSH2 | ENST00000233146 | 0.86879 | 0.0334 | 0.86869 | 0.93642 | 0.99326 | 0.7397474304 | 0.95455 | 0.9992799163 | 0.553836 | 7.838752128 | 0 |
| 2 | 47416375 | T | C | L | P | MSH2 | ENST00000233146 | 0.88892 | 0.03006 | 0.89845 | 0.93706 | 0.99652 | 0.8912934507 | 0.97629 | 0.9995081425 | 0.554356 | 9.980201395 | 1 |
| 2 | 47416375 | T | G | L | R | MSH2 | ENST00000233146 | 0.88523 | 0.0315 | 0.87627 | 0.95035 | 0.99414 | 0.8621651514 | 0.96708 | 0.9992433786 | 0.544511 | 5.694555479 | 0 |
| 2 | 47416377 | G | A | V | I | MSH2 | ENST00000233146 | 0.37856 | 0.00659 | 0.20943 | 0.69149 | 0.82556 | 0.1993827933 | 0.95704 | 0.7067524195 | 0.0963997 | 2.182731245 | 0 |
| 2 | 47416377 | G | C | V | L | MSH2 | ENST00000233146 | 0.31049 | 0.00586 | 0.38272 | 0.43095 | 0.68908 | 0.364111935 | 0.85167 | 0.3782807887 | -0.0671167 | 1.144387543 | 0 |

| | | | | | | | | | | | | | | | | | | |
|---|---|---|---|---|---|---|---|---|---|---|---|---|---|---|---|---|---|---|
| 2 | 47416377 | G | T | V | F | MSH2 | ENST00000233146 | 0.71762 | 0.01791 | 0.35889 | 0.87122 | 0.93044 | 0.7078060393 | 0.90565 | 0.9319403768 | 0.444859 | 5.344601766 | 0 |
| 2 | 47416378 | T | A | V | D | MSH2 | ENST00000233146 | 0.77497 | 0.02602 | 0.41332 | 0.97043 | 0.98904 | 0.8499056946 | 0.96829 | 0.9986619949 | 0.532905 | 7.200627268 | 1 |
| 2 | 47416378 | T | C | V | A | MSH2 | ENST00000233146 | 0.55312 | 0.01108 | 0.40884 | 0.83961 | 0.95572 | 0.5283081929 | 0.89896 | 0.9636291862 | 0.303811 | 7.200627268 | 0 |
| 2 | 47416378 | T | G | V | G | MSH2 | ENST00000233146 | 0.75009 | 0.02484 | 0.26011 | 0.91048 | 0.98728 | 0.7140500604 | 0.95689 | 0.9975766765 | 0.475078 | 7.200627268 | 1 |
| 2 | 47416380 | A | C | N | H | MSH2 | ENST00000233146 | 0.38782 | 0.00566 | 0.67939 | 0.11566 | 0.60123 | 0.3912771045 | 0.9422 | 0.7646433711 | 0.123892 | 4.523370222 | 0 |
| 2 | 47416380 | A | G | N | D | MSH2 | ENST00000233146 | 0.43136 | 0.00608 | 0.73208 | 0.74097 | 0.80178 | 0.4972345697 | 0.92862 | 0.9663241506 | 0.137008 | 6.047934142 | 0 |
| 2 | 47416380 | A | T | N | Y | MSH2 | ENST00000233146 | 0.37436 | 0.00543 | 0.7166 | 0.39055 | 0.83004 | 0.5062385403 | 0.96133 | 0.9248754382 | 0.250145 | 5.104039603 | 0 |
| 2 | 47416381 | A | C | N | T | MSH2 | ENST00000233146 | 0.37109 | 0.00566 | 0.64128 | 0.1531 | 0.74446 | 0.3307339909 | 0.93659 | 0.7646433711 | 0.139716 | 3.512311266 | 0 |
| 2 | 47416381 | A | G | N | S | MSH2 | ENST00000233146 | 0.32383 | 0.00608 | 0.58342 | 0.61176 | 0.60243 | 0.2762870113 | 0.90237 | 0.6468669772 | 0.0990923 | 2.626031932 | 0 |
| 2 | 47416381 | A | T | N | I | MSH2 | ENST00000233146 | 0.43058 | 0.00705 | 0.67005 | 0.45285 | 0.89963 | 0.497210553 | 0.92039 | 0.9708390832 | 0.245085 | 6.047934142 | 0 |
| 2 | 47416382 | C | A | N | K | MSH2 | ENST00000233146 | 0.33217 | 0.00608 | 0.76192 | 0.7547 | 0.73148 | 0.4399830087 | 0.89555 | 0.462705642 | 0.0888991 | 2.538925439 | 0 |
| 2 | 47416382 | C | G | N | K | MSH2 | ENST00000233146 | 0.33785 | 0.00608 | 0.76192 | 0.75444 | 0.73148 | 0.4399830087 | 0.89574 | 0.462705642 | 0.0888971 | 2.538925439 | 0 |
| 2 | 47416383 | C | A | Q | K | MSH2 | ENST00000233146 | 0.67939 | 0.02193 | 0.70146 | 0.85149 | 0.94318 | 0.6187504946 | 0.98004 | 0.9914814234 | 0.45427 | 4.567493676 | 0 |
| 2 | 47416383 | C | G | Q | E | MSH2 | ENST00000233146 | 0.66114 | 0.02335 | 0.64058 | 0.87048 | 0.93397 | 0.5417974995 | 0.95717 | 0.9578715563 | 0.350711 | 7.233486152 | 0 |
| 2 | 47416384 | A | C | Q | P | MSH2 | ENST00000233146 | 0.82295 | 0.03231 | 0.65643 | 0.89731 | 0.98657 | 0.8115396201 | 0.97516 | 0.9976266026 | 0.501746 | 9.723865478 | 1 |
| 2 | 47416384 | A | G | Q | R | MSH2 | ENST00000233146 | 0.61716 | 0.01025 | 0.6963 | 0.81951 | 0.93863 | 0.6439140304 | 0.98773 | 0.9751445651 | 0.434746 | 5.105531244 | 0 |
| 2 | 47416384 | A | T | Q | L | MSH2 | ENST00000233146 | 0.83905 | 0.03334 | 0.63388 | 0.93521 | 0.96919 | 0.6153407176 | 0.94429 | 0.9952778816 | 0.446151 | 7.427650269 | 0 |
| 2 | 47416385 | G | C | Q | H | MSH2 | ENST00000233146 | 0.60874 | 0.03697 | 0.69675 | 0.88562 | 0.96444 | 0.5902938224 | 0.98164 | 0.944653511 | 0.278115 | 6.573497033 | 0 |
| 2 | 47416385 | G | T | Q | H | MSH2 | ENST00000233146 | 0.6028 | 0.03697 | 0.69675 | 0.8913 | 0.96444 | 0.5902938224 | 0.98202 | 0.9963173866 | 0.442874 | 6.573497033 | 0 |
| 2 | 47416386 | T | A | W | R | MSH2 | ENST00000233146 | 0.896 | 0.02857 | 0.96356 | 0.92548 | 0.98621 | 0.8457564957 | 0.96168 | 0.999747932 | 0.537292 | 9.06468846 | 1 |
| 2 | 47416386 | T | C | W | R | MSH2 | ENST00000233146 | 0.90625 | 0.02857 | 0.96356 | 0.91866 | 0.98621 | 0.8457564957 | 0.96211 | 0.999747932 | 0.537291 | 9.06468846 | 1 |
| 2 | 47416386 | T | G | W | G | MSH2 | ENST00000233146 | 0.9253 | 0.02851 | 0.92248 | 0.96018 | 0.99498 | 0.7795340422 | 0.98371 | 0.9997624755 | 0.544952 | 9.984271254 | 1 |
| 2 | 47416387 | G | C | W | S | MSH2 | ENST00000233146 | 0.93842 | 0.02989 | 0.93965 | 0.92089 | 0.9755 | 0.8144193913 | 0.97278 | 0.9998836517 | 0.547778 | 7.312093693 | 1 |
| 2 | 47416387 | G | T | W | L | MSH2 | ENST00000233146 | 0.92446 | 0.02844 | 0.94768 | 0.92833 | 0.97698 | 0.7083239817 | 0.96479 | 0.9998732805 | 0.53828 | 7.852346394 | 0 |
| 2 | 47416388 | G | C | W | C | MSH2 | ENST00000233146 | 0.95546 | 0.02589 | 0.9438 | 0.94438 | 0.97327 | 0.8249152331 | 0.98437 | 0.99987638 | 0.546164 | 7.188049508 | 1 |
| 2 | 47416388 | G | T | W | C | MSH2 | ENST00000233146 | 0.95516 | 0.02589 | 0.9438 | 0.94438 | 0.97327 | 0.8249152331 | 0.98444 | 0.99987638 | 0.546164 | 7.188049508 | 1 |
| 2 | 47416389 | A | C | I | L | MSH2 | ENST00000233146 | 0.45181 | 0.00681 | 0.61016 | 0.65797 | 0.81124 | 0.4514618821 | 0.84934 | 0.7035981417 | 0.207272 | 3.335469504 | 0 |
| 2 | 47416389 | A | G | I | V | MSH2 | ENST00000233146 | 0.41316 | 0.00812 | 0.49819 | 0.76295 | 0.80058 | 0.2625293347 | 0.85943 | 0.7702945471 | 0.214971 | 2.304499667 | 0 |
| 2 | 47416389 | A | T | I | F | MSH2 | ENST00000233146 | 0.85115 | 0.02628 | 0.62645 | 0.90483 | 0.9833 | 0.693144556 | 0.97475 | 0.9927850962 | 0.48278 | 8.809899203 | 0 |
| 2 | 47416390 | T | A | I | N | MSH2 | ENST00000233146 | 0.87173 | 0.0275 | 0.72253 | 0.95175 | 0.98657 | 0.8154243325 | 0.93658 | 0.9968242645 | 0.468656 | 8.809899203 | 1 |
| 2 | 47416390 | T | C | I | T | MSH2 | ENST00000233146 | 0.80929 | 0.02883 | 0.74625 | 0.93257 | 0.98728 | 0.6403940025 | 0.97534 | 0.9945560694 | 0.50866 | 7.172939761 | 0 |
| 2 | 47416390 | T | G | I | S | MSH2 | ENST00000233146 | 0.87643 | 0.02979 | 0.63817 | 0.95192 | 0.99138 | 0.8048633951 | 0.98281 | 0.9978208542 | 0.514925 | 8.809899203 | 0 |
| 2 | 47416391 | T | G | I | M | MSH2 | ENST00000233146 | 0.49486 | 0.03409 | 0.56538 | 0.89462 | 0.95243 | 0.5187036262 | 0.97596 | 0.9387248755 | 0.43781 | 4.755625565 | 0 |
| 2 | 47416392 | A | C | K | Q | MSH2 | ENST00000233146 | 0.8249 | 0.03379 | 0.55017 | 0.92394 | 0.97252 | 0.5033591684 | 0.96332 | 0.996905148 | 0.40393 | 5.307469365 | 0 |
| 2 | 47416392 | A | G | K | E | MSH2 | ENST00000233146 | 0.8498 | 0.02802 | 0.60033 | 0.92552 | 0.97141 | 0.6118772749 | 0.98868 | 0.9978181124 | 0.495137 | 4.355534843 | 0 |
| 2 | 47416393 | A | C | K | T | MSH2 | ENST00000233146 | 0.83498 | 0.03293 | 0.6085 | 0.94061 | 0.96919 | 0.5619944429 | 0.9587 | 0.9980739355 | 0.483441 | 6.268215769 | 0 |
| 2 | 47416393 | A | G | K | R | MSH2 | ENST00000233146 | 0.51468 | 0.0133 | 0.51661 | 0.82129 | 0.91015 | 0.4218180365 | 0.97115 | 0.9292574525 | 0.356756 | 5.716266597 | 0 |
| 2 | 47416393 | A | T | K | M | MSH2 | ENST00000233146 | 0.86699 | 0.03197 | 0.52246 | 0.94964 | 0.97997 | 0.6154284115 | 0.94803 | 0.9983458519 | 0.483213 | 7.845922694 | 0 |
| 2 | 47416394 | G | C | K | N | MSH2 | ENST00000233146 | 0.56221 | 0.0151 | 0.63175 | 0.93601 | 0.92682 | 0.5961589473 | 0.95261 | 0.9936156273 | 0.179536 | 4.899117409 | 0 |
| 2 | 47416394 | G | T | K | N | MSH2 | ENST00000233146 | 0.55663 | 0.0151 | 0.63175 | 0.93699 | 0.92682 | 0.5961589473 | 0.95293 | 0.9936156273 | 0.179531 | 4.899117409 | 0 |
| 2 | 47416395 | C | A | Q | K | MSH2 | ENST00000233146 | 0.71121 | 0.03293 | 0.71211 | 0.921 | 0.96993 | 0.6806019942 | 0.97513 | 0.9969131947 | 0.490489 | 4.735919389 | 0 |
| 2 | 47416395 | C | G | Q | E | MSH2 | ENST00000233146 | 0.66359 | 0.02972 | 0.66361 | 0.86776 | 0.95353 | 0.6575734919 | 0.95461 | 0.9842718244 | 0.418117 | 5.282890124 | 0 |
| 2 | 47416396 | A | C | Q | P | MSH2 | ENST00000233146 | 0.8228 | 0.03419 | 0.673 | 0.9277 | 0.98404 | 0.8291959041 | 0.96082 | 0.9979426265 | 0.492937 | 9.488480577 | 1 |
| 2 | 47416396 | A | G | Q | R | MSH2 | ENST00000233146 | 0.80163 | 0.03587 | 0.707 | 0.89111 | 0.97624 | 0.7150865924 | 0.98854 | 0.9778523445 | 0.488001 | 4.698895754 | 0 |
| 2 | 47416396 | A | T | Q | L | MSH2 | ENST00000233146 | 0.84816 | 0.03735 | 0.65115 | 0.93099 | 0.98585 | 0.6638346442 | 0.96137 | 0.9972685575 | 0.48362 | 8.457558919 | 0 |
| 2 | 47416398 | C | A | P | T | MSH2 | ENST00000233146 | 0.68571 | 0.03552 | 0.79268 | 0.98467 | 0.99962 | 0.7559227773 | 0.98202 | 0.9994627833 | 0.587603 | 10.19018705 | 1 |
| 2 | 47416398 | C | G | P | A | MSH2 | ENST00000233146 | 0.66454 | 0.03761 | 0.7484 | 0.98414 | 0.9999 | 0.697967007 | 0.9846 | 0.992474854 | 0.37143 | 10.19018705 | 0 |
| 2 | 47416398 | C | T | P | S | MSH2 | ENST00000233146 | 0.71626 | 0.03706 | 0.75962 | 0.98424 | 0.99906 | 0.7742598186 | 0.98437 | 0.9994627833 | 0.587602 | 10.19018705 | 1 |
| 2 | 47416399 | C | A | P | H | MSH2 | ENST00000233146 | 0.61978 | 0.03503 | 0.80725 | 0.98491 | 0.9986 | 0.7999762202 | 0.97396 | 0.9994639754 | 0.587404 | 10.19018705 | 1 |
| 2 | 47416399 | C | G | P | R | MSH2 | ENST00000233146 | 0.62362 | 0.03633 | 0.82913 | 0.98476 | 0.99919 | 0.8206243027 | 0.97926 | 0.9994639754 | 0.587404 | 10.19018705 | 1 |
| 2 | 47416399 | C | T | P | L | MSH2 | ENST00000233146 | 0.64143 | 0.03692 | 0.82376 | 0.98404 | 0.99943 | 0.8062611275 | 0.99164 | 0.9999060631 | 0.587403 | 10.19018705 | 1 |
| 2 | 47416401 | C | A | L | I | MSH2 | ENST00000233146 | 0.63298 | 0.03602 | 0.6579 | 0.91252 | 0.93709 | 0.3792149453 | 0.88911 | 0.9977814555 | 0.374671 | 8.349534461 | 0 |
| 2 | 47416401 | C | G | L | V | MSH2 | ENST00000233146 | 0.62856 | 0.03706 | 0.58661 | 0.91378 | 0.96664 | 0.5200730179 | 0.95047 | 0.9964259267 | 0.508617 | 8.341185778 | 0 |
| 2 | 47416402 | T | A | L | H | MSH2 | ENST00000233146 | 0.725 | 0.0334 | 0.74137 | 0.94883 | 0.99414 | 0.7896478342 | 0.96919 | 0.9995288849 | 0.545381 | 8.275576791 | 0 |
| 2 | 47416402 | T | C | L | P | MSH2 | ENST00000233146 | 0.74978 | 0.03006 | 0.78669 | 0.94806 | 0.99296 | 0.8554286919 | 0.97335 | 0.9996453524 | 0.540961 | 9.359827572 | 0 |
| 2 | 47416402 | T | G | L | R | MSH2 | ENST00000233146 | 0.74523 | 0.0315 | 0.7544 | 0.94766 | 0.99498 | 0.8475035304 | 0.97519 | 0.9994131327 | 0.544514 | 7.661326042 | 0 |
| 2 | 47416404 | A | C | M | L | MSH2 | ENST00000233146 | 0.29889 | 0.0058 | 0.51377 | 0.58394 | 0.71705 | 0.3001428359 | 0.9156 | 0.1642862111 | 0.0854141 | 1.417422507 | 0 |
| 2 | 47416404 | A | G | M | V | MSH2 | ENST00000233146 | 0.287 | 0.0058 | 0.44097 | 0.76143 | 0.84853 | 0.3788819066 | 0.94294 | 0.06405485828 | 0.0524361 | 2.388590273 | 0 |
| 2 | 47416404 | A | T | M | L | MSH2 | ENST00000233146 | 0.29462 | 0.0058 | 0.51377 | 0.58394 | 0.71705 | 0.3001428359 | 0.91279 | 0.1642862111 | 0.0854246 | 1.417422507 | 0 |
| 2 | 47416405 | T | A | M | K | MSH2 | ENST00000233146 | 0.48272 | 0.00754 | 0.62227 | 0.85885 | 0.88823 | 0.6360570769 | 0.82764 | 0.9485078454 | 0.260248 | 4.628910225 | 0 |
| 2 | 47416405 | T | C | M | T | MSH2 | ENST00000233146 | 0.45856 | 0.00551 | 0.66819 | 0.75838 | 0.7872 | 0.5095543162 | 0.859 | 0.8730852008 | 0.202022 | 4.582454619 | 0 |
| 2 | 47416405 | T | G | M | R | MSH2 | ENST00000233146 | 0.50374 | 0.01048 | 0.59903 | 0.83368 | 0.91526 | 0.6992391614 | 0.84163 | 0.9676423669 | 0.266304 | 4.743775696 | 0 |
| 2 | 47416406 | G | A | M | I | MSH2 | ENST00000233146 | 0.50792 | 0.00556 | 0.52602 | 0.76245 | 0.80118 | 0.3276591943 | 0.88659 | 0.5359046788 | 0.17327 | 2.876350968 | 0 |
| 2 | 47416406 | G | C | M | I | MSH2 | ENST00000233146 | 0.50524 | 0.00556 | 0.52602 | 0.76213 | 0.80118 | 0.3276591943 | 0.88479 | 0.4610322171 | 0.173274 | 2.876350968 | 0 |
| 2 | 47416406 | G | T | M | I | MSH2 | ENST00000233146 | 0.49997 | 0.00556 | 0.52602 | 0.76213 | 0.80118 | 0.3276591943 | 0.88792 | 0.4380809289 | 0.0555252 | 2.876350968 | 0 |
| 2 | 47416407 | G | A | D | N | MSH2 | ENST00000233146 | 0.72427 | 0.03747 | 0.77291 | 0.8496 | 0.89873 | 0.4411120545 | 0.88773 | 0.960926652 | 0.123915 | 4.063163078 | 0 |
| 2 | 47416407 | G | C | D | H | MSH2 | ENST00000233146 | 0.87429 | 0.03465 | 0.79315 | 0.90909 | 0.97401 | 0.6109093915 | 0.98026 | 0.9947579503 | 0.521364 | 5.91454805 | 0 |
| 2 | 47416407 | G | T | D | Y | MSH2 | ENST00000233146 | 0.88112 | 0.03409 | 0.77937 | 0.94234 | 0.98367 | 0.7086188884 | 0.98575 | 0.9970929623 | 0.473829 | 7.635999981 | 0 |
| 2 | 47416408 | A | C | D | A | MSH2 | ENST00000233146 | 0.86886 | 0.03388 | 0.71736 | 0.93178 | 0.9917 | 0.6382926941 | 0.95706 | 0.9970807312 | 0.530011 | 7.255357687 | 0 |
| 2 | 47416408 | A | G | D | G | MSH2 | ENST00000233146 | 0.87349 | 0.03304 | 0.7269 | 0.92658 | 0.98146 | 0.6142628587 | 0.98034 | 0.9973487854 | 0.521353 | 8.853388518 | 0 |
| 2 | 47416408 | A | T | D | V | MSH2 | ENST00000233146 | 0.86233 | 0.0318 | 0.71283 | 0.94902 | 0.98764 | 0.6763588079 | 0.98782 | 0.9996447563 | 0.545534 | 6.375011558 | 0 |
| 2 | 47416409 | T | A | D | E | MSH2 | ENST00000233146 | 0.68803 | 0.03815 | 0.74866 | 0.83461 | 0.94618 | 0.3887165184 | 0.97652 | 0.9876266122 | 0.434071 | 5.600399773 | 0 |
| 2 | 47416409 | T | G | D | E | MSH2 | ENST00000233146 | 0.69628 | 0.03815 | 0.74866 | 0.83344 | 0.94618 | 0.3887165184 | 0.97629 | 0.9876266122 | 0.434074 | 5.600399773 | 0 |
| 2 | 47416410 | A | C | K | Q | MSH2 | ENST00000233146 | 0.56734 | 0.0071 | 0.6084 | 0.70689 | 0.87394 | 0.3398877628 | 0.90731 | 0.8793563843 | 0.243888 | 3.787443241 | 0 |
| 2 | 47416410 | A | G | K | E | MSH2 | ENST00000233146 | 0.59929 | 0.00718 | 0.65813 | 0.76886 | 0.88401 | 0.4461229622 | 0.93765 | 0.8719242811 | 0.304316 | 4.787889818 | 0 |
| 2 | 47416411 | A | C | K | T | MSH2 | ENST00000233146 | 0.48767 | 0.00566 | 0.66949 | 0.74181 | 0.80118 | 0.389165702 | 0.9281 | 0.7132158279 | 0.198913 | 5.223046114 | 0 |
| 2 | 47416411 | A | G | K | R | MSH2 | ENST00000233146 | 0.47488 | 0.00615 | 0.56842 | 0.65297 | 0.74587 | 0.2928255692 | 0.96253 | 0.505762279 | 0.187941 | 3.695322267 | 0 |
| 2 | 47416411 | A | T | K | M | MSH2 | ENST00000233146 | 0.57765 | 0.01574 | 0.57942 | 0.85005 | 0.92151 | 0.454611717 | 0.9148 | 0.9644213319 | 0.253198 | 6.023785967 | 0 |
| 2 | 47416412 | G | C | K | N | MSH2 | ENST00000233146 | 0.42109 | 0.01197 | 0.69391 | 0.80265 | 0.8345 | 0.4570591603 | 0.86732 | 0.9514986277 | 0.12749 | 6.956618859 | 0 |
| 2 | 47416412 | G | T | K | N | MSH2 | ENST00000233146 | 0.41629 | 0.01197 | 0.69391 | 0.80239 | 0.8345 | 0.4570591603 | 0.86693 | 0.957619369 | 0.127491 | 6.956618859 | 0 |
| 2 | 47416413 | A | C | N | H | MSH2 | ENST00000233146 | 0.78107 | 0.00715 | 0.57895 | 0.88105 | 0.86951 | 0.1902671809 | 0.89931 | 0.9480932951 | 0.16247 | 5.572022292 | 0 |
| 2 | 47416413 | A | G | N | D | MSH2 | ENST00000233146 | 0.58796 | 0.01053 | 0.63819 | 0.79542 | 0.82443 | 0.1986717259 | 0.88224 | 0.9301264882 | 0.147532 | 3.903250699 | 0 |
| 2 | 47416413 | A | T | N | Y | MSH2 | ENST00000233146 | 0.92747 | 0.01529 | 0.62629 | 0.93861 | 0.95243 | 0.303254291 | 0.86918 | 0.9916125536 | 0.304141 | 5.917469029 | 0 |
| 2 | 47416414 | A | C | N | T | MSH2 | ENST00000233146 | 0.46432 | 0.00586 | 0.54613 | 0.76575 | 0.79516 | 0.1273315153 | 0.87624 | 0.7118239403 | 0.128727 | 3.770785925 | 0 |
| 2 | 47416414 | A | G | N | S | MSH2 | ENST00000233146 | 0.42217 | 0.00638 | 0.48864 | 0.68261 | 0.79577 | 0.1056824852 | 0.87773 | 0.7657416463 | 0.118133 | 3.693565209 | 0 |
| 2 | 47416414 | A | T | N | I | MSH2 | ENST00000233146 | 0.8356 | 0.00773 | 0.5939 | 0.90822 | 0.93123 | 0.3022067284 | 0.89027 | 0.963049829 | 0.250752 | 4.477086312 | 0 |
| 2 | 47416415 | C | A | N | K | MSH2 | ENST00000233146 | 0.2462 | 0.00608 | 0.66366 | 0.79584 | 0.81588 | 0.2281811937 | 0.86436 | 0.8069195151 | 0.11775 | 3.871222608 | 0 |
| 2 | 47416415 | C | G | N | K | MSH2 | ENST00000233146 | 0.25089 | 0.00608 | 0.66366 | 0.79576 | 0.81588 | 0.2281811937 | 0.86491 | 0.8194927573 | 0.11775 | 3.871222608 | 0 |
| 2 | 47416416 | A | G | R | G | MSH2 | ENST00000233146 | 0.70784 | 0.01351 | 0.56039 | 0.85851 | 0.93358 | 0.3838263965 | 0.96972 | 0.9849925041 | 0.377714 | 5.872761324 | 0 |
| 2 | 47416417 | G | A | R | K | MSH2 | ENST00000233146 | 0.43786 | 0.00574 | 0.63634 | 0.59536 | 0.70345 | 0.2706445365 | 0.9216 | 0.7720746398 | 0.156832 | 1.296928331 | 0 |
| 2 | 47416417 | G | C | R | T | MSH2 | ENST00000233146 | 0.52586 | 0.01115 | 0.6151 | 0.75084 | 0.89513 | 0.3519425821 | 0.92554 | 0.9760388732 | 0.327254 | 5.201063226 | 0 |
| 2 | 47416417 | G | T | R | I | MSH2 | ENST00000233146 | 0.5785 | 0.00492 | 0.59574 | 0.80711 | 0.84747 | 0.361956176 | 0.93968 | 0.9431647062 | 0.26214 | 6.106744899 | 0 |
| 2 | 47416418 | A | C | R | S | MSH2 | ENST00000233146 | 0.49183 | 0.00928 | 0.56559 | 0.85222 | 0.90494 | 0.3687654706 | 0.96555 | 0.9247114658 | 0.331794 | 4.728926928 | 0 |
| 2 | 47416418 | A | T | R | S | MSH2 | ENST00000233146 | 0.48646 | 0.00928 | 0.56559 | 0.85324 | 0.90494 | 0.3687654706 | 0.96574 | 0.9247114658 | 0.331177 | 4.728926928 | 0 |
| 2 | 47416419 | A | C | I | L | MSH2 | ENST00000233146 | 0.66699 | 0.00791 | 0.71224 | 0.91014 | 0.93004 | 0.6146315919 | 0.92725 | 0.9350223541 | 0.351268 | 5.677817053 | 0 |
| 2 | 47416419 | A | G | I | V | MSH2 | ENST00000233146 | 0.74269 | 0.00949 | 0.61357 | 0.89309 | 0.91484 | 0.4435554079 | 0.90305 | 0.9669064879 | 0.33307 | 5.925290636 | 0 |
| 2 | 47416419 | A | T | I | L | MSH2 | ENST00000233146 | 0.66035 | 0.00791 | 0.71224 | 0.91008 | 0.93004 | 0.6146315919 | 0.92738 | 0.9308875203 | 0.351268 | 5.677817053 | 0 |
| 2 | 47416420 | T | A | I | K | MSH2 | ENST00000233146 | 0.76267 | 0.02068 | 0.7842 | 0.9649 | 0.99876 | 0.8834752627 | 0.98595 | 0.9984189272 | 0.575934 | 9.396233439 | 1 |
| 2 | 47416420 | T | C | I | T | MSH2 | ENST00000233146 | 0.74802 | 0.02687 | 0.82441 | 0.9426 | 0.99443 | 0.7615348044 | 0.97355 | 0.9915178418 | 0.505971 | 9.396233439 | 0 |
| 2 | 47416420 | T | G | I | R | MSH2 | ENST00000233146 | 0.77601 | 0.00832 | 0.76405 | 0.95821 | 0.99843 | 0.8868237585 | 0.9841 | 0.9951235652 | 0.575939 | 9.396233439 | 1 |
| 2 | 47416421 | A | G | I | M | MSH2 | ENST00000233146 | 0.73445 | 0.03379 | 0.67093 | 0.95096 | 0.98034 | 0.6823436995 | 0.9648 | 0.9893866777 | 0.513144 | 6.82405082 | 0 |

| | | | | | | | | | | | | | | | | | | |
|---|---|---|---|---|---|---|---|---|---|---|---|---|---|---|---|---|---|---|
| 2 | 47416422 | G | A | E | K | MSH2 | ENST00000233146 | 0.58931 | 0.00763 | 0.76974 | 0.77731 | 0.91986 | 0.5380695703 | 0.97019 | 0.956561029 | 0.437025 | 5.525414903 | 0 |
| 2 | 47416422 | G | C | E | Q | MSH2 | ENST00000233146 | 0.57449 | 0.00983 | 0.6377 | 0.75708 | 0.91694 | 0.3915919792 | 0.95281 | 0.8918374777 | 0.357049 | 5.920098449 | 0 |
| 2 | 47416423 | A | C | E | A | MSH2 | ENST00000233146 | 0.56734 | 0.00638 | 0.65678 | 0.88161 | 0.96226 | 0.5064652666 | 0.95907 | 0.9084382653 | 0.447051 | 7.23397428 | 0 |
| 2 | 47416423 | A | G | E | G | MSH2 | ENST00000233146 | 0.89707 | 0.01909 | 0.59303 | 0.93574 | 0.9729 | 0.5207025607 | 0.93783 | 0.9972321391 | 0.47416 | 7.928495363 | 0 |
| 2 | 47416423 | A | T | E | V | MSH2 | ENST00000233146 | 0.85574 | 0.00923 | 0.62855 | 0.91684 | 0.97252 | 0.5043779909 | 0.95928 | 0.9886168838 | 0.445594 | 6.363178802 | 0 |
| 2 | 47416424 | G | C | E | D | MSH2 | ENST00000233146 | 0.27039 | 0.00726 | 0.64689 | 0.82054 | 0.89963 | 0.3688190123 | 0.93995 | 0.9708932638 | 0.294691 | 6.096775686 | 0 |
| 2 | 47416424 | G | T | E | D | MSH2 | ENST00000233146 | 0.2666 | 0.00726 | 0.64689 | 0.82445 | 0.89963 | 0.3688190123 | 0.94057 | 0.9708932638 | 0.294688 | 6.096775686 | 0 |
| 2 | 47416425 | G | A | E | K | MSH2 | ENST00000233146 | 0.79626 | 0.02802 | 0.83037 | 0.90155 | 0.91229 | 0.5465937993 | 0.96533 | 0.9893398881 | 0.426236 | 4.758702294 | 0 |
| 2 | 47416425 | G | C | E | Q | MSH2 | ENST00000233146 | 0.88285 | 0.03322 | 0.72153 | 0.85495 | 0.87877 | 0.4093194319 | 0.92311 | 0.9492339492 | 0.315191 | 5.243087345 | 0 |
| 2 | 47416426 | A | C | E | A | MSH2 | ENST00000233146 | 0.855 | 0.03053 | 0.7419 | 0.9386 | 0.96444 | 0.5068907915 | 0.91672 | 0.998167634 | 0.430002 | 5.228709448 | 0 |
| 2 | 47416426 | A | G | E | G | MSH2 | ENST00000233146 | 0.90655 | 0.03099 | 0.68771 | 0.93912 | 0.97735 | 0.5144634145 | 0.90368 | 0.9988401532 | 0.467362 | 7.010087917 | 0 |
| 2 | 47416426 | A | T | E | V | MSH2 | ENST00000233146 | 0.89394 | 0.03274 | 0.72234 | 0.94811 | 0.98257 | 0.5168935064 | 0.94604 | 0.9970554113 | 0.460488 | 8.268327726 | 0 |
| 2 | 47416427 | G | C | E | D | MSH2 | ENST00000233146 | 0.44832 | 0.01276 | 0.73981 | 0.82864 | 0.91442 | 0.354006447 | 0.95669 | 0.9094134569 | 0.238592 | 5.36672848 | 0 |
| 2 | 47416427 | G | T | E | D | MSH2 | ENST00000233146 | 0.44332 | 0.01276 | 0.73981 | 0.83097 | 0.91442 | 0.354006447 | 0.95703 | 0.9802688956 | 0.306015 | 5.36672848 | 0 |
| 2 | 47416428 | A | G | R | G | MSH2 | ENST00000233146 | 0.89823 | 0.02646 | 0.75524 | 0.95812 | 0.99906 | 0.8633471056 | 0.99363 | 0.9990000129 | 0.588397 | 10.19018264 | 1 |
| 2 | 47416429 | G | A | R | K | MSH2 | ENST00000233146 | 0.87147 | 0.02582 | 0.81743 | 0.889 | 0.97401 | 0.8267705357 | 0.96 | 0.9743282795 | 0.451881 | 10.19018264 | 0 |
| 2 | 47416429 | G | C | R | T | MSH2 | ENST00000233146 | 0.87623 | 0.02673 | 0.79632 | 0.95587 | 0.99138 | 0.8610198145 | 0.9865 | 0.9978203773 | 0.58814 | 10.19018264 | 1 |
| 2 | 47416429 | G | T | R | I | MSH2 | ENST00000233146 | 0.91016 | 0.02687 | 0.7857 | 0.94351 | 0.98834 | 0.8113724948 | 0.98093 | 0.9993977547 | 0.589237 | 10.19018264 | 1 |
| 2 | 47429742 | A | C | R | S | MSH2 | ENST00000233146 | 0.72292 | 0.00945 | 0.76495 | 0.96098 | 0.96298 | 0.865782031 | 0.99648 | 0.9974591136 | 0.515798 | 10.19018264 | 1 |
| 2 | 47429742 | A | T | R | S | MSH2 | ENST00000233146 | 0.71618 | 0.00945 | 0.76495 | 0.96079 | 0.96298 | 0.865782031 | 0.99653 | 0.9974591136 | 0.515795 | 10.19018264 | 1 |
| 2 | 47429743 | T | A | L | M | MSH2 | ENST00000233146 | 0.41499 | 0.03137 | 0.76229 | 0.91982 | 0.89918 | 0.3502882264 | 0.91705 | 0.9766398668 | 0.288074 | 9.105944894 | 0 |
| 2 | 47429743 | T | G | L | V | MSH2 | ENST00000233146 | 0.28101 | 0.01211 | 0.72854 | 0.86736 | 0.91058 | 0.4446972332 | 0.97343 | 0.7112305164 | 0.392464 | 8.37312619 | 0 |
| 2 | 47429744 | T | C | L | S | MSH2 | ENST00000233146 | 0.89149 | 0.02547 | 0.79417 | 0.94536 | 0.97661 | 0.6620086188 | 0.97619 | 0.9914137125 | 0.479451 | 7.481670984 | 0 |
| 2 | 47429744 | T | G | L | W | MSH2 | ENST00000233146 | 0.89971 | 0.02696 | 0.85081 | 0.96604 | 0.98034 | 0.7104636286 | 0.95334 | 0.9915225506 | 0.486096 | 9.595025842 | 0 |
| 2 | 47429745 | G | C | L | F | MSH2 | ENST00000233146 | 0.47778 | 0.02193 | 0.77297 | 0.91498 | 0.93044 | 0.4654785505 | 0.97112 | 0.7790603042 | 0.387914 | 6.996311848 | 0 |
| 2 | 47429745 | G | T | L | F | MSH2 | ENST00000233146 | 0.47263 | 0.02193 | 0.77297 | 0.91486 | 0.93083 | 0.4654785505 | 0.97203 | 0.7852822542 | 0.387901 | 6.996311848 | 0 |
| 2 | 47429746 | A | C | N | H | MSH2 | ENST00000233146 | 0.70623 | 0.02869 | 0.52785 | 0.90617 | 0.91399 | 0.2172115319 | 0.93427 | 0.9623648524 | 0.169821 | 5.827413127 | 0 |
| 2 | 47429746 | A | G | N | D | MSH2 | ENST00000233146 | 0.31546 | 0.00556 | 0.59137 | 0.49365 | 0.60952 | 0.1630922891 | 0.89291 | 0.4206269085 | 0.0780382 | 1.651016012 | 0 |
| 2 | 47429746 | A | T | N | Y | MSH2 | ENST00000233146 | 0.75591 | 0.02679 | 0.5775 | 0.94096 | 0.96008 | 0.3246172417 | 0.93767 | 0.9926198125 | 0.303019 | 7.330039924 | 0 |
| 2 | 47429747 | A | C | N | T | MSH2 | ENST00000233146 | 0.77188 | 0.01315 | 0.49842 | 0.8827 | 0.90842 | 0.1670970023 | 0.8406 | 0.9823547602 | 0.171003 | 5.086397928 | 0 |
| 2 | 47429747 | A | G | N | S | MSH2 | ENST00000233146 | 0.43865 | 0.00769 | 0.42972 | 0.81149 | 0.79577 | 0.1422946669 | 0.84927 | 0.864552021 | 0.130174 | 4.491573904 | 0 |
| 2 | 47429747 | A | T | N | I | MSH2 | ENST00000233146 | 0.83368 | 0.00599 | 0.52262 | 0.915 | 0.95681 | 0.2854043725 | 0.88096 | 0.9873422384 | 0.281406 | 7.627216981 | 0 |
| 2 | 47429748 | T | A | N | K | MSH2 | ENST00000233146 | 0.52194 | 0.00669 | 0.62427 | 0.86898 | 0.90755 | 0.261380734 | 0.90583 | 0.9728426337 | 0.137044 | 4.639865578 | 0 |
| 2 | 47429748 | T | G | N | K | MSH2 | ENST00000233146 | 0.52879 | 0.00669 | 0.62427 | 0.86868 | 0.90755 | 0.261380734 | 0.90589 | 0.973312676 | 0.137047 | 4.639865578 | 0 |
| 2 | 47429749 | T | A | L | I | MSH2 | ENST00000233146 | 0.285 | 0.00633 | 0.66333 | 0.75725 | 0.78223 | 0.175972558 | 0.91467 | 0.5412176251 | 0.206207 | 1.900383816 | 0 |
| 2 | 47429749 | T | G | L | V | MSH2 | ENST00000233146 | 0.30647 | 0.00696 | 0.57767 | 0.80587 | 0.85274 | 0.3168489874 | 0.97498 | 0.3806839287 | 0.32878 | 1.820592567 | 0 |
| 2 | 47429750 | T | C | L | S | MSH2 | ENST00000233146 | 0.86396 | 0.02965 | 0.67774 | 0.93397 | 0.98404 | 0.5842842531 | 0.97722 | 0.9985491633 | 0.491056 | 4.193477791 | 0 |
| 2 | 47429751 | A | C | L | F | MSH2 | ENST00000233146 | 0.51711 | 0.03083 | 0.6277 | 0.89881 | 0.93397 | 0.4251714487 | 0.97022 | 0.9670422673 | 0.379453 | 4.957613271 | 0 |
| 2 | 47429751 | A | T | L | F | MSH2 | ENST00000233146 | 0.51158 | 0.03083 | 0.6277 | 0.89881 | 0.93397 | 0.4251714487 | 0.9705 | 0.9670422673 | 0.379439 | 4.957613271 | 0 |
| 2 | 47429752 | G | A | V | M | MSH2 | ENST00000233146 | 0.87847 | 0.03547 | 0.53967 | 0.89593 | 0.97997 | 0.7085754958 | 0.96455 | 0.9987483025 | 0.511781 | 9.886071983 | 0 |
| 2 | 47429752 | G | C | V | L | MSH2 | ENST00000233146 | 0.84605 | 0.01236 | 0.49853 | 0.82896 | 0.99039 | 0.7107467611 | 0.98618 | 0.9914487004 | 0.558902 | 6.545595318 | 0 |
| 2 | 47429752 | G | T | V | L | MSH2 | ENST00000233146 | 0.83982 | 0.01236 | 0.49853 | 0.831 | 0.99039 | 0.7107467611 | 0.9863 | 0.9938243628 | 0.364368 | 6.545595318 | 0 |
| 2 | 47429753 | T | C | V | A | MSH2 | ENST00000233146 | 0.73942 | 0.03224 | 0.51315 | 0.95105 | 0.99202 | 0.7151372285 | 0.97389 | 0.9982603192 | 0.556255 | 8.754550938 | 0 |
| 2 | 47429753 | T | G | V | G | MSH2 | ENST00000233146 | 0.77481 | 0.0325 | 0.36926 | 0.97261 | 0.99552 | 0.833243817 | 0.98402 | 0.9990612864 | 0.560843 | 9.886071983 | 0 |
| 2 | 47429755 | G | C | E | Q | MSH2 | ENST00000233146 | 0.71081 | 0.0142 | 0.61282 | 0.79156 | 0.91945 | 0.4703003818 | 0.95639 | 0.968586266 | 0.400857 | 4.198522319 | 0 |
| 2 | 47429756 | A | C | E | A | MSH2 | ENST00000233146 | 0.83921 | 0.02883 | 0.63177 | 0.93035 | 0.98693 | 0.5666902555 | 0.96312 | 0.9965330362 | 0.462835 | 4.637484038 | 0 |
| 2 | 47429756 | A | G | E | G | MSH2 | ENST00000233146 | 0.73301 | 0.02927 | 0.57478 | 0.93722 | 0.97923 | 0.5595211101 | 0.95292 | 0.9955587387 | 0.482692 | 5.169921984 | 0 |
| 2 | 47429756 | A | T | E | V | MSH2 | ENST00000233146 | 0.89884 | 0.02809 | 0.6154 | 0.96078 | 0.98938 | 0.5808219121 | 0.98362 | 0.9981964231 | 0.520027 | 8.728279003 | 0 |
| 2 | 47429757 | A | C | E | D | MSH2 | ENST00000233146 | 0.44423 | 0.00715 | 0.63434 | 0.79014 | 0.86503 | 0.4304112998 | 0.98218 | 0.8146080971 | 0.287642 | 4.419736587 | 0 |
| 2 | 47429757 | A | T | E | D | MSH2 | ENST00000233146 | 0.43914 | 0.00715 | 0.63434 | 0.79014 | 0.86503 | 0.4304112998 | 0.98233 | 0.8343690038 | 0.287636 | 4.419736587 | 0 |
| 2 | 47429758 | G | A | A | T | MSH2 | ENST00000233146 | 0.44065 | 0.00586 | 0.47678 | 0.54952 | 0.74093 | 0.1297438012 | 0.87481 | 0.5162234735 | 0.195444 | 2.385918133 | 0 |
| 2 | 47429758 | G | C | A | P | MSH2 | ENST00000233146 | 0.65696 | 0.02143 | 0.55482 | 0.79443 | 0.9367 | 0.5745934522 | 0.911 | 0.9890649319 | 0.391071 | 7.739604095 | 0 |
| 2 | 47429758 | G | T | A | S | MSH2 | ENST00000233146 | 0.41275 | 0.00627 | 0.43689 | 0.5923 | 0.78035 | 0.1147635184 | 0.87757 | 0.4523436319 | 0.181412 | 2.044341551 | 0 |
| 2 | 47429759 | C | G | A | G | MSH2 | ENST00000233146 | 0.3617 | 0.00622 | 0.46241 | 0.70049 | 0.85843 | 0.1428792749 | 0.86963 | 0.9840919971 | 0.264865 | 4.689445613 | 0 |
| 2 | 47429759 | C | T | A | V | MSH2 | ENST00000233146 | 0.32162 | 0.00622 | 0.50675 | 0.64812 | 0.78223 | 0.193723628 | 0.84502 | 0.7069445848 | 0.226725 | 2.641000889 | 0 |
| 2 | 47429761 | T | A | F | I | MSH2 | ENST00000233146 | 0.73381 | 0.01475 | 0.76261 | 0.91658 | 0.97698 | 0.5821515055 | 0.95782 | 0.9951562881 | 0.471661 | 3.777469251 | 0 |
| 2 | 47429761 | T | C | F | L | MSH2 | ENST00000233146 | 0.53401 | 0.0073 | 0.79938 | 0.80679 | 0.92275 | 0.4995324734 | 0.93618 | 0.9277346134 | 0.372786 | 2.405684403 | 0 |
| 2 | 47429761 | T | G | F | V | MSH2 | ENST00000233146 | 0.88666 | 0.02186 | 0.75229 | 0.95077 | 0.98657 | 0.5110713333 | 0.95369 | 0.9968575239 | 0.488791 | 6.449322357 | 0 |
| 2 | 47429762 | T | A | F | Y | MSH2 | ENST00000233146 | 0.81716 | 0.03409 | 0.78582 | 0.92719 | 0.96773 | 0.4834023781 | 0.91829 | 0.9853919148 | 0.395239 | 7.707159544 | 0 |
| 2 | 47429762 | T | C | F | S | MSH2 | ENST00000233146 | 0.89388 | 0.02775 | 0.761 | 0.95502 | 0.98938 | 0.7116060874 | 0.9778 | 0.9985414743 | 0.507391 | 8.316569053 | 0 |
| 2 | 47429762 | T | G | F | C | MSH2 | ENST00000233146 | 0.89491 | 0.03124 | 0.78963 | 0.95488 | 0.98764 | 0.6201512291 | 0.96498 | 0.9983479977 | 0.511511 | 7.141124873 | 0 |
| 2 | 47429763 | T | A | F | L | MSH2 | ENST00000233146 | 0.38914 | 0.0073 | 0.79938 | 0.7892 | 0.85222 | 0.4995324734 | 0.95011 | 0.829595685 | 0.334625 | 2.405684403 | 0 |
| 2 | 47429763 | T | G | F | L | MSH2 | ENST00000233146 | 0.3951 | 0.0073 | 0.79938 | 0.79001 | 0.85222 | 0.4995324734 | 0.94962 | 0.6575289369 | 0.334626 | 2.405684403 | 0 |
| 2 | 47429764 | G | A | V | I | MSH2 | ENST00000233146 | 0.53314 | 0.00687 | 0.56961 | 0.64014 | 0.82892 | 0.1984124173 | 0.89444 | 0.3049256285 | 0.167076 | 3.369766971 | 0 |
| 2 | 47429764 | G | C | V | L | MSH2 | ENST00000233146 | 0.56082 | 0.00687 | 0.74635 | 0.67407 | 0.84535 | 0.3905266382 | 0.94769 | 0.7426790343 | 0.360728 | 2.998525304 | 0 |
| 2 | 47429764 | G | T | V | L | MSH2 | ENST00000233146 | 0.55518 | 0.00687 | 0.74635 | 0.67407 | 0.84535 | 0.3905266382 | 0.94834 | 0.7518487049 | 0.360726 | 2.998525304 | 0 |
| 2 | 47429765 | T | A | V | E | MSH2 | ENST00000233146 | 0.65743 | 0.02506 | 0.75492 | 0.93731 | 0.96737 | 0.6745776073 | 0.96939 | 0.9988712668 | 0.471121 | 7.0304153 | 0 |
| 2 | 47429765 | T | C | V | A | MSH2 | ENST00000233146 | 0.35206 | 0.00715 | 0.76281 | 0.80344 | 0.87247 | 0.4060472689 | 0.94868 | 0.3486561249 | 0.273647 | 4.625981596 | 0 |
| 2 | 47429765 | T | G | V | G | MSH2 | ENST00000233146 | 0.64679 | 0.02408 | 0.62616 | 0.93102 | 0.97178 | 0.5804673217 | 0.97001 | 0.9961853623 | 0.476797 | 6.516846979 | 0 |
| 2 | 47429767 | G | A | E | K | MSH2 | ENST00000233146 | 0.37182 | 0.00569 | 0.68721 | 0.62842 | 0.74164 | 0.1891367856 | 0.96678 | 0.6796734333 | 0.215176 | 3.112445909 | 0 |
| 2 | 47429767 | G | C | E | Q | MSH2 | ENST00000233146 | 0.35223 | 0.01075 | 0.54904 | 0.60536 | 0.74446 | 0.1905187777 | 0.93345 | 0.5734176636 | 0.20587 | 3.172460376 | 0 |
| 2 | 47429768 | A | C | E | A | MSH2 | ENST00000233146 | 0.41843 | 0.00726 | 0.57688 | 0.79851 | 0.84375 | 0.2412988476 | 0.92059 | 0.8444725871 | 0.212019 | 3.734472593 | 0 |
| 2 | 47429768 | A | G | E | G | MSH2 | ENST00000233146 | 0.47288 | 0.00547 | 0.51458 | 0.7852 | 0.81356 | 0.2286556471 | 0.96484 | 0.8539948463 | 0.203141 | 3.531178308 | 0 |
| 2 | 47429768 | A | T | E | V | MSH2 | ENST00000233146 | 0.4574 | 0.00547 | 0.53893 | 0.71907 | 0.79456 | 0.2829760572 | 0.96207 | 0.6665301919 | 0.158806 | 3.586450061 | 0 |
| 2 | 47429769 | A | C | E | D | MSH2 | ENST00000233146 | 0.33973 | 0.00592 | 0.57497 | 0.58245 | 0.77205 | 0.1495776024 | 0.97706 | 0.07687764048 | 0.180852 | 1.122739186 | 0 |
| 2 | 47429769 | A | T | E | D | MSH2 | ENST00000233146 | 0.33521 | 0.00592 | 0.57497 | 0.58512 | 0.77205 | 0.1495776024 | 0.97699 | 0.08121996532 | 0.180849 | 1.122739186 | 0 |
| 2 | 47429770 | G | A | D | N | MSH2 | ENST00000233146 | 0.48921 | 0.01151 | 0.70623 | 0.68847 | 0.72246 | 0.4192778453 | 0.81534 | 0.9653660059 | 0.0725106 | 3.069580426 | 0 |
| 2 | 47429770 | G | C | D | H | MSH2 | ENST00000233146 | 0.83614 | 0.03129 | 0.73122 | 0.85093 | 0.96883 | 0.5871217885 | 0.97333 | 0.9972834587 | 0.475526 | 4.937091858 | 0 |
| 2 | 47429770 | G | T | D | Y | MSH2 | ENST00000233146 | 0.85242 | 0.03076 | 0.71466 | 0.85009 | 0.97624 | 0.6849953122 | 0.97776 | 0.998016715 | 0.482584 | 7.570020448 | 0 |
| 2 | 47429771 | A | C | D | A | MSH2 | ENST00000233146 | 0.52334 | 0.01803 | 0.64269 | 0.93249 | 0.96993 | 0.595878569 | 0.9762 | 0.9908754826 | 0.466828 | 6.123513874 | 0 |
| 2 | 47429771 | A | G | D | G | MSH2 | ENST00000233146 | 0.81174 | 0.02119 | 0.64677 | 0.91556 | 0.96407 | 0.5950826583 | 0.98969 | 0.9951666594 | 0.475583 | 6.664386764 | 0 |
| 2 | 47429771 | A | T | D | V | MSH2 | ENST00000233146 | 0.82662 | 0.02043 | 0.64343 | 0.94998 | 0.9796 | 0.6408941389 | 0.98603 | 0.9980587363 | 0.497237 | 9.616423852 | 0 |
| 2 | 47429772 | T | A | D | E | MSH2 | ENST00000233146 | 0.41955 | 0.0199 | 0.67483 | 0.85284 | 0.91143 | 0.3233609377 | 0.98609 | 0.9694945812 | 0.345977 | 5.895765938 | 0 |
| 2 | 47429772 | T | G | D | E | MSH2 | ENST00000233146 | 0.42567 | 0.0199 | 0.67483 | 0.84194 | 0.91143 | 0.3233609377 | 0.98597 | 0.9727175236 | 0.345947 | 5.895765938 | 0 |
| 2 | 47429773 | G | A | A | T | MSH2 | ENST00000233146 | 0.34435 | 0.0058 | 0.38257 | 0.36792 | 0.71391 | 0.09428145951 | 0.93414 | 0.1512480229 | -0.018032 | 0.09723755136 | 0 |
| 2 | 47429773 | G | C | A | P | MSH2 | ENST00000233146 | 0.43744 | 0.00535 | 0.48173 | 0.57987 | 0.74021 | 0.4067430779 | 0.98385 | 0.7233910561 | 0.203398 | 0.7955242812 | 0 |
| 2 | 47429773 | G | T | A | S | MSH2 | ENST00000233146 | 0.30514 | 0.00586 | 0.36076 | 0.45185 | 0.67661 | 0.0893069625 | 0.88162 | 0.446089983 | 0.0777771 | 0.2844878146 | 0 |
| 2 | 47429774 | C | A | A | E | MSH2 | ENST00000233146 | 0.25417 | 0.00644 | 0.48148 | 0.55318 | 0.74235 | 0.2437573766 | 0.89459 | 0.8036850095 | 0.197955 | 3.841219096 | 0 |
| 2 | 47429774 | C | G | A | G | MSH2 | ENST00000233146 | 0.26192 | 0.00574 | 0.3866 | 0.4602 | 0.68381 | 0.1067214409 | 0.93836 | 0.7566257119 | 0.162246 | 2.058477352 | 0 |
| 2 | 47429774 | C | T | A | V | MSH2 | ENST00000233146 | 0.25597 | 0.0058 | 0.43733 | 0.45263 | 0.70994 | 0.1558856864 | 0.941 | 0.3872723723 | 0.19101 | 1.815488754 | 0 |
| 2 | 47429776 | G | A | E | K | MSH2 | ENST00000233146 | 0.79737 | 0.00735 | 0.79516 | 0.80021 | 0.89009 | 0.4030180682 | 0.99066 | 0.9783356786 | 0.44838 | 6.580347687 | 0 |
| 2 | 47429776 | G | C | E | Q | MSH2 | ENST00000233146 | 0.62339 | 0.01145 | 0.67183 | 0.69541 | 0.91694 | 0.3087061471 | 0.95889 | 0.7522021616 | 0.381265 | 6.029823392 | 0 |
| 2 | 47429777 | A | C | E | A | MSH2 | ENST00000233146 | 0.73877 | 0.01902 | 0.69549 | 0.89555 | 0.93004 | 0.3886774362 | 0.9677 | 0.9864131212 | 0.437464 | 5.43199465 | 0 |
| 2 | 47429777 | A | G | E | G | MSH2 | ENST00000233146 | 0.92588 | 0.02218 | 0.6276 | 0.91995 | 0.96371 | 0.3964265212 | 0.97018 | 0.9701288342 | 0.373643 | 4.779890711 | 0 |
| 2 | 47429777 | A | T | E | V | MSH2 | ENST00000233146 | 0.76235 | 0.00855 | 0.66611 | 0.88132 | 0.9211 | 0.4222100841 | 0.98755 | 0.9876137972 | 0.430327 | 4.517246941 | 0 |
| 2 | 47429778 | A | C | E | D | MSH2 | ENST00000233146 | 0.27971 | 0.00788 | 0.69244 | 0.7402 | 0.84482 | 0.2535335717 | 0.98809 | 0.7432305217 | 0.320009 | 4.565732562 | 0 |
| 2 | 47429778 | A | T | E | D | MSH2 | ENST00000233146 | 0.27569 | 0.00788 | 0.69244 | 0.73779 | 0.84482 | 0.2535335717 | 0.98821 | 0.7432305217 | 0.320005 | 4.565732562 | 0 |
| 2 | 47429779 | T | A | L | M | MSH2 | ENST00000233146 | 0.44655 | 0.03534 | 0.64643 | 0.8933 | 0.88823 | 0.3498484759 | 0.86386 | 0.956261158 | 0.245562 | 5.679509047 | 0 |
| 2 | 47429779 | T | G | L | V | MSH2 | ENST00000233146 | 0.29512 | 0.03587 | 0.61475 | 0.84179 | 0.85222 | 0.4073807319 | 0.93595 | 0.9304890037 | 0.26174 | 5.295628482 | 0 |
| 2 | 47429780 | T | C | L | S | MSH2 | ENST00000233146 | 0.89 | 0.03189 | 0.6797 | 0.93277 | 0.97997 | 0.6661975059 | 0.93996 | 0.9954218268 | 0.450827 | 6.228816427 | 0 |
| 2 | 47429780 | T | G | L | W | MSH2 | ENST00000233146 | 0.89757 | 0.02984 | 0.74322 | 0.94981 | 0.97773 | 0.7181917679 | 0.91727 | 0.9972387552 | 0.474229 | 8.234252031 | 0 |

| | | | | | | | | | | | | | | | | | |
|---|---|---|---|---|---|---|---|---|---|---|---|---|---|---|---|---|---|
| 2 | 47429781 | G | C | L | F | MSH2 | ENST00000233146 | 0.62817 | 0.0357 | 0.65287 | 0.87995 | 0.94318 | 0.4586117756 | 0.94136 | 0.9840330482 | 0.379025 | 6.365825016 | 0 |
| 2 | 47429781 | G | T | L | F | MSH2 | ENST00000233146 | 0.62203 | 0.0357 | 0.65287 | 0.88103 | 0.94318 | 0.4586117756 | 0.94172 | 0.9840330482 | 0.37902 | 6.365825016 | 0 |
| 2 | 47429782 | A | G | R | G | MSH2 | ENST00000233146 | 0.87617 | 0.02844 | 0.66201 | 0.96808 | 0.98404 | 0.7520814299 | 0.99175 | 0.9945828319 | 0.491481 | 8.371023249 | 0 |
| 2 | 47429782 | A | T | R | W | MSH2 | ENST00000233146 | 0.92667 | 0.0364 | 0.76348 | 0.96883 | 0.97773 | 0.7474441167 | 0.95711 | 0.9972444773 | 0.501219 | 9.337309951 | 0 |
| 2 | 47429783 | G | A | R | K | MSH2 | ENST00000233146 | 0.91333 | 0.02582 | 0.73829 | 0.8431 | 0.96664 | 0.7012011757 | 0.97745 | 0.9917590022 | 0.494456 | 5.882222937 | 0 |
| 2 | 47429783 | G | C | R | T | MSH2 | ENST00000233146 | 0.92155 | 0.0142 | 0.70966 | 0.89818 | 0.99006 | 0.7517867848 | 0.9779 | 0.9984394908 | 0.520944 | 6.702507188 | 0 |
| 2 | 47429784 | G | C | R | S | MSH2 | ENST00000233146 | 0.69243 | 0.02796 | 0.67926 | 0.90317 | 0.93554 | 0.7644211857 | 0.99035 | 0.9873260856 | 0.457154 | 6.426153043 | 0 |
| 2 | 47429784 | G | T | R | S | MSH2 | ENST00000233146 | 0.68595 | 0.02796 | 0.67926 | 0.9083 | 0.93554 | 0.7644211857 | 0.9907 | 0.9891324639 | 0.457145 | 6.426153043 | 0 |
| 2 | 47429785 | C | A | Q | K | MSH2 | ENST00000233146 | 0.4622 | 0.00535 | 0.62856 | 0.67618 | 0.78473 | 0.4749171762 | 0.92818 | 0.7270466685 | 0.209496 | 4.941814118 | 0 |
| 2 | 47429786 | A | C | Q | P | MSH2 | ENST00000233146 | 0.76792 | 0.01628 | 0.58584 | 0.91739 | 0.95645 | 0.7148398292 | 0.98006 | 0.9926453829 | 0.467154 | 8.147708658 | 0 |
| 2 | 47429786 | A | G | Q | R | MSH2 | ENST00000233146 | 0.45089 | 0.00574 | 0.62117 | 0.81902 | 0.80889 | 0.4806326375 | 0.97226 | 0.276063248 | -0.00736017 | 6.842013262 | 0 |
| 2 | 47429786 | A | T | Q | L | MSH2 | ENST00000233146 | 0.52107 | 0.01063 | 0.564 | 0.91879 | 0.90842 | 0.4671421741 | 0.94291 | 0.9875734448 | 0.312899 | 5.473088273 | 0 |
| 2 | 47429787 | G | C | Q | H | MSH2 | ENST00000233146 | 0.39568 | 0.02595 | 0.61751 | 0.86909 | 0.90096 | 0.424151676 | 0.94103 | 0.9392675757 | 0.189628 | 6.349786123 | 0 |
| 2 | 47429787 | G | T | Q | H | MSH2 | ENST00000233146 | 0.39098 | 0.02595 | 0.61751 | 0.87486 | 0.89963 | 0.424151676 | 0.94096 | 0.9884666204 | 0.233901 | 6.349786123 | 0 |
| 2 | 47429788 | A | C | T | P | MSH2 | ENST00000233146 | 0.32399 | 0.01108 | 0.26702 | 0.83437 | 0.83997 | 0.5027494834 | 0.961 | 0.9133605361 | 0.206717 | 5.705580982 | 0 |
| 2 | 47429788 | A | G | T | A | MSH2 | ENST00000233146 | 0.2504 | 0.0058 | 0.26126 | 0.57646 | 0.70262 | 0.1122110572 | 0.97163 | 0.1235943884 | 0.0934806 | 1.866875194 | 0 |
| 2 | 47429788 | A | T | T | S | MSH2 | ENST00000233146 | 0.18932 | 0.00592 | 0.24111 | 0.43722 | 0.64157 | 0.08857048758 | 0.85978 | 0.04372894066 | -0.00827873 | 0.02701984319 | 0 |
| 2 | 47429789 | C | A | T | N | MSH2 | ENST00000233146 | 0.15511 | 0.00551 | 0.30539 | 0.51484 | 0.68995 | 0.2286260669 | 0.90605 | 0.1262614727 | 0.050781 | 1.221605963 | 0 |
| 2 | 47429789 | C | G | T | S | MSH2 | ENST00000233146 | 0.11428 | 0.00592 | 0.24111 | 0.35474 | 0.66449 | 0.08857048758 | 0.77573 | 0.06891833246 | 0.0195683 | 0.02701984319 | 0 |
| 2 | 47429789 | C | T | T | I | MSH2 | ENST00000233146 | 0.18346 | 0.00566 | 0.33876 | 0.61398 | 0.72398 | 0.2057659982 | 0.95694 | 0.07178551704 | -0.040551 | 2.885575813 | 0 |
| 2 | 47429791 | T | A | L | I | MSH2 | ENST00000233146 | 0.38668 | 0.00633 | 0.62845 | 0.78099 | 0.79334 | 0.1254481336 | 0.8923 | 0.2978926301 | 0.122637 | 2.583164334 | 0 |
| 2 | 47429791 | T | G | L | V | MSH2 | ENST00000233146 | 0.3907 | 0.00648 | 0.55541 | 0.82041 | 0.86803 | 0.2447710349 | 0.95431 | 0.600448966 | 0.22734 | 3.35137763 | 0 |
| 2 | 47429792 | T | C | L | S | MSH2 | ENST00000233146 | 0.89993 | 0.02691 | 0.63448 | 0.94704 | 0.96993 | 0.562665051 | 0.90939 | 0.9865765572 | 0.237242 | 7.832088703 | 0 |
| 2 | 47429793 | A | C | L | F | MSH2 | ENST00000233146 | 0.71842 | 0.02595 | 0.6071 | 0.88939 | 0.92275 | 0.3675956074 | 0.92462 | 0.9586647749 | 0.246057 | 4.564628584 | 0 |
| 2 | 47429793 | A | T | L | F | MSH2 | ENST00000233146 | 0.71169 | 0.02595 | 0.6071 | 0.88949 | 0.92316 | 0.3675956074 | 0.92409 | 0.9586647749 | 0.246023 | 4.564628584 | 0 |
| 2 | 47429794 | C | A | Q | K | MSH2 | ENST00000233146 | 0.38209 | 0.00747 | 0.70382 | 0.70084 | 0.85996 | 0.5415192971 | 0.97959 | 0.9698809385 | 0.402701 | 4.328862223 | 0 |
| 2 | 47429794 | C | G | Q | E | MSH2 | ENST00000233146 | 0.36475 | 0.00941 | 0.65691 | 0.73709 | 0.9045 | 0.4832423036 | 0.951 | 0.6466322304 | 0.259665 | 4.998885811 | 0 |
| 2 | 47429795 | A | C | Q | P | MSH2 | ENST00000233146 | 0.63305 | 0.01893 | 0.66096 | 0.92103 | 0.97438 | 0.754222991 | 0.98352 | 0.9926453829 | 0.47817 | 7.175218826 | 0 |
| 2 | 47429795 | A | G | Q | R | MSH2 | ENST00000233146 | 0.43738 | 0.00586 | 0.69929 | 0.82336 | 0.83943 | 0.5632409694 | 0.99394 | 0.5793538814 | 0.333525 | 3.457774995 | 0 |
| 2 | 47429795 | A | T | Q | L | MSH2 | ENST00000233146 | 0.47357 | 0.00596 | 0.6431 | 0.87515 | 0.89739 | 0.5263889376 | 0.97209 | 0.9855325818 | 0.401261 | 4.901826479 | 0 |
| 2 | 47429796 | A | C | Q | H | MSH2 | ENST00000233146 | 0.32504 | 0.00608 | 0.69777 | 0.88388 | 0.91652 | 0.5031951416 | 0.97411 | 0.9700712562 | 0.280044 | 4.267459979 | 0 |
| 2 | 47429796 | A | T | Q | H | MSH2 | ENST00000233146 | 0.32062 | 0.00608 | 0.69777 | 0.88457 | 0.91652 | 0.5031951416 | 0.97433 | 0.969694376 | 0.280037 | 4.267459979 | 0 |
| 2 | 47429797 | G | A | E | K | MSH2 | ENST00000233146 | 0.72588 | 0.02943 | 0.81048 | 0.84364 | 0.94876 | 0.3503557871 | 0.97575 | 0.9939892292 | 0.435141 | 5.3604026 | 0 |
| 2 | 47429797 | G | C | E | Q | MSH2 | ENST00000233146 | 0.6313 | 0.03426 | 0.70343 | 0.61069 | 0.8124 | 0.2802592524 | 0.94589 | 0.9829685092 | 0.298449 | 4.263664458 | 0 |
| 2 | 47429798 | A | C | E | A | MSH2 | ENST00000233146 | 0.7076 | 0.02349 | 0.72084 | 0.88548 | 0.93319 | 0.3672705697 | 0.9404 | 0.9903484583 | 0.307505 | 5.048943956 | 0 |
| 2 | 47429798 | A | G | E | G | MSH2 | ENST00000233146 | 0.59711 | 0.00722 | 0.67557 | 0.86119 | 0.86251 | 0.3661880149 | 0.89088 | 0.9818088412 | 0.264132 | 5.822443511 | 0 |
| 2 | 47429798 | A | T | E | V | MSH2 | ENST00000233146 | 0.87617 | 0.03287 | 0.70713 | 0.93051 | 0.96117 | 0.4007851776 | 0.95006 | 0.9980086684 | 0.398661 | 6.692898829 | 0 |
| 2 | 47429799 | A | C | E | D | MSH2 | ENST00000233146 | 0.37171 | 0.00696 | 0.72001 | 0.65447 | 0.79577 | 0.2140673706 | 0.97273 | 0.3235140145 | 0.212624 | 2.491947366 | 0 |
| 2 | 47429799 | A | T | E | D | MSH2 | ENST00000233146 | 0.36699 | 0.00696 | 0.72001 | 0.65447 | 0.79577 | 0.2140673706 | 0.97276 | 0.3235140145 | 0.212622 | 2.491947366 | 0 |
| 2 | 47429800 | G | C | D | H | MSH2 | ENST00000233146 | 0.88367 | 0.02989 | 0.69983 | 0.81154 | 0.97141 | 0.5619558703 | 0.96718 | 0.9963201284 | 0.456278 | 8.397367131 | 0 |
| 2 | 47429800 | G | T | D | Y | MSH2 | ENST00000233146 | 0.89 | 0.02934 | 0.68498 | 0.79618 | 0.9755 | 0.6339478639 | 0.97789 | 0.9873971343 | 0.430333 | 8.397367131 | 0 |
| 2 | 47429801 | A | C | D | A | MSH2 | ENST00000233146 | 0.84507 | 0.01562 | 0.60415 | 0.90586 | 0.95535 | 0.5690992364 | 0.97172 | 0.9921632409 | 0.45819 | 3.643246371 | 0 |
| 2 | 47429801 | A | G | D | G | MSH2 | ENST00000233146 | 0.85931 | 0.01533 | 0.61923 | 0.91589 | 0.96883 | 0.5513521737 | 0.98578 | 0.9923106432 | 0.460408 | 5.050330752 | 0 |
| 2 | 47429801 | A | T | D | V | MSH2 | ENST00000233146 | 0.69444 | 0.02005 | 0.60543 | 0.93624 | 0.97624 | 0.626434539 | 0.98407 | 0.9969046712 | 0.47629 | 8.397367131 | 0 |
| 2 | 47429802 | T | A | D | E | MSH2 | ENST00000233146 | 0.31483 | 0.00615 | 0.63823 | 0.37489 | 0.80178 | 0.3146273477 | 0.98333 | 0.2093174905 | -0.094935 | 1.723683825 | 0 |
| 2 | 47429802 | T | G | D | E | MSH2 | ENST00000233146 | 0.32014 | 0.00615 | 0.63823 | 0.40197 | 0.80178 | 0.3146273477 | 0.98327 | 0.2093174905 | -0.102797 | 1.723683825 | 0 |
| 2 | 47429803 | T | A | L | I | MSH2 | ENST00000233146 | 0.42591 | 0.00592 | 0.76493 | 0.86822 | 0.76024 | 0.1991719449 | 0.89665 | 0.4858054221 | 0.019262 | 3.041560144 | 0 |
| 2 | 47429803 | T | G | L | V | MSH2 | ENST00000233146 | 0.4104 | 0.00608 | 0.71029 | 0.50054 | 0.75485 | 0.3399357273 | 0.9041 | 0.3270928562 | -0.063913 | 3.425343342 | 0 |
| 2 | 47429804 | T | C | L | S | MSH2 | ENST00000233146 | 0.48208 | 0.00547 | 0.7814 | 0.38109 | 0.83888 | 0.596875165 | 0.94992 | 0.6928948706 | 0.0712269 | 7.460666095 | 0 |
| 2 | 47429805 | A | C | L | F | MSH2 | ENST00000233146 | 0.34506 | 0.01108 | 0.75405 | 0.15012 | 0.82273 | 0.3904951026 | 0.84496 | 0.2134543822 | 0.199493 | 4.551853717 | 0 |
| 2 | 47429805 | A | T | L | F | MSH2 | ENST00000233146 | 0.34049 | 0.01108 | 0.75405 | 0.15101 | 0.82273 | 0.3904951026 | 0.84491 | 0.2241738552 | 0.199488 | 4.551853717 | 0 |
| 2 | 47429806 | C | A | L | I | MSH2 | ENST00000233146 | 0.65226 | 0.03527 | 0.80746 | 0.87695 | 0.95972 | 0.3404183745 | 0.89712 | 0.9904977679 | 0.400772 | 5.974422092 | 0 |
| 2 | 47429806 | C | G | L | V | MSH2 | ENST00000233146 | 0.64937 | 0.03633 | 0.762 | 0.61206 | 0.97141 | 0.5408791368 | 0.93401 | 0.9944280386 | 0.47389 | 8.12454041 | 0 |
| 2 | 47429806 | C | T | L | F | MSH2 | ENST00000233146 | 0.69115 | 0.03582 | 0.80377 | 0.92009 | 0.9781 | 0.5855286589 | 0.95114 | 0.9932328463 | 0.531045 | 6.040235915 | 0 |
| 2 | 47429807 | T | A | L | H | MSH2 | ENST00000233146 | 0.75503 | 0.0334 | 0.88031 | 0.97491 | 0.99876 | 0.7529430009 | 0.97516 | 0.9985741377 | 0.479302 | 8.765826786 | 0 |
| 2 | 47429807 | T | C | L | P | MSH2 | ENST00000233146 | 0.77964 | 0.03001 | 0.90872 | 0.96917 | 0.99906 | 0.8906848123 | 0.97981 | 0.9990832806 | 0.472276 | 8.765826786 | 0 |
| 2 | 47429807 | T | G | L | R | MSH2 | ENST00000233146 | 0.77497 | 0.03137 | 0.8878 | 0.97018 | 0.99676 | 0.8656304946 | 0.97828 | 0.998883307 | 0.479302 | 8.765826786 | 0 |
| 2 | 47429809 | C | A | R | S | MSH2 | ENST00000233146 | 0.8434 | 0.0203 | 0.75345 | 0.88789 | 0.9328 | 0.5626119912 | 0.94829 | 0.9955272079 | 0.619807 | 4.403625019 | 0 |
| 2 | 47429809 | C | G | R | G | MSH2 | ENST00000233146 | 0.87355 | 0.02294 | 0.74083 | 0.87849 | 0.9458 | 0.5407321172 | 0.97422 | 0.989364028 | 0.593777 | 5.273786542 | 0 |
| 2 | 47429809 | C | T | R | C | MSH2 | ENST00000233146 | 0.93813 | 0.02955 | 0.83469 | 0.8463 | 0.94318 | 0.5164999164 | 0.94124 | 0.8917925954 | 0.325276 | 8.877379535 | 0 |
| 2 | 47429810 | G | C | R | P | MSH2 | ENST00000233146 | 0.78709 | 0.02732 | 0.7895 | 0.64931 | 0.96773 | 0.7564254339 | 0.97188 | 0.9929972291 | 0.45481 | 8.877379535 | 0 |
| 2 | 47429810 | G | T | R | L | MSH2 | ENST00000233146 | 0.5679 | 0.02324 | 0.7751 | 0.80612 | 0.92275 | 0.5754400128 | 0.9637 | 0.9852579236 | 0.418486 | 8.877379535 | 0 |
| 2 | 47429812 | C | G | R | G | MSH2 | ENST00000233146 | 0.61343 | 0.00633 | 0.79092 | 0.8259 | 0.86149 | 0.5952793775 | 0.98079 | 0.9809906483 | 0.393907 | 4.202531668 | 0 |
| 2 | 47429813 | G | A | R | Q | MSH2 | ENST00000233146 | 0.66771 | 0.01803 | 0.73455 | 0.74849 | 0.92682 | 0.5233418346 | 0.93458 | 0.683195138 | 0.42892 | 4.350902849 | 0 |
| 2 | 47429813 | G | C | R | P | MSH2 | ENST00000233146 | 0.83352 | 0.02646 | 0.83664 | 0.85235 | 0.97327 | 0.7923721992 | 0.94548 | 0.9971165657 | 0.602235 | 5.510110899 | 0 |
| 2 | 47429813 | G | T | R | L | MSH2 | ENST00000233146 | 0.7737 | 0.01799 | 0.82066 | 0.76837 | 0.94949 | 0.6263127204 | 0.97225 | 0.9892328978 | 0.473163 | 6.941815199 | 0 |
| 2 | 47429815 | T | A | F | I | MSH2 | ENST00000233146 | 0.3509 | 0.00569 | 0.86921 | 0.41071 | 0.77972 | 0.528259347 | 0.95671 | 0.7487897277 | 0.0133355 | 2.503847005 | 0 |
| 2 | 47429815 | T | G | F | V | MSH2 | ENST00000233146 | 0.38628 | 0.0054 | 0.84555 | 0.5259 | 0.78286 | 0.5458913251 | 0.96492 | 0.7716366649 | 0.0511883 | 1.954737828 | 0 |
| 2 | 47429816 | T | A | F | Y | MSH2 | ENST00000233146 | 0.80116 | 0.01053 | 0.88179 | 0.76029 | 0.94091 | 0.4779061517 | 0.91869 | 0.9060040116 | 0.318676 | 7.527785398 | 0 |
| 2 | 47429816 | T | C | F | S | MSH2 | ENST00000233146 | 0.87329 | 0.00889 | 0.86849 | 0.65717 | 0.98183 | 0.6990762225 | 0.98085 | 0.9840855598 | 0.457277 | 6.445277599 | 0 |
| 2 | 47429816 | T | G | F | C | MSH2 | ENST00000233146 | 0.89532 | 0.02469 | 0.88958 | 0.54705 | 0.97364 | 0.6486014151 | 0.9802 | 0.990267694 | 0.442667 | 6.515841534 | 0 |
| 2 | 47429818 | C | A | P | T | MSH2 | ENST00000233146 | 0.68299 | 0.03313 | 0.82545 | 0.88916 | 0.96627 | 0.6616996932 | 0.96824 | 0.9988242984 | 0.499991 | 7.407787067 | 0 |
| 2 | 47429818 | C | G | P | A | MSH2 | ENST00000233146 | 0.65516 | 0.03261 | 0.77651 | 0.85995 | 0.9528 | 0.5773773908 | 0.94229 | 0.9936881661 | 0.327704 | 7.047799945 | 0 |
| 2 | 47429818 | C | T | P | S | MSH2 | ENST00000233146 | 0.71137 | 0.03353 | 0.79046 | 0.82376 | 0.93319 | 0.6720829431 | 0.94231 | 0.9962717295 | 0.451404 | 5.462887549 | 0 |
| 2 | 47429819 | C | A | P | Q | MSH2 | ENST00000233146 | 0.58602 | 0.01881 | 0.79858 | 0.67348 | 0.94802 | 0.6743152553 | 0.96359 | 0.997433722 | 0.475307 | 5.53949761 | 0 |
| 2 | 47429819 | C | G | P | R | MSH2 | ENST00000233146 | 0.63466 | 0.0337 | 0.85942 | 0.64366 | 0.9648 | 0.7441564199 | 0.96991 | 0.9988242984 | 0.50746 | 5.522587485 | 0 |
| 2 | 47429819 | C | T | P | L | MSH2 | ENST00000233146 | 0.4935 | 0.0334 | 0.86155 | 0.74502 | 0.9367 | 0.6916279657 | 0.97726 | 0.9994136095 | 0.486588 | 6.198430169 | 0 |
| 2 | 47429821 | G | A | D | N | MSH2 | ENST00000233146 | 0.88952 | 0.03713 | 0.85172 | 0.92582 | 0.97512 | 0.717317197 | 0.95306 | 0.9974288344 | 0.28026 | 10.19769288 | 0 |
| 2 | 47429821 | G | C | D | H | MSH2 | ENST00000233146 | 0.88807 | 0.03436 | 0.87255 | 0.71698 | 0.99906 | 0.789763139 | 0.98246 | 0.9958902529 | 0.58563 | 10.19769288 | 0 |
| 2 | 47429821 | G | T | D | Y | MSH2 | ENST00000233146 | 0.89423 | 0.0337 | 0.85647 | 0.70452 | 0.99786 | 0.8537938635 | 0.99284 | 0.9989053011 | 0.584967 | 10.19769288 | 0 |
| 2 | 47429822 | A | C | D | A | MSH2 | ENST00000233146 | 0.85301 | 0.0325 | 0.78642 | 0.77479 | 0.99652 | 0.8124960735 | 0.98342 | 0.9987957478 | 0.582091 | 10.19769288 | 0 |
| 2 | 47429822 | A | G | D | G | MSH2 | ENST00000233146 | 0.86169 | 0.03261 | 0.79983 | 0.77691 | 0.99676 | 0.7953747116 | 0.99081 | 0.971873058 | 0.524621 | 10.19769288 | 0 |
| 2 | 47429822 | A | T | D | V | MSH2 | ENST00000233146 | 0.84883 | 0.03088 | 0.7847 | 0.83943 | 0.99744 | 0.8243952251 | 0.99352 | 0.9991446733 | 0.583261 | 10.19769288 | 0 |
| 2 | 47429823 | T | A | D | E | MSH2 | ENST00000233146 | 0.60816 | 0.03728 | 0.82671 | 0.62819 | 0.97698 | 0.6457193717 | 0.98692 | 0.9969444871 | 0.507163 | 10.19769288 | 0 |
| 2 | 47429823 | T | G | D | E | MSH2 | ENST00000233146 | 0.61581 | 0.03728 | 0.82671 | 0.62104 | 0.97698 | 0.6457193717 | 0.98716 | 0.9969444871 | 0.507165 | 10.19769288 | 0 |
| 2 | 47429824 | C | A | L | I | MSH2 | ENST00000233146 | 0.39081 | 0.01888 | 0.79785 | 0.78728 | 0.84052 | 0.2699885808 | 0.79331 | 0.9047791362 | 0.167592 | 1.970001145 | 0 |
| 2 | 47429824 | C | G | L | V | MSH2 | ENST00000233146 | 0.63099 | 0.01699 | 0.73713 | 0.80493 | 0.92151 | 0.4556178976 | 0.92433 | 0.8898431063 | 0.432968 | 4.480251962 | 0 |
| 2 | 47429824 | C | T | L | F | MSH2 | ENST00000233146 | 0.37177 | 0.00603 | 0.78355 | 0.76213 | 0.77011 | 0.539181485 | 0.88295 | 0.09896976653 | 0.0142387 | 0.5353733849 | 0 |
| 2 | 47429825 | T | A | L | H | MSH2 | ENST00000233146 | 0.76887 | 0.02595 | 0.87421 | 0.66485 | 0.98477 | 0.7132871038 | 0.9642 | 0.9989515543 | 0.514586 | 7.028178829 | 0 |
| 2 | 47429825 | T | C | L | P | MSH2 | ENST00000233146 | 0.80068 | 0.02802 | 0.90539 | 0.96744 | 0.98728 | 0.8809398775 | 0.97754 | 0.9977787137 | 0.511321 | 7.028178829 | 0 |
| 2 | 47429825 | T | G | L | R | MSH2 | ENST00000233146 | 0.79255 | 0.02741 | 0.88162 | 0.81386 | 0.98294 | 0.8549328658 | 0.98601 | 0.998225987 | 0.514762 | 7.028178829 | 1 |
| 2 | 47429827 | A | C | N | H | MSH2 | ENST00000233146 | 0.43683 | 0.00569 | 0.67525 | 0.24296 | 0.67752 | 0.2301452461 | 0.89518 | 0.7513012886 | -0.0986769 | 2.708055792 | 0 |
| 2 | 47429827 | A | G | N | D | MSH2 | ENST00000233146 | 0.49795 | 0.00556 | 0.73087 | 0.6307 | 0.62213 | 0.2538497822 | 0.90487 | 0.8273188472 | 0.0963092 | 4.081240222 | 0 |
| 2 | 47429827 | A | T | N | Y | MSH2 | ENST00000233146 | 0.47319 | 0.00872 | 0.72631 | 0.52487 | 0.84321 | 0.3611641212 | 0.95311 | 0.8928705454 | -0.0669942 | 5.509031572 | 0 |
| 2 | 47429828 | A | C | N | T | MSH2 | ENST00000233146 | 0.41098 | 0.00592 | 0.6441 | 0.14922 | 0.75005 | 0.1635679819 | 0.88436 | 0.7946441174 | 0.138842 | 5.509031572 | 0 |
| 2 | 47429828 | A | G | N | S | MSH2 | ENST00000233146 | 0.36236 | 0.00622 | 0.57406 | 0.13648 | 0.57356 | 0.1423435541 | 0.82524 | 0.5002197027 | 0.0975014 | 1.96642593 | 0 |
| 2 | 47429828 | A | T | N | I | MSH2 | ENST00000233146 | 0.46283 | 0.01229 | 0.67553 | 0.49712 | 0.86753 | 0.3249728786 | 0.86771 | 0.97522223 | 0.241074 | 5.509031572 | 0 |
| 2 | 47429829 | C | A | N | K | MSH2 | ENST00000233146 | 0.35267 | 0.00705 | 0.76495 | 0.20851 | 0.67478 | 0.275973913 | 0.8336 | 0.751224637 | 0.0983446 | 3.833397941 | 0 |
| 2 | 47429829 | C | G | N | K | MSH2 | ENST00000233146 | 0.35853 | 0.00705 | 0.76495 | 0.16812 | 0.67478 | 0.275973913 | 0.83375 | 0.61657691 | 0.0983476 | 3.833397941 | 0 |
| 2 | 47429830 | C | G | R | G | MSH2 | ENST00000233146 | 0.84203 | 0.03274 | 0.67477 | 0.95087 | 0.97923 | 0.7100949918 | 0.98003 | 0.9920344949 | 0.499479 | 8.918245968 | 0 |
| 2 | 47429831 | G | A | R | Q | MSH2 | ENST00000233146 | 0.43701 | 0.03419 | 0.59138 | 0.87358 | 0.9659 | 0.6246858104 | 0.96732 | 0.9822849035 | 0.290318 | 8.918245968 | 0 |

| | | | | | | | | | | | | | | | | | | |
|---|---|---|---|---|---|---|---|---|---|---|---|---|---|---|---|---|---|---|
| 2 | 47429831 | G | C | R | P | MSH2 | ENST00000233146 | 0.76475 | 0.03137 | 0.7402 | 0.95699 | 0.98972 | 0.852137332 | 0.97036 | 0.9966783524 | 0.546084 | 8.918245968 | 0 |
| 2 | 47429831 | G | T | R | L | MSH2 | ENST00000233146 | 0.54793 | 0.0296 | 0.71959 | 0.95058 | 0.97885 | 0.7073065776 | 0.97162 | 0.9978511333 | 0.547798 | 8.918245968 | 0 |
| 2 | 47429833 | C | A | L | I | MSH2 | ENST00000233146 | 0.41848 | 0.00681 | 0.66677 | 0.5072 | 0.80713 | 0.3104573683 | 0.8809 | 0.7033234239 | 0.103834 | 2.38868754 | 0 |
| 2 | 47429833 | C | G | L | V | MSH2 | ENST00000233146 | 0.5525 | 0.0129 | 0.58824 | 0.80165 | 0.91399 | 0.5050164023 | 0.98784 | 0.9567139745 | 0.374621 | 5.612240409 | 0 |
| 2 | 47429833 | C | T | L | F | MSH2 | ENST00000233146 | 0.58595 | 0.00855 | 0.65623 | | 0.88543 | 0.5789064713 | 0.9106 | 0.1131552989 | -0.116395 | 7.93827828 | 0 |
| 2 | 47429834 | T | A | L | H | MSH2 | ENST00000233146 | 0.89884 | 0.032 | 0.75118 | 0.85662 | 0.96444 | 0.7393438702 | 0.92323 | 0.9982823133 | 0.437118 | 8.200034636 | 0 |
| 2 | 47429834 | T | C | L | P | MSH2 | ENST00000233146 | 0.91428 | 0.02875 | 0.79773 | 0.95911 | 0.98404 | 0.8970299955 | 0.96699 | 0.9981815815 | 0.511205 | 8.200034636 | 0 |
| 2 | 47429834 | T | G | L | R | MSH2 | ENST00000233146 | 0.91073 | 0.02883 | 0.75555 | 0.83452 | 0.98294 | 0.8753976558 | 0.97455 | 0.9979404807 | 0.508754 | 8.200034636 | 0 |
| 2 | 47429836 | G | A | A | T | MSH2 | ENST00000233146 | 0.6385 | 0.01965 | 0.67949 | 0.73407 | 0.85012 | 0.4335635309 | 0.98581 | 0.9489936233 | 0.401192 | 4.471352431 | 0 |
| 2 | 47429836 | G | C | A | P | MSH2 | ENST00000233146 | 0.85669 | 0.02914 | 0.72982 | 0.54776 | 0.97848 | 0.8067150759 | 0.98454 | 0.991430223 | 0.473853 | 8.81275069 | 0 |
| 2 | 47429836 | G | T | A | S | MSH2 | ENST00000233146 | 0.44801 | 0.00741 | 0.6224 | 0.31926 | 0.79516 | 0.3715240842 | 0.95295 | 0.6453558803 | 0.280305 | 2.847277602 | 0 |
| 2 | 47429837 | C | A | A | D | MSH2 | ENST00000233146 | 0.62339 | 0.02001 | 0.7858 | 0.74664 | 0.96189 | 0.8098894095 | 0.97744 | 0.9940475225 | 0.468051 | 8.81275069 | 0 |
| 2 | 47429837 | C | G | A | G | MSH2 | ENST00000233146 | 0.48767 | 0.01699 | 0.65129 | 0.45263 | 0.93319 | 0.4209099057 | 0.95863 | 0.9738465548 | 0.403529 | 5.927009931 | 0 |
| 2 | 47429837 | C | T | A | V | MSH2 | ENST00000233146 | 0.47426 | 0.02857 | 0.69401 | 0.61422 | 0.9211 | 0.4826227034 | 0.96326 | 0.9740584493 | 0.421001 | 4.612572028 | 0 |
| 2 | 47429839 | A | C | K | Q | MSH2 | ENST00000233146 | 0.82709 | 0.03313 | 0.66285 | 0.91003 | 0.97178 | 0.4930813772 | 0.95603 | 0.9908285141 | 0.360496 | 8.442280716 | 0 |
| 2 | 47429840 | A | C | K | T | MSH2 | ENST00000233146 | 0.83875 | 0.02943 | 0.72138 | 0.93183 | 0.97364 | 0.5434656813 | 0.9512 | 0.9973276854 | 0.466549 | 4.403199654 | 0 |
| 2 | 47429840 | A | G | K | R | MSH2 | ENST00000233146 | 0.5242 | 0.00881 | 0.63209 | 0.69254 | 0.85894 | 0.4010419247 | 0.97786 | 0.8867269158 | 0.296379 | 6.273158276 | 0 |
| 2 | 47429840 | A | T | K | M | MSH2 | ENST00000233146 | 0.87315 | 0.03189 | 0.63338 | 0.9205 | 0.98404 | 0.6311052057 | 0.9208 | 0.982399404 | 0.423483 | 6.880522665 | 0 |
| 2 | 47429841 | G | C | K | N | MSH2 | ENST00000233146 | 0.73349 | 0.00935 | 0.74427 | 0.70884 | 0.88823 | 0.5818077804 | 0.9005 | 0.9934138656 | 0.171615 | 6.225356915 | 0 |
| 2 | 47429841 | G | T | K | N | MSH2 | ENST00000233146 | 0.727 | 0.00935 | 0.74427 | 0.70884 | 0.88823 | 0.5818077804 | 0.90085 | 0.9934138656 | 0.171612 | 6.225356915 | 0 |
| 2 | 47429842 | A | C | K | Q | MSH2 | ENST00000233146 | 0.82794 | 0.03404 | 0.71676 | 0.72621 | 0.98034 | 0.6678768009 | 0.96752 | 0.9833825231 | 0.418066 | 9.413260012 | 0 |
| 2 | 47429843 | A | C | K | T | MSH2 | ENST00000233146 | 0.80684 | 0.03353 | 0.76352 | 0.66974 | 0.97735 | 0.6918227432 | 0.96586 | 0.9959773421 | 0.493729 | 9.413260012 | 0 |
| 2 | 47429843 | A | G | K | R | MSH2 | ENST00000233146 | 0.49421 | 0.01621 | 0.68497 | 0.48513 | 0.87345 | 0.5899039782 | 0.95564 | 0.929998219 | 0.277297 | 2.696532294 | 0 |
| 2 | 47429843 | A | T | K | M | MSH2 | ENST00000233146 | 0.84165 | 0.0325 | 0.68455 | 0.79371 | 0.98972 | 0.7483241408 | 0.98192 | 0.9977787137 | 0.512386 | 9.413260012 | 0 |
| 2 | 47429844 | G | C | K | N | MSH2 | ENST00000233146 | 0.57547 | 0.03334 | 0.78416 | 0.61833 | 0.94949 | 0.7174466172 | 0.95273 | 0.9956625104 | 0.211707 | 9.413260012 | 0 |
| 2 | 47429844 | G | T | K | N | MSH2 | ENST00000233146 | 0.56978 | 0.03334 | 0.78416 | 0.58453 | 0.94949 | 0.7174466172 | 0.95292 | 0.9963967204 | 0.211701 | 9.413260012 | 0 |
| 2 | 47429845 | T | A | F | I | MSH2 | ENST00000233146 | 0.60412 | 0.02063 | 0.81025 | 0.72844 | 0.90362 | 0.5941807519 | 0.91156 | 0.9920457602 | 0.317488 | 2.879294788 | 0 |
| 2 | 47429845 | T | C | F | L | MSH2 | ENST00000233146 | 0.45524 | 0.00586 | 0.84278 | 0.68803 | 0.8543 | 0.5308301513 | 0.92629 | 0.9207078815 | 0.291244 | 3.312606555 | 0 |
| 2 | 47429845 | T | G | F | V | MSH2 | ENST00000233146 | 0.51586 | 0.02119 | 0.79198 | 0.58095 | 0.94243 | 0.534638159 | 0.96312 | 0.9929868579 | 0.442708 | 3.608351089 | 0 |
| 2 | 47429846 | T | A | F | Y | MSH2 | ENST00000233146 | 0.50608 | 0.0073 | 0.82456 | 0.93506 | 0.94281 | 0.5076942488 | 0.95961 | 0.9468365312 | 0.356066 | 8.213178888 | 0 |
| 2 | 47429846 | T | C | F | S | MSH2 | ENST00000233146 | 0.89113 | 0.02094 | 0.8109 | 0.87373 | 0.9822 | 0.7326049181 | 0.97859 | 0.9984336495 | 0.477455 | 8.213178888 | 0 |
| 2 | 47429846 | T | G | F | C | MSH2 | ENST00000233146 | 0.90675 | 0.02965 | 0.83805 | 0.68395 | 0.97512 | 0.6420822625 | 0.9483 | 0.9972562194 | 0.48207 | 8.213178888 | 0 |
| 2 | 47429847 | T | A | F | L | MSH2 | ENST00000233146 | 0.33228 | 0.00586 | 0.84278 | 0.24947 | 0.77909 | 0.5308301513 | 0.95553 | 0.760139823 | 0.262084 | 3.312606555 | 0 |
| 2 | 47429847 | T | G | F | L | MSH2 | ENST00000233146 | 0.33779 | 0.00586 | 0.84278 | 0.35761 | 0.77909 | 0.5308301513 | 0.95259 | 0.1813773997 | -0.0675741 | 3.312606555 | 0 |
| 2 | 47429848 | C | A | Q | K | MSH2 | ENST00000233146 | 0.46531 | 0.00556 | 0.58157 | 0.55073 | 0.85843 | 0.4785881208 | 0.93606 | 0.9361437559 | 0.176853 | 7.817076151 | 0 |
| 2 | 47429848 | C | G | Q | E | MSH2 | ENST00000233146 | 0.42675 | 0.00566 | 0.5201 | 0.45362 | 0.78473 | 0.4214691474 | 0.93873 | 0.8308686018 | 0.07292 | 7.817076151 | 0 |
| 2 | 47429849 | A | C | Q | P | MSH2 | ENST00000233146 | 0.70744 | 0.01676 | 0.51425 | 0.85414 | 0.9528 | 0.7343037267 | 0.96754 | 0.9943290353 | 0.362633 | 7.014207902 | 0 |
| 2 | 47429849 | A | G | Q | R | MSH2 | ENST00000233146 | 0.4349 | 0.01075 | 0.57629 | 0.64589 | 0.85843 | 0.4931040443 | 0.94918 | 0.4181759027 | -0.111611 | 7.242773655 | 0 |
| 2 | 47429849 | A | T | Q | L | MSH2 | ENST00000233146 | 0.44435 | 0.00586 | 0.50179 | 0.8518 | 0.8124 | 0.4755785301 | 0.92157 | 0.9871459007 | 0.17593 | 6.836399095 | 0 |
| 2 | 47429850 | A | C | Q | H | MSH2 | ENST00000233146 | 0.34244 | 0.00599 | 0.56821 | 0.38759 | 0.79938 | 0.4166648879 | 0.95325 | 0.3978202343 | -0.109334 | 4.880782248 | 0 |
| 2 | 47429850 | A | T | Q | H | MSH2 | ENST00000233146 | 0.3379 | 0.00599 | 0.56821 | 0.45655 | 0.79938 | 0.4166648879 | 0.95345 | 0.4908911884 | -0.101198 | 4.880782248 | 0 |
| 2 | 47429851 | A | G | R | G | MSH2 | ENST00000233146 | 0.73301 | 0.02574 | 0.61623 | 0.94769 | 0.95133 | 0.5652682723 | 0.96991 | 0.996538043 | 0.424054 | 7.33871856 | 0 |
| 2 | 47429852 | G | A | R | K | MSH2 | ENST00000233146 | 0.42764 | 0.00648 | 0.69804 | 0.61181 | 0.80595 | 0.4498024036 | 0.93576 | 0.8153717518 | 0.133837 | 3.911441325 | 0 |
| 2 | 47429852 | G | C | R | T | MSH2 | ENST00000233146 | 0.74085 | 0.02017 | 0.68535 | 0.86348 | 0.96044 | 0.6163756108 | 0.94379 | 0.995439589 | 0.382039 | 4.103006468 | 0 |
| 2 | 47429853 | A | C | R | S | MSH2 | ENST00000233146 | 0.57174 | 0.01096 | 0.63468 | 0.86217 | 0.92763 | 0.5973580876 | 0.97378 | 0.9864674211 | 0.186995 | 4.122943553 | 0 |
| 2 | 47429853 | A | T | R | S | MSH2 | ENST00000233146 | 0.56582 | 0.01096 | 0.63468 | 0.86217 | 0.92763 | 0.5973580876 | 0.97418 | 0.9864674211 | 0.190431 | 4.122943553 | 0 |
| 2 | 47429854 | C | A | Q | K | MSH2 | ENST00000233146 | 0.3497 | 0.00532 | 0.5159 | 0.28764 | 0.7285 | 0.3763670265 | 0.95829 | 0.8878097534 | -0.0458167 | 1.073532878 | 0 |
| 2 | 47429854 | C | G | Q | E | MSH2 | ENST00000233146 | 0.43592 | 0.00722 | 0.4595 | 0.45623 | 0.76024 | 0.3135519125 | 0.89543 | 0.06523539115 | -0.174596 | 3.613719323 | 0 |
| 2 | 47429855 | A | C | Q | P | MSH2 | ENST00000233146 | 0.7262 | 0.01145 | 0.45634 | 0.8075 | 0.92844 | 0.7115478084 | 0.93372 | 0.9649157524 | 0.23787 | 6.757652322 | 0 |
| 2 | 47429855 | A | G | Q | R | MSH2 | ENST00000233146 | 0.42247 | 0.00569 | 0.51071 | 0.60153 | 0.68292 | 0.3369530727 | 0.97591 | 0.6539762397 | 0.0718569 | 2.172271092 | 0 |
| 2 | 47429855 | A | T | Q | L | MSH2 | ENST00000233146 | 0.48889 | 0.00747 | 0.44278 | 0.9076 | 0.84641 | 0.387732578 | 0.89193 | 0.9726143479 | 0.128424 | 5.152572254 | 0 |
| 2 | 47429856 | A | C | Q | H | MSH2 | ENST00000233146 | 0.34277 | 0.00592 | 0.49933 | 0.43534 | 0.79212 | 0.4064894017 | 0.9869 | 0.2583595514 | -0.073172 | 2.311399992 | 0 |
| 2 | 47429856 | A | T | Q | H | MSH2 | ENST00000233146 | 0.33823 | 0.00592 | 0.49933 | 0.43862 | 0.79212 | 0.4064894017 | 0.98689 | 0.3446485698 | -0.0472815 | 2.311399992 | 0 |
| 2 | 47429857 | G | A | A | T | MSH2 | ENST00000233146 | 0.48107 | 0.00586 | 0.29966 | 0.61616 | 0.63525 | 0.2200388767 | 0.93351 | 0.5552829504 | 0.145447 | 1.013319876 | 0 |
| 2 | 47429857 | G | C | A | P | MSH2 | ENST00000233146 | 0.52579 | 0.01121 | 0.37114 | 0.35746 | 0.82836 | 0.5331883032 | 0.91573 | 0.7272934318 | 0.253144 | 5.927831505 | 0 |
| 2 | 47429857 | G | T | A | S | MSH2 | ENST00000233146 | 0.44065 | 0.00586 | 0.25701 | 0.12255 | 0.62213 | 0.2018774161 | 0.89289 | 0.8336168528 | 0.09496 | 0.9780991598 | 0 |
| 2 | 47429858 | C | A | A | E | MSH2 | ENST00000233146 | 0.29492 | 0.00705 | 0.39487 | 0.32827 | 0.75552 | 0.3557519807 | 0.86576 | 0.686337173 | 0.218742 | 5.927831505 | 0 |
| 2 | 47429858 | C | G | A | G | MSH2 | ENST00000233146 | 0.31478 | 0.00741 | 0.28152 | 0.31409 | 0.77141 | 0.2355080037 | 0.87262 | 0.6549128294 | 0.184481 | 4.173461761 | 0 |
| 2 | 47429858 | C | T | A | V | MSH2 | ENST00000233146 | 0.31655 | 0.0058 | 0.34311 | 0.16639 | 0.74306 | 0.2672608969 | 0.87977 | 0.6408271194 | 0.205246 | 3.295613427 | 0 |
| 2 | 47429860 | G | A | A | T | MSH2 | ENST00000233146 | 0.92003 | 0.03413 | 0.53872 | 0.89258 | 0.96517 | 0.5436099329 | 0.90851 | 0.9924300313 | 0.313734 | 7.354676094 | 0 |
| 2 | 47429860 | G | C | A | P | MSH2 | ENST00000233146 | 0.93174 | 0.03243 | 0.6035 | 0.65081 | 0.97178 | 0.7759118745 | 0.95584 | 0.9959197044 | 0.483717 | 8.978896718 | 0 |
| 2 | 47429860 | G | T | A | S | MSH2 | ENST00000233146 | 0.59276 | 0.01604 | 0.48746 | 0.42116 | 0.86403 | 0.4495062437 | 0.87868 | 0.866915882 | 0.244521 | 5.369807885 | 0 |
| 2 | 47429861 | C | A | A | E | MSH2 | ENST00000233146 | 0.63696 | 0.03257 | 0.61741 | 0.76776 | 0.97735 | 0.7628346799 | 0.97217 | 0.9983152151 | 0.5008 | 8.978896718 | 0 |
| 2 | 47429861 | C | G | A | G | MSH2 | ENST00000233146 | 0.4742 | 0.01221 | 0.51149 | 0.49051 | 0.87636 | 0.4617151195 | 0.92262 | 0.9739471674 | 0.274839 | 6.874462099 | 0 |
| 2 | 47429861 | C | T | A | V | MSH2 | ENST00000233146 | 0.64283 | 0.03231 | 0.56528 | 0.74672 | 0.9608 | 0.5625882775 | 0.9442 | 0.9954715967 | 0.345513 | 4.120426664 | 0 |
| 2 | 47429863 | A | C | N | H | MSH2 | ENST00000233146 | 0.57195 | 0.02574 | 0.34859 | 0.67486 | 0.87877 | 0.4105480836 | 0.93105 | 0.9612686634 | 0.166563 | 3.955525354 | 0 |
| 2 | 47429863 | A | G | N | D | MSH2 | ENST00000233146 | 0.54535 | 0.01063 | 0.41781 | 0.88967 | 0.85064 | 0.3419035943 | 0.87318 | 0.8996646404 | 0.121884 | 5.034013515 | 0 |
| 2 | 47429863 | A | T | N | Y | MSH2 | ENST00000233146 | 0.74372 | 0.02475 | 0.40294 | 0.72409 | 0.92438 | 0.4671477513 | 0.91736 | 0.9909723997 | 0.275854 | 6.860791179 | 0 |
| 2 | 47429864 | A | C | N | T | MSH2 | ENST00000233146 | 0.27379 | 0.00574 | 0.31928 | 0.06192 | 0.68292 | 0.263071101 | 0.9249 | 0.2639541626 | 0.0863874 | 0.5303516404 | 0 |
| 2 | 47429864 | A | G | N | S | MSH2 | ENST00000233146 | 0.27875 | 0.00648 | 0.2651 | 0.12753 | 0.63525 | 0.2072513145 | 0.86077 | 0.2828012407 | 0.0663843 | 2.474366684 | 0 |
| 2 | 47429864 | A | T | N | I | MSH2 | ENST00000233146 | 0.54351 | 0.01516 | 0.36156 | 0.50995 | 0.90581 | 0.5247252451 | 0.92383 | 0.9883325696 | 0.215111 | 6.860791179 | 0 |
| 2 | 47429865 | C | A | N | K | MSH2 | ENST00000233146 | 0.29904 | 0.00574 | 0.45954 | 0.08682 | 0.79998 | 0.413988749 | 0.88483 | 0.9539754987 | 0.0562808 | 5.862154045 | 0 |
| 2 | 47429865 | C | G | N | K | MSH2 | ENST00000233146 | 0.30447 | 0.00574 | 0.45954 | 0.05752 | 0.80058 | 0.413988749 | 0.88258 | 0.6625140309 | -0.0700831 | 5.862154045 | 0 |
| 2 | 47429866 | T | A | L | I | MSH2 | ENST00000233146 | 0.59197 | 0.03124 | 0.6047 | 0.89973 | 0.91904 | 0.2662125631 | 0.84383 | 0.9857459068 | 0.252959 | 7.779386722 | 0 |
| 2 | 47429866 | T | G | L | V | MSH2 | ENST00000233146 | 0.5876 | 0.03212 | 0.53752 | 0.92296 | 0.93786 | 0.4585140701 | 0.91683 | 0.9902138114 | 0.301332 | 8.870533153 | 0 |
| 2 | 47429867 | T | C | L | S | MSH2 | ENST00000233146 | 0.9056 | 0.03144 | 0.62471 | 0.78026 | 0.97438 | 0.7122686669 | 0.96318 | 0.9984397292 | 0.512502 | 6.715932324 | 0 |
| 2 | 47429868 | A | C | L | F | MSH2 | ENST00000233146 | 0.67653 | 0.01445 | 0.59498 | 0.63487 | 0.90755 | 0.5058757974 | 0.96608 | 0.9932966828 | 0.326072 | 8.870533153 | 0 |
| 2 | 47429868 | A | T | L | F | MSH2 | ENST00000233146 | 0.66976 | 0.01445 | 0.59498 | 0.65847 | 0.90755 | 0.5058757974 | 0.96502 | 0.9932966828 | 0.32607 | 8.870533153 | 0 |
| 2 | 47429869 | C | A | Q | K | MSH2 | ENST00000233146 | 0.70158 | 0.02475 | 0.60357 | 0.48957 | 0.94469 | 0.5595099968 | 0.98435 | 0.9410895705 | 0.343926 | 3.249526552 | 0 |
| 2 | 47429869 | C | G | Q | E | MSH2 | ENST00000233146 | 0.38295 | 0.02277 | 0.55076 | 0.43083 | 0.81182 | 0.4539109917 | 0.95801 | 0.6138963103 | 0.144297 | 5.126196033 | 0 |
| 2 | 47429870 | A | C | Q | P | MSH2 | ENST00000233146 | 0.82264 | 0.03053 | 0.55546 | 0.89886 | 0.99138 | 0.7807901258 | 0.98147 | 0.993750155 | 0.372972 | 6.65864236 | 0 |
| 2 | 47429870 | A | G | Q | R | MSH2 | ENST00000233146 | 0.65908 | 0.02875 | 0.60028 | 0.90831 | 0.97735 | 0.5752089486 | 0.98932 | 0.9627716541 | 0.311008 | 3.106123758 | 0 |
| 2 | 47429870 | A | T | Q | L | MSH2 | ENST00000233146 | 0.8066 | 0.02342 | 0.53982 | 0.9728 | 0.96773 | 0.5602119065 | 0.97564 | 0.996308744 | 0.385941 | 7.728034735 | 0 |
| 2 | 47429871 | A | C | Q | H | MSH2 | ENST00000233146 | 0.44138 | 0.01048 | 0.59432 | 0.88875 | 0.88401 | 0.5276525126 | 0.98632 | 0.9735661149 | 0.0514041 | 7.470556648 | 0 |
| 2 | 47429871 | A | T | Q | H | MSH2 | ENST00000233146 | 0.43623 | 0.01048 | 0.59432 | 0.89003 | 0.88401 | 0.5276525126 | 0.98627 | 0.9739843607 | 0.0739322 | 7.470556648 | 0 |
| 2 | 47429872 | G | A | D | N | MSH2 | ENST00000233146 | 0.77172 | 0.01002 | 0.5646 | 0.77265 | 0.93123 | 0.6861569127 | 0.94672 | 0.9940599203 | 0.197564 | 9.011184286 | 0 |
| 2 | 47429872 | G | C | D | H | MSH2 | ENST00000233146 | 0.81724 | 0.0325 | 0.59295 | 0.57999 | 0.97215 | 0.7390608342 | 0.98828 | 0.9981934428 | 0.508005 | 9.584434224 | 0 |
| 2 | 47429872 | G | T | D | Y | MSH2 | ENST00000233146 | 0.82584 | 0.0325 | 0.56985 | 0.61502 | 0.98071 | 0.7952013205 | 0.99201 | 0.9994540811 | 0.521164 | 9.584434224 | 0 |
| 2 | 47429873 | A | C | D | A | MSH2 | ENST00000233146 | 0.8701 | 0.02896 | 0.49575 | 0.80372 | 0.98869 | 0.7543656865 | 0.97203 | 0.9985550046 | 0.515569 | 9.584434224 | 0 |
| 2 | 47429873 | A | G | D | G | MSH2 | ENST00000233146 | 0.87563 | 0.02637 | 0.51484 | 0.75104 | 0.99072 | 0.7417745955 | 0.98799 | 0.9964485168 | 0.512246 | 9.584434224 | 0 |
| 2 | 47429873 | A | T | D | V | MSH2 | ENST00000233146 | 0.86354 | 0.02719 | 0.48114 | 0.88324 | 0.99006 | 0.7748621499 | 0.99088 | 0.9990719557 | 0.529165 | 9.584434224 | 0 |
| 2 | 47429874 | T | A | D | E | MSH2 | ENST00000233146 | 0.44783 | 0.01654 | 0.53513 | 0.42302 | 0.93004 | 0.5840334826 | 0.94877 | 0.9872788191 | 0.264643 | 4.250314339 | 0 |
| 2 | 47429874 | T | G | D | E | MSH2 | ENST00000233146 | 0.45413 | 0.01654 | 0.53513 | 0.44729 | 0.93044 | 0.5840334826 | 0.94868 | 0.9839830399 | 0.264644 | 4.250314339 | 0 |
| 2 | 47429875 | T | A | C | S | MSH2 | ENST00000233146 | 0.67987 | 0.02391 | 0.68826 | 0.91805 | 0.97401 | 0.5683217375 | 0.94241 | 0.9801863432 | 0.433471 | 8.66636744 | 0 |
| 2 | 47429875 | T | C | C | R | MSH2 | ENST00000233146 | 0.89143 | 0.02828 | 0.79997 | 0.95672 | 0.97997 | 0.820274422 | 0.96258 | 0.998526454 | 0.461443 | 8.66636744 | 1 |
| 2 | 47429875 | T | G | C | G | MSH2 | ENST00000233146 | 0.87925 | 0.03224 | 0.64886 | 0.8099 | 0.98071 | 0.6033297728 | 0.95902 | 0.9982819557 | 0.461128 | 8.66636744 | 0 |
| 2 | 47429876 | G | A | C | Y | MSH2 | ENST00000233146 | 0.88916 | 0.02908 | 0.81764 | 0.56337 | 0.98367 | 0.6948825156 | 0.95879 | 0.9976690412 | 0.480863 | 7.397928951 | 0 |
| 2 | 47429876 | G | C | C | S | MSH2 | ENST00000233146 | 0.72124 | 0.02391 | 0.68826 | 0.42141 | 0.97364 | 0.5683217375 | 0.93865 | 0.9882115722 | 0.444313 | 8.66636744 | 0 |
| 2 | 47429876 | G | T | C | F | MSH2 | ENST00000233146 | 0.90411 | 0.03013 | 0.71149 | 0.3581 | 0.98034 | 0.6574676202 | 0.95377 | 0.9978838563 | 0.459841 | 4.991884884 | 0 |
| 2 | 47429877 | T | G | C | W | MSH2 | ENST00000233146 | 0.71008 | 0.03257 | 0.76607 | 0.6686 | 0.94543 | 0.7224481468 | 0.97984 | 0.998953104 | 0.42799 | 8.66636744 | 0 |
| 2 | 47429878 | T | A | Y | N | MSH2 | ENST00000233146 | 0.90122 | 0.02785 | 0.55889 | 0.9448 | 0.95681 | 0.679821989 | 0.96853 | 0.9960891008 | 0.425961 | 5.217666226 | 0 |

| | | | | | | | | | | | | | | | | |
|---|---|---|---|---|---|---|---|---|---|---|---|---|---|---|---|---|
| 2 | 47429878 | T | C | Y | H | MSH2 | ENST00000233146 | 0.90804 | 0.03112 | 0.56153 | 0.87864 | 0.9659 | 0.5659307692 | 0.95272 | 0.9946318865 | 0.422922 | 4.804400097 | 0 |
| 2 | 47429878 | T | G | Y | D | MSH2 | ENST00000233146 | 0.9067 | 0.02679 | 0.55259 | 0.94753 | 0.97438 | 0.7752031895 | 0.99023 | 0.9924756885 | 0.462543 | 6.9859218 | 0 |
| 2 | 47429879 | A | C | Y | S | MSH2 | ENST00000233146 | 0.75837 | 0.0279 | 0.45947 | 0.86855 | 0.93863 | 0.6444593083 | 0.98001 | 0.9940851927 | 0.356505 | 7.327502973 | 0 |
| 2 | 47429879 | A | G | Y | C | MSH2 | ENST00000233146 | 0.93994 | 0.03161 | 0.56971 | 0.84266 | 0.9781 | 0.602047235 | 0.97061 | 0.9817532301 | 0.35337 | 7.474672033 | 0 |
| 2 | 47429881 | C | G | R | G | MSH2 | ENST00000233146 | 0.79887 | 0.01479 | 0.19647 | 0.88369 | 0.95133 | 0.6931620589 | 0.97362 | 0.9947510362 | 0.417904 | 4.528186237 | 0 |
| 2 | 47429882 | G | A | R | Q | MSH2 | ENST00000233146 | 0.48366 | 0.00574 | 0.14012 | 0.63536 | 0.89009 | 0.5890109679 | 0.95976 | 0.2421281189 | 0.0947051 | 3.018466795 | 0 |
| 2 | 47429882 | G | C | R | P | MSH2 | ENST00000233146 | 0.82061 | 0.02381 | 0.2537 | 0.85988 | 0.9659 | 0.8596825094 | 0.9834 | 0.9959018826 | 0.40971 | 7.368447297 | 0 |
| 2 | 47429882 | G | T | R | L | MSH2 | ENST00000233146 | 0.59567 | 0.0058 | 0.22388 | 0.86054 | 0.93786 | 0.7002629689 | 0.9846 | 0.9916833639 | 0.287675 | 5.755261418 | 0 |
| 2 | 47429884 | C | A | L | I | MSH2 | ENST00000233146 | 0.28724 | 0.00556 | 0.37974 | 0.52999 | 0.67478 | 0.202284543 | 0.80003 | 0.3643614054 | -0.0961289 | 0.8686538121 | 0 |
| 2 | 47429884 | C | G | L | V | MSH2 | ENST00000233146 | 0.26669 | 0.00569 | 0.30698 | 0.48523 | 0.72016 | 0.2980230881 | 0.85221 | 0.08314263821 | -0.246464 | 0.07516041708 | 0 |
| 2 | 47429884 | C | T | L | F | MSH2 | ENST00000233146 | 0.66913 | 0.00904 | 0.37016 | 0.85137 | 0.89829 | 0.4971487624 | 0.83516 | 0.7516057491 | 0.185715 | 2.31982249 | 0 |
| 2 | 47429885 | T | A | L | H | MSH2 | ENST00000233146 | 0.72668 | 0.01516 | 0.47086 | 0.78768 | 0.9781 | 0.6226613995 | 0.90183 | 0.9886615276 | 0.346203 | 5.913741719 | 0 |
| 2 | 47429885 | T | C | L | P | MSH2 | ENST00000233146 | 0.75193 | 0.0118 | 0.52427 | 0.82707 | 0.9729 | 0.8375483394 | 0.97181 | 0.977978766 | 0.472917 | 5.913741719 | 1 |
| 2 | 47429885 | T | G | L | R | MSH2 | ENST00000233146 | 0.72981 | 0.00812 | 0.47894 | 0.74572 | 0.97624 | 0.7965855712 | 0.9825 | 0.9638521671 | 0.491333 | 5.913741719 | 1 |
| 2 | 47429887 | T | A | Y | N | MSH2 | ENST00000233146 | 0.90031 | 0.02844 | 0.66878 | 0.95967 | 0.9729 | 0.7749125381 | 0.94898 | 0.9995273352 | 0.464877 | 6.469061233 | 0 |
| 2 | 47429887 | T | C | Y | H | MSH2 | ENST00000233146 | 0.8979 | 0.01731 | 0.67794 | 0.94374 | 0.97923 | 0.6770238112 | 0.91223 | 0.998077035 | 0.421693 | 6.656966379 | 0 |
| 2 | 47429887 | T | G | Y | D | MSH2 | ENST00000233146 | 0.86976 | 0.02738 | 0.65969 | 0.955 | 0.96189 | 0.8327334212 | 0.97981 | 0.9996380806 | 0.489256 | 8.491854674 | 0 |
| 2 | 47429888 | A | C | Y | S | MSH2 | ENST00000233146 | 0.91002 | 0.02927 | 0.58681 | 0.91503 | 0.9528 | 0.7393258302 | 0.97405 | 0.9992796779 | 0.368824 | 4.736885928 | 0 |
| 2 | 47429888 | A | G | Y | C | MSH2 | ENST00000233146 | 0.92864 | 0.03231 | 0.66918 | 0.91593 | 0.95572 | 0.7203287295 | 0.96799 | 0.9991750121 | 0.363176 | 5.237508262 | 0 |
| 2 | 47429888 | A | T | Y | F | MSH2 | ENST00000233146 | 0.67318 | 0.01015 | 0.56295 | 0.88292 | 0.89694 | 0.427671883 | 0.89163 | 0.928919673 | 0.167007 | 6.301703031 | 0 |
| 2 | 47429890 | C | A | Q | K | MSH2 | ENST00000233146 | 0.39463 | 0.00566 | 0.51713 | 0.83646 | 0.93162 | 0.67943965 | 0.98784 | 0.8932765126 | 0.447355 | 5.170792783 | 0 |
| 2 | 47429890 | C | G | Q | E | MSH2 | ENST00000233146 | 0.39754 | 0.01281 | 0.45207 | 0.76394 | 0.91442 | 0.5946493252 | 0.95902 | 0.9733518362 | 0.339833 | 5.523350654 | 0 |
| 2 | 47429891 | A | C | Q | P | MSH2 | ENST00000233146 | 0.79002 | 0.02494 | 0.46339 | 0.94846 | 0.98834 | 0.8456816363 | 0.98335 | 0.9947762489 | 0.488934 | 9.376607244 | 0 |
| 2 | 47429891 | A | G | Q | R | MSH2 | ENST00000233146 | 0.47363 | 0.01471 | 0.5129 | 0.84397 | 0.96117 | 0.7002849199 | 0.99155 | 0.9802752137 | 0.465078 | 4.781461989 | 0 |
| 2 | 47429891 | A | T | Q | L | MSH2 | ENST00000233146 | 0.49228 | 0.00754 | 0.45074 | 0.7925 | 0.89739 | 0.66721827 | 0.96655 | 0.9701536298 | 0.405369 | 5.90991512 | 0 |
| 2 | 47429892 | G | C | Q | H | MSH2 | ENST00000233146 | 0.76998 | 0.0317 | 0.50451 | 0.73498 | 0.95023 | 0.6511163916 | 0.97399 | 0.9949106574 | 0.449439 | 4.854619619 | 0 |
| 2 | 47429892 | G | T | Q | H | MSH2 | ENST00000233146 | 0.76347 | 0.0317 | 0.50451 | 0.73235 | 0.95023 | 0.6511163916 | 0.9739 | 0.9949106574 | 0.449435 | 4.854619619 | 0 |
| 2 | 47429893 | G | A | G | S | MSH2 | ENST00000233146 | 0.52712 | 0.00603 | 0.3585 | 0.38152 | 0.67843 | 0.4250588154 | 0.92042 | 0.8278850913 | 0.126155 | 0.3164053749 | 0 |
| 2 | 47429893 | G | C | G | R | MSH2 | ENST00000233146 | 0.73077 | 0.02037 | 0.49038 | 0.20616 | 0.81414 | 0.7185209369 | 0.95504 | 0.8910418153 | 0.241588 | 4.941041312 | 0 |
| 2 | 47429893 | G | T | G | C | MSH2 | ENST00000233146 | 0.56408 | 0.02883 | 0.4348 | 0.10058 | 0.79697 | 0.5299593399 | 0.88105 | 0.7672605515 | 0.208137 | 4.941041312 | 0 |
| 2 | 47429894 | G | A | G | D | MSH2 | ENST00000233146 | 0.65672 | 0.01253 | 0.528 | 0.20378 | 0.8356 | 0.7585856823 | 0.93556 | 0.9048444033 | 0.246104 | 4.941041312 | 1 |
| 2 | 47429894 | G | C | G | A | MSH2 | ENST00000233146 | 0.37816 | 0.00556 | 0.38613 | 0.01026 | 0.73148 | 0.374663701 | 0.81452 | 0.5875073671 | 0.0915411 | -1.53918517 | 0 |
| 2 | 47429894 | G | T | G | V | MSH2 | ENST00000233146 | 0.48723 | 0.00627 | 0.3988 | 0.06726 | 0.74867 | 0.5356559349 | 0.91243 | 0.84882164 | 0.199963 | 2.206481602 | 0 |
| 2 | 47429896 | A | C | I | L | MSH2 | ENST00000233146 | 0.3318 | 0.0058 | 0.41378 | 0.73029 | 0.77141 | 0.4030560074 | 0.95022 | 0.6768519282 | 0.141875 | 1.830998694 | 0 |
| 2 | 47429896 | A | G | I | V | MSH2 | ENST00000233146 | 0.21165 | 0.00603 | 0.30514 | 0.50866 | 0.65575 | 0.1410706836 | 0.81231 | 0.6298763752 | 0.0819931 | 0.469802977 | 0 |
| 2 | 47429896 | A | T | I | L | MSH2 | ENST00000233146 | 0.32733 | 0.0058 | 0.41378 | 0.73029 | 0.77141 | 0.4030560074 | 0.95001 | 0.3093633207 | 0.0154961 | 1.830998694 | 0 |
| 2 | 47429897 | T | A | I | K | MSH2 | ENST00000233146 | 0.59392 | 0.00722 | 0.5046 | 0.79657 | 0.96627 | 0.7377862836 | 0.96365 | 0.9886264205 | 0.403887 | 7.206616996 | 1 |
| 2 | 47429897 | T | C | I | T | MSH2 | ENST00000233146 | 0.57631 | 0.00747 | 0.56662 | 0.63188 | 0.93358 | 0.4644591772 | 0.95313 | 0.9581299424 | 0.330259 | 5.228858241 | 0 |
| 2 | 47429897 | T | G | I | R | MSH2 | ENST00000233146 | 0.58183 | 0.00497 | 0.47159 | 0.83147 | 0.95753 | 0.7372434708 | 0.97646 | 0.9899569154 | 0.385181 | 7.206616996 | 1 |
| 2 | 47429898 | A | G | I | M | MSH2 | ENST00000233146 | 0.38559 | 0.01705 | 0.38227 | 0.75203 | 0.87974 | 0.4389612944 | 0.92905 | 0.7250044942 | 0.214879 | 4.517606244 | 0 |
| 2 | 47429899 | A | C | N | H | MSH2 | ENST00000233146 | 0.48806 | 0.0058 | 0.35623 | 0.32271 | 0.7727 | 0.2800605364 | 0.93057 | 0.576428473 | 0.122007 | 3.080433145 | 0 |
| 2 | 47429899 | A | G | N | D | MSH2 | ENST00000233146 | 0.44826 | 0.00551 | 0.4075 | 0.75864 | 0.70179 | 0.3587955583 | 0.87342 | 0.6481030583 | 0.105183 | 2.042824548 | 0 |
| 2 | 47429899 | A | T | N | Y | MSH2 | ENST00000233146 | 0.67199 | 0.00866 | 0.3936 | 0.45515 | 0.89422 | 0.3789900643 | 0.9482 | 0.9834656715 | 0.27494 | 4.355126348 | 0 |
| 2 | 47429900 | A | C | N | T | MSH2 | ENST00000233146 | 0.3277 | 0.00669 | 0.33163 | 0.76225 | 0.78286 | 0.210875892 | 0.86149 | 0.6831107736 | 0.1314 | 4.072574644 | 0 |
| 2 | 47429900 | A | G | N | S | MSH2 | ENST00000233146 | 0.26688 | 0.00627 | 0.28002 | 0.66635 | 0.57225 | 0.1563550721 | 0.79794 | 0.6626456976 | 0.089305 | 2.608190717 | 0 |
| 2 | 47429900 | A | T | N | I | MSH2 | ENST00000233146 | 0.39969 | 0.00861 | 0.35138 | 0.82802 | 0.8306 | 0.3469174076 | 0.91127 | 0.6824865937 | 0.220877 | 4.540682444 | 0 |
| 2 | 47429902 | C | A | Q | K | MSH2 | ENST00000233146 | 0.33553 | 0.0058 | 0.38048 | 0.66639 | 0.73734 | 0.5415609958 | 0.95426 | 0.5678053452 | 0.237112 | 2.00056565 | 0 |
| 2 | 47429902 | C | G | Q | E | MSH2 | ENST00000233146 | 0.3689 | 0.01203 | 0.32649 | 0.65245 | 0.79818 | 0.5245704449 | 0.92153 | 0.8562172651 | 0.216543 | 4.074432941 | 0 |
| 2 | 47429903 | A | C | Q | P | MSH2 | ENST00000233146 | 0.54569 | 0.0262 | 0.33919 | 0.91048 | 0.96664 | 0.7901016999 | 0.97288 | 0.9366737008 | 0.253234 | 5.006662999 | 0 |
| 2 | 47429903 | A | G | Q | R | MSH2 | ENST00000233146 | 0.31821 | 0.00615 | 0.37488 | 0.66813 | 0.79028 | 0.5666688829 | 0.97158 | 0.5747087598 | 0.25143 | 2.785224767 | 0 |
| 2 | 47429903 | A | T | Q | L | MSH2 | ENST00000233146 | 0.42424 | 0.01009 | 0.31099 | 0.86686 | 0.8933 | 0.5571005065 | 0.94975 | 0.9393747449 | 0.279421 | 2.957890379 | 0 |
| 2 | 47429904 | A | C | Q | H | MSH2 | ENST00000233146 | 0.27251 | 0.00664 | 0.37394 | 0.69275 | 0.77463 | 0.5673125548 | 0.93851 | 0.1535624117 | 0.10683 | 2.835743149 | 0 |
| 2 | 47429904 | A | T | Q | H | MSH2 | ENST00000233146 | 0.26855 | 0.00664 | 0.37394 | 0.69275 | 0.77591 | 0.5673125548 | 0.93865 | 0.1535624117 | 0.106829 | 2.835743149 | 0 |
| 2 | 47429905 | C | A | L | I | MSH2 | ENST00000233146 | 0.39882 | 0.00687 | 0.50834 | 0.64547 | 0.75822 | 0.2302362314 | 0.9036 | 0.5402325392 | 0.117315 | 1.591218827 | 0 |
| 2 | 47429905 | C | G | L | V | MSH2 | ENST00000233146 | 0.42144 | 0.00773 | 0.43387 | 0.65605 | 0.84375 | 0.390447923 | 0.96337 | 0.6999246478 | 0.207226 | 3.208018206 | 0 |
| 2 | 47429906 | T | A | L | Q | MSH2 | ENST00000233146 | 0.55243 | 0.01044 | 0.52067 | 0.92796 | 0.96008 | 0.6947310342 | 0.98477 | 0.9922560453 | 0.473011 | 7.766799184 | 0 |
| 2 | 47429906 | T | C | L | P | MSH2 | ENST00000233146 | 0.75495 | 0.02714 | 0.62864 | 0.95452 | 0.97252 | 0.8615533457 | 0.96099 | 0.9979404807 | 0.472388 | 7.766799184 | 1 |
| 2 | 47429906 | T | G | L | R | MSH2 | ENST00000233146 | 0.73605 | 0.0245 | 0.58327 | 0.96065 | 0.97997 | 0.8456292788 | 0.98256 | 0.997597158 | 0.498324 | 7.766799184 | 1 |
| 2 | 47429908 | C | A | P | T | MSH2 | ENST00000233146 | 0.65877 | 0.01562 | 0.60221 | 0.87894 | 0.97401 | 0.5116501024 | 0.98139 | 0.9986762404 | 0.485954 | 6.24098815 | 0 |
| 2 | 47429908 | C | G | P | A | MSH2 | ENST00000233146 | 0.48354 | 0.01244 | 0.54649 | 0.85911 | 0.93824 | 0.4426002902 | 0.94644 | 0.9816958904 | 0.378737 | 6.14047556 | 0 |
| 2 | 47429908 | C | T | P | S | MSH2 | ENST00000233146 | 0.66438 | 0.02927 | 0.56172 | 0.82575 | 0.93939 | 0.5577118991 | 0.9347 | 0.9975389242 | 0.37361 | 5.497795428 | 0 |
| 2 | 47429909 | C | A | P | H | MSH2 | ENST00000233146 | 0.63512 | 0.03281 | 0.62014 | 0.91183 | 0.96553 | 0.5588635569 | 0.92924 | 0.999311924 | 0.43851 | 7.12932375 | 0 |
| 2 | 47429909 | C | G | P | R | MSH2 | ENST00000233146 | 0.6355 | 0.03212 | 0.63508 | 0.87925 | 0.97661 | 0.6775263989 | 0.9779 | 0.9982787371 | 0.484724 | 6.675427454 | 0 |
| 2 | 47429909 | C | T | P | L | MSH2 | ENST00000233146 | 0.65008 | 0.03137 | 0.64 | 0.9016 | 0.96883 | 0.6433810914 | 0.98073 | 0.9993738532 | 0.484627 | 7.861346681 | 0 |
| 2 | 47429911 | A | C | N | H | MSH2 | ENST00000233146 | 0.33254 | 0.00569 | 0.15315 | 0.70078 | 0.70262 | 0.2623087227 | 0.94348 | 0.3575619459 | 0.0998727 | 2.108430008 | 0 |
| 2 | 47429911 | A | G | N | D | MSH2 | ENST00000233146 | 0.30946 | 0.00556 | 0.19461 | 0.71065 | 0.69339 | 0.3489502327 | 0.9235 | 0.4932090044 | 0.0880442 | 3.070552223 | 0 |
| 2 | 47429911 | A | T | N | Y | MSH2 | ENST00000233146 | 0.35774 | 0.00543 | 0.18644 | 0.72539 | 0.77845 | 0.3768139792 | 0.99308 | 0.4087138474 | 0.205516 | 2.167957675 | 0 |
| 2 | 47429912 | A | C | N | T | MSH2 | ENST00000233146 | 0.32204 | 0.00569 | 0.12477 | 0.68714 | 0.76223 | 0.1921968196 | 0.97478 | 0.1111910045 | 0.0804476 | 2.465504916 | 0 |
| 2 | 47429912 | A | G | N | S | MSH2 | ENST00000233146 | 0.2764 | 0.00608 | 0.08963 | 0.66022 | 0.62657 | 0.1461486424 | 0.86666 | 0.1193650886 | 0.0475703 | 2.243484017 | 0 |
| 2 | 47429912 | A | T | N | I | MSH2 | ENST00000233146 | 0.39585 | 0.00622 | 0.16969 | 0.82344 | 0.85689 | 0.3619155377 | 0.96268 | 0.4735962749 | 0.167024 | 3.943351729 | 0 |
| 2 | 47429913 | T | A | N | K | MSH2 | ENST00000233146 | 0.26428 | 0.00556 | 0.21908 | 0.66529 | 0.70262 | 0.2892371819 | 0.92851 | 0.1609524786 | 0.0729596 | 2.963357545 | 0 |
| 2 | 47429913 | T | G | N | K | MSH2 | ENST00000233146 | 0.26907 | 0.00556 | 0.21908 | 0.662 | 0.70427 | 0.2892371819 | 0.92853 | 0.1430921257 | 0.0729304 | 2.963357545 | 0 |
| 2 | 47429914 | G | A | V | I | MSH2 | ENST00000233146 | 0.4534 | 0.00681 | 0.30388 | 0.63906 | 0.75552 | 0.1513313712 | 0.72224 | 0.4988424454 | 0.131626 | 2.343113726 | 0 |
| 2 | 47429914 | G | C | V | L | MSH2 | ENST00000233146 | 0.36856 | 0.00592 | 0.467 | 0.47845 | 0.74446 | 0.3173785112 | 0.86438 | 0.4821297824 | 0.154176 | 1.083156495 | 0 |
| 2 | 47429914 | G | T | V | F | MSH2 | ENST00000233146 | 0.50413 | 0.00586 | 0.44804 | 0.6851 | 0.80713 | 0.6518815929 | 0.95565 | 0.7248495817 | 0.381002 | 3.099194951 | 0 |
| 2 | 47429915 | T | C | V | A | MSH2 | ENST00000233146 | 0.69564 | 0.01533 | 0.4915 | 0.86677 | 0.94765 | 0.4742031343 | 0.94044 | 0.9296595454 | 0.292413 | 6.201556635 | 0 |
| 2 | 47429915 | T | G | V | G | MSH2 | ENST00000233146 | 0.74269 | 0.02875 | 0.35976 | 0.92092 | 0.98549 | 0.6787846583 | 0.97472 | 0.9942933564 | 0.459608 | 7.324297451 | 0 |
| 2 | 47429917 | A | C | I | L | MSH2 | ENST00000233146 | 0.41623 | 0.0058 | 0.37901 | 0.71394 | 0.68113 | 0.2984159757 | 0.95237 | 0.6854972243 | 0.158118 | 0.8438435325 | 0 |
| 2 | 47429917 | A | G | I | V | MSH2 | ENST00000233146 | 0.3386 | 0.00615 | 0.28302 | 0.51987 | 0.64157 | 0.1202805666 | 0.89636 | 0.5128861666 | 0.100389 | 0.122587136 | 0 |
| 2 | 47429917 | A | T | I | L | MSH2 | ENST00000233146 | 0.41127 | 0.0058 | 0.37901 | 0.72924 | 0.68023 | 0.2984159757 | 0.95239 | 0.380331099 | 0.147333 | 0.8438435325 | 0 |
| 2 | 47429918 | T | A | I | K | MSH2 | ENST00000233146 | 0.52479 | 0.01108 | 0.45975 | 0.89089 | 0.89648 | 0.6527917343 | 0.96166 | 0.9879698753 | 0.337747 | 2.016298862 | 0 |
| 2 | 47429918 | T | C | I | T | MSH2 | ENST00000233146 | 0.37481 | 0.0054 | 0.51392 | 0.78576 | 0.84535 | 0.3385662805 | 0.98234 | 0.8389956355 | 0.30916 | 2.448170541 | 0 |
| 2 | 47429918 | T | G | I | R | MSH2 | ENST00000233146 | 0.51488 | 0.01533 | 0.43819 | 0.87845 | 0.9045 | 0.6268863942 | 0.96596 | 0.9839083552 | 0.346846 | 2.290335167 | 0 |
| 2 | 47429920 | C | G | Q | E | MSH2 | ENST00000233146 | 0.08922 | 0.00556 | 0.08871 | 0.31852 | 0.79028 | 0.171888317 | 0.97186 | 0.03864667856 | 0.143372 | 0.5223871682 | 0 |
| 2 | 47429921 | A | C | Q | P | MSH2 | ENST00000233146 | 0.16744 | 0.00654 | 0.10281 | 0.56669 | 0.85117 | 0.6204932465 | 0.99186 | 0.462108016 | 0.286553 | 6.189572894 | 0 |
| 2 | 47429921 | A | G | Q | R | MSH2 | ENST00000233146 | 0.13805 | 0.00556 | 0.11557 | 0.47094 | 0.80058 | 0.2298560406 | 0.98831 | 0.07344754788 | 0.165874 | 3.542096876 | 0 |
| 2 | 47429921 | A | T | Q | L | MSH2 | ENST00000233146 | 0.16225 | 0.00574 | 0.07888 | 0.55283 | 0.75552 | 0.3012112892 | 0.98002 | 0.194094643 | 0.144557 | 2.547560896 | 0 |
| 2 | 47429922 | G | C | Q | H | MSH2 | ENST00000233146 | 0.26716 | 0.00586 | 0.11598 | 0.58957 | 0.81761 | 0.2224015869 | 0.99283 | 0.08874852857 | 0.174161 | 3.793628764 | 0 |
| 2 | 47429922 | G | T | Q | H | MSH2 | ENST00000233146 | 0.2634 | 0.00586 | 0.11598 | 0.59184 | 0.81761 | 0.2224015869 | 0.99289 | 0.09153947331 | 0.17415 | 3.793628764 | 0 |
| 2 | 47429923 | G | A | A | T | MSH2 | ENST00000233146 | 0.54109 | 0.00596 | 0.4754 | 0.77858 | 0.80713 | 0.3772341885 | 0.92616 | 0.8063271642 | 0.267908 | 1.644825613 | 0 |
| 2 | 47429923 | G | C | A | P | MSH2 | ENST00000233146 | 0.89342 | 0.0178 | 0.54742 | 0.84901 | 0.94802 | 0.7744322733 | 0.94087 | 0.8891355395 | 0.373038 | 7.718855889 | 0 |
| 2 | 47429923 | G | T | A | S | MSH2 | ENST00000233146 | 0.50661 | 0.00644 | 0.42117 | 0.77188 | 0.82216 | 0.3364838109 | 0.88606 | 0.5353733409 | 0.111028 | 3.600210059 | 0 |
| 2 | 47429924 | C | A | A | D | MSH2 | ENST00000233146 | 0.44917 | 0.01258 | 0.59488 | 0.86139 | 0.911 | 0.784804644 | 0.9754 | 0.9717209935 | 0.408326 | 5.942500078 | 0 |
| 2 | 47429924 | C | G | A | G | MSH2 | ENST00000233146 | 0.34653 | 0.01025 | 0.44767 | 0.8142 | 0.89873 | 0.3724355063 | 0.90415 | 0.9591637254 | 0.222342 | 5.549555717 | 0 |
| 2 | 47429924 | C | T | A | V | MSH2 | ENST00000233146 | 0.29974 | 0.00574 | 0.49045 | 0.76302 | 0.77141 | 0.4055189156 | 0.96769 | 0.5532855392 | 0.192902 | 1.781005693 | 0 |
| 2 | 47429926 | C | A | L | M | MSH2 | ENST00000233146 | 0.57202 | 0.03001 | 0.40906 | 0.84253 | 0.90972 | 0.463963475 | 0.83665 | 0.9197323918 | 0.189752 | 5.547784533 | 0 |
| 2 | 47429926 | C | G | L | V | MSH2 | ENST00000233146 | 0.64136 | 0.02094 | 0.3731 | 0.89564 | 0.93436 | 0.538287855 | 0.95172 | 0.8481086493 | 0.277612 | 5.245137436 | 0 |
| 2 | 47429927 | T | C | L | P | MSH2 | ENST00000233146 | 0.91772 | 0.02951 | 0.5626 | 0.96697 | 0.99552 | 0.904808702 | 0.99359 | 0.9983958602 | 0.534216 | 9.093901029 | 1 |
| 2 | 47429927 | T | G | L | R | MSH2 | ENST00000233146 | 0.91401 | 0.0307 | 0.52318 | 0.962 | 0.99356 | 0.8817864243 | 0.98553 | 0.9983428717 | 0.532578 | 9.093901029 | 1 |
| 2 | 47429929 | G | A | E | K | MSH2 | ENST00000233146 | 0.59762 | 0.0058 | 0.3287 | 0.72233 | 0.84428 | 0.396620197 | 0.9679 | 0.873619616 | 0.259691 | 3.860902917 | 0 |
| 2 | 47429929 | G | C | E | Q | MSH2 | ENST00000233146 | 0.52872 | 0.00726 | 0.22788 | 0.7118 | 0.81124 | 0.3285025049 | 0.91755 | 0.7588079572 | 0.238989 | 3.81020377 | 0 |

| | | | | | | | | | | | | | | | | | | |
|---|---|---|---|---|---|---|---|---|---|---|---|---|---|---|---|---|---|---|
| 2 | 47429930 | A | C | E | A | MSH2 | ENST00000233146 | 0.51001 | 0.00816 | 0.23942 | 0.82011 | 0.85326 | 0.4088083888 | 0.93727 | 0.9542557001 | 0.252875 | 3.842078845 | 0 |
| 2 | 47429930 | A | G | E | G | MSH2 | ENST00000233146 | 0.53697 | 0.00551 | 0.20774 | 0.72525 | 0.77591 | 0.4324770238 | 0.92794 | 0.9667315483 | 0.1917 | 4.493429567 | 0 |
| 2 | 47429930 | A | T | E | V | MSH2 | ENST00000233146 | 0.54008 | 0.00592 | 0.21843 | 0.82005 | 0.86352 | 0.448085252 | 0.98025 | 0.9818521738 | 0.294095 | 6.068505016 | 0 |
| 2 | 47429931 | A | C | E | D | MSH2 | ENST00000233146 | 0.44259 | 0.00599 | 0.2371 | 0.59116 | 0.76553 | 0.3027590157 | 0.96559 | 0.3154656887 | 0.182092 | 3.454925912 | 0 |
| 2 | 47429931 | A | T | E | D | MSH2 | ENST00000233146 | 0.43744 | 0.00599 | 0.2371 | 0.5947 | 0.76553 | 0.3027590157 | 0.96554 | 0.3154656887 | 0.182089 | 3.454925912 | 0 |
| 2 | 47429932 | A | C | K | Q | MSH2 | ENST00000233146 | 0.46245 | 0.00648 | 0.30247 | 0.7614 | 0.81933 | 0.2160683294 | 0.91234 | 0.8295739889 | 0.211576 | 3.585567205 | 0 |
| 2 | 47429932 | A | G | K | E | MSH2 | ENST00000233146 | 0.45573 | 0.00532 | 0.33679 | 0.73748 | 0.7675 | 0.243216175 | 0.97051 | 0.3962533772 | 0.244989 | 3.511972921 | 0 |
| 2 | 47429933 | A | C | K | T | MSH2 | ENST00000233146 | 0.42764 | 0.00763 | 0.35147 | 0.77688 | 0.84906 | 0.3182142477 | 0.91351 | 0.5939602256 | 0.216473 | 3.432578045 | 0 |
| 2 | 47429933 | A | G | K | R | MSH2 | ENST00000233146 | 0.37312 | 0.00608 | 0.28143 | 0.72536 | 0.69594 | 0.1497110575 | 0.95088 | 0.3596110756 | 0.138017 | 1.409054294 | 0 |
| 2 | 47429933 | A | T | K | I | MSH2 | ENST00000233146 | 0.58953 | 0.01604 | 0.32446 | 0.89194 | 0.91736 | 0.3899371691 | 0.86644 | 0.9755342007 | 0.24914 | 6.087609295 | 0 |
| 2 | 47429935 | C | G | H | D | MSH2 | ENST00000233146 | 0.35836 | 0.00687 | 0.26201 | 0.81197 | 0.84959 | 0.5895696725 | 0.93818 | 0.6128479838 | 0.185817 | 3.134886695 | 0 |
| 2 | 47429935 | C | T | H | Y | MSH2 | ENST00000233146 | 0.23699 | 0.00547 | 0.23353 | 0.67993 | 0.72926 | 0.3192068914 | 0.83387 | 0.02840566571 | 0.0227937 | -0.8701009194 | 0 |
| 2 | 47429936 | A | C | H | P | MSH2 | ENST00000233146 | 0.59053 | 0.01315 | 0.24393 | 0.86339 | 0.85534 | 0.637839751 | 0.89162 | 0.9679549336 | 0.24262 | 3.940941263 | 0 |
| 2 | 47429936 | A | G | H | R | MSH2 | ENST00000233146 | 0.5123 | 0.01009 | 0.2606 | 0.73257 | 0.81414 | 0.4073681039 | 0.92598 | 0.8263409138 | 0.219216 | 3.720094472 | 0 |
| 2 | 47429936 | A | T | H | L | MSH2 | ENST00000233146 | 0.51896 | 0.00654 | 0.22777 | 0.7627 | 0.79334 | 0.4698119507 | 0.94693 | 0.7664489746 | 0.210141 | 2.20890013 | 0 |
| 2 | 47429938 | G | A | E | K | MSH2 | ENST00000233146 | 0.40301 | 0.00566 | 0.16346 | 0.75079 | 0.82668 | 0.3373652398 | 0.93143 | 0.73072505 | 0.202173 | 3.414522389 | 0 |
| 2 | 47429938 | G | C | E | Q | MSH2 | ENST00000233146 | 0.36197 | 0.0058 | 0.08721 | 0.67059 | 0.8124 | 0.2870741793 | 0.96565 | 0.2767286003 | 0.2008 | 1.77103602 | 0 |
| 2 | 47429939 | A | C | E | A | MSH2 | ENST00000233146 | 0.31608 | 0.00569 | 0.09998 | 0.71985 | 0.80417 | 0.2848369508 | 0.95459 | 0.5193296075 | 0.178939 | 3.982206827 | 0 |
| 2 | 47429939 | A | G | E | G | MSH2 | ENST00000233146 | 0.38038 | 0.00547 | 0.07773 | 0.79079 | 0.83725 | 0.3652838088 | 0.92436 | 0.6745920181 | 0.200491 | 2.457256633 | 0 |
| 2 | 47429939 | A | T | E | V | MSH2 | ENST00000233146 | 0.37736 | 0.00855 | 0.08161 | 0.85692 | 0.848 | 0.4132003848 | 0.97408 | 0.8132549524 | 0.22327 | 4.599215992 | 0 |
| 2 | 47429940 | A | C | E | D | MSH2 | ENST00000233146 | 0.39672 | 0.00592 | 0.09675 | 0.65205 | 0.77011 | 0.1998165946 | 0.9414 | 0.1761632714 | 0.177222 | 2.305220868 | 0 |
| 2 | 47429940 | A | T | E | D | MSH2 | ENST00000233146 | 0.39185 | 0.00592 | 0.09675 | 0.6601 | 0.77011 | 0.1998165946 | 0.94154 | 0.1907264768 | 0.177717 | 2.305220868 | 0 |
| 2 | 47429941 | G | A | G | R | MSH2 | ENST00000233146 | 0.68371 | 0.02679 | 0.37433 | 0.87703 | 0.89239 | 0.626169722 | 0.94976 | 0.9901408553 | 0.394794 | 6.898178892 | 0 |
| 2 | 47429941 | G | C | G | R | MSH2 | ENST00000233146 | 0.68051 | 0.02679 | 0.37433 | | 0.89239 | 0.626169722 | 0.94936 | 0.9901408553 | 0.394797 | 6.898178892 | 0 |
| 2 | 47445548 | G | A | G | E | MSH2 | ENST00000233146 | 0.42091 | 0.00696 | 0.34825 | 0.7997 | 0.90052 | 0.5824573326 | 0.94736 | 0.9589352012 | 0.363117 | 4.418187637 | 0 |
| 2 | 47445548 | G | C | G | A | MSH2 | ENST00000233146 | 0.4286 | 0.0167 | 0.27204 | 0.79801 | 0.86302 | 0.518910054 | 0.9051 | 0.9587358832 | 0.261342 | 3.956986332 | 0 |
| 2 | 47445548 | G | T | G | V | MSH2 | ENST00000233146 | 0.46861 | 0.00705 | 0.27062 | 0.86212 | 0.9528 | 0.5924724551 | 0.9307 | 0.9872456789 | 0.438328 | 5.593518042 | 0 |
| 2 | 47445550 | A | G | K | E | MSH2 | ENST00000233146 | 0.23857 | 0.00517 | 0.19332 | 0.30863 | 0.76553 | 0.2262298794 | 0.97984 | 0.4716933072 | 0.156113 | 1.642265752 | 0 |
| 2 | 47445551 | A | C | K | T | MSH2 | ENST00000233146 | 0.28256 | 0.00543 | 0.22064 | 0.31146 | 0.80238 | 0.2439139533 | 0.96911 | 0.09138980508 | 0.111986 | 1.329935922 | 0 |
| 2 | 47445551 | A | G | K | R | MSH2 | ENST00000233146 | 0.28641 | 0.00608 | 0.1461 | 0.34408 | 0.79577 | 0.1807423348 | 0.95793 | 0.1711273789 | 0.0906417 | 2.908312175 | 0 |
| 2 | 47445551 | A | T | K | I | MSH2 | ENST00000233146 | 0.34298 | 0.00603 | 0.18254 | 0.62234 | 0.82836 | 0.343098989 | 0.95893 | 0.6028981209 | 0.161634 | 6.072239055 | 0 |
| 2 | 47445552 | A | C | K | N | MSH2 | ENST00000233146 | 0.39417 | 0.00543 | 0.21323 | 0.47736 | 0.78411 | 0.2314188878 | 0.96489 | 0.3173699975 | 0.0909076 | 0.9994404619 | 0 |
| 2 | 47445552 | A | T | K | N | MSH2 | ENST00000233146 | 0.38932 | 0.00543 | 0.21323 | 0.52141 | 0.78411 | 0.2314188878 | 0.9658 | 0.3173699975 | 0.0949586 | 0.9994404619 | 0 |
| 2 | 47445553 | C | A | H | N | MSH2 | ENST00000233146 | 0.36147 | 0.00562 | 0.48261 | 0.70985 | 0.84159 | 0.4068870679 | 0.88753 | 0.9206370115 | 0.204952 | 2.985960546 | 0 |
| 2 | 47445553 | C | G | H | D | MSH2 | ENST00000233146 | 0.41884 | 0.00808 | 0.4774 | 0.7996 | 0.9252 | 0.6348781386 | 0.96439 | 0.8539948463 | 0.409345 | 7.658150922 | 0 |
| 2 | 47445553 | C | T | H | Y | MSH2 | ENST00000233146 | 0.34456 | 0.00547 | 0.48545 | 0.68856 | 0.78223 | 0.4561121952 | 0.93904 | 0.8105466366 | 0.23051 | 3.998044764 | 0 |
| 2 | 47445554 | A | C | H | P | MSH2 | ENST00000233146 | 0.77734 | 0.01485 | 0.41931 | 0.83111 | 0.94431 | 0.7321073018 | 0.9663 | 0.9935817719 | 0.427872 | 7.658150922 | 0 |
| 2 | 47445554 | A | G | H | R | MSH2 | ENST00000233146 | 0.59233 | 0.00556 | 0.48691 | 0.72638 | 0.92397 | 0.5138041263 | 0.92465 | 0.9552599788 | 0.358238 | 3.245492212 | 0 |
| 2 | 47445554 | A | T | H | L | MSH2 | ENST00000233146 | 0.75224 | 0.00836 | 0.44658 | 0.84633 | 0.94618 | 0.5598030169 | 0.9689 | 0.9712178707 | 0.416442 | 4.044491928 | 0 |
| 2 | 47445555 | C | A | H | Q | MSH2 | ENST00000233146 | 0.48152 | 0.0109 | 0.45723 | 0.73113 | 0.86603 | 0.3400450417 | 0.93431 | 0.903631568 | 0.185156 | 7.658150922 | 0 |
| 2 | 47445555 | C | G | H | Q | MSH2 | ENST00000233146 | 0.48825 | 0.0109 | 0.45723 | 0.72792 | 0.86603 | 0.3400450417 | 0.93412 | 0.8900739551 | 0.242716 | 7.658150922 | 0 |
| 2 | 47445556 | C | A | Q | K | MSH2 | ENST00000233146 | 0.29138 | 0.00535 | 0.27332 | 0.68485 | 0.76685 | 0.4779170308 | 0.97684 | 0.6839348078 | 0.299116 | 2.060838378 | 0 |
| 2 | 47445556 | C | G | Q | E | MSH2 | ENST00000233146 | 0.26855 | 0.00556 | 0.22767 | 0.55325 | 0.7609 | 0.4134607538 | 0.95424 | 0.5280215144 | 0.255119 | 3.457219814 | 0 |
| 2 | 47445557 | A | C | Q | P | MSH2 | ENST00000233146 | 0.58382 | 0.01075 | 0.24293 | 0.81901 | 0.93515 | 0.7339591204 | 0.97791 | 0.9438509345 | 0.437792 | 7.162777396 | 0 |
| 2 | 47445557 | A | G | Q | R | MSH2 | ENST00000233146 | 0.3922 | 0.00562 | 0.26037 | 0.65069 | 0.8345 | 0.4790629874 | 0.9911 | 0.7756100893 | 0.367748 | 2.389959629 | 0 |
| 2 | 47445557 | A | T | Q | L | MSH2 | ENST00000233146 | 0.45862 | 0.00681 | 0.21399 | 0.75614 | 0.8933 | 0.5053315524 | 0.96913 | 0.9433086514 | 0.366583 | 3.390637202 | 0 |
| 2 | 47445558 | G | C | Q | H | MSH2 | ENST00000233146 | 0.27841 | 0.01505 | 0.26284 | 0.7657 | 0.90274 | 0.4432697691 | 0.97218 | 0.9503231645 | 0.311167 | 3.236103407 | 0 |
| 2 | 47445558 | G | T | Q | H | MSH2 | ENST00000233146 | 0.2745 | 0.01505 | 0.26284 | 0.7657 | 0.90274 | 0.4432697691 | 0.97233 | 0.9503231645 | 0.311167 | 3.236103407 | 0 |
| 2 | 47445559 | A | C | K | Q | MSH2 | ENST00000233146 | 0.175 | 0.00569 | 0.0484 | 0.62229 | 0.85894 | 0.1942550797 | 0.96954 | 0.05166134969 | 0.0624127 | 0.9314389938 | 0 |
| 2 | 47445559 | A | G | K | E | MSH2 | ENST00000233146 | 0.1607 | 0.0054 | 0.06006 | 0.55178 | 0.84106 | 0.2133462642 | 0.98464 | 0.06086681411 | 0.0539062 | 0.3115130282 | 0 |
| 2 | 47445560 | A | C | K | T | MSH2 | ENST00000233146 | 0.11606 | 0.0054 | 0.06089 | 0.33833 | 0.76356 | 0.2215640463 | 0.83737 | 0.03310628059 | 0.0448128 | -0.4592354375 | 0 |
| 2 | 47445560 | A | G | K | R | MSH2 | ENST00000233146 | 0.15817 | 0.00687 | 0.05696 | 0.48901 | 0.78286 | 0.1953866043 | 0.90606 | 0.06402401618 | 0.0666502 | 1.046250201 | 0 |
| 2 | 47445560 | A | T | K | I | MSH2 | ENST00000233146 | 0.1887 | 0.00509 | 0.06663 | 0.55009 | 0.76289 | 0.247627354 | 0.91754 | 0.07739273676 | 0.0860128 | 2.107544997 | 0 |
| 2 | 47445561 | A | C | K | N | MSH2 | ENST00000233146 | 0.22484 | 0.00543 | 0.06359 | 0.40783 | 0.75143 | 0.2345524316 | 0.90175 | 0.0340455962 | 0.0232795 | 1.001435514 | 0 |
| 2 | 47445561 | A | T | K | N | MSH2 | ENST00000233146 | 0.22143 | 0.00543 | 0.06359 | 0.40783 | 0.75143 | 0.2345524316 | 0.90248 | 0.03579158396 | 0.0232865 | 1.001435514 | 0 |
| 2 | 47445562 | T | A | L | I | MSH2 | ENST00000233146 | 0.38559 | 0.01888 | 0.42478 | 0.78126 | 0.84588 | 0.2583850245 | 0.82155 | 0.9010534883 | 0.186003 | 5.779543164 | 0 |
| 2 | 47445562 | T | G | L | V | MSH2 | ENST00000233146 | 0.34844 | 0.00935 | 0.34954 | 0.7769 | 0.85689 | 0.4121534419 | 0.86392 | 0.6038036346 | 0.211108 | 5.680120377 | 0 |
| 2 | 47445563 | T | C | L | S | MSH2 | ENST00000233146 | 0.8498 | 0.02068 | 0.43151 | 0.85509 | 0.94167 | 0.6215723035 | 0.96847 | 0.9920210242 | 0.449927 | 5.421246648 | 0 |
| 2 | 47445565 | T | A | L | M | MSH2 | ENST00000233146 | 0.39626 | 0.01108 | 0.37801 | 0.70367 | 0.86653 | 0.3950276595 | 0.84025 | 0.9248481989 | 0.134298 | 4.862086111 | 0 |
| 2 | 47445565 | T | G | L | V | MSH2 | ENST00000233146 | 0.27369 | 0.00866 | 0.34064 | 0.66393 | 0.87925 | 0.4268144433 | 0.92572 | 0.243377924 | 0.169498 | 3.990791232 | 0 |
| 2 | 47445566 | T | C | L | S | MSH2 | ENST00000233146 | 0.84423 | 0.00687 | 0.42632 | 0.85491 | 0.94281 | 0.7142143213 | 0.96413 | 0.9945411086 | 0.46994 | 8.341056519 | 0 |
| 2 | 47445566 | T | G | L | W | MSH2 | ENST00000233146 | 0.85033 | 0.00627 | 0.49373 | 0.86925 | 0.93397 | 0.758320879 | 0.88855 | 0.9959812164 | 0.359311 | 8.341056519 | 0 |
| 2 | 47445567 | G | C | L | F | MSH2 | ENST00000233146 | 0.38932 | 0.00669 | 0.38981 | 0.81632 | 0.85586 | 0.535494442 | 0.9318 | 0.9690483809 | 0.195597 | 7.939942156 | 0 |
| 2 | 47445567 | G | T | L | F | MSH2 | ENST00000233146 | 0.38467 | 0.00669 | 0.38981 | 0.81474 | 0.85586 | 0.535494442 | 0.93214 | 0.9690483809 | 0.195594 | 7.939942156 | 0 |
| 2 | 47445568 | T | A | L | M | MSH2 | ENST00000233146 | 0.28896 | 0.00592 | 0.28383 | 0.54227 | 0.76946 | 0.229052969 | 0.90661 | 0.08990001678 | 0.089573 | 1.347078648 | 0 |
| 2 | 47445568 | T | G | L | V | MSH2 | ENST00000233146 | 0.2548 | 0.00574 | 0.25091 | 0.52044 | 0.79212 | 0.2368965371 | 0.95319 | 0.1431755573 | 0.124917 | 3.123594938 | 0 |
| 2 | 47445569 | T | C | L | S | MSH2 | ENST00000233146 | 0.29617 | 0.00522 | 0.33019 | 0.49784 | 0.8233 | 0.3618945919 | 0.98681 | 0.2802387774 | 0.333384 | 3.638280724 | 0 |
| 2 | 47445569 | T | G | L | W | MSH2 | ENST00000233146 | 0.64633 | 0.01604 | 0.39719 | 0.77843 | 0.90229 | 0.4581746692 | 0.83208 | 0.9411389828 | 0.24038 | 6.466653331 | 0 |
| 2 | 47445570 | G | C | L | F | MSH2 | ENST00000233146 | 0.49261 | 0.00566 | 0.29827 | 0.63006 | 0.862 | 0.2281854214 | 0.81235 | 0.7099663019 | 0.209547 | 5.841759119 | 0 |
| 2 | 47445570 | G | T | L | F | MSH2 | ENST00000233146 | 0.48742 | 0.00566 | 0.29827 | 0.63549 | 0.862 | 0.2281854214 | 0.81337 | 0.7099663019 | 0.209535 | 5.841759119 | 0 |
| 2 | 47445571 | G | A | A | T | MSH2 | ENST00000233146 | 0.3793 | 0.00608 | 0.44598 | 0.78338 | 0.8705 | 0.3129266387 | 0.88272 | 0.8927301168 | 0.328726 | 3.685432977 | 0 |
| 2 | 47445571 | G | C | A | P | MSH2 | ENST00000233146 | 0.58268 | 0.01649 | 0.51793 | 0.87156 | 0.91862 | 0.7484276715 | 0.91002 | 0.9657600522 | 0.433608 | 7.607190199 | 0 |
| 2 | 47445571 | G | T | A | S | MSH2 | ENST00000233146 | 0.34347 | 0.00622 | 0.40217 | 0.76275 | 0.86047 | 0.2871076291 | 0.80195 | 0.8505029678 | 0.246238 | 3.559344621 | 0 |
| 2 | 47445572 | C | A | A | E | MSH2 | ENST00000233146 | 0.23771 | 0.00551 | 0.51831 | 0.73919 | 0.8233 | 0.6218829091 | 0.92846 | 0.717245698 | 0.391082 | 1.350434523 | 0 |
| 2 | 47445572 | C | G | A | G | MSH2 | ENST00000233146 | 0.30802 | 0.00586 | 0.42549 | 0.81639 | 0.90318 | 0.3193244272 | 0.85495 | 0.9317771792 | 0.331704 | 4.006564986 | 0 |
| 2 | 47445572 | C | T | A | V | MSH2 | ENST00000233146 | 0.34124 | 0.00965 | 0.4679 | 0.81131 | 0.90842 | 0.4325762716 | 0.86193 | 0.7296528816 | 0.353256 | 4.517221325 | 0 |
| 2 | 47445574 | G | A | V | I | MSH2 | ENST00000233146 | 0.21581 | 0.00615 | 0.09878 | 0.35506 | 0.77076 | 0.1768967321 | 0.94027 | 0.191066429 | 0.109663 | 2.354086026 | 0 |
| 2 | 47445574 | G | C | V | L | MSH2 | ENST00000233146 | 0.24738 | 0.0058 | 0.19603 | 0.46744 | 0.79456 | 0.3787349421 | 0.97956 | 0.1263088584 | 0.201828 | 1.928445273 | 0 |
| 2 | 47445575 | T | A | V | D | MSH2 | ENST00000233146 | 0.68515 | 0.01351 | 0.21329 | 0.82149 | 0.91862 | 0.7444953664 | 0.90967 | 0.9767382741 | 0.295731 | 2.938702616 | 0 |
| 2 | 47445575 | T | C | V | A | MSH2 | ENST00000233146 | 0.37177 | 0.00543 | 0.2144 | 0.47246 | 0.78906 | 0.3646121679 | 0.93224 | 0.6589068174 | 0.198387 | 2.404225479 | 0 |
| 2 | 47445575 | T | G | V | G | MSH2 | ENST00000233146 | 0.40851 | 0.00543 | 0.14124 | 0.65577 | 0.87684 | 0.5384154161 | 0.92814 | 0.8452355862 | 0.27811 | 3.350161642 | 0 |
| 2 | 47445577 | T | A | F | I | MSH2 | ENST00000233146 | 0.59828 | 0.02249 | 0.59325 | 0.82832 | 0.92804 | 0.6214000278 | 0.81681 | 0.9901491404 | 0.271699 | 5.305142456 | 0 |
| 2 | 47445577 | T | C | F | L | MSH2 | ENST00000233146 | 0.50419 | 0.00788 | 0.63761 | 0.80246 | 0.92275 | 0.558026223 | 0.79882 | 0.9797177911 | 0.250605 | 4.987381023 | 0 |
| 2 | 47445577 | T | G | F | V | MSH2 | ENST00000233146 | 0.77893 | 0.02628 | 0.57363 | 0.88675 | 0.96517 | 0.575011651 | 0.87895 | 0.9947326183 | 0.386309 | 5.774505774 | 0 |
| 2 | 47445578 | T | C | F | S | MSH2 | ENST00000233146 | 0.86112 | 0.02628 | 0.59572 | 0.92038 | 0.95717 | 0.7127767079 | 0.88241 | 0.9975377321 | 0.32994 | 5.804469139 | 0 |
| 2 | 47445578 | T | G | F | C | MSH2 | ENST00000233146 | 0.8609 | 0.03083 | 0.62586 | 0.8778 | 0.96226 | 0.6396288563 | 0.93196 | 0.9971434474 | 0.444368 | 7.713734266 | 0 |
| 2 | 47445579 | T | A | F | L | MSH2 | ENST00000233146 | 0.38593 | 0.00788 | 0.63761 | 0.79721 | 0.91986 | 0.558026223 | 0.82767 | 0.980302453 | 0.231161 | 4.987381023 | 0 |
| 2 | 47445579 | T | G | F | L | MSH2 | ENST00000233146 | 0.39179 | 0.00788 | 0.63761 | 0.79475 | 0.91986 | 0.558026223 | 0.82791 | 0.9796332121 | 0.231163 | 4.987381023 | 0 |
| 2 | 47445580 | G | A | V | M | MSH2 | ENST00000233146 | 0.54786 | 0.01365 | 0.47844 | 0.55783 | 0.79089 | 0.2904566759 | 0.84606 | 0.7659979463 | 0.197029 | 2.711254387 | 0 |
| 2 | 47445580 | G | C | V | L | MSH2 | ENST00000233146 | 0.31764 | 0.00574 | 0.44495 | 0.39055 | 0.70671 | 0.2743798056 | 0.94703 | 0.3773491085 | 0.0862471 | 1.177605839 | 0 |
| 2 | 47445580 | G | T | V | L | MSH2 | ENST00000233146 | 0.31343 | 0.00574 | 0.44495 | 0.41369 | 0.70671 | 0.2743798056 | 0.94671 | 0.6565946341 | 0.1563 | 1.177605839 | 0 |
| 2 | 47445581 | T | A | V | E | MSH2 | ENST00000233146 | 0.39035 | 0.00898 | 0.44968 | 0.693 | 0.90052 | 0.5961405767 | 0.97543 | 0.6067175865 | 0.267275 | 4.545426727 | 0 |
| 2 | 47445581 | T | C | V | A | MSH2 | ENST00000233146 | 0.31069 | 0.00543 | 0.47272 | 0.36914 | 0.8124 | 0.3453041998 | 0.96675 | 0.1499689668 | 0.178483 | 2.065669147 | 0 |
| 2 | 47445581 | T | G | V | G | MSH2 | ENST00000233146 | 0.37493 | 0.00763 | 0.35841 | 0.69257 | 0.87588 | 0.531909043 | 0.96722 | 0.8946675658 | 0.270711 | 2.920109517 | 0 |
| 2 | 47445583 | A | C | T | P | MSH2 | ENST00000233146 | 0.4215 | 0.00527 | 0.29597 | 0.71818 | 0.91186 | 0.5037233159 | 0.92335 | 0.9486892223 | 0.271364 | 5.702432507 | 0 |
| 2 | 47445583 | A | G | T | A | MSH2 | ENST00000233146 | 0.31743 | 0.0058 | 0.28723 | 0.55621 | 0.7528 | 0.2491764577 | 0.95149 | 0.7807002664 | 0.200532 | 3.245604419 | 0 |
| 2 | 47445583 | A | T | T | S | MSH2 | ENST00000233146 | 0.29467 | 0.00592 | 0.29954 | 0.54051 | 0.71861 | 0.1730023111 | 0.92169 | 0.5361206532 | 0.159738 | 0.9378922489 | 0 |
| 2 | 47445584 | C | A | T | N | MSH2 | ENST00000233146 | 0.18208 | 0.00547 | 0.36717 | 0.5815 | 0.68908 | 0.2480926498 | 0.8186 | 0.7579315901 | 0.115621 | 0.8746228433 | 0 |
| 2 | 47445584 | C | G | T | S | MSH2 | ENST00000233146 | 0.17917 | 0.00592 | 0.29954 | 0.59376 | 0.69339 | 0.1730023111 | 0.85313 | 0.7950780988 | 0.157975 | 0.9378922489 | 0 |
| 2 | 47445584 | C | T | T | I | MSH2 | ENST00000233146 | 0.26539 | 0.00687 | 0.43227 | 0.66828 | 0.85534 | 0.3530668631 | 0.82866 | 0.9275615811 | 0.253615 | 3.743951413 | 0 |
| 2 | 47445586 | C | A | P | T | MSH2 | ENST00000233146 | 0.6072 | 0.03538 | 0.465 | 0.8916 | 0.91652 | 0.5443960937 | 0.92638 | 0.9951386452 | 0.45165 | 5.339863152 | 0 |

| | | | | | | | | | | | | | | | | | |
|---|---|---|---|---|---|---|---|---|---|---|---|---|---|---|---|---|---|
| 2 | 47445586 | C | G | P | A | MSH2 | ENST00000233146 | 0.5876 | 0.03747 | 0.41667 | 0.87224 | 0.93201 | 0.467142601 | 0.84866 | 0.9448879957 | 0.22297 | 4.754159585 | 0 |
| 2 | 47445586 | C | T | P | S | MSH2 | ENST00000233146 | 0.62696 | 0.0368 | 0.42044 | 0.82751 | 0.90624 | 0.5619854229 | 0.8902 | 0.9775868058 | 0.380901 | 4.439924145 | 0 |
| 2 | 47445587 | C | A | P | H | MSH2 | ENST00000233146 | 0.7254 | 0.03497 | 0.50215 | 0.88535 | 0.94913 | 0.5804930322 | 0.87426 | 0.9924936295 | 0.456638 | 6.476289486 | 0 |
| 2 | 47445587 | C | G | P | R | MSH2 | ENST00000233146 | 0.72941 | 0.03626 | 0.51698 | 0.87381 | 0.94986 | 0.6471703981 | 0.90657 | 0.9951386452 | 0.462007 | 5.023037112 | 0 |
| 2 | 47445587 | C | T | P | L | MSH2 | ENST00000233146 | 0.74802 | 0.0368 | 0.52057 | 0.87663 | 0.95535 | 0.6409128125 | 0.93443 | 0.9814528263 | 0.356641 | 6.643831173 | 0 |
| 2 | 47445589 | C | A | L | I | MSH2 | ENST00000233146 | 0.43142 | 0.00696 | 0.22803 | 0.63372 | 0.84853 | 0.3327687163 | 0.76079 | 0.8330113888 | 0.151265 | 2.559880915 | 0 |
| 2 | 47445589 | C | G | L | V | MSH2 | ENST00000233146 | 0.5702 | 0.00812 | 0.16872 | 0.81976 | 0.91229 | 0.5179838831 | 0.8898 | 0.9833759665 | 0.307607 | 3.183120397 | 0 |
| 2 | 47445590 | T | A | L | H | MSH2 | ENST00000233146 | 0.72276 | 0.02414 | 0.28795 | 0.90397 | 0.96773 | 0.7472625869 | 0.91744 | 0.996742785 | 0.420059 | 6.257595918 | 1 |
| 2 | 47445590 | T | C | L | P | MSH2 | ENST00000233146 | 0.74754 | 0.02628 | 0.34093 | 0.92314 | 0.99039 | 0.8866284164 | 0.96852 | 0.9966984391 | 0.530146 | 7.78361185 | 1 |
| 2 | 47445590 | T | G | L | R | MSH2 | ENST00000233146 | 0.743 | 0.0262 | 0.2923 | 0.90761 | 0.98657 | 0.8611512315 | 0.94078 | 0.997320354 | 0.492398 | 5.292731673 | 1 |
| 2 | 47445592 | A | C | T | P | MSH2 | ENST00000233146 | 0.25394 | 0.00735 | 0.12758 | 0.64295 | 0.82948 | 0.4528584027 | 0.99788 | 0.06411779329 | 0.2176 | 4.778470781 | 0 |
| 2 | 47445592 | A | G | T | A | MSH2 | ENST00000233146 | 0.20455 | 0.00586 | 0.11138 | 0.41904 | 0.80713 | 0.1352815473 | 0.98014 | 0.08527670801 | 0.0976397 | 1.594154772 | 0 |
| 2 | 47445592 | A | T | T | S | MSH2 | ENST00000233146 | 0.18126 | 0.00592 | 0.11776 | 0.40062 | 0.80713 | 0.1375150025 | 0.96432 | 0.07604215294 | 0.0849658 | 1.255246227 | 0 |
| 2 | 47445593 | C | A | T | N | MSH2 | ENST00000233146 | 0.20602 | 0.00547 | 0.15233 | 0.43405 | 0.7528 | 0.1893696262 | 0.89691 | 0.05496713147 | 0.034696 | -0.2402578783 | 0 |
| 2 | 47445593 | C | G | T | S | MSH2 | ENST00000233146 | 0.19831 | 0.00592 | 0.11776 | 0.43989 | 0.80948 | 0.1375150025 | 0.96691 | 0.07025887817 | 0.0556466 | 1.255246227 | 0 |
| 2 | 47445593 | C | T | T | I | MSH2 | ENST00000233146 | 0.24699 | 0.00556 | 0.15947 | 0.38515 | 0.71548 | 0.1329896616 | 0.947 | 0.1975623071 | 0.0646014 | 0.8205985442 | 0 |
| 2 | 47445595 | G | A | D | N | MSH2 | ENST00000233146 | 0.49596 | 0.00627 | 0.45664 | 0.78972 | 0.81472 | 0.4675607309 | 0.87935 | 0.9221608639 | 0.114262 | 4.425451713 | 0 |
| 2 | 47445595 | G | C | D | H | MSH2 | ENST00000233146 | 0.82342 | 0.02442 | 0.47839 | 0.82898 | 0.93436 | 0.5951497645 | 0.97066 | 0.9866951704 | 0.442241 | 5.943953041 | 0 |
| 2 | 47445595 | G | T | D | Y | MSH2 | ENST00000233146 | 0.83375 | 0.02403 | 0.46812 | 0.82371 | 0.95608 | 0.6561907837 | 0.9831 | 0.9954751134 | 0.464988 | 6.833906152 | 0 |
| 2 | 47445596 | A | C | D | A | MSH2 | ENST00000233146 | 0.68755 | 0.00619 | 0.40333 | 0.74984 | 0.91314 | 0.5822648548 | 0.92844 | 0.9424923658 | 0.382633 | 4.516419651 | 0 |
| 2 | 47445596 | A | T | D | V | MSH2 | ENST00000233146 | 0.84043 | 0.01408 | 0.39298 | 0.8808 | 0.93747 | 0.6536186574 | 0.96977 | 0.9949982166 | 0.439924 | 5.919152571 | 0 |
| 2 | 47445597 | T | A | D | E | MSH2 | ENST00000233146 | 0.26678 | 0.00592 | 0.42714 | 0.43251 | 0.76289 | 0.2964776476 | 0.94633 | 0.2490714788 | 0.176602 | 1.158755478 | 0 |
| 2 | 47445597 | T | G | D | E | MSH2 | ENST00000233146 | 0.27161 | 0.00592 | 0.42714 | 0.44059 | 0.76289 | 0.2964776476 | 0.94639 | 0.2490714788 | 0.176574 | 1.158755478 | 0 |
| 2 | 47445598 | C | G | L | V | MSH2 | ENST00000233146 | 0.51942 | 0.00898 | 0.34158 | 0.69372 | 0.85274 | 0.4082560739 | 0.91848 | 0.9344694614 | 0.263969 | 5.455057482 | 0 |
| 2 | 47445599 | T | A | L | H | MSH2 | ENST00000233146 | 0.63788 | 0.03019 | 0.5124 | 0.8364 | 0.94431 | 0.6688592127 | 0.89497 | 0.9855529666 | 0.342294 | 5.383565352 | 0 |
| 2 | 47445599 | T | C | L | P | MSH2 | ENST00000233146 | 0.79958 | 0.0262 | 0.56624 | 0.90619 | 0.97698 | 0.8487971952 | 0.94231 | 0.9953029156 | 0.470643 | 8.130643918 | 1 |
| 2 | 47445599 | T | G | L | R | MSH2 | ENST00000233146 | 0.79112 | 0.0194 | 0.51153 | 0.87346 | 0.96044 | 0.7991031091 | 0.89334 | 0.993829906 | 0.394745 | 4.363870315 | 0 |
| 2 | 47445601 | C | A | R | S | MSH2 | ENST00000233146 | 0.33613 | 0.00556 | 0.1792 | 0.22581 | 0.70914 | 0.2987582135 | 0.94273 | 0.8307970166 | 0.155348 | 1.883165748 | 0 |
| 2 | 47445601 | C | G | R | G | MSH2 | ENST00000233146 | 0.38364 | 0.00535 | 0.17843 | 0.49032 | 0.7609 | 0.2792053414 | 0.97514 | 0.5932536125 | 0.215818 | 3.003543216 | 0 |
| 2 | 47445601 | C | T | R | C | MSH2 | ENST00000233146 | 0.44807 | 0.00527 | 0.25338 | 0.35183 | 0.71705 | 0.3213627193 | 0.97854 | 0.7544518113 | 0.187378 | 1.595000094 | 0 |
| 2 | 47445602 | G | A | R | H | MSH2 | ENST00000233146 | 0.29647 | 0.00535 | 0.18508 | 0.3565 | 0.77076 | 0.235171894 | 0.9855 | 0.4734270275 | 0.157984 | 2.950470809 | 0 |
| 2 | 47445602 | G | C | R | P | MSH2 | ENST00000233146 | 0.47382 | 0.00952 | 0.21298 | 0.58704 | 0.84159 | 0.6665708937 | 0.98079 | 0.6583163738 | 0.264335 | 2.980652513 | 0 |
| 2 | 47445602 | G | T | R | L | MSH2 | ENST00000233146 | 0.27779 | 0.00535 | 0.19991 | 0.36125 | 0.75485 | 0.3001833778 | 0.99023 | 0.1758965701 | 0.0463865 | -1.27165319 | 0 |
| 2 | 47445604 | T | A | S | T | MSH2 | ENST00000233146 | 0.36175 | 0.00599 | 0.4615 | 0.58704 | 0.73661 | 0.198920114 | 0.97804 | 0.570230186 | 0.255093 | 2.839577837 | 0 |
| 2 | 47445604 | T | C | S | P | MSH2 | ENST00000233146 | 0.64742 | 0.01133 | 0.47484 | 0.78863 | 0.92151 | 0.645541629 | 0.9831 | 0.909951508 | 0.420655 | 4.087984192 | 0 |
| 2 | 47445604 | T | G | S | A | MSH2 | ENST00000233146 | 0.3801 | 0.00615 | 0.43608 | 0.55532 | 0.69846 | 0.1729851248 | 0.97162 | 0.7589767575 | 0.239513 | 2.654414552 | 0 |
| 2 | 47445605 | C | A | S | Y | MSH2 | ENST00000233146 | 0.42377 | 0.00556 | 0.5383 | 0.68225 | 0.83997 | 0.397466014 | 0.99014 | 0.9266517162 | 0.421052 | 3.481024346 | 0 |
| 2 | 47445607 | G | A | D | N | MSH2 | ENST00000233146 | 0.78376 | 0.03442 | 0.64416 | 0.8639 | 0.84213 | 0.6057492196 | 0.90678 | 0.988697052 | 0.0722579 | 6.49823888 | 0 |
| 2 | 47445607 | G | C | D | H | MSH2 | ENST00000233146 | 0.79144 | 0.03313 | 0.66849 | 0.8753 | 0.93824 | 0.6839320506 | 0.96853 | 0.9962604046 | 0.429438 | 7.381955202 | 0 |
| 2 | 47445607 | G | T | D | Y | MSH2 | ENST00000233146 | 0.79966 | 0.03013 | 0.64719 | 0.87693 | 0.9648 | 0.7415015159 | 0.97758 | 0.9948550463 | 0.467612 | 6.613482071 | 0 |
| 2 | 47445608 | A | C | D | A | MSH2 | ENST00000233146 | 0.8119 | 0.02687 | 0.57334 | 0.87713 | 0.9579 | 0.6854562287 | 0.94613 | 0.9947528839 | 0.424007 | 6.197664894 | 0 |
| 2 | 47445608 | A | G | D | G | MSH2 | ENST00000233146 | 0.81826 | 0.03019 | 0.58182 | 0.90057 | 0.96517 | 0.6711815462 | 0.97145 | 0.9605276585 | 0.442338 | 6.667654468 | 0 |
| 2 | 47445608 | A | T | D | V | MSH2 | ENST00000233146 | 0.80345 | 0.02083 | 0.56581 | 0.91129 | 0.97252 | 0.7293193697 | 0.97655 | 0.9984489679 | 0.463987 | 5.974196952 | 0 |
| 2 | 47445609 | C | A | D | E | MSH2 | ENST00000233146 | 0.37391 | 0.02658 | 0.61342 | 0.79381 | 0.89648 | 0.4815075939 | 0.95417 | 0.9925149083 | 0.334074 | 5.199545244 | 0 |
| 2 | 47445609 | C | G | D | E | MSH2 | ENST00000233146 | 0.37998 | 0.02658 | 0.61342 | 0.79012 | 0.89603 | 0.4815075939 | 0.95422 | 0.9925149083 | 0.334076 | 5.199545244 | 0 |
| 2 | 47445610 | T | A | F | I | MSH2 | ENST00000233146 | 0.78193 | 0.02602 | 0.66642 | 0.8741 | 0.9579 | 0.6393529949 | 0.86109 | 0.9755900502 | 0.305553 | 5.819429425 | 0 |
| 2 | 47445610 | T | C | F | L | MSH2 | ENST00000233146 | 0.46264 | 0.0103 | 0.71815 | 0.73141 | 0.91271 | 0.5656686238 | 0.95744 | 0.9432709813 | 0.385 | 4.098355625 | 0 |
| 2 | 47445610 | T | G | F | V | MSH2 | ENST00000233146 | 0.78614 | 0.02442 | 0.65248 | 0.88197 | 0.9681 | 0.5774049191 | 0.95329 | 0.9941840768 | 0.443164 | 5.261008965 | 0 |
| 2 | 47445611 | T | A | F | Y | MSH2 | ENST00000233146 | 0.77822 | 0.03231 | 0.69745 | 0.83577 | 0.94015 | 0.5301549988 | 0.9066 | 0.9848541021 | 0.329553 | 6.454842306 | 0 |
| 2 | 47445611 | T | C | F | S | MSH2 | ENST00000233146 | 0.86276 | 0.02324 | 0.67242 | 0.90496 | 0.97252 | 0.7319117531 | 0.94892 | 0.986409353 | 0.408314 | 7.802253596 | 0 |
| 2 | 47445611 | T | G | F | C | MSH2 | ENST00000233146 | 0.8203 | 0.00904 | 0.70768 | 0.87173 | 0.96444 | 0.658502183 | 0.97457 | 0.9886052012 | 0.441034 | 4.820667399 | 0 |
| 2 | 47445612 | C | A | F | L | MSH2 | ENST00000233146 | 0.37335 | 0.0103 | 0.71815 | 0.72623 | 0.88963 | 0.5656686238 | 0.96823 | 0.9567060471 | 0.361239 | 4.098355625 | 0 |
| 2 | 47445612 | C | G | F | L | MSH2 | ENST00000233146 | 0.37936 | 0.0103 | 0.71815 | 0.72416 | 0.89009 | 0.5656686238 | 0.96824 | 0.814723134 | 0.361243 | 4.098355625 | 0 |
| 2 | 47445613 | T | A | S | T | MSH2 | ENST00000233146 | 0.40471 | 0.00608 | 0.4932 | 0.58564 | 0.77399 | 0.2318098199 | 0.9792 | 0.5057569146 | 0.24249 | 3.094587003 | 0 |
| 2 | 47445613 | T | C | S | P | MSH2 | ENST00000233146 | 0.68147 | 0.01133 | 0.50962 | 0.73398 | 0.90885 | 0.7162363223 | 0.98117 | 0.9421342611 | 0.416778 | 3.816058343 | 0 |
| 2 | 47445613 | T | G | S | A | MSH2 | ENST00000233146 | 0.40763 | 0.00615 | 0.46923 | 0.55655 | 0.73661 | 0.1935245263 | 0.9608 | 0.5919252038 | 0.208295 | 1.961402416 | 0 |
| 2 | 47445614 | C | G | S | C | MSH2 | ENST00000233146 | 0.72372 | 0.0262 | 0.50014 | 0.73448 | 0.8933 | 0.2957082736 | 0.92828 | 0.8642144203 | 0.387419 | 6.600154394 | 0 |
| 2 | 47445614 | C | T | S | F | MSH2 | ENST00000233146 | 0.72981 | 0.01731 | 0.5423 | 0.83494 | 0.92275 | 0.4244209273 | 0.98568 | 0.9835885763 | 0.427165 | 7.10734755 | 0 |
| 2 | 47445616 | A | C | K | Q | MSH2 | ENST00000233146 | 0.75248 | 0.03083 | 0.59351 | 0.83727 | 0.9517 | 0.5295610954 | 0.94614 | 0.9941518989 | 0.314315 | 5.44597865 | 0 |
| 2 | 47445616 | A | G | K | E | MSH2 | ENST00000233146 | 0.7965 | 0.02556 | 0.64381 | 0.82931 | 0.96627 | 0.6042187344 | 0.96479 | 0.9966763258 | 0.463895 | 3.421880993 | 0 |
| 2 | 47445617 | A | C | K | T | MSH2 | ENST00000233146 | 0.78535 | 0.02943 | 0.66149 | 0.90067 | 0.97141 | 0.5801139156 | 0.95069 | 0.9974508882 | 0.47365 | 5.427758759 | 0 |
| 2 | 47445617 | A | G | K | R | MSH2 | ENST00000233146 | 0.77362 | 0.01044 | 0.56332 | 0.75129 | 0.90007 | 0.4554020229 | 0.93603 | 0.9747723341 | 0.296335 | 6.398977808 | 0 |
| 2 | 47445617 | A | T | K | M | MSH2 | ENST00000233146 | 0.8025 | 0.03189 | 0.56451 | 0.89319 | 0.97178 | 0.6573915904 | 0.93802 | 0.9970274568 | 0.441533 | 8.088936675 | 0 |
| 2 | 47445618 | G | C | K | N | MSH2 | ENST00000233146 | 0.46208 | 0.00935 | 0.68051 | 0.77262 | 0.85843 | 0.6019654213 | 0.86348 | 0.9387853146 | 0.111964 | 3.591163497 | 0 |
| 2 | 47445618 | G | T | K | N | MSH2 | ENST00000233146 | 0.45709 | 0.00935 | 0.68051 | 0.77262 | 0.85843 | 0.6019654213 | 0.8628 | 0.9893178344 | 0.11196 | 3.591163497 | 0 |
| 2 | 47445619 | T | A | F | I | MSH2 | ENST00000233146 | 0.85793 | 0.00627 | 0.78864 | 0.77967 | 0.9528 | 0.6391707353 | 0.90167 | 0.9889668822 | 0.327621 | 3.132300678 | 0 |
| 2 | 47445619 | T | C | F | L | MSH2 | ENST00000233146 | 0.54169 | 0.00556 | 0.81899 | 0.66037 | 0.86803 | 0.5617924494 | 0.93568 | 0.8392144442 | 0.314515 | 3.348751595 | 0 |
| 2 | 47445619 | T | G | F | V | MSH2 | ENST00000233146 | 0.86664 | 0.00844 | 0.7608 | 0.80733 | 0.95572 | 0.570008221 | 0.94946 | 0.9898838997 | 0.442134 | 2.878632048 | 0 |
| 2 | 47445620 | T | A | F | Y | MSH2 | ENST00000233146 | 0.45052 | 0.00562 | 0.80764 | 0.66211 | 0.7609 | 0.4493497497 | 0.93375 | 0.7069396377 | 0.249435 | 2.563338021 | 0 |
| 2 | 47445620 | T | C | F | S | MSH2 | ENST00000233146 | 0.91613 | 0.0134 | 0.78768 | 0.91515 | 0.97364 | 0.6959492337 | 0.96411 | 0.9957911968 | 0.453366 | 7.528637479 | 0 |
| 2 | 47445620 | T | G | F | C | MSH2 | ENST00000233146 | 0.91622 | 0.02494 | 0.80952 | 0.91343 | 0.96226 | 0.6316004671 | 0.96529 | 0.9973457456 | 0.45854 | 6.548601935 | 0 |
| 2 | 47445621 | T | A | F | L | MSH2 | ENST00000233146 | 0.38181 | 0.00556 | 0.81899 | 0.5818 | 0.85638 | 0.5617924494 | 0.95904 | 0.8210543394 | 0.298444 | 3.348751595 | 0 |
| 2 | 47445621 | T | G | F | L | MSH2 | ENST00000233146 | 0.38771 | 0.00556 | 0.81899 | 0.5818 | 0.85638 | 0.5617924494 | 0.95903 | 0.8210543394 | 0.298445 | 3.348751595 | 0 |
| 2 | 47445622 | C | A | Q | K | MSH2 | ENST00000233146 | 0.38352 | 0.00562 | 0.5808 | 0.6269 | 0.79212 | 0.5940183639 | 0.93752 | 0.7274678946 | 0.241973 | 4.96037804 | 0 |
| 2 | 47445622 | C | G | Q | E | MSH2 | ENST00000233146 | 0.36979 | 0.01082 | 0.52174 | 0.65612 | 0.76553 | 0.5294057719 | 0.8628 | 0.6680257916 | 0.162439 | 4.637075188 | 0 |
| 2 | 47445623 | A | C | Q | P | MSH2 | ENST00000233146 | 0.83174 | 0.02186 | 0.52969 | 0.81558 | 0.95426 | 0.8054524786 | 0.9389 | 0.9925578833 | 0.393625 | 8.415821538 | 0 |
| 2 | 47445623 | A | G | Q | R | MSH2 | ENST00000233146 | 0.49267 | 0.00608 | 0.57122 | 0.5968 | 0.81703 | 0.6211687157 | 0.96978 | 0.7828049064 | 0.258259 | 4.241756588 | 0 |
| 2 | 47445623 | A | T | Q | L | MSH2 | ENST00000233146 | 0.52281 | 0.00586 | 0.50687 | 0.61759 | 0.77011 | 0.5519604403 | 0.92779 | 0.9267843962 | 0.167941 | 4.096662558 | 0 |
| 2 | 47445624 | G | C | Q | H | MSH2 | ENST00000233146 | 0.58013 | 0.02951 | 0.57398 | 0.85505 | 0.93515 | 0.5445736161 | 0.92253 | 0.9890099168 | 0.245237 | 4.243374136 | 0 |
| 2 | 47445624 | G | T | Q | H | MSH2 | ENST00000233146 | 0.57441 | 0.02951 | 0.57398 | 0.85519 | 0.93515 | 0.5445736161 | 0.92224 | 0.9890099168 | 0.245235 | 4.243374136 | 0 |
| 2 | 47445625 | G | A | E | K | MSH2 | ENST00000233146 | 0.67286 | 0.00654 | 0.77204 | 0.85993 | 0.95972 | 0.5006898324 | 0.98988 | 0.9598232662 | 0.472462 | 4.553219234 | 0 |
| 2 | 47445625 | G | C | E | Q | MSH2 | ENST00000233146 | 0.45487 | 0.00904 | 0.6639 | 0.75855 | 0.90842 | 0.4003410342 | 0.96521 | 0.937030077 | 0.404361 | 3.795418398 | 0 |
| 2 | 47445626 | A | C | E | A | MSH2 | ENST00000233146 | 0.58631 | 0.01037 | 0.675 | 0.85252 | 0.94356 | 0.4887686473 | 0.9767 | 0.9864556789 | 0.426567 | 4.564713215 | 0 |
| 2 | 47445626 | A | G | E | G | MSH2 | ENST00000233146 | 0.80195 | 0.01436 | 0.62405 | 0.87472 | 0.96008 | 0.5028987614 | 0.97678 | 0.9979140162 | 0.47387 | 5.190785862 | 0 |
| 2 | 47445626 | A | T | E | V | MSH2 | ENST00000233146 | 0.78788 | 0.00726 | 0.64488 | 0.90396 | 0.9781 | 0.5266270939 | 0.97396 | 0.9940142035 | 0.453946 | 5.981043758 | 0 |
| 2 | 47445627 | A | C | E | D | MSH2 | ENST00000233146 | 0.32542 | 0.00627 | 0.67631 | 0.68695 | 0.86902 | 0.3517097552 | 0.98493 | 0.7548262477 | 0.321407 | 3.585354588 | 0 |
| 2 | 47445627 | A | T | E | D | MSH2 | ENST00000233146 | 0.32094 | 0.00627 | 0.67631 | 0.68761 | 0.86852 | 0.3517097552 | 0.98487 | 0.7627209425 | 0.321401 | 3.585354588 | 0 |
| 2 | 47445628 | A | C | M | L | MSH2 | ENST00000233146 | 0.38972 | 0.00592 | 0.68024 | 0.63531 | 0.81182 | 0.5822570467 | 0.95308 | 0.8296313286 | 0.323883 | 2.396329407 | 0 |
| 2 | 47445628 | A | G | M | V | MSH2 | ENST00000233146 | 0.62993 | 0.00705 | 0.61103 | 0.89275 | 0.95462 | 0.6695570048 | 0.97657 | 0.972443521 | 0.468288 | 4.870452084 | 0 |
| 2 | 47445628 | A | T | M | L | MSH2 | ENST00000233146 | 0.3849 | 0.00592 | 0.68024 | 0.63531 | 0.81182 | 0.5822570467 | 0.95336 | 0.8296313286 | 0.323882 | 2.396329407 | 0 |
| 2 | 47445629 | T | C | M | T | MSH2 | ENST00000233146 | 0.71008 | 0.01009 | 0.79361 | 0.88954 | 0.9659 | 0.7205395778 | 0.97678 | 0.993240118 | 0.476993 | 5.459939024 | 0 |
| 2 | 47445629 | T | G | M | R | MSH2 | ENST00000233146 | 0.76586 | 0.00904 | 0.74293 | 0.87568 | 0.95499 | 0.8886796809 | 0.97842 | 0.9957209229 | 0.471763 | 5.414022045 | 0 |
| 2 | 47445630 | G | A | M | I | MSH2 | ENST00000233146 | 0.69716 | 0.00599 | 0.68988 | 0.84955 | 0.93044 | 0.6910192309 | 0.94828 | 0.976290226 | 0.406934 | 4.354449783 | 0 |
| 2 | 47445630 | G | C | M | I | MSH2 | ENST00000233146 | 0.69388 | 0.00599 | 0.68988 | 0.84802 | 0.93044 | 0.6910192309 | 0.94746 | 0.9767905474 | 0.40694 | 4.354449783 | 0 |
| 2 | 47445630 | G | T | M | I | MSH2 | ENST00000233146 | 0.68731 | 0.00599 | 0.68988 | 0.84841 | 0.93044 | 0.6910192309 | 0.94768 | 0.976290226 | 0.406938 | 4.354449783 | 0 |
| 2 | 47445631 | A | C | I | L | MSH2 | ENST00000233146 | 0.42383 | 0.00627 | 0.63799 | 0.6532 | 0.86047 | 0.5329923689 | 0.92439 | 0.8670005798 | 0.267437 | 4.030035919 | 0 |
| 2 | 47445631 | A | G | I | V | MSH2 | ENST00000233146 | 0.2735 | 0.00608 | 0.5149 | 0.53565 | 0.81761 | 0.2755021587 | 0.81472 | 0.2482731245 | 0.178329 | 2.371292822 | 0 |
| 2 | 47445631 | A | T | I | L | MSH2 | ENST00000233146 | 0.41884 | 0.00627 | 0.63799 | 0.6532 | 0.86047 | 0.5329923689 | 0.92454 | 0.596064508 | 0.267435 | 4.030035919 | 0 |
| 2 | 47445632 | T | A | I | K | MSH2 | ENST00000233146 | 0.69732 | 0.01058 | 0.7244 | 0.92579 | 0.97438 | 0.8413453579 | 0.95963 | 0.9981942773 | 0.495044 | 8.757730332 | 1 |
| 2 | 47445632 | T | C | I | T | MSH2 | ENST00000233146 | 0.67271 | 0.01211 | 0.77088 | 0.88824 | 0.97141 | 0.6360888191 | 0.97027 | 0.9623117663 | 0.482482 | 7.626283829 | 0 |
| 2 | 47445632 | T | G | I | R | MSH2 | ENST00000233146 | 0.71233 | 0.00527 | 0.6947 | 0.91631 | 0.97252 | 0.8433746713 | 0.95523 | 0.9967219234 | 0.497105 | 8.757730332 | 1 |

| | | | | | | | | | | | | | | | | |
|---|---|---|---|---|---|---|---|---|---|---|---|---|---|---|---|---|
| 2 | 47445634 | G | C | E | Q | MSH2 | ENST00000233146 | 0.79934 | 0.02904 | 0.6971 | 0.89116 | 0.95753 | 0.5067759549 | 0.95712 | 0.9941796064 | 0.435224 | 5.396822367 | 0 |
| 2 | 47445635 | A | C | E | A | MSH2 | ENST00000233146 | 0.79761 | 0.02277 | 0.72104 | 0.84168 | 0.97848 | 0.5897984645 | 0.97927 | 0.9963831902 | 0.493735 | 5.220514543 | 0 |
| 2 | 47445635 | A | G | E | G | MSH2 | ENST00000233146 | 0.85308 | 0.02193 | 0.66369 | 0.8495 | 0.97401 | 0.5925438028 | 0.97476 | 0.9985414743 | 0.49391 | 5.678081631 | 0 |
| 2 | 47445635 | A | T | E | V | MSH2 | ENST00000233146 | 0.83244 | 0.02488 | 0.69015 | 0.91131 | 0.98034 | 0.5994311516 | 0.98233 | 0.9984051585 | 0.488904 | 7.481537618 | 0 |
| 2 | 47445637 | A | C | T | P | MSH2 | ENST00000233146 | 0.72989 | 0.02072 | 0.68919 | 0.87198 | 0.96117 | 0.6880858904 | 0.98185 | 0.9924351573 | 0.463164 | 8.600490157 | 1 |
| 2 | 47445637 | A | G | T | A | MSH2 | ENST00000233146 | 0.53107 | 0.00872 | 0.68092 | 0.72661 | 0.91399 | 0.3709327284 | 0.96514 | 0.4251970947 | 0.316737 | 3.786140248 | 0 |
| 2 | 47445637 | A | T | T | S | MSH2 | ENST00000233146 | 0.37725 | 0.00615 | 0.70032 | 0.75552 | 0.90885 | 0.2359203222 | 0.95024 | 0.930891037 | 0.297708 | 2.955820383 | 0 |
| 2 | 47445638 | C | G | T | R | MSH2 | ENST00000233146 | 0.46911 | 0.00648 | 0.78993 | 0.80255 | 0.87636 | 0.5790670278 | 0.9778 | 0.9775492549 | 0.430245 | 3.960748077 | 0 |
| 2 | 47445638 | C | T | T | I | MSH2 | ENST00000233146 | 0.60412 | 0.01408 | 0.82866 | 0.83757 | 0.94431 | 0.4299101443 | 0.97588 | 0.97477597 | 0.455292 | 6.183963994 | 0 |
| 2 | 47445640 | A | C | T | P | MSH2 | ENST00000233146 | 0.82981 | 0.00687 | 0.72701 | 0.88503 | 0.95935 | 0.821017507 | 0.89425 | 0.9939050078 | 0.337798 | 8.716773362 | 1 |
| 2 | 47445640 | A | G | T | A | MSH2 | ENST00000233146 | 0.7775 | 0.00763 | 0.72854 | 0.76363 | 0.93709 | 0.6244207042 | 0.94239 | 0.979880631 | 0.249828 | 3.666858402 | 1 |
| 2 | 47445640 | A | T | T | S | MSH2 | ENST00000233146 | 0.74858 | 0.01164 | 0.7385 | 0.82968 | 0.93201 | 0.4732163708 | 0.91071 | 0.9938204885 | 0.298237 | 4.126892164 | 0 |
| 2 | 47445641 | C | A | T | N | MSH2 | ENST00000233146 | 0.57519 | 0.02741 | 0.80954 | 0.90608 | 0.93593 | 0.7463994735 | 0.86985 | 0.9967795014 | 0.217133 | 7.848576018 | 0 |
| 2 | 47445641 | C | G | T | S | MSH2 | ENST00000233146 | 0.59798 | 0.01164 | 0.7385 | 0.83336 | 0.92356 | 0.4732163708 | 0.88005 | 0.9924542308 | 0.293027 | 4.126892164 | 0 |
| 2 | 47445641 | C | T | T | I | MSH2 | ENST00000233146 | 0.48494 | 0.01324 | 0.84643 | 0.88029 | 0.93709 | 0.6994595392 | 0.93199 | 0.9962931275 | 0.415906 | 8.868635034 | 0 |
| 2 | 47445643 | T | A | L | I | MSH2 | ENST00000233146 | 0.32225 | 0.0058 | 0.68309 | 0.56759 | 0.78782 | 0.3224606426 | 0.85057 | 0.4207020998 | 0.171124 | 2.338708777 | 0 |
| 2 | 47445643 | T | G | L | V | MSH2 | ENST00000233146 | 0.33839 | 0.00592 | 0.62455 | 0.65193 | 0.83615 | 0.4444375666 | 0.94716 | 0.1315458417 | 0.242329 | 2.91454887 | 0 |
| 2 | 47445644 | T | C | L | S | MSH2 | ENST00000233146 | 0.8701 | 0.00603 | 0.68948 | 0.89435 | 0.9833 | 0.8258028313 | 0.98595 | 0.9945985079 | 0.50407 | 7.631477117 | 1 |
| 2 | 47445645 | A | C | L | F | MSH2 | ENST00000233146 | 0.67112 | 0.02115 | 0.66408 | 0.84499 | 0.93436 | 0.6205234464 | 0.9372 | 0.9735873938 | 0.348008 | 6.394678202 | 0 |
| 2 | 47445645 | A | T | L | F | MSH2 | ENST00000233146 | 0.66438 | 0.02115 | 0.66408 | 0.85228 | 0.93436 | 0.6205234464 | 0.93695 | 0.9735873938 | 0.348004 | 6.394678202 | 0 |
| 2 | 47445647 | A | C | D | A | MSH2 | ENST00000233146 | 0.86368 | 0.02366 | 0.72705 | 0.85314 | 0.97327 | 0.8022316264 | 0.97137 | 0.9994736314 | 0.500737 | 5.781422111 | 0 |
| 2 | 47445647 | A | G | D | G | MSH2 | ENST00000233146 | 0.86858 | 0.02307 | 0.73448 | 0.84897 | 0.97475 | 0.7876566114 | 0.983 | 0.9990272522 | 0.506547 | 8.774866108 | 1 |
| 2 | 47445647 | A | T | D | V | MSH2 | ENST00000233146 | 0.85581 | 0.02469 | 0.71887 | 0.9041 | 0.9729 | 0.8146379927 | 0.98689 | 0.9994736314 | 0.498721 | 4.686216631 | 0 |
| 2 | 47445648 | T | A | D | E | MSH2 | ENST00000233146 | 0.63996 | 0.0084 | 0.76122 | 0.85345 | 0.95133 | 0.6249176595 | 0.97411 | 0.9964137673 | 0.421691 | 5.584680969 | 0 |
| 2 | 47445648 | T | G | D | E | MSH2 | ENST00000233146 | 0.64789 | 0.0084 | 0.76122 | 0.85292 | 0.95133 | 0.6249176595 | 0.97427 | 0.9964137673 | 0.421693 | 5.584680969 | 0 |
| 2 | 47445649 | A | C | M | L | MSH2 | ENST00000233146 | 0.3647 | 0.0058 | 0.57631 | 0.3873 | 0.71548 | 0.4364575826 | 0.88546 | 0.6770573258 | 0.159006 | 1.913085592 | 0 |
| 2 | 47445649 | A | G | M | V | MSH2 | ENST00000233146 | 0.44911 | 0.00569 | 0.4959 | 0.71462 | 0.88776 | 0.5380708235 | 0.97215 | 0.2251659053 | 0.11819 | 6.722830963 | 0 |
| 2 | 47445649 | A | T | M | L | MSH2 | ENST00000233146 | 0.36002 | 0.0058 | 0.57631 | 0.3873 | 0.71548 | 0.4364575826 | 0.88523 | 0.6770573258 | 0.158997 | 1.913085592 | 0 |
| 2 | 47445650 | T | A | M | K | MSH2 | ENST00000233146 | 0.61045 | 0.00674 | 0.67839 | 0.8223 | 0.92316 | 0.7927802074 | 0.96599 | 0.9909217954 | 0.450834 | 6.085087487 | 0 |
| 2 | 47445650 | T | C | M | T | MSH2 | ENST00000233146 | 0.55161 | 0.00551 | 0.71393 | 0.77584 | 0.89009 | 0.6362751412 | 0.96502 | 0.9883844852 | 0.439346 | 5.240922728 | 0 |
| 2 | 47445650 | T | G | M | R | MSH2 | ENST00000233146 | 0.64789 | 0.00916 | 0.65309 | 0.81024 | 0.93631 | 0.8279635755 | 0.96947 | 0.9916371703 | 0.452814 | 6.586765538 | 0 |
| 2 | 47445651 | G | A | M | I | MSH2 | ENST00000233146 | 0.48012 | 0.00566 | 0.58 | 0.71887 | 0.85638 | 0.5242448288 | 0.92366 | 0.590512792 | 0.149545 | 7.589249697 | 0 |
| 2 | 47445651 | G | C | M | I | MSH2 | ENST00000233146 | 0.47759 | 0.00566 | 0.58 | 0.71589 | 0.85638 | 0.5242448288 | 0.9232 | 0.8741495013 | 0.267293 | 7.589249697 | 0 |
| 2 | 47445651 | G | T | M | I | MSH2 | ENST00000233146 | 0.47244 | 0.00566 | 0.58 | 0.71553 | 0.85638 | 0.5242448288 | 0.92333 | 0.8741495013 | 0.267291 | 7.589249697 | 0 |
| 2 | 47445652 | G | A | D | N | MSH2 | ENST00000233146 | 0.42734 | 0.0058 | 0.50004 | 0.47073 | 0.61988 | 0.2189801734 | 0.91523 | 0.5437200069 | 0.116921 | 0.4041562342 | 0 |
| 2 | 47445652 | G | C | D | H | MSH2 | ENST00000233146 | 0.47854 | 0.00562 | 0.52964 | 0.6612 | 0.87684 | 0.3932824013 | 0.96791 | 0.9435999393 | 0.271527 | 4.486132924 | 0 |
| 2 | 47445652 | G | T | D | Y | MSH2 | ENST00000233146 | 0.79049 | 0.01108 | 0.52751 | 0.78649 | 0.93631 | 0.4809746962 | 0.96302 | 0.9920553565 | 0.339147 | 6.386162025 | 0 |
| 2 | 47445653 | A | C | D | A | MSH2 | ENST00000233146 | 0.37742 | 0.00562 | 0.44574 | 0.75727 | 0.86453 | 0.3945755289 | 0.91721 | 0.3352580369 | 0.108843 | 3.268986506 | 0 |
| 2 | 47445653 | A | G | D | G | MSH2 | ENST00000233146 | 0.50393 | 0.00638 | 0.44454 | 0.83915 | 0.91314 | 0.3852280262 | 0.96143 | 0.9789050221 | 0.281971 | 4.310790546 | 0 |
| 2 | 47445653 | A | T | D | V | MSH2 | ENST00000233146 | 0.62939 | 0.00522 | 0.42647 | 0.8934 | 0.95023 | 0.4630632212 | 0.96816 | 0.9866776466 | 0.348238 | 6.386162025 | 0 |
| 2 | 47445655 | C | A | Q | K | MSH2 | ENST00000233146 | 0.36103 | 0.00535 | 0.43561 | 0.57633 | 0.76289 | 0.5401389574 | 0.9649 | 0.7391446829 | 0.1939 | 3.25326725 | 0 |
| 2 | 47445655 | C | G | Q | E | MSH2 | ENST00000233146 | 0.3413 | 0.0058 | 0.38808 | 0.61592 | 0.74093 | 0.5067648141 | 0.93225 | 0.6216987967 | 0.154856 | 6.329822175 | 0 |
| 2 | 47445656 | A | C | Q | P | MSH2 | ENST00000233146 | 0.47043 | 0.01369 | 0.39907 | 0.84559 | 0.93901 | 0.7841134069 | 0.95026 | 0.9813007116 | 0.387818 | 2.202404506 | 0 |
| 2 | 47445656 | A | G | Q | R | MSH2 | ENST00000233146 | 0.40553 | 0.00569 | 0.42154 | 0.6936 | 0.82159 | 0.5519818543 | 0.98359 | 0.8478660583 | 0.263847 | 4.309162876 | 0 |
| 2 | 47445656 | A | T | Q | L | MSH2 | ENST00000233146 | 0.45358 | 0.0058 | 0.36634 | 0.70236 | 0.86503 | 0.5560699649 | 0.96925 | 0.9288541079 | 0.255028 | 5.772142162 | 0 |
| 2 | 47463031 | G | A | V | M | MSH2 | ENST00000233146 | 0.68363 | 0.03293 | 0.63039 | 0.88752 | 0.89239 | 0.4224606756 | 0.80334 | 0.9424949288 | 0.23257 | 5.926799366 | 0 |
| 2 | 47463031 | G | C | V | L | MSH2 | ENST00000233146 | 0.4887 | 0.01086 | 0.58821 | 0.75016 | 0.85741 | 0.3815184262 | 0.85155 | 0.7481732368 | 0.215489 | 4.297469467 | 0 |
| 2 | 47463031 | G | T | V | L | MSH2 | ENST00000233146 | 0.48354 | 0.01086 | 0.58821 | 0.76695 | 0.85741 | 0.3815184262 | 0.8522 | 0.7481732368 | 0.215486 | 4.297469467 | 0 |
| 2 | 47463032 | T | A | V | E | MSH2 | ENST00000233146 | 0.76195 | 0.0275 | 0.59599 | 0.939 | 0.96371 | 0.6785089897 | 0.90389 | 0.9978348613 | 0.329485 | 8.537700689 | 0 |
| 2 | 47463032 | T | C | V | A | MSH2 | ENST00000233146 | 0.40197 | 0.00741 | 0.61912 | 0.78155 | 0.92356 | 0.4154593302 | 0.87464 | 0.9538843632 | 0.25965 | 5.125427365 | 0 |
| 2 | 47463032 | T | G | V | G | MSH2 | ENST00000233146 | 0.74817 | 0.02679 | 0.46115 | 0.93784 | 0.97735 | 0.6005785267 | 0.96536 | 0.9955793023 | 0.474952 | 8.537700689 | 0 |
| 2 | 47463034 | G | A | E | K | MSH2 | ENST00000233146 | 0.8759 | 0.00687 | 0.82013 | 0.84183 | 0.96773 | 0.3432024645 | 0.97109 | 0.9796216488 | 0.421674 | 6.91458052 | 0 |
| 2 | 47463035 | A | C | E | A | MSH2 | ENST00000233146 | 0.52314 | 0.00638 | 0.72681 | 0.83713 | 0.9182 | 0.3997277411 | 0.90969 | 0.9647075534 | 0.271865 | 4.137134388 | 0 |
| 2 | 47463035 | A | G | E | G | MSH2 | ENST00000233146 | 0.7266 | 0.00547 | 0.67232 | 0.90413 | 0.94986 | 0.3938792379 | 0.90633 | 0.9725344777 | 0.215051 | 5.720255875 | 0 |
| 2 | 47463035 | A | T | E | V | MSH2 | ENST00000233146 | 0.71185 | 0.01562 | 0.70011 | 0.93808 | 0.9648 | 0.4508980408 | 0.9427 | 0.996653378 | 0.373274 | 4.318538977 | 0 |
| 2 | 47463036 | A | C | E | D | MSH2 | ENST00000233146 | 0.35383 | 0.00608 | 0.72254 | 0.72654 | 0.84747 | 0.2469922752 | 0.99048 | 0.5081710219 | 0.320147 | 5.277712733 | 0 |
| 2 | 47463036 | A | T | E | D | MSH2 | ENST00000233146 | 0.34921 | 0.00608 | 0.72254 | 0.73171 | 0.84747 | 0.2469922752 | 0.99048 | 0.3765872717 | 0.320143 | 5.277712733 | 0 |
| 2 | 47463037 | A | C | N | H | MSH2 | ENST00000233146 | 0.50056 | 0.0058 | 0.77043 | 0.67912 | 0.76816 | 0.2886757231 | 0.93772 | 0.7357383966 | 0.133683 | 2.41619946 | 0 |
| 2 | 47463037 | A | G | N | D | MSH2 | ENST00000233146 | 0.71072 | 0.00622 | 0.81216 | 0.82791 | 0.88637 | 0.2776178165 | 0.93167 | 0.968126297 | 0.165361 | 7.035109644 | 0 |
| 2 | 47463037 | A | T | N | Y | MSH2 | ENST00000233146 | 0.75201 | 0.01786 | 0.79873 | 0.90121 | 0.95316 | 0.3943071039 | 0.95162 | 0.9877540469 | 0.311891 | 7.035109644 | 0 |
| 2 | 47463038 | A | C | N | T | MSH2 | ENST00000233146 | 0.48341 | 0.01721 | 0.74783 | 0.89977 | 0.92642 | 0.2697754189 | 0.90052 | 0.9847865701 | 0.20524 | 2.791199993 | 0 |
| 2 | 47463038 | A | G | N | S | MSH2 | ENST00000233146 | 0.45175 | 0.01211 | 0.69489 | 0.82162 | 0.87491 | 0.192359428 | 0.91885 | 0.9711334705 | 0.164233 | 4.303304008 | 0 |
| 2 | 47463038 | A | T | N | I | MSH2 | ENST00000233146 | 0.84134 | 0.02475 | 0.76049 | 0.94783 | 0.95863 | 0.4080890313 | 0.89619 | 0.997325182 | 0.300632 | 6.723638803 | 0 |
| 2 | 47463039 | C | A | N | K | MSH2 | ENST00000233146 | 0.26786 | 0.00596 | 0.83506 | 0.76347 | 0.7562 | 0.2689598157 | 0.874 | 0.3320160806 | 0.0866937 | 2.686669974 | 0 |
| 2 | 47463039 | C | G | N | K | MSH2 | ENST00000233146 | 0.27284 | 0.00596 | 0.83506 | 0.72396 | 0.7562 | 0.2689598157 | 0.87545 | 0.2761504352 | 0.0875019 | 2.686669974 | 0 |
| 2 | 47463040 | C | A | H | N | MSH2 | ENST00000233146 | 0.55435 | 0.01108 | 0.84168 | 0.84338 | 0.91986 | 0.5772481083 | 0.92027 | 0.9916570187 | 0.178692 | 7.102189557 | 0 |
| 2 | 47463040 | C | G | H | D | MSH2 | ENST00000233146 | 0.6138 | 0.02077 | 0.83991 | 0.90454 | 0.92601 | 0.7240704848 | 0.91809 | 0.9987410903 | 0.396583 | 3.554832692 | 0 |
| 2 | 47463040 | C | T | H | Y | MSH2 | ENST00000233146 | 0.72804 | 0.00726 | 0.83863 | 0.84154 | 0.94506 | 0.6714461153 | 0.9153 | 0.9937707782 | 0.288673 | 8.194441835 | 0 |
| 2 | 47463041 | A | C | H | P | MSH2 | ENST00000233146 | 0.83936 | 0.0278 | 0.77635 | 0.90808 | 0.94802 | 0.8149067482 | 0.93072 | 0.9973298311 | 0.439997 | 8.194441835 | 0 |
| 2 | 47463041 | A | G | H | R | MSH2 | ENST00000233146 | 0.79057 | 0.02691 | 0.83952 | 0.88803 | 0.95608 | 0.6769375494 | 0.94706 | 0.9427734613 | 0.191033 | 8.194441835 | 0 |
| 2 | 47463041 | A | T | H | L | MSH2 | ENST00000233146 | 0.84567 | 0.02172 | 0.80202 | 0.93014 | 0.94506 | 0.6883178221 | 0.9482 | 0.9994361997 | 0.441082 | 8.194441835 | 0 |
| 2 | 47463042 | T | A | H | Q | MSH2 | ENST00000233146 | 0.47728 | 0.0114 | 0.81424 | 0.85298 | 0.90494 | 0.578536664 | 0.90325 | 0.9938752055 | 0.258886 | 7.098196311 | 0 |
| 2 | 47463042 | T | G | H | Q | MSH2 | ENST00000233146 | 0.48379 | 0.0114 | 0.81424 | 0.85298 | 0.90494 | 0.578536664 | 0.90288 | 0.9937746525 | 0.258889 | 7.098196311 | 0 |
| 2 | 47463043 | G | C | E | Q | MSH2 | ENST00000233146 | 0.77703 | 0.01616 | 0.8124 | 0.81434 | 0.96044 | 0.6229279103 | 0.95563 | 0.9566629529 | 0.374536 | 7.703845885 | 0 |
| 2 | 47463044 | A | C | E | A | MSH2 | ENST00000233146 | 0.89634 | 0.02864 | 0.82894 | 0.93116 | 0.96956 | 0.6828698219 | 0.96864 | 0.997092247 | 0.471847 | 5.630270582 | 0 |
| 2 | 47463044 | A | G | E | G | MSH2 | ENST00000233146 | 0.92501 | 0.03189 | 0.78084 | 0.9369 | 0.96956 | 0.7298418978 | 0.95561 | 0.9976705909 | 0.467965 | 3.343527984 | 0 |
| 2 | 47463044 | A | T | E | V | MSH2 | ENST00000233146 | 0.91556 | 0.02972 | 0.80666 | 0.9509 | 0.97252 | 0.6960734346 | 0.98619 | 0.997944057 | 0.477633 | 7.703845885 | 0 |
| 2 | 47463045 | A | C | E | D | MSH2 | ENST00000233146 | 0.48755 | 0.03059 | 0.82524 | 0.83097 | 0.92316 | 0.5629963408 | 0.96504 | 0.9503136873 | 0.3124 | 5.89074811 | 0 |
| 2 | 47463045 | A | T | E | D | MSH2 | ENST00000233146 | 0.48221 | 0.03059 | 0.82524 | 0.83208 | 0.92316 | 0.5629963408 | 0.96496 | 0.9503136873 | 0.312396 | 5.89074811 | 0 |
| 2 | 47463046 | T | A | F | I | MSH2 | ENST00000233146 | 0.93383 | 0.03076 | 0.87148 | 0.91803 | 0.96627 | 0.6760247915 | 0.91701 | 0.9990985394 | 0.339769 | 7.644949167 | 0 |
| 2 | 47463046 | T | C | F | L | MSH2 | ENST00000233146 | 0.93768 | 0.02802 | 0.89041 | 0.88687 | 0.96919 | 0.6342287379 | 0.90383 | 0.996329844 | 0.322855 | 6.645120518 | 0 |
| 2 | 47463046 | T | G | F | V | MSH2 | ENST00000233146 | 0.9353 | 0.03001 | 0.84225 | 0.93353 | 0.98257 | 0.6599767991 | 0.97228 | 0.9979471564 | 0.458375 | 6.647705297 | 0 |
| 2 | 47463047 | T | A | F | Y | MSH2 | ENST00000233146 | 0.56978 | 0.00722 | 0.87882 | 0.79046 | 0.7816 | 0.5171063652 | 0.87029 | 0.6366531849 | 0.176591 | 2.109547646 | 0 |
| 2 | 47463047 | T | C | F | S | MSH2 | ENST00000233146 | 0.95217 | 0.02765 | 0.87117 | 0.9532 | 0.9579 | 0.7476944741 | 0.97893 | 0.9972444773 | 0.463614 | 4.47976979 | 1 |
| 2 | 47463047 | T | G | F | C | MSH2 | ENST00000233146 | 0.95219 | 0.03124 | 0.89162 | 0.94438 | 0.96044 | 0.7212183107 | 0.97538 | 0.9983538389 | 0.46754 | 7.644949167 | 0 |
| 2 | 47463048 | C | A | F | L | MSH2 | ENST00000233146 | 0.63912 | 0.02802 | 0.89041 | 0.90339 | 0.94543 | 0.6342287379 | 0.9122 | 0.9893464446 | 0.26891 | 6.645120518 | 0 |
| 2 | 47463048 | C | G | F | L | MSH2 | ENST00000233146 | 0.64734 | 0.02802 | 0.89041 | 0.91195 | 0.94543 | 0.6342287379 | 0.91146 | 0.9868980646 | 0.268913 | 6.645120518 | 0 |
| 2 | 47463049 | C | A | L | I | MSH2 | ENST00000233146 | 0.6028 | 0.01115 | 0.75794 | 0.77315 | 0.87877 | 0.2922445555 | 0.80244 | 0.9488411546 | 0.150106 | 4.068676708 | 0 |
| 2 | 47463049 | C | G | L | V | MSH2 | ENST00000233146 | 0.56435 | 0.00696 | 0.7006 | 0.82918 | 0.86753 | 0.4152011096 | 0.89517 | 0.6141152978 | 0.233843 | 2.730085605 | 0 |
| 2 | 47463049 | C | T | L | F | MSH2 | ENST00000233146 | 0.64711 | 0.00877 | 0.74409 | 0.83106 | 0.90972 | 0.5209854729 | 0.81719 | 0.9791193604 | 0.247693 | 5.524740153 | 0 |
| 2 | 47463050 | T | A | L | H | MSH2 | ENST00000233146 | 0.82264 | 0.03019 | 0.83418 | 0.94624 | 0.98477 | 0.7403260617 | 0.95129 | 0.9985907674 | 0.465959 | 9.199641392 | 0 |
| 2 | 47463050 | T | C | L | P | MSH2 | ENST00000233146 | 0.8465 | 0.0262 | 0.86444 | 0.94675 | 0.99296 | 0.8896082342 | 0.98185 | 0.9983251691 | 0.491713 | 9.199641392 | 0 |
| 2 | 47463050 | T | G | L | R | MSH2 | ENST00000233146 | 0.84195 | 0.0262 | 0.83461 | 0.92026 | 0.97401 | 0.8236436846 | 0.97116 | 0.995375216 | 0.464203 | 9.199641392 | 0 |
| 2 | 47463052 | G | A | V | I | MSH2 | ENST00000233146 | 0.31915 | 0.00654 | 0.49941 | 0.52999 | 0.76881 | 0.2698993858 | 0.89238 | 0.8745487928 | 0.142462 | 1.51806605 | 0 |
| 2 | 47463052 | G | C | V | L | MSH2 | ENST00000233146 | 0.69981 | 0.01151 | 0.681 | 0.83871 | 0.93709 | 0.5792165262 | 0.92516 | 0.9559895992 | 0.263061 | 3.572021872 | 0 |
| 2 | 47463052 | G | T | V | L | MSH2 | ENST00000233146 | 0.69332 | 0.01151 | 0.681 | 0.83727 | 0.93709 | 0.5792165262 | 0.92544 | 0.9638642669 | 0.263057 | 3.572021872 | 0 |
| 2 | 47463053 | T | A | V | E | MSH2 | ENST00000233146 | 0.7575 | 0.0218 | 0.68986 | 0.94798 | 0.99296 | 0.8598398985 | 0.97334 | 0.9992015958 | 0.477947 | 7.410591398 | 1 |
| 2 | 47463053 | T | C | V | A | MSH2 | ENST00000233146 | 0.71097 | 0.0129 | 0.70502 | 0.89957 | 0.98257 | 0.6524945554 | 0.95145 | 0.9872411489 | 0.359648 | 7.410591398 | 0 |
| 2 | 47463053 | T | G | V | G | MSH2 | ENST00000233146 | 0.75185 | 0.02408 | 0.56169 | 0.94058 | 0.99265 | 0.8636021864 | 0.98535 | 0.9989910722 | 0.488471 | 7.410591398 | 1 |
| 2 | 47463055 | A | C | K | Q | MSH2 | ENST00000233146 | 0.82147 | 0.03313 | 0.79182 | 0.88677 | 0.97698 | 0.7057295325 | 0.97307 | 0.9963947033 | 0.416448 | 5.376989596 | 0 |
| 2 | 47463055 | A | G | K | E | MSH2 | ENST00000233146 | 0.84861 | 0.02714 | 0.83015 | 0.89822 | 0.98257 | 0.7654085688 | 0.98608 | 0.9972815514 | 0.496535 | 7.469774055 | 0 |

| | | | | | | | | | | | | | | | | | |
|---|---|---|---|---|---|---|---|---|---|---|---|---|---|---|---|---|---|
| 2 | 47463056 | A | C | K | T | MSH2 | ENST00000233146 | 0.8336 | 0.0317 | 0.83966 | 0.90404 | 0.98257 | 0.7183357027 | 0.96328 | 0.9979154468 | 0.488813 | 9.290103647 | 0 |
| 2 | 47463056 | A | G | K | R | MSH2 | ENST00000233146 | 0.67215 | 0.01044 | 0.7542 | 0.79536 | 0.9517 | 0.5882191196 | 0.95624 | 0.9755077958 | 0.323752 | 3.403245528 | 0 |
| 2 | 47463056 | A | T | K | I | MSH2 | ENST00000233146 | 0.87617 | 0.02366 | 0.80243 | 0.92283 | 0.9833 | 0.7407443294 | 0.96623 | 0.9997314811 | 0.50017 | 9.290103647 | 0 |
| 2 | 47463057 | A | C | K | N | MSH2 | ENST00000233146 | 0.51474 | 0.01172 | 0.85438 | 0.80694 | 0.90052 | 0.7213453485 | 0.92381 | 0.7815865278 | 0.140416 | 6.051576686 | 0 |
| 2 | 47463057 | A | T | K | N | MSH2 | ENST00000233146 | 0.50922 | 0.01172 | 0.85438 | 0.80895 | 0.90052 | 0.7213453485 | 0.92365 | 0.9718304276 | 0.151193 | 6.051576686 | 0 |
| 2 | 47463058 | C | A | P | T | MSH2 | ENST00000233146 | 0.55353 | 0.02335 | 0.50854 | 0.91369 | 0.93631 | 0.4860150325 | 0.92088 | 0.9923436046 | 0.274187 | 4.063826521 | 0 |
| 2 | 47463058 | C | G | P | A | MSH2 | ENST00000233146 | 0.34839 | 0.00627 | 0.49185 | 0.56383 | 0.74306 | 0.3126117365 | 0.89336 | 0.6361060739 | 0.110341 | 0.9293845118 | 0 |
| 2 | 47463058 | C | T | P | S | MSH2 | ENST00000233146 | 0.43232 | 0.0084 | 0.48045 | 0.78799 | 0.90096 | 0.4256188007 | 0.87988 | 0.9740573168 | 0.200899 | 1.997948922 | 0 |
| 2 | 47463059 | C | A | P | H | MSH2 | ENST00000233146 | 0.49151 | 0.03281 | 0.57742 | 0.92419 | 0.94506 | 0.4769990123 | 0.83853 | 0.9973166585 | 0.299108 | 6.015481975 | 0 |
| 2 | 47463059 | C | G | P | R | MSH2 | ENST00000233146 | 0.65008 | 0.03076 | 0.60766 | 0.89932 | 0.95353 | 0.5198504504 | 0.85961 | 0.9972578883 | 0.312993 | 4.777638422 | 0 |
| 2 | 47463059 | C | T | P | L | MSH2 | ENST00000233146 | 0.63267 | 0.02421 | 0.61954 | 0.91732 | 0.94506 | 0.4991969421 | 0.86339 | 0.9938959479 | 0.276139 | 5.355730714 | 1 |
| 2 | 47463061 | T | A | S | T | MSH2 | ENST00000233146 | 0.45095 | 0.0114 | 0.64668 | 0.80577 | 0.89739 | 0.3649238202 | 0.95919 | 0.8699323535 | 0.297795 | 3.213912166 | 0 |
| 2 | 47463061 | T | C | S | P | MSH2 | ENST00000233146 | 0.8536 | 0.03231 | 0.65905 | 0.89579 | 0.96407 | 0.664180008 | 0.97098 | 0.9827225208 | 0.465955 | 6.844422828 | 0 |
| 2 | 47463061 | T | G | S | A | MSH2 | ENST00000233146 | 0.64656 | 0.02133 | 0.62196 | 0.85052 | 0.92844 | 0.2798371977 | 0.94747 | 0.9750458598 | 0.314217 | 4.549318455 | 0 |
| 2 | 47463062 | C | T | S | L | MSH2 | ENST00000233146 | 0.68811 | 0.03287 | 0.6643 | 0.83438 | 0.97587 | 0.4303136494 | 0.98296 | 0.9685069919 | 0.344806 | 5.836699964 | 0 |
| 2 | 47463064 | T | A | F | I | MSH2 | ENST00000233146 | 0.90422 | 0.01611 | 0.75872 | 0.86989 | 0.91399 | 0.6808454523 | 0.9117 | 0.9937380552 | 0.248554 | 6.045011589 | 0 |
| 2 | 47463064 | T | C | F | L | MSH2 | ENST00000233146 | 0.92836 | 0.02673 | 0.79778 | 0.88146 | 0.96627 | 0.6250083969 | 0.94296 | 0.9958482981 | 0.325284 | 8.634506635 | 0 |
| 2 | 47463064 | T | G | F | V | MSH2 | ENST00000233146 | 0.92333 | 0.02796 | 0.74351 | 0.93147 | 0.97327 | 0.6655620828 | 0.97828 | 0.9975065589 | 0.457956 | 6.259497155 | 0 |
| 2 | 47463065 | T | A | F | Y | MSH2 | ENST00000233146 | 0.69788 | 0.02172 | 0.77986 | 0.81772 | 0.92924 | 0.5497846444 | 0.90738 | 0.9572948217 | 0.277155 | 3.597324644 | 0 |
| 2 | 47463065 | T | C | F | S | MSH2 | ENST00000233146 | 0.90715 | 0.0279 | 0.7611 | 0.93443 | 0.98367 | 0.7572466557 | 0.95798 | 0.9993336797 | 0.465547 | 8.634506635 | 0 |
| 2 | 47463065 | T | G | F | C | MSH2 | ENST00000233146 | 0.90715 | 0.03137 | 0.79389 | 0.9372 | 0.98477 | 0.7374758294 | 0.98011 | 0.9993336797 | 0.497956 | 6.078356574 | 0 |
| 2 | 47463066 | T | A | F | L | MSH2 | ENST00000233146 | 0.69548 | 0.02673 | 0.79778 | 0.92228 | 0.95826 | 0.6250083969 | 0.95437 | 0.9948865771 | 0.308044 | 8.634506635 | 0 |
| 2 | 47463066 | T | G | F | L | MSH2 | ENST00000233146 | 0.70374 | 0.02673 | 0.79778 | 0.92298 | 0.95826 | 0.6250083969 | 0.95422 | 0.9948865771 | 0.308046 | 8.634506635 | 0 |
| 2 | 47463067 | G | A | D | N | MSH2 | ENST00000233146 | 0.75089 | 0.03099 | 0.67702 | 0.86557 | 0.92844 | 0.6220037224 | 0.92647 | 0.990175426 | 0.166759 | 4.144380677 | 0 |
| 2 | 47463067 | G | C | D | H | MSH2 | ENST00000233146 | 0.89042 | 0.03458 | 0.7023 | 0.91936 | 0.97215 | 0.7215316156 | 0.94975 | 0.998781383 | 0.485697 | 4.593874255 | 0 |
| 2 | 47463067 | G | T | D | Y | MSH2 | ENST00000233146 | 0.89639 | 0.03398 | 0.68323 | 0.93046 | 0.98404 | 0.8058210053 | 0.98288 | 0.9997233748 | 0.511226 | 9.598522586 | 0 |
| 2 | 47463068 | A | C | D | A | MSH2 | ENST00000233146 | 0.72788 | 0.0337 | 0.61266 | 0.90113 | 0.97252 | 0.7443916292 | 0.92335 | 0.9932247996 | 0.451678 | 9.417502917 | 0 |
| 2 | 47463068 | A | G | D | G | MSH2 | ENST00000233146 | 0.86969 | 0.03281 | 0.63133 | 0.92153 | 0.9796 | 0.7406495847 | 0.95305 | 0.9978532195 | 0.493061 | 9.598522586 | 0 |
| 2 | 47463068 | A | T | D | V | MSH2 | ENST00000233146 | 0.85814 | 0.0318 | 0.60909 | 0.94691 | 0.98183 | 0.7608422177 | 0.97913 | 0.9990116358 | 0.502005 | 9.598522586 | 0 |
| 2 | 47463069 | T | A | D | E | MSH2 | ENST00000233146 | 0.58688 | 0.03761 | 0.65134 | 0.85988 | 0.96153 | 0.568820249 | 0.95437 | 0.9899072647 | 0.294386 | 7.950789521 | 0 |
| 2 | 47463069 | T | G | D | E | MSH2 | ENST00000233146 | 0.59421 | 0.03761 | 0.65134 | 0.85796 | 0.96153 | 0.568820249 | 0.95435 | 0.9899072647 | 0.294388 | 7.950789521 | 0 |
| 2 | 47463070 | C | A | P | T | MSH2 | ENST00000233146 | 0.6412 | 0.02943 | 0.64059 | 0.85536 | 0.92151 | 0.4134955119 | 0.82426 | 0.9827804565 | 0.281887 | 5.394740944 | 0 |
| 2 | 47463070 | C | G | P | A | MSH2 | ENST00000233146 | 0.48723 | 0.02556 | 0.58969 | 0.82489 | 0.9328 | 0.3604151284 | 0.86875 | 0.9878985286 | 0.286152 | 4.198179155 | 0 |
| 2 | 47463070 | C | T | P | S | MSH2 | ENST00000233146 | 0.39702 | 0.01766 | 0.60731 | 0.82181 | 0.89784 | 0.4431388754 | 0.89957 | 0.9734607339 | 0.286208 | 4.258053171 | 0 |
| 2 | 47463071 | C | A | P | H | MSH2 | ENST00000233146 | 0.62856 | 0.0337 | 0.66746 | 0.87043 | 0.93554 | 0.4473428968 | 0.86272 | 0.9885546565 | 0.363249 | 5.918102622 | 0 |
| 2 | 47463071 | C | G | P | R | MSH2 | ENST00000233146 | 0.62309 | 0.03398 | 0.68187 | 0.83928 | 0.9367 | 0.5177626927 | 0.91873 | 0.9781954288 | 0.456679 | 5.928372398 | 0 |
| 2 | 47463071 | C | T | P | L | MSH2 | ENST00000233146 | 0.64492 | 0.03447 | 0.6924 | 0.8385 | 0.93709 | 0.5481393914 | 0.91507 | 0.9941707253 | 0.406172 | 6.840533508 | 0 |
| 2 | 47463073 | A | C | N | H | MSH2 | ENST00000233146 | 0.49017 | 0.00615 | 0.34691 | 0.66056 | 0.70671 | 0.3392495029 | 0.89504 | 0.4755556583 | 0.110891 | 5.392808261 | 0 |
| 2 | 47463073 | A | G | N | D | MSH2 | ENST00000233146 | 0.42181 | 0.00556 | 0.41225 | 0.6033 | 0.60362 | 0.3615476858 | 0.81307 | 0.7979963422 | 0.0842021 | 3.282838775 | 1 |
| 2 | 47463073 | A | T | N | Y | MSH2 | ENST00000233146 | 0.68715 | 0.01334 | 0.39716 | 0.82522 | 0.90581 | 0.4378151725 | 0.92795 | 0.977730155 | 0.255365 | 4.785134643 | 0 |
| 2 | 47463074 | A | C | N | T | MSH2 | ENST00000233146 | 0.37919 | 0.00603 | 0.31524 | 0.62433 | 0.74658 | 0.2573662021 | 0.89044 | 0.6044293046 | 0.134572 | 3.316778756 | 0 |
| 2 | 47463074 | A | G | N | S | MSH2 | ENST00000233146 | 0.30519 | 0.00654 | 0.26164 | 0.55682 | 0.58006 | 0.2265314788 | 0.74484 | 0.7666309476 | 0.067171 | 4.27107866 | 0 |
| 2 | 47463074 | A | T | N | I | MSH2 | ENST00000233146 | 0.43853 | 0.00855 | 0.34432 | 0.74362 | 0.86352 | 0.3713579366 | 0.87618 | 0.9203644991 | 0.208126 | 5.870930138 | 0 |
| 2 | 47463075 | T | A | N | K | MSH2 | ENST00000233146 | 0.38209 | 0.0058 | 0.44662 | 0.70193 | 0.73661 | 0.3682956717 | 0.83276 | 0.358697772 | 0.0696152 | 3.783203746 | 0 |
| 2 | 47463075 | T | G | N | K | MSH2 | ENST00000233146 | 0.38794 | 0.0058 | 0.44662 | 0.65169 | 0.73661 | 0.3682956717 | 0.83295 | 0.3003613651 | 0.0696104 | 3.783203746 | 0 |
| 2 | 47463076 | C | A | L | I | MSH2 | ENST00000233146 | 0.52446 | 0.03527 | 0.48678 | 0.9191 | 0.93004 | 0.4647216806 | 0.87683 | 0.9790710211 | 0.333253 | 5.597690615 | 0 |
| 2 | 47463076 | C | G | L | V | MSH2 | ENST00000233146 | 0.52194 | 0.03633 | 0.41167 | 0.92318 | 0.96993 | 0.6480726349 | 0.9798 | 0.9804887772 | 0.490321 | 5.335740143 | 1 |
| 2 | 47463076 | C | T | L | F | MSH2 | ENST00000233146 | 0.54732 | 0.03582 | 0.46858 | 0.92212 | 0.9659 | 0.6921134497 | 0.97938 | 0.9379917979 | 0.33827 | 10.3849885 | 0 |
| 2 | 47463077 | T | A | L | H | MSH2 | ENST00000233146 | 0.90969 | 0.0334 | 0.58259 | 0.94108 | 0.99138 | 0.8281872033 | 0.98492 | 0.9991133809 | 0.55427 | 10.3849885 | 1 |
| 2 | 47463077 | T | C | L | P | MSH2 | ENST00000233146 | 0.92299 | 0.03001 | 0.62786 | 0.94058 | 0.99296 | 0.9353264987 | 0.98698 | 0.9993674159 | 0.554224 | 10.3849885 | 1 |
| 2 | 47463077 | T | G | L | R | MSH2 | ENST00000233146 | 0.92076 | 0.03144 | 0.58778 | 0.94057 | 0.99471 | 0.923973713 | 0.98672 | 0.9992833734 | 0.553949 | 10.3849885 | 1 |
| 2 | 47463079 | A | C | S | R | MSH2 | ENST00000233146 | 0.44429 | 0.00696 | 0.38894 | 0.6757 | 0.8153 | 0.4974282318 | 0.93744 | 0.2092620597 | 0.0691616 | 3.056963117 | 0 |
| 2 | 47463079 | A | G | S | G | MSH2 | ENST00000233146 | 0.44954 | 0.00586 | 0.29063 | 0.71519 | 0.81065 | 0.2711941522 | 0.93151 | 0.8377597928 | 0.189638 | 4.366499333 | 0 |
| 2 | 47463079 | A | T | S | C | MSH2 | ENST00000233146 | 0.8228 | 0.01871 | 0.34053 | 0.8579 | 0.92844 | 0.3012787122 | 0.864 | 0.9638922811 | 0.237129 | 3.494650613 | 0 |
| 2 | 47463080 | G | A | S | N | MSH2 | ENST00000233146 | 0.27436 | 0.00747 | 0.37472 | 0.81506 | 0.77399 | 0.3156655322 | 0.82924 | 0.2591524319 | 0.0588135 | 2.648787665 | 0 |
| 2 | 47463080 | G | C | S | T | MSH2 | ENST00000233146 | 0.22628 | 0.00586 | 0.32545 | 0.72768 | 0.70833 | 0.2346266712 | 0.92429 | 0.589738965 | 0.128274 | -0.4266605905 | 0 |
| 2 | 47463080 | G | T | S | I | MSH2 | ENST00000233146 | 0.34921 | 0.00747 | 0.38907 | 0.82825 | 0.85689 | 0.4330299721 | 0.93386 | 0.8752772212 | 0.240976 | 4.058144718 | 0 |
| 2 | 47463081 | T | A | S | R | MSH2 | ENST00000233146 | 0.37431 | 0.00696 | 0.38894 | 0.6964 | 0.81298 | 0.4974282318 | 0.95733 | 0.2138568611 | 0.196107 | 3.056963117 | 0 |
| 2 | 47463081 | T | G | S | R | MSH2 | ENST00000233146 | 0.38015 | 0.00696 | 0.38894 | 0.67138 | 0.81298 | 0.4974282318 | 0.95737 | 0.1431872475 | 0.128714 | 3.056963117 | 0 |
| 2 | 47463082 | G | C | E | Q | MSH2 | ENST00000233146 | 0.6221 | 0.0071 | 0.49016 | 0.71493 | 0.83997 | 0.4652950927 | 0.94135 | 0.9104485512 | 0.269044 | 4.189820074 | 0 |
| 2 | 47463083 | A | C | E | A | MSH2 | ENST00000233146 | 0.49126 | 0.00615 | 0.504 | 0.79314 | 0.85996 | 0.5518371483 | 0.94506 | 0.8746090531 | 0.257583 | 3.597794321 | 0 |
| 2 | 47463083 | A | G | E | G | MSH2 | ENST00000233146 | 0.53778 | 0.00586 | 0.46104 | 0.85179 | 0.85012 | 0.57499379 | 0.89781 | 0.9514428377 | 0.234881 | 5.9176935 | 0 |
| 2 | 47463083 | A | T | E | V | MSH2 | ENST00000233146 | 0.5324 | 0.00674 | 0.48161 | 0.85325 | 0.90842 | 0.5691235422 | 0.9562 | 0.9601259828 | 0.303081 | 4.269889321 | 0 |
| 2 | 47463084 | A | C | E | D | MSH2 | ENST00000233146 | 0.38335 | 0.00619 | 0.49984 | 0.6732 | 0.8367 | 0.4484956426 | 0.97589 | 0.2443708479 | 0.21989 | 4.775455922 | 0 |
| 2 | 47463084 | A | T | E | D | MSH2 | ENST00000233146 | 0.3785 | 0.00619 | 0.49984 | 0.67355 | 0.8367 | 0.4484956426 | 0.97575 | 0.2443708479 | 0.219888 | 4.775455922 | 0 |
| 2 | 47463085 | T | A | L | I | MSH2 | ENST00000233146 | 0.40899 | 0.01253 | 0.64733 | 0.70316 | 0.84159 | 0.3163278914 | 0.77198 | 0.7940846682 | 0.125629 | 5.014721026 | 0 |
| 2 | 47463085 | T | G | L | V | MSH2 | ENST00000233146 | 0.66652 | 0.01959 | 0.59116 | 0.80255 | 0.90755 | 0.4983106934 | 0.93272 | 0.9332769513 | 0.262299 | 6.151766535 | 0 |
| 2 | 47463086 | T | C | L | S | MSH2 | ENST00000233146 | 0.90498 | 0.02608 | 0.66165 | 0.89404 | 0.98071 | 0.7513112159 | 0.95239 | 0.9965566397 | 0.45637 | 8.482024537 | 0 |
| 2 | 47463087 | A | C | L | F | MSH2 | ENST00000233146 | 0.47886 | 0.03106 | 0.63085 | 0.8723 | 0.93747 | 0.5807964907 | 0.8926 | 0.9766606688 | 0.266742 | 4.690633807 | 0 |
| 2 | 47463087 | A | T | L | F | MSH2 | ENST00000233146 | 0.47357 | 0.03106 | 0.63085 | 0.87155 | 0.93747 | 0.5807964907 | 0.89301 | 0.9766606688 | 0.266732 | 4.690633807 | 0 |
| 2 | 47463088 | A | G | R | G | MSH2 | ENST00000233146 | 0.93035 | 0.0315 | 0.5184 | 0.93888 | 0.98034 | 0.6706356349 | 0.98664 | 0.9977370501 | 0.469688 | 8.492895433 | 0 |
| 2 | 47463089 | G | A | R | K | MSH2 | ENST00000233146 | 0.54129 | 0.0233 | 0.59121 | 0.83269 | 0.93241 | 0.5916061331 | 0.95426 | 0.9414090514 | 0.363254 | 3.791619852 | 0 |
| 2 | 47463089 | G | C | R | T | MSH2 | ENST00000233146 | 0.87749 | 0.03013 | 0.56598 | 0.88547 | 0.98799 | 0.6791216355 | 0.97083 | 0.9966286243 | 0.4933 | 6.24141782 | 0 |
| 2 | 47463090 | A | C | R | S | MSH2 | ENST00000233146 | 0.65374 | 0.02589 | 0.52618 | 0.94455 | 0.95243 | 0.6741063349 | 0.99395 | 0.9947347641 | 0.423364 | 6.127539038 | 0 |
| 2 | 47463090 | A | T | R | S | MSH2 | ENST00000233146 | 0.64718 | 0.02589 | 0.52618 | 0.94817 | 0.95243 | 0.6741063349 | 0.99403 | 0.9956252575 | 0.423357 | 6.127539038 | 0 |
| 2 | 47463091 | G | A | E | K | MSH2 | ENST00000233146 | 0.59443 | 0.0063 | 0.53452 | 0.52827 | 0.80772 | 0.4822054962 | 0.92098 | 0.8235049844 | 0.0518448 | 3.789330291 | 0 |
| 2 | 47463091 | G | C | E | Q | MSH2 | ENST00000233146 | 0.57603 | 0.01959 | 0.43015 | 0.85265 | 0.88022 | 0.3915722178 | 0.89546 | 0.9302378893 | 0.27346 | 4.526378313 | 0 |
| 2 | 47463092 | A | C | E | A | MSH2 | ENST00000233146 | 0.59951 | 0.0114 | 0.44004 | 0.61482 | 0.86503 | 0.4801889898 | 0.90957 | 0.8851950765 | 0.0478718 | 3.401587891 | 0 |
| 2 | 47463092 | A | G | E | G | MSH2 | ENST00000233146 | 0.65241 | 0.00608 | 0.40225 | 0.84522 | 0.87636 | 0.499079007 | 0.93658 | 0.9463761449 | 0.104301 | 4.998971679 | 0 |
| 2 | 47463092 | A | T | E | V | MSH2 | ENST00000233146 | 0.64867 | 0.00893 | 0.41627 | 0.87729 | 0.87925 | 0.5030737087 | 0.92879 | 0.9685891867 | 0.133837 | 5.516604764 | 0 |
| 2 | 47463093 | A | C | E | D | MSH2 | ENST00000233146 | 0.37879 | 0.00638 | 0.4397 | 0.08433 | 0.79697 | 0.3501646727 | 0.97365 | 0.1873952299 | 0.199979 | 3.30092358 | 0 |
| 2 | 47463093 | A | T | E | D | MSH2 | ENST00000233146 | 0.37403 | 0.00638 | 0.4397 | 0.09337 | 0.79697 | 0.3501646727 | 0.97378 | 0.2079651356 | 0.199976 | 3.30092358 | 0 |
| 2 | 47463094 | A | C | I | L | MSH2 | ENST00000233146 | 0.33828 | 0.00574 | 0.21281 | 0.06427 | 0.73443 | 0.320298301 | 0.86562 | 0.5460788608 | 0.0680126 | 1.120743101 | 0 |
| 2 | 47463094 | A | G | I | V | MSH2 | ENST00000233146 | 0.28232 | 0.00603 | 0.15909 | 0.02497 | 0.64876 | 0.1882506608 | 0.78328 | 0.3429844677 | 0.0511925 | 0.442099958 | 0 |
| 2 | 47463094 | A | T | I | L | MSH2 | ENST00000233146 | 0.33371 | 0.00574 | 0.21281 | 0.06726 | 0.73443 | 0.320298301 | 0.86159 | 0.5460788608 | 0.0679918 | 1.120743101 | 0 |
| 2 | 47463095 | T | A | I | K | MSH2 | ENST00000233146 | 0.24497 | 0.00476 | 0.25346 | 0.06726 | 0.6757 | 0.3936891838 | 0.7532 | 0.3993217647 | 0.0462448 | -1.313858727 | 0 |
| 2 | 47463095 | T | C | I | T | MSH2 | ENST00000233146 | 0.30025 | 0.00535 | 0.30404 | 0.50222 | 0.77335 | 0.24650927 | 0.9038 | 0.4139681756 | 0.0452233 | 1.812450292 | 0 |
| 2 | 47463095 | T | G | I | R | MSH2 | ENST00000233146 | 0.3243 | 0.0048 | 0.23722 | 0.11094 | 0.74376 | 0.3761391985 | 0.93467 | 0.8112578392 | 0.128439 | 0.6837586163 | 0 |
| 2 | 47463096 | A | G | I | M | MSH2 | ENST00000233146 | 0.4609 | 0.00735 | 0.19554 | 0.12846 | 0.74021 | 0.3084562579 | 0.82594 | 0.2400661409 | 0.0678936 | 1.686238185 | 0 |
| 2 | 47463097 | A | C | M | L | MSH2 | ENST00000233146 | 0.44399 | 0.00638 | 0.47858 | 0.33731 | 0.80298 | 0.5084284102 | 0.87122 | 0.307261616 | 0.169725 | 4.068711393 | 0 |
| 2 | 47463097 | A | G | M | V | MSH2 | ENST00000233146 | 0.55236 | 0.00928 | 0.41218 | 0.6884 | 0.90755 | 0.5952370454 | 0.9446 | 0.8199888468 | 0.190089 | 5.210554874 | 0 |
| 2 | 47463097 | A | T | M | L | MSH2 | ENST00000233146 | 0.43883 | 0.00638 | 0.47858 | 0.40571 | 0.80298 | 0.5084284102 | 0.87377 | 0.7751297355 | 0.214032 | 4.068711393 | 0 |
| 2 | 47463098 | T | A | M | K | MSH2 | ENST00000233146 | 0.75623 | 0.02561 | 0.5553 | 0.48703 | 0.94356 | 0.8332861629 | 0.97028 | 0.9940199852 | 0.430389 | 5.028523967 | 0 |
| 2 | 47463098 | T | C | M | T | MSH2 | ENST00000233146 | 0.88442 | 0.02556 | 0.60732 | 0.94213 | 0.9703 | 0.6766392413 | 0.95522 | 0.9942653775 | 0.423202 | 4.725335816 | 0 |
| 2 | 47463098 | T | G | M | R | MSH2 | ENST00000233146 | 0.92809 | 0.02732 | 0.53654 | 0.66355 | 0.96117 | 0.8551581737 | 0.96983 | 0.9974217415 | 0.442455 | 5.422764785 | 0 |
| 2 | 47463100 | A | C | N | H | MSH2 | ENST00000233146 | 0.76339 | 0.01774 | 0.12493 | 0.58394 | 0.81646 | 0.3418387052 | 0.83662 | 0.968875885 | 0.280655 | 3.223646487 | 0 |
| 2 | 47463100 | A | G | N | D | MSH2 | ENST00000233146 | 0.37736 | 0.00547 | 0.11378 | 0.19576 | 0.61531 | 0.2676233092 | 0.68023 | 0.06580427732 | -0.249411 | -1.518906116 | 0 |
| 2 | 47463100 | A | T | N | Y | MSH2 | ENST00000233146 | 0.82006 | 0.01945 | 0.12695 | 0.70173 | 0.87247 | 0.4356376139 | 0.91147 | 0.9960482717 | 0.373183 | 6.268086666 | 0 |
| 2 | 47463101 | A | C | N | T | MSH2 | ENST00000233146 | 0.55518 | 0.00586 | 0.11226 | 0.43047 | 0.7255 | 0.2858193076 | 0.90994 | 0.9419525266 | 0.020412 | 1.161245501 | 0 |
| 2 | 47463101 | A | G | N | S | MSH2 | ENST00000233146 | 0.516 | 0.00627 | 0.09583 | 0.57714 | 0.70508 | 0.2528594796 | 0.94875 | 0.9238553047 | 0.127594 | 1.526169597 | 0 |
| 2 | 47463101 | A | T | N | I | MSH2 | ENST00000233146 | 0.74738 | 0.00935 | 0.1196 | 0.61242 | 0.83116 | 0.3908165323 | 0.93043 | 0.988065064 | 0.241289 | 2.962741602 | 0 |

| | | | | | | | | | | | | | | | | | | |
|---|---|---|---|---|---|---|---|---|---|---|---|---|---|---|---|---|---|---|
| 2 | 47463102 | T | A | N | K | MSH2 | ENST00000233146 | 0.52999 | 0.0073 | 0.13493 | 0.32488 | 0.79089 | 0.3509268719 | 0.8931 | 0.9650942683 | 0.101072 | 1.712183661 | 0 |
| 2 | 47463102 | T | G | N | K | MSH2 | ENST00000233146 | 0.5369 | 0.0073 | 0.13493 | 0.30096 | 0.79151 | 0.3509268719 | 0.89296 | 0.9650942683 | 0.101057 | 1.712183661 | 0 |
| 2 | 47463103 | G | A | D | N | MSH2 | ENST00000233146 | 0.38972 | 0.0058 | 0.09851 | 0.514 | 0.62325 | 0.2878828833 | 0.88048 | 0.5030593872 | 0.063161 | 0.500784621 | 0 |
| 2 | 47463103 | G | C | D | H | MSH2 | ENST00000233146 | 0.52825 | 0.0138 | 0.10657 | 0.26741 | 0.83339 | 0.4030342629 | 0.95334 | 0.9330071211 | 0.207771 | 2.082393798 | 0 |
| 2 | 47463103 | G | T | D | Y | MSH2 | ENST00000233146 | 0.50419 | 0.01795 | 0.11061 | 0.14982 | 0.80772 | 0.4901864028 | 0.96475 | 0.9454058409 | 0.215144 | 4.492193804 | 0 |
| 2 | 47463104 | A | C | D | A | MSH2 | ENST00000233146 | 0.25866 | 0.00535 | 0.07977 | 0.05752 | 0.74797 | 0.37453048 | 0.97098 | 0.1109714971 | 0.120511 | 2.732383896 | 0 |
| 2 | 47463104 | A | G | D | G | MSH2 | ENST00000233146 | 0.2621 | 0.00527 | 0.0818 | 0.12877 | 0.74376 | 0.4140598626 | 0.98678 | 0.3043192029 | 0.11664 | 0.9981689126 | 0 |
| 2 | 47463104 | A | T | D | V | MSH2 | ENST00000233146 | 0.2634 | 0.00522 | 0.06827 | 0.22705 | 0.77527 | 0.460754645 | 0.99033 | 0.1386841373 | 0.165441 | 5.233324137 | 0 |
| 2 | 47463105 | C | A | D | E | MSH2 | ENST00000233146 | 0.16267 | 0.0058 | 0.08543 | 0.18402 | 0.74728 | 0.1943810985 | 0.89332 | 0.05596676735 | 0.0238699 | -0.5209789893 | 0 |
| 2 | 47463105 | C | G | D | E | MSH2 | ENST00000233146 | 0.16633 | 0.0058 | 0.08543 | 0.17472 | 0.74728 | 0.1943810985 | 0.89359 | 0.04681950768 | -0.113727 | -0.5209789893 | 0 |
| 2 | 47463106 | T | A | L | M | MSH2 | ENST00000233146 | 0.4622 | 0.02464 | 0.3583 | 0.75131 | 0.88259 | 0.4494921422 | 0.83169 | 0.8443574905 | 0.110767 | 6.455624652 | 0 |
| 2 | 47463106 | T | G | L | V | MSH2 | ENST00000233146 | 0.32041 | 0.01158 | 0.32589 | 0.77277 | 0.90318 | 0.4806137712 | 0.93501 | 0.1884815842 | 0.105903 | 4.091192506 | 0 |
| 2 | 47463107 | T | C | L | S | MSH2 | ENST00000233146 | 0.86197 | 0.03124 | 0.41825 | 0.91577 | 0.96553 | 0.7477414608 | 0.86653 | 0.9960910678 | 0.318919 | 3.876724412 | 0 |
| 2 | 47463107 | T | G | L | W | MSH2 | ENST00000233146 | 0.8747 | 0.02965 | 0.48561 | 0.93139 | 0.97141 | 0.7948882912 | 0.89331 | 0.9983729124 | 0.330915 | 7.175490544 | 0 |
| 2 | 47463108 | G | C | L | F | MSH2 | ENST00000233146 | 0.55881 | 0.03442 | 0.37565 | 0.89194 | 0.89422 | 0.5896812041 | 0.90482 | 0.9898634553 | 0.199236 | 7.175490544 | 0 |
| 2 | 47463108 | G | T | L | F | MSH2 | ENST00000233146 | 0.55319 | 0.03442 | 0.37565 | 0.89255 | 0.89422 | 0.5896812041 | 0.90507 | 0.9898634553 | 0.199231 | 7.175490544 | 0 |
| 2 | 47463109 | G | A | E | K | MSH2 | ENST00000233146 | 0.78535 | 0.03124 | 0.49139 | 0.8273 | 0.92438 | 0.684804005 | 0.92401 | 0.9254186153 | 0.331902 | 3.223774515 | 0 |
| 2 | 47463109 | G | C | E | Q | MSH2 | ENST00000233146 | 0.69556 | 0.02089 | 0.3768 | 0.80733 | 0.8933 | 0.5425363706 | 0.89137 | 0.9059966207 | 0.253305 | 4.414729126 | 0 |
| 2 | 47463110 | A | C | E | A | MSH2 | ENST00000233146 | 0.91369 | 0.02755 | 0.38178 | 0.90109 | 0.92151 | 0.6212514384 | 0.87734 | 0.9302425867 | 0.287251 | 4.650925735 | 0 |
| 2 | 47463110 | A | G | E | G | MSH2 | ENST00000233146 | 0.66359 | 0.01229 | 0.34839 | 0.8653 | 0.84694 | 0.6417261022 | 0.82856 | 0.9881240726 | 0.217455 | 5.874362261 | 0 |
| 2 | 47463110 | A | T | E | V | MSH2 | ENST00000233146 | 0.92709 | 0.02835 | 0.36342 | 0.89749 | 0.92561 | 0.6071170827 | 0.92472 | 0.991938889 | 0.304464 | 7.348116162 | 0 |
| 2 | 47463111 | A | C | E | D | MSH2 | ENST00000233146 | 0.43454 | 0.0134 | 0.38604 | 0.69529 | 0.85378 | 0.5308120011 | 0.93756 | 0.6747639775 | 0.203202 | 5.293105851 | 0 |
| 2 | 47463111 | A | T | E | D | MSH2 | ENST00000233146 | 0.42945 | 0.0134 | 0.38604 | 0.69514 | 0.85378 | 0.5308120011 | 0.93757 | 0.6747639775 | 0.203201 | 5.293105851 | 0 |
| 2 | 47463112 | A | C | K | Q | MSH2 | ENST00000233146 | 0.43653 | 0.01075 | 0.18989 | 0.71478 | 0.83834 | 0.3498978313 | 0.90705 | 0.8415017724 | 0.185201 | 3.040528984 | 0 |
| 2 | 47463112 | A | G | K | E | MSH2 | ENST00000233146 | 0.35482 | 0.00517 | 0.21809 | 0.4483 | 0.72322 | 0.374154035 | 0.83977 | 0.690440774 | 0.0910252 | 0.9662393117 | 0 |
| 2 | 47463113 | A | C | K | T | MSH2 | ENST00000233146 | 0.51488 | 0.00909 | 0.23042 | 0.78332 | 0.88496 | 0.4305756571 | 0.86912 | 0.8374931812 | 0.201626 | 3.236887963 | 0 |
| 2 | 47463113 | A | G | K | R | MSH2 | ENST00000233146 | 0.52612 | 0.00638 | 0.17158 | 0.67113 | 0.87345 | 0.3107107415 | 0.94537 | 0.8138408661 | 0.210836 | 2.931368676 | 0 |
| 2 | 47463113 | A | T | K | M | MSH2 | ENST00000233146 | 0.86375 | 0.02552 | 0.17216 | 0.87132 | 0.89829 | 0.5257064038 | 0.81569 | 0.9802231789 | 0.192927 | 6.153507518 | 1 |
| 2 | 47463114 | G | C | K | N | MSH2 | ENST00000233146 | 0.47533 | 0.00916 | 0.24397 | 0.80915 | 0.81298 | 0.4340422809 | 0.83613 | 0.7558869123 | 0.0991579 | 2.459299584 | 0 |
| 2 | 47463114 | G | T | K | N | MSH2 | ENST00000233146 | 0.47024 | 0.00916 | 0.24397 | 0.80915 | 0.81298 | 0.4340422809 | 0.83584 | 0.7558869123 | 0.0991539 | 2.459299584 | 0 |
| 2 | 47463115 | A | C | K | Q | MSH2 | ENST00000233146 | 0.49074 | 0.00648 | 0.30971 | 0.68991 | 0.78348 | 0.3528508369 | 0.92184 | 0.7066867948 | 0.172089 | 2.418524384 | 0 |
| 2 | 47463115 | A | G | K | E | MSH2 | ENST00000233146 | 0.4901 | 0.00556 | 0.35264 | 0.70667 | 0.73516 | 0.3934576377 | 0.92544 | 0.8029796481 | 0.176736 | 2.119980599 | 0 |
| 2 | 47463116 | A | C | K | T | MSH2 | ENST00000233146 | 0.48799 | 0.01324 | 0.36391 | 0.78741 | 0.84482 | 0.4235530767 | 0.9054 | 0.7895508409 | 0.196645 | 5.401303037 | 0 |
| 2 | 47463116 | A | G | K | R | MSH2 | ENST00000233146 | 0.46103 | 0.00654 | 0.28905 | 0.69112 | 0.79938 | 0.3232465555 | 0.92362 | 0.8306639791 | 0.170388 | 5.075480367 | 0 |
| 2 | 47463116 | A | T | K | M | MSH2 | ENST00000233146 | 0.83019 | 0.02937 | 0.29432 | 0.88565 | 0.90052 | 0.511883425 | 0.842 | 0.9758732319 | 0.217537 | 3.210609328 | 0 |
| 2 | 47463117 | G | C | K | N | MSH2 | ENST00000233146 | 0.56179 | 0.00586 | 0.38377 | 0.5323 | 0.6847 | 0.4173745093 | 0.91439 | 0.7641544342 | 0.104672 | 2.584848755 | 0 |
| 2 | 47463117 | G | T | K | N | MSH2 | ENST00000233146 | 0.55614 | 0.00586 | 0.38377 | 0.5323 | 0.6847 | 0.4173745093 | 0.91529 | 0.9640986919 | -0.0827972 | 2.584848755 | 0 |
| 2 | 47463118 | A | C | M | L | MSH2 | ENST00000233146 | 0.41369 | 0.00592 | 0.57418 | 0.69056 | 0.78286 | 0.5834379817 | 0.95863 | 0.7947259545 | 0.251728 | 2.744757597 | 0 |
| 2 | 47463118 | A | G | M | V | MSH2 | ENST00000233146 | 0.53414 | 0.00821 | 0.5094 | 0.85277 | 0.88307 | 0.6340849546 | 0.93041 | 0.9802787304 | 0.280929 | 4.110148909 | 0 |
| 2 | 47463118 | A | T | M | L | MSH2 | ENST00000233146 | 0.40875 | 0.00592 | 0.57418 | 0.69056 | 0.78286 | 0.5834379817 | 0.95889 | 0.4886205861 | 0.251726 | 2.744757597 | 0 |
| 2 | 47463119 | T | A | M | K | MSH2 | ENST00000233146 | 0.82014 | 0.02037 | 0.64803 | 0.91588 | 0.98183 | 0.8827212848 | 0.94228 | 0.9988071918 | 0.456215 | 7.677380358 | 0 |
| 2 | 47463119 | T | C | M | T | MSH2 | ENST00000233146 | 0.75559 | 0.01705 | 0.68406 | 0.92037 | 0.97885 | 0.7427321093 | 0.95142 | 0.9912075996 | 0.450665 | 7.677380358 | 0 |
| 2 | 47463119 | T | G | M | R | MSH2 | ENST00000233146 | 0.84233 | 0.02307 | 0.62815 | 0.91121 | 0.98728 | 0.9029048014 | 0.96708 | 0.9992489219 | 0.46799 | 7.677380358 | 0 |
| 2 | 47463120 | G | A | M | I | MSH2 | ENST00000233146 | 0.45746 | 0.00566 | 0.57538 | 0.83714 | 0.80058 | 0.6566958777 | 0.92532 | 0.7065237761 | 0.252969 | 1.789288237 | 0 |
| 2 | 47463120 | G | C | M | I | MSH2 | ENST00000233146 | 0.45493 | 0.00566 | 0.57538 | 0.83745 | 0.80058 | 0.6566958777 | 0.92447 | 0.7065237761 | 0.252969 | 1.789288237 | 0 |
| 2 | 47463120 | G | T | M | I | MSH2 | ENST00000233146 | 0.44997 | 0.00566 | 0.57538 | 0.83624 | 0.80058 | 0.6566958777 | 0.92476 | 0.7065237761 | 0.252969 | 1.789288237 | 0 |
| 2 | 47463121 | C | A | Q | K | MSH2 | ENST00000233146 | 0.36342 | 0.00556 | 0.39723 | 0.76423 | 0.77463 | 0.5763801613 | 0.95892 | 0.7590888143 | 0.252736 | 1.902010495 | 0 |
| 2 | 47463121 | C | G | Q | E | MSH2 | ENST00000233146 | 0.34696 | 0.00569 | 0.34191 | 0.75228 | 0.79758 | 0.4510131254 | 0.89108 | 0.7747261524 | 0.22209 | 5.412289902 | 0 |
| 2 | 47463122 | A | C | Q | P | MSH2 | ENST00000233146 | 0.46308 | 0.00551 | 0.35686 | 0.76484 | 0.86352 | 0.7954746067 | 0.98428 | 0.8803138733 | 0.407431 | 3.437712981 | 0 |
| 2 | 47463122 | A | G | Q | R | MSH2 | ENST00000233146 | 0.44174 | 0.00586 | 0.392 | 0.75114 | 0.76685 | 0.5788337741 | 0.95454 | 0.7903909683 | 0.213891 | 5.000329727 | 0 |
| 2 | 47463122 | A | T | Q | L | MSH2 | ENST00000233146 | 0.76721 | 0.01376 | 0.33512 | 0.84742 | 0.91611 | 0.5676725985 | 0.94379 | 0.9423244596 | 0.172843 | 3.837919179 | 0 |
| 2 | 47463124 | T | A | S | T | MSH2 | ENST00000233146 | 0.38857 | 0.00608 | 0.29642 | 0.71782 | 0.77972 | 0.2246504519 | 0.92925 | 0.4712767899 | 0.119163 | 3.160777919 | 0 |
| 2 | 47463124 | T | C | S | P | MSH2 | ENST00000233146 | 0.62962 | 0.01351 | 0.30981 | 0.79245 | 0.87877 | 0.6779306063 | 0.95787 | 0.420737944 | 0.0264962 | 7.007630219 | 0 |
| 2 | 47463124 | T | G | S | A | MSH2 | ENST00000233146 | 0.38455 | 0.00608 | 0.27084 | 0.48313 | 0.69846 | 0.1635216837 | 0.81868 | 0.4981611967 | 0.0560824 | 1.470922577 | 0 |
| 2 | 47463125 | C | T | S | L | MSH2 | ENST00000233146 | 0.37431 | 0.00836 | 0.29842 | 0.79035 | 0.79758 | 0.3502667022 | 0.87952 | 0.8646963835 | 0.152119 | 4.755224608 | 0 |
| 2 | 47463127 | A | C | T | P | MSH2 | ENST00000233146 | 0.43877 | 0.00696 | 0.13581 | 0.55804 | 0.83725 | 0.6398021923 | 0.93537 | 0.5276654363 | 0.141154 | 3.985489735 | 0 |
| 2 | 47463127 | A | G | T | A | MSH2 | ENST00000233146 | 0.33688 | 0.00574 | 0.1201 | 0.45362 | 0.71627 | 0.2162567447 | 0.89772 | 0.3209779263 | 0.0621996 | 0.4108709248 | 0 |
| 2 | 47463127 | A | T | T | S | MSH2 | ENST00000233146 | 0.32728 | 0.00592 | 0.12738 | 0.42091 | 0.69762 | 0.2062106787 | 0.92708 | 0.4471231401 | 0.0701548 | 2.820624511 | 0 |
| 2 | 47463128 | C | A | T | K | MSH2 | ENST00000233146 | 0.24906 | 0.00599 | 0.19786 | 0.58866 | 0.77463 | 0.4820406844 | 0.89321 | 0.4055313766 | 0.142471 | 3.006263131 | 0 |
| 2 | 47463128 | C | G | T | R | MSH2 | ENST00000233146 | 0.25142 | 0.00638 | 0.18626 | 0.63724 | 0.76619 | 0.5398686596 | 0.91412 | 0.3633368015 | 0.147511 | 3.985489735 | 0 |
| 2 | 47463128 | C | T | T | I | MSH2 | ENST00000233146 | 0.24219 | 0.00566 | 0.22295 | 0.54066 | 0.76289 | 0.2108507599 | 0.94222 | 0.380589962 | 0.129225 | 0.9057065595 | 0 |
| 2 | 47463130 | T | A | L | I | MSH2 | ENST00000233146 | 0.42103 | 0.01355 | 0.43289 | 0.78292 | 0.88069 | 0.3366144843 | 0.90651 | 0.9491766691 | 0.215373 | 4.545275654 | 0 |
| 2 | 47463130 | T | G | L | V | MSH2 | ENST00000233146 | 0.5185 | 0.02094 | 0.36952 | 0.84068 | 0.89558 | 0.4829736445 | 0.97191 | 0.9814413786 | 0.342483 | 5.951166749 | 0 |
| 2 | 47463131 | T | C | L | S | MSH2 | ENST00000233146 | 0.74986 | 0.02824 | 0.45784 | 0.89152 | 0.93631 | 0.6994445701 | 0.97202 | 0.9879669547 | 0.424821 | 7.432003503 | 0 |
| 2 | 47463132 | A | C | L | F | MSH2 | ENST00000233146 | 0.40383 | 0.00777 | 0.4202 | 0.81587 | 0.88354 | 0.4969795164 | 0.99466 | 0.4166243374 | 0.324656 | 7.571493715 | 0 |
| 2 | 47463132 | A | T | L | F | MSH2 | ENST00000233146 | 0.39894 | 0.00777 | 0.4202 | 0.81587 | 0.88354 | 0.4969795164 | 0.99469 | 0.4166243374 | 0.324664 | 7.571493715 | 0 |
| 2 | 47463133 | A | C | I | L | MSH2 | ENST00000233146 | 0.24116 | 0.00574 | 0.04871 | 0.64517 | 0.70671 | 0.2285658462 | 0.83462 | 0.04671342671 | 0.0290216 | 0.2446889757 | 0 |
| 2 | 47463133 | A | G | I | V | MSH2 | ENST00000233146 | 0.20501 | 0.00603 | 0.02608 | 0.59037 | 0.70345 | 0.1338714347 | 0.86224 | 0.01919392267 | -0.0304871 | -0.08843778197 | 0 |
| 2 | 47463133 | A | T | I | L | MSH2 | ENST00000233146 | 0.2375 | 0.00574 | 0.04871 | 0.6587 | 0.70671 | 0.2285658462 | 0.83371 | 0.04671342671 | 0.0290195 | 0.2446889757 | 0 |
| 2 | 47463134 | T | A | I | K | MSH2 | ENST00000233146 | 0.15159 | 0.00476 | 0.07269 | 0.63461 | 0.69425 | 0.3379032878 | 0.82777 | 0.04574255834 | 0.0428948 | -2.563263657 | 0 |
| 2 | 47463134 | T | C | I | T | MSH2 | ENST00000233146 | 0.17183 | 0.00532 | 0.10001 | 0.53977 | 0.77076 | 0.1781018353 | 0.89365 | 0.02284995402 | 0.0604738 | -0.955124191 | 0 |
| 2 | 47463134 | T | G | I | R | MSH2 | ENST00000233146 | 0.18654 | 0.00512 | 0.06202 | 0.66325 | 0.78658 | 0.313938672 | 0.92649 | 0.0966296196 | 0.0855513 | -0.5929440261 | 0 |
| 2 | 47463135 | A | G | I | M | MSH2 | ENST00000233146 | 0.36772 | 0.0058 | 0.04111 | 0.65427 | 0.74376 | 0.2174343885 | 0.88399 | 0.1441125125 | 0.060957 | 0.765831514 | 0 |
| 2 | 47463136 | A | C | S | R | MSH2 | ENST00000233146 | 0.40564 | 0.00551 | 0.24896 | 0.62147 | 0.79938 | 0.4225796023 | 0.95949 | 0.3907755315 | 0.186959 | 1.394584595 | 0 |
| 2 | 47463136 | A | G | S | G | MSH2 | ENST00000233146 | 0.41451 | 0.00586 | 0.1756 | 0.74737 | 0.74164 | 0.2591228628 | 0.95319 | 0.4336839318 | 0.102688 | 4.757222198 | 0 |
| 2 | 47463136 | A | T | S | C | MSH2 | ENST00000233146 | 0.63038 | 0.01807 | 0.21166 | 0.82319 | 0.87829 | 0.2888670854 | 0.94094 | 0.9689722657 | 0.231018 | 6.536205699 | 0 |
| 2 | 47463137 | G | A | S | N | MSH2 | ENST00000233146 | 0.37357 | 0.00586 | 0.23935 | 0.64388 | 0.66733 | 0.2736555732 | 0.8949 | 0.3501623571 | 0.0854148 | 3.332981824 | 0 |
| 2 | 47463137 | G | C | S | T | MSH2 | ENST00000233146 | 0.3637 | 0.00592 | 0.20224 | 0.63372 | 0.7609 | 0.2315473853 | 0.93969 | 0.3742465377 | 0.136678 | 1.497747885 | 0 |
| 2 | 47463137 | G | T | S | I | MSH2 | ENST00000233146 | 0.44053 | 0.00669 | 0.23619 | 0.70178 | 0.78967 | 0.3952207845 | 0.96684 | 0.9110174775 | 0.201179 | 4.858153447 | 0 |
| 2 | 47463138 | T | A | S | R | MSH2 | ENST00000233146 | 0.42998 | 0.00551 | 0.24896 | 0.648 | 0.80831 | 0.4225796023 | 0.94176 | 0.4138995409 | 0.182302 | 1.394584595 | 0 |
| 2 | 47463138 | T | G | S | R | MSH2 | ENST00000233146 | 0.43611 | 0.00551 | 0.24896 | 0.64366 | 0.80831 | 0.4225796023 | 0.94127 | 0.3515062332 | 0.18227 | 1.394584595 | 0 |
| 2 | 47463139 | G | A | A | T | MSH2 | ENST00000233146 | 0.50628 | 0.01009 | 0.30328 | 0.72416 | 0.82046 | 0.4894835843 | 0.89043 | 0.9702388048 | 0.229051 | 4.025813528 | 0 |
| 2 | 47463139 | G | C | A | P | MSH2 | ENST00000233146 | 0.84635 | 0.03124 | 0.37618 | 0.89368 | 0.94167 | 0.8150843532 | 0.86253 | 0.9909332991 | 0.300012 | 6.889372156 | 1 |
| 2 | 47463139 | G | T | A | S | MSH2 | ENST00000233146 | 0.77766 | 0.03059 | 0.26754 | 0.88338 | 0.89784 | 0.4646683454 | 0.86819 | 0.9914904237 | 0.232369 | 6.889372156 | 0 |
| 2 | 47463140 | C | A | A | E | MSH2 | ENST00000233146 | 0.68027 | 0.03118 | 0.37691 | 0.8461 | 0.94281 | 0.798761009 | 0.85957 | 0.9983010888 | 0.359287 | 2.736504899 | 0 |
| 2 | 47463140 | C | G | A | G | MSH2 | ENST00000233146 | 0.67795 | 0.02943 | 0.29159 | 0.81517 | 0.93593 | 0.4901276969 | 0.84976 | 0.997343123 | 0.27948 | 6.889372156 | 0 |
| 2 | 47463140 | C | T | A | V | MSH2 | ENST00000233146 | 0.38742 | 0.01244 | 0.32939 | 0.78189 | 0.86852 | 0.4820354652 | 0.79733 | 0.9910932183 | 0.226423 | 2.77491041 | 0 |
| 2 | 47463142 | G | A | A | T | MSH2 | ENST00000233146 | 0.89468 | 0.03633 | 0.35459 | 0.88695 | 0.95863 | 0.5206438135 | 0.92278 | 0.9945408702 | 0.388215 | 6.686328196 | 0 |
| 2 | 47463142 | G | C | A | P | MSH2 | ENST00000233146 | 0.91477 | 0.03313 | 0.42686 | 0.88425 | 0.97252 | 0.8310577755 | 0.96392 | 0.9977453351 | 0.480437 | 7.386165614 | 1 |
| 2 | 47463142 | G | T | A | S | MSH2 | ENST00000233146 | 0.72292 | 0.02844 | 0.31223 | 0.85451 | 0.89009 | 0.4324204386 | 0.8932 | 0.9655680656 | 0.287422 | 5.262496076 | 0 |
| 2 | 47463143 | C | A | A | D | MSH2 | ENST00000233146 | 0.69909 | 0.03076 | 0.47749 | 0.91372 | 0.97401 | 0.8101696087 | 0.95252 | 0.9994749427 | 0.476424 | 7.386165614 | 1 |
| 2 | 47463143 | C | G | A | G | MSH2 | ENST00000233146 | 0.52107 | 0.02218 | 0.33361 | 0.82474 | 0.89422 | 0.4587479849 | 0.90599 | 0.9880946279 | 0.332059 | 4.533871953 | 0 |
| 2 | 47463143 | C | T | A | V | MSH2 | ENST00000233146 | 0.69307 | 0.03674 | 0.38287 | 0.90023 | 0.95316 | 0.5895981923 | 0.93465 | 0.9981716871 | 0.467921 | 7.031645426 | 0 |
| 2 | 47463145 | A | G | R | G | MSH2 | ENST00000233146 | 0.59871 | 0.01959 | 0.18418 | 0.79545 | 0.89009 | 0.4554255576 | 0.98067 | 0.9953857064 | 0.337043 | 6.800598837 | 0 |
| 2 | 47463146 | G | A | R | K | MSH2 | ENST00000233146 | 0.41534 | 0.00718 | 0.23425 | 0.49603 | 0.73589 | 0.3580628154 | 0.93169 | 0.1288528442 | -0.0454826 | 2.016617125 | 0 |
| 2 | 47463146 | G | C | R | T | MSH2 | ENST00000233146 | 0.42914 | 0.00726 | 0.22354 | 0.65674 | 0.81818 | 0.4699807423 | 0.97483 | 0.906352222 | 0.292672 | 3.938632675 | 0 |
| 2 | 47463146 | G | T | R | I | MSH2 | ENST00000233146 | 0.76737 | 0.02381 | 0.22429 | 0.80972 | 0.89422 | 0.4652864308 | 0.92531 | 0.9930745363 | 0.283177 | 5.817602958 | 0 |
| 2 | 47463147 | A | C | R | S | MSH2 | ENST00000233146 | 0.46606 | 0.00718 | 0.19035 | 0.69272 | 0.8278 | 0.471309234 | 0.93698 | 0.4029690325 | 0.204768 | 4.76906533 | 0 |
| 2 | 47463147 | A | T | R | S | MSH2 | ENST00000233146 | 0.46084 | 0.00718 | 0.19035 | 0.69667 | 0.82836 | 0.471309234 | 0.93732 | 0.4029690325 | 0.204767 | 4.76906533 | 0 |
| 2 | 47463148 | G | A | D | N | MSH2 | ENST00000233146 | 0.9384 | 0.02243 | 0.14406 | 0.81161 | 0.80298 | 0.3136814291 | 0.89272 | 0.9886948466 | 0.135538 | 4.022530197 | 0 |
| 2 | 47463148 | G | C | D | H | MSH2 | ENST00000233146 | 0.94364 | 0.02937 | 0.15686 | 0.84887 | 0.84641 | 0.4389321273 | 0.82775 | 0.9959671497 | 0.191698 | 4.676455449 | 0 |

| | | | | | | | | | | | | | | | | |
|---|---|---|---|---|---|---|---|---|---|---|---|---|---|---|---|---|
| 2 | 47463148 | G | T | D | Y | MSH2 | ENST00000233146 | 0.94629 | 0.03046 | 0.15674 | 0.86157 | 0.88354 | 0.5191075582 | 0.89773 | 0.9953304529 | 0.279273 | 5.823971242 | 0 |
| 2 | 47463149 | A | C | D | A | MSH2 | ENST00000233146 | 0.52999 | 0.01485 | 0.12118 | 0.8703 | 0.80595 | 0.4264511772 | 0.89233 | 0.9894475937 | 0.22947 | 2.806781444 | 0 |
| 2 | 47463149 | A | G | D | G | MSH2 | ENST00000233146 | 0.5183 | 0.00532 | 0.11565 | 0.74823 | 0.74867 | 0.4773378211 | 0.91003 | 0.9176360965 | -0.0161781 | 5.465731026 | 0 |
| 2 | 47463149 | A | T | D | V | MSH2 | ENST00000233146 | 0.52785 | 0.00821 | 0.10156 | 0.87152 | 0.85482 | 0.4802240371 | 0.94965 | 0.9965774417 | 0.239383 | 3.594628464 | 0 |
| 2 | 47463150 | T | A | D | E | MSH2 | ENST00000233146 | 0.15875 | 0.00608 | 0.12137 | 0.64078 | 0.57877 | 0.1859393304 | 0.78563 | 0.8036691546 | -0.0175466 | -0.5037387925 | 0 |
| 2 | 47463150 | T | G | D | E | MSH2 | ENST00000233146 | 0.16228 | 0.00608 | 0.12137 | 0.64164 | 0.57877 | 0.1859393304 | 0.78442 | 0.8036691546 | -0.0175571 | -0.5037387925 | 0 |
| 2 | 47463151 | C | A | L | I | MSH2 | ENST00000233146 | 0.55683 | 0.03602 | 0.32441 | 0.87924 | 0.86503 | 0.3140788061 | 0.86182 | 0.9892718792 | 0.207157 | 8.00617642 | 0 |
| 2 | 47463151 | C | G | L | V | MSH2 | ENST00000233146 | 0.55442 | 0.03706 | 0.26515 | 0.8861 | 0.89285 | 0.4981962424 | 0.895 | 0.9914585948 | 0.286441 | 6.908548282 | 0 |
| 2 | 47463152 | T | A | L | H | MSH2 | ENST00000233146 | 0.73069 | 0.0334 | 0.39624 | 0.92787 | 0.91694 | 0.7181906387 | 0.96377 | 0.9795548955 | 0.480379 | 8.00617642 | 0 |
| 2 | 47463152 | T | C | L | P | MSH2 | ENST00000233146 | 0.75543 | 0.03006 | 0.43698 | 0.94079 | 0.92884 | 0.856992682 | 0.98504 | 0.9992142916 | 0.501303 | 8.00617642 | 1 |
| 2 | 47463152 | T | G | L | R | MSH2 | ENST00000233146 | 0.75089 | 0.0315 | 0.39024 | 0.94447 | 0.92964 | 0.8191280385 | 0.98076 | 0.9993357062 | 0.501044 | 8.00617642 | 1 |
| 2 | 47463154 | G | A | G | S | MSH2 | ENST00000233146 | 0.50922 | 0.01591 | 0.49502 | 0.73843 | 0.83834 | 0.49995092 | 0.90224 | 0.9845246673 | 0.213018 | 2.516909126 | 0 |
| 2 | 47463154 | G | C | G | R | MSH2 | ENST00000233146 | 0.88672 | 0.03144 | 0.60001 | 0.7817 | 0.93593 | 0.5714060912 | 0.94692 | 0.9939269423 | 0.408216 | 6.284167622 | 0 |
| 2 | 47463154 | G | T | G | C | MSH2 | ENST00000233146 | 0.60742 | 0.03602 | 0.55497 | 0.85077 | 0.93863 | 0.5700069816 | 0.96387 | 0.9943888783 | 0.435745 | 7.229405957 | 0 |
| 2 | 47466658 | G | A | G | D | MSH2 | ENST00000233146 | 0.48934 | 0.03029 | 0.61578 | 0.69803 | 0.92438 | 0.5013074082 | 0.92956 | 0.9486067295 | 0.283963 | 5.814929412 | 0 |
| 2 | 47466658 | G | C | G | A | MSH2 | ENST00000233146 | 0.75312 | 0.02896 | 0.50186 | 0.78633 | 0.90885 | 0.4764543817 | 0.88915 | 0.9953736663 | 0.29774 | 5.169732463 | 0 |
| 2 | 47466658 | G | T | G | V | MSH2 | ENST00000233146 | 0.80835 | 0.03219 | 0.50658 | 0.82641 | 0.95499 | 0.6336971347 | 0.93171 | 0.997695148 | 0.459859 | 7.289479849 | 0 |
| 2 | 47466660 | T | A | L | M | MSH2 | ENST00000233146 | 0.63 | 0.03653 | 0.30165 | 0.8167 | 0.85064 | 0.4114074512 | 0.86497 | 0.7773485184 | 0.111045 | 7.574744048 | 0 |
| 2 | 47466660 | T | G | L | V | MSH2 | ENST00000233146 | 0.70704 | 0.01888 | 0.2675 | 0.86612 | 0.90842 | 0.4312547343 | 0.94375 | 0.627833724 | 0.223682 | 3.508373899 | 0 |
| 2 | 47466661 | T | C | L | S | MSH2 | ENST00000233146 | 0.93474 | 0.03161 | 0.34647 | 0.90895 | 0.97512 | 0.7622960809 | 0.92835 | 0.9969512224 | 0.439841 | 7.640436644 | 0 |
| 2 | 47466662 | G | C | L | F | MSH2 | ENST00000233146 | 0.78661 | 0.03442 | 0.31481 | 0.86041 | 0.92438 | 0.5401324851 | 0.87262 | 0.9857926965 | 0.240762 | 7.640436644 | 0 |
| 2 | 47466662 | G | T | L | F | MSH2 | ENST00000233146 | 0.78019 | 0.03442 | 0.31481 | 0.86089 | 0.92438 | 0.5401324851 | 0.87427 | 0.9854835272 | 0.240757 | 7.640436644 | 0 |
| 2 | 47466663 | G | A | D | N | MSH2 | ENST00000233146 | 0.89988 | 0.01948 | 0.17191 | 0.80968 | 0.825 | 0.2835572224 | 0.87503 | 0.9836072922 | 0.157567 | 1.275595305 | 0 |
| 2 | 47466663 | G | C | D | H | MSH2 | ENST00000233146 | 0.9058 | 0.02775 | 0.18088 | 0.87688 | 0.89101 | 0.4600946131 | 0.84429 | 0.9943233728 | 0.28885 | 2.176952229 | 0 |
| 2 | 47466663 | G | T | D | Y | MSH2 | ENST00000233146 | 0.9145 | 0.03046 | 0.18648 | 0.89987 | 0.92884 | 0.5586671958 | 0.97187 | 0.9969898462 | 0.449366 | 5.626025028 | 0 |
| 2 | 47466664 | A | C | D | A | MSH2 | ENST00000233146 | 0.76355 | 0.01485 | 0.14577 | 0.8852 | 0.86603 | 0.4537025946 | 0.83816 | 0.9814890027 | 0.260369 | 2.97962489 | 0 |
| 2 | 47466664 | A | G | D | G | MSH2 | ENST00000233146 | 0.77877 | 0.0167 | 0.14098 | 0.88446 | 0.89009 | 0.5027620013 | 0.88155 | 0.9826697111 | 0.266351 | 2.376387826 | 0 |
| 2 | 47466664 | A | T | D | V | MSH2 | ENST00000233146 | 0.77473 | 0.02396 | 0.12743 | 0.93046 | 0.92844 | 0.5246434865 | 0.92483 | 0.9924069047 | 0.326178 | 5.626025028 | 0 |
| 2 | 47466665 | C | A | D | E | MSH2 | ENST00000233146 | 0.11606 | 0.00596 | 0.14518 | 0.62517 | 0.76487 | 0.1784918172 | 0.84065 | 0.04822326069 | 0.0224567 | -1.872039476 | 0 |
| 2 | 47466665 | C | G | D | E | MSH2 | ENST00000233146 | 0.11938 | 0.00596 | 0.14518 | 0.6262 | 0.76421 | 0.1784918172 | 0.84046 | 0.04940690286 | 0.0224706 | -1.872039476 | 0 |
| 2 | 47466666 | C | A | P | T | MSH2 | ENST00000233146 | 0.41848 | 0.00562 | 0.08869 | 0.73324 | 0.7816 | 0.3649063034 | 0.89 | 0.9237390161 | 0.163942 | 6.895209801 | 0 |
| 2 | 47466666 | C | G | P | A | MSH2 | ENST00000233146 | 0.32696 | 0.00574 | 0.08707 | 0.39125 | 0.68203 | 0.2182188555 | 0.6733 | 0.5178459883 | 0.070883 | -0.3262245883 | 0 |
| 2 | 47466666 | C | T | P | S | MSH2 | ENST00000233146 | 0.46016 | 0.00574 | 0.07776 | 0.67727 | 0.72474 | 0.3028484311 | 0.86922 | 0.7307716608 | 0.105769 | 4.49969339 | 0 |
| 2 | 47466667 | C | A | P | H | MSH2 | ENST00000233146 | 0.51501 | 0.0177 | 0.11455 | 0.77221 | 0.82443 | 0.3555144166 | 0.72113 | 0.9323232174 | 0.18965 | 6.895209801 | 0 |
| 2 | 47466667 | C | G | P | R | MSH2 | ENST00000233146 | 0.39185 | 0.00777 | 0.13016 | 0.66459 | 0.7217 | 0.3812225831 | 0.81376 | 0.8160836697 | 0.218737 | 6.895209801 | 0 |
| 2 | 47466667 | C | T | P | L | MSH2 | ENST00000233146 | 0.38851 | 0.00669 | 0.13548 | 0.81458 | 0.71627 | 0.3610913275 | 0.80363 | 0.84902215 | 0.146708 | 6.895209801 | 0 |
| 2 | 47466669 | G | A | G | S | MSH2 | ENST00000233146 | 0.6712 | 0.01365 | 0.37943 | 0.73948 | 0.84159 | 0.5533975166 | 0.85032 | 0.9489505291 | 0.162952 | 2.734894402 | 0 |
| 2 | 47466669 | G | C | G | R | MSH2 | ENST00000233146 | 0.94775 | 0.02984 | 0.49727 | 0.89945 | 0.94281 | 0.6515079847 | 0.9386 | 0.9942130446 | 0.404364 | 7.629996171 | 0 |
| 2 | 47466669 | G | T | G | C | MSH2 | ENST00000233146 | 0.95289 | 0.03518 | 0.43483 | 0.91937 | 0.96262 | 0.6289516638 | 0.94178 | 0.9953161478 | 0.403526 | 7.629996171 | 0 |
| 2 | 47466670 | G | A | G | D | MSH2 | ENST00000233146 | 0.66138 | 0.02687 | 0.51571 | 0.72893 | 0.86951 | 0.5898914741 | 0.91537 | 0.9477601647 | 0.326768 | 7.629996171 | 0 |
| 2 | 47466670 | G | C | G | A | MSH2 | ENST00000233146 | 0.58545 | 0.00832 | 0.38508 | 0.59076 | 0.73589 | 0.5301898533 | 0.85349 | 0.8994409442 | 0.162614 | 4.421317425 | 0 |
| 2 | 47466670 | G | T | G | V | MSH2 | ENST00000233146 | 0.8203 | 0.0292 | 0.4056 | 0.83527 | 0.93631 | 0.6145178019 | 0.92083 | 0.9961530566 | 0.420394 | 7.629996171 | 0 |
| 2 | 47466672 | A | C | K | Q | MSH2 | ENST00000233146 | 0.88573 | 0.03613 | 0.40086 | 0.88744 | 0.96371 | 0.6406804125 | 0.96136 | 0.9981472492 | 0.348426 | 9.744706171 | 0 |
| 2 | 47466672 | A | G | K | E | MSH2 | ENST00000233146 | 0.9057 | 0.032 | 0.4435 | 0.8996 | 0.96153 | 0.689108073 | 0.97281 | 0.9981472492 | 0.491881 | 9.744706171 | 0 |
| 2 | 47466673 | A | C | K | T | MSH2 | ENST00000233146 | 0.89867 | 0.03426 | 0.4551 | 0.91114 | 0.96407 | 0.6758163393 | 0.95753 | 0.9988979101 | 0.484519 | 9.744706171 | 0 |
| 2 | 47466673 | A | G | K | R | MSH2 | ENST00000233146 | 0.88715 | 0.03985 | 0.36828 | 0.86927 | 0.93631 | 0.5360516593 | 0.96406 | 0.9973927736 | 0.356299 | 9.325234958 | 0 |
| 2 | 47466673 | A | T | K | I | MSH2 | ENST00000233146 | 0.92552 | 0.02857 | 0.41122 | 0.92417 | 0.9648 | 0.6996668346 | 0.94621 | 0.999524951 | 0.455047 | 9.744706171 | 0 |
| 2 | 47466674 | A | C | K | N | MSH2 | ENST00000233146 | 0.80795 | 0.03458 | 0.47024 | 0.89526 | 0.88354 | 0.6638234209 | 0.96123 | 0.999281466 | 0.184547 | 9.744706171 | 0 |
| 2 | 47466674 | A | T | K | N | MSH2 | ENST00000233146 | 0.80124 | 0.03458 | 0.47024 | 0.8972 | 0.88354 | 0.6638234209 | 0.96134 | 0.9993848801 | 0.184541 | 9.744706171 | 0 |
| 2 | 47466675 | C | A | Q | K | MSH2 | ENST00000233146 | 0.49991 | 0.00687 | 0.20153 | 0.59932 | 0.74235 | 0.4935676875 | 0.94489 | 0.8996169567 | 0.149399 | -0.3744003996 | 0 |
| 2 | 47466675 | C | G | Q | E | MSH2 | ENST00000233146 | 0.49782 | 0.00664 | 0.16598 | 0.58095 | 0.75005 | 0.473943385 | 0.92934 | 0.8805832863 | 0.140851 | 3.130297539 | 0 |
| 2 | 47466676 | A | C | Q | P | MSH2 | ENST00000233146 | 0.62863 | 0.00965 | 0.16511 | 0.75019 | 0.82556 | 0.7446261912 | 0.96913 | 0.842127502 | 0.226652 | 2.048809794 | 0 |
| 2 | 47466676 | A | G | Q | R | MSH2 | ENST00000233146 | 0.58881 | 0.0073 | 0.19346 | 0.66124 | 0.76024 | 0.4845863703 | 0.96191 | 0.7268619537 | 0.172738 | 3.130297539 | 0 |
| 2 | 47466676 | A | T | Q | L | MSH2 | ENST00000233146 | 0.6209 | 0.00681 | 0.15894 | 0.69996 | 0.77845 | 0.5572464381 | 0.97714 | 0.8400850296 | 0.195166 | 3.130297539 | 0 |
| 2 | 47466677 | G | C | Q | H | MSH2 | ENST00000233146 | 0.62597 | 0.00592 | 0.19409 | 0.68856 | 0.74517 | 0.5656267874 | 0.96442 | 0.2520363033 | 0.0967242 | 2.879710005 | 0 |
| 2 | 47466677 | G | T | Q | H | MSH2 | ENST00000233146 | 0.61985 | 0.00592 | 0.19409 | 0.68847 | 0.74517 | 0.5656267874 | 0.9644 | 0.838280797 | 0.107488 | 2.879710005 | 0 |
| 2 | 47466678 | A | C | I | L | MSH2 | ENST00000233146 | 0.38255 | 0.00615 | 0.37174 | 0.47032 | 0.68734 | 0.4129547371 | 0.83069 | 0.756757319 | 0.121978 | 1.292957365 | 0 |
| 2 | 47466678 | A | G | I | V | MSH2 | ENST00000233146 | 0.35897 | 0.00633 | 0.28263 | 0.46295 | 0.66639 | 0.2308149866 | 0.85455 | 0.8699114919 | 0.102962 | 1.4695502 | 0 |
| 2 | 47466678 | A | T | I | F | MSH2 | ENST00000233146 | 0.77378 | 0.0233 | 0.38901 | 0.88129 | 0.92924 | 0.6518649299 | 0.87288 | 0.9821182489 | 0.25362 | 6.144851263 | 0 |
| 2 | 47466679 | T | A | I | N | MSH2 | ENST00000233146 | 0.83951 | 0.02556 | 0.46902 | 0.91625 | 0.93824 | 0.7466219446 | 0.83253 | 0.9963102937 | 0.288025 | 6.144851263 | 1 |
| 2 | 47466679 | T | C | I | T | MSH2 | ENST00000233146 | 0.77077 | 0.02307 | 0.49732 | 0.89756 | 0.95316 | 0.6373916416 | 0.89262 | 0.9885187745 | 0.347207 | 6.144851263 | 0 |
| 2 | 47466679 | T | G | I | S | MSH2 | ENST00000233146 | 0.8349 | 0.02366 | 0.39681 | 0.9202 | 0.96298 | 0.8088990105 | 0.91808 | 0.9955300689 | 0.406577 | 6.144851263 | 1 |
| 2 | 47466680 | T | G | I | M | MSH2 | ENST00000233146 | 0.6666 | 0.01075 | 0.33788 | 0.71887 | 0.8517 | 0.4822703027 | 0.88108 | 0.8019164205 | 0.178032 | 2.714649228 | 0 |
| 2 | 47466681 | A | C | K | Q | MSH2 | ENST00000233146 | 0.84748 | 0.03621 | 0.53454 | 0.84496 | 0.93593 | 0.6651539369 | 0.96382 | 0.9936722517 | 0.384789 | 10.13582754 | 0 |
| 2 | 47466681 | A | G | K | E | MSH2 | ENST00000233146 | 0.87072 | 0.03219 | 0.57592 | 0.85229 | 0.94053 | 0.7273658933 | 0.97655 | 0.992660284 | 0.461606 | 10.13582754 | 1 |
| 2 | 47466682 | A | C | K | T | MSH2 | ENST00000233146 | 0.85545 | 0.03458 | 0.59212 | 0.86794 | 0.95097 | 0.6554654082 | 0.94516 | 0.9962986112 | 0.450439 | 10.13582754 | 0 |
| 2 | 47466682 | A | G | K | R | MSH2 | ENST00000233146 | 0.84816 | 0.0401 | 0.49814 | 0.79496 | 0.87491 | 0.6039141466 | 0.96675 | 0.9953623414 | 0.336708 | 5.68312642 | 0 |
| 2 | 47466682 | A | T | K | I | MSH2 | ENST00000233146 | 0.89509 | 0.02864 | 0.5424 | 0.88669 | 0.96371 | 0.6948111954 | 0.9272 | 0.9782656604 | 0.422264 | 10.13582754 | 0 |
| 2 | 47466684 | C | A | L | M | MSH2 | ENST00000233146 | 0.56048 | 0.03761 | 0.49292 | 0.89674 | 0.92028 | 0.4847737528 | 0.87658 | 0.9891459942 | 0.259544 | 7.98332729 | 0 |
| 2 | 47466684 | C | G | L | V | MSH2 | ENST00000233146 | 0.54015 | 0.03674 | 0.45762 | 0.93096 | 0.94655 | 0.5028417769 | 0.93103 | 0.9851562977 | 0.351641 | 3.637465143 | 0 |
| 2 | 47466685 | T | A | L | Q | MSH2 | ENST00000233146 | 0.91917 | 0.03646 | 0.53193 | 0.92096 | 0.98367 | 0.8254676994 | 0.97862 | 0.9989538193 | 0.484264 | 8.615982805 | 0 |
| 2 | 47466685 | T | C | L | P | MSH2 | ENST00000233146 | 0.92667 | 0.03001 | 0.64047 | 0.92061 | 0.98834 | 0.9076483444 | 0.98155 | 0.9990763664 | 0.484544 | 8.615982805 | 1 |
| 2 | 47466685 | T | G | L | R | MSH2 | ENST00000233146 | 0.9246 | 0.03144 | 0.61021 | 0.92201 | 0.98513 | 0.8847647686 | 0.98044 | 0.9989538193 | 0.484244 | 8.615982805 | 0 |
| 2 | 47466687 | G | A | D | N | MSH2 | ENST00000233146 | 0.88164 | 0.01628 | 0.36553 | 0.81108 | 0.83888 | 0.3640731145 | 0.86981 | 0.9716863632 | 0.141794 | 2.763291196 | 0 |
| 2 | 47466687 | G | C | D | H | MSH2 | ENST00000233146 | 0.70495 | 0.02802 | 0.39024 | 0.83893 | 0.89239 | 0.4707817345 | 0.90735 | 0.7934236201 | 0.285839 | 5.279251404 | 0 |
| 2 | 47466687 | G | T | D | Y | MSH2 | ENST00000233146 | 0.90009 | 0.02943 | 0.37738 | 0.84827 | 0.93709 | 0.5344339071 | 0.96102 | 0.9978019595 | 0.358887 | 5.279251404 | 0 |
| 2 | 47466688 | A | C | D | A | MSH2 | ENST00000233146 | 0.74285 | 0.01082 | 0.31982 | 0.84253 | 0.90494 | 0.4704112524 | 0.91113 | 0.9551829696 | 0.276332 | 3.619683458 | 0 |
| 2 | 47466688 | A | G | D | G | MSH2 | ENST00000233146 | 0.71201 | 0.0167 | 0.31292 | 0.85143 | 0.90537 | 0.5772405342 | 0.94338 | 0.99184376 | 0.293575 | 0.3997593433 | 0 |
| 2 | 47466688 | A | T | D | V | MSH2 | ENST00000233146 | 0.89131 | 0.01823 | 0.29753 | 0.90679 | 0.95863 | 0.5247723696 | 0.96777 | 0.9939816594 | 0.361401 | 5.279251404 | 0 |
| 2 | 47466690 | T | A | S | T | MSH2 | ENST00000233146 | 0.46444 | 0.00855 | 0.33288 | 0.47418 | 0.74164 | 0.2716626153 | 0.91087 | 0.7555652261 | 0.18242 | 3.328896812 | 0 |
| 2 | 47466690 | T | C | S | P | MSH2 | ENST00000233146 | 0.87782 | 0.03039 | 0.34731 | 0.77078 | 0.93515 | 0.7145928044 | 0.94441 | 0.9829909801 | 0.386319 | 6.566594621 | 0 |
| 2 | 47466690 | T | G | S | A | MSH2 | ENST00000233146 | 0.52879 | 0.01096 | 0.31917 | 0.69342 | 0.85534 | 0.2086033684 | 0.90905 | 0.9445254207 | 0.217991 | 6.566594621 | 0 |
| 2 | 47466691 | C | A | S | Y | MSH2 | ENST00000233146 | 0.40952 | 0.03175 | 0.40137 | 0.78342 | 0.88823 | 0.4557804096 | 0.96304 | 0.9766051769 | 0.363377 | 2.608704153 | 0 |
| 2 | 47466691 | C | G | S | C | MSH2 | ENST00000233146 | 0.31946 | 0.00958 | 0.34608 | 0.70253 | 0.79212 | 0.3708934947 | 0.872 | 0.5434202666 | 0.201376 | 3.86604199 | 0 |
| 2 | 47466691 | C | T | S | F | MSH2 | ENST00000233146 | 0.43841 | 0.03212 | 0.37276 | 0.85922 | 0.89648 | 0.4224082493 | 0.96716 | 0.9693604708 | 0.381142 | 2.590133979 | 0 |
| 2 | 47466693 | A | C | S | R | MSH2 | ENST00000233146 | 0.64012 | 0.00696 | 0.28701 | 0.70882 | 0.79212 | 0.451380151 | 0.92426 | 0.8092009793 | 0.252101 | 5.401848594 | 0 |
| 2 | 47466693 | A | G | S | G | MSH2 | ENST00000233146 | 0.6075 | 0.00687 | 0.21169 | 0.65392 | 0.69762 | 0.3073620529 | 0.91654 | 0.4443015867 | 0.148258 | 5.401848594 | 0 |
| 2 | 47466693 | A | T | S | C | MSH2 | ENST00000233146 | 0.82918 | 0.022 | 0.2424 | 0.82527 | 0.91271 | 0.3109892469 | 0.88322 | 0.9833879471 | 0.242749 | 5.401848594 | 0 |
| 2 | 47466694 | G | A | S | N | MSH2 | ENST00000233146 | 0.23275 | 0.0058 | 0.26736 | 0.50178 | 0.613 | 0.1999433019 | 0.67091 | 0.2012773752 | 0.00992353 | -1.753180355 | 0 |
| 2 | 47466694 | G | C | S | T | MSH2 | ENST00000233146 | 0.30264 | 0.00603 | 0.23337 | 0.71672 | 0.76487 | 0.2417398237 | 0.92758 | 0.2542241812 | 0.0478137 | 2.315173497 | 0 |
| 2 | 47466694 | G | T | S | I | MSH2 | ENST00000233146 | 0.46749 | 0.01145 | 0.25429 | 0.8607 | 0.83779 | 0.4633019283 | 0.95718 | 0.9822173715 | 0.217024 | 5.401848594 | 0 |
| 2 | 47466695 | T | A | S | R | MSH2 | ENST00000233146 | 0.40418 | 0.00696 | 0.28701 | 0.8302 | 0.84428 | 0.451380151 | 0.9477 | 0.8515301943 | 0.195513 | 5.401848594 | 0 |
| 2 | 47466695 | T | G | S | R | MSH2 | ENST00000233146 | 0.41022 | 0.00696 | 0.28701 | 0.82684 | 0.84428 | 0.451380151 | 0.94735 | 0.7850497961 | 0.195497 | 5.401848594 | 0 |
| 2 | 47466696 | G | A | A | T | MSH2 | ENST00000233146 | 0.25797 | 0.00592 | 0.06847 | 0.58144 | 0.7147 | 0.3140950697 | 0.87882 | 0.1596284807 | 0.084358 | 3.780590997 | 0 |
| 2 | 47466696 | G | C | A | P | MSH2 | ENST00000233146 | 0.28553 | 0.0058 | 0.09379 | 0.80272 | 0.82556 | 0.5665171123 | 0.97681 | 0.2335573137 | 0.203612 | 6.175723411 | 0 |
| 2 | 47466696 | G | T | A | S | MSH2 | ENST00000233146 | 0.20869 | 0.00599 | 0.05907 | 0.61916 | 0.6757 | 0.2413602389 | 0.76896 | 0.1096399873 | 0.0555463 | 0.9247971463 | 0 |
| 2 | 47466697 | C | A | A | E | MSH2 | ENST00000233146 | 0.27114 | 0.01151 | 0.10085 | 0.6895 | 0.7589 | 0.4850423706 | 0.85606 | 0.7757754326 | 0.225673 | 5.618255991 | 0 |
| 2 | 47466697 | C | G | A | G | MSH2 | ENST00000233146 | 0.29109 | 0.01053 | 0.07232 | 0.57848 | 0.73 | 0.2346063177 | 0.80816 | 0.8754310608 | 0.137265 | 6.115826362 | 0 |
| 2 | 47466697 | C | T | A | V | MSH2 | ENST00000233146 | 0.31049 | 0.01211 | 0.08608 | 0.62248 | 0.77527 | 0.3592215791 | 0.83987 | 0.8635677099 | 0.222203 | 6.175723411 | 0 |
| 2 | 47466699 | C | A | Q | K | MSH2 | ENST00000233146 | 0.44442 | 0.00821 | 0.2956 | 0.76071 | 0.77909 | 0.486935034 | 0.94761 | 0.6168690324 | 0.259358 | 5.24057849 | 0 |
| 2 | 47466699 | C | G | Q | E | MSH2 | ENST00000233146 | 0.69139 | 0.02108 | 0.24219 | 0.69571 | 0.82273 | 0.4963018162 | 0.85369 | 0.845353961 | 0.218858 | 6.296642975 | 0 |

| | | | | | | | | | | | | | | | | |
|---|---|---|---|---|---|---|---|---|---|---|---|---|---|---|---|---|
| 2 | 47466700 | A | C | Q | P | MSH2 | ENST00000233146 | 0.86162 | 0.03083 | 0.25515 | 0.82967 | 0.94318 | 0.7435105446 | 0.95507 | 0.9885292649 | 0.408594 | 7.314970333 | 0 |
| 2 | 47466700 | A | G | Q | R | MSH2 | ENST00000233146 | 0.83004 | 0.02494 | 0.28949 | 0.78836 | 0.90624 | 0.5079562702 | 0.94529 | 0.9771186709 | 0.306583 | 7.314970333 | 0 |
| 2 | 47466700 | A | T | Q | L | MSH2 | ENST00000233146 | 0.71834 | 0.01549 | 0.23915 | 0.8044 | 0.86302 | 0.4788558297 | 0.93126 | 0.7684792876 | 0.118914 | 6.263999195 | 0 |
| 2 | 47466701 | G | C | Q | H | MSH2 | ENST00000233146 | 0.33441 | 0.01015 | 0.28551 | 0.74387 | 0.88259 | 0.5695887966 | 0.97077 | 0.9210538864 | 0.244087 | 7.314970333 | 0 |
| 2 | 47466701 | G | T | Q | H | MSH2 | ENST00000233146 | 0.33009 | 0.01015 | 0.28551 | 0.74387 | 0.88259 | 0.5695887966 | 0.97143 | 0.9353161454 | 0.244082 | 7.314970333 | 0 |
| 2 | 47466702 | T | A | F | I | MSH2 | ENST00000233146 | 0.46768 | 0.00527 | 0.39482 | 0.60671 | 0.68646 | 0.352480396 | 0.90972 | 0.2224411113 | 0.0742796 | 1.985473432 | 0 |
| 2 | 47466702 | T | C | F | L | MSH2 | ENST00000233146 | 0.41416 | 0.00543 | 0.42547 | 0.52198 | 0.68558 | 0.3059141938 | 0.85305 | 0.5802417397 | 0.0579446 | -0.01983797559 | 0 |
| 2 | 47466702 | T | G | F | V | MSH2 | ENST00000233146 | 0.50101 | 0.00527 | 0.34901 | 0.64601 | 0.78348 | 0.4233672091 | 0.95518 | 0.323439151 | 0.125479 | 5.226442901 | 0 |
| 2 | 47466703 | T | A | F | Y | MSH2 | ENST00000233146 | 0.34462 | 0.00556 | 0.41592 | 0.6262 | 0.64977 | 0.3532026593 | 0.85938 | 0.3832938969 | 0.0890982 | 1.955332013 | 0 |
| 2 | 47466703 | T | C | F | S | MSH2 | ENST00000233146 | 0.45193 | 0.00492 | 0.38808 | 0.71302 | 0.80653 | 0.5676569959 | 0.95223 | 0.7644256353 | 0.227374 | 3.41481121 | 0 |
| 2 | 47466703 | T | G | F | C | MSH2 | ENST00000233146 | 0.76634 | 0.02231 | 0.42127 | 0.82111 | 0.87198 | 0.4927273101 | 0.86622 | 0.9669643044 | 0.236654 | 5.226442901 | 0 |
| 2 | 47466704 | T | A | F | L | MSH2 | ENST00000233146 | 0.30463 | 0.00543 | 0.42547 | 0.5135 | 0.71312 | 0.3059141938 | 0.87717 | 0.469652921 | 0.0316949 | -0.01983797559 | 0 |
| 2 | 47466704 | T | G | F | L | MSH2 | ENST00000233146 | 0.30992 | 0.00543 | 0.42547 | 0.51291 | 0.71312 | 0.3059141938 | 0.87788 | 0.469652921 | 0.0316726 | -0.01983797559 | 0 |
| 2 | 47466705 | G | A | G | R | MSH2 | ENST00000233146 | 0.90906 | 0.03442 | 0.59081 | 0.95496 | 0.99498 | 0.810217587 | 0.95339 | 0.9984519482 | 0.488572 | 8.502966506 | 0 |
| 2 | 47466705 | G | C | G | R | MSH2 | ENST00000233146 | 0.9071 | 0.03442 | 0.59081 | 0.95376 | 0.99498 | 0.810217587 | 0.95213 | 0.9984519482 | 0.488576 | 8.502966506 | 0 |
| 2 | 47466706 | G | C | G | A | MSH2 | ENST00000233146 | 0.74221 | 0.03595 | 0.49133 | 0.90022 | 0.98146 | 0.7668531228 | 0.97373 | 0.9978656173 | 0.534382 | 9.651408919 | 0 |
| 2 | 47466706 | G | T | G | V | MSH2 | ENST00000233146 | 0.77378 | 0.0337 | 0.49752 | 0.92035 | 0.98693 | 0.8056172077 | 0.97347 | 0.9993671775 | 0.540362 | 9.651408919 | 0 |
| 2 | 47466708 | T | A | Y | N | MSH2 | ENST00000233146 | 0.85691 | 0.01861 | 0.39946 | 0.94973 | 0.91986 | 0.6873864182 | 0.9062 | 0.9888044 | 0.301273 | 6.134528495 | 0 |
| 2 | 47466708 | T | C | Y | H | MSH2 | ENST00000233146 | 0.53839 | 0.00592 | 0.3969 | 0.40862 | 0.75755 | 0.5883212181 | 0.83394 | 0.8701520562 | 0.135626 | 1.623714805 | 0 |
| 2 | 47466708 | T | G | Y | D | MSH2 | ENST00000233146 | 0.86453 | 0.02172 | 0.39187 | 0.95179 | 0.92884 | 0.7635467663 | 0.92107 | 0.9974344373 | 0.335252 | 6.134528495 | 1 |
| 2 | 47466709 | A | C | Y | S | MSH2 | ENST00000233146 | 0.87892 | 0.01973 | 0.32349 | 0.95413 | 0.92723 | 0.7016879818 | 0.92816 | 0.9907034636 | 0.299355 | 6.134528495 | 0 |
| 2 | 47466709 | A | G | Y | C | MSH2 | ENST00000233146 | 0.90339 | 0.00687 | 0.38408 | 0.93994 | 0.90624 | 0.6555262073 | 0.90402 | 0.9926803708 | 0.233068 | 6.134528495 | 0 |
| 2 | 47466709 | A | T | Y | F | MSH2 | ENST00000233146 | 0.48082 | 0.00615 | 0.30112 | 0.65518 | 0.71391 | 0.3375278619 | 0.86536 | 0.735322535 | 0.0926459 | 1.541383488 | 0 |
| 2 | 47466711 | T | A | Y | N | MSH2 | ENST00000233146 | 0.79721 | 0.02143 | 0.44116 | 0.78254 | 0.911 | 0.6621968964 | 0.90113 | 0.9856548905 | 0.276563 | 6.966746127 | 0 |
| 2 | 47466711 | T | C | Y | H | MSH2 | ENST00000233146 | 0.45592 | 0.00592 | 0.44441 | 0.36203 | 0.68113 | 0.5609619734 | 0.82907 | 0.5485011935 | 0.115287 | 2.875397999 | 0 |
| 2 | 47466711 | T | G | Y | D | MSH2 | ENST00000233146 | 0.81944 | 0.02374 | 0.43082 | 0.82108 | 0.92964 | 0.7413320496 | 0.95476 | 0.9919272661 | 0.401105 | 6.966746127 | 1 |
| 2 | 47466712 | A | C | Y | S | MSH2 | ENST00000233146 | 0.89155 | 0.02237 | 0.35968 | 0.73472 | 0.91442 | 0.661711137 | 0.96041 | 0.9835661054 | 0.364433 | 5.126452913 | 0 |
| 2 | 47466712 | A | G | Y | C | MSH2 | ENST00000233146 | 0.79911 | 0.03144 | 0.43777 | 0.6774 | 0.8887 | 0.5780228667 | 0.93177 | 0.9619368911 | 0.333523 | 4.365852669 | 0 |
| 2 | 47466712 | A | T | Y | F | MSH2 | ENST00000233146 | 0.4585 | 0.0063 | 0.34398 | 0.40992 | 0.55468 | 0.3132319425 | 0.83672 | 0.7529487014 | 0.0930852 | 0.767443288 | 0 |
| 2 | 47466714 | T | A | F | I | MSH2 | ENST00000233146 | 0.82428 | 0.02908 | 0.64306 | 0.84242 | 0.92561 | 0.5754100195 | 0.82924 | 0.9919369221 | 0.264273 | 3.433862498 | 1 |
| 2 | 47466714 | T | C | F | L | MSH2 | ENST00000233146 | 0.52227 | 0.01096 | 0.68998 | 0.59459 | 0.75552 | 0.5539791225 | 0.76438 | 0.7673378587 | 0.157986 | 3.461966432 | 0 |
| 2 | 47466714 | T | G | F | V | MSH2 | ENST00000233146 | 0.81591 | 0.02994 | 0.6139 | 0.8954 | 0.94913 | 0.573608355 | 0.92973 | 0.9894467592 | 0.399901 | 7.616104673 | 1 |
| 2 | 47466715 | T | A | F | Y | MSH2 | ENST00000233146 | 0.92337 | 0.03431 | 0.66692 | 0.89135 | 0.92233 | 0.5512417989 | 0.8811 | 0.9814838767 | 0.25306 | 6.846852442 | 0 |
| 2 | 47466715 | T | C | F | S | MSH2 | ENST00000233146 | 0.94924 | 0.0279 | 0.64794 | 0.92406 | 0.96737 | 0.7928749246 | 0.93841 | 0.9968252182 | 0.455529 | 8.81279031 | 1 |
| 2 | 47466715 | T | G | F | C | MSH2 | ENST00000233146 | 0.94922 | 0.03137 | 0.69063 | 0.92864 | 0.95826 | 0.7186740846 | 0.92558 | 0.9964050055 | 0.445438 | 8.81279031 | 0 |
| 2 | 47466716 | T | A | F | L | MSH2 | ENST00000233146 | 0.32558 | 0.01096 | 0.68998 | 0.66147 | 0.79998 | 0.5539791225 | 0.79391 | 0.8941802382 | 0.130388 | 3.461966432 | 0 |
| 2 | 47466716 | T | G | F | L | MSH2 | ENST00000233146 | 0.33099 | 0.01096 | 0.68998 | 0.66147 | 0.79998 | 0.5539791225 | 0.79476 | 0.8809123039 | 0.13038 | 3.461966432 | 0 |
| 2 | 47466717 | C | A | R | S | MSH2 | ENST00000233146 | 0.82841 | 0.03527 | 0.56929 | 0.93421 | 0.96664 | 0.8178472221 | 0.95543 | 0.9916525483 | 0.44857 | 7.68846026 | 1 |
| 2 | 47466717 | C | G | R | G | MSH2 | ENST00000233146 | 0.86003 | 0.03334 | 0.55219 | 0.94793 | 0.9648 | 0.8541800171 | 0.98008 | 0.9873972436 | 0.45456 | 8.357279596 | 1 |
| 2 | 47466718 | G | A | R | H | MSH2 | ENST00000233146 | 0.83058 | 0.03388 | 0.59151 | 0.9114 | 0.97923 | 0.7219048106 | 0.96778 | 0.9728737106 | 0.359136 | 8.357279596 | 0 |
| 2 | 47466718 | G | C | R | P | MSH2 | ENST00000233146 | 0.85077 | 0.03166 | 0.60836 | 0.93144 | 0.98834 | 0.9100355395 | 0.98495 | 0.9941191673 | 0.495147 | 8.357279596 | 1 |
| 2 | 47466718 | G | T | R | L | MSH2 | ENST00000233146 | 0.83275 | 0.03379 | 0.60169 | 0.92826 | 0.9822 | 0.8149892092 | 0.98973 | 0.9966384172 | 0.494465 | 8.357279596 | 1 |
| 2 | 47466720 | G | A | V | I | MSH2 | ENST00000233146 | 0.40039 | 0.00872 | 0.48626 | 0.50416 | 0.71073 | 0.2765462235 | 0.83658 | 0.5317208767 | 0.155181 | 1.842716107 | 0 |
| 2 | 47466720 | G | C | V | L | MSH2 | ENST00000233146 | 0.4614 | 0.01479 | 0.67473 | 0.75337 | 0.80948 | 0.5890063684 | 0.96653 | 0.7580405474 | 0.335797 | 5.390308119 | 0 |
| 2 | 47466720 | G | T | V | L | MSH2 | ENST00000233146 | 0.45635 | 0.01479 | 0.67473 | 0.75314 | 0.80948 | 0.5890063684 | 0.96734 | 0.7580405474 | 0.335793 | 5.390308119 | 0 |
| 2 | 47466721 | T | A | V | E | MSH2 | ENST00000233146 | 0.76721 | 0.02943 | 0.67789 | 0.94209 | 0.98904 | 0.879450485 | 0.98276 | 0.9971621633 | 0.494659 | 8.268376931 | 1 |
| 2 | 47466721 | T | C | V | A | MSH2 | ENST00000233146 | 0.7169 | 0.03059 | 0.69338 | 0.84147 | 0.96517 | 0.6498567413 | 0.96347 | 0.9891232848 | 0.441865 | 8.268376931 | 0 |
| 2 | 47466721 | T | G | V | G | MSH2 | ENST00000233146 | 0.76235 | 0.03237 | 0.56162 | 0.94255 | 0.98549 | 0.8761026226 | 0.96847 | 0.9969153404 | 0.494946 | 8.268376931 | 0 |
| 2 | 47466723 | A | C | T | P | MSH2 | ENST00000233146 | 0.83236 | 0.02068 | 0.66824 | 0.88939 | 0.97698 | 0.7274734589 | 0.96767 | 0.9960923791 | 0.472712 | 5.918700637 | 0 |
| 2 | 47466723 | A | G | T | A | MSH2 | ENST00000233146 | 0.79255 | 0.03547 | 0.67308 | 0.89164 | 0.97661 | 0.5947465762 | 0.97087 | 0.994369328 | 0.457559 | 9.316613766 | 0 |
| 2 | 47466723 | A | T | T | S | MSH2 | ENST00000233146 | 0.5924 | 0.02927 | 0.69131 | 0.7416 | 0.90972 | 0.4295744065 | 0.93653 | 0.9442421794 | 0.28986 | 3.752883678 | 0 |
| 2 | 47466724 | C | A | T | N | MSH2 | ENST00000233146 | 0.58638 | 0.03484 | 0.77287 | 0.92725 | 0.9506 | 0.6449302914 | 0.93094 | 0.9966142774 | 0.22097 | 9.316613766 | 0 |
| 2 | 47466724 | C | G | T | S | MSH2 | ENST00000233146 | 0.44381 | 0.02927 | 0.69131 | 0.82226 | 0.90494 | 0.4295744065 | 0.91988 | 0.969951272 | 0.278824 | 3.752883678 | 0 |
| 2 | 47466724 | C | T | T | I | MSH2 | ENST00000233146 | 0.62635 | 0.03595 | 0.81305 | 0.94255 | 0.96956 | 0.6721360855 | 0.96542 | 0.9879012108 | 0.412523 | 9.316613766 | 0 |
| 2 | 47466726 | T | A | C | S | MSH2 | ENST00000233146 | 0.53435 | 0.01705 | 0.81365 | 0.77886 | 0.91694 | 0.6211654933 | 0.92048 | 0.9803433418 | 0.350905 | 3.7583139 | 0 |
| 2 | 47466726 | T | C | C | R | MSH2 | ENST00000233146 | 0.45912 | 0.00483 | 0.89715 | 0.4963 | 0.74867 | 0.7188147557 | 0.9022 | 0.8668646812 | 0.203304 | 1.034691365 | 0 |
| 2 | 47466726 | T | G | C | G | MSH2 | ENST00000233146 | 0.71185 | 0.01924 | 0.7818 | 0.8372 | 0.93123 | 0.6685256232 | 0.92567 | 0.9893767834 | 0.374522 | 6.202451163 | 0 |
| 2 | 47466727 | G | A | C | Y | MSH2 | ENST00000233146 | 0.5324 | 0.00574 | 0.9083 | 0.76189 | 0.81124 | 0.6620744995 | 0.8675 | 0.9660961628 | 0.220936 | 3.423446779 | 0 |
| 2 | 47466727 | G | C | C | S | MSH2 | ENST00000233146 | 0.54163 | 0.01705 | 0.81365 | 0.76856 | 0.91442 | 0.6211654933 | 0.91771 | 0.9807597995 | 0.350993 | 3.7583139 | 0 |
| 2 | 47466727 | G | T | C | F | MSH2 | ENST00000233146 | 0.69668 | 0.01385 | 0.83065 | 0.83682 | 0.9252 | 0.6571259609 | 0.95011 | 0.9912507534 | 0.39469 | 6.202451163 | 0 |
| 2 | 47466728 | T | G | C | W | MSH2 | ENST00000233146 | 0.81693 | 0.02785 | 0.86946 | 0.87968 | 0.88069 | 0.7152584962 | 0.93035 | 0.9947614074 | 0.319265 | 6.202451163 | 0 |
| 2 | 47466729 | A | C | K | Q | MSH2 | ENST00000233146 | 0.92104 | 0.03442 | 0.66307 | 0.8019 | 0.89193 | 0.6315301936 | 0.93934 | 0.8621074834 | 0.278518 | 7.931764895 | 0 |
| 2 | 47466730 | A | C | K | T | MSH2 | ENST00000233146 | 0.65382 | 0.02126 | 0.7273 | 0.81456 | 0.89422 | 0.6689208106 | 0.95125 | 0.9855318666 | 0.315628 | 4.225989728 | 0 |
| 2 | 47466730 | A | G | K | R | MSH2 | ENST00000233146 | 0.62173 | 0.01523 | 0.62456 | 0.73545 | 0.83228 | 0.5480573987 | 0.94122 | 0.962854743 | 0.235467 | 3.346129535 | 0 |
| 2 | 47466730 | A | T | K | M | MSH2 | ENST00000233146 | 0.8437 | 0.03243 | 0.62687 | 0.91226 | 0.93977 | 0.7263416864 | 0.93387 | 0.9973561764 | 0.335561 | 7.931764895 | 0 |
| 2 | 47466731 | G | C | K | N | MSH2 | ENST00000233146 | 0.61537 | 0.03362 | 0.75194 | 0.81907 | 0.83505 | 0.6733379387 | 0.89512 | 0.9892271757 | 0.111497 | 7.931764895 | 0 |
| 2 | 47466731 | G | T | K | N | MSH2 | ENST00000233146 | 0.60927 | 0.03362 | 0.75194 | 0.81996 | 0.83615 | 0.6733379387 | 0.89534 | 0.9892271757 | 0.111495 | 7.931764895 | 0 |
| 2 | 47466732 | G | A | E | K | MSH2 | ENST00000233146 | 0.92221 | 0.01855 | 0.82686 | 0.89202 | 0.95207 | 0.6693687272 | 0.96483 | 0.9941799641 | 0.43068 | 6.916996686 | 0 |
| 2 | 47466732 | G | C | E | Q | MSH2 | ENST00000233146 | 0.85974 | 0.0279 | 0.72208 | 0.73514 | 0.90885 | 0.5843022243 | 0.94498 | 0.9858645201 | 0.302929 | 5.055862398 | 0 |
| 2 | 47466733 | A | C | E | A | MSH2 | ENST00000233146 | 0.9285 | 0.02172 | 0.7365 | 0.88107 | 0.95243 | 0.6417061893 | 0.92127 | 0.9949064851 | 0.336239 | 5.682834095 | 0 |
| 2 | 47466733 | A | G | E | G | MSH2 | ENST00000233146 | 0.94522 | 0.01827 | 0.69465 | 0.90764 | 0.9681 | 0.6402550941 | 0.88912 | 0.9955028892 | 0.341594 | 7.949483909 | 0 |
| 2 | 47466733 | A | T | E | V | MSH2 | ENST00000233146 | 0.94091 | 0.02403 | 0.71969 | 0.90861 | 0.96298 | 0.6480531352 | 0.9427 | 0.9968281388 | 0.397016 | 7.886076731 | 0 |
| 2 | 47466734 | A | C | E | D | MSH2 | ENST00000233146 | 0.45205 | 0.00638 | 0.73678 | 0.68925 | 0.86902 | 0.4816461994 | 0.93496 | 0.8921419382 | 0.206774 | 3.806831486 | 0 |
| 2 | 47466734 | A | T | E | D | MSH2 | ENST00000233146 | 0.44686 | 0.00638 | 0.73678 | 0.68925 | 0.86852 | 0.4816461994 | 0.93492 | 0.8921419382 | 0.206765 | 3.806831486 | 0 |
| 2 | 47466735 | G | A | E | K | MSH2 | ENST00000233146 | 0.92519 | 0.03518 | 0.8382 | 0.77213 | 0.90362 | 0.6316626021 | 0.95031 | 0.9913086891 | 0.478848 | 4.529310391 | 0 |
| 2 | 47466735 | G | C | E | Q | MSH2 | ENST00000233146 | 0.88486 | 0.03728 | 0.75668 | 0.71421 | 0.88117 | 0.5261643045 | 0.93762 | 0.9184215047 | 0.423065 | 6.024303016 | 0 |
| 2 | 47466736 | A | C | E | A | MSH2 | ENST00000233146 | 0.73549 | 0.03602 | 0.76709 | 0.87112 | 0.92804 | 0.5894387375 | 0.94564 | 0.9922425747 | 0.284997 | 5.486586677 | 0 |
| 2 | 47466736 | A | G | E | G | MSH2 | ENST00000233146 | 0.91319 | 0.03465 | 0.73036 | 0.91503 | 0.95023 | 0.6032777183 | 0.89106 | 0.995519042 | 0.476756 | 8.945142482 | 0 |
| 2 | 47466736 | A | T | E | V | MSH2 | ENST00000233146 | 0.90196 | 0.03475 | 0.74595 | 0.93568 | 0.96883 | 0.6137102546 | 0.93566 | 0.995519042 | 0.476523 | 5.366195282 | 0 |
| 2 | 47466737 | A | C | E | D | MSH2 | ENST00000233146 | 0.46786 | 0.02864 | 0.76376 | 0.48976 | 0.85894 | 0.4830577442 | 0.94406 | 0.9136181474 | 0.208252 | 8.945142482 | 0 |
| 2 | 47466737 | A | T | E | D | MSH2 | ENST00000233146 | 0.46258 | 0.02864 | 0.76376 | 0.48109 | 0.85894 | 0.4830577442 | 0.94504 | 0.9395419359 | 0.208246 | 8.945142482 | 0 |
| 2 | 47466738 | A | G | K | E | MSH2 | ENST00000233146 | 0.87703 | 0.03219 | 0.75905 | 0.71558 | 0.9211 | 0.6589170078 | 0.94092 | 0.9434123061 | 0.39714 | 5.968822535 | 1 |
| 2 | 47466739 | A | C | K | T | MSH2 | ENST00000233146 | 0.84415 | 0.03458 | 0.76748 | 0.87874 | 0.91862 | 0.6299168893 | 0.94861 | 0.9804850817 | 0.372726 | 7.150818464 | 0 |
| 2 | 47466739 | A | G | K | R | MSH2 | ENST00000233146 | 0.85566 | 0.0401 | 0.69423 | 0.78776 | 0.86149 | 0.5118998814 | 0.95093 | 0.9109231234 | 0.310652 | 6.21774888 | 0 |
| 2 | 47466739 | A | T | K | I | MSH2 | ENST00000233146 | 0.90009 | 0.02864 | 0.73268 | 0.78238 | 0.95462 | 0.6800503403 | 0.93265 | 0.9932211637 | 0.420457 | 8.929247288 | 0 |
| 2 | 47466740 | A | C | K | N | MSH2 | ENST00000233146 | 0.801 | 0.03484 | 0.78307 | 0.66452 | 0.87 | 0.6583071989 | 0.94393 | 0.9862298965 | 0.130133 | 4.999086619 | 0 |
| 2 | 47466740 | A | T | K | N | MSH2 | ENST00000233146 | 0.79437 | 0.03484 | 0.78307 | 0.67449 | 0.87 | 0.6583071989 | 0.94387 | 0.9862298965 | 0.130124 | 4.999086619 | 0 |
| 2 | 47466741 | G | A | V | I | MSH2 | ENST00000233146 | 0.42633 | 0.00861 | 0.44735 | 0.39432 | 0.72093 | 0.2480105882 | 0.86791 | 0.5598629117 | 0.0407318 | 5.109381638 | 0 |
| 2 | 47466741 | G | C | V | L | MSH2 | ENST00000233146 | 0.44181 | 0.00586 | 0.60525 | 0.37534 | 0.73877 | 0.3946782058 | 0.92074 | 0.3776926631 | -0.00998624 | 2.139329615 | 0 |
| 2 | 47466741 | G | T | V | F | MSH2 | ENST00000233146 | 0.66019 | 0.01574 | 0.59054 | 0.70675 | 0.86098 | 0.5869845739 | 0.8735 | 0.8108725548 | 0.235476 | 4.067466881 | 0 |
| 2 | 47466742 | T | A | V | D | MSH2 | ENST00000233146 | 0.57195 | 0.00923 | 0.63292 | 0.75016 | 0.85945 | 0.6626484762 | 0.94958 | 0.9102215171 | 0.0435409 | 4.112710179 | 0 |
| 2 | 47466742 | T | C | V | A | MSH2 | ENST00000233146 | 0.3193 | 0.0054 | 0.62509 | 0.4839 | 0.7562 | 0.3117336264 | 0.92725 | 0.05978666618 | 0.0688703 | 0.7170580608 | 0 |
| 2 | 47466742 | T | G | V | G | MSH2 | ENST00000233146 | 0.44114 | 0.00535 | 0.49453 | 0.69797 | 0.84747 | 0.447153008 | 0.91004 | 0.4361017048 | -0.0373408 | 3.429453153 | 0 |
| 2 | 47466744 | C | A | L | I | MSH2 | ENST00000233146 | 0.50909 | 0.02582 | 0.77764 | 0.60791 | 0.79697 | 0.2570201084 | 0.81352 | 0.879771769 | 0.177285 | 3.570734776 | 0 |
| 2 | 47466744 | C | G | L | V | MSH2 | ENST00000233146 | 0.54929 | 0.03053 | 0.72848 | 0.64134 | 0.84428 | 0.4369939644 | 0.84899 | 0.8491779566 | 0.223689 | 3.47831773 | 0 |
| 2 | 47466744 | C | T | L | F | MSH2 | ENST00000233146 | 0.72324 | 0.03563 | 0.76746 | 0.65538 | 0.91945 | 0.5276782645 | 0.89043 | 0.9851544499 | 0.312998 | 5.709953586 | 0 |
| 2 | 47466745 | T | A | L | H | MSH2 | ENST00000233146 | 0.86566 | 0.03334 | 0.84467 | 0.92555 | 0.97067 | 0.6814479901 | 0.94047 | 0.996842742 | 0.465705 | 8.826576697 | 0 |
| 2 | 47466745 | T | C | L | P | MSH2 | ENST00000233146 | 0.8861 | 0.02994 | 0.87094 | 0.91531 | 0.97923 | 0.8384176498 | 0.96811 | 0.9973409772 | 0.476209 | 8.826576697 | 1 |
| 2 | 47466745 | T | G | L | R | MSH2 | ENST00000233146 | 0.88246 | 0.03137 | 0.84478 | 0.91567 | 0.97438 | 0.7934060872 | 0.95487 | 0.9969889522 | 0.475642 | 7.725737228 | 1 |
| 2 | 47466747 | C | A | R | S | MSH2 | ENST00000233146 | 0.82553 | 0.03327 | 0.55486 | 0.76707 | 0.95023 | 0.7054274236 | 0.95847 | 0.9991497993 | 0.411568 | 6.870925685 | 0 |
| 2 | 47466747 | C | G | R | G | MSH2 | ENST00000233146 | 0.85865 | 0.03327 | 0.54046 | 0.75705 | 0.95535 | 0.6719187923 | 0.97033 | 0.9950436354 | 0.445374 | 8.700917721 | 0 |

| | | | | | | | | | | | | | | | | | |
|---|---|---|---|---|---|---|---|---|---|---|---|---|---|---|---|---|---|
| 2 | 47466748 | G | A | R | H | MSH2 | ENST00000233146 | 0.84786 | 0.03388 | 0.57052 | 0.90272 | 0.90052 | 0.6025289045 | 0.92901 | 0.9624638557 | 0.178581 | 8.325780964 | 0 |
| 2 | 47466748 | G | C | R | P | MSH2 | ENST00000233146 | 0.8699 | 0.03166 | 0.5938 | 0.91373 | 0.94655 | 0.8098001043 | 0.90357 | 0.9989590645 | 0.465872 | 3.553653188 | 1 |
| 2 | 47466748 | G | T | R | L | MSH2 | ENST00000233146 | 0.85301 | 0.03388 | 0.58004 | 0.92677 | 0.94876 | 0.6831008585 | 0.92937 | 0.9956364036 | 0.274577 | 8.737088124 | 0 |
| 2 | 47466750 | A | T | N | Y | MSH2 | ENST00000233146 | 0.85471 | 0.03212 | 0.70388 | 0.73709 | 0.96189 | 0.5835799198 | 0.93568 | 0.9911415577 | 0.386787 | 8.683414817 | 0 |
| 2 | 47466751 | A | C | N | T | MSH2 | ENST00000233146 | 0.59879 | 0.02011 | 0.64738 | 0.52431 | 0.83004 | 0.4664885319 | 0.89541 | 0.992244184 | 0.167271 | 8.277601806 | 0 |
| 2 | 47466751 | A | G | N | S | MSH2 | ENST00000233146 | 0.42687 | 0.00696 | 0.59664 | 0.7057 | 0.70097 | 0.3585266513 | 0.86248 | 0.9658139348 | 0.101955 | 7.071108379 | 0 |
| 2 | 47466751 | A | T | N | I | MSH2 | ENST00000233146 | 0.82256 | 0.02864 | 0.66812 | 0.66895 | 0.95316 | 0.5938188354 | 0.92696 | 0.9990339279 | 0.321948 | 4.734216879 | 0 |
| 2 | 47466752 | C | A | N | K | MSH2 | ENST00000233146 | 0.31774 | 0.01924 | 0.73496 | 0.80577 | 0.77845 | 0.5259730661 | 0.86624 | 0.9910065532 | 0.101255 | 5.312074628 | 0 |
| 2 | 47466752 | C | G | N | K | MSH2 | ENST00000233146 | 0.32331 | 0.01924 | 0.73496 | 0.7998 | 0.77845 | 0.5259730661 | 0.86603 | 0.988935411 | 0.101258 | 5.312074628 | 0 |
| 2 | 47466753 | A | C | N | H | MSH2 | ENST00000233146 | 0.80843 | 0.0364 | 0.60796 | 0.90315 | 0.92601 | 0.5554869268 | 0.94798 | 0.9903685451 | 0.201148 | 8.458681036 | 0 |
| 2 | 47466753 | A | G | N | D | MSH2 | ENST00000233146 | 0.8319 | 0.03447 | 0.65264 | 0.89831 | 0.91058 | 0.5134489713 | 0.9241 | 0.9940808415 | 0.175453 | 4.40459632 | 0 |
| 2 | 47466753 | A | T | N | Y | MSH2 | ENST00000233146 | 0.85647 | 0.03398 | 0.63741 | 0.94429 | 0.97178 | 0.6206005917 | 0.94293 | 0.9967243075 | 0.433448 | 7.834643621 | 0 |
| 2 | 47466754 | A | C | N | T | MSH2 | ENST00000233146 | 0.78463 | 0.03527 | 0.5869 | 0.81834 | 0.86098 | 0.5210645874 | 0.83453 | 0.9793300033 | 0.16933 | 5.322331256 | 0 |
| 2 | 47466754 | A | G | N | S | MSH2 | ENST00000233146 | 0.55669 | 0.0245 | 0.53418 | 0.66628 | 0.80417 | 0.4180490559 | 0.83897 | 0.7220090032 | 0.0983451 | 5.52703165 | 0 |
| 2 | 47466754 | A | T | N | I | MSH2 | ENST00000233146 | 0.83159 | 0.03353 | 0.60767 | 0.93277 | 0.9506 | 0.6333393357 | 0.89617 | 0.9937145114 | 0.331141 | 7.158796508 | 0 |
| 2 | 47466755 | T | A | N | K | MSH2 | ENST00000233146 | 0.45438 | 0.03353 | 0.67428 | 0.76582 | 0.80595 | 0.576405803 | 0.89915 | 0.9938649535 | 0.0738417 | 5.401850412 | 0 |
| 2 | 47466755 | T | G | N | K | MSH2 | ENST00000233146 | 0.46072 | 0.03353 | 0.67428 | 0.7663 | 0.80595 | 0.576405803 | 0.89875 | 0.9911965132 | 0.0738452 | 5.401850412 | 0 |
| 2 | 47466756 | A | C | K | Q | MSH2 | ENST00000233146 | 0.84906 | 0.03137 | 0.37182 | 0.77467 | 0.89193 | 0.479598419 | 0.91555 | 0.9858535528 | 0.256432 | 3.935691647 | 0 |
| 2 | 47466756 | A | G | K | E | MSH2 | ENST00000233146 | 0.8539 | 0.01897 | 0.42096 | 0.79307 | 0.91186 | 0.5709136665 | 0.93727 | 0.9888730645 | 0.298779 | 3.316584672 | 0 |
| 2 | 47466757 | A | C | K | T | MSH2 | ENST00000233146 | 0.64812 | 0.00935 | 0.43215 | 0.66045 | 0.82046 | 0.5557304951 | 0.93973 | 0.9000492096 | 0.203583 | 5.161666934 | 0 |
| 2 | 47466757 | A | G | K | R | MSH2 | ENST00000233146 | 0.48608 | 0.00669 | 0.33735 | 0.60869 | 0.71938 | 0.3847234004 | 0.93093 | 0.7611738443 | 0.187234 | 3.715478317 | 0 |
| 2 | 47466757 | A | T | K | I | MSH2 | ENST00000233146 | 0.88189 | 0.02231 | 0.39187 | 0.89814 | 0.89784 | 0.5914375481 | 0.91041 | 0.9928852916 | 0.28798 | 5.878488988 | 0 |
| 2 | 47466758 | A | C | K | N | MSH2 | ENST00000233146 | 0.76649 | 0.02414 | 0.45491 | 0.86148 | 0.88069 | 0.5317952805 | 0.89999 | 0.9824280143 | 0.139279 | 4.135046542 | 0 |
| 2 | 47466758 | A | T | K | N | MSH2 | ENST00000233146 | 0.75981 | 0.02414 | 0.45491 | 0.8627 | 0.88069 | 0.5317952805 | 0.89973 | 0.9853695035 | 0.139275 | 4.135046542 | 0 |
| 2 | 47466759 | A | C | N | H | MSH2 | ENST00000233146 | 0.47137 | 0.02702 | 0.52227 | 0.75733 | 0.76356 | 0.3640455173 | 0.87213 | 0.7774628401 | 0.152381 | 2.921733511 | 0 |
| 2 | 47466759 | A | G | N | D | MSH2 | ENST00000233146 | 0.51803 | 0.00945 | 0.56462 | 0.64125 | 0.74658 | 0.3269227607 | 0.85544 | 0.7734425068 | 0.137706 | 2.471894874 | 0 |
| 2 | 47466759 | A | T | N | Y | MSH2 | ENST00000233146 | 0.83081 | 0.02755 | 0.55048 | 0.88034 | 0.9182 | 0.401674993 | 0.93318 | 0.972497642 | 0.297516 | 2.984234817 | 0 |
| 2 | 47466760 | A | C | N | T | MSH2 | ENST00000233146 | 0.53596 | 0.00983 | 0.51263 | 0.74467 | 0.8278 | 0.3382807381 | 0.88255 | 0.9516640306 | 0.199229 | 2.164418309 | 0 |
| 2 | 47466760 | A | G | N | S | MSH2 | ENST00000233146 | 0.47162 | 0.00674 | 0.45565 | 0.70631 | 0.7147 | 0.2509087024 | 0.85365 | 0.9234945774 | 0.121949 | 3.843812553 | 0 |
| 2 | 47466760 | A | T | N | I | MSH2 | ENST00000233146 | 0.8891 | 0.02231 | 0.52647 | 0.91723 | 0.95023 | 0.4426216491 | 0.90618 | 0.9935804605 | 0.279599 | 5.977268033 | 0 |
| 2 | 47466761 | C | A | N | K | MSH2 | ENST00000233146 | 0.27152 | 0.00551 | 0.57115 | 0.39584 | 0.66828 | 0.2821284638 | 0.78939 | 0.5279870629 | 0.0479755 | -0.1631657469 | 0 |
| 2 | 47466761 | C | G | N | K | MSH2 | ENST00000233146 | 0.27655 | 0.00551 | 0.57115 | 0.367 | 0.66828 | 0.2821284638 | 0.78937 | 0.3234805763 | 0.0479804 | -0.1631657469 | 0 |
| 2 | 47466762 | T | A | F | I | MSH2 | ENST00000233146 | 0.93015 | 0.02574 | 0.65016 | 0.84006 | 0.95645 | 0.6726635023 | 0.93086 | 0.9963102937 | 0.319756 | 7.768017253 | 0 |
| 2 | 47466762 | T | C | F | L | MSH2 | ENST00000233146 | 0.93189 | 0.01953 | 0.68954 | 0.81409 | 0.94876 | 0.6249983991 | 0.93247 | 0.9904112816 | 0.321153 | 7.768017253 | 0 |
| 2 | 47466762 | T | G | F | V | MSH2 | ENST00000233146 | 0.93271 | 0.02646 | 0.61643 | 0.87873 | 0.97401 | 0.6604164945 | 0.94905 | 0.996604085 | 0.419655 | 7.768017253 | 0 |
| 2 | 47466763 | T | A | F | Y | MSH2 | ENST00000233146 | 0.473 | 0.0058 | 0.67217 | 0.49612 | 0.73661 | 0.519082247 | 0.80578 | 0.8268728256 | 0.101911 | 0.3822778452 | 0 |
| 2 | 47466763 | T | C | F | S | MSH2 | ENST00000233146 | 0.94907 | 0.02556 | 0.64552 | 0.90398 | 0.91186 | 0.7488788794 | 0.92361 | 0.9977450371 | 0.328696 | 7.768017253 | 1 |
| 2 | 47466763 | T | G | F | C | MSH2 | ENST00000233146 | 0.94931 | 0.03039 | 0.67516 | 0.90144 | 0.94431 | 0.7266047269 | 0.95378 | 0.9972018003 | 0.416636 | 7.768017253 | 0 |
| 2 | 47466764 | T | A | F | L | MSH2 | ENST00000233146 | 0.52446 | 0.01953 | 0.68954 | 0.89574 | 0.87974 | 0.6249983991 | 0.92481 | 0.992519021 | 0.24866 | 7.768017253 | 0 |
| 2 | 47466764 | T | G | F | L | MSH2 | ENST00000233146 | 0.53133 | 0.01953 | 0.68954 | 0.89574 | 0.87974 | 0.6249983991 | 0.92468 | 0.992519021 | 0.248663 | 7.768017253 | 0 |
| 2 | 47466765 | A | C | S | R | MSH2 | ENST00000233146 | 0.44193 | 0.00551 | 0.32898 | 0.60864 | 0.65772 | 0.4071373151 | 0.85468 | 0.8988748789 | 0.0918084 | 1.991030222 | 0 |
| 2 | 47466765 | A | G | S | G | MSH2 | ENST00000233146 | 0.43938 | 0.00586 | 0.22116 | 0.6553 | 0.55468 | 0.3599919996 | 0.84992 | 0.9110274911 | 0.0760973 | 5.047647165 | 0 |
| 2 | 47466765 | A | T | S | C | MSH2 | ENST00000233146 | 0.78827 | 0.01556 | 0.26466 | 0.78441 | 0.80476 | 0.341337894 | 0.85263 | 0.9239446521 | 0.13124 | 5.449578616 | 0 |
| 2 | 47466766 | G | A | S | N | MSH2 | ENST00000233146 | 0.26178 | 0.00586 | 0.30538 | 0.5352 | 0.56961 | 0.2607762407 | 0.69748 | 0.3538999856 | 0.00084547 | 1.678586743 | 0 |
| 2 | 47466766 | G | C | S | T | MSH2 | ENST00000233146 | 0.20436 | 0.00586 | 0.26699 | 0.49457 | 0.59638 | 0.2180455746 | 0.68796 | 0.09125682712 | -0.00348262 | 0.7236017462 | 0 |
| 2 | 47466766 | G | T | S | I | MSH2 | ENST00000233146 | 0.30699 | 0.00543 | 0.3021 | 0.59348 | 0.73443 | 0.3780452611 | 0.94715 | 0.4999575615 | 0.0833681 | 1.242658109 | 0 |
| 2 | 47466767 | T | A | S | R | MSH2 | ENST00000233146 | 0.39446 | 0.00551 | 0.32898 | 0.543 | 0.74164 | 0.4071373151 | 0.88253 | 0.4668495059 | 0.0420956 | 1.991030222 | 0 |
| 2 | 47466767 | T | G | S | R | MSH2 | ENST00000233146 | 0.40045 | 0.00551 | 0.32898 | 0.49612 | 0.74235 | 0.4071373151 | 0.88261 | 0.4403785467 | 0.0420942 | 1.991030222 | 0 |
| 2 | 47466768 | A | C | T | P | MSH2 | ENST00000233146 | 0.78399 | 0.02904 | 0.30335 | 0.8378 | 0.86403 | 0.6656129392 | 0.89582 | 0.9915178418 | 0.2693 | 6.318685548 | 0 |
| 2 | 47466768 | A | G | T | A | MSH2 | ENST00000233146 | 0.44594 | 0.01459 | 0.29586 | 0.7858 | 0.7872 | 0.38672816 | 0.90657 | 0.9654548168 | 0.197964 | 6.318685548 | 0 |
| 2 | 47466768 | A | T | T | S | MSH2 | ENST00000233146 | 0.67748 | 0.02243 | 0.30998 | 0.76441 | 0.80417 | 0.3410587138 | 0.8575 | 0.9628980756 | 0.154529 | 6.318685548 | 0 |
| 2 | 47466769 | C | A | T | N | MSH2 | ENST00000233146 | 0.47759 | 0.02891 | 0.38259 | 0.82955 | 0.77718 | 0.4832180115 | 0.83895 | 0.9838455915 | 0.109154 | 6.318685548 | 0 |
| 2 | 47466769 | C | G | T | S | MSH2 | ENST00000233146 | 0.45076 | 0.02243 | 0.30998 | 0.77419 | 0.80772 | 0.3410587138 | 0.86816 | 0.9413973093 | 0.134367 | 6.318685548 | 0 |
| 2 | 47466769 | C | T | T | I | MSH2 | ENST00000233146 | 0.23376 | 0.00608 | 0.44205 | 0.50755 | 0.77782 | 0.3732617546 | 0.91866 | 0.8711096048 | 0.158805 | 2.407235275 | 0 |
| 2 | 47466771 | G | A | V | I | MSH2 | ENST00000233146 | 0.26004 | 0.00608 | 0.26193 | 0.36235 | 0.71938 | 0.2100577824 | 0.84694 | 0.3401665986 | 0.0400941 | -1.702966353 | 0 |
| 2 | 47466771 | G | C | V | L | MSH2 | ENST00000233146 | 0.25471 | 0.0058 | 0.42034 | 0.43511 | 0.71627 | 0.338751556 | 0.82471 | 0.1951794922 | 0.0313955 | -1.731917365 | 0 |
| 2 | 47466771 | G | T | V | L | MSH2 | ENST00000233146 | 0.25102 | 0.0058 | 0.42034 | 0.44874 | 0.71627 | 0.338751556 | 0.82388 | 0.1951794922 | 0.0313899 | -1.731917365 | 0 |
| 2 | 47466772 | T | A | V | E | MSH2 | ENST00000233146 | 0.68691 | 0.0158 | 0.43678 | 0.88407 | 0.80831 | 0.6969555796 | 0.96597 | 0.9541463852 | 0.249726 | 3.42734552 | 0 |
| 2 | 47466772 | T | C | V | A | MSH2 | ENST00000233146 | 0.51028 | 0.00804 | 0.46416 | 0.68113 | 0.77654 | 0.5019323516 | 0.94059 | 0.587204814 | 0.189025 | 3.42734552 | 0 |
| 2 | 47466772 | T | G | V | G | MSH2 | ENST00000233146 | 0.67382 | 0.01211 | 0.33069 | 0.77402 | 0.84321 | 0.7438633447 | 0.95121 | 0.9535112977 | 0.254304 | 3.42734552 | 0 |
| 2 | 47466774 | G | C | D | H | MSH2 | ENST00000233146 | 0.9075 | 0.03379 | 0.76042 | 0.87537 | 0.94655 | 0.6022345349 | 0.9361 | 0.9977501035 | 0.449878 | 6.861614551 | 0 |
| 2 | 47466774 | G | T | D | Y | MSH2 | ENST00000233146 | 0.91259 | 0.03346 | 0.74794 | 0.88487 | 0.94949 | 0.682401111 | 0.97107 | 0.9965211153 | 0.463599 | 6.861614551 | 0 |
| 2 | 47466775 | A | C | D | A | MSH2 | ENST00000233146 | 0.92471 | 0.02955 | 0.68189 | 0.88139 | 0.9579 | 0.6242453163 | 0.93292 | 0.9937525392 | 0.389251 | 6.861614551 | 0 |
| 2 | 47466775 | A | G | D | G | MSH2 | ENST00000233146 | 0.91423 | 0.03083 | 0.69368 | 0.9006 | 0.96298 | 0.6550190064 | 0.96332 | 0.9962902069 | 0.436867 | 6.861614551 | 0 |
| 2 | 47466775 | A | T | D | V | MSH2 | ENST00000233146 | 0.93089 | 0.03076 | 0.67962 | 0.91349 | 0.97401 | 0.6992882826 | 0.98114 | 0.9972558618 | 0.455748 | 6.861614551 | 0 |
| 2 | 47466776 | T | A | D | E | MSH2 | ENST00000233146 | 0.30483 | 0.00773 | 0.71576 | 0.6712 | 0.8306 | 0.4419527487 | 0.90307 | 0.717777133 | 0.131933 | 2.659071711 | 0 |
| 2 | 47466776 | T | G | D | E | MSH2 | ENST00000233146 | 0.31013 | 0.00773 | 0.71576 | 0.67493 | 0.8306 | 0.4419527487 | 0.90256 | 0.7081938982 | 0.131942 | 2.659071711 | 0 |
| 2 | 47466777 | A | C | I | L | MSH2 | ENST00000233146 | 0.61581 | 0.00586 | 0.33887 | 0.75254 | 0.78967 | 0.3441540706 | 0.8992 | 0.7443493009 | 0.126935 | 4.532590581 | 0 |
| 2 | 47466777 | A | G | I | V | MSH2 | ENST00000233146 | 0.41516 | 0.00599 | 0.25834 | 0.5259 | 0.63202 | 0.1730586484 | 0.86073 | 0.8627997041 | 0.0746666 | 1.386643194 | 0 |
| 2 | 47466777 | A | T | I | F | MSH2 | ENST00000233146 | 0.67629 | 0.00497 | 0.34864 | 0.77728 | 0.91314 | 0.4174752857 | 0.92164 | 0.8014041185 | 0.227145 | 4.404640389 | 0 |
| 2 | 47466778 | T | A | I | N | MSH2 | ENST00000233146 | 0.56637 | 0.0058 | 0.42763 | 0.75794 | 0.8153 | 0.5101869162 | 0.90504 | 0.8329164386 | 0.21521 | 4.532590581 | 1 |
| 2 | 47466778 | T | C | I | T | MSH2 | ENST00000233146 | 0.29094 | 0.00527 | 0.45296 | 0.47347 | 0.70262 | 0.3223699052 | 0.79408 | 0.2249372571 | 0.104181 | -0.3665545432 | 0 |
| 2 | 47466778 | T | G | I | S | MSH2 | ENST00000233146 | 0.4141 | 0.00556 | 0.34883 | 0.62391 | 0.70589 | 0.5259990245 | 0.93897 | 0.6488870382 | 0.219436 | 1.984428499 | 0 |
| 2 | 47466779 | C | G | I | M | MSH2 | ENST00000233146 | 0.49628 | 0.01168 | 0.30567 | 0.7519 | 0.80772 | 0.3546107338 | 0.84213 | 0.8624621034 | 0.0950317 | 4.400135869 | 0 |
| 2 | 47466780 | C | A | Q | K | MSH2 | ENST00000233146 | 0.37047 | 0.00664 | 0.74149 | 0.76372 | 0.76024 | 0.5725970653 | 0.94538 | 0.9589082599 | 0.28221 | 3.214321817 | 1 |
| 2 | 47466781 | A | C | Q | P | MSH2 | ENST00000233146 | 0.69556 | 0.02844 | 0.70378 | 0.90577 | 0.9648 | 0.835634207 | 0.90361 | 0.992626965 | 0.316848 | 6.175566879 | 0 |
| 2 | 47466781 | A | G | Q | R | MSH2 | ENST00000233146 | 0.43598 | 0.00644 | 0.73528 | 0.8075 | 0.86047 | 0.635054339 | 0.98819 | 0.9173527956 | 0.364444 | 3.360790378 | 0 |
| 2 | 47466781 | A | T | Q | L | MSH2 | ENST00000233146 | 0.52035 | 0.00795 | 0.68411 | 0.84909 | 0.89739 | 0.6110543867 | 0.95708 | 0.9859722853 | 0.282904 | 2.460199143 | 0 |
| 2 | 47466782 | G | C | Q | H | MSH2 | ENST00000233146 | 0.6395 | 0.03353 | 0.73647 | 0.86363 | 0.94765 | 0.6592107853 | 0.91131 | 0.9893355966 | 0.308699 | 6.175566879 | 0 |
| 2 | 47466782 | G | T | Q | H | MSH2 | ENST00000233146 | 0.63321 | 0.03353 | 0.73647 | 0.86342 | 0.94765 | 0.6592107853 | 0.9119 | 0.9893355966 | 0.308697 | 6.175566879 | 0 |
| 2 | 47466783 | A | C | K | Q | MSH2 | ENST00000233146 | 0.84612 | 0.03587 | 0.77957 | 0.89252 | 0.98183 | 0.7223183685 | 0.9614 | 0.9903795719 | 0.432224 | 8.500296972 | 0 |
| 2 | 47466783 | A | G | K | E | MSH2 | ENST00000233146 | 0.86935 | 0.03166 | 0.81249 | 0.89302 | 0.98728 | 0.7680757566 | 0.96952 | 0.9948443174 | 0.509531 | 8.500296972 | 1 |
| 2 | 47466784 | A | C | K | T | MSH2 | ENST00000233146 | 0.8516 | 0.03409 | 0.82269 | 0.90169 | 0.98869 | 0.7508771116 | 0.95387 | 0.9977080822 | 0.512422 | 3.450366287 | 0 |
| 2 | 47466784 | A | G | K | R | MSH2 | ENST00000233146 | 0.84423 | 0.0394 | 0.75192 | 0.84284 | 0.9648 | 0.6457272901 | 0.95709 | 0.9907750487 | 0.418625 | 6.589461181 | 0 |
| 2 | 47466784 | A | T | K | M | MSH2 | ENST00000233146 | 0.88266 | 0.03269 | 0.74824 | 0.91067 | 0.99105 | 0.7816295116 | 0.96321 | 0.9992040992 | 0.528499 | 8.500296972 | 0 |
| 2 | 47466785 | G | C | K | N | MSH2 | ENST00000233146 | 0.71522 | 0.02499 | 0.83603 | 0.90422 | 0.9328 | 0.737212844 | 0.9162 | 0.9935192466 | 0.186811 | 5.698576724 | 0 |
| 2 | 47466785 | G | T | K | N | MSH2 | ENST00000233146 | 0.70864 | 0.02499 | 0.83603 | 0.90558 | 0.9328 | 0.737212844 | 0.91697 | 0.9935192466 | 0.186808 | 5.698576724 | 0 |
| 2 | 47466786 | A | C | N | H | MSH2 | ENST00000233146 | 0.51086 | 0.01621 | 0.68516 | 0.80521 | 0.90318 | 0.5737696276 | 0.95641 | 0.9758367538 | 0.305219 | 6.055372234 | 0 |
| 2 | 47466786 | A | T | N | Y | MSH2 | ENST00000233146 | 0.85062 | 0.03265 | 0.71018 | 0.91451 | 0.9844 | 0.6236145203 | 0.96779 | 0.9954640269 | 0.494317 | 7.471243467 | 0 |
| 2 | 47466787 | A | C | N | T | MSH2 | ENST00000233146 | 0.51982 | 0.03442 | 0.65968 | 0.84395 | 0.92601 | 0.5299055227 | 0.8936 | 0.9926710129 | 0.236351 | 5.128668677 | 0 |
| 2 | 47466787 | A | G | N | S | MSH2 | ENST00000233146 | 0.4515 | 0.03076 | 0.61569 | 0.77507 | 0.86403 | 0.4016307329 | 0.88797 | 0.8607504368 | 0.0340914 | 4.416264344 | 0 |
| 2 | 47466787 | A | T | N | I | MSH2 | ENST00000233146 | 0.85851 | 0.03281 | 0.68456 | 0.93481 | 0.98183 | 0.6495737741 | 0.94453 | 0.9966601133 | 0.450806 | 7.471243467 | 0 |
| 2 | 47466788 | T | A | N | K | MSH2 | ENST00000233146 | 0.37879 | 0.03083 | 0.74399 | 0.92201 | 0.91143 | 0.5734151612 | 0.92449 | 0.984847486 | 0.133671 | 7.471243467 | 0 |
| 2 | 47466788 | T | G | N | K | MSH2 | ENST00000233146 | 0.38467 | 0.03083 | 0.74399 | 0.92257 | 0.91143 | 0.5734151612 | 0.92449 | 0.9809199572 | 0.133674 | 7.471243467 | 0 |
| 2 | 47466789 | G | A | G | S | MSH2 | ENST00000233146 | 0.93089 | 0.03728 | 0.73988 | 0.91503 | 0.9833 | 0.7655167128 | 0.96456 | 0.9994128942 | 0.485148 | 4.067986087 | 0 |
| 2 | 47466789 | G | C | G | R | MSH2 | ENST00000233146 | 0.94151 | 0.03452 | 0.83062 | 0.94394 | 0.99138 | 0.8571875284 | 0.98119 | 0.9997314811 | 0.499196 | 9.250414989 | 0 |
| 2 | 47466789 | G | T | G | C | MSH2 | ENST00000233146 | 0.94784 | 0.03626 | 0.79078 | 0.94369 | 0.98585 | 0.7591712129 | 0.97735 | 0.9995581508 | 0.499616 | 4.821607406 | 0 |
| 2 | 47466790 | G | A | G | D | MSH2 | ENST00000233146 | 0.84415 | 0.03327 | 0.83961 | 0.90343 | 0.97661 | 0.8513185339 | 0.97289 | 0.9993878603 | 0.499322 | 9.250414989 | 1 |
| 2 | 47466790 | G | C | G | A | MSH2 | ENST00000233146 | 0.8152 | 0.03608 | 0.74447 | 0.88079 | 0.95572 | 0.7489711316 | 0.96336 | 0.9994629025 | 0.49287 | 4.232333264 | 0 |
| 2 | 47466790 | G | T | G | V | MSH2 | ENST00000233146 | 0.84567 | 0.03379 | 0.75305 | 0.9132 | 0.96627 | 0.8037720268 | 0.97273 | 0.9997548461 | 0.498921 | 9.250414989 | 0 |
| 2 | 47466792 | G | A | V | I | MSH2 | ENST00000233146 | 0.74483 | 0.03388 | 0.5089 | 0.73985 | 0.84641 | 0.3539412687 | 0.84486 | 0.927406013 | 0.264856 | 4.14425896 | 0 |

| | | | | | | | | | | | | | | | | |
|---|---|---|---|---|---|---|---|---|---|---|---|---|---|---|---|---|
| 2 | 47466792 | G | C | V | L | MSH2 | ENST00000233146 | 0.8948 | 0.03582 | 0.68103 | 0.86287 | 0.93824 | 0.6272452122 | 0.93965 | 0.9942682981 | 0.437105 | 6.9223606 | 0 |
| 2 | 47466792 | G | T | V | F | MSH2 | ENST00000233146 | 0.92202 | 0.03281 | 0.66367 | 0.91159 | 0.93939 | 0.8058314299 | 0.94508 | 0.9987490177 | 0.494831 | 7.337469906 | 0 |
| 2 | 47466793 | T | A | V | D | MSH2 | ENST00000233146 | 0.84748 | 0.02994 | 0.70654 | 0.92466 | 0.97104 | 0.8890046635 | 0.97574 | 0.9990587831 | 0.48953 | 7.041350818 | 1 |
| 2 | 47466793 | T | C | V | A | MSH2 | ENST00000233146 | 0.79406 | 0.03419 | 0.70406 | 0.8674 | 0.97587 | 0.6258591714 | 0.93192 | 0.9971997738 | 0.451333 | 7.337469906 | 0 |
| 2 | 47466793 | T | G | V | G | MSH2 | ENST00000233146 | 0.83259 | 0.03313 | 0.57262 | 0.90784 | 0.97438 | 0.8506315796 | 0.9666 | 0.9972912669 | 0.490467 | 7.337469906 | 1 |
| 2 | 47466795 | A | C | K | Q | MSH2 | ENST00000233146 | 0.83706 | 0.01391 | 0.46977 | 0.81486 | 0.88165 | 0.4815761599 | 0.90376 | 0.9417096376 | 0.214262 | 6.21758238 | 0 |
| 2 | 47466795 | A | G | K | E | MSH2 | ENST00000233146 | 0.9313 | 0.01827 | 0.52683 | 0.79801 | 0.89963 | 0.5563326474 | 0.91824 | 0.9530900717 | 0.252695 | 6.21758238 | 1 |
| 2 | 47466796 | A | C | K | T | MSH2 | ENST00000233146 | 0.89763 | 0.02914 | 0.53866 | 0.83529 | 0.88259 | 0.5153954106 | 0.86489 | 0.9786207676 | 0.225264 | 6.21758238 | 0 |
| 2 | 47466796 | A | G | K | R | MSH2 | ENST00000233146 | 0.44752 | 0.00638 | 0.44035 | 0.49079 | 0.6993 | 0.3669547999 | 0.90807 | 0.9232416749 | 0.0869666 | -0.104407316 | 0 |
| 2 | 47466796 | A | T | K | I | MSH2 | ENST00000233146 | 0.93139 | 0.02637 | 0.49789 | 0.86214 | 0.89101 | 0.5393025048 | 0.88484 | 0.9897202849 | 0.232658 | 6.21758238 | 0 |
| 2 | 47466797 | A | C | K | N | MSH2 | ENST00000233146 | 0.83421 | 0.02937 | 0.56065 | 0.84016 | 0.86852 | 0.5274869297 | 0.87214 | 0.959423244 | 0.101494 | 6.21758238 | 0 |
| 2 | 47466797 | A | T | K | N | MSH2 | ENST00000233146 | 0.82763 | 0.02937 | 0.56065 | 0.84016 | 0.86852 | 0.5274869297 | 0.87212 | 0.959423244 | 0.101493 | 6.21758238 | 0 |
| 2 | 47466798 | T | A | F | I | MSH2 | ENST00000233146 | 0.9069 | 0.03224 | 0.85944 | 0.90759 | 0.98869 | 0.7824461753 | 0.96748 | 0.9988177419 | 0.528927 | 10.25730634 | 0 |
| 2 | 47466798 | T | C | F | L | MSH2 | ENST00000233146 | 0.9141 | 0.03491 | 0.88399 | 0.88308 | 0.97773 | 0.7630079057 | 0.98184 | 0.9979727864 | 0.526315 | 10.25730634 | 0 |
| 2 | 47466798 | T | G | F | V | MSH2 | ENST00000233146 | 0.91087 | 0.03313 | 0.84051 | 0.91656 | 0.98834 | 0.7561723636 | 0.96413 | 0.9981344342 | 0.527199 | 10.25730634 | 0 |
| 2 | 47466799 | T | A | F | Y | MSH2 | ENST00000233146 | 0.89423 | 0.03527 | 0.87005 | 0.87981 | 0.95133 | 0.6706835305 | 0.92405 | 0.9961012602 | 0.357008 | 4.278227039 | 1 |
| 2 | 47466799 | T | C | F | S | MSH2 | ENST00000233146 | 0.93852 | 0.02864 | 0.86223 | 0.92972 | 0.99443 | 0.8499116574 | 0.98253 | 0.9992048144 | 0.531539 | 10.25730634 | 0 |
| 2 | 47466799 | T | G | F | C | MSH2 | ENST00000233146 | 0.93835 | 0.03175 | 0.88042 | 0.93199 | 0.99039 | 0.820190584 | 0.96456 | 0.9992048144 | 0.527539 | 10.25730634 | 0 |
| 2 | 47466800 | T | A | F | L | MSH2 | ENST00000233146 | 0.76966 | 0.03491 | 0.88399 | 0.93226 | 0.97624 | 0.7630079057 | 0.99049 | 0.9954202771 | 0.483319 | 10.25730634 | 0 |
| 2 | 47466800 | T | G | F | L | MSH2 | ENST00000233146 | 0.77782 | 0.03491 | 0.88399 | 0.93138 | 0.97624 | 0.7630079057 | 0.9904 | 0.9954202771 | 0.483323 | 10.25730634 | 0 |
| 2 | 47466801 | A | C | T | P | MSH2 | ENST00000233146 | 0.66494 | 0.01906 | 0.52314 | 0.88498 | 0.94839 | 0.7296803222 | 0.98769 | 0.9930577278 | 0.45065 | 7.367242511 | 1 |
| 2 | 47466801 | A | G | T | A | MSH2 | ENST00000233146 | 0.43896 | 0.00592 | 0.53507 | 0.69741 | 0.85843 | 0.5464548358 | 0.96615 | 0.9636057019 | 0.261226 | 5.058892825 | 0 |
| 2 | 47466801 | A | T | T | S | MSH2 | ENST00000233146 | 0.41351 | 0.00644 | 0.54731 | 0.72966 | 0.82556 | 0.4158416692 | 0.9403 | 0.8539551497 | 0.16991 | 3.351744744 | 0 |
| 2 | 47466802 | C | A | T | N | MSH2 | ENST00000233146 | 0.36884 | 0.0058 | 0.63791 | 0.70178 | 0.87394 | 0.6050312805 | 0.88284 | 0.9828625321 | 0.187255 | 4.087134579 | 0 |
| 2 | 47466802 | C | G | T | S | MSH2 | ENST00000233146 | 0.35547 | 0.00644 | 0.54731 | 0.68379 | 0.83943 | 0.4158416692 | 0.8918 | 0.9686425328 | 0.177924 | 3.351744744 | 0 |
| 2 | 47466802 | C | T | T | I | MSH2 | ENST00000233146 | 0.40057 | 0.00705 | 0.6915 | 0.76758 | 0.92438 | 0.6098691413 | 0.92966 | 0.9851947427 | 0.316137 | 3.265058827 | 0 |
| 2 | 47466804 | A | C | N | H | MSH2 | ENST00000233146 | 0.8402 | 0.03646 | 0.5496 | 0.83115 | 0.9211 | 0.6762792114 | 0.96534 | 0.9947810769 | 0.225576 | 6.150220695 | 0 |
| 2 | 47466804 | A | G | N | D | MSH2 | ENST00000233146 | 0.67621 | 0.01721 | 0.59543 | 0.73873 | 0.83339 | 0.6581999501 | 0.94138 | 0.9718521833 | 0.187106 | 4.642226554 | 0 |
| 2 | 47466804 | A | T | N | Y | MSH2 | ENST00000233146 | 0.88278 | 0.03398 | 0.5783 | 0.87883 | 0.96517 | 0.7310305033 | 0.95382 | 0.9879230857 | 0.424653 | 7.298422422 | 0 |
| 2 | 47466805 | A | C | N | T | MSH2 | ENST00000233146 | 0.59762 | 0.03475 | 0.51618 | 0.77088 | 0.85012 | 0.562517398 | 0.89227 | 0.9379463792 | 0.137929 | 3.214099584 | 0 |
| 2 | 47466805 | A | G | N | S | MSH2 | ENST00000233146 | 0.53334 | 0.02537 | 0.46678 | 0.79262 | 0.77972 | 0.4907726357 | 0.86828 | 0.9678611159 | 0.105522 | 3.739304474 | 0 |
| 2 | 47466805 | A | T | N | I | MSH2 | ENST00000233146 | 0.91569 | 0.03257 | 0.53037 | 0.91863 | 0.9367 | 0.7449643803 | 0.92275 | 0.9945244789 | 0.304101 | 4.481207267 | 1 |
| 2 | 47466806 | C | A | N | K | MSH2 | ENST00000233146 | 0.65 | 0.03362 | 0.62497 | 0.8838 | 0.90274 | 0.7110626686 | 0.92766 | 0.9964877367 | 0.166217 | 7.298422422 | 1 |
| 2 | 47466806 | C | G | N | K | MSH2 | ENST00000233146 | 0.6583 | 0.03362 | 0.62497 | 0.87576 | 0.90318 | 0.7110626686 | 0.92754 | 0.9964877367 | 0.166217 | 7.298422422 | 1 |
| 2 | 47466807 | A | C | S | R | MSH2 | ENST00000233146 | 0.60237 | 0.01881 | 0.5475 | 0.72832 | 0.91399 | 0.5111322702 | 0.94901 | 0.933372736 | 0.372013 | 7.588606986 | 0 |
| 2 | 47466807 | A | G | S | G | MSH2 | ENST00000233146 | 0.55012 | 0.00615 | 0.44179 | 0.58353 | 0.82668 | 0.3043036989 | 0.9641 | 0.8681555986 | 0.225654 | 6.285299935 | 0 |
| 2 | 47466808 | G | A | S | N | MSH2 | ENST00000233146 | 0.53873 | 0.00669 | 0.53914 | 0.63876 | 0.78782 | 0.3493613531 | 0.81652 | 0.6645611525 | 0.107814 | 3.496143131 | 0 |
| 2 | 47466808 | G | T | S | I | MSH2 | ENST00000233146 | 0.62673 | 0.0073 | 0.52246 | 0.65455 | 0.8873 | 0.4823133096 | 0.90813 | 0.9908229113 | 0.264381 | 7.184475638 | 0 |
| 2 | 47470965 | C | A | S | R | MSH2 | ENST00000233146 | 0.25684 | 0.01881 | 0.5475 | 0.84651 | 0.89558 | 0.5111322702 | 0.96043 | 0.836532414 | 0.327299 | 7.588606986 | 0 |
| 2 | 47470965 | C | G | S | R | MSH2 | ENST00000233146 | 0.26169 | 0.01881 | 0.5475 | 0.84681 | 0.89558 | 0.5111322702 | 0.96033 | 0.8203029037 | 0.327316 | 7.588606986 | 0 |
| 2 | 47470966 | A | C | K | Q | MSH2 | ENST00000233146 | 0.38249 | 0.00648 | 0.29564 | 0.7757 | 0.83394 | 0.4503590012 | 0.95299 | 0.5598365068 | 0.187699 | 3.272157272 | 0 |
| 2 | 47470966 | A | G | K | E | MSH2 | ENST00000233146 | 0.38478 | 0.00522 | 0.34049 | 0.73438 | 0.77782 | 0.5153837465 | 0.95115 | 0.656658411 | 0.184072 | 2.059194149 | 0 |
| 2 | 47470967 | A | C | K | T | MSH2 | ENST00000233146 | 0.44223 | 0.00592 | 0.35722 | 0.6174 | 0.84321 | 0.5349869105 | 0.92212 | 0.9689296484 | 0.1957 | 3.32418983 | 0 |
| 2 | 47470967 | A | G | K | R | MSH2 | ENST00000233146 | 0.42663 | 0.00627 | 0.25991 | 0.63984 | 0.77205 | 0.3911120036 | 0.96435 | 0.8564565182 | 0.183092 | 3.511103564 | 0 |
| 2 | 47470967 | A | T | K | I | MSH2 | ENST00000233146 | 0.63428 | 0.01058 | 0.32346 | 0.86966 | 0.92844 | 0.6094563408 | 0.94615 | 0.9958360195 | 0.281217 | 6.255302337 | 0 |
| 2 | 47470968 | A | C | K | N | MSH2 | ENST00000233146 | 0.41629 | 0.00569 | 0.37259 | 0.63802 | 0.7816 | 0.5459525511 | 0.91154 | 0.8141691685 | 0.110608 | 4.531976608 | 0 |
| 2 | 47470968 | A | T | K | N | MSH2 | ENST00000233146 | 0.41127 | 0.00569 | 0.37259 | 0.64014 | 0.7816 | 0.5459525511 | 0.91182 | 0.8141691685 | 0.110603 | 4.531976608 | 0 |
| 2 | 47470969 | T | A | L | M | MSH2 | ENST00000233146 | 0.46943 | 0.03747 | 0.62853 | 0.88629 | 0.90929 | 0.5788560694 | 0.89857 | 0.9483724833 | 0.353057 | 7.863927656 | 0 |
| 2 | 47470969 | T | G | L | V | MSH2 | ENST00000233146 | 0.55216 | 0.02653 | 0.58898 | 0.93981 | 0.94765 | 0.6554065095 | 0.92129 | 0.9875121713 | 0.421801 | 3.582883397 | 0 |
| 2 | 47470970 | T | C | L | S | MSH2 | ENST00000233146 | 0.91547 | 0.03212 | 0.6795 | 0.9376 | 0.99006 | 0.8505309271 | 0.97918 | 0.9992890358 | 0.524349 | 7.927668123 | 1 |
| 2 | 47470970 | T | G | L | W | MSH2 | ENST00000233146 | 0.91622 | 0.02984 | 0.73833 | 0.93669 | 0.98513 | 0.8637473331 | 0.95949 | 0.9994108677 | 0.524364 | 7.927668123 | 1 |
| 2 | 47470971 | G | C | L | F | MSH2 | ENST00000233146 | 0.59668 | 0.0357 | 0.64259 | 0.91847 | 0.95389 | 0.7065051723 | 0.93928 | 0.9973516464 | 0.472339 | 7.927668123 | 0 |
| 2 | 47470971 | G | T | L | F | MSH2 | ENST00000233146 | 0.59082 | 0.0357 | 0.64259 | 0.92461 | 0.95389 | 0.7065051723 | 0.93991 | 0.9973516464 | 0.472336 | 7.927668123 | 0 |
| 2 | 47470972 | A | C | T | P | MSH2 | ENST00000233146 | 0.50088 | 0.00638 | 0.16496 | 0.90381 | 0.85534 | 0.606271431 | 0.95839 | 0.6614687443 | 0.191406 | 3.005655749 | 1 |
| 2 | 47470972 | A | G | T | A | MSH2 | ENST00000233146 | 0.35448 | 0.00574 | 0.15664 | 0.74637 | 0.74235 | 0.1877576578 | 0.90795 | 0.5250893831 | 0.0640208 | 1.449916876 | 0 |
| 2 | 47470972 | A | T | T | S | MSH2 | ENST00000233146 | 0.24677 | 0.00586 | 0.15706 | 0.67689 | 0.68646 | 0.1666335932 | 0.79218 | 0.07490328699 | 0.0393325 | -2.683782245 | 0 |
| 2 | 47470973 | C | A | T | N | MSH2 | ENST00000233146 | 0.38055 | 0.00608 | 0.19411 | 0.70054 | 0.60599 | 0.2988674729 | 0.84392 | 0.7580502033 | 0.077083 | 0.8818997891 | 0 |
| 2 | 47470973 | C | G | T | S | MSH2 | ENST00000233146 | 0.287 | 0.00586 | 0.15706 | 0.41344 | 0.5927 | 0.1666335932 | 0.81014 | 0.479736656 | 0.0567006 | -2.683782245 | 0 |
| 2 | 47470973 | C | T | T | I | MSH2 | ENST00000233146 | 0.53785 | 0.00687 | 0.23051 | 0.74904 | 0.81703 | 0.3019267633 | 0.94641 | 0.7764939666 | 0.200093 | 2.699880924 | 0 |
| 2 | 47470975 | T | A | S | T | MSH2 | ENST00000233146 | 0.38651 | 0.00608 | 0.29506 | 0.70901 | 0.76553 | 0.3193534703 | 0.9299 | 0.5864265561 | 0.153606 | 3.893731567 | 0 |
| 2 | 47470975 | T | C | S | P | MSH2 | ENST00000233146 | 0.64376 | 0.01414 | 0.30891 | 0.83546 | 0.92924 | 0.74353861 | 0.96753 | 0.9730479121 | 0.354085 | 6.485518288 | 0 |
| 2 | 47470975 | T | G | S | A | MSH2 | ENST00000233146 | 0.40728 | 0.00615 | 0.27963 | 0.59795 | 0.66449 | 0.2443925794 | 0.86447 | 0.3497256637 | 0.080818 | 2.703083876 | 0 |
| 2 | 47470976 | C | A | S | Y | MSH2 | ENST00000233146 | 0.71386 | 0.02475 | 0.3803 | 0.81623 | 0.91015 | 0.513764328 | 0.92076 | 0.9845558405 | 0.28185 | 6.485518288 | 0 |
| 2 | 47470976 | C | G | S | C | MSH2 | ENST00000233146 | 0.74499 | 0.02702 | 0.29886 | 0.75403 | 0.89376 | 0.3748057768 | 0.86218 | 0.9831712842 | 0.262477 | 6.485518288 | 0 |
| 2 | 47470976 | C | T | S | F | MSH2 | ENST00000233146 | 0.75845 | 0.02366 | 0.34223 | 0.81259 | 0.91314 | 0.4649650561 | 0.93394 | 0.9911875129 | 0.29497 | 6.485518288 | 0 |
| 2 | 47470978 | T | A | L | I | MSH2 | ENST00000233146 | 0.32722 | 0.00603 | 0.36542 | 0.77552 | 0.76946 | 0.3768884529 | 0.88393 | 0.07222554204 | 0.0820245 | 2.101652255 | 0 |
| 2 | 47470978 | T | G | L | V | MSH2 | ENST00000233146 | 0.3399 | 0.00615 | 0.29753 | 0.76184 | 0.87247 | 0.5536559439 | 0.95808 | 0.1067584413 | 0.181522 | 2.939123606 | 0 |
| 2 | 47470979 | T | C | L | S | MSH2 | ENST00000233146 | 0.76816 | 0.02427 | 0.38854 | 0.92836 | 0.93631 | 0.7962805014 | 0.88649 | 0.9924635887 | 0.326448 | 6.62306892 | 0 |
| 2 | 47470980 | A | C | L | F | MSH2 | ENST00000233146 | 0.34555 | 0.00627 | 0.34307 | 0.83111 | 0.77141 | 0.6054339318 | 0.90805 | 0.2885275483 | 0.102709 | 2.235789769 | 0 |
| 2 | 47470980 | A | T | L | F | MSH2 | ENST00000233146 | 0.34092 | 0.00627 | 0.34307 | 0.83124 | 0.77141 | 0.6054339318 | 0.90717 | 0.2867601216 | 0.102724 | 2.235789769 | 0 |
| 2 | 47470981 | A | C | N | H | MSH2 | ENST00000233146 | 0.79587 | 0.0364 | 0.56975 | 0.81282 | 0.91271 | 0.6983698924 | 0.91949 | 0.9930127263 | 0.1933 | 7.594445966 | 0 |
| 2 | 47470981 | A | G | N | D | MSH2 | ENST00000233146 | 0.81528 | 0.03322 | 0.61499 | 0.79113 | 0.8887 | 0.7378639798 | 0.89646 | 0.9909895658 | 0.169703 | 7.594445966 | 0 |
| 2 | 47470981 | A | T | N | Y | MSH2 | ENST00000233146 | 0.84279 | 0.03388 | 0.60289 | 0.8836 | 0.9608 | 0.7695041846 | 0.92062 | 0.9968729615 | 0.414841 | 7.594445966 | 0 |
| 2 | 47470982 | A | C | N | T | MSH2 | ENST00000233146 | 0.69067 | 0.03475 | 0.53867 | 0.82322 | 0.87733 | 0.6258554257 | 0.82038 | 0.988021493 | 0.149586 | 7.594445966 | 0 |
| 2 | 47470982 | A | G | N | S | MSH2 | ENST00000233146 | 0.36749 | 0.01662 | 0.49002 | 0.66474 | 0.73805 | 0.569626667 | 0.83396 | 0.9313694239 | 0.0877992 | 5.5306225 | 0 |
| 2 | 47470982 | A | T | N | I | MSH2 | ENST00000233146 | 0.76323 | 0.03334 | 0.5615 | 0.89576 | 0.93939 | 0.7572437087 | 0.88234 | 0.9977476001 | 0.30114 | 7.594445966 | 0 |
| 2 | 47470983 | T | A | N | K | MSH2 | ENST00000233146 | 0.46507 | 0.03304 | 0.63859 | 0.8724 | 0.86098 | 0.75284261 | 0.91583 | 0.9986188412 | 0.0882858 | 7.263373946 | 0 |
| 2 | 47470983 | T | G | N | K | MSH2 | ENST00000233146 | 0.47149 | 0.03304 | 0.63859 | 0.8723 | 0.86098 | 0.75284261 | 0.91578 | 0.9968217611 | 0.0882886 | 7.263373946 | 0 |
| 2 | 47470984 | G | A | E | K | MSH2 | ENST00000233146 | 0.6218 | 0.01048 | 0.49387 | 0.89305 | 0.81703 | 0.5225123638 | 0.92765 | 0.6125287414 | 0.30431 | 6.067837781 | 0 |
| 2 | 47470984 | G | C | E | Q | MSH2 | ENST00000233146 | 0.73517 | 0.03155 | 0.37588 | 0.83779 | 0.87247 | 0.4514972736 | 0.89301 | 0.9195533395 | 0.273161 | 5.330300371 | 0 |
| 2 | 47470985 | A | C | E | A | MSH2 | ENST00000233146 | 0.78487 | 0.029 | 0.39417 | 0.88577 | 0.93083 | 0.5352223401 | 0.83305 | 0.9825655222 | 0.276197 | 4.246863459 | 0 |
| 2 | 47470985 | A | G | E | G | MSH2 | ENST00000233146 | 0.65085 | 0.02574 | 0.3571 | 0.91697 | 0.94469 | 0.5559555276 | 0.87214 | 0.9934444427 | 0.328751 | 5.418604087 | 0 |
| 2 | 47470985 | A | T | E | V | MSH2 | ENST00000233146 | 0.82887 | 0.032 | 0.37171 | 0.91918 | 0.95863 | 0.5676689354 | 0.89516 | 0.9935558438 | 0.346986 | 6.714719641 | 0 |
| 2 | 47470986 | A | C | E | D | MSH2 | ENST00000233146 | 0.34953 | 0.00664 | 0.38779 | 0.60494 | 0.77527 | 0.3884349558 | 0.90863 | 0.3726660311 | 0.131918 | 2.742713832 | 0 |
| 2 | 47470986 | A | T | E | D | MSH2 | ENST00000233146 | 0.34489 | 0.00664 | 0.38779 | 0.59139 | 0.77527 | 0.3884349558 | 0.90778 | 0.3726660311 | 0.131902 | 2.742713832 | 0 |
| 2 | 47470987 | G | C | E | Q | MSH2 | ENST00000233146 | 0.86879 | 0.01115 | 0.52257 | 0.71488 | 0.89513 | 0.5115500971 | 0.96813 | 0.6838962362 | 0.371918 | 3.668068896 | 0 |
| 2 | 47470988 | A | C | E | A | MSH2 | ENST00000233146 | 0.47244 | 0.00681 | 0.53484 | 0.83084 | 0.91736 | 0.6032060676 | 0.96998 | 0.9781445861 | 0.447439 | 5.963550573 | 0 |
| 2 | 47470988 | A | G | E | G | MSH2 | ENST00000233146 | 0.68619 | 0.0048 | 0.49487 | 0.8454 | 0.98071 | 0.6203684672 | 0.96105 | 0.9822571874 | 0.471428 | 7.333481272 | 0 |
| 2 | 47470988 | A | T | E | V | MSH2 | ENST00000233146 | 0.76235 | 0.02427 | 0.50761 | 0.87404 | 0.98034 | 0.6148298122 | 0.97787 | 0.7774521997 | 0.326361 | 4.894177593 | 0 |
| 2 | 47470989 | G | C | E | D | MSH2 | ENST00000233146 | 0.26884 | 0.00633 | 0.53336 | 0.67226 | 0.8517 | 0.4716242877 | 0.97872 | 0.2661579549 | 0.28629 | 2.926019628 | 0 |
| 2 | 47470989 | G | T | E | D | MSH2 | ENST00000233146 | 0.26507 | 0.00633 | 0.53336 | 0.675 | 0.8517 | 0.4716242877 | 0.97883 | 0.3107789457 | 0.286286 | 2.926019628 | 0 |
| 2 | 47470990 | T | A | Y | N | MSH2 | ENST00000233146 | 0.85515 | 0.02802 | 0.82666 | 0.90627 | 0.98657 | 0.8135905127 | 0.93613 | 0.9978206646 | 0.461312 | 7.130996564 | 0 |
| 2 | 47470990 | T | C | Y | H | MSH2 | ENST00000233146 | 0.70687 | 0.01184 | 0.83294 | 0.77413 | 0.95535 | 0.7274491255 | 0.89669 | 0.971965313 | 0.370792 | 4.162543804 | 0 |
| 2 | 47470991 | A | C | Y | S | MSH2 | ENST00000233146 | 0.84921 | 0.03053 | 0.77742 | 0.91239 | 0.9917 | 0.7906313339 | 0.97208 | 0.9967103212 | 0.47021 | 7.130996564 | 0 |
| 2 | 47470991 | A | G | Y | C | MSH2 | ENST00000233146 | 0.86474 | 0.03322 | 0.82541 | 0.88959 | 0.9833 | 0.7572637593 | 0.92559 | 0.997420311 | 0.476508 | 7.130996564 | 0 |
| 2 | 47470991 | A | T | Y | F | MSH2 | ENST00000233146 | 0.45635 | 0.01082 | 0.75371 | 0.74603 | 0.85586 | 0.5073850753 | 0.88874 | 0.8807734847 | 0.225995 | 3.139578679 | 0 |
| 2 | 47470993 | A | C | T | P | MSH2 | ENST00000233146 | 0.4301 | 0.00916 | 0.29325 | 0.77432 | 0.87 | 0.6956348961 | 0.94096 | 0.7682419419 | 0.221773 | 4.78350112 | 1 |
| 2 | 47470993 | A | G | T | A | MSH2 | ENST00000233146 | 0.32004 | 0.0058 | 0.27952 | 0.54467 | 0.77909 | 0.3228679622 | 0.9942 | 0.04866816344 | -0.0629601 | 4.329606195 | 0 |
| 2 | 47470993 | A | T | T | S | MSH2 | ENST00000233146 | 0.30193 | 0.00603 | 0.29681 | 0.56996 | 0.73296 | 0.2700178718 | 0.95322 | 0.3808999956 | 0.130796 | 2.835687718 | 0 |
| 2 | 47470994 | C | A | T | N | MSH2 | ENST00000233146 | 0.20028 | 0.00769 | 0.3535 | 0.60126 | 0.71153 | 0.4632619235 | 0.86114 | 0.7814159989 | 0.0828898 | 4.78350112 | 0 |
| 2 | 47470994 | C | G | T | S | MSH2 | ENST00000233146 | 0.17081 | 0.00603 | 0.29681 | 0.43804 | 0.72322 | 0.2700178718 | 0.95226 | 0.417744875 | 0.12391 | 2.835687718 | 0 |

| | | | | | | | | | | | | | | | | |
|---|---|---|---|---|---|---|---|---|---|---|---|---|---|---|---|---|
| 2 | 47470994 | C | T | T | I | MSH2 | ENST00000233146 | 0.23119 | 0.00562 | 0.39798 | 0.28682 | 0.70833 | 0.3374520661 | 0.95418 | 0.6515750885 | 0.139778 | 0.7923226462 | 0 |
| 2 | 47470996 | A | C | K | Q | MSH2 | ENST00000233146 | 0.45875 | 0.00923 | 0.45083 | 0.66022 | 0.75074 | 0.4135328727 | 0.92979 | 0.8926347494 | 0.216353 | 2.680841962 | 0 |
| 2 | 47470996 | A | G | K | E | MSH2 | ENST00000233146 | 0.45322 | 0.00556 | 0.4997 | 0.70435 | 0.7255 | 0.4375138072 | 0.95435 | 0.708027184 | 0.252569 | 3.472992871 | 0 |
| 2 | 47470997 | A | C | K | T | MSH2 | ENST00000233146 | 0.38616 | 0.00754 | 0.51771 | 0.74291 | 0.80831 | 0.5018210954 | 0.93242 | 0.7462809682 | 0.216672 | 3.786815138 | 0 |
| 2 | 47470997 | A | G | K | R | MSH2 | ENST00000233146 | 0.37617 | 0.00633 | 0.41517 | 0.61803 | 0.65277 | 0.3461857801 | 0.9385 | 0.755679965 | 0.133494 | 2.949829819 | 0 |
| 2 | 47470998 | A | C | K | N | MSH2 | ENST00000233146 | 0.53995 | 0.01746 | 0.53325 | 0.82317 | 0.8199 | 0.4914653876 | 0.85519 | 0.7463318706 | 0.0909584 | 2.88734185 | 0 |
| 2 | 47470998 | A | T | K | N | MSH2 | ENST00000233146 | 0.53428 | 0.01746 | 0.53325 | 0.82254 | 0.8199 | 0.4914653876 | 0.85481 | 0.7463318706 | 0.0909556 | 2.88734185 | 0 |
| 2 | 47470999 | A | C | N | H | MSH2 | ENST00000233146 | 0.50114 | 0.01774 | 0.47277 | 0.7347 | 0.73734 | 0.4750508241 | 0.87509 | 0.6689828038 | 0.109571 | 3.760169514 | 0 |
| 2 | 47470999 | A | G | N | D | MSH2 | ENST00000233146 | 0.43568 | 0.00715 | 0.52314 | 0.67604 | 0.71861 | 0.5937377928 | 0.9011 | 0.7343242764 | 0.122134 | 4.477768788 | 0 |
| 2 | 47470999 | A | T | N | Y | MSH2 | ENST00000233146 | 0.34773 | 0.00556 | 0.50192 | 0.59088 | 0.67843 | 0.5725693718 | 0.96515 | 0.6357099414 | 0.224828 | 1.636118348 | 0 |
| 2 | 47471000 | A | C | N | T | MSH2 | ENST00000233146 | 0.42992 | 0.00741 | 0.45766 | 0.70862 | 0.78223 | 0.3814898789 | 0.95684 | 0.408764869 | 0.138226 | 4.477768788 | 0 |
| 2 | 47471000 | A | G | N | S | MSH2 | ENST00000233146 | 0.38238 | 0.00615 | 0.40103 | 0.53761 | 0.61416 | 0.3340234641 | 0.87897 | 0.4921176434 | 0.0596099 | 0.5158662753 | 0 |
| 2 | 47471000 | A | T | N | I | MSH2 | ENST00000233146 | 0.45573 | 0.00535 | 0.46943 | 0.59162 | 0.70262 | 0.4865570053 | 0.9462 | 0.4765219092 | 0.0998421 | 3.407668393 | 0 |
| 2 | 47471001 | T | A | N | K | MSH2 | ENST00000233146 | 0.30396 | 0.0071 | 0.54215 | 0.70167 | 0.73734 | 0.5315171056 | 0.90616 | 0.5912937522 | 0.0927531 | 4.477768788 | 0 |
| 2 | 47471001 | T | G | N | K | MSH2 | ENST00000233146 | 0.3092 | 0.0071 | 0.54215 | 0.70069 | 0.73734 | 0.5315171056 | 0.90624 | 0.5912937522 | 0.0927566 | 4.477768788 | 0 |
| 2 | 47471002 | A | C | K | Q | MSH2 | ENST00000233146 | 0.55059 | 0.01063 | 0.33728 | 0.73123 | 0.76487 | 0.4317932231 | 0.92064 | 0.7619180679 | 0.159353 | 2.74225145 | 0 |
| 2 | 47471002 | A | G | K | E | MSH2 | ENST00000233146 | 0.72244 | 0.01459 | 0.3892 | 0.76111 | 0.83615 | 0.4600595382 | 0.9175 | 0.747064352 | 0.201047 | 2.391752696 | 0 |
| 2 | 47471003 | A | C | K | T | MSH2 | ENST00000233146 | 0.54521 | 0.02574 | 0.40958 | 0.77576 | 0.83172 | 0.5124549642 | 0.90595 | 0.9307413697 | 0.176232 | 4.706330523 | 0 |
| 2 | 47471003 | A | G | K | R | MSH2 | ENST00000233146 | 0.30483 | 0.00648 | 0.32295 | 0.35506 | 0.65673 | 0.3489042099 | 0.89177 | 0.5648334622 | 0.080841 | -0.07157965515 | 0 |
| 2 | 47471003 | A | T | K | I | MSH2 | ENST00000233146 | 0.75121 | 0.02349 | 0.36819 | 0.85815 | 0.82836 | 0.5486848345 | 0.9027 | 0.9931682348 | 0.197746 | 6.902390076 | 0 |
| 2 | 47471004 | A | C | K | N | MSH2 | ENST00000233146 | 0.73101 | 0.02381 | 0.42706 | 0.76231 | 0.80417 | 0.4871774593 | 0.88708 | 0.8713357449 | 0.112608 | 3.921028114 | 0 |
| 2 | 47471004 | A | T | K | N | MSH2 | ENST00000233146 | 0.7242 | 0.02381 | 0.42706 | 0.76231 | 0.80417 | 0.4871774593 | 0.88682 | 0.9041958451 | 0.112604 | 3.921028114 | 0 |
| 2 | 47471005 | A | C | T | P | MSH2 | ENST00000233146 | 0.33769 | 0.00664 | 0.13947 | 0.68149 | 0.83725 | 0.625634373 | 0.93706 | 0.188467942 | 0.151041 | 4.088173043 | 0 |
| 2 | 47471005 | A | G | T | A | MSH2 | ENST00000233146 | 0.26405 | 0.00574 | 0.12052 | 0.40048 | 0.67386 | 0.2880963206 | 0.83555 | 0.1884779334 | 0.0438827 | 2.088371733 | 0 |
| 2 | 47471005 | A | T | T | S | MSH2 | ENST00000233146 | 0.25399 | 0.00592 | 0.12541 | 0.36899 | 0.72626 | 0.2302753549 | 0.90036 | 0.200800851 | 0.0628617 | 0.8668654308 | 0 |
| 2 | 47471006 | C | A | T | K | MSH2 | ENST00000233146 | 0.20459 | 0.00543 | 0.18458 | 0.48656 | 0.70833 | 0.4280793679 | 0.87671 | 0.6771598458 | 0.0658907 | 1.248460099 | 0 |
| 2 | 47471006 | C | G | T | R | MSH2 | ENST00000233146 | 0.21741 | 0.00551 | 0.17528 | 0.49811 | 0.74658 | 0.4573581846 | 0.92904 | 0.1558984071 | 0.0782039 | 4.088173043 | 0 |
| 2 | 47471006 | C | T | T | I | MSH2 | ENST00000233146 | 0.22847 | 0.00674 | 0.21003 | 0.63544 | 0.78967 | 0.3226909631 | 0.91994 | 0.3380539119 | 0.104565 | 3.951037764 | 1 |
| 2 | 47471008 | G | C | E | Q | MSH2 | ENST00000233146 | 0.5185 | 0.00974 | 0.4799 | 0.64291 | 0.83004 | 0.5763421946 | 0.94622 | 0.9247573614 | 0.288606 | 4.403117164 | 0 |
| 2 | 47471009 | A | C | E | A | MSH2 | ENST00000233146 | 0.50942 | 0.00648 | 0.48919 | 0.70359 | 0.81065 | 0.6533396976 | 0.93242 | 0.8515248299 | 0.288449 | 5.080409374 | 0 |
| 2 | 47471009 | A | G | E | G | MSH2 | ENST00000233146 | 0.71762 | 0.0167 | 0.4509 | 0.81275 | 0.90362 | 0.6717245656 | 0.90758 | 0.9381818175 | 0.362094 | 5.705794519 | 0 |
| 2 | 47471009 | A | T | E | V | MSH2 | ENST00000233146 | 0.55511 | 0.01121 | 0.46107 | 0.78721 | 0.86902 | 0.6648347932 | 0.9556 | 0.9488615394 | 0.360771 | 7.086489932 | 0 |
| 2 | 47471010 | A | C | E | D | MSH2 | ENST00000233146 | 0.2441 | 0.00674 | 0.48785 | 0.83605 | 0.79028 | 0.5533912095 | 0.92014 | 0.1689806134 | 0.180526 | 2.726111202 | 0 |
| 2 | 47471010 | A | T | E | D | MSH2 | ENST00000233146 | 0.24046 | 0.00674 | 0.48785 | 0.83743 | 0.79028 | 0.5533912095 | 0.92034 | 0.1689806134 | 0.180522 | 2.726111202 | 0 |
| 2 | 47471011 | T | A | Y | N | MSH2 | ENST00000233146 | 0.86914 | 0.02979 | 0.79577 | 0.94341 | 0.95753 | 0.8449937043 | 0.9508 | 0.9997050166 | 0.498347 | 9.05256758 | 0 |
| 2 | 47471011 | T | C | Y | H | MSH2 | ENST00000233146 | 0.86375 | 0.03327 | 0.8033 | 0.90936 | 0.95499 | 0.7678899853 | 0.90256 | 0.99955942712 | 0.434656 | 9.05256758 | 0 |
| 2 | 47471011 | T | G | Y | D | MSH2 | ENST00000233146 | 0.8749 | 0.02857 | 0.79177 | 0.94303 | 0.96153 | 0.8829884979 | 0.97759 | 0.9999427795 | 0.498348 | 9.05256758 | 1 |
| 2 | 47471012 | A | C | Y | S | MSH2 | ENST00000233146 | 0.87024 | 0.03189 | 0.74453 | 0.9338 | 0.96117 | 0.8210187877 | 0.97603 | 0.9996628761 | 0.506508 | 4.32180463 | 0 |
| 2 | 47471012 | A | T | Y | F | MSH2 | ENST00000233146 | 0.79729 | 0.03175 | 0.72331 | 0.89636 | 0.91904 | 0.5698135537 | 0.93287 | 0.9972699285 | 0.30615 | 9.05256758 | 0 |
| 2 | 47471014 | G | A | E | K | MSH2 | ENST00000233146 | 0.59842 | 0.00893 | 0.48517 | 0.72792 | 0.86653 | 0.719894694 | 0.92751 | 0.9435659647 | 0.337208 | 3.57412097 | 0 |
| 2 | 47471014 | G | C | E | Q | MSH2 | ENST00000233146 | 0.53853 | 0.00619 | 0.3591 | 0.57461 | 0.79151 | 0.602837596 | 0.90953 | 0.9282397032 | 0.219846 | 3.216235622 | 0 |
| 2 | 47471015 | A | C | E | A | MSH2 | ENST00000233146 | 0.41831 | 0.00592 | 0.37044 | 0.71874 | 0.81065 | 0.6776201521 | 0.94307 | 0.9247546196 | 0.257255 | 3.032476565 | 0 |
| 2 | 47471015 | A | G | E | G | MSH2 | ENST00000233146 | 0.49641 | 0.00881 | 0.32466 | 0.7861 | 0.88963 | 0.6934657102 | 0.93276 | 0.9884859324 | 0.361432 | 4.632783217 | 0 |
| 2 | 47471015 | A | T | E | V | MSH2 | ENST00000233146 | 0.46974 | 0.00674 | 0.34451 | 0.81671 | 0.89147 | 0.676778191 | 0.95555 | 0.9810957313 | 0.333657 | 3.252145976 | 0 |
| 2 | 47471016 | A | C | E | D | MSH2 | ENST00000233146 | 0.28432 | 0.00615 | 0.36609 | 0.63557 | 0.74376 | 0.5813023972 | 0.9489 | 0.1717749834 | 0.172608 | 7.644972202 | 0 |
| 2 | 47471016 | A | T | E | D | MSH2 | ENST00000233146 | 0.2802 | 0.00615 | 0.36609 | 0.68238 | 0.74376 | 0.5813023972 | 0.94875 | 0.1717749834 | 0.172603 | 7.644972202 | 0 |
| 2 | 47471017 | G | A | E | K | MSH2 | ENST00000233146 | 0.76816 | 0.01263 | 0.50647 | 0.75367 | 0.92682 | 0.6295884088 | 0.95058 | 0.9333893657 | 0.400547 | 4.966597625 | 0 |
| 2 | 47471017 | G | C | E | Q | MSH2 | ENST00000233146 | 0.67819 | 0.00705 | 0.39003 | 0.70288 | 0.86503 | 0.5391611038 | 0.94117 | 0.9285126925 | 0.293095 | 4.601226003 | 0 |
| 2 | 47471018 | A | C | E | A | MSH2 | ENST00000233146 | 0.70559 | 0.00754 | 0.39685 | 0.70502 | 0.89009 | 0.6131219285 | 0.89952 | 0.4182120313 | 0.20228 | 4.621357016 | 0 |
| 2 | 47471018 | A | G | E | G | MSH2 | ENST00000233146 | 0.63727 | 0.00603 | 0.36035 | 0.72208 | 0.90362 | 0.6307872187 | 0.93734 | 0.9570909142 | 0.403291 | 5.738021424 | 0 |
| 2 | 47471018 | A | T | E | V | MSH2 | ENST00000233146 | 0.89 | 0.02001 | 0.37344 | 0.84961 | 0.96226 | 0.6300624492 | 0.94456 | 0.9830500391 | 0.408217 | 5.931244184 | 0 |
| 2 | 47471020 | G | A | A | T | MSH2 | ENST00000233146 | 0.43653 | 0.00627 | 0.57261 | 0.56092 | 0.70914 | 0.5389059266 | 0.86462 | 0.2470756001 | 0.00344128 | 1.978823724 | 0 |
| 2 | 47471020 | G | C | A | P | MSH2 | ENST00000233146 | 0.65335 | 0.02469 | 0.63726 | 0.75188 | 0.93436 | 0.8478220131 | 0.91978 | 0.964871943 | 0.418623 | 6.066583034 | 1 |
| 2 | 47471020 | G | T | A | S | MSH2 | ENST00000233146 | 0.41116 | 0.01191 | 0.52628 | 0.64651 | 0.77141 | 0.5046648553 | 0.84476 | 0.8904906511 | 0.223976 | 5.348377786 | 0 |
| 2 | 47471021 | C | A | A | D | MSH2 | ENST00000233146 | 0.44405 | 0.01408 | 0.6704 | 0.79998 | 0.90052 | 0.8262490679 | 0.91241 | 0.9730179906 | 0.427026 | 6.066583034 | 0 |
| 2 | 47471021 | C | G | A | G | MSH2 | ENST00000233146 | 0.46525 | 0.01818 | 0.54782 | 0.69661 | 0.89009 | 0.5274440605 | 0.851 | 0.9673558474 | 0.256309 | 6.066583034 | 0 |
| 2 | 47471021 | C | T | A | V | MSH2 | ENST00000233146 | 0.44826 | 0.00722 | 0.58006 | 0.62938 | 0.70014 | 0.5641816 | 0.93035 | 0.8253393769 | 0.246592 | 2.692842509 | 0 |
| 2 | 47471023 | C | A | Q | K | MSH2 | ENST00000233146 | 0.71818 | 0.03362 | 0.62566 | 0.89498 | 0.96553 | 0.8835341338 | 0.96219 | 0.9989227653 | 0.480148 | 8.151505238 | 1 |
| 2 | 47471024 | A | C | Q | P | MSH2 | ENST00000233146 | 0.82724 | 0.03419 | 0.56675 | 0.94957 | 0.96773 | 0.9490249598 | 0.97763 | 0.9987377524 | 0.509658 | 8.151505238 | 1 |
| 2 | 47471024 | A | G | Q | R | MSH2 | ENST00000233146 | 0.80629 | 0.03613 | 0.6177 | 0.9447 | 0.9528 | 0.9015960712 | 0.96056 | 0.9989652634 | 0.348294 | 8.151505238 | 1 |
| 2 | 47471024 | A | T | Q | L | MSH2 | ENST00000233146 | 0.85257 | 0.03753 | 0.55428 | 0.96042 | 0.95753 | 0.8585346661 | 0.94614 | 0.9995239973 | 0.45418 | 8.151505238 | 1 |
| 2 | 47471025 | G | C | Q | H | MSH2 | ENST00000233146 | 0.80929 | 0.03809 | 0.60977 | 0.93117 | 0.95717 | 0.8731586509 | 0.95666 | 0.999461472 | 0.372234 | 8.151505238 | 1 |
| 2 | 47471025 | G | T | Q | H | MSH2 | ENST00000233146 | 0.8029 | 0.03809 | 0.60977 | 0.93366 | 0.95717 | 0.8731586509 | 0.95692 | 0.999461472 | 0.372231 | 8.151505238 | 1 |
| 2 | 47471026 | G | A | D | N | MSH2 | ENST00000233146 | 0.28354 | 0.0058 | 0.20977 | 0.60436 | 0.62213 | 0.3726062947 | 0.85511 | 0.3845091164 | 0.0665939 | -0.2506418936 | 0 |
| 2 | 47471026 | G | C | D | H | MSH2 | ENST00000233146 | 0.56526 | 0.01965 | 0.23573 | 0.79128 | 0.86553 | 0.5188125964 | 0.94995 | 0.8558990359 | 0.225784 | 2.060213609 | 0 |
| 2 | 47471027 | A | C | D | A | MSH2 | ENST00000233146 | 0.57498 | 0.00574 | 0.18084 | 0.70516 | 0.73148 | 0.5365523053 | 0.91543 | 0.856554985 | 0.232091 | 1.886620808 | 0 |
| 2 | 47471027 | A | G | D | G | MSH2 | ENST00000233146 | 0.6061 | 0.00535 | 0.19455 | 0.71138 | 0.73949 | 0.5405201121 | 0.95101 | 0.4378726184 | 0.236348 | 2.63702325 | 0 |
| 2 | 47471027 | A | T | D | V | MSH2 | ENST00000233146 | 0.61432 | 0.0099 | 0.1656 | 0.77312 | 0.83228 | 0.5899863427 | 0.95298 | 0.7120870352 | 0.240968 | 4.661920543 | 0 |
| 2 | 47471029 | G | A | A | T | MSH2 | ENST00000233146 | 0.50393 | 0.01676 | 0.51402 | 0.63971 | 0.79395 | 0.462995027 | 0.93051 | 0.8962858319 | 0.335386 | 2.979786761 | 0 |
| 2 | 47471029 | G | C | A | P | MSH2 | ENST00000233146 | 0.73581 | 0.02702 | 0.59206 | 0.77739 | 0.9458 | 0.8144822698 | 0.93048 | 0.9802543521 | 0.462839 | 7.475031304 | 1 |
| 2 | 47471029 | G | T | A | S | MSH2 | ENST00000233146 | 0.45444 | 0.0073 | 0.45786 | 0.6686 | 0.76223 | 0.4373965055 | 0.8743 | 0.9054971337 | 0.257809 | 2.413898247 | 0 |
| 2 | 47471030 | C | A | A | D | MSH2 | ENST00000233146 | 0.41884 | 0.00596 | 0.64121 | 0.69856 | 0.79395 | 0.7490610289 | 0.92846 | 0.8795490861 | 0.397309 | 3.235791402 | 0 |
| 2 | 47471030 | C | G | A | G | MSH2 | ENST00000233146 | 0.417 | 0.00615 | 0.48736 | 0.66067 | 0.87 | 0.4580799107 | 0.90255 | 0.8388499022 | 0.266956 | 5.451363871 | 0 |
| 2 | 47471030 | C | T | A | V | MSH2 | ENST00000233146 | 0.44655 | 0.00881 | 0.53178 | 0.6287 | 0.80536 | 0.5612642889 | 0.90827 | 0.945453167 | 0.254432 | 3.362282402 | 0 |
| 2 | 47471032 | A | C | I | L | MSH2 | ENST00000233146 | 0.40828 | 0.00622 | 0.49023 | 0.61636 | 0.70589 | 0.6157231943 | 0.93468 | 0.6450563073 | 0.240478 | 2.735609093 | 0 |
| 2 | 47471032 | A | G | I | V | MSH2 | ENST00000233146 | 0.30249 | 0.00622 | 0.37199 | 0.56132 | 0.67386 | 0.4141324911 | 0.85604 | 0.1901368072 | -0.0484219 | 1.670232405 | 0 |
| 2 | 47471032 | A | T | I | F | MSH2 | ENST00000233146 | 0.45413 | 0.00532 | 0.49561 | 0.66777 | 0.87148 | 0.7917936781 | 0.98913 | 0.8941227794 | 0.423174 | 6.563944952 | 0 |
| 2 | 47471033 | T | A | I | N | MSH2 | ENST00000233146 | 0.92927 | 0.02484 | 0.62084 | 0.94327 | 0.98367 | 0.8776903678 | 0.91629 | 0.9974971414 | 0.385133 | 8.105069259 | 0 |
| 2 | 47471033 | T | C | I | T | MSH2 | ENST00000233146 | 0.88776 | 0.01505 | 0.64845 | 0.90367 | 0.98294 | 0.7393625162 | 0.98247 | 0.4472008944 | 0.25288 | 5.231055625 | 0 |
| 2 | 47471033 | T | G | I | S | MSH2 | ENST00000233146 | 0.92854 | 0.02143 | 0.51893 | 0.9338 | 0.9833 | 0.8674217481 | 0.98757 | 0.9969671369 | 0.474789 | 8.105069259 | 0 |
| 2 | 47471034 | T | G | I | M | MSH2 | ENST00000233146 | 0.6281 | 0.0278 | 0.44034 | 0.83693 | 0.91442 | 0.6611116701 | 0.97594 | 0.940656364 | 0.389828 | 5.192950371 | 0 |
| 2 | 47471035 | G | A | V | I | MSH2 | ENST00000233146 | 0.64213 | 0.03872 | 0.48989 | 0.7136 | 0.88823 | 0.5486387567 | 0.84017 | 0.9487538338 | 0.252277 | 7.465863306 | 0 |
| 2 | 47471035 | G | C | V | L | MSH2 | ENST00000233146 | 0.8242 | 0.03557 | 0.65455 | 0.89915 | 0.94543 | 0.7333417511 | 0.94882 | 0.9773434401 | 0.401395 | 5.429396214 | 1 |
| 2 | 47471035 | G | T | V | F | MSH2 | ENST00000233146 | 0.85669 | 0.03281 | 0.62654 | 0.91543 | 0.967 | 0.8800149047 | 0.94296 | 0.9972487092 | 0.490754 | 3.008799243 | 0 |
| 2 | 47471036 | T | C | V | A | MSH2 | ENST00000233146 | 0.63038 | 0.01556 | 0.6705 | 0.75533 | 0.91186 | 0.7409606035 | 0.93386 | 0.9253144264 | 0.406867 | 4.014820154 | 0 |
| 2 | 47471036 | T | G | V | G | MSH2 | ENST00000233146 | 0.85945 | 0.03212 | 0.55053 | 0.93707 | 0.98693 | 0.8399537673 | 0.97637 | 0.997797966 | 0.496599 | 7.465863306 | 0 |
| 2 | 47471038 | A | C | K | Q | MSH2 | ENST00000233146 | 0.49216 | 0.02824 | 0.58667 | 0.7084 | 0.85012 | 0.6305671163 | 0.87852 | 0.9016029835 | 0.232264 | 3.60665635 | 0 |
| 2 | 47471038 | A | G | K | E | MSH2 | ENST00000233146 | 0.45899 | 0.01586 | 0.63333 | 0.66676 | 0.8367 | 0.7034423427 | 0.93218 | 0.9331838489 | 0.291728 | 4.142636975 | 0 |
| 2 | 47471039 | A | C | K | T | MSH2 | ENST00000233146 | 0.40975 | 0.00916 | 0.63784 | 0.75225 | 0.85638 | 0.6880261103 | 0.93901 | 0.9278866053 | 0.284031 | 2.919278742 | 0 |
| 2 | 47471039 | A | G | K | R | MSH2 | ENST00000233146 | 0.41298 | 0.00763 | 0.55702 | 0.63296 | 0.79028 | 0.5696883612 | 0.9489 | 0.9206876755 | 0.203514 | 4.863121288 | 0 |
| 2 | 47471039 | A | T | K | I | MSH2 | ENST00000233146 | 0.63 | 0.01906 | 0.60468 | 0.88524 | 0.94281 | 0.7519123216 | 0.93551 | 0.9927020073 | 0.302793 | 3.83920251 | 0 |
| 2 | 47471040 | A | C | K | N | MSH2 | ENST00000233146 | 0.35041 | 0.00747 | 0.66024 | 0.75886 | 0.75211 | 0.7072517276 | 0.89232 | 0.7210600376 | 0.114412 | 3.328457323 | 0 |
| 2 | 47471040 | A | T | K | N | MSH2 | ENST00000233146 | 0.34576 | 0.00747 | 0.66024 | 0.75886 | 0.75211 | 0.7072517276 | 0.89249 | 0.7210600376 | 0.114409 | 3.328457323 | 0 |
| 2 | 47471041 | G | A | E | K | MSH2 | ENST00000233146 | 0.90047 | 0.02955 | 0.79281 | 0.84261 | 0.96773 | 0.7618478062 | 0.97597 | 0.9879844189 | 0.469082 | 5.129182454 | 0 |
| 2 | 47471041 | G | C | E | Q | MSH2 | ENST00000233146 | 0.85705 | 0.03692 | 0.69054 | 0.85073 | 0.95316 | 0.6641221008 | 0.95721 | 0.9865677357 | 0.423331 | 4.938529813 | 0 |
| 2 | 47471042 | A | C | E | A | MSH2 | ENST00000233146 | 0.8507 | 0.02658 | 0.69994 | 0.92925 | 0.98799 | 0.7397341138 | 0.96257 | 0.9959115982 | 0.508731 | 5.74821393 | 0 |
| 2 | 47471042 | A | G | E | G | MSH2 | ENST00000233146 | 0.89365 | 0.03362 | 0.66662 | 0.93333 | 0.99006 | 0.7534127997 | 0.95153 | 0.9979338646 | 0.523121 | 4.920615418 | 0 |
| 2 | 47471042 | A | T | E | V | MSH2 | ENST00000233146 | 0.87958 | 0.03431 | 0.68293 | 0.9377 | 0.98904 | 0.7384892536 | 0.97282 | 0.9971097112 | 0.534159 | 8.617323975 | 0 |
| 2 | 47471043 | A | C | E | D | MSH2 | ENST00000233146 | 0.77948 | 0.02828 | 0.69924 | 0.86332 | 0.95023 | 0.6424982854 | 0.9748 | 0.9650657177 | 0.433838 | 5.024799629 | 0 |
| 2 | 47471043 | A | T | E | D | MSH2 | ENST00000233146 | 0.77275 | 0.02828 | 0.69924 | 0.86822 | 0.95023 | 0.6424982854 | 0.97518 | 0.9897498488 | 0.433833 | 5.024799629 | 0 |
| 2 | 47471044 | A | C | I | L | MSH2 | ENST00000233146 | 0.8434 | 0.01385 | 0.59039 | 0.79485 | 0.90929 | 0.6703726081 | 0.90874 | 0.9667550325 | 0.281261 | 4.022674536 | 0 |

| | | | | | | | | | | | | | | | | | | |
|---|---|---|---|---|---|---|---|---|---|---|---|---|---|---|---|---|---|---|
| 2 | 47471044 | A | G | I | V | MSH2 | ENST00000233146 | 0.4249 | 0.01158 | 0.50025 | 0.60293 | 0.78597 | 0.4629921076 | 0.86648 | 0.4902228415 | 0.22626 | 2.559970204 | 0 |
| 2 | 47471044 | A | T | I | F | MSH2 | ENST00000233146 | 0.87569 | 0.02979 | 0.60666 | 0.90686 | 0.98621 | 0.8280556624 | 0.96226 | 0.995877611 | 0.486451 | 8.051623462 | 0 |
| 2 | 47471045 | T | A | I | N | MSH2 | ENST00000233146 | 0.82506 | 0.02835 | 0.69236 | 0.9447 | 0.98404 | 0.8999288396 | 0.9313 | 0.9982784986 | 0.463923 | 9.199292537 | 0 |
| 2 | 47471045 | T | C | I | T | MSH2 | ENST00000233146 | 0.74173 | 0.03231 | 0.71502 | 0.89886 | 0.98764 | 0.7765088076 | 0.96932 | 0.9975958467 | 0.465104 | 7.315901874 | 0 |
| 2 | 47471045 | T | G | I | S | MSH2 | ENST00000233146 | 0.8291 | 0.03093 | 0.62191 | 0.94361 | 0.99202 | 0.8887341591 | 0.98137 | 0.9988873601 | 0.491388 | 9.199292537 | 0 |
| 2 | 47471046 | T | G | I | M | MSH2 | ENST00000233146 | 0.63099 | 0.0364 | 0.55763 | 0.90898 | 0.91694 | 0.7130222455 | 0.9575 | 0.9888429642 | 0.364488 | 9.199292537 | 0 |
| 2 | 47471047 | G | A | V | I | MSH2 | ENST00000233146 | 0.27123 | 0.00608 | 0.39552 | 0.47488 | 0.63202 | 0.4745448625 | 0.85223 | 0.8460903764 | 0.091995 | -1.03443255 | 0 |
| 2 | 47471047 | G | C | V | L | MSH2 | ENST00000233146 | 0.38272 | 0.0058 | 0.56087 | 0.70115 | 0.72093 | 0.6305497097 | 0.97343 | 0.7443181872 | 0.197397 | 1.013504142 | 0 |
| 2 | 47471048 | T | C | V | A | MSH2 | ENST00000233146 | 0.78114 | 0.00763 | 0.58139 | 0.83784 | 0.89239 | 0.731152253 | 0.894 | 0.9885768294 | 0.295302 | 4.90701303 | 0 |
| 2 | 47471048 | T | G | V | G | MSH2 | ENST00000233146 | 0.93725 | 0.0178 | 0.46984 | 0.94472 | 0.9579 | 0.8462096316 | 0.87416 | 0.9966797829 | 0.337769 | 4.90701303 | 0 |
| 2 | 47471050 | A | C | N | H | MSH2 | ENST00000233146 | 0.67581 | 0.02702 | 0.56204 | 0.73648 | 0.88963 | 0.4873978647 | 0.90731 | 0.9859128594 | 0.178652 | 5.914126143 | 0 |
| 2 | 47471050 | A | G | N | D | MSH2 | ENST00000233146 | 0.56325 | 0.00945 | 0.61821 | 0.62773 | 0.77463 | 0.5177696471 | 0.86704 | 0.845345974 | 0.139163 | 3.004517559 | 0 |
| 2 | 47471050 | A | T | N | Y | MSH2 | ENST00000233146 | 0.89231 | 0.02755 | 0.60175 | 0.83829 | 0.95972 | 0.6077083971 | 0.94817 | 0.9940108657 | 0.387901 | 6.544131193 | 0 |
| 2 | 47471051 | A | C | N | T | MSH2 | ENST00000233146 | 0.54196 | 0.00627 | 0.53357 | 0.63323 | 0.73148 | 0.4313530457 | 0.90437 | 0.3559029812 | 0.115415 | 3.894430209 | 0 |
| 2 | 47471051 | A | G | N | S | MSH2 | ENST00000233146 | 0.44869 | 0.00622 | 0.4685 | 0.47428 | 0.62213 | 0.3563521361 | 0.79217 | 0.02149474275 | -0.0868911 | 2.258867402 | 0 |
| 2 | 47471051 | A | T | N | I | MSH2 | ENST00000233146 | 0.88157 | 0.01983 | 0.54202 | 0.8535 | 0.93319 | 0.6155145356 | 0.93127 | 0.952780664 | 0.232593 | 6.497686774 | 0 |
| 2 | 47471052 | T | A | N | K | MSH2 | ENST00000233146 | 0.3211 | 0.00603 | 0.65137 | 0.72816 | 0.67752 | 0.5293933366 | 0.88213 | 0.7332004905 | 0.0824101 | 3.130483868 | 0 |
| 2 | 47471052 | T | G | N | K | MSH2 | ENST00000233146 | 0.32653 | 0.00603 | 0.65137 | 0.72778 | 0.67752 | 0.5293933366 | 0.88165 | 0.6285248399 | 0.0824296 | 3.130483868 | 0 |
| 2 | 47471053 | A | C | I | L | MSH2 | ENST00000233146 | 0.63428 | 0.02017 | 0.76986 | 0.8172 | 0.90581 | 0.5933349488 | 0.88075 | 0.9792439342 | 0.282432 | 5.835479574 | 0 |
| 2 | 47471053 | A | G | I | V | MSH2 | ENST00000233146 | 0.40541 | 0.00935 | 0.68131 | 0.63138 | 0.74164 | 0.3789013237 | 0.80911 | 0.3746136237 | 0.157808 | 3.34257884 | 0 |
| 2 | 47471053 | A | T | I | F | MSH2 | ENST00000233146 | 0.82076 | 0.02891 | 0.78339 | 0.88123 | 0.9506 | 0.7246870002 | 0.92625 | 0.9945382476 | 0.432034 | 5.570770618 | 0 |
| 2 | 47471054 | T | A | I | N | MSH2 | ENST00000233146 | 0.94132 | 0.02815 | 0.85164 | 0.92214 | 0.97475 | 0.8106277022 | 0.89757 | 0.9976161718 | 0.439387 | 5.635777446 | 0 |
| 2 | 47471054 | T | C | I | T | MSH2 | ENST00000233146 | 0.88215 | 0.03053 | 0.8688 | 0.8223 | 0.95572 | 0.6320341996 | 0.84354 | 0.9877743125 | 0.383106 | 4.497811879 | 0 |
| 2 | 47471054 | T | G | I | S | MSH2 | ENST00000233146 | 0.94128 | 0.03006 | 0.79468 | 0.90991 | 0.97997 | 0.8017095527 | 0.87167 | 0.9980702996 | 0.430954 | 5.351852589 | 0 |
| 2 | 47471055 | T | G | I | M | MSH2 | ENST00000233146 | 0.72356 | 0.03595 | 0.73627 | 0.92202 | 0.94129 | 0.6306041603 | 0.88933 | 0.9843771458 | 0.296082 | 5.797658264 | 0 |
| 2 | 47471056 | T | A | S | T | MSH2 | ENST00000233146 | 0.659 | 0.01191 | 0.54715 | 0.78467 | 0.76157 | 0.7337303008 | 0.88988 | 0.7922057509 | 0.163868 | 3.43602248 | 0 |
| 2 | 47471056 | T | C | S | P | MSH2 | ENST00000233146 | 0.91909 | 0.02494 | 0.58278 | 0.91863 | 0.92924 | 0.9190619706 | 0.91784 | 0.9626523852 | 0.286515 | 6.460336351 | 0 |
| 2 | 47471056 | T | G | S | A | MSH2 | ENST00000233146 | 0.4446 | 0.00608 | 0.53724 | 0.57161 | 0.65673 | 0.6065317586 | 0.76588 | 0.8742099404 | 0.0819193 | 0.04456868981 | 0 |
| 2 | 47471057 | C | A | S | Y | MSH2 | ENST00000233146 | 0.85055 | 0.02883 | 0.63398 | 0.91853 | 0.94167 | 0.8423121258 | 0.9007 | 0.9934590459 | 0.314536 | 6.460336351 | 0 |
| 2 | 47471057 | C | G | S | C | MSH2 | ENST00000233146 | 0.87543 | 0.032 | 0.54941 | 0.89835 | 0.91058 | 0.7355914011 | 0.74485 | 0.972679317 | 0.265124 | 4.903140385 | 0 |
| 2 | 47471059 | T | A | S | T | MSH2 | ENST00000233146 | 0.47993 | 0.01945 | 0.49204 | 0.73207 | 0.86653 | 0.5348863682 | 0.91036 | 0.947563827 | 0.222978 | 5.710118738 | 0 |
| 2 | 47471059 | T | C | S | P | MSH2 | ENST00000233146 | 0.75647 | 0.03039 | 0.51305 | 0.85207 | 0.93824 | 0.8409422109 | 0.95849 | 0.9852821827 | 0.390619 | 7.660083269 | 0 |
| 2 | 47471059 | T | G | S | A | MSH2 | ENST00000233146 | 0.4154 | 0.00664 | 0.46601 | 0.57621 | 0.67933 | 0.4082221696 | 0.90735 | 0.5156990886 | 0.101752 | 0.9442579457 | 0 |
| 2 | 47471060 | C | T | S | L | MSH2 | ENST00000233146 | 0.76531 | 0.01591 | 0.51521 | 0.76714 | 0.87247 | 0.6653018791 | 0.93336 | 0.9867048264 | 0.28605 | 4.372261384 | 0 |
| 2 | 47471062 | G | A | G | S | MSH2 | ENST00000233146 | 0.72893 | 0.03728 | 0.71633 | 0.78754 | 0.87733 | 0.6726866377 | 0.95875 | 0.9904798865 | 0.350149 | 5.357476044 | 0 |
| 2 | 47471062 | G | T | G | C | MSH2 | ENST00000233146 | 0.91508 | 0.03626 | 0.76591 | 0.87715 | 0.96444 | 0.71784124 | 0.96966 | 0.9981831858 | 0.475845 | 8.710930011 | 0 |
| 2 | 47475025 | G | A | G | D | MSH2 | ENST00000233146 | 0.79128 | 0.03327 | 0.82223 | 0.8868 | 0.93824 | 0.7459309967 | 0.97291 | 0.9984242916 | 0.472447 | 5.144646162 | 0 |
| 2 | 47475025 | G | C | G | A | MSH2 | ENST00000233146 | 0.75925 | 0.03608 | 0.7208 | 0.86886 | 0.92028 | 0.6513535701 | 0.95474 | 0.9975814819 | 0.46667 | 4.997970047 | 0 |
| 2 | 47475025 | G | T | G | V | MSH2 | ENST00000233146 | 0.79286 | 0.03379 | 0.73095 | 0.87161 | 0.93786 | 0.7339074652 | 0.96911 | 0.9991111159 | 0.476899 | 7.000352318 | 0 |
| 2 | 47475027 | T | A | Y | N | MSH2 | ENST00000233146 | 0.89296 | 0.02965 | 0.71759 | 0.93623 | 0.97067 | 0.8528992007 | 0.97153 | 0.9993882179 | 0.486811 | 5.156123049 | 0 |
| 2 | 47475027 | T | G | Y | D | MSH2 | ENST00000233146 | 0.89785 | 0.02844 | 0.70674 | 0.94044 | 0.97512 | 0.8692749713 | 0.98554 | 0.9997941852 | 0.486813 | 6.991839908 | 0 |
| 2 | 47475028 | A | C | Y | S | MSH2 | ENST00000233146 | 0.89095 | 0.0315 | 0.65555 | 0.93524 | 0.98071 | 0.8422933008 | 0.98193 | 0.9991372228 | 0.49431 | 5.068613847 | 0 |
| 2 | 47475028 | A | G | Y | C | MSH2 | ENST00000233146 | 0.75177 | 0.02561 | 0.71951 | 0.92886 | 0.9506 | 0.8085141651 | 0.97611 | 0.9988713861 | 0.49647 | 6.906941878 | 0 |
| 2 | 47475028 | A | T | Y | F | MSH2 | ENST00000233146 | 0.645 | 0.03815 | 0.62936 | 0.86419 | 0.94205 | 0.6236382478 | 0.91148 | 0.9922202229 | 0.277396 | 5.472778594 | 0 |
| 2 | 47475030 | G | A | V | I | MSH2 | ENST00000233146 | 0.53428 | 0.01492 | 0.24316 | 0.60577 | 0.78658 | 0.4646787426 | 0.87731 | 0.9281440973 | 0.17242 | 2.868674631 | 0 |
| 2 | 47475030 | G | C | V | L | MSH2 | ENST00000233146 | 0.54082 | 0.00654 | 0.41808 | 0.6321 | 0.80831 | 0.6432144619 | 0.96456 | 0.8293877244 | 0.254998 | 3.536920457 | 0 |
| 2 | 47475030 | G | T | V | L | MSH2 | ENST00000233146 | 0.53535 | 0.00654 | 0.41808 | 0.62906 | 0.80831 | 0.6432144619 | 0.96496 | 0.8293877244 | 0.254996 | 3.536920457 | 0 |
| 2 | 47475031 | T | A | V | E | MSH2 | ENST00000233146 | 0.41316 | 0.00791 | 0.42481 | 0.80799 | 0.84641 | 0.8147605687 | 0.97496 | 0.937876761 | 0.267472 | 7.029535136 | 0 |
| 2 | 47475031 | T | C | V | A | MSH2 | ENST00000233146 | 0.34576 | 0.00562 | 0.44117 | 0.51105 | 0.76356 | 0.5998379617 | 0.97657 | 0.5313361883 | 0.196563 | 2.001339873 | 0 |
| 2 | 47475031 | T | G | V | G | MSH2 | ENST00000233146 | 0.40999 | 0.01479 | 0.30488 | 0.83727 | 0.86352 | 0.7218724073 | 0.97432 | 0.9618492126 | 0.277017 | 4.227547783 | 0 |
| 2 | 47475033 | G | A | E | K | MSH2 | ENST00000233146 | 0.63935 | 0.02857 | 0.62104 | 0.8335 | 0.88684 | 0.5352622899 | 0.94027 | 0.9590912461 | 0.314953 | 4.066233854 | 0 |
| 2 | 47475033 | G | C | E | Q | MSH2 | ENST00000233146 | 0.73045 | 0.03452 | 0.49513 | 0.78278 | 0.88022 | 0.4891160078 | 0.93898 | 0.9181472063 | 0.293206 | 4.885529143 | 0 |
| 2 | 47475034 | A | C | E | A | MSH2 | ENST00000233146 | 0.5465 | 0.02595 | 0.51557 | 0.7762 | 0.87588 | 0.5232172067 | 0.9203 | 0.9727239609 | 0.272634 | 4.80049217 | 0 |
| 2 | 47475034 | A | G | E | G | MSH2 | ENST00000233146 | 0.57498 | 0.00971 | 0.46685 | 0.69649 | 0.86902 | 0.5188359659 | 0.95306 | 0.9542102218 | 0.284437 | 4.949246456 | 0 |
| 2 | 47475034 | A | T | E | V | MSH2 | ENST00000233146 | 0.86304 | 0.03137 | 0.48915 | 0.91385 | 0.96117 | 0.5729083045 | 0.9636 | 0.9920312166 | 0.355327 | 5.405535156 | 0 |
| 2 | 47475035 | A | C | E | D | MSH2 | ENST00000233146 | 0.40068 | 0.00696 | 0.51294 | 0.41149 | 0.77463 | 0.4213640882 | 0.97565 | 0.09086900204 | 0.0219922 | 2.146834275 | 0 |
| 2 | 47475035 | A | T | E | D | MSH2 | ENST00000233146 | 0.39574 | 0.00696 | 0.51294 | 0.41162 | 0.77463 | 0.4213640882 | 0.97649 | 0.437625736 | 0.148336 | 2.146834275 | 0 |
| 2 | 47475036 | C | A | P | T | MSH2 | ENST00000233146 | 0.41092 | 0.02927 | 0.64245 | 0.80578 | 0.9182 | 0.6462102678 | 0.91117 | 0.980320096 | 0.393304 | 5.709036569 | 0 |
| 2 | 47475036 | C | G | P | A | MSH2 | ENST00000233146 | 0.55298 | 0.03099 | 0.59323 | 0.76815 | 0.91904 | 0.5937176871 | 0.85671 | 0.9816195965 | 0.291148 | 4.754956533 | 0 |
| 2 | 47475036 | C | T | P | S | MSH2 | ENST00000233146 | 0.74412 | 0.03613 | 0.6109 | 0.86835 | 0.93319 | 0.6855698398 | 0.80395 | 0.977832377 | 0.29522 | 5.45617243 | 0 |
| 2 | 47475037 | C | A | P | Q | MSH2 | ENST00000233146 | 0.67477 | 0.03802 | 0.616 | 0.85619 | 0.94205 | 0.6727835307 | 0.85914 | 0.9922096729 | 0.390386 | 5.394510578 | 0 |
| 2 | 47475037 | C | G | P | R | MSH2 | ENST00000233146 | 0.5131 | 0.03608 | 0.67459 | 0.85102 | 0.91058 | 0.8010667622 | 0.92544 | 0.9527254105 | 0.439499 | 4.012583934 | 0 |
| 2 | 47475037 | C | T | P | L | MSH2 | ENST00000233146 | 0.42484 | 0.03595 | 0.68352 | 0.80706 | 0.93824 | 0.7509180804 | 0.91138 | 0.9518386511 | 0.392752 | 5.270108319 | 0 |
| 2 | 47475039 | A | C | M | L | MSH2 | ENST00000233146 | 0.43732 | 0.00596 | 0.36146 | 0.4695 | 0.61875 | 0.5663868934 | 0.79707 | 0.5864928365 | 0.0701269 | 1.746508773 | 0 |
| 2 | 47475039 | A | G | M | V | MSH2 | ENST00000233146 | 0.48806 | 0.00615 | 0.28928 | 0.65483 | 0.70345 | 0.6012873631 | 0.92059 | 0.4754370749 | -0.0678316 | 2.279311774 | 0 |
| 2 | 47475039 | A | T | M | L | MSH2 | ENST00000233146 | 0.43226 | 0.00596 | 0.36146 | 0.46158 | 0.61875 | 0.5663868934 | 0.79578 | 0.5864928365 | 0.0701346 | 1.746508773 | 0 |
| 2 | 47475040 | T | A | M | K | MSH2 | ENST00000233146 | 0.64571 | 0.01391 | 0.46499 | 0.86925 | 0.97512 | 0.8831084574 | 0.9643 | 0.9898070693 | 0.397864 | 6.739289064 | 1 |
| 2 | 47475040 | T | C | M | T | MSH2 | ENST00000233146 | 0.59125 | 0.01556 | 0.51851 | 0.7335 | 0.9045 | 0.7623088054 | 0.93902 | 0.9870525002 | 0.269854 | 5.263383542 | 0 |
| 2 | 47475040 | T | G | M | R | MSH2 | ENST00000233146 | 0.64851 | 0.00599 | 0.43392 | 0.66625 | 0.92723 | 0.9095224169 | 0.96639 | 0.9799904823 | 0.386867 | 6.739289064 | 0 |
| 2 | 47475041 | G | A | M | I | MSH2 | ENST00000233146 | 0.4894 | 0.0058 | 0.38377 | 0.2599 | 0.613 | 0.58529944 | 0.88027 | 0.2457222628 | 0.0171191 | 0.8536638908 | 0 |
| 2 | 47475041 | G | C | M | I | MSH2 | ENST00000233146 | 0.48685 | 0.0058 | 0.38377 | 0.246 | 0.613 | 0.58529944 | 0.88031 | 0.6166172624 | 0.0845536 | 0.8536638908 | 0 |
| 2 | 47475041 | G | T | M | I | MSH2 | ENST00000233146 | 0.48164 | 0.0058 | 0.38377 | 0.2762 | 0.613 | 0.58529944 | 0.88031 | 0.6166172624 | 0.0845523 | 0.8536638908 | 0 |
| 2 | 47475042 | C | G | Q | E | MSH2 | ENST00000233146 | 0.37964 | 0.0073 | 0.52711 | 0.68343 | 0.79818 | 0.6474534892 | 0.96174 | 0.8844792247 | 0.276889 | 5.250127853 | 0 |
| 2 | 47475043 | A | C | Q | P | MSH2 | ENST00000233146 | 0.66874 | 0.00615 | 0.53868 | 0.79133 | 0.93709 | 0.8751523524 | 0.99073 | 0.9768189788 | 0.450739 | 7.178328873 | 0 |
| 2 | 47475043 | A | G | Q | R | MSH2 | ENST00000233146 | 0.50505 | 0.00696 | 0.57818 | 0.79219 | 0.87588 | 0.7352554842 | 0.99223 | 0.6847536546 | 0.420219 | 4.602235586 | 0 |
| 2 | 47475043 | A | T | Q | L | MSH2 | ENST00000233146 | 0.55854 | 0.0063 | 0.51714 | 0.7935 | 0.92356 | 0.7088748781 | 0.98501 | 0.9764965177 | 0.407369 | 2.798235159 | 0 |
| 2 | 47475044 | G | C | Q | H | MSH2 | ENST00000233146 | 0.4812 | 0.00726 | 0.576 | 0.8839 | 0.93747 | 0.6839894896 | 0.99332 | 0.974788785 | 0.393927 | 6.711667335 | 0 |
| 2 | 47475044 | G | T | Q | H | MSH2 | ENST00000233146 | 0.47608 | 0.00726 | 0.576 | 0.88369 | 0.93747 | 0.6839894896 | 0.99344 | 0.974788785 | 0.393925 | 6.711667335 | 0 |
| 2 | 47475045 | A | C | T | P | MSH2 | ENST00000233146 | 0.48411 | 0.01281 | 0.22172 | 0.71006 | 0.89376 | 0.7938241345 | 0.98107 | 0.965058744 | 0.326313 | 6.995354411 | 0 |
| 2 | 47475045 | A | G | T | A | MSH2 | ENST00000233146 | 0.39731 | 0.0058 | 0.21629 | 0.38928 | 0.76157 | 0.4556211338 | 0.96569 | 0.1886535904 | 0.0899143 | 2.610705736 | 0 |
| 2 | 47475045 | A | T | T | S | MSH2 | ENST00000233146 | 0.35041 | 0.00603 | 0.22843 | 0.3073 | 0.76553 | 0.3699057948 | 0.96384 | 0.7532280087 | 0.151713 | 2.077760443 | 0 |
| 2 | 47475046 | C | A | T | K | MSH2 | ENST00000233146 | 0.19054 | 0.00681 | 0.32564 | 0.59184 | 0.78348 | 0.665150921 | 0.97343 | 0.5420939326 | 0.213917 | 2.431364165 | 0 |
| 2 | 47475046 | C | G | T | R | MSH2 | ENST00000233146 | 0.19002 | 0.00556 | 0.31236 | 0.58739 | 0.71548 | 0.7006888106 | 0.95976 | 0.4986300468 | 0.164841 | 3.065894954 | 0 |
| 2 | 47475046 | C | T | T | I | MSH2 | ENST00000233146 | 0.1894 | 0.00562 | 0.36878 | 0.59636 | 0.79577 | 0.5326627317 | 0.95892 | 0.8513907194 | 0.199916 | 2.590559658 | 0 |
| 2 | 47475048 | C | A | L | I | MSH2 | ENST00000233146 | 0.28783 | 0.00978 | 0.37839 | 0.70659 | 0.8345 | 0.5123403066 | 0.88355 | 0.9005830884 | 0.174579 | 2.813587541 | 0 |
| 2 | 47475048 | C | G | L | V | MSH2 | ENST00000233146 | 0.39336 | 0.00821 | 0.31389 | 0.74908 | 0.86047 | 0.652769921 | 0.95624 | 0.9238613248 | 0.235354 | 3.545346534 | 0 |
| 2 | 47475048 | C | T | L | F | MSH2 | ENST00000233146 | 0.32966 | 0.00799 | 0.36386 | 0.7489 | 0.87394 | 0.7073778152 | 0.93684 | 0.9726760387 | 0.228282 | 7.161399761 | 0 |
| 2 | 47475049 | T | A | L | H | MSH2 | ENST00000233146 | 0.69243 | 0.03099 | 0.46284 | 0.95274 | 0.97215 | 0.8294429977 | 0.89892 | 0.9984893799 | 0.447835 | 7.577211096 | 0 |
| 2 | 47475049 | T | C | L | P | MSH2 | ENST00000233146 | 0.7141 | 0.0278 | 0.50997 | 0.94517 | 0.99138 | 0.9378232423 | 0.94432 | 0.9964684248 | 0.48745 | 7.577211096 | 1 |
| 2 | 47475049 | T | G | L | R | MSH2 | ENST00000233146 | 0.70599 | 0.02828 | 0.46414 | 0.95995 | 0.997 | 0.9280992808 | 0.96443 | 0.9975866079 | 0.507016 | 6.121141914 | 1 |
| 2 | 47475051 | A | C | N | H | MSH2 | ENST00000233146 | 0.78471 | 0.03518 | 0.47609 | 0.81876 | 0.91569 | 0.5740431066 | 0.87579 | 0.9769932628 | 0.207101 | 7.143668509 | 0 |
| 2 | 47475051 | A | G | N | D | MSH2 | ENST00000233146 | 0.64906 | 0.02608 | 0.51911 | 0.77108 | 0.89603 | 0.6700577472 | 0.8973 | 0.9904920459 | 0.199622 | 7.143668509 | 0 |
| 2 | 47475051 | A | T | N | Y | MSH2 | ENST00000233146 | 0.83352 | 0.03281 | 0.50987 | 0.88676 | 0.9608 | 0.6668127268 | 0.89316 | 0.9896875024 | 0.326215 | 7.143668509 | 0 |
| 2 | 47475052 | A | C | N | T | MSH2 | ENST00000233146 | 0.55257 | 0.02724 | 0.44534 | 0.80347 | 0.8873 | 0.4854844346 | 0.88739 | 0.9798348546 | 0.221559 | 6.541022954 | 0 |
| 2 | 47475052 | A | G | N | S | MSH2 | ENST00000233146 | 0.32764 | 0.00754 | 0.38927 | 0.62902 | 0.70508 | 0.3723113604 | 0.81853 | 0.03873228481 | -0.153687 | 1.606077761 | 0 |
| 2 | 47475052 | A | T | N | I | MSH2 | ENST00000233146 | 0.6194 | 0.01327 | 0.4736 | 0.89401 | 0.93083 | 0.6585938705 | 0.87926 | 0.9887256026 | 0.280619 | 7.143668509 | 0 |
| 2 | 47475053 | T | A | N | K | MSH2 | ENST00000233146 | 0.40899 | 0.02595 | 0.55517 | 0.80285 | 0.86149 | 0.6422604046 | 0.90569 | 0.9502564073 | 0.0873685 | 7.143668509 | 0 |
| 2 | 47475053 | T | G | N | K | MSH2 | ENST00000233146 | 0.41505 | 0.02595 | 0.55517 | 0.79966 | 0.86149 | 0.6422604046 | 0.90522 | 0.9830998182 | 0.154797 | 7.143668509 | 0 |
| 2 | 47475054 | G | A | D | N | MSH2 | ENST00000233146 | 0.52939 | 0.01676 | 0.53272 | 0.76649 | 0.75822 | 0.6482908507 | 0.83307 | 0.8576668501 | 0.0991951 | 4.170892431 | 0 |
| 2 | 47475054 | G | C | D | H | MSH2 | ENST00000233146 | 0.86297 | 0.03322 | 0.56428 | 0.79719 | 0.95133 | 0.7285576111 | 0.95487 | 0.9811001134 | 0.434855 | 4.652526097 | 0 |
| 2 | 47475054 | G | T | D | Y | MSH2 | ENST00000233146 | 0.68946 | 0.03313 | 0.5399 | 0.75618 | 0.95681 | 0.7826882976 | 0.97594 | 0.9912870526 | 0.464457 | 5.655681853 | 0 |

| | | | | | | | | | | | | | | | | |
|---|---|---|---|---|---|---|---|---|---|---|---|---|---|---|---|---|
| 2 | 47475055 | A | C | D | A | MSH2 | ENST00000233146 | 0.57744 | 0.01562 | 0.46889 | 0.77259 | 0.88637 | 0.738000437 | 0.97156 | 0.7164806724 | 0.279037 | 4.665186535 | 0 |
| 2 | 47475055 | A | G | D | G | MSH2 | ENST00000233146 | 0.56062 | 0.00935 | 0.48353 | 0.72842 | 0.86352 | 0.7250259605 | 0.98333 | 0.9300513268 | 0.419313 | 5.081663207 | 0 |
| 2 | 47475055 | A | T | D | V | MSH2 | ENST00000233146 | 0.73293 | 0.02857 | 0.46495 | 0.83394 | 0.9579 | 0.781711766 | 0.97957 | 0.9781630635 | 0.433953 | 6.068363541 | 0 |
| 2 | 47475056 | T | A | D | E | MSH2 | ENST00000233146 | 0.22504 | 0.00777 | 0.5054 | 0.63706 | 0.79395 | 0.5292491677 | 0.95652 | 0.7288821936 | 0.142458 | 2.65416017 | 0 |
| 2 | 47475056 | T | G | D | E | MSH2 | ENST00000233146 | 0.22931 | 0.00777 | 0.5054 | 0.62587 | 0.79395 | 0.5292491677 | 0.95654 | 0.7288821936 | 0.142458 | 2.65416017 | 0 |
| 2 | 47475057 | G | A | V | M | MSH2 | ENST00000233146 | 0.82436 | 0.03484 | 0.64524 | 0.82904 | 0.94506 | 0.6665646279 | 0.94926 | 0.7982715368 | 0.287222 | 5.845498855 | 0 |
| 2 | 47475057 | G | C | V | L | MSH2 | ENST00000233146 | 0.47829 | 0.00681 | 0.60681 | 0.73927 | 0.8306 | 0.6804091308 | 0.96353 | 0.8244167566 | 0.301118 | 3.464470634 | 0 |
| 2 | 47475057 | G | T | V | L | MSH2 | ENST00000233146 | 0.47313 | 0.00681 | 0.60681 | 0.73927 | 0.8306 | 0.6804091308 | 0.96364 | 0.8244167566 | 0.301118 | 3.464470634 | 0 |
| 2 | 47475058 | T | A | V | E | MSH2 | ENST00000233146 | 0.53893 | 0.0275 | 0.61376 | 0.90056 | 0.96517 | 0.8589784652 | 0.98169 | 0.9958800077 | 0.465093 | 6.018414827 | 0 |
| 2 | 47475058 | T | C | V | A | MSH2 | ENST00000233146 | 0.50354 | 0.01236 | 0.62799 | 0.83723 | 0.91652 | 0.6706277716 | 0.95549 | 0.9821637869 | 0.419007 | 4.039625227 | 0 |
| 2 | 47475058 | T | G | V | G | MSH2 | ENST00000233146 | 0.55477 | 0.02646 | 0.48727 | 0.91819 | 0.96298 | 0.7874513404 | 0.96859 | 0.9969545603 | 0.488181 | 5.705641132 | 0 |
| 2 | 47475060 | T | A | L | I | MSH2 | ENST00000233146 | 0.21921 | 0.00569 | 0.55463 | 0.66748 | 0.73949 | 0.3851598595 | 0.85599 | 0.1230182275 | 0.0538066 | -0.6792802019 | 0 |
| 2 | 47475060 | T | G | L | V | MSH2 | ENST00000233146 | 0.27899 | 0.00596 | 0.48379 | 0.73808 | 0.81414 | 0.5238515969 | 0.93926 | 0.07923428717 | 0.112129 | 1.132030509 | 0 |
| 2 | 47475061 | T | C | L | S | MSH2 | ENST00000233146 | 0.37992 | 0.0071 | 0.57113 | 0.82499 | 0.85064 | 0.8310437841 | 0.97099 | 0.8146242499 | 0.323726 | 6.354823186 | 0 |
| 2 | 47475062 | A | C | L | F | MSH2 | ENST00000233146 | 0.43016 | 0.02172 | 0.55008 | 0.84023 | 0.8887 | 0.7015768082 | 0.94114 | 0.9682156444 | 0.184963 | 6.354823186 | 0 |
| 2 | 47475062 | A | T | L | F | MSH2 | ENST00000233146 | 0.42514 | 0.02172 | 0.55008 | 0.84596 | 0.8887 | 0.7015768082 | 0.94074 | 0.9682156444 | 0.184937 | 6.354823186 | 0 |
| 2 | 47475063 | G | A | A | T | MSH2 | ENST00000233146 | 0.94298 | 0.03633 | 0.54394 | 0.93892 | 0.98693 | 0.7724930913 | 0.98751 | 0.9839071035 | 0.442669 | 9.276522718 | 1 |
| 2 | 47475063 | G | C | A | P | MSH2 | ENST00000233146 | 0.94665 | 0.03313 | 0.61367 | 0.93451 | 0.99326 | 0.9278472571 | 0.98805 | 0.9968132135 | 0.563006 | 9.520860822 | 0 |
| 2 | 47475063 | G | T | A | S | MSH2 | ENST00000233146 | 0.90833 | 0.02844 | 0.48916 | 0.83882 | 0.94281 | 0.6673965499 | 0.95718 | 0.885078438 | 0.399228 | 3.932132463 | 0 |
| 2 | 47475064 | C | A | A | D | MSH2 | ENST00000233146 | 0.6632 | 0.03076 | 0.66094 | 0.95046 | 0.99265 | 0.9291819733 | 0.9798 | 0.9974283576 | 0.553977 | 9.520860822 | 1 |
| 2 | 47475064 | C | G | A | G | MSH2 | ENST00000233146 | 0.48158 | 0.02218 | 0.51013 | 0.86796 | 0.93631 | 0.6870379284 | 0.96417 | 0.9766334891 | 0.480459 | 5.641724845 | 0 |
| 2 | 47475064 | C | T | A | V | MSH2 | ENST00000233146 | 0.65664 | 0.03692 | 0.5624 | 0.944 | 0.98799 | 0.8238522194 | 0.98844 | 0.9966087341 | 0.548641 | 9.520860822 | 1 |
| 2 | 47475066 | C | A | Q | K | MSH2 | ENST00000233146 | 0.45752 | 0.00547 | 0.40787 | 0.68186 | 0.82159 | 0.7354435002 | 0.96464 | 0.7229643464 | 0.258943 | 4.647349391 | 0 |
| 2 | 47475066 | C | G | Q | E | MSH2 | ENST00000233146 | 0.44174 | 0.00596 | 0.35092 | 0.67106 | 0.76356 | 0.6631039394 | 0.94875 | 0.6444220543 | 0.202991 | 3.348125277 | 0 |
| 2 | 47475067 | A | C | Q | P | MSH2 | ENST00000233146 | 0.63137 | 0.01568 | 0.36097 | 0.81703 | 0.95899 | 0.8792638187 | 0.97283 | 0.9804777503 | 0.438773 | 8.47857887 | 0 |
| 2 | 47475067 | A | G | Q | R | MSH2 | ENST00000233146 | 0.46097 | 0.00586 | 0.40104 | 0.72382 | 0.85741 | 0.7595981838 | 0.92475 | 0.4867554307 | 0.0539604 | 3.665838215 | 0 |
| 2 | 47475067 | A | T | Q | L | MSH2 | ENST00000233146 | 0.49274 | 0.00812 | 0.3429 | 0.77812 | 0.90052 | 0.7095438887 | 0.94328 | 0.9621250033 | 0.271185 | 5.091198877 | 0 |
| 2 | 47475068 | G | C | Q | H | MSH2 | ENST00000233146 | 0.31193 | 0.00586 | 0.39478 | 0.52383 | 0.74164 | 0.690396849 | 0.92878 | 0.07513552159 | 0.139685 | 3.830835806 | 0 |
| 2 | 47475068 | G | T | Q | H | MSH2 | ENST00000233146 | 0.30771 | 0.00586 | 0.39478 | 0.52141 | 0.74164 | 0.690396849 | 0.92917 | 0.07513552159 | 0.139684 | 3.830835806 | 0 |
| 2 | 47475069 | C | A | L | I | MSH2 | ENST00000233146 | 0.36649 | 0.02011 | 0.6946 | 0.83615 | 0.85894 | 0.5640296366 | 0.87561 | 0.9417203069 | 0.230235 | 2.903486562 | 0 |
| 2 | 47475069 | C | G | L | V | MSH2 | ENST00000233146 | 0.47237 | 0.02785 | 0.62827 | 0.88751 | 0.93593 | 0.7299061947 | 0.89743 | 0.8176030517 | 0.265827 | 4.036188324 | 0 |
| 2 | 47475070 | T | A | L | Q | MSH2 | ENST00000233146 | 0.72116 | 0.03595 | 0.69723 | 0.94853 | 0.99039 | 0.8835238606 | 0.98491 | 0.9970929623 | 0.546381 | 8.904920179 | 0 |
| 2 | 47475070 | T | C | L | P | MSH2 | ENST00000233146 | 0.74507 | 0.0296 | 0.79934 | 0.9494 | 0.98904 | 0.9584870927 | 0.98776 | 0.9977996945 | 0.55458 | 8.904920179 | 1 |
| 2 | 47475070 | T | G | L | R | MSH2 | ENST00000233146 | 0.72284 | 0.03099 | 0.76765 | 0.93288 | 0.98693 | 0.9553712584 | 0.98647 | 0.9959067106 | 0.553144 | 8.904920179 | 1 |
| 2 | 47475072 | G | A | D | N | MSH2 | ENST00000233146 | 0.94206 | 0.03741 | 0.67348 | 0.97112 | 0.9796 | 0.9109428303 | 0.95187 | 0.9953262806 | 0.266638 | 9.999870856 | 1 |
| 2 | 47475072 | G | C | D | H | MSH2 | ENST00000233146 | 0.94093 | 0.03458 | 0.70424 | 0.97313 | 0.99765 | 0.9318110428 | 0.98269 | 0.9969329834 | 0.574687 | 9.999870856 | 1 |
| 2 | 47475072 | G | T | D | Y | MSH2 | ENST00000233146 | 0.94282 | 0.03398 | 0.68833 | 0.96691 | 0.99356 | 0.9506697171 | 0.97988 | 0.9982085228 | 0.560591 | 9.999870856 | 1 |
| 2 | 47475073 | A | C | D | A | MSH2 | ENST00000233146 | 0.87329 | 0.0337 | 0.6072 | 0.97458 | 0.99676 | 0.9411508476 | 0.9642 | 0.9990025163 | 0.571942 | 9.999870856 | 0 |
| 2 | 47475073 | A | G | D | G | MSH2 | ENST00000233146 | 0.87951 | 0.03299 | 0.62574 | 0.97269 | 0.99982 | 0.9324737738 | 0.98747 | 0.9969078898 | 0.568902 | 9.999870856 | 1 |
| 2 | 47475073 | A | T | D | V | MSH2 | ENST00000233146 | 0.86762 | 0.0318 | 0.60391 | 0.98051 | 0.99953 | 0.941373897 | 0.9914 | 0.9990025163 | 0.583905 | 9.999870856 | 1 |
| 2 | 47475074 | T | A | D | E | MSH2 | ENST00000233146 | 0.52147 | 0.03281 | 0.64816 | 0.9699 | 0.95972 | 0.8872451247 | 0.99333 | 0.9891080856 | 0.5119 | 9.999870856 | 0 |
| 2 | 47475074 | T | G | D | E | MSH2 | ENST00000233146 | 0.52825 | 0.03281 | 0.64816 | 0.96751 | 0.95972 | 0.8872451247 | 0.9933 | 0.9902796149 | 0.511903 | 9.999870856 | 0 |
| 2 | 47475075 | G | A | A | T | MSH2 | ENST00000233146 | 0.85219 | 0.0325 | 0.61184 | 0.83548 | 0.95207 | 0.6211817845 | 0.95476 | 0.9503545165 | 0.437906 | 5.217014184 | 0 |
| 2 | 47475075 | G | C | A | P | MSH2 | ENST00000233146 | 0.88125 | 0.03261 | 0.68586 | 0.89595 | 0.9833 | 0.892384998 | 0.97501 | 0.9753596783 | 0.473057 | 8.362034198 | 1 |
| 2 | 47475075 | G | T | A | S | MSH2 | ENST00000233146 | 0.82981 | 0.03667 | 0.57299 | 0.87522 | 0.95426 | 0.6313199512 | 0.9067 | 0.9613044262 | 0.327746 | 5.277889519 | 0 |
| 2 | 47475076 | C | A | A | D | MSH2 | ENST00000233146 | 0.66067 | 0.03046 | 0.73269 | 0.91333 | 0.97773 | 0.9090876654 | 0.9747 | 0.993706286 | 0.470264 | 8.362034198 | 1 |
| 2 | 47475076 | C | G | A | G | MSH2 | ENST00000233146 | 0.64275 | 0.03534 | 0.60188 | 0.87375 | 0.96444 | 0.6608596326 | 0.92864 | 0.9788579345 | 0.396047 | 6.051947522 | 0 |
| 2 | 47475076 | C | T | A | V | MSH2 | ENST00000233146 | 0.26688 | 0.00722 | 0.64896 | 0.73672 | 0.76619 | 0.5890478054 | 0.85923 | 0.8045729399 | 0.217404 | 1.645168338 | 0 |
| 2 | 47475078 | G | A | V | I | MSH2 | ENST00000233146 | 0.3593 | 0.00608 | 0.29087 | 0.39803 | 0.64261 | 0.3430838408 | 0.80405 | 0.2842795551 | 0.0646884 | 1.64907059 | 0 |
| 2 | 47475078 | G | C | V | L | MSH2 | ENST00000233146 | 0.32357 | 0.00574 | 0.48124 | 0.45892 | 0.67661 | 0.5397796574 | 0.87355 | 0.2126120031 | -0.00778236 | 0.8314085975 | 0 |
| 2 | 47475078 | G | T | V | F | MSH2 | ENST00000233146 | 0.4316 | 0.00527 | 0.45995 | 0.62324 | 0.77335 | 0.8179620174 | 0.97695 | 0.1531597002 | 0.0723924 | 2.95404317 | 0 |
| 2 | 47475079 | T | A | V | D | MSH2 | ENST00000233146 | 0.5807 | 0.01177 | 0.51393 | 0.88052 | 0.93709 | 0.9203225022 | 0.98501 | 0.9179212929 | 0.404875 | 6.825281285 | 0 |
| 2 | 47475079 | T | C | V | A | MSH2 | ENST00000233146 | 0.311 | 0.0054 | 0.51128 | 0.53618 | 0.80536 | 0.661003471 | 0.95476 | 0.4103729725 | 0.0724033 | 4.90175551 | 0 |
| 2 | 47475079 | T | G | V | G | MSH2 | ENST00000233146 | 0.43726 | 0.00722 | 0.35859 | 0.83372 | 0.90096 | 0.8092580902 | 0.92996 | 0.7050873449 | 0.253217 | 6.825281285 | 0 |
| 2 | 47475081 | G | A | V | I | MSH2 | ENST00000233146 | 0.74882 | 0.02595 | 0.46939 | 0.68959 | 0.8199 | 0.4848106178 | 0.773 | 0.8462148905 | 0.179445 | 4.182462997 | 0 |
| 2 | 47475081 | G | C | V | L | MSH2 | ENST00000233146 | 0.4553 | 0.00687 | 0.68007 | 0.68277 | 0.85689 | 0.6820813613 | 0.94457 | 0.8253477812 | 0.321034 | 3.088249477 | 0 |
| 2 | 47475081 | G | T | V | F | MSH2 | ENST00000233146 | 0.82724 | 0.02844 | 0.65054 | 0.90574 | 0.96846 | 0.8522886344 | 0.92434 | 0.9944807291 | 0.451699 | 6.441588039 | 0 |
| 2 | 47475082 | T | A | V | D | MSH2 | ENST00000233146 | 0.89949 | 0.02835 | 0.70828 | 0.91748 | 0.98071 | 0.9157195339 | 0.97252 | 0.9919422865 | 0.336875 | 8.364531577 | 0 |
| 2 | 47475082 | T | C | V | A | MSH2 | ENST00000233146 | 0.5369 | 0.0099 | 0.70074 | 0.70398 | 0.85792 | 0.6630195643 | 0.83403 | 0.870757997 | 0.222269 | 2.990533448 | 0 |
| 2 | 47475082 | T | G | V | G | MSH2 | ENST00000233146 | 0.88529 | 0.02864 | 0.54413 | 0.90867 | 0.97475 | 0.7846718912 | 0.96106 | 0.996493876 | 0.467372 | 7.133797315 | 0 |
| 2 | 47475084 | A | C | S | R | MSH2 | ENST00000233146 | 0.84703 | 0.03118 | 0.7131 | 0.94375 | 0.99824 | 0.9538533461 | 0.98262 | 0.9990756512 | 0.555256 | 9.142370635 | 0 |
| 2 | 47475084 | A | G | S | G | MSH2 | ENST00000233146 | 0.82186 | 0.03821 | 0.59197 | 0.92922 | 0.99806 | 0.8161695245 | 0.95478 | 0.8283501649 | 0.368934 | 4.756366423 | 0 |
| 2 | 47475084 | A | T | S | C | MSH2 | ENST00000233146 | 0.85974 | 0.03761 | 0.65417 | 0.9413 | 0.99676 | 0.8248459923 | 0.97251 | 0.9989263415 | 0.55475 | 4.494997411 | 0 |
| 2 | 47475085 | G | A | S | N | MSH2 | ENST00000233146 | 0.82537 | 0.03809 | 0.69859 | 0.90708 | 0.97624 | 0.8938263296 | 0.93315 | 0.9966448545 | 0.283542 | 5.418382944 | 0 |
| 2 | 47475085 | G | C | S | T | MSH2 | ENST00000233146 | 0.81928 | 0.03832 | 0.64826 | 0.90852 | 0.98972 | 0.8343393304 | 0.97512 | 0.9925729036 | 0.532449 | 5.724630131 | 0 |
| 2 | 47475085 | G | T | S | I | MSH2 | ENST00000233146 | 0.89113 | 0.03458 | 0.67781 | 0.94014 | 0.99471 | 0.9057641502 | 0.98482 | 0.9989584684 | 0.562335 | 9.142370635 | 0 |
| 2 | 47475086 | C | A | S | R | MSH2 | ENST00000233146 | 0.50949 | 0.03118 | 0.7131 | 0.96781 | 0.99552 | 0.9538533461 | 0.9909 | 0.996476233 | 0.495265 | 9.142370635 | 0 |
| 2 | 47475086 | C | G | S | R | MSH2 | ENST00000233146 | 0.51645 | 0.03118 | 0.7131 | 0.9659 | 0.99552 | 0.9538533461 | 0.99075 | 0.9937081337 | 0.495266 | 9.142370635 | 0 |
| 2 | 47475087 | T | A | F | I | MSH2 | ENST00000233146 | 0.82802 | 0.02809 | 0.73463 | 0.89256 | 0.9659 | 0.8554416917 | 0.90045 | 0.9956790805 | 0.359191 | 9.154343295 | 0 |
| 2 | 47475087 | T | C | F | L | MSH2 | ENST00000233146 | 0.52759 | 0.0071 | 0.77519 | 0.75283 | 0.86453 | 0.8075231877 | 0.7157 | 0.9561344385 | 0.138917 | 2.943571277 | 0 |
| 2 | 47475087 | T | G | F | V | MSH2 | ENST00000233146 | 0.83437 | 0.02883 | 0.71927 | 0.91592 | 0.97997 | 0.8117143608 | 0.92737 | 0.9957348704 | 0.464626 | 8.082856485 | 0 |
| 2 | 47475088 | T | A | F | Y | MSH2 | ENST00000233146 | 0.85705 | 0.01523 | 0.75987 | 0.91403 | 0.96407 | 0.7746672071 | 0.90734 | 0.9936777949 | 0.333256 | 6.276556196 | 0 |
| 2 | 47475088 | T | C | F | S | MSH2 | ENST00000233146 | 0.92364 | 0.02765 | 0.74256 | 0.93306 | 0.98764 | 0.9065321194 | 0.93337 | 0.9989258647 | 0.491349 | 9.154343295 | 1 |
| 2 | 47475088 | T | G | F | C | MSH2 | ENST00000233146 | 0.92345 | 0.03124 | 0.76723 | 0.92563 | 0.98799 | 0.8636474243 | 0.93508 | 0.9981178045 | 0.506018 | 9.154343295 | 0 |
| 2 | 47475089 | T | A | F | L | MSH2 | ENST00000233146 | 0.45279 | 0.0071 | 0.77519 | 0.78557 | 0.82443 | 0.8075231877 | 0.72247 | 0.9735376835 | 0.0921788 | 2.943571277 | 0 |
| 2 | 47475089 | T | G | F | L | MSH2 | ENST00000233146 | 0.45912 | 0.0071 | 0.77519 | 0.7864 | 0.82443 | 0.8075231877 | 0.72387 | 0.9674093723 | 0.0921753 | 2.943571277 | 0 |
| 2 | 47475090 | G | A | A | T | MSH2 | ENST00000233146 | 0.88266 | 0.03674 | 0.6099 | 0.92089 | 0.99552 | 0.8386918173 | 0.97716 | 0.9968421459 | 0.578418 | 6.706607858 | 0 |
| 2 | 47475090 | G | C | A | P | MSH2 | ENST00000233146 | 0.89196 | 0.03313 | 0.67193 | 0.92322 | 0.99578 | 0.9581776988 | 0.97878 | 0.9969818022 | 0.581882 | 8.549236215 | 1 |
| 2 | 47475090 | G | T | A | S | MSH2 | ENST00000233146 | 0.71113 | 0.0317 | 0.55386 | 0.90781 | 0.96627 | 0.7445972858 | 0.89166 | 0.8619325161 | 0.256694 | 3.256519025 | 0 |
| 2 | 47475091 | C | A | A | D | MSH2 | ENST00000233146 | 0.65728 | 0.03088 | 0.72005 | 0.93921 | 0.99843 | 0.9594755224 | 0.97622 | 0.9974283576 | 0.575963 | 8.549236215 | 1 |
| 2 | 47475091 | C | G | A | G | MSH2 | ENST00000233146 | 0.63214 | 0.03582 | 0.58275 | 0.92804 | 0.98294 | 0.7672443579 | 0.93854 | 0.9973276284 | 0.531702 | 4.538213455 | 0 |
| 2 | 47475091 | C | T | A | V | MSH2 | ENST00000233146 | 0.65163 | 0.03674 | 0.62478 | 0.94027 | 0.99552 | 0.8753212541 | 0.97502 | 0.9966087341 | 0.569646 | 8.549236215 | 1 |
| 2 | 47475093 | C | A | H | N | MSH2 | ENST00000233146 | 0.30238 | 0.00566 | 0.548 | 0.68749 | 0.65077 | 0.6460759726 | 0.92433 | 0.5698308945 | 0.0965512 | 4.138209385 | 0 |
| 2 | 47475093 | C | G | H | D | MSH2 | ENST00000233146 | 0.3486 | 0.00754 | 0.54304 | 0.75261 | 0.85117 | 0.7978428528 | 0.98235 | 0.7120203972 | 0.236836 | 4.04630009 | 0 |
| 2 | 47475093 | C | T | H | Y | MSH2 | ENST00000233146 | 0.23359 | 0.00556 | 0.54902 | 0.57405 | 0.73949 | 0.6855263784 | 0.91104 | 0.3776645958 | 0.105486 | 3.467826847 | 0 |
| 2 | 47475094 | A | C | H | P | MSH2 | ENST00000233146 | 0.34141 | 0.01211 | 0.45238 | 0.82601 | 0.90755 | 0.8825966707 | 0.9196 | 0.9895692468 | 0.296916 | 7.590799281 | 1 |
| 2 | 47475094 | A | G | H | R | MSH2 | ENST00000233146 | 0.28159 | 0.00783 | 0.55446 | 0.73312 | 0.80118 | 0.71847143 | 0.9187 | 0.9667599797 | 0.236333 | 4.595236728 | 0 |
| 2 | 47475094 | A | T | H | L | MSH2 | ENST00000233146 | 0.27005 | 0.0054 | 0.49243 | 0.74655 | 0.80536 | 0.7195144069 | 0.87177 | 0.9651315808 | 0.160507 | 3.054951998 | 0 |
| 2 | 47475095 | C | A | H | Q | MSH2 | ENST00000233146 | 0.14094 | 0.00596 | 0.50981 | 0.81056 | 0.76619 | 0.5533819691 | 0.94152 | 0.1556936075 | 0.0607244 | 3.209074033 | 0 |
| 2 | 47475095 | C | G | H | Q | MSH2 | ENST00000233146 | 0.14444 | 0.00596 | 0.50981 | 0.80471 | 0.76619 | 0.5533819691 | 0.94186 | 0.5097686648 | 0.0607196 | 3.209074033 | 0 |
| 2 | 47475096 | G | C | V | L | MSH2 | ENST00000233146 | 0.58091 | 0.00681 | 0.49037 | 0.63706 | 0.8278 | 0.7412481564 | 0.88053 | 0.7545436234 | 0.236083 | 4.124528317 | 0 |
| 2 | 47475096 | G | T | V | L | MSH2 | ENST00000233146 | 0.57519 | 0.00681 | 0.49037 | 0.63706 | 0.8278 | 0.7412481564 | 0.88111 | 0.7507561482 | 0.236081 | 4.124528317 | 0 |
| 2 | 47475097 | T | A | V | E | MSH2 | ENST00000233146 | 0.42615 | 0.00726 | 0.50265 | 0.90455 | 0.94243 | 0.9017866485 | 0.97573 | 0.9913800359 | 0.422174 | 7.998508319 | 0 |
| 2 | 47475097 | T | C | V | A | MSH2 | ENST00000233146 | 0.21212 | 0.00535 | 0.51953 | 0.58727 | 0.67386 | 0.6800559176 | 0.76731 | 0.1089444235 | 0.0957091 | 1.498207359 | 0 |
| 2 | 47475097 | T | G | V | G | MSH2 | ENST00000233146 | 0.40999 | 0.00608 | 0.36172 | 0.90796 | 0.93319 | 0.7921520142 | 0.95874 | 0.9950352907 | 0.414515 | 7.998508319 | 0 |
| 2 | 47475099 | T | A | S | T | MSH2 | ENST00000233146 | 0.65877 | 0.01408 | 0.60169 | 0.89706 | 0.96627 | 0.8737232771 | 0.94009 | 0.9821464419 | 0.334476 | 7.278741233 | 0 |
| 2 | 47475099 | T | C | S | P | MSH2 | ENST00000233146 | 0.90595 | 0.02294 | 0.61669 | 0.92131 | 0.98146 | 0.9629306061 | 0.98558 | 0.9971358776 | 0.497974 | 8.687353316 | 0 |
| 2 | 47475099 | T | G | S | A | MSH2 | ENST00000233146 | 0.48602 | 0.00622 | 0.57883 | 0.65608 | 0.70914 | 0.7907369164 | 0.8838 | 0.7176583409 | 0.0955086 | 1.83485516 | 0 |
| 2 | 47475100 | C | T | S | L | MSH2 | ENST00000233146 | 0.5318 | 0.00735 | 0.62277 | 0.92399 | 0.96773 | 0.9273753926 | 0.96977 | 0.9949102998 | 0.508963 | 8.687353316 | 0 |
| 2 | 47475102 | A | C | N | H | MSH2 | ENST00000233146 | 0.36873 | 0.00562 | 0.3497 | 0.63282 | 0.56561 | 0.4580771841 | 0.91995 | 0.08577171854 | -0.109534 | 2.350443701 | 0 |
| 2 | 47475102 | A | G | N | D | MSH2 | ENST00000233146 | 0.34975 | 0.00547 | 0.40583 | 0.69242 | 0.58006 | 0.5129352015 | 0.91004 | 0.762547791 | 0.0770016 | 3.348768724 | 0 |

| | | | | | | | | | | | | | | | | | | |
|---|---|---|---|---|---|---|---|---|---|---|---|---|---|---|---|---|---|---|
| 2 | 47475102 | A | T | N | Y | MSH2 | ENST00000233146 | 0.39929 | 0.00566 | 0.39371 | 0.75373 | 0.80238 | 0.5447487286 | 0.96843 | 0.7259976268 | 0.202684 | 3.322588603 | 0 |
| 2 | 47475103 | A | C | N | T | MSH2 | ENST00000233146 | 0.31395 | 0.00566 | 0.31977 | 0.56824 | 0.68292 | 0.4316665158 | 0.88152 | 0.726121068 | 0.0801706 | 2.507041982 | 0 |
| 2 | 47475103 | A | G | N | S | MSH2 | ENST00000233146 | 0.27199 | 0.00608 | 0.26588 | 0.59252 | 0.58771 | 0.3367664063 | 0.85952 | 0.7169870734 | 0.0889869 | 1.956829222 | 0 |
| 2 | 47475103 | A | T | N | I | MSH2 | ENST00000233146 | 0.38164 | 0.00532 | 0.34764 | 0.58041 | 0.72398 | 0.5017703491 | 0.95237 | 0.8760910034 | 0.161653 | 3.411575848 | 0 |
| 2 | 47475104 | T | A | N | K | MSH2 | ENST00000233146 | 0.24271 | 0.00562 | 0.45202 | 0.63884 | 0.71938 | 0.4722423868 | 0.87434 | 0.5021634102 | 0.0647943 | 4.450760634 | 0 |
| 2 | 47475104 | T | G | N | K | MSH2 | ENST00000233146 | 0.24721 | 0.00562 | 0.45202 | 0.63884 | 0.71938 | 0.4722423868 | 0.87462 | 0.5021634102 | 0.0647866 | 4.450760634 | 0 |
| 2 | 47475105 | G | A | G | R | MSH2 | ENST00000233146 | 0.34337 | 0.00543 | 0.39235 | 0.38687 | 0.73148 | 0.6521948839 | 0.9692 | 0.3240126607 | 0.100284 | 2.557450714 | 0 |
| 2 | 47475105 | G | C | G | R | MSH2 | ENST00000233146 | 0.34114 | 0.00543 | 0.39235 | 0.39195 | 0.73148 | 0.6521948839 | 0.9687 | 0.4426496241 | 0.167722 | 2.557450714 | 0 |
| 2 | 47475106 | G | A | G | E | MSH2 | ENST00000233146 | 0.27175 | 0.00532 | 0.38323 | 0.53686 | 0.78348 | 0.6080167857 | 0.88988 | 0.354439497 | 0.133907 | 2.099796018 | 0 |
| 2 | 47475106 | G | C | G | A | MSH2 | ENST00000233146 | 0.25802 | 0.00556 | 0.28373 | 0.53414 | 0.74164 | 0.4855385547 | 0.82885 | 0.04567916842 | 0.071057 | 0.3861249553 | 0 |
| 2 | 47475106 | G | T | G | V | MSH2 | ENST00000233146 | 0.31467 | 0.00574 | 0.30059 | 0.54409 | 0.79089 | 0.6530594708 | 0.87667 | 0.2248167247 | 0.145605 | 3.52651232 | 0 |
| 2 | 47475108 | G | A | A | T | MSH2 | ENST00000233146 | 0.86664 | 0.03327 | 0.42203 | 0.92692 | 0.9506 | 0.8301578108 | 0.93757 | 0.9964125752 | 0.437799 | 8.866737473 | 0 |
| 2 | 47475108 | G | C | A | P | MSH2 | ENST00000233146 | 0.88246 | 0.03261 | 0.49366 | 0.84602 | 0.97327 | 0.932585994 | 0.93295 | 0.9758378863 | 0.510645 | 8.866737473 | 0 |
| 2 | 47475108 | G | T | A | S | MSH2 | ENST00000233146 | 0.82521 | 0.01786 | 0.37326 | 0.81867 | 0.91399 | 0.7619160968 | 0.90851 | 0.9659867883 | 0.313351 | 8.866737473 | 0 |
| 2 | 47475109 | C | A | A | E | MSH2 | ENST00000233146 | 0.63788 | 0.03334 | 0.50473 | 0.92257 | 0.95572 | 0.9307735331 | 0.9674 | 0.9970059991 | 0.508152 | 8.866737473 | 0 |
| 2 | 47475109 | C | G | A | G | MSH2 | ENST00000233146 | 0.6426 | 0.0132 | 0.39926 | 0.84916 | 0.93863 | 0.7714232196 | 0.9056 | 0.9871067405 | 0.369425 | 8.866737473 | 0 |
| 2 | 47475109 | C | T | A | V | MSH2 | ENST00000233146 | 0.65202 | 0.03293 | 0.44928 | 0.90692 | 0.94015 | 0.8523621166 | 0.91742 | 0.9948551594 | 0.433352 | 8.866737473 | 0 |
| 2 | 47475111 | C | A | P | T | MSH2 | ENST00000233146 | 0.67613 | 0.03518 | 0.5853 | 0.90628 | 0.92804 | 0.7503961582 | 0.90654 | 0.9972519517 | 0.453891 | 8.814623571 | 0 |
| 2 | 47475111 | C | G | P | A | MSH2 | ENST00000233146 | 0.65374 | 0.03706 | 0.52465 | 0.84435 | 0.91904 | 0.695552504 | 0.85294 | 0.9976812601 | 0.337174 | 8.814623571 | 0 |
| 2 | 47475111 | C | T | P | S | MSH2 | ENST00000233146 | 0.70567 | 0.0366 | 0.5385 | 0.86722 | 0.90711 | 0.7623515782 | 0.8655 | 0.9972798824 | 0.337859 | 5.234149191 | 0 |
| 2 | 47475112 | C | A | P | H | MSH2 | ENST00000233146 | 0.64781 | 0.03484 | 0.60218 | 0.9113 | 0.90274 | 0.7469881439 | 0.88566 | 0.9995526671 | 0.424079 | 8.814623571 | 0 |
| 2 | 47475112 | C | G | P | R | MSH2 | ENST00000233146 | 0.61328 | 0.03602 | 0.62315 | 0.88627 | 0.9506 | 0.7694116433 | 0.93779 | 0.9533476233 | 0.274422 | 8.814623571 | 0 |
| 2 | 47475112 | C | T | P | L | MSH2 | ENST00000233146 | 0.61761 | 0.0366 | 0.6236 | 0.87222 | 0.93004 | 0.7878654767 | 0.92439 | 0.9988731742 | 0.447247 | 8.814623571 | 0 |
| 2 | 47475114 | G | A | V | I | MSH2 | ENST00000233146 | 0.25893 | 0.00608 | 0.16066 | 0.55206 | 0.70427 | 0.5012647456 | 0.78754 | 0.3234169185 | 0.088629 | 1.275136099 | 0 |
| 2 | 47475114 | G | C | V | L | MSH2 | ENST00000233146 | 0.31337 | 0.0058 | 0.31971 | 0.73639 | 0.78348 | 0.727311484 | 0.9327 | 0.1143580964 | 0.182462 | 3.750990693 | 0 |
| 2 | 47475114 | G | T | V | F | MSH2 | ENST00000233146 | 0.46568 | 0.00804 | 0.29749 | 0.85365 | 0.9045 | 0.8659370378 | 0.90014 | 0.9554789066 | 0.237912 | 4.352542271 | 0 |
| 2 | 47475115 | T | A | V | D | MSH2 | ENST00000233146 | 0.56124 | 0.01108 | 0.35114 | 0.86936 | 0.94167 | 0.9043499338 | 0.87536 | 0.9950837493 | 0.294188 | 5.777135378 | 0 |
| 2 | 47475115 | T | C | V | A | MSH2 | ENST00000233146 | 0.38565 | 0.00551 | 0.34454 | 0.6922 | 0.82216 | 0.7557597558 | 0.81949 | 0.9478392601 | 0.247046 | 4.67744901 | 0 |
| 2 | 47475115 | T | G | V | G | MSH2 | ENST00000233146 | 0.55278 | 0.00795 | 0.2161 | 0.81202 | 0.9248 | 0.8709355515 | 0.814 | 0.9291217327 | 0.191822 | 4.353365091 | 0 |
| 2 | 47475117 | C | A | P | T | MSH2 | ENST00000233146 | 0.35747 | 0.0132 | 0.58523 | 0.86755 | 0.88401 | 0.810831362 | 0.86357 | 0.9790719151 | 0.279965 | 8.157722996 | 0 |
| 2 | 47475117 | C | G | P | A | MSH2 | ENST00000233146 | 0.3334 | 0.00627 | 0.52857 | 0.63601 | 0.78411 | 0.7569424673 | 0.84405 | 0.929197371 | 0.169448 | 4.150096597 | 0 |
| 2 | 47475117 | C | T | P | S | MSH2 | ENST00000233146 | 0.38983 | 0.00664 | 0.54457 | 0.76303 | 0.86302 | 0.807187863 | 0.80466 | 0.9465928078 | 0.176348 | 3.559466236 | 0 |
| 2 | 47475118 | C | A | P | Q | MSH2 | ENST00000233146 | 0.56415 | 0.02158 | 0.5478 | 0.80906 | 0.89147 | 0.8038757365 | 0.79464 | 0.9795748591 | 0.291774 | 4.309423958 | 0 |
| 2 | 47475118 | C | G | P | R | MSH2 | ENST00000233146 | 0.58027 | 0.02056 | 0.62737 | 0.80462 | 0.91652 | 0.854362139 | 0.88055 | 0.9744073749 | 0.374032 | 4.246131247 | 0 |
| 2 | 47475118 | C | T | P | L | MSH2 | ENST00000233146 | 0.43926 | 0.01361 | 0.631 | 0.87866 | 0.92028 | 0.8451327504 | 0.8756 | 0.9980461597 | 0.339616 | 7.21371842 | 0 |
| 2 | 47475120 | T | A | Y | N | MSH2 | ENST00000233146 | 0.91259 | 0.0296 | 0.64359 | 0.92664 | 0.96262 | 0.9428997276 | 0.93255 | 0.9997962117 | 0.458826 | 9.173670836 | 0 |
| 2 | 47475120 | T | C | Y | H | MSH2 | ENST00000233146 | 0.90896 | 0.03304 | 0.64965 | 0.88709 | 0.96189 | 0.9211330378 | 0.88375 | 0.9987183809 | 0.380061 | 9.173670836 | 0 |
| 2 | 47475120 | T | G | Y | D | MSH2 | ENST00000233146 | 0.91644 | 0.02844 | 0.63076 | 0.92442 | 0.97923 | 0.9628843355 | 0.97007 | 0.9999350309 | 0.484586 | 9.173670836 | 1 |
| 2 | 47475121 | A | C | Y | S | MSH2 | ENST00000233146 | 0.91942 | 0.03144 | 0.55711 | 0.93236 | 0.9648 | 0.9484565328 | 0.9463 | 0.9992245436 | 0.468871 | 9.173670836 | 0 |
| 2 | 47475121 | A | G | Y | C | MSH2 | ENST00000233146 | 0.9338 | 0.03353 | 0.64702 | 0.9054 | 0.9755 | 0.9342674786 | 0.95605 | 0.9993085861 | 0.471491 | 9.173670836 | 0 |
| 2 | 47475121 | A | T | Y | F | MSH2 | ENST00000233146 | 0.6918 | 0.01823 | 0.53458 | 0.76851 | 0.85792 | 0.7945512038 | 0.82685 | 0.9680703878 | 0.146672 | 3.348746441 | 0 |
| 2 | 47475123 | G | A | V | I | MSH2 | ENST00000233146 | 0.47658 | 0.0132 | 0.47963 | 0.62281 | 0.79273 | 0.7016386264 | 0.78977 | 0.7470520735 | 0.160401 | 2.570177749 | 0 |
| 2 | 47475123 | G | C | V | L | MSH2 | ENST00000233146 | 0.79571 | 0.02628 | 0.67543 | 0.83428 | 0.94986 | 0.8703027862 | 0.95401 | 0.9964410663 | 0.439656 | 7.120933832 | 0 |
| 2 | 47475123 | G | T | V | L | MSH2 | ENST00000233146 | 0.78931 | 0.02628 | 0.67543 | 0.83371 | 0.94949 | 0.8703027862 | 0.95423 | 0.9971113205 | 0.439655 | 7.120933832 | 0 |
| 2 | 47475124 | T | A | V | E | MSH2 | ENST00000233146 | 0.76459 | 0.02883 | 0.68154 | 0.92687 | 0.99006 | 0.9630893999 | 0.95875 | 0.9992663264 | 0.46689 | 6.521079589 | 0 |
| 2 | 47475124 | T | C | V | A | MSH2 | ENST00000233146 | 0.72188 | 0.03059 | 0.6976 | 0.86994 | 0.96773 | 0.872900141 | 0.8942 | 0.9925568899 | 0.380667 | 5.643627643 | 0 |
| 2 | 47475124 | T | G | V | G | MSH2 | ENST00000233146 | 0.75981 | 0.03205 | 0.55618 | 0.92233 | 0.98869 | 0.9568037765 | 0.94713 | 0.9994026423 | 0.472563 | 8.704192668 | 0 |
| 2 | 47475126 | C | G | R | G | MSH2 | ENST00000233146 | 0.8336 | 0.03327 | 0.57829 | 0.91335 | 0.94802 | 0.9557926658 | 0.96073 | 0.9978173971 | 0.450013 | 9.374475341 | 1 |
| 2 | 47475127 | G | C | R | P | MSH2 | ENST00000233146 | 0.83321 | 0.03166 | 0.63475 | 0.91982 | 0.98938 | 0.9713681465 | 0.94385 | 0.9981765747 | 0.482202 | 7.886016148 | 1 |
| 2 | 47475127 | G | T | R | L | MSH2 | ENST00000233146 | 0.81158 | 0.03362 | 0.61241 | 0.89489 | 0.98257 | 0.9429691061 | 0.97335 | 0.9982222915 | 0.476928 | 7.24515593 | 1 |
| 2 | 47475129 | C | A | P | T | MSH2 | ENST00000233146 | 0.64976 | 0.03547 | 0.54322 | 0.95846 | 0.99525 | 0.9337106744 | 0.9696 | 0.99990201 | 0.578501 | 9.937758368 | 1 |
| 2 | 47475129 | C | G | P | A | MSH2 | ENST00000233146 | 0.63229 | 0.03753 | 0.48901 | 0.93762 | 0.98869 | 0.9106003584 | 0.93631 | 0.9998642206 | 0.52685 | 9.937758368 | 0 |
| 2 | 47475129 | C | T | P | S | MSH2 | ENST00000233146 | 0.68147 | 0.03686 | 0.50083 | 0.93917 | 0.997 | 0.9420533048 | 0.97274 | 0.9999204874 | 0.578168 | 9.564133128 | 0 |
| 2 | 47475130 | C | A | P | Q | MSH2 | ENST00000233146 | 0.5998 | 0.03821 | 0.51673 | 0.93594 | 0.99386 | 0.9557654956 | 0.97458 | 0.9998595715 | 0.578046 | 9.937758368 | 1 |
| 2 | 47475130 | C | G | P | R | MSH2 | ENST00000233146 | 0.61425 | 0.03626 | 0.59886 | 0.92757 | 0.99386 | 0.9640197159 | 0.9749 | 0.9998508692 | 0.578046 | 9.937758368 | 1 |
| 2 | 47475130 | C | T | P | L | MSH2 | ENST00000233146 | 0.63183 | 0.03692 | 0.59928 | 0.92708 | 0.99386 | 0.9524720838 | 0.98563 | 0.9999738932 | 0.577713 | 9.937758368 | 1 |
| 2 | 47475132 | G | A | A | T | MSH2 | ENST00000233146 | 0.29939 | 0.0058 | 0.04174 | 0.52415 | 0.40608 | 0.4668261977 | 0.67872 | 0.09156002849 | -0.00703866 | -2.216984269 | 0 |
| 2 | 47475132 | G | C | A | P | MSH2 | ENST00000233146 | 0.35405 | 0.00715 | 0.05895 | 0.71436 | 0.75005 | 0.8283065569 | 0.86409 | 0.225300163 | 0.0705161 | 3.53974796 | 1 |
| 2 | 47475132 | G | T | A | S | MSH2 | ENST00000233146 | 0.26549 | 0.00592 | 0.02886 | 0.58441 | 0.54344 | 0.5185495697 | 0.82118 | 0.1189229488 | 0.0131691 | -0.8344730566 | 0 |
| 2 | 47475133 | C | A | A | D | MSH2 | ENST00000233146 | 0.13825 | 0.0054 | 0.0723 | 0.77147 | 0.70179 | 0.665185748 | 0.76759 | 0.2223833501 | 0.0302364 | 0.1201535477 | 0 |
| 2 | 47475133 | C | G | A | G | MSH2 | ENST00000233146 | 0.15223 | 0.00619 | 0.01916 | 0.70774 | 0.56829 | 0.5978051643 | 0.83353 | 0.1643658578 | 0.026649 | 1.965192608 | 0 |
| 2 | 47475133 | C | T | A | V | MSH2 | ENST00000233146 | 0.14963 | 0.00574 | 0.03638 | 0.63945 | 0.43577 | 0.3994905075 | 0.63327 | 0.1687226146 | -0.0286297 | -3.441747873 | 0 |
| 2 | 47475135 | A | C | I | L | MSH2 | ENST00000233146 | 0.398 | 0.00586 | 0.50633 | 0.47236 | 0.58644 | 0.6047094376 | 0.85111 | 0.4820145369 | 0.0641823 | 1.715360821 | 0 |
| 2 | 47475135 | A | G | I | V | MSH2 | ENST00000233146 | 0.39318 | 0.00644 | 0.37934 | 0.43156 | 0.64876 | 0.4671134339 | 0.86663 | 0.4009704292 | 0.0805966 | 2.649066296 | 0 |
| 2 | 47475135 | A | T | I | F | MSH2 | ENST00000233146 | 0.58796 | 0.0233 | 0.51665 | 0.74253 | 0.82892 | 0.7795457889 | 0.9687 | 0.8165527582 | 0.223171 | 3.696067978 | 0 |
| 2 | 47475136 | T | A | I | N | MSH2 | ENST00000233146 | 0.77766 | 0.02582 | 0.62031 | 0.86469 | 0.96883 | 0.8970496605 | 0.85317 | 0.9986822009 | 0.301083 | 8.19725499 | 1 |
| 2 | 47475136 | T | C | I | T | MSH2 | ENST00000233146 | 0.69107 | 0.02056 | 0.6543 | 0.80629 | 0.95935 | 0.8136493701 | 0.92496 | 0.9899779558 | 0.368883 | 7.090712516 | 0 |
| 2 | 47475136 | T | G | I | S | MSH2 | ENST00000233146 | 0.7737 | 0.02366 | 0.5361 | 0.84315 | 0.96846 | 0.9264603624 | 0.93023 | 0.9977745414 | 0.424979 | 8.19725499 | 1 |
| 2 | 47475137 | T | G | I | M | MSH2 | ENST00000233146 | 0.27688 | 0.01358 | 0.45488 | 0.69779 | 0.83283 | 0.6502089493 | 0.92798 | 0.495764792 | 0.13506 | 3.075521377 | 0 |
| 2 | 47475138 | T | A | L | M | MSH2 | ENST00000233146 | 0.54082 | 0.03595 | 0.53859 | 0.73104 | 0.77011 | 0.6585358669 | 0.84084 | 0.9309242964 | 0.0904091 | 6.642779912 | 0 |
| 2 | 47475138 | T | G | L | V | MSH2 | ENST00000233146 | 0.34882 | 0.01019 | 0.50106 | 0.52214 | 0.79938 | 0.6738551099 | 0.91242 | 0.3814520836 | 0.109579 | 4.090060441 | 0 |
| 2 | 47475139 | T | C | L | S | MSH2 | ENST00000233146 | 0.6372 | 0.02574 | 0.58402 | 0.72947 | 0.77463 | 0.8961564575 | 0.87479 | 0.7718957663 | 0.250162 | 2.701953451 | 0 |
| 2 | 47475139 | T | G | L | W | MSH2 | ENST00000233146 | 0.83391 | 0.02891 | 0.65746 | 0.86316 | 0.87733 | 0.8689517789 | 0.78667 | 0.9839857221 | 0.210186 | 7.833770253 | 0 |
| 2 | 47475140 | G | C | L | F | MSH2 | ENST00000233146 | 0.57603 | 0.00974 | 0.55179 | 0.76321 | 0.76487 | 0.7460071918 | 0.75428 | 0.9540264606 | 0.143931 | 6.408973053 | 0 |
| 2 | 47475140 | G | T | L | F | MSH2 | ENST00000233146 | 0.57027 | 0.00974 | 0.55179 | 0.76363 | 0.76487 | 0.7460071918 | 0.75444 | 0.9540264606 | 0.143931 | 6.408973053 | 0 |
| 2 | 47475141 | G | A | E | K | MSH2 | ENST00000233146 | 0.66945 | 0.00556 | 0.45476 | 0.68443 | 0.83725 | 0.6001485812 | 0.90475 | 0.9312953949 | 0.242633 | 4.109596581 | 0 |
| 2 | 47475141 | G | C | E | Q | MSH2 | ENST00000233146 | 0.65995 | 0.00795 | 0.33422 | 0.58906 | 0.78844 | 0.5582949922 | 0.87929 | 0.9430843592 | 0.204657 | 4.269653086 | 0 |
| 2 | 47475142 | A | C | E | A | MSH2 | ENST00000233146 | 0.41611 | 0.00566 | 0.34721 | 0.56039 | 0.613 | 0.5901814094 | 0.80326 | 0.6523233056 | 0.112994 | 2.913049872 | 0 |
| 2 | 47475142 | A | G | E | G | MSH2 | ENST00000233146 | 0.63008 | 0.00547 | 0.31099 | 0.65463 | 0.7727 | 0.6506057479 | 0.72791 | 0.928761363 | 0.129365 | 5.425160328 | 0 |
| 2 | 47475142 | A | T | E | V | MSH2 | ENST00000233146 | 0.65101 | 0.00633 | 0.32975 | 0.70567 | 0.88448 | 0.6716146315 | 0.92464 | 0.9587146044 | 0.263064 | 5.357409178 | 0 |
| 2 | 47475143 | G | C | E | D | MSH2 | ENST00000233146 | 0.29128 | 0.00592 | 0.34819 | 0.48947 | 0.73223 | 0.4666615099 | 0.96349 | 0.4930919111 | 0.115787 | 2.718942229 | 0 |
| 2 | 47475143 | G | T | E | D | MSH2 | ENST00000233146 | 0.28729 | 0.00592 | 0.34819 | 0.49106 | 0.73148 | 0.4666615099 | 0.96179 | 0.4930919111 | 0.115805 | 2.718942229 | 0 |
| 2 | 47475144 | A | C | K | Q | MSH2 | ENST00000233146 | 0.55958 | 0.00783 | 0.39225 | 0.61417 | 0.77205 | 0.5722358511 | 0.91574 | 0.7174558043 | 0.171895 | 2.817532071 | 0 |
| 2 | 47475144 | A | G | K | E | MSH2 | ENST00000233146 | 0.56269 | 0.00586 | 0.44385 | 0.63314 | 0.73805 | 0.6209101095 | 0.83711 | 0.9281675816 | 0.136692 | 3.575544708 | 0 |
| 2 | 47475145 | A | C | K | T | MSH2 | ENST00000233146 | 0.51197 | 0.00681 | 0.45067 | 0.58198 | 0.73074 | 0.6599840564 | 0.85335 | 0.8465977907 | 0.184416 | 4.042353194 | 0 |
| 2 | 47475145 | A | G | K | R | MSH2 | ENST00000233146 | 0.64843 | 0.01699 | 0.35603 | 0.56669 | 0.76421 | 0.5164951103 | 0.89497 | 0.7245783806 | 0.157986 | 3.703725126 | 0 |
| 2 | 47475145 | A | T | K | I | MSH2 | ENST00000233146 | 0.86382 | 0.00763 | 0.4156 | 0.75207 | 0.87 | 0.6939841208 | 0.85826 | 0.983297348 | 0.218849 | 7.134690268 | 0 |
| 2 | 47475146 | A | C | K | N | MSH2 | ENST00000233146 | 0.76426 | 0.01848 | 0.47682 | 0.68571 | 0.77845 | 0.6348317715 | 0.90285 | 0.9515377283 | 0.0874991 | 3.653899083 | 0 |
| 2 | 47475146 | A | T | K | N | MSH2 | ENST00000233146 | 0.7575 | 0.01848 | 0.47682 | 0.68571 | 0.77845 | 0.6348317715 | 0.90296 | 0.9515377283 | 0.0874908 | 3.653899083 | 0 |
| 2 | 47475147 | G | A | G | R | MSH2 | ENST00000233146 | 0.88304 | 0.03442 | 0.56029 | 0.81966 | 0.848 | 0.8695925727 | 0.82154 | 0.9869143367 | 0.163191 | 8.454955023 | 0 |
| 2 | 47475147 | G | C | G | R | MSH2 | ENST00000233146 | 0.88042 | 0.03442 | 0.56029 | 0.80843 | 0.84694 | 0.8695925727 | 0.81829 | 0.9974480867 | 0.289573 | 8.454955023 | 0 |
| 2 | 47475148 | G | A | G | E | MSH2 | ENST00000233146 | 0.55504 | 0.03155 | 0.54215 | 0.73005 | 0.87 | 0.8441449128 | 0.86886 | 0.9820528626 | 0.380368 | 7.946670291 | 0 |
| 2 | 47475148 | G | C | G | A | MSH2 | ENST00000233146 | 0.66494 | 0.03547 | 0.43687 | 0.7608 | 0.8705 | 0.8312176985 | 0.89918 | 0.9958119392 | 0.345852 | 4.816844255 | 0 |
| 2 | 47475148 | G | T | G | V | MSH2 | ENST00000233146 | 0.69933 | 0.03362 | 0.45213 | 0.79717 | 0.91229 | 0.8810718591 | 0.93897 | 0.9985983968 | 0.433953 | 5.256570581 | 0 |
| 2 | 47475150 | C | A | Q | K | MSH2 | ENST00000233146 | 0.19024 | 0.00532 | 0.05869 | 0.5759 | 0.77399 | 0.5680943227 | 0.95602 | 0.3901615441 | 0.136383 | 2.531108821 | 0 |
| 2 | 47475150 | C | G | Q | E | MSH2 | ENST00000233146 | 0.14229 | 0.00556 | 0.0454 | 0.48816 | 0.77972 | 0.5896661524 | 0.89405 | 0.08150086552 | 0.0768164 | 1.116199645 | 0 |
| 2 | 47475151 | A | C | Q | P | MSH2 | ENST00000233146 | 0.22431 | 0.00535 | 0.04959 | 0.65297 | 0.76619 | 0.8228966749 | 0.94768 | 0.08430998027 | 0.109201 | 5.598993987 | 0 |
| 2 | 47475151 | A | G | Q | R | MSH2 | ENST00000233146 | 0.19581 | 0.00556 | 0.05539 | | 0.79395 | 0.5668618839 | 0.99686 | 0.009369518153 | -0.116592 | 3.064581989 | 0 |
| 2 | 47475151 | A | T | Q | L | MSH2 | ENST00000233146 | 0.24029 | 0.00622 | 0.04221 | 0.74062 | 0.77463 | 0.6303325818 | 0.9297 | 0.3567739725 | 0.126887 | 6.27954904 | 0 |
| 2 | 47475152 | A | C | Q | H | MSH2 | ENST00000233146 | 0.28788 | 0.00592 | 0.05925 | 0.69372 | 0.78411 | 0.6359670281 | 0.97841 | 0.2971500754 | 0.131798 | 3.854915014 | 0 |
| 2 | 47475152 | A | T | Q | H | MSH2 | ENST00000233146 | 0.28379 | 0.00592 | 0.05925 | 0.69372 | 0.78411 | 0.6359670281 | 0.97784 | 0.5659429431 | 0.131841 | 3.854915014 | 0 |

| | | | | | | | | | | | | | | | | | | |
|---|---|---|---|---|---|---|---|---|---|---|---|---|---|---|---|---|---|---|
| 2 | 47475153 | G | A | G | R | MSH2 | ENST00000233146 | 0.61918 | 0.00681 | 0.42239 | 0.83415 | 0.90494 | 0.6737236379 | 0.79286 | 0.9675624967 | 0.145858 | 3.275745676 | 0 |
| 2 | 47475153 | G | C | G | R | MSH2 | ENST00000233146 | 0.61611 | 0.00681 | 0.42239 | 0.82842 | 0.90494 | 0.6737236379 | 0.77309 | 0.9675624967 | 0.14589 | 3.275745676 | 0 |
| 2 | 47475154 | G | A | G | E | MSH2 | ENST00000233146 | 0.71586 | 0.02574 | 0.40035 | 0.89456 | 0.94876 | 0.6622344914 | 0.89928 | 0.9981151819 | 0.366274 | 5.105335817 | 0 |
| 2 | 47475154 | G | C | G | A | MSH2 | ENST00000233146 | 0.66153 | 0.02506 | 0.30753 | 0.83977 | 0.89285 | 0.5824427353 | 0.8631 | 0.9959765077 | 0.271301 | 6.753354075 | 0 |
| 2 | 47475154 | G | T | G | V | MSH2 | ENST00000233146 | 0.71378 | 0.03039 | 0.31969 | 0.86232 | 0.94356 | 0.7034813561 | 0.89884 | 0.9969260097 | 0.404245 | 8.726782785 | 0 |
| 2 | 47475156 | A | G | R | G | MSH2 | ENST00000233146 | 0.4843 | 0.00527 | 0.17948 | 0.63421 | 0.7589 | 0.9156693798 | 0.9407 | 0.5307907462 | 0.162877 | 2.489508602 | 0 |
| 2 | 47475157 | G | A | R | K | MSH2 | ENST00000233146 | 0.23596 | 0.00569 | 0.21673 | 0.41727 | 0.68558 | 0.8567650606 | 0.83332 | 0.2986882329 | 0.0682765 | 2.303963638 | 0 |
| 2 | 47475157 | G | C | R | T | MSH2 | ENST00000233146 | 0.28524 | 0.00569 | 0.22473 | 0.65694 | 0.82836 | 0.871482055 | 0.94755 | 0.4925475121 | 0.238228 | 3.131733244 | 0 |
| 2 | 47475157 | G | T | R | I | MSH2 | ENST00000233146 | 0.33153 | 0.0048 | 0.20577 | 0.70115 | 0.80298 | 0.8493583924 | 0.91686 | 0.2552731162 | 0.0745695 | 2.033731134 | 0 |
| 2 | 47475158 | A | C | R | S | MSH2 | ENST00000233146 | 0.22611 | 0.00556 | 0.18771 | 0.60404 | 0.78782 | 0.8851051472 | 0.9766 | 0.307838738 | 0.2058 | 3.85896838 | 0 |
| 2 | 47475158 | A | T | R | S | MSH2 | ENST00000233146 | 0.22269 | 0.00556 | 0.18771 | 0.60404 | 0.78782 | 0.8851051472 | 0.97285 | 0.2078441828 | 0.205856 | 3.85896838 | 0 |
| 2 | 47475159 | A | C | I | L | MSH2 | ENST00000233146 | 0.32717 | 0.0058 | 0.31708 | 0.46051 | 0.64876 | 0.578965012 | 0.84534 | 0.3733632863 | 0.0455633 | 1.17554574 | 0 |
| 2 | 47475159 | A | G | I | V | MSH2 | ENST00000233146 | 0.35146 | 0.00615 | 0.23191 | 0.598 | 0.74867 | 0.4087429213 | 0.86768 | 0.1614759173 | -0.08362 | 3.993721556 | 0 |
| 2 | 47475159 | A | T | I | F | MSH2 | ENST00000233146 | 0.42968 | 0.01533 | 0.32947 | 0.73639 | 0.91862 | 0.7799952341 | 0.92031 | 0.8940740824 | 0.190989 | 5.725614519 | 0 |
| 2 | 47475160 | T | A | I | N | MSH2 | ENST00000233146 | 0.71153 | 0.02166 | 0.41025 | 0.87632 | 0.9367 | 0.8987328252 | 0.84481 | 0.992775321 | 0.270367 | 8.236195206 | 0 |
| 2 | 47475160 | T | C | I | T | MSH2 | ENST00000233146 | 0.368 | 0.01401 | 0.4473 | 0.54662 | 0.8705 | 0.8116864469 | 0.83613 | 0.9494357109 | 0.245362 | 6.103509908 | 0 |
| 2 | 47475160 | T | G | I | S | MSH2 | ENST00000233146 | 0.70334 | 0.01715 | 0.34305 | 0.86462 | 0.93475 | 0.9278455357 | 0.92948 | 0.9913707972 | 0.402961 | 8.236195206 | 0 |
| 2 | 47475161 | T | G | I | M | MSH2 | ENST00000233146 | 0.13317 | 0.00763 | 0.28373 | 0.67662 | 0.82216 | 0.6070039949 | 0.90404 | 0.4794430435 | 0.0639331 | 4.244992071 | 0 |
| 2 | 47475162 | A | C | I | L | MSH2 | ENST00000233146 | 0.12457 | 0.00574 | 0.04745 | 0.61242 | 0.50616 | 0.5091297552 | 0.85515 | 0.1143157184 | 0.0252964 | 1.969642546 | 0 |
| 2 | 47475162 | A | G | I | V | MSH2 | ENST00000233146 | 0.09779 | 0.00603 | 0.02633 | 0.47508 | 0.47225 | 0.3701120161 | 0.72265 | 0.03406080926 | -0.133821 | 0.1142448735 | 0 |
| 2 | 47475162 | A | T | I | L | MSH2 | ENST00000233146 | 0.12187 | 0.00574 | 0.04745 | 0.61242 | 0.50147 | 0.5091297552 | 0.85908 | 0.1291255206 | 0.0253012 | 1.969642546 | 0 |
| 2 | 47475163 | T | C | I | T | MSH2 | ENST00000233146 | 0.1135 | 0.00532 | 0.10297 | 0.48196 | 0.60123 | 0.5558479621 | 0.75069 | 0.01099758415 | -0.0079026 | 1.144050954 | 0 |
| 2 | 47475163 | T | G | I | R | MSH2 | ENST00000233146 | 0.12712 | 0.00476 | 0.06745 | 0.59669 | 0.72926 | 0.7132236055 | 0.87271 | 0.02016978474 | 0.0166631 | 1.133295706 | 0 |
| 2 | 47475165 | T | A | L | I | MSH2 | ENST00000233146 | 0.21355 | 0.0203 | 0.44551 | 0.623 | 0.79938 | 0.7223599846 | 0.77283 | 0.7403700352 | 0.145074 | 4.013561915 | 0 |
| 2 | 47475165 | T | G | L | V | MSH2 | ENST00000233146 | 0.20764 | 0.0099 | 0.38475 | 0.61482 | 0.8345 | 0.8222989018 | 0.95394 | 0.9673613906 | 0.268814 | 4.965217806 | 0 |
| 2 | 47475166 | T | C | L | S | MSH2 | ENST00000233146 | 0.89405 | 0.03029 | 0.46145 | 0.85247 | 0.96262 | 0.9796292386 | 0.97256 | 0.9994537234 | 0.473636 | 9.308120606 | 0 |
| 2 | 47475167 | A | C | L | F | MSH2 | ENST00000233146 | 0.53093 | 0.03293 | 0.42954 | 0.85594 | 0.93162 | 0.9015564243 | 0.9038 | 0.9869000316 | 0.260765 | 8.093779081 | 0 |
| 2 | 47475167 | A | T | L | F | MSH2 | ENST00000233146 | 0.52532 | 0.03293 | 0.42954 | 0.85768 | 0.93162 | 0.9015564243 | 0.9017 | 0.9869000316 | 0.260777 | 8.093779081 | 0 |
| 2 | 47475168 | A | C | K | Q | MSH2 | ENST00000233146 | 0.3211 | 0.00592 | 0.26972 | 0.44128 | 0.70097 | 0.6336006961 | 0.93422 | 0.4618730843 | 0.0874783 | 3.692484316 | 0 |
| 2 | 47475168 | A | G | K | E | MSH2 | ENST00000233146 | 0.30244 | 0.00664 | 0.31138 | 0.55709 | 0.77205 | 0.7280165059 | 0.94598 | 0.5540707111 | 0.148841 | 3.368441454 | 0 |
| 2 | 47475169 | A | C | K | T | MSH2 | ENST00000233146 | 0.35168 | 0.00543 | 0.32113 | 0.58957 | 0.72775 | 0.6726754831 | 0.94284 | 0.1102239415 | 0.0796839 | 2.667199602 | 0 |
| 2 | 47475169 | A | G | K | R | MSH2 | ENST00000233146 | 0.35875 | 0.00615 | 0.23625 | 0.4474 | 0.69678 | 0.6213553408 | 0.93159 | 0.1154875302 | 0.0644441 | 3.365909613 | 0 |
| 2 | 47475169 | A | T | K | I | MSH2 | ENST00000233146 | 0.42085 | 0.00483 | 0.29808 | 0.66672 | 0.77972 | 0.6830123383 | 0.97511 | 0.2673011124 | 0.156091 | 2.986571596 | 0 |
| 2 | 47475171 | G | A | A | T | MSH2 | ENST00000233146 | 0.38679 | 0.0084 | 0.16527 | 0.5411 | 0.72626 | 0.5004171659 | 0.80607 | 0.5993599463 | 0.119429 | 6.420202438 | 0 |
| 2 | 47475171 | G | C | A | P | MSH2 | ENST00000233146 | 0.42138 | 0.0129 | 0.22473 | 0.68421 | 0.86251 | 0.8755609665 | 0.9519 | 0.4135669709 | 0.376499 | 6.420202438 | 1 |
| 2 | 47475171 | G | T | A | S | MSH2 | ENST00000233146 | 0.33169 | 0.00599 | 0.13868 | 0.40143 | 0.65178 | 0.445288589 | 0.80141 | 0.5043087602 | 0.078678 | 2.07355362 | 0 |
| 2 | 47475172 | C | A | A | E | MSH2 | ENST00000233146 | 0.17063 | 0.00556 | 0.21697 | 0.40197 | 0.72016 | 0.656106684 | 0.88902 | 0.3866191208 | 0.134483 | 1.190326053 | 0 |
| 2 | 47475172 | C | G | A | G | MSH2 | ENST00000233146 | 0.20962 | 0.00574 | 0.15259 | 0.13464 | 0.60481 | 0.5088145775 | 0.71386 | 0.641551733 | 0.0242514 | 0.4037513545 | 0 |
| 2 | 47475172 | C | T | A | V | MSH2 | ENST00000233146 | 0.27091 | 0.00945 | 0.18153 | 0.58582 | 0.81414 | 0.5783002566 | 0.82288 | 0.7299249172 | 0.145114 | 5.456077321 | 0 |
| 2 | 47475174 | T | A | S | T | MSH2 | ENST00000233146 | 0.52387 | 0.01191 | 0.22391 | 0.64215 | 0.78473 | 0.7973731504 | 0.90335 | 0.9567789435 | 0.13286 | 6.435060109 | 0 |
| 2 | 47475174 | T | C | S | P | MSH2 | ENST00000233146 | 0.75185 | 0.02494 | 0.23272 | 0.75918 | 0.87974 | 0.9609970014 | 0.93554 | 0.9730326533 | 0.249058 | 6.435060109 | 0 |
| 2 | 47475174 | T | G | S | A | MSH2 | ENST00000233146 | 0.32336 | 0.00615 | 0.20089 | 0.19415 | 0.62547 | 0.7016831895 | 0.75517 | 0.6405918598 | 0.0322351 | 0.506860098 | 0 |
| 2 | 47475175 | C | G | S | C | MSH2 | ENST00000233146 | 0.39585 | 0.00603 | 0.22139 | 0.39817 | 0.66828 | 0.8309710042 | 0.89386 | 0.7369026542 | 0.117195 | 3.523987173 | 0 |
| 2 | 47475175 | C | T | S | F | MSH2 | ENST00000233146 | 0.70784 | 0.02696 | 0.25655 | 0.81922 | 0.91058 | 0.8859174592 | 0.84866 | 0.9939959049 | 0.229881 | 6.435060109 | 0 |
| 2 | 47475177 | A | G | R | G | MSH2 | ENST00000233146 | 0.83151 | 0.0334 | 0.42851 | 0.93933 | 0.99039 | 0.9734765883 | 0.98405 | 0.9989055395 | 0.451593 | 9.486754443 | 0 |
| 2 | 47475177 | A | T | R | W | MSH2 | ENST00000233146 | 0.89143 | 0.0366 | 0.53513 | 0.92941 | 0.97848 | 0.9633401292 | 0.95885 | 0.9998703003 | 0.450526 | 6.647235371 | 0 |
| 2 | 47475178 | G | A | R | K | MSH2 | ENST00000233146 | 0.82934 | 0.03692 | 0.50379 | 0.83459 | 0.95316 | 0.9461408645 | 0.977 | 0.9994394183 | 0.475772 | 7.094158584 | 0 |
| 2 | 47475178 | G | C | R | T | MSH2 | ENST00000233146 | 0.83283 | 0.03137 | 0.48603 | 0.89056 | 0.97252 | 0.9641530609 | 0.98414 | 0.9995638728 | 0.48627 | 9.486754443 | 0 |
| 2 | 47475179 | G | C | R | S | MSH2 | ENST00000233146 | 0.58254 | 0.03538 | 0.44445 | 0.94802 | 0.97215 | 0.9674519673 | 0.9938 | 0.9985582232 | 0.414363 | 9.486754443 | 0 |
| 2 | 47475179 | G | T | R | S | MSH2 | ENST00000233146 | 0.5768 | 0.03538 | 0.44445 | 0.94775 | 0.97215 | 0.9674519673 | 0.99312 | 0.9988029003 | 0.414391 | 9.486754443 | 0 |
| 2 | 47475180 | C | A | H | N | MSH2 | ENST00000233146 | 0.6735 | 0.03582 | 0.55242 | 0.89044 | 0.92233 | 0.9591102458 | 0.94993 | 0.9994289279 | 0.335973 | 9.999862403 | 0 |
| 2 | 47475180 | C | G | H | D | MSH2 | ENST00000233146 | 0.70984 | 0.03243 | 0.54611 | 0.88753 | 0.98294 | 0.9861708931 | 0.97982 | 0.9999426603 | 0.52047 | 9.999862403 | 0 |
| 2 | 47475181 | A | C | H | P | MSH2 | ENST00000233146 | 0.77251 | 0.03059 | 0.48382 | 0.90814 | 0.99443 | 0.9855126652 | 0.98475 | 0.999938488 | 0.532146 | 9.999862403 | 0 |
| 2 | 47475181 | A | G | H | R | MSH2 | ENST00000233146 | 0.72612 | 0.03419 | 0.5608 | 0.88642 | 0.99676 | 0.9833385907 | 0.98933 | 0.999828577 | 0.529651 | 9.999862403 | 0 |
| 2 | 47475181 | A | T | H | L | MSH2 | ENST00000233146 | 0.77561 | 0.03398 | 0.51607 | 0.92665 | 0.99414 | 0.9751406112 | 0.9885 | 0.999938488 | 0.530494 | 9.999862403 | 0 |
| 2 | 47475182 | T | A | H | Q | MSH2 | ENST00000233146 | 0.55813 | 0.03713 | 0.52751 | 0.9524 | 0.9703 | 0.9695864776 | 0.99335 | 0.9998170733 | 0.473344 | 9.999862403 | 0 |
| 2 | 47475182 | T | G | H | Q | MSH2 | ENST00000233146 | 0.56526 | 0.03713 | 0.52751 | 0.94916 | 0.96993 | 0.9695864776 | 0.99329 | 0.9997863173 | 0.473345 | 9.999862403 | 0 |
| 2 | 47475183 | G | A | A | T | MSH2 | ENST00000233146 | 0.64641 | 0.00916 | 0.22182 | 0.78459 | 0.84428 | 0.7666119369 | 0.8343 | 0.7207899094 | 0.187102 | 5.455753684 | 0 |
| 2 | 47475183 | G | C | A | P | MSH2 | ENST00000233146 | 0.29829 | 0.00527 | 0.26414 | 0.23631 | 0.62657 | 0.7829315492 | 0.44327 | 0.3783602417 | 0.00306416 | -1.779001724 | 0 |
| 2 | 47475183 | G | T | A | S | MSH2 | ENST00000233146 | 0.49647 | 0.00747 | 0.1854 | 0.78015 | 0.82046 | 0.7456506322 | 0.78505 | 0.223961393 | -0.0289967 | 5.455753684 | 0 |
| 2 | 47475184 | C | G | A | G | MSH2 | ENST00000233146 | 0.68403 | 0.00791 | 0.19424 | 0.67794 | 0.82556 | 0.7069108945 | 0.7897 | 0.9093763828 | 0.173485 | 5.455753684 | 0 |
| 2 | 47475184 | C | T | A | V | MSH2 | ENST00000233146 | 0.68835 | 0.01019 | 0.23194 | 0.70871 | 0.83888 | 0.7355937559 | 0.84067 | 0.9093524218 | 0.21328 | 5.455753684 | 0 |
| 2 | 47475186 | T | A | C | S | MSH2 | ENST00000233146 | 0.81693 | 0.03413 | 0.53356 | 0.91787 | 0.98367 | 0.9417696903 | 0.93184 | 0.9849963359 | 0.452054 | 6.977597013 | 0 |
| 2 | 47475186 | T | C | C | R | MSH2 | ENST00000233146 | 0.87709 | 0.02864 | 0.66297 | 0.91173 | 0.97997 | 0.9806682356 | 0.96059 | 0.9845529201 | 0.472981 | 8.035177439 | 0 |
| 2 | 47475186 | T | G | C | G | MSH2 | ENST00000233146 | 0.86311 | 0.03394 | 0.48849 | 0.91372 | 0.94091 | 0.9439428559 | 0.92903 | 0.9809368395 | 0.468605 | 8.035177439 | 0 |
| 2 | 47475187 | G | A | C | Y | MSH2 | ENST00000233146 | 0.87827 | 0.02934 | 0.68694 | 0.866 | 0.97475 | 0.9580278149 | 0.94967 | 0.9997144341 | 0.346545 | 8.035177439 | 0 |
| 2 | 47475187 | G | C | C | S | MSH2 | ENST00000233146 | 0.86061 | 0.03413 | 0.53356 | 0.84792 | 0.96956 | 0.9417696903 | 0.93178 | 0.9999034405 | 0.461 | 6.977597013 | 0 |
| 2 | 47475187 | G | T | C | F | MSH2 | ENST00000233146 | 0.89515 | 0.03083 | 0.56019 | 0.87936 | 0.96846 | 0.9582379747 | 0.95779 | 0.9999238253 | 0.462344 | 8.035177439 | 0 |
| 2 | 47475189 | G | A | V | I | MSH2 | ENST00000233146 | 0.30735 | 0.00654 | 0.1224 | 0.45241 | 0.61646 | 0.3228880266 | 0.84937 | 0.1318331734 | -0.087604 | -0.2303477922 | 0 |
| 2 | 47475189 | G | C | V | L | MSH2 | ENST00000233146 | 0.25944 | 0.00592 | 0.25536 | 0.44908 | 0.62876 | 0.469627653 | 0.85117 | 0.835080862 | 0.068988 | 0.4710622751 | 0 |
| 2 | 47475189 | G | T | V | F | MSH2 | ENST00000233146 | 0.66059 | 0.02307 | 0.23574 | 0.90687 | 0.9182 | 0.7962488969 | 0.93399 | 0.9841622114 | 0.269014 | 5.890191345 | 0 |
| 2 | 47475190 | T | A | V | D | MSH2 | ENST00000233146 | 0.80692 | 0.02719 | 0.27723 | 0.92789 | 0.99202 | 0.8910060493 | 0.95983 | 0.9996436834 | 0.462002 | 5.890191345 | 0 |
| 2 | 47475190 | T | C | V | A | MSH2 | ENST00000233146 | 0.72989 | 0.01533 | 0.28702 | 0.80564 | 0.91862 | 0.6455268251 | 0.90658 | 0.9862546921 | 0.253461 | 4.760109094 | 0 |
| 2 | 47475190 | T | G | V | G | MSH2 | ENST00000233146 | 0.78994 | 0.02696 | 0.17115 | 0.91086 | 0.98367 | 0.8079714211 | 0.93892 | 0.9987108707 | 0.431099 | 5.890191345 | 0 |
| 2 | 47475192 | G | A | E | K | MSH2 | ENST00000233146 | 0.9112 | 0.03542 | 0.68474 | 0.90039 | 0.96846 | 0.9608744117 | 0.99158 | 0.9992917776 | 0.409988 | 9.349921155 | 0 |
| 2 | 47475192 | G | C | E | Q | MSH2 | ENST00000233146 | 0.86212 | 0.03735 | 0.55593 | 0.87402 | 0.95097 | 0.9199001033 | 0.97383 | 0.9997014403 | 0.442202 | 7.022512385 | 0 |
| 2 | 47475193 | A | C | E | A | MSH2 | ENST00000233146 | 0.84127 | 0.03626 | 0.57491 | 0.90617 | 0.98034 | 0.9409331275 | 0.97684 | 0.9992819428 | 0.480594 | 8.240779798 | 0 |
| 2 | 47475193 | A | G | E | G | MSH2 | ENST00000233146 | 0.88323 | 0.03484 | 0.522 | 0.92678 | 0.99498 | 0.9525669938 | 0.98049 | 0.9998810291 | 0.486816 | 9.349921155 | 0 |
| 2 | 47475193 | A | T | E | V | MSH2 | ENST00000233146 | 0.8681 | 0.03484 | 0.54602 | 0.90884 | 0.99202 | 0.9434666123 | 0.97683 | 0.9997450709 | 0.486229 | 9.349921155 | 0 |
| 2 | 47475194 | A | C | E | D | MSH2 | ENST00000233146 | 0.56221 | 0.03864 | 0.572 | 0.91855 | 0.95535 | 0.8945924673 | 0.9816 | 0.997009635 | 0.425174 | 7.748278629 | 0 |
| 2 | 47475194 | A | T | E | D | MSH2 | ENST00000233146 | 0.55642 | 0.03864 | 0.572 | 0.92244 | 0.95535 | 0.8945924673 | 0.98176 | 0.9975163937 | 0.425169 | 7.748278629 | 0 |
| 2 | 47475195 | G | A | V | I | MSH2 | ENST00000233146 | 0.31307 | 0.00638 | 0.1679 | 0.51384 | 0.58263 | 0.4535769339 | 0.84665 | 0.7761220336 | 0.0818233 | 3.108333104 | 0 |
| 2 | 47475195 | G | C | V | L | MSH2 | ENST00000233146 | 0.32199 | 0.00592 | 0.285 | 0.54748 | 0.6331 | 0.6006175538 | 0.9104 | 0.8295842409 | 0.103868 | 2.985058405 | 0 |
| 2 | 47475195 | G | T | V | F | MSH2 | ENST00000233146 | 0.50354 | 0.0178 | 0.2655 | 0.78857 | 0.88916 | 0.7724547665 | 0.87227 | 0.9739269614 | 0.222076 | 4.334414111 | 0 |
| 2 | 47475196 | T | A | V | D | MSH2 | ENST00000233146 | 0.51738 | 0.02056 | 0.31443 | 0.86541 | 0.90885 | 0.8114995326 | 0.8858 | 0.9407285452 | 0.25656 | 5.92511075 | 0 |
| 2 | 47475196 | T | C | V | A | MSH2 | ENST00000233146 | 0.31452 | 0.00556 | 0.31117 | 0.37594 | 0.59393 | 0.6066350689 | 0.82263 | 0.7866684794 | 0.0759949 | 2.600398817 | 0 |
| 2 | 47475196 | T | G | V | G | MSH2 | ENST00000233146 | 0.47627 | 0.01244 | 0.20818 | 0.83475 | 0.86553 | 0.75942628 | 0.89201 | 0.9855291247 | 0.251864 | 7.351010159 | 0 |
| 2 | 47475198 | C | A | Q | K | MSH2 | ENST00000233146 | 0.79729 | 0.03124 | 0.36362 | 0.84192 | 0.87733 | 0.7352237971 | 0.87865 | 0.9904971123 | 0.278115 | 7.620857319 | 0 |
| 2 | 47475198 | C | G | Q | E | MSH2 | ENST00000233146 | 0.75312 | 0.02683 | 0.3145 | 0.78524 | 0.84106 | 0.7148361661 | 0.86105 | 0.8916656971 | 0.113149 | 7.607185686 | 0 |
| 2 | 47475199 | A | C | Q | P | MSH2 | ENST00000233146 | 0.87788 | 0.0334 | 0.32747 | 0.86899 | 0.98146 | 0.8855217623 | 0.91336 | 0.9972593784 | 0.449019 | 6.850484117 | 0 |
| 2 | 47475199 | A | G | Q | R | MSH2 | ENST00000233146 | 0.86075 | 0.03484 | 0.35863 | 0.84601 | 0.94281 | 0.7626780757 | 0.92428 | 0.9897719026 | 0.294531 | 8.491275417 | 0 |
| 2 | 47475199 | A | T | Q | L | MSH2 | ENST00000233146 | 0.8944 | 0.01253 | 0.30748 | 0.83665 | 0.93241 | 0.7278063874 | 0.85218 | 0.9875213504 | 0.264853 | 3.438357347 | 0 |
| 2 | 47475200 | A | C | Q | H | MSH2 | ENST00000233146 | 0.59588 | 0.03741 | 0.35748 | 0.83478 | 0.89147 | 0.7756713772 | 0.91897 | 0.996312201 | 0.230968 | 7.147908855 | 0 |
| 2 | 47475200 | A | T | Q | H | MSH2 | ENST00000233146 | 0.58981 | 0.03741 | 0.35748 | 0.84698 | 0.89147 | 0.7756713772 | 0.91897 | 0.9969367981 | 0.230968 | 7.147908855 | 0 |
| 2 | 47475201 | G | A | D | N | MSH2 | ENST00000233146 | 0.91789 | 0.02896 | 0.40208 | 0.8295 | 0.85222 | 0.6145369298 | 0.88573 | 0.9977293611 | 0.128624 | 5.21596178 | 0 |
| 2 | 47475201 | G | T | D | Y | MSH2 | ENST00000233146 | 0.92584 | 0.02796 | 0.41599 | 0.83814 | 0.94356 | 0.7871117467 | 0.94327 | 0.9992038608 | 0.422391 | 8.052839085 | 0 |
| 2 | 47475202 | A | C | D | A | MSH2 | ENST00000233146 | 0.50753 | 0.00928 | 0.33759 | 0.66344 | 0.81818 | 0.7221924462 | 0.75929 | 0.5923501253 | 0.00222951 | 4.407867522 | 0 |
| 2 | 47475202 | A | G | D | G | MSH2 | ENST00000233146 | 0.81434 | 0.02307 | 0.35501 | 0.84506 | 0.93397 | 0.7027041877 | 0.94354 | 0.9965617061 | 0.322016 | 4.551404222 | 0 |
| 2 | 47475202 | A | T | D | V | MSH2 | ENST00000233146 | 0.80756 | 0.02469 | 0.33858 | 0.85701 | 0.94469 | 0.7779665955 | 0.95774 | 0.9974478483 | 0.37775 | 5.773898689 | 0 |
| 2 | 47475203 | T | A | D | E | MSH2 | ENST00000233146 | 0.41375 | 0.00855 | 0.37809 | 0.68433 | 0.84321 | 0.5274668928 | 0.88107 | 0.7516719699 | 0.00311692 | 3.961856299 | 0 |
| 2 | 47475203 | T | G | D | E | MSH2 | ENST00000233146 | 0.41979 | 0.00855 | 0.37809 | 0.68401 | 0.84321 | 0.5274668928 | 0.88024 | 0.9345520139 | 0.120877 | 3.961856299 | 0 |

| | | | | | | | | | | | | | | | | | | |
|---|---|---|---|---|---|---|---|---|---|---|---|---|---|---|---|---|---|---|
| 2 | 47475204 | G | A | E | K | MSH2 | ENST00000233146 | 0.61798 | 0.00777 | 0.41298 | 0.71977 | 0.81876 | 0.5522550441 | 0.97514 | 0.9777937531 | 0.374708 | 3.314541277 | 0 |
| 2 | 47475204 | G | C | E | Q | MSH2 | ENST00000233146 | 0.57259 | 0.01459 | 0.29609 | 0.69081 | 0.78223 | 0.5347333857 | 0.89613 | 0.6056358814 | 0.136147 | 3.681122957 | 0 |
| 2 | 47475205 | A | C | E | A | MSH2 | ENST00000233146 | 0.47949 | 0.00791 | 0.31126 | 0.70939 | 0.80178 | 0.5745150537 | 0.89683 | 0.981349647 | 0.22462 | 3.594143644 | 0 |
| 2 | 47475205 | A | G | E | G | MSH2 | ENST00000233146 | 0.36716 | 0.00547 | 0.2793 | 0.65847 | 0.74797 | 0.544490866 | 0.77559 | 0.946498096 | 0.18523 | 2.410113884 | 0 |
| 2 | 47475205 | A | T | E | V | MSH2 | ENST00000233146 | 0.52919 | 0.01591 | 0.29056 | 0.8173 | 0.90972 | 0.6361954895 | 0.93581 | 0.9956197143 | 0.269816 | 4.882815887 | 0 |
| 2 | 47475206 | A | C | E | D | MSH2 | ENST00000233146 | 0.19402 | 0.00586 | 0.30946 | 0.29446 | 0.7562 | 0.3931710349 | 0.92412 | 0.4456298649 | 0.0291281 | -0.3831611384 | 0 |
| 2 | 47475206 | A | T | E | D | MSH2 | ENST00000233146 | 0.19087 | 0.00586 | 0.30946 | 0.29446 | 0.7562 | 0.3931710349 | 0.92349 | 0.4456298649 | 0.0291309 | -0.3831611384 | 0 |
| 2 | 47475207 | A | C | I | L | MSH2 | ENST00000233146 | 0.34893 | 0.00586 | 0.06388 | 0.65651 | 0.68023 | 0.453884331 | 0.83011 | 0.664714694 | 0.0627997 | 4.811338542 | 0 |
| 2 | 47475207 | A | G | I | V | MSH2 | ENST00000233146 | 0.15835 | 0.00603 | 0.03967 | 0.61577 | 0.55468 | 0.3392253622 | 0.54631 | 0.04209226836 | -0.0713934 | -3.039491967 | 0 |
| 2 | 47475207 | A | T | I | F | MSH2 | ENST00000233146 | 0.49061 | 0.01184 | 0.07346 | 0.82628 | 0.78286 | 0.6088514068 | 0.78523 | 0.9903646111 | 0.104878 | 3.785418741 | 0 |
| 2 | 47475208 | T | C | I | T | MSH2 | ENST00000233146 | 0.29104 | 0.00543 | 0.11365 | 0.52535 | 0.70427 | 0.4840871015 | 0.64387 | 0.7817421556 | 0.0587327 | 0.4243701365 | 0 |
| 2 | 47475208 | T | G | I | S | MSH2 | ENST00000233146 | 0.45021 | 0.00754 | 0.0698 | 0.6917 | 0.73296 | 0.6429613778 | 0.82678 | 0.9651299119 | 0.126475 | 1.388551031 | 0 |
| 2 | 47475209 | T | G | I | M | MSH2 | ENST00000233146 | 0.35437 | 0.01848 | 0.06604 | 0.76234 | 0.80653 | 0.4828394182 | 0.86452 | 0.6557116508 | 0.0462914 | 3.414972655 | 0 |
| 2 | 47475210 | G | A | A | T | MSH2 | ENST00000233146 | 0.48844 | 0.02186 | 0.3162 | 0.69903 | 0.81065 | 0.6516468381 | 0.76515 | 0.9285941124 | 0.114163 | 4.756354262 | 0 |
| 2 | 47475210 | G | C | A | P | MSH2 | ENST00000233146 | 0.5508 | 0.03001 | 0.36419 | 0.80399 | 0.86703 | 0.8683940244 | 0.8774 | 0.9932308197 | 0.277865 | 4.210192498 | 0 |
| 2 | 47475210 | G | T | A | S | MSH2 | ENST00000233146 | 0.2895 | 0.00696 | 0.28281 | 0.65137 | 0.78223 | 0.5935859121 | 0.71771 | 0.3171975315 | 0.0787699 | 2.677362142 | 0 |
| 2 | 47475211 | C | A | A | E | MSH2 | ENST00000233146 | 0.30432 | 0.02037 | 0.36524 | 0.74745 | 0.81006 | 0.8172393411 | 0.77401 | 0.9084615111 | 0.225327 | 4.39628433 | 0 |
| 2 | 47475211 | C | G | A | G | MSH2 | ENST00000233146 | 0.23069 | 0.00718 | 0.30121 | 0.63134 | 0.67108 | 0.6374430764 | 0.72274 | 0.9007906318 | 0.0903674 | 4.553135908 | 0 |
| 2 | 47475211 | C | T | A | V | MSH2 | ENST00000233146 | 0.36431 | 0.03019 | 0.32273 | 0.72272 | 0.79998 | 0.7189581535 | 0.76774 | 0.9605284333 | 0.202372 | 5.071437073 | 0 |
| 2 | 47475213 | T | A | F | I | MSH2 | ENST00000233146 | 0.76251 | 0.0315 | 0.51571 | 0.90721 | 0.98585 | 0.9425961727 | 0.96536 | 0.9983121157 | 0.473791 | 9.920611652 | 0 |
| 2 | 47475213 | T | C | F | L | MSH2 | ENST00000233146 | 0.73677 | 0.021 | 0.55717 | 0.88348 | 0.9506 | 0.9182175996 | 0.90455 | 0.99402529 | 0.27931 | 9.920611652 | 0 |
| 2 | 47475213 | T | G | F | V | MSH2 | ENST00000233146 | 0.7524 | 0.03237 | 0.48909 | 0.91473 | 0.96664 | 0.936636069 | 0.95259 | 0.9967706203 | 0.460776 | 9.920611652 | 0 |
| 2 | 47475214 | T | A | F | Y | MSH2 | ENST00000233146 | 0.65586 | 0.03491 | 0.53422 | 0.80636 | 0.90929 | 0.87724577 | 0.86286 | 0.9279463291 | 0.242933 | 4.453798554 | 0 |
| 2 | 47475214 | T | C | F | S | MSH2 | ENST00000233146 | 0.9276 | 0.02844 | 0.51567 | 0.92231 | 0.99296 | 0.95725862 | 0.95648 | 0.9998127818 | 0.482981 | 9.920611652 | 0 |
| 2 | 47475214 | T | G | F | C | MSH2 | ENST00000233146 | 0.92736 | 0.03166 | 0.54771 | 0.91944 | 0.98513 | 0.9582765336 | 0.9404 | 0.9994462132 | 0.484306 | 9.920611652 | 0 |
| 2 | 47475215 | T | A | F | L | MSH2 | ENST00000233146 | 0.6938 | 0.021 | 0.55717 | 0.87702 | 0.91484 | 0.9182175996 | 0.92607 | 0.9862404466 | 0.247807 | 9.920611652 | 0 |
| 2 | 47475215 | T | G | F | L | MSH2 | ENST00000233146 | 0.70206 | 0.021 | 0.55717 | 0.8757 | 0.91484 | 0.9182175996 | 0.92632 | 0.9862404466 | 0.247805 | 9.920611652 | 0 |
| 2 | 47475216 | A | C | I | L | MSH2 | ENST00000233146 | 0.76235 | 0.03718 | 0.59115 | 0.90594 | 0.9528 | 0.8383470457 | 0.92598 | 0.998462677 | 0.380298 | 9.719005091 | 0 |
| 2 | 47475216 | A | G | I | V | MSH2 | ENST00000233146 | 0.6946 | 0.03967 | 0.50129 | 0.76164 | 0.85378 | 0.7137836735 | 0.88069 | 0.9546818733 | 0.267293 | 4.150789582 | 0 |
| 2 | 47475216 | A | T | I | F | MSH2 | ENST00000233146 | 0.7783 | 0.03039 | 0.60391 | 0.90599 | 0.98367 | 0.9499322231 | 0.97009 | 0.999532938 | 0.485362 | 9.719005091 | 0 |
| 2 | 47475217 | T | A | I | N | MSH2 | ENST00000233146 | 0.83699 | 0.02864 | 0.68456 | 0.92672 | 0.98146 | 0.9718376837 | 0.90724 | 0.9998053908 | 0.431612 | 9.719005091 | 0 |
| 2 | 47475217 | T | C | I | T | MSH2 | ENST00000233146 | 0.78201 | 0.0337 | 0.70944 | 0.9095 | 0.98585 | 0.9455932869 | 0.96962 | 0.9982904792 | 0.447621 | 6.87724545 | 0 |
| 2 | 47475217 | T | G | I | S | MSH2 | ENST00000233146 | 0.84567 | 0.03155 | 0.61746 | 0.92325 | 0.99265 | 0.9807783488 | 0.96046 | 0.9998046756 | 0.477895 | 9.719005091 | 0 |
| 2 | 47475218 | T | G | I | M | MSH2 | ENST00000233146 | 0.66422 | 0.03697 | 0.55747 | 0.89726 | 0.92804 | 0.8901884354 | 0.95473 | 0.9949538112 | 0.421603 | 9.719005091 | 0 |
| 2 | 47475219 | C | A | P | T | MSH2 | ENST00000233146 | 0.52952 | 0.03413 | 0.57277 | 0.78212 | 0.94129 | 0.8920393726 | 0.96397 | 0.9972826242 | 0.411897 | 6.453909967 | 0 |
| 2 | 47475219 | C | G | P | A | MSH2 | ENST00000233146 | 0.31613 | 0.01591 | 0.5241 | 0.74831 | 0.89239 | 0.8571460914 | 0.93083 | 0.9652509689 | 0.36548 | 3.873912549 | 0 |
| 2 | 47475219 | C | T | P | S | MSH2 | ENST00000233146 | 0.67485 | 0.02702 | 0.53951 | 0.78485 | 0.94318 | 0.8885246052 | 0.92031 | 0.9895114303 | 0.415454 | 3.63431917 | 0 |
| 2 | 47475220 | C | A | P | H | MSH2 | ENST00000233146 | 0.81442 | 0.03484 | 0.59852 | 0.90074 | 0.9648 | 0.9157011909 | 0.91384 | 0.9993603826 | 0.463498 | 9.170947599 | 0 |
| 2 | 47475220 | C | G | P | R | MSH2 | ENST00000233146 | 0.81803 | 0.03576 | 0.60866 | 0.91976 | 0.96773 | 0.9447123538 | 0.94757 | 0.9989871383 | 0.453107 | 7.707386673 | 0 |
| 2 | 47475220 | C | T | P | L | MSH2 | ENST00000233146 | 0.83521 | 0.03613 | 0.6155 | 0.87781 | 0.9648 | 0.9314291582 | 0.95286 | 0.9995738864 | 0.440823 | 9.170947599 | 0 |
| 2 | 47475222 | A | C | N | H | MSH2 | ENST00000233146 | 0.75758 | 0.03674 | 0.41606 | 0.89431 | 0.97698 | 0.9409756662 | 0.96486 | 0.9991894364 | 0.310606 | 9.999859679 | 0 |
| 2 | 47475222 | A | T | N | Y | MSH2 | ENST00000233146 | 0.80519 | 0.03419 | 0.45788 | 0.90746 | 0.98799 | 0.9587394193 | 0.96594 | 0.9998608828 | 0.520348 | 9.999859679 | 0 |
| 2 | 47475223 | A | C | N | T | MSH2 | ENST00000233146 | 0.78233 | 0.03692 | 0.38735 | 0.90093 | 0.97438 | 0.9150541589 | 0.94469 | 0.9995968938 | 0.333672 | 9.999859679 | 0 |
| 2 | 47475223 | A | G | N | S | MSH2 | ENST00000233146 | 0.73277 | 0.04051 | 0.32994 | 0.90522 | 0.96298 | 0.8992395442 | 0.95823 | 0.9995119572 | 0.241042 | 9.999859679 | 0 |
| 2 | 47475223 | A | T | N | I | MSH2 | ENST00000233146 | 0.82475 | 0.03388 | 0.42037 | 0.91622 | 0.97735 | 0.9498824408 | 0.92777 | 0.999868989 | 0.389982 | 9.999859679 | 0 |
| 2 | 47475224 | T | A | N | K | MSH2 | ENST00000233146 | 0.58133 | 0.03509 | 0.50198 | 0.95057 | 0.95681 | 0.9636143931 | 0.96351 | 0.9995852113 | 0.204293 | 9.999859679 | 0 |
| 2 | 47475224 | T | G | N | K | MSH2 | ENST00000233146 | 0.58867 | 0.03509 | 0.50198 | 0.94747 | 0.95681 | 0.9636143931 | 0.96338 | 0.9992573857 | 0.204295 | 9.999859679 | 0 |
| 2 | 47475225 | G | A | D | N | MSH2 | ENST00000233146 | 0.87315 | 0.02249 | 0.54201 | 0.80168 | 0.90185 | 0.9106661289 | 0.92483 | 0.9969287515 | 0.140207 | 3.229957911 | 0 |
| 2 | 47475225 | G | C | D | H | MSH2 | ENST00000233146 | 0.87302 | 0.03442 | 0.57167 | 0.84367 | 0.97252 | 0.9242811238 | 0.96732 | 0.9992499948 | 0.485391 | 5.989605539 | 0 |
| 2 | 47475225 | G | T | D | Y | MSH2 | ENST00000233146 | 0.8799 | 0.03394 | 0.5513 | 0.84678 | 0.9703 | 0.9555666281 | 0.98448 | 0.9998949766 | 0.492566 | 8.328695147 | 0 |
| 2 | 47475226 | A | C | D | A | MSH2 | ENST00000233146 | 0.91319 | 0.03334 | 0.47376 | 0.91712 | 0.98693 | 0.9400282879 | 0.93883 | 0.9995552897 | 0.482813 | 7.540908331 | 0 |
| 2 | 47475226 | A | G | D | G | MSH2 | ENST00000233146 | 0.91729 | 0.03197 | 0.49284 | 0.86531 | 0.9796 | 0.940914633 | 0.96119 | 0.9984832406 | 0.471847 | 5.472705603 | 0 |
| 2 | 47475226 | A | T | D | V | MSH2 | ENST00000233146 | 0.90896 | 0.03166 | 0.46691 | 0.89028 | 0.98938 | 0.9519751698 | 0.97764 | 0.9997119308 | 0.491241 | 8.328695147 | 0 |
| 2 | 47475227 | C | A | D | E | MSH2 | ENST00000233146 | 0.53596 | 0.03144 | 0.51146 | 0.88761 | 0.89739 | 0.8780905816 | 0.93446 | 0.9968342185 | 0.244694 | 6.154713053 | 0 |
| 2 | 47475227 | C | G | D | E | MSH2 | ENST00000233146 | 0.54311 | 0.03144 | 0.51146 | 0.88249 | 0.89739 | 0.8780905816 | 0.93448 | 0.9968342185 | 0.244692 | 6.154713053 | 0 |
| 2 | 47475228 | G | A | V | I | MSH2 | ENST00000233146 | 0.20386 | 0.00659 | 0.08069 | 0.44469 | 0.68734 | 0.3892337722 | 0.78862 | 0.02519593873 | -0.214172 | 2.056162321 | 0 |
| 2 | 47475228 | G | C | V | L | MSH2 | ENST00000233146 | 0.34707 | 0.00799 | 0.1464 | 0.75039 | 0.86852 | 0.6905296568 | 0.93178 | 0.7689136267 | 0.158446 | 4.029314497 | 0 |
| 2 | 47475228 | G | T | V | L | MSH2 | ENST00000233146 | 0.34261 | 0.00799 | 0.1464 | 0.75014 | 0.86852 | 0.6905296568 | 0.93291 | 0.7689136267 | 0.158443 | 4.029314497 | 0 |
| 2 | 47475229 | T | A | V | E | MSH2 | ENST00000233146 | 0.72459 | 0.02262 | 0.16391 | 0.86907 | 0.97104 | 0.9374278346 | 0.94924 | 0.9989930987 | 0.465083 | 7.950084373 | 1 |
| 2 | 47475229 | T | C | V | A | MSH2 | ENST00000233146 | 0.3915 | 0.00615 | 0.17278 | 0.60293 | 0.86703 | 0.7699386057 | 0.89291 | 0.8414405584 | 0.239171 | 2.362963569 | 0 |
| 2 | 47475229 | T | G | V | G | MSH2 | ENST00000233146 | 0.70951 | 0.02049 | 0.10672 | 0.85288 | 0.96117 | 0.940149776 | 0.93658 | 0.9964878559 | 0.464589 | 7.950084373 | 0 |
| 2 | 47475231 | T | A | Y | N | MSH2 | ENST00000233146 | 0.20986 | 0.00492 | 0.18177 | 0.44024 | 0.5046 | 0.5131533072 | 0.55218 | 0.2087144107 | -0.0335279 | 0.8038514265 | 0 |
| 2 | 47475231 | T | G | Y | D | MSH2 | ENST00000233146 | 0.20463 | 0.00476 | 0.18499 | 0.44572 | 0.68292 | 0.5816746421 | 0.73392 | 0.1927789748 | -0.00448719 | 0.856954098 | 0 |
| 2 | 47475232 | A | C | Y | S | MSH2 | ENST00000233146 | 0.1737 | 0.00512 | 0.13694 | 0.43441 | 0.39643 | 0.5125774851 | 0.5863 | 0.1744730026 | -0.0293279 | -0.5045597708 | 0 |
| 2 | 47475232 | A | G | Y | C | MSH2 | ENST00000233146 | 0.24445 | 0.00535 | 0.14704 | 0.60351 | 0.69846 | 0.5665496808 | 0.90552 | 0.1054821163 | -0.00688542 | 1.971359609 | 0 |
| 2 | 47475232 | A | T | Y | F | MSH2 | ENST00000233146 | 0.19083 | 0.00664 | 0.15819 | 0.51367 | 0.41179 | 0.3857018186 | 0.83678 | 0.1801770478 | 0.0207657 | 0.6980737195 | 0 |
| 2 | 47475234 | T | A | F | I | MSH2 | ENST00000233146 | 0.86375 | 0.02875 | 0.63415 | 0.82493 | 0.9328 | 0.8190069498 | 0.94208 | 0.9796685576 | 0.384733 | 4.668059512 | 0 |
| 2 | 47475234 | T | C | F | L | MSH2 | ENST00000233146 | 0.65194 | 0.01108 | 0.66391 | 0.74143 | 0.82948 | 0.7734310502 | 0.95114 | 0.726454854 | 0.351873 | 2.963915209 | 0 |
| 2 | 47475234 | T | G | F | V | MSH2 | ENST00000233146 | 0.86983 | 0.0296 | 0.60317 | 0.8818 | 0.95243 | 0.8327690744 | 0.95738 | 0.98962152 | 0.402181 | 5.622340937 | 0 |
| 2 | 47475235 | T | A | F | Y | MSH2 | ENST00000233146 | 0.85367 | 0.03461 | 0.65112 | 0.87743 | 0.92069 | 0.7933952494 | 0.8685 | 0.9779528975 | 0.277604 | 8.733673796 | 0 |
| 2 | 47475235 | T | C | F | S | MSH2 | ENST00000233146 | 0.91852 | 0.02815 | 0.63157 | 0.91483 | 0.98693 | 0.9294224706 | 0.96302 | 0.9943490624 | 0.421436 | 8.733673796 | 0 |
| 2 | 47475235 | T | G | F | C | MSH2 | ENST00000233146 | 0.91831 | 0.0315 | 0.66453 | 0.91 | 0.97885 | 0.9001894702 | 0.93059 | 0.9943490624 | 0.424342 | 8.733673796 | 0 |
| 2 | 47475236 | T | A | F | L | MSH2 | ENST00000233146 | 0.41016 | 0.01108 | 0.66391 | 0.82369 | 0.79878 | 0.7734310502 | 0.95318 | 0.330094099 | 0.308925 | 2.963915209 | 0 |
| 2 | 47475236 | T | G | F | L | MSH2 | ENST00000233146 | 0.41623 | 0.01108 | 0.66391 | 0.82254 | 0.79878 | 0.7734310502 | 0.9527 | 0.4246459901 | 0.308929 | 2.963915209 | 0 |
| 2 | 47475237 | G | C | E | Q | MSH2 | ENST00000233146 | 0.66193 | 0.01221 | 0.37423 | 0.63606 | 0.79334 | 0.5424263676 | 0.88993 | 0.8588995934 | 0.171234 | 4.512546466 | 0 |
| 2 | 47475238 | A | C | E | A | MSH2 | ENST00000233146 | 0.40021 | 0.00574 | 0.38898 | 0.47597 | 0.59516 | 0.5930354288 | 0.80156 | 0.6981802583 | 0.0883025 | 4.446879511 | 0 |
| 2 | 47475238 | A | G | E | G | MSH2 | ENST00000233146 | 0.49692 | 0.00705 | 0.34674 | 0.59855 | 0.74797 | 0.6895779682 | 0.82684 | 0.9322408438 | 0.167221 | 3.961662607 | 0 |
| 2 | 47475238 | A | T | E | V | MSH2 | ENST00000233146 | 0.45709 | 0.00556 | 0.36105 | 0.53345 | 0.5927 | 0.6128076322 | 0.84605 | 0.6498541832 | 0.0894505 | 5.353224011 | 0 |
| 2 | 47475239 | A | C | E | D | MSH2 | ENST00000233146 | 0.33142 | 0.00596 | 0.38148 | 0.34004 | 0.74306 | 0.5347538908 | 0.93403 | 0.2238001972 | 0.0651284 | 1.850034261 | 0 |
| 2 | 47475239 | A | T | E | D | MSH2 | ENST00000233146 | 0.32696 | 0.00596 | 0.38148 | 0.36344 | 0.74235 | 0.5347538908 | 0.93403 | 0.2238001972 | 0.0651521 | 1.850034261 | 0 |
| 2 | 47475240 | A | C | K | Q | MSH2 | ENST00000233146 | 0.71522 | 0.01396 | 0.34043 | 0.56132 | 0.71705 | 0.6289139587 | 0.84988 | 0.8043689132 | 0.103132 | 3.119793797 | 0 |
| 2 | 47475240 | A | G | K | E | MSH2 | ENST00000233146 | 0.90218 | 0.02063 | 0.37787 | 0.6844 | 0.82613 | 0.7302046544 | 0.8939 | 0.9701240063 | 0.211243 | 4.115723716 | 0 |
| 2 | 47475241 | A | C | K | T | MSH2 | ENST00000233146 | 0.80148 | 0.02556 | 0.38892 | 0.74924 | 0.86453 | 0.7249406764 | 0.86594 | 0.9862232804 | 0.189552 | 4.37813849 | 0 |
| 2 | 47475241 | A | G | K | R | MSH2 | ENST00000233146 | 0.41848 | 0.00681 | 0.31891 | 0.49502 | 0.73516 | 0.604618659 | 0.90448 | 0.2276680829 | 0.0165155 | 1.985699528 | 0 |
| 2 | 47475241 | A | T | K | I | MSH2 | ENST00000233146 | 0.85952 | 0.02696 | 0.35747 | 0.81541 | 0.88307 | 0.7621356897 | 0.82548 | 0.9963743091 | 0.217693 | 6.352888607 | 0 |
| 2 | 47475242 | A | C | K | N | MSH2 | ENST00000233146 | 0.79698 | 0.00836 | 0.40579 | 0.75813 | 0.82668 | 0.734332577 | 0.86934 | 0.9517382979 | 0.100982 | 3.87440945 | 0 |
| 2 | 47475242 | A | T | K | N | MSH2 | ENST00000233146 | 0.79034 | 0.00836 | 0.40579 | 0.75779 | 0.82668 | 0.734332577 | 0.86929 | 0.9702650309 | 0.100981 | 3.87440945 | 0 |
| 2 | 47475243 | G | A | D | N | MSH2 | ENST00000233146 | 0.40781 | 0.00596 | 0.222 | 0.64626 | 0.54344 | 0.5751012729 | 0.77789 | 0.6437517405 | 0.0414454 | 3.049836983 | 0 |
| 2 | 47475243 | G | C | D | H | MSH2 | ENST00000233146 | 0.7157 | 0.02294 | 0.23514 | 0.90447 | 0.87394 | 0.6830633049 | 0.87136 | 0.9838529825 | 0.176478 | 4.572481966 | 0 |
| 2 | 47475243 | G | T | D | Y | MSH2 | ENST00000233146 | 0.73365 | 0.02708 | 0.23641 | 0.92722 | 0.90624 | 0.7782807128 | 0.85792 | 0.9922351241 | 0.214119 | 6.320961512 | 0 |
| 2 | 47475244 | A | C | D | A | MSH2 | ENST00000233146 | 0.45868 | 0.00935 | 0.19017 | 0.79299 | 0.78844 | 0.7087165112 | 0.85076 | 0.9696608782 | 0.117378 | 2.693752541 | 0 |
| 2 | 47475244 | A | T | D | V | MSH2 | ENST00000233146 | 0.70976 | 0.01662 | 0.1824 | 0.9106 | 0.89739 | 0.7858377057 | 0.86506 | 0.9890220165 | 0.192966 | 5.425886867 | 0 |
| 2 | 47475245 | T | A | D | E | MSH2 | ENST00000233146 | 0.34065 | 0.00596 | 0.20363 | 0.34674 | 0.56695 | 0.5756190913 | 0.8098 | 0.1143977728 | 0.0284618 | 1.947816416 | 0 |
| 2 | 47475245 | T | G | D | E | MSH2 | ENST00000233146 | 0.3462 | 0.00596 | 0.20363 | 0.33698 | 0.56695 | 0.5756190913 | 0.8112 | 0.1722867191 | 0.0284688 | 1.947816416 | 0 |
| 2 | 47475246 | A | C | K | Q | MSH2 | ENST00000233146 | 0.47778 | 0.00705 | 0.28906 | 0.41279 | 0.65575 | 0.5734948129 | 0.85301 | 0.4278840125 | 0.0840956 | 3.897923269 | 0 |
| 2 | 47475246 | A | G | K | E | MSH2 | ENST00000233146 | 0.40541 | 0.00517 | 0.32498 | 0.33698 | 0.66639 | 0.6563409017 | 0.82241 | 0.6923207641 | 0.0708899 | 1.809316493 | 0 |
| 2 | 47475247 | A | C | K | T | MSH2 | ENST00000233146 | 0.51395 | 0.0063 | 0.34523 | 0.5775 | 0.78286 | 0.6842237762 | 0.87332 | 0.6985955238 | 0.176994 | 3.466201035 | 0 |
| 2 | 47475247 | A | G | K | R | MSH2 | ENST00000233146 | 0.50694 | 0.00659 | 0.26126 | 0.42488 | 0.70914 | 0.5117654899 | 0.91736 | 0.6383520365 | 0.100741 | 3.85995388 | 0 |
| 2 | 47475247 | A | T | K | I | MSH2 | ENST00000233146 | 0.86594 | 0.01334 | 0.30339 | 0.79219 | 0.90362 | 0.7025175489 | 0.8348 | 0.987940073 | 0.212264 | 6.371986179 | 0 |
| 2 | 47475248 | A | C | K | N | MSH2 | ENST00000233146 | 0.63298 | 0.01044 | 0.35938 | 0.67358 | 0.79697 | 0.654467201 | 0.84807 | 0.770940423 | 0.0749368 | 4.028340263 | 0 |
| 2 | 47475248 | A | T | K | N | MSH2 | ENST00000233146 | 0.62657 | 0.01044 | 0.35938 | 0.67358 | 0.79697 | 0.654467201 | 0.84622 | 0.770940423 | 0.0749576 | 4.028340263 | 0 |

| | | | | | | | | | | | | | | | | | | |
|---|---|---|---|---|---|---|---|---|---|---|---|---|---|---|---|---|---|---|
| 2 | 47475249 | C | A | Q | K | MSH2 | ENST00000233146 | 0.36587 | 0.00535 | 0.28586 | 0.34324 | 0.63525 | 0.6243472633 | 0.87384 | 0.6223836541 | 0.0827812 | 2.46698356 | 0 |
| 2 | 47475249 | C | G | Q | E | MSH2 | ENST00000233146 | 0.34964 | 0.00586 | 0.24937 | 0.42377 | 0.63525 | 0.6692878224 | 0.88345 | 0.4796945711 | 0.106928 | 2.997004942 | 0 |
| 2 | 47475250 | A | C | Q | P | MSH2 | ENST00000233146 | 0.61261 | 0.01871 | 0.25581 | 0.54712 | 0.90274 | 0.8539158198 | 0.9344 | 0.9635486007 | 0.316428 | 7.784357551 | 0 |
| 2 | 47475250 | A | G | Q | R | MSH2 | ENST00000233146 | 0.43261 | 0.00586 | 0.28154 | 0.35746 | 0.74517 | 0.6361855882 | 0.88851 | 0.8465899825 | 0.149484 | 3.427520085 | 0 |
| 2 | 47475250 | A | T | Q | L | MSH2 | ENST00000233146 | 0.5016 | 0.0114 | 0.23563 | 0.5759 | 0.86753 | 0.6537435332 | 0.89748 | 0.9555752277 | 0.229584 | 5.487984284 | 0 |
| 2 | 47475251 | G | C | Q | H | MSH2 | ENST00000233146 | 0.39405 | 0.00638 | 0.28746 | 0.52343 | 0.77141 | 0.690641216 | 0.92358 | 0.9277459979 | 0.179262 | 2.758098973 | 0 |
| 2 | 47475251 | G | T | Q | H | MSH2 | ENST00000233146 | 0.38937 | 0.00638 | 0.28746 | 0.52351 | 0.77141 | 0.690641216 | 0.92365 | 0.9277459979 | 0.179262 | 2.758098973 | 0 |
| 2 | 47475252 | A | C | M | L | MSH2 | ENST00000233146 | 0.48614 | 0.00763 | 0.35702 | 0.43722 | 0.73148 | 0.5049317242 | 0.85852 | 0.8660898209 | 0.100007 | 5.236290729 | 0 |
| 2 | 47475252 | A | G | M | V | MSH2 | ENST00000233146 | 0.45438 | 0.00741 | 0.30191 | 0.47914 | 0.74164 | 0.5386133609 | 0.84319 | 0.3078678846 | -0.0203319 | 6.959577504 | 0 |
| 2 | 47475252 | A | T | M | L | MSH2 | ENST00000233146 | 0.48075 | 0.00763 | 0.35702 | 0.43511 | 0.73148 | 0.5049317242 | 0.85901 | 0.4006976434 | 0.100006 | 5.236290729 | 0 |
| 2 | 47475253 | T | A | M | K | MSH2 | ENST00000233146 | 0.40559 | 0.00638 | 0.43826 | 0.51291 | 0.7872 | 0.6953214535 | 0.92718 | 0.6868884563 | 0.180435 | 4.577299494 | 0 |
| 2 | 47475253 | T | C | M | T | MSH2 | ENST00000233146 | 0.37002 | 0.00547 | 0.48711 | 0.47134 | 0.68113 | 0.5411070612 | 0.80724 | 0.290361762 | 0.0627587 | 3.29405729 | 0 |
| 2 | 47475253 | T | G | M | R | MSH2 | ENST00000233146 | 0.45795 | 0.00517 | 0.41103 | 0.48845 | 0.78473 | 0.7450442798 | 0.93837 | 0.6670140028 | 0.20885 | 3.794133431 | 0 |
| 2 | 47475254 | G | A | M | I | MSH2 | ENST00000233146 | 0.58133 | 0.00808 | 0.36299 | 0.51029 | 0.70994 | 0.516995894 | 0.79396 | 0.3643713758 | 0.119749 | 5.93994378 | 0 |
| 2 | 47475254 | G | C | M | I | MSH2 | ENST00000233146 | 0.57843 | 0.00808 | 0.36299 | 0.50995 | 0.70994 | 0.516995894 | 0.80185 | 0.8040552735 | 0.119707 | 5.93994378 | 0 |
| 2 | 47475254 | G | T | M | I | MSH2 | ENST00000233146 | 0.57266 | 0.00808 | 0.36299 | 0.50995 | 0.70994 | 0.516995894 | 0.79993 | 0.3628569223 | 0.119729 | 5.93994378 | 0 |
| 2 | 47475255 | T | A | F | I | MSH2 | ENST00000233146 | 0.91157 | 0.03039 | 0.76829 | 0.80568 | 0.91442 | 0.8763169688 | 0.91933 | 0.9970934391 | 0.372389 | 4.143593416 | 0 |
| 2 | 47475255 | T | C | F | L | MSH2 | ENST00000233146 | 0.779 | 0.01799 | 0.79741 | 0.61492 | 0.91015 | 0.8472270769 | 0.92151 | 0.9839885831 | 0.367172 | 4.838877461 | 0 |
| 2 | 47475255 | T | G | F | V | MSH2 | ENST00000233146 | 0.91622 | 0.03124 | 0.74684 | 0.82379 | 0.93824 | 0.8677197257 | 0.93264 | 0.9955073595 | 0.410071 | 4.626222044 | 0 |
| 2 | 47475256 | T | A | F | Y | MSH2 | ENST00000233146 | 0.90175 | 0.03447 | 0.78009 | 0.86294 | 0.93901 | 0.8046722531 | 0.88031 | 0.9966889024 | 0.318383 | 9.560118775 | 0 |
| 2 | 47475256 | T | C | F | S | MSH2 | ENST00000233146 | 0.94145 | 0.02702 | 0.76222 | 0.90363 | 0.98257 | 0.93352858 | 0.94734 | 0.9989054203 | 0.449145 | 8.621626159 | 0 |
| 2 | 47475256 | T | G | F | C | MSH2 | ENST00000233146 | 0.94145 | 0.03144 | 0.78408 | 0.87323 | 0.97735 | 0.9199699775 | 0.94225 | 0.9977161884 | 0.442877 | 6.589087617 | 0 |
| 2 | 47475257 | C | A | F | L | MSH2 | ENST00000233146 | 0.48883 | 0.01799 | 0.79741 | 0.66162 | 0.8306 | 0.8472270769 | 0.93658 | 0.9722366929 | 0.330232 | 4.838877461 | 0 |
| 2 | 47475257 | C | G | F | L | MSH2 | ENST00000233146 | 0.49563 | 0.01799 | 0.79741 | 0.64622 | 0.8306 | 0.8472270769 | 0.93613 | 0.9667438865 | 0.330233 | 4.838877461 | 0 |
| 2 | 47475258 | C | A | H | N | MSH2 | ENST00000233146 | 0.42156 | 0.01538 | 0.73498 | 0.58716 | 0.81414 | 0.6960541827 | 0.77802 | 0.9765520096 | 0.16128 | 5.215803275 | 0 |
| 2 | 47475258 | C | G | H | D | MSH2 | ENST00000233146 | 0.60339 | 0.01621 | 0.73404 | 0.55511 | 0.87148 | 0.8348083167 | 0.84011 | 0.9857429266 | 0.323993 | 5.979442238 | 0 |
| 2 | 47475258 | C | T | H | Y | MSH2 | ENST00000233146 | 0.31524 | 0.00562 | 0.73659 | 0.35442 | 0.67661 | 0.6439654928 | 0.65796 | 0.8229641914 | 0.0974083 | 3.469709228 | 0 |
| 2 | 47475259 | A | C | H | P | MSH2 | ENST00000233146 | 0.7165 | 0.02037 | 0.65643 | 0.74789 | 0.94691 | 0.895623546 | 0.85561 | 0.9820654988 | 0.364155 | 7.944685316 | 0 |
| 2 | 47475259 | A | G | H | R | MSH2 | ENST00000233146 | 0.46918 | 0.01586 | 0.74029 | 0.66233 | 0.89009 | 0.7779636485 | 0.87788 | 0.9754577875 | 0.248273 | 7.944685316 | 0 |
| 2 | 47475259 | A | T | H | L | MSH2 | ENST00000233146 | 0.45217 | 0.00547 | 0.70132 | 0.5821 | 0.81124 | 0.7173691414 | 0.91922 | 0.863676846 | 0.235279 | 2.477455434 | 0 |
| 2 | 47475260 | C | A | H | Q | MSH2 | ENST00000233146 | 0.29357 | 0.00633 | 0.7059 | 0.49263 | 0.75957 | 0.6719320951 | 0.88638 | 0.8050234914 | 0.167263 | 4.476869387 | 0 |
| 2 | 47475260 | C | G | H | Q | MSH2 | ENST00000233146 | 0.29889 | 0.00633 | 0.7059 | 0.47766 | 0.75957 | 0.6719320951 | 0.88573 | 0.6385458112 | 0.167264 | 4.476869387 | 0 |
| 2 | 47475261 | A | C | I | L | MSH2 | ENST00000233146 | 0.85996 | 0.03334 | 0.75445 | 0.73916 | 0.87099 | 0.8345868372 | 0.88097 | 0.9503187537 | 0.260607 | 4.258151911 | 0 |
| 2 | 47475261 | A | G | I | V | MSH2 | ENST00000233146 | 0.77924 | 0.02683 | 0.66487 | 0.5875 | 0.8517 | 0.6912714887 | 0.85817 | 0.9176089168 | 0.231902 | 5.449539941 | 0 |
| 2 | 47475261 | A | T | I | F | MSH2 | ENST00000233146 | 0.87523 | 0.0254 | 0.76732 | 0.84191 | 0.9844 | 0.9546178038 | 0.95854 | 0.9990527034 | 0.483611 | 9.762655568 | 0 |
| 2 | 47475262 | T | C | I | T | MSH2 | ENST00000233146 | 0.92174 | 0.03334 | 0.85125 | 0.88845 | 0.98657 | 0.9599771461 | 0.94201 | 0.9995954633 | 0.481492 | 9.762655568 | 0 |
| 2 | 47475262 | T | G | I | S | MSH2 | ENST00000233146 | 0.94448 | 0.03129 | 0.77472 | 0.91323 | 0.99296 | 0.9868370393 | 0.95356 | 0.9996187687 | 0.483192 | 9.762655568 | 0 |
| 2 | 47475263 | C | G | I | M | MSH2 | ENST00000233146 | 0.69332 | 0.03667 | 0.72192 | 0.77787 | 0.87829 | 0.8712379909 | 0.90237 | 0.9905003905 | 0.378152 | 7.29543705 | 0 |
| 2 | 47475264 | A | C | I | L | MSH2 | ENST00000233146 | 0.84883 | 0.00935 | 0.72111 | 0.56907 | 0.90185 | 0.8150032556 | 0.84651 | 0.9058679342 | 0.234083 | 4.256308088 | 0 |
| 2 | 47475264 | A | G | I | V | MSH2 | ENST00000233146 | 0.60676 | 0.00648 | 0.62629 | 0.39111 | 0.81818 | 0.6525197013 | 0.76641 | 0.5535231281 | 0.129874 | 3.72973292 | 0 |
| 2 | 47475264 | A | T | I | F | MSH2 | ENST00000233146 | 0.86664 | 0.02381 | 0.72955 | 0.83255 | 0.96226 | 0.9469711202 | 0.97009 | 0.9986380935 | 0.469933 | 8.453881731 | 0 |
| 2 | 47475265 | T | A | I | N | MSH2 | ENST00000233146 | 0.93177 | 0.02663 | 0.81601 | 0.84754 | 0.98257 | 0.9792286392 | 0.91964 | 0.9994514585 | 0.463431 | 8.453881731 | 1 |
| 2 | 47475265 | T | C | I | T | MSH2 | ENST00000233146 | 0.90255 | 0.02414 | 0.83424 | 0.8001 | 0.98549 | 0.9530382248 | 0.96388 | 0.9987623692 | 0.489212 | 8.453881731 | 0 |
| 2 | 47475265 | T | G | I | S | MSH2 | ENST00000233146 | 0.93508 | 0.02614 | 0.74568 | 0.8553 | 0.99072 | 0.9843933968 | 0.95466 | 0.9996187687 | 0.496529 | 8.453881731 | 1 |
| 2 | 47475266 | T | G | I | M | MSH2 | ENST00000233146 | 0.77648 | 0.032 | 0.68198 | 0.87688 | 0.9528 | 0.8516745289 | 0.93828 | 0.996675849 | 0.422232 | 8.453881731 | 0 |
| 2 | 47475267 | A | C | T | P | MSH2 | ENST00000233146 | 0.85945 | 0.03293 | 0.72305 | 0.84583 | 0.99105 | 0.974017184 | 0.97022 | 0.9996298552 | 0.515813 | 9.647122977 | 1 |
| 2 | 47475267 | A | G | T | A | MSH2 | ENST00000233146 | 0.82404 | 0.03509 | 0.72014 | 0.82925 | 0.9917 | 0.9422628035 | 0.98103 | 0.9996644258 | 0.512562 | 9.647122977 | 0 |
| 2 | 47475267 | A | T | T | S | MSH2 | ENST00000233146 | 0.78574 | 0.03753 | 0.73398 | 0.84274 | 0.97698 | 0.8767066202 | 0.96508 | 0.9989708662 | 0.403331 | 8.479952956 | 0 |
| 2 | 47475268 | C | A | T | N | MSH2 | ENST00000233146 | 0.60618 | 0.03465 | 0.80129 | 0.80637 | 0.93319 | 0.9639020149 | 0.93938 | 0.999853611 | 0.294379 | 9.647122977 | 0 |
| 2 | 47475268 | C | G | T | S | MSH2 | ENST00000233146 | 0.60149 | 0.03753 | 0.73398 | 0.82995 | 0.94091 | 0.8767066202 | 0.95108 | 0.9993461967 | 0.393263 | 8.479952956 | 0 |
| 2 | 47475268 | C | T | T | I | MSH2 | ENST00000233146 | 0.64757 | 0.03394 | 0.84039 | 0.8276 | 0.97773 | 0.9566762843 | 0.97733 | 0.999853611 | 0.505043 | 9.647122977 | 0 |
| 2 | 47475270 | G | A | G | S | MSH2 | ENST00000233146 | 0.89818 | 0.03728 | 0.87765 | 0.94122 | 0.99953 | 0.9636520018 | 0.98932 | 0.999673605 | 0.594176 | 9.746731204 | 0 |
| 2 | 47475270 | G | C | G | R | MSH2 | ENST00000233146 | 0.91785 | 0.03452 | 0.93229 | 0.93332 | 0.99953 | 0.9806198166 | 0.98693 | 0.9998612404 | 0.594479 | 9.746731204 | 1 |
| 2 | 47475270 | G | T | G | C | MSH2 | ENST00000233146 | 0.93143 | 0.03626 | 0.90529 | 0.93512 | 0.99919 | 0.9721009583 | 0.98706 | 0.9998612404 | 0.594479 | 9.746731204 | 0 |
| 2 | 47476367 | G | A | G | D | MSH2 | ENST00000233146 | 0.87106 | 0.03327 | 0.93228 | 0.95768 | 0.99824 | 0.9829429488 | 0.979 | 0.9994976521 | 0.577047 | 9.746731204 | 1 |
| 2 | 47476367 | G | C | G | A | MSH2 | ENST00000233146 | 0.84393 | 0.03608 | 0.87764 | 0.95577 | 0.99386 | 0.9674396559 | 0.96679 | 0.9993927479 | 0.57678 | 8.503868944 | 0 |
| 2 | 47476369 | C | A | P | T | MSH2 | ENST00000233146 | 0.73981 | 0.03552 | 0.76298 | 0.90226 | 0.97848 | 0.9283084215 | 0.96661 | 0.9998764992 | 0.497157 | 9.999854852 | 0 |
| 2 | 47476369 | C | G | P | A | MSH2 | ENST00000233146 | 0.7185 | 0.03761 | 0.71644 | 0.87313 | 0.97252 | 0.9001750932 | 0.96425 | 0.999221921 | 0.481143 | 9.999854852 | 0 |
| 2 | 47476369 | C | T | P | S | MSH2 | ENST00000233146 | 0.77006 | 0.03706 | 0.73346 | 0.89952 | 0.98071 | 0.9328580821 | 0.96412 | 0.9998664856 | 0.496453 | 9.999854852 | 0 |
| 2 | 47476370 | C | A | P | H | MSH2 | ENST00000233146 | 0.69732 | 0.03503 | 0.78446 | 0.90504 | 0.96846 | 0.9395638873 | 0.94945 | 0.9998764992 | 0.498104 | 9.999854852 | 0 |
| 2 | 47476370 | C | G | P | R | MSH2 | ENST00000233146 | 0.7015 | 0.03633 | 0.79496 | 0.89362 | 0.97438 | 0.962929353 | 0.96861 | 0.9998764992 | 0.497062 | 9.999854852 | 0 |
| 2 | 47476370 | C | T | P | L | MSH2 | ENST00000233146 | 0.72019 | 0.03692 | 0.80641 | 0.90214 | 0.90624 | 0.947617987 | 0.98267 | 0.9999871254 | 0.452397 | 9.999854852 | 0 |
| 2 | 47476372 | A | G | N | D | MSH2 | ENST00000233146 | 0.86935 | 0.0337 | 0.7348 | 0.92602 | 0.97252 | 0.9606318487 | 0.96432 | 0.998073101 | 0.276389 | 9.86719115 | 0 |
| 2 | 47476372 | A | T | N | Y | MSH2 | ENST00000233146 | 0.8869 | 0.03388 | 0.7162 | 0.94598 | 0.98146 | 0.9782255433 | 0.97682 | 0.9997231364 | 0.505981 | 9.86719115 | 1 |
| 2 | 47476373 | A | C | N | T | MSH2 | ENST00000233146 | 0.85952 | 0.03542 | 0.64918 | 0.9332 | 0.97104 | 0.9574478753 | 0.93139 | 0.998989284 | 0.326287 | 9.86719115 | 1 |
| 2 | 47476373 | A | G | N | S | MSH2 | ENST00000233146 | 0.79974 | 0.02403 | 0.60017 | 0.93832 | 0.92561 | 0.9430282665 | 0.92944 | 0.9961330891 | 0.204839 | 9.645005311 | 0 |
| 2 | 47476373 | A | T | N | I | MSH2 | ENST00000233146 | 0.89503 | 0.03353 | 0.67673 | 0.94253 | 0.97178 | 0.9780423334 | 0.93553 | 0.9996765852 | 0.373677 | 9.86719115 | 1 |
| 2 | 47476374 | T | A | N | K | MSH2 | ENST00000233146 | 0.62068 | 0.0337 | 0.75926 | 0.95646 | 0.94129 | 0.9802971062 | 0.96643 | 0.9979453683 | 0.177233 | 9.86719115 | 1 |
| 2 | 47476374 | T | G | N | K | MSH2 | ENST00000233146 | 0.62848 | 0.0337 | 0.75926 | 0.95615 | 0.94129 | 0.9802971062 | 0.96637 | 0.9979453683 | 0.177236 | 9.86719115 | 1 |
| 2 | 47476375 | A | C | M | L | MSH2 | ENST00000233146 | 0.84665 | 0.03815 | 0.72479 | 0.93349 | 0.9755 | 0.8486277701 | 0.96444 | 0.9957854152 | 0.469457 | 8.390531957 | 0 |
| 2 | 47476375 | A | G | M | V | MSH2 | ENST00000233146 | 0.80392 | 0.03686 | 0.66153 | 0.95923 | 0.97327 | 0.9027590488 | 0.96559 | 0.9932131767 | 0.408365 | 9.587189686 | 0 |
| 2 | 47476375 | A | T | M | L | MSH2 | ENST00000233146 | 0.84028 | 0.03815 | 0.72479 | 0.93349 | 0.9755 | 0.8486277701 | 0.96466 | 0.9957854152 | 0.469457 | 8.390531957 | 0 |
| 2 | 47476376 | T | A | M | K | MSH2 | ENST00000233146 | 0.9112 | 0.03065 | 0.80826 | 0.96456 | 0.98367 | 0.9713162159 | 0.95377 | 0.999348104 | 0.47935 | 9.587189686 | 0 |
| 2 | 47476376 | T | C | M | T | MSH2 | ENST00000233146 | 0.87676 | 0.03503 | 0.8497 | 0.96353 | 0.98034 | 0.9348860872 | 0.93957 | 0.9987367988 | 0.463594 | 9.587189686 | 0 |
| 2 | 47476376 | T | G | M | R | MSH2 | ENST00000233146 | 0.92052 | 0.03093 | 0.78825 | 0.96432 | 0.98657 | 0.9680980078 | 0.97339 | 0.999348104 | 0.484977 | 9.587189686 | 0 |
| 2 | 47476377 | G | A | M | I | MSH2 | ENST00000233146 | 0.91569 | 0.03563 | 0.73114 | 0.90788 | 0.97067 | 0.9051744745 | 0.97625 | 0.998298347 | 0.481477 | 9.587189686 | 0 |
| 2 | 47476377 | G | C | M | I | MSH2 | ENST00000233146 | 0.91392 | 0.03563 | 0.73114 | 0.90343 | 0.97067 | 0.9051744745 | 0.97589 | 0.998298347 | 0.481477 | 9.587189686 | 0 |
| 2 | 47476377 | G | T | M | I | MSH2 | ENST00000233146 | 0.91012 | 0.03563 | 0.73114 | 0.90343 | 0.97067 | 0.9051744745 | 0.97599 | 0.998298347 | 0.481477 | 9.587189686 | 0 |
| 2 | 47476378 | G | A | G | R | MSH2 | ENST00000233146 | 0.94618 | 0.03452 | 0.91954 | 0.94185 | 0.97178 | 0.9844985249 | 0.98367 | 0.9998831749 | 0.476566 | 9.126578355 | 0 |
| 2 | 47476378 | G | C | G | R | MSH2 | ENST00000233146 | 0.94552 | 0.03452 | 0.91954 | 0.9388 | 0.97178 | 0.9844985249 | 0.98344 | 0.9998831749 | 0.476566 | 9.126578355 | 0 |
| 2 | 47476379 | G | A | G | E | MSH2 | ENST00000233146 | 0.86297 | 0.03334 | 0.9077 | 0.93936 | 0.97141 | 0.9871719509 | 0.97152 | 0.9998831749 | 0.476566 | 9.126578355 | 1 |
| 2 | 47476379 | G | C | G | A | MSH2 | ENST00000233146 | 0.82856 | 0.03608 | 0.8508 | 0.78496 | 0.9328 | 0.959188121 | 0.96868 | 0.9982261062 | 0.425489 | 3.667634884 | 0 |
| 2 | 47476379 | G | T | G | V | MSH2 | ENST00000233146 | 0.85851 | 0.03379 | 0.86262 | 0.93871 | 0.97475 | 0.976629866 | 0.97233 | 0.9998736382 | 0.475909 | 9.126578355 | 0 |
| 2 | 47476381 | G | A | G | S | MSH2 | ENST00000233146 | 0.93978 | 0.03728 | 0.82107 | 0.96125 | 0.99906 | 0.9363613369 | 0.9921 | 0.999725163 | 0.58487 | 9.999860499 | 0 |
| 2 | 47476381 | G | C | G | R | MSH2 | ENST00000233146 | 0.94725 | 0.03452 | 0.9022 | 0.96574 | 0.99919 | 0.9705254969 | 0.9897 | 0.9998831749 | 0.585236 | 9.999860499 | 1 |
| 2 | 47476381 | G | T | G | C | MSH2 | ENST00000233146 | 0.95142 | 0.03626 | 0.87021 | 0.96337 | 0.99765 | 0.9615407232 | 0.98971 | 0.9997038245 | 0.585236 | 9.999860499 | 0 |
| 2 | 47476382 | G | A | G | D | MSH2 | ENST00000233146 | 0.86531 | 0.03327 | 0.91156 | 0.95482 | 0.99932 | 0.9563374204 | 0.98746 | 0.9996045232 | 0.58514 | 9.999860499 | 1 |
| 2 | 47476382 | G | C | G | A | MSH2 | ENST00000233146 | 0.8376 | 0.03608 | 0.83172 | 0.95883 | 0.99976 | 0.9498925901 | 0.9902 | 0.999725163 | 0.58487 | 9.999860499 | 1 |
| 2 | 47476382 | G | T | G | V | MSH2 | ENST00000233146 | 0.86678 | 0.03379 | 0.84166 | 0.9565 | 0.99552 | 0.9726439915 | 0.98462 | 0.9998831749 | 0.585236 | 9.999860499 | 1 |
| 2 | 47476384 | A | G | K | E | MSH2 | ENST00000233146 | 0.93289 | 0.03219 | 0.80553 | 0.97977 | 0.99525 | 0.9907477102 | 0.99028 | 0.9996218681 | 0.583508 | 9.991767589 | 1 |
| 2 | 47476385 | A | C | K | T | MSH2 | ENST00000233146 | 0.92715 | 0.03458 | 0.81636 | 0.97218 | 0.99722 | 0.9850480856 | 0.98568 | 0.9996103644 | 0.584333 | 9.991767589 | 1 |
| 2 | 47476385 | A | G | K | R | MSH2 | ENST00000233146 | 0.92504 | 0.0401 | 0.74455 | 0.95973 | 0.99072 | 0.9783062966 | 0.99419 | 0.999417305 | 0.582901 | 9.991767589 | 1 |
| 2 | 47476385 | A | T | K | I | MSH2 | ENST00000233146 | 0.94284 | 0.02864 | 0.7768 | 0.97575 | 0.99326 | 0.9835656062 | 0.9741 | 0.9998438358 | 0.582756 | 9.991767589 | 1 |
| 2 | 47476386 | A | C | K | N | MSH2 | ENST00000233146 | 0.7954 | 0.03484 | 0.82729 | 0.97544 | 0.97215 | 0.988023111 | 0.97688 | 0.9993845224 | 0.402403 | 9.991767589 | 1 |
| 2 | 47476386 | A | T | K | N | MSH2 | ENST00000233146 | 0.78875 | 0.03484 | 0.82729 | 0.97544 | 0.97215 | 0.988023111 | 0.97699 | 0.9993845224 | 0.402398 | 9.991767589 | 1 |
| 2 | 47476387 | T | A | S | T | MSH2 | ENST00000233146 | 0.83213 | 0.03857 | 0.87037 | 0.93828 | 0.98549 | 0.9353364965 | 0.97928 | 0.9938124418 | 0.502412 | 9.986804285 | 0 |
| 2 | 47476387 | T | C | S | P | MSH2 | ENST00000233146 | 0.90916 | 0.0337 | 0.88114 | 0.95441 | 0.99676 | 0.9846489872 | 0.98874 | 0.9993483424 | 0.537714 | 9.986804285 | 1 |
| 2 | 47476387 | T | G | S | A | MSH2 | ENST00000233146 | 0.87133 | 0.0399 | 0.85125 | 0.95524 | 0.98972 | 0.9145679449 | 0.97772 | 0.9989256263 | 0.531344 | 9.986804285 | 1 |
| 2 | 47476388 | C | T | S | L | MSH2 | ENST00000233146 | 0.74906 | 0.03894 | 0.88106 | 0.90588 | 0.99202 | 0.965789046 | 0.98796 | 0.9996999502 | 0.53623 | 9.986804285 | 1 |
| 2 | 47476390 | A | C | T | P | MSH2 | ENST00000233146 | 0.87821 | 0.03327 | 0.67857 | 0.94333 | 0.99843 | 0.9744032549 | 0.98857 | 0.9996103644 | 0.540611 | 9.423498013 | 1 |
| 2 | 47476390 | A | G | T | A | MSH2 | ENST00000233146 | 0.84507 | 0.03747 | 0.68072 | 0.95505 | 0.99443 | 0.9328225941 | 0.9902 | 0.9965135455 | 0.536387 | 9.423498013 | 0 |

| | | | | | | | | | | | | | | | | | | |
|---|---|---|---|---|---|---|---|---|---|---|---|---|---|---|---|---|---|---|
| 2 | 47476390 | A | T | T | S | MSH2 | ENST00000233146 | 0.80898 | 0.03882 | 0.69247 | 0.94408 | 0.96846 | 0.8623900875 | 0.96641 | 0.9961155653 | 0.44598 | 7.596091865 | 0 |
| 2 | 47476391 | C | A | T | K | MSH2 | ENST00000233146 | 0.68979 | 0.03442 | 0.79541 | 0.91431 | 0.99105 | 0.9762655395 | 0.98378 | 0.9998425245 | 0.532067 | 9.423498013 | 1 |
| 2 | 47476391 | C | G | T | R | MSH2 | ENST00000233146 | 0.677 | 0.03518 | 0.77592 | 0.93821 | 0.99202 | 0.9850273877 | 0.98746 | 0.9997094274 | 0.532068 | 9.423498013 | 1 |
| 2 | 47476391 | C | T | T | I | MSH2 | ENST00000233146 | 0.66929 | 0.03608 | 0.82291 | 0.92364 | 0.99138 | 0.9553839278 | 0.98642 | 0.9968941212 | 0.528177 | 9.423498013 | 0 |
| 2 | 47476393 | T | C | Y | H | MSH2 | ENST00000233146 | 0.93786 | 0.03313 | 0.8679 | 0.90146 | 0.96993 | 0.9106248709 | 0.89253 | 0.9985123277 | 0.334845 | 8.079809597 | 0 |
| 2 | 47476393 | T | G | Y | D | MSH2 | ENST00000233146 | 0.94323 | 0.02851 | 0.85153 | 0.93514 | 0.98071 | 0.9685958169 | 0.95429 | 0.9995419979 | 0.481112 | 8.079809597 | 1 |
| 2 | 47476394 | A | C | Y | S | MSH2 | ENST00000233146 | 0.94372 | 0.03166 | 0.79752 | 0.92365 | 0.95426 | 0.9444448928 | 0.91057 | 0.9992134571 | 0.379956 | 8.079809597 | 1 |
| 2 | 47476394 | A | G | Y | C | MSH2 | ENST00000233146 | 0.94971 | 0.03379 | 0.85994 | 0.92471 | 0.95572 | 0.9388920261 | 0.88763 | 0.9957019687 | 0.417095 | 8.079809597 | 0 |
| 2 | 47476394 | A | T | Y | F | MSH2 | ENST00000233146 | 0.75591 | 0.023 | 0.77008 | 0.59948 | 0.80595 | 0.7857919996 | 0.81601 | 0.8960491419 | 0.113214 | 2.047685573 | 0 |
| 2 | 47476396 | A | C | I | L | MSH2 | ENST00000233146 | 0.71746 | 0.0276 | 0.79829 | 0.76514 | 0.911 | 0.8487040066 | 0.88676 | 0.9033383131 | 0.31091 | 4.033723572 | 0 |
| 2 | 47476396 | A | G | I | V | MSH2 | ENST00000233146 | 0.79302 | 0.03735 | 0.70061 | 0.83627 | 0.91357 | 0.7151058305 | 0.88769 | 0.8938782412 | 0.296717 | 7.656571807 | 0 |
| 2 | 47476396 | A | T | I | F | MSH2 | ENST00000233146 | 0.8755 | 0.03019 | 0.80827 | 0.93321 | 0.98657 | 0.9531587627 | 0.9642 | 0.9910740256 | 0.462725 | 8.75911898 | 0 |
| 2 | 47476397 | T | A | I | N | MSH2 | ENST00000233146 | 0.91626 | 0.02857 | 0.88698 | 0.94595 | 0.98513 | 0.9799676618 | 0.9162 | 0.9989677668 | 0.422106 | 8.75911898 | 0 |
| 2 | 47476397 | T | C | I | T | MSH2 | ENST00000233146 | 0.87794 | 0.03353 | 0.89482 | 0.94057 | 0.98728 | 0.9388915579 | 0.94584 | 0.9956839085 | 0.460207 | 8.75911898 | 0 |
| 2 | 47476397 | T | G | I | S | MSH2 | ENST00000233146 | 0.92104 | 0.0315 | 0.82033 | 0.94555 | 0.98728 | 0.9753132449 | 0.96957 | 0.9993494153 | 0.467906 | 8.75911898 | 0 |
| 2 | 47476398 | T | G | I | M | MSH2 | ENST00000233146 | 0.73277 | 0.0368 | 0.75649 | 0.92035 | 0.9248 | 0.8794305721 | 0.93755 | 0.9556355476 | 0.354206 | 3.991877361 | 0 |
| 2 | 47476399 | C | G | R | G | MSH2 | ENST00000233146 | 0.9342 | 0.03334 | 0.81453 | 0.91697 | 0.9579 | 0.9465283677 | 0.98249 | 0.9988732934 | 0.314681 | 9.986800735 | 0 |
| 2 | 47476400 | G | A | R | Q | MSH2 | ENST00000233146 | 0.8624 | 0.03491 | 0.76009 | 0.9096 | 0.97624 | 0.9080540804 | 0.98407 | 0.9991658926 | 0.490435 | 9.986800735 | 0 |
| 2 | 47476400 | G | C | R | P | MSH2 | ENST00000233146 | 0.87516 | 0.03166 | 0.86102 | 0.88825 | 0.97735 | 0.9779517751 | 0.9822 | 0.9984611273 | 0.494323 | 9.986800735 | 1 |
| 2 | 47476400 | G | T | R | L | MSH2 | ENST00000233146 | 0.85873 | 0.03388 | 0.85059 | 0.89621 | 0.9781 | 0.9374303134 | 0.98668 | 0.999121964 | 0.493497 | 9.986800735 | 0 |
| 2 | 47476402 | C | A | Q | K | MSH2 | ENST00000233146 | 0.73829 | 0.029 | 0.78948 | 0.92593 | 0.96226 | 0.9783483534 | 0.98346 | 0.9974835515 | 0.451678 | 7.856226172 | 0 |
| 2 | 47476402 | C | G | Q | E | MSH2 | ENST00000233146 | 0.69836 | 0.02943 | 0.74004 | 0.86211 | 0.94129 | 0.9710489615 | 0.97344 | 0.990783453 | 0.455764 | 7.856226172 | 0 |
| 2 | 47476403 | A | C | Q | P | MSH2 | ENST00000233146 | 0.84868 | 0.03313 | 0.74616 | 0.92134 | 0.98257 | 0.9918881446 | 0.97583 | 0.997987628 | 0.466899 | 7.856226172 | 1 |
| 2 | 47476403 | A | G | Q | R | MSH2 | ENST00000233146 | 0.82864 | 0.0337 | 0.784 | 0.91979 | 0.9781 | 0.9840665689 | 0.98635 | 0.9972572923 | 0.466051 | 7.856226172 | 1 |
| 2 | 47476403 | A | T | Q | L | MSH2 | ENST00000233146 | 0.86025 | 0.03046 | 0.72936 | 0.93994 | 0.98585 | 0.9688198303 | 0.98752 | 0.9906864762 | 0.466609 | 7.856226172 | 0 |
| 2 | 47476404 | A | C | Q | H | MSH2 | ENST00000233146 | 0.43418 | 0.03741 | 0.78533 | 0.72156 | 0.862 | 0.9689385642 | 0.97623 | 0.983591795 | 0.269761 | 7.856226172 | 0 |
| 2 | 47476404 | A | T | Q | H | MSH2 | ENST00000233146 | 0.42914 | 0.03741 | 0.78533 | 0.72423 | 0.862 | 0.9689385642 | 0.97626 | 0.9992799163 | 0.396131 | 7.856226172 | 0 |
| 2 | 47476405 | A | C | T | P | MSH2 | ENST00000233146 | 0.70935 | 0.02193 | 0.57533 | 0.84808 | 0.90537 | 0.9185627568 | 0.94858 | 0.9210376143 | 0.307157 | 6.800508058 | 0 |
| 2 | 47476405 | A | G | T | A | MSH2 | ENST00000233146 | 0.36203 | 0.0058 | 0.57214 | 0.54336 | 0.71548 | 0.6633841386 | 0.96976 | 0.520863235 | 0.166924 | 3.514816164 | 0 |
| 2 | 47476405 | A | T | T | S | MSH2 | ENST00000233146 | 0.49261 | 0.01019 | 0.58868 | 0.71103 | 0.81472 | 0.6575793584 | 0.96315 | 0.7208108306 | 0.208791 | 4.891445262 | 0 |
| 2 | 47476406 | C | A | T | N | MSH2 | ENST00000233146 | 0.51876 | 0.02307 | 0.67943 | 0.67376 | 0.81006 | 0.84530022 | 0.86464 | 0.9657834768 | 0.177083 | 5.275269313 | 0 |
| 2 | 47476406 | C | G | T | S | MSH2 | ENST00000233146 | 0.38027 | 0.01019 | 0.58868 | 0.61043 | 0.77141 | 0.6575793584 | 0.93653 | 0.8769236803 | 0.214178 | 4.891445262 | 0 |
| 2 | 47476406 | C | T | T | I | MSH2 | ENST00000233146 | 0.22406 | 0.00569 | 0.74488 | 0.54365 | 0.71783 | 0.6467475447 | 0.91857 | 0.8279083967 | 0.183459 | 1.974908899 | 0 |
| 2 | 47476408 | G | A | G | R | MSH2 | ENST00000233146 | 0.92917 | 0.03447 | 0.94204 | 0.91396 | 0.96371 | 0.9837349899 | 0.97315 | 0.9992679954 | 0.472484 | 8.684862259 | 1 |
| 2 | 47476408 | G | C | G | R | MSH2 | ENST00000233146 | 0.92778 | 0.03447 | 0.94204 | 0.91396 | 0.96371 | 0.9837349899 | 0.97283 | 0.9992679954 | 0.472484 | 8.684862259 | 1 |
| 2 | 47476409 | G | A | G | E | MSH2 | ENST00000233146 | 0.79792 | 0.03322 | 0.92956 | 0.92709 | 0.97475 | 0.9822945534 | 0.96302 | 0.9998736382 | 0.472665 | 8.684862259 | 0 |
| 2 | 47476409 | G | C | G | A | MSH2 | ENST00000233146 | 0.62589 | 0.03313 | 0.87499 | 0.8127 | 0.91526 | 0.9234669802 | 0.94966 | 0.9891127944 | 0.40696 | 3.614492961 | 0 |
| 2 | 47476409 | G | T | G | V | MSH2 | ENST00000233146 | 0.7931 | 0.0337 | 0.8844 | 0.91385 | 0.96956 | 0.9596537885 | 0.96285 | 0.9989221096 | 0.455937 | 8.684862259 | 0 |
| 2 | 47476411 | G | A | V | M | MSH2 | ENST00000233146 | 0.80274 | 0.0197 | 0.66602 | 0.85259 | 0.93901 | 0.7822855499 | 0.90232 | 0.9673397485 | 0.348772 | 5.193200088 | 0 |
| 2 | 47476411 | G | C | V | L | MSH2 | ENST00000233146 | 0.87201 | 0.00747 | 0.62453 | 0.74523 | 0.89603 | 0.7742689075 | 0.9043 | 0.9392703772 | 0.233772 | 3.258157155 | 0 |
| 2 | 47476411 | G | T | V | L | MSH2 | ENST00000233146 | 0.86636 | 0.00747 | 0.62453 | 0.76161 | 0.89603 | 0.7742689075 | 0.90426 | 0.9392703772 | 0.233774 | 3.258157155 | 0 |
| 2 | 47476412 | T | A | V | E | MSH2 | ENST00000233146 | 0.87342 | 0.02815 | 0.63243 | 0.9337 | 0.9833 | 0.9526753443 | 0.94625 | 0.9905390974 | 0.464641 | 7.264299978 | 1 |
| 2 | 47476412 | T | C | V | A | MSH2 | ENST00000233146 | 0.82358 | 0.02414 | 0.64875 | 0.87266 | 0.96407 | 0.8427245132 | 0.89186 | 0.968993187 | 0.34027 | 4.406715369 | 0 |
| 2 | 47476412 | T | G | V | G | MSH2 | ENST00000233146 | 0.869 | 0.03124 | 0.50602 | 0.91819 | 0.9822 | 0.9244497363 | 0.94748 | 0.9769558311 | 0.465134 | 7.264299978 | 0 |
| 2 | 47476414 | A | C | I | L | MSH2 | ENST00000233146 | 0.45832 | 0.00603 | 0.37428 | 0.67155 | 0.82103 | 0.8200401354 | 0.93316 | 0.6579895616 | 0.181892 | 4.06026883 | 0 |
| 2 | 47476414 | A | G | I | V | MSH2 | ENST00000233146 | 0.28106 | 0.00608 | 0.27313 | 0.58675 | 0.71153 | 0.6142266409 | 0.88111 | 0.3496392965 | 0.106371 | 1.485286071 | 0 |
| 2 | 47476414 | A | T | I | L | MSH2 | ENST00000233146 | 0.4531 | 0.00603 | 0.37428 | 0.67155 | 0.82103 | 0.8200401354 | 0.93403 | 0.6579895616 | 0.181888 | 4.06026883 | 0 |
| 2 | 47476415 | T | C | I | T | MSH2 | ENST00000233146 | 0.41558 | 0.00983 | 0.55328 | 0.76744 | 0.86653 | 0.8869995596 | 0.88665 | 0.8736730218 | 0.36481 | 4.064996291 | 0 |
| 2 | 47476415 | T | G | I | R | MSH2 | ENST00000233146 | 0.72356 | 0.01649 | 0.44725 | 0.93483 | 0.95426 | 0.9687823869 | 0.9461 | 0.9968768358 | 0.432113 | 7.412092408 | 1 |
| 2 | 47476417 | G | A | V | I | MSH2 | ENST00000233146 | 0.82934 | 0.0357 | 0.42055 | 0.74022 | 0.79028 | 0.7047635909 | 0.83515 | 0.9161680937 | 0.210192 | 4.07893306 | 0 |
| 2 | 47476417 | G | C | V | L | MSH2 | ENST00000233146 | 0.87234 | 0.02994 | 0.62387 | 0.83118 | 0.89603 | 0.8541063008 | 0.92512 | 0.938075006 | 0.314398 | 7.067819573 | 0 |
| 2 | 47476417 | G | T | V | L | MSH2 | ENST00000233146 | 0.86671 | 0.02994 | 0.62387 | 0.83222 | 0.89603 | 0.8541063008 | 0.92553 | 0.938075006 | 0.314398 | 7.067819573 | 0 |
| 2 | 47476418 | T | A | V | E | MSH2 | ENST00000233146 | 0.75742 | 0.02883 | 0.62961 | 0.93827 | 0.9659 | 0.9542547718 | 0.95297 | 0.9944061041 | 0.43664 | 8.627173672 | 0 |
| 2 | 47476418 | T | C | V | A | MSH2 | ENST00000233146 | 0.48215 | 0.01391 | 0.64763 | 0.56205 | 0.76356 | 0.8032062163 | 0.7527 | 0.4679705501 | 0.164946 | 3.41010616 | 0 |
| 2 | 47476418 | T | G | V | G | MSH2 | ENST00000233146 | 0.74986 | 0.03099 | 0.49098 | 0.88835 | 0.96044 | 0.8896206006 | 0.92464 | 0.9821313437 | 0.430947 | 7.631733659 | 0 |
| 2 | 47476420 | C | A | L | I | MSH2 | ENST00000233146 | 0.72292 | 0.03527 | 0.84627 | 0.75773 | 0.82948 | 0.6935134682 | 0.82738 | 0.9595156908 | 0.23906 | 4.165453692 | 0 |
| 2 | 47476420 | C | G | L | V | MSH2 | ENST00000233146 | 0.71506 | 0.03313 | 0.80426 | 0.74217 | 0.86603 | 0.8318163391 | 0.90723 | 0.9906898737 | 0.302299 | 5.011271057 | 0 |
| 2 | 47476420 | C | T | L | F | MSH2 | ENST00000233146 | 0.75687 | 0.03582 | 0.84566 | 0.86941 | 0.911 | 0.8969362836 | 0.90355 | 0.9955376387 | 0.412149 | 7.909087578 | 0 |
| 2 | 47476421 | T | A | L | H | MSH2 | ENST00000233146 | 0.79982 | 0.0334 | 0.91525 | 0.94036 | 0.99265 | 0.9491328145 | 0.97042 | 0.9991675615 | 0.483121 | 6.731079148 | 0 |
| 2 | 47476421 | T | C | L | P | MSH2 | ENST00000233146 | 0.82412 | 0.03001 | 0.9376 | 0.94914 | 0.98904 | 0.983434382 | 0.97848 | 0.9994055033 | 0.483117 | 9.058079743 | 1 |
| 2 | 47476421 | T | G | L | R | MSH2 | ENST00000233146 | 0.81967 | 0.03144 | 0.91592 | 0.93765 | 0.99072 | 0.9850277044 | 0.97791 | 0.9992813468 | 0.482395 | 9.058079743 | 1 |
| 2 | 47476423 | A | C | M | L | MSH2 | ENST00000233146 | 0.61566 | 0.00909 | 0.81926 | 0.61577 | 0.88401 | 0.9218529461 | 0.93516 | 0.7816397548 | 0.2882 | 3.339263228 | 0 |
| 2 | 47476423 | A | G | M | V | MSH2 | ENST00000233146 | 0.77561 | 0.03088 | 0.75668 | 0.89566 | 0.97364 | 0.9537384682 | 0.98511 | 0.9962003827 | 0.460327 | 9.32421286 | 0 |
| 2 | 47476423 | A | T | M | L | MSH2 | ENST00000233146 | 0.60941 | 0.00909 | 0.81926 | 0.61577 | 0.88401 | 0.9218529461 | 0.93571 | 0.7671789527 | 0.288198 | 3.339263228 | 0 |
| 2 | 47476424 | T | A | M | K | MSH2 | ENST00000233146 | 0.8755 | 0.03023 | 0.88764 | 0.94077 | 0.99326 | 0.9935603889 | 0.96723 | 0.9991950393 | 0.471698 | 9.32421286 | 1 |
| 2 | 47476424 | T | C | M | T | MSH2 | ENST00000233146 | 0.83089 | 0.03436 | 0.91577 | 0.94402 | 0.9844 | 0.9745533868 | 0.95034 | 0.9985980392 | 0.469031 | 9.32421286 | 0 |
| 2 | 47476424 | T | G | M | R | MSH2 | ENST00000233146 | 0.8891 | 0.03053 | 0.87023 | 0.93466 | 0.9917 | 0.9943420466 | 0.96824 | 0.9991950393 | 0.472236 | 9.32421286 | 1 |
| 2 | 47476425 | G | A | M | I | MSH2 | ENST00000233146 | 0.87834 | 0.03129 | 0.81669 | 0.88989 | 0.94356 | 0.9559237661 | 0.98326 | 0.9324326515 | 0.216087 | 7.201661186 | 0 |
| 2 | 47476425 | G | C | M | I | MSH2 | ENST00000233146 | 0.87563 | 0.03129 | 0.81669 | 0.88889 | 0.94394 | 0.9559237661 | 0.98284 | 0.9730467485 | 0.454687 | 7.201661186 | 0 |
| 2 | 47476425 | G | T | M | I | MSH2 | ENST00000233146 | 0.87017 | 0.03129 | 0.81669 | 0.88989 | 0.94394 | 0.9559237661 | 0.983 | 0.9733350978 | 0.454687 | 7.201661186 | 0 |
| 2 | 47476426 | G | A | A | T | MSH2 | ENST00000233146 | 0.88373 | 0.02927 | 0.81221 | 0.87201 | 0.96737 | 0.953991208 | 0.96958 | 0.9979174733 | 0.494111 | 6.204915065 | 0 |
| 2 | 47476426 | G | C | A | P | MSH2 | ENST00000233146 | 0.89388 | 0.03313 | 0.86569 | 0.86119 | 0.98034 | 0.9898848447 | 0.97495 | 0.9996891022 | 0.497976 | 9.077249099 | 1 |
| 2 | 47476426 | G | T | A | S | MSH2 | ENST00000233146 | 0.85189 | 0.03753 | 0.77513 | 0.85545 | 0.92642 | 0.9074156139 | 0.93727 | 0.9941412807 | 0.399738 | 4.918410488 | 0 |
| 2 | 47476427 | C | A | A | D | MSH2 | ENST00000233146 | 0.74682 | 0.03093 | 0.89613 | 0.90803 | 0.9729 | 0.9912761323 | 0.97059 | 0.9998425245 | 0.494923 | 9.077249099 | 1 |
| 2 | 47476427 | C | G | A | G | MSH2 | ENST00000233146 | 0.73845 | 0.0366 | 0.79384 | 0.90032 | 0.967 | 0.9070964564 | 0.96124 | 0.9959540367 | 0.490618 | 5.341874594 | 0 |
| 2 | 47476427 | C | T | A | V | MSH2 | ENST00000233146 | 0.74117 | 0.03626 | 0.83046 | 0.8922 | 0.97327 | 0.965734196 | 0.96704 | 0.9998743534 | 0.493371 | 9.077249099 | 0 |
| 2 | 47476429 | C | A | Q | K | MSH2 | ENST00000233146 | 0.7575 | 0.03362 | 0.76489 | 0.92407 | 0.97624 | 0.9844299727 | 0.97261 | 0.9993513227 | 0.447687 | 9.597402494 | 1 |
| 2 | 47476429 | C | G | Q | E | MSH2 | ENST00000233146 | 0.71634 | 0.03595 | 0.70651 | 0.87375 | 0.96153 | 0.9803467232 | 0.97751 | 0.9991763234 | 0.455252 | 9.597402494 | 1 |
| 2 | 47476430 | A | C | Q | P | MSH2 | ENST00000233146 | 0.86685 | 0.03419 | 0.71666 | 0.94047 | 0.97773 | 0.995131719 | 0.96503 | 0.9993166924 | 0.467152 | 9.597402494 | 1 |
| 2 | 47476430 | A | G | Q | R | MSH2 | ENST00000233146 | 0.84688 | 0.03613 | 0.7564 | 0.94062 | 0.98972 | 0.9893840335 | 0.98093 | 0.9991630316 | 0.461159 | 9.597402494 | 1 |
| 2 | 47476430 | A | T | Q | L | MSH2 | ENST00000233146 | 0.88952 | 0.03753 | 0.70092 | 0.95819 | 0.98549 | 0.9756675323 | 0.9684 | 0.9988161325 | 0.470773 | 9.597402494 | 0 |
| 2 | 47476431 | A | C | Q | H | MSH2 | ENST00000233146 | 0.44801 | 0.03809 | 0.75601 | 0.86468 | 0.91143 | 0.9823661491 | 0.97984 | 0.9901592135 | 0.355041 | 5.356749647 | 0 |
| 2 | 47476431 | A | T | Q | H | MSH2 | ENST00000233146 | 0.44283 | 0.03809 | 0.75601 | 0.86717 | 0.91186 | 0.9823661491 | 0.97981 | 0.9919025302 | 0.355037 | 5.356749647 | 0 |
| 2 | 47476432 | A | C | I | L | MSH2 | ENST00000233146 | 0.83806 | 0.02349 | 0.79988 | 0.82887 | 0.87829 | 0.8761113122 | 0.9101 | 0.8798521161 | 0.251187 | 2.646480864 | 0 |
| 2 | 47476432 | A | G | I | V | MSH2 | ENST00000233146 | 0.63897 | 0.03006 | 0.71305 | 0.5942 | 0.79028 | 0.7686148926 | 0.84207 | 0.7682339837 | 0.202412 | 3.093634802 | 0 |
| 2 | 47476432 | A | T | I | F | MSH2 | ENST00000233146 | 0.87742 | 0.03006 | 0.81327 | 0.93811 | 0.96846 | 0.961576101 | 0.95145 | 0.9984973669 | 0.464124 | 8.251395709 | 0 |
| 2 | 47476433 | T | A | I | N | MSH2 | ENST00000233146 | 0.93457 | 0.02851 | 0.87898 | 0.95781 | 0.96956 | 0.9804047682 | 0.88253 | 0.9980649352 | 0.373207 | 8.251395709 | 0 |
| 2 | 47476433 | T | C | I | T | MSH2 | ENST00000233146 | 0.78756 | 0.03334 | 0.89351 | 0.85414 | 0.96846 | 0.9545780881 | 0.94818 | 0.9727089405 | 0.411888 | 5.078959577 | 0 |
| 2 | 47476433 | T | G | I | S | MSH2 | ENST00000233146 | 0.93758 | 0.03129 | 0.81897 | 0.91237 | 0.97997 | 0.9799291855 | 0.94908 | 0.9889040589 | 0.447979 | 7.445978739 | 0 |
| 2 | 47476434 | T | G | I | M | MSH2 | ENST00000233146 | 0.73509 | 0.03431 | 0.76613 | 0.87078 | 0.86753 | 0.9144872742 | 0.87794 | 0.9235622883 | 0.137801 | 2.804006999 | 0 |
| 2 | 47476435 | G | A | G | R | MSH2 | ENST00000233146 | 0.9368 | 0.03452 | 0.94522 | 0.94316 | 0.99414 | 0.971596966 | 0.98544 | 0.9998831749 | 0.558029 | 9.986793403 | 1 |
| 2 | 47476435 | G | C | G | R | MSH2 | ENST00000233146 | 0.93573 | 0.03452 | 0.94522 | 0.93794 | 0.99414 | 0.971596966 | 0.98526 | 0.9998831749 | 0.55803 | 9.986793403 | 1 |
| 2 | 47476436 | G | A | G | E | MSH2 | ENST00000233146 | 0.87423 | 0.03334 | 0.93183 | 0.9226 | 0.99138 | 0.9703898384 | 0.9768 | 0.9998831749 | 0.557559 | 9.986793403 | 1 |
| 2 | 47476436 | G | C | G | A | MSH2 | ENST00000233146 | 0.84127 | 0.03608 | 0.87914 | 0.90822 | 0.98869 | 0.9507586642 | 0.97746 | 0.999725163 | 0.555825 | 9.986793403 | 0 |
| 2 | 47476436 | G | T | G | V | MSH2 | ENST00000233146 | 0.8701 | 0.03379 | 0.88331 | 0.92841 | 0.98904 | 0.9727780525 | 0.97797 | 0.9998831749 | 0.55803 | 9.986793403 | 1 |
| 2 | 47476438 | T | A | C | S | MSH2 | ENST00000233146 | 0.58006 | 0.00827 | 0.45783 | 0.48608 | 0.82387 | 0.948754056 | 0.76026 | 0.7482958436 | 0.203625 | 3.14844422 | 0 |
| 2 | 47476438 | T | C | C | R | MSH2 | ENST00000233146 | 0.94854 | 0.0276 | 0.58392 | 0.94847 | 0.96371 | 0.9853341926 | 0.94722 | 0.9921660222 | 0.445619 | 9.235179491 | 1 |
| 2 | 47476438 | T | G | C | G | MSH2 | ENST00000233146 | 0.94102 | 0.02955 | 0.41564 | 0.88773 | 0.93004 | 0.9659037725 | 0.91436 | 0.9952414036 | 0.419136 | 9.235179491 | 0 |
| 2 | 47476439 | G | C | C | S | MSH2 | ENST00000233146 | 0.63804 | 0.00827 | 0.45783 | 0.48826 | 0.82159 | 0.948754056 | 0.77834 | 0.8212917447 | 0.217581 | 3.14844422 | 0 |
| 2 | 47476439 | G | T | C | F | MSH2 | ENST00000233146 | 0.95338 | 0.03001 | 0.4775 | 0.92467 | 0.95935 | 0.9779327435 | 0.93178 | 0.9951891899 | 0.453388 | 8.093563197 | 1 |
| 2 | 47476440 | T | G | C | W | MSH2 | ENST00000233146 | 0.72764 | 0.03265 | 0.52836 | 0.94931 | 0.94506 | 0.9819477988 | 0.93591 | 0.9989473224 | 0.418582 | 9.235179491 | 1 |

| | | | | | | | | | | | | | | | | | | |
|---|---|---|---|---|---|---|---|---|---|---|---|---|---|---|---|---|---|---|
| 2 | 47476441 | T | A | F | I | MSH2 | ENST00000233146 | 0.9056 | 0.0318 | 0.71928 | 0.93872 | 0.98513 | 0.9670993461 | 0.94233 | 0.9993228912 | 0.482833 | 8.769741362 | 1 |
| 2 | 47476441 | T | C | F | L | MSH2 | ENST00000233146 | 0.91405 | 0.03362 | 0.75375 | 0.91561 | 0.98938 | 0.9516020712 | 0.95649 | 0.9934257269 | 0.476224 | 6.427751247 | 0 |
| 2 | 47476441 | T | G | F | V | MSH2 | ENST00000233146 | 0.90964 | 0.03274 | 0.68317 | 0.95196 | 0.98657 | 0.9617169649 | 0.93974 | 0.9990255833 | 0.482637 | 8.769741362 | 1 |
| 2 | 47476442 | T | A | F | Y | MSH2 | ENST00000233146 | 0.89101 | 0.0175 | 0.72846 | 0.84454 | 0.90494 | 0.9219975695 | 0.8945 | 0.981400013 | 0.283841 | 2.734548041 | 0 |
| 2 | 47476442 | T | C | F | S | MSH2 | ENST00000233146 | 0.93867 | 0.02844 | 0.71349 | 0.95415 | 0.99296 | 0.9762464391 | 0.96397 | 0.9996949434 | 0.483609 | 8.506619268 | 1 |
| 2 | 47476443 | T | A | F | L | MSH2 | ENST00000233146 | 0.71281 | 0.03362 | 0.75375 | 0.9422 | 0.97698 | 0.9516020712 | 0.96961 | 0.995621264 | 0.440017 | 6.427751247 | 0 |
| 2 | 47476443 | T | G | F | L | MSH2 | ENST00000233146 | 0.721 | 0.03362 | 0.75375 | 0.9435 | 0.97698 | 0.9516020712 | 0.96945 | 0.995621264 | 0.440019 | 6.427751247 | 0 |
| 2 | 47476444 | G | A | V | M | MSH2 | ENST00000233146 | 0.90271 | 0.03697 | 0.81961 | 0.87975 | 0.96664 | 0.9468640779 | 0.96317 | 0.9995442033 | 0.497798 | 9.436122802 | 1 |
| 2 | 47476444 | G | C | V | L | MSH2 | ENST00000233146 | 0.88411 | 0.03582 | 0.78711 | 0.8775 | 0.96627 | 0.9332727417 | 0.97693 | 0.9901407361 | 0.356725 | 9.436122802 | 0 |
| 2 | 47476444 | G | T | V | L | MSH2 | ENST00000233146 | 0.87892 | 0.03582 | 0.78711 | 0.8775 | 0.96627 | 0.9332727417 | 0.97712 | 0.9948195815 | 0.479144 | 9.436122802 | 0 |
| 2 | 47476445 | T | A | V | E | MSH2 | ENST00000233146 | 0.89249 | 0.02989 | 0.79278 | 0.96111 | 0.98621 | 0.9898333961 | 0.97427 | 0.9993228912 | 0.493755 | 9.436122802 | 1 |
| 2 | 47476445 | T | C | V | A | MSH2 | ENST00000233146 | 0.8629 | 0.03409 | 0.80452 | 0.94128 | 0.98513 | 0.9387621515 | 0.94606 | 0.9993026257 | 0.490989 | 8.97617134 | 0 |
| 2 | 47476445 | T | G | V | G | MSH2 | ENST00000233146 | 0.88892 | 0.03313 | 0.6845 | 0.95023 | 0.98834 | 0.9869444672 | 0.95908 | 0.9989577532 | 0.494022 | 9.436122802 | 1 |
| 2 | 47476447 | C | A | P | T | MSH2 | ENST00000233146 | 0.75057 | 0.03552 | 0.82894 | 0.91975 | 0.98071 | 0.9545014936 | 0.94339 | 0.9998764992 | 0.49387 | 9.991366733 | 1 |
| 2 | 47476447 | C | G | P | A | MSH2 | ENST00000233146 | 0.72933 | 0.03761 | 0.78148 | 0.92 | 0.97401 | 0.9437695612 | 0.93079 | 0.9998368025 | 0.491617 | 7.977053946 | 0 |
| 2 | 47476447 | C | T | P | S | MSH2 | ENST00000233146 | 0.78075 | 0.03706 | 0.79116 | 0.93006 | 0.98513 | 0.9595776071 | 0.94541 | 0.9998664856 | 0.492747 | 7.492298706 | 0 |
| 2 | 47476448 | C | A | P | Q | MSH2 | ENST00000233146 | 0.68195 | 0.03832 | 0.79604 | 0.89503 | 0.96846 | 0.9727489268 | 0.93588 | 0.9998764992 | 0.494665 | 9.991366733 | 1 |
| 2 | 47476448 | C | G | P | R | MSH2 | ENST00000233146 | 0.69764 | 0.03633 | 0.84928 | 0.89808 | 0.97661 | 0.9831893402 | 0.94567 | 0.9998764992 | 0.494303 | 9.991366733 | 1 |
| 2 | 47476448 | C | T | P | L | MSH2 | ENST00000233146 | 0.71634 | 0.03692 | 0.85581 | 0.9189 | 0.98367 | 0.9706380476 | 0.97028 | 0.9999756813 | 0.494296 | 9.991366733 | 1 |
| 2 | 47476450 | T | A | C | S | MSH2 | ENST00000233146 | 0.87736 | 0.03261 | 0.75504 | 0.90787 | 0.97438 | 0.9652287577 | 0.92611 | 0.993332684 | 0.430156 | 7.024230103 | 0 |
| 2 | 47476450 | T | C | C | R | MSH2 | ENST00000233146 | 0.91958 | 0.02844 | 0.859 | 0.92266 | 0.96919 | 0.992010156 | 0.96262 | 0.9992105961 | 0.459906 | 8.75102474 | 1 |
| 2 | 47476450 | T | G | C | G | MSH2 | ENST00000233146 | 0.91016 | 0.03243 | 0.71591 | 0.93479 | 0.97067 | 0.9760002543 | 0.94731 | 0.9990398288 | 0.442249 | 8.75102474 | 0 |
| 2 | 47476451 | G | A | C | Y | MSH2 | ENST00000233146 | 0.92135 | 0.02914 | 0.87386 | 0.8945 | 0.95572 | 0.9846197238 | 0.94482 | 0.9994367957 | 0.472428 | 8.75102474 | 1 |
| 2 | 47476451 | G | C | C | S | MSH2 | ENST00000233146 | 0.91129 | 0.03261 | 0.75504 | 0.87371 | 0.94543 | 0.9652287577 | 0.91838 | 0.9967343807 | 0.436943 | 7.024230103 | 0 |
| 2 | 47476451 | G | T | C | F | MSH2 | ENST00000233146 | 0.93117 | 0.03065 | 0.77384 | 0.90482 | 0.95133 | 0.9852300285 | 0.96283 | 0.9994693398 | 0.472429 | 8.75102474 | 1 |
| 2 | 47476452 | T | G | C | W | MSH2 | ENST00000233146 | 0.5038 | 0.03274 | 0.82478 | 0.97469 | 0.92151 | 0.9878677737 | 0.97845 | 0.999103725 | 0.382414 | 8.75102474 | 1 |
| 2 | 47476453 | G | A | E | K | MSH2 | ENST00000233146 | 0.72059 | 0.0071 | 0.67487 | 0.67545 | 0.7872 | 0.7853230683 | 0.88924 | 0.7495583892 | 0.209577 | 2.30127826 | 0 |
| 2 | 47476453 | G | C | E | Q | MSH2 | ENST00000233146 | 0.63046 | 0.00638 | 0.53247 | 0.61186 | 0.73443 | 0.7457396206 | 0.9258 | 0.5804255387 | 0.18615 | 1.908524322 | 0 |
| 2 | 47476454 | A | C | E | A | MSH2 | ENST00000233146 | 0.38725 | 0.00566 | 0.54144 | 0.68916 | 0.79637 | 0.8031858903 | 0.89551 | 0.8322399259 | 0.165933 | 2.195190811 | 0 |
| 2 | 47476454 | A | G | E | G | MSH2 | ENST00000233146 | 0.43932 | 0.00547 | 0.48995 | 0.75725 | 0.84375 | 0.7693533643 | 0.93152 | 0.9075749516 | 0.228609 | 4.219260034 | 0 |
| 2 | 47476454 | A | T | E | V | MSH2 | ENST00000233146 | 0.56707 | 0.00791 | 0.51305 | 0.86 | 0.92682 | 0.8290725018 | 0.9513 | 0.9191943407 | 0.307886 | 5.466487577 | 0 |
| 2 | 47476455 | G | C | E | D | MSH2 | ENST00000233146 | 0.19956 | 0.00592 | 0.54525 | 0.39515 | 0.77718 | 0.6556218609 | 0.96795 | 0.0846170634 | 0.0971162 | -0.5432630214 | 0 |
| 2 | 47476455 | G | T | E | D | MSH2 | ENST00000233146 | 0.19652 | 0.00592 | 0.54525 | 0.39515 | 0.77718 | 0.6556218609 | 0.96811 | 0.0846170634 | 0.0971151 | -0.5432630214 | 0 |
| 2 | 47476456 | T | A | S | T | MSH2 | ENST00000233146 | 0.39649 | 0.00608 | 0.42564 | 0.57516 | 0.79516 | 0.5569080838 | 0.93161 | 0.2261073291 | 0.0502773 | 3.377854688 | 0 |
| 2 | 47476456 | T | C | S | P | MSH2 | ENST00000233146 | 0.52413 | 0.01549 | 0.42983 | 0.6913 | 0.83283 | 0.8921202499 | 0.95064 | 0.9182096124 | 0.238118 | 7.898740902 | 1 |
| 2 | 47476456 | T | G | S | A | MSH2 | ENST00000233146 | 0.42986 | 0.00622 | 0.38563 | 0.52866 | 0.70262 | 0.4772869233 | 0.9169 | 0.3663762212 | 0.0575102 | 3.463209744 | 0 |
| 2 | 47476457 | C | T | S | L | MSH2 | ENST00000233146 | 0.25259 | 0.00622 | 0.42951 | 0.71785 | 0.79577 | 0.6900548867 | 0.94233 | 0.6629170179 | 0.14516 | 5.933595616 | 0 |
| 2 | 47476459 | G | A | A | T | MSH2 | ENST00000233146 | 0.90935 | 0.03741 | 0.66807 | 0.91115 | 0.95863 | 0.9462721989 | 0.95006 | 0.9997059703 | 0.480815 | 8.871343396 | 0 |
| 2 | 47476459 | G | C | A | P | MSH2 | ENST00000233146 | 0.91613 | 0.03322 | 0.74411 | 0.91802 | 0.96993 | 0.990738759 | 0.95337 | 0.999725163 | 0.484239 | 8.871343396 | 0 |
| 2 | 47476459 | G | T | A | S | MSH2 | ENST00000233146 | 0.8824 | 0.03832 | 0.62724 | 0.87207 | 0.89603 | 0.915348556 | 0.90161 | 0.999507308 | 0.337918 | 7.750867704 | 0 |
| 2 | 47476460 | C | A | A | E | MSH2 | ENST00000233146 | 0.6966 | 0.03484 | 0.74278 | 0.89134 | 0.96737 | 0.9904143272 | 0.94493 | 0.9997094274 | 0.474774 | 8.871343396 | 1 |
| 2 | 47476460 | C | G | A | G | MSH2 | ENST00000233146 | 0.70848 | 0.03697 | 0.65242 | 0.89537 | 0.95097 | 0.9226683291 | 0.91541 | 0.995079875 | 0.450911 | 5.334823085 | 0 |
| 2 | 47476460 | C | T | A | V | MSH2 | ENST00000233146 | 0.7031 | 0.03334 | 0.69749 | 0.84991 | 0.95753 | 0.9564192066 | 0.94451 | 0.9971014857 | 0.455851 | 6.211379578 | 0 |
| 2 | 47476462 | G | A | E | K | MSH2 | ENST00000233146 | 0.66494 | 0.02243 | 0.78669 | 0.75339 | 0.92682 | 0.8768487235 | 0.97849 | 0.7422119379 | 0.306392 | 4.286946736 | 0 |
| 2 | 47476462 | G | C | E | Q | MSH2 | ENST00000233146 | 0.60676 | 0.0142 | 0.6671 | 0.67303 | 0.88212 | 0.7827241161 | 0.96185 | 0.8474153876 | 0.228645 | 2.955643418 | 0 |
| 2 | 47476463 | A | C | E | A | MSH2 | ENST00000233146 | 0.93167 | 0.02023 | 0.67957 | 0.78329 | 0.91399 | 0.8527488073 | 0.90943 | 0.8869137764 | 0.130834 | 5.949091338 | 0 |
| 2 | 47476463 | A | G | E | G | MSH2 | ENST00000233146 | 0.94886 | 0.02775 | 0.63677 | 0.80593 | 0.93554 | 0.8927607958 | 0.90748 | 0.9903548956 | 0.31158 | 6.300749673 | 0 |
| 2 | 47476463 | A | T | E | V | MSH2 | ENST00000233146 | 0.94439 | 0.02984 | 0.66273 | 0.83763 | 0.94394 | 0.8530920365 | 0.95659 | 0.9879167676 | 0.326283 | 4.417951249 | 0 |
| 2 | 47476464 | A | C | E | D | MSH2 | ENST00000233146 | 0.42615 | 0.00909 | 0.68135 | 0.66548 | 0.83615 | 0.7800918792 | 0.97725 | 0.441318661 | 0.178097 | 2.915426628 | 0 |
| 2 | 47476464 | A | T | E | D | MSH2 | ENST00000233146 | 0.42109 | 0.00909 | 0.68135 | 0.6831 | 0.83615 | 0.7800918792 | 0.97746 | 0.441318661 | 0.178091 | 2.915426628 | 0 |
| 2 | 47476465 | G | A | V | M | MSH2 | ENST00000233146 | 0.64992 | 0.01401 | 0.51727 | 0.78649 | 0.82836 | 0.7168793883 | 0.8471 | 0.8154340982 | 0.0922686 | 3.639564324 | 0 |
| 2 | 47476465 | G | C | V | L | MSH2 | ENST00000233146 | 0.43968 | 0.00615 | 0.47921 | 0.47587 | 0.70014 | 0.6358968233 | 0.84603 | 0.1987267584 | 0.047106 | 1.809802787 | 0 |
| 2 | 47476465 | G | T | V | L | MSH2 | ENST00000233146 | 0.43478 | 0.00615 | 0.47921 | 0.48168 | 0.69762 | 0.6358968233 | 0.84562 | 0.1987267584 | 0.0471422 | 1.809802787 | 0 |
| 2 | 47476466 | T | A | V | E | MSH2 | ENST00000233146 | 0.81048 | 0.02714 | 0.48481 | 0.92356 | 0.9703 | 0.9503068503 | 0.93789 | 0.9933101751 | 0.398491 | 7.465415561 | 1 |
| 2 | 47476466 | T | C | V | A | MSH2 | ENST00000233146 | 0.73517 | 0.01533 | 0.50601 | 0.85545 | 0.91314 | 0.8286893916 | 0.86667 | 0.9605942408 | 0.229581 | 7.465415561 | 0 |
| 2 | 47476466 | T | G | V | G | MSH2 | ENST00000233146 | 0.80353 | 0.02696 | 0.37735 | 0.91331 | 0.97252 | 0.9595110242 | 0.90981 | 0.9926379051 | 0.408196 | 7.465415561 | 1 |
| 2 | 47476468 | T | A | S | T | MSH2 | ENST00000233146 | 0.43798 | 0.00791 | 0.53019 | 0.49069 | 0.78348 | 0.7886195786 | 0.91877 | 0.7367096543 | 0.146145 | 1.54637011 | 0 |
| 2 | 47476468 | T | C | S | P | MSH2 | ENST00000233146 | 0.70262 | 0.03189 | 0.54381 | 0.80583 | 0.92356 | 0.9444210964 | 0.94616 | 0.9420740604 | 0.297586 | 3.443628614 | 0 |
| 2 | 47476468 | T | G | S | A | MSH2 | ENST00000233146 | 0.54203 | 0.0144 | 0.50569 | 0.7391 | 0.81703 | 0.7520365914 | 0.91069 | 0.7243754864 | 0.144775 | 3.651866636 | 0 |
| 2 | 47476469 | C | G | S | C | MSH2 | ENST00000233146 | 0.51619 | 0.01037 | 0.53781 | 0.68539 | 0.79818 | 0.86377471 | 0.91176 | 0.5595413457 | 0.219967 | 3.322229042 | 0 |
| 2 | 47476469 | C | T | S | F | MSH2 | ENST00000233146 | 0.8495 | 0.03431 | 0.59028 | 0.89979 | 0.89963 | 0.9155169069 | 0.91261 | 0.9609058499 | 0.331002 | 6.433585944 | 0 |
| 2 | 47476471 | A | C | I | L | MSH2 | ENST00000233146 | 0.47457 | 0.00885 | 0.46761 | 0.61572 | 0.71861 | 0.5483871323 | 0.84557 | 0.5813964605 | 0.16187 | 2.321523643 | 0 |
| 2 | 47476471 | A | G | I | V | MSH2 | ENST00000233146 | 0.41682 | 0.00935 | 0.34362 | 0.41676 | 0.63418 | 0.3401870487 | 0.77936 | 0.3649833574 | 0.0336112 | 1.642293135 | 0 |
| 2 | 47476471 | A | T | I | F | MSH2 | ENST00000233146 | 0.85092 | 0.02891 | 0.48024 | 0.84942 | 0.9517 | 0.8009285148 | 0.90393 | 0.9933622333 | 0.306176 | 7.876082025 | 0 |
| 2 | 47476472 | T | C | I | T | MSH2 | ENST00000233146 | 0.90882 | 0.03053 | 0.63385 | 0.89709 | 0.96553 | 0.7673105553 | 0.89582 | 0.982956171 | 0.293299 | 5.961217638 | 0 |
| 2 | 47476472 | T | G | I | S | MSH2 | ENST00000233146 | 0.9392 | 0.03006 | 0.49516 | 0.91927 | 0.96846 | 0.895557445 | 0.8984 | 0.985857143 | 0.387226 | 7.876082025 | 0 |
| 2 | 47476473 | T | G | I | M | MSH2 | ENST00000233146 | 0.55141 | 0.03557 | 0.42926 | 0.80828 | 0.87781 | 0.6505856146 | 0.88123 | 0.9443529844 | 0.221202 | 4.596502282 | 0 |
| 2 | 47476474 | G | A | V | M | MSH2 | ENST00000233146 | 0.88958 | 0.0114 | 0.71216 | 0.79107 | 0.90096 | 0.7912207479 | 0.88179 | 0.8683417744 | 0.220901 | 5.698334754 | 0 |
| 2 | 47476474 | G | C | V | L | MSH2 | ENST00000233146 | 0.51619 | 0.00687 | 0.66706 | 0.72299 | 0.79577 | 0.7543259433 | 0.95557 | 0.6573503613 | 0.246727 | 5.060490396 | 0 |
| 2 | 47476474 | G | T | V | L | MSH2 | ENST00000233146 | 0.5108 | 0.00687 | 0.66706 | 0.72384 | 0.79577 | 0.7543259433 | 0.95637 | 0.6573503613 | 0.246722 | 5.060490396 | 0 |
| 2 | 47476475 | T | C | V | A | MSH2 | ENST00000233146 | 0.84241 | 0.01616 | 0.69405 | 0.75543 | 0.87099 | 0.8188217156 | 0.91801 | 0.9416363273 | 0.307662 | 4.756094399 | 0 |
| 2 | 47476475 | T | G | V | G | MSH2 | ENST00000233146 | 0.89243 | 0.02765 | 0.53391 | 0.90278 | 0.94728 | 0.9382244476 | 0.9366 | 0.9935293794 | 0.432801 | 7.94118974 | 0 |
| 2 | 47476477 | G | C | D | H | MSH2 | ENST00000233146 | 0.92956 | 0.03465 | 0.87847 | 0.91822 | 0.97885 | 0.9695669227 | 0.97931 | 0.9974933863 | 0.481783 | 9.643300417 | 0 |
| 2 | 47476477 | G | T | D | Y | MSH2 | ENST00000233146 | 0.93283 | 0.03409 | 0.86487 | 0.90482 | 0.97364 | 0.9819579343 | 0.98326 | 0.9998831749 | 0.488868 | 9.643300417 | 0 |
| 2 | 47476478 | A | C | D | A | MSH2 | ENST00000233146 | 0.91405 | 0.03388 | 0.80114 | 0.93797 | 0.9844 | 0.977432731 | 0.97456 | 0.9996567965 | 0.483653 | 9.643300417 | 0 |
| 2 | 47476478 | A | G | D | G | MSH2 | ENST00000233146 | 0.91763 | 0.03304 | 0.81095 | 0.94276 | 0.98549 | 0.9689249722 | 0.98071 | 0.9993323684 | 0.48556 | 9.643300417 | 0 |
| 2 | 47476478 | A | T | D | V | MSH2 | ENST00000233146 | 0.90906 | 0.0318 | 0.80027 | 0.95883 | 0.98109 | 0.9802903859 | 0.98357 | 0.9998342991 | 0.486595 | 9.643300417 | 0 |
| 2 | 47476480 | T | A | C | S | MSH2 | ENST00000233146 | 0.54082 | 0.00574 | 0.54011 | 0.53693 | 0.75687 | 0.657968762 | 0.76425 | 0.7994480729 | 0.120899 | 1.742196486 | 0 |
| 2 | 47476480 | T | C | C | R | MSH2 | ENST00000233146 | 0.83197 | 0.00633 | 0.71213 | 0.80523 | 0.83615 | 0.8446418544 | 0.87327 | 0.9793813825 | 0.192836 | 2.771212377 | 0 |
| 2 | 47476480 | T | G | C | G | MSH2 | ENST00000233146 | 0.83844 | 0.0058 | 0.50767 | 0.76209 | 0.88069 | 0.7374662586 | 0.8982 | 0.9669257402 | 0.257163 | 4.838369323 | 0 |
| 2 | 47476481 | G | A | C | Y | MSH2 | ENST00000233146 | 0.92566 | 0.0254 | 0.73727 | 0.83765 | 0.90007 | 0.802363732 | 0.87275 | 0.2946763337 | 0.0487755 | 6.31779877 | 0 |
| 2 | 47476481 | G | C | C | S | MSH2 | ENST00000233146 | 0.51783 | 0.00574 | 0.54011 | 0.61537 | 0.73223 | 0.657968762 | 0.68044 | 0.3652232232 | 0.0219168 | 1.742196486 | 0 |
| 2 | 47476481 | G | T | C | F | MSH2 | ENST00000233146 | 0.93457 | 0.02663 | 0.59538 | 0.86503 | 0.91611 | 0.793836432 | 0.8821 | 0.9895454049 | 0.37554 | 7.351727284 | 0 |
| 2 | 47476482 | C | G | C | W | MSH2 | ENST00000233146 | 0.71402 | 0.03112 | 0.65723 | 0.8959 | 0.8933 | 0.8380084848 | 0.90973 | 0.9678850174 | 0.337302 | 7.234473084 | 0 |
| 2 | 47476483 | A | C | I | L | MSH2 | ENST00000233146 | 0.78051 | 0.02283 | 0.80609 | 0.88107 | 0.91357 | 0.8648315405 | 0.8414 | 0.8877914478 | 0.223116 | 6.45932603 | 0 |
| 2 | 47476483 | A | G | I | V | MSH2 | ENST00000233146 | 0.54419 | 0.01048 | 0.69617 | 0.65601 | 0.79151 | 0.7328563359 | 0.74779 | 0.3778414452 | 0.0933797 | 2.991312152 | 0 |
| 2 | 47476483 | A | T | I | F | MSH2 | ENST00000233146 | 0.80716 | 0.02927 | 0.82585 | 0.94251 | 0.95826 | 0.9671759131 | 0.97257 | 0.9981088638 | 0.44594 | 8.917828296 | 0 |
| 2 | 47476484 | T | A | I | N | MSH2 | ENST00000233146 | 0.93813 | 0.02828 | 0.90392 | 0.94151 | 0.97215 | 0.9852530261 | 0.93871 | 0.9978600144 | 0.449062 | 8.917828296 | 1 |
| 2 | 47476484 | T | C | I | T | MSH2 | ENST00000233146 | 0.91347 | 0.03212 | 0.91999 | 0.91415 | 0.98693 | 0.9640374393 | 0.9715 | 0.9956348538 | 0.354435 | 8.917828296 | 0 |
| 2 | 47476484 | T | G | I | S | MSH2 | ENST00000233146 | 0.94073 | 0.03046 | 0.83766 | 0.93666 | 0.98549 | 0.9885916481 | 0.97339 | 0.9985123277 | 0.483236 | 8.917828296 | 0 |
| 2 | 47476485 | C | G | I | M | MSH2 | ENST00000233146 | 0.78193 | 0.03621 | 0.76421 | 0.92732 | 0.94091 | 0.9090606191 | 0.93873 | 0.9943648577 | 0.417532 | 8.917828296 | 0 |
| 2 | 47476486 | T | A | L | I | MSH2 | ENST00000233146 | 0.47778 | 0.03527 | 0.91401 | 0.82486 | 0.87148 | 0.7512049173 | 0.7964 | 0.9528326392 | 0.197488 | 7.319230647 | 0 |
| 2 | 47476486 | T | G | L | V | MSH2 | ENST00000233146 | 0.47526 | 0.03633 | 0.87996 | 0.92147 | 0.94802 | 0.8378317198 | 0.92736 | 0.996768117 | 0.381126 | 7.319230647 | 0 |
| 2 | 47476487 | T | C | L | S | MSH2 | ENST00000233146 | 0.90978 | 0.03189 | 0.92621 | 0.91508 | 0.98257 | 0.9661689062 | 0.95914 | 0.9977381229 | 0.464175 | 7.319230647 | 0 |
| 2 | 47476488 | A | C | L | F | MSH2 | ENST00000233146 | 0.80171 | 0.03582 | 0.90807 | 0.8744 | 0.85117 | 0.8463365867 | 0.92946 | 0.9805220366 | 0.337757 | 2.70037699 | 0 |
| 2 | 47476488 | A | T | L | F | MSH2 | ENST00000233146 | 0.795 | 0.03582 | 0.90807 | 0.8747 | 0.85117 | 0.8463365867 | 0.93034 | 0.9805220366 | 0.337751 | 2.70037699 | 0 |
| 2 | 47476489 | G | A | A | T | MSH2 | ENST00000233146 | 0.90391 | 0.03595 | 0.88063 | 0.85351 | 0.91694 | 0.8850774537 | 0.88643 | 0.9907826781 | 0.382081 | 3.005752727 | 0 |
| 2 | 47476489 | G | C | A | P | MSH2 | ENST00000233146 | 0.91934 | 0.03334 | 0.92144 | 0.89114 | 0.95863 | 0.9798205183 | 0.95335 | 0.9976515174 | 0.455126 | 8.085461809 | 0 |
| 2 | 47476489 | G | T | A | S | MSH2 | ENST00000233146 | 0.87925 | 0.03821 | 0.84649 | 0.82103 | 0.9182 | 0.8448195007 | 0.88461 | 0.982011795 | 0.334591 | 5.893955155 | 0 |
| 2 | 47476490 | C | A | A | D | MSH2 | ENST00000233146 | 0.72788 | 0.03106 | 0.9393 | 0.91441 | 0.96919 | 0.9770827815 | 0.95436 | 0.9987648726 | 0.457598 | 8.085461809 | 0 |
| 2 | 47476490 | C | G | A | G | MSH2 | ENST00000233146 | 0.66921 | 0.03706 | 0.86575 | 0.82342 | 0.92884 | 0.8720910514 | 0.90477 | 0.86968837 | 0.290458 | 8.085461809 | 0 |

| | | | | | | | | | | | | | | | | | | |
|---|---|---|---|---|---|---|---|---|---|---|---|---|---|---|---|---|---|---|
| 2 | 47476490 | C | T | A | V | MSH2 | ENST00000233146 | 0.72156 | 0.03702 | 0.88888 | 0.86175 | 0.94394 | 0.918438845 | 0.93332 | 0.9937717319 | 0.414623 | 8.020229924 | 0 |
| 2 | 47476492 | C | G | R | G | MSH2 | ENST00000233146 | 0.92537 | 0.0334 | 0.92791 | 0.93999 | 0.94949 | 0.9860005729 | 0.98583 | 0.9993390441 | 0.490601 | 9.986804952 | 0 |
| 2 | 47476493 | G | A | R | Q | MSH2 | ENST00000233146 | 0.89382 | 0.03509 | 0.8919 | 0.92378 | 0.97735 | 0.9648383351 | 0.98817 | 0.9997006655 | 0.453184 | 9.986804952 | 0 |
| 2 | 47476493 | G | C | R | P | MSH2 | ENST00000233146 | 0.90391 | 0.03175 | 0.95631 | 0.92963 | 0.98109 | 0.9910935284 | 0.98814 | 0.9994741082 | 0.524814 | 9.986804952 | 1 |
| 2 | 47476493 | G | T | R | L | MSH2 | ENST00000233146 | 0.89078 | 0.03394 | 0.94996 | 0.94118 | 0.98621 | 0.9785828328 | 0.99106 | 0.9997400641 | 0.523264 | 9.986804952 | 1 |
| 2 | 47476495 | G | A | V | I | MSH2 | ENST00000233146 | 0.73205 | 0.03608 | 0.77221 | 0.60531 | 0.82668 | 0.7428930652 | 0.84075 | 0.8967114091 | 0.207098 | 3.97460447 | 0 |
| 2 | 47476495 | G | C | V | L | MSH2 | ENST00000233146 | 0.90068 | 0.03582 | 0.89422 | 0.81777 | 0.94543 | 0.9085524128 | 0.9628 | 0.984408915 | 0.429728 | 9.77979123 | 0 |
| 2 | 47476495 | G | T | V | L | MSH2 | ENST00000233146 | 0.89617 | 0.03582 | 0.89422 | 0.81978 | 0.94543 | 0.9085524128 | 0.96309 | 0.984408915 | 0.429728 | 9.77979123 | 0 |
| 2 | 47476496 | T | A | V | E | MSH2 | ENST00000233146 | 0.8263 | 0.02989 | 0.89579 | 0.95995 | 0.9917 | 0.9832159321 | 0.98344 | 0.9997435212 | 0.478269 | 9.77979123 | 0 |
| 2 | 47476496 | T | C | V | A | MSH2 | ENST00000233146 | 0.79041 | 0.03404 | 0.90123 | 0.92591 | 0.98477 | 0.9409586315 | 0.95847 | 0.9991281629 | 0.469768 | 9.77979123 | 0 |
| 2 | 47476496 | T | G | V | G | MSH2 | ENST00000233146 | 0.82163 | 0.03313 | 0.81142 | 0.94993 | 0.99265 | 0.9827988487 | 0.97328 | 0.9989677668 | 0.478539 | 9.77979123 | 0 |
| 2 | 47476498 | G | A | G | R | MSH2 | ENST00000233146 | 0.94084 | 0.03452 | 0.97243 | 0.91748 | 0.95899 | 0.9767689672 | 0.97471 | 0.9998736382 | 0.498677 | 9.730645972 | 0 |
| 2 | 47476498 | G | C | G | R | MSH2 | ENST00000233146 | 0.93992 | 0.03452 | 0.97243 | 0.91565 | 0.95899 | 0.9767689672 | 0.97432 | 0.9998736382 | 0.498679 | 9.730645972 | 0 |
| 2 | 47476499 | G | A | G | E | MSH2 | ENST00000233146 | 0.90838 | 0.03334 | 0.96807 | 0.93019 | 0.96334 | 0.9788833856 | 0.9751 | 0.9998748302 | 0.430764 | 9.730645972 | 0 |
| 2 | 47476499 | G | C | G | A | MSH2 | ENST00000233146 | 0.88272 | 0.03608 | 0.93149 | 0.9186 | 0.95863 | 0.9658557391 | 0.97313 | 0.9997028708 | 0.494587 | 9.730645972 | 0 |
| 2 | 47476499 | G | T | G | V | MSH2 | ENST00000233146 | 0.90534 | 0.03379 | 0.93564 | 0.93034 | 0.96773 | 0.9808077499 | 0.97853 | 0.9998736382 | 0.498586 | 9.730645972 | 0 |
| 2 | 47476501 | G | A | A | T | MSH2 | ENST00000233146 | 0.92412 | 0.03753 | 0.76495 | 0.89619 | 0.94543 | 0.9445174937 | 0.9792 | 0.9879293442 | 0.464406 | 6.978422751 | 0 |
| 2 | 47476501 | G | C | A | P | MSH2 | ENST00000233146 | 0.9294 | 0.03334 | 0.83134 | 0.9002 | 0.97252 | 0.9788788136 | 0.97305 | 0.9975094795 | 0.486528 | 8.14288164 | 1 |
| 2 | 47476501 | G | T | A | S | MSH2 | ENST00000233146 | 0.899 | 0.03674 | 0.72327 | 0.84884 | 0.90711 | 0.8929324104 | 0.94402 | 0.9631525278 | 0.348332 | 4.432800612 | 0 |
| 2 | 47476502 | C | A | A | D | MSH2 | ENST00000233146 | 0.72941 | 0.03106 | 0.86879 | 0.93222 | 0.97698 | 0.9801123402 | 0.97594 | 0.9991355538 | 0.483973 | 8.14288164 | 0 |
| 2 | 47476502 | C | G | A | G | MSH2 | ENST00000233146 | 0.72075 | 0.03686 | 0.75037 | 0.91209 | 0.94839 | 0.8891109897 | 0.94148 | 0.9979943037 | 0.419126 | 8.14288164 | 0 |
| 2 | 47476502 | C | T | A | V | MSH2 | ENST00000233146 | 0.72364 | 0.03718 | 0.79527 | 0.92813 | 0.95863 | 0.9552373489 | 0.97197 | 0.9988414645 | 0.476231 | 8.14288164 | 0 |
| 2 | 47476504 | G | A | G | S | MSH2 | ENST00000233146 | 0.80471 | 0.03718 | 0.91056 | 0.82554 | 0.87491 | 0.8327150919 | 0.95952 | 0.9378755093 | 0.355226 | 3.733213873 | 0 |
| 2 | 47476504 | G | C | G | R | MSH2 | ENST00000233146 | 0.93996 | 0.03447 | 0.96004 | 0.82365 | 0.94015 | 0.9290188416 | 0.9617 | 0.9938107133 | 0.414709 | 6.646810274 | 0 |
| 2 | 47476504 | G | T | G | C | MSH2 | ENST00000233146 | 0.94694 | 0.03626 | 0.9382 | 0.83701 | 0.95426 | 0.9072880941 | 0.96316 | 0.9997061749 | 0.459492 | 8.240542013 | 0 |
| 2 | 47476505 | G | A | G | D | MSH2 | ENST00000233146 | 0.62294 | 0.03313 | 0.96111 | 0.77114 | 0.90755 | 0.9095471497 | 0.95782 | 0.983897388 | 0.393966 | 8.240542013 | 0 |
| 2 | 47476505 | G | C | G | A | MSH2 | ENST00000233146 | 0.78899 | 0.03595 | 0.9117 | 0.81778 | 0.92151 | 0.8511060606 | 0.95297 | 0.9959301353 | 0.390291 | 3.801372474 | 0 |
| 2 | 47476505 | G | T | G | V | MSH2 | ENST00000233146 | 0.83414 | 0.03379 | 0.91922 | 0.85275 | 0.95023 | 0.9011852399 | 0.95777 | 0.9970170259 | 0.437454 | 7.239944719 | 0 |
| 2 | 47476507 | G | A | D | N | MSH2 | ENST00000233146 | 0.93712 | 0.03753 | 0.81913 | 0.93033 | 0.92438 | 0.9639459032 | 0.97966 | 0.9993035793 | 0.23175 | 9.986802395 | 0 |
| 2 | 47476507 | G | C | D | H | MSH2 | ENST00000233146 | 0.93533 | 0.03465 | 0.84602 | 0.91212 | 0.99006 | 0.9744958927 | 0.98969 | 0.9996045232 | 0.531539 | 9.986802395 | 0 |
| 2 | 47476507 | G | T | D | Y | MSH2 | ENST00000233146 | 0.93805 | 0.03409 | 0.82525 | 0.92295 | 0.98549 | 0.9853741699 | 0.99224 | 0.9998831749 | 0.532344 | 9.986802395 | 0 |
| 2 | 47476508 | A | C | D | A | MSH2 | ENST00000233146 | 0.92229 | 0.03394 | 0.75799 | 0.94217 | 0.99072 | 0.9814634853 | 0.98373 | 0.9998342991 | 0.52892 | 9.986802395 | 0 |
| 2 | 47476508 | A | G | D | G | MSH2 | ENST00000233146 | 0.92537 | 0.03313 | 0.76857 | 0.93937 | 0.99386 | 0.9736432866 | 0.98949 | 0.9996801615 | 0.529557 | 9.986802395 | 0 |
| 2 | 47476508 | A | T | D | V | MSH2 | ENST00000233146 | 0.91793 | 0.03189 | 0.7531 | 0.95978 | 0.99386 | 0.982763168 | 0.99016 | 0.9998342991 | 0.531096 | 9.986802395 | 0 |
| 2 | 47476510 | A | C | S | R | MSH2 | ENST00000233146 | 0.93114 | 0.03001 | 0.8119 | 0.82317 | 0.9648 | 0.9170622511 | 0.97436 | 0.9934971333 | 0.396253 | 5.899077867 | 0 |
| 2 | 47476510 | A | G | S | G | MSH2 | ENST00000233146 | 0.93451 | 0.01812 | 0.7105 | 0.81792 | 0.93475 | 0.819394687 | 0.93901 | 0.9493832588 | 0.205212 | 7.48702802 | 0 |
| 2 | 47476510 | A | T | S | C | MSH2 | ENST00000233146 | 0.67176 | 0.00627 | 0.75681 | 0.76245 | 0.83997 | 0.8451231658 | 0.883 | 0.873370111 | 0.176205 | 5.13495189 | 0 |
| 2 | 47476511 | G | A | S | N | MSH2 | ENST00000233146 | 0.5807 | 0.00735 | 0.80013 | 0.65585 | 0.78348 | 0.8414450877 | 0.88042 | 0.7677489122 | 0.13392 | 3.345083217 | 0 |
| 2 | 47476511 | G | C | S | T | MSH2 | ENST00000233146 | 0.87755 | 0.02396 | 0.75893 | 0.78617 | 0.89784 | 0.8156082034 | 0.942 | 0.8984856606 | 0.258266 | 6.515171851 | 0 |
| 2 | 47476511 | G | T | S | I | MSH2 | ENST00000233146 | 0.92663 | 0.02567 | 0.78493 | 0.88946 | 0.93977 | 0.9089043041 | 0.96818 | 0.992649138 | 0.432081 | 7.48702802 | 0 |
| 2 | 47476512 | T | A | S | R | MSH2 | ENST00000233146 | 0.77227 | 0.03001 | 0.8119 | 0.84547 | 0.93044 | 0.9170622511 | 0.97674 | 0.9905691147 | 0.366636 | 5.899077867 | 0 |
| 2 | 47476512 | T | G | S | R | MSH2 | ENST00000233146 | 0.78043 | 0.03001 | 0.8119 | 0.84823 | 0.93044 | 0.9170622511 | 0.97665 | 0.9905691147 | 0.366639 | 5.899077867 | 0 |
| 2 | 47476513 | C | A | Q | K | MSH2 | ENST00000233146 | 0.85756 | 0.03112 | 0.8349 | 0.81673 | 0.90624 | 0.9338929891 | 0.94997 | 0.9956052899 | 0.418229 | 8.145063867 | 0 |
| 2 | 47476513 | C | G | Q | E | MSH2 | ENST00000233146 | 0.8199 | 0.03465 | 0.78247 | 0.75772 | 0.91357 | 0.9354569379 | 0.94206 | 0.9874973297 | 0.384046 | 8.145063867 | 0 |
| 2 | 47476514 | A | C | Q | P | MSH2 | ENST00000233146 | 0.94001 | 0.03409 | 0.78525 | 0.8849 | 0.95972 | 0.9618650861 | 0.95586 | 0.9969352484 | 0.430192 | 8.145063867 | 0 |
| 2 | 47476514 | A | G | Q | R | MSH2 | ENST00000233146 | 0.93454 | 0.03547 | 0.82908 | 0.84219 | 0.94091 | 0.9419344745 | 0.9617 | 0.9955843091 | 0.4394 | 8.145063867 | 0 |
| 2 | 47476514 | A | T | Q | L | MSH2 | ENST00000233146 | 0.938 | 0.03718 | 0.77073 | 0.87396 | 0.9367 | 0.8805700701 | 0.95275 | 0.9957685471 | 0.357614 | 3.531392633 | 0 |
| 2 | 47476515 | A | C | Q | H | MSH2 | ENST00000233146 | 0.5106 | 0.03076 | 0.82649 | 0.87656 | 0.87877 | 0.9190077127 | 0.95812 | 0.9960330129 | 0.321368 | 8.145063867 | 0 |
| 2 | 47476515 | A | T | Q | H | MSH2 | ENST00000233146 | 0.50505 | 0.03076 | 0.82649 | 0.87681 | 0.87877 | 0.9190077127 | 0.95818 | 0.9960330129 | 0.321363 | 8.145063867 | 0 |
| 2 | 47476516 | T | A | L | M | MSH2 | ENST00000233146 | 0.3666 | 0.0099 | 0.7462 | 0.70741 | 0.84694 | 0.634894416 | 0.9062 | 0.7699735761 | 0.10621 | 6.730772769 | 0 |
| 2 | 47476516 | T | G | L | V | MSH2 | ENST00000233146 | 0.33105 | 0.00654 | 0.70829 | 0.61794 | 0.86352 | 0.6722718541 | 0.97818 | 0.2030360997 | 0.152734 | 2.755295462 | 0 |
| 2 | 47476517 | T | C | L | S | MSH2 | ENST00000233146 | 0.62726 | 0.02133 | 0.79764 | 0.82328 | 0.88963 | 0.8271514976 | 0.90065 | 0.9738000631 | 0.260779 | 3.528974978 | 0 |
| 2 | 47476517 | T | G | L | W | MSH2 | ENST00000233146 | 0.81387 | 0.02796 | 0.85366 | 0.87201 | 0.92924 | 0.8375702629 | 0.87691 | 0.9916808009 | 0.283005 | 7.88285917 | 0 |
| 2 | 47476518 | G | C | L | F | MSH2 | ENST00000233146 | 0.61268 | 0.02663 | 0.76079 | 0.82145 | 0.88776 | 0.7116867994 | 0.89736 | 0.9172460135 | 0.19113 | 5.204043762 | 0 |
| 2 | 47476518 | G | T | L | F | MSH2 | ENST00000233146 | 0.60669 | 0.02663 | 0.76079 | 0.82803 | 0.88776 | 0.7116867994 | 0.89727 | 0.9321584702 | 0.191131 | 5.204043762 | 0 |
| 2 | 47476519 | A | C | K | Q | MSH2 | ENST00000233146 | 0.87834 | 0.03362 | 0.82823 | 0.75314 | 0.89963 | 0.8422148335 | 0.92176 | 0.9462745786 | 0.252594 | 4.225069155 | 0 |
| 2 | 47476519 | A | G | K | E | MSH2 | ENST00000233146 | 0.906 | 0.03161 | 0.86037 | 0.88656 | 0.96153 | 0.9091192561 | 0.9316 | 0.9242804646 | 0.20462 | 7.004346951 | 0 |
| 2 | 47476520 | A | C | K | T | MSH2 | ENST00000233146 | 0.89042 | 0.03442 | 0.8701 | 0.87445 | 0.95972 | 0.9017180274 | 0.92787 | 0.9933331609 | 0.369877 | 4.22268462 | 0 |
| 2 | 47476520 | A | G | K | R | MSH2 | ENST00000233146 | 0.87423 | 0.03761 | 0.79624 | 0.72744 | 0.87099 | 0.7832893895 | 0.91558 | 0.9469355941 | 0.212767 | 3.68825001 | 0 |
| 2 | 47476520 | A | T | K | I | MSH2 | ENST00000233146 | 0.91678 | 0.02857 | 0.83897 | 0.90665 | 0.94949 | 0.9036533811 | 0.89568 | 0.9944972396 | 0.340697 | 8.45919737 | 0 |
| 2 | 47476521 | A | C | K | N | MSH2 | ENST00000233146 | 0.77766 | 0.03465 | 0.8828 | 0.87695 | 0.89147 | 0.8925986419 | 0.89566 | 0.9901870489 | 0.116229 | 7.301756924 | 0 |
| 2 | 47476521 | A | T | K | N | MSH2 | ENST00000233146 | 0.77101 | 0.03465 | 0.8828 | 0.874 | 0.89147 | 0.8925986419 | 0.89616 | 0.9901870489 | 0.116222 | 7.301756924 | 0 |
| 2 | 47476522 | G | A | G | R | MSH2 | ENST00000233146 | 0.93488 | 0.03452 | 0.97494 | 0.94684 | 0.96956 | 0.9516876444 | 0.9738 | 0.9998736382 | 0.480424 | 8.147544014 | 0 |
| 2 | 47476522 | G | C | G | R | MSH2 | ENST00000233146 | 0.93371 | 0.03452 | 0.97494 | 0.94321 | 0.96919 | 0.9516876444 | 0.97331 | 0.9998736382 | 0.480426 | 8.147544014 | 0 |
| 2 | 47476523 | G | A | G | E | MSH2 | ENST00000233146 | 0.81897 | 0.03334 | 0.96864 | 0.94621 | 0.9659 | 0.9462471631 | 0.9589 | 0.9992679954 | 0.476949 | 9.57016555 | 0 |
| 2 | 47476523 | G | C | G | A | MSH2 | ENST00000233146 | 0.78304 | 0.03608 | 0.93171 | 0.89726 | 0.94986 | 0.9234547102 | 0.96925 | 0.9996370077 | 0.476036 | 9.57016555 | 0 |
| 2 | 47476523 | G | T | G | V | MSH2 | ENST00000233146 | 0.81426 | 0.03379 | 0.94247 | 0.9248 | 0.96553 | 0.9581390435 | 0.96287 | 0.9998736382 | 0.480575 | 9.57016555 | 0 |
| 2 | 47476525 | G | A | V | I | MSH2 | ENST00000233146 | 0.70222 | 0.02083 | 0.73464 | 0.58553 | 0.78411 | 0.6945774459 | 0.82077 | 0.2553777797 | 0.0543924 | 4.484132169 | 0 |
| 2 | 47476525 | G | C | V | L | MSH2 | ENST00000233146 | 0.68627 | 0.03281 | 0.87546 | 0.6339 | 0.85378 | 0.8569690096 | 0.8894 | 0.9256662726 | 0.316729 | 6.204042136 | 0 |
| 2 | 47476525 | G | T | V | F | MSH2 | ENST00000233146 | 0.91036 | 0.03265 | 0.85676 | 0.88183 | 0.89513 | 0.946212887 | 0.9407 | 0.9912624359 | 0.442826 | 7.708927574 | 0 |
| 2 | 47476526 | T | A | V | D | MSH2 | ENST00000233146 | 0.81111 | 0.02989 | 0.89002 | 0.93167 | 0.95753 | 0.961228341 | 0.96027 | 0.9932668209 | 0.433644 | 5.541714297 | 0 |
| 2 | 47476526 | T | C | V | A | MSH2 | ENST00000233146 | 0.76411 | 0.03379 | 0.88496 | 0.87981 | 0.93436 | 0.8870090891 | 0.913 | 0.9849204421 | 0.381615 | 7.708927574 | 0 |
| 2 | 47476526 | T | G | V | G | MSH2 | ENST00000233146 | 0.79626 | 0.03304 | 0.79021 | 0.9202 | 0.96262 | 0.9428293301 | 0.93034 | 0.991832261 | 0.447129 | 7.708927574 | 0 |
| 2 | 47476528 | T | A | S | T | MSH2 | ENST00000233146 | 0.81716 | 0.03857 | 0.90077 | 0.90523 | 0.96444 | 0.9730262066 | 0.9769 | 0.9993405938 | 0.497449 | 9.986807702 | 0 |
| 2 | 47476528 | T | C | S | P | MSH2 | ENST00000233146 | 0.89927 | 0.0337 | 0.91205 | 0.91291 | 0.97401 | 0.9862125367 | 0.97835 | 0.9996001124 | 0.501263 | 9.986807702 | 0 |
| 2 | 47476528 | T | G | S | A | MSH2 | ENST00000233146 | 0.85778 | 0.0399 | 0.88965 | 0.92121 | 0.9608 | 0.9607779869 | 0.94942 | 0.9993405938 | 0.471288 | 9.986807702 | 0 |
| 2 | 47476529 | C | A | S | Y | MSH2 | ENST00000233146 | 0.79382 | 0.03346 | 0.94005 | 0.92831 | 0.96444 | 0.9870305499 | 0.97925 | 0.9996891022 | 0.501291 | 9.986807702 | 1 |
| 2 | 47476529 | C | G | S | C | MSH2 | ENST00000233146 | 0.82553 | 0.03761 | 0.90524 | 0.90006 | 0.94986 | 0.9805853753 | 0.95661 | 0.999706924 | 0.500936 | 9.986807702 | 0 |
| 2 | 47476529 | C | T | S | F | MSH2 | ENST00000233146 | 0.84271 | 0.03582 | 0.92496 | 0.92831 | 0.96044 | 0.9894963776 | 0.98295 | 0.9996788502 | 0.501288 | 9.986807702 | 1 |
| 2 | 47476531 | A | C | T | P | MSH2 | ENST00000233146 | 0.93649 | 0.03304 | 0.63512 | 0.91957 | 0.96262 | 0.9839287208 | 0.97057 | 0.9986627102 | 0.489656 | 9.986806179 | 1 |
| 2 | 47476531 | A | G | T | A | MSH2 | ENST00000233146 | 0.92096 | 0.03534 | 0.64416 | 0.9086 | 0.9648 | 0.9707078254 | 0.9761 | 0.9918090701 | 0.478752 | 9.986806179 | 0 |
| 2 | 47476531 | A | T | T | S | MSH2 | ENST00000233146 | 0.89774 | 0.03189 | 0.65228 | 0.89113 | 0.9458 | 0.9283802374 | 0.9374 | 0.992320478 | 0.419703 | 9.986806179 | 0 |
| 2 | 47476532 | C | A | T | K | MSH2 | ENST00000233146 | 0.62369 | 0.03409 | 0.77319 | 0.88591 | 0.90668 | 0.9888011744 | 0.94964 | 0.9996327162 | 0.467395 | 9.986806179 | 0 |
| 2 | 47476532 | C | G | T | R | MSH2 | ENST00000233146 | 0.61179 | 0.03509 | 0.75318 | 0.91869 | 0.9211 | 0.991665825 | 0.9635 | 0.9986195564 | 0.468352 | 9.986806179 | 0 |
| 2 | 47476534 | T | A | F | I | MSH2 | ENST00000233146 | 0.89423 | 0.03224 | 0.95341 | 0.9125 | 0.97773 | 0.9810540174 | 0.97402 | 0.999584496 | 0.512964 | 9.63699004 | 0 |
| 2 | 47476534 | T | C | F | L | MSH2 | ENST00000233146 | 0.90799 | 0.03491 | 0.96974 | 0.91694 | 0.9833 | 0.9666977828 | 0.97503 | 0.9995598197 | 0.507656 | 8.469465247 | 0 |
| 2 | 47476534 | T | G | F | V | MSH2 | ENST00000233146 | 0.89884 | 0.03313 | 0.94727 | 0.92558 | 0.9796 | 0.9756236716 | 0.95494 | 0.9994020462 | 0.513236 | 9.63699004 | 0 |
| 2 | 47476535 | T | A | F | Y | MSH2 | ENST00000233146 | 0.88454 | 0.03527 | 0.96182 | 0.93441 | 0.9579 | 0.9454946725 | 0.93756 | 0.9993417859 | 0.451998 | 9.63699004 | 0 |
| 2 | 47476535 | T | C | F | S | MSH2 | ENST00000233146 | 0.93426 | 0.02864 | 0.95445 | 0.94783 | 0.98034 | 0.9857512621 | 0.9697 | 0.9998125434 | 0.51624 | 9.63699004 | 0 |
| 2 | 47476535 | T | G | F | C | MSH2 | ENST00000233146 | 0.93403 | 0.03175 | 0.96668 | 0.93403 | 0.97698 | 0.9840680148 | 0.96542 | 0.9998125434 | 0.517853 | 9.63699004 | 0 |
| 2 | 47476536 | C | A | F | L | MSH2 | ENST00000233146 | 0.63443 | 0.03491 | 0.96974 | 0.88029 | 0.9681 | 0.9666977828 | 0.98136 | 0.9988822341 | 0.4627 | 8.469465247 | 0 |
| 2 | 47476536 | C | G | F | L | MSH2 | ENST00000233146 | 0.6426 | 0.03491 | 0.96974 | 0.88004 | 0.9681 | 0.9666977828 | 0.98118 | 0.9988822341 | 0.462703 | 8.469465247 | 0 |
| 2 | 47476537 | A | C | M | L | MSH2 | ENST00000233146 | 0.82887 | 0.03815 | 0.83291 | 0.82503 | 0.94205 | 0.9451004353 | 0.95314 | 0.9749728396 | 0.41509 | 5.386537916 | 0 |
| 2 | 47476537 | A | G | M | V | MSH2 | ENST00000233146 | 0.78851 | 0.03686 | 0.77865 | 0.93288 | 0.95207 | 0.9733542464 | 0.94146 | 0.9989251494 | 0.435828 | 9.225227557 | 0 |
| 2 | 47476537 | A | T | M | L | MSH2 | ENST00000233146 | 0.82233 | 0.03815 | 0.83291 | 0.82568 | 0.94205 | 0.9451004353 | 0.95331 | 0.9714458585 | 0.41509 | 5.386537916 | 0 |
| 2 | 47476538 | T | A | M | K | MSH2 | ENST00000233146 | 0.88361 | 0.03065 | 0.89022 | 0.92491 | 0.96008 | 0.9950625471 | 0.95306 | 0.9965253472 | 0.462151 | 9.225227557 | 0 |
| 2 | 47476538 | T | C | M | T | MSH2 | ENST00000233146 | 0.84073 | 0.03503 | 0.91938 | 0.94316 | 0.96334 | 0.9847745928 | 0.93767 | 0.9994931221 | 0.464768 | 9.225227557 | 0 |
| 2 | 47476538 | T | G | M | R | MSH2 | ENST00000233146 | 0.89645 | 0.03093 | 0.87853 | 0.94652 | 0.96153 | 0.995509004 | 0.95129 | 0.9996478558 | 0.46524 | 9.225227557 | 0 |
| 2 | 47476540 | G | A | A | T | MSH2 | ENST00000233146 | 0.81951 | 0.03274 | 0.73988 | 0.63785 | 0.84375 | 0.8086629749 | 0.8877 | 0.9202910066 | 0.34498 | 7.02571254 | 0 |
| 2 | 47476540 | G | C | A | P | MSH2 | ENST00000233146 | 0.94102 | 0.03212 | 0.8088 | 0.8077 | 0.85064 | 0.962056655 | 0.93154 | 0.9700189254 | 0.424852 | 7.02571254 | 0 |
| 2 | 47476540 | G | T | A | S | MSH2 | ENST00000233146 | 0.60383 | 0.01445 | 0.70127 | 0.42647 | 0.71391 | 0.7533332169 | 0.71343 | 0.8155581951 | 0.133929 | 2.410113055 | 0 |

| | | | | | | | | | | | | | | | | | |
|---|---|---|---|---|---|---|---|---|---|---|---|---|---|---|---|---|---|
| 2 | 47476541 | C | A | A | D | MSH2 | ENST00000233146 | 0.71962 | 0.02914 | 0.86293 | 0.85442 | 0.90185 | 0.9638073528 | 0.94921 | 0.9903370142 | 0.442581 | 7.02571254 | 0 |
| 2 | 47476541 | C | G | A | G | MSH2 | ENST00000233146 | 0.69885 | 0.02979 | 0.72905 | 0.69251 | 0.825 | 0.7904663982 | 0.80102 | 0.9707087874 | 0.26154 | 7.02571254 | 0 |
| 2 | 47476541 | C | T | A | V | MSH2 | ENST00000233146 | 0.41801 | 0.02017 | 0.77912 | 0.69342 | 0.86047 | 0.828813028 | 0.90587 | 0.9541061521 | 0.357871 | 3.054234619 | 0 |
| 2 | 47476543 | G | A | E | K | MSH2 | ENST00000233146 | 0.9427 | 0.03542 | 0.91868 | 0.9114 | 0.96553 | 0.9970041665 | 0.98428 | 0.9997082353 | 0.512364 | 9.98679641 | 1 |
| 2 | 47476543 | G | C | E | Q | MSH2 | ENST00000233146 | 0.91772 | 0.03735 | 0.83703 | 0.92246 | 0.95572 | 0.9869166359 | 0.98712 | 0.9995468259 | 0.513631 | 9.98679641 | 0 |
| 2 | 47476544 | A | C | E | A | MSH2 | ENST00000233146 | 0.91185 | 0.03626 | 0.84489 | 0.92311 | 0.97624 | 0.9927189787 | 0.96846 | 0.9993540645 | 0.514549 | 9.98679641 | 0 |
| 2 | 47476544 | A | G | E | G | MSH2 | ENST00000233146 | 0.93406 | 0.03484 | 0.80707 | 0.92906 | 0.98183 | 0.9923725958 | 0.97342 | 0.9994868167 | 0.515736 | 9.98679641 | 0 |
| 2 | 47476544 | A | T | E | V | MSH2 | ENST00000233146 | 0.92677 | 0.03484 | 0.82678 | 0.93471 | 0.9822 | 0.9915362534 | 0.98503 | 0.9990704656 | 0.516639 | 9.98679641 | 0 |
| 2 | 47476546 | A | C | M | L | MSH2 | ENST00000233146 | 0.84965 | 0.03815 | 0.92613 | 0.87263 | 0.93554 | 0.9644388236 | 0.93321 | 0.9696737528 | 0.379642 | 8.298784233 | 0 |
| 2 | 47476546 | A | G | M | V | MSH2 | ENST00000233146 | 0.80708 | 0.03686 | 0.88971 | 0.90901 | 0.95316 | 0.9809796125 | 0.96207 | 0.9920176268 | 0.471283 | 9.445632176 | 0 |
| 2 | 47476546 | A | T | M | L | MSH2 | ENST00000233146 | 0.84332 | 0.03815 | 0.92613 | 0.87263 | 0.93554 | 0.9644388236 | 0.93347 | 0.9696737528 | 0.379642 | 8.298784233 | 0 |
| 2 | 47476547 | T | A | M | K | MSH2 | ENST00000233146 | 0.8969 | 0.03065 | 0.96067 | 0.95295 | 0.96371 | 0.9975603098 | 0.95232 | 0.9996478558 | 0.473958 | 9.445632176 | 1 |
| 2 | 47476547 | T | C | M | T | MSH2 | ENST00000233146 | 0.85742 | 0.03503 | 0.97479 | 0.9445 | 0.96993 | 0.989418282 | 0.9385 | 0.999584496 | 0.472354 | 9.445632176 | 0 |
| 2 | 47476547 | T | G | M | R | MSH2 | ENST00000233146 | 0.90814 | 0.03093 | 0.95427 | 0.93808 | 0.96444 | 0.997933312 | 0.94842 | 0.9996478558 | 0.474484 | 9.445632176 | 1 |
| 2 | 47476548 | G | A | M | I | MSH2 | ENST00000233146 | 0.90116 | 0.03563 | 0.92816 | 0.86114 | 0.89829 | 0.9812010507 | 0.95045 | 0.994489789 | 0.456487 | 9.445632176 | 0 |
| 2 | 47476548 | G | C | M | I | MSH2 | ENST00000233146 | 0.89895 | 0.03563 | 0.92816 | 0.86169 | 0.89873 | 0.9812010507 | 0.94995 | 0.994489789 | 0.456487 | 9.445632176 | 0 |
| 2 | 47476548 | G | T | M | I | MSH2 | ENST00000233146 | 0.8944 | 0.03563 | 0.92816 | 0.86114 | 0.89873 | 0.9812010507 | 0.9501 | 0.994489789 | 0.456487 | 9.445632176 | 0 |
| 2 | 47476549 | T | A | L | M | MSH2 | ENST00000233146 | 0.65101 | 0.03753 | 0.90664 | 0.86208 | 0.86352 | 0.8301832459 | 0.81676 | 0.9272450805 | 0.210589 | 5.406526934 | 0 |
| 2 | 47476549 | T | G | L | V | MSH2 | ENST00000233146 | 0.4819 | 0.03587 | 0.89085 | 0.81463 | 0.89101 | 0.8970508586 | 0.8782 | 0.9372064471 | 0.287809 | 6.173531512 | 0 |
| 2 | 47476550 | T | C | L | S | MSH2 | ENST00000233146 | 0.92379 | 0.03224 | 0.93452 | 0.92041 | 0.9517 | 0.9552747648 | 0.95486 | 0.9976174235 | 0.423959 | 4.743979931 | 0 |
| 2 | 47476551 | G | C | L | F | MSH2 | ENST00000233146 | 0.6475 | 0.03582 | 0.91487 | 0.92322 | 0.95535 | 0.913800196 | 0.90843 | 0.9836059809 | 0.357377 | 7.875317552 | 0 |
| 2 | 47476551 | G | T | L | F | MSH2 | ENST00000233146 | 0.64112 | 0.03582 | 0.91487 | 0.92344 | 0.95535 | 0.913800196 | 0.90913 | 0.9844361507 | 0.357374 | 7.875317552 | 0 |
| 2 | 47476552 | G | C | E | Q | MSH2 | ENST00000233146 | 0.94284 | 0.03735 | 0.8313 | 0.89487 | 0.91229 | 0.9747696471 | 0.97153 | 0.9937927723 | 0.47798 | 9.986805547 | 0 |
| 2 | 47476553 | A | C | E | A | MSH2 | ENST00000233146 | 0.93938 | 0.03626 | 0.83716 | 0.94555 | 0.97624 | 0.9862765031 | 0.97087 | 0.9993540645 | 0.498216 | 9.986805547 | 0 |
| 2 | 47476553 | A | G | E | G | MSH2 | ENST00000233146 | 0.94946 | 0.03484 | 0.80303 | 0.95256 | 0.96919 | 0.9844667276 | 0.96661 | 0.9996173382 | 0.498875 | 9.986805547 | 0 |
| 2 | 47476553 | A | T | E | V | MSH2 | ENST00000233146 | 0.94644 | 0.03484 | 0.82166 | 0.95679 | 0.9729 | 0.9841853027 | 0.98323 | 0.9991241097 | 0.500027 | 9.986805547 | 0 |
| 2 | 47476554 | A | C | E | D | MSH2 | ENST00000233146 | 0.69804 | 0.03864 | 0.83863 | 0.92094 | 0.97401 | 0.969758106 | 0.9795 | 0.9891282916 | 0.421923 | 9.986805547 | 0 |
| 2 | 47476554 | A | T | E | D | MSH2 | ENST00000233146 | 0.69123 | 0.03864 | 0.83863 | 0.92332 | 0.97401 | 0.969758106 | 0.97967 | 0.9891282916 | 0.421916 | 9.986805547 | 0 |
| 2 | 47476555 | A | C | T | P | MSH2 | ENST00000233146 | 0.92401 | 0.03327 | 0.69095 | 0.94299 | 0.96956 | 0.9861018038 | 0.94192 | 0.9992663264 | 0.466693 | 9.613711236 | 1 |
| 2 | 47476555 | A | G | T | A | MSH2 | ENST00000233146 | 0.90302 | 0.03747 | 0.70109 | 0.8821 | 0.95133 | 0.9457851312 | 0.93289 | 0.9947270751 | 0.368149 | 5.075645387 | 0 |
| 2 | 47476555 | A | T | T | S | MSH2 | ENST00000233146 | 0.87261 | 0.03882 | 0.71742 | 0.9244 | 0.93475 | 0.898147088 | 0.90637 | 0.9860420823 | 0.367015 | 6.51608039 | 0 |
| 2 | 47476556 | C | G | T | S | MSH2 | ENST00000233146 | 0.68715 | 0.03882 | 0.71742 | 0.77272 | 0.84375 | 0.898147088 | 0.88407 | 0.9814507961 | 0.360017 | 6.51608039 | 0 |
| 2 | 47476556 | C | T | T | I | MSH2 | ENST00000233146 | 0.72957 | 0.03608 | 0.83533 | 0.79486 | 0.89963 | 0.9648236414 | 0.89145 | 0.9929805398 | 0.408486 | 7.529992328 | 0 |
| 2 | 47476558 | G | A | A | T | MSH2 | ENST00000233146 | 0.94752 | 0.03261 | 0.75591 | 0.83432 | 0.95462 | 0.9292561165 | 0.91849 | 0.9466522932 | 0.140667 | 7.26027107 | 0 |
| 2 | 47476558 | G | C | A | P | MSH2 | ENST00000233146 | 0.94989 | 0.03166 | 0.82682 | 0.87628 | 0.97923 | 0.988766196 | 0.92182 | 0.9981880784 | 0.446263 | 8.905925228 | 1 |
| 2 | 47476558 | G | T | A | S | MSH2 | ENST00000233146 | 0.64579 | 0.01096 | 0.71852 | 0.5674 | 0.78967 | 0.858945456 | 0.74003 | 0.7247504592 | 0.107257 | 2.544285414 | 0 |
| 2 | 47476559 | C | A | A | D | MSH2 | ENST00000233146 | 0.75615 | 0.02819 | 0.87007 | 0.91901 | 0.91058 | 0.9887531411 | 0.91435 | 0.9994182587 | 0.463965 | 7.911476545 | 0 |
| 2 | 47476559 | C | G | A | G | MSH2 | ENST00000233146 | 0.74069 | 0.02951 | 0.74818 | 0.75436 | 0.82103 | 0.8912957781 | 0.8538 | 0.9827711889 | 0.276209 | 7.813587943 | 0 |
| 2 | 47476559 | C | T | A | V | MSH2 | ENST00000233146 | 0.73629 | 0.03353 | 0.79244 | 0.87419 | 0.85222 | 0.9600619206 | 0.9011 | 0.9965077043 | 0.393295 | 8.905925228 | 0 |
| 2 | 47476561 | T | A | S | T | MSH2 | ENST00000233146 | 0.52354 | 0.00735 | 0.60356 | 0.76154 | 0.79334 | 0.8089138557 | 0.9189 | 0.8601224422 | 0.148853 | 4.440566538 | 0 |
| 2 | 47476561 | T | C | S | P | MSH2 | ENST00000233146 | 0.92295 | 0.02255 | 0.62528 | 0.90904 | 0.9458 | 0.9665426108 | 0.9502 | 0.9732974172 | 0.361262 | 7.932907865 | 0 |
| 2 | 47476561 | T | G | S | A | MSH2 | ENST00000233146 | 0.51356 | 0.00619 | 0.57416 | 0.67896 | 0.61416 | 0.7043330532 | 0.86141 | 0.6799814105 | 0.05889 | 2.000810951 | 0 |
| 2 | 47476562 | C | A | S | Y | MSH2 | ENST00000233146 | 0.40916 | 0.00648 | 0.6894 | 0.79837 | 0.86703 | 0.9198409154 | 0.92862 | 0.9624795318 | 0.239127 | 4.219492669 | 0 |
| 2 | 47476562 | C | G | S | C | MSH2 | ENST00000233146 | 0.74682 | 0.02943 | 0.60782 | 0.91263 | 0.911 | 0.8235248268 | 0.78637 | 0.9764529974 | 0.226539 | 7.932907865 | 0 |
| 2 | 47476562 | C | T | S | F | MSH2 | ENST00000233146 | 0.74611 | 0.00681 | 0.66665 | 0.85148 | 0.92028 | 0.9128889804 | 0.91262 | 0.981097579 | 0.2511 | 7.627198402 | 0 |
| 2 | 47476564 | A | C | I | L | MSH2 | ENST00000233146 | 0.8533 | 0.0357 | 0.80484 | 0.8823 | 0.90929 | 0.9484057452 | 0.84487 | 0.9693627954 | 0.278894 | 8.139510332 | 0 |
| 2 | 47476564 | A | G | I | V | MSH2 | ENST00000233146 | 0.78543 | 0.03802 | 0.70112 | 0.75918 | 0.89963 | 0.8805486149 | 0.91048 | 0.3652283963 | 0.15377 | 9.740515632 | 0 |
| 2 | 47476564 | A | T | I | F | MSH2 | ENST00000233146 | 0.87058 | 0.03013 | 0.807 | 0.87348 | 0.95389 | 0.9868297545 | 0.95509 | 0.9983959794 | 0.474781 | 9.807811757 | 0 |
| 2 | 47476565 | T | A | I | N | MSH2 | ENST00000233146 | 0.95015 | 0.02828 | 0.89534 | 0.935 | 0.94243 | 0.9943059802 | 0.90616 | 0.9981825352 | 0.415794 | 9.807811757 | 0 |
| 2 | 47476565 | T | C | I | T | MSH2 | ENST00000233146 | 0.93847 | 0.0334 | 0.9053 | 0.90386 | 0.95316 | 0.9811959003 | 0.93595 | 0.9984340072 | 0.467898 | 7.729990025 | 0 |
| 2 | 47476565 | T | G | I | S | MSH2 | ENST00000233146 | 0.95126 | 0.03118 | 0.8213 | 0.92662 | 0.9579 | 0.9931320271 | 0.93825 | 0.9919286966 | 0.467398 | 9.807811757 | 0 |
| 2 | 47476566 | C | G | I | M | MSH2 | ENST00000233146 | 0.53556 | 0.0368 | 0.76151 | 0.92154 | 0.93123 | 0.9635539245 | 0.88878 | 0.9722619653 | 0.312465 | 9.807811757 | 0 |
| 2 | 47476567 | C | A | L | I | MSH2 | ENST00000233146 | 0.57498 | 0.0357 | 0.8696 | 0.77897 | 0.82556 | 0.8543240897 | 0.78872 | 0.9521583319 | 0.223511 | 5.029272321 | 0 |
| 2 | 47476567 | C | G | L | V | MSH2 | ENST00000233146 | 0.71706 | 0.03653 | 0.82578 | 0.83239 | 0.84906 | 0.9373280359 | 0.93556 | 0.980014801 | 0.441127 | 5.161518171 | 0 |
| 2 | 47476567 | C | T | L | F | MSH2 | ENST00000233146 | 0.76697 | 0.03587 | 0.86301 | 0.89775 | 0.87733 | 0.9541449891 | 0.92526 | 0.9942570329 | 0.462098 | 9.813137104 | 0 |
| 2 | 47476568 | T | A | L | H | MSH2 | ENST00000233146 | 0.91556 | 0.0334 | 0.92683 | 0.91335 | 0.97735 | 0.9813990093 | 0.98344 | 0.9996520281 | 0.501522 | 9.813137104 | 0 |
| 2 | 47476568 | T | C | L | P | MSH2 | ENST00000233146 | 0.92768 | 0.03001 | 0.94662 | 0.91091 | 0.9781 | 0.9948570557 | 0.98713 | 0.9996380806 | 0.501512 | 9.813137104 | 1 |
| 2 | 47476568 | T | G | L | R | MSH2 | ENST00000233146 | 0.92563 | 0.03137 | 0.92823 | 0.91346 | 0.97735 | 0.9956600447 | 0.98359 | 0.9996001124 | 0.501239 | 9.813137104 | 1 |
| 2 | 47476570 | A | G | R | G | MSH2 | ENST00000233146 | 0.91725 | 0.02582 | 0.50982 | 0.93605 | 0.9528 | 0.9267666165 | 0.98331 | 0.9806565642 | 0.398359 | 7.635069276 | 0 |
| 2 | 47476570 | A | T | R | W | MSH2 | ENST00000233146 | 0.94478 | 0.0364 | 0.62573 | 0.92343 | 0.95462 | 0.9343278233 | 0.90982 | 0.9816248172 | 0.38984 | 8.903473365 | 0 |
| 2 | 47476571 | G | A | R | K | MSH2 | ENST00000233146 | 0.45119 | 0.00754 | 0.58446 | 0.7011 | 0.81588 | 0.8878817734 | 0.9168 | 0.3495832086 | 0.17697 | 2.878677289 | 0 |
| 2 | 47476571 | G | C | R | T | MSH2 | ENST00000233146 | 0.84233 | 0.02225 | 0.5719 | 0.92537 | 0.96008 | 0.9369463029 | 0.96236 | 0.9800936014 | 0.381878 | 4.565704242 | 0 |
| 2 | 47478272 | G | C | R | S | MSH2 | ENST00000233146 | 0.70133 | 0.01068 | 0.51911 | 0.86262 | 0.87781 | 0.9293857295 | 0.98031 | 0.9648150802 | 0.311089 | 4.691985256 | 0 |
| 2 | 47478272 | G | T | R | S | MSH2 | ENST00000233146 | 0.69477 | 0.01068 | 0.51911 | 0.86358 | 0.87781 | 0.9293857295 | 0.98021 | 0.9594750404 | 0.31109 | 4.691985256 | 0 |
| 2 | 47478273 | T | A | S | T | MSH2 | ENST00000233146 | 0.46047 | 0.00696 | 0.44211 | 0.65616 | 0.74936 | 0.7799652683 | 0.96278 | 0.7526881099 | 0.163522 | 1.777807525 | 0 |
| 2 | 47478273 | T | C | S | P | MSH2 | ENST00000233146 | 0.75694 | 0.00759 | 0.45206 | 0.83942 | 0.91986 | 0.961576156 | 0.97966 | 0.9645030499 | 0.366355 | 7.470746061 | 0 |
| 2 | 47478273 | T | G | S | A | MSH2 | ENST00000233146 | 0.56568 | 0.0071 | 0.41229 | 0.73808 | 0.75485 | 0.7208451157 | 0.94476 | 0.8170324564 | 0.121846 | 5.561223635 | 0 |
| 2 | 47478274 | C | A | S | Y | MSH2 | ENST00000233146 | 0.63 | 0.03175 | 0.5142 | 0.86074 | 0.86902 | 0.9112435484 | 0.93467 | 0.9907714128 | 0.285079 | 4.429770906 | 0 |
| 2 | 47478276 | G | A | A | T | MSH2 | ENST00000233146 | 0.90122 | 0.03735 | 0.70377 | 0.88001 | 0.98183 | 0.9484865537 | 0.92786 | 0.9920560718 | 0.426336 | 7.162919386 | 0 |
| 2 | 47478276 | G | C | A | P | MSH2 | ENST00000233146 | 0.90877 | 0.03322 | 0.76432 | 0.92229 | 0.98764 | 0.9854084735 | 0.94597 | 0.9983947873 | 0.482981 | 9.11305963 | 0 |
| 2 | 47478276 | G | T | A | S | MSH2 | ENST00000233146 | 0.86748 | 0.03452 | 0.6556 | 0.82731 | 0.93786 | 0.9101668738 | 0.88297 | 0.9795410038 | 0.28346 | 5.017631088 | 0 |
| 2 | 47478277 | C | A | A | E | MSH2 | ENST00000233146 | 0.67971 | 0.0347 | 0.76836 | 0.94277 | 0.99138 | 0.989546752 | 0.96771 | 0.9985681772 | 0.475996 | 9.11305963 | 1 |
| 2 | 47478277 | C | G | A | G | MSH2 | ENST00000233146 | 0.67565 | 0.02708 | 0.67872 | 0.90808 | 0.97587 | 0.9104443602 | 0.931 | 0.9848495464 | 0.317245 | 6.285996455 | 0 |
| 2 | 47478277 | C | T | A | V | MSH2 | ENST00000233146 | 0.69228 | 0.03713 | 0.71911 | 0.92015 | 0.96627 | 0.9554555647 | 0.91648 | 0.9898418784 | 0.301793 | 7.971585285 | 0 |
| 2 | 47478279 | A | C | T | P | MSH2 | ENST00000233146 | 0.83267 | 0.03212 | 0.52794 | 0.9638 | 0.94167 | 0.9531606631 | 0.95733 | 0.9925419092 | 0.330089 | 8.039535396 | 0 |
| 2 | 47478279 | A | G | T | A | MSH2 | ENST00000233146 | 0.78962 | 0.0279 | 0.53039 | 0.93171 | 0.93241 | 0.9228688078 | 0.95998 | 0.9889656305 | 0.254046 | 8.039535396 | 0 |
| 2 | 47478279 | A | T | T | S | MSH2 | ENST00000233146 | 0.45395 | 0.00923 | 0.53594 | 0.82121 | 0.82836 | 0.8183485979 | 0.91537 | 0.478859216 | 0.109735 | 1.675903779 | 0 |
| 2 | 47478280 | C | A | T | N | MSH2 | ENST00000233146 | 0.59089 | 0.03205 | 0.63105 | 0.8499 | 0.90274 | 0.9327581732 | 0.87903 | 0.9672780721 | 0.173811 | 6.273175797 | 0 |
| 2 | 47478280 | C | G | T | S | MSH2 | ENST00000233146 | 0.32908 | 0.00923 | 0.53594 | 0.49319 | 0.83615 | 0.8183485979 | 0.84272 | 0.7848694921 | 0.124096 | 1.675903779 | 0 |
| 2 | 47478280 | C | T | T | I | MSH2 | ENST00000233146 | 0.63351 | 0.03436 | 0.70593 | 0.9272 | 0.9328 | 0.9469202638 | 0.86418 | 0.9875705838 | 0.284482 | 8.039535396 | 0 |
| 2 | 47478282 | A | C | K | Q | MSH2 | ENST00000233146 | 0.42998 | 0.00569 | 0.41629 | 0.67331 | 0.81182 | 0.6951317438 | 0.95048 | 0.1773136705 | 0.1419 | 2.783339463 | 0 |
| 2 | 47478282 | A | G | K | E | MSH2 | ENST00000233146 | 0.36682 | 0.00517 | 0.4613 | 0.50642 | 0.68381 | 0.7626709147 | 0.88702 | 0.2604521215 | 0.0501851 | -0.4050458887 | 0 |
| 2 | 47478283 | A | C | K | T | MSH2 | ENST00000233146 | 0.54209 | 0.00687 | 0.49261 | 0.78036 | 0.86047 | 0.7911875184 | 0.87151 | 0.7230312228 | 0.179787 | 4.390996059 | 0 |
| 2 | 47478283 | A | G | K | R | MSH2 | ENST00000233146 | 0.50036 | 0.00608 | 0.39363 | 0.61998 | 0.73734 | 0.6402023786 | 0.94049 | 0.5740855932 | 0.14089 | 2.76792944 | 0 |
| 2 | 47478283 | A | T | K | I | MSH2 | ENST00000233146 | 0.76467 | 0.0099 | 0.4435 | 0.83532 | 0.90229 | 0.792122131 | 0.83574 | 0.9211308956 | 0.208474 | 7.183122528 | 0 |
| 2 | 47478284 | A | C | K | N | MSH2 | ENST00000233146 | 0.71466 | 0.00687 | 0.50877 | 0.76376 | 0.79577 | 0.7531275878 | 0.89562 | 0.6390925646 | 0.0741877 | 3.645455531 | 0 |
| 2 | 47478284 | A | T | K | N | MSH2 | ENST00000233146 | 0.70792 | 0.00687 | 0.50877 | 0.76245 | 0.79637 | 0.7531275878 | 0.89572 | 0.6953299642 | 0.0741919 | 3.645455531 | 0 |
| 2 | 47478285 | G | A | D | N | MSH2 | ENST00000233146 | 0.47658 | 0.00586 | 0.50366 | 0.39529 | 0.66161 | 0.5412497153 | 0.81629 | 0.3182888347 | 0.0304327 | -0.02679660045 | 0 |
| 2 | 47478285 | G | C | D | H | MSH2 | ENST00000233146 | 0.80574 | 0.0279 | 0.53849 | 0.79703 | 0.84213 | 0.7356616747 | 0.89983 | 0.7869810694 | 0.187453 | 2.702742753 | 0 |
| 2 | 47478285 | G | T | D | Y | MSH2 | ENST00000233146 | 0.92405 | 0.02943 | 0.5396 | 0.82954 | 0.85378 | 0.8104091421 | 0.9289 | 0.979175806 | 0.228373 | 6.996136661 | 0 |
| 2 | 47478286 | A | C | D | A | MSH2 | ENST00000233146 | 0.79049 | 0.00608 | 0.45792 | 0.76256 | 0.82387 | 0.7496162357 | 0.88518 | 0.7317880392 | 0.179984 | 5.170530859 | 0 |
| 2 | 47478286 | A | G | D | G | MSH2 | ENST00000233146 | 0.82662 | 0.01063 | 0.45067 | 0.7955 | 0.85222 | 0.7372087815 | 0.87105 | 0.9561433792 | 0.216396 | 6.700208451 | 0 |
| 2 | 47478286 | A | T | D | V | MSH2 | ENST00000233146 | 0.92548 | 0.01731 | 0.43429 | 0.8956 | 0.91945 | 0.7828197421 | 0.94304 | 0.9665460587 | 0.322889 | 6.996136661 | 0 |
| 2 | 47478287 | T | A | D | E | MSH2 | ENST00000233146 | 0.44722 | 0.00741 | 0.47849 | 0.65916 | 0.79516 | 0.5599760913 | 0.94898 | 0.160444364 | 0.118685 | 4.481902401 | 0 |
| 2 | 47478287 | T | G | D | E | MSH2 | ENST00000233146 | 0.45352 | 0.00741 | 0.47849 | 0.69776 | 0.79516 | 0.5599760913 | 0.94932 | 0.160444364 | 0.118682 | 4.481902401 | 0 |
| 2 | 47478288 | T | A | S | T | MSH2 | ENST00000233146 | 0.86425 | 0.03832 | 0.77429 | 0.91903 | 0.98621 | 0.9660952174 | 0.97445 | 0.9898256019 | 0.478584 | 9.75288704 | 0 |
| 2 | 47478288 | T | C | S | P | MSH2 | ENST00000233146 | 0.92712 | 0.03362 | 0.79811 | 0.94856 | 0.9917 | 0.9915004487 | 0.98222 | 0.9991795421 | 0.487583 | 9.75288704 | 1 |
| 2 | 47478288 | T | G | S | A | MSH2 | ENST00000233146 | 0.89752 | 0.03864 | 0.76782 | 0.9164 | 0.97475 | 0.9534639013 | 0.93946 | 0.9983170033 | 0.375323 | 8.549516046 | 0 |
| 2 | 47478289 | C | T | S | L | MSH2 | ENST00000233146 | 0.79856 | 0.03882 | 0.8051 | 0.93472 | 0.98071 | 0.9840430066 | 0.98111 | 0.9995410442 | 0.485065 | 9.75288704 | 1 |
| 2 | 47478291 | T | A | L | I | MSH2 | ENST00000233146 | 0.68923 | 0.03595 | 0.77346 | 0.93825 | 0.86503 | 0.8466442592 | 0.93288 | 0.989408493 | 0.28522 | 8.946191218 | 0 |
| 2 | 47478291 | T | G | L | V | MSH2 | ENST00000233146 | 0.68435 | 0.03697 | 0.71327 | 0.93406 | 0.88684 | 0.904995575 | 0.95771 | 0.9916346073 | 0.321872 | 9.605602935 | 0 |

| | | | | | | | | | | | | | | | | | |
|---|---|---|---|---|---|---|---|---|---|---|---|---|---|---|---|---|---|
| 2 | 47478292 | T | C | L | S | MSH2 | ENST00000233146 | 0.90359 | 0.03224 | 0.80203 | 0.94872 | 0.98034 | 0.9920347373 | 0.96532 | 0.9987400174 | 0.502531 | 9.605602935 | 1 |
| 2 | 47478293 | A | C | L | F | MSH2 | ENST00000233146 | 0.761 | 0.03595 | 0.76778 | 0.93061 | 0.91442 | 0.9444585123 | 0.96156 | 0.9708057566 | 0.387742 | 6.7422286 | 0 |
| 2 | 47478293 | A | T | L | F | MSH2 | ENST00000233146 | 0.75424 | 0.03595 | 0.76778 | 0.93266 | 0.91442 | 0.9444585123 | 0.96164 | 0.9721549629 | 0.387763 | 6.7422286 | 0 |
| 2 | 47478294 | A | C | I | L | MSH2 | ENST00000233146 | 0.81434 | 0.02083 | 0.8929 | 0.8641 | 0.92924 | 0.8477827106 | 0.85309 | 0.9566477537 | 0.202728 | 4.07446997 | 0 |
| 2 | 47478294 | A | G | I | V | MSH2 | ENST00000233146 | 0.41404 | 0.00971 | 0.81825 | 0.63174 | 0.74658 | 0.6457191376 | 0.84254 | 0.9175797105 | 0.101921 | 2.943525794 | 0 |
| 2 | 47478294 | A | T | I | L | MSH2 | ENST00000233146 | 0.80764 | 0.02083 | 0.8929 | 0.8643 | 0.92844 | 0.8477827106 | 0.85274 | 0.9566477537 | 0.202731 | 4.07446997 | 0 |
| 2 | 47478295 | T | A | I | K | MSH2 | ENST00000233146 | 0.91083 | 0.02628 | 0.93914 | 0.96303 | 0.98764 | 0.982150233 | 0.96954 | 0.9993788004 | 0.472192 | 8.898529793 | 1 |
| 2 | 47478295 | T | C | I | T | MSH2 | ENST00000233146 | 0.87931 | 0.01906 | 0.95719 | 0.9539 | 0.98513 | 0.9388320945 | 0.97627 | 0.9986573458 | 0.462678 | 8.898529793 | 0 |
| 2 | 47478295 | T | G | I | R | MSH2 | ENST00000233146 | 0.92015 | 0.02775 | 0.92661 | 0.9629 | 0.98549 | 0.9840035801 | 0.95983 | 0.9993570447 | 0.472636 | 8.898529793 | 1 |
| 2 | 47478296 | A | G | I | M | MSH2 | ENST00000233146 | 0.81716 | 0.02421 | 0.8633 | 0.90745 | 0.92844 | 0.8799533619 | 0.93342 | 0.9568358055 | 0.30395 | 6.720719509 | 0 |
| 2 | 47478297 | A | C | I | L | MSH2 | ENST00000233146 | 0.65704 | 0.02775 | 0.89618 | 0.75986 | 0.88496 | 0.9071083133 | 0.84871 | 0.9254059792 | 0.196677 | 4.75834926 | 0 |
| 2 | 47478297 | A | G | I | V | MSH2 | ENST00000233146 | 0.75949 | 0.0262 | 0.82378 | 0.77106 | 0.90624 | 0.8068759216 | 0.85799 | 0.8882484514 | 0.214186 | 5.568697558 | 0 |
| 2 | 47478297 | A | T | I | F | MSH2 | ENST00000233146 | 0.85441 | 0.0296 | 0.9028 | 0.90725 | 0.99296 | 0.9784918062 | 0.94872 | 0.9986224174 | 0.457356 | 9.958149124 | 0 |
| 2 | 47478298 | T | A | I | N | MSH2 | ENST00000233146 | 0.9077 | 0.0276 | 0.95146 | 0.95844 | 0.98294 | 0.9920718502 | 0.92963 | 0.99880898 | 0.418671 | 9.958149124 | 1 |
| 2 | 47478298 | T | C | I | T | MSH2 | ENST00000233146 | 0.82436 | 0.01475 | 0.95898 | 0.9434 | 0.96189 | 0.9804628544 | 0.90347 | 0.9974076152 | 0.313426 | 9.73914642 | 0 |
| 2 | 47478298 | T | G | I | S | MSH2 | ENST00000233146 | 0.91162 | 0.02869 | 0.90989 | 0.96078 | 0.98657 | 0.9941846849 | 0.96684 | 0.9991216063 | 0.464951 | 9.958149124 | 0 |
| 2 | 47478299 | C | G | I | M | MSH2 | ENST00000233146 | 0.58881 | 0.03626 | 0.86933 | 0.89084 | 0.9517 | 0.9306694378 | 0.89088 | 0.9970862269 | 0.265216 | 9.958149124 | 0 |
| 2 | 47478300 | A | C | I | L | MSH2 | ENST00000233146 | 0.80558 | 0.03334 | 0.87506 | 0.7847 | 0.84267 | 0.8778061271 | 0.81159 | 0.9253824949 | 0.191787 | 3.18196126 | 0 |
| 2 | 47478300 | A | G | I | V | MSH2 | ENST00000233146 | 0.59161 | 0.02225 | 0.79054 | 0.59981 | 0.84641 | 0.7496712234 | 0.79652 | 0.7915478945 | 0.168663 | 6.783137612 | 0 |
| 2 | 47478300 | A | T | I | L | MSH2 | ENST00000233146 | 0.79895 | 0.03334 | 0.87506 | 0.79484 | 0.84267 | 0.8778061271 | 0.81197 | 0.9253824949 | 0.191787 | 3.18196126 | 0 |
| 2 | 47478301 | T | C | I | T | MSH2 | ENST00000233146 | 0.74906 | 0.03334 | 0.94726 | 0.9366 | 0.97438 | 0.9715505163 | 0.91902 | 0.9954845905 | 0.36084 | 8.644138424 | 0 |
| 2 | 47478301 | T | G | I | R | MSH2 | ENST00000233146 | 0.77505 | 0.02851 | 0.91063 | 0.94352 | 0.97735 | 0.9920731585 | 0.97068 | 0.9991480112 | 0.464209 | 8.644138424 | 1 |
| 2 | 47478302 | A | G | I | M | MSH2 | ENST00000233146 | 0.65539 | 0.03667 | 0.83933 | 0.91371 | 0.87198 | 0.9047918463 | 0.92631 | 0.9764134288 | 0.229741 | 5.7331481 | 0 |
| 2 | 47478303 | G | A | D | N | MSH2 | ENST00000233146 | 0.92015 | 0.03741 | 0.94433 | 0.92415 | 0.99072 | 0.9847552445 | 0.98115 | 0.9993304014 | 0.284504 | 9.986798665 | 0 |
| 2 | 47478303 | G | C | D | H | MSH2 | ENST00000233146 | 0.91746 | 0.03458 | 0.95625 | 0.93296 | 0.99953 | 0.9864858642 | 0.99085 | 0.9986652136 | 0.583664 | 9.986798665 | 1 |
| 2 | 47478303 | G | T | D | Y | MSH2 | ENST00000233146 | 0.92166 | 0.03398 | 0.94762 | 0.93458 | 0.99843 | 0.9933114223 | 0.9949 | 0.9996054769 | 0.583761 | 9.986798665 | 1 |
| 2 | 47478304 | A | C | D | A | MSH2 | ENST00000233146 | 0.89577 | 0.02802 | 0.90299 | 0.97192 | 0.99234 | 0.990796198 | 0.99145 | 0.9996803999 | 0.579972 | 9.986798665 | 1 |
| 2 | 47478304 | A | G | D | G | MSH2 | ENST00000233146 | 0.90391 | 0.0325 | 0.91268 | 0.9686 | 0.99652 | 0.9915385944 | 0.99335 | 0.9991549253 | 0.580325 | 9.986798665 | 1 |
| 2 | 47478304 | A | T | D | V | MSH2 | ENST00000233146 | 0.89371 | 0.03166 | 0.90705 | 0.97751 | 0.99471 | 0.9909667247 | 0.99548 | 0.9996988773 | 0.583232 | 9.986798665 | 1 |
| 2 | 47478305 | T | A | D | E | MSH2 | ENST00000233146 | 0.59545 | 0.03783 | 0.92791 | 0.96889 | 0.97885 | 0.9823622794 | 0.99096 | 0.9985194802 | 0.531853 | 9.986798665 | 0 |
| 2 | 47478305 | T | G | D | E | MSH2 | ENST00000233146 | 0.60295 | 0.03783 | 0.92791 | 0.96849 | 0.97885 | 0.9823622794 | 0.9909 | 0.9985194802 | 0.531855 | 9.986798665 | 0 |
| 2 | 47478306 | G | A | E | K | MSH2 | ENST00000233146 | 0.92979 | 0.03175 | 0.96973 | 0.91436 | 0.99786 | 0.9952069226 | 0.99415 | 0.9997194409 | 0.571454 | 9.98679786 | 1 |
| 2 | 47478306 | G | C | E | Q | MSH2 | ENST00000233146 | 0.89628 | 0.03713 | 0.91807 | 0.93195 | 0.99234 | 0.9801199832 | 0.98863 | 0.9984708428 | 0.572213 | 9.98679786 | 1 |
| 2 | 47478307 | A | C | E | A | MSH2 | ENST00000233146 | 0.88946 | 0.03552 | 0.92825 | 0.95876 | 0.99765 | 0.9875153592 | 0.98774 | 0.9991713762 | 0.572334 | 9.98679786 | 1 |
| 2 | 47478307 | A | G | E | G | MSH2 | ENST00000233146 | 0.91975 | 0.03452 | 0.8923 | 0.9609 | 0.99552 | 0.9890592574 | 0.97951 | 0.999319911 | 0.574453 | 9.98679786 | 1 |
| 2 | 47478307 | A | T | E | V | MSH2 | ENST00000233146 | 0.90978 | 0.0347 | 0.91478 | 0.96437 | 0.99786 | 0.9894622943 | 0.9924 | 0.9988077879 | 0.575489 | 9.98679786 | 1 |
| 2 | 47478308 | A | C | E | D | MSH2 | ENST00000233146 | 0.58817 | 0.03832 | 0.92863 | 0.98324 | 0.92642 | 0.9753319184 | 0.97769 | 0.9984513521 | 0.371617 | 9.98679786 | 1 |
| 2 | 47478308 | A | T | E | D | MSH2 | ENST00000233146 | 0.58211 | 0.03832 | 0.92863 | 0.98325 | 0.92642 | 0.9753319184 | 0.97766 | 0.9984513521 | 0.371614 | 9.98679786 | 1 |
| 2 | 47478309 | T | A | L | M | MSH2 | ENST00000233146 | 0.61492 | 0.03761 | 0.91235 | 0.93941 | 0.87442 | 0.881498472 | 0.93408 | 0.9329544469 | 0.284587 | 8.912014119 | 0 |
| 2 | 47478309 | T | G | L | V | MSH2 | ENST00000233146 | 0.58988 | 0.03686 | 0.88993 | 0.86685 | 0.87148 | 0.9108662496 | 0.91149 | 0.9645665884 | 0.196883 | 3.971225911 | 0 |
| 2 | 47478310 | T | C | L | S | MSH2 | ENST00000233146 | 0.93451 | 0.032 | 0.9339 | 0.94315 | 0.97698 | 0.9806729177 | 0.97233 | 0.9987400174 | 0.476928 | 8.912014119 | 0 |
| 2 | 47478310 | T | G | L | W | MSH2 | ENST00000233146 | 0.9351 | 0.02984 | 0.96075 | 0.94652 | 0.97438 | 0.9778529955 | 0.95629 | 0.9994977713 | 0.476945 | 8.912014119 | 1 |
| 2 | 47478311 | G | C | L | F | MSH2 | ENST00000233146 | 0.88164 | 0.03587 | 0.91603 | 0.88813 | 0.93162 | 0.9480976459 | 0.91759 | 0.9758835435 | 0.378362 | 8.858396404 | 0 |
| 2 | 47478311 | G | T | L | F | MSH2 | ENST00000233146 | 0.8763 | 0.03587 | 0.91603 | 0.89093 | 0.93241 | 0.9480976459 | 0.91706 | 0.9758835435 | 0.378367 | 8.858396404 | 0 |
| 2 | 47478312 | G | A | G | R | MSH2 | ENST00000233146 | 0.92802 | 0.03452 | 0.9757 | 0.9354 | 0.99722 | 0.9818235703 | 0.98937 | 0.9996510744 | 0.557401 | 9.977961334 | 1 |
| 2 | 47478312 | G | C | G | R | MSH2 | ENST00000233146 | 0.9266 | 0.03452 | 0.9757 | 0.93578 | 0.997 | 0.9818235703 | 0.98921 | 0.9996510744 | 0.557401 | 9.977961334 | 1 |
| 2 | 47478313 | G | A | G | E | MSH2 | ENST00000233146 | 0.91125 | 0.03334 | 0.97216 | 0.93529 | 0.99722 | 0.9811158493 | 0.98149 | 0.9996510744 | 0.55693 | 9.977961334 | 1 |
| 2 | 47478313 | G | C | G | A | MSH2 | ENST00000233146 | 0.88653 | 0.03608 | 0.94185 | 0.94228 | 0.99603 | 0.9684556002 | 0.98407 | 0.9991809726 | 0.555132 | 9.977961334 | 0 |
| 2 | 47478313 | G | T | G | V | MSH2 | ENST00000233146 | 0.90833 | 0.03379 | 0.94475 | 0.93961 | 0.99498 | 0.9824712634 | 0.97978 | 0.9996510744 | 0.557401 | 9.922045703 | 0 |
| 2 | 47478315 | A | G | R | G | MSH2 | ENST00000233146 | 0.91148 | 0.03313 | 0.91075 | 0.95607 | 0.98146 | 0.9749682254 | 0.97402 | 0.998287499 | 0.489454 | 9.986810905 | 1 |
| 2 | 47478316 | G | A | R | K | MSH2 | ENST00000233146 | 0.85375 | 0.03509 | 0.9492 | 0.88523 | 0.97773 | 0.9543573247 | 0.97034 | 0.9816122651 | 0.428279 | 9.986810905 | 0 |
| 2 | 47478316 | G | C | R | T | MSH2 | ENST00000233146 | 0.85771 | 0.03112 | 0.93927 | 0.91071 | 0.99105 | 0.9737335007 | 0.97974 | 0.9994462132 | 0.524326 | 9.986810905 | 1 |
| 2 | 47478316 | G | T | R | I | MSH2 | ENST00000233146 | 0.89411 | 0.02824 | 0.93327 | 0.92625 | 0.98869 | 0.9599350068 | 0.97316 | 0.9998717308 | 0.525545 | 9.986810905 | 0 |
| 2 | 47478317 | A | C | R | S | MSH2 | ENST00000233146 | 0.82615 | 0.03503 | 0.91658 | 0.96802 | 0.97997 | 0.9751151486 | 0.99115 | 0.9982513785 | 0.481746 | 9.986810905 | 1 |
| 2 | 47478317 | A | T | R | S | MSH2 | ENST00000233146 | 0.81959 | 0.03503 | 0.91658 | 0.96895 | 0.97997 | 0.9751151486 | 0.99113 | 0.9982513785 | 0.481747 | 9.986810905 | 1 |
| 2 | 47478318 | G | A | G | R | MSH2 | ENST00000233146 | 0.92733 | 0.03452 | 0.99305 | 0.92264 | 0.99676 | 0.9712333831 | 0.9905 | 0.9996510744 | 0.557401 | 9.986795116 | 1 |
| 2 | 47478318 | G | C | G | R | MSH2 | ENST00000233146 | 0.92591 | 0.03452 | 0.99305 | 0.92264 | 0.99676 | 0.9712333831 | 0.99036 | 0.9996510744 | 0.557401 | 9.986795116 | 1 |
| 2 | 47478319 | G | A | G | E | MSH2 | ENST00000233146 | 0.80408 | 0.03334 | 0.99093 | 0.92256 | 0.997 | 0.9706870449 | 0.985 | 0.9996510744 | 0.55693 | 9.986795116 | 1 |
| 2 | 47478319 | G | C | G | A | MSH2 | ENST00000233146 | 0.76808 | 0.03608 | 0.97891 | 0.9088 | 0.99722 | 0.9571231681 | 0.98982 | 0.9989991784 | 0.555132 | 9.986795116 | 0 |
| 2 | 47478319 | G | T | G | V | MSH2 | ENST00000233146 | 0.79934 | 0.03379 | 0.97905 | 0.94909 | 0.99932 | 0.9742218629 | 0.9839 | 0.9996510744 | 0.557401 | 9.986795116 | 0 |
| 2 | 47478321 | A | C | T | P | MSH2 | ENST00000233146 | 0.85077 | 0.02442 | 0.72 | 0.94793 | 0.98621 | 0.980094369 | 0.97993 | 0.9988908172 | 0.488843 | 9.986801234 | 1 |
| 2 | 47478321 | A | G | T | A | MSH2 | ENST00000233146 | 0.82514 | 0.03633 | 0.72722 | 0.91938 | 0.98367 | 0.9702972833 | 0.98232 | 0.9711617827 | 0.301604 | 9.986801234 | 0 |
| 2 | 47478321 | A | T | T | S | MSH2 | ENST00000233146 | 0.78431 | 0.03475 | 0.74056 | 0.90678 | 0.97997 | 0.9264861418 | 0.97109 | 0.9956958294 | 0.433765 | 9.986801234 | 0 |
| 2 | 47478322 | C | A | T | N | MSH2 | ENST00000233146 | 0.59039 | 0.03497 | 0.81958 | 0.9093 | 0.96627 | 0.9756924854 | 0.92112 | 0.9988580942 | 0.29661 | 9.986801234 | 1 |
| 2 | 47478322 | C | G | T | S | MSH2 | ENST00000233146 | 0.58304 | 0.03475 | 0.74056 | 0.88871 | 0.97327 | 0.9264861418 | 0.9561 | 0.9939288488 | 0.433054 | 9.986801234 | 0 |
| 2 | 47478322 | C | T | T | I | MSH2 | ENST00000233146 | 0.63114 | 0.03595 | 0.86858 | 0.91443 | 0.98728 | 0.9800703661 | 0.97202 | 0.9987882972 | 0.481116 | 9.986801234 | 1 |
| 2 | 47478324 | T | A | S | T | MSH2 | ENST00000233146 | 0.79144 | 0.03832 | 0.88229 | 0.88964 | 0.97698 | 0.9483106149 | 0.94187 | 0.9895037349 | 0.389339 | 10.01367662 | 0 |
| 2 | 47478324 | T | C | S | P | MSH2 | ENST00000233146 | 0.88002 | 0.03362 | 0.8965 | 0.91878 | 0.99603 | 0.9810301521 | 0.97242 | 0.9991795421 | 0.463082 | 10.01367662 | 0 |
| 2 | 47478324 | T | G | S | A | MSH2 | ENST00000233146 | 0.83283 | 0.03901 | 0.87763 | 0.8817 | 0.95899 | 0.9259571826 | 0.91932 | 0.9847015627 | 0.296359 | 5.351612597 | 0 |
| 2 | 47478325 | C | A | S | Y | MSH2 | ENST00000233146 | 0.65366 | 0.03334 | 0.9244 | 0.89621 | 0.98477 | 0.9759344425 | 0.97677 | 0.9987882972 | 0.461565 | 10.01367662 | 1 |
| 2 | 47478325 | C | G | S | C | MSH2 | ENST00000233146 | 0.68571 | 0.03753 | 0.88899 | 0.90221 | 0.97624 | 0.9628424302 | 0.93462 | 0.9985787868 | 0.457192 | 10.01367662 | 0 |
| 2 | 47478325 | C | T | S | F | MSH2 | ENST00000233146 | 0.70342 | 0.03563 | 0.91143 | 0.89302 | 0.98938 | 0.9795515837 | 0.97728 | 0.9991251826 | 0.46128 | 10.01367662 | 0 |
| 2 | 47478327 | A | C | T | P | MSH2 | ENST00000233146 | 0.83598 | 0.03322 | 0.74524 | 0.93439 | 0.98367 | 0.9770136922 | 0.93925 | 0.9959634542 | 0.462713 | 9.736790706 | 1 |
| 2 | 47478327 | A | G | T | A | MSH2 | ENST00000233146 | 0.79919 | 0.03713 | 0.74668 | 0.88795 | 0.98034 | 0.9416044654 | 0.94833 | 0.9944168329 | 0.31717 | 9.736790706 | 0 |
| 2 | 47478327 | A | T | T | S | MSH2 | ENST00000233146 | 0.76243 | 0.03852 | 0.7594 | 0.89685 | 0.95645 | 0.8796351822 | 0.92698 | 0.9920824766 | 0.251817 | 9.736790706 | 0 |
| 2 | 47478328 | C | A | T | N | MSH2 | ENST00000233146 | 0.5906 | 0.03503 | 0.84331 | 0.88084 | 0.95426 | 0.9679405911 | 0.90336 | 0.9934753776 | 0.20449 | 7.101232527 | 0 |
| 2 | 47478328 | C | G | T | S | MSH2 | ENST00000233146 | 0.58745 | 0.03852 | 0.7594 | 0.88317 | 0.94802 | 0.8796351822 | 0.91004 | 0.9290035963 | 0.0461835 | 9.736790706 | 0 |
| 2 | 47478328 | C | T | T | I | MSH2 | ENST00000233146 | 0.62817 | 0.03353 | 0.88682 | 0.9023 | 0.97885 | 0.9640717291 | 0.97168 | 0.9930784106 | 0.461937 | 9.736790706 | 0 |
| 2 | 47478330 | T | A | Y | N | MSH2 | ENST00000233146 | 0.84514 | 0.02979 | 0.97047 | 0.93106 | 0.97438 | 0.9245254495 | 0.93535 | 0.9985377789 | 0.424104 | 9.821126326 | 0 |
| 2 | 47478330 | T | C | Y | H | MSH2 | ENST00000233146 | 0.83928 | 0.03327 | 0.97314 | 0.88265 | 0.9659 | 0.866931513 | 0.8836 | 0.9953092933 | 0.305312 | 8.292691271 | 0 |
| 2 | 47478330 | T | G | Y | D | MSH2 | ENST00000233146 | 0.8516 | 0.02857 | 0.96936 | 0.94975 | 0.98477 | 0.9440446377 | 0.97691 | 0.9975960851 | 0.444255 | 9.821126326 | 0 |
| 2 | 47478331 | A | C | Y | S | MSH2 | ENST00000233146 | 0.84111 | 0.03189 | 0.9401 | 0.95226 | 0.96883 | 0.9171038396 | 0.96141 | 0.9982675314 | 0.381118 | 7.885483806 | 0 |
| 2 | 47478331 | A | G | Y | C | MSH2 | ENST00000233146 | 0.87092 | 0.03379 | 0.97171 | 0.94886 | 0.95462 | 0.9153550697 | 0.90015 | 0.9995137453 | 0.34019 | 9.821126326 | 0 |
| 2 | 47478331 | A | T | Y | F | MSH2 | ENST00000233146 | 0.62772 | 0.02809 | 0.93144 | 0.77091 | 0.91569 | 0.7478526895 | 0.9047 | 0.9623948336 | 0.208239 | 4.57130708 | 0 |
| 2 | 47478333 | G | A | D | N | MSH2 | ENST00000233146 | 0.90735 | 0.03741 | 0.92006 | 0.9122 | 0.95753 | 0.95534205 | 0.96021 | 0.9992992878 | 0.191027 | 9.778977546 | 0 |
| 2 | 47478333 | G | C | D | H | MSH2 | ENST00000233146 | 0.90427 | 0.03458 | 0.93506 | 0.91801 | 0.98972 | 0.9659195265 | 0.98418 | 0.9986652136 | 0.497376 | 9.778977546 | 1 |
| 2 | 47478333 | G | T | D | Y | MSH2 | ENST00000233146 | 0.90944 | 0.03398 | 0.92689 | 0.91233 | 0.98728 | 0.9788678794 | 0.98867 | 0.9997151494 | 0.498299 | 9.778977546 | 1 |
| 2 | 47478334 | A | C | D | A | MSH2 | ENST00000233146 | 0.88733 | 0.03362 | 0.87171 | 0.94945 | 0.98657 | 0.9733053041 | 0.9655 | 0.9983383417 | 0.489403 | 9.778977546 | 0 |
| 2 | 47478334 | A | G | D | G | MSH2 | ENST00000233146 | 0.86893 | 0.02432 | 0.88113 | 0.94581 | 0.99202 | 0.9692991174 | 0.98531 | 0.9980017543 | 0.485965 | 9.778977546 | 1 |
| 2 | 47478334 | A | T | D | V | MSH2 | ENST00000233146 | 0.88132 | 0.0317 | 0.87353 | 0.95633 | 0.99386 | 0.9750272205 | 0.98607 | 0.9997014403 | 0.49672 | 9.778977546 | 0 |
| 2 | 47478335 | T | A | D | E | MSH2 | ENST00000233146 | 0.47451 | 0.03767 | 0.90066 | 0.87611 | 0.92275 | 0.9327956029 | 0.98723 | 0.9645124674 | 0.382523 | 6.792474639 | 0 |
| 2 | 47478335 | T | G | D | E | MSH2 | ENST00000233146 | 0.48101 | 0.03767 | 0.90066 | 0.87393 | 0.92275 | 0.9327956029 | 0.98718 | 0.9645124674 | 0.382525 | 6.792474639 | 0 |
| 2 | 47478336 | G | A | G | R | MSH2 | ENST00000233146 | 0.91926 | 0.03313 | 0.94046 | 0.94326 | 0.99296 | 0.9855395738 | 0.98841 | 0.9996652603 | 0.554679 | 9.986800713 | 1 |
| 2 | 47478336 | G | C | G | R | MSH2 | ENST00000233146 | 0.91754 | 0.03313 | 0.94046 | 0.94294 | 0.99296 | 0.9855395738 | 0.98797 | 0.9996652603 | 0.554684 | 9.986800713 | 1 |
| 2 | 47478337 | G | A | G | E | MSH2 | ENST00000233146 | 0.78946 | 0.03322 | 0.93431 | 0.92268 | 0.99765 | 0.9858047213 | 0.98439 | 0.9996054769 | 0.556882 | 9.986800713 | 1 |
| 2 | 47478337 | G | C | G | A | MSH2 | ENST00000233146 | 0.75328 | 0.03595 | 0.87958 | 0.93985 | 0.99806 | 0.9610129483 | 0.98497 | 0.9985890985 | 0.554801 | 9.986800713 | 0 |
| 2 | 47478337 | G | T | G | V | MSH2 | ENST00000233146 | 0.78471 | 0.0337 | 0.8866 | 0.9278 | 0.99824 | 0.9751125734 | 0.98465 | 0.9996054769 | 0.557352 | 9.986800713 | 1 |
| 2 | 47478339 | T | A | F | I | MSH2 | ENST00000233146 | 0.87689 | 0.0318 | 0.85339 | 0.87955 | 0.97512 | 0.920733843 | 0.9515 | 0.9912291765 | 0.377619 | 4.265174647 | 0 |
| 2 | 47478339 | T | C | F | L | MSH2 | ENST00000233146 | 0.86996 | 0.02875 | 0.89563 | 0.79937 | 0.97401 | 0.8915200944 | 0.96048 | 0.9872276783 | 0.364479 | 3.908293783 | 0 |
| 2 | 47478339 | T | G | F | V | MSH2 | ENST00000233146 | 0.88093 | 0.03274 | 0.84973 | 0.91346 | 0.97997 | 0.8863413454 | 0.95843 | 0.9912291765 | 0.404683 | 4.916140665 | 0 |
| 2 | 47478340 | T | A | F | Y | MSH2 | ENST00000233146 | 0.8658 | 0.03509 | 0.87796 | 0.87922 | 0.90798 | 0.8285670085 | 0.90254 | 0.9730266929 | 0.251476 | 6.124807339 | 0 |

| | | | | | | | | | | | | | | | | |
|---|---|---|---|---|---|---|---|---|---|---|---|---|---|---|---|---|
| 2 | 47478340 | T | C | F | S | MSH2 | ENST00000233146 | 0.92438 | 0.02857 | 0.86324 | 0.96598 | 0.96117 | 0.9340524301 | 0.95824 | 0.9952347875 | 0.422248 | 8.602315115 | 0 |
| 2 | 47478340 | T | G | F | C | MSH2 | ENST00000233146 | 0.60786 | 0.03175 | 0.8951 | 0.92865 | 0.90798 | 0.9109215265 | 0.86274 | 0.9827521737 | 0.254945 | 8.602315115 | 0 |
| 2 | 47478341 | T | A | F | L | MSH2 | ENST00000233146 | 0.6666 | 0.02875 | 0.89563 | 0.84206 | 0.92192 | 0.8915200944 | 0.97299 | 0.9794576168 | 0.334098 | 3.908293783 | 0 |
| 2 | 47478341 | T | G | F | L | MSH2 | ENST00000233146 | 0.67477 | 0.02875 | 0.89563 | 0.84156 | 0.92192 | 0.8915200944 | 0.97274 | 0.9794576168 | 0.3341 | 3.908293783 | 0 |
| 2 | 47478342 | G | A | G | R | MSH2 | ENST00000233146 | 0.92264 | 0.03452 | 0.9517 | 0.92823 | 0.99552 | 0.9796637213 | 0.9789 | 0.9968089461 | 0.468104 | 8.675753292 | 1 |
| 2 | 47478342 | G | C | G | R | MSH2 | ENST00000233146 | 0.92108 | 0.03452 | 0.9517 | 0.92616 | 0.99552 | 0.9796637213 | 0.97872 | 0.9968089461 | 0.468104 | 8.675753292 | 1 |
| 2 | 47478343 | G | A | G | E | MSH2 | ENST00000233146 | 0.7984 | 0.03334 | 0.94631 | 0.90683 | 0.99006 | 0.9782832714 | 0.9608 | 0.9996516705 | 0.47243 | 8.675753292 | 1 |
| 2 | 47478343 | G | C | G | A | MSH2 | ENST00000233146 | 0.76227 | 0.03608 | 0.89278 | 0.84499 | 0.97215 | 0.9254766561 | 0.9732 | 0.9954782128 | 0.416644 | 3.554036895 | 1 |
| 2 | 47478343 | G | T | G | V | MSH2 | ENST00000233146 | 0.79358 | 0.03379 | 0.90273 | 0.90177 | 0.99296 | 0.9584814328 | 0.97103 | 0.9968089461 | 0.467265 | 8.675753292 | 1 |
| 2 | 47478345 | T | A | L | I | MSH2 | ENST00000233146 | 0.38409 | 0.03602 | 0.87345 | 0.90916 | 0.95097 | 0.680201684 | 0.89052 | 0.7966502309 | 0.138556 | 3.689394867 | 0 |
| 2 | 47478345 | T | G | L | V | MSH2 | ENST00000233146 | 0.51698 | 0.03706 | 0.82738 | 0.92785 | 0.89239 | 0.8238538306 | 0.96339 | 0.9531087875 | 0.326473 | 9.172460628 | 0 |
| 2 | 47478346 | T | C | L | S | MSH2 | ENST00000233146 | 0.94368 | 0.03231 | 0.88596 | 0.94917 | 0.98799 | 0.9565399373 | 0.98503 | 0.9996407032 | 0.473047 | 9.483022908 | 0 |
| 2 | 47478347 | A | C | L | F | MSH2 | ENST00000233146 | 0.76586 | 0.03595 | 0.86873 | 0.95185 | 0.9458 | 0.8994987839 | 0.97005 | 0.9984782338 | 0.409076 | 9.483022908 | 0 |
| 2 | 47478347 | A | T | L | F | MSH2 | ENST00000233146 | 0.75917 | 0.03595 | 0.86873 | 0.95292 | 0.9458 | 0.8994987839 | 0.97013 | 0.9984782338 | 0.409071 | 9.483022908 | 0 |
| 2 | 47478348 | G | A | A | T | MSH2 | ENST00000233146 | 0.89949 | 0.03758 | 0.8093 | 0.9143 | 0.9648 | 0.9233541128 | 0.9462 | 0.998306632 | 0.40501 | 9.986788358 | 0 |
| 2 | 47478348 | G | C | A | P | MSH2 | ENST00000233146 | 0.90725 | 0.03334 | 0.86032 | 0.9158 | 0.9822 | 0.9853564741 | 0.95678 | 0.9990723133 | 0.495123 | 9.986788358 | 1 |
| 2 | 47478348 | G | T | A | S | MSH2 | ENST00000233146 | 0.86969 | 0.03848 | 0.76256 | 0.88002 | 0.93241 | 0.864664126 | 0.93999 | 0.986297909 | 0.318426 | 9.986788358 | 0 |
| 2 | 47478349 | C | A | A | E | MSH2 | ENST00000233146 | 0.66502 | 0.03484 | 0.86326 | 0.93949 | 0.9833 | 0.9864176287 | 0.97308 | 0.9987955093 | 0.485309 | 9.986788358 | 1 |
| 2 | 47478349 | C | G | A | G | MSH2 | ENST00000233146 | 0.67668 | 0.03706 | 0.7815 | 0.93099 | 0.96117 | 0.870129161 | 0.94888 | 0.9981606603 | 0.380175 | 9.986788358 | 0 |
| 2 | 47478349 | C | T | A | V | MSH2 | ENST00000233146 | 0.67931 | 0.03718 | 0.81962 | 0.94559 | 0.9608 | 0.9434311656 | 0.9378 | 0.9977529645 | 0.321964 | 9.986788358 | 0 |
| 2 | 47478351 | T | A | W | R | MSH2 | ENST00000233146 | 0.91805 | 0.02857 | 0.94539 | 0.91729 | 0.97773 | 0.9768116298 | 0.9605 | 0.9990175962 | 0.433281 | 5.137838512 | 0 |
| 2 | 47478351 | T | C | W | R | MSH2 | ENST00000233146 | 0.92555 | 0.02857 | 0.94539 | 0.91597 | 0.9781 | 0.9768116298 | 0.96107 | 0.9990175962 | 0.433277 | 5.137838512 | 0 |
| 2 | 47478351 | T | G | W | G | MSH2 | ENST00000233146 | 0.93884 | 0.02857 | 0.86982 | 0.95429 | 0.97698 | 0.947489999 | 0.97899 | 0.9988284707 | 0.445245 | 7.738821174 | 0 |
| 2 | 47478352 | G | C | W | S | MSH2 | ENST00000233146 | 0.94735 | 0.02994 | 0.89773 | 0.90817 | 0.96226 | 0.9609638821 | 0.96375 | 0.9995724559 | 0.457041 | 9.33208677 | 0 |
| 2 | 47478352 | G | T | W | L | MSH2 | ENST00000233146 | 0.93595 | 0.02381 | 0.90843 | 0.90627 | 0.96773 | 0.926569608 | 0.97538 | 0.9996689558 | 0.454473 | 9.33208677 | 0 |
| 2 | 47478353 | G | C | W | C | MSH2 | ENST00000233146 | 0.94948 | 0.03029 | 0.90093 | 0.94467 | 0.95389 | 0.974765984 | 0.97176 | 0.9995523095 | 0.463639 | 8.375703319 | 0 |
| 2 | 47478353 | G | T | W | C | MSH2 | ENST00000233146 | 0.94851 | 0.03029 | 0.90093 | 0.94485 | 0.95389 | 0.974765984 | 0.972 | 0.9995523095 | 0.463639 | 8.375703319 | 0 |
| 2 | 47478354 | G | A | A | T | MSH2 | ENST00000233146 | 0.8984 | 0.03718 | 0.87488 | 0.93544 | 0.98071 | 0.9096111299 | 0.9581 | 0.9899625182 | 0.472921 | 10.04439208 | 0 |
| 2 | 47478354 | G | C | A | P | MSH2 | ENST00000233146 | 0.90705 | 0.03322 | 0.91919 | 0.95046 | 0.98693 | 0.9819760432 | 0.97393 | 0.9987015724 | 0.482954 | 10.04439208 | 1 |
| 2 | 47478354 | G | T | A | S | MSH2 | ENST00000233146 | 0.70109 | 0.03452 | 0.83799 | 0.85466 | 0.92192 | 0.8371063581 | 0.92551 | 0.9648966789 | 0.275874 | 4.752185281 | 0 |
| 2 | 47478355 | C | A | A | D | MSH2 | ENST00000233146 | 0.68555 | 0.03099 | 0.93601 | 0.94864 | 0.99386 | 0.983282336 | 0.96034 | 0.9988458157 | 0.482887 | 10.04439208 | 1 |
| 2 | 47478355 | C | G | A | G | MSH2 | ENST00000233146 | 0.63275 | 0.02708 | 0.85913 | 0.88986 | 0.96737 | 0.8506521397 | 0.95414 | 0.9696423662 | 0.394549 | 7.846424187 | 0 |
| 2 | 47478355 | C | T | A | V | MSH2 | ENST00000233146 | 0.67987 | 0.03713 | 0.88284 | 0.93839 | 0.98621 | 0.9387228627 | 0.963 | 0.9981383085 | 0.418367 | 10.04439208 | 0 |
| 2 | 47478357 | A | C | I | L | MSH2 | ENST00000233146 | 0.84408 | 0.03667 | 0.69974 | 0.88732 | 0.94986 | 0.8738689608 | 0.91522 | 0.9756493568 | 0.285484 | 5.852521509 | 0 |
| 2 | 47478357 | A | G | I | V | MSH2 | ENST00000233146 | 0.77569 | 0.03911 | 0.58427 | 0.77764 | 0.88022 | 0.711542658 | 0.89812 | 0.8492775559 | 0.118205 | 3.897644343 | 0 |
| 2 | 47478357 | A | T | I | L | MSH2 | ENST00000233146 | 0.8376 | 0.03667 | 0.69974 | 0.88279 | 0.94986 | 0.8738689608 | 0.91539 | 0.9756493568 | 0.285484 | 5.852521509 | 0 |
| 2 | 47478358 | T | C | I | T | MSH2 | ENST00000233146 | 0.71137 | 0.03362 | 0.82561 | 0.92417 | 0.96993 | 0.9312617025 | 0.97901 | 0.9933027029 | 0.424896 | 6.763690481 | 0 |
| 2 | 47478358 | T | G | I | R | MSH2 | ENST00000233146 | 0.73717 | 0.02851 | 0.75244 | 0.94567 | 0.9755 | 0.983507699 | 0.97997 | 0.9990468621 | 0.4594 | 9.177626641 | 1 |
| 2 | 47478359 | A | G | I | M | MSH2 | ENST00000233146 | 0.67231 | 0.03686 | 0.65498 | 0.92381 | 0.92069 | 0.903346948 | 0.96004 | 0.9803379774 | 0.357937 | 5.297417607 | 0 |
| 2 | 47478360 | T | A | S | T | MSH2 | ENST00000233146 | 0.87716 | 0.03783 | 0.80488 | 0.8428 | 0.92561 | 0.8612544592 | 0.94656 | 0.9796941876 | 0.286897 | 4.651820965 | 0 |
| 2 | 47478360 | T | C | S | P | MSH2 | ENST00000233146 | 0.93437 | 0.03353 | 0.82206 | 0.90608 | 0.97773 | 0.9770513285 | 0.93967 | 0.9956653714 | 0.430422 | 8.803903776 | 0 |
| 2 | 47478360 | T | G | S | A | MSH2 | ENST00000233146 | 0.55965 | 0.02687 | 0.78651 | 0.75425 | 0.86503 | 0.7887431323 | 0.931 | 0.9592651725 | 0.219149 | 3.746117444 | 0 |
| 2 | 47478361 | C | T | S | L | MSH2 | ENST00000233146 | 0.69356 | 0.03848 | 0.82377 | 0.89759 | 0.93824 | 0.9289396719 | 0.95175 | 0.994717896 | 0.393119 | 4.690410875 | 0 |
| 2 | 47478363 | G | A | E | K | MSH2 | ENST00000233146 | 0.94828 | 0.00898 | 0.80589 | 0.72274 | 0.94129 | 0.8994591096 | 0.94627 | 0.9432387352 | 0.287395 | 5.605881278 | 0 |
| 2 | 47478363 | G | C | E | Q | MSH2 | ENST00000233146 | 0.80132 | 0.00935 | 0.6831 | 0.62815 | 0.911 | 0.8252421437 | 0.94809 | 0.7739057609 | 0.224699 | 5.317702724 | 0 |
| 2 | 47478364 | A | C | E | A | MSH2 | ENST00000233146 | 0.77275 | 0.01562 | 0.69547 | 0.80203 | 0.94986 | 0.878599022 | 0.92202 | 0.995831523 | 0.297039 | 7.594762614 | 0 |
| 2 | 47478364 | A | G | E | G | MSH2 | ENST00000233146 | 0.8203 | 0.01827 | 0.66177 | 0.86692 | 0.96262 | 0.8820893182 | 0.89833 | 0.99828583 | 0.308254 | 7.594762614 | 0 |
| 2 | 47478364 | A | T | E | V | MSH2 | ENST00000233146 | 0.81072 | 0.02809 | 0.67442 | 0.86034 | 0.96226 | 0.8803393502 | 0.93899 | 0.9972843742 | 0.325553 | 6.568752571 | 0 |
| 2 | 47478365 | A | C | E | D | MSH2 | ENST00000233146 | 0.35443 | 0.00638 | 0.69461 | 0.54029 | 0.87539 | 0.791839329 | 0.95795 | 0.7050864697 | 0.134151 | 3.606312291 | 0 |
| 2 | 47478365 | A | T | E | D | MSH2 | ENST00000233146 | 0.34975 | 0.00638 | 0.69461 | 0.54103 | 0.87539 | 0.791839329 | 0.9586 | 0.7376309633 | 0.134133 | 3.606312291 | 0 |
| 2 | 47478366 | T | A | Y | N | MSH2 | ENST00000233146 | 0.84195 | 0.02283 | 0.83869 | 0.91248 | 0.95097 | 0.7899314899 | 0.88927 | 0.9810124636 | 0.267611 | 6.92447234 | 0 |
| 2 | 47478366 | T | C | Y | H | MSH2 | ENST00000233146 | 0.39469 | 0.00547 | 0.84777 | 0.48845 | 0.73661 | 0.7144043478 | 0.84169 | 0.7080486417 | 0.0759455 | 1.75301022 | 0 |
| 2 | 47478366 | T | G | Y | D | MSH2 | ENST00000233146 | 0.71738 | 0.02342 | 0.83531 | 0.8894 | 0.96956 | 0.8523976459 | 0.91315 | 0.9891379476 | 0.346483 | 8.092355223 | 0 |
| 2 | 47478367 | A | C | Y | S | MSH2 | ENST00000233146 | 0.83637 | 0.01133 | 0.78645 | 0.87702 | 0.94986 | 0.7756225726 | 0.92524 | 0.9831761122 | 0.321066 | 6.988450275 | 0 |
| 2 | 47478367 | A | G | Y | C | MSH2 | ENST00000233146 | 0.90993 | 0.03212 | 0.84444 | 0.90636 | 0.97624 | 0.7979624543 | 0.89474 | 0.9937935073 | 0.330381 | 8.092355223 | 0 |
| 2 | 47478367 | A | T | Y | F | MSH2 | ENST00000233146 | 0.63773 | 0.03287 | 0.76663 | 0.80081 | 0.92397 | 0.6068329449 | 0.89072 | 0.9694856091 | 0.223167 | 3.330139562 | 0 |
| 2 | 47478369 | A | C | I | L | MSH2 | ENST00000233146 | 0.42591 | 0.01385 | 0.65935 | 0.67563 | 0.80476 | 0.7967933898 | 0.75329 | 0.948087275 | 0.115749 | 4.360631025 | 0 |
| 2 | 47478369 | A | G | I | V | MSH2 | ENST00000233146 | 0.40488 | 0.01158 | 0.54534 | 0.63097 | 0.73 | 0.6485275473 | 0.77866 | 0.1825806017 | -0.102732 | 3.871025109 | 0 |
| 2 | 47478369 | A | T | I | F | MSH2 | ENST00000233146 | 0.78946 | 0.02951 | 0.6656 | 0.88122 | 0.95535 | 0.9339273478 | 0.90817 | 0.9811457992 | 0.313959 | 9.268066786 | 0 |
| 2 | 47478370 | T | C | I | T | MSH2 | ENST00000233146 | 0.86699 | 0.03231 | 0.78615 | 0.91267 | 0.99628 | 0.9116160686 | 0.97417 | 0.9955095053 | 0.332906 | 8.171605227 | 0 |
| 2 | 47478370 | T | G | I | S | MSH2 | ENST00000233146 | 0.91508 | 0.03093 | 0.68404 | 0.94561 | 0.99676 | 0.9681948595 | 0.96409 | 0.9939991236 | 0.466208 | 9.268066786 | 0 |
| 2 | 47478371 | T | G | I | M | MSH2 | ENST00000233146 | 0.511 | 0.0364 | 0.61117 | 0.90325 | 0.89558 | 0.835151601 | 0.93182 | 0.9738433957 | 0.235211 | 6.689927242 | 0 |
| 2 | 47478372 | G | A | A | T | MSH2 | ENST00000233146 | 0.91007 | 0.03542 | 0.64469 | 0.85333 | 0.92561 | 0.8306737839 | 0.91291 | 0.9674347043 | 0.3558 | 5.44683005 | 0 |
| 2 | 47478372 | G | C | A | P | MSH2 | ENST00000233146 | 0.9175 | 0.03293 | 0.72061 | 0.8688 | 0.9608 | 0.9655184314 | 0.95958 | 0.9961924553 | 0.422724 | 8.150936354 | 0 |
| 2 | 47478372 | G | T | A | S | MSH2 | ENST00000233146 | 0.72308 | 0.01871 | 0.59432 | 0.67254 | 0.88401 | 0.7755306647 | 0.79615 | 0.847666502 | 0.189187 | 2.699391277 | 0 |
| 2 | 47478373 | C | A | A | E | MSH2 | ENST00000233146 | 0.74237 | 0.03353 | 0.71847 | 0.89755 | 0.97848 | 0.9655276855 | 0.93927 | 0.991187036 | 0.432291 | 7.200860174 | 0 |
| 2 | 47478373 | C | G | A | G | MSH2 | ENST00000233146 | 0.75169 | 0.03491 | 0.62295 | 0.87802 | 0.95753 | 0.8009042364 | 0.82136 | 0.9718921389 | 0.267169 | 5.864435477 | 0 |
| 2 | 47478373 | C | T | A | V | MSH2 | ENST00000233146 | 0.62098 | 0.02702 | 0.67086 | 0.81802 | 0.91862 | 0.8428184455 | 0.89506 | 0.9713303447 | 0.317685 | 3.539984907 | 0 |
| 2 | 47478375 | A | C | T | P | MSH2 | ENST00000233146 | 0.75702 | 0.02469 | 0.46032 | 0.81229 | 0.89422 | 0.8047662129 | 0.8758 | 0.9696210027 | 0.26288 | 7.854591528 | 1 |
| 2 | 47478375 | A | G | T | A | MSH2 | ENST00000233146 | 0.41222 | 0.00622 | 0.46016 | 0.70078 | 0.82948 | 0.4475897977 | 0.93725 | 0.7457924485 | 0.185298 | 5.820872688 | 0 |
| 2 | 47478375 | A | T | T | S | MSH2 | ENST00000233146 | 0.36019 | 0.00638 | 0.46567 | 0.44526 | 0.68292 | 0.3558169937 | 0.84904 | 0.6771199107 | 0.0791159 | 1.792776472 | 0 |
| 2 | 47478376 | C | A | T | K | MSH2 | ENST00000233146 | 0.2693 | 0.00586 | 0.58147 | 0.56357 | 0.7528 | 0.6238916073 | 0.91369 | 0.7961423397 | 0.182711 | 2.873320255 | 0 |
| 2 | 47478376 | C | G | T | R | MSH2 | ENST00000233146 | 0.31758 | 0.01866 | 0.57 | 0.69186 | 0.86852 | 0.661652073 | 0.90481 | 0.8197193146 | 0.0889498 | 5.013090757 | 0 |
| 2 | 47478376 | C | T | T | I | MSH2 | ENST00000233146 | 0.3406 | 0.00759 | 0.62083 | 0.66233 | 0.86453 | 0.5488225036 | 0.84272 | 0.9588689208 | 0.222345 | 5.767656152 | 0 |
| 2 | 47478378 | A | C | K | Q | MSH2 | ENST00000233146 | 0.39742 | 0.00615 | 0.45028 | 0.69517 | 0.77011 | 0.6782792889 | 0.98593 | 0.115406774 | 0.129925 | 3.548650281 | 0 |
| 2 | 47478378 | A | G | K | E | MSH2 | ENST00000233146 | 0.35278 | 0.00522 | 0.51979 | 0.68626 | 0.73877 | 0.7566405512 | 0.97701 | 0.042919207 | 0.0841429 | 3.8578659 | 0 |
| 2 | 47478379 | A | C | K | T | MSH2 | ENST00000233146 | 0.31962 | 0.00627 | 0.53749 | 0.63645 | 0.78286 | 0.7769145435 | 0.98564 | 0.1126528913 | 0.149722 | 4.695639205 | 0 |
| 2 | 47478379 | A | G | K | R | MSH2 | ENST00000233146 | 0.30755 | 0.00615 | 0.41941 | 0.64639 | 0.71705 | 0.6107994711 | 0.96567 | 0.05946811431 | 0.0569735 | 2.290698199 | 0 |
| 2 | 47478379 | A | T | K | M | MSH2 | ENST00000233146 | 0.60713 | 0.02231 | 0.44389 | 0.80859 | 0.84321 | 0.8029863617 | 0.9581 | 0.9253619909 | 0.197872 | 6.96049263 | 0 |
| 2 | 47478381 | A | C | I | L | MSH2 | ENST00000233146 | 0.56623 | 0.00681 | 0.68177 | 0.64396 | 0.81065 | 0.7734114678 | 0.8729 | 0.7863903046 | 0.108256 | 2.537269035 | 0 |
| 2 | 47478381 | A | G | I | V | MSH2 | ENST00000233146 | 0.63989 | 0.01203 | 0.60062 | 0.64717 | 0.83505 | 0.6267707588 | 0.86624 | 0.7903256416 | 0.162156 | 3.028151829 | 0 |
| 2 | 47478381 | A | T | I | F | MSH2 | ENST00000233146 | 0.89679 | 0.02628 | 0.69084 | 0.88077 | 0.94728 | 0.8902064617 | 0.88085 | 0.8903000355 | 0.148605 | 8.25839984 | 0 |
| 2 | 47478382 | T | A | I | N | MSH2 | ENST00000233146 | 0.94346 | 0.02785 | 0.77251 | 0.90838 | 0.96883 | 0.9390657615 | 0.82978 | 0.9913678765 | 0.280463 | 6.349073222 | 0 |
| 2 | 47478382 | T | C | I | T | MSH2 | ENST00000233146 | 0.77386 | 0.02366 | 0.79329 | 0.82431 | 0.96298 | 0.8576731502 | 0.93712 | 0.9949641228 | 0.382732 | 5.754181943 | 0 |
| 2 | 47478382 | T | G | I | S | MSH2 | ENST00000233146 | 0.94073 | 0.02979 | 0.70463 | 0.9107 | 0.98834 | 0.9412458815 | 0.93522 | 0.9949641228 | 0.4105 | 5.501981068 | 0 |
| 2 | 47478384 | G | A | G | S | MSH2 | ENST00000233146 | 0.48037 | 0.00608 | 0.43209 | 0.72299 | 0.82613 | 0.6712703418 | 0.93753 | 0.7427527308 | 0.118968 | 2.846646016 | 0 |
| 2 | 47478384 | G | C | G | R | MSH2 | ENST00000233146 | 0.38369 | 0.00543 | 0.54095 | 0.55546 | 0.77972 | 0.7252973325 | 0.93678 | 0.08844733288 | 0.0790281 | 2.13947674 | 0 |
| 2 | 47478384 | G | T | G | C | MSH2 | ENST00000233146 | 0.46599 | 0.00586 | 0.49988 | 0.60223 | 0.8233 | 0.7934095575 | 0.95828 | 0.8342577815 | 0.167029 | 2.062025083 | 0 |
| 2 | 47478385 | G | A | G | D | MSH2 | ENST00000233146 | 0.46513 | 0.00741 | 0.55859 | 0.6584 | 0.84052 | 0.7238656544 | 0.88656 | 0.9227118492 | 0.194574 | 6.470321237 | 0 |
| 2 | 47478385 | G | C | G | A | MSH2 | ENST00000233146 | 0.32262 | 0.00705 | 0.45556 | 0.6076 | 0.84052 | 0.6875717213 | 0.87862 | 0.747102797 | 0.154347 | 3.532147632 | 0 |
| 2 | 47478385 | G | T | G | V | MSH2 | ENST00000233146 | 0.4986 | 0.01015 | 0.45404 | 0.767 | 0.88776 | 0.7941051737 | 0.84748 | 0.9506778121 | 0.239211 | 6.593956307 | 0 |
| 2 | 47478387 | G | A | A | T | MSH2 | ENST00000233146 | 0.75145 | 0.0129 | 0.41512 | 0.72685 | 0.90537 | 0.6272851896 | 0.88215 | 0.9221340418 | 0.207178 | 5.152819939 | 0 |
| 2 | 47478387 | G | C | A | P | MSH2 | ENST00000233146 | 0.60618 | 0.00747 | 0.48914 | 0.71452 | 0.9328 | 0.8573194136 | 0.96479 | 0.9308298826 | 0.383241 | 4.029780392 | 0 |
| 2 | 47478387 | G | T | A | S | MSH2 | ENST00000233146 | 0.51969 | 0.00773 | 0.37185 | 0.62824 | 0.78658 | 0.5857114556 | 0.8862 | 0.7733242512 | 0.123896 | 2.146251678 | 0 |
| 2 | 47478388 | C | G | A | G | MSH2 | ENST00000233146 | 0.34817 | 0.00669 | 0.39963 | 0.55269 | 0.85378 | 0.626989732 | 0.88969 | 0.7746701837 | 0.173866 | 2.54736971 | 0 |
| 2 | 47478388 | C | T | A | V | MSH2 | ENST00000233146 | 0.62696 | 0.0245 | 0.43515 | 0.80072 | 0.92561 | 0.6522198095 | 0.90728 | 0.886074198 | 0.264411 | 4.555153106 | 0 |
| 2 | 47478390 | T | A | F | I | MSH2 | ENST00000233146 | 0.89463 | 0.03197 | 0.71416 | 0.90381 | 0.96993 | 0.9044211713 | 0.86427 | 0.990756554 | 0.27887 | 6.263247222 | 0 |
| 2 | 47478390 | T | C | F | L | MSH2 | ENST00000233146 | 0.90695 | 0.03409 | 0.75014 | 0.85525 | 0.93004 | 0.8770890969 | 0.8201 | 0.9893802404 | 0.222257 | 5.20396468 | 0 |
| 2 | 47478390 | T | G | F | V | MSH2 | ENST00000233146 | 0.899 | 0.03243 | 0.68939 | 0.93712 | 0.97178 | 0.8945840532 | 0.93326 | 0.9936327024 | 0.419772 | 7.233038583 | 0 |
| 2 | 47478391 | T | A | F | Y | MSH2 | ENST00000233146 | 0.70671 | 0.02589 | 0.73261 | 0.77091 | 0.89147 | 0.8143497787 | 0.8261 | 0.9288473725 | 0.189028 | 3.765401352 | 0 |
| 2 | 47478391 | T | C | F | S | MSH2 | ENST00000233146 | 0.93342 | 0.02802 | 0.72202 | 0.95014 | 0.93083 | 0.9397135647 | 0.95378 | 0.9840020112 | 0.405954 | 7.233038583 | 0 |

| | | | | | | | | | | | | | | | | | | |
|---|---|---|---|---|---|---|---|---|---|---|---|---|---|---|---|---|---|---|
| 2 | 47478391 | T | G | F | C | MSH2 | ENST00000233146 | 0.93351 | 0.03166 | 0.74597 | 0.94531 | 0.94431 | 0.930075438 | 0.85526 | 0.9975314311 | 0.30662 | 7.233038583 | 0 |
| 2 | 47478392 | T | A | F | L | MSH2 | ENST00000233146 | 0.78368 | 0.03409 | 0.75014 | 0.87338 | 0.85689 | 0.8770890969 | 0.8179 | 0.9866248369 | 0.174669 | 5.20396468 | 0 |
| 2 | 47478392 | T | G | F | L | MSH2 | ENST00000233146 | 0.79183 | 0.03409 | 0.75014 | 0.87338 | 0.85689 | 0.8770890969 | 0.81727 | 0.9866248369 | 0.174671 | 5.20396468 | 0 |
| 2 | 47478393 | T | A | C | S | MSH2 | ENST00000233146 | 0.64399 | 0.01574 | 0.71522 | 0.79506 | 0.9367 | 0.9064280655 | 0.8863 | 0.9834144711 | 0.30012 | 4.285337243 | 0 |
| 2 | 47478393 | T | C | C | R | MSH2 | ENST00000233146 | 0.94662 | 0.02653 | 0.83495 | 0.92602 | 0.9703 | 0.9781711752 | 0.96609 | 0.9980304837 | 0.403894 | 7.82843701 | 1 |
| 2 | 47478393 | T | G | C | G | MSH2 | ENST00000233146 | 0.82973 | 0.02741 | 0.67667 | 0.85216 | 0.95207 | 0.936391399 | 0.94448 | 0.9975485206 | 0.37918 | 7.82843701 | 0 |
| 2 | 47478394 | G | C | C | S | MSH2 | ENST00000233146 | 0.70423 | 0.01574 | 0.71522 | 0.72131 | 0.93824 | 0.9064280655 | 0.86871 | 0.9837101698 | 0.314004 | 4.285337243 | 0 |
| 2 | 47478394 | G | T | C | F | MSH2 | ENST00000233146 | 0.95219 | 0.02955 | 0.73724 | 0.88887 | 0.98869 | 0.9506822212 | 0.94158 | 0.9946607947 | 0.428411 | 7.82843701 | 0 |
| 2 | 47478395 | C | G | C | W | MSH2 | ENST00000233146 | 0.76594 | 0.03257 | 0.78971 | 0.91625 | 0.93863 | 0.9621067127 | 0.92564 | 0.9921129942 | 0.414746 | 7.82843701 | 1 |
| 2 | 47478396 | A | C | M | L | MSH2 | ENST00000233146 | 0.3652 | 0.00586 | 0.49635 | 0.43298 | 0.69678 | 0.6554639347 | 0.76652 | 0.2824585736 | 0.0343876 | 0.5828938255 | 0 |
| 2 | 47478396 | A | G | M | V | MSH2 | ENST00000233146 | 0.55745 | 0.00599 | 0.41552 | 0.71871 | 0.82216 | 0.6933154269 | 0.95134 | 0.7105050683 | 0.171537 | 3.079823595 | 0 |
| 2 | 47478396 | A | T | M | L | MSH2 | ENST00000233146 | 0.36053 | 0.00586 | 0.49635 | 0.4523 | 0.69678 | 0.6554639347 | 0.76648 | 0.2824585736 | 0.0344085 | 0.5828938255 | 0 |
| 2 | 47478397 | T | A | M | K | MSH2 | ENST00000233146 | 0.76164 | 0.01533 | 0.62205 | 0.87946 | 0.99006 | 0.9576953546 | 0.96599 | 0.9859977018 | 0.418795 | 6.672236603 | 0 |
| 2 | 47478397 | T | C | M | T | MSH2 | ENST00000233146 | 0.70495 | 0.01236 | 0.66835 | 0.82076 | 0.98257 | 0.8772463759 | 0.95919 | 0.9774370193 | 0.358474 | 6.672236603 | 0 |
| 2 | 47478397 | T | G | M | R | MSH2 | ENST00000233146 | 0.89955 | 0.02043 | 0.58699 | 0.86821 | 0.99525 | 0.9738130009 | 0.97829 | 0.9914829408 | 0.423038 | 6.672236603 | 1 |
| 2 | 47478398 | G | A | M | I | MSH2 | ENST00000233146 | 0.66644 | 0.00696 | 0.51107 | 0.60692 | 0.74093 | 0.6430409331 | 0.90839 | 0.6889499426 | 0.141018 | 4.087867256 | 0 |
| 2 | 47478398 | G | C | M | I | MSH2 | ENST00000233146 | 0.6632 | 0.00696 | 0.51107 | 0.60382 | 0.74093 | 0.6430409331 | 0.9091 | 0.8363871574 | 0.141015 | 4.087867256 | 0 |
| 2 | 47478398 | G | T | M | I | MSH2 | ENST00000233146 | 0.6568 | 0.00696 | 0.51107 | 0.60671 | 0.74093 | 0.6430409331 | 0.90893 | 0.8844825625 | 0.141016 | 4.087867256 | 0 |
| 2 | 47478399 | T | A | F | I | MSH2 | ENST00000233146 | 0.89729 | 0.03224 | 0.76278 | 0.96049 | 0.99443 | 0.9505173406 | 0.96436 | 0.9975352287 | 0.47472 | 9.699257667 | 0 |
| 2 | 47478399 | T | C | F | L | MSH2 | ENST00000233146 | 0.91059 | 0.03491 | 0.79743 | 0.93726 | 0.99234 | 0.9376179851 | 0.96359 | 0.9980117083 | 0.4629 | 9.699257667 | 0 |
| 2 | 47478399 | T | G | F | V | MSH2 | ENST00000233146 | 0.90175 | 0.03313 | 0.74476 | 0.96351 | 0.98904 | 0.9402939862 | 0.95964 | 0.9950419664 | 0.471843 | 7.107327167 | 0 |
| 2 | 47478400 | T | A | F | Y | MSH2 | ENST00000233146 | 0.88511 | 0.03527 | 0.77613 | 0.94817 | 0.97067 | 0.9051734692 | 0.95222 | 0.9993189573 | 0.435493 | 9.699257667 | 0 |
| 2 | 47478400 | T | C | F | S | MSH2 | ENST00000233146 | 0.93451 | 0.02864 | 0.76696 | 0.97687 | 0.99525 | 0.9740738793 | 0.97782 | 0.9998369217 | 0.479526 | 9.699257667 | 0 |
| 2 | 47478400 | T | G | F | C | MSH2 | ENST00000233146 | 0.93429 | 0.03175 | 0.80103 | 0.97158 | 0.99498 | 0.9671776758 | 0.97214 | 0.9998010993 | 0.480959 | 9.699257667 | 0 |
| 2 | 47478401 | T | A | F | L | MSH2 | ENST00000233146 | 0.7548 | 0.03491 | 0.79743 | 0.9452 | 0.96993 | 0.9376179851 | 0.97136 | 0.9977545142 | 0.430606 | 9.699257667 | 0 |
| 2 | 47478401 | T | G | F | L | MSH2 | ENST00000233146 | 0.76299 | 0.03491 | 0.79743 | 0.94659 | 0.96993 | 0.9376179851 | 0.97119 | 0.9977545142 | 0.430609 | 9.699257667 | 0 |
| 2 | 47478402 | G | A | A | T | MSH2 | ENST00000233146 | 0.90974 | 0.03767 | 0.70669 | 0.93159 | 0.96444 | 0.9167154276 | 0.93395 | 0.9970381669 | 0.425559 | 8.789281701 | 0 |
| 2 | 47478402 | G | C | A | P | MSH2 | ENST00000233146 | 0.91648 | 0.03334 | 0.77496 | 0.96315 | 0.98764 | 0.9868125682 | 0.96915 | 0.9995937943 | 0.475343 | 10.00680778 | 0 |
| 2 | 47478402 | G | T | A | S | MSH2 | ENST00000233146 | 0.88291 | 0.03857 | 0.6626 | 0.92125 | 0.93786 | 0.874549911 | 0.9214 | 0.9920294285 | 0.313811 | 4.906576251 | 0 |
| 2 | 47478403 | C | A | A | E | MSH2 | ENST00000233146 | 0.68283 | 0.03491 | 0.77632 | 0.97331 | 0.99039 | 0.9857400525 | 0.96588 | 0.9995189905 | 0.465095 | 10.00680778 | 0 |
| 2 | 47478403 | C | G | A | G | MSH2 | ENST00000233146 | 0.69468 | 0.03713 | 0.69032 | 0.96519 | 0.97141 | 0.8936010079 | 0.94501 | 0.9992432594 | 0.462007 | 10.00680778 | 0 |
| 2 | 47478403 | C | T | A | V | MSH2 | ENST00000233146 | 0.6974 | 0.03728 | 0.73006 | 0.94431 | 0.96737 | 0.9330583129 | 0.93323 | 0.9956490397 | 0.433317 | 10.00680778 | 0 |
| 2 | 47478405 | A | C | T | P | MSH2 | ENST00000233146 | 0.93217 | 0.03304 | 0.55648 | 0.97647 | 0.98585 | 0.981605988 | 0.98532 | 0.9991099238 | 0.49215 | 9.98680166 | 1 |
| 2 | 47478405 | A | G | T | A | MSH2 | ENST00000233146 | 0.91441 | 0.03534 | 0.5661 | 0.96723 | 0.97773 | 0.9584535051 | 0.98725 | 0.9991099238 | 0.486573 | 9.98680166 | 0 |
| 2 | 47478405 | A | T | T | S | MSH2 | ENST00000233146 | 0.88517 | 0.03189 | 0.57854 | 0.96099 | 0.96993 | 0.9065854133 | 0.93797 | 0.9897213578 | 0.309453 | 9.98680166 | 0 |
| 2 | 47478406 | C | A | T | N | MSH2 | ENST00000233146 | 0.61306 | 0.03458 | 0.66401 | 0.9813 | 0.96044 | 0.9737751718 | 0.9348 | 0.9987970591 | 0.290681 | 9.98680166 | 0 |
| 2 | 47478406 | C | G | T | S | MSH2 | ENST00000233146 | 0.60353 | 0.03189 | 0.57854 | 0.96177 | 0.97104 | 0.9065854133 | 0.9253 | 0.9873883995 | 0.313156 | 9.98680166 | 0 |
| 2 | 47478406 | C | T | T | I | MSH2 | ENST00000233146 | 0.65531 | 0.03582 | 0.72128 | 0.97667 | 0.99296 | 0.9704934379 | 0.97748 | 0.9987234473 | 0.49514 | 9.98680166 | 1 |
| 2 | 47478408 | C | A | H | N | MSH2 | ENST00000233146 | 0.75694 | 0.03582 | 0.73786 | 0.95663 | 0.98693 | 0.9550386191 | 0.96847 | 0.998459816 | 0.551902 | 9.702263997 | 0 |
| 2 | 47478408 | C | G | H | D | MSH2 | ENST00000233146 | 0.79286 | 0.03243 | 0.72928 | 0.94816 | 0.99765 | 0.9832544359 | 0.98403 | 0.9995474219 | 0.522626 | 9.702263997 | 0 |
| 2 | 47478408 | C | T | H | Y | MSH2 | ENST00000233146 | 0.74898 | 0.03497 | 0.7452 | 0.95877 | 0.99806 | 0.9685229268 | 0.98786 | 0.99887079 | 0.497794 | 8.443988985 | 0 |
| 2 | 47478409 | A | C | H | P | MSH2 | ENST00000233146 | 0.85566 | 0.03059 | 0.66643 | 0.95596 | 0.99962 | 0.9832400314 | 0.99189 | 0.9999572039 | 0.468606 | 9.702263997 | 0 |
| 2 | 47478409 | A | G | H | R | MSH2 | ENST00000233146 | 0.81103 | 0.03419 | 0.74651 | 0.95693 | 0.99953 | 0.9814738686 | 0.99496 | 0.999855876 | 0.459096 | 9.702263997 | 1 |
| 2 | 47478409 | A | T | H | L | MSH2 | ENST00000233146 | 0.85851 | 0.03398 | 0.70911 | 0.97165 | 0.99953 | 0.9637893678 | 0.99385 | 0.9999657869 | 0.530894 | 9.702263997 | 0 |
| 2 | 47478410 | T | A | H | Q | MSH2 | ENST00000233146 | 0.66169 | 0.03713 | 0.71327 | 0.8856 | 0.9781 | 0.9579449821 | 0.99685 | 0.9997656941 | 0.541295 | 9.702263997 | 0 |
| 2 | 47478410 | T | G | H | Q | MSH2 | ENST00000233146 | 0.66984 | 0.03713 | 0.71327 | 0.88721 | 0.9781 | 0.9579449821 | 0.99682 | 0.9997656941 | 0.541298 | 9.702263997 | 0 |
| 2 | 47478411 | T | A | F | I | MSH2 | ENST00000233146 | 0.89509 | 0.03224 | 0.76273 | 0.95978 | 0.99498 | 0.9540678575 | 0.97165 | 0.9995732903 | 0.475241 | 8.416217105 | 0 |
| 2 | 47478411 | T | C | F | L | MSH2 | ENST00000233146 | 0.90867 | 0.03491 | 0.80751 | 0.939 | 0.98869 | 0.9290510521 | 0.95045 | 0.9980117083 | 0.450069 | 8.416217105 | 0 |
| 2 | 47478411 | T | G | F | V | MSH2 | ENST00000233146 | 0.89966 | 0.03313 | 0.75949 | 0.88396 | 0.99676 | 0.9520432263 | 0.97792 | 0.9993810654 | 0.474989 | 8.416217105 | 0 |
| 2 | 47478412 | T | A | F | Y | MSH2 | ENST00000233146 | 0.88316 | 0.03527 | 0.78576 | 0.87093 | 0.92069 | 0.9021196183 | 0.92543 | 0.9894894958 | 0.509493 | 3.055489109 | 0 |
| 2 | 47478412 | T | C | F | S | MSH2 | ENST00000233146 | 0.93363 | 0.02864 | 0.77489 | 0.92748 | 0.99578 | 0.9674807487 | 0.98041 | 0.9998371601 | 0.507212 | 8.416217105 | 0 |
| 2 | 47478412 | T | G | F | C | MSH2 | ENST00000233146 | 0.93339 | 0.03175 | 0.8096 | 0.84923 | 0.99676 | 0.9659125997 | 0.97768 | 0.9990566373 | 0.477567 | 8.416217105 | 1 |
| 2 | 47478413 | T | A | F | L | MSH2 | ENST00000233146 | 0.69477 | 0.03491 | 0.80751 | 0.86 | 0.96664 | 0.9290510521 | 0.95901 | 0.9977198243 | 0.411249 | 8.416217105 | 0 |
| 2 | 47478413 | T | G | F | L | MSH2 | ENST00000233146 | 0.70302 | 0.03491 | 0.80751 | 0.9202 | 0.96664 | 0.9290510521 | 0.95463 | 0.9977198243 | 0.445143 | 8.416217105 | 0 |
| 2 | 47478414 | C | A | H | N | MSH2 | ENST00000233146 | 0.76021 | 0.03582 | 0.75241 | 0.912 | 0.96371 | 0.9237708656 | 0.94343 | 0.9911337495 | 0.504901 | 9.517710919 | 0 |
| 2 | 47478414 | C | G | H | D | MSH2 | ENST00000233146 | 0.79618 | 0.03243 | 0.74741 | 0.92764 | 0.99105 | 0.9712159077 | 0.96051 | 0.9995482564 | 0.490405 | 9.517710919 | 0 |
| 2 | 47478414 | C | T | H | Y | MSH2 | ENST00000233146 | 0.74754 | 0.03497 | 0.75832 | 0.94996 | 0.99202 | 0.9492790079 | 0.96255 | 0.9934989214 | 0.463806 | 7.328433751 | 0 |
| 2 | 47478415 | A | C | H | P | MSH2 | ENST00000233146 | 0.86119 | 0.03059 | 0.67755 | 0.82637 | 0.99471 | 0.9742755424 | 0.977 | 0.9948105812 | 0.316718 | 9.517710919 | 0 |
| 2 | 47478415 | A | G | H | R | MSH2 | ENST00000233146 | 0.81669 | 0.03419 | 0.76286 | 0.86317 | 0.99722 | 0.9669526984 | 0.98327 | 0.9991440773 | 0.464701 | 9.517710919 | 0 |
| 2 | 47478415 | A | T | H | L | MSH2 | ENST00000233146 | 0.86382 | 0.03398 | 0.72158 | 0.84542 | 0.99676 | 0.9490443219 | 0.98207 | 0.9998354912 | 0.460267 | 9.517710919 | 0 |
| 2 | 47478416 | T | A | H | Q | MSH2 | ENST00000233146 | 0.63229 | 0.03713 | 0.72826 | 0.84425 | 0.9608 | 0.9058063308 | 0.98072 | 0.9991939664 | 0.424394 | 9.517710919 | 0 |
| 2 | 47478416 | T | G | H | Q | MSH2 | ENST00000233146 | 0.6402 | 0.03713 | 0.72826 | 0.83406 | 0.9608 | 0.9058063308 | 0.9806 | 0.9990584254 | 0.424397 | 9.517710919 | 0 |
| 2 | 47478417 | G | C | E | Q | MSH2 | ENST00000233146 | 0.89718 | 0.03735 | 0.76717 | 0.88528 | 0.97215 | 0.9053370828 | 0.979 | 0.9934555888 | 0.467619 | 6.212598254 | 0 |
| 2 | 47478418 | A | C | E | A | MSH2 | ENST00000233146 | 0.88958 | 0.03626 | 0.77985 | 0.84026 | 0.99652 | 0.9371065839 | 0.97874 | 0.9967896342 | 0.47786 | 9.145965111 | 0 |
| 2 | 47478418 | A | G | E | G | MSH2 | ENST00000233146 | 0.91958 | 0.03484 | 0.72999 | 0.88405 | 0.99676 | 0.9349873181 | 0.96955 | 0.9980869293 | 0.482329 | 9.145965111 | 0 |
| 2 | 47478418 | A | T | E | V | MSH2 | ENST00000233146 | 0.90944 | 0.03484 | 0.75949 | 0.89184 | 0.99628 | 0.9339962443 | 0.98788 | 0.9994206429 | 0.483341 | 9.145965111 | 0 |
| 2 | 47478419 | A | C | E | D | MSH2 | ENST00000233146 | 0.58724 | 0.03864 | 0.78228 | 0.84329 | 0.95353 | 0.8911575306 | 0.98716 | 0.9862996936 | 0.403457 | 9.145965111 | 0 |
| 2 | 47478419 | A | T | E | D | MSH2 | ENST00000233146 | 0.58126 | 0.03864 | 0.78228 | 0.85013 | 0.95353 | 0.8911575306 | 0.98719 | 0.9858486652 | 0.28571 | 9.145965111 | 0 |
| 2 | 47478420 | C | A | L | I | MSH2 | ENST00000233146 | 0.50478 | 0.03527 | 0.74065 | 0.79649 | 0.93044 | 0.7328262324 | 0.86957 | 0.9222740918 | 0.257667 | 6.169662153 | 0 |
| 2 | 47478420 | C | G | L | V | MSH2 | ENST00000233146 | 0.66771 | 0.03633 | 0.68716 | 0.81953 | 0.96993 | 0.8643563571 | 0.95894 | 0.9715491453 | 0.430513 | 9.566875758 | 0 |
| 2 | 47478420 | C | T | L | F | MSH2 | ENST00000233146 | 0.71249 | 0.03582 | 0.7246 | 0.91986 | 0.97252 | 0.9002717659 | 0.96473 | 0.9965991378 | 0.457131 | 9.566875758 | 0 |
| 2 | 47478421 | T | A | L | H | MSH2 | ENST00000233146 | 0.88086 | 0.0334 | 0.80976 | 0.8932 | 0.99676 | 0.9573430227 | 0.98321 | 0.9994257689 | 0.496961 | 9.566875758 | 0 |
| 2 | 47478421 | T | C | L | P | MSH2 | ENST00000233146 | 0.89927 | 0.03001 | 0.84064 | 0.97511 | 0.99443 | 0.9870592487 | 0.98358 | 0.9996283054 | 0.49696 | 9.566875758 | 1 |
| 2 | 47478421 | T | G | L | R | MSH2 | ENST00000233146 | 0.896 | 0.03137 | 0.81317 | 0.84514 | 0.99603 | 0.9872319375 | 0.98519 | 0.9990623593 | 0.496413 | 9.566875758 | 1 |
| 2 | 47478423 | A | C | T | P | MSH2 | ENST00000233146 | 0.86403 | 0.03327 | 0.51314 | 0.89896 | 0.99296 | 0.9545012045 | 0.96974 | 0.9956353075 | 0.462623 | 8.942822606 | 0 |
| 2 | 47478423 | A | G | T | A | MSH2 | ENST00000233146 | 0.82887 | 0.03747 | 0.51533 | 0.91433 | 0.98513 | 0.870104263 | 0.95607 | 0.9956704378 | 0.4123 | 5.96198986 | 0 |
| 2 | 47478423 | A | T | T | S | MSH2 | ENST00000233146 | 0.79239 | 0.03882 | 0.52989 | 0.89963 | 0.96919 | 0.7719963563 | 0.9266 | 0.9940032363 | 0.465616 | 5.779888016 | 0 |
| 2 | 47478424 | C | A | T | N | MSH2 | ENST00000233146 | 0.60083 | 0.03518 | 0.6207 | 0.86507 | 0.93475 | 0.92905791 | 0.88136 | 0.9958301187 | 0.516176 | 5.601421608 | 0 |
| 2 | 47478424 | C | G | T | S | MSH2 | ENST00000233146 | 0.59769 | 0.03882 | 0.52989 | 0.8744 | 0.95899 | 0.7719963563 | 0.89756 | 0.9890588374 | 0.483456 | 5.779888016 | 0 |
| 2 | 47478424 | C | T | T | I | MSH2 | ENST00000233146 | 0.6419 | 0.03608 | 0.68452 | 0.92143 | 0.98071 | 0.9148618051 | 0.94041 | 0.9943659306 | 0.45884 | 5.461810771 | 0 |
| 2 | 47478426 | G | A | A | T | MSH2 | ENST00000233146 | 0.57167 | 0.021 | 0.59816 | 0.69096 | 0.90755 | 0.6431706837 | 0.94085 | 0.8870930076 | 0.352718 | 4.197596606 | 0 |
| 2 | 47478426 | G | C | A | P | MSH2 | ENST00000233146 | 0.89834 | 0.03265 | 0.66783 | 0.82534 | 0.96773 | 0.8972810965 | 0.94125 | 0.9865312207 | 0.438646 | 7.501003605 | 0 |
| 2 | 47478426 | G | T | A | S | MSH2 | ENST00000233146 | 0.51041 | 0.01475 | 0.55162 | 0.68966 | 0.84375 | 0.5701927804 | 0.93701 | 0.7873528004 | 0.288487 | 3.08920543 | 0 |
| 2 | 47478427 | C | A | A | D | MSH2 | ENST00000233146 | 0.47476 | 0.03026 | 0.71662 | 0.55648 | 0.91015 | 0.848945399 | 0.92823 | 0.9813868403 | 0.378197 | 4.035588911 | 0 |
| 2 | 47478427 | C | G | A | G | MSH2 | ENST00000233146 | 0.60544 | 0.0325 | 0.57799 | 0.50858 | 0.90624 | 0.5856051845 | 0.93008 | 0.9282681346 | 0.338637 | 5.575666108 | 0 |
| 2 | 47478427 | C | T | A | V | MSH2 | ENST00000233146 | 0.46724 | 0.02484 | 0.62856 | 0.58429 | 0.90624 | 0.7099982916 | 0.92821 | 0.9653258324 | 0.363689 | 4.047234504 | 0 |
| 2 | 47478429 | T | A | L | M | MSH2 | ENST00000233146 | 0.63481 | 0.03653 | 0.6176 | 0.85069 | 0.89784 | 0.7274962224 | 0.88026 | 0.9041213099 | 0.240657 | 5.390796566 | 0 |
| 2 | 47478429 | T | G | L | V | MSH2 | ENST00000233146 | 0.61261 | 0.03633 | 0.57982 | 0.94182 | 0.93863 | 0.8052527022 | 0.90011 | 0.9801636143 | 0.271088 | 8.689136882 | 0 |
| 2 | 47478430 | T | C | L | S | MSH2 | ENST00000233146 | 0.929 | 0.03212 | 0.66229 | 0.82303 | 0.99072 | 0.933626313 | 0.96116 | 0.9922586083 | 0.465226 | 9.221706584 | 0 |
| 2 | 47478430 | T | G | L | W | MSH2 | ENST00000233146 | 0.92953 | 0.02984 | 0.73089 | 0.87274 | 0.97401 | 0.9349870978 | 0.90943 | 0.9995860457 | 0.394236 | 9.221706584 | 0 |
| 2 | 47478431 | G | C | L | F | MSH2 | ENST00000233146 | 0.79414 | 0.03582 | 0.6335 | 0.8616 | 0.94281 | 0.8409303953 | 0.93599 | 0.9843575008 | 0.410841 | 9.221706584 | 0 |
| 2 | 47478431 | G | T | L | F | MSH2 | ENST00000233146 | 0.78772 | 0.03582 | 0.6335 | 0.8276 | 0.94243 | 0.8409303953 | 0.94093 | 0.9837755533 | 0.410828 | 9.221706584 | 0 |
| 2 | 47478432 | G | C | A | P | MSH2 | ENST00000233146 | 0.94947 | 0.03243 | 0.67755 | 0.67845 | 0.97624 | 0.927931467 | 0.93187 | 0.9877764382 | 0.421964 | 7.729688814 | 0 |
| 2 | 47478432 | G | T | A | S | MSH2 | ENST00000233146 | 0.80542 | 0.01649 | 0.55846 | 0.67751 | 0.90406 | 0.7005309119 | 0.8655 | 0.8623700142 | 0.220869 | 3.283469122 | 0 |
| 2 | 47478433 | C | A | A | D | MSH2 | ENST00000233146 | 0.51481 | 0.01244 | 0.72297 | 0.67777 | 0.90798 | 0.9279798721 | 0.81291 | 0.9806116223 | 0.247246 | 4.591725325 | 0 |
| 2 | 47478433 | C | G | A | G | MSH2 | ENST00000233146 | 0.66882 | 0.03293 | 0.58246 | 0.45999 | 0.91357 | 0.6934224141 | 0.85904 | 0.956420881 | 0.252963 | 5.685904306 | 0 |
| 2 | 47478433 | C | T | A | V | MSH2 | ENST00000233146 | 0.6712 | 0.03576 | 0.62921 | 0.52519 | 0.9211 | 0.8092478721 | 0.85082 | 0.974966526 | 0.272213 | 8.161802668 | 0 |
| 2 | 47478435 | A | C | N | H | MSH2 | ENST00000233146 | 0.49835 | 0.00569 | 0.09883 | 0.44526 | 0.61416 | 0.445031828 | 0.91859 | 0.3194538636 | 0.0615884 | 2.820886782 | 0 |
| 2 | 47478435 | A | G | N | D | MSH2 | ENST00000233146 | 0.31426 | 0.00547 | 0.12145 | 0.34707 | 0.60123 | 0.3961378528 | 0.80008 | 0.09243871516 | -0.00805506 | -2.026247945 | 0 |
| 2 | 47478435 | A | T | N | Y | MSH2 | ENST00000233146 | 0.5446 | 0.00747 | 0.12159 | 0.70909 | 0.77782 | 0.5278471387 | 0.94686 | 0.8917310834 | 0.131302 | 4.787139852 | 0 |
| 2 | 47478436 | A | C | N | T | MSH2 | ENST00000233146 | 0.27793 | 0.0063 | 0.08926 | 0.36313 | 0.73661 | 0.4233168071 | 0.92486 | 0.390361293 | 0.165633 | 2.066583453 | 0 |

| | | | | | | | | | | | | | | | | | |
|---|---|---|---|---|---|---|---|---|---|---|---|---|---|---|---|---|---|
| 2 | 47478436 | A | G | N | S | MSH2 | ENST00000233146 | 0.17649 | 0.00608 | 0.07687 | 0.13032 | 0.62325 | 0.3195787644 | 0.85463 | 0.09015730023 | -0.15847 | 0.6246602973 | 0 |
| 2 | 47478436 | A | T | N | I | MSH2 | ENST00000233146 | 0.31634 | 0.00777 | 0.11058 | 0.64194 | 0.77527 | 0.5145010508 | 0.91214 | 0.9464151859 | 0.264085 | 3.467232925 | 0 |
| 2 | 47478437 | T | A | N | K | MSH2 | ENST00000233146 | 0.39539 | 0.00608 | 0.13302 | 0.37399 | 0.68995 | 0.3941044081 | 0.87754 | 0.3337592483 | 0.0637184 | 1.509960732 | 0 |
| 2 | 47478437 | T | G | N | K | MSH2 | ENST00000233146 | 0.40133 | 0.00608 | 0.13302 | 0.36807 | 0.68995 | 0.3941044081 | 0.87771 | 0.5971556306 | 0.150132 | 1.509960732 | 0 |
| 2 | 47478438 | C | A | Q | K | MSH2 | ENST00000233146 | 0.48487 | 0.0054 | 0.48422 | 0.60536 | 0.73443 | 0.741497302 | 0.95212 | 0.7451350093 | 0.186245 | 2.851725287 | 0 |
| 2 | 47478438 | C | G | Q | E | MSH2 | ENST00000233146 | 0.34375 | 0.00556 | 0.41103 | 0.15757 | 0.66639 | 0.7376397598 | 0.89525 | 0.3416785447 | -0.0598257 | 2.060679079 | 0 |
| 2 | 47478439 | A | C | Q | P | MSH2 | ENST00000233146 | 0.71249 | 0.0132 | 0.43017 | 0.80575 | 0.93824 | 0.9331930487 | 0.96484 | 0.9632985592 | 0.349568 | 6.023020213 | 0 |
| 2 | 47478439 | A | G | Q | R | MSH2 | ENST00000233146 | 0.53758 | 0.00816 | 0.47751 | 0.48264 | 0.80178 | 0.7734055325 | 0.93904 | 0.7799216509 | 0.215155 | 3.695307887 | 0 |
| 2 | 47478439 | A | T | Q | L | MSH2 | ENST00000233146 | 0.70639 | 0.00747 | 0.40545 | 0.56225 | 0.84588 | 0.75752691 | 0.94086 | 0.9733062387 | 0.230498 | 3.84875948 | 0 |
| 2 | 47478440 | G | C | Q | H | MSH2 | ENST00000233146 | 0.31519 | 0.00615 | 0.46279 | 0.3479 | 0.80238 | 0.7759449939 | 0.96934 | 0.7775027156 | 0.153886 | 3.118836169 | 0 |
| 2 | 47478440 | G | T | Q | H | MSH2 | ENST00000233146 | 0.311 | 0.00615 | 0.46279 | 0.35793 | 0.80238 | 0.7759449939 | 0.96936 | 0.2590015952 | 0.143099 | 3.118836169 | 0 |
| 2 | 47478441 | A | C | I | L | MSH2 | ENST00000233146 | 0.36542 | 0.00574 | 0.45346 | 0.06666 | 0.70671 | 0.483050239 | 0.86066 | 0.1723011298 | 0.0379165 | 0.7031748119 | 0 |
| 2 | 47478441 | A | G | I | V | MSH2 | ENST00000233146 | 0.24659 | 0.00603 | 0.34492 | 0.31501 | 0.65476 | 0.3510741531 | 0.7999 | 0.06319021153 | 0.030762 | -0.7162127396 | 0 |
| 2 | 47478441 | A | T | I | L | MSH2 | ENST00000233146 | 0.36075 | 0.00574 | 0.45346 | 0.06996 | 0.70671 | 0.483050239 | 0.86106 | 0.1821608186 | 0.0379388 | 0.7031748119 | 0 |
| 2 | 47478442 | T | C | I | T | MSH2 | ENST00000233146 | 0.60742 | 0.00747 | 0.61883 | 0.38471 | 0.84482 | 0.6004206968 | 0.74682 | 0.4707397223 | 0.153402 | 5.997391787 | 0 |
| 2 | 47478442 | T | G | I | R | MSH2 | ENST00000233146 | 0.48608 | 0.00497 | 0.51778 | 0.41727 | 0.80713 | 0.739743244 | 0.82339 | 0.3772703426 | 0.116079 | 3.331738601 | 0 |
| 2 | 47478443 | A | G | I | M | MSH2 | ENST00000233146 | 0.53569 | 0.01295 | 0.41224 | 0.28187 | 0.82273 | 0.5183564309 | 0.87963 | 0.3266387941 | 0.0745184 | 2.817799964 | 1 |
| 2 | 47478444 | C | A | P | T | MSH2 | ENST00000233146 | 0.74452 | 0.00898 | 0.67223 | 0.6317 | 0.9182 | 0.8050640941 | 0.91433 | 0.9813396335 | 0.35833 | 3.29702531 | 0 |
| 2 | 47478444 | C | G | P | A | MSH2 | ENST00000233146 | 0.72324 | 0.00687 | 0.61086 | 0.53573 | 0.89648 | 0.7492901927 | 0.86655 | 0.9780678153 | 0.316616 | 3.311499625 | 0 |
| 2 | 47478444 | C | T | P | S | MSH2 | ENST00000233146 | 0.59161 | 0.00715 | 0.63128 | 0.76264 | 0.89558 | 0.7979042441 | 0.86558 | 0.9857909679 | 0.307349 | 2.38547727 | 0 |
| 2 | 47478445 | C | A | P | Q | MSH2 | ENST00000233146 | 0.6624 | 0.02864 | 0.63099 | 0.78009 | 0.96444 | 0.8347953719 | 0.89583 | 0.9947682103 | 0.398666 | 4.339856402 | 0 |
| 2 | 47478445 | C | G | P | R | MSH2 | ENST00000233146 | 0.67692 | 0.02225 | 0.70723 | 0.74303 | 0.98071 | 0.8528843969 | 0.94129 | 0.9920542065 | 0.432167 | 6.124251875 | 0 |
| 2 | 47478445 | C | T | P | L | MSH2 | ENST00000233146 | 0.6962 | 0.01915 | 0.70979 | 0.90238 | 0.96407 | 0.8709971769 | 0.91112 | 0.9959387779 | 0.419141 | 6.723289303 | 0 |
| 2 | 47478447 | A | C | T | P | MSH2 | ENST00000233146 | 0.86204 | 0.02998 | 0.41262 | 0.82182 | 0.89873 | 0.9007215416 | 0.88319 | 0.9401049018 | 0.328223 | 7.713430539 | 0 |
| 2 | 47478447 | A | G | T | A | MSH2 | ENST00000233146 | 0.4824 | 0.00681 | 0.41792 | 0.74902 | 0.78658 | 0.7357667752 | 0.92802 | 0.3995046914 | 0.115296 | 3.690507699 | 0 |
| 2 | 47478447 | A | T | T | S | MSH2 | ENST00000233146 | 0.44411 | 0.00791 | 0.42607 | 0.43954 | 0.77527 | 0.6156126404 | 0.91075 | 0.3458469328 | -0.00914875 | 1.993456743 | 0 |
| 2 | 47478448 | C | A | T | N | MSH2 | ENST00000233146 | 0.39702 | 0.01221 | 0.52314 | 0.50522 | 0.74728 | 0.7885081847 | 0.79375 | 0.7089007497 | 0.276508 | 3.759650036 | 0 |
| 2 | 47478448 | C | G | T | S | MSH2 | ENST00000233146 | 0.26776 | 0.00791 | 0.42607 | 0.41305 | 0.7609 | 0.6156126404 | 0.89735 | 0.2983458139 | -0.113069 | 1.993456743 | 0 |
| 2 | 47478448 | C | T | T | I | MSH2 | ENST00000233146 | 0.54068 | 0.03129 | 0.59863 | 0.82271 | 0.8543 | 0.8023549874 | 0.87485 | 0.9639981985 | 0.211435 | 6.201548117 | 0 |
| 2 | 47478450 | G | A | V | I | MSH2 | ENST00000233146 | 0.82287 | 0.03783 | 0.59717 | 0.78466 | 0.86852 | 0.6171624051 | 0.84001 | 0.9024502551 | 0.187917 | 4.142436192 | 0 |
| 2 | 47478450 | G | C | V | L | MSH2 | ENST00000233146 | 0.85632 | 0.03527 | 0.78826 | 0.86855 | 0.94876 | 0.8343572189 | 0.95919 | 0.9663136981 | 0.396958 | 4.659679671 | 0 |
| 2 | 47478450 | G | T | V | F | MSH2 | ENST00000233146 | 0.88843 | 0.03281 | 0.76464 | 0.91734 | 0.97661 | 0.9464806923 | 0.96231 | 0.9943522793 | 0.472507 | 8.001550049 | 0 |
| 2 | 47478451 | T | A | V | D | MSH2 | ENST00000233146 | 0.7935 | 0.02984 | 0.81523 | 0.93449 | 0.96956 | 0.9749550741 | 0.98419 | 0.9990262985 | 0.46959 | 9.048567907 | 1 |
| 2 | 47478451 | T | C | V | A | MSH2 | ENST00000233146 | 0.58304 | 0.02115 | 0.80656 | 0.92024 | 0.94167 | 0.8426224423 | 0.96114 | 0.9337893724 | 0.385951 | 5.346840073 | 0 |
| 2 | 47478451 | T | G | V | G | MSH2 | ENST00000233146 | 0.77766 | 0.03243 | 0.66633 | 0.93783 | 0.97215 | 0.9600068227 | 0.9528 | 0.997677505 | 0.440472 | 9.048567907 | 0 |
| 2 | 47478453 | A | C | N | H | MSH2 | ENST00000233146 | 0.50779 | 0.00574 | 0.41815 | 0.27979 | 0.613 | 0.5674914681 | 0.88939 | 0.7031762004 | -0.00858833 | 3.196040718 | 0 |
| 2 | 47478453 | A | G | N | D | MSH2 | ENST00000233146 | 0.52107 | 0.00566 | 0.47947 | 0.77489 | 0.75005 | 0.6235056052 | 0.86106 | 0.8020032048 | 0.0621843 | 5.780215738 | 0 |
| 2 | 47478453 | A | T | N | Y | MSH2 | ENST00000233146 | 0.72909 | 0.00592 | 0.46223 | 0.75112 | 0.88448 | 0.6821646761 | 0.88839 | 0.9564487934 | 0.209354 | 6.450362811 | 0 |
| 2 | 47478454 | A | C | N | T | MSH2 | ENST00000233146 | 0.3286 | 0.00574 | 0.38434 | 0.34274 | 0.72093 | 0.5311733255 | 0.8995 | 0.7485255599 | -0.0113953 | 2.711121915 | 0 |
| 2 | 47478454 | A | G | N | S | MSH2 | ENST00000233146 | 0.27904 | 0.00615 | 0.32625 | 0.35199 | 0.63202 | 0.4669798549 | 0.81275 | 0.6758718491 | -0.0671167 | 3.03069941 | 0 |
| 2 | 47478454 | A | T | N | I | MSH2 | ENST00000233146 | 0.41997 | 0.00861 | 0.41642 | 0.51333 | 0.83505 | 0.6469851226 | 0.92688 | 0.9368072748 | 0.0963997 | 4.416066399 | 0 |
| 2 | 47478455 | T | A | N | K | MSH2 | ENST00000233146 | 0.25054 | 0.00556 | 0.51058 | 0.34021 | 0.69846 | 0.611880511 | 0.87342 | 0.1732860223 | -0.0807394 | 2.790467158 | 0 |
| 2 | 47478455 | T | G | N | K | MSH2 | ENST00000233146 | 0.25516 | 0.00556 | 0.51058 | 0.40584 | 0.69846 | 0.611880511 | 0.87309 | 0.1081718785 | -0.104302 | 2.790467158 | 0 |
| 2 | 47478456 | A | C | N | H | MSH2 | ENST00000233146 | 0.8115 | 0.03674 | 0.71818 | 0.95393 | 0.97364 | 0.86769962 | 0.9535 | 0.9964665174 | 0.240315 | 9.44561291 | 0 |
| 2 | 47478456 | A | G | N | D | MSH2 | ENST00000233146 | 0.83798 | 0.03527 | 0.76067 | 0.90625 | 0.97773 | 0.8288472766 | 0.9543 | 0.9996378422 | 0.476027 | 9.44561291 | 0 |
| 2 | 47478456 | A | T | N | Y | MSH2 | ENST00000233146 | 0.85756 | 0.03419 | 0.74831 | 0.95131 | 0.99525 | 0.8857254497 | 0.95728 | 0.9991663694 | 0.47708 | 9.44561291 | 0 |
| 2 | 47478457 | A | C | N | T | MSH2 | ENST00000233146 | 0.82311 | 0.03692 | 0.69537 | 0.90343 | 0.96993 | 0.8283615309 | 0.92175 | 0.9967896342 | 0.468606 | 8.258426464 | 0 |
| 2 | 47478457 | A | G | N | S | MSH2 | ENST00000233146 | 0.77188 | 0.04051 | 0.6492 | 0.9255 | 0.9659 | 0.7510342116 | 0.94389 | 0.9984062314 | 0.457529 | 9.44561291 | 0 |
| 2 | 47478457 | A | T | N | I | MSH2 | ENST00000233146 | 0.86382 | 0.03388 | 0.70702 | 0.94215 | 0.98146 | 0.9134992851 | 0.90968 | 0.9998342991 | 0.480936 | 9.44561291 | 0 |
| 2 | 47478458 | T | A | N | K | MSH2 | ENST00000233146 | 0.59697 | 0.03509 | 0.7854 | 0.93053 | 0.92924 | 0.9002107739 | 0.95453 | 0.9960842133 | 0.447546 | 9.44561291 | 0 |
| 2 | 47478458 | T | G | N | K | MSH2 | ENST00000233146 | 0.60449 | 0.03509 | 0.7854 | 0.93955 | 0.92964 | 0.9002107739 | 0.95448 | 0.9960842133 | 0.435904 | 9.44561291 | 0 |
| 2 | 47478459 | C | A | L | I | MSH2 | ENST00000233146 | 0.58909 | 0.03419 | 0.77596 | 0.84375 | 0.89784 | 0.5943542394 | 0.83881 | 0.9101217049 | 0.238479 | 4.541363272 | 0 |
| 2 | 47478459 | C | G | L | V | MSH2 | ENST00000233146 | 0.55326 | 0.03484 | 0.71538 | 0.82835 | 0.88165 | 0.6979844549 | 0.94158 | 0.8714232431 | 0.461397 | 3.994466539 | 0 |
| 2 | 47478460 | T | A | L | Q | MSH2 | ENST00000233146 | 0.59647 | 0.03587 | 0.78911 | 0.74781 | 0.97475 | 0.9134138634 | 0.96635 | 0.981119381 | 0.39477 | 7.326104609 | 0 |
| 2 | 47478460 | T | C | L | P | MSH2 | ENST00000233146 | 0.78344 | 0.02979 | 0.87423 | 0.78137 | 0.98799 | 0.9605927665 | 0.9726 | 0.991884461 | 0.442567 | 8.258312938 | 1 |
| 2 | 47478460 | T | G | L | R | MSH2 | ENST00000233146 | 0.59697 | 0.01812 | 0.84601 | 0.76395 | 0.97438 | 0.9456138195 | 0.97341 | 0.9786900878 | 0.396099 | 6.349430776 | 0 |
| 2 | 47478462 | C | A | H | N | MSH2 | ENST00000233146 | 0.73877 | 0.03582 | 0.91055 | 0.84792 | 0.92884 | 0.8739383255 | 0.91431 | 0.9941912524 | 0.267451 | 4.155680704 | 0 |
| 2 | 47478462 | C | T | H | Y | MSH2 | ENST00000233146 | 0.73086 | 0.03497 | 0.90937 | 0.90408 | 0.98294 | 0.8900119321 | 0.96415 | 0.9932767749 | 0.445026 | 7.031146234 | 0 |
| 2 | 47478463 | A | C | H | P | MSH2 | ENST00000233146 | 0.8402 | 0.03059 | 0.85744 | 0.90185 | 0.98799 | 0.964771339 | 0.97141 | 0.9999494553 | 0.474671 | 9.43894004 | 1 |
| 2 | 47478463 | A | G | H | R | MSH2 | ENST00000233146 | 0.79334 | 0.03419 | 0.91136 | 0.83129 | 0.99138 | 0.9271132059 | 0.98647 | 0.9985810518 | 0.44246 | 5.337695964 | 0 |
| 2 | 47478463 | A | T | H | L | MSH2 | ENST00000233146 | 0.84302 | 0.03398 | 0.88713 | 0.89746 | 0.98294 | 0.9194042634 | 0.98035 | 0.9999595881 | 0.474009 | 9.43894004 | 0 |
| 2 | 47478464 | T | A | H | Q | MSH2 | ENST00000233146 | 0.56152 | 0.03713 | 0.88929 | 0.83159 | 0.94839 | 0.8913589182 | 0.98973 | 0.9987111092 | 0.387906 | 7.50386611 | 0 |
| 2 | 47478464 | T | G | H | Q | MSH2 | ENST00000233146 | 0.56866 | 0.03713 | 0.88929 | 0.83485 | 0.94839 | 0.8913589182 | 0.98965 | 0.9987111092 | 0.387908 | 7.50386611 | 0 |
| 2 | 47478465 | G | A | V | I | MSH2 | ENST00000233146 | 0.83721 | 0.03917 | 0.64585 | 0.87761 | 0.89467 | 0.5956314478 | 0.88848 | 0.9735664725 | 0.479081 | 4.631320478 | 0 |
| 2 | 47478465 | G | C | V | L | MSH2 | ENST00000233146 | 0.86845 | 0.03602 | 0.80921 | 0.90075 | 0.94691 | 0.8000835516 | 0.94582 | 0.9920294285 | 0.476027 | 8.620384097 | 0 |
| 2 | 47478465 | G | T | V | F | MSH2 | ENST00000233146 | 0.89255 | 0.03265 | 0.78716 | 0.89613 | 0.97178 | 0.9254047989 | 0.96542 | 0.9979921579 | 0.481856 | 7.469555411 | 0 |
| 2 | 47478466 | T | C | V | A | MSH2 | ENST00000233146 | 0.93348 | 0.03334 | 0.8237 | 0.90346 | 0.97848 | 0.8178268685 | 0.94275 | 0.9969257712 | 0.456739 | 6.022133645 | 0 |
| 2 | 47478466 | T | G | V | G | MSH2 | ENST00000233146 | 0.94307 | 0.03189 | 0.7033 | 0.9475 | 0.99072 | 0.9482044403 | 0.95121 | 0.9995686412 | 0.466743 | 9.680169969 | 0 |
| 2 | 47478468 | A | C | T | P | MSH2 | ENST00000233146 | 0.91604 | 0.02972 | 0.6151 | 0.90179 | 0.95133 | 0.9175251644 | 0.93974 | 0.9868191239 | 0.409549 | 8.883534653 | 0 |
| 2 | 47478468 | A | G | T | A | MSH2 | ENST00000233146 | 0.69885 | 0.00909 | 0.62044 | 0.76538 | 0.85274 | 0.7608387199 | 0.93868 | 0.9359182119 | 0.214289 | 3.72069236 | 0 |
| 2 | 47478468 | A | T | T | S | MSH2 | ENST00000233146 | 0.52912 | 0.00821 | 0.63901 | 0.72994 | 0.73074 | 0.6385477199 | 0.87481 | 0.8512459993 | 0.100723 | 3.980835428 | 0 |
| 2 | 47478469 | C | A | T | K | MSH2 | ENST00000233146 | 0.62734 | 0.02547 | 0.73398 | 0.78918 | 0.93319 | 0.8771216654 | 0.90605 | 0.9802983403 | 0.574014 | 5.694527106 | 0 |
| 2 | 47478469 | C | G | T | R | MSH2 | ENST00000233146 | 0.6281 | 0.03019 | 0.7167 | 0.85686 | 0.93436 | 0.9030437374 | 0.92907 | 0.9830449094 | 0.476027 | 5.225860506 | 0 |
| 2 | 47478469 | C | T | T | I | MSH2 | ENST00000233146 | 0.6246 | 0.03219 | 0.76739 | 0.92113 | 0.95097 | 0.8331506147 | 0.87509 | 0.9929962477 | 0.481856 | 8.883534653 | 0 |
| 2 | 47478471 | G | A | A | T | MSH2 | ENST00000233146 | 0.89509 | 0.03686 | 0.67247 | 0.82636 | 0.96334 | 0.8317232332 | 0.96315 | 0.9932462325 | 0.426988 | 8.948539409 | 0 |
| 2 | 47478471 | G | C | A | P | MSH2 | ENST00000233146 | 0.90359 | 0.03299 | 0.74024 | 0.80857 | 0.98972 | 0.9653805007 | 0.98349 | 0.9953411818 | 0.47357 | 8.948539409 | 0 |
| 2 | 47478471 | G | T | A | S | MSH2 | ENST00000233146 | 0.86418 | 0.03767 | 0.62165 | 0.81565 | 0.95316 | 0.7927244898 | 0.9426 | 0.9852935672 | 0.342664 | 8.948539409 | 0 |
| 2 | 47478472 | C | A | A | E | MSH2 | ENST00000233146 | 0.66043 | 0.03436 | 0.74512 | 0.82137 | 0.97885 | 0.9557625899 | 0.98106 | 0.9952644374 | 0.454944 | 8.948539409 | 0 |
| 2 | 47478472 | C | G | A | G | MSH2 | ENST00000233146 | 0.67184 | 0.03633 | 0.64488 | 0.79949 | 0.95572 | 0.8020812467 | 0.93068 | 0.9949018553 | 0.374037 | 8.948539409 | 0 |
| 2 | 47478472 | C | T | A | V | MSH2 | ENST00000233146 | 0.6704 | 0.03353 | 0.68968 | 0.74627 | 0.93747 | 0.8383640391 | 0.97467 | 0.9902826548 | 0.390148 | 3.592454977 | 0 |
| 2 | 47478474 | C | A | L | I | MSH2 | ENST00000233146 | 0.31764 | 0.01244 | 0.7104 | 0.77046 | 0.85274 | 0.5001819842 | 0.86802 | 0.5523231908 | 0.0108599 | 4.874245332 | 0 |
| 2 | 47478474 | C | G | L | V | MSH2 | ENST00000233146 | 0.31255 | 0.02005 | 0.65463 | 0.7563 | 0.87394 | 0.6196744346 | 0.91949 | 0.7258220315 | -0.0242361 | 5.107285377 | 0 |
| 2 | 47478474 | C | T | L | F | MSH2 | ENST00000233146 | 0.37459 | 0.03426 | 0.69442 | 0.8494 | 0.85534 | 0.6766119058 | 0.93807 | 0.7868142128 | 0.145488 | 8.61679722 | 0 |
| 2 | 47478475 | T | A | L | H | MSH2 | ENST00000233146 | 0.57301 | 0.02687 | 0.79511 | 0.64522 | 0.90711 | 0.8252581869 | 0.96304 | 0.9644133449 | 0.371255 | 6.306209752 | 0 |
| 2 | 47478475 | T | C | L | P | MSH2 | ENST00000233146 | 0.72973 | 0.02951 | 0.82222 | 0.76583 | 0.95207 | 0.9473138399 | 0.97394 | 0.9889896665 | 0.420703 | 8.61679722 | 0 |
| 2 | 47478475 | T | G | L | R | MSH2 | ENST00000233146 | 0.72451 | 0.0307 | 0.79638 | 0.61542 | 0.9328 | 0.9006448507 | 0.96959 | 0.9810535312 | 0.392986 | 4.635016342 | 0 |
| 2 | 47478477 | A | C | T | P | MSH2 | ENST00000233146 | 0.8356 | 0.03243 | 0.60139 | 0.89331 | 0.91778 | 0.8827594443 | 0.93518 | 0.9886997342 | 0.360213 | 8.231571207 | 0 |
| 2 | 47478477 | A | G | T | A | MSH2 | ENST00000233146 | 0.60295 | 0.01479 | 0.6045 | 0.82116 | 0.88637 | 0.7171438886 | 0.90272 | 0.9442935586 | 0.207681 | 7.897294388 | 0 |
| 2 | 47478477 | A | T | T | S | MSH2 | ENST00000233146 | 0.75193 | 0.03497 | 0.61506 | 0.8383 | 0.86352 | 0.6341864195 | 0.90765 | 0.9454602003 | 0.20988 | 8.231571207 | 0 |
| 2 | 47478478 | C | A | T | N | MSH2 | ENST00000233146 | 0.72083 | 0.03426 | 0.68824 | 0.74103 | 0.81876 | 0.801876857 | 0.78295 | 0.9536153004 | 0.37876 | 6.239259605 | 0 |
| 2 | 47478478 | C | G | T | S | MSH2 | ENST00000233146 | 0.70799 | 0.03497 | 0.61506 | 0.78454 | 0.84321 | 0.6341864195 | 0.83113 | 0.8986323476 | 0.269953 | 8.231571207 | 0 |
| 2 | 47478478 | C | T | T | I | MSH2 | ENST00000233146 | 0.59255 | 0.02614 | 0.73463 | 0.70313 | 0.87198 | 0.7543459801 | 0.88419 | 0.8162218105 | 0.352512 | 7.186995751 | 0 |
| 2 | 47478480 | A | C | T | P | MSH2 | ENST00000233146 | 0.66509 | 0.01915 | 0.38657 | 0.75884 | 0.83172 | 0.7721086729 | 0.93806 | 0.8701225519 | 0.187174 | 7.962587464 | 0 |
| 2 | 47478480 | A | G | T | A | MSH2 | ENST00000233146 | 0.35168 | 0.00592 | 0.38594 | 0.54322 | 0.77718 | 0.5512959742 | 0.98055 | 0.2362889647 | 0.137622 | 6.008181403 | 0 |
| 2 | 47478480 | A | T | T | S | MSH2 | ENST00000233146 | 0.24436 | 0.00596 | 0.38072 | 0.56105 | 0.68908 | 0.4105376589 | 0.92116 | 0.08662633598 | 0.0407235 | 1.257516503 | 0 |
| 2 | 47478481 | C | A | T | N | MSH2 | ENST00000233146 | 0.24964 | 0.01574 | 0.46856 | 0.51509 | 0.66258 | 0.53462162 | 0.85838 | 0.6765239835 | 0.223038 | 4.206592722 | 0 |
| 2 | 47478481 | C | G | T | S | MSH2 | ENST00000233146 | 0.16783 | 0.00596 | 0.38072 | 0.14409 | 0.66449 | 0.4105376589 | 0.8805 | 0.3448708951 | -0.16437 | 1.257516503 | 0 |
| 2 | 47478481 | C | T | T | I | MSH2 | ENST00000233146 | 0.50101 | 0.02056 | 0.53793 | 0.72785 | 0.75417 | 0.6253094455 | 0.88241 | 0.8057974571 | 0.162983 | 4.582827751 | 0 |
| 2 | 47478483 | G | C | E | Q | MSH2 | ENST00000233146 | 0.44132 | 0.00615 | 0.21044 | 0.43686 | 0.74093 | 0.4281941632 | 0.95201 | 0.356581067 | 0.111578 | 2.509973349 | 0 |
| 2 | 47478484 | A | C | E | A | MSH2 | ENST00000233146 | 0.36325 | 0.00586 | 0.22708 | 0.23269 | 0.81933 | 0.48354685 | 0.88498 | 0.6892009237 | 0.140229 | 3.936252412 | 0 |
| 2 | 47478484 | A | G | E | G | MSH2 | ENST00000233146 | 0.3861 | 0.00547 | 0.20036 | 0.10121 | 0.73949 | 0.4403276427 | 0.93214 | 0.8012987971 | 0.133962 | 0.6286061955 | 0 |

| | | | | | | | | | | | | | | | | | | |
|---|---|---|---|---|---|---|---|---|---|---|---|---|---|---|---|---|---|---|
| 2 | 47478484 | A | T | E | V | MSH2 | ENST00000233146 | 0.52999 | 0.00799 | 0.20992 | 0.52439 | 0.82836 | 0.5358702535 | 0.89574 | 0.9096192718 | 0.162527 | 5.787931526 | 0 |
| 2 | 47478485 | A | C | E | D | MSH2 | ENST00000233146 | 0.20912 | 0.00592 | 0.22384 | 0.02267 | 0.70014 | 0.3579358877 | 0.90663 | 0.03834005818 | -0.00066313 | -0.4235386477 | 0 |
| 2 | 47478485 | A | T | E | D | MSH2 | ENST00000233146 | 0.20583 | 0.00592 | 0.22384 | 0.02113 | 0.70014 | 0.3579358877 | 0.91216 | 0.03834005818 | -0.000658257 | -0.4235386477 | 0 |
| 2 | 47478486 | G | A | E | K | MSH2 | ENST00000233146 | 0.51619 | 0.00556 | 0.47705 | 0.50729 | 0.72775 | 0.4665367994 | 0.88432 | 0.05381216809 | -0.244059 | 3.132737964 | 0 |
| 2 | 47478486 | G | C | E | Q | MSH2 | ENST00000233146 | 0.52008 | 0.00836 | 0.34987 | 0.14258 | 0.77527 | 0.481011699 | 0.87557 | 0.3372330631 | 0.107114 | 3.46386081 | 0 |
| 2 | 47478487 | A | C | E | A | MSH2 | ENST00000233146 | 0.40952 | 0.00726 | 0.37179 | 0.70675 | 0.79758 | 0.5324357437 | 0.7991 | 0.415670393 | 0.091074 | 5.733771658 | 0 |
| 2 | 47478487 | A | G | E | G | MSH2 | ENST00000233146 | 0.41392 | 0.00543 | 0.33562 | 0.04757 | 0.73589 | 0.4865686006 | 0.86754 | 0.6152704954 | 0.113132 | 1.098767226 | 0 |
| 2 | 47478487 | A | T | E | V | MSH2 | ENST00000233146 | 0.68643 | 0.01158 | 0.34868 | 0.5639 | 0.84641 | 0.5955168452 | 0.83079 | 0.9261860847 | 0.163125 | 6.3444837 | 0 |
| 2 | 47478488 | G | C | E | D | MSH2 | ENST00000233146 | 0.24853 | 0.00592 | 0.36944 | 0.00529 | 0.73805 | 0.3676826128 | 0.89435 | 0.1243965998 | 0.0294782 | 0.3033837704 | 0 |
| 2 | 47478488 | G | T | E | D | MSH2 | ENST00000233146 | 0.24497 | 0.00592 | 0.36944 | 0.00491 | 0.73805 | 0.3676826128 | 0.8879 | 0.1702376157 | 0.0295242 | 0.3033837704 | 0 |
| 2 | 47478489 | A | C | T | P | MSH2 | ENST00000233146 | 0.74269 | 0.0197 | 0.43127 | 0.76645 | 0.90842 | 0.8618909977 | 0.87924 | 0.9515022039 | 0.292435 | 8.73844469 | 0 |
| 2 | 47478489 | A | G | T | A | MSH2 | ENST00000233146 | 0.48285 | 0.00592 | 0.43875 | 0.5349 | 0.70179 | 0.7034653266 | 0.92531 | 0.6283882856 | 0.139244 | 7.375105683 | 0 |
| 2 | 47478489 | A | T | T | S | MSH2 | ENST00000233146 | 0.47306 | 0.00654 | 0.45062 | 0.42902 | 0.78597 | 0.5641173718 | 0.88441 | 0.472996825 | -0.0136843 | 4.876385913 | 0 |
| 2 | 47478490 | C | A | T | N | MSH2 | ENST00000233146 | 0.25788 | 0.01523 | 0.53271 | 0.64308 | 0.67293 | 0.7173579455 | 0.81004 | 0.7996080518 | 0.31553 | 4.680272746 | 0 |
| 2 | 47478490 | C | G | T | S | MSH2 | ENST00000233146 | 0.24471 | 0.00654 | 0.45062 | 0.53992 | 0.74867 | 0.5641173718 | 0.848 | 0.6339709759 | 0.0505889 | 4.876385913 | 0 |
| 2 | 47478490 | C | T | T | I | MSH2 | ENST00000233146 | 0.41819 | 0.01645 | 0.59311 | 0.80451 | 0.89603 | 0.750413675 | 0.82876 | 0.9766018987 | 0.413555 | 8.73844469 | 0 |
| 2 | 47478492 | T | A | L | I | MSH2 | ENST00000233146 | 0.42633 | 0.02212 | 0.66047 | 0.64228 | 0.77782 | 0.587335391 | 0.93847 | 0.645250082 | 0.150824 | 3.447313709 | 0 |
| 2 | 47478492 | T | G | L | V | MSH2 | ENST00000233146 | 0.40261 | 0.0178 | 0.61032 | 0.6388 | 0.81472 | 0.7177129903 | 0.9613 | 0.7284029722 | 0.161204 | 3.714836268 | 0 |
| 2 | 47478493 | T | C | L | S | MSH2 | ENST00000233146 | 0.85931 | 0.03118 | 0.68345 | 0.97101 | 0.96153 | 0.9601570234 | 0.98191 | 0.9921410084 | 0.4516 | 8.609008174 | 0 |
| 2 | 47478494 | A | C | L | F | MSH2 | ENST00000233146 | 0.43683 | 0.02056 | 0.65482 | 0.4558 | 0.84906 | 0.8400066481 | 0.99296 | 0.9548441768 | 0.339234 | 6.329163439 | 0 |
| 2 | 47478494 | A | T | L | F | MSH2 | ENST00000233146 | 0.43172 | 0.02056 | 0.65482 | 0.45774 | 0.84906 | 0.8400066481 | 0.99256 | 0.9548441768 | 0.339255 | 6.329163439 | 0 |
| 2 | 47478495 | A | C | T | P | MSH2 | ENST00000233146 | 0.8249 | 0.00898 | 0.48894 | 0.87619 | 0.99039 | 0.9045107244 | 0.95909 | 0.9966359735 | 0.463328 | 9.198012565 | 0 |
| 2 | 47478495 | A | G | T | A | MSH2 | ENST00000233146 | 0.81536 | 0.02614 | 0.50328 | 0.82552 | 0.96444 | 0.7791885407 | 0.92451 | 0.9948742114 | 0.339326 | 6.499046217 | 0 |
| 2 | 47478495 | A | T | T | S | MSH2 | ENST00000233146 | 0.75392 | 0.03413 | 0.51503 | 0.60718 | 0.9248 | 0.6724687524 | 0.8984 | 0.9805448055 | 0.256741 | 9.198012565 | 0 |
| 2 | 47478496 | C | A | T | N | MSH2 | ENST00000233146 | 0.50628 | 0.0337 | 0.60262 | 0.84691 | 0.92924 | 0.8483241464 | 0.85602 | 0.9922713041 | 0.152326 | 9.198012565 | 0 |
| 2 | 47478496 | C | G | T | S | MSH2 | ENST00000233146 | 0.48278 | 0.03413 | 0.51503 | 0.72196 | 0.89558 | 0.6724687524 | 0.86777 | 0.9677889943 | 0.227939 | 9.198012565 | 0 |
| 2 | 47478496 | C | T | T | I | MSH2 | ENST00000233146 | 0.54216 | 0.0114 | 0.66635 | 0.7963 | 0.89513 | 0.8178768711 | 0.89999 | 0.9915450811 | 0.254574 | 3.974566371 | 0 |
| 2 | 47478498 | A | C | M | L | MSH2 | ENST00000233146 | 0.37453 | 0.00596 | 0.71336 | 0.61237 | 0.8233 | 0.7943275484 | 0.94772 | 0.7347988486 | 0.303359 | 1.822802129 | 0 |
| 2 | 47478498 | A | G | M | V | MSH2 | ENST00000233146 | 0.58945 | 0.00615 | 0.64311 | 0.83421 | 0.93977 | 0.8263466357 | 0.98021 | 0.7347769141 | 0.38764 | 6.668765309 | 0 |
| 2 | 47478498 | A | T | M | L | MSH2 | ENST00000233146 | 0.36974 | 0.00596 | 0.71336 | 0.58911 | 0.8233 | 0.7943275484 | 0.94776 | 0.7347988486 | 0.303359 | 1.822802129 | 0 |
| 2 | 47478499 | T | C | M | T | MSH2 | ENST00000233146 | 0.6412 | 0.0134 | 0.82138 | 0.88464 | 0.9659 | 0.9071340651 | 0.97275 | 0.9860007167 | 0.436235 | 6.668765309 | 0 |
| 2 | 47478499 | T | G | M | R | MSH2 | ENST00000233146 | 0.8624 | 0.021 | 0.77289 | 0.90704 | 0.98034 | 0.9736945011 | 0.97137 | 0.9955285192 | 0.443761 | 6.668765309 | 0 |
| 2 | 47478500 | G | A | M | I | MSH2 | ENST00000233146 | 0.74587 | 0.00909 | 0.71119 | 0.72401 | 0.91442 | 0.8429092792 | 0.97122 | 0.7482044101 | 0.405854 | 6.668765309 | 0 |
| 2 | 47478500 | G | C | M | I | MSH2 | ENST00000233146 | 0.7426 | 0.00909 | 0.71119 | 0.72203 | 0.91399 | 0.8429092792 | 0.97114 | 0.8757263283 | 0.405854 | 6.668765309 | 0 |
| 2 | 47478500 | G | T | M | I | MSH2 | ENST00000233146 | 0.73613 | 0.00909 | 0.71119 | 0.72265 | 0.91442 | 0.8429092792 | 0.97115 | 0.881901646 | 0.405854 | 6.668765309 | 0 |
| 2 | 47478501 | C | A | L | I | MSH2 | ENST00000233146 | 0.73365 | 0.03388 | 0.83274 | 0.82595 | 0.93241 | 0.6455103963 | 0.90444 | 0.9595555067 | 0.283934 | 7.007800313 | 0 |
| 2 | 47478501 | C | G | L | V | MSH2 | ENST00000233146 | 0.70927 | 0.03484 | 0.7891 | 0.8211 | 0.94053 | 0.7735094349 | 0.97889 | 0.9496572018 | 0.406188 | 9.321626384 | 0 |
| 2 | 47478501 | C | T | L | F | MSH2 | ENST00000233146 | 0.74531 | 0.02037 | 0.82027 | 0.85993 | 0.94949 | 0.8065249668 | 0.95706 | 0.994083941 | 0.379067 | 9.321626384 | 0 |
| 2 | 47478502 | T | A | L | H | MSH2 | ENST00000233146 | 0.8456 | 0.03313 | 0.88205 | 0.94214 | 0.99072 | 0.9156802314 | 0.96801 | 0.9940044284 | 0.467472 | 9.321626384 | 0 |
| 2 | 47478502 | T | C | L | P | MSH2 | ENST00000233146 | 0.8665 | 0.01753 | 0.9042 | 0.94743 | 0.99525 | 0.9702690114 | 0.9812 | 0.9989259839 | 0.466707 | 9.321626384 | 0 |
| 2 | 47478502 | T | G | L | R | MSH2 | ENST00000233146 | 0.86382 | 0.03118 | 0.88707 | 0.93782 | 0.99471 | 0.9623974054 | 0.98077 | 0.9932768941 | 0.465331 | 8.119305328 | 0 |
| 2 | 47478504 | T | A | Y | N | MSH2 | ENST00000233146 | 0.92979 | 0.0296 | 0.83059 | 0.93883 | 0.98869 | 0.9542753044 | 0.95259 | 0.9997739196 | 0.467207 | 8.968763661 | 0 |
| 2 | 47478504 | T | C | Y | H | MSH2 | ENST00000233146 | 0.92719 | 0.03304 | 0.83982 | 0.85118 | 0.97885 | 0.930121929 | 0.94621 | 0.9939302206 | 0.39071 | 3.786874879 | 0 |
| 2 | 47478504 | T | G | Y | D | MSH2 | ENST00000233146 | 0.9325 | 0.02844 | 0.82275 | 0.94202 | 0.98834 | 0.9662764442 | 0.97851 | 0.9999085665 | 0.467207 | 8.968763661 | 1 |
| 2 | 47478505 | A | C | Y | S | MSH2 | ENST00000233146 | 0.92754 | 0.03124 | 0.76679 | 0.9562 | 0.96737 | 0.9520393979 | 0.97198 | 0.9997615218 | 0.472084 | 8.968763661 | 0 |
| 2 | 47478505 | A | G | Y | C | MSH2 | ENST00000233146 | 0.9395 | 0.03353 | 0.83266 | 0.95087 | 0.96334 | 0.9439177238 | 0.98033 | 0.9996803999 | 0.475995 | 8.968763661 | 0 |
| 2 | 47478505 | A | T | Y | F | MSH2 | ENST00000233146 | 0.85152 | 0.03686 | 0.74154 | 0.84741 | 0.92316 | 0.8000428169 | 0.93641 | 0.9863103628 | 0.263174 | 8.968763661 | 0 |
| 2 | 47478507 | C | A | Q | K | MSH2 | ENST00000233146 | 0.39835 | 0.00532 | 0.51809 | 0.46369 | 0.69168 | 0.4357826504 | 0.93504 | 0.5600671768 | 0.127629 | -0.7761027043 | 0 |
| 2 | 47478507 | C | G | Q | E | MSH2 | ENST00000233146 | 0.49673 | 0.00808 | 0.47498 | 0.59162 | 0.76816 | 0.472464073 | 0.89506 | 0.4953184426 | 0.150118 | 3.019175274 | 0 |
| 2 | 47478508 | A | C | Q | P | MSH2 | ENST00000233146 | 0.74946 | 0.01871 | 0.48972 | 0.83151 | 0.91143 | 0.8741858048 | 0.95954 | 0.9552569389 | 0.309181 | 5.468478027 | 0 |
| 2 | 47478508 | A | G | Q | R | MSH2 | ENST00000233146 | 0.31821 | 0.00562 | 0.49803 | 0.24947 | 0.70671 | 0.4573956556 | 0.93263 | 0.173733758 | -0.133278 | -1.983325926 | 0 |
| 2 | 47478508 | A | T | Q | L | MSH2 | ENST00000233146 | 0.73813 | 0.0114 | 0.45343 | 0.82097 | 0.88259 | 0.6018804953 | 0.96061 | 0.9307518005 | 0.228074 | 5.468478027 | 0 |
| 2 | 47478509 | G | C | Q | H | MSH2 | ENST00000233146 | 0.54176 | 0.0063 | 0.51177 | 0.60062 | 0.84213 | 0.5280145393 | 0.96609 | 0.7533830404 | 0.161563 | 3.242923309 | 0 |
| 2 | 47478509 | G | T | Q | H | MSH2 | ENST00000233146 | 0.53623 | 0.0063 | 0.51177 | 0.60164 | 0.84213 | 0.5280145393 | 0.96618 | 0.7533830404 | 0.161544 | 3.242923309 | 0 |
| 2 | 47478510 | G | A | V | M | MSH2 | ENST00000233146 | 0.89125 | 0.03653 | 0.77109 | 0.80113 | 0.98034 | 0.7886781055 | 0.96888 | 0.9898673811 | 0.44888 | 5.612520457 | 1 |
| 2 | 47478510 | G | C | V | L | MSH2 | ENST00000233146 | 0.83012 | 0.03001 | 0.7372 | 0.7399 | 0.93631 | 0.7333879529 | 0.98643 | 0.9554970647 | 0.397064 | 5.597338696 | 0 |
| 2 | 47478510 | G | T | V | L | MSH2 | ENST00000233146 | 0.82381 | 0.03001 | 0.7372 | 0.74287 | 0.93631 | 0.7333879529 | 0.98645 | 0.9541325559 | 0.397064 | 5.597338696 | 0 |
| 2 | 47478511 | T | C | V | A | MSH2 | ENST00000233146 | 0.61015 | 0.01676 | 0.75671 | 0.90211 | 0.9781 | 0.7913023 | 0.98422 | 0.9882375598 | 0.459174 | 9.428850282 | 0 |
| 2 | 47478511 | T | G | V | G | MSH2 | ENST00000233146 | 0.8199 | 0.03189 | 0.62038 | 0.9444 | 0.98477 | 0.9432931798 | 0.98029 | 0.998487711 | 0.474073 | 9.428850282 | 1 |
| 2 | 47478513 | A | C | K | Q | MSH2 | ENST00000233146 | 0.82942 | 0.01391 | 0.7415 | 0.62667 | 0.89873 | 0.6282827771 | 0.90775 | 0.8282747865 | 0.200765 | 5.28238538 | 0 |
| 2 | 47478513 | A | G | K | E | MSH2 | ENST00000233146 | 0.5819 | 0.01258 | 0.78466 | 0.61587 | 0.88448 | 0.7228120463 | 0.95116 | 0.8716990948 | 0.222812 | 4.533641653 | 0 |
| 2 | 47478514 | A | C | K | T | MSH2 | ENST00000233146 | 0.82506 | 0.02914 | 0.79009 | 0.74572 | 0.9014 | 0.6628276787 | 0.89142 | 0.9564566612 | 0.210065 | 6.010959956 | 0 |
| 2 | 47478514 | A | G | K | R | MSH2 | ENST00000233146 | 0.49576 | 0.00741 | 0.70617 | 0.53161 | 0.79334 | 0.5607916533 | 0.97282 | 0.5355378012 | 0.158527 | 3.289277955 | 0 |
| 2 | 47478514 | A | T | K | M | MSH2 | ENST00000233146 | 0.84801 | 0.03144 | 0.70872 | 0.752 | 0.90972 | 0.743563026 | 0.8532 | 0.966042161 | 0.211038 | 4.425322464 | 0 |
| 2 | 47478515 | G | C | K | N | MSH2 | ENST00000233146 | 0.45364 | 0.00849 | 0.81244 | 0.71969 | 0.86047 | 0.6935819791 | 0.84601 | 0.8871047497 | 0.0708593 | 6.372838231 | 0 |
| 2 | 47478515 | G | T | K | N | MSH2 | ENST00000233146 | 0.44863 | 0.00849 | 0.81244 | 0.71685 | 0.86098 | 0.6935819791 | 0.84613 | 0.8871047497 | 0.0708885 | 6.372838231 | 0 |
| 2 | 47478516 | A | C | K | Q | MSH2 | ENST00000233146 | 0.52035 | 0.00596 | 0.49086 | 0.63832 | 0.79516 | 0.4763719758 | 0.8908 | 0.3822744964 | 0.0986559 | 2.981064044 | 0 |
| 2 | 47478516 | A | G | K | E | MSH2 | ENST00000233146 | 0.43075 | 0.00517 | 0.55321 | 0.4546 | 0.74587 | 0.5133900313 | 0.90163 | 0.2698091774 | 0.058968 | 2.440161797 | 1 |
| 2 | 47478517 | A | C | K | T | MSH2 | ENST00000233146 | 0.60941 | 0.00543 | 0.56227 | 0.64354 | 0.76157 | 0.5342299305 | 0.89718 | 0.506764104 | 0.0928346 | 4.501180014 | 0 |
| 2 | 47478517 | A | G | K | R | MSH2 | ENST00000233146 | 0.61723 | 0.00615 | 0.45784 | 0.57497 | 0.79758 | 0.3856090431 | 0.86713 | 0.4909524104 | 0.0999667 | 3.210328842 | 0 |
| 2 | 47478517 | A | T | K | I | MSH2 | ENST00000233146 | 0.66968 | 0.00754 | 0.5264 | 0.78515 | 0.83505 | 0.5907164416 | 0.90366 | 0.9569591284 | 0.189945 | 6.286839015 | 0 |
| 2 | 47478519 | G | A | G | S | MSH2 | ENST00000233146 | 0.9061 | 0.03728 | 0.77504 | 0.87317 | 0.97848 | 0.9089126906 | 0.96866 | 0.9996495247 | 0.482631 | 9.375832193 | 0 |
| 2 | 47478519 | G | C | G | R | MSH2 | ENST00000233146 | 0.92386 | 0.03452 | 0.88038 | 0.86632 | 0.98728 | 0.9278650355 | 0.96976 | 0.9998509884 | 0.485377 | 9.375832193 | 0 |
| 2 | 47478519 | G | T | G | C | MSH2 | ENST00000233146 | 0.93579 | 0.03626 | 0.83195 | 0.9012 | 0.97885 | 0.9183613279 | 0.97688 | 0.99985075 | 0.488186 | 9.375832193 | 0 |
| 2 | 47480696 | G | A | G | D | MSH2 | ENST00000233146 | 0.90339 | 0.03327 | 0.88691 | 0.88958 | 0.98404 | 0.932058343 | 0.96567 | 0.9997068048 | 0.482467 | 8.278464459 | 0 |
| 2 | 47480696 | G | C | G | A | MSH2 | ENST00000233146 | 0.88157 | 0.03608 | 0.78396 | 0.91882 | 0.97141 | 0.9190120781 | 0.95975 | 0.9997100234 | 0.483677 | 7.814239694 | 0 |
| 2 | 47480696 | G | T | G | V | MSH2 | ENST00000233146 | 0.90442 | 0.03379 | 0.79744 | 0.92958 | 0.97587 | 0.9371584181 | 0.96616 | 0.9998764992 | 0.487297 | 9.375832193 | 0 |
| 2 | 47480698 | G | A | V | I | MSH2 | ENST00000233146 | 0.53442 | 0.00849 | 0.31721 | 0.67063 | 0.8199 | 0.4071436773 | 0.82995 | 0.6306868792 | 0.137633 | 6.643377247 | 0 |
| 2 | 47480698 | G | C | V | L | MSH2 | ENST00000233146 | 0.74173 | 0.0114 | 0.50848 | 0.78472 | 0.90711 | 0.6651654908 | 0.83906 | 0.9017099738 | 0.223204 | 7.575494156 | 0 |
| 2 | 47480698 | G | T | V | F | MSH2 | ENST00000233146 | 0.78233 | 0.02658 | 0.48502 | 0.88557 | 0.9211 | 0.8325814578 | 0.86895 | 0.9853236675 | 0.309131 | 6.139219255 | 0 |
| 2 | 47480699 | T | A | V | D | MSH2 | ENST00000233146 | 0.83298 | 0.02574 | 0.54794 | 0.93375 | 0.97327 | 0.8715698866 | 0.93598 | 0.9959611297 | 0.386925 | 7.575494156 | 0 |
| 2 | 47480699 | T | C | V | A | MSH2 | ENST00000233146 | 0.59784 | 0.00747 | 0.5395 | 0.80025 | 0.87636 | 0.6521013235 | 0.86828 | 0.8825586023 | 0.173016 | 2.612431319 | 0 |
| 2 | 47480699 | T | G | V | G | MSH2 | ENST00000233146 | 0.8181 | 0.02243 | 0.3892 | 0.90286 | 0.93397 | 0.8220689944 | 0.93955 | 0.9925563931 | 0.354117 | 3.072835217 | 0 |
| 2 | 47480701 | T | A | C | S | MSH2 | ENST00000233146 | 0.7477 | 0.02288 | 0.81311 | 0.75484 | 0.94949 | 0.8122065651 | 0.90902 | 0.986418426 | 0.36695 | 4.784115999 | 0 |
| 2 | 47480701 | T | C | C | R | MSH2 | ENST00000233146 | 0.9186 | 0.01841 | 0.89969 | 0.90291 | 0.98109 | 0.9075412058 | 0.96428 | 0.9988359809 | 0.446534 | 8.698328125 | 0 |
| 2 | 47480701 | T | G | C | G | MSH2 | ENST00000233146 | 0.91805 | 0.03265 | 0.78373 | 0.90067 | 0.97587 | 0.8475150155 | 0.95496 | 0.9983721375 | 0.403805 | 7.329906729 | 0 |
| 2 | 47480702 | G | A | C | Y | MSH2 | ENST00000233146 | 0.92712 | 0.029 | 0.90855 | 0.9076 | 0.98404 | 0.8698497192 | 0.93837 | 0.9992156029 | 0.451895 | 8.698328125 | 0 |
| 2 | 47480702 | G | C | C | S | MSH2 | ENST00000233146 | 0.7954 | 0.02288 | 0.81311 | 0.77175 | 0.94802 | 0.8122065651 | 0.90068 | 0.9164348841 | 0.258997 | 4.784115999 | 0 |
| 2 | 47480702 | G | T | C | F | MSH2 | ENST00000233146 | 0.93565 | 0.03013 | 0.82446 | 0.89904 | 0.98657 | 0.8844349936 | 0.95575 | 0.9994902611 | 0.46053 | 8.698328125 | 0 |
| 2 | 47480703 | T | G | C | W | MSH2 | ENST00000233146 | 0.70567 | 0.03257 | 0.86884 | 0.97129 | 0.94053 | 0.9010188858 | 0.97722 | 0.9994276166 | 0.413805 | 8.698328125 | 0 |
| 2 | 47480704 | G | C | D | H | MSH2 | ENST00000233146 | 0.93283 | 0.03465 | 0.7966 | 0.90139 | 0.98621 | 0.8071177272 | 0.96887 | 0.9936971497 | 0.456432 | 9.411281505 | 0 |
| 2 | 47480704 | G | T | D | Y | MSH2 | ENST00000233146 | 0.93579 | 0.03409 | 0.7795 | 0.92892 | 0.98146 | 0.8339366515 | 0.97462 | 0.9977461696 | 0.471357 | 9.411281505 | 0 |
| 2 | 47480705 | A | C | D | A | MSH2 | ENST00000233146 | 0.92159 | 0.03394 | 0.71389 | 0.9238 | 0.9917 | 0.8059482358 | 0.95081 | 0.9987362027 | 0.441434 | 8.441608502 | 0 |
| 2 | 47480705 | A | G | D | G | MSH2 | ENST00000233146 | 0.92482 | 0.03313 | 0.72322 | 0.92243 | 0.9917 | 0.7572893735 | 0.96363 | 0.9975941777 | 0.446934 | 8.344564106 | 0 |
| 2 | 47480705 | A | T | D | V | MSH2 | ENST00000233146 | 0.91733 | 0.03189 | 0.7134 | 0.92186 | 0.98257 | 0.8351394688 | 0.9696 | 0.9989637136 | 0.463629 | 9.411281505 | 0 |
| 2 | 47480706 | T | A | D | E | MSH2 | ENST00000233146 | 0.66636 | 0.03821 | 0.75029 | 0.85823 | 0.90185 | 0.6189185838 | 0.973 | 0.9956698418 | 0.349424 | 8.094635134 | 0 |
| 2 | 47480706 | T | G | D | E | MSH2 | ENST00000233146 | 0.67445 | 0.03821 | 0.75029 | 0.86387 | 0.90318 | 0.6189185838 | 0.97297 | 0.9956698418 | 0.349427 | 8.094635134 | 0 |
| 2 | 47480707 | C | A | Q | K | MSH2 | ENST00000233146 | 0.61963 | 0.02255 | 0.63298 | 0.78626 | 0.93201 | 0.6860032968 | 0.96148 | 0.9296303988 | 0.361851 | 3.974472405 | 0 |
| 2 | 47480707 | C | G | Q | E | MSH2 | ENST00000233146 | 0.7532 | 0.03166 | 0.56999 | 0.86095 | 0.94543 | 0.7013660288 | 0.94646 | 0.8713657237 | 0.325289 | 5.344964427 | 0 |
| 2 | 47480708 | A | C | Q | P | MSH2 | ENST00000233146 | 0.90004 | 0.03353 | 0.58518 | 0.88527 | 0.98869 | 0.8899170496 | 0.97176 | 0.9964111108 | 0.44627 | 9.608345519 | 1 |
| 2 | 47480708 | A | G | Q | R | MSH2 | ENST00000233146 | 0.71378 | 0.01597 | 0.62659 | 0.6811 | 0.94015 | 0.7031077754 | 0.98203 | 0.9260953665 | 0.38406 | 4.365965364 | 0 |

| | | | | | | | | | | | | | | | | | | |
|---|---|---|---|---|---|---|---|---|---|---|---|---|---|---|---|---|---|---|
| 2 | 47480708 | A | T | Q | L | MSH2 | ENST00000233146 | 0.91578 | 0.03595 | 0.56365 | 0.91306 | 0.98109 | 0.7087175027 | 0.94619 | 0.9968058033 | 0.426773 | 9.608345519 | 0 |
| 2 | 47480709 | A | C | Q | H | MSH2 | ENST00000233146 | 0.52347 | 0.03767 | 0.62516 | 0.68584 | 0.86098 | 0.7634496805 | 0.96262 | 0.9967761636 | 0.349886 | 5.728236616 | 0 |
| 2 | 47480709 | A | T | Q | H | MSH2 | ENST00000233146 | 0.5179 | 0.03767 | 0.62516 | 0.69394 | 0.86098 | 0.7634496805 | 0.96298 | 0.9967761636 | 0.349882 | 5.728236616 | 0 |
| 2 | 47480710 | A | C | S | R | MSH2 | ENST00000233146 | 0.91324 | 0.03547 | 0.81567 | 0.91823 | 0.99356 | 0.9419946539 | 0.97779 | 0.9996204376 | 0.4894 | 9.986808399 | 0 |
| 2 | 47480710 | A | G | S | G | MSH2 | ENST00000233146 | 0.89645 | 0.03848 | 0.70586 | 0.88944 | 0.99265 | 0.8660112223 | 0.97043 | 0.9995931983 | 0.486686 | 9.986808399 | 0 |
| 2 | 47480710 | A | T | S | C | MSH2 | ENST00000233146 | 0.91884 | 0.03761 | 0.75578 | 0.93362 | 0.98513 | 0.8703493461 | 0.95528 | 0.9996417761 | 0.49071 | 9.986808399 | 0 |
| 2 | 47480711 | G | A | S | N | MSH2 | ENST00000233146 | 0.9059 | 0.03826 | 0.80257 | 0.92948 | 0.95133 | 0.8917027121 | 0.93431 | 0.9991944432 | 0.24691 | 9.986808399 | 1 |
| 2 | 47480711 | G | C | S | T | MSH2 | ENST00000233146 | 0.90186 | 0.03857 | 0.75436 | 0.94413 | 0.97997 | 0.8538891591 | 0.97061 | 0.9991944432 | 0.487514 | 9.986808399 | 0 |
| 2 | 47480711 | G | T | S | I | MSH2 | ENST00000233146 | 0.93701 | 0.03465 | 0.78476 | 0.96202 | 0.99552 | 0.9290226011 | 0.97711 | 0.9996127486 | 0.491604 | 9.986808399 | 0 |
| 2 | 47480712 | T | A | S | R | MSH2 | ENST00000233146 | 0.69893 | 0.03547 | 0.81567 | 0.95949 | 0.97141 | 0.9419946539 | 0.98742 | 0.9988910556 | 0.433865 | 9.986808399 | 0 |
| 2 | 47480712 | T | G | S | R | MSH2 | ENST00000233146 | 0.7072 | 0.03547 | 0.81567 | 0.9572 | 0.97141 | 0.9419946539 | 0.98733 | 0.9988910556 | 0.433868 | 9.986808399 | 0 |
| 2 | 47480713 | T | A | F | I | MSH2 | ENST00000233146 | 0.92656 | 0.0315 | 0.84638 | 0.88765 | 0.98728 | 0.8422505005 | 0.94992 | 0.9995163679 | 0.458917 | 9.103548497 | 0 |
| 2 | 47480713 | T | C | F | L | MSH2 | ENST00000233146 | 0.93161 | 0.02506 | 0.87313 | 0.85761 | 0.98257 | 0.7805033027 | 0.943 | 0.9884866476 | 0.420841 | 9.103548497 | 0 |
| 2 | 47480713 | T | G | F | V | MSH2 | ENST00000233146 | 0.92937 | 0.032 | 0.8266 | 0.91925 | 0.98367 | 0.8073629755 | 0.95196 | 0.9983945489 | 0.459894 | 9.103548497 | 0 |
| 2 | 47480714 | T | A | F | Y | MSH2 | ENST00000233146 | 0.91797 | 0.03491 | 0.85233 | 0.73321 | 0.94129 | 0.6977877081 | 0.91026 | 0.9490197897 | 0.285765 | 3.635576001 | 0 |
| 2 | 47480714 | T | C | F | S | MSH2 | ENST00000233146 | 0.94837 | 0.02828 | 0.84341 | 0.92665 | 0.98693 | 0.8564118749 | 0.9635 | 0.9979255199 | 0.466516 | 9.103548497 | 0 |
| 2 | 47480714 | T | G | F | C | MSH2 | ENST00000233146 | 0.94828 | 0.03166 | 0.8718 | 0.93594 | 0.98869 | 0.8348589528 | 0.96348 | 0.9981588721 | 0.468812 | 9.103548497 | 0 |
| 2 | 47480715 | T | A | F | L | MSH2 | ENST00000233146 | 0.73781 | 0.02506 | 0.87313 | 0.85628 | 0.94431 | 0.7805033027 | 0.96098 | 0.987654388 | 0.375385 | 9.103548497 | 0 |
| 2 | 47480715 | T | G | F | L | MSH2 | ENST00000233146 | 0.74603 | 0.02506 | 0.87313 | 0.85279 | 0.94431 | 0.7805033027 | 0.96081 | 0.987654388 | 0.375386 | 9.103548497 | 0 |
| 2 | 47480716 | G | A | G | R | MSH2 | ENST00000233146 | 0.9285 | 0.03447 | 0.88492 | 0.96649 | 0.99525 | 0.9490379734 | 0.98087 | 0.9997717738 | 0.55782 | 9.986799191 | 0 |
| 2 | 47480716 | G | C | G | R | MSH2 | ENST00000233146 | 0.92709 | 0.03447 | 0.88492 | 0.96416 | 0.99525 | 0.9490379734 | 0.98046 | 0.9997717738 | 0.557821 | 9.986799191 | 0 |
| 2 | 47480717 | G | C | G | A | MSH2 | ENST00000233146 | 0.92116 | 0.03547 | 0.80475 | 0.93287 | 0.97698 | 0.8917550834 | 0.96459 | 0.9958050251 | 0.523956 | 9.986799191 | 0 |
| 2 | 47480717 | G | T | G | V | MSH2 | ENST00000233146 | 0.93451 | 0.03362 | 0.81464 | 0.96785 | 0.99765 | 0.9219411358 | 0.97378 | 0.9997717738 | 0.557821 | 9.986799191 | 0 |
| 2 | 47480719 | A | C | I | L | MSH2 | ENST00000233146 | 0.67979 | 0.01841 | 0.83896 | 0.71723 | 0.91778 | 0.6713127704 | 0.89718 | 0.8008119153 | 0.293886 | 8.469657525 | 0 |
| 2 | 47480719 | A | G | I | V | MSH2 | ENST00000233146 | 0.77632 | 0.03269 | 0.76254 | 0.80588 | 0.90711 | 0.491466751 | 0.86934 | 0.8699914174 | 0.252889 | 4.88249525 | 0 |
| 2 | 47480719 | A | T | I | F | MSH2 | ENST00000233146 | 0.86169 | 0.02979 | 0.84881 | 0.91011 | 0.98477 | 0.8433723853 | 0.95854 | 0.995467576 | 0.457069 | 9.612682357 | 0 |
| 2 | 47480720 | T | A | I | N | MSH2 | ENST00000233146 | 0.93086 | 0.02844 | 0.91023 | 0.9478 | 0.97624 | 0.9136243261 | 0.92255 | 0.9996409416 | 0.440232 | 9.612682357 | 0 |
| 2 | 47480720 | T | C | I | T | MSH2 | ENST00000233146 | 0.90122 | 0.03304 | 0.92079 | 0.92887 | 0.98034 | 0.8026894582 | 0.96629 | 0.9978542924 | 0.454615 | 9.612682357 | 0 |
| 2 | 47480720 | T | G | I | S | MSH2 | ENST00000233146 | 0.93429 | 0.03112 | 0.85428 | 0.95158 | 0.99105 | 0.9067076864 | 0.96654 | 0.9983028173 | 0.475214 | 9.612682357 | 0 |
| 2 | 47480721 | T | G | I | M | MSH2 | ENST00000233146 | 0.79879 | 0.02972 | 0.80763 | 0.91506 | 0.91569 | 0.7314954272 | 0.91569 | 0.9824730158 | 0.360994 | 9.612682357 | 0 |
| 2 | 47480722 | C | A | H | N | MSH2 | ENST00000233146 | 0.74746 | 0.03509 | 0.89399 | 0.80564 | 0.90052 | 0.7218254068 | 0.84985 | 0.9766180306 | 0.207478 | 6.502600275 | 0 |
| 2 | 47480722 | C | G | H | D | MSH2 | ENST00000233146 | 0.77916 | 0.0318 | 0.89014 | 0.8544 | 0.96334 | 0.8560202818 | 0.93581 | 0.989451468 | 0.406898 | 6.804401351 | 0 |
| 2 | 47480722 | C | T | H | Y | MSH2 | ENST00000233146 | 0.74826 | 0.03461 | 0.8957 | 0.88641 | 0.96226 | 0.815418893 | 0.93253 | 0.9886344671 | 0.414239 | 5.413133665 | 0 |
| 2 | 47480723 | A | C | H | P | MSH2 | ENST00000233146 | 0.85137 | 0.03023 | 0.83636 | 0.90535 | 0.96993 | 0.9123463053 | 0.93167 | 0.9998600483 | 0.464983 | 9.309475638 | 0 |
| 2 | 47480723 | A | G | H | R | MSH2 | ENST00000233146 | 0.80471 | 0.03322 | 0.89542 | 0.86578 | 0.9781 | 0.841927611 | 0.95736 | 0.9947692752 | 0.435976 | 9.309475638 | 0 |
| 2 | 47480723 | A | T | H | L | MSH2 | ENST00000233146 | 0.8536 | 0.03299 | 0.87233 | 0.90376 | 0.96993 | 0.818103983 | 0.9609 | 0.9996982813 | 0.452516 | 9.309475638 | 0 |
| 2 | 47480724 | T | A | H | Q | MSH2 | ENST00000233146 | 0.31074 | 0.02708 | 0.8745 | 0.7286 | 0.81588 | 0.6787005999 | 0.94196 | 0.9878296852 | 0.293054 | 4.333643106 | 0 |
| 2 | 47480724 | T | G | H | Q | MSH2 | ENST00000233146 | 0.31603 | 0.02708 | 0.8745 | 0.71866 | 0.81588 | 0.6787005999 | 0.94204 | 0.9287405014 | 0.170636 | 4.333643106 | 0 |
| 2 | 47480725 | G | A | V | I | MSH2 | ENST00000233146 | 0.82615 | 0.03713 | 0.52699 | 0.84077 | 0.89963 | 0.6574598396 | 0.89279 | 0.9511841536 | 0.275553 | 9.778991533 | 0 |
| 2 | 47480725 | G | C | V | L | MSH2 | ENST00000233146 | 0.86226 | 0.02243 | 0.70928 | 0.8764 | 0.95717 | 0.8164256795 | 0.93968 | 0.9906348547 | 0.37539 | 9.778991533 | 0 |
| 2 | 47480725 | G | T | V | F | MSH2 | ENST00000233146 | 0.88554 | 0.0325 | 0.67446 | 0.9558 | 0.98764 | 0.9258955848 | 0.96542 | 0.9994064569 | 0.494196 | 9.778991533 | 0 |
| 2 | 47480726 | T | A | V | D | MSH2 | ENST00000233146 | 0.79453 | 0.02989 | 0.73167 | 0.95981 | 0.97624 | 0.9605727296 | 0.96692 | 0.9995556474 | 0.486683 | 9.778991533 | 0 |
| 2 | 47480726 | T | C | V | A | MSH2 | ENST00000233146 | 0.74802 | 0.03379 | 0.72403 | 0.90316 | 0.95133 | 0.8107749972 | 0.92333 | 0.9969367981 | 0.425946 | 9.778991533 | 0 |
| 2 | 47480726 | T | G | V | G | MSH2 | ENST00000233146 | 0.77956 | 0.03304 | 0.59023 | 0.95983 | 0.9729 | 0.8933824065 | 0.9393 | 0.9994679093 | 0.486272 | 9.778991533 | 0 |
| 2 | 47480728 | G | A | A | T | MSH2 | ENST00000233146 | 0.8929 | 0.03718 | 0.61128 | 0.95725 | 0.9833 | 0.8010136335 | 0.95996 | 0.9996241331 | 0.495801 | 9.696074802 | 0 |
| 2 | 47480728 | G | C | A | P | MSH2 | ENST00000233146 | 0.90116 | 0.03313 | 0.68632 | 0.95218 | 0.98585 | 0.9404802533 | 0.9698 | 0.9995133877 | 0.498271 | 9.696074802 | 0 |
| 2 | 47480728 | G | T | A | S | MSH2 | ENST00000233146 | 0.86147 | 0.03783 | 0.56928 | 0.94972 | 0.97475 | 0.7106352019 | 0.95274 | 0.9991944432 | 0.467781 | 6.694601227 | 0 |
| 2 | 47480729 | C | G | A | G | MSH2 | ENST00000233146 | 0.6735 | 0.03646 | 0.59385 | 0.92113 | 0.96553 | 0.7158307102 | 0.94997 | 0.9915344119 | 0.479644 | 6.444275445 | 0 |
| 2 | 47480729 | C | T | A | V | MSH2 | ENST00000233146 | 0.67613 | 0.03595 | 0.63408 | 0.95839 | 0.97067 | 0.8332287653 | 0.94716 | 0.999229908 | 0.425102 | 9.696074802 | 0 |
| 2 | 47480731 | G | A | E | K | MSH2 | ENST00000233146 | 0.61261 | 0.03518 | 0.72516 | 0.7146 | 0.93083 | 0.6375882093 | 0.96449 | 0.9639465809 | 0.380302 | 4.1826398 | 0 |
| 2 | 47480731 | G | C | E | Q | MSH2 | ENST00000233146 | 0.85227 | 0.03728 | 0.60149 | 0.68514 | 0.93319 | 0.5337137784 | 0.96189 | 0.8486532569 | 0.354958 | 4.421172882 | 0 |
| 2 | 47480732 | A | C | E | A | MSH2 | ENST00000233146 | 0.83729 | 0.03602 | 0.60416 | 0.85345 | 0.96956 | 0.6281976721 | 0.96012 | 0.9896259089 | 0.399727 | 7.191300518 | 0 |
| 2 | 47480732 | A | G | E | G | MSH2 | ENST00000233146 | 0.90949 | 0.03465 | 0.55492 | 0.90459 | 0.98477 | 0.6587980812 | 0.95069 | 0.9929701668 | 0.445221 | 7.388475149 | 0 |
| 2 | 47480732 | A | T | E | V | MSH2 | ENST00000233146 | 0.86453 | 0.03475 | 0.57829 | 0.90618 | 0.98146 | 0.6481450155 | 0.96233 | 0.9937510278 | 0.445471 | 9.172978178 | 0 |
| 2 | 47480733 | G | C | E | D | MSH2 | ENST00000233146 | 0.70696 | 0.03189 | 0.60767 | 0.87512 | 0.94129 | 0.5425097237 | 0.95609 | 0.9634181261 | 0.356613 | 9.172978178 | 0 |
| 2 | 47480733 | G | T | E | D | MSH2 | ENST00000233146 | 0.70045 | 0.03189 | 0.60767 | 0.87294 | 0.94129 | 0.5425097237 | 0.95648 | 0.9417327642 | 0.230238 | 9.172978178 | 0 |
| 2 | 47480734 | C | A | L | I | MSH2 | ENST00000233146 | 0.48889 | 0.01285 | 0.75995 | 0.73391 | 0.86098 | 0.4228880872 | 0.88906 | 0.8270692353 | 0.228373 | 5.09891245 | 0 |
| 2 | 47480734 | C | G | L | V | MSH2 | ENST00000233146 | 0.60786 | 0.01906 | 0.71531 | 0.81388 | 0.91357 | 0.5937173979 | 0.95271 | 0.8783699175 | 0.342684 | 8.587647758 | 0 |
| 2 | 47480734 | C | T | L | F | MSH2 | ENST00000233146 | 0.6594 | 0.0194 | 0.75138 | 0.78278 | 0.91186 | 0.6225597555 | 0.93454 | 0.9429619312 | 0.361485 | 8.587647758 | 0 |
| 2 | 47480735 | T | A | L | H | MSH2 | ENST00000233146 | 0.77156 | 0.03243 | 0.82302 | 0.90938 | 0.98764 | 0.7629587293 | 0.958 | 0.9971681237 | 0.467635 | 8.587647758 | 0 |
| 2 | 47480735 | T | C | L | P | MSH2 | ENST00000233146 | 0.79618 | 0.02864 | 0.85252 | 0.9184 | 0.98904 | 0.9066127213 | 0.97177 | 0.9989783764 | 0.467634 | 8.587647758 | 0 |
| 2 | 47480735 | T | G | L | R | MSH2 | ENST00000233146 | 0.79128 | 0.02696 | 0.83062 | 0.87897 | 0.98294 | 0.8777926178 | 0.97707 | 0.9953383675 | 0.455912 | 6.037228734 | 0 |
| 2 | 47480737 | G | A | A | T | MSH2 | ENST00000233146 | 0.89012 | 0.03552 | 0.67436 | 0.90579 | 0.95608 | 0.7716937378 | 0.94501 | 0.5758918524 | 0.251908 | 8.103717954 | 0 |
| 2 | 47480737 | G | C | A | P | MSH2 | ENST00000233146 | 0.899 | 0.03287 | 0.73059 | 0.88044 | 0.98657 | 0.8947165994 | 0.96191 | 0.9980325103 | 0.46756 | 8.103717954 | 1 |
| 2 | 47480737 | G | T | A | S | MSH2 | ENST00000233146 | 0.85822 | 0.03735 | 0.62183 | 0.84486 | 0.94356 | 0.6778399791 | 0.92605 | 0.9479112768 | 0.372276 | 5.501213626 | 0 |
| 2 | 47480738 | C | A | A | D | MSH2 | ENST00000233146 | 0.63965 | 0.03053 | 0.7761 | 0.9181 | 0.95753 | 0.9025088291 | 0.95344 | 0.9994249344 | 0.45264 | 8.103717954 | 1 |
| 2 | 47480738 | C | G | A | G | MSH2 | ENST00000233146 | 0.62962 | 0.03595 | 0.64464 | 0.90089 | 0.93631 | 0.6475668248 | 0.90977 | 0.9964506626 | 0.397837 | 8.103717954 | 0 |
| 2 | 47480738 | C | T | A | V | MSH2 | ENST00000233146 | 0.4507 | 0.01096 | 0.68039 | 0.67906 | 0.86047 | 0.7633783603 | 0.96697 | 0.9320503473 | 0.373344 | 4.484969829 | 0 |
| 2 | 47480740 | A | C | N | H | MSH2 | ENST00000233146 | 0.38352 | 0.00562 | 0.26156 | 0.80772 | 0.59638 | 0.2040645181 | 0.8253 | 0.1340437624 | -0.0798649 | 0.3258680102 | 0 |
| 2 | 47480740 | A | G | N | D | MSH2 | ENST00000233146 | 0.39393 | 0.00547 | 0.31559 | 0.86569 | 0.62876 | 0.1287234777 | 0.84008 | 0.3257885359 | 0.0441577 | 2.224247029 | 0 |
| 2 | 47480740 | A | T | N | Y | MSH2 | ENST00000233146 | 0.74874 | 0.00664 | 0.29409 | 0.90739 | 0.825 | 0.2355604989 | 0.89542 | 0.7965923548 | 0.194246 | 5.31507259 | 0 |
| 2 | 47480741 | A | C | N | T | MSH2 | ENST00000233146 | 0.44649 | 0.0063 | 0.24522 | 0.64816 | 0.77076 | 0.1614452871 | 0.89631 | 0.3169228293 | 0.0859717 | 5.31507259 | 0 |
| 2 | 47480741 | A | G | N | S | MSH2 | ENST00000233146 | 0.37725 | 0.00608 | 0.19666 | 0.52375 | 0.56426 | 0.07712082262 | 0.80972 | 0.1384350654 | -0.0883032 | 2.546864865 | 0 |
| 2 | 47480741 | A | T | N | I | MSH2 | ENST00000233146 | 0.62339 | 0.00543 | 0.27659 | 0.8104 | 0.84694 | 0.359072246 | 0.90068 | 0.740133822 | 0.175154 | 5.31507259 | 0 |
| 2 | 47480742 | T | A | N | K | MSH2 | ENST00000233146 | 0.33425 | 0.00608 | 0.34482 | 0.6368 | 0.67843 | 0.2057511531 | 0.81185 | 0.3077068627 | 0.0466897 | 2.439387463 | 0 |
| 2 | 47480742 | T | G | N | K | MSH2 | ENST00000233146 | 0.33979 | 0.00608 | 0.34482 | 0.6361 | 0.67843 | 0.2057511531 | 0.81122 | 0.3077068627 | 0.0466917 | 2.439387463 | 0 |
| 2 | 47480743 | T | A | F | I | MSH2 | ENST00000233146 | 0.8714 | 0.03224 | 0.68744 | 0.87418 | 0.92642 | 0.7679524232 | 0.93415 | 0.9984028935 | 0.328366 | 4.380175599 | 0 |
| 2 | 47480743 | T | C | F | L | MSH2 | ENST00000233146 | 0.88157 | 0.03491 | 0.72298 | 0.79418 | 0.95133 | 0.7043162663 | 0.95003 | 0.9915956855 | 0.390817 | 3.686961476 | 0 |
| 2 | 47480743 | T | G | F | V | MSH2 | ENST00000233146 | 0.87703 | 0.03313 | 0.66537 | 0.88765 | 0.95753 | 0.7686350947 | 0.95761 | 0.996552825 | 0.412198 | 4.231969481 | 0 |
| 2 | 47480744 | T | A | F | Y | MSH2 | ENST00000233146 | 0.86962 | 0.03527 | 0.70325 | 0.86092 | 0.95535 | 0.6890333926 | 0.92092 | 0.99133396239 | 0.328562 | 8.957162682 | 0 |
| 2 | 47480744 | T | C | F | S | MSH2 | ENST00000233146 | 0.92778 | 0.02864 | 0.68836 | 0.90371 | 0.97923 | 0.8581516523 | 0.96062 | 0.9990311861 | 0.462055 | 8.957162682 | 1 |
| 2 | 47480744 | T | G | F | C | MSH2 | ENST00000233146 | 0.92754 | 0.03175 | 0.71915 | 0.90113 | 0.9822 | 0.8313011647 | 0.96088 | 0.9990261793 | 0.451634 | 8.957162682 | 0 |
| 2 | 47480745 | C | A | F | L | MSH2 | ENST00000233146 | 0.61723 | 0.03491 | 0.72298 | 0.83465 | 0.87247 | 0.7043162663 | 0.95673 | 0.9901459813 | 0.353751 | 3.686961476 | 0 |
| 2 | 47480745 | C | G | F | L | MSH2 | ENST00000233146 | 0.62521 | 0.03491 | 0.72298 | 0.83751 | 0.87247 | 0.7043162663 | 0.95649 | 0.9901459813 | 0.353752 | 3.686961476 | 0 |
| 2 | 47480746 | C | A | P | T | MSH2 | ENST00000233146 | 0.71891 | 0.03527 | 0.59947 | 0.90786 | 0.98834 | 0.7546125323 | 0.95662 | 0.9997380376 | 0.540352 | 7.400248302 | 0 |
| 2 | 47480746 | C | G | P | A | MSH2 | ENST00000233146 | 0.69732 | 0.03713 | 0.5416 | 0.91261 | 0.99105 | 0.7038685562 | 0.951 | 0.9997385144 | 0.539332 | 9.347354098 | 0 |
| 2 | 47480746 | C | T | P | S | MSH2 | ENST00000233146 | 0.74802 | 0.03674 | 0.55851 | 0.91013 | 0.97067 | 0.7766726278 | 0.91593 | 0.9984778762 | 0.444047 | 7.231917911 | 0 |
| 2 | 47480747 | C | A | P | H | MSH2 | ENST00000233146 | 0.67461 | 0.03484 | 0.62313 | 0.93209 | 0.98109 | 0.785491791 | 0.93196 | 0.9997380376 | 0.541215 | 9.347354098 | 0 |
| 2 | 47480747 | C | G | P | R | MSH2 | ENST00000233146 | 0.67859 | 0.03189 | 0.63988 | 0.90511 | 0.9844 | 0.8040892013 | 0.94974 | 0.9997380376 | 0.540714 | 7.133731914 | 0 |
| 2 | 47480747 | C | T | P | L | MSH2 | ENST00000233146 | 0.69724 | 0.03626 | 0.64099 | 0.91239 | 0.96334 | 0.800959293 | 0.93146 | 0.9999763966 | 0.517193 | 9.347354098 | 0 |
| 2 | 47480749 | A | C | K | Q | MSH2 | ENST00000233146 | 0.37036 | 0.00586 | 0.03126 | 0.46671 | 0.69846 | 0.2784972898 | 0.92067 | 0.5359835625 | 0.0543949 | 1.58935159 | 0 |
| 2 | 47480749 | A | G | K | E | MSH2 | ENST00000233146 | 0.3541 | 0.00522 | 0.05537 | 0.47776 | 0.64876 | 0.2752334583 | 0.84243 | 0.4054689705 | 0.034946 | 2.173346297 | 0 |
| 2 | 47480750 | A | C | K | T | MSH2 | ENST00000233146 | 0.33656 | 0.00722 | 0.06929 | 0.58465 | 0.76223 | 0.3634339319 | 0.94787 | 0.1032524809 | 0.0865997 | 3.507276361 | 0 |
| 2 | 47480750 | A | G | K | R | MSH2 | ENST00000233146 | 0.30147 | 0.00608 | 0.03669 | 0.28661 | 0.64468 | 0.228890099 | 0.82394 | 0.1244743615 | 0.02499 | 2.044124068 | 0 |
| 2 | 47480750 | A | T | K | M | MSH2 | ENST00000233146 | 0.51041 | 0.01628 | 0.03698 | 0.80906 | 0.83172 | 0.429832875 | 0.88897 | 0.824582994 | 0.0925227 | 5.188631068 | 0 |
| 2 | 47480751 | G | C | K | N | MSH2 | ENST00000233146 | 0.36164 | 0.00586 | 0.06591 | 0.6601 | 0.64261 | 0.3307737617 | 0.81747 | 0.1748281941 | 0.0138012 | 3.42554702 | 0 |
| 2 | 47480751 | G | T | K | N | MSH2 | ENST00000233146 | 0.35713 | 0.00586 | 0.06591 | 0.70268 | 0.64261 | 0.3307737617 | 0.81742 | 0.1795695683 | 0.0138102 | 3.42554702 | 0 |
| 2 | 47480752 | C | A | H | N | MSH2 | ENST00000233146 | 0.41186 | 0.00608 | 0.53292 | 0.58204 | 0.60717 | 0.3688267517 | 0.83739 | 0.3967167109 | 0.0796227 | 3.742168252 | 0 |
| 2 | 47480752 | C | G | H | D | MSH2 | ENST00000233146 | 0.40109 | 0.00543 | 0.52682 | 0.63129 | 0.73296 | 0.5321468412 | 0.94528 | 0.3679832369 | 0.269704 | 3.759491399 | 0 |
| 2 | 47480752 | C | T | H | Y | MSH2 | ENST00000233146 | 0.74507 | 0.02574 | 0.53297 | 0.67148 | 0.85222 | 0.4637766709 | 0.88433 | 0.7904328704 | 0.208542 | 7.227635165 | 0 |
| 2 | 47480753 | A | C | H | P | MSH2 | ENST00000233146 | 0.61164 | 0.02023 | 0.45823 | 0.73779 | 0.88496 | 0.7508364457 | 0.92243 | 0.9161273241 | 0.313333 | 4.174934469 | 0 |

| | | | | | | | | | | | | | | | | | |
|---|---|---|---|---|---|---|---|---|---|---|---|---|---|---|---|---|---|
| 2 | 47480753 | A | G | H | R | MSH2 | ENST00000233146 | 0.42663 | 0.0109 | 0.53408 | 0.6665 | 0.82948 | 0.4695355661 | 0.97323 | 0.1404672622 | 0.121834 | 4.873842032 | 0 |
| 2 | 47480753 | A | T | H | L | MSH2 | ENST00000233146 | 0.47955 | 0.01086 | 0.49317 | 0.78778 | 0.87974 | 0.5469194705 | 0.94165 | 0.8836714029 | 0.308299 | 5.282506402 | 0 |
| 2 | 47480754 | T | A | H | Q | MSH2 | ENST00000233146 | 0.36766 | 0.00718 | 0.50013 | 0.66363 | 0.77205 | 0.2809219283 | 0.96582 | 0.1764945227 | 0.142973 | 4.40138282 | 0 |
| 2 | 47480754 | T | G | H | Q | MSH2 | ENST00000233146 | 0.37346 | 0.00718 | 0.50013 | 0.64915 | 0.77205 | 0.2809219283 | 0.96591 | 0.3243579865 | 0.153751 | 4.40138282 | 0 |
| 2 | 47480755 | G | A | V | I | MSH2 | ENST00000233146 | 0.82771 | 0.03848 | 0.35866 | 0.69003 | 0.86951 | 0.5196496412 | 0.81501 | 0.8093033412 | 0.22961 | 4.829543694 | 0 |
| 2 | 47480755 | G | C | V | L | MSH2 | ENST00000233146 | 0.86039 | 0.03088 | 0.54693 | 0.81606 | 0.96517 | 0.7309644987 | 0.9308 | 0.9696900249 | 0.445699 | 5.168896285 | 0 |
| 2 | 47480755 | G | T | V | L | MSH2 | ENST00000233146 | 0.85441 | 0.03088 | 0.54693 | 0.81606 | 0.96627 | 0.7309644987 | 0.93097 | 0.9696900249 | 0.445699 | 5.168896285 | 0 |
| 2 | 47480756 | T | A | V | E | MSH2 | ENST00000233146 | 0.78067 | 0.02989 | 0.55032 | 0.93824 | 0.99202 | 0.9106878045 | 0.96841 | 0.9997320771 | 0.483769 | 9.833145824 | 1 |
| 2 | 47480756 | T | C | V | A | MSH2 | ENST00000233146 | 0.74141 | 0.03419 | 0.56885 | 0.87835 | 0.98257 | 0.7587613731 | 0.94839 | 0.9903770089 | 0.45508 | 9.200280818 | 0 |
| 2 | 47480756 | T | G | V | G | MSH2 | ENST00000233146 | 0.77585 | 0.03313 | 0.42109 | 0.93399 | 0.99356 | 0.860345447 | 0.95385 | 0.9994285703 | 0.484038 | 9.833145824 | 0 |
| 2 | 47480758 | A | C | I | L | MSH2 | ENST00000233146 | 0.47652 | 0.00659 | 0.29485 | 0.6748 | 0.74376 | 0.5263553363 | 0.86854 | 0.6286564469 | 0.0813172 | 2.57856607 | 0 |
| 2 | 47480758 | A | G | I | V | MSH2 | ENST00000233146 | 0.38553 | 0.00654 | 0.21122 | 0.42793 | 0.65377 | 0.3063382728 | 0.83671 | 0.4634926021 | 0.0667505 | 2.189161006 | 0 |
| 2 | 47480758 | A | T | I | L | MSH2 | ENST00000233146 | 0.47124 | 0.00659 | 0.29485 | 0.67525 | 0.74376 | 0.5263553363 | 0.86681 | 0.6286564469 | 0.081322 | 2.57856607 | 0 |
| 2 | 47480759 | T | A | I | K | MSH2 | ENST00000233146 | 0.74786 | 0.02547 | 0.37829 | 0.92878 | 0.97587 | 0.8577212249 | 0.90503 | 0.9995704293 | 0.434396 | 8.260760668 | 0 |
| 2 | 47480759 | T | C | I | T | MSH2 | ENST00000233146 | 0.72853 | 0.02225 | 0.43442 | 0.88074 | 0.96189 | 0.6521439036 | 0.90294 | 0.9858635068 | 0.340292 | 3.84119514 | 0 |
| 2 | 47480759 | T | G | I | R | MSH2 | ENST00000233146 | 0.76187 | 0.02614 | 0.35116 | 0.91817 | 0.97624 | 0.8587266894 | 0.8921 | 0.9989913106 | 0.424672 | 8.260760668 | 0 |
| 2 | 47480760 | A | G | I | M | MSH2 | ENST00000233146 | 0.68275 | 0.03118 | 0.26339 | 0.86492 | 0.911 | 0.5868500034 | 0.91838 | 0.959896028 | 0.225707 | 8.260760668 | 0 |
| 2 | 47480761 | G | A | E | K | MSH2 | ENST00000233146 | 0.64641 | 0.00893 | 0.31293 | 0.61363 | 0.81818 | 0.4304860627 | 0.92217 | 0.8130729198 | 0.245581 | 3.52368124 | 0 |
| 2 | 47480761 | G | C | E | Q | MSH2 | ENST00000233146 | 0.57787 | 0.0071 | 0.19998 | 0.62272 | 0.70589 | 0.3622064232 | 0.91479 | 0.8134903312 | 0.208442 | 3.23338272 | 0 |
| 2 | 47480762 | A | C | E | A | MSH2 | ENST00000233146 | 0.38983 | 0.00569 | 0.21472 | 0.5474 | 0.66449 | 0.4302221684 | 0.8127 | 0.6598694921 | 0.0709352 | 2.908716153 | 0 |
| 2 | 47480762 | A | G | E | G | MSH2 | ENST00000233146 | 0.57787 | 0.00881 | 0.17943 | 0.72768 | 0.85117 | 0.4421845289 | 0.85537 | 0.9055601954 | 0.150398 | 5.461658971 | 0 |
| 2 | 47480762 | A | T | E | V | MSH2 | ENST00000233146 | 0.69058 | 0.0124 | 0.19477 | 0.78623 | 0.90007 | 0.4951463989 | 0.92015 | 0.9875176549 | 0.255086 | 5.332642092 | 0 |
| 2 | 47480763 | G | C | E | D | MSH2 | ENST00000233146 | 0.25729 | 0.00608 | 0.21389 | 0.57473 | 0.76024 | 0.3865686088 | 0.95963 | 0.3552083671 | 0.130932 | 2.464958943 | 0 |
| 2 | 47480763 | G | T | E | D | MSH2 | ENST00000233146 | 0.25363 | 0.00608 | 0.21389 | 0.56612 | 0.76024 | 0.3865686088 | 0.95975 | 0.5628438592 | 0.130932 | 2.464958943 | 0 |
| 2 | 47480764 | T | A | C | S | MSH2 | ENST00000233146 | 0.44265 | 0.0054 | 0.20118 | 0.39279 | 0.56156 | 0.2351056141 | 0.70837 | 0.58975178 | 0.026145 | -0.8536246914 | 0 |
| 2 | 47480764 | T | C | C | R | MSH2 | ENST00000233146 | 0.5027 | 0.00608 | 0.295 | 0.54343 | 0.73734 | 0.4982099721 | 0.91959 | 0.7223767638 | 0.128607 | -0.0785472022 | 0 |
| 2 | 47480764 | T | G | C | G | MSH2 | ENST00000233146 | 0.49248 | 0.00599 | 0.19764 | 0.59116 | 0.71938 | 0.2989614326 | 0.94006 | 0.2842644262 | 0.133436 | 2.312203953 | 0 |
| 2 | 47480765 | G | A | C | Y | MSH2 | ENST00000233146 | 0.29803 | 0.00492 | 0.31849 | 0.51029 | 0.68734 | 0.4135752599 | 0.80303 | 0.04137761518 | -0.14925 | 0.922916536 | 0 |
| 2 | 47480765 | G | C | C | S | MSH2 | ENST00000233146 | 0.34904 | 0.0054 | 0.20118 | 0.41929 | 0.57748 | 0.2351056141 | 0.68639 | 0.1577216055 | 0.0117621 | -0.8536246914 | 0 |
| 2 | 47480765 | G | T | C | F | MSH2 | ENST00000233146 | 0.33543 | 0.00504 | 0.24467 | 0.47786 | 0.74093 | 0.3453803536 | 0.93923 | 0.1526456922 | 0.0984793 | -0.636434881 | 0 |
| 2 | 47480766 | T | G | C | W | MSH2 | ENST00000233146 | 0.51303 | 0.02108 | 0.26179 | 0.76247 | 0.79028 | 0.4544340431 | 0.85842 | 0.7899316549 | 0.0594122 | 3.238820453 | 0 |
| 2 | 47480767 | G | A | A | T | MSH2 | ENST00000233146 | 0.89971 | 0.03633 | 0.47344 | 0.90137 | 0.97661 | 0.7732636633 | 0.9603 | 0.9993433356 | 0.483128 | 9.047170483 | 0 |
| 2 | 47480767 | G | C | A | P | MSH2 | ENST00000233146 | 0.9074 | 0.02315 | 0.53634 | 0.88861 | 0.98799 | 0.9274621639 | 0.97145 | 0.999194324 | 0.501485 | 9.047170483 | 1 |
| 2 | 47480767 | G | T | A | S | MSH2 | ENST00000233146 | 0.8658 | 0.0317 | 0.42802 | 0.83262 | 0.95353 | 0.6751883939 | 0.94718 | 0.9887645638 | 0.443696 | 3.836595257 | 0 |
| 2 | 47480768 | C | A | A | D | MSH2 | ENST00000233146 | 0.69131 | 0.03076 | 0.583 | 0.90702 | 0.98728 | 0.9325638088 | 0.96905 | 0.999555409 | 0.489921 | 9.047170483 | 0 |
| 2 | 47480768 | C | G | A | G | MSH2 | ENST00000233146 | 0.68131 | 0.03582 | 0.45182 | 0.87318 | 0.95243 | 0.681329752 | 0.9222 | 0.9961966276 | 0.349771 | 9.047170483 | 0 |
| 2 | 47480768 | C | T | A | V | MSH2 | ENST00000233146 | 0.68555 | 0.02663 | 0.49629 | 0.89839 | 0.97923 | 0.8100952314 | 0.95651 | 0.9995242357 | 0.488027 | 9.047170483 | 0 |
| 2 | 47480770 | A | C | K | Q | MSH2 | ENST00000233146 | 0.44606 | 0.00909 | 0.13694 | 0.67676 | 0.89467 | 0.4613214057 | 0.87787 | 0.8629508615 | 0.0526823 | 3.864089447 | 0 |
| 2 | 47480770 | A | G | K | E | MSH2 | ENST00000233146 | 0.59929 | 0.00827 | 0.17759 | 0.72243 | 0.93977 | 0.5192235929 | 0.98361 | 0.9503465295 | 0.366285 | 4.268045647 | 0 |
| 2 | 47480771 | A | C | K | T | MSH2 | ENST00000233146 | 0.53636 | 0.02307 | 0.19569 | 0.81287 | 0.89873 | 0.548082035 | 0.84604 | 0.9799388647 | 0.212678 | 4.73462286 | 0 |
| 2 | 47480771 | A | G | K | R | MSH2 | ENST00000233146 | 0.25875 | 0.00633 | 0.12879 | 0.42339 | 0.75349 | 0.3543496763 | 0.87307 | 0.3364725113 | 0.0621676 | 1.006876801 | 0 |
| 2 | 47480771 | A | T | K | I | MSH2 | ENST00000233146 | 0.57336 | 0.02442 | 0.1726 | 0.89945 | 0.88165 | 0.6123444985 | 0.87916 | 0.9923089743 | 0.217455 | 7.022363572 | 0 |
| 2 | 47480772 | A | C | K | N | MSH2 | ENST00000233146 | 0.62294 | 0.02403 | 0.19626 | 0.81828 | 0.87394 | 0.5375685526 | 0.81843 | 0.9959964752 | 0.0897227 | 3.703279673 | 0 |
| 2 | 47480772 | A | T | K | N | MSH2 | ENST00000233146 | 0.61656 | 0.02403 | 0.19626 | 0.82342 | 0.87442 | 0.5375685526 | 0.81808 | 0.9959964752 | 0.0897185 | 3.703279673 | 0 |
| 2 | 47480773 | C | A | Q | K | MSH2 | ENST00000233146 | 0.34669 | 0.00556 | 0.24302 | 0.55729 | 0.7285 | 0.3148893141 | 0.90461 | 0.636100471 | 0.134582 | 2.131154626 | 0 |
| 2 | 47480773 | C | G | Q | E | MSH2 | ENST00000233146 | 0.26814 | 0.00562 | 0.20318 | 0.28208 | 0.61761 | 0.1960478068 | 0.8072 | 0.6920224428 | 0.0402111 | 0.4204632435 | 0 |
| 2 | 47480774 | A | C | Q | P | MSH2 | ENST00000233146 | 0.77742 | 0.01871 | 0.21358 | 0.87417 | 0.89376 | 0.6381334046 | 0.89457 | 0.9612085223 | 0.237165 | 6.453011805 | 0 |
| 2 | 47480774 | A | G | Q | R | MSH2 | ENST00000233146 | 0.35936 | 0.00592 | 0.23188 | 0.64177 | 0.65673 | 0.2607881802 | 0.89882 | 0.06323017737 | -0.134929 | 2.847586157 | 0 |
| 2 | 47480774 | A | T | Q | L | MSH2 | ENST00000233146 | 0.62233 | 0.00996 | 0.19679 | 0.83314 | 0.81703 | 0.3674820377 | 0.92962 | 0.9870382547 | 0.202479 | 3.8585048 | 0 |
| 2 | 47480775 | G | C | Q | H | MSH2 | ENST00000233146 | 0.28276 | 0.02802 | 0.23374 | 0.78134 | 0.862 | 0.2859247661 | 0.87352 | 0.9596774578 | 0.131923 | 3.642528217 | 0 |
| 2 | 47480775 | G | T | Q | H | MSH2 | ENST00000233146 | 0.27884 | 0.02802 | 0.23374 | 0.79357 | 0.862 | 0.2859247661 | 0.87276 | 0.9707334638 | 0.131938 | 3.642528217 | 0 |
| 2 | 47480776 | A | G | K | E | MSH2 | ENST00000233146 | 0.87221 | 0.03033 | 0.59206 | 0.84083 | 0.97215 | 0.7803677544 | 0.95181 | 0.9943586081 | 0.455196 | 8.740317177 | 0 |
| 2 | 47480777 | A | C | K | T | MSH2 | ENST00000233146 | 0.85829 | 0.03404 | 0.59867 | 0.87922 | 0.96993 | 0.7233358689 | 0.92462 | 0.9943551234 | 0.419126 | 8.740317177 | 0 |
| 2 | 47480777 | A | G | K | R | MSH2 | ENST00000233146 | 0.8291 | 0.02403 | 0.5271 | 0.72582 | 0.90007 | 0.6611557649 | 0.94256 | 0.9667453487 | 0.274999 | 7.649405632 | 0 |
| 2 | 47480777 | A | T | K | I | MSH2 | ENST00000233146 | 0.89555 | 0.02785 | 0.56186 | 0.8659 | 0.94543 | 0.7340892979 | 0.95234 | 0.9973662496 | 0.389203 | 3.437385372 | 0 |
| 2 | 47480778 | A | C | K | N | MSH2 | ENST00000233146 | 0.63835 | 0.0334 | 0.61963 | 0.87543 | 0.91229 | 0.7587479326 | 0.93955 | 0.9927224151 | 0.131673 | 8.740317177 | 0 |
| 2 | 47480778 | A | T | K | N | MSH2 | ENST00000233146 | 0.63191 | 0.0334 | 0.61963 | 0.87512 | 0.91186 | 0.7587479326 | 0.93939 | 0.9927224151 | 0.13167 | 8.740317177 | 0 |
| 2 | 47480779 | G | A | A | T | MSH2 | ENST00000233146 | 0.87958 | 0.03099 | 0.47929 | 0.81326 | 0.95207 | 0.5758845005 | 0.93258 | 0.8988275963 | 0.307548 | 8.509457727 | 0 |
| 2 | 47480779 | G | C | A | P | MSH2 | ENST00000233146 | 0.89834 | 0.03313 | 0.53599 | 0.841 | 0.9796 | 0.8455862031 | 0.94196 | 0.9990869761 | 0.457878 | 8.509457727 | 0 |
| 2 | 47480779 | G | T | A | S | MSH2 | ENST00000233146 | 0.85508 | 0.03718 | 0.43196 | 0.76069 | 0.93319 | 0.4881250304 | 0.94851 | 0.9566956758 | 0.36066 | 5.595473282 | 0 |
| 2 | 47480780 | C | A | A | D | MSH2 | ENST00000233146 | 0.66327 | 0.03093 | 0.57997 | 0.87927 | 0.9703 | 0.8549500934 | 0.93959 | 0.9998111129 | 0.455638 | 8.509457727 | 0 |
| 2 | 47480780 | C | G | A | G | MSH2 | ENST00000233146 | 0.65453 | 0.03646 | 0.45303 | 0.81816 | 0.92601 | 0.5073180654 | 0.87226 | 0.992927134 | 0.37301 | 8.509457727 | 0 |
| 2 | 47480780 | C | T | A | V | MSH2 | ENST00000233146 | 0.53569 | 0.03595 | 0.49943 | 0.7991 | 0.92723 | 0.6172987796 | 0.88663 | 0.9956249595 | 0.389294 | 4.787732289 | 0 |
| 2 | 47480782 | C | G | L | V | MSH2 | ENST00000233146 | 0.2203 | 0.00574 | 0.1886 | 0.54154 | 0.75005 | 0.2989802301 | 0.91728 | 0.1864119757 | -0.0467812 | 4.739736366 | 0 |
| 2 | 47480783 | T | A | L | Q | MSH2 | ENST00000233146 | 0.41837 | 0.00586 | 0.25495 | 0.71892 | 0.72398 | 0.3023394536 | 0.92821 | 0.8449502587 | 0.127453 | 3.265307844 | 0 |
| 2 | 47480783 | T | C | L | P | MSH2 | ENST00000233146 | 0.61454 | 0.01191 | 0.37192 | 0.86536 | 0.88823 | 0.7324842012 | 0.96558 | 0.8381578922 | 0.323838 | 7.788147417 | 0 |
| 2 | 47480783 | T | G | L | R | MSH2 | ENST00000233146 | 0.44869 | 0.0073 | 0.33475 | 0.77475 | 0.79089 | 0.4829698437 | 0.9798 | 0.655839622 | 0.263313 | 3.006221772 | 0 |
| 2 | 47480785 | G | A | E | K | MSH2 | ENST00000233146 | 0.93164 | 0.0218 | 0.62085 | 0.7872 | 0.89193 | 0.6972176148 | 0.93447 | 0.9200357199 | 0.286226 | 5.211696688 | 0 |
| 2 | 47480785 | G | C | E | Q | MSH2 | ENST00000233146 | 0.89673 | 0.01791 | 0.511 | 0.62143 | 0.84694 | 0.5545457455 | 0.88201 | 0.9260586405 | 0.206694 | 4.912048359 | 0 |
| 2 | 47480786 | A | C | E | A | MSH2 | ENST00000233146 | 0.85865 | 0.03269 | 0.52021 | 0.90891 | 0.91526 | 0.6412180612 | 0.78378 | 0.9888648987 | 0.189576 | 6.004223931 | 0 |
| 2 | 47480786 | A | G | E | G | MSH2 | ENST00000233146 | 0.90009 | 0.03353 | 0.473 | 0.92646 | 0.9528 | 0.6414304243 | 0.82212 | 0.9937484264 | 0.29911 | 6.330290065 | 0 |
| 2 | 47480786 | A | T | E | V | MSH2 | ENST00000233146 | 0.88622 | 0.0325 | 0.49147 | 0.93348 | 0.96298 | 0.6531622302 | 0.91124 | 0.9945381284 | 0.371963 | 6.400611177 | 0 |
| 2 | 47480787 | A | C | E | D | MSH2 | ENST00000233146 | 0.33212 | 0.01479 | 0.52091 | 0.78314 | 0.85012 | 0.5297950515 | 0.93404 | 0.9277665615 | 0.19102 | 5.386792174 | 0 |
| 2 | 47480787 | A | T | E | D | MSH2 | ENST00000233146 | 0.32764 | 0.01479 | 0.52091 | 0.78314 | 0.85012 | 0.5297950515 | 0.93444 | 0.9277665615 | 0.191014 | 5.386792174 | 0 |
| 2 | 47480788 | C | A | L | I | MSH2 | ENST00000233146 | 0.54821 | 0.02582 | 0.66144 | 0.89318 | 0.90494 | 0.4162365537 | 0.82597 | 0.7615415741 | 0.0508035 | 5.621983671 | 0 |
| 2 | 47480788 | C | G | L | V | MSH2 | ENST00000233146 | 0.54569 | 0.03362 | 0.61127 | 0.89789 | 0.92356 | 0.6001919462 | 0.8976 | 0.9911025763 | 0.314563 | 9.604530311 | 0 |
| 2 | 47480788 | C | T | L | F | MSH2 | ENST00000233146 | 0.56873 | 0.03547 | 0.64542 | 0.88759 | 0.92642 | 0.6198942891 | 0.90462 | 0.9943962097 | 0.304522 | 5.256114732 | 0 |
| 2 | 47480789 | T | A | L | H | MSH2 | ENST00000233146 | 0.73245 | 0.03327 | 0.72261 | 0.90218 | 0.9729 | 0.7785153437 | 0.94901 | 0.9996936321 | 0.488957 | 8.520201286 | 0 |
| 2 | 47480789 | T | C | L | P | MSH2 | ENST00000233146 | 0.75726 | 0.02994 | 0.74563 | 0.91118 | 0.9822 | 0.9043244711 | 0.97023 | 0.9994346499 | 0.488955 | 9.604530311 | 0 |
| 2 | 47480789 | T | G | L | R | MSH2 | ENST00000233146 | 0.75264 | 0.03137 | 0.72456 | 0.90247 | 0.98034 | 0.8887697572 | 0.96497 | 0.9977707863 | 0.488312 | 9.604530311 | 0 |
| 2 | 47480791 | G | A | E | K | MSH2 | ENST00000233146 | 0.89712 | 0.03419 | 0.60079 | 0.84268 | 0.93786 | 0.7983363929 | 0.94506 | 0.9964453578 | 0.448401 | 8.308236736 | 0 |
| 2 | 47480791 | G | C | E | Q | MSH2 | ENST00000233146 | 0.84188 | 0.03702 | 0.49029 | 0.75265 | 0.87588 | 0.6658078269 | 0.91982 | 0.9761593938 | 0.327664 | 7.862340696 | 0 |
| 2 | 47480792 | A | C | E | A | MSH2 | ENST00000233146 | 0.8349 | 0.03497 | 0.49534 | 0.88451 | 0.94986 | 0.7344746666 | 0.90482 | 0.9965956807 | 0.387516 | 7.440413065 | 0 |
| 2 | 47480792 | A | G | E | G | MSH2 | ENST00000233146 | 0.87788 | 0.03394 | 0.4513 | 0.88844 | 0.95207 | 0.7200092828 | 0.88887 | 0.9996386766 | 0.398119 | 7.413229389 | 0 |
| 2 | 47480792 | A | T | E | V | MSH2 | ENST00000233146 | 0.8624 | 0.03442 | 0.47688 | 0.90475 | 0.95462 | 0.7414339828 | 0.93547 | 0.9994282126 | 0.445333 | 7.267347626 | 0 |
| 2 | 47480793 | G | C | E | D | MSH2 | ENST00000233146 | 0.46855 | 0.03795 | 0.50186 | 0.8668 | 0.86352 | 0.6091320467 | 0.92126 | 0.9257910252 | 0.10803 | 6.760821284 | 0 |
| 2 | 47480793 | G | T | E | D | MSH2 | ENST00000233146 | 0.46351 | 0.03795 | 0.50186 | 0.87124 | 0.86352 | 0.6091320467 | 0.92171 | 0.9834678173 | 0.2344 | 6.760821284 | 0 |
| 2 | 47480794 | G | A | E | K | MSH2 | ENST00000233146 | 0.67621 | 0.00718 | 0.62931 | 0.83126 | 0.91569 | 0.4489524278 | 0.96424 | 0.9462117553 | 0.339544 | 4.060940471 | 0 |
| 2 | 47480794 | G | C | E | Q | MSH2 | ENST00000233146 | 0.51342 | 0.0084 | 0.52447 | 0.68433 | 0.86653 | 0.379978439 | 0.93069 | 0.9086577892 | 0.235331 | 4.029889183 | 0 |
| 2 | 47480795 | A | C | E | A | MSH2 | ENST00000233146 | 0.51527 | 0.00735 | 0.54067 | 0.80352 | 0.90185 | 0.4595721125 | 0.90169 | 0.5619866252 | 0.113759 | 3.994822592 | 0 |
| 2 | 47480795 | A | G | E | G | MSH2 | ENST00000233146 | 0.70919 | 0.00596 | 0.49679 | 0.87637 | 0.93939 | 0.4570690342 | 0.94345 | 0.864913073 | 0.316878 | 4.413356319 | 0 |
| 2 | 47480795 | A | T | E | V | MSH2 | ENST00000233146 | 0.82521 | 0.01126 | 0.51242 | 0.90359 | 0.94543 | 0.4977839927 | 0.95492 | 0.9902423024 | 0.363267 | 6.084590971 | 0 |
| 2 | 47480796 | G | C | E | D | MSH2 | ENST00000233146 | 0.19439 | 0.00603 | 0.53613 | 0.56039 | 0.79334 | 0.3165209751 | 0.94573 | 0.1345592439 | 0.139364 | 1.766355922 | 0 |
| 2 | 47480796 | G | T | E | D | MSH2 | ENST00000233146 | 0.19138 | 0.00603 | 0.53613 | 0.55931 | 0.79334 | 0.3165209751 | 0.94644 | 0.1345592439 | 0.13936 | 1.766355922 | 0 |
| 2 | 47480797 | T | A | F | I | MSH2 | ENST00000233146 | 0.65015 | 0.00866 | 0.68223 | 0.80415 | 0.90624 | 0.5513329356 | 0.90035 | 0.9847664833 | 0.320903 | 7.5773908 | 0 |
| 2 | 47480797 | T | C | F | L | MSH2 | ENST00000233146 | 0.53435 | 0.01676 | 0.71171 | 0.79347 | 0.90624 | 0.4718060378 | 0.93229 | 0.9789034128 | 0.378611 | 6.200116382 | 0 |
| 2 | 47480797 | T | G | F | V | MSH2 | ENST00000233146 | 0.81024 | 0.00821 | 0.66438 | 0.88766 | 0.94091 | 0.5168358333 | 0.95776 | 0.9792349935 | 0.439552 | 6.121564478 | 0 |
| 2 | 47480798 | T | A | F | Y | MSH2 | ENST00000233146 | 0.47024 | 0.00669 | 0.70177 | 0.75773 | 0.80058 | 0.3624308773 | 0.93746 | 0.6606459022 | 0.315793 | 2.845800472 | 0 |
| 2 | 47480798 | T | C | F | S | MSH2 | ENST00000233146 | 0.75941 | 0.01158 | 0.68422 | 0.90227 | 0.95207 | 0.5639025713 | 0.96947 | 0.9888384342 | 0.454269 | 4.079587147 | 0 |
| 2 | 47480798 | T | G | F | C | MSH2 | ENST00000233146 | 0.57617 | 0.00777 | 0.70277 | 0.82874 | 0.88916 | 0.5368576229 | 0.96831 | 0.7754665017 | 0.437239 | 8.523899393 | 0 |
| 2 | 47480799 | T | A | F | L | MSH2 | ENST00000233146 | 0.37549 | 0.01676 | 0.71171 | 0.84871 | 0.8933 | 0.4718060378 | 0.95371 | 0.9741723537 | 0.336222 | 6.200116382 | 0 |

| | | | | | | | | | | | | | | | | | | |
|---|---|---|---|---|---|---|---|---|---|---|---|---|---|---|---|---|---|---|
| 2 | 47480799 | T | G | F | L | MSH2 | ENST00000233146 | 0.38135 | 0.01676 | 0.71171 | 0.84816 | 0.8933 | 0.4718060378 | 0.95347 | 0.9741723537 | 0.336223 | 6.200116382 | 0 |
| 2 | 47480800 | C | G | Q | E | MSH2 | ENST00000233146 | 0.36114 | 0.00622 | 0.33694 | 0.6946 | 0.86403 | 0.5000120221 | 0.94889 | 0.4376940131 | 0.32917 | 4.288965892 | 0 |
| 2 | 47480801 | A | C | Q | P | MSH2 | ENST00000233146 | 0.78851 | 0.02243 | 0.34965 | 0.86318 | 0.88776 | 0.7458830322 | 0.95906 | 0.9090460539 | 0.442005 | 5.763972713 | 0 |
| 2 | 47480801 | A | G | Q | R | MSH2 | ENST00000233146 | 0.57202 | 0.01795 | 0.37423 | 0.83594 | 0.87 | 0.5703159486 | 0.97059 | 0.8442080021 | 0.431834 | 5.61498355 | 0 |
| 2 | 47480801 | A | T | Q | L | MSH2 | ENST00000233146 | 0.77354 | 0.01002 | 0.32081 | 0.86054 | 0.91986 | 0.5406541181 | 0.93975 | 0.859023869 | 0.373104 | 6.53049499 | 0 |
| 2 | 47480802 | G | C | Q | H | MSH2 | ENST00000233146 | 0.34071 | 0.00638 | 0.37869 | 0.75421 | 0.81646 | 0.565770736 | 0.97038 | 0.608217895 | 0.374353 | 4.384161459 | 0 |
| 2 | 47480802 | G | T | Q | H | MSH2 | ENST00000233146 | 0.33634 | 0.00638 | 0.37869 | 0.75833 | 0.81646 | 0.565770736 | 0.97064 | 0.608217895 | 0.37435 | 4.384161459 | 0 |
| 2 | 47480803 | T | A | Y | N | MSH2 | ENST00000233146 | 0.30203 | 0.00487 | 0.23601 | 0.72 | 0.56426 | 0.2198080053 | 0.47394 | 0.0399114415 | 0.0119814 | -3.308501356 | 0 |
| 2 | 47480803 | T | C | Y | H | MSH2 | ENST00000233146 | 0.29313 | 0.00527 | 0.22755 | 0.59937 | 0.50304 | 0.2541183174 | 0.569 | 0.04413749278 | 0.0104826 | -0.3659573879 | 0 |
| 2 | 47480803 | T | G | Y | D | MSH2 | ENST00000233146 | 0.33062 | 0.00476 | 0.25685 | 0.60239 | 0.59146 | 0.2834004667 | 0.56658 | 0.02770021184 | 0.0302545 | -1.190572723 | 0 |
| 2 | 47480804 | A | C | Y | S | MSH2 | ENST00000233146 | 0.36036 | 0.00512 | 0.18533 | 0.63888 | 0.57487 | 0.2536017108 | 0.73544 | 0.09179183841 | 0.0404374 | -2.27779974 | 0 |
| 2 | 47480804 | A | G | Y | C | MSH2 | ENST00000233146 | 0.42353 | 0.00527 | 0.22481 | 0.73111 | 0.71783 | 0.2571761067 | 0.89388 | 0.08816445521 | 0.0370962 | 1.091505242 | 0 |
| 2 | 47480804 | A | T | Y | F | MSH2 | ENST00000233146 | 0.29467 | 0.00586 | 0.20691 | 0.65794 | 0.60717 | 0.1768296809 | 0.85889 | 0.09238830209 | 0.0492745 | 0.07967099116 | 0 |
| 2 | 47480806 | A | C | I | L | MSH2 | ENST00000233146 | 0.29934 | 0.00569 | 0.14351 | 0.65988 | 0.74093 | 0.2293632579 | 0.90553 | 0.05248201931 | 0.0762107 | 2.17245833 | 0 |
| 2 | 47480806 | A | G | I | V | MSH2 | ENST00000233146 | 0.22881 | 0.00599 | 0.0841 | 0.51971 | 0.79697 | 0.1646156272 | 0.95721 | 0.04870323092 | 0.0908303 | 1.488653866 | 0 |
| 2 | 47480806 | A | T | I | F | MSH2 | ENST00000233146 | 0.31183 | 0.00497 | 0.14614 | 0.75016 | 0.79212 | 0.2957598324 | 0.97191 | 0.07684708688 | 0.125508 | 4.460624623 | 0 |
| 2 | 47480807 | T | A | I | N | MSH2 | ENST00000233146 | 0.49087 | 0.00476 | 0.21007 | 0.69929 | 0.78535 | 0.3818000714 | 0.87591 | 0.2209413089 | 0.190659 | 2.707190671 | 0 |
| 2 | 47480807 | T | C | I | T | MSH2 | ENST00000233146 | 0.423 | 0.00532 | 0.23823 | 0.59002 | 0.75755 | 0.2614299517 | 0.91678 | 0.1922658272 | 0.20854 | 2.56078709 | 0 |
| 2 | 47480807 | T | G | I | S | MSH2 | ENST00000233146 | 0.48793 | 0.00509 | 0.16133 | 0.59286 | 0.76223 | 0.4230399955 | 0.88722 | 0.2067543395 | 0.172583 | 2.899177172 | 0 |
| 2 | 47480808 | T | G | I | M | MSH2 | ENST00000233146 | 0.32913 | 0.0071 | 0.11711 | 0.70288 | 0.8233 | 0.2586583382 | 0.87163 | 0.06785723109 | 0.0470334 | 5.321886857 | 0 |
| 2 | 47480809 | G | A | G | R | MSH2 | ENST00000233146 | 0.65179 | 0.00586 | 0.35132 | 0.75637 | 0.85326 | 0.4315045822 | 0.98341 | 0.739671886 | 0.108173 | 4.142993198 | 0 |
| 2 | 47480809 | G | C | G | R | MSH2 | ENST00000233146 | 0.64859 | 0.00586 | 0.35132 | 0.75648 | 0.85326 | 0.4315045822 | 0.98227 | 0.9647094607 | 0.328279 | 4.142993198 | 0 |
| 2 | 47480810 | G | A | G | E | MSH2 | ENST00000233146 | 0.50094 | 0.00574 | 0.32838 | 0.57799 | 0.77527 | 0.3976195886 | 0.83577 | 0.7981852889 | 0.228257 | 3.190792351 | 0 |
| 2 | 47480810 | G | C | G | A | MSH2 | ENST00000233146 | 0.35459 | 0.00569 | 0.24466 | 0.55525 | 0.75485 | 0.2716388189 | 0.92771 | 0.78645432 | 0.219672 | 2.74520758 | 0 |
| 2 | 47480810 | G | T | G | V | MSH2 | ENST00000233146 | 0.49081 | 0.0058 | 0.2573 | 0.56983 | 0.81703 | 0.3341676606 | 0.96581 | 0.8538715243 | 0.38522 | 3.766241067 | 0 |
| 2 | 47480812 | G | A | E | K | MSH2 | ENST00000233146 | 0.12244 | 0.00547 | 0.02251 | 0.72161 | 0.74658 | 0.1966100505 | 0.81932 | 0.03841996716 | 0.00710682 | -1.638738966 | 0 |
| 2 | 47480812 | G | C | E | Q | MSH2 | ENST00000233146 | 0.11769 | 0.00574 | 0.01714 | 0.68176 | 0.69846 | 0.2284662678 | 0.87767 | 0.02424916252 | 0.0471951 | 0.8753081532 | 0 |
| 2 | 47480813 | A | C | E | A | MSH2 | ENST00000233146 | 0.08528 | 0.00556 | 0.01746 | 0.56521 | 0.7285 | 0.2393827734 | 0.70085 | 0.06366663218 | -0.00991036 | -0.5233202381 | 0 |
| 2 | 47480813 | A | G | E | G | MSH2 | ENST00000233146 | 0.11894 | 0.0054 | 0.01907 | 0.50347 | 0.7217 | 0.2117809244 | 0.68172 | 0.05889750585 | 0.0207712 | 0.3894711084 | 0 |
| 2 | 47480813 | A | T | E | V | MSH2 | ENST00000233146 | 0.11494 | 0.00543 | 0.01395 | 0.63988 | 0.68908 | 0.3022819182 | 0.74594 | 0.04483589952 | 0.0061301 | -0.738026002 | 0 |
| 2 | 47480814 | A | C | E | D | MSH2 | ENST00000233146 | 0.12543 | 0.00586 | 0.02221 | 0.6489 | 0.72775 | 0.2115785178 | 0.86423 | 0.0445510362 | 0.0255393 | 0.9737820555 | 0 |
| 2 | 47480814 | A | T | E | D | MSH2 | ENST00000233146 | 0.12276 | 0.00586 | 0.02221 | 0.6489 | 0.72775 | 0.2115785178 | 0.87001 | 0.03812126065 | 0.025595 | 0.9737820555 | 0 |
| 2 | 47480815 | T | A | S | T | MSH2 | ENST00000233146 | 0.16612 | 0.00608 | 0.06232 | 0.73168 | 0.80238 | 0.1931989102 | 0.95635 | 0.02075541533 | 0.0650497 | 1.896632259 | 0 |
| 2 | 47480815 | T | C | S | P | MSH2 | ENST00000233146 | 0.22525 | 0.00527 | 0.06596 | 0.62386 | 0.76356 | 0.2850360662 | 0.87957 | 0.0681598261 | 0.0397266 | 1.576527583 | 0 |
| 2 | 47480815 | T | G | S | A | MSH2 | ENST00000233146 | 0.17447 | 0.00603 | 0.05153 | 0.62119 | 0.72701 | 0.1408566955 | 0.94279 | 0.0336846821 | 0.050905 | 2.405628748 | 0 |
| 2 | 47480816 | C | G | S | W | MSH2 | ENST00000233146 | 0.27645 | 0.01645 | 0.06989 | 0.64833 | 0.76487 | 0.30723496 | 0.92425 | 0.7659276535 | 0.215092 | 7.082208038 | 0 |
| 2 | 47480816 | C | T | S | L | MSH2 | ENST00000233146 | 0.13033 | 0.00592 | 0.06283 | 0.53753 | 0.71312 | 0.2442090115 | 0.9114 | 0.148910163 | 0.157502 | 3.371134716 | 0 |
| 2 | 47480818 | C | A | Q | K | MSH2 | ENST00000233146 | 0.12636 | 0.00527 | 0.02097 | 0.60164 | 0.77011 | 0.2356942156 | 0.93036 | 0.03459539282 | 0.0771485 | 0.8173707745 | 0 |
| 2 | 47480818 | C | G | Q | E | MSH2 | ENST00000233146 | 0.08166 | 0.00556 | 0.01712 | 0.64861 | 0.70752 | 0.1781993205 | 0.84564 | 0.03677266364 | 0.0646578 | 0.738434176 | 0 |
| 2 | 47480819 | A | C | Q | P | MSH2 | ENST00000233146 | 0.27265 | 0.00556 | 0.01813 | 0.74611 | 0.82103 | 0.4526758263 | 0.96857 | 0.112700738 | 0.181279 | 2.404153261 | 0 |
| 2 | 47480819 | A | G | Q | R | MSH2 | ENST00000233146 | 0.24559 | 0.00556 | 0.01891 | 0.66529 | 0.78348 | 0.1765876963 | 0.94942 | 0.03213586658 | 0.0517181 | 1.276761862 | 0 |
| 2 | 47480819 | A | T | Q | L | MSH2 | ENST00000233146 | 0.28447 | 0.00574 | 0.01494 | 0.69143 | 0.72474 | 0.2908583219 | 0.92704 | 0.04200716689 | 0.0632341 | 1.131912906 | 0 |
| 2 | 47480820 | A | C | Q | H | MSH2 | ENST00000233146 | 0.2744 | 0.00664 | 0.02062 | 0.67676 | 0.84213 | 0.3126534902 | 0.95788 | 0.09664179647 | 0.156666 | 2.465289889 | 0 |
| 2 | 47480820 | A | T | Q | H | MSH2 | ENST00000233146 | 0.27043 | 0.00664 | 0.02062 | 0.67483 | 0.84213 | 0.3126534902 | 0.95814 | 0.1025598803 | 0.156655 | 2.465289889 | 0 |
| 2 | 47480821 | G | A | G | R | MSH2 | ENST00000233146 | 0.37024 | 0.00556 | 0.04028 | 0.67497 | 0.72474 | 0.2818200752 | 0.90719 | 0.3962336779 | 0.11925 | 1.725520369 | 0 |
| 2 | 47480821 | G | C | G | R | MSH2 | ENST00000233146 | 0.36794 | 0.00556 | 0.04028 | 0.68682 | 0.72474 | 0.2818200752 | 0.90708 | 0.5906744003 | 0.119256 | 1.725520369 | 0 |
| 2 | 47480822 | G | C | G | A | MSH2 | ENST00000233146 | 0.1563 | 0.00556 | 0.02486 | 0.58377 | 0.71233 | 0.220543957 | 0.92614 | 0.04425969223 | 0.0526141 | 0.4640125648 | 0 |
| 2 | 47480822 | G | T | G | V | MSH2 | ENST00000233146 | 0.1915 | 0.00547 | 0.02357 | 0.72257 | 0.76685 | 0.2665468275 | 0.95648 | 0.09494053968 | 0.112643 | 1.54246854 | 0 |
| 2 | 47480824 | T | A | Y | N | MSH2 | ENST00000233146 | 0.14572 | 0.00492 | 0.05351 | 0.63767 | 0.71627 | 0.2734831047 | 0.81893 | 0.04186878223 | 0.0172507 | -0.4456485003 | 0 |
| 2 | 47480824 | T | C | Y | H | MSH2 | ENST00000233146 | 0.14252 | 0.00527 | 0.05181 | 0.60035 | 0.65077 | 0.2899056968 | 0.83381 | 0.04918958896 | 0.0131719 | 0.6383787678 | 0 |
| 2 | 47480824 | T | G | Y | D | MSH2 | ENST00000233146 | 0.15061 | 0.00476 | 0.05842 | 0.64194 | 0.77141 | 0.3124223985 | 0.92002 | 0.0434602169 | 0.0397377 | -1.968431614 | 0 |
| 2 | 47480825 | A | C | Y | S | MSH2 | ENST00000233146 | 0.13698 | 0.00512 | 0.03792 | 0.70066 | 0.65575 | 0.2869208388 | 0.53731 | 0.02942700811 | -0.00609952 | -2.572085794 | 0 |
| 2 | 47480825 | A | G | Y | C | MSH2 | ENST00000233146 | 0.19294 | 0.00527 | 0.04555 | 0.62338 | 0.72398 | 0.2567825581 | 0.68562 | 0.06556033343 | 0.019413 | 0.6697875885 | 0 |
| 2 | 47480825 | A | T | Y | F | MSH2 | ENST00000233146 | 0.11597 | 0.00599 | 0.04318 | 0.68385 | 0.64571 | 0.1828476918 | 0.80898 | 0.03658529788 | 0.0183569 | 0.9480130913 | 0 |
| 2 | 47480827 | G | A | D | N | MSH2 | ENST00000233146 | 0.4894 | 0.00608 | 0.10077 | 0.72505 | 0.62985 | 0.2331187017 | 0.86952 | 0.6008536816 | 0.108543 | 3.304104495 | 0 |
| 2 | 47480827 | G | C | D | H | MSH2 | ENST00000233146 | 0.51698 | 0.00791 | 0.10594 | 0.74801 | 0.80058 | 0.3727867641 | 0.92255 | 0.8911274076 | 0.237108 | 4.048153358 | 0 |
| 2 | 47480827 | G | T | D | Y | MSH2 | ENST00000233146 | 0.63451 | 0.00799 | 0.10417 | 0.78856 | 0.82273 | 0.4281143876 | 0.95882 | 0.9394643903 | 0.273526 | 4.446857546 | 0 |
| 2 | 47480828 | A | C | D | A | MSH2 | ENST00000233146 | 0.38209 | 0.00532 | 0.07877 | 0.81976 | 0.81933 | 0.3305525851 | 0.96346 | 0.1797551039 | 0.205578 | 2.777192718 | 0 |
| 2 | 47480828 | A | G | D | G | MSH2 | ENST00000233146 | 0.28916 | 0.00522 | 0.08304 | 0.74761 | 0.72246 | 0.3172842898 | 0.95417 | 0.1123382964 | 0.114463 | 0.9764851895 | 0 |
| 2 | 47480828 | A | T | D | V | MSH2 | ENST00000233146 | 0.37947 | 0.00532 | 0.07298 | 0.79922 | 0.80476 | 0.4246540021 | 0.9757 | 0.3190639913 | 0.203634 | 3.330466196 | 0 |
| 2 | 47480830 | A | C | I | L | MSH2 | ENST00000233146 | 0.25031 | 0.00569 | 0.06959 | 0.75283 | 0.66733 | 0.2406920958 | 0.80372 | 0.1232089254 | 0.0615807 | 0.0279345431 | 0 |
| 2 | 47480830 | A | G | I | V | MSH2 | ENST00000233146 | 0.21629 | 0.00599 | 0.04986 | 0.75624 | 0.73734 | 0.2049519786 | 0.87379 | 0.08719490469 | 0.0789718 | 0.159146857 | 0 |
| 2 | 47480830 | A | T | I | F | MSH2 | ENST00000233146 | 0.36649 | 0.00509 | 0.06588 | 0.77693 | 0.75687 | 0.2667257683 | 0.87032 | 0.1599217505 | 0.0937361 | 2.422079836 | 0 |
| 2 | 47480831 | T | A | I | N | MSH2 | ENST00000233146 | 0.1501 | 0.00476 | 0.08284 | 0.68891 | 0.78906 | 0.280229562 | 0.84981 | 0.05102606863 | 0.0910587 | 0.375095102 | 0 |
| 2 | 47480831 | T | C | I | T | MSH2 | ENST00000233146 | 0.12203 | 0.00527 | 0.08934 | 0.66953 | 0.79395 | 0.2255233702 | 0.94118 | 0.01343616658 | 0.0141723 | -0.6741817629 | 0 |
| 2 | 47480831 | T | G | I | S | MSH2 | ENST00000233146 | 0.13854 | 0.00509 | 0.06664 | 0.64509 | 0.79697 | 0.3171967886 | 0.87113 | 0.02675579478 | 0.086789 | 0.006825568336 | 0 |
| 2 | 47480832 | C | G | I | M | MSH2 | ENST00000233146 | 0.09833 | 0.00696 | 0.06488 | 0.81467 | 0.82836 | 0.2459982544 | 0.88571 | 0.107498151 | 0.0413431 | 0.5200956762 | 0 |
| 2 | 47480833 | A | C | M | L | MSH2 | ENST00000233146 | 0.13861 | 0.0058 | 0.07786 | 0.50142 | 0.73877 | 0.2792094589 | 0.87272 | 0.06435962021 | 0.0349077 | 0.07004865112 | 0 |
| 2 | 47480833 | A | G | M | V | MSH2 | ENST00000233146 | 0.12219 | 0.00566 | 0.06119 | 0.4694 | 0.78348 | 0.3268338687 | 0.95341 | 0.070950048 | 0.0546065 | 0.03568788234 | 0 |
| 2 | 47480833 | A | T | M | L | MSH2 | ENST00000233146 | 0.13587 | 0.0058 | 0.07786 | 0.50142 | 0.73877 | 0.2792094589 | 0.87218 | 0.06435962021 | 0.034907 | 0.07004865112 | 0 |
| 2 | 47480834 | T | A | M | K | MSH2 | ENST00000233146 | 0.12438 | 0.00497 | 0.10819 | 0.72503 | 0.78906 | 0.4816963535 | 0.89561 | 0.02247961179 | 0.0393228 | -0.3611045967 | 0 |
| 2 | 47480834 | T | C | M | T | MSH2 | ENST00000233146 | 0.12668 | 0.00543 | 0.12739 | 0.70539 | 0.72775 | 0.3256515565 | 0.7115 | 0.02383623007 | 0.0114997 | -0.377806223 | 0 |
| 2 | 47480834 | T | G | M | R | MSH2 | ENST00000233146 | 0.15763 | 0.005 | 0.09836 | 0.74249 | 0.7337 | 0.5334742174 | 0.78702 | 0.02078699879 | 0.0266734 | 0.2739478579 | 0 |
| 2 | 47480835 | G | A | M | I | MSH2 | ENST00000233146 | 0.23665 | 0.00551 | 0.09232 | 0.62395 | 0.75211 | 0.2903218161 | 0.89353 | 0.05503798522 | 0.0175403 | 0.2702586212 | 0 |
| 2 | 47480835 | G | C | M | I | MSH2 | ENST00000233146 | 0.2349 | 0.00551 | 0.09232 | 0.62471 | 0.75211 | 0.2903218161 | 0.89751 | 0.05784445366 | 0.0175793 | 0.2702586212 | 0 |
| 2 | 47480835 | G | T | M | I | MSH2 | ENST00000233146 | 0.23148 | 0.00551 | 0.09232 | 0.62471 | 0.75211 | 0.2903218161 | 0.89832 | 0.05810600828 | 0.0175877 | 0.2702586212 | 0 |
| 2 | 47480836 | G | A | E | K | MSH2 | ENST00000233146 | 0.63989 | 0.00718 | 0.33037 | 0.74266 | 0.85894 | 0.4652301761 | 0.94981 | 0.8359869123 | 0.2745 | 3.748387743 | 0 |
| 2 | 47480836 | G | C | E | Q | MSH2 | ENST00000233146 | 0.69485 | 0.00586 | 0.24804 | 0.63584 | 0.78286 | 0.361353528 | 0.96032 | 0.8361097574 | 0.233667 | 4.143095136 | 0 |
| 2 | 47480837 | A | C | E | A | MSH2 | ENST00000233146 | 0.59857 | 0.00574 | 0.25692 | 0.64984 | 0.76421 | 0.4227249831 | 0.92068 | 0.6831371784 | 0.22158 | 2.852660292 | 0 |
| 2 | 47480837 | A | G | E | G | MSH2 | ENST00000233146 | 0.51257 | 0.00543 | 0.22621 | 0.6302 | 0.7337 | 0.4269646853 | 0.86274 | 0.7843002677 | 0.132397 | 2.174791305 | 0 |
| 2 | 47480837 | A | T | E | V | MSH2 | ENST00000233146 | 0.65148 | 0.00816 | 0.23501 | 0.73637 | 0.83116 | 0.4805611245 | 0.96342 | 0.9234418869 | 0.304254 | 4.069226839 | 0 |
| 2 | 47480838 | A | C | E | D | MSH2 | ENST00000233146 | 0.33607 | 0.00603 | 0.25243 | 0.68711 | 0.82836 | 0.3530516185 | 0.9537 | 0.1116852552 | 0.175453 | 3.747337369 | 0 |
| 2 | 47480838 | A | T | E | D | MSH2 | ENST00000233146 | 0.33158 | 0.00603 | 0.25243 | 0.72773 | 0.82836 | 0.3530516185 | 0.95174 | 0.1135974125 | 0.175461 | 3.747337369 | 0 |
| 2 | 47480839 | C | A | P | T | MSH2 | ENST00000233146 | 0.40523 | 0.00556 | 0.29986 | 0.77247 | 0.84641 | 0.388058139 | 0.86065 | 0.7679955959 | 0.259719 | 4.933256092 | 0 |
| 2 | 47480839 | C | G | P | A | MSH2 | ENST00000233146 | 0.39387 | 0.0058 | 0.25274 | 0.70741 | 0.8233 | 0.3175575622 | 0.74898 | 0.6614386439 | 0.163615 | 4.690603397 | 0 |
| 2 | 47480839 | C | T | P | S | MSH2 | ENST00000233146 | 0.43738 | 0.0058 | 0.26542 | 0.76635 | 0.81876 | 0.4271852146 | 0.82314 | 0.4991802275 | 0.176266 | 2.884202471 | 0 |
| 2 | 47480840 | C | A | P | Q | MSH2 | ENST00000233146 | 0.51902 | 0.0099 | 0.2723 | 0.59286 | 0.81933 | 0.3834413446 | 0.86434 | 0.8337942362 | 0.271367 | 4.818088204 | 0 |
| 2 | 47480840 | C | G | P | R | MSH2 | ENST00000233146 | 0.52134 | 0.00696 | 0.32289 | 0.665 | 0.86852 | 0.4906709504 | 0.93074 | 0.8908071518 | 0.397004 | 5.117290126 | 0 |
| 2 | 47480840 | C | T | P | L | MSH2 | ENST00000233146 | 0.4111 | 0.00654 | 0.32835 | 0.69176 | 0.82836 | 0.4913005759 | 0.88206 | 0.9008256197 | 0.27153 | 4.868969502 | 0 |
| 2 | 47480842 | G | A | A | T | MSH2 | ENST00000233146 | 0.31956 | 0.00574 | 0.06865 | 0.64539 | 0.72016 | 0.1939384836 | 0.95108 | 0.52073282 | 0.135357 | 3.298597881 | 0 |
| 2 | 47480842 | G | C | A | P | MSH2 | ENST00000233146 | 0.34027 | 0.00527 | 0.09524 | 0.59387 | 0.76289 | 0.5103674407 | 0.95202 | 0.4925591946 | 0.173691 | 2.083694727 | 0 |
| 2 | 47480842 | G | T | A | S | MSH2 | ENST00000233146 | 0.28636 | 0.00586 | 0.05563 | 0.68113 | 0.75349 | 0.2024086063 | 0.88268 | 0.3062232733 | 0.104529 | 3.648878978 | 0 |
| 2 | 47480843 | C | A | A | E | MSH2 | ENST00000233146 | 0.19892 | 0.00543 | 0.09957 | 0.68106 | 0.74164 | 0.4267279199 | 0.88485 | 0.4496029317 | 0.122448 | 2.400073574 | 0 |
| 2 | 47480843 | C | G | A | G | MSH2 | ENST00000233146 | 0.24445 | 0.00569 | 0.06465 | 0.71215 | 0.75687 | 0.2220600928 | 0.85488 | 0.6802921891 | 0.125642 | 4.302094748 | 0 |
| 2 | 47480843 | C | T | A | V | MSH2 | ENST00000233146 | 0.23969 | 0.0058 | 0.08102 | 0.58681 | 0.74936 | 0.2082369349 | 0.95046 | 0.7501828671 | 0.200043 | 4.132431831 | 0 |
| 2 | 47480845 | G | A | A | T | MSH2 | ENST00000233146 | 0.41724 | 0.0058 | 0.18904 | 0.64668 | 0.7609 | 0.3643981109 | 0.93889 | 0.5099180937 | 0.147138 | 3.979278161 | 0 |
| 2 | 47480845 | G | C | A | P | MSH2 | ENST00000233146 | 0.44546 | 0.01818 | 0.24036 | 0.68859 | 0.89376 | 0.7143968151 | 0.91382 | 0.8614560366 | 0.252231 | 4.619068156 | 0 |
| 2 | 47480845 | G | T | A | S | MSH2 | ENST00000233146 | 0.38983 | 0.00599 | 0.16442 | 0.68274 | 0.80948 | 0.3038495302 | 0.88584 | 0.3600551486 | 0.16122 | 3.483572025 | 0 |
| 2 | 47480846 | C | A | A | E | MSH2 | ENST00000233146 | 0.30106 | 0.00872 | 0.24764 | 0.68587 | 0.8153 | 0.6542518083 | 0.92997 | 0.4689026094 | 0.243457 | 3.917701679 | 0 |
| 2 | 47480846 | C | G | A | G | MSH2 | ENST00000233146 | 0.31983 | 0.00821 | 0.18016 | 0.65506 | 0.77718 | 0.3370872163 | 0.90632 | 0.5105850697 | 0.187024 | 5.265445989 | 0 |
| 2 | 47480846 | C | T | A | V | MSH2 | ENST00000233146 | 0.31028 | 0.0058 | 0.21484 | 0.64614 | 0.75143 | 0.4164933972 | 0.94968 | 0.2930160522 | 0.115304 | 2.088938309 | 0 |

| | | | | | | | | | | | | | | | | |
|---|---|---|---|---|---|---|---|---|---|---|---|---|---|---|---|---|
| 2 | 47480848 | A | C | K | Q | MSH2 | ENST00000233146 | 0.6632 | 0.03129 | 0.37696 | 0.83245 | 0.86149 | 0.4327938402 | 0.92846 | 0.9257779394 | 0.302292 | 5.496933072 | 0 |
| 2 | 47480848 | A | G | K | E | MSH2 | ENST00000233146 | 0.68891 | 0.01002 | 0.42393 | 0.82324 | 0.88165 | 0.5077077995 | 0.95881 | 0.8449257612 | 0.428586 | 5.002965938 | 0 |
| 2 | 47480849 | A | C | K | T | MSH2 | ENST00000233146 | 0.8609 | 0.03076 | 0.43012 | 0.88923 | 0.91357 | 0.4920885682 | 0.93624 | 0.9855776985 | 0.424337 | 5.210921878 | 0 |
| 2 | 47480849 | A | G | K | R | MSH2 | ENST00000233146 | 0.54209 | 0.03413 | 0.34937 | 0.81003 | 0.84482 | 0.3369378557 | 0.93512 | 0.8392524414 | 0.295224 | 5.293200355 | 0 |
| 2 | 47480849 | A | T | K | M | MSH2 | ENST00000233146 | 0.89651 | 0.03155 | 0.35587 | 0.89964 | 0.93201 | 0.5803718334 | 0.95189 | 0.983504653 | 0.439953 | 6.872553668 | 0 |
| 2 | 47480850 | G | C | K | N | MSH2 | ENST00000233146 | 0.7832 | 0.02844 | 0.45136 | 0.8828 | 0.86047 | 0.5201647467 | 0.91754 | 0.9672868848 | 0.149974 | 5.713250036 | 0 |
| 2 | 47480850 | G | T | K | N | MSH2 | ENST00000233146 | 0.77679 | 0.02844 | 0.45136 | 0.88534 | 0.86098 | 0.5201647467 | 0.91775 | 0.9553073049 | 0.149974 | 5.713250036 | 0 |
| 2 | 47480851 | A | C | K | Q | MSH2 | ENST00000233146 | 0.58347 | 0.00909 | 0.07729 | 0.78201 | 0.77141 | 0.3287047188 | 0.8978 | 0.9045447707 | 0.152745 | 3.818536611 | 0 |
| 2 | 47480851 | A | G | K | E | MSH2 | ENST00000233146 | 0.59363 | 0.00543 | 0.11667 | 0.77002 | 0.77141 | 0.3424545183 | 0.87137 | 0.8068934083 | 0.161985 | 3.114029789 | 0 |
| 2 | 47480852 | A | C | K | T | MSH2 | ENST00000233146 | 0.49081 | 0.01002 | 0.13802 | 0.83197 | 0.81356 | 0.406437719 | 0.86756 | 0.9866216183 | 0.115393 | 4.123506349 | 0 |
| 2 | 47480852 | A | G | K | R | MSH2 | ENST00000233146 | 0.27918 | 0.00608 | 0.08822 | 0.7294 | 0.69425 | 0.2199909122 | 0.74092 | 0.02617056109 | 0.0388522 | 0.1921147771 | 0 |
| 2 | 47480852 | A | T | K | M | MSH2 | ENST00000233146 | 0.66043 | 0.00574 | 0.09209 | 0.84777 | 0.83116 | 0.4632607805 | 0.89629 | 0.8206254244 | 0.161514 | 4.632743785 | 0 |
| 2 | 47480853 | G | C | K | N | MSH2 | ENST00000233146 | 0.51738 | 0.01009 | 0.13315 | 0.73921 | 0.7147 | 0.3630567158 | 0.87094 | 0.9831053615 | 0.0719725 | 3.534366896 | 0 |
| 2 | 47480853 | G | T | K | N | MSH2 | ENST00000233146 | 0.51197 | 0.01009 | 0.13315 | 0.74341 | 0.7147 | 0.3630567158 | 0.87133 | 0.9891751409 | 0.0719711 | 3.534366896 | 0 |
| 2 | 47480854 | T | A | C | S | MSH2 | ENST00000233146 | 0.3857 | 0.0054 | 0.11214 | 0.75256 | 0.8124 | 0.2719262342 | 0.93877 | 0.3778048456 | 0.186974 | 0.3510597703 | 0 |
| 2 | 47480854 | T | C | C | R | MSH2 | ENST00000233146 | 0.45242 | 0.00476 | 0.17792 | 0.51063 | 0.70914 | 0.4146566855 | 0.77412 | 0.5709781051 | 0.0654744 | -1.199611692 | 0 |
| 2 | 47480854 | T | G | C | G | MSH2 | ENST00000233146 | 0.46363 | 0.00543 | 0.09616 | 0.7913 | 0.82443 | 0.303473278 | 0.937 | 0.2981978655 | 0.19573 | 2.437333813 | 0 |
| 2 | 47480855 | G | C | C | S | MSH2 | ENST00000233146 | 0.4392 | 0.0054 | 0.11214 | 0.6949 | 0.74093 | 0.2719262342 | 0.88178 | 0.3073160218 | 0.217562 | 0.3510597703 | 0 |
| 2 | 47480855 | G | T | C | F | MSH2 | ENST00000233146 | 0.55834 | 0.005 | 0.12612 | 0.54453 | 0.76356 | 0.3369525632 | 0.87088 | 0.5347945578 | 0.233662 | 1.917871595 | 0 |
| 2 | 47480856 | C | G | C | W | MSH2 | ENST00000233146 | 0.70744 | 0.00916 | 0.15952 | 0.75041 | 0.85012 | 0.4300940014 | 0.88611 | 0.7290013834 | 0.251967 | 5.655401975 | 0 |
| 2 | 47480857 | T | A | Y | N | MSH2 | ENST00000233146 | 0.45082 | 0.005 | 0.12808 | 0.73556 | 0.79151 | 0.3577974888 | 0.96177 | 0.5133218765 | 0.184624 | 2.882818943 | 0 |
| 2 | 47480857 | T | C | Y | H | MSH2 | ENST00000233146 | 0.41736 | 0.00527 | 0.12496 | 0.58814 | 0.66064 | 0.2861561333 | 0.85439 | 0.2679453492 | 0.077026 | 3.325406923 | 0 |
| 2 | 47480857 | T | G | Y | D | MSH2 | ENST00000233146 | 0.46899 | 0.00487 | 0.11995 | 0.71161 | 0.8153 | 0.4634039166 | 0.97028 | 0.5895818472 | 0.213914 | 2.728097374 | 0 |
| 2 | 47480858 | A | C | Y | S | MSH2 | ENST00000233146 | 0.28588 | 0.00522 | 0.08495 | 0.72996 | 0.81414 | 0.3640370068 | 0.96951 | 0.08287455027 | 0.156983 | 2.703721012 | 0 |
| 2 | 47480858 | A | T | Y | F | MSH2 | ENST00000233146 | 0.21909 | 0.00586 | 0.07131 | 0.56311 | 0.66639 | 0.178732893 | 0.86243 | 0.01956883445 | 0.0243767 | 3.603148839 | 0 |
| 2 | 47480860 | C | G | L | V | MSH2 | ENST00000233146 | 0.26814 | 0.00574 | 0.08185 | 0.51742 | 0.68646 | 0.2006719686 | 0.84755 | 0.1885080242 | 0.0809092 | 1.95242662 | 0 |
| 2 | 47480861 | T | A | L | Q | MSH2 | ENST00000233146 | 0.45187 | 0.00812 | 0.11991 | 0.72647 | 0.76024 | 0.1899945144 | 0.88479 | 0.1426892378 | 0.0918662 | 2.110185159 | 0 |
| 2 | 47480861 | T | C | L | P | MSH2 | ENST00000233146 | 0.51619 | 0.01009 | 0.19424 | 0.75603 | 0.8278 | 0.6079767257 | 0.96054 | 0.4146272242 | 0.215176 | 3.178238656 | 0 |
| 2 | 47480861 | T | G | L | R | MSH2 | ENST00000233146 | 0.45113 | 0.00509 | 0.15169 | 0.45008 | 0.75349 | 0.3043784343 | 0.89318 | 0.1218475608 | 0.0787232 | 1.428936011 | 0 |
| 2 | 47480863 | G | C | E | Q | MSH2 | ENST00000233146 | 0.74539 | 0.01015 | 0.28731 | 0.74566 | 0.8356 | 0.3875316723 | 0.90098 | 0.9587672949 | 0.306154 | 3.336925922 | 0 |
| 2 | 47480864 | A | C | E | A | MSH2 | ENST00000233146 | 0.51975 | 0.00562 | 0.29792 | 0.82393 | 0.80058 | 0.4446888329 | 0.87867 | 0.86499542 | 0.313339 | 2.359998616 | 0 |
| 2 | 47480864 | A | G | E | G | MSH2 | ENST00000233146 | 0.73109 | 0.00638 | 0.26178 | 0.83444 | 0.90842 | 0.4407276774 | 0.93091 | 0.9703570008 | 0.426476 | 5.509635166 | 0 |
| 2 | 47480864 | A | T | E | V | MSH2 | ENST00000233146 | 0.55374 | 0.00599 | 0.27788 | 0.84526 | 0.88496 | 0.4707925723 | 0.95177 | 0.9664917588 | 0.418147 | 3.174091589 | 0 |
| 2 | 47480865 | A | C | E | D | MSH2 | ENST00000233146 | 0.3243 | 0.00586 | 0.29964 | 0.7065 | 0.81472 | 0.2943742047 | 0.9244 | 0.1941134185 | 0.178801 | 2.971439343 | 0 |
| 2 | 47480865 | A | T | E | D | MSH2 | ENST00000233146 | 0.31983 | 0.00586 | 0.29964 | 0.70785 | 0.81472 | 0.2943742047 | 0.91904 | 0.1941134185 | 0.178828 | 2.971439343 | 0 |
| 2 | 47480866 | A | G | R | G | MSH2 | ENST00000233146 | 0.63428 | 0.00586 | 0.21672 | 0.70884 | 0.81761 | 0.5121328184 | 0.9656 | 0.9203829765 | 0.355376 | 4.190157996 | 0 |
| 2 | 47480867 | G | A | R | K | MSH2 | ENST00000233146 | 0.35802 | 0.00569 | 0.26325 | 0.45241 | 0.69168 | 0.38947292 | 0.89051 | 0.85730654 | 0.105336 | -0.1698935434 | 0 |
| 2 | 47480867 | G | C | R | T | MSH2 | ENST00000233146 | 0.45481 | 0.00556 | 0.25715 | 0.69862 | 0.82103 | 0.4559774456 | 0.9724 | 0.8128324151 | 0.280576 | 3.727661125 | 0 |
| 2 | 47480867 | G | T | R | I | MSH2 | ENST00000233146 | 0.50283 | 0.00487 | 0.2497 | 0.70645 | 0.85586 | 0.4210131334 | 0.95222 | 0.8534117341 | 0.274669 | 3.141911369 | 0 |
| 2 | 47480868 | A | C | R | S | MSH2 | ENST00000233146 | 0.47363 | 0.00551 | 0.22435 | 0.72343 | 0.78597 | 0.4887397832 | 0.962 | 0.8273588419 | 0.302605 | 3.226087333 | 0 |
| 2 | 47480868 | A | T | R | S | MSH2 | ENST00000233146 | 0.46836 | 0.00551 | 0.22435 | 0.76006 | 0.79577 | 0.4887397832 | 0.96263 | 0.8273588419 | 0.30453 | 3.226087333 | 0 |
| 2 | 47480869 | G | A | E | K | MSH2 | ENST00000233146 | 0.6132 | 0.00562 | 0.29332 | 0.64018 | 0.7675 | 0.3571595044 | 0.94011 | 0.5220869456 | 0.25809 | 2.725491053 | 0 |
| 2 | 47480869 | G | C | E | Q | MSH2 | ENST00000233146 | 0.50883 | 0.0058 | 0.2012 | 0.66184 | 0.69594 | 0.2651246101 | 0.88358 | 0.3554863296 | 0.125592 | 0.7529199948 | 0 |
| 2 | 47480870 | A | C | E | A | MSH2 | ENST00000233146 | 0.5316 | 0.00586 | 0.20999 | 0.74151 | 0.81124 | 0.3538504074 | 0.8967 | 0.8990316391 | 0.284208 | 3.900281231 | 0 |
| 2 | 47480870 | A | G | E | G | MSH2 | ENST00000233146 | 0.72083 | 0.00696 | 0.17799 | 0.82742 | 0.89376 | 0.3680137511 | 0.92434 | 0.964147985 | 0.336133 | 4.242349653 | 0 |
| 2 | 47480870 | A | T | E | V | MSH2 | ENST00000233146 | 0.57956 | 0.0058 | 0.18818 | 0.85424 | 0.9182 | 0.3933486675 | 0.94758 | 0.9661477804 | 0.38621 | 4.666673581 | 0 |
| 2 | 47482779 | C | A | Q | K | MSH2 | ENST00000233146 | 0.44655 | 0.00535 | 0.13943 | 0.85581 | 0.79334 | 0.2857218637 | 0.93657 | 0.6463161111 | 0.21045 | 2.1694689 | 0 |
| 2 | 47482779 | C | G | Q | E | MSH2 | ENST00000233146 | 0.30925 | 0.00556 | 0.10878 | 0.66026 | 0.63947 | 0.1576463958 | 0.76116 | 0.2617073357 | 0.0874887 | -1.350584394 | 0 |
| 2 | 47482780 | A | C | Q | P | MSH2 | ENST00000233146 | 0.58304 | 0.00592 | 0.12441 | 0.8825 | 0.87925 | 0.59600036 | 0.89845 | 0.9487865567 | 0.256139 | 2.306974951 | 0 |
| 2 | 47482780 | A | G | Q | R | MSH2 | ENST00000233146 | 0.558 | 0.00556 | 0.13189 | 0.86686 | 0.8199 | 0.2354654374 | 0.90645 | 0.8589084744 | 0.216218 | 3.099766255 | 0 |
| 2 | 47482780 | A | T | Q | L | MSH2 | ENST00000233146 | 0.47867 | 0.00574 | 0.10053 | 0.80875 | 0.81356 | 0.3109360357 | 0.94769 | 0.92408216 | 0.228524 | 1.21754647 | 0 |
| 2 | 47482781 | A | C | Q | H | MSH2 | ENST00000233146 | 0.69957 | 0.00715 | 0.1329 | 0.92717 | 0.88496 | 0.2563088207 | 0.85606 | 0.8627454042 | 0.232928 | 4.252629613 | 0 |
| 2 | 47482781 | A | T | Q | H | MSH2 | ENST00000233146 | 0.69275 | 0.00715 | 0.1329 | 0.9276 | 0.88496 | 0.2563088207 | 0.85619 | 0.8627454042 | 0.232926 | 4.252629613 | 0 |
| 2 | 47482782 | G | A | G | S | MSH2 | ENST00000233146 | 0.75997 | 0.02126 | 0.41682 | 0.90527 | 0.95608 | 0.6655715435 | 0.97666 | 0.9933080673 | 0.409461 | 5.670127672 | 0 |
| 2 | 47482782 | G | C | G | R | MSH2 | ENST00000233146 | 0.92108 | 0.03243 | 0.52635 | 0.94573 | 0.95972 | 0.8147299419 | 0.98731 | 0.9924306273 | 0.462775 | 6.494688325 | 0 |
| 2 | 47482782 | G | T | G | C | MSH2 | ENST00000233146 | 0.92663 | 0.0357 | 0.46564 | 0.93956 | 0.96553 | 0.6982704655 | 0.97981 | 0.9988510609 | 0.470782 | 7.862338326 | 0 |
| 2 | 47482783 | G | A | G | D | MSH2 | ENST00000233146 | 0.81998 | 0.0318 | 0.54487 | 0.92497 | 0.94618 | 0.8142680752 | 0.97251 | 0.9978906512 | 0.472567 | 5.965749938 | 0 |
| 2 | 47482783 | G | C | G | A | MSH2 | ENST00000233146 | 0.60061 | 0.01628 | 0.41453 | 0.83908 | 0.90842 | 0.6654605214 | 0.93777 | 0.9931750894 | 0.377595 | 5.452758212 | 0 |
| 2 | 47482783 | G | T | G | V | MSH2 | ENST00000233146 | 0.81842 | 0.03189 | 0.42968 | 0.93266 | 0.94205 | 0.7469371773 | 0.97998 | 0.9975569248 | 0.47355 | 5.495975152 | 0 |
| 2 | 47482785 | G | A | E | K | MSH2 | ENST00000233146 | 0.77219 | 0.02315 | 0.41146 | 0.90391 | 0.90972 | 0.4997078888 | 0.88913 | 0.974016726 | 0.280497 | 5.084024896 | 0 |
| 2 | 47482785 | G | C | E | Q | MSH2 | ENST00000233146 | 0.87821 | 0.0118 | 0.3044 | 0.83938 | 0.88916 | 0.3767958152 | 0.91846 | 0.941397965 | 0.270252 | 4.940700377 | 0 |
| 2 | 47482786 | A | C | E | A | MSH2 | ENST00000233146 | 0.74698 | 0.01459 | 0.31121 | 0.91373 | 0.92601 | 0.4669621316 | 0.86605 | 0.9763981104 | 0.27766 | 5.076412473 | 0 |
| 2 | 47482786 | A | G | E | G | MSH2 | ENST00000233146 | 0.91213 | 0.02719 | 0.27614 | 0.92494 | 0.94356 | 0.4958003302 | 0.87145 | 0.9948439598 | 0.324536 | 5.800386336 | 0 |
| 2 | 47482786 | A | T | E | V | MSH2 | ENST00000233146 | 0.894 | 0.00877 | 0.28703 | 0.91802 | 0.94431 | 0.4882002753 | 0.9244 | 0.9870826006 | 0.348578 | 5.079683197 | 0 |
| 2 | 47482787 | A | C | E | D | MSH2 | ENST00000233146 | 0.37833 | 0.00674 | 0.31697 | 0.84164 | 0.89147 | 0.3764151149 | 0.92699 | 0.6480677724 | 0.221098 | 4.502441912 | 0 |
| 2 | 47482787 | A | T | E | D | MSH2 | ENST00000233146 | 0.37352 | 0.00674 | 0.31697 | 0.84302 | 0.89147 | 0.3764151149 | 0.92765 | 0.6480677724 | 0.221088 | 4.502441912 | 0 |
| 2 | 47482788 | A | C | K | Q | MSH2 | ENST00000233146 | 0.47652 | 0.00596 | 0.2325 | 0.76254 | 0.7675 | 0.2516416657 | 0.93317 | 0.6942853928 | 0.166374 | 2.466335996 | 0 |
| 2 | 47482788 | A | G | K | E | MSH2 | ENST00000233146 | 0.46295 | 0.00522 | 0.26398 | 0.78971 | 0.76946 | 0.2819353112 | 0.95825 | 0.6874234676 | 0.238489 | 1.783521308 | 0 |
| 2 | 47482789 | A | C | K | T | MSH2 | ENST00000233146 | 0.48621 | 0.00551 | 0.27932 | 0.8079 | 0.78535 | 0.3351322252 | 0.91359 | 0.2583366772 | 0.146561 | 3.525582237 | 0 |
| 2 | 47482789 | A | G | K | R | MSH2 | ENST00000233146 | 0.4871 | 0.00608 | 0.21047 | 0.7697 | 0.71783 | 0.200369598 | 0.96233 | 0.7096261978 | 0.174213 | 3.042073577 | 0 |
| 2 | 47482789 | A | T | K | I | MSH2 | ENST00000233146 | 0.56831 | 0.00763 | 0.25489 | 0.86527 | 0.88496 | 0.4306855499 | 0.91554 | 0.9149051309 | 0.225511 | 4.104458862 | 0 |
| 2 | 47482790 | A | C | K | N | MSH2 | ENST00000233146 | 0.41075 | 0.00556 | 0.28719 | 0.82419 | 0.74797 | 0.3095304675 | 0.91373 | 0.4715281427 | 0.109815 | 3.180443133 | 0 |
| 2 | 47482790 | A | T | K | N | MSH2 | ENST00000233146 | 0.40582 | 0.00556 | 0.28719 | 0.82532 | 0.74797 | 0.3095304675 | 0.91369 | 0.4715281427 | 0.109814 | 3.180443133 | 0 |
| 2 | 47482791 | A | C | I | L | MSH2 | ENST00000233146 | 0.473 | 0.0058 | 0.53314 | 0.71871 | 0.72926 | 0.2846660798 | 0.86215 | 0.6958315372 | 0.127774 | 2.661917541 | 0 |
| 2 | 47482791 | A | G | I | V | MSH2 | ENST00000233146 | 0.62339 | 0.00965 | 0.44612 | 0.76084 | 0.82668 | 0.186271956 | 0.89433 | 0.8032169938 | 0.197499 | 5.112659294 | 0 |
| 2 | 47482791 | A | T | I | F | MSH2 | ENST00000233146 | 0.70984 | 0.01741 | 0.54133 | 0.87638 | 0.90318 | 0.4157076771 | 0.87931 | 0.9752946496 | 0.224849 | 6.855325926 | 0 |
| 2 | 47482792 | T | A | I | N | MSH2 | ENST00000233146 | 0.81024 | 0.02023 | 0.62137 | 0.95426 | 0.94913 | 0.5995510836 | 0.87334 | 0.9970548153 | 0.296036 | 5.010089903 | 0 |
| 2 | 47482792 | T | C | I | T | MSH2 | ENST00000233146 | 0.59747 | 0.01823 | 0.64366 | 0.92706 | 0.93358 | 0.3520789979 | 0.91896 | 0.9829477668 | 0.318369 | 4.728430227 | 0 |
| 2 | 47482792 | T | G | I | S | MSH2 | ENST00000233146 | 0.7897 | 0.01346 | 0.55283 | 0.93061 | 0.94691 | 0.6076149056 | 0.94436 | 0.995680213 | 0.39102 | 5.003717031 | 0 |
| 2 | 47482793 | T | G | I | M | MSH2 | ENST00000233146 | 0.72676 | 0.02049 | 0.49391 | 0.73141 | 0.84321 | 0.2970079701 | 0.85433 | 0.3238017529 | 0.00659112 | 6.107234089 | 0 |
| 2 | 47482794 | A | C | I | L | MSH2 | ENST00000233146 | 0.45702 | 0.00603 | 0.49199 | 0.76871 | 0.81414 | 0.4183947089 | 0.8038 | 0.753352046 | 0.177932 | 4.973039467 | 0 |
| 2 | 47482794 | A | G | I | V | MSH2 | ENST00000233146 | 0.43166 | 0.00638 | 0.41193 | 0.69087 | 0.79818 | 0.2260230384 | 0.79325 | 0.8574028015 | 0.158465 | 4.43925265 | 0 |
| 2 | 47482794 | A | T | I | F | MSH2 | ENST00000233146 | 0.62203 | 0.02083 | 0.51203 | 0.88945 | 0.93747 | 0.6072526723 | 0.92736 | 0.9718040824 | 0.338647 | 6.68209178 | 0 |
| 2 | 47482795 | T | A | I | N | MSH2 | ENST00000233146 | 0.91839 | 0.0245 | 0.59425 | 0.95842 | 0.967 | 0.7353513444 | 0.92474 | 0.995803535 | 0.386507 | 4.876424717 | 0 |
| 2 | 47482795 | T | C | I | T | MSH2 | ENST00000233146 | 0.8761 | 0.01568 | 0.61649 | 0.91679 | 0.96044 | 0.5144038273 | 0.95295 | 0.9867622256 | 0.437849 | 4.963755043 | 0 |
| 2 | 47482795 | T | G | I | S | MSH2 | ENST00000233146 | 0.92194 | 0.01731 | 0.51354 | 0.94971 | 0.97698 | 0.729041181 | 0.97992 | 0.9974210262 | 0.457825 | 5.017119952 | 0 |
| 2 | 47482796 | T | G | I | M | MSH2 | ENST00000233146 | 0.64508 | 0.01172 | 0.45806 | 0.86165 | 0.89147 | 0.4699554587 | 0.85858 | 0.7266754508 | 0.231726 | 6.107586881 | 0 |
| 2 | 47482797 | C | A | Q | K | MSH2 | ENST00000233146 | 0.37289 | 0.00532 | 0.15068 | 0.75959 | 0.74021 | 0.3131044365 | 0.92771 | 0.7442184091 | 0.202123 | 1.449990396 | 0 |
| 2 | 47482797 | C | G | Q | E | MSH2 | ENST00000233146 | 0.33866 | 0.00556 | 0.10792 | 0.71325 | 0.73516 | 0.232916653 | 0.90201 | 0.6902896762 | 0.131525 | 1.698062163 | 0 |
| 2 | 47482798 | A | C | Q | P | MSH2 | ENST00000233146 | 0.70744 | 0.00763 | 0.11438 | 0.83009 | 0.92233 | 0.6565470541 | 0.98089 | 0.9259892702 | 0.345761 | 4.566603984 | 0 |
| 2 | 47482798 | A | G | Q | R | MSH2 | ENST00000233146 | 0.52566 | 0.00556 | 0.1471 | 0.72813 | 0.73296 | 0.2842510346 | 0.96049 | 0.8185175061 | 0.166286 | 2.152709481 | 0 |
| 2 | 47482798 | A | T | Q | L | MSH2 | ENST00000233146 | 0.58183 | 0.00574 | 0.09732 | 0.74592 | 0.80118 | 0.3525904817 | 0.95046 | 0.907019794 | 0.122011 | 4.25666338 | 0 |
| 2 | 47482800 | G | A | E | K | MSH2 | ENST00000233146 | 0.52354 | 0.00551 | 0.35697 | 0.79079 | 0.77972 | 0.2402950907 | 0.959 | 0.7880734801 | 0.176437 | 1.624891817 | 0 |
| 2 | 47482800 | G | C | E | Q | MSH2 | ENST00000233146 | 0.46905 | 0.00654 | 0.25408 | 0.74226 | 0.7872 | 0.2117730061 | 0.95905 | 0.3383261263 | 0.0895727 | 2.857728179 | 0 |
| 2 | 47482801 | A | C | E | A | MSH2 | ENST00000233146 | 0.38415 | 0.00556 | 0.2627 | 0.79023 | 0.77335 | 0.274438484 | 0.94128 | 0.755028069 | 0.154867 | 2.445812474 | 0 |
| 2 | 47482801 | A | G | E | G | MSH2 | ENST00000233146 | 0.43196 | 0.00543 | 0.23345 | 0.81587 | 0.8345 | 0.2552080882 | 0.98754 | 0.866979003 | 0.31309 | 2.176038545 | 0 |
| 2 | 47482801 | A | T | E | V | MSH2 | ENST00000233146 | 0.52705 | 0.0058 | 0.2379 | 0.86244 | 0.8543 | 0.3220545623 | 0.96987 | 0.9682307839 | 0.24512 | 4.043451239 | 0 |
| 2 | 47482802 | G | C | E | D | MSH2 | ENST00000233146 | 0.25049 | 0.00586 | 0.26408 | 0.45666 | 0.79938 | 0.1717995214 | 0.94001 | 0.04664817705 | 0.0568635 | -0.4403949028 | 0 |
| 2 | 47482802 | G | T | E | D | MSH2 | ENST00000233146 | 0.2469 | 0.00586 | 0.26408 | 0.46369 | 0.79998 | 0.1717995214 | 0.94002 | 0.06149888411 | 0.0568544 | -0.4403949028 | 0 |
| 2 | 47482803 | T | A | F | I | MSH2 | ENST00000233146 | 0.85456 | 0.02732 | 0.57742 | 0.87949 | 0.90668 | 0.6119877048 | 0.87173 | 0.9877361655 | 0.287083 | 3.901933716 | 0 |
| 2 | 47482803 | T | C | F | L | MSH2 | ENST00000233146 | 0.56221 | 0.00791 | 0.60923 | 0.82309 | 0.8345 | 0.5574185072 | 0.80679 | 0.9563437104 | 0.207624 | 3.908970862 | 0 |

| | | | | | | | | | | | | | | | | | | |
|---|---|---|---|---|---|---|---|---|---|---|---|---|---|---|---|---|---|---|
| 2 | 47482803 | T | G | F | V | MSH2 | ENST00000233146 | 0.85353 | 0.02809 | 0.5537 | 0.89208 | 0.92723 | 0.5280793734 | 0.89545 | 0.9865654111 | 0.370139 | 3.742835305 | 0 |
| 2 | 47482804 | T | A | F | Y | MSH2 | ENST00000233146 | 0.60156 | 0.01126 | 0.59922 | 0.77778 | 0.8705 | 0.4182711276 | 0.86008 | 0.8515325189 | 0.244357 | 7.945839849 | 0 |
| 2 | 47482804 | T | C | F | S | MSH2 | ENST00000233146 | 0.83637 | 0.02741 | 0.58351 | 0.94548 | 0.95753 | 0.682543049 | 0.9518 | 0.9949843884 | 0.43452 | 6.769453161 | 0 |
| 2 | 47482804 | T | G | F | C | MSH2 | ENST00000233146 | 0.83767 | 0.03106 | 0.60894 | 0.94385 | 0.95462 | 0.6025969196 | 0.93475 | 0.9974210262 | 0.434905 | 7.17386277 | 0 |
| 2 | 47482805 | C | A | F | L | MSH2 | ENST00000233146 | 0.33222 | 0.00791 | 0.60923 | 0.79514 | 0.81356 | 0.5574185072 | 0.82803 | 0.5406110287 | 0.151697 | 3.908970862 | 0 |
| 2 | 47482805 | C | G | F | L | MSH2 | ENST00000233146 | 0.3379 | 0.00791 | 0.60923 | 0.79507 | 0.81414 | 0.5574185072 | 0.82824 | 0.08988741039 | 0.0253218 | 3.908970862 | 0 |
| 2 | 47482806 | C | G | L | V | MSH2 | ENST00000233146 | 0.35036 | 0.00889 | 0.37223 | 0.71278 | 0.78782 | 0.4591186736 | 0.91154 | 0.4380898774 | 0.183673 | 3.241177964 | 0 |
| 2 | 47482807 | T | A | L | Q | MSH2 | ENST00000233146 | 0.76036 | 0.03547 | 0.45471 | 0.9364 | 0.96444 | 0.6722249086 | 0.9713 | 0.9972634315 | 0.465038 | 6.131182013 | 0 |
| 2 | 47482807 | T | C | L | P | MSH2 | ENST00000233146 | 0.77593 | 0.0292 | 0.56693 | 0.93806 | 0.96956 | 0.8615492006 | 0.98363 | 0.9966283441 | 0.473145 | 8.527099296 | 0 |
| 2 | 47482807 | T | G | L | R | MSH2 | ENST00000233146 | 0.76737 | 0.03059 | 0.52799 | 0.93597 | 0.96919 | 0.8233914819 | 0.97897 | 0.9978142977 | 0.475319 | 5.29348625 | 0 |
| 2 | 47482809 | T | A | S | T | MSH2 | ENST00000233146 | 0.17881 | 0.00586 | 0.14932 | 0.62195 | 0.62101 | 0.1371011554 | 0.87628 | 0.02241795696 | 0.0562383 | 2.944620046 | 0 |
| 2 | 47482809 | T | C | S | P | MSH2 | ENST00000233146 | 0.25498 | 0.00592 | 0.16384 | 0.73353 | 0.85689 | 0.5707170713 | 0.96816 | 0.07595074922 | 0.18081 | 6.189495477 | 0 |
| 2 | 47482809 | T | G | S | A | MSH2 | ENST00000233146 | 0.18346 | 0.00603 | 0.13558 | 0.5346 | 0.5108 | 0.1146863869 | 0.7799 | 0.02727485262 | 0.0171184 | 2.55514464 | 0 |
| 2 | 47482810 | C | A | S | Y | MSH2 | ENST00000233146 | 0.22665 | 0.00527 | 0.20242 | 0.69409 | 0.66922 | 0.2791056528 | 0.92717 | 0.1580034941 | 0.0945124 | 7.032555581 | 0 |
| 2 | 47482810 | C | T | S | F | MSH2 | ENST00000233146 | 0.25875 | 0.00556 | 0.1798 | 0.77216 | 0.72926 | 0.2200397168 | 0.92962 | 0.123241514 | 0.107803 | 5.785448283 | 0 |
| 2 | 47482812 | A | C | K | Q | MSH2 | ENST00000233146 | 0.53745 | 0.0058 | 0.26627 | 0.69984 | 0.80889 | 0.3152854516 | 0.93382 | 0.749045074 | 0.177448 | 2.273597064 | 0 |
| 2 | 47482812 | A | G | K | E | MSH2 | ENST00000233146 | 0.57055 | 0.00527 | 0.31405 | 0.81081 | 0.87442 | 0.3752762779 | 0.97189 | 0.8668890595 | 0.29244 | 2.388384092 | 0 |
| 2 | 47482813 | A | C | K | T | MSH2 | ENST00000233146 | 0.8044 | 0.00855 | 0.30748 | 0.87158 | 0.89873 | 0.409946551 | 0.90867 | 0.9779499173 | 0.269538 | 3.635391257 | 0 |
| 2 | 47482813 | A | G | K | R | MSH2 | ENST00000233146 | 0.49068 | 0.00615 | 0.24001 | 0.73987 | 0.7562 | 0.2625322955 | 0.96061 | 0.8719171882 | 0.193439 | 2.994985481 | 0 |
| 2 | 47482813 | A | T | K | M | MSH2 | ENST00000233146 | 0.85894 | 0.01977 | 0.24824 | 0.91835 | 0.92844 | 0.5035904943 | 0.8962 | 0.9865216613 | 0.293801 | 5.01273861 | 0 |
| 2 | 47482814 | G | C | K | N | MSH2 | ENST00000233146 | 0.7657 | 0.00592 | 0.34019 | 0.76807 | 0.84321 | 0.4175595232 | 0.87673 | 0.9384729266 | 0.156235 | 3.26446093 | 0 |
| 2 | 47482814 | G | T | K | N | MSH2 | ENST00000233146 | 0.75925 | 0.00592 | 0.34019 | 0.76712 | 0.84321 | 0.4175595232 | 0.87687 | 0.9384729266 | 0.15623 | 3.26446093 | 0 |
| 2 | 47482815 | G | A | V | M | MSH2 | ENST00000233146 | 0.69516 | 0.01505 | 0.50394 | 0.86333 | 0.89467 | 0.4133676203 | 0.89672 | 0.9617182016 | 0.246153 | 5.659318035 | 0 |
| 2 | 47482815 | G | C | V | L | MSH2 | ENST00000233146 | 0.50179 | 0.0058 | 0.47375 | 0.80867 | 0.77527 | 0.4007085693 | 0.93582 | 0.851841867 | 0.244106 | 3.387868929 | 0 |
| 2 | 47482815 | G | T | V | L | MSH2 | ENST00000233146 | 0.49654 | 0.0058 | 0.47375 | 0.80835 | 0.77527 | 0.4007085693 | 0.93628 | 0.851841867 | 0.244106 | 3.387868929 | 0 |
| 2 | 47482816 | T | A | V | E | MSH2 | ENST00000233146 | 0.64571 | 0.01108 | 0.48182 | 0.90805 | 0.93083 | 0.6790652982 | 0.95133 | 0.9691332579 | 0.39237 | 6.044788541 | 0 |
| 2 | 47482816 | T | C | V | A | MSH2 | ENST00000233146 | 0.44181 | 0.00543 | 0.49606 | 0.75725 | 0.76157 | 0.3987815058 | 0.87049 | 0.7382870317 | 0.207558 | 4.29864191 | 0 |
| 2 | 47482816 | T | G | V | G | MSH2 | ENST00000233146 | 0.62385 | 0.00788 | 0.37365 | 0.88078 | 0.90798 | 0.5727709522 | 0.9748 | 0.904196322 | 0.309113 | 7.028532146 | 0 |
| 2 | 47482818 | A | C | K | Q | MSH2 | ENST00000233146 | 0.84545 | 0.02556 | 0.53935 | 0.89062 | 0.93162 | 0.4477056121 | 0.9576 | 0.946401 | 0.312673 | 4.441473278 | 0 |
| 2 | 47482818 | A | G | K | E | MSH2 | ENST00000233146 | 0.83151 | 0.00889 | 0.58503 | 0.88993 | 0.92682 | 0.5430587335 | 0.98221 | 0.9273487926 | 0.442023 | 3.537158444 | 0 |
| 2 | 47482819 | A | C | K | T | MSH2 | ENST00000233146 | 0.83506 | 0.01953 | 0.59366 | 0.92078 | 0.94655 | 0.5151132559 | 0.97562 | 0.9870969057 | 0.442905 | 4.895102815 | 0 |
| 2 | 47482819 | A | G | K | R | MSH2 | ENST00000233146 | 0.51034 | 0.00664 | 0.51238 | 0.79021 | 0.85534 | 0.3428411677 | 0.98321 | 0.8700868487 | 0.307863 | 2.573464252 | 0 |
| 2 | 47482819 | A | T | K | I | MSH2 | ENST00000233146 | 0.89605 | 0.02403 | 0.55522 | 0.94825 | 0.9781 | 0.5763645587 | 0.97515 | 0.9967081547 | 0.444222 | 6.296008668 | 0 |
| 2 | 47482820 | A | C | K | N | MSH2 | ENST00000233146 | 0.60083 | 0.00769 | 0.61041 | 0.86348 | 0.78348 | 0.5240005858 | 0.89773 | 0.9344509244 | 0.13866 | 3.645956322 | 0 |
| 2 | 47482820 | A | T | K | N | MSH2 | ENST00000233146 | 0.59465 | 0.00769 | 0.61041 | 0.86348 | 0.78348 | 0.5240005858 | 0.89765 | 0.9344509244 | 0.138657 | 3.645956322 | 0 |
| 2 | 47482821 | C | A | Q | K | MSH2 | ENST00000233146 | 0.33613 | 0.00532 | 0.18397 | 0.70711 | 0.73734 | 0.2000073097 | 0.95036 | 0.6231957674 | 0.188306 | 0.5609120612 | 0 |
| 2 | 47482821 | C | G | Q | E | MSH2 | ENST00000233146 | 0.30463 | 0.00556 | 0.15629 | 0.60309 | 0.65772 | 0.1866196883 | 0.82656 | 0.4512955546 | 0.076224 | 0.9621319436 | 0 |
| 2 | 47482822 | A | C | Q | P | MSH2 | ENST00000233146 | 0.43914 | 0.00535 | 0.15963 | 0.76409 | 0.77076 | 0.505967306 | 0.98271 | 0.7283630967 | 0.258593 | 5.991382718 | 0 |
| 2 | 47482822 | A | G | Q | R | MSH2 | ENST00000233146 | 0.38909 | 0.00556 | 0.17691 | 0.70966 | 0.72926 | 0.1780665952 | 0.98073 | 0.6018841267 | 0.224717 | 3.001314165 | 0 |
| 2 | 47482822 | A | T | Q | L | MSH2 | ENST00000233146 | 0.45346 | 0.00574 | 0.14371 | 0.83139 | 0.80889 | 0.2632517907 | 0.9528 | 0.7765287161 | 0.218149 | 2.546576959 | 0 |
| 2 | 47482823 | A | C | Q | H | MSH2 | ENST00000233146 | 0.34065 | 0.01177 | 0.17595 | 0.83482 | 0.84906 | 0.2779512683 | 0.96416 | 0.5455995202 | 0.2053 | 3.872544492 | 0 |
| 2 | 47482823 | A | T | Q | H | MSH2 | ENST00000233146 | 0.33613 | 0.01177 | 0.17595 | 0.81743 | 0.84906 | 0.2779512683 | 0.96424 | 0.5455995202 | 0.205298 | 3.872544492 | 0 |
| 2 | 47482824 | A | C | M | L | MSH2 | ENST00000233146 | 0.25862 | 0.0058 | 0.12968 | 0.59263 | 0.75755 | 0.1862674941 | 0.73121 | 0.2457869351 | 0.0498823 | -1.56302815 | 0 |
| 2 | 47482824 | A | G | M | V | MSH2 | ENST00000233146 | 0.29472 | 0.00566 | 0.10512 | 0.51617 | 0.72926 | 0.2010641815 | 0.72634 | 0.3216216862 | 0.0433585 | 0.448056734 | 0 |
| 2 | 47482824 | A | T | M | L | MSH2 | ENST00000233146 | 0.25475 | 0.0058 | 0.12968 | 0.59263 | 0.75755 | 0.1862674941 | 0.72946 | 0.2457869351 | 0.0498774 | -1.56302815 | 0 |
| 2 | 47482825 | T | A | M | K | MSH2 | ENST00000233146 | 0.70919 | 0.00509 | 0.16461 | 0.76812 | 0.77591 | 0.5510962666 | 0.88316 | 0.8256617785 | 0.183482 | 2.761803493 | 0 |
| 2 | 47482825 | T | C | M | T | MSH2 | ENST00000233146 | 0.66051 | 0.00556 | 0.19727 | 0.74066 | 0.77399 | 0.3713256987 | 0.90592 | 0.5708243018 | 0.184954 | 1.889241418 | 0 |
| 2 | 47482825 | T | G | M | R | MSH2 | ENST00000233146 | 0.72973 | 0.00522 | 0.15847 | 0.74962 | 0.77972 | 0.5618454541 | 0.85236 | 0.8186761737 | 0.164885 | 5.164785659 | 0 |
| 2 | 47482826 | G | A | M | I | MSH2 | ENST00000233146 | 0.41081 | 0.00551 | 0.12073 | 0.66533 | 0.74658 | 0.1681855062 | 0.78698 | 0.1509507447 | 0.0761878 | 0.2421538194 | 0 |
| 2 | 47482826 | G | C | M | I | MSH2 | ENST00000233146 | 0.40845 | 0.00551 | 0.12073 | 0.62615 | 0.74658 | 0.1681855062 | 0.78644 | 0.1229968891 | 0.0761906 | 0.2421538194 | 0 |
| 2 | 47482826 | G | T | M | I | MSH2 | ENST00000233146 | 0.40366 | 0.00551 | 0.12073 | 0.64915 | 0.74658 | 0.1681855062 | 0.78646 | 0.1229968891 | 0.0761822 | 0.2421538194 | 0 |
| 2 | 47482827 | C | A | P | T | MSH2 | ENST00000233146 | 0.58974 | 0.00718 | 0.45756 | 0.84547 | 0.87877 | 0.3758011885 | 0.90215 | 0.9303317666 | 0.267417 | 5.450257207 | 0 |
| 2 | 47482827 | C | G | P | A | MSH2 | ENST00000233146 | 0.41028 | 0.0058 | 0.40584 | 0.81931 | 0.81703 | 0.2966642864 | 0.77608 | 0.8184132576 | 0.139494 | 4.262611147 | 0 |
| 2 | 47482827 | C | T | P | S | MSH2 | ENST00000233146 | 0.46134 | 0.0058 | 0.41882 | 0.81881 | 0.80889 | 0.3930276784 | 0.86038 | 0.2971926715 | 0.136596 | 3.956500811 | 0 |
| 2 | 47482828 | C | A | P | H | MSH2 | ENST00000233146 | 0.68619 | 0.01637 | 0.47493 | 0.90357 | 0.911 | 0.4024145526 | 0.82253 | 0.9905044436 | 0.283509 | 6.306533727 | 0 |
| 2 | 47482828 | C | G | P | R | MSH2 | ENST00000233146 | 0.65798 | 0.00844 | 0.48715 | 0.8257 | 0.92069 | 0.3839409715 | 0.88529 | 0.9772712588 | 0.332466 | 6.850365213 | 0 |
| 2 | 47482828 | C | T | P | L | MSH2 | ENST00000233146 | 0.53448 | 0.00855 | 0.48909 | 0.8742 | 0.91526 | 0.4348446915 | 0.84408 | 0.9861400127 | 0.252392 | 7.901541058 | 0 |
| 2 | 47482830 | T | A | F | I | MSH2 | ENST00000233146 | 0.48857 | 0.00517 | 0.3309 | 0.76589 | 0.66064 | 0.2275499983 | 0.85361 | 0.5434065461 | 0.0972844 | 0.01932688953 | 0 |
| 2 | 47482830 | T | C | F | L | MSH2 | ENST00000233146 | 0.43768 | 0.00543 | 0.36213 | 0.52646 | 0.672 | 0.1935279691 | 0.78971 | 0.3343954384 | 0.0794062 | -1.138856553 | 0 |
| 2 | 47482830 | T | G | F | V | MSH2 | ENST00000233146 | 0.48152 | 0.00527 | 0.29612 | 0.75035 | 0.74164 | 0.2712542077 | 0.90618 | 0.483014524 | 0.0878542 | -0.01896912673 | 0 |
| 2 | 47482831 | T | A | F | Y | MSH2 | ENST00000233146 | 0.36314 | 0.00569 | 0.36106 | 0.807 | 0.74306 | 0.2082424571 | 0.80964 | 0.6087676883 | 0.125475 | 3.331243922 | 0 |
| 2 | 47482831 | T | C | F | S | MSH2 | ENST00000233146 | 0.49036 | 0.00487 | 0.33017 | 0.80149 | 0.77399 | 0.3495431032 | 0.9234 | 0.6441073418 | 0.232073 | 2.410277626 | 0 |
| 2 | 47482832 | T | A | F | L | MSH2 | ENST00000233146 | 0.22906 | 0.00543 | 0.36213 | 0.68427 | 0.71705 | 0.1935279691 | 0.84887 | 0.07651089877 | 0.0619511 | -1.138856553 | 0 |
| 2 | 47482832 | T | G | F | L | MSH2 | ENST00000233146 | 0.23334 | 0.00543 | 0.36213 | 0.68392 | 0.71705 | 0.1935279691 | 0.84871 | 0.07651089877 | 0.0619476 | -1.138856553 | 0 |
| 2 | 47482833 | A | C | T | P | MSH2 | ENST00000233146 | 0.27908 | 0.00527 | 0.12282 | 0.4017 | 0.80772 | 0.406413303 | 0.96911 | 0.0748025775 | 0.0793741 | 4.035762167 | 0 |
| 2 | 47482833 | A | G | T | A | MSH2 | ENST00000233146 | 0.2276 | 0.00574 | 0.11287 | 0.37549 | 0.70097 | 0.2130740659 | 0.84288 | 0.03696306435 | 0.0305782 | 1.124489729 | 0 |
| 2 | 47482833 | A | T | T | S | MSH2 | ENST00000233146 | 0.19741 | 0.00586 | 0.11949 | 0.45384 | 0.69762 | 0.1713386875 | 0.86986 | 0.04322162271 | 0.0596523 | 1.039033773 | 0 |
| 2 | 47482834 | C | A | T | N | MSH2 | ENST00000233146 | 0.22321 | 0.00547 | 0.16726 | 0.62611 | 0.60599 | 0.219053449 | 0.87805 | 0.1068031713 | 0.0479212 | 2.077157208 | 0 |
| 2 | 47482834 | C | G | T | S | MSH2 | ENST00000233146 | 0.21029 | 0.00586 | 0.11949 | 0.63702 | 0.59393 | 0.1713386875 | 0.84783 | 0.1091085076 | 0.0609744 | 1.039033773 | 0 |
| 2 | 47482834 | C | T | T | I | MSH2 | ENST00000233146 | 0.28083 | 0.00556 | 0.22215 | 0.65961 | 0.7675 | 0.2454464078 | 0.95145 | 0.1292499709 | 0.0849505 | 2.94425152 | 0 |
| 2 | 47482836 | G | C | E | Q | MSH2 | ENST00000233146 | 0.34527 | 0.0058 | 0.21198 | 0.74811 | 0.74867 | 0.2416439773 | 0.86467 | 0.3338583112 | 0.117682 | 2.256887828 | 0 |
| 2 | 47482837 | A | C | E | A | MSH2 | ENST00000233146 | 0.28759 | 0.00556 | 0.22272 | 0.76928 | 0.79151 | 0.2616383901 | 0.90824 | 0.07546383888 | 0.101651 | 2.646547687 | 0 |
| 2 | 47482837 | A | G | E | G | MSH2 | ENST00000233146 | 0.3413 | 0.00543 | 0.20556 | 0.69589 | 0.7872 | 0.2229079065 | 0.91831 | 0.4468585253 | 0.126271 | 1.763362824 | 0 |
| 2 | 47482837 | A | T | E | V | MSH2 | ENST00000233146 | 0.3413 | 0.0058 | 0.20304 | 0.77159 | 0.80889 | 0.3446102636 | 0.96447 | 0.4339003563 | 0.214545 | 3.4087922 | 0 |
| 2 | 47482838 | A | C | E | D | MSH2 | ENST00000233146 | 0.35625 | 0.00586 | 0.22394 | 0.72527 | 0.7285 | 0.1546975353 | 0.82187 | 0.05977465186 | 0.0521288 | 0.08083938711 | 0 |
| 2 | 47482838 | A | T | E | D | MSH2 | ENST00000233146 | 0.35157 | 0.00586 | 0.22394 | 0.72561 | 0.7285 | 0.1546975353 | 0.82228 | 0.06066971876 | 0.0521351 | 0.08083938711 | 0 |
| 2 | 47482839 | A | C | M | L | MSH2 | ENST00000233146 | 0.43586 | 0.0058 | 0.50096 | 0.70788 | 0.71783 | 0.3645591356 | 0.87522 | 0.6042556167 | 0.146646 | 1.76100003 | 0 |
| 2 | 47482839 | A | G | M | V | MSH2 | ENST00000233146 | 0.57716 | 0.0058 | 0.44137 | 0.74073 | 0.79089 | 0.4635072407 | 0.95289 | 0.9360664487 | 0.257736 | 4.916766994 | 0 |
| 2 | 47482839 | A | T | M | L | MSH2 | ENST00000233146 | 0.43075 | 0.0058 | 0.50096 | 0.70788 | 0.71783 | 0.3645591356 | 0.87942 | 0.6042556167 | 0.146639 | 1.76100003 | 0 |
| 2 | 47482840 | T | A | M | K | MSH2 | ENST00000233146 | 0.49932 | 0.00543 | 0.57627 | 0.86117 | 0.89467 | 0.7100219915 | 0.96897 | 0.9846748114 | 0.399002 | 5.487227431 | 0 |
| 2 | 47482840 | T | C | M | T | MSH2 | ENST00000233146 | 0.46519 | 0.00556 | 0.61747 | 0.82238 | 0.87345 | 0.5327680663 | 0.94441 | 0.9092741013 | 0.289474 | 4.544748808 | 0 |
| 2 | 47482840 | T | G | M | R | MSH2 | ENST00000233146 | 0.50244 | 0.00504 | 0.55491 | 0.79667 | 0.84052 | 0.718750266 | 0.96833 | 0.8458268642 | 0.408589 | 5.883620892 | 0 |
| 2 | 47482841 | G | A | M | I | MSH2 | ENST00000233146 | 0.66335 | 0.00586 | 0.4989 | 0.76595 | 0.75552 | 0.4388799627 | 0.90859 | 0.9705881476 | 0.206227 | 3.772033656 | 0 |
| 2 | 47482841 | G | C | M | I | MSH2 | ENST00000233146 | 0.66011 | 0.00586 | 0.4989 | 0.76571 | 0.75552 | 0.4388799627 | 0.90543 | 0.9482005835 | 0.206232 | 3.772033656 | 0 |
| 2 | 47482841 | G | T | M | I | MSH2 | ENST00000233146 | 0.65374 | 0.00586 | 0.4989 | 0.76623 | 0.75552 | 0.4388799627 | 0.90608 | 0.9674603939 | 0.206232 | 3.772033656 | 0 |
| 2 | 47482842 | T | A | S | T | MSH2 | ENST00000233146 | 0.43484 | 0.00619 | 0.45365 | 0.74331 | 0.80178 | 0.3093007391 | 0.94109 | 0.5097052455 | 0.215131 | 1.871035009 | 0 |
| 2 | 47482842 | T | C | S | P | MSH2 | ENST00000233146 | 0.52453 | 0.02494 | 0.46937 | 0.89737 | 0.88165 | 0.4873274947 | 0.9513 | 0.9684791565 | 0.368323 | 6.622066076 | 0 |
| 2 | 47482842 | T | G | S | A | MSH2 | ENST00000233146 | 0.4871 | 0.00659 | 0.43126 | 0.76082 | 0.79456 | 0.2235993639 | 0.8964 | 0.6212716103 | 0.140553 | 3.729625835 | 0 |
| 2 | 47482843 | C | T | S | L | MSH2 | ENST00000233146 | 0.61522 | 0.01731 | 0.46861 | 0.89426 | 0.89101 | 0.3881172305 | 0.90883 | 0.9765641689 | 0.228808 | 7.771135176 | 0 |
| 2 | 47482845 | G | A | E | K | MSH2 | ENST00000233146 | 0.51982 | 0.00551 | 0.43831 | 0.81816 | 0.84052 | 0.299699505 | 0.96354 | 0.8798330426 | 0.270238 | 4.0090162 | 0 |
| 2 | 47482845 | G | C | E | Q | MSH2 | ENST00000233146 | 0.6284 | 0.00747 | 0.33101 | 0.77056 | 0.88543 | 0.2549514099 | 0.92967 | 0.831574142 | 0.240787 | 2.991677366 | 0 |
| 2 | 47482846 | A | C | E | A | MSH2 | ENST00000233146 | 0.44472 | 0.00556 | 0.34166 | 0.77741 | 0.83834 | 0.2920847426 | 0.92927 | 0.8367358446 | 0.233203 | 2.92484971 | 0 |
| 2 | 47482846 | A | G | E | G | MSH2 | ENST00000233146 | 0.50589 | 0.00562 | 0.30811 | 0.82132 | 0.87296 | 0.2859682826 | 0.87475 | 0.9174721837 | 0.248726 | 5.016670667 | 0 |
| 2 | 47482846 | A | T | E | V | MSH2 | ENST00000233146 | 0.50628 | 0.00644 | 0.32682 | 0.85462 | 0.90318 | 0.3504948745 | 0.93294 | 0.9432455301 | 0.278145 | 3.654790829 | 0 |
| 2 | 47482847 | A | C | E | D | MSH2 | ENST00000233146 | 0.36214 | 0.00586 | 0.34506 | 0.68533 | 0.79456 | 0.1828174506 | 0.97427 | 0.2636418045 | 0.178266 | 1.8691586 | 0 |
| 2 | 47482847 | A | T | E | D | MSH2 | ENST00000233146 | 0.35747 | 0.00586 | 0.34506 | 0.68571 | 0.79456 | 0.1828174506 | 0.97424 | 0.2636418045 | 0.178268 | 1.8691586 | 0 |
| 2 | 47482848 | G | A | E | K | MSH2 | ENST00000233146 | 0.58382 | 0.00556 | 0.40172 | 0.76532 | 0.74376 | 0.2755878421 | 0.96625 | 0.9012296796 | 0.227665 | 4.521226864 | 0 |
| 2 | 47482848 | G | C | E | Q | MSH2 | ENST00000233146 | 0.53663 | 0.00586 | 0.32002 | 0.68729 | 0.73734 | 0.2285432755 | 0.95527 | 0.7527410388 | 0.227839 | 4.123506127 | 0 |
| 2 | 47482849 | A | C | E | A | MSH2 | ENST00000233146 | 0.44181 | 0.00627 | 0.32483 | 0.81173 | 0.81761 | 0.2979119972 | 0.94837 | 0.8323920965 | 0.21924 | 5.665794106 | 0 |
| 2 | 47482849 | A | G | E | G | MSH2 | ENST00000233146 | 0.5027 | 0.00547 | 0.30314 | 0.82661 | 0.80713 | 0.2884911086 | 0.93457 | 0.9447715878 | 0.190772 | 5.693433806 | 0 |
| 2 | 47482849 | A | T | E | V | MSH2 | ENST00000233146 | 0.47835 | 0.00556 | 0.30218 | 0.88039 | 0.88022 | 0.3605877483 | 0.97217 | 0.9668362737 | 0.260987 | 4.426990142 | 0 |

| | | | | | | | | | | | | | | | | | | |
|---|---|---|---|---|---|---|---|---|---|---|---|---|---|---|---|---|---|---|
| 2 | 47482850 | A | C | E | D | MSH2 | ENST00000233146 | 0.38329 | 0.00586 | 0.32331 | 0.68935 | 0.79151 | 0.1927289048 | 0.97816 | 0.1976639479 | 0.180082 | 2.445981806 | 0 |
| 2 | 47482850 | A | T | E | D | MSH2 | ENST00000233146 | 0.37845 | 0.00586 | 0.32331 | 0.7053 | 0.79151 | 0.1927289048 | 0.97818 | 0.2038565874 | 0.18008 | 2.445981806 | 0 |
| 2 | 47482851 | A | C | N | H | MSH2 | ENST00000233146 | 0.50576 | 0.0071 | 0.22915 | 0.7869 | 0.65575 | 0.1610528677 | 0.88319 | 0.4248191416 | 0.104174 | 1.52683123 | 0 |
| 2 | 47482851 | A | G | N | D | MSH2 | ENST00000233146 | 0.35936 | 0.00547 | 0.24848 | 0.51946 | 0.54344 | 0.09929396457 | 0.7882 | 0.6480092406 | 0.0396883 | -2.344133945 | 0 |
| 2 | 47482851 | A | T | N | Y | MSH2 | ENST00000233146 | 0.54487 | 0.00622 | 0.242 | 0.86348 | 0.79818 | 0.2088631314 | 0.94218 | 0.3361340155 | 0.0193363 | 2.821667302 | 0 |
| 2 | 47482852 | A | C | N | T | MSH2 | ENST00000233146 | 0.31167 | 0.00566 | 0.22869 | 0.66522 | 0.64157 | 0.1170038592 | 0.90805 | 0.1407314241 | 0.0761711 | 1.921419542 | 0 |
| 2 | 47482852 | A | G | N | S | MSH2 | ENST00000233146 | 0.28 | 0.00608 | 0.20067 | 0.58814 | 0.61068 | 0.08993076282 | 0.79583 | 0.1141658649 | 0.038147 | 1.172805394 | 0 |
| 2 | 47482852 | A | T | N | I | MSH2 | ENST00000233146 | 0.44844 | 0.00543 | 0.23931 | 0.78751 | 0.74797 | 0.1875101923 | 0.94201 | 0.1670459309 | 0.112718 | 2.780296012 | 0 |
| 2 | 47482853 | C | A | N | K | MSH2 | ENST00000233146 | 0.34784 | 0.00547 | 0.264 | 0.71512 | 0.55468 | 0.1331140002 | 0.84526 | 0.5537416339 | 0.0632815 | 0.4395704616 | 0 |
| 2 | 47482853 | C | G | N | K | MSH2 | ENST00000233146 | 0.35366 | 0.00547 | 0.264 | 0.71381 | 0.55746 | 0.1331140002 | 0.84519 | 0.3468002379 | 0.0632822 | 0.4395704616 | 0 |
| 2 | 47482854 | A | C | I | L | MSH2 | ENST00000233146 | 0.38989 | 0.00574 | 0.35574 | 0.66813 | 0.77845 | 0.2931049422 | 0.90274 | 0.6889771223 | 0.140059 | 1.958960152 | 0 |
| 2 | 47482854 | A | G | I | V | MSH2 | ENST00000233146 | 0.35674 | 0.00603 | 0.26894 | 0.6091 | 0.67843 | 0.09791697129 | 0.82816 | 0.433685571 | 0.080747 | 1.305756562 | 1 |
| 2 | 47482854 | A | T | I | F | MSH2 | ENST00000233146 | 0.47432 | 0.00608 | 0.36608 | 0.79257 | 0.89422 | 0.4235324064 | 0.95748 | 0.9009417892 | 0.257273 | 5.006159284 | 0 |
| 2 | 47482855 | T | A | I | N | MSH2 | ENST00000233146 | 0.57463 | 0.00483 | 0.47024 | 0.86097 | 0.89101 | 0.5326237459 | 0.84068 | 0.9679479599 | 0.265286 | 4.927538575 | 0 |
| 2 | 47482855 | T | C | I | T | MSH2 | ENST00000233146 | 0.6489 | 0.00586 | 0.50552 | 0.79854 | 0.862 | 0.2728135019 | 0.92596 | 0.8225210905 | 0.275576 | 3.372133129 | 0 |
| 2 | 47482855 | T | G | I | S | MSH2 | ENST00000233146 | 0.7153 | 0.0058 | 0.38773 | 0.83216 | 0.89873 | 0.4981776102 | 0.95654 | 0.9314830899 | 0.311603 | 3.404731629 | 0 |
| 2 | 47482857 | A | C | T | P | MSH2 | ENST00000233146 | 0.264 | 0.0058 | 0.14239 | 0.7123 | 0.75687 | 0.5214694145 | 0.93797 | 0.1685954332 | 0.0997883 | 2.968182559 | 0 |
| 2 | 47482857 | A | G | T | A | MSH2 | ENST00000233146 | 0.21208 | 0.00574 | 0.12006 | 0.54132 | 0.65772 | 0.1424828481 | 0.84188 | 0.06773500144 | 0.057962 | 0.1192425686 | 0 |
| 2 | 47482857 | A | T | T | S | MSH2 | ENST00000233146 | 0.20249 | 0.00586 | 0.13274 | 0.53322 | 0.64157 | 0.1205979201 | 0.80178 | 0.06213982403 | 0.0325873 | 0.3194987722 | 0 |
| 2 | 47482858 | C | A | T | K | MSH2 | ENST00000233146 | 0.13717 | 0.00535 | 0.17109 | 0.4179 | 0.70262 | 0.2720230721 | 0.88789 | 0.190402627 | 0.0726881 | -1.644579834 | 0 |
| 2 | 47482858 | C | G | T | R | MSH2 | ENST00000233146 | 0.13522 | 0.00543 | 0.15731 | 0.40796 | 0.79395 | 0.2914525145 | 0.99692 | 0.04676571315 | 0.172639 | -0.3339283886 | 0 |
| 2 | 47482858 | C | T | T | I | MSH2 | ENST00000233146 | 0.13156 | 0.00556 | 0.19908 | 0.45924 | 0.74021 | 0.1844631994 | 0.94208 | 0.0309920661 | 0.0126984 | -0.032949011 | 0 |
| 2 | 47482860 | A | C | I | L | MSH2 | ENST00000233146 | 0.12168 | 0.00569 | 0.03928 | 0.62018 | 0.62876 | 0.1338303557 | 0.41925 | 0.04851135239 | -0.00569296 | 0.1599109238 | 0 |
| 2 | 47482860 | A | G | I | V | MSH2 | ENST00000233146 | 0.09631 | 0.00599 | 0.02355 | 0.65065 | 0.59021 | 0.08304458458 | 0.33793 | 0.06988883018 | -0.0122906 | 0.1251826295 | 0 |
| 2 | 47482860 | A | T | I | L | MSH2 | ENST00000233146 | 0.119 | 0.00569 | 0.03928 | 0.64354 | 0.62876 | 0.1338303557 | 0.41742 | 0.04851135239 | -0.00569505 | 0.1599109238 | 0 |
| 2 | 47482861 | T | A | I | K | MSH2 | ENST00000233146 | 0.09935 | 0.00476 | 0.06251 | 0.47073 | 0.6331 | 0.1986731718 | 0.32887 | 0.02982699019 | -0.00304055 | -0.449239272 | 0 |
| 2 | 47482861 | T | C | I | T | MSH2 | ENST00000233146 | 0.11675 | 0.00527 | 0.09557 | 0.46816 | 0.63418 | 0.09376970033 | 0.34274 | 0.03604845063 | -0.00442802 | -1.073440509 | 0 |
| 2 | 47482861 | T | G | I | R | MSH2 | ENST00000233146 | 0.13217 | 0.00476 | 0.05383 | 0.41032 | 0.67478 | 0.1765641478 | 0.37259 | 0.03255820274 | 0.00708942 | -0.03499929407 | 0 |
| 2 | 47482863 | A | C | K | Q | MSH2 | ENST00000233146 | 0.50531 | 0.01025 | 0.44099 | 0.79304 | 0.80417 | 0.2485900044 | 0.72235 | 0.7994909286 | 0.159429 | 3.357900179 | 0 |
| 2 | 47482863 | A | G | K | E | MSH2 | ENST00000233146 | 0.47024 | 0.00517 | 0.47959 | 0.76595 | 0.70589 | 0.2970614155 | 0.78818 | 0.7065977454 | 0.12982 | 1.034154691 | 0 |
| 2 | 47482864 | A | C | K | T | MSH2 | ENST00000233146 | 0.53482 | 0.0114 | 0.49234 | 0.82759 | 0.87099 | 0.3245143445 | 0.82925 | 0.9509428144 | 0.232145 | 3.533901979 | 0 |
| 2 | 47482864 | A | G | K | R | MSH2 | ENST00000233146 | 0.52952 | 0.00633 | 0.40907 | 0.76273 | 0.83505 | 0.2226102318 | 0.77451 | 0.8407219648 | 0.149787 | 3.854769293 | 0 |
| 2 | 47482865 | G | C | K | N | MSH2 | ENST00000233146 | 0.78185 | 0.01145 | 0.50227 | 0.78225 | 0.82724 | 0.3455557415 | 0.75157 | 0.9368715286 | 0.146553 | 5.326442283 | 0 |
| 2 | 47482865 | G | T | K | N | MSH2 | ENST00000233146 | 0.77537 | 0.01145 | 0.50227 | 0.78225 | 0.82724 | 0.3455557415 | 0.75162 | 0.9368715286 | 0.146553 | 5.326442283 | 0 |
| 2 | 47482866 | T | A | L | I | MSH2 | ENST00000233146 | 0.27218 | 0.00603 | 0.53724 | 0.81011 | 0.79273 | 0.2209694957 | 0.71243 | 0.385815531 | 0.122243 | 2.570792493 | 0 |
| 2 | 47482866 | T | G | L | V | MSH2 | ENST00000233146 | 0.27312 | 0.00615 | 0.48658 | 0.793 | 0.82046 | 0.3589687979 | 0.80013 | 0.2736750245 | 0.181521 | 2.394738416 | 0 |
| 2 | 47482867 | T | C | L | S | MSH2 | ENST00000233146 | 0.7222 | 0.02356 | 0.55406 | 0.9249 | 0.95826 | 0.6284996571 | 0.89329 | 0.9873411059 | 0.410158 | 6.196632828 | 0 |
| 2 | 47482868 | A | C | L | F | MSH2 | ENST00000233146 | 0.29507 | 0.00741 | 0.52596 | 0.84299 | 0.83779 | 0.4386267271 | 0.69015 | 0.5492987633 | 0.112643 | 5.088947986 | 1 |
| 2 | 47482868 | A | T | L | F | MSH2 | ENST00000233146 | 0.29084 | 0.00741 | 0.52596 | 0.84686 | 0.83779 | 0.4386267271 | 0.69022 | 0.5492987633 | 0.112645 | 5.088947986 | 1 |
| 2 | 47482869 | A | C | K | Q | MSH2 | ENST00000233146 | 0.46686 | 0.00556 | 0.22798 | 0.59531 | 0.73074 | 0.2446756981 | 0.84734 | 0.5624150634 | 0.133116 | 1.793099775 | 0 |
| 2 | 47482869 | A | G | K | E | MSH2 | ENST00000233146 | 0.49886 | 0.00517 | 0.25944 | 0.67912 | 0.77845 | 0.2550800864 | 0.9269 | 0.5953922868 | 0.229183 | 2.386746542 | 0 |
| 2 | 47482870 | A | C | K | T | MSH2 | ENST00000233146 | 0.40722 | 0.00562 | 0.27662 | 0.75749 | 0.83615 | 0.3236650573 | 0.87586 | 0.6932452321 | 0.205712 | 4.625709402 | 0 |
| 2 | 47482870 | A | G | K | R | MSH2 | ENST00000233146 | 0.35112 | 0.00603 | 0.20936 | 0.58675 | 0.74867 | 0.1867910964 | 0.80179 | 0.293520242 | 0.0958496 | 3.034135653 | 0 |
| 2 | 47482870 | A | T | K | I | MSH2 | ENST00000233146 | 0.45813 | 0.00608 | 0.24709 | 0.81418 | 0.86603 | 0.4138227392 | 0.86705 | 0.8211560845 | 0.236877 | 6.526018979 | 0 |
| 2 | 47482871 | A | C | K | N | MSH2 | ENST00000233146 | 0.30874 | 0.00596 | 0.2843 | 0.74655 | 0.75552 | 0.3194068331 | 0.76872 | 0.2215017527 | 0.106464 | 1.942112439 | 0 |
| 2 | 47482871 | A | T | K | N | MSH2 | ENST00000233146 | 0.30442 | 0.00596 | 0.2843 | 0.74655 | 0.75552 | 0.3194068331 | 0.76896 | 0.2189797005 | 0.10646 | 1.942112439 | 0 |
| 2 | 47482872 | C | A | Q | K | MSH2 | ENST00000233146 | 0.36064 | 0.00527 | 0.23109 | 0.74329 | 0.73443 | 0.2337179207 | 0.78884 | 0.2251179516 | 0.0548724 | 1.085802234 | 0 |
| 2 | 47482872 | C | G | Q | E | MSH2 | ENST00000233146 | 0.29607 | 0.00556 | 0.19821 | 0.65542 | 0.70914 | 0.1935260549 | 0.70858 | 0.1471109858 | 0.0888922 | 1.813322797 | 0 |
| 2 | 47482873 | A | C | Q | P | MSH2 | ENST00000233146 | 0.49978 | 0.00783 | 0.21149 | 0.81703 | 0.89694 | 0.6194312106 | 0.85921 | 0.7827838659 | 0.274623 | 6.198177403 | 0 |
| 2 | 47482873 | A | G | Q | R | MSH2 | ENST00000233146 | 0.43172 | 0.00556 | 0.22413 | 0.68667 | 0.73949 | 0.199019417 | 0.82786 | 0.6937984824 | 0.173232 | 2.318333235 | 0 |
| 2 | 47482873 | A | T | Q | L | MSH2 | ENST00000233146 | 0.49248 | 0.0058 | 0.18401 | 0.80184 | 0.8124 | 0.3142898196 | 0.85344 | 0.8964663744 | 0.211127 | 4.929561295 | 0 |
| 2 | 47482874 | G | C | Q | H | MSH2 | ENST00000233146 | 0.30274 | 0.01401 | 0.22709 | 0.80789 | 0.87247 | 0.2234208775 | 0.80859 | 0.9010909796 | 0.192123 | 3.92465549 | 0 |
| 2 | 47482874 | G | T | Q | H | MSH2 | ENST00000233146 | 0.29864 | 0.01401 | 0.22709 | 0.80789 | 0.87247 | 0.2234208775 | 0.80908 | 0.9010909796 | 0.19212 | 3.92465549 | 0 |
| 2 | 47482875 | C | A | L | I | MSH2 | ENST00000233146 | 0.36041 | 0.00916 | 0.46592 | 0.80102 | 0.79697 | 0.2619522182 | 0.62088 | 0.9279207587 | 0.130008 | 3.887030586 | 0 |
| 2 | 47482875 | C | G | L | V | MSH2 | ENST00000233146 | 0.48171 | 0.01369 | 0.38625 | 0.81629 | 0.89055 | 0.4049880284 | 0.73901 | 0.8926256895 | 0.211896 | 5.249124635 | 0 |
| 2 | 47482876 | T | A | L | Q | MSH2 | ENST00000233146 | 0.75822 | 0.03293 | 0.47394 | 0.95465 | 0.95353 | 0.6111541303 | 0.83419 | 0.9969345331 | 0.3586 | 6.700904088 | 0 |
| 2 | 47482876 | T | C | L | P | MSH2 | ENST00000233146 | 0.92166 | 0.02796 | 0.57327 | 0.955 | 0.96737 | 0.8323051145 | 0.8664 | 0.9972634315 | 0.435739 | 7.95719211 | 0 |
| 2 | 47482876 | T | G | L | R | MSH2 | ENST00000233146 | 0.91534 | 0.02732 | 0.53277 | 0.95789 | 0.97735 | 0.7813314651 | 0.9133 | 0.7972609997 | 0.283315 | 8.101496531 | 0 |
| 2 | 47482878 | A | C | K | Q | MSH2 | ENST00000233146 | 0.79713 | 0.02835 | 0.45233 | 0.89322 | 0.91945 | 0.4131146739 | 0.87988 | 0.9336572886 | 0.244363 | 3.952973439 | 0 |
| 2 | 47482878 | A | G | K | E | MSH2 | ENST00000233146 | 0.79579 | 0.02056 | 0.49186 | 0.89955 | 0.92601 | 0.4826415009 | 0.94598 | 0.9557011724 | 0.298053 | 5.624444104 | 0 |
| 2 | 47482879 | A | C | K | T | MSH2 | ENST00000233146 | 0.75623 | 0.02869 | 0.50941 | 0.92412 | 0.9252 | 0.4637537835 | 0.90964 | 0.9872041345 | 0.249913 | 5.381976938 | 0 |
| 2 | 47482879 | A | G | K | R | MSH2 | ENST00000233146 | 0.40757 | 0.00654 | 0.41957 | 0.70995 | 0.73949 | 0.3016003071 | 0.9301 | 0.7717584968 | 0.138443 | 2.71711546 | 0 |
| 2 | 47482879 | A | T | K | I | MSH2 | ENST00000233146 | 0.8089 | 0.01115 | 0.47247 | 0.94815 | 0.93554 | 0.5464518888 | 0.89033 | 0.9896358848 | 0.258628 | 5.033497178 | 0 |
| 2 | 47482880 | A | C | K | N | MSH2 | ENST00000233146 | 0.66541 | 0.02891 | 0.52759 | 0.87336 | 0.8887 | 0.4798389852 | 0.8933 | 0.9746175408 | 0.144249 | 5.607387241 | 0 |
| 2 | 47482880 | A | T | K | N | MSH2 | ENST00000233146 | 0.65877 | 0.02891 | 0.52759 | 0.88736 | 0.8887 | 0.4798389852 | 0.89305 | 0.9746175408 | 0.144242 | 5.607387241 | 0 |
| 2 | 47482881 | G | A | A | T | MSH2 | ENST00000233146 | 0.39731 | 0.00586 | 0.13841 | 0.75444 | 0.7872 | 0.1669219449 | 0.95324 | 0.3352116942 | 0.200534 | 3.285870258 | 0 |
| 2 | 47482881 | G | C | A | P | MSH2 | ENST00000233146 | 0.46196 | 0.01044 | 0.19513 | 0.76804 | 0.84375 | 0.6146381332 | 0.96569 | 0.7204585671 | 0.263251 | 5.908298616 | 0 |
| 2 | 47482881 | G | T | A | S | MSH2 | ENST00000233146 | 0.34778 | 0.00596 | 0.11202 | 0.62709 | 0.66354 | 0.1727230758 | 0.9073 | 0.3506679237 | 0.110652 | 1.956073595 | 0 |
| 2 | 47482882 | C | A | A | D | MSH2 | ENST00000233146 | 0.29035 | 0.00504 | 0.22061 | 0.7372 | 0.7816 | 0.4180091888 | 0.97071 | 0.4985866547 | 0.245077 | 2.362928045 | 0 |
| 2 | 47482882 | C | G | A | G | MSH2 | ENST00000233146 | 0.31748 | 0.00674 | 0.13245 | 0.75232 | 0.7872 | 0.1917685955 | 0.89867 | 0.3337257918 | 0.149467 | 4.26805019 | 0 |
| 2 | 47482882 | C | T | A | V | MSH2 | ENST00000233146 | 0.33131 | 0.00735 | 0.15268 | 0.76042 | 0.80476 | 0.260793482 | 0.90673 | 0.6405146718 | 0.204418 | 5.328170562 | 0 |
| 2 | 47482884 | G | A | E | K | MSH2 | ENST00000233146 | 0.58888 | 0.00566 | 0.44851 | 0.70749 | 0.7675 | 0.302618262 | 0.9769 | 0.9174355865 | 0.262976 | 2.866431084 | 0 |
| 2 | 47482884 | G | C | E | Q | MSH2 | ENST00000233146 | 0.56825 | 0.00659 | 0.33479 | 0.67805 | 0.76816 | 0.2524917103 | 0.93278 | 0.8787643313 | 0.220401 | 2.645986905 | 0 |
| 2 | 47482885 | A | C | E | A | MSH2 | ENST00000233146 | 0.61529 | 0.00799 | 0.3487 | 0.81702 | 0.85274 | 0.328643121 | 0.9089 | 0.9364677072 | 0.233895 | 4.70626007 | 0 |
| 2 | 47482885 | A | G | E | G | MSH2 | ENST00000233146 | 0.52493 | 0.00569 | 0.31088 | 0.76705 | 0.83172 | 0.3475583804 | 0.94722 | 0.9779266715 | 0.240611 | 3.490930856 | 0 |
| 2 | 47482885 | A | T | E | V | MSH2 | ENST00000233146 | 0.79579 | 0.01471 | 0.32472 | 0.90492 | 0.9045 | 0.3819376854 | 0.93137 | 0.9793942571 | 0.284521 | 5.97479676 | 0 |
| 2 | 47482886 | A | C | E | D | MSH2 | ENST00000233146 | 0.30915 | 0.00608 | 0.34916 | 0.58173 | 0.79878 | 0.22466067 | 0.96025 | 0.2256473154 | 0.122813 | 2.622198073 | 0 |
| 2 | 47482886 | A | T | E | D | MSH2 | ENST00000233146 | 0.30478 | 0.00608 | 0.34916 | 0.58173 | 0.79878 | 0.22466067 | 0.96039 | 0.23428002 | 0.122801 | 2.622198073 | 0 |
| 2 | 47482887 | G | A | V | I | MSH2 | ENST00000233146 | 0.45998 | 0.00627 | 0.30218 | 0.72193 | 0.79758 | 0.245860007 | 0.79724 | 0.6511089802 | 0.18237 | 3.535172376 | 0 |
| 2 | 47482887 | G | C | V | L | MSH2 | ENST00000233146 | 0.49357 | 0.00586 | 0.44439 | 0.75033 | 0.77141 | 0.3957282609 | 0.79292 | 0.7117798924 | 0.169972 | 2.655908348 | 0 |
| 2 | 47482887 | G | T | V | L | MSH2 | ENST00000233146 | 0.48838 | 0.00586 | 0.44439 | 0.75147 | 0.77205 | 0.3957282609 | 0.79184 | 0.7020775676 | 0.169978 | 2.655908348 | 0 |
| 2 | 47482888 | T | A | V | E | MSH2 | ENST00000233146 | 0.73837 | 0.00608 | 0.44929 | 0.91279 | 0.93515 | 0.736615787 | 0.84827 | 0.9847103953 | 0.28266 | 6.749136754 | 0 |
| 2 | 47482888 | T | C | V | A | MSH2 | ENST00000233146 | 0.42323 | 0.00556 | 0.46658 | 0.77831 | 0.8705 | 0.4877516839 | 0.87616 | 0.9346564412 | 0.261672 | 3.769856309 | 0 |
| 2 | 47482888 | T | G | V | G | MSH2 | ENST00000233146 | 0.73989 | 0.01211 | 0.35656 | 0.92125 | 0.95681 | 0.6632854141 | 0.90688 | 0.993461132 | 0.363519 | 7.168953633 | 0 |
| 2 | 47482890 | A | C | I | L | MSH2 | ENST00000233146 | 0.21589 | 0.00569 | 0.20598 | 0.4371 | 0.73 | 0.1948570254 | 0.7612 | 0.0157052269 | -0.00372968 | -1.958146442 | 0 |
| 2 | 47482890 | A | G | I | V | MSH2 | ENST00000233146 | 0.21501 | 0.00599 | 0.13565 | 0.41853 | 0.75349 | 0.0936835625 | 0.8027 | 0.02301904032 | 0.0413368 | -0.8102664533 | 0 |
| 2 | 47482890 | A | T | I | L | MSH2 | ENST00000233146 | 0.21256 | 0.00569 | 0.20598 | 0.43581 | 0.73 | 0.1948570254 | 0.76103 | 0.02630697299 | 0.0405773 | -1.958146442 | 0 |
| 2 | 47482891 | T | A | I | K | MSH2 | ENST00000233146 | 0.28622 | 0.00476 | 0.26466 | 0.73346 | 0.77527 | 0.3894823945 | 0.94678 | 0.7322980762 | 0.207378 | -0.6350954325 | 0 |
| 2 | 47482891 | T | C | I | T | MSH2 | ENST00000233146 | 0.26447 | 0.00532 | 0.33012 | 0.71907 | 0.76421 | 0.1682243405 | 0.90175 | 0.5344392061 | 0.204563 | 0.9755570745 | 0 |
| 2 | 47482891 | T | G | I | R | MSH2 | ENST00000233146 | 0.30905 | 0.00476 | 0.24573 | 0.74428 | 0.79456 | 0.3519653731 | 0.93411 | 0.5314692855 | 0.238716 | 0.4122187465 | 0 |
| 2 | 47482893 | G | A | A | T | MSH2 | ENST00000233146 | 0.50348 | 0.00608 | 0.36086 | 0.67208 | 0.80713 | 0.1951715145 | 0.94601 | 0.223390139 | 0.100134 | 4.676861249 | 0 |
| 2 | 47482893 | G | C | A | P | MSH2 | ENST00000233146 | 0.81127 | 0.01376 | 0.44025 | 0.79631 | 0.88354 | 0.6435960985 | 0.89876 | 0.9242274165 | 0.278603 | 7.683453298 | 0 |
| 2 | 47482893 | G | T | A | S | MSH2 | ENST00000233146 | 0.42843 | 0.00599 | 0.32012 | 0.70589 | 0.66064 | 0.1953031656 | 0.72802 | 0.4777084291 | 0.0939276 | 2.564763361 | 0 |
| 2 | 47482894 | C | A | A | E | MSH2 | ENST00000233146 | 0.33505 | 0.00574 | 0.43693 | 0.80412 | 0.82556 | 0.5001868454 | 0.95768 | 0.4797611833 | 0.25773 | 2.344062244 | 0 |
| 2 | 47482894 | C | G | A | G | MSH2 | ENST00000233146 | 0.34931 | 0.0058 | 0.3442 | 0.75381 | 0.78597 | 0.2196785576 | 0.92082 | 0.6060836315 | 0.192418 | 3.843208159 | 0 |
| 2 | 47482894 | C | T | A | V | MSH2 | ENST00000233146 | 0.46302 | 0.00923 | 0.38921 | 0.78663 | 0.85482 | 0.2440550926 | 0.87485 | 0.5764276981 | 0.140195 | 7.270979479 | 0 |
| 2 | 47482896 | A | C | K | Q | MSH2 | ENST00000233146 | 0.38386 | 0.00566 | 0.31552 | 0.67943 | 0.74164 | 0.2628065456 | 0.76173 | 0.08487540483 | 0.0841645 | 1.835581945 | 0 |
| 2 | 47482896 | A | G | K | E | MSH2 | ENST00000233146 | 0.39347 | 0.00527 | 0.36187 | 0.70979 | 0.7872 | 0.3323916103 | 0.8852 | 0.09075158089 | 0.108948 | 3.433847541 | 0 |
| 2 | 47482897 | A | C | K | T | MSH2 | ENST00000233146 | 0.5072 | 0.00562 | 0.37337 | 0.71204 | 0.81761 | 0.344482496 | 0.8354 | 0.7327850461 | 0.163936 | 4.60494828 | 0 |
| 2 | 47482897 | A | G | K | R | MSH2 | ENST00000233146 | 0.50563 | 0.00608 | 0.28021 | 0.58125 | 0.71705 | 0.233751164 | 0.8865 | 0.5346093774 | 0.123548 | 2.212908947 | 0 |
| 2 | 47482898 | G | C | K | N | MSH2 | ENST00000233146 | 0.47344 | 0.00674 | 0.39914 | 0.7896 | 0.75687 | 0.3804635374 | 0.75038 | 0.5355038047 | 0.112897 | 4.34419699 | 0 |

| | | | | | | | | | | | | | | | | | | |
|---|---|---|---|---|---|---|---|---|---|---|---|---|---|---|---|---|---|---|
| 2 | 47482898 | G | T | K | N | MSH2 | ENST00000233146 | 0.4683 | 0.00674 | 0.39914 | 0.78938 | 0.75687 | 0.3804635374 | 0.74961 | 0.5355038047 | 0.112888 | 4.34419699 | 0 |
| 2 | 47482899 | A | C | N | H | MSH2 | ENST00000233146 | 0.74794 | 0.03261 | 0.60308 | 0.90193 | 0.86553 | 0.5430016526 | 0.90271 | 0.9661539793 | 0.180793 | 5.330742441 | 0 |
| 2 | 47482899 | A | G | N | D | MSH2 | ENST00000233146 | 0.43768 | 0.00574 | 0.6306 | 0.77773 | 0.68381 | 0.6026186228 | 0.90921 | 0.8461475968 | 0.141248 | 3.300895726 | 0 |
| 2 | 47482900 | A | C | N | T | MSH2 | ENST00000233146 | 0.54923 | 0.00916 | 0.58793 | 0.78543 | 0.83725 | 0.4505600308 | 0.84537 | 0.9400666952 | 0.181545 | 5.468703354 | 0 |
| 2 | 47482900 | A | G | N | S | MSH2 | ENST00000233146 | 0.50289 | 0.0084 | 0.53878 | 0.7838 | 0.77527 | 0.379912641 | 0.86169 | 0.8273311853 | 0.131202 | 4.064449901 | 0 |
| 2 | 47482901 | T | A | N | K | MSH2 | ENST00000233146 | 0.39765 | 0.02319 | 0.65697 | 0.8129 | 0.79151 | 0.6024042353 | 0.87391 | 0.9881101847 | 0.0864368 | 2.97873533 | 0 |
| 2 | 47482901 | T | G | N | K | MSH2 | ENST00000233146 | 0.4036 | 0.02319 | 0.65697 | 0.81276 | 0.79151 | 0.6024042353 | 0.87379 | 0.9737499952 | 0.0864375 | 2.97873533 | 0 |
| 2 | 47482902 | A | G | N | D | MSH2 | ENST00000233146 | 0.77251 | 0.01959 | 0.55099 | 0.88905 | 0.90581 | 0.6348031829 | 0.90682 | 0.9773234129 | 0.173447 | 4.886927816 | 0 |
| 2 | 47482902 | A | T | N | Y | MSH2 | ENST00000233146 | 0.84665 | 0.03388 | 0.54096 | 0.95396 | 0.95972 | 0.7053725322 | 0.96485 | 0.9984542131 | 0.469456 | 8.895647533 | 0 |
| 2 | 47482903 | A | C | N | T | MSH2 | ENST00000233146 | 0.80843 | 0.03626 | 0.48517 | 0.92085 | 0.95316 | 0.5399460528 | 0.89744 | 0.9951298033 | 0.212989 | 7.82536709 | 0 |
| 2 | 47482903 | A | G | N | S | MSH2 | ENST00000233146 | 0.71153 | 0.03901 | 0.42792 | 0.90649 | 0.91694 | 0.4989018977 | 0.92721 | 0.9838012017 | 0.13936 | 4.42178107 | 0 |
| 2 | 47482903 | A | T | N | I | MSH2 | ENST00000233146 | 0.85412 | 0.03362 | 0.49625 | 0.95771 | 0.96117 | 0.6966964638 | 0.93822 | 0.9985282421 | 0.393029 | 5.480967322 | 0 |
| 2 | 47482904 | T | A | N | K | MSH2 | ENST00000233146 | 0.70752 | 0.03447 | 0.59128 | 0.92472 | 0.86603 | 0.6765407922 | 0.9078 | 0.9930297732 | 0.109582 | 4.763984194 | 0 |
| 2 | 47482904 | T | G | N | K | MSH2 | ENST00000233146 | 0.71578 | 0.03447 | 0.59128 | 0.92415 | 0.86603 | 0.6765407922 | 0.90757 | 0.9914206266 | 0.109583 | 4.763984194 | 0 |
| 2 | 47482905 | A | C | S | R | MSH2 | ENST00000233146 | 0.36041 | 0.00551 | 0.26293 | 0.65372 | 0.82103 | 0.3052215383 | 0.96053 | 0.1167733371 | 0.159596 | 2.935589012 | 0 |
| 2 | 47482905 | A | G | S | G | MSH2 | ENST00000233146 | 0.35547 | 0.00586 | 0.1628 | 0.69127 | 0.77141 | 0.1562907614 | 0.91913 | 0.10798634 | 0.0924914 | 2.177965475 | 0 |
| 2 | 47482905 | A | T | S | C | MSH2 | ENST00000233146 | 0.45187 | 0.01244 | 0.20826 | 0.84681 | 0.85894 | 0.1688815494 | 0.89286 | 0.6873736382 | 0.165648 | 6.635562869 | 0 |
| 2 | 47482906 | G | A | S | N | MSH2 | ENST00000233146 | 0.35069 | 0.00664 | 0.2441 | 0.75499 | 0.66733 | 0.180186646 | 0.85425 | 0.66381073 | 0.114076 | 2.931401541 | 0 |
| 2 | 47482906 | G | C | S | T | MSH2 | ENST00000233146 | 0.33591 | 0.00586 | 0.20804 | 0.66752 | 0.69509 | 0.1362127446 | 0.90824 | 0.6700635552 | 0.126933 | 2.520574742 | 0 |
| 2 | 47482906 | G | T | S | I | MSH2 | ENST00000233146 | 0.43496 | 0.00543 | 0.22785 | 0.77792 | 0.82216 | 0.2775831273 | 0.85005 | 0.7790813446 | 0.176882 | 6.635562869 | 0 |
| 2 | 47482907 | C | A | S | R | MSH2 | ENST00000233146 | 0.29492 | 0.00551 | 0.26293 | 0.6961 | 0.81065 | 0.3052215383 | 0.95853 | 0.2091832906 | 0.132877 | 2.935589012 | 0 |
| 2 | 47482907 | C | G | S | R | MSH2 | ENST00000233146 | 0.30025 | 0.00551 | 0.26293 | 0.6404 | 0.81065 | 0.3052215383 | 0.95898 | 0.1871647984 | 0.132867 | 2.935589012 | 0 |
| 2 | 47482908 | T | A | F | I | MSH2 | ENST00000233146 | 0.90993 | 0.01574 | 0.6255 | 0.8875 | 0.93123 | 0.6279306517 | 0.86991 | 0.969832971 | 0.279435 | 4.637541985 | 0 |
| 2 | 47482908 | T | C | F | L | MSH2 | ENST00000233146 | 0.78867 | 0.00777 | 0.66322 | 0.81203 | 0.89603 | 0.5212912311 | 0.83409 | 0.9493826032 | 0.264679 | 4.482408093 | 0 |
| 2 | 47482908 | T | G | F | V | MSH2 | ENST00000233146 | 0.90745 | 0.01133 | 0.58343 | 0.89347 | 0.94431 | 0.5762983889 | 0.9384 | 0.9833684564 | 0.404304 | 3.995598439 | 0 |
| 2 | 47482909 | T | A | F | Y | MSH2 | ENST00000233146 | 0.45175 | 0.00551 | 0.64307 | 0.74278 | 0.71938 | 0.430546132 | 0.74965 | 0.8189247847 | 0.127246 | 1.301013314 | 0 |
| 2 | 47482909 | T | C | F | S | MSH2 | ENST00000233146 | 0.9404 | 0.01621 | 0.62506 | 0.93185 | 0.94431 | 0.6573620653 | 0.88837 | 0.9910180569 | 0.334229 | 5.802086884 | 0 |
| 2 | 47482909 | T | G | F | C | MSH2 | ENST00000233146 | 0.94398 | 0.02442 | 0.65965 | 0.94619 | 0.94913 | 0.6110086531 | 0.9137 | 0.9929910898 | 0.393966 | 8.061413063 | 0 |
| 2 | 47482910 | T | A | F | L | MSH2 | ENST00000233146 | 0.48538 | 0.00777 | 0.66322 | 0.85917 | 0.89193 | 0.5212912311 | 0.85291 | 0.8584894538 | 0.217551 | 4.482408093 | 0 |
| 2 | 47482910 | T | G | F | L | MSH2 | ENST00000233146 | 0.4919 | 0.00777 | 0.66322 | 0.8593 | 0.89193 | 0.5212912311 | 0.85208 | 0.8584894538 | 0.217553 | 4.482408093 | 0 |
| 2 | 47482911 | G | A | V | I | MSH2 | ENST00000233146 | 0.33575 | 0.00659 | 0.45036 | 0.67166 | 0.7255 | 0.1723188822 | 0.79482 | 0.531068027 | 0.130946 | 1.459502079 | 0 |
| 2 | 47482911 | G | C | V | L | MSH2 | ENST00000233146 | 0.4496 | 0.00654 | 0.64354 | 0.81909 | 0.78223 | 0.3932470097 | 0.8926 | 0.8233166933 | 0.231999 | 4.265492603 | 0 |
| 2 | 47482911 | G | T | V | L | MSH2 | ENST00000233146 | 0.44466 | 0.00654 | 0.64354 | 0.81889 | 0.78223 | 0.3932470097 | 0.89341 | 0.8288613558 | 0.231997 | 4.265492603 | 0 |
| 2 | 47482912 | T | A | V | E | MSH2 | ENST00000233146 | 0.76658 | 0.02589 | 0.64759 | 0.95632 | 0.9796 | 0.7533786338 | 0.97678 | 0.9922127128 | 0.471083 | 8.118158845 | 1 |
| 2 | 47482912 | T | C | V | A | MSH2 | ENST00000233146 | 0.69485 | 0.01715 | 0.66706 | 0.87889 | 0.91694 | 0.479071195 | 0.80666 | 0.968546927 | 0.27515 | 8.118158845 | 0 |
| 2 | 47482912 | T | G | V | G | MSH2 | ENST00000233146 | 0.76179 | 0.02864 | 0.53397 | 0.94285 | 0.96189 | 0.6784853173 | 0.93996 | 0.9938236475 | 0.460822 | 8.118158845 | 0 |
| 2 | 47482914 | A | C | N | H | MSH2 | ENST00000233146 | 0.48966 | 0.00566 | 0.43582 | 0.67576 | 0.59146 | 0.1981507401 | 0.94211 | 0.7831796408 | 0.136716 | 3.774667507 | 0 |
| 2 | 47482914 | A | G | N | D | MSH2 | ENST00000233146 | 0.46699 | 0.00551 | 0.49239 | 0.77592 | 0.69425 | 0.2531969663 | 0.90116 | 0.7760589719 | 0.132144 | 3.949701912 | 0 |
| 2 | 47482914 | A | T | N | Y | MSH2 | ENST00000233146 | 0.67302 | 0.00754 | 0.47157 | 0.84584 | 0.89193 | 0.3109944937 | 0.9656 | 0.9791353345 | 0.296021 | 7.437547085 | 0 |
| 2 | 47482915 | A | C | N | T | MSH2 | ENST00000233146 | 0.40086 | 0.00633 | 0.40203 | 0.76084 | 0.81646 | 0.1384495875 | 0.87359 | 0.6597067714 | 0.140497 | 3.044797249 | 0 |
| 2 | 47482915 | A | G | N | S | MSH2 | ENST00000233146 | 0.34544 | 0.00619 | 0.34741 | 0.59008 | 0.57748 | 0.1017481832 | 0.84789 | 0.2966425419 | 0.0400771 | 4.006381211 | 0 |
| 2 | 47482915 | A | T | N | I | MSH2 | ENST00000233146 | 0.45352 | 0.00543 | 0.42543 | 0.80595 | 0.86149 | 0.3171247248 | 0.94715 | 0.849722147 | 0.241804 | 7.437547085 | 0 |
| 2 | 47482916 | T | A | N | K | MSH2 | ENST00000233146 | 0.34745 | 0.00547 | 0.51922 | 0.77579 | 0.74446 | 0.201054528 | 0.90825 | 0.3325391412 | 0.0955218 | 2.665251843 | 0 |
| 2 | 47482916 | T | G | N | K | MSH2 | ENST00000233146 | 0.35311 | 0.00547 | 0.51922 | 0.75238 | 0.74376 | 0.201054528 | 0.90615 | 0.2247868925 | 0.0955378 | 2.665251843 | 0 |
| 2 | 47482917 | G | A | E | K | MSH2 | ENST00000233146 | 0.59849 | 0.00783 | 0.70493 | 0.82343 | 0.86951 | 0.3983615581 | 0.92689 | 0.9264680743 | 0.367578 | 3.795204914 | 0 |
| 2 | 47482917 | G | C | E | Q | MSH2 | ENST00000233146 | 0.55039 | 0.0063 | 0.58354 | 0.79854 | 0.80713 | 0.2938388419 | 0.92128 | 0.7891772985 | 0.275535 | 4.226927809 | 0 |
| 2 | 47482918 | A | C | E | A | MSH2 | ENST00000233146 | 0.4769 | 0.00599 | 0.59603 | 0.81373 | 0.88259 | 0.3781451287 | 0.87974 | 0.8418254852 | 0.263723 | 5.001499682 | 0 |
| 2 | 47482918 | A | G | E | G | MSH2 | ENST00000233146 | 0.53643 | 0.00547 | 0.53927 | 0.80039 | 0.81182 | 0.3813443466 | 0.92586 | 0.9191184044 | 0.281931 | 2.225421975 | 0 |
| 2 | 47482919 | A | C | E | D | MSH2 | ENST00000233146 | 0.37697 | 0.00586 | 0.59489 | 0.78342 | 0.73661 | 0.2727911653 | 0.93427 | 0.3817827404 | 0.190748 | 1.659892813 | 0 |
| 2 | 47482919 | A | T | E | D | MSH2 | ENST00000233146 | 0.37222 | 0.00586 | 0.59489 | 0.7965 | 0.73661 | 0.2727911653 | 0.93279 | 0.4623948336 | 0.190763 | 1.659892813 | 0 |
| 2 | 47482920 | A | C | I | L | MSH2 | ENST00000233146 | 0.39754 | 0.00574 | 0.69582 | 0.76644 | 0.74446 | 0.3492478936 | 0.81359 | 0.6162768006 | 0.13116 | 1.636492907 | 0 |
| 2 | 47482920 | A | G | I | V | MSH2 | ENST00000233146 | 0.39127 | 0.00608 | 0.59541 | 0.63706 | 0.7217 | 0.1522433855 | 0.84308 | 0.4934398234 | 0.123298 | 1.933390221 | 0 |
| 2 | 47482920 | A | T | I | F | MSH2 | ENST00000233146 | 0.77108 | 0.00608 | 0.71086 | 0.819 | 0.91058 | 0.5711056072 | 0.84892 | 0.8020679951 | 0.249675 | 7.610583229 | 0 |
| 2 | 47482921 | T | C | I | T | MSH2 | ENST00000233146 | 0.40716 | 0.00971 | 0.8132 | 0.85681 | 0.89055 | 0.4534642593 | 0.83799 | 0.9730695486 | 0.269376 | 7.610583229 | 0 |
| 2 | 47482921 | T | G | I | S | MSH2 | ENST00000233146 | 0.60222 | 0.01211 | 0.72262 | 0.92821 | 0.94205 | 0.7005089609 | 0.92453 | 0.9948042035 | 0.329461 | 7.610583229 | 0 |
| 2 | 47482923 | A | C | I | L | MSH2 | ENST00000233146 | 0.37115 | 0.00574 | 0.49606 | 0.70005 | 0.73 | 0.253598089 | 0.82349 | 0.3760069609 | 0.0938148 | 0.8906934563 | 0 |
| 2 | 47482923 | A | G | I | V | MSH2 | ENST00000233146 | 0.42252 | 0.00603 | 0.36409 | 0.69161 | 0.76157 | 0.1548012312 | 0.94695 | 0.4770435095 | 0.174678 | 1.513284383 | 0 |
| 2 | 47482923 | A | T | I | F | MSH2 | ENST00000233146 | 0.49203 | 0.00941 | 0.5059 | 0.85147 | 0.88684 | 0.4448578448 | 0.89701 | 0.6324715614 | 0.236198 | 7.056047148 | 0 |
| 2 | 47482924 | T | A | I | N | MSH2 | ENST00000233146 | 0.84347 | 0.01818 | 0.62079 | 0.95347 | 0.94015 | 0.629318111 | 0.82884 | 0.9967008233 | 0.276232 | 5.544288183 | 0 |
| 2 | 47482924 | T | C | I | T | MSH2 | ENST00000233146 | 0.61447 | 0.00754 | 0.65623 | 0.86951 | 0.90229 | 0.3739439441 | 0.89572 | 0.9578610659 | 0.30854 | 7.056047148 | 0 |
| 2 | 47482924 | T | G | I | S | MSH2 | ENST00000233146 | 0.69123 | 0.0129 | 0.52643 | 0.94095 | 0.93939 | 0.6297080791 | 0.8868 | 0.978705585 | 0.307084 | 7.056047148 | 0 |
| 2 | 47482925 | T | G | I | M | MSH2 | ENST00000233146 | 0.32653 | 0.01529 | 0.4409 | 0.83493 | 0.85996 | 0.2726459773 | 0.83934 | 0.8463779092 | 0.149212 | 3.159754242 | 0 |
| 2 | 47482926 | T | A | S | T | MSH2 | ENST00000233146 | 0.37691 | 0.00696 | 0.43561 | 0.79451 | 0.85638 | 0.1706220153 | 0.91717 | 0.5414139628 | 0.180699 | 5.185790903 | 0 |
| 2 | 47482926 | T | C | S | P | MSH2 | ENST00000233146 | 0.44655 | 0.00836 | 0.45245 | 0.84091 | 0.88543 | 0.6141509415 | 0.9722 | 0.7431228161 | 0.275372 | 7.31561987 | 0 |
| 2 | 47482926 | T | G | S | A | MSH2 | ENST00000233146 | 0.35058 | 0.00603 | 0.40997 | 0.67229 | 0.74376 | 0.1044708407 | 0.8771 | 0.1445281059 | 0.0975458 | 1.096472169 | 0 |
| 2 | 47482927 | C | T | S | L | MSH2 | ENST00000233146 | 0.41617 | 0.00669 | 0.45711 | 0.85927 | 0.84106 | 0.3035552983 | 0.90484 | 0.8421655297 | 0.210758 | 3.460486754 | 0 |
| 2 | 47482929 | C | G | R | G | MSH2 | ENST00000233146 | 0.3468 | 0.00586 | 0.51212 | 0.85136 | 0.80536 | 0.4225250965 | 0.82595 | 0.2608197629 | 0.150449 | 7.81182756 | 0 |
| 2 | 47482930 | G | C | R | P | MSH2 | ENST00000233146 | 0.8115 | 0.02691 | 0.57014 | 0.91157 | 0.90885 | 0.7083202084 | 0.89139 | 0.9741718173 | 0.300757 | 7.81182756 | 0 |
| 2 | 47482930 | G | T | R | L | MSH2 | ENST00000233146 | 0.76331 | 0.0248 | 0.55037 | 0.90575 | 0.90624 | 0.449023748 | 0.90958 | 0.9706676602 | 0.277332 | 7.81182756 | 0 |
| 2 | 47482932 | A | C | I | L | MSH2 | ENST00000233146 | 0.34358 | 0.00569 | 0.49361 | 0.66378 | 0.74446 | 0.1860214333 | 0.87653 | 0.2781584263 | 0.084459 | 3.257963962 | 0 |
| 2 | 47482932 | A | G | I | V | MSH2 | ENST00000233146 | 0.26977 | 0.00599 | 0.40082 | 0.58739 | 0.73223 | 0.1043675304 | 0.83959 | 0.06099519408 | 0.0294258 | 2.110158589 | 0 |
| 2 | 47482932 | A | T | I | L | MSH2 | ENST00000233146 | 0.33903 | 0.00569 | 0.49361 | 0.6609 | 0.74446 | 0.1860214333 | 0.87864 | 0.4095658362 | 0.0844457 | 3.257963962 | 0 |
| 2 | 47482933 | T | A | I | K | MSH2 | ENST00000233146 | 0.21013 | 0.00522 | 0.56573 | 0.85916 | 0.80653 | 0.4598973155 | 0.92668 | 0.8204690218 | 0.212666 | 1.034301164 | 0 |
| 2 | 47482933 | T | C | I | T | MSH2 | ENST00000233146 | 0.17928 | 0.00527 | 0.60106 | 0.53022 | 0.69594 | 0.1736561323 | 0.79144 | 0.1727554947 | 0.0999636 | 0.2956625522 | 0 |
| 2 | 47482933 | T | G | I | R | MSH2 | ENST00000233146 | 0.29607 | 0.00586 | 0.54454 | 0.85634 | 0.81703 | 0.4582287618 | 0.91446 | 0.8044463992 | 0.216292 | 5.845531522 | 0 |
| 2 | 47482935 | A | G | K | E | MSH2 | ENST00000233146 | 0.4094 | 0.00804 | 0.60537 | 0.83978 | 0.85741 | 0.6440369369 | 0.97486 | 0.3671518564 | 0.260407 | 3.941294915 | 0 |
| 2 | 47482936 | A | C | K | T | MSH2 | ENST00000233146 | 0.67279 | 0.03175 | 0.61659 | 0.92622 | 0.95389 | 0.6258863279 | 0.96645 | 0.9316573143 | 0.40279 | 2.679424595 | 0 |
| 2 | 47482936 | A | G | K | R | MSH2 | ENST00000233146 | 0.5332 | 0.03327 | 0.52139 | 0.83977 | 0.911 | 0.5141979365 | 0.92016 | 0.9509626627 | 0.253575 | 7.267250504 | 0 |
| 2 | 47482936 | A | T | K | I | MSH2 | ENST00000233146 | 0.73709 | 0.02785 | 0.57181 | 0.95295 | 0.92924 | 0.6208997812 | 0.93541 | 0.9453088045 | 0.31595 | 4.243724336 | 0 |
| 2 | 47482937 | A | C | K | N | MSH2 | ENST00000233146 | 0.60083 | 0.032 | 0.63717 | 0.89143 | 0.81124 | 0.6229944657 | 0.87618 | 0.7574799657 | 0.12469 | 3.292200275 | 0 |
| 2 | 47482937 | A | T | K | N | MSH2 | ENST00000233146 | 0.59472 | 0.032 | 0.63717 | 0.89143 | 0.81182 | 0.6229944657 | 0.87637 | 0.7721853852 | 0.124688 | 3.292200275 | 0 |
| 2 | 47482938 | G | A | V | I | MSH2 | ENST00000233146 | 0.43406 | 0.00615 | 0.38063 | 0.70539 | 0.68646 | 0.4094880307 | 0.82623 | 0.3056566715 | 0.064563 | 2.924106712 | 0 |
| 2 | 47482938 | G | C | V | L | MSH2 | ENST00000233146 | 0.46574 | 0.00619 | 0.55638 | 0.73696 | 0.71783 | 0.5418325055 | 0.80675 | 0.5349941254 | 0.133807 | 3.965958988 | 0 |
| 2 | 47482938 | G | T | V | F | MSH2 | ENST00000233146 | 0.63973 | 0.00928 | 0.52648 | 0.7697 | 0.79334 | 0.6791349245 | 0.754 | 0.8481290936 | 0.173808 | 7.043587156 | 0 |
| 2 | 47482939 | T | A | V | D | MSH2 | ENST00000233146 | 0.44497 | 0.00741 | 0.58324 | 0.82795 | 0.85792 | 0.7474645116 | 0.91986 | 0.557069838 | 0.218857 | 7.043587156 | 0 |
| 2 | 47482939 | T | C | V | A | MSH2 | ENST00000233146 | 0.18313 | 0.00543 | 0.57961 | 0.72811 | 0.77591 | 0.4646233417 | 0.88862 | 0.07502485607 | 0.0886708 | 2.580395344 | 0 |
| 2 | 47482939 | T | G | V | G | MSH2 | ENST00000233146 | 0.22735 | 0.00527 | 0.4453 | 0.85355 | 0.79212 | 0.589040369 | 0.81579 | 0.2126401869 | 0.122295 | 7.043587156 | 0 |
| 2 | 47482941 | A | C | T | P | MSH2 | ENST00000233146 | 0.34239 | 0.00681 | 0.47688 | 0.57386 | 0.7562 | 0.4085236451 | 0.92307 | 0.135322541 | 0.0767196 | 6.780586483 | 0 |
| 2 | 47482941 | A | G | T | A | MSH2 | ENST00000233146 | 0.31697 | 0.00586 | 0.4748 | 0.51122 | 0.7589 | 0.3077465677 | 0.9583 | 0.1024808663 | 0.0835045 | 2.222820998 | 0 |
| 2 | 47482941 | A | T | T | S | MSH2 | ENST00000233146 | 0.20606 | 0.00586 | 0.49629 | 0.35889 | 0.71938 | 0.253655404 | 0.89262 | 0.106228747 | 0.0357904 | 1.561801656 | 0 |
| 2 | 47482942 | C | A | T | N | MSH2 | ENST00000233146 | 0.24778 | 0.00586 | 0.57962 | 0.68205 | 0.62767 | 0.3831073006 | 0.81982 | 0.1983325034 | 0.0859251 | 6.780586483 | 0 |
| 2 | 47482942 | C | G | T | S | MSH2 | ENST00000233146 | 0.16846 | 0.00586 | 0.49629 | 0.53814 | 0.63736 | 0.253655404 | 0.86456 | 0.1905269921 | 0.0559 | 1.561801656 | 0 |
| 2 | 47482942 | C | T | T | I | MSH2 | ENST00000233146 | 0.27312 | 0.0058 | 0.64259 | 0.70586 | 0.76289 | 0.3394257859 | 0.93121 | 0.2439454066 | 0.143664 | 6.780586483 | 0 |
| 2 | 47482944 | A | C | T | P | MSH2 | ENST00000233146 | 0.15416 | 0.00783 | 0.16062 | 0.6577 | 0.70914 | 0.3679863743 | 0.62364 | 0.06540242338 | 0.0321035 | 6.850962808 | 0 |
| 2 | 47482944 | A | G | T | A | MSH2 | ENST00000233146 | 0.13799 | 0.00599 | 0.14413 | 0.60755 | 0.58006 | 0.3023560753 | 0.68116 | 0.04459992747 | 0.0123789 | 6.850962808 | 0 |
| 2 | 47482944 | A | T | T | S | MSH2 | ENST00000233146 | 0.05439 | 0.00592 | 0.14573 | 0.65412 | 0.66064 | 0.2296609326 | 0.72083 | 0.0464086719 | 0.0126142 | 6.850962808 | 0 |
| 2 | 47482945 | C | A | T | K | MSH2 | ENST00000233146 | 0.10488 | 0.00687 | 0.21674 | 0.75977 | 0.63094 | 0.4120424749 | 0.64652 | 0.1522808794 | -0.00212163 | 6.850962808 | 0 |
| 2 | 47482945 | C | G | T | R | MSH2 | ENST00000233146 | 0.10161 | 0.00909 | 0.21067 | 0.77934 | 0.62767 | 0.447223075 | 0.67052 | 0.1648442108 | 0.072638 | 6.850962808 | 0 |

| Variants with functional readout data from deep mutational scan experiments of PTEN | | | | | | | | | | | | | | | | | | |
|---|---|---|---|---|---|---|---|---|---|---|---|---|---|---|---|---|---|---|
| chrom | pos | ref | alt | ref_aa | alt_aa | genename | transcript_id | CADD_raw_rankscore | MPC_rankscore | PrimateAI_rankscore | M-CAP_rankscore | REVEL_rankscore | gMVP_rankscore | MVP_rankscore | ClinPred | BayesDel | EVmutation | target |
| 10 | 87864473 | A | C | T | P | PTEN | ENST00000371953 | 0.69997 | 0.96506 | 0.74547 | 0.89577 | 0.67933 | 0.8001761067 | 0.92742 | 0.922716856 | 0.185203 | 3.107209863 | 0 |
| 10 | 87864474 | C | A | T | K | PTEN | ENST00000371953 | 0.66399 | 0.95965 | 0.84098 | 0.86654 | 0.7217 | 0.8291754678 | 0.89159 | 0.9724084735 | 0.204216 | 2.960490852 | 0 |
| 10 | 87864474 | C | G | T | R | PTEN | ENST00000371953 | 0.65374 | 0.9734 | 0.82064 | 0.91893 | 0.82387 | 0.8419832322 | 0.93324 | 0.9679777026 | 0.220998 | 3.897564803 | 0 |
| 10 | 87864477 | C | A | A | D | PTEN | ENST00000371953 | 0.5322 | 0.97803 | 0.88782 | 0.91499 | 0.76553 | 0.8633167542 | 0.98497 | 0.8985434175 | 0.36532 | 3.762288291 | 0 |
| 10 | 87864477 | C | T | A | V | PTEN | ENST00000371953 | 0.5154 | 0.92739 | 0.81491 | 0.93216 | 0.80653 | 0.717773638 | 0.94922 | 0.7825034857 | 0.195822 | 4.030344402 | 0 |
| 10 | 87864485 | A | C | K | Q | PTEN | ENST00000371953 | 0.74507 | 0.98843 | 0.85628 | 0.97858 | 0.93475 | 0.8810268829 | 0.95801 | 0.9848486781 | 0.215077 | 4.658916025 | 0 |
| 10 | 87864487 | A | T | K | N | PTEN | ENST00000371953 | 0.63152 | 0.99403 | 0.91569 | 0.94228 | 0.78597 | 0.9159337011 | 0.96963 | 0.9906716347 | 0.128687 | 7.892279782 | 0 |
| 10 | 87864488 | G | C | E | Q | PTEN | ENST00000371953 | 0.56589 | 0.92721 | 0.90649 | 0.80497 | 0.69339 | 0.8147797379 | 0.87718 | 0.7808659673 | 0.153036 | 2.777687838 | 0 |
| 10 | 87864489 | A | C | E | A | PTEN | ENST00000371953 | 0.57125 | 0.93824 | 0.91681 | 0.8808 | 0.76024 | 0.857665645 | 0.98209 | 0.8607254028 | 0.303727 | 3.530785759 | 0 |
| 10 | 87864489 | A | G | E | G | PTEN | ENST00000371953 | 0.62704 | 0.95171 | 0.88585 | 0.92513 | 0.7562 | 0.8737260175 | 0.97687 | 0.8923626542 | 0.298186 | 1.900477683 | 0 |
| 10 | 87864489 | A | T | E | V | PTEN | ENST00000371953 | 0.65422 | 0.95201 | 0.90294 | 0.95621 | 0.87636 | 0.8819446397 | 0.99266 | 0.9393098354 | 0.361657 | 3.560620918 | 0 |
| 10 | 87864490 | G | T | E | D | PTEN | ENST00000371953 | 0.50114 | 0.91059 | 0.91442 | 0.83819 | 0.60243 | 0.831695939 | 0.86133 | 0.7163307667 | 0.137234 | 5.720445546 | 0 |
| 10 | 87864491 | A | C | I | L | PTEN | ENST00000371953 | 0.45364 | 0.92685 | 0.91803 | 0.77362 | 0.5927 | 0.7945640108 | 0.74109 | 0.4026416242 | 0.0750516 | 1.032618364 | 0 |
| 10 | 87864491 | A | G | I | V | PTEN | ENST00000371953 | 0.48234 | 0.89442 | 0.8575 | 0.80712 | 0.54769 | 0.6813782811 | 0.74915 | 0.4234788716 | 0.0589597 | 2.561065712 | 0 |
| 10 | 87864491 | A | T | I | F | PTEN | ENST00000371953 | 0.58938 | 0.98161 | 0.9232 | 0.82434 | 0.56156 | 0.8829649632 | 0.84377 | 0.7834416032 | 0.1141 | 2.383438763 | 0 |
| 10 | 87864492 | T | C | I | T | PTEN | ENST00000371953 | 0.74834 | 0.96121 | 0.97026 | 0.84602 | 0.70671 | 0.8423799482 | 0.90054 | 0.7834008932 | 0.177928 | 3.216900983 | 0 |
| 10 | 87864492 | T | G | I | S | PTEN | ENST00000371953 | 0.82287 | 0.97816 | 0.92832 | 0.91293 | 0.85222 | 0.9508363053 | 0.94437 | 0.9715384841 | 0.218198 | 5.063062094 | 0 |
| 10 | 87864493 | C | G | I | M | PTEN | ENST00000371953 | 0.46649 | 0.9318 | 0.89606 | 0.77008 | 0.52745 | 0.771139013 | 0.65714 | 0.528057158 | 0.031761 | 1.512349297 | 0 |
| 10 | 87864494 | G | A | V | I | PTEN | ENST00000371953 | 0.83898 | 0.99204 | 0.83645 | 0.95942 | 0.92438 | 0.8984354673 | 0.91729 | 0.9834154844 | 0.27174 | 4.790937216 | 0 |
| 10 | 87864494 | G | C | V | L | PTEN | ENST00000371953 | 0.87173 | 0.9966 | 0.94875 | 0.98393 | 0.97252 | 0.9666695108 | 0.95535 | 0.9959199429 | 0.401285 | 6.132773499 | 0 |
| 10 | 87864494 | G | T | V | F | PTEN | ENST00000371953 | 0.894 | 0.99941 | 0.93862 | 0.9916 | 0.9822 | 0.9938352863 | 0.95622 | 0.9983794689 | 0.464713 | 6.577190187 | 0 |
| 10 | 87864497 | A | G | S | G | PTEN | ENST00000371953 | 0.6402 | 0.96506 | 0.86934 | 0.99308 | 0.93709 | 0.8484815355 | 0.97702 | 0.993357718 | 0.37051 | 5.275477337 | 0 |
| 10 | 87864498 | G | A | S | N | PTEN | ENST00000371953 | 0.76251 | 0.99228 | 0.93692 | 0.9798 | 0.88776 | 0.9433345343 | 0.94443 | 0.9927595854 | 0.237548 | 6.362142969 | 0 |
| 10 | 87864498 | G | C | S | T | PTEN | ENST00000371953 | 0.75655 | 0.99282 | 0.90612 | 0.97996 | 0.91314 | 0.873881823 | 0.95252 | 0.9920634627 | 0.31263 | 6.21007677 | 0 |
| 10 | 87864498 | G | T | S | I | PTEN | ENST00000371953 | 0.85581 | 0.99831 | 0.92591 | 0.98667 | 0.97923 | 0.9542802344 | 0.98597 | 0.9974096014 | 0.53901 | 7.931603188 | 1 |
| 10 | 87864500 | A | G | R | G | PTEN | ENST00000371953 | 0.80369 | 0.96273 | 0.92372 | 0.94911 | 0.87539 | 0.9155801848 | 0.99808 | 0.9145017862 | 0.372128 | 3.634001775 | 0 |
| 10 | 87864501 | G | C | R | T | PTEN | ENST00000371953 | 0.79152 | 0.9771 | 0.95346 | 0.94561 | 0.862 | 0.9197729415 | 0.97729 | 0.973392725 | 0.298613 | 6.665554667 | 0 |
| 10 | 87864501 | G | T | R | I | PTEN | ENST00000371953 | 0.83606 | 0.99078 | 0.94275 | 0.96923 | 0.90537 | 0.9054615179 | 0.98772 | 0.9937185049 | 0.353153 | 7.660724808 | 0 |
| 10 | 87864502 | A | C | R | S | PTEN | ENST00000371953 | 0.68411 | 0.94702 | 0.93625 | 0.94977 | 0.82836 | 0.9054167896 | 0.99739 | 0.9342246652 | 0.317885 | 4.634290369 | 0 |
| 10 | 87864502 | A | T | R | S | PTEN | ENST00000371953 | 0.6774 | 0.94702 | 0.93625 | 0.94996 | 0.82836 | 0.9054167896 | 0.99739 | 0.9342246652 | 0.317883 | 4.634290369 | 0 |
| 10 | 87864503 | A | C | N | H | PTEN | ENST00000371953 | 0.50589 | 0.93539 | 0.92491 | 0.94181 | 0.76881 | 0.8898475059 | 0.94057 | 0.8774110675 | 0.16699 | 4.724551588 | 0 |
| 10 | 87864503 | A | T | N | Y | PTEN | ENST00000371953 | 0.67184 | 0.98228 | 0.94413 | 0.9873 | 0.90362 | 0.938626011 | 0.95571 | 0.9729329944 | 0.284042 | 7.069124356 | 0 |
| 10 | 87864504 | A | C | N | T | PTEN | ENST00000371953 | 0.60273 | 0.94385 | 0.90851 | 0.97938 | 0.7727 | 0.9025954351 | 0.92568 | 0.9467374682 | 0.166029 | 6.03886789 | 0 |
| 10 | 87864504 | A | G | N | S | PTEN | ENST00000371953 | 0.55895 | 0.8844 | 0.87691 | 0.93829 | 0.60952 | 0.8425497726 | 0.89251 | 0.8470689058 | 0.0238887 | 4.931603426 | 0 |
| 10 | 87864504 | A | T | N | I | PTEN | ENST00000371953 | 0.79658 | 0.98003 | 0.92556 | 0.98881 | 0.89009 | 0.9564663999 | 0.92906 | 0.9907502532 | 0.239289 | 8.051847937 | 1 |
| 10 | 87864505 | C | A | N | K | PTEN | ENST00000371953 | 0.4043 | 0.94935 | 0.96144 | 0.94778 | 0.59516 | 0.91336174 | 0.69626 | 0.6326789856 | 0.048719 | 1.905974363 | 0 |
| 10 | 87864506 | A | C | K | Q | PTEN | ENST00000371953 | 0.68539 | 0.98348 | 0.89626 | 0.98998 | 0.92356 | 0.9403665871 | 0.96199 | 0.9847491384 | 0.271936 | 5.4640496 | 0 |
| 10 | 87864506 | A | G | K | E | PTEN | ENST00000371953 | 0.69477 | 0.99912 | 0.92466 | 0.98963 | 0.93786 | 0.9637612474 | 0.98934 | 0.9851188064 | 0.440461 | 6.169820997 | 1 |
| 10 | 87864507 | A | C | K | T | PTEN | ENST00000371953 | 0.80961 | 0.99806 | 0.93234 | 0.99486 | 0.94691 | 0.9453849587 | 0.95075 | 0.9933477044 | 0.299967 | 4.30036389 | 0 |
| 10 | 87864507 | A | G | K | R | PTEN | ENST00000371953 | 0.59342 | 0.90703 | 0.8686 | 0.94869 | 0.83283 | 0.852317705 | 0.91837 | 0.8639878035 | 0.174232 | 2.88310965 | 0 |
| 10 | 87864507 | A | T | K | I | PTEN | ENST00000371953 | 0.86068 | 0.99994 | 0.90937 | 0.99741 | 0.95572 | 0.9608782262 | 0.94498 | 0.9956226945 | 0.323356 | 7.269024842 | 0 |
| 10 | 87864508 | A | T | K | N | PTEN | ENST00000371953 | 0.90493 | 0.99787 | 0.94466 | 0.98397 | 0.90185 | 0.9524035592 | 0.96695 | 0.9860972762 | 0.228051 | 5.178768692 | 0 |
| 10 | 87864509 | A | G | R | G | PTEN | ENST00000371953 | 0.82155 | 0.97855 | 0.89737 | 0.98479 | 0.93977 | 0.978616021 | 0.99645 | 0.9862827659 | 0.495357 | 5.65257259 | 0 |
| 10 | 87864509 | A | T | R | W | PTEN | ENST00000371953 | 0.95379 | 0.98981 | 0.9581 | 0.98679 | 0.9729 | 0.9317551048 | 0.97934 | 0.9948506951 | 0.50584 | 7.211788752 | 0 |
| 10 | 87864510 | G | C | R | T | PTEN | ENST00000371953 | 0.83035 | 0.98043 | 0.93241 | 0.9796 | 0.95097 | 0.9553449833 | 0.97538 | 0.9653961658 | 0.453149 | 4.173577591 | 0 |
| 10 | 87864510 | G | T | R | M | PTEN | ENST00000371953 | 0.94836 | 0.9893 | 0.93131 | 0.98249 | 0.97698 | 0.9321159059 | 0.96581 | 0.9931911826 | 0.4498 | 5.823871074 | 0 |
| 10 | 87864511 | G | C | R | S | PTEN | ENST00000371953 | 0.73901 | 0.95413 | 0.91328 | 0.98518 | 0.9211 | 0.9605551715 | 0.98364 | 0.9755109549 | 0.482116 | 5.039237773 | 0 |
| 10 | 87864511 | G | T | R | S | PTEN | ENST00000371953 | 0.73245 | 0.95413 | 0.91328 | 0.98525 | 0.92438 | 0.9605551715 | 0.98364 | 0.9755109549 | 0.482115 | 5.039237773 | 0 |
| 10 | 87864513 | G | T | R | I | PTEN | ENST00000371953 | 0.88055 | 0.99994 | 0.88733 | 0.99091 | 0.97401 | 0.9846109654 | 0.99284 | 0.9988765121 | 0.570971 | 7.277317082 | 1 |
| 10 | 87864514 | A | C | R | S | PTEN | ENST00000371953 | 0.70591 | 0.98952 | 0.88611 | 0.99231 | 0.93515 | 0.9929074628 | 0.99837 | 0.996342361 | 0.526306 | 6.795463104 | 1 |
| 10 | 87864514 | A | T | R | S | PTEN | ENST00000371953 | 0.69909 | 0.98952 | 0.88611 | 0.9923 | 0.93515 | 0.9929074628 | 0.99837 | 0.996342361 | 0.526313 | 6.795463104 | 1 |
| 10 | 87864515 | T | A | Y | N | PTEN | ENST00000371953 | 0.71089 | 0.99912 | 0.96747 | 0.99618 | 0.94431 | 0.9644889915 | 0.98749 | 0.9951443076 | 0.444453 | 7.16248576 | 0 |
| 10 | 87864515 | T | C | Y | H | PTEN | ENST00000371953 | 0.5638 | 0.96763 | 0.97091 | 0.9858 | 0.96008 | 0.9068567991 | 0.99637 | 0.9759083986 | 0.424072 | 3.464166721 | 0 |
| 10 | 87864515 | T | G | Y | D | PTEN | ENST00000371953 | 0.72268 | 0.9997 | 0.96544 | 0.99614 | 0.95207 | 0.9817594661 | 0.99717 | 0.9963687658 | 0.526731 | 5.566336938 | 1 |
| 10 | 87864516 | A | C | Y | S | PTEN | ENST00000371953 | 0.78399 | 0.99691 | 0.93963 | 0.99194 | 0.95863 | 0.965052392 | 0.99726 | 0.9953665733 | 0.531272 | 3.958871871 | 1 |
| 10 | 87864516 | A | G | Y | C | PTEN | ENST00000371953 | 0.8504 | 0.9977 | 0.96652 | 0.99121 | 0.96956 | 0.9636432572 | 0.98741 | 0.9941357374 | 0.537917 | 5.618418808 | 0 |
| 10 | 87864516 | A | T | Y | F | PTEN | ENST00000371953 | 0.54021 | 0.91337 | 0.92471 | 0.93756 | 0.60952 | 0.782777768 | 0.92266 | 0.8246917725 | 0.150571 | 1.582416972 | 0 |
| 10 | 87864518 | C | A | Q | K | PTEN | ENST00000371953 | 0.33361 | 0.96146 | 0.82233 | 0.93837 | 0.73296 | 0.7968436541 | 0.97863 | 0.7046899199 | 0.244751 | 2.085428858 | 0 |
| 10 | 87864518 | C | G | Q | E | PTEN | ENST00000371953 | 0.39121 | 0.94063 | 0.76596 | 0.93639 | 0.78473 | 0.8199503345 | 0.96002 | 0.6986112595 | 0.208026 | 4.221060054 | 0 |
| 10 | 87864519 | A | C | Q | P | PTEN | ENST00000371953 | 0.58738 | 0.9583 | 0.77084 | 0.98161 | 0.92642 | 0.9637869441 | 0.99585 | 0.9282952547 | 0.449281 | 6.084619363 | 0 |
| 10 | 87864519 | A | T | Q | L | PTEN | ENST00000371953 | 0.60047 | 0.92614 | 0.75885 | 0.98341 | 0.85689 | 0.827272545 | 0.99173 | 0.9363764524 | 0.317109 | 5.25390705 | 0 |
| 10 | 87864520 | A | T | Q | H | PTEN | ENST00000371953 | 0.44041 | 0.91839 | 0.81092 | 0.94726 | 0.86753 | 0.8000013661 | 0.9799 | 0.3835706115 | 0.261915 | 5.666064264 | 0 |
| 10 | 87864521 | G | A | E | K | PTEN | ENST00000371953 | 0.60551 | 0.94684 | 0.93176 | 0.96562 | 0.92151 | 0.9215865592 | 0.99457 | 0.9760802984 | 0.437441 | 4.001586677 | 0 |
| 10 | 87864522 | A | C | E | A | PTEN | ENST00000371953 | 0.66422 | 0.9401 | 0.84822 | 0.9766 | 0.91611 | 0.889438217 | 0.97821 | 0.9602661133 | 0.352451 | 4.313646715 | 0 |
| 10 | 87864522 | A | G | E | G | PTEN | ENST00000371953 | 0.85137 | 0.95331 | 0.8 | 0.98313 | 0.94015 | 0.9237279275 | 0.97593 | 0.9886666536 | 0.394748 | 4.720629673 | 0 |
| 10 | 87864522 | A | T | E | V | PTEN | ENST00000371953 | 0.84058 | 0.95441 | 0.82565 | 0.98421 | 0.93901 | 0.9141447473 | 0.99138 | 0.9901325703 | 0.414716 | 5.101412424 | 0 |
| 10 | 87864523 | G | T | E | D | PTEN | ENST00000371953 | 0.60039 | 0.91099 | 0.85057 | 0.94434 | 0.77076 | 0.8368275359 | 0.9185 | 0.8973933458 | 0.187555 | 3.125701857 | 0 |
| 10 | 87864524 | G | A | D | N | PTEN | ENST00000371953 | 0.78535 | 0.94829 | 0.87095 | 0.97166 | 0.87877 | 0.9006881331 | 0.94899 | 0.971816361 | 0.0799206 | 3.76041679 | 0 |
| 10 | 87864524 | G | C | D | H | PTEN | ENST00000371953 | 0.91162 | 0.9756 | 0.89274 | 0.97352 | 0.95462 | 0.9531251201 | 0.96005 | 0.9903531671 | 0.407054 | 4.84580391 | 0 |
| 10 | 87864524 | G | T | D | Y | PTEN | ENST00000371953 | 0.91712 | 0.98906 | 0.88027 | 0.97883 | 0.967 | 0.9696693517 | 0.98492 | 0.9947421551 | 0.521347 | 5.145936449 | 0 |
| 10 | 87864525 | A | C | D | A | PTEN | ENST00000371953 | 0.93073 | 0.98533 | 0.81186 | 0.98785 | 0.95572 | 0.9684230317 | 0.96248 | 0.9869465232 | 0.372348 | 4.642161024 | 0 |
| 10 | 87864525 | A | T | D | V | PTEN | ENST00000371953 | 0.9421 | 0.99941 | 0.80844 | 0.99472 | 0.97067 | 0.984021028 | 0.98533 | 0.9948811531 | 0.533502 | 7.788742989 | 0 |
| 10 | 87864527 | G | A | G | R | PTEN | ENST00000371953 | 0.91561 | 0.98348 | 0.91731 | 0.99411 | 0.98904 | 0.9501559749 | 0.97759 | 0.9922786951 | 0.522662 | 4.938570892 | 0 |
| 10 | 87864528 | G | A | G | E | PTEN | ENST00000371953 | 0.91716 | 0.97521 | 0.90479 | 0.98175 | 0.98071 | 0.9626483963 | 0.98309 | 0.9858499169 | 0.533028 | 3.610255459 | 0 |
| 10 | 87864528 | G | C | G | A | PTEN | ENST00000371953 | 0.74388 | 0.9632 | 0.8376 | 0.98576 | 0.97215 | 0.9410236721 | 0.96854 | 0.986004889 | 0.412872 | 5.233979512 | 0 |
| 10 | 87864528 | G | T | G | V | PTEN | ENST00000371953 | 0.92116 | 0.99565 | 0.84798 | 0.98594 | 0.98404 | 0.978027681 | 0.98867 | 0.9982690811 | 0.584141 | 5.445700238 | 0 |
| 10 | 87864530 | T | A | F | I | PTEN | ENST00000371953 | 0.74802 | 0.98491 | 0.93643 | 0.98438 | 0.89193 | 0.9417363231 | 0.9528 | 0.9903134108 | 0.27347 | 5.849445213 | 0 |
| 10 | 87864530 | T | G | F | V | PTEN | ENST00000371953 | 0.74834 | 0.97662 | 0.92199 | 0.98632 | 0.91652 | 0.9248253275 | 0.97183 | 0.9897750616 | 0.36996 | 5.671957785 | 0 |
| 10 | 87864531 | T | C | F | S | PTEN | ENST00000371953 | 0.89779 | 0.99114 | 0.93677 | 0.98933 | 0.87877 | 0.9600367472 | 0.98959 | 0.9904967546 | 0.367843 | 6.04519127 | 1 |
| 10 | 87864531 | T | G | F | C | PTEN | ENST00000371953 | 0.92534 | 0.9907 | 0.95261 | 0.99504 | 0.94691 | 0.9593319869 | 0.93975 | 0.9950937033 | 0.29 | 6.915055629 | 0 |
| 10 | 87864532 | C | G | F | L | PTEN | ENST00000371953 | 0.71145 | 0.96093 | 0.95634 | 0.98001 | 0.79456 | 0.8949886187 | 0.92335 | 0.9872125983 | 0.198742 | 5.736516908 | 0 |
| 10 | 87864533 | G | A | D | N | PTEN | ENST00000371953 | 0.61679 | 0.9303 | 0.85816 | 0.98009 | 0.79273 | 0.9084830757 | 0.91742 | 0.8890939951 | 0.0925965 | 3.108547261 | 0 |
| 10 | 87864533 | G | C | D | H | PTEN | ENST00000371953 | 0.93268 | 0.99351 | 0.88426 | 0.98917 | 0.99525 | 0.9294226497 | 0.99693 | 0.9916760921 | 0.559866 | 5.551762526 | 0 |
| 10 | 87864533 | G | T | D | Y | PTEN | ENST00000371953 | 0.93562 | 0.99467 | 0.86594 | 0.98974 | 0.99296 | 0.9692767946 | 0.9977 | 0.9952215552 | 0.560333 | 7.344846663 | 0 |
| 10 | 87864534 | A | C | D | A | PTEN | ENST00000371953 | 0.85763 | 0.9856 | 0.80408 | 0.98953 | 0.95863 | 0.9563570304 | 0.9945 | 0.9917631149 | 0.562375 | 5.948624696 | 0 |
| 10 | 87864534 | A | T | D | V | PTEN | ENST00000371953 | 0.86636 | 0.99691 | 0.79437 | 0.99222 | 0.97438 | 0.9692419676 | 0.99813 | 0.9955130219 | 0.571489 | 5.899530829 | 0 |
| 10 | 87864536 | T | A | L | I | PTEN | ENST00000371953 | 0.84906 | 0.99797 | 0.89397 | 0.98758 | 0.94618 | 0.9050512788 | 0.92652 | 0.9884294271 | 0.355273 | 4.864864432 | 0 |
| 10 | 87864536 | T | G | L | V | PTEN | ENST00000371953 | 0.84446 | 0.99707 | 0.83104 | 0.99433 | 0.96773 | 0.943689882 | 0.98844 | 0.9903991818 | 0.522326 | 5.136437293 | 1 |
| 10 | 87864537 | T | C | L | S | PTEN | ENST00000371953 | 0.9519 | 0.99941 | 0.89996 | 0.99744 | 0.99603 | 0.997194138 | 0.99614 | 0.9980772734 | 0.581197 | 7.105480155 | 1 |
| 10 | 87864539 | G | C | D | H | PTEN | ENST00000371953 | 0.92048 | 0.99863 | 0.95281 | 0.99309 | 0.9986 | 0.9839894373 | 0.99758 | 0.9989632368 | 0.574627 | 6.29863792 | 1 |
| 10 | 87864539 | G | T | D | Y | PTEN | ENST00000371953 | 0.93051 | 0.99898 | 0.94328 | 0.99391 | 0.9997 | 0.9944188338 | 0.99845 | 0.9997455478 | 0.575747 | 6.405711083 | 1 |
| 10 | 87864540 | A | C | D | A | PTEN | ENST00000371953 | 0.87228 | 0.99884 | 0.90024 | 0.99239 | 0.99414 | 0.9915776904 | 0.99692 | 0.9984887838 | 0.572951 | 6.405711083 | 1 |
| 10 | 87864540 | A | G | D | G | PTEN | ENST00000371953 | 0.86297 | 0.9977 | 0.90992 | 0.99391 | 0.99138 | 0.9935395257 | 0.99789 | 0.9975899458 | 0.571733 | 5.230954909 | 1 |
| 10 | 87864540 | A | T | D | V | PTEN | ENST00000371953 | 0.94471 | 0.99964 | 0.89594 | 0.99675 | 0.99993 | 0.9933261573 | 0.99812 | 0.9994446635 | 0.573818 | 6.405711083 | 1 |
| 10 | 87864541 | C | A | D | E | PTEN | ENST00000371953 | 0.49596 | 0.97582 | 0.92442 | 0.98989 | 0.96298 | 0.979922479 | 0.9966 | 0.9956071973 | 0.537859 | 4.567480064 | 0 |
| 10 | 87864542 | T | A | L | M | PTEN | ENST00000371953 | 0.80692 | 0.97893 | 0.90616 | 0.97693 | 0.92151 | 0.8589590343 | 0.96854 | 0.8559685349 | 0.322017 | 3.986818582 | 0 |
| 10 | 87864543 | T | G | L | W | PTEN | ENST00000371953 | 0.94328 | 0.99994 | 0.96298 | 0.99495 | 0.99072 | 0.9852802239 | 0.9942 | 0.9927874207 | 0.56372 | 7.145738739 | 1 |
| 10 | 87864545 | A | C | T | P | PTEN | ENST00000371953 | 0.65664 | 0.99499 | 0.89962 | 0.99754 | 0.98693 | 0.9925753605 | 0.99728 | 0.9886465669 | 0.559022 | 7.491812387 | 1 |
| 10 | 87864545 | A | G | T | A | PTEN | ENST00000371953 | 0.47911 | 0.928 | 0.89542 | 0.99312 | 0.95316 | 0.9652759923 | 0.99728 | 0.9256635904 | 0.456619 | 3.386005864 | 0 |

| | | | | | | | | | | | | | | | | | |
|---|---|---|---|---|---|---|---|---|---|---|---|---|---|---|---|---|---|
| 10 | 87864545 | A | T | T | S | PTEN | ENST00000371953 | 0.43502 | 0.90959 | 0.90574 | 0.9802 | 0.862 | 0.906454919 | 0.99168 | 0.8193434477 | 0.323651 | 2.231704048 | 0 |
| 10 | 87864546 | C | A | T | N | PTEN | ENST00000371953 | 0.60889 | 0.98468 | 0.94996 | 0.98331 | 0.86553 | 0.989542841 | 0.92551 | 0.9790964723 | 0.249795 | 5.163666015 | 0 |
| 10 | 87864546 | C | G | T | S | PTEN | ENST00000371953 | 0.46289 | 0.90959 | 0.90574 | 0.97523 | 0.84694 | 0.906454919 | 0.98483 | 0.8522011638 | 0.310933 | 2.231704048 | 0 |
| 10 | 87864546 | C | T | T | I | PTEN | ENST00000371953 | 0.64883 | 0.94843 | 0.96865 | 0.98879 | 0.9917 | 0.9846912643 | 0.99414 | 0.988258481 | 0.537829 | 5.750690181 | 0 |
| 10 | 87864548 | T | A | Y | N | PTEN | ENST00000371953 | 0.84165 | 0.99941 | 0.94249 | 0.99664 | 0.9986 | 0.9931868771 | 0.98452 | 0.9986227751 | 0.489042 | 6.315303753 | 1 |
| 10 | 87864548 | T | G | Y | D | PTEN | ENST00000371953 | 0.81818 | 0.99441 | 0.94001 | 0.99536 | 0.97773 | 0.9967922028 | 0.99683 | 0.9976174235 | 0.550746 | 3.140030351 | 1 |
| 10 | 87894025 | A | C | Y | S | PTEN | ENST00000371953 | 0.76283 | 0.98736 | 0.89334 | 0.999 | 0.99552 | 0.9915090418 | 0.99589 | 0.9985634685 | 0.571835 | 5.130528918 | 0 |
| 10 | 87894025 | A | T | Y | F | PTEN | ENST00000371953 | 0.57139 | 0.9234 | 0.88011 | 0.98986 | 0.93631 | 0.9279800099 | 0.93113 | 0.8981945515 | 0.257158 | 3.730890928 | 0 |
| 10 | 87894027 | A | C | I | L | PTEN | ENST00000371953 | 0.77505 | 0.94242 | 0.85555 | 0.98663 | 0.97624 | 0.9630580295 | 0.96839 | 0.9719469547 | 0.399005 | 5.225661269 | 0 |
| 10 | 87894027 | A | G | I | V | PTEN | ENST00000371953 | 0.70423 | 0.93227 | 0.77204 | 0.97116 | 0.93319 | 0.9032506747 | 0.93004 | 0.9156270623 | 0.26577 | 4.172506226 | 0 |
| 10 | 87894028 | T | A | I | N | PTEN | ENST00000371953 | 0.92715 | 0.99994 | 0.92989 | 0.98932 | 0.99806 | 0.9987729733 | 0.98209 | 0.9988589287 | 0.535331 | 7.918098726 | 1 |
| 10 | 87894029 | T | G | I | M | PTEN | ENST00000371953 | 0.78994 | 0.99381 | 0.82099 | 0.98349 | 0.97885 | 0.9712470716 | 0.99204 | 0.9869861007 | 0.543292 | 6.97973929 | 0 |
| 10 | 87894030 | T | C | Y | H | PTEN | ENST00000371953 | 0.65335 | 0.97099 | 0.96611 | 0.93463 | 0.96444 | 0.9545158568 | 0.96922 | 0.9179483056 | 0.456957 | 4.810822079 | 0 |
| 10 | 87894030 | T | G | Y | D | PTEN | ENST00000371953 | 0.70792 | 0.99565 | 0.96178 | 0.96564 | 0.97624 | 0.9920162979 | 0.99457 | 0.9661349654 | 0.504675 | 4.749105174 | 0 |
| 10 | 87894031 | A | C | Y | S | PTEN | ENST00000371953 | 0.7901 | 0.97686 | 0.92558 | 0.97351 | 0.95826 | 0.9718244773 | 0.99051 | 0.8913347721 | 0.501919 | 4.011547522 | 0 |
| 10 | 87894031 | A | G | Y | C | PTEN | ENST00000371953 | 0.63927 | 0.96067 | 0.96527 | 0.95987 | 0.9182 | 0.9857804429 | 0.98889 | 0.9608865976 | 0.491331 | 2.558761045 | 0 |
| 10 | 87894031 | A | T | Y | F | PTEN | ENST00000371953 | 0.68043 | 0.92565 | 0.91006 | 0.9461 | 0.91611 | 0.9307999046 | 0.90894 | 0.6505821943 | 0.168873 | 6.02351746 | 0 |
| 10 | 87894033 | C | A | P | T | PTEN | ENST00000371953 | 0.5508 | 0.97893 | 0.94844 | 0.98229 | 0.95608 | 0.9544588861 | 0.90577 | 0.9862543344 | 0.345897 | 5.46014578 | 0 |
| 10 | 87894033 | C | G | P | A | PTEN | ENST00000371953 | 0.53522 | 0.9813 | 0.92834 | 0.98686 | 0.95717 | 0.9328643204 | 0.86158 | 0.9928032756 | 0.258474 | 5.349656097 | 0 |
| 10 | 87894033 | C | T | P | S | PTEN | ENST00000371953 | 0.57568 | 0.95467 | 0.93073 | 0.97069 | 0.92356 | 0.9464246718 | 0.87049 | 0.9833940864 | 0.237286 | 4.498470145 | 0 |
| 10 | 87894034 | C | A | P | Q | PTEN | ENST00000371953 | 0.82241 | 0.97243 | 0.9369 | 0.98843 | 0.95097 | 0.9416440985 | 0.8938 | 0.9881958961 | 0.308014 | 5.721915311 | 0 |
| 10 | 87894034 | C | G | P | R | PTEN | ENST00000371953 | 0.71016 | 0.96597 | 0.96485 | 0.97567 | 0.96008 | 0.9655176878 | 0.9202 | 0.9890385866 | 0.375926 | 4.549238559 | 0 |
| 10 | 87894034 | C | T | P | L | PTEN | ENST00000371953 | 0.858 | 0.97205 | 0.96631 | 0.99396 | 0.99471 | 0.9599018186 | 0.97506 | 0.988082281 | 0.525583 | 6.831053709 | 0 |
| 10 | 87894036 | A | C | N | H | PTEN | ENST00000371953 | 0.9066 | 0.99441 | 0.8435 | 0.97534 | 0.9755 | 0.943997706 | 0.97277 | 0.9920177728 | 0.284774 | 4.876449421 | 0 |
| 10 | 87894036 | A | G | N | D | PTEN | ENST00000371953 | 0.9204 | 0.95358 | 0.88888 | 0.97778 | 0.9796 | 0.9611862017 | 0.96575 | 0.9940469265 | 0.27005 | 6.254941999 | 0 |
| 10 | 87894036 | A | T | N | Y | PTEN | ENST00000371953 | 0.93098 | 0.9975 | 0.88202 | 0.99074 | 0.99628 | 0.9635555081 | 0.98684 | 0.9981317603 | 0.550775 | 7.821140163 | 0 |
| 10 | 87894037 | A | C | N | T | PTEN | ENST00000371953 | 0.89243 | 0.93946 | 0.8114 | 0.98583 | 0.9781 | 0.9570210834 | 0.91737 | 0.982390523 | 0.261554 | 4.945407612 | 0 |
| 10 | 87894037 | A | G | N | S | PTEN | ENST00000371953 | 0.7185 | 0.88881 | 0.75653 | 0.98606 | 0.96553 | 0.9159565334 | 0.94805 | 0.9767223001 | 0.218213 | 4.736988339 | 0 |
| 10 | 87894037 | A | T | N | I | PTEN | ENST00000371953 | 0.92202 | 0.99616 | 0.8438 | 0.9966 | 0.97773 | 0.9769947571 | 0.95686 | 0.9977373647 | 0.398947 | 7.821140163 | 1 |
| 10 | 87894038 | C | A | N | K | PTEN | ENST00000371953 | 0.67485 | 0.95189 | 0.90885 | 0.92273 | 0.90318 | 0.9467507699 | 0.93684 | 0.9830394387 | 0.190955 | 4.076272947 | 0 |
| 10 | 87894039 | A | C | I | L | PTEN | ENST00000371953 | 0.7438 | 0.93432 | 0.8895 | 0.93089 | 0.93123 | 0.9440921064 | 0.95318 | 0.8832771778 | 0.343942 | 2.432004342 | 0 |
| 10 | 87894039 | A | G | I | V | PTEN | ENST00000371953 | 0.70487 | 0.92085 | 0.80764 | 0.92617 | 0.90711 | 0.8580822738 | 0.88935 | 0.8549491763 | 0.226147 | 3.031575176 | 0 |
| 10 | 87894040 | T | A | I | N | PTEN | ENST00000371953 | 0.9338 | 0.99994 | 0.95134 | 0.98405 | 0.99326 | 0.9978815605 | 0.98069 | 0.9987502098 | 0.527924 | 7.235393488 | 1 |
| 10 | 87894040 | T | G | I | S | PTEN | ENST00000371953 | 0.93678 | 0.99941 | 0.9069 | 0.98385 | 0.99578 | 0.998611563 | 0.99714 | 0.9983391762 | 0.578835 | 7.235393488 | 1 |
| 10 | 87894041 | T | G | I | M | PTEN | ENST00000371953 | 0.75089 | 0.99448 | 0.85743 | 0.96006 | 0.92397 | 0.9453832373 | 0.99188 | 0.9606838822 | 0.524576 | 4.480788948 | 0 |
| 10 | 87894042 | A | C | I | L | PTEN | ENST00000371953 | 0.48666 | 0.96774 | 0.91664 | 0.95557 | 0.91694 | 0.9459423964 | 0.99069 | 0.9233031273 | 0.38045 | 3.67369616 | 0 |
| 10 | 87894042 | A | G | I | V | PTEN | ENST00000371953 | 0.4457 | 0.95014 | 0.84428 | 0.95379 | 0.91442 | 0.8660335314 | 0.93792 | 0.8963773251 | 0.296835 | 3.214615274 | 0 |
| 10 | 87894042 | A | T | I | F | PTEN | ENST00000371953 | 0.81889 | 0.99982 | 0.92606 | 0.98615 | 0.99744 | 0.9877588586 | 0.99708 | 0.9962269068 | 0.580704 | 6.705545007 | 1 |
| 10 | 87894043 | T | A | I | N | PTEN | ENST00000371953 | 0.94125 | 0.99994 | 0.96504 | 0.99433 | 0.99105 | 0.9979872118 | 0.97514 | 0.9987502098 | 0.453419 | 7.469790189 | 1 |
| 10 | 87894043 | T | C | I | T | PTEN | ENST00000371953 | 0.91383 | 0.99898 | 0.9721 | 0.9933 | 0.997 | 0.9889295343 | 0.99717 | 0.998123467 | 0.576726 | 7.469790189 | 0 |
| 10 | 87894043 | T | G | I | S | PTEN | ENST00000371953 | 0.94288 | 0.9997 | 0.93005 | 0.99362 | 0.98972 | 0.9986720454 | 0.99691 | 0.9986737967 | 0.582812 | 7.425548957 | 1 |
| 10 | 87894044 | T | G | I | M | PTEN | ENST00000371953 | 0.76816 | 0.99679 | 0.89238 | 0.98399 | 0.96919 | 0.9472222625 | 0.9923 | 0.9799619913 | 0.527237 | 5.199782954 | 0 |
| 10 | 87894045 | G | A | A | T | PTEN | ENST00000371953 | 0.83097 | 0.99523 | 0.8953 | 0.99181 | 0.98869 | 0.9681090108 | 0.96851 | 0.9946742058 | 0.407011 | 5.50058622 | 0 |
| 10 | 87894045 | G | C | A | P | PTEN | ENST00000371953 | 0.84241 | 0.99941 | 0.93836 | 0.99156 | 0.99443 | 0.9986794542 | 0.96897 | 0.9978535771 | 0.531341 | 7.811876813 | 1 |
| 10 | 87894046 | C | A | A | D | PTEN | ENST00000371953 | 0.81983 | 0.99982 | 0.95665 | 0.994 | 0.99652 | 0.9990292109 | 0.98625 | 0.9991320968 | 0.579854 | 7.837105361 | 1 |
| 10 | 87894046 | C | G | A | G | PTEN | ENST00000371953 | 0.80937 | 0.99707 | 0.88216 | 0.9898 | 0.9755 | 0.963169988 | 0.94833 | 0.9950187541 | 0.353676 | 4.792882395 | 0 |
| 10 | 87894048 | A | T | M | L | PTEN | ENST00000371953 | 0.68763 | 0.9851 | 0.90219 | 0.97853 | 0.98657 | 0.9635516247 | 0.9924 | 0.9857136011 | 0.497121 | 5.337174242 | 0 |
| 10 | 87894049 | T | A | M | K | PTEN | ENST00000371953 | 0.77402 | 0.99982 | 0.94541 | 0.99114 | 0.9986 | 0.998720244 | 0.99711 | 0.9992274046 | 0.582714 | 7.981319897 | 1 |
| 10 | 87894049 | T | C | M | T | PTEN | ENST00000371953 | 0.72564 | 0.9975 | 0.96336 | 0.98366 | 0.99072 | 0.9926944111 | 0.99819 | 0.9951183796 | 0.546932 | 5.187579124 | 1 |
| 10 | 87894049 | T | G | M | R | PTEN | ENST00000371953 | 0.79437 | 0.99982 | 0.93384 | 0.99087 | 0.99976 | 0.998779845 | 0.99714 | 0.9983659387 | 0.578261 | 7.915688765 | 1 |
| 10 | 87894050 | G | T | M | I | PTEN | ENST00000371953 | 0.76459 | 0.99696 | 0.91076 | 0.97564 | 0.98972 | 0.9785619146 | 0.99569 | 0.995539248 | 0.55305 | 5.656352917 | 0 |
| 10 | 87894051 | G | A | G | R | PTEN | ENST00000371953 | 0.85624 | 0.99707 | 0.9887 | 0.99369 | 0.99995 | 0.990104837 | 0.99687 | 0.9974539042 | 0.595039 | 7.043303201 | 1 |
| 10 | 87894052 | G | C | G | A | PTEN | ENST00000371953 | 0.86803 | 0.99616 | 0.96758 | 0.9754 | 0.98477 | 0.9586905459 | 0.97362 | 0.9968690276 | 0.435396 | 4.335885945 | 0 |
| 10 | 87894054 | T | G | F | V | PTEN | ENST00000371953 | 0.80447 | 0.99912 | 0.88569 | 0.98229 | 0.99722 | 0.9747580794 | 0.98406 | 0.9961601496 | 0.56961 | 5.110182015 | 0 |
| 10 | 87894055 | T | A | F | Y | PTEN | ENST00000371953 | 0.75049 | 0.97175 | 0.91558 | 0.96781 | 0.95097 | 0.9119883685 | 0.94356 | 0.8520365357 | 0.330389 | 3.27902329 | 0 |
| 10 | 87894055 | T | C | F | S | PTEN | ENST00000371953 | 0.939 | 0.99994 | 0.90294 | 0.98913 | 0.99765 | 0.9849048668 | 0.99119 | 0.9992796779 | 0.574326 | 6.930132049 | 0 |
| 10 | 87894055 | T | G | F | C | PTEN | ENST00000371953 | 0.93886 | 0.9997 | 0.92415 | 0.99009 | 0.99443 | 0.9857016175 | 0.9859 | 0.9992796779 | 0.569282 | 6.331779361 | 0 |
| 10 | 87894056 | T | G | F | L | PTEN | ENST00000371953 | 0.83151 | 0.99114 | 0.92586 | 0.97086 | 0.98404 | 0.9484832211 | 0.97589 | 0.9959942102 | 0.470354 | 4.700213682 | 0 |
| 10 | 87894057 | C | A | P | T | PTEN | ENST00000371953 | 0.56985 | 0.99831 | 0.9261 | 0.99413 | 0.99138 | 0.9926370548 | 0.98973 | 0.9993609786 | 0.563571 | 6.088475567 | 1 |
| 10 | 87894058 | C | A | P | H | PTEN | ENST00000371953 | 0.73942 | 0.99863 | 0.94195 | 0.99339 | 0.99072 | 0.9942125713 | 0.99136 | 0.9991400242 | 0.568372 | 5.701143769 | 1 |
| 10 | 87894058 | C | G | P | R | PTEN | ENST00000371953 | 0.74029 | 0.9977 | 0.94521 | 0.99285 | 0.98938 | 0.997291871 | 0.99009 | 0.9993609786 | 0.563892 | 6.317572843 | 1 |
| 10 | 87894058 | C | T | P | L | PTEN | ENST00000371953 | 0.76195 | 0.99721 | 0.9514 | 0.99251 | 0.98109 | 0.9952959523 | 0.98663 | 0.9994702935 | 0.550002 | 5.582644051 | 1 |
| 10 | 87894060 | G | T | A | S | PTEN | ENST00000371953 | 0.76816 | 0.94913 | 0.79289 | 0.94104 | 0.9045 | 0.9155998361 | 0.91805 | 0.9407246113 | 0.293875 | 3.525648876 | 0 |
| 10 | 87894061 | C | A | A | E | PTEN | ENST00000371953 | 0.70895 | 0.99842 | 0.88842 | 0.98737 | 0.97067 | 0.9962996129 | 0.98844 | 0.9982541203 | 0.565099 | 6.278184638 | 1 |
| 10 | 87894063 | G | A | E | K | PTEN | ENST00000371953 | 0.74754 | 0.95395 | 0.9286 | 0.96021 | 0.96008 | 0.9385480256 | 0.98802 | 0.9811717272 | 0.484918 | 4.03454022 | 0 |
| 10 | 87894063 | G | C | E | Q | PTEN | ENST00000371953 | 0.83283 | 0.93642 | 0.84372 | 0.9442 | 0.9045 | 0.8939273539 | 0.95165 | 0.9712679386 | 0.299665 | 3.737755842 | 0 |
| 10 | 87894064 | A | C | E | A | PTEN | ENST00000371953 | 0.71762 | 0.9401 | 0.85052 | 0.93171 | 0.96737 | 0.9237144732 | 0.98477 | 0.9863562584 | 0.44676 | 4.20365587 | 0 |
| 10 | 87894064 | A | G | E | G | PTEN | ENST00000371953 | 0.89955 | 0.95316 | 0.81378 | 0.95198 | 0.97401 | 0.9169260143 | 0.98737 | 0.9883648157 | 0.481655 | 3.859215792 | 0 |
| 10 | 87894065 | A | T | E | D | PTEN | ENST00000371953 | 0.3892 | 0.9108 | 0.85756 | 0.83224 | 0.8153 | 0.8573920558 | 0.95354 | 0.239848718 | 0.129747 | 1.792530492 | 0 |
| 10 | 87894066 | A | G | R | G | PTEN | ENST00000371953 | 0.83352 | 0.96288 | 0.86218 | 0.93851 | 0.89558 | 0.9140882447 | 0.99534 | 0.9561590552 | 0.459154 | 4.541147374 | 0 |
| 10 | 87894067 | G | A | R | K | PTEN | ENST00000371953 | 0.39958 | 0.93076 | 0.92043 | 0.9143 | 0.79577 | 0.8518725701 | 0.94184 | 0.711699605 | 0.224443 | 1.772181376 | 0 |
| 10 | 87894067 | G | C | R | T | PTEN | ENST00000371953 | 0.82178 | 0.97972 | 0.90472 | 0.94941 | 0.8887 | 0.9019167572 | 0.97289 | 0.9658820033 | 0.350841 | 3.752816624 | 0 |
| 10 | 87894068 | A | C | R | S | PTEN | ENST00000371953 | 0.82965 | 0.94799 | 0.87195 | 0.94109 | 0.89739 | 0.9060387721 | 0.99663 | 0.9616332054 | 0.471761 | 3.427828056 | 0 |
| 10 | 87894068 | A | T | R | S | PTEN | ENST00000371953 | 0.82311 | 0.94799 | 0.87195 | 0.94109 | 0.89739 | 0.9060387721 | 0.99662 | 0.9616332054 | 0.471758 | 3.427828056 | 0 |
| 10 | 87894069 | C | A | L | I | PTEN | ENST00000371953 | 0.40372 | 0.94989 | 0.89119 | 0.92574 | 0.77782 | 0.7715151964 | 0.81018 | 0.8030474782 | 0.186955 | 3.166092499 | 0 |
| 10 | 87894069 | C | G | L | V | PTEN | ENST00000371953 | 0.41327 | 0.9404 | 0.84586 | 0.94018 | 0.7872 | 0.8453119804 | 0.86666 | 0.7743067145 | 0.212476 | 3.752856853 | 0 |
| 10 | 87894069 | C | T | L | F | PTEN | ENST00000371953 | 0.61052 | 0.94631 | 0.87994 | 0.95678 | 0.86251 | 0.8729299415 | 0.87439 | 0.9667819142 | 0.296113 | 4.14967869 | 0 |
| 10 | 87894070 | T | A | L | H | PTEN | ENST00000371953 | 0.90804 | 0.99889 | 0.94062 | 0.98538 | 0.98367 | 0.9375642367 | 0.98663 | 0.9985041618 | 0.569477 | 6.03538899 | 0 |
| 10 | 87894070 | T | C | L | P | PTEN | ENST00000371953 | 0.91934 | 0.99707 | 0.95383 | 0.98063 | 0.99006 | 0.9830349256 | 0.99607 | 0.9950180054 | 0.541735 | 6.221728472 | 1 |
| 10 | 87894072 | G | A | E | K | PTEN | ENST00000371953 | 0.8712 | 0.97228 | 0.72359 | 0.98619 | 0.99443 | 0.9665085275 | 0.99865 | 0.9960856438 | 0.575332 | 6.150565402 | 0 |
| 10 | 87894072 | G | C | E | Q | PTEN | ENST00000371953 | 0.81206 | 0.96033 | 0.57524 | 0.97797 | 0.96993 | 0.9269297757 | 0.9776 | 0.9893375635 | 0.409244 | 4.796576101 | 0 |
| 10 | 87894073 | A | C | E | A | PTEN | ENST00000371953 | 0.91283 | 0.9893 | 0.58922 | 0.98744 | 0.99265 | 0.9481069138 | 0.99607 | 0.9980590069 | 0.578257 | 6.124133315 | 0 |
| 10 | 87894073 | A | G | E | G | PTEN | ENST00000371953 | 0.93488 | 0.98843 | 0.53113 | 0.99204 | 0.99578 | 0.9531870209 | 0.99101 | 0.9992909431 | 0.592518 | 6.028709488 | 0 |
| 10 | 87894073 | A | T | E | V | PTEN | ENST00000371953 | 0.92785 | 0.99806 | 0.57356 | 0.99364 | 0.99786 | 0.9605376135 | 0.9976 | 0.9987342954 | 0.59017 | 7.261271468 | 0 |
| 10 | 87894074 | A | T | E | D | PTEN | ENST00000371953 | 0.77837 | 0.99134 | 0.59231 | 0.97662 | 0.95863 | 0.9028821756 | 0.9756 | 0.9939693213 | 0.373796 | 5.239363779 | 0 |
| 10 | 87894075 | G | C | G | R | PTEN | ENST00000371953 | 0.78764 | 0.96209 | 0.93813 | 0.97221 | 0.94356 | 0.972501117 | 0.98662 | 0.9892815351 | 0.515637 | 3.878659993 | 0 |
| 10 | 87894075 | G | T | G | C | PTEN | ENST00000371953 | 0.81747 | 0.99616 | 0.91417 | 0.98239 | 0.96883 | 0.9552471263 | 0.97808 | 0.9988823533 | 0.542388 | 7.909644552 | 0 |
| 10 | 87894076 | G | C | G | A | PTEN | ENST00000371953 | 0.72668 | 0.94463 | 0.86347 | 0.95257 | 0.91778 | 0.9106286717 | 0.96229 | 0.9799474478 | 0.366855 | 4.267662594 | 0 |
| 10 | 87894076 | G | T | G | V | PTEN | ENST00000371953 | 0.78399 | 0.98196 | 0.87286 | 0.9755 | 0.96846 | 0.9516477635 | 0.98343 | 0.9968139529 | 0.555756 | 6.515769443 | 0 |
| 10 | 87894078 | G | A | V | I | PTEN | ENST00000371953 | 0.60332 | 0.89023 | 0.71622 | 0.90713 | 0.80772 | 0.8351405567 | 0.82435 | 0.5773968697 | 0.198508 | 3.521574899 | 0 |
| 10 | 87894078 | G | C | V | L | PTEN | ENST00000371953 | 0.6628 | 0.92711 | 0.87105 | 0.91815 | 0.91484 | 0.9341562913 | 0.95771 | 0.9270121455 | 0.354489 | 3.456629989 | 0 |
| 10 | 87894078 | G | T | V | L | PTEN | ENST00000371953 | 0.65633 | 0.92711 | 0.87105 | 0.91778 | 0.91484 | 0.9341562913 | 0.95797 | 0.9270121455 | 0.354494 | 3.456629989 | 0 |
| 10 | 87894079 | T | C | V | A | PTEN | ENST00000371953 | 0.77006 | 0.95585 | 0.89095 | 0.96742 | 0.95389 | 0.9307078728 | 0.98816 | 0.9790938497 | 0.42146 | 5.486726159 | 0 |
| 10 | 87894079 | T | G | V | G | PTEN | ENST00000371953 | 0.84271 | 0.98449 | 0.77049 | 0.97948 | 0.97773 | 0.969987435 | 0.99258 | 0.9933100939 | 0.554228 | 5.911464306 | 0 |
| 10 | 87894081 | T | C | Y | H | PTEN | ENST00000371953 | 0.92345 | 0.99815 | 0.97117 | 0.98895 | 0.99525 | 0.9798560751 | 0.99072 | 0.9954297543 | 0.584741 | 5.999556293 | 1 |
| 10 | 87894081 | T | G | Y | D | PTEN | ENST00000371953 | 0.9318 | 0.99994 | 0.96817 | 0.99378 | 0.99824 | 0.9960303341 | 0.99743 | 0.9998886585 | 0.585474 | 7.328305178 | 1 |
| 10 | 87894082 | A | C | Y | S | PTEN | ENST00000371953 | 0.92573 | 0.99964 | 0.93333 | 0.99198 | 0.99953 | 0.9905521477 | 0.99731 | 0.9997600913 | 0.593212 | 7.201114737 | 1 |
| 10 | 87894082 | A | G | Y | C | PTEN | ENST00000371953 | 0.93712 | 0.99912 | 0.96901 | 0.9907 | 0.99932 | 0.9896944464 | 0.99386 | 0.9992465973 | 0.594295 | 6.959730636 | 0 |
| 10 | 87894082 | A | T | Y | F | PTEN | ENST00000371953 | 0.74269 | 0.97175 | 0.91923 | 0.97135 | 0.96773 | 0.9320231856 | 0.93734 | 0.9769774675 | 0.272577 | 3.521980545 | 0 |
| 10 | 87894084 | A | G | R | G | PTEN | ENST00000371953 | 0.90364 | 0.99941 | 0.93039 | 0.99235 | 0.99326 | 0.9918363655 | 0.99555 | 0.9982985854 | 0.566617 | 5.741642698 | 1 |
| 10 | 87894085 | G | C | R | T | PTEN | ENST00000371953 | 0.91595 | 0.99982 | 0.95413 | 0.99197 | 0.99843 | 0.9913297567 | 0.98969 | 0.9984542131 | 0.590925 | 6.634335501 | 1 |

| | | | | | | | | | | | | | | | | | |
|---|---|---|---|---|---|---|---|---|---|---|---|---|---|---|---|---|---|
| 10 | 87894085 | G | T | R | M | PTEN | ENST00000371953 | 0.93327 | 0.99834 | 0.9514 | 0.99203 | 0.99414 | 0.9807501457 | 0.98738 | 0.9997466207 | 0.589796 | 7.097356873 | 0 |
| 10 | 87894086 | G | C | R | S | PTEN | ENST00000371953 | 0.82545 | 0.99842 | 0.93561 | 0.98377 | 0.97067 | 0.9909646315 | 0.99825 | 0.9972735047 | 0.551899 | 6.030668955 | 1 |
| 10 | 87894086 | G | T | R | S | PTEN | ENST00000371953 | 0.81904 | 0.99842 | 0.93561 | 0.98337 | 0.97067 | 0.9909646315 | 0.99824 | 0.9976655245 | 0.551894 | 6.030668955 | 1 |
| 10 | 87894087 | A | T | N | Y | PTEN | ENST00000371953 | 0.79318 | 0.99889 | 0.98062 | 0.99558 | 0.98764 | 0.9967334555 | 0.98139 | 0.9996747971 | 0.507147 | 6.157962589 | 1 |
| 10 | 87894088 | A | C | N | T | PTEN | ENST00000371953 | 0.88227 | 0.99707 | 0.96249 | 0.99217 | 0.9796 | 0.9913575742 | 0.97463 | 0.9972828526 | 0.321734 | 6.116863449 | 1 |
| 10 | 87894088 | A | G | N | S | PTEN | ENST00000371953 | 0.7023 | 0.98613 | 0.94266 | 0.9891 | 0.97923 | 0.9859746145 | 0.98294 | 0.998139739 | 0.271757 | 5.680807038 | 1 |
| 10 | 87894088 | A | T | N | I | PTEN | ENST00000371953 | 0.91204 | 0.99912 | 0.97066 | 0.99327 | 0.9833 | 0.9972818457 | 0.9766 | 0.999907136 | 0.464122 | 6.974349607 | 1 |
| 10 | 87894089 | C | A | N | K | PTEN | ENST00000371953 | 0.79199 | 0.99842 | 0.98713 | 0.98043 | 0.96262 | 0.996425246 | 0.98047 | 0.9982738495 | 0.270623 | 6.445561433 | 1 |
| 10 | 87894090 | A | G | N | D | PTEN | ENST00000371953 | 0.68795 | 0.94989 | 0.9662 | 0.95108 | 0.90052 | 0.9376223367 | 0.94967 | 0.9559425116 | 0.207458 | 3.54794416 | 0 |
| 10 | 87894091 | A | G | N | S | PTEN | ENST00000371953 | 0.56172 | 0.88762 | 0.8947 | 0.95641 | 0.82103 | 0.8700011592 | 0.91986 | 0.9804539084 | 0.0911102 | 4.154931723 | 0 |
| 10 | 87894091 | A | T | N | I | PTEN | ENST00000371953 | 0.93044 | 0.99499 | 0.94661 | 0.99366 | 0.98764 | 0.9748458009 | 0.96646 | 0.995562911 | 0.425794 | 7.566458133 | 1 |
| 10 | 87894092 | T | A | N | K | PTEN | ENST00000371953 | 0.64851 | 0.95356 | 0.97551 | 0.94376 | 0.86047 | 0.9518585567 | 0.93952 | 0.8283479214 | 0.136174 | 4.167520783 | 0 |
| 10 | 87894093 | A | C | I | L | PTEN | ENST00000371953 | 0.67271 | 0.93125 | 0.92202 | 0.97518 | 0.93747 | 0.9326139767 | 0.93311 | 0.9125747681 | 0.330418 | 3.709204136 | 0 |
| 10 | 87894093 | A | G | I | V | PTEN | ENST00000371953 | 0.61499 | 0.925 | 0.85317 | 0.95993 | 0.90972 | 0.8391211293 | 0.88908 | 0.9219469428 | 0.232431 | 4.490133994 | 0 |
| 10 | 87894093 | A | T | I | F | PTEN | ENST00000371953 | 0.8336 | 0.98906 | 0.92642 | 0.98673 | 0.96773 | 0.9750428644 | 0.98284 | 0.9459557533 | 0.546485 | 4.483799726 | 0 |
| 10 | 87894094 | T | G | I | S | PTEN | ENST00000371953 | 0.9381 | 0.99448 | 0.93507 | 0.98797 | 0.99414 | 0.9930800001 | 0.99441 | 0.9944148064 | 0.565102 | 7.678924736 | 0 |
| 10 | 87894095 | T | G | I | M | PTEN | ENST00000371953 | 0.58211 | 0.94204 | 0.89634 | 0.85885 | 0.87491 | 0.9353626752 | 0.97322 | 0.6364311576 | 0.397777 | 2.824186815 | 0 |
| 10 | 87894096 | G | C | D | H | PTEN | ENST00000371953 | 0.92764 | 0.98254 | 0.97637 | 0.97709 | 0.95207 | 0.9164727544 | 0.97976 | 0.9959546328 | 0.534278 | 4.996965507 | 0 |
| 10 | 87894096 | G | T | D | Y | PTEN | ENST00000371953 | 0.93229 | 0.9975 | 0.96917 | 0.98264 | 0.96993 | 0.9506867518 | 0.99165 | 0.9991990924 | 0.564972 | 6.089433969 | 0 |
| 10 | 87894097 | A | C | D | A | PTEN | ENST00000371953 | 0.8681 | 0.96823 | 0.93777 | 0.9741 | 0.97141 | 0.9298217342 | 0.98231 | 0.9910633564 | 0.522886 | 4.664913768 | 0 |
| 10 | 87894097 | A | G | D | G | PTEN | ENST00000371953 | 0.87636 | 0.98069 | 0.94471 | 0.9808 | 0.967 | 0.9435288436 | 0.9866 | 0.9956733584 | 0.530308 | 5.249827767 | 0 |
| 10 | 87894097 | A | T | D | V | PTEN | ENST00000371953 | 0.86474 | 0.98018 | 0.9378 | 0.9861 | 0.97475 | 0.9549952954 | 0.99552 | 0.9981921315 | 0.546258 | 5.332817367 | 0 |
| 10 | 87894098 | T | A | D | E | PTEN | ENST00000371953 | 0.5429 | 0.92096 | 0.95178 | 0.93971 | 0.83615 | 0.8139633497 | 0.94609 | 0.8706271052 | 0.184968 | 2.632767239 | 0 |
| 10 | 87894099 | G | A | D | N | PTEN | ENST00000371953 | 0.94781 | 0.99419 | 0.95032 | 0.98889 | 0.96773 | 0.9512985714 | 0.98391 | 0.9975693822 | 0.197736 | 4.948710026 | 0 |
| 10 | 87894099 | G | C | D | H | PTEN | ENST00000371953 | 0.94748 | 0.9948 | 0.96245 | 0.98698 | 0.99296 | 0.966076778 | 0.99319 | 0.9980500937 | 0.565231 | 3.666392932 | 0 |
| 10 | 87894100 | A | C | D | A | PTEN | ENST00000371953 | 0.88461 | 0.9977 | 0.91533 | 0.98503 | 0.98938 | 0.970652218 | 0.98874 | 0.9976582527 | 0.533237 | 3.945764482 | 0 |
| 10 | 87894100 | A | G | D | G | PTEN | ENST00000371953 | 0.8897 | 0.99941 | 0.92253 | 0.98939 | 0.98728 | 0.9773503252 | 0.99603 | 0.9989118576 | 0.555607 | 5.802826379 | 0 |
| 10 | 87894100 | A | T | D | V | PTEN | ENST00000371953 | 0.87768 | 0.99842 | 0.91316 | 0.98852 | 0.99072 | 0.9804856317 | 0.99337 | 0.9993488193 | 0.537392 | 4.179088456 | 0 |
| 10 | 87894101 | T | A | D | E | PTEN | ENST00000371953 | 0.61544 | 0.96823 | 0.93528 | 0.90039 | 0.90274 | 0.9053277461 | 0.96952 | 0.9119766355 | 0.300885 | 2.064373328 | 0 |
| 10 | 87894103 | T | A | V | E | PTEN | ENST00000371953 | 0.91428 | 0.99994 | 0.98903 | 0.99597 | 0.99932 | 0.9987537352 | 0.99594 | 0.9997785687 | 0.586871 | 7.300172085 | 1 |
| 10 | 87894103 | T | C | V | A | PTEN | ENST00000371953 | 0.89018 | 0.99826 | 0.99011 | 0.99356 | 0.99039 | 0.9822946912 | 0.96107 | 0.9974704981 | 0.457058 | 5.092327106 | 0 |
| 10 | 87894103 | T | G | V | G | PTEN | ENST00000371953 | 0.91148 | 0.99941 | 0.95766 | 0.99628 | 0.99932 | 0.9954573351 | 0.99276 | 0.9997828603 | 0.587137 | 6.345097456 | 1 |
| 10 | 87894105 | G | A | V | I | PTEN | ENST00000371953 | 0.63076 | 0.92025 | 0.91354 | 0.97132 | 0.84641 | 0.8392892736 | 0.87496 | 0.655379355 | 0.167576 | 2.680196738 | 0 |
| 10 | 87894105 | G | C | V | L | PTEN | ENST00000371953 | 0.66763 | 0.94046 | 0.97977 | 0.97587 | 0.8859 | 0.9495843668 | 0.96434 | 0.9455212951 | 0.411406 | 3.39456753 | 0 |
| 10 | 87894105 | G | T | V | L | PTEN | ENST00000371953 | 0.66114 | 0.94046 | 0.97977 | 0.97582 | 0.8859 | 0.9495843668 | 0.96429 | 0.9455212951 | 0.411405 | 3.39456753 | 0 |
| 10 | 87894106 | T | A | V | E | PTEN | ENST00000371953 | 0.90111 | 0.99175 | 0.98039 | 0.98756 | 0.98621 | 0.9824169917 | 0.99034 | 0.9783026576 | 0.510772 | 4.509968092 | 0 |
| 10 | 87894106 | T | C | V | A | PTEN | ENST00000371953 | 0.75081 | 0.97027 | 0.98339 | 0.97836 | 0.93123 | 0.9168195091 | 0.94993 | 0.9580947161 | 0.368793 | 3.761588386 | 0 |
| 10 | 87894106 | T | G | V | G | PTEN | ENST00000371953 | 0.90004 | 0.99842 | 0.93812 | 0.99312 | 0.9796 | 0.9622294539 | 0.98316 | 0.9963376522 | 0.525277 | 5.272474718 | 0 |
| 10 | 87894108 | A | G | R | G | PTEN | ENST00000371953 | 0.8665 | 0.9977 | 0.90809 | 0.98199 | 0.95499 | 0.9468894855 | 0.99372 | 0.991471827 | 0.513243 | 5.345858991 | 0 |
| 10 | 87894108 | A | T | R | W | PTEN | ENST00000371953 | 0.91746 | 0.99735 | 0.96483 | 0.98103 | 0.96664 | 0.9198241836 | 0.96486 | 0.9974597096 | 0.542719 | 7.519406045 | 0 |
| 10 | 87894109 | G | A | R | K | PTEN | ENST00000371953 | 0.62309 | 0.95014 | 0.95167 | 0.91808 | 0.84052 | 0.8857777108 | 0.93044 | 0.9119116664 | 0.232488 | 2.033861395 | 0 |
| 10 | 87894109 | G | C | R | T | PTEN | ENST00000371953 | 0.82295 | 0.98685 | 0.93967 | 0.97271 | 0.98109 | 0.9396717007 | 0.98156 | 0.9823519588 | 0.495132 | 5.927633027 | 0 |
| 10 | 87894109 | G | T | R | M | PTEN | ENST00000371953 | 0.86347 | 0.99707 | 0.93612 | 0.96445 | 0.9781 | 0.9092409232 | 0.97638 | 0.9978739023 | 0.498126 | 6.196399457 | 0 |
| 10 | 87925513 | G | C | R | S | PTEN | ENST00000371953 | 0.87376 | 0.98311 | 0.917 | 0.96206 | 0.95899 | 0.9391222365 | 0.97634 | 0.9889150262 | 0.481477 | 4.760608039 | 0 |
| 10 | 87925513 | G | T | R | S | PTEN | ENST00000371953 | 0.86817 | 0.98311 | 0.917 | 0.96239 | 0.9608 | 0.9391222365 | 0.97638 | 0.9889150262 | 0.481477 | 4.760608039 | 0 |
| 10 | 87925514 | T | A | F | I | PTEN | ENST00000371953 | 0.82475 | 0.98348 | 0.98431 | 0.98583 | 0.99628 | 0.9799706776 | 0.96751 | 0.994117074 | 0.537114 | 6.687460232 | 1 |
| 10 | 87925515 | T | A | F | Y | PTEN | ENST00000371953 | 0.8044 | 0.96564 | 0.98726 | 0.98302 | 0.95316 | 0.9078881945 | 0.90846 | 0.9341565371 | 0.332073 | 5.27803422 | 0 |
| 10 | 87925515 | T | C | F | S | PTEN | ENST00000371953 | 0.94707 | 0.99294 | 0.98486 | 0.99486 | 0.99943 | 0.9875894611 | 0.98862 | 0.9979348183 | 0.570177 | 7.17704442 | 1 |
| 10 | 87925515 | T | G | F | C | PTEN | ENST00000371953 | 0.94695 | 0.98638 | 0.98971 | 0.99437 | 0.99892 | 0.9779236133 | 0.98819 | 0.9963931441 | 0.565393 | 7.305434542 | 0 |
| 10 | 87925516 | T | G | F | L | PTEN | ENST00000371953 | 0.77742 | 0.95345 | 0.99019 | 0.94266 | 0.95681 | 0.9516441968 | 0.93873 | 0.9850971699 | 0.350803 | 3.943512663 | 0 |
| 10 | 87925517 | T | A | L | M | PTEN | ENST00000371953 | 0.82397 | 0.96387 | 0.96453 | 0.9902 | 0.94129 | 0.9242951565 | 0.93715 | 0.9649174213 | 0.338482 | 3.35582917 | 0 |
| 10 | 87925517 | T | G | L | V | PTEN | ENST00000371953 | 0.80187 | 0.96697 | 0.94825 | 0.98828 | 0.9648 | 0.9637726773 | 0.98216 | 0.9639527798 | 0.515672 | 5.558817213 | 0 |
| 10 | 87925518 | T | C | L | S | PTEN | ENST00000371953 | 0.93641 | 0.9862 | 0.97387 | 0.99315 | 0.99786 | 0.994278149 | 0.99297 | 0.9973356128 | 0.569269 | 6.746634299 | 1 |
| 10 | 87925518 | T | G | L | W | PTEN | ENST00000371953 | 0.93683 | 0.99204 | 0.98936 | 0.99384 | 0.99525 | 0.9932934924 | 0.98918 | 0.9992513061 | 0.569433 | 6.746634299 | 1 |
| 10 | 87925519 | G | T | L | F | PTEN | ENST00000371953 | 0.75726 | 0.95036 | 0.9648 | 0.97134 | 0.9367 | 0.9689920233 | 0.95771 | 0.945528388 | 0.455467 | 2.086623007 | 0 |
| 10 | 87925520 | G | A | D | N | PTEN | ENST00000371953 | 0.75495 | 0.82937 | 0.96905 | 0.94461 | 0.79151 | 0.8959999634 | 0.93656 | 0.8073309373 | 0.0762323 | 2.728956367 | 0 |
| 10 | 87925520 | G | C | D | H | PTEN | ENST00000371953 | 0.88253 | 0.95345 | 0.97801 | 0.9822 | 0.96517 | 0.9341804181 | 0.9921 | 0.9943929911 | 0.552038 | 5.252341725 | 0 |
| 10 | 87925520 | G | T | D | Y | PTEN | ENST00000371953 | 0.88892 | 0.97924 | 0.9699 | 0.97893 | 0.9781 | 0.9614026548 | 0.9946 | 0.9966735003 | 0.560591 | 5.896898372 | 0 |
| 10 | 87925521 | A | C | D | A | PTEN | ENST00000371953 | 0.93201 | 0.97099 | 0.94212 | 0.98878 | 0.9822 | 0.9443316536 | 0.98525 | 0.9914329891 | 0.530326 | 4.832507347 | 0 |
| 10 | 87925521 | A | G | D | G | PTEN | ENST00000371953 | 0.93633 | 0.97893 | 0.94835 | 0.9838 | 0.98146 | 0.9558128404 | 0.9935 | 0.9926244617 | 0.543798 | 5.677664453 | 0 |
| 10 | 87925521 | A | T | D | V | PTEN | ENST00000371953 | 0.93244 | 0.97143 | 0.93771 | 0.98724 | 0.98728 | 0.9618039703 | 0.99528 | 0.9962924584 | 0.558473 | 6.779145505 | 0 |
| 10 | 87925523 | T | A | S | T | PTEN | ENST00000371953 | 0.57533 | 0.78232 | 0.91549 | 0.94163 | 0.84159 | 0.7401405246 | 0.90373 | 0.6555985808 | 0.205361 | 2.642241717 | 0 |
| 10 | 87925523 | T | C | S | P | PTEN | ENST00000371953 | 0.85412 | 0.97822 | 0.93317 | 0.98287 | 0.97178 | 0.9648839861 | 0.97037 | 0.9615248442 | 0.509737 | 7.194467085 | 1 |
| 10 | 87925523 | T | G | S | A | PTEN | ENST00000371953 | 0.62741 | 0.77624 | 0.90675 | 0.96129 | 0.90798 | 0.6872231902 | 0.91375 | 0.7181429863 | 0.224639 | 3.160547136 | 0 |
| 10 | 87925524 | C | T | S | L | PTEN | ENST00000371953 | 0.5558 | 0.79191 | 0.93205 | 0.93643 | 0.85996 | 0.8365569902 | 0.98366 | 0.9577630758 | 0.361407 | 2.691962636 | 0 |
| 10 | 87925526 | A | C | K | Q | PTEN | ENST00000371953 | 0.8349 | 0.93164 | 0.96551 | 0.97004 | 0.93786 | 0.8712939151 | 0.96663 | 0.956657052 | 0.268872 | 5.393572255 | 0 |
| 10 | 87925526 | A | G | K | E | PTEN | ENST00000371953 | 0.85894 | 0.96965 | 0.97699 | 0.98674 | 0.96334 | 0.901561258 | 0.96933 | 0.975482285 | 0.472161 | 6.052977836 | 0 |
| 10 | 87925527 | A | C | K | T | PTEN | ENST00000371953 | 0.9217 | 0.97662 | 0.98102 | 0.99084 | 0.9729 | 0.9080815812 | 0.94921 | 0.9932383409 | 0.403717 | 5.709288828 | 0 |
| 10 | 87925527 | A | G | K | R | PTEN | ENST00000371953 | 0.79705 | 0.75521 | 0.95394 | 0.94555 | 0.87442 | 0.746525451 | 0.93216 | 0.7874617577 | 0.21604 | 2.926732058 | 0 |
| 10 | 87925527 | A | T | K | M | PTEN | ENST00000371953 | 0.93527 | 0.9663 | 0.95316 | 0.99341 | 0.96846 | 0.9018439085 | 0.95482 | 0.998072667 | 0.401624 | 4.670932588 | 0 |
| 10 | 87925528 | G | T | K | N | PTEN | ENST00000371953 | 0.73981 | 0.91035 | 0.98406 | 0.9599 | 0.8543 | 0.9062749454 | 0.93566 | 0.8110949402 | 0.15286 | 4.093258595 | 0 |
| 10 | 87925529 | C | G | H | D | PTEN | ENST00000371953 | 0.7606 | 0.9858 | 0.98532 | 0.98316 | 0.99326 | 0.9977835933 | 0.97579 | 0.9972762465 | 0.561331 | 7.057672525 | 1 |
| 10 | 87925529 | C | T | H | Y | PTEN | ENST00000371953 | 0.71137 | 0.97475 | 0.98707 | 0.97932 | 0.99443 | 0.9896671246 | 0.98602 | 0.9934393764 | 0.526314 | 3.969125092 | 0 |
| 10 | 87925530 | A | C | H | P | PTEN | ENST00000371953 | 0.82568 | 0.99056 | 0.97402 | 0.99005 | 0.99919 | 0.9963262736 | 0.98006 | 0.9998933077 | 0.574611 | 7.076355484 | 1 |
| 10 | 87925530 | A | G | H | R | PTEN | ENST00000371953 | 0.77972 | 0.97972 | 0.9873 | 0.98592 | 0.99876 | 0.9945981052 | 0.99517 | 0.9990193844 | 0.574329 | 6.259282121 | 1 |
| 10 | 87925530 | A | T | H | L | PTEN | ENST00000371953 | 0.82864 | 0.98043 | 0.9795 | 0.98836 | 0.99265 | 0.9919373623 | 0.97505 | 0.999024272 | 0.549933 | 5.390450414 | 1 |
| 10 | 87925531 | T | A | H | Q | PTEN | ENST00000371953 | 0.807 | 0.96909 | 0.98259 | 0.98985 | 0.99552 | 0.9888634057 | 0.98813 | 0.9968237877 | 0.568544 | 7.01244777 | 1 |
| 10 | 87925532 | A | C | K | Q | PTEN | ENST00000371953 | 0.70752 | 0.89472 | 0.89371 | 0.94354 | 0.87636 | 0.872821536 | 0.95584 | 0.8184551597 | 0.246124 | 3.796574505 | 0 |
| 10 | 87925532 | A | G | K | E | PTEN | ENST00000371953 | 0.86803 | 0.96209 | 0.92016 | 0.96369 | 0.93786 | 0.9219057856 | 0.98798 | 0.8742137556 | 0.481911 | 4.43114681 | 0 |
| 10 | 87925533 | A | C | K | T | PTEN | ENST00000371953 | 0.78669 | 0.91266 | 0.93091 | 0.97842 | 0.95608 | 0.9108332542 | 0.95502 | 0.8985706568 | 0.381785 | 5.152888097 | 0 |
| 10 | 87925533 | A | G | K | R | PTEN | ENST00000371953 | 0.63061 | 0.78767 | 0.86758 | 0.96184 | 0.89009 | 0.7688360141 | 0.95146 | 0.7877039909 | 0.238795 | 3.174382726 | 0 |
| 10 | 87925533 | A | T | K | I | PTEN | ENST00000371953 | 0.83936 | 0.9927 | 0.89759 | 0.98823 | 0.97215 | 0.9304532876 | 0.93286 | 0.9891249278 | 0.380633 | 7.616530039 | 0 |
| 10 | 87925534 | A | T | K | N | PTEN | ENST00000371953 | 0.91414 | 0.92813 | 0.93937 | 0.95976 | 0.92151 | 0.9175588897 | 0.93409 | 0.9459942579 | 0.19014 | 4.452220709 | 0 |
| 10 | 87925535 | A | C | N | H | PTEN | ENST00000371953 | 0.80724 | 0.95776 | 0.937 | 0.94028 | 0.92561 | 0.8279967086 | 0.94772 | 0.932471931 | 0.17678 | 4.338200888 | 0 |
| 10 | 87925535 | A | G | N | D | PTEN | ENST00000371953 | 0.40582 | 0.82827 | 0.95283 | 0.82745 | 0.70179 | 0.8330835635 | 0.79288 | 0.9624093175 | 0.0864284 | 0.9037429305 | 0 |
| 10 | 87925535 | A | T | N | Y | PTEN | ENST00000371953 | 0.8536 | 0.97054 | 0.94876 | 0.9785 | 0.94091 | 0.9051724914 | 0.9431 | 0.9709373116 | 0.337198 | 5.955360605 | 0 |
| 10 | 87925536 | A | C | N | T | PTEN | ENST00000371953 | 0.65798 | 0.84214 | 0.92527 | 0.93385 | 0.83615 | 0.8503081806 | 0.91953 | 0.8541115522 | 0.197014 | 4.22137059 | 0 |
| 10 | 87925536 | A | G | N | S | PTEN | ENST00000371953 | 0.48417 | 0.72775 | 0.89459 | 0.88717 | 0.71548 | 0.7445175653 | 0.91386 | 0.6741245985 | 0.0626897 | 2.799843304 | 0 |
| 10 | 87925536 | A | T | N | I | PTEN | ENST00000371953 | 0.87221 | 0.9632 | 0.92916 | 0.97964 | 0.96444 | 0.9238379306 | 0.94785 | 0.9863218069 | 0.354596 | 6.249528879 | 0 |
| 10 | 87925537 | C | A | N | K | PTEN | ENST00000371953 | 0.54637 | 0.84731 | 0.96513 | 0.85431 | 0.76223 | 0.8747884253 | 0.91074 | 0.9715967774 | 0.125764 | 3.638083207 | 0 |
| 10 | 87925538 | C | A | H | N | PTEN | ENST00000371953 | 0.42717 | 0.89106 | 0.98312 | 0.93644 | 0.83004 | 0.9254636425 | 0.91535 | 0.977416873 | 0.176718 | 2.890617827 | 0 |
| 10 | 87925538 | C | G | H | D | PTEN | ENST00000371953 | 0.74372 | 0.96836 | 0.98191 | 0.95671 | 0.91271 | 0.9806114163 | 0.89689 | 0.9972633123 | 0.300676 | 5.046581964 | 0 |
| 10 | 87925538 | C | T | H | Y | PTEN | ENST00000371953 | 0.72051 | 0.95362 | 0.98359 | 0.98383 | 0.95353 | 0.9456915707 | 0.94821 | 0.9974347949 | 0.280316 | 5.695251572 | 0 |
| 10 | 87925539 | A | C | H | P | PTEN | ENST00000371953 | 0.92401 | 0.98981 | 0.96319 | 0.9881 | 0.9608 | 0.9868132292 | 0.93065 | 0.9965157509 | 0.381301 | 7.67387357 | 1 |
| 10 | 87925539 | A | G | H | R | PTEN | ENST00000371953 | 0.77227 | 0.92663 | 0.98284 | 0.96903 | 0.95535 | 0.9487004867 | 0.92174 | 0.9391655326 | 0.224944 | 4.146239721 | 0 |
| 10 | 87925539 | A | T | H | L | PTEN | ENST00000371953 | 0.92493 | 0.97475 | 0.97359 | 0.98817 | 0.93747 | 0.9641321013 | 0.94462 | 0.9996685982 | 0.310533 | 6.748582614 | 0 |
| 10 | 87925540 | T | A | H | Q | PTEN | ENST00000371953 | 0.8877 | 0.92245 | 0.97563 | 0.98227 | 0.94543 | 0.9266531707 | 0.94151 | 0.9872841835 | 0.227818 | 4.347896376 | 0 |
| 10 | 87925541 | T | C | Y | H | PTEN | ENST00000371953 | 0.87918 | 0.98258 | 0.99159 | 0.99233 | 0.99765 | 0.9878592081 | 0.99271 | 0.9991000891 | 0.574613 | 5.159625049 | 0 |
| 10 | 87925541 | T | G | Y | D | PTEN | ENST00000371953 | 0.8894 | 0.99403 | 0.9893 | 0.99306 | 0.99765 | 0.9988319684 | 0.99669 | 0.9999461174 | 0.580582 | 6.858089654 | 1 |
| 10 | 87925542 | A | C | Y | S | PTEN | ENST00000371953 | 0.93818 | 0.98719 | 0.97324 | 0.99172 | 0.99962 | 0.9955619261 | 0.9963 | 0.9996811152 | 0.587757 | 5.888617826 | 0 |
| 10 | 87925542 | A | G | Y | C | PTEN | ENST00000371953 | 0.9463 | 0.98086 | 0.99014 | 0.99029 | 0.9999 | 0.9963288625 | 0.99421 | 0.9997105002 | 0.587757 | 4.558801189 | 0 |

| | | | | | | | | | | | | | | | | | |
|---|---|---|---|---|---|---|---|---|---|---|---|---|---|---|---|---|---|
| 10 | 87925542 | A | T | Y | F | PTEN | ENST00000371953 | 0.90323 | 0.95541 | 0.96835 | 0.97982 | 0.97438 | 0.9469874251 | 0.92275 | 0.9699678421 | 0.26714 | 2.718883609 | 0 |
| 10 | 87925544 | A | C | K | Q | PTEN | ENST00000371953 | 0.8366 | 0.97175 | 0.9057 | 0.98482 | 0.95645 | 0.9388169877 | 0.98138 | 0.9729180932 | 0.406834 | 4.841772061 | 0 |
| 10 | 87925545 | A | C | K | T | PTEN | ENST00000371953 | 0.92907 | 0.96043 | 0.93692 | 0.98316 | 0.96664 | 0.9378087277 | 0.97536 | 0.9942313433 | 0.530073 | 5.527430893 | 0 |
| 10 | 87925545 | A | G | K | R | PTEN | ENST00000371953 | 0.83035 | 0.85203 | 0.88566 | 0.97569 | 0.91399 | 0.8845689995 | 0.96021 | 0.846894443 | 0.332272 | 3.534162396 | 0 |
| 10 | 87925545 | A | T | K | M | PTEN | ENST00000371953 | 0.94035 | 0.9756 | 0.88554 | 0.98654 | 0.95972 | 0.9358001397 | 0.98008 | 0.9969291091 | 0.489784 | 4.188452238 | 0 |
| 10 | 87925546 | G | T | K | N | PTEN | ENST00000371953 | 0.9294 | 0.97742 | 0.9468 | 0.97882 | 0.93201 | 0.9593742365 | 0.97109 | 0.9962162375 | 0.285981 | 7.256230369 | 0 |
| 10 | 87925547 | A | C | I | L | PTEN | ENST00000371953 | 0.74698 | 0.93681 | 0.96819 | 0.96723 | 0.93863 | 0.9497967023 | 0.98741 | 0.9120895267 | 0.42332 | 3.474917284 | 0 |
| 10 | 87925547 | A | G | I | V | PTEN | ENST00000371953 | 0.39585 | 0.85117 | 0.92403 | 0.85569 | 0.84482 | 0.8264865907 | 0.93836 | 0.7737334967 | 0.258905 | 1.911827459 | 0 |
| 10 | 87925547 | A | T | I | L | PTEN | ENST00000371953 | 0.74029 | 0.93681 | 0.96819 | 0.96709 | 0.93747 | 0.9497967023 | 0.98747 | 0.9120895267 | 0.42332 | 3.474917284 | 0 |
| 10 | 87925548 | T | A | I | K | PTEN | ENST00000371953 | 0.92264 | 0.99399 | 0.9862 | 0.98839 | 0.99824 | 0.997707288 | 0.9963 | 0.9977884293 | 0.576604 | 6.860583691 | 1 |
| 10 | 87925548 | T | C | I | T | PTEN | ENST00000371953 | 0.9076 | 0.98161 | 0.99239 | 0.98606 | 0.99552 | 0.9793729183 | 0.99554 | 0.9982201457 | 0.569301 | 5.848953351 | 0 |
| 10 | 87925549 | A | G | I | M | PTEN | ENST00000371953 | 0.84423 | 0.9695 | 0.95141 | 0.98144 | 0.9755 | 0.9546719653 | 0.98046 | 0.9893831611 | 0.491047 | 5.105611035 | 0 |
| 10 | 87925550 | T | A | Y | N | PTEN | ENST00000371953 | 0.8619 | 0.99192 | 0.98398 | 0.99133 | 0.98764 | 0.9957543488 | 0.95662 | 0.9994612336 | 0.497822 | 8.00185113 | 1 |
| 10 | 87925550 | T | C | Y | H | PTEN | ENST00000371953 | 0.85778 | 0.98311 | 0.98637 | 0.98996 | 0.99962 | 0.9812184435 | 0.98563 | 0.9986030459 | 0.553706 | 6.461190131 | 1 |
| 10 | 87925550 | T | G | Y | D | PTEN | ENST00000371953 | 0.87072 | 0.99403 | 0.98229 | 0.99154 | 0.99919 | 0.998321545 | 0.99347 | 0.999835372 | 0.561544 | 7.989007724 | 1 |
| 10 | 87925551 | A | C | Y | S | PTEN | ENST00000371953 | 0.93781 | 0.98685 | 0.9616 | 0.99315 | 0.99943 | 0.9945439161 | 0.99036 | 0.9990904331 | 0.566879 | 7.592793264 | 1 |
| 10 | 87925551 | A | G | Y | C | PTEN | ENST00000371953 | 0.94605 | 0.98161 | 0.98482 | 0.99181 | 0.99932 | 0.9935849702 | 0.99124 | 0.999168992 | 0.579343 | 6.471435269 | 1 |
| 10 | 87925553 | A | C | N | H | PTEN | ENST00000371953 | 0.77322 | 0.97027 | 0.818 | 0.99084 | 0.9822 | 0.9874564467 | 0.98864 | 0.9969288707 | 0.364443 | 4.289500776 | 0 |
| 10 | 87925553 | A | G | N | D | PTEN | ENST00000371953 | 0.7999 | 0.97662 | 0.85601 | 0.98954 | 0.98513 | 0.9875915819 | 0.98317 | 0.9982017279 | 0.307928 | 4.529372212 | 0 |
| 10 | 87925553 | A | T | N | Y | PTEN | ENST00000371953 | 0.82069 | 0.98028 | 0.84678 | 0.99269 | 0.99892 | 0.9938280565 | 0.98889 | 0.9998419285 | 0.584278 | 6.499729608 | 1 |
| 10 | 87925554 | A | C | N | T | PTEN | ENST00000371953 | 0.89497 | 0.96909 | 0.78537 | 0.98647 | 0.97661 | 0.9764095019 | 0.97235 | 0.9959794283 | 0.343194 | 5.754078079 | 0 |
| 10 | 87925554 | A | G | N | S | PTEN | ENST00000371953 | 0.8536 | 0.95057 | 0.74106 | 0.98742 | 0.98585 | 0.9699011733 | 0.98074 | 0.9952093959 | 0.280048 | 5.547692369 | 0 |
| 10 | 87925554 | A | T | N | I | PTEN | ENST00000371953 | 0.92108 | 0.9813 | 0.81044 | 0.99172 | 0.9781 | 0.9942136179 | 0.96331 | 0.999879241 | 0.403214 | 6.540258149 | 1 |
| 10 | 87925555 | T | A | N | K | PTEN | ENST00000371953 | 0.84681 | 0.97717 | 0.88457 | 0.98653 | 0.96737 | 0.9947092236 | 0.97322 | 0.9974877834 | 0.260481 | 5.850175938 | 0 |
| 10 | 87925556 | C | A | L | I | PTEN | ENST00000371953 | 0.7836 | 0.97295 | 0.87704 | 0.9765 | 0.95645 | 0.9349543089 | 0.91908 | 0.9879410863 | 0.337788 | 3.762727003 | 0 |
| 10 | 87925556 | C | G | L | V | PTEN | ENST00000371953 | 0.78059 | 0.96909 | 0.8211 | 0.97539 | 0.98513 | 0.9682334046 | 0.96621 | 0.9886926413 | 0.426167 | 3.849459324 | 0 |
| 10 | 87925556 | C | T | L | F | PTEN | ENST00000371953 | 0.82201 | 0.97175 | 0.86398 | 0.9713 | 0.97698 | 0.978468423 | 0.98129 | 0.9970228076 | 0.500334 | 3.747546391 | 0 |
| 10 | 87925557 | T | C | L | P | PTEN | ENST00000371953 | 0.86126 | 0.99155 | 0.95833 | 0.9948 | 0.99962 | 0.9983904553 | 0.99624 | 0.9997569919 | 0.580591 | 6.509236185 | 1 |
| 10 | 87925557 | T | G | L | R | PTEN | ENST00000371953 | 0.85713 | 0.9882 | 0.93954 | 0.99527 | 0.99932 | 0.9985908514 | 0.99538 | 0.9997617602 | 0.580328 | 4.428035105 | 1 |
| 10 | 87931047 | T | C | C | R | PTEN | ENST00000371953 | 0.86061 | 0.99294 | 0.99641 | 0.98552 | 0.99525 | 0.9934121299 | 0.99611 | 0.9998315573 | 0.534591 | 4.629299858 | 0 |
| 10 | 87931048 | G | C | C | S | PTEN | ENST00000371953 | 0.93259 | 0.98069 | 0.98367 | 0.96906 | 0.9648 | 0.9825762675 | 0.95933 | 0.9998848438 | 0.50857 | 5.872796636 | 0 |
| 10 | 87931049 | T | G | C | W | PTEN | ENST00000371953 | 0.94222 | 0.9851 | 0.99358 | 0.99345 | 0.98938 | 0.9906889078 | 0.99414 | 0.9999142885 | 0.559398 | 8.156412798 | 1 |
| 10 | 87931050 | G | A | A | T | PTEN | ENST00000371953 | 0.69981 | 0.93519 | 0.94555 | 0.95865 | 0.89467 | 0.9069163038 | 0.92089 | 0.8532361388 | 0.330064 | 3.191661143 | 0 |
| 10 | 87931050 | G | C | A | P | PTEN | ENST00000371953 | 0.85566 | 0.9551 | 0.9762 | 0.94952 | 0.94394 | 0.9826434977 | 0.98321 | 0.9754194021 | 0.518254 | 4.610372912 | 0 |
| 10 | 87931050 | G | T | A | S | PTEN | ENST00000371953 | 0.38708 | 0.79294 | 0.9223 | 0.94839 | 0.83725 | 0.850029689 | 0.95055 | 0.8945817947 | 0.307439 | 1.576680569 | 0 |
| 10 | 87931051 | C | A | A | D | PTEN | ENST00000371953 | 0.84279 | 0.93493 | 0.98479 | 0.96563 | 0.93397 | 0.9797900016 | 0.97834 | 0.8768858314 | 0.492353 | 4.019686707 | 0 |
| 10 | 87931051 | C | G | A | G | PTEN | ENST00000371953 | 0.81755 | 0.84381 | 0.9395 | 0.94831 | 0.90007 | 0.8764353308 | 0.93646 | 0.6598733394 | 0.329257 | 4.323658296 | 0 |
| 10 | 87931051 | C | T | A | V | PTEN | ENST00000371953 | 0.83498 | 0.86371 | 0.95845 | 0.9659 | 0.95353 | 0.9448449275 | 0.9694 | 0.9639178514 | 0.43457 | 4.793773116 | 0 |
| 10 | 87931053 | G | A | E | K | PTEN | ENST00000371953 | 0.90447 | 0.97582 | 0.97962 | 0.9792 | 0.99105 | 0.989000882 | 0.99625 | 0.9986398816 | 0.581011 | 5.85930429 | 0 |
| 10 | 87931053 | G | C | E | Q | PTEN | ENST00000371953 | 0.85234 | 0.96812 | 0.92364 | 0.97376 | 0.97661 | 0.9616884726 | 0.98722 | 0.996781826 | 0.511559 | 5.058250818 | 0 |
| 10 | 87931054 | A | C | E | A | PTEN | ENST00000371953 | 0.92857 | 0.9727 | 0.93591 | 0.98492 | 0.99786 | 0.972894404 | 0.98795 | 0.9995544553 | 0.567913 | 7.0771641 | 0 |
| 10 | 87931054 | A | G | E | G | PTEN | ENST00000371953 | 0.9439 | 0.9771 | 0.9024 | 0.98459 | 0.99386 | 0.9835017361 | 0.98886 | 0.9990098476 | 0.559837 | 4.5517615 | 0 |
| 10 | 87931054 | A | T | E | V | PTEN | ENST00000371953 | 0.93912 | 0.97816 | 0.92626 | 0.99008 | 0.99765 | 0.9847772782 | 0.99365 | 0.9998602867 | 0.583105 | 7.720404793 | 0 |
| 10 | 87931055 | A | T | E | D | PTEN | ENST00000371953 | 0.77505 | 0.96093 | 0.93922 | 0.98832 | 0.98183 | 0.9503638899 | 0.98091 | 0.9962562323 | 0.392109 | 4.845250592 | 0 |
| 10 | 87931056 | A | G | R | G | PTEN | ENST00000371953 | 0.92314 | 0.97295 | 0.96244 | 0.98564 | 0.97997 | 0.9842090991 | 0.99479 | 0.9955788255 | 0.510482 | 6.496011358 | 0 |
| 10 | 87931057 | G | A | R | K | PTEN | ENST00000371953 | 0.86559 | 0.93589 | 0.98331 | 0.96632 | 0.94318 | 0.9455569038 | 0.95737 | 0.981836319 | 0.373871 | 4.212222312 | 0 |
| 10 | 87931057 | G | C | R | T | PTEN | ENST00000371953 | 0.87335 | 0.98834 | 0.97682 | 0.98554 | 0.98404 | 0.9734290782 | 0.9772 | 0.9989494681 | 0.547175 | 5.779485951 | 0 |
| 10 | 87931058 | A | C | R | S | PTEN | ENST00000371953 | 0.80558 | 0.97491 | 0.96635 | 0.98982 | 0.98972 | 0.9774691279 | 0.99152 | 0.9985338449 | 0.544087 | 6.440963448 | 0 |
| 10 | 87931058 | A | T | R | S | PTEN | ENST00000371953 | 0.79895 | 0.97491 | 0.96635 | 0.98983 | 0.98972 | 0.9774691279 | 0.9915 | 0.9985338449 | 0.544084 | 6.440963448 | 0 |
| 10 | 87931059 | C | A | H | N | PTEN | ENST00000371953 | 0.3756 | 0.81811 | 0.98303 | 0.94083 | 0.72626 | 0.8240901691 | 0.89322 | 0.9454477429 | 0.191592 | 1.370793961 | 0 |
| 10 | 87931059 | C | G | H | D | PTEN | ENST00000371953 | 0.41943 | 0.87447 | 0.98049 | 0.92395 | 0.82724 | 0.9358578954 | 0.97782 | 0.8044421673 | 0.469167 | 2.947134331 | 0 |
| 10 | 87931059 | C | T | H | Y | PTEN | ENST00000371953 | 0.44035 | 0.83418 | 0.98409 | 0.94537 | 0.86803 | 0.8743114379 | 0.89608 | 0.8922733665 | 0.311591 | 3.763386107 | 0 |
| 10 | 87931060 | A | C | H | P | PTEN | ENST00000371953 | 0.59995 | 0.9015 | 0.96531 | 0.94774 | 0.90885 | 0.961315429 | 0.98023 | 0.944246769 | 0.483984 | 4.187862243 | 0 |
| 10 | 87931060 | A | G | H | R | PTEN | ENST00000371953 | 0.52546 | 0.84646 | 0.98312 | 0.90614 | 0.83779 | 0.8449711887 | 0.96191 | 0.8973745704 | 0.336012 | 2.982672203 | 0 |
| 10 | 87931060 | A | T | H | L | PTEN | ENST00000371953 | 0.53374 | 0.85023 | 0.97483 | 0.94773 | 0.86503 | 0.9248543844 | 0.987 | 0.8782185316 | 0.464811 | 3.969236001 | 0 |
| 10 | 87931061 | T | A | H | Q | PTEN | ENST00000371953 | 0.49978 | 0.79288 | 0.97749 | 0.94665 | 0.83615 | 0.840361087 | 0.96148 | 0.8191937804 | 0.329423 | 2.678968831 | 0 |
| 10 | 87931062 | T | A | Y | N | PTEN | ENST00000371953 | 0.88616 | 0.99204 | 0.95924 | 0.99139 | 0.98869 | 0.9936458794 | 0.9756 | 0.999891758 | 0.533646 | 5.849263549 | 0 |
| 10 | 87931062 | T | C | Y | H | PTEN | ENST00000371953 | 0.88132 | 0.98329 | 0.96362 | 0.98597 | 0.99105 | 0.9866975663 | 0.96809 | 0.9995607734 | 0.502984 | 5.494388375 | 0 |
| 10 | 87931063 | A | C | Y | S | PTEN | ENST00000371953 | 0.94284 | 0.98736 | 0.91484 | 0.9927 | 0.99356 | 0.9941765876 | 0.98967 | 0.9998051524 | 0.577017 | 5.196154283 | 0 |
| 10 | 87931063 | A | G | Y | C | PTEN | ENST00000371953 | 0.94914 | 0.98161 | 0.95888 | 0.99234 | 0.99552 | 0.9950545323 | 0.97675 | 0.9999405146 | 0.583128 | 6.717981959 | 0 |
| 10 | 87931063 | A | T | Y | F | PTEN | ENST00000371953 | 0.91525 | 0.96192 | 0.90417 | 0.99134 | 0.98904 | 0.9571932351 | 0.97088 | 0.9968764782 | 0.374623 | 4.710943389 | 0 |
| 10 | 87931065 | G | A | D | N | PTEN | ENST00000371953 | 0.91002 | 0.89862 | 0.96631 | 0.9665 | 0.90972 | 0.9464244514 | 0.9783 | 0.9968906045 | 0.185392 | 3.066188697 | 0 |
| 10 | 87931065 | G | C | D | H | PTEN | ENST00000371953 | 0.91073 | 0.96618 | 0.97369 | 0.983 | 0.98764 | 0.9677939847 | 0.98692 | 0.9976670742 | 0.578117 | 5.48520694 | 0 |
| 10 | 87931065 | G | T | D | Y | PTEN | ENST00000371953 | 0.91547 | 0.97662 | 0.96822 | 0.99066 | 0.98621 | 0.9899396534 | 0.99141 | 0.9997078776 | 0.583849 | 7.710993093 | 0 |
| 10 | 87931066 | A | C | D | A | PTEN | ENST00000371953 | 0.8974 | 0.96697 | 0.93499 | 0.9872 | 0.98183 | 0.9800622688 | 0.96997 | 0.9997122884 | 0.553006 | 4.932988746 | 0 |
| 10 | 87931066 | A | G | D | G | PTEN | ENST00000371953 | 0.90186 | 0.96273 | 0.94045 | 0.98606 | 0.98972 | 0.9819026711 | 0.99149 | 0.9981102943 | 0.571408 | 5.975813074 | 0 |
| 10 | 87931066 | A | T | D | V | PTEN | ENST00000371953 | 0.89119 | 0.98043 | 0.93805 | 0.98997 | 0.99386 | 0.9864048492 | 0.99138 | 0.9999155998 | 0.583128 | 6.010468808 | 0 |
| 10 | 87931067 | C | A | D | E | PTEN | ENST00000371953 | 0.58631 | 0.9415 | 0.95408 | 0.97729 | 0.94431 | 0.9383115219 | 0.9671 | 0.9965091348 | 0.327141 | 4.127867197 | 0 |
| 10 | 87931068 | A | C | T | P | PTEN | ENST00000371953 | 0.42717 | 0.86159 | 0.68987 | 0.83543 | 0.75822 | 0.9655428887 | 0.97033 | 0.8726570606 | 0.240637 | 1.184205938 | 0 |
| 10 | 87931068 | A | G | T | A | PTEN | ENST00000371953 | 0.36175 | 0.78576 | 0.69459 | 0.83148 | 0.73877 | 0.8654725683 | 0.80807 | 0.6116379499 | 0.0763583 | 0.6802535122 | 0 |
| 10 | 87931068 | A | T | T | S | PTEN | ENST00000371953 | 0.37103 | 0.76057 | 0.70517 | 0.85144 | 0.75005 | 0.8114903473 | 0.84781 | 0.6116379499 | 0.12246 | 2.711085454 | 0 |
| 10 | 87931069 | C | A | T | N | PTEN | ENST00000371953 | 0.40893 | 0.86648 | 0.80001 | 0.94132 | 0.82103 | 0.9423737843 | 0.81771 | 0.4410320384 | 0.160698 | 3.875583279 | 0 |
| 10 | 87931069 | C | G | T | S | PTEN | ENST00000371953 | 0.36626 | 0.76057 | 0.70517 | 0.85477 | 0.73949 | 0.8114903473 | 0.78078 | 0.9433323145 | 0.14456 | 2.711085454 | 0 |
| 10 | 87931069 | C | T | T | I | PTEN | ENST00000371953 | 0.41069 | 0.81726 | 0.86059 | 0.88897 | 0.73516 | 0.9107169716 | 0.82554 | 0.8516389728 | 0.193839 | 2.518419949 | 0 |
| 10 | 87931071 | G | A | A | T | PTEN | ENST00000371953 | 0.49235 | 0.78278 | 0.84408 | 0.76695 | 0.77654 | 0.7524006012 | 0.98081 | 0.1317448169 | 0.0865203 | 1.911498021 | 0 |
| 10 | 87931071 | G | C | A | P | PTEN | ENST00000371953 | 0.73926 | 0.94115 | 0.91377 | 0.911 | 0.91229 | 0.9628175459 | 0.93603 | 0.9316857457 | 0.3029 | 5.052362688 | 0 |
| 10 | 87931071 | G | T | A | S | PTEN | ENST00000371953 | 0.41233 | 0.76532 | 0.80402 | 0.79987 | 0.77845 | 0.7116713208 | 0.86572 | 0.9231678247 | 0.152346 | 0.5467338627 | 0 |
| 10 | 87931072 | C | A | A | D | PTEN | ENST00000371953 | 0.51507 | 0.89404 | 0.94208 | 0.88655 | 0.80476 | 0.9280774812 | 0.93931 | 0.7893813252 | 0.289211 | 2.371019842 | 0 |
| 10 | 87931072 | C | G | A | G | PTEN | ENST00000371953 | 0.52141 | 0.7976 | 0.83646 | 0.89118 | 0.80595 | 0.7770684762 | 0.84844 | 0.6961876154 | 0.176719 | 2.295501557 | 0 |
| 10 | 87931072 | C | T | A | V | PTEN | ENST00000371953 | 0.63643 | 0.81056 | 0.87554 | 0.90201 | 0.81818 | 0.8452523932 | 0.85696 | 0.8700892329 | 0.189893 | 3.087540079 | 0 |
| 10 | 87931074 | A | C | K | Q | PTEN | ENST00000371953 | 0.83806 | 0.94569 | 0.82533 | 0.9416 | 0.93201 | 0.9278086431 | 0.93459 | 0.9820865393 | 0.14318 | 5.826520256 | 0 |
| 10 | 87931075 | A | C | K | T | PTEN | ENST00000371953 | 0.84325 | 0.9756 | 0.87616 | 0.94155 | 0.93515 | 0.9521021663 | 0.91174 | 0.9969310164 | 0.288472 | 5.57510427 | 0 |
| 10 | 87931075 | A | G | K | R | PTEN | ENST00000371953 | 0.69676 | 0.7829 | 0.79531 | 0.76754 | 0.80358 | 0.8345156547 | 0.89357 | 0.8651938438 | 0.100755 | 3.105697465 | 0 |
| 10 | 87931075 | A | T | K | I | PTEN | ENST00000371953 | 0.88417 | 0.98449 | 0.83652 | 0.92443 | 0.9248 | 0.9614846751 | 0.89307 | 0.9991387129 | 0.265593 | 4.478779187 | 0 |
| 10 | 87931076 | A | T | K | N | PTEN | ENST00000371953 | 0.86119 | 0.92538 | 0.8948 | 0.92056 | 0.89147 | 0.9563501035 | 0.93154 | 0.9890817404 | 0.102724 | 4.743152262 | 0 |
| 10 | 87931077 | T | A | F | I | PTEN | ENST00000371953 | 0.8536 | 0.98591 | 0.97468 | 0.92816 | 0.96226 | 0.9929217709 | 0.89589 | 0.9981340766 | 0.292094 | 5.147277554 | 0 |
| 10 | 87931078 | T | A | F | Y | PTEN | ENST00000371953 | 0.92642 | 0.97662 | 0.97831 | 0.95745 | 0.94765 | 0.9651437078 | 0.89268 | 0.9954876304 | 0.263628 | 5.787430645 | 0 |
| 10 | 87931078 | T | C | F | S | PTEN | ENST00000371953 | 0.9502 | 0.99406 | 0.97406 | 0.96764 | 0.98513 | 0.9960269188 | 0.95678 | 0.9998602867 | 0.441671 | 5.763746914 | 1 |
| 10 | 87931079 | T | G | F | L | PTEN | ENST00000371953 | 0.92482 | 0.97613 | 0.98351 | 0.90678 | 0.96189 | 0.9794629809 | 0.90054 | 0.9956794381 | 0.265499 | 3.294338659 | 0 |
| 10 | 87931080 | A | C | N | H | PTEN | ENST00000371953 | 0.46805 | 0.80172 | 0.86196 | 0.90547 | 0.74587 | 0.8258461137 | 0.85281 | 0.7913182974 | 0.176886 | 1.814530134 | 0 |
| 10 | 87931080 | A | G | N | D | PTEN | ENST00000371953 | 0.59298 | 0.82971 | 0.89354 | 0.90307 | 0.82159 | 0.8257821059 | 0.91985 | 0.9322582483 | 0.17404 | 3.343690032 | 0 |
| 10 | 87931080 | A | T | N | Y | PTEN | ENST00000371953 | 0.73701 | 0.84808 | 0.89325 | 0.94284 | 0.92804 | 0.890720796 | 0.95528 | 0.8924735785 | 0.33529 | 3.07486674 | 0 |
| 10 | 87931081 | A | C | N | T | PTEN | ENST00000371953 | 0.52679 | 0.82747 | 0.83224 | 0.89754 | 0.88684 | 0.8358772795 | 0.93753 | 0.9644807577 | 0.221861 | 4.866703691 | 0 |
| 10 | 87931081 | A | G | N | S | PTEN | ENST00000371953 | 0.46158 | 0.72306 | 0.80086 | 0.87704 | 0.75485 | 0.6925039274 | 0.88171 | 0.8637894392 | 0.0669064 | 3.413095962 | 0 |
| 10 | 87931081 | A | T | N | I | PTEN | ENST00000371953 | 0.72212 | 0.90984 | 0.85366 | 0.954 | 0.94281 | 0.9530257345 | 0.96139 | 0.9886455536 | 0.349345 | 6.680168985 | 0 |
| 10 | 87931082 | T | A | N | K | PTEN | ENST00000371953 | 0.34637 | 0.83503 | 0.91886 | 0.89024 | 0.71861 | 0.888316401 | 0.82773 | 0.2305511981 | 0.0985002 | 2.822224291 | 0 |
| 10 | 87931083 | T | A | C | S | PTEN | ENST00000371953 | 0.43744 | 0.84808 | 0.89452 | 0.87328 | 0.8278 | 0.9523522345 | 0.86745 | 0.849611342 | 0.202948 | 3.387275051 | 0 |
| 10 | 87931083 | T | C | C | R | PTEN | ENST00000371953 | 0.58874 | 0.92513 | 0.96541 | 0.87698 | 0.87148 | 0.9849066846 | 0.96822 | 0.8058370352 | 0.35752 | 4.099814809 | 0 |
| 10 | 87931083 | T | G | C | G | PTEN | ENST00000371953 | 0.45407 | 0.85069 | 0.86724 | 0.86733 | 0.79089 | 0.9662418101 | 0.95506 | 0.7688049078 | 0.331585 | 2.794089776 | 0 |
| 10 | 87931084 | G | A | C | Y | PTEN | ENST00000371953 | 0.69852 | 0.91795 | 0.97176 | 0.84745 | 0.84853 | 0.9785598627 | 0.93568 | 0.9435546994 | 0.365212 | 3.766190292 | 0 |
| 10 | 87931084 | G | C | C | S | PTEN | ENST00000371953 | 0.56352 | 0.84808 | 0.89452 | 0.86063 | 0.825 | 0.9523522345 | 0.85298 | 0.8746880293 | 0.213318 | 3.387275051 | 0 |

| | | | | | | | | | | | | | | | | |
|---|---|---|---|---|---|---|---|---|---|---|---|---|---|---|---|---|
| 10 | 87931084 | G | T | C | F | PTEN | ENST00000371953 | 0.77045 | 0.8993 | 0.91172 | 0.85143 | 0.88776 | 0.9757186917 | 0.96201 | 0.8783234954 | 0.37169 | 4.653197178 | 0 |
| 10 | 87931085 | C | G | C | W | PTEN | ENST00000371953 | 0.7282 | 0.9478 | 0.94611 | 0.84121 | 0.87148 | 0.981293716 | 0.96097 | 0.9633542895 | 0.348464 | 6.78733478 | 0 |
| 10 | 87931087 | G | A | R | K | PTEN | ENST00000371953 | 0.7741 | 0.83147 | 0.97616 | 0.85217 | 0.85741 | 0.9591275697 | 0.96273 | 0.9498429298 | 0.266086 | 2.967791228 | 0 |
| 10 | 87931087 | G | C | R | T | PTEN | ENST00000371953 | 0.91868 | 0.96697 | 0.9697 | 0.93744 | 0.94205 | 0.9764274593 | 0.93946 | 0.9970074296 | 0.391409 | 4.521514006 | 0 |
| 10 | 87931087 | G | T | R | I | PTEN | ENST00000371953 | 0.94201 | 0.99192 | 0.95981 | 0.94207 | 0.94469 | 0.9705953849 | 0.95176 | 0.9994914532 | 0.418409 | 4.486115212 | 0 |
| 10 | 87931088 | A | C | R | S | PTEN | ENST00000371953 | 0.88486 | 0.92685 | 0.95316 | 0.95981 | 0.96956 | 0.979231476 | 0.97695 | 0.9962323308 | 0.418393 | 3.989755723 | 0 |
| 10 | 87931088 | A | T | R | S | PTEN | ENST00000371953 | 0.87951 | 0.92685 | 0.95316 | 0.95993 | 0.97067 | 0.979231476 | 0.97694 | 0.9962323308 | 0.418392 | 3.989755723 | 0 |
| 10 | 87931089 | G | C | V | L | PTEN | ENST00000371953 | 0.80376 | 0.95887 | 0.95269 | 0.98664 | 0.9796 | 0.9709664316 | 0.96752 | 0.9971354008 | 0.507088 | 5.427630844 | 0 |
| 10 | 87931089 | G | T | V | F | PTEN | ENST00000371953 | 0.82879 | 0.9811 | 0.93968 | 0.98616 | 0.98728 | 0.9944182968 | 0.9677 | 0.9918612242 | 0.530982 | 4.637830286 | 0 |
| 10 | 87933013 | T | A | V | D | PTEN | ENST00000371953 | 0.92778 | 0.99163 | 0.96134 | 0.99264 | 0.9999 | 0.9990262639 | 0.99063 | 0.9994037151 | 0.586488 | 7.395875797 | 0 |
| 10 | 87933013 | T | C | V | A | PTEN | ENST00000371953 | 0.90223 | 0.97597 | 0.95777 | 0.99101 | 0.99603 | 0.9676918311 | 0.94286 | 0.9925757176 | 0.444155 | 4.831785794 | 0 |
| 10 | 87933013 | T | G | V | G | PTEN | ENST00000371953 | 0.92076 | 0.98348 | 0.88628 | 0.98813 | 0.99722 | 0.9968386387 | 0.98564 | 0.9995108843 | 0.582951 | 7.418799268 | 0 |
| 10 | 87933015 | G | A | A | T | PTEN | ENST00000371953 | 0.82334 | 0.89018 | 0.94767 | 0.9819 | 0.92192 | 0.888880738 | 0.88032 | 0.9789258838 | 0.348997 | 5.679468423 | 0 |
| 10 | 87933015 | G | C | A | P | PTEN | ENST00000371953 | 0.84468 | 0.98069 | 0.97069 | 0.98699 | 0.98904 | 0.9942268932 | 0.98848 | 0.992816925 | 0.548541 | 7.651372678 | 1 |
| 10 | 87933015 | G | T | A | S | PTEN | ENST00000371953 | 0.64929 | 0.8119 | 0.92202 | 0.96923 | 0.87247 | 0.8747755494 | 0.90173 | 0.9677501321 | 0.3188 | 4.751436248 | 0 |
| 10 | 87933016 | C | A | A | E | PTEN | ENST00000371953 | 0.83019 | 0.88639 | 0.97182 | 0.9738 | 0.96298 | 0.9873615367 | 0.98692 | 0.9879498482 | 0.528893 | 4.190199597 | 0 |
| 10 | 87933016 | C | G | A | G | PTEN | ENST00000371953 | 0.84583 | 0.89656 | 0.93589 | 0.97186 | 0.95243 | 0.9166505109 | 0.92939 | 0.976755619 | 0.380185 | 5.164152131 | 0 |
| 10 | 87933016 | C | T | A | V | PTEN | ENST00000371953 | 0.66763 | 0.87234 | 0.95714 | 0.96633 | 0.93123 | 0.9185008147 | 0.94491 | 0.9804387093 | 0.374161 | 3.950112956 | 0 |
| 10 | 87933018 | C | A | Q | K | PTEN | ENST00000371953 | 0.38771 | 0.85798 | 0.88534 | 0.80096 | 0.84106 | 0.8439378929 | 0.94049 | 0.8586404324 | 0.215932 | 4.342350296 | 0 |
| 10 | 87933018 | C | G | Q | E | PTEN | ENST00000371953 | 0.35311 | 0.81614 | 0.8323 | 0.73581 | 0.82948 | 0.838401276 | 0.9361 | 0.8939223886 | 0.209493 | 3.947399677 | 0 |
| 10 | 87933019 | A | C | Q | P | PTEN | ENST00000371953 | 0.68403 | 0.91936 | 0.84319 | 0.82211 | 0.94876 | 0.9817338796 | 0.97909 | 0.9363433123 | 0.396152 | 6.707565362 | 0 |
| 10 | 87933019 | A | G | Q | R | PTEN | ENST00000371953 | 0.60259 | 0.8144 | 0.87706 | 0.76044 | 0.8859 | 0.8255134193 | 0.98353 | 0.9519307613 | 0.346643 | 3.887268282 | 0 |
| 10 | 87933019 | A | T | Q | L | PTEN | ENST00000371953 | 0.53462 | 0.80294 | 0.83006 | 0.84326 | 0.90624 | 0.8777796867 | 0.96205 | 0.9573414922 | 0.295068 | 4.781964973 | 0 |
| 10 | 87933020 | A | T | Q | H | PTEN | ENST00000371953 | 0.3699 | 0.77455 | 0.8786 | 0.74224 | 0.77591 | 0.8275404467 | 0.97324 | 0.6436983943 | 0.184147 | 3.261337099 | 0 |
| 10 | 87933021 | T | A | Y | N | PTEN | ENST00000371953 | 0.8456 | 0.98981 | 0.98988 | 0.98652 | 0.99356 | 0.9918618695 | 0.97104 | 0.999515295 | 0.521462 | 7.413292902 | 1 |
| 10 | 87933021 | T | C | Y | H | PTEN | ENST00000371953 | 0.83974 | 0.98196 | 0.99116 | 0.97299 | 0.99676 | 0.9801508303 | 0.99207 | 0.9972701669 | 0.56364 | 7.123603624 | 0 |
| 10 | 87933021 | T | G | Y | D | PTEN | ENST00000371953 | 0.85204 | 0.99282 | 0.98949 | 0.98854 | 0.99676 | 0.9974396066 | 0.99674 | 0.9997261167 | 0.581237 | 8.522902786 | 1 |
| 10 | 87933022 | A | T | Y | F | PTEN | ENST00000371953 | 0.51435 | 0.78215 | 0.96967 | 0.8893 | 0.79516 | 0.9336910506 | 0.89171 | 0.9504178166 | 0.218349 | 2.89191738 | 0 |
| 10 | 87933024 | C | A | P | T | PTEN | ENST00000371953 | 0.67247 | 0.97507 | 0.97161 | 0.98495 | 0.99471 | 0.9660259491 | 0.9824 | 0.9962180257 | 0.556945 | 6.674460811 | 0 |
| 10 | 87933024 | C | G | P | A | PTEN | ENST00000371953 | 0.37346 | 0.94498 | 0.95505 | 0.97434 | 0.97104 | 0.9422118782 | 0.95677 | 0.8960469365 | 0.440627 | 3.678413705 | 0 |
| 10 | 87933024 | C | T | P | S | PTEN | ENST00000371953 | 0.69516 | 0.96823 | 0.96173 | 0.9744 | 0.98621 | 0.9688620936 | 0.9587 | 0.994370997 | 0.530071 | 4.932592692 | 0 |
| 10 | 87933025 | C | A | P | H | PTEN | ENST00000371953 | 0.83583 | 0.97742 | 0.97894 | 0.98745 | 0.99105 | 0.9599156172 | 0.96918 | 0.9991735816 | 0.577476 | 6.873575073 | 0 |
| 10 | 87933025 | C | G | P | R | PTEN | ENST00000371953 | 0.83905 | 0.97054 | 0.98227 | 0.98701 | 0.99578 | 0.9757413175 | 0.98835 | 0.9971820116 | 0.562167 | 5.895197251 | 0 |
| 10 | 87933025 | C | T | P | L | PTEN | ENST00000371953 | 0.84816 | 0.94728 | 0.98356 | 0.98369 | 0.99105 | 0.9737871113 | 0.97542 | 0.9978122711 | 0.538869 | 5.78284911 | 0 |
| 10 | 87933027 | T | A | F | I | PTEN | ENST00000371953 | 0.85189 | 0.98003 | 0.90299 | 0.97093 | 0.97475 | 0.9912040272 | 0.95878 | 0.9978300929 | 0.398264 | 4.05556549 | 0 |
| 10 | 87933027 | T | G | F | V | PTEN | ENST00000371953 | 0.85793 | 0.98258 | 0.88965 | 0.98217 | 0.99356 | 0.9911979541 | 0.9709 | 0.9986608028 | 0.523678 | 5.073515208 | 0 |
| 10 | 87933028 | T | A | F | Y | PTEN | ENST00000371953 | 0.92092 | 0.9764 | 0.91823 | 0.98537 | 0.99006 | 0.968394691 | 0.96643 | 0.9954168797 | 0.47021 | 6.749709078 | 0 |
| 10 | 87933028 | T | C | F | S | PTEN | ENST00000371953 | 0.9488 | 0.99351 | 0.89947 | 0.99111 | 0.99919 | 0.9964143944 | 0.9924 | 0.9997716546 | 0.588228 | 6.582825505 | 0 |
| 10 | 87933028 | T | G | F | C | PTEN | ENST00000371953 | 0.9487 | 0.98704 | 0.92635 | 0.98881 | 0.99892 | 0.9964064623 | 0.98436 | 0.9997010231 | 0.584913 | 6.636197628 | 0 |
| 10 | 87933029 | T | G | F | L | PTEN | ENST00000371953 | 0.83529 | 0.95105 | 0.93112 | 0.96969 | 0.98904 | 0.9766865338 | 0.98071 | 0.996850431 | 0.528699 | 5.487530711 | 0 |
| 10 | 87933030 | G | A | E | K | PTEN | ENST00000371953 | 0.76934 | 0.83016 | 0.95065 | 0.96286 | 0.95097 | 0.947855675 | 0.96778 | 0.9646247029 | 0.40825 | 4.137780934 | 0 |
| 10 | 87933030 | G | C | E | Q | PTEN | ENST00000371953 | 0.73573 | 0.8511 | 0.87415 | 0.95944 | 0.90798 | 0.8981726746 | 0.95169 | 0.9237063527 | 0.334503 | 4.303650928 | 0 |
| 10 | 87933031 | A | C | E | A | PTEN | ENST00000371953 | 0.86032 | 0.90467 | 0.88692 | 0.97732 | 0.9755 | 0.9220094402 | 0.97022 | 0.9853500128 | 0.397901 | 4.951215944 | 0 |
| 10 | 87933031 | A | G | E | G | PTEN | ENST00000371953 | 0.89684 | 0.94186 | 0.84458 | 0.9831 | 0.97475 | 0.9397543545 | 0.98432 | 0.9943625927 | 0.512172 | 5.573862734 | 0 |
| 10 | 87933031 | A | T | E | V | PTEN | ENST00000371953 | 0.76761 | 0.90338 | 0.87013 | 0.97414 | 0.9755 | 0.9509320828 | 0.98865 | 0.9915931225 | 0.493064 | 4.299510291 | 0 |
| 10 | 87933032 | A | T | E | D | PTEN | ENST00000371953 | 0.36041 | 0.76381 | 0.88819 | 0.87625 | 0.81646 | 0.8528978236 | 0.96338 | 0.8088243008 | 0.261004 | 2.156707104 | 0 |
| 10 | 87933033 | G | A | D | N | PTEN | ENST00000371953 | 0.85945 | 0.96723 | 0.95114 | 0.99619 | 0.98146 | 0.973899483 | 0.98771 | 0.9992693067 | 0.289706 | 6.554277007 | 0 |
| 10 | 87933033 | G | C | D | H | PTEN | ENST00000371953 | 0.85471 | 0.97893 | 0.96065 | 0.99651 | 0.99876 | 0.9857349297 | 0.99469 | 0.9998083711 | 0.595751 | 7.777346394 | 1 |
| 10 | 87933033 | G | T | D | Y | PTEN | ENST00000371953 | 0.86219 | 0.98086 | 0.95175 | 0.99602 | 0.99386 | 0.9939206529 | 0.99492 | 0.9999057055 | 0.595772 | 8.584320286 | 1 |
| 10 | 87933034 | A | C | D | A | PTEN | ENST00000371953 | 0.91699 | 0.98134 | 0.91494 | 0.99422 | 0.9997 | 0.9905017595 | 0.99517 | 0.9997549653 | 0.594973 | 5.939529007 | 1 |
| 10 | 87933034 | A | G | D | G | PTEN | ENST00000371953 | 0.9204 | 0.98373 | 0.92348 | 0.99218 | 0.99806 | 0.9875518249 | 0.99537 | 0.9994540811 | 0.576798 | 5.92145838 | 1 |
| 10 | 87933034 | A | T | D | V | PTEN | ENST00000371953 | 0.91218 | 0.9875 | 0.91645 | 0.99506 | 0.99919 | 0.9935006363 | 0.99058 | 0.9997691512 | 0.594972 | 6.741277927 | 1 |
| 10 | 87933035 | C | A | D | E | PTEN | ENST00000371953 | 0.64352 | 0.96387 | 0.9371 | 0.98695 | 0.97141 | 0.9639787333 | 0.998 | 0.9944531322 | 0.552338 | 6.109882464 | 1 |
| 10 | 87933036 | C | A | H | N | PTEN | ENST00000371953 | 0.66454 | 0.97114 | 0.93714 | 0.98352 | 0.96919 | 0.9795348932 | 0.9715 | 0.9983530045 | 0.331461 | 5.695461354 | 0 |
| 10 | 87933036 | C | G | H | D | PTEN | ENST00000371953 | 0.70109 | 0.9858 | 0.93364 | 0.98547 | 0.99356 | 0.9971228866 | 0.98278 | 0.9997199178 | 0.585379 | 7.026023209 | 1 |
| 10 | 87933036 | C | T | H | Y | PTEN | ENST00000371953 | 0.65672 | 0.97553 | 0.93872 | 0.98713 | 0.99765 | 0.9888667934 | 0.99444 | 0.9978973866 | 0.579418 | 6.288537765 | 1 |
| 10 | 87933037 | A | C | H | P | PTEN | ENST00000371953 | 0.86996 | 0.99048 | 0.88669 | 0.99335 | 0.99744 | 0.9955170463 | 0.98159 | 0.9999091625 | 0.594973 | 8.039277099 | 1 |
| 10 | 87933037 | A | G | H | R | PTEN | ENST00000371953 | 0.82193 | 0.97582 | 0.939 | 0.98874 | 0.99603 | 0.9900926634 | 0.99649 | 0.9990561604 | 0.590493 | 6.222103808 | 0 |
| 10 | 87933037 | A | T | H | L | PTEN | ENST00000371953 | 0.87255 | 0.98069 | 0.91375 | 0.99157 | 0.99414 | 0.9914277376 | 0.97957 | 0.9996641874 | 0.539235 | 6.573673381 | 1 |
| 10 | 87933038 | T | A | H | Q | PTEN | ENST00000371953 | 0.72764 | 0.96883 | 0.9202 | 0.98531 | 0.96993 | 0.9854246132 | 0.99111 | 0.9989715815 | 0.524637 | 7.198347385 | 0 |
| 10 | 87933039 | A | C | N | H | PTEN | ENST00000371953 | 0.45009 | 0.88537 | 0.79144 | 0.83279 | 0.81124 | 0.943043387 | 0.91721 | 0.9868152142 | 0.112591 | 4.189555482 | 0 |
| 10 | 87933039 | A | G | N | D | PTEN | ENST00000371953 | 0.76044 | 0.97175 | 0.83138 | 0.93216 | 0.91357 | 0.9555472385 | 0.96098 | 0.9964677095 | 0.114873 | 7.155405175 | 0 |
| 10 | 87933039 | A | T | N | Y | PTEN | ENST00000371953 | 0.78717 | 0.97448 | 0.82488 | 0.94765 | 0.95097 | 0.9743307367 | 0.9436 | 0.9967669249 | 0.31904 | 6.160995798 | 0 |
| 10 | 87933040 | A | C | N | T | PTEN | ENST00000371953 | 0.85115 | 0.92856 | 0.76967 | 0.90027 | 0.90494 | 0.9313598762 | 0.92978 | 0.9924135804 | 0.143901 | 5.772716863 | 0 |
| 10 | 87933040 | A | G | N | S | PTEN | ENST00000371953 | 0.80061 | 0.91476 | 0.71652 | 0.8906 | 0.88022 | 0.8982003406 | 0.94854 | 0.9913724661 | -0.0126171 | 4.997210068 | 0 |
| 10 | 87933041 | C | A | N | K | PTEN | ENST00000371953 | 0.6243 | 0.93746 | 0.86499 | 0.93994 | 0.86453 | 0.9611631489 | 0.94608 | 0.9932717681 | 0.0896677 | 6.326127973 | 0 |
| 10 | 87933042 | C | A | P | T | PTEN | ENST00000371953 | 0.64143 | 0.97143 | 0.85938 | 0.974 | 0.95572 | 0.9850215212 | 0.92646 | 0.9970267415 | 0.416644 | 6.120889164 | 0 |
| 10 | 87933042 | C | G | P | A | PTEN | ENST00000371953 | 0.49589 | 0.86197 | 0.81266 | 0.95371 | 0.92438 | 0.9640297826 | 0.88145 | 0.9828936458 | 0.32851 | 4.277316259 | 0 |
| 10 | 87933042 | C | T | P | S | PTEN | ENST00000371953 | 0.66652 | 0.90632 | 0.82777 | 0.97494 | 0.94543 | 0.9827439023 | 0.89467 | 0.9944977164 | 0.368501 | 5.257736248 | 0 |
| 10 | 87933043 | C | A | P | Q | PTEN | ENST00000371953 | 0.79239 | 0.96146 | 0.82968 | 0.99248 | 0.97141 | 0.9896855227 | 0.9493 | 0.99896276 | 0.514729 | 8.441714795 | 1 |
| 10 | 87933043 | C | G | P | R | PTEN | ENST00000371953 | 0.80787 | 0.97054 | 0.89107 | 0.99294 | 0.97178 | 0.9937112643 | 0.97139 | 0.9989136457 | 0.580146 | 8.441714795 | 1 |
| 10 | 87933043 | C | T | P | L | PTEN | ENST00000371953 | 0.82607 | 0.96273 | 0.89375 | 0.98303 | 0.9833 | 0.9901468524 | 0.98492 | 0.9992215633 | 0.574774 | 6.692582019 | 1 |
| 10 | 87933045 | C | G | P | A | PTEN | ENST00000371953 | 0.61529 | 0.94445 | 0.77005 | 0.9871 | 0.96226 | 0.9923654073 | 0.96869 | 0.9964942336 | 0.316509 | 5.353058653 | 0 |
| 10 | 87933046 | C | G | P | R | PTEN | ENST00000371953 | 0.81567 | 0.97198 | 0.84955 | 0.99637 | 0.99552 | 0.9989508813 | 0.99487 | 0.9997708201 | 0.584427 | 7.707176055 | 1 |
| 10 | 87933046 | C | T | P | L | PTEN | ENST00000371953 | 0.83375 | 0.96948 | 0.86561 | 0.99439 | 0.98477 | 0.9979236998 | 0.98124 | 0.9998832941 | 0.543527 | 6.271279409 | 1 |
| 10 | 87933048 | C | A | Q | K | PTEN | ENST00000371953 | 0.28237 | 0.85872 | 0.88565 | 0.91137 | 0.7816 | 0.9238420343 | 0.97984 | 0.7311950922 | 0.318529 | 2.734189117 | 0 |
| 10 | 87933048 | C | G | Q | E | PTEN | ENST00000371953 | 0.3365 | 0.82641 | 0.84314 | 0.89605 | 0.89193 | 0.9207314193 | 0.97009 | 0.6925677657 | 0.339717 | 4.350780427 | 0 |
| 10 | 87933049 | A | C | Q | P | PTEN | ENST00000371953 | 0.69163 | 0.84619 | 0.84724 | 0.94231 | 0.90318 | 0.9502824618 | 0.96435 | 0.8928151131 | 0.358328 | 3.433463017 | 0 |
| 10 | 87933049 | A | G | Q | R | PTEN | ENST00000371953 | 0.47696 | 0.81593 | 0.88067 | 0.88557 | 0.85117 | 0.9001426211 | 0.99524 | 0.7787133455 | 0.411654 | 2.903967433 | 0 |
| 10 | 87933049 | A | T | Q | L | PTEN | ENST00000371953 | 0.74356 | 0.78709 | 0.83166 | 0.95672 | 0.9517 | 0.9253092279 | 0.98211 | 0.9550766349 | 0.442911 | 4.59324022 | 0 |
| 10 | 87933050 | G | T | Q | H | PTEN | ENST00000371953 | 0.57843 | 0.79446 | 0.87978 | 0.92833 | 0.91862 | 0.9141875752 | 0.96796 | 0.8696704507 | 0.328786 | 4.899090737 | 0 |
| 10 | 87933051 | C | A | L | I | PTEN | ENST00000371953 | 0.50192 | 0.8288 | 0.90088 | 0.76788 | 0.84906 | 0.9253784549 | 0.90243 | 0.8124549389 | 0.176621 | 2.785437891 | 0 |
| 10 | 87933051 | C | G | L | V | PTEN | ENST00000371953 | 0.54896 | 0.89831 | 0.85171 | 0.87449 | 0.92233 | 0.9675299939 | 0.95727 | 0.9807850122 | 0.299477 | 4.010209018 | 0 |
| 10 | 87933052 | T | A | L | Q | PTEN | ENST00000371953 | 0.85938 | 0.96965 | 0.90302 | 0.96227 | 0.98071 | 0.993431974 | 0.98718 | 0.995562911 | 0.47147 | 7.374851656 | 0 |
| 10 | 87933052 | T | C | L | P | PTEN | ENST00000371953 | 0.87315 | 0.99078 | 0.95822 | 0.94605 | 0.97698 | 0.9992350466 | 0.98621 | 0.995562911 | 0.472118 | 7.374851656 | 0 |
| 10 | 87933052 | T | G | L | R | PTEN | ENST00000371953 | 0.869 | 0.98736 | 0.94685 | 0.96333 | 0.97364 | 0.9987580868 | 0.98988 | 0.9964444041 | 0.474689 | 7.350959732 | 1 |
| 10 | 87933054 | G | A | E | K | PTEN | ENST00000371953 | 0.87834 | 0.89783 | 0.92343 | 0.87639 | 0.9248 | 0.9439359153 | 0.98765 | 0.9652585983 | 0.42454 | 5.502276021 | 0 |
| 10 | 87933054 | G | C | E | Q | PTEN | ENST00000371953 | 0.67883 | 0.81956 | 0.83564 | 0.83265 | 0.83339 | 0.9082002186 | 0.94587 | 0.8026047945 | 0.241626 | 4.486847754 | 0 |
| 10 | 87933055 | A | C | E | A | PTEN | ENST00000371953 | 0.90925 | 0.84903 | 0.84326 | 0.85009 | 0.94913 | 0.9331252264 | 0.96593 | 0.983453095 | 0.377783 | 5.309371498 | 0 |
| 10 | 87933055 | A | G | E | G | PTEN | ENST00000371953 | 0.9325 | 0.85586 | 0.80261 | 0.85325 | 0.95535 | 0.940439188 | 0.96082 | 0.9831910729 | 0.3911 | 5.419903657 | 0 |
| 10 | 87933055 | A | T | E | V | PTEN | ENST00000371953 | 0.92705 | 0.92004 | 0.83007 | 0.93391 | 0.9781 | 0.9484545911 | 0.97426 | 0.9969784021 | 0.430294 | 6.157837645 | 0 |
| 10 | 87933056 | A | T | E | D | PTEN | ENST00000371953 | 0.63558 | 0.77256 | 0.84916 | 0.77937 | 0.848 | 0.8878261384 | 0.96216 | 0.7696455717 | 0.197818 | 4.074742179 | 0 |
| 10 | 87933057 | C | A | L | I | PTEN | ENST00000371953 | 0.75543 | 0.93697 | 0.88644 | 0.85895 | 0.80118 | 0.8547573402 | 0.80371 | 0.9255649447 | 0.158056 | 4.281520384 | 0 |
| 10 | 87933057 | C | G | L | V | PTEN | ENST00000371953 | 0.62053 | 0.92739 | 0.8487 | 0.9129 | 0.87974 | 0.9219924054 | 0.89249 | 0.9719527364 | 0.263335 | 4.549192667 | 0 |
| 10 | 87933057 | C | T | L | F | PTEN | ENST00000371953 | 0.66256 | 0.90901 | 0.87942 | 0.90299 | 0.88496 | 0.9197243573 | 0.87476 | 0.9914535284 | 0.315276 | 6.218691791 | 0 |
| 10 | 87933058 | T | A | L | H | PTEN | ENST00000371953 | 0.84763 | 0.98228 | 0.93327 | 0.96356 | 0.98621 | 0.9682786562 | 0.95821 | 0.9985205531 | 0.448055 | 6.591302502 | 0 |
| 10 | 87933058 | T | C | L | P | PTEN | ENST00000371953 | 0.86727 | 0.99114 | 0.95443 | 0.96537 | 0.99443 | 0.9966986286 | 0.9766 | 0.9979052544 | 0.461771 | 8.419848496 | 1 |
| 10 | 87933058 | T | G | L | R | PTEN | ENST00000371953 | 0.86608 | 0.9858 | 0.94123 | 0.96027 | 0.96444 | 0.9886222612 | 0.97311 | 0.9977789521 | 0.432721 | 5.889267096 | 0 |
| 10 | 87933060 | A | C | I | L | PTEN | ENST00000371953 | 0.74906 | 0.84735 | 0.89886 | 0.93711 | 0.93123 | 0.9582112313 | 0.93351 | 0.9203950763 | 0.32406 | 4.314068856 | 1 |
| 10 | 87933060 | A | T | I | F | PTEN | ENST00000371953 | 0.76942 | 0.97569 | 0.909 | 0.97661 | 0.98183 | 0.990518326 | 0.99063 | 0.9935035642 | 0.560212 | 4.731535081 | 1 |
| 10 | 87933061 | T | A | I | N | PTEN | ENST00000371953 | 0.91987 | 0.99351 | 0.9525 | 0.9875 | 0.99234 | 0.9981673783 | 0.97372 | 0.9995384216 | 0.476135 | 7.285895026 | 1 |

| | | | | | | | | | | | | | | | | |
|---|---|---|---|---|---|---|---|---|---|---|---|---|---|---|---|---|
| 10 | 87933061 | T | C | I | T | PTEN | ENST00000371953 | 0.88354 | 0.97924 | 0.9592 | 0.97769 | 0.99876 | 0.9832664993 | 0.99571 | 0.9974023413 | 0.574281 | 6.149980507 | 1 |
| 10 | 87933061 | T | G | I | S | PTEN | ENST00000371953 | 0.92198 | 0.97924 | 0.91321 | 0.98547 | 0.99824 | 0.9976686603 | 0.99644 | 0.9986258745 | 0.585677 | 7.763599678 | 1 |
| 10 | 87933063 | A | C | K | Q | PTEN | ENST00000371953 | 0.56505 | 0.81831 | 0.84665 | 0.65686 | 0.86149 | 0.9209884419 | 0.95369 | 0.7946170568 | 0.189879 | 3.624218323 | 0 |
| 10 | 87933063 | A | G | K | E | PTEN | ENST00000371953 | 0.61067 | 0.88308 | 0.88427 | 0.74471 | 0.89558 | 0.9446484837 | 0.95142 | 0.8841635585 | 0.234897 | 4.175106886 | 0 |
| 10 | 87933064 | A | C | K | T | PTEN | ENST00000371953 | 0.81928 | 0.85125 | 0.88644 | 0.83466 | 0.93319 | 0.9420267404 | 0.96358 | 0.9792449474 | 0.325145 | 5.353549124 | 0 |
| 10 | 87933064 | A | T | K | I | PTEN | ENST00000371953 | 0.87133 | 0.928 | 0.8511 | 0.86335 | 0.90274 | 0.9520562812 | 0.94962 | 0.9857586622 | 0.256238 | 4.993230824 | 1 |
| 10 | 87933066 | C | A | P | T | PTEN | ENST00000371953 | 0.46712 | 0.84503 | 0.89419 | 0.96653 | 0.92964 | 0.9040553713 | 0.97543 | 0.9561649561 | 0.450083 | 4.777583468 | 0 |
| 10 | 87933066 | C | G | P | A | PTEN | ENST00000371953 | 0.59349 | 0.80193 | 0.84993 | 0.9637 | 0.88543 | 0.8632021103 | 0.93502 | 0.9834854603 | 0.357014 | 4.701995065 | 0 |
| 10 | 87933066 | C | T | P | S | PTEN | ENST00000371953 | 0.64291 | 0.8243 | 0.87033 | 0.96282 | 0.90885 | 0.9061188094 | 0.94328 | 0.980656147 | 0.3697 | 4.798062793 | 0 |
| 10 | 87933067 | C | A | P | H | PTEN | ENST00000371953 | 0.77029 | 0.86927 | 0.90655 | 0.9703 | 0.92884 | 0.8925365069 | 0.91912 | 0.9955139756 | 0.387158 | 6.129447594 | 0 |
| 10 | 87933067 | C | G | P | R | PTEN | ENST00000371953 | 0.64081 | 0.83617 | 0.9111 | 0.96581 | 0.91694 | 0.9400113771 | 0.96824 | 0.9536993504 | 0.484333 | 5.177911327 | 0 |
| 10 | 87933069 | T | A | F | I | PTEN | ENST00000371953 | 0.79334 | 0.98196 | 0.95418 | 0.89849 | 0.96919 | 0.9864583497 | 0.97865 | 0.9973262548 | 0.421144 | 5.859872645 | 1 |
| 10 | 87933070 | T | A | F | Y | PTEN | ENST00000371953 | 0.86935 | 0.9764 | 0.96197 | 0.93569 | 0.97141 | 0.9473361903 | 0.93975 | 0.9943962653 | 0.33868 | 5.572477819 | 0 |
| 10 | 87933070 | T | C | F | S | PTEN | ENST00000371953 | 0.92782 | 0.99329 | 0.95588 | 0.9701 | 0.98869 | 0.9954035454 | 0.99104 | 0.9996826649 | 0.474169 | 7.550734342 | 1 |
| 10 | 87933071 | T | G | F | L | PTEN | ENST00000371953 | 0.73021 | 0.95345 | 0.97012 | 0.9008 | 0.91357 | 0.9710028009 | 0.97916 | 0.9960933328 | 0.397532 | 4.732748162 | 0 |
| 10 | 87933072 | T | C | C | R | PTEN | ENST00000371953 | 0.83798 | 0.99403 | 0.97054 | 0.96393 | 0.97178 | 0.9967948055 | 0.98901 | 0.9997389913 | 0.473053 | 8.62930684 | 1 |
| 10 | 87933072 | T | G | C | G | PTEN | ENST00000371953 | 0.82201 | 0.98134 | 0.88793 | 0.96251 | 0.9703 | 0.99035642 | 0.98973 | 0.9991233945 | 0.469457 | 7.756964569 | 1 |
| 10 | 87933073 | G | C | C | S | PTEN | ENST00000371953 | 0.90473 | 0.98069 | 0.91783 | 0.94649 | 0.98693 | 0.9867160746 | 0.98093 | 0.9998127818 | 0.454293 | 5.870118257 | 0 |
| 10 | 87933074 | T | G | C | W | PTEN | ENST00000371953 | 0.75002 | 0.9851 | 0.95457 | 0.98006 | 0.92924 | 0.9941266538 | 0.99488 | 0.9998269677 | 0.433636 | 8.62930684 | 1 |
| 10 | 87933075 | G | C | E | Q | PTEN | ENST00000371953 | 0.79374 | 0.80741 | 0.90115 | 0.97201 | 0.90052 | 0.8789536812 | 0.96297 | 0.8841635585 | 0.352403 | 5.006320324 | 0 |
| 10 | 87933076 | A | C | E | A | PTEN | ENST00000371953 | 0.89679 | 0.82678 | 0.91789 | 0.96565 | 0.95972 | 0.9141859778 | 0.98549 | 0.9886543751 | 0.486447 | 5.099203019 | 0 |
| 10 | 87933076 | A | T | E | V | PTEN | ENST00000371953 | 0.91797 | 0.88559 | 0.90014 | 0.97733 | 0.9703 | 0.9263830656 | 0.98658 | 0.9928082228 | 0.48574 | 5.334637747 | 0 |
| 10 | 87933078 | G | A | D | N | PTEN | ENST00000371953 | 0.71081 | 0.89783 | 0.95444 | 0.89069 | 0.83339 | 0.9422289681 | 0.95305 | 0.9965041876 | 0.0789808 | 5.636351931 | 0 |
| 10 | 87933078 | G | C | D | H | PTEN | ENST00000371953 | 0.83936 | 0.94597 | 0.9636 | 0.93902 | 0.93863 | 0.9599560902 | 0.96068 | 0.9988184571 | 0.443362 | 5.666333471 | 0 |
| 10 | 87933078 | G | T | D | Y | PTEN | ENST00000371953 | 0.84726 | 0.97972 | 0.95785 | 0.95845 | 0.9458 | 0.9784873306 | 0.98765 | 0.999570787 | 0.475209 | 8.361261092 | 1 |
| 10 | 87933079 | A | C | D | A | PTEN | ENST00000371953 | 0.90872 | 0.9521 | 0.92131 | 0.9166 | 0.95316 | 0.9622724746 | 0.95722 | 0.9986673594 | 0.417453 | 6.524990134 | 0 |
| 10 | 87933079 | A | G | D | G | PTEN | ENST00000371953 | 0.91264 | 0.97893 | 0.92797 | 0.9416 | 0.98585 | 0.9703923861 | 0.97865 | 0.997759819 | 0.439971 | 7.429137381 | 0 |
| 10 | 87933079 | A | T | D | V | PTEN | ENST00000371953 | 0.90329 | 0.98605 | 0.92499 | 0.95315 | 0.95863 | 0.9775924475 | 0.98719 | 0.9996792078 | 0.466003 | 7.528658277 | 1 |
| 10 | 87933080 | T | A | D | E | PTEN | ENST00000371953 | 0.78558 | 0.90217 | 0.94174 | 0.85535 | 0.88637 | 0.9137307349 | 0.95908 | 0.9814985394 | 0.2128 | 5.786740249 | 0 |
| 10 | 87933082 | T | A | L | H | PTEN | ENST00000371953 | 0.91793 | 0.98086 | 0.96237 | 0.98583 | 0.99552 | 0.9779439945 | 0.99082 | 0.9970688224 | 0.545358 | 7.82071933 | 1 |
| 10 | 87933082 | T | C | L | P | PTEN | ENST00000371953 | 0.92867 | 0.98906 | 0.97502 | 0.98188 | 0.99105 | 0.9969257267 | 0.99749 | 0.9923692942 | 0.539203 | 7.82071933 | 1 |
| 10 | 87933082 | T | G | L | R | PTEN | ENST00000371953 | 0.92599 | 0.98406 | 0.96432 | 0.97912 | 0.99296 | 0.9949034779 | 0.99496 | 0.998067677 | 0.543623 | 7.82071933 | 1 |
| 10 | 87933084 | G | A | D | N | PTEN | ENST00000371953 | 0.86587 | 0.90115 | 0.95568 | 0.98051 | 0.85378 | 0.9343853311 | 0.96583 | 0.9864814281 | 0.178415 | 5.38651971 | 0 |
| 10 | 87933084 | G | C | D | H | PTEN | ENST00000371953 | 0.68731 | 0.8636 | 0.96533 | 0.96701 | 0.88401 | 0.9551378807 | 0.96626 | 0.9645726681 | 0.485099 | 4.109593935 | 0 |
| 10 | 87933084 | G | T | D | Y | PTEN | ENST00000371953 | 0.87133 | 0.91778 | 0.9576 | 0.98179 | 0.9506 | 0.9740757522 | 0.99313 | 0.996468544 | 0.537193 | 5.551058877 | 0 |
| 10 | 87933085 | A | C | D | A | PTEN | ENST00000371953 | 0.82553 | 0.88909 | 0.92307 | 0.98679 | 0.97252 | 0.9609903362 | 0.97667 | 0.9945078492 | 0.529682 | 5.068429692 | 0 |
| 10 | 87933085 | A | G | D | G | PTEN | ENST00000371953 | 0.92836 | 0.9234 | 0.92835 | 0.99103 | 0.97773 | 0.9725071901 | 0.98578 | 0.9981677532 | 0.535418 | 5.943869465 | 0 |
| 10 | 87933085 | A | T | D | V | PTEN | ENST00000371953 | 0.92371 | 0.96424 | 0.91971 | 0.98971 | 0.97923 | 0.9747694956 | 0.99254 | 0.9969803691 | 0.542317 | 6.214021158 | 0 |
| 10 | 87933086 | C | A | D | E | PTEN | ENST00000371953 | 0.48183 | 0.82614 | 0.94315 | 0.93997 | 0.87829 | 0.9009470148 | 0.95818 | 0.8131477833 | 0.313665 | 5.030262958 | 0 |
| 10 | 87933087 | C | A | Q | K | PTEN | ENST00000371953 | 0.39145 | 0.857 | 0.71666 | 0.85529 | 0.65772 | 0.8081057438 | 0.80769 | 0.5790032148 | 0.103951 | 1.775054367 | 0 |
| 10 | 87933087 | C | G | Q | E | PTEN | ENST00000371953 | 0.31566 | 0.81464 | 0.65959 | 0.82293 | 0.7217 | 0.7564504696 | 0.78055 | 0.1180594149 | 0.00830777 | 2.535852204 | 0 |
| 10 | 87933088 | A | C | Q | P | PTEN | ENST00000371953 | 0.62772 | 0.912 | 0.66835 | 0.92282 | 0.91271 | 0.9695746345 | 0.97671 | 0.9147921205 | 0.353114 | 7.048430791 | 1 |
| 10 | 87933088 | A | G | Q | R | PTEN | ENST00000371953 | 0.46768 | 0.81651 | 0.70654 | 0.84512 | 0.74021 | 0.7234848302 | 0.91342 | 0.7550731897 | 0.106503 | 3.080140582 | 0 |
| 10 | 87933088 | A | T | Q | L | PTEN | ENST00000371953 | 0.5001 | 0.78607 | 0.64821 | 0.91395 | 0.75349 | 0.8462637379 | 0.92918 | 0.8999769092 | 0.184171 | 3.196433242 | 0 |
| 10 | 87933090 | T | A | W | R | PTEN | ENST00000371953 | 0.85085 | 0.99306 | 0.99494 | 0.98458 | 0.99414 | 0.9921273199 | 0.97714 | 0.9990443587 | 0.563053 | 7.802812799 | 1 |
| 10 | 87933090 | T | C | W | R | PTEN | ENST00000371953 | 0.86481 | 0.99306 | 0.99494 | 0.98433 | 0.99414 | 0.9921273199 | 0.9772 | 0.998840034 | 0.563049 | 7.802812799 | 1 |
| 10 | 87933090 | T | G | W | G | PTEN | ENST00000371953 | 0.88868 | 0.99282 | 0.98071 | 0.99292 | 0.99471 | 0.9860104881 | 0.99613 | 0.9997330308 | 0.576478 | 7.802812799 | 1 |
| 10 | 87933091 | G | C | W | S | PTEN | ENST00000371953 | 0.91595 | 0.98981 | 0.98611 | 0.98265 | 0.9796 | 0.9924574392 | 0.99113 | 0.9999229908 | 0.584223 | 7.802812799 | 1 |
| 10 | 87933092 | G | T | W | C | PTEN | ENST00000371953 | 0.95421 | 0.99011 | 0.98781 | 0.98506 | 0.98109 | 0.9932501688 | 0.99705 | 0.9998455048 | 0.588279 | 6.693396121 | 1 |
| 10 | 87933093 | C | G | L | V | PTEN | ENST00000371953 | 0.77045 | 0.9685 | 0.70011 | 0.97302 | 0.96956 | 0.9709622039 | 0.98591 | 0.9911366701 | 0.557447 | 5.43973403 | 0 |
| 10 | 87933094 | T | G | L | R | PTEN | ENST00000371953 | 0.91652 | 0.98834 | 0.8453 | 0.99126 | 0.99932 | 0.9991828544 | 0.99227 | 0.9997759461 | 0.586847 | 7.803417098 | 1 |
| 10 | 87933096 | A | T | S | C | PTEN | ENST00000371953 | 0.64252 | 0.80914 | 0.62175 | 0.97208 | 0.94876 | 0.8407436188 | 0.94519 | 0.9696310759 | 0.377918 | 4.1598911 | 0 |
| 10 | 87933097 | G | A | S | N | PTEN | ENST00000371953 | 0.74627 | 0.79187 | 0.68499 | 0.9597 | 0.74658 | 0.8346041473 | 0.89129 | 0.9165326357 | 0.159241 | 3.179319254 | 0 |
| 10 | 87933097 | G | C | S | T | PTEN | ENST00000371953 | 0.72148 | 0.78576 | 0.6154 | 0.95847 | 0.80948 | 0.7821349912 | 0.91178 | 0.886936307 | 0.241643 | 3.371220486 | 0 |
| 10 | 87933097 | G | T | S | I | PTEN | ENST00000371953 | 0.92726 | 0.90831 | 0.66853 | 0.98011 | 0.93436 | 0.8905912657 | 0.97258 | 0.9842810631 | 0.482685 | 5.294579098 | 0 |
| 10 | 87933099 | G | C | E | Q | PTEN | ENST00000371953 | 0.46494 | 0.79411 | 0.76757 | 0.93437 | 0.75143 | 0.826623764 | 0.91324 | 0.9805608392 | 0.219303 | 2.786754387 | 0 |
| 10 | 87933100 | A | C | E | A | PTEN | ENST00000371953 | 0.46593 | 0.81476 | 0.78346 | 0.93691 | 0.88401 | 0.8667798664 | 0.95522 | 0.927230835 | 0.324716 | 2.937958269 | 0 |
| 10 | 87933100 | A | G | E | G | PTEN | ENST00000371953 | 0.67239 | 0.86095 | 0.73162 | 0.96538 | 0.92763 | 0.8751140276 | 0.95382 | 0.9794363379 | 0.355773 | 4.198809594 | 0 |
| 10 | 87933102 | G | A | D | N | PTEN | ENST00000371953 | 0.56318 | 0.79743 | 0.89423 | 0.6987 | 0.70914 | 0.8651021413 | 0.90638 | 0.9171503186 | -0.0346822 | 2.447001419 | 0 |
| 10 | 87933102 | G | C | D | H | PTEN | ENST00000371953 | 0.55032 | 0.85648 | 0.91069 | 0.7467 | 0.80948 | 0.9287310408 | 0.95384 | 0.8687913418 | 0.187058 | 3.12247111 | 0 |
| 10 | 87933102 | G | T | D | Y | PTEN | ENST00000371953 | 0.88666 | 0.97466 | 0.8971 | 0.91855 | 0.90885 | 0.9776037811 | 0.95412 | 0.9964764714 | 0.286435 | 7.212919215 | 0 |
| 10 | 87933103 | A | G | D | G | PTEN | ENST00000371953 | 0.72917 | 0.88592 | 0.85393 | 0.80334 | 0.88069 | 0.9552357377 | 0.91742 | 0.988048315 | 0.215628 | 4.422022896 | 0 |
| 10 | 87933103 | A | T | D | V | PTEN | ENST00000371953 | 0.87749 | 0.98138 | 0.83629 | 0.94474 | 0.94091 | 0.9709851878 | 0.95006 | 0.9983971715 | 0.32849 | 7.541138643 | 0 |
| 10 | 87933105 | G | C | D | H | PTEN | ENST00000371953 | 0.67995 | 0.94843 | 0.94719 | 0.96531 | 0.84853 | 0.76680794 | 0.87257 | 0.9181579947 | 0.243028 | 4.208657849 | 0 |
| 10 | 87933105 | G | T | D | Y | PTEN | ENST00000371953 | 0.8366 | 0.95201 | 0.93595 | 0.97173 | 0.92438 | 0.8182106535 | 0.92601 | 0.9799664617 | 0.30239 | 6.551534476 | 0 |
| 10 | 87933106 | A | G | D | G | PTEN | ENST00000371953 | 0.56249 | 0.86473 | 0.89405 | 0.87028 | 0.80831 | 0.7656150793 | 0.87264 | 0.7886818647 | 0.185803 | 3.673159798 | 0 |
| 10 | 87933106 | A | T | D | V | PTEN | ENST00000371953 | 0.57568 | 0.88855 | 0.88229 | 0.91051 | 0.87781 | 0.8343878732 | 0.94166 | 0.925365746 | 0.269437 | 3.431597995 | 0 |
| 10 | 87933108 | A | C | N | H | PTEN | ENST00000371953 | 0.68107 | 0.80715 | 0.82843 | 0.81399 | 0.7562 | 0.7550910619 | 0.89461 | 0.4949758351 | -0.0398003 | 3.881743722 | 0 |
| 10 | 87933108 | A | G | N | D | PTEN | ENST00000371953 | 0.47162 | 0.82827 | 0.86182 | 0.79054 | 0.54201 | 0.7803361085 | 0.80187 | 0.888898015 | 0.0291329 | 2.527841858 | 0 |
| 10 | 87933108 | A | T | N | Y | PTEN | ENST00000371953 | 0.86789 | 0.88228 | 0.8563 | 0.88363 | 0.94129 | 0.847320899 | 0.96178 | 0.9665042162 | 0.273485 | 5.306537251 | 0 |
| 10 | 87933109 | A | C | N | T | PTEN | ENST00000371953 | 0.3933 | 0.84291 | 0.79998 | 0.83609 | 0.64977 | 0.7889562941 | 0.84098 | 0.8461395502 | 0.0769146 | 4.293244372 | 0 |
| 10 | 87933109 | A | G | N | S | PTEN | ENST00000371953 | 0.3303 | 0.7218 | 0.75425 | 0.75 | 0.59516 | 0.644137493 | 0.83123 | 0.2598228148 | -0.134763 | 3.517545428 | 0 |
| 10 | 87933110 | T | A | N | K | PTEN | ENST00000371953 | 0.35388 | 0.83394 | 0.89214 | 0.77319 | 0.63736 | 0.7784442301 | 0.80287 | 0.7630900741 | 0.0381163 | 2.47563991 | 0 |
| 10 | 87933111 | C | A | H | N | PTEN | ENST00000371953 | 0.36347 | 0.81899 | 0.94101 | 0.76804 | 0.71627 | 0.8766109253 | 0.83278 | 0.3551863447 | 0.0835017 | 1.691898739 | 0 |
| 10 | 87933111 | C | G | H | D | PTEN | ENST00000371953 | 0.66153 | 0.91466 | 0.93517 | 0.87382 | 0.90755 | 0.9798031805 | 0.97332 | 0.8813062906 | 0.394899 | 7.08809635 | 1 |
| 10 | 87933111 | C | T | H | Y | PTEN | ENST00000371953 | 0.78764 | 0.90175 | 0.9429 | 0.84373 | 0.93824 | 0.9577759289 | 0.96929 | 0.9612150192 | 0.398407 | 4.828996967 | 0 |
| 10 | 87933112 | A | G | H | R | PTEN | ENST00000371953 | 0.80937 | 0.86936 | 0.94449 | 0.79555 | 0.94015 | 0.9521109522 | 0.97698 | 0.8698869944 | 0.347008 | 4.575590993 | 0 |
| 10 | 87933113 | T | A | H | Q | PTEN | ENST00000371953 | 0.7141 | 0.94658 | 0.92666 | 0.89447 | 0.91569 | 0.9318967124 | 0.98255 | 0.9130898154 | 0.352176 | 5.760822339 | 0 |
| 10 | 87933114 | G | A | V | I | PTEN | ENST00000371953 | 0.47501 | 0.93854 | 0.78972 | 0.96591 | 0.90711 | 0.8875689781 | 0.96421 | 0.8095270395 | 0.381964 | 3.42581284 | 0 |
| 10 | 87933114 | G | C | V | L | PTEN | ENST00000371953 | 0.78915 | 0.96653 | 0.923 | 0.97664 | 0.98109 | 0.9747241338 | 0.96891 | 0.9948160052 | 0.428526 | 5.930040819 | 0 |
| 10 | 87933115 | T | A | V | D | PTEN | ENST00000371953 | 0.92143 | 0.99163 | 0.93049 | 0.99282 | 0.99843 | 0.9996281821 | 0.99195 | 0.9984853864 | 0.585245 | 7.832713917 | 1 |
| 10 | 87933115 | T | C | V | A | PTEN | ENST00000371953 | 0.88529 | 0.97054 | 0.92898 | 0.97991 | 0.98657 | 0.9753999886 | 0.95082 | 0.9881455778 | 0.416953 | 5.407628583 | 0 |
| 10 | 87933115 | T | G | V | G | PTEN | ENST00000371953 | 0.91347 | 0.98329 | 0.83755 | 0.98678 | 0.99932 | 0.9981613052 | 0.9892 | 0.998758316 | 0.584892 | 7.69859402 | 1 |
| 10 | 87933117 | G | C | A | P | PTEN | ENST00000371953 | 0.85382 | 0.98273 | 0.94288 | 0.96358 | 0.97475 | 0.9946735291 | 0.934 | 0.9990609288 | 0.461015 | 8.106830144 | 1 |
| 10 | 87933117 | G | T | A | S | PTEN | ENST00000371953 | 0.80195 | 0.94282 | 0.86914 | 0.97179 | 0.9328 | 0.9139948358 | 0.90903 | 0.9934252501 | 0.269857 | 7.225877703 | 0 |
| 10 | 87933118 | C | G | A | G | PTEN | ENST00000371953 | 0.81331 | 0.89993 | 0.89191 | 0.96547 | 0.95243 | 0.9320039337 | 0.9002 | 0.9951230884 | 0.349925 | 5.268648776 | 0 |
| 10 | 87933118 | C | T | A | V | PTEN | ENST00000371953 | 0.67987 | 0.8416 | 0.91832 | 0.84736 | 0.83228 | 0.8940524086 | 0.8209 | 0.8823848963 | 0.1737 | 4.024979718 | 0 |
| 10 | 87933120 | G | A | A | T | PTEN | ENST00000371953 | 0.83513 | 0.96618 | 0.79343 | 0.96277 | 0.94394 | 0.9244000504 | 0.89855 | 0.9899086952 | 0.29768 | 6.94900646 | 0 |
| 10 | 87933121 | C | G | A | G | PTEN | ENST00000371953 | 0.82186 | 0.95367 | 0.77755 | 0.95119 | 0.93004 | 0.9494492867 | 0.88125 | 0.9896572232 | 0.260498 | 4.889867488 | 0 |
| 10 | 87933123 | A | C | I | L | PTEN | ENST00000371953 | 0.73229 | 0.91566 | 0.91934 | 0.82736 | 0.87442 | 0.9399691551 | 0.90689 | 0.9165934324 | 0.209741 | 5.263101096 | 0 |
| 10 | 87933123 | A | G | I | V | PTEN | ENST00000371953 | 0.34565 | 0.78647 | 0.84454 | 0.58299 | 0.72626 | 0.7888021412 | 0.85014 | 0.8405969739 | 0.105172 | 3.38182415 | 0 |
| 10 | 87933123 | A | T | I | F | PTEN | ENST00000371953 | 0.78399 | 0.99048 | 0.92596 | 0.94671 | 0.96846 | 0.9846914709 | 0.95651 | 0.9949624538 | 0.423677 | 6.795769531 | 1 |
| 10 | 87933124 | T | A | I | N | PTEN | ENST00000371953 | 0.9112 | 0.99381 | 0.97241 | 0.97381 | 0.98477 | 0.9972098094 | 0.93167 | 0.9976327676 | 0.414283 | 8.031859011 | 1 |
| 10 | 87933124 | T | G | I | S | PTEN | ENST00000371953 | 0.9153 | 0.98685 | 0.93771 | 0.97001 | 0.98693 | 0.9979302686 | 0.97386 | 0.9980862141 | 0.471392 | 8.031859011 | 1 |
| 10 | 87933126 | C | A | H | N | PTEN | ENST00000371953 | 0.67819 | 0.97427 | 0.98237 | 0.98847 | 0.96044 | 0.9950306947 | 0.96467 | 0.9990558028 | 0.417264 | 4.38790663 | 0 |
| 10 | 87933126 | C | G | H | D | PTEN | ENST00000371953 | 0.7149 | 0.9858 | 0.97869 | 0.99283 | 0.9986 | 0.9991526132 | 0.99091 | 0.9997205138 | 0.58409 | 7.089636514 | 1 |
| 10 | 87933126 | C | T | H | Y | PTEN | ENST00000371953 | 0.67032 | 0.97769 | 0.98307 | 0.9918 | 0.99652 | 0.9941753757 | 0.99104 | 0.9993009567 | 0.582387 | 6.380638481 | 1 |
| 10 | 87933127 | A | C | H | P | PTEN | ENST00000371953 | 0.88916 | 0.99056 | 0.96334 | 0.98971 | 0.99722 | 0.9989960365 | 0.98949 | 0.9999527931 | 0.593698 | 7.089636514 | 1 |
| 10 | 87933127 | A | G | H | R | PTEN | ENST00000371953 | 0.84935 | 0.98043 | 0.98283 | 0.98681 | 0.99932 | 0.998086859 | 0.99568 | 0.9996040463 | 0.593413 | 6.964699545 | 1 |
| 10 | 87933127 | A | T | H | L | PTEN | ENST00000371953 | 0.89137 | 0.9811 | 0.97186 | 0.98994 | 0.99006 | 0.9966161952 | 0.99221 | 0.9998924732 | 0.593413 | 5.901741474 | 1 |
| 10 | 87933129 | T | C | C | R | PTEN | ENST00000371953 | 0.86075 | 0.99403 | 0.99204 | 0.98846 | 0.99552 | 0.99780845 | 0.99545 | 0.999864459 | 0.58708 | 7.335935889 | 1 |

| | | | | | | | | | | | | | | | | | | |
|---|---|---|---|---|---|---|---|---|---|---|---|---|---|---|---|---|---|---|
| 10 | 87933129 | T | G | C | G | PTEN | ENST00000371953 | 0.8456 | 0.9811 | 0.95125 | 0.9883 | 0.99471 | 0.9975716846 | 0.99066 | 0.9995476604 | 0.586803 | 6.451624912 | 1 |
| 10 | 87933130 | G | C | C | S | PTEN | ENST00000371953 | 0.93238 | 0.98069 | 0.96715 | 0.99264 | 0.99386 | 0.9960321381 | 0.98857 | 0.9998905659 | 0.594156 | 5.373138085 | 1 |
| 10 | 87933130 | G | T | C | F | PTEN | ENST00000371953 | 0.94423 | 0.98981 | 0.97579 | 0.99054 | 0.98869 | 0.9965060269 | 0.98428 | 0.9999815226 | 0.594437 | 7.335935889 | 1 |
| 10 | 87933132 | A | C | K | Q | PTEN | ENST00000371953 | 0.79911 | 0.97311 | 0.91496 | 0.98566 | 0.97104 | 0.9844840791 | 0.98096 | 0.9951704604 | 0.415531 | 5.845449202 | 0 |
| 10 | 87933132 | A | G | K | E | PTEN | ENST00000371953 | 0.82428 | 0.98652 | 0.93839 | 0.99008 | 0.99105 | 0.9928418301 | 0.98234 | 0.9990925789 | 0.537502 | 7.676103474 | 1 |
| 10 | 87933133 | A | G | K | R | PTEN | ENST00000371953 | 0.80321 | 0.95441 | 0.89551 | 0.98651 | 0.96371 | 0.9526938801 | 0.96978 | 0.9907628894 | 0.376273 | 5.646946558 | 0 |
| 10 | 87933133 | A | T | K | I | PTEN | ENST00000371953 | 0.85301 | 0.99403 | 0.92001 | 0.99176 | 0.97661 | 0.9896723438 | 0.96344 | 0.9997195601 | 0.444667 | 6.045899857 | 0 |
| 10 | 87933135 | G | C | A | P | PTEN | ENST00000371953 | 0.85382 | 0.98329 | 0.96232 | 0.99651 | 0.99628 | 0.9921756838 | 0.995 | 0.9997521043 | 0.593765 | 8.180054579 | 1 |
| 10 | 87933135 | G | T | A | S | PTEN | ENST00000371953 | 0.80519 | 0.96146 | 0.90692 | 0.99557 | 0.96846 | 0.8982268774 | 0.97396 | 0.9943780303 | 0.424339 | 6.551092837 | 0 |
| 10 | 87933136 | C | A | A | D | PTEN | ENST00000371953 | 0.83882 | 0.98885 | 0.97605 | 0.99288 | 0.99386 | 0.991097398 | 0.98991 | 0.998641789 | 0.567064 | 5.235364928 | 0 |
| 10 | 87933136 | C | G | A | G | PTEN | ENST00000371953 | 0.83058 | 0.96415 | 0.9248 | 0.99183 | 0.96444 | 0.8945093453 | 0.96815 | 0.9956210256 | 0.40877 | 5.603836057 | 0 |
| 10 | 87933138 | G | A | G | R | PTEN | ENST00000371953 | 0.88423 | 0.97893 | 0.99567 | 0.99605 | 0.99414 | 0.9922458197 | 0.99569 | 0.9999057055 | 0.594481 | 7.156893804 | 1 |
| 10 | 87933139 | G | A | G | E | PTEN | ENST00000371953 | 0.91746 | 0.98138 | 0.9947 | 0.99542 | 0.99202 | 0.9962493624 | 0.9884 | 0.9994146824 | 0.594481 | 6.658451022 | 1 |
| 10 | 87933139 | G | C | G | A | PTEN | ENST00000371953 | 0.89807 | 0.97295 | 0.98655 | 0.99603 | 0.99039 | 0.9872326949 | 0.99175 | 0.9993995428 | 0.594079 | 7.336292637 | 1 |
| 10 | 87933141 | A | C | K | Q | PTEN | ENST00000371953 | 0.79334 | 0.97295 | 0.96429 | 0.95223 | 0.96956 | 0.9927437803 | 0.96986 | 0.9984189272 | 0.341652 | 5.616716044 | 0 |
| 10 | 87933141 | A | G | K | E | PTEN | ENST00000371953 | 0.8185 | 0.98652 | 0.9757 | 0.9689 | 0.99105 | 0.9967173847 | 0.9921 | 0.9990925789 | 0.477018 | 7.219369841 | 1 |
| 10 | 87933142 | A | T | K | M | PTEN | ENST00000371953 | 0.93051 | 0.98432 | 0.95469 | 0.96324 | 0.98367 | 0.9918177609 | 0.94644 | 0.9992762208 | 0.418019 | 5.587338643 | 0 |
| 10 | 87933145 | G | A | G | E | PTEN | ENST00000371953 | 0.92182 | 0.98311 | 0.99847 | 0.98259 | 0.98728 | 0.9981102698 | 0.97734 | 0.9997332692 | 0.481762 | 5.697940776 | 1 |
| 10 | 87933145 | G | C | G | A | PTEN | ENST00000371953 | 0.90063 | 0.97295 | 0.99575 | 0.9598 | 0.96919 | 0.9853419181 | 0.96282 | 0.9986763597 | 0.449144 | 5.347938559 | 0 |
| 10 | 87933145 | G | T | G | V | PTEN | ENST00000371953 | 0.91938 | 0.98161 | 0.9956 | 0.98046 | 0.98549 | 0.9934011406 | 0.97981 | 0.9997332692 | 0.482229 | 6.423529342 | 1 |
| 10 | 87933147 | C | G | R | G | PTEN | ENST00000371953 | 0.87716 | 0.98291 | 0.97263 | 0.99232 | 0.99443 | 0.9976320982 | 0.99566 | 0.999138236 | 0.570271 | 7.746178054 | 1 |
| 10 | 87933148 | G | C | R | P | PTEN | ENST00000371953 | 0.94605 | 0.98772 | 0.98456 | 0.993 | 0.99919 | 0.9993776181 | 0.99376 | 0.9992620349 | 0.594398 | 6.112516852 | 1 |
| 10 | 87933148 | G | T | R | L | PTEN | ENST00000371953 | 0.94174 | 0.9813 | 0.98112 | 0.99152 | 0.99953 | 0.9962879626 | 0.99711 | 0.9995791316 | 0.594396 | 6.851616994 | 1 |
| 10 | 87933150 | A | C | T | P | PTEN | ENST00000371953 | 0.78391 | 0.98348 | 0.93524 | 0.99524 | 0.99953 | 0.9974695449 | 0.99299 | 0.9995458722 | 0.593391 | 7.252554301 | 1 |
| 10 | 87933151 | C | A | T | N | PTEN | ENST00000371953 | 0.73893 | 0.97686 | 0.97274 | 0.99045 | 0.97252 | 0.9959500902 | 0.96851 | 0.9993413091 | 0.335033 | 8.22698104 | 1 |
| 10 | 87933153 | G | A | G | S | PTEN | ENST00000371953 | 0.84726 | 0.96803 | 0.97982 | 0.99616 | 0.99265 | 0.9890120641 | 0.98929 | 0.9991213679 | 0.587129 | 5.288096217 | 1 |
| 10 | 87933153 | G | C | G | R | PTEN | ENST00000371953 | 0.87603 | 0.97924 | 0.99524 | 0.99651 | 0.99603 | 0.9979466148 | 0.99474 | 0.9998931885 | 0.594467 | 7.235607737 | 1 |
| 10 | 87933153 | G | T | G | C | PTEN | ENST00000371953 | 0.89862 | 0.9727 | 0.9909 | 0.9969 | 0.99552 | 0.9936694279 | 0.97499 | 0.9998931885 | 0.539819 | 6.899168886 | 1 |
| 10 | 87933154 | G | A | G | D | PTEN | ENST00000371953 | 0.89884 | 0.98161 | 0.99606 | 0.99511 | 0.99982 | 0.9988024708 | 0.99297 | 0.999818027 | 0.594322 | 7.319539768 | 1 |
| 10 | 87933154 | G | C | G | A | PTEN | ENST00000371953 | 0.87996 | 0.97332 | 0.98072 | 0.99229 | 0.99525 | 0.9890836872 | 0.99185 | 0.9989527464 | 0.588762 | 4.788293748 | 1 |
| 10 | 87933154 | G | T | G | V | PTEN | ENST00000371953 | 0.90318 | 0.98161 | 0.98403 | 0.99219 | 0.9997 | 0.9953334096 | 0.99146 | 0.9997836947 | 0.594485 | 7.337458513 | 1 |
| 10 | 87933156 | G | C | V | L | PTEN | ENST00000371953 | 0.79911 | 0.86153 | 0.93709 | 0.96702 | 0.95572 | 0.9718062995 | 0.94897 | 0.9490692019 | 0.381292 | 5.201892279 | 0 |
| 10 | 87933156 | G | T | V | L | PTEN | ENST00000371953 | 0.79263 | 0.86153 | 0.93709 | 0.96727 | 0.95572 | 0.9718062995 | 0.94887 | 0.9535477161 | 0.38129 | 5.201892279 | 0 |
| 10 | 87933157 | T | C | V | A | PTEN | ENST00000371953 | 0.88795 | 0.95965 | 0.94614 | 0.98382 | 0.98693 | 0.9662413281 | 0.95181 | 0.985406518 | 0.498239 | 6.751406935 | 0 |
| 10 | 87933159 | A | T | M | L | PTEN | ENST00000371953 | 0.72228 | 0.92225 | 0.97951 | 0.80208 | 0.88684 | 0.9726078426 | 0.90978 | 0.9857305288 | 0.244158 | 5.012120984 | 0 |
| 10 | 87933160 | T | A | M | K | PTEN | ENST00000371953 | 0.79595 | 0.98981 | 0.99235 | 0.96693 | 0.99628 | 0.9991504236 | 0.98218 | 0.9992623925 | 0.471483 | 8.605814437 | 1 |
| 10 | 87933160 | T | G | M | R | PTEN | ENST00000371953 | 0.81371 | 0.98939 | 0.98907 | 0.96568 | 0.99722 | 0.9993498006 | 0.98193 | 0.9990996122 | 0.472045 | 8.417679607 | 1 |
| 10 | 87933162 | A | C | I | L | PTEN | ENST00000371953 | 0.76339 | 0.95467 | 0.97404 | 0.9692 | 0.96008 | 0.9552297886 | 0.93293 | 0.9629337788 | 0.359343 | 5.297283891 | 0 |
| 10 | 87933162 | A | G | I | V | PTEN | ENST00000371953 | 0.53879 | 0.89442 | 0.93304 | 0.94419 | 0.94318 | 0.8313154453 | 0.9665 | 0.9042348862 | 0.432191 | 3.421893892 | 0 |
| 10 | 87933162 | A | T | I | L | PTEN | ENST00000371953 | 0.75663 | 0.95467 | 0.97404 | 0.96848 | 0.96008 | 0.9552297886 | 0.9324 | 0.9681044817 | 0.35934 | 5.297283891 | 0 |
| 10 | 87933163 | T | A | I | K | PTEN | ENST00000371953 | 0.88119 | 0.99399 | 0.99128 | 0.99177 | 0.99976 | 0.9969963584 | 0.99532 | 0.9995521903 | 0.586135 | 8.208296319 | 1 |
| 10 | 87933163 | T | C | I | T | PTEN | ENST00000371953 | 0.87483 | 0.97662 | 0.99531 | 0.98632 | 0.99906 | 0.9714269075 | 0.99097 | 0.9961392931 | 0.554562 | 5.815928567 | 0 |
| 10 | 87933163 | T | G | I | R | PTEN | ENST00000371953 | 0.89509 | 0.99399 | 0.98944 | 0.99286 | 0.99993 | 0.9958606336 | 0.99452 | 0.9989449382 | 0.586188 | 8.191416518 | 1 |
| 10 | 87933164 | A | G | I | M | PTEN | ENST00000371953 | 0.76171 | 0.96763 | 0.95828 | 0.9906 | 0.97364 | 0.9584808957 | 0.97532 | 0.9904604554 | 0.486136 | 7.272068161 | 0 |
| 10 | 87933165 | T | C | C | R | PTEN | ENST00000371953 | 0.85204 | 0.98196 | 0.99219 | 0.99272 | 0.99982 | 0.9956160187 | 0.99218 | 0.9998015761 | 0.581897 | 8.887735438 | 1 |
| 10 | 87933165 | T | G | C | G | PTEN | ENST00000371953 | 0.84111 | 0.94813 | 0.94781 | 0.97958 | 0.98294 | 0.9797351516 | 0.98447 | 0.9989350438 | 0.564264 | 6.684474515 | 0 |
| 10 | 87933166 | G | A | C | Y | PTEN | ENST00000371953 | 0.92983 | 0.97924 | 0.99414 | 0.99416 | 0.99744 | 0.9905742915 | 0.97299 | 0.9995206594 | 0.575805 | 8.887735438 | 1 |
| 10 | 87933166 | G | C | C | S | PTEN | ENST00000371953 | 0.92202 | 0.90611 | 0.96317 | 0.97019 | 0.98799 | 0.9704678513 | 0.95445 | 0.993507266 | 0.519077 | 5.110558139 | 0 |
| 10 | 87933166 | G | T | C | F | PTEN | ENST00000371953 | 0.93973 | 0.98712 | 0.97271 | 0.99267 | 0.99552 | 0.9874916041 | 0.98295 | 0.9997451901 | 0.589254 | 8.697493224 | 1 |
| 10 | 87933167 | T | G | C | W | PTEN | ENST00000371953 | 0.89486 | 0.98481 | 0.98636 | 0.99369 | 0.99202 | 0.9920484257 | 0.99525 | 0.9998773336 | 0.559598 | 8.05528912 | 1 |
| 10 | 87933168 | G | A | A | T | PTEN | ENST00000371953 | 0.80764 | 0.96288 | 0.96462 | 0.97274 | 0.97923 | 0.9006561981 | 0.97941 | 0.9802144766 | 0.492181 | 4.716555018 | 0 |
| 10 | 87933168 | G | C | A | P | PTEN | ENST00000371953 | 0.84035 | 0.98311 | 0.98351 | 0.98913 | 0.99296 | 0.9931689748 | 0.98709 | 0.9941473007 | 0.559154 | 7.56448991 | 1 |
| 10 | 87933168 | G | T | A | S | PTEN | ENST00000371953 | 0.46463 | 0.88902 | 0.94238 | 0.96696 | 0.88448 | 0.8508821435 | 0.94181 | 0.8568759561 | 0.332656 | 2.956080454 | 0 |
| 10 | 87933169 | C | A | A | E | PTEN | ENST00000371953 | 0.78043 | 0.97698 | 0.98353 | 0.99108 | 0.99386 | 0.9921680684 | 0.99099 | 0.996154964 | 0.55148 | 7.56448991 | 0 |
| 10 | 87933169 | C | G | A | G | PTEN | ENST00000371953 | 0.63842 | 0.95708 | 0.95632 | 0.98568 | 0.967 | 0.8903643741 | 0.93646 | 0.9890683293 | 0.4053 | 6.014542129 | 0 |
| 10 | 87933169 | C | T | A | V | PTEN | ENST00000371953 | 0.78415 | 0.96033 | 0.97276 | 0.98901 | 0.9822 | 0.9379707439 | 0.97466 | 0.9838985801 | 0.502382 | 4.569817645 | 0 |
| 10 | 87933171 | T | A | Y | N | PTEN | ENST00000371953 | 0.81072 | 0.99186 | 0.98789 | 0.98717 | 0.97997 | 0.9968188773 | 0.95184 | 0.9999070168 | 0.403723 | 8.161157873 | 1 |
| 10 | 87933172 | A | T | Y | F | PTEN | ENST00000371953 | 0.85632 | 0.93777 | 0.95862 | 0.92655 | 0.94913 | 0.9524920518 | 0.89404 | 0.969741995 | 0.226128 | 4.40282096 | 0 |
| 10 | 87933174 | T | A | L | I | PTEN | ENST00000371953 | 0.82139 | 0.97295 | 0.93144 | 0.92133 | 0.91357 | 0.9081579966 | 0.87718 | 0.9410497546 | 0.273927 | 5.157649245 | 0 |
| 10 | 87933174 | T | G | L | V | PTEN | ENST00000371953 | 0.81975 | 0.96909 | 0.90447 | 0.94957 | 0.9517 | 0.9633052884 | 0.97417 | 0.9783664942 | 0.46289 | 5.988716228 | 0 |
| 10 | 87933175 | T | C | L | S | PTEN | ENST00000371953 | 0.91691 | 0.98626 | 0.94684 | 0.98373 | 0.99552 | 0.9939509492 | 0.98774 | 0.9997072816 | 0.506568 | 8.091299748 | 0 |
| 10 | 87933176 | A | T | L | F | PTEN | ENST00000371953 | 0.72909 | 0.97392 | 0.93181 | 0.9617 | 0.95316 | 0.9689378343 | 0.97657 | 0.9922072291 | 0.447267 | 5.23503485 | 0 |
| 10 | 87933177 | T | A | L | I | PTEN | ENST00000371953 | 0.57554 | 0.93076 | 0.96131 | 0.78797 | 0.81588 | 0.7947032773 | 0.88502 | 0.6686311364 | 0.129901 | 4.010445165 | 0 |
| 10 | 87933177 | T | G | L | V | PTEN | ENST00000371953 | 0.58567 | 0.92161 | 0.93425 | 0.86303 | 0.90755 | 0.8802389456 | 0.97844 | 0.8081582189 | 0.317171 | 4.667589942 | 0 |
| 10 | 87933178 | T | C | L | S | PTEN | ENST00000371953 | 0.91387 | 0.98591 | 0.96789 | 0.96009 | 0.97997 | 0.9743996607 | 0.98367 | 0.9976415634 | 0.461907 | 7.106057853 | 1 |
| 10 | 87933179 | A | T | L | F | PTEN | ENST00000371953 | 0.70423 | 0.97362 | 0.95874 | 0.9437 | 0.92233 | 0.9214481879 | 0.97895 | 0.9813981652 | 0.396613 | 6.777358627 | 1 |
| 10 | 87933180 | C | T | H | Y | PTEN | ENST00000371953 | 0.52619 | 0.85487 | 0.97567 | 0.85599 | 0.8887 | 0.9298244058 | 0.90026 | 0.9748650789 | 0.249435 | 3.788043228 | 0 |
| 10 | 87933181 | A | C | H | P | PTEN | ENST00000371953 | 0.87194 | 0.98959 | 0.94266 | 0.96444 | 0.9822 | 0.9849405062 | 0.9562 | 0.9997550845 | 0.449796 | 8.257581067 | 1 |
| 10 | 87933181 | A | T | H | L | PTEN | ENST00000371953 | 0.8784 | 0.95057 | 0.96213 | 0.92203 | 0.93397 | 0.9558419386 | 0.95245 | 0.9990029931 | 0.403152 | 4.168967297 | 0 |
| 10 | 87933182 | T | A | H | Q | PTEN | ENST00000371953 | 0.76649 | 0.96564 | 0.96503 | 0.88127 | 0.87636 | 0.9042315304 | 0.94081 | 0.9973515272 | 0.348484 | 6.667450809 | 0 |
| 10 | 87933183 | C | G | R | G | PTEN | ENST00000371953 | 0.68859 | 0.88398 | 0.98707 | 0.83933 | 0.90711 | 0.9374392783 | 0.98321 | 0.9344701767 | 0.368302 | 5.552702886 | 0 |
| 10 | 87933183 | C | T | R | W | PTEN | ENST00000371953 | 0.91674 | 0.96653 | 0.99671 | 0.88411 | 0.90052 | 0.9309766007 | 0.94517 | 0.9855616689 | 0.38862 | 7.929752294 | 0 |
| 10 | 87933184 | G | A | R | Q | PTEN | ENST00000371953 | 0.77569 | 0.85979 | 0.97678 | 0.8056 | 0.87877 | 0.8681851041 | 0.93009 | 0.750588119 | 0.216342 | 4.164100796 | 0 |
| 10 | 87933184 | G | C | R | P | PTEN | ENST00000371953 | 0.8312 | 0.9811 | 0.99269 | 0.91093 | 0.96407 | 0.9762544951 | 0.97261 | 0.9713009 | 0.394189 | 6.813617666 | 1 |
| 10 | 87933184 | G | T | R | L | PTEN | ENST00000371953 | 0.79049 | 0.86384 | 0.9918 | 0.82678 | 0.92561 | 0.9356152635 | 0.97956 | 0.9161081314 | 0.383602 | 5.004838317 | 0 |
| 10 | 87933186 | G | A | G | S | PTEN | ENST00000371953 | 0.72764 | 0.81235 | 0.96559 | 0.842 | 0.88117 | 0.8578050905 | 0.9001 | 0.9123991728 | 0.265257 | 4.055779819 | 0 |
| 10 | 87933186 | G | C | G | R | PTEN | ENST00000371953 | 0.56221 | 0.84841 | 0.9899 | 0.77834 | 0.80595 | 0.8674999263 | 0.95499 | 0.8109999299 | 0.336419 | 1.32468623 | 0 |
| 10 | 87933186 | G | T | G | C | PTEN | ENST00000371953 | 0.93943 | 0.96469 | 0.98101 | 0.88457 | 0.96883 | 0.9476620405 | 0.95205 | 0.9934667945 | 0.404397 | 4.847462029 | 0 |
| 10 | 87933187 | G | A | G | D | PTEN | ENST00000371953 | 0.70823 | 0.87764 | 0.99107 | 0.85871 | 0.94053 | 0.8909965472 | 0.94454 | 0.9200455546 | 0.407932 | 4.306696768 | 0 |
| 10 | 87933189 | A | C | K | Q | PTEN | ENST00000371953 | 0.53026 | 0.81408 | 0.89024 | 0.69702 | 0.72626 | 0.8437853236 | 0.902 | 0.9269853234 | 0.101824 | 3.675953648 | 0 |
| 10 | 87933189 | A | G | K | E | PTEN | ENST00000371953 | 0.4698 | 0.88308 | 0.91839 | 0.77892 | 0.89829 | 0.9115090815 | 0.96657 | 0.9193065763 | 0.291143 | 4.366363244 | 0 |
| 10 | 87933190 | A | C | K | T | PTEN | ENST00000371953 | 0.64594 | 0.85125 | 0.92602 | 0.72733 | 0.88916 | 0.9011803099 | 0.91331 | 0.9039580822 | 0.206462 | 4.447378546 | 0 |
| 10 | 87933190 | A | G | K | R | PTEN | ENST00000371953 | 0.46736 | 0.7299 | 0.86053 | 0.54511 | 0.47888 | 0.7279095875 | 0.81057 | 0.7549402714 | -0.00619629 | 2.918054567 | 0 |
| 10 | 87933190 | A | T | K | I | PTEN | ENST00000371953 | 0.72676 | 0.928 | 0.8949 | 0.83827 | 0.86503 | 0.8906072952 | 0.91833 | 0.8541042805 | 0.199086 | 2.604225221 | 0 |
| 10 | 87933192 | T | A | F | I | PTEN | ENST00000371953 | 0.81324 | 0.9695 | 0.99301 | 0.93548 | 0.94913 | 0.9783245019 | 0.8905 | 0.9932442307 | 0.265951 | 4.44724883 | 0 |
| 10 | 87933192 | T | G | F | V | PTEN | ENST00000371953 | 0.8181 | 0.95615 | 0.99186 | 0.93732 | 0.95935 | 0.9775039961 | 0.93926 | 0.9903711677 | 0.390499 | 5.061047402 | 0 |
| 10 | 87933193 | T | A | F | Y | PTEN | ENST00000371953 | 0.77972 | 0.86867 | 0.99518 | 0.87856 | 0.88448 | 0.9203639806 | 0.89176 | 0.8968297243 | 0.224044 | 3.922318002 | 0 |
| 10 | 87933193 | T | C | F | S | PTEN | ENST00000371953 | 0.94238 | 0.9799 | 0.99406 | 0.9327 | 0.95863 | 0.9832742662 | 0.97441 | 0.9954763055 | 0.404071 | 3.728118786 | 0 |
| 10 | 87933194 | T | G | F | L | PTEN | ENST00000371953 | 0.7015 | 0.92762 | 0.99642 | 0.89013 | 0.88307 | 0.9477899046 | 0.85857 | 0.9899922013 | 0.228505 | 4.760070769 | 0 |
| 10 | 87933195 | T | A | L | I | PTEN | ENST00000371953 | 0.36086 | 0.83244 | 0.90269 | 0.79036 | 0.67108 | 0.3749133974 | 0.75477 | 0.135319236 | 0.0921391 | 3.287461957 | 0 |
| 10 | 87933195 | T | G | L | V | PTEN | ENST00000371953 | 0.34642 | 0.8023 | 0.85886 | 0.77002 | 0.672 | 0.5447053912 | 0.78371 | 0.1040740972 | 0.0848453 | 3.284697988 | 0 |
| 10 | 87933196 | T | C | L | S | PTEN | ENST00000371953 | 0.41281 | 0.87982 | 0.9132 | 0.79373 | 0.72701 | 0.5494467583 | 0.85185 | 0.1917297843 | 0.150574 | 0.7800675585 | 0 |
| 10 | 87933197 | A | T | L | F | PTEN | ENST00000371953 | 0.49971 | 0.8184 | 0.89797 | 0.84512 | 0.77591 | 0.4716108472 | 0.85959 | 0.6399381161 | 0.179096 | 3.340898629 | 0 |
| 10 | 87933198 | A | C | K | Q | PTEN | ENST00000371953 | 0.4029 | 0.82388 | 0.89401 | 0.89294 | 0.80417 | 0.6656867658 | 0.89124 | 0.6379618049 | 0.184085 | 3.019711758 | 0 |
| 10 | 87933198 | A | G | K | E | PTEN | ENST00000371953 | 0.36358 | 0.87875 | 0.92101 | 0.82695 | 0.70671 | 0.7516673762 | 0.79473 | 0.6317436099 | 0.11153 | 0.5772243337 | 0 |
| 10 | 87933199 | A | C | K | T | PTEN | ENST00000371953 | 0.39969 | 0.84062 | 0.93002 | 0.72761 | 0.72398 | 0.7049117121 | 0.79633 | 0.8021227717 | 0.159163 | 0.5029412219 | 0 |
| 10 | 87933199 | A | G | K | R | PTEN | ENST00000371953 | 0.48691 | 0.73196 | 0.86304 | 0.81795 | 0.71548 | 0.4998144766 | 0.86698 | 0.4613159001 | -0.00162447 | 3.48101862 | 0 |
| 10 | 87933199 | A | T | K | M | PTEN | ENST00000371953 | 0.85705 | 0.96749 | 0.8614 | 0.95296 | 0.88259 | 0.7365734273 | 0.87978 | 0.9217951894 | 0.213856 | 4.157570155 | 0 |
| 10 | 87933200 | G | T | K | N | PTEN | ENST00000371953 | 0.50544 | 0.83722 | 0.94072 | 0.80819 | 0.66639 | 0.7108137571 | 0.82534 | 0.639588058 | 0.0843887 | 2.079198725 | 0 |
| 10 | 87933201 | G | A | A | T | PTEN | ENST00000371953 | 0.74922 | 0.88681 | 0.9539 | 0.86688 | 0.90972 | 0.8890291622 | 0.89057 | 0.8996598125 | 0.252581 | 3.146962557 | 0 |
| 10 | 87933201 | G | C | A | P | PTEN | ENST00000371953 | 0.58084 | 0.88466 | 0.97297 | 0.83711 | 0.83339 | 0.9834960624 | 0.85116 | 0.9174442887 | 0.133951 | 2.909618132 | 0 |

| | | | | | | | | | | | | | | | | | |
|---|---|---|---|---|---|---|---|---|---|---|---|---|---|---|---|---|---|
| 10 | 87933201 | G | T | A | S | PTEN | ENST00000371953 | 0.69444 | 0.87797 | 0.9309 | 0.89712 | 0.86703 | 0.8157049036 | 0.87734 | 0.9298599362 | 0.188118 | 3.282818991 | 0 |
| 10 | 87933202 | C | A | A | E | PTEN | ENST00000371953 | 0.77085 | 0.97114 | 0.97655 | 0.94395 | 0.96262 | 0.9855057659 | 0.97254 | 0.9949194789 | 0.442252 | 5.521715365 | 1 |
| 10 | 87933202 | C | G | A | G | PTEN | ENST00000371953 | 0.76792 | 0.86544 | 0.94525 | 0.90229 | 0.90406 | 0.8432674087 | 0.86534 | 0.9744374156 | 0.196571 | 4.564544722 | 0 |
| 10 | 87933202 | C | T | A | V | PTEN | ENST00000371953 | 0.76737 | 0.89172 | 0.96126 | 0.89126 | 0.94949 | 0.9227998975 | 0.89884 | 0.9679275751 | 0.266808 | 3.650653966 | 0 |
| 10 | 87933204 | C | A | Q | K | PTEN | ENST00000371953 | 0.446 | 0.86874 | 0.96479 | 0.82438 | 0.8367 | 0.8668598761 | 0.9356 | 0.791685164 | 0.204727 | 3.156473765 | 0 |
| 10 | 87933204 | C | G | Q | E | PTEN | ENST00000371953 | 0.3057 | 0.82029 | 0.94064 | 0.75116 | 0.75074 | 0.7875276044 | 0.8816 | 0.6609999537 | 0.104643 | 2.819356488 | 0 |
| 10 | 87933205 | A | C | Q | P | PTEN | ENST00000371953 | 0.72236 | 0.87307 | 0.94586 | 0.86249 | 0.94015 | 0.9819136604 | 0.9803 | 0.9424378276 | 0.403238 | 5.80191147 | 0 |
| 10 | 87933205 | A | G | Q | R | PTEN | ENST00000371953 | 0.67835 | 0.82789 | 0.96156 | 0.79931 | 0.85012 | 0.8093392567 | 0.96546 | 0.6509478268 | 0.251676 | 4.297061683 | 0 |
| 10 | 87933205 | A | T | Q | L | PTEN | ENST00000371953 | 0.73269 | 0.82109 | 0.93757 | 0.85688 | 0.90229 | 0.8852513267 | 0.93082 | 0.9580360651 | 0.253611 | 4.484605892 | 0 |
| 10 | 87933206 | A | T | Q | H | PTEN | ENST00000371953 | 0.70069 | 0.81112 | 0.96106 | 0.85607 | 0.86553 | 0.7756302018 | 0.90546 | 0.8040681481 | 0.14589 | 4.749613922 | 0 |
| 10 | 87933207 | G | A | E | K | PTEN | ENST00000371953 | 0.64953 | 0.90331 | 0.9719 | 0.9764 | 0.90842 | 0.9597945835 | 0.99548 | 0.9648420811 | 0.528159 | 3.174247784 | 0 |
| 10 | 87933207 | G | C | E | Q | PTEN | ENST00000371953 | 0.78439 | 0.86294 | 0.92469 | 0.97146 | 0.92763 | 0.9145102581 | 0.97983 | 0.9150946736 | 0.387988 | 3.844796178 | 0 |
| 10 | 87933208 | A | C | E | A | PTEN | ENST00000371953 | 0.86403 | 0.87593 | 0.9284 | 0.95708 | 0.9608 | 0.9377051557 | 0.98841 | 0.9856535792 | 0.491472 | 3.998270738 | 0 |
| 10 | 87933208 | A | G | E | G | PTEN | ENST00000371953 | 0.90442 | 0.87427 | 0.90097 | 0.97047 | 0.9822 | 0.9457288076 | 0.98491 | 0.9890669584 | 0.551141 | 4.693014993 | 0 |
| 10 | 87933208 | A | T | E | V | PTEN | ENST00000371953 | 0.9009 | 0.96564 | 0.91841 | 0.98539 | 0.9796 | 0.949409089 | 0.99514 | 0.9980354905 | 0.569469 | 5.592751834 | 0 |
| 10 | 87933209 | G | T | E | D | PTEN | ENST00000371953 | 0.31836 | 0.77 | 0.92987 | 0.70401 | 0.83004 | 0.8873505557 | 0.97 | 0.3806678951 | 0.267394 | 1.805284886 | 0 |
| 10 | 87933210 | G | A | A | T | PTEN | ENST00000371953 | 0.81559 | 0.96308 | 0.91383 | 0.96404 | 0.96189 | 0.9507060451 | 0.95138 | 0.9988408685 | 0.434126 | 6.175855534 | 0 |
| 10 | 87933210 | G | T | A | S | PTEN | ENST00000371953 | 0.77742 | 0.9253 | 0.88973 | 0.92855 | 0.91945 | 0.8896125858 | 0.93301 | 0.985386014 | 0.310439 | 4.442595565 | 0 |
| 10 | 87933211 | C | A | A | D | PTEN | ENST00000371953 | 0.76467 | 0.98906 | 0.9631 | 0.98576 | 0.98693 | 0.9978657238 | 0.98354 | 0.9995978475 | 0.521188 | 7.620422344 | 1 |
| 10 | 87933211 | C | T | A | V | PTEN | ENST00000371953 | 0.75885 | 0.9634 | 0.92577 | 0.95928 | 0.96773 | 0.9693433362 | 0.95112 | 0.9962285757 | 0.455604 | 4.596542503 | 0 |
| 10 | 87933213 | C | G | L | V | PTEN | ENST00000371953 | 0.7465 | 0.94498 | 0.93193 | 0.92932 | 0.93515 | 0.9244668537 | 0.93882 | 0.9712187648 | 0.36865 | 4.839095893 | 0 |
| 10 | 87933214 | T | C | L | P | PTEN | ENST00000371953 | 0.89949 | 0.99134 | 0.99091 | 0.98233 | 0.99105 | 0.9979429242 | 0.98662 | 0.9997733235 | 0.478371 | 6.924895272 | 1 |
| 10 | 87933214 | T | G | L | R | PTEN | ENST00000371953 | 0.89634 | 0.98626 | 0.98742 | 0.97425 | 0.99234 | 0.9968781753 | 0.98917 | 0.9991809726 | 0.459993 | 3.261348848 | 1 |
| 10 | 87933216 | G | C | D | H | PTEN | ENST00000371953 | 0.50968 | 0.8895 | 0.97981 | 0.95672 | 0.86453 | 0.9170800433 | 0.99202 | 0.8990286589 | 0.509146 | 2.955985259 | 0 |
| 10 | 87933216 | G | T | D | Y | PTEN | ENST00000371953 | 0.83844 | 0.95942 | 0.975 | 0.96964 | 0.96262 | 0.9522472304 | 0.99417 | 0.9919906259 | 0.524083 | 4.707219968 | 0 |
| 10 | 87933217 | A | C | D | A | PTEN | ENST00000371953 | 0.74643 | 0.88909 | 0.94202 | 0.93816 | 0.93515 | 0.9280008317 | 0.98133 | 0.7436078191 | 0.489571 | 3.610237342 | 0 |
| 10 | 87933217 | A | T | D | V | PTEN | ENST00000371953 | 0.89656 | 0.96424 | 0.94175 | 0.98338 | 0.97141 | 0.9548134902 | 0.99068 | 0.9860317707 | 0.52628 | 5.019330404 | 0 |
| 10 | 87933220 | T | C | F | S | PTEN | ENST00000371953 | 0.92104 | 0.97054 | 0.9823 | 0.95469 | 0.98294 | 0.9758647335 | 0.97236 | 0.9857165813 | 0.424608 | 6.772929143 | 0 |
| 10 | 87933220 | T | G | F | C | PTEN | ENST00000371953 | 0.81818 | 0.95345 | 0.98787 | 0.95707 | 0.9729 | 0.9593836283 | 0.95689 | 0.9939629436 | 0.445626 | 5.713831655 | 0 |
| 10 | 87933221 | C | G | F | L | PTEN | ENST00000371953 | 0.52446 | 0.84354 | 0.98902 | 0.82092 | 0.8356 | 0.9140107551 | 0.95562 | 0.9104966521 | 0.221317 | 5.572634139 | 1 |
| 10 | 87933222 | T | A | Y | N | PTEN | ENST00000371953 | 0.81143 | 0.99195 | 0.96935 | 0.96745 | 0.97661 | 0.9970054886 | 0.93196 | 0.9997848868 | 0.41672 | 7.66670154 | 1 |
| 10 | 87933222 | T | C | Y | H | PTEN | ENST00000371953 | 0.80535 | 0.98311 | 0.97298 | 0.93539 | 0.98938 | 0.9894231294 | 0.9762 | 0.9987056255 | 0.461077 | 6.01362737 | 1 |
| 10 | 87933223 | A | C | Y | S | PTEN | ENST00000371953 | 0.91763 | 0.98725 | 0.93843 | 0.96662 | 0.99105 | 0.9958984351 | 0.98123 | 0.9998099208 | 0.478997 | 7.66670154 | 1 |
| 10 | 87933223 | A | G | Y | C | PTEN | ENST00000371953 | 0.93238 | 0.98161 | 0.97078 | 0.95457 | 0.99603 | 0.9944217395 | 0.97919 | 0.999594152 | 0.463472 | 6.386085202 | 1 |
| 10 | 87933223 | A | T | Y | F | PTEN | ENST00000371953 | 0.86782 | 0.95573 | 0.92395 | 0.82137 | 0.92069 | 0.947676569 | 0.88885 | 0.9826269746 | 0.147327 | 4.817656816 | 0 |
| 10 | 87933225 | G | T | G | W | PTEN | ENST00000371953 | 0.95202 | 0.97822 | 0.97816 | 0.94841 | 0.95899 | 0.9830370876 | 0.94342 | 0.9980040193 | 0.362707 | 8.117983091 | 1 |
| 10 | 87933226 | G | A | G | E | PTEN | ENST00000371953 | 0.79231 | 0.97539 | 0.97808 | 0.91974 | 0.94876 | 0.9898348145 | 0.88938 | 0.9935981035 | 0.373266 | 6.598096632 | 0 |
| 10 | 87933226 | G | C | G | A | PTEN | ENST00000371953 | 0.61134 | 0.85063 | 0.94585 | 0.76282 | 0.87877 | 0.9272445678 | 0.8662 | 0.9721998572 | 0.212066 | 3.654773205 | 0 |
| 10 | 87933226 | G | T | G | V | PTEN | ENST00000371953 | 0.80084 | 0.97582 | 0.94954 | 0.91875 | 0.95389 | 0.9647113111 | 0.93142 | 0.9974892139 | 0.42932 | 6.601410866 | 1 |
| 10 | 87933228 | G | A | E | K | PTEN | ENST00000371953 | 0.51982 | 0.8553 | 0.93187 | 0.85059 | 0.84588 | 0.8876944047 | 0.97671 | 0.8225185871 | 0.372216 | 3.349767923 | 0 |
| 10 | 87933229 | A | C | E | A | PTEN | ENST00000371953 | 0.74898 | 0.84201 | 0.85187 | 0.85366 | 0.93593 | 0.8682551022 | 0.95723 | 0.9474079013 | 0.388793 | 4.1697156 | 0 |
| 10 | 87933229 | A | G | E | G | PTEN | ENST00000371953 | 0.81575 | 0.87276 | 0.80658 | 0.95647 | 0.97475 | 0.8956666768 | 0.97303 | 0.9397303462 | 0.366396 | 5.620071156 | 0 |
| 10 | 87933229 | A | T | E | V | PTEN | ENST00000371953 | 0.77734 | 0.87815 | 0.83609 | 0.88141 | 0.94318 | 0.888491362 | 0.97705 | 0.9829624891 | 0.40045 | 4.521677235 | 0 |
| 10 | 87933230 | A | T | E | D | PTEN | ENST00000371953 | 0.6372 | 0.79756 | 0.85016 | 0.82483 | 0.90007 | 0.8324121567 | 0.93924 | 0.8071362376 | 0.23334 | 3.553969291 | 0 |
| 10 | 87933231 | G | A | V | I | PTEN | ENST00000371953 | 0.57751 | 0.75045 | 0.68328 | 0.93746 | 0.92438 | 0.6460391351 | 0.94812 | 0.9261336923 | 0.349977 | 4.176853967 | 0 |
| 10 | 87933231 | G | C | V | L | PTEN | ENST00000371953 | 0.58453 | 0.79275 | 0.8487 | 0.95621 | 0.90972 | 0.8381698951 | 0.92317 | 0.6429623342 | 0.320548 | 5.08134921 | 0 |
| 10 | 87933231 | G | T | V | L | PTEN | ENST00000371953 | 0.57872 | 0.79275 | 0.8487 | 0.95695 | 0.91143 | 0.8381698951 | 0.92315 | 0.6469274632 | 0.320547 | 5.08134921 | 0 |
| 10 | 87933232 | T | A | V | E | PTEN | ENST00000371953 | 0.70519 | 0.91936 | 0.85934 | 0.97452 | 0.96334 | 0.9168807902 | 0.98748 | 0.8321698308 | 0.503479 | 2.480403516 | 0 |
| 10 | 87933232 | T | C | V | A | PTEN | ENST00000371953 | 0.47658 | 0.84681 | 0.87047 | 0.91227 | 0.84375 | 0.7955937675 | 0.97225 | 0.8280348182 | 0.340352 | 2.064229343 | 0 |
| 10 | 87933232 | T | G | V | G | PTEN | ENST00000371953 | 0.75503 | 0.92349 | 0.73682 | 0.97316 | 0.9703 | 0.885700469 | 0.99234 | 0.9438125491 | 0.51514 | 5.947787113 | 0 |
| 10 | 87933234 | A | G | R | G | PTEN | ENST00000371953 | 0.91884 | 0.98273 | 0.95136 | 0.99576 | 0.99234 | 0.9958673401 | 0.99653 | 0.9981081486 | 0.559511 | 7.401297522 | 1 |
| 10 | 87933234 | A | T | R | W | PTEN | ENST00000371953 | 0.94394 | 0.9713 | 0.98317 | 0.99473 | 0.98904 | 0.9906625776 | 0.98649 | 0.9998947382 | 0.559877 | 8.390425943 | 1 |
| 10 | 87933235 | G | A | R | K | PTEN | ENST00000371953 | 0.86949 | 0.96618 | 0.97596 | 0.98823 | 0.99414 | 0.9856466573 | 0.99592 | 0.989007473 | 0.569861 | 5.956743329 | 1 |
| 10 | 87933235 | G | C | R | T | PTEN | ENST00000371953 | 0.8784 | 0.98858 | 0.96901 | 0.99224 | 0.99843 | 0.9952276067 | 0.99571 | 0.99917382 | 0.593857 | 6.789457683 | 1 |
| 10 | 87933235 | G | T | R | M | PTEN | ENST00000371953 | 0.90605 | 0.9756 | 0.96212 | 0.99217 | 0.99525 | 0.9897527391 | 0.99178 | 0.9995172024 | 0.594138 | 8.390425943 | 1 |
| 10 | 87933236 | G | C | R | S | PTEN | ENST00000371953 | 0.90468 | 0.97613 | 0.95467 | 0.99145 | 0.9986 | 0.9953136619 | 0.99498 | 0.9994273186 | 0.593857 | 6.642359083 | 1 |
| 10 | 87933236 | G | T | R | S | PTEN | ENST00000371953 | 0.90036 | 0.97613 | 0.95467 | 0.99128 | 0.9986 | 0.9953136619 | 0.995 | 0.9994273186 | 0.593855 | 6.642359083 | 1 |
| 10 | 87933237 | A | C | T | P | PTEN | ENST00000371953 | 0.82412 | 0.98348 | 0.8419 | 0.99036 | 0.96334 | 0.9521327105 | 0.97085 | 0.9991213679 | 0.474317 | 6.894061483 | 1 |
| 10 | 87933237 | A | G | T | A | PTEN | ENST00000371953 | 0.78709 | 0.96723 | 0.84863 | 0.98949 | 0.9755 | 0.9129606862 | 0.97287 | 0.9979793429 | 0.415492 | 5.422224749 | 0 |
| 10 | 87933237 | A | T | T | S | PTEN | ENST00000371953 | 0.75009 | 0.9583 | 0.8587 | 0.98415 | 0.96226 | 0.8290739891 | 0.96526 | 0.9955226183 | 0.347371 | 5.728289433 | 0 |
| 10 | 87933238 | C | G | T | S | PTEN | ENST00000371953 | 0.80314 | 0.9583 | 0.8587 | 0.97398 | 0.93083 | 0.8290739891 | 0.94967 | 0.9953415394 | 0.338049 | 5.728289433 | 0 |
| 10 | 87933241 | G | A | R | K | PTEN | ENST00000371953 | 0.56887 | 0.79826 | 0.98994 | 0.92111 | 0.80948 | 0.8404746292 | 0.99015 | 0.8380841613 | 0.446701 | 3.619749417 | 0 |
| 10 | 87933241 | G | C | R | T | PTEN | ENST00000371953 | 0.62985 | 0.8947 | 0.98649 | 0.96228 | 0.91736 | 0.8893723914 | 0.9445 | 0.9374494553 | 0.345002 | 3.573189581 | 0 |
| 10 | 87933241 | G | T | R | I | PTEN | ENST00000371953 | 0.68387 | 0.93051 | 0.98423 | 0.96972 | 0.91399 | 0.8930040336 | 0.97141 | 0.7723765398 | 0.39144 | 3.725626288 | 0 |
| 10 | 87933242 | A | C | R | S | PTEN | ENST00000371953 | 0.55202 | 0.83065 | 0.97952 | 0.94806 | 0.89239 | 0.8966074175 | 0.99379 | 0.7854293585 | 0.45678 | 4.622130321 | 0 |
| 10 | 87933242 | A | T | R | S | PTEN | ENST00000371953 | 0.54623 | 0.83065 | 0.97952 | 0.94859 | 0.89193 | 0.8966074175 | 0.99385 | 0.7854293585 | 0.456776 | 4.622130321 | 0 |
| 10 | 87933243 | G | A | D | N | PTEN | ENST00000371953 | 0.53515 | 0.87117 | 0.94409 | 0.94741 | 0.76685 | 0.8124197407 | 0.87397 | 0.7180814568 | 0.0885977 | 2.609772018 | 0 |
| 10 | 87933243 | G | C | D | H | PTEN | ENST00000371953 | 0.88297 | 0.97834 | 0.95636 | 0.9832 | 0.9681 | 0.9022079044 | 0.9568 | 0.9935848117 | 0.419778 | 6.470451647 | 0 |
| 10 | 87933244 | A | C | D | A | PTEN | ENST00000371953 | 0.92504 | 0.97295 | 0.90145 | 0.97791 | 0.96371 | 0.9275887748 | 0.93628 | 0.9944624091 | 0.389829 | 4.982615989 | 0 |
| 10 | 87933244 | A | G | D | G | PTEN | ENST00000371953 | 0.92397 | 0.97114 | 0.91389 | 0.97503 | 0.95753 | 0.9443756107 | 0.97055 | 0.9944089489 | 0.411602 | 6.557840185 | 0 |
| 10 | 87933244 | A | T | D | V | PTEN | ENST00000371953 | 0.9239 | 0.98449 | 0.90429 | 0.98213 | 0.96956 | 0.9529585181 | 0.97843 | 0.9978824258 | 0.456799 | 5.831330544 | 0 |
| 10 | 87933245 | C | A | D | E | PTEN | ENST00000371953 | 0.62241 | 0.96223 | 0.92866 | 0.94895 | 0.90318 | 0.8359774775 | 0.99223 | 0.959817946 | 0.373012 | 5.71782124 | 0 |
| 10 | 87933246 | A | C | K | Q | PTEN | ENST00000371953 | 0.63145 | 0.8153 | 0.85725 | 0.90427 | 0.85689 | 0.8810519462 | 0.91716 | 0.9265007377 | 0.223032 | 3.722283069 | 0 |
| 10 | 87933246 | A | G | K | E | PTEN | ENST00000371953 | 0.46177 | 0.87958 | 0.89069 | 0.89632 | 0.86653 | 0.9352772397 | 0.98617 | 0.8831144571 | 0.399863 | 3.171485861 | 0 |
| 10 | 87933247 | A | C | K | T | PTEN | ENST00000371953 | 0.71498 | 0.85005 | 0.90064 | 0.92745 | 0.91484 | 0.9237114987 | 0.91551 | 0.9501562119 | 0.307991 | 4.477134436 | 0 |
| 10 | 87933247 | A | G | K | R | PTEN | ENST00000371953 | 0.68875 | 0.72862 | 0.82788 | 0.9001 | 0.82556 | 0.7663465553 | 0.91504 | 0.872505486 | 0.199617 | 3.074438292 | 0 |
| 10 | 87933247 | A | T | K | I | PTEN | ENST00000371953 | 0.9025 | 0.9579 | 0.86715 | 0.96417 | 0.95426 | 0.9440554204 | 0.92452 | 0.9824062586 | 0.336599 | 6.997172207 | 0 |
| 10 | 87933249 | A | G | K | E | PTEN | ENST00000371953 | 0.82763 | 0.97228 | 0.97517 | 0.98166 | 0.96044 | 0.9601884076 | 0.99075 | 0.9947452545 | 0.527394 | 5.26712534 | 0 |
| 10 | 87933250 | A | C | K | T | PTEN | ENST00000371953 | 0.80787 | 0.97725 | 0.97749 | 0.98518 | 0.97587 | 0.9571410154 | 0.96333 | 0.9975668192 | 0.409281 | 8.336584449 | 0 |
| 10 | 87933250 | A | G | K | R | PTEN | ENST00000371953 | 0.80148 | 0.9013 | 0.94803 | 0.96112 | 0.92069 | 0.8250714517 | 0.96058 | 0.9902009368 | 0.268647 | 4.589015781 | 0 |
| 10 | 87933250 | A | T | K | M | PTEN | ENST00000371953 | 0.84111 | 0.98348 | 0.9506 | 0.98806 | 0.97252 | 0.9368302543 | 0.96857 | 0.99949193 | 0.394188 | 6.633443478 | 0 |
| 10 | 87933251 | G | T | K | N | PTEN | ENST00000371953 | 0.86439 | 0.95858 | 0.98213 | 0.98421 | 0.95462 | 0.9544436553 | 0.97009 | 0.996696949 | 0.238113 | 4.972591402 | 0 |
| 10 | 87952118 | G | A | G | R | PTEN | ENST00000371953 | 0.88739 | 0.97893 | 0.99353 | 0.99061 | 0.98904 | 0.9767243903 | 0.97626 | 0.9996316433 | 0.521329 | 7.044029319 | 1 |
| 10 | 87952119 | G | A | G | E | PTEN | ENST00000371953 | 0.93235 | 0.98311 | 0.99171 | 0.9893 | 0.99386 | 0.9851302849 | 0.97988 | 0.9991237521 | 0.589823 | 8.233515645 | 1 |
| 10 | 87952119 | G | C | G | A | PTEN | ENST00000371953 | 0.91543 | 0.96909 | 0.97802 | 0.97229 | 0.97252 | 0.9618567684 | 0.97008 | 0.9971033931 | 0.390309 | 6.289215895 | 0 |
| 10 | 87952119 | G | T | G | V | PTEN | ENST00000371953 | 0.93044 | 0.98161 | 0.97896 | 0.98785 | 0.99138 | 0.9893283847 | 0.98694 | 0.999330163 | 0.581476 | 7.168571635 | 1 |
| 10 | 87952121 | G | A | V | I | PTEN | ENST00000371953 | 0.7548 | 0.88253 | 0.76726 | 0.98208 | 0.87636 | 0.7057727736 | 0.90788 | 0.7928836566 | 0.296516 | 5.844105756 | 0 |
| 10 | 87952122 | T | A | V | E | PTEN | ENST00000371953 | 0.89611 | 0.99148 | 0.91242 | 0.99014 | 0.99652 | 0.9807649083 | 0.99215 | 0.9996600151 | 0.58131 | 8.807274987 | 1 |
| 10 | 87952122 | T | G | V | G | PTEN | ENST00000371953 | 0.89266 | 0.98311 | 0.81653 | 0.99177 | 0.99628 | 0.9729498049 | 0.98627 | 0.9993242025 | 0.581691 | 8.097946139 | 1 |
| 10 | 87952124 | A | C | T | P | PTEN | ENST00000371953 | 0.81166 | 0.98311 | 0.77436 | 0.9845 | 0.97215 | 0.8927867817 | 0.96932 | 0.9970566034 | 0.453114 | 7.298184569 | 1 |
| 10 | 87952124 | A | G | T | A | PTEN | ENST00000371953 | 0.74348 | 0.95933 | 0.77833 | 0.97599 | 0.95572 | 0.8318314184 | 0.96388 | 0.9914610386 | 0.26522 | 7.127497022 | 0 |
| 10 | 87952124 | A | T | T | S | PTEN | ENST00000371953 | 0.72356 | 0.94702 | 0.79329 | 0.9703 | 0.94839 | 0.7030196546 | 0.95526 | 0.9908487797 | 0.290023 | 6.042961755 | 0 |
| 10 | 87952125 | C | A | T | N | PTEN | ENST00000371953 | 0.75877 | 0.97597 | 0.87017 | 0.98782 | 0.91569 | 0.8701185849 | 0.95334 | 0.9965636134 | 0.284762 | 6.491084898 | 0 |
| 10 | 87952125 | C | T | T | I | PTEN | ENST00000371953 | 0.60215 | 0.95226 | 0.91608 | 0.97849 | 0.89055 | 0.885889146 | 0.97602 | 0.9954404831 | 0.435985 | 6.728791399 | 0 |
| 10 | 87952127 | A | C | I | L | PTEN | ENST00000371953 | 0.75893 | 0.94829 | 0.89956 | 0.97374 | 0.89193 | 0.7991470249 | 0.95065 | 0.9748044014 | 0.391794 | 6.645373514 | 0 |
| 10 | 87952127 | A | G | I | V | PTEN | ENST00000371953 | 0.56366 | 0.94028 | 0.8346 | 0.97561 | 0.83834 | 0.6945715519 | 0.89601 | 0.9373706579 | 0.277301 | 6.176292548 | 0 |
| 10 | 87952127 | A | T | I | F | PTEN | ENST00000371953 | 0.81135 | 0.99078 | 0.91406 | 0.9891 | 0.96444 | 0.9127796797 | 0.98962 | 0.9971407652 | 0.57594 | 7.617532526 | 0 |
| 10 | 87952128 | T | A | I | N | PTEN | ENST00000371953 | 0.93657 | 0.99172 | 0.96203 | 0.98269 | 0.97661 | 0.9590276471 | 0.9584 | 0.9970909299 | 0.425487 | 6.798696504 | 0 |
| 10 | 87952128 | T | G | I | S | PTEN | ENST00000371953 | 0.79666 | 0.98291 | 0.92044 | 0.97788 | 0.92601 | 0.9715449253 | 0.99278 | 0.9973086119 | 0.561618 | 6.987043792 | 0 |
| 10 | 87952129 | T | G | I | M | PTEN | ENST00000371953 | 0.82139 | 0.9695 | 0.87777 | 0.97578 | 0.93436 | 0.8037630343 | 0.95998 | 0.9818373322 | 0.421342 | 7.153971541 | 0 |
| 10 | 87952130 | C | G | P | A | PTEN | ENST00000371953 | 0.67732 | 0.96192 | 0.93657 | 0.97974 | 0.99386 | 0.9059245139 | 0.97359 | 0.9901244772 | 0.557473 | 4.847987451 | 0 |

| | | | | | | | | | | | | | | | | | | |
|---|---|---|---|---|---|---|---|---|---|---|---|---|---|---|---|---|---|---|
| 10 | 87952130 | C | T | P | S | PTEN | ENST00000371953 | 0.72764 | 0.9632 | 0.945 | 0.97993 | 0.99386 | 0.9545219712 | 0.98011 | 0.9987134933 | 0.581831 | 5.768978304 | 0 |
| 10 | 87952131 | C | A | P | H | PTEN | ENST00000371953 | 0.89018 | 0.97749 | 0.96734 | 0.98602 | 0.99386 | 0.944004798 | 0.97118 | 0.9998264909 | 0.584537 | 8.02279217 | 0 |
| 10 | 87952131 | C | G | P | R | PTEN | ENST00000371953 | 0.89319 | 0.97054 | 0.97311 | 0.98348 | 0.99676 | 0.9726837072 | 0.98471 | 0.9993195534 | 0.581886 | 5.976778048 | 0 |
| 10 | 87952131 | C | T | P | L | PTEN | ENST00000371953 | 0.90271 | 0.9362 | 0.97527 | 0.97611 | 0.99443 | 0.9643778317 | 0.98956 | 0.9998264909 | 0.572788 | 6.477582092 | 0 |
| 10 | 87952133 | A | G | S | G | PTEN | ENST00000371953 | 0.8356 | 0.96288 | 0.84518 | 0.99171 | 0.99138 | 0.9628742413 | 0.98421 | 0.999153018 | 0.54136 | 7.122370229 | 0 |
| 10 | 87952133 | A | T | S | C | PTEN | ENST00000371953 | 0.8701 | 0.96697 | 0.88991 | 0.99443 | 0.99498 | 0.9699774097 | 0.9898 | 0.9993109703 | 0.579336 | 5.332097498 | 0 |
| 10 | 87952134 | G | A | S | N | PTEN | ENST00000371953 | 0.92819 | 0.96387 | 0.92593 | 0.9819 | 0.96008 | 0.9902426162 | 0.9702 | 0.9958161712 | 0.315889 | 6.465958321 | 0 |
| 10 | 87952134 | G | T | S | I | PTEN | ENST00000371953 | 0.94749 | 0.97893 | 0.90626 | 0.98717 | 0.99578 | 0.9919549892 | 0.99602 | 0.999674201 | 0.593936 | 7.941205299 | 1 |
| 10 | 87952135 | T | A | S | R | PTEN | ENST00000371953 | 0.89143 | 0.97582 | 0.93307 | 0.99376 | 0.9917 | 0.9960478646 | 0.99738 | 0.9996731281 | 0.557809 | 7.941205299 | 1 |
| 10 | 87952136 | C | A | Q | K | PTEN | ENST00000371953 | 0.72124 | 0.98161 | 0.92796 | 0.99302 | 0.99498 | 0.996282041 | 0.99682 | 0.9985758066 | 0.578175 | 8.017356875 | 1 |
| 10 | 87952136 | C | G | Q | E | PTEN | ENST00000371953 | 0.66937 | 0.95226 | 0.88999 | 0.99381 | 0.99676 | 0.9918763428 | 0.99446 | 0.9950775504 | 0.577073 | 7.97418484 | 1 |
| 10 | 87952137 | A | C | Q | P | PTEN | ENST00000371953 | 0.9275 | 0.98012 | 0.89575 | 0.99493 | 0.99722 | 0.999679672 | 0.99593 | 0.9986103773 | 0.588917 | 8.017356875 | 1 |
| 10 | 87952137 | A | G | Q | R | PTEN | ENST00000371953 | 0.91337 | 0.97213 | 0.92517 | 0.99365 | 0.99765 | 0.99583126 | 0.99806 | 0.9992750287 | 0.587574 | 7.558906525 | 1 |
| 10 | 87952137 | A | T | Q | L | PTEN | ENST00000371953 | 0.81959 | 0.96604 | 0.88246 | 0.99505 | 0.98904 | 0.9938838429 | 0.99665 | 0.9992005229 | 0.587054 | 7.951928469 | 1 |
| 10 | 87952138 | G | T | Q | H | PTEN | ENST00000371953 | 0.6731 | 0.9634 | 0.92184 | 0.99314 | 0.98693 | 0.9938630349 | 0.9975 | 0.9982460737 | 0.549544 | 5.691023319 | 0 |
| 10 | 87952139 | A | G | R | G | PTEN | ENST00000371953 | 0.76539 | 0.97027 | 0.9569 | 0.98078 | 0.94949 | 0.9709486395 | 0.99768 | 0.9926540852 | 0.530226 | 7.433957131 | 0 |
| 10 | 87952139 | A | T | R | W | PTEN | ENST00000371953 | 0.94771 | 0.97087 | 0.98509 | 0.98684 | 0.9755 | 0.9461693981 | 0.95351 | 0.9977787137 | 0.505031 | 9.037851044 | 0 |
| 10 | 87952140 | G | A | R | K | PTEN | ENST00000371953 | 0.59747 | 0.85203 | 0.98007 | 0.97166 | 0.91569 | 0.9376577696 | 0.99102 | 0.7606939673 | 0.394619 | 4.89558244 | 0 |
| 10 | 87952140 | G | C | R | T | PTEN | ENST00000371953 | 0.7438 | 0.97924 | 0.9753 | 0.98108 | 0.96371 | 0.9650325893 | 0.99476 | 0.9726492763 | 0.544989 | 7.382843244 | 0 |
| 10 | 87952140 | G | T | R | M | PTEN | ENST00000371953 | 0.92874 | 0.9725 | 0.96818 | 0.98946 | 0.97698 | 0.9309797818 | 0.98683 | 0.9883588552 | 0.550423 | 6.444134634 | 0 |
| 10 | 87952141 | G | C | R | S | PTEN | ENST00000371953 | 0.54136 | 0.90185 | 0.96178 | 0.96435 | 0.87442 | 0.9703779816 | 0.99871 | 0.9458548427 | 0.488608 | 6.353556347 | 0 |
| 10 | 87952141 | G | T | R | S | PTEN | ENST00000371953 | 0.53583 | 0.90185 | 0.96178 | 0.96435 | 0.87442 | 0.9703779816 | 0.99871 | 0.9393530488 | 0.488607 | 6.353556347 | 0 |
| 10 | 87952142 | C | G | R | G | PTEN | ENST00000371953 | 0.78527 | 0.9826 | 0.95391 | 0.99128 | 0.99676 | 0.9930271882 | 0.99611 | 0.9989889264 | 0.580964 | 7.628095543 | 1 |
| 10 | 87952143 | G | C | R | P | PTEN | ENST00000371953 | 0.94901 | 0.98772 | 0.97201 | 0.99376 | 0.99986 | 0.997901253 | 0.99371 | 0.9982702362 | 0.591127 | 7.172742867 | 1 |
| 10 | 87952143 | G | T | R | L | PTEN | ENST00000371953 | 0.66747 | 0.96738 | 0.96909 | 0.99203 | 0.94691 | 0.9900219353 | 0.99474 | 0.9941907525 | 0.582555 | 5.618877385 | 0 |
| 10 | 87952145 | T | A | Y | N | PTEN | ENST00000371953 | 0.83521 | 0.99179 | 0.98716 | 0.99345 | 0.98938 | 0.9975910054 | 0.96488 | 0.9997693896 | 0.456488 | 8.342410544 | 1 |
| 10 | 87952145 | T | G | Y | D | PTEN | ENST00000371953 | 0.8453 | 0.99399 | 0.98605 | 0.99422 | 0.99786 | 0.9993839665 | 0.99581 | 0.9998928308 | 0.582258 | 8.435011945 | 1 |
| 10 | 87952146 | A | C | Y | S | PTEN | ENST00000371953 | 0.91383 | 0.9856 | 0.96977 | 0.99016 | 0.99786 | 0.9967620855 | 0.9951 | 0.9997487664 | 0.58887 | 6.442696768 | 0 |
| 10 | 87952146 | A | G | Y | C | PTEN | ENST00000371953 | 0.93562 | 0.98161 | 0.98717 | 0.99029 | 0.99824 | 0.996854737 | 0.9877 | 0.9997333884 | 0.590403 | 6.566819872 | 0 |
| 10 | 87952146 | A | T | Y | F | PTEN | ENST00000371953 | 0.86439 | 0.94902 | 0.96083 | 0.97736 | 0.97698 | 0.9642185559 | 0.93958 | 0.993876636 | 0.324046 | 5.392630784 | 0 |
| 10 | 87952148 | G | A | V | M | PTEN | ENST00000371953 | 0.83073 | 0.96506 | 0.97744 | 0.97556 | 0.9681 | 0.9030296909 | 0.98518 | 0.9922429919 | 0.583055 | 6.43518205 | 1 |
| 10 | 87952148 | G | T | V | L | PTEN | ENST00000371953 | 0.80013 | 0.91241 | 0.9679 | 0.95696 | 0.9252 | 0.9254487698 | 0.93766 | 0.8927685485 | 0.371327 | 5.699227995 | 0 |
| 10 | 87952149 | T | A | V | E | PTEN | ENST00000371953 | 0.89884 | 0.99155 | 0.96752 | 0.9949 | 0.99919 | 0.9923652696 | 0.99508 | 0.9996127486 | 0.585875 | 8.355337236 | 1 |
| 10 | 87952149 | T | C | V | A | PTEN | ENST00000371953 | 0.87024 | 0.97717 | 0.97191 | 0.99153 | 0.9844 | 0.9301049356 | 0.97195 | 0.9898047609 | 0.546359 | 6.695135802 | 0 |
| 10 | 87952149 | T | G | V | G | PTEN | ENST00000371953 | 0.89543 | 0.98311 | 0.90281 | 0.99534 | 0.99906 | 0.9786543044 | 0.99082 | 0.9993995428 | 0.586159 | 8.624849295 | 1 |
| 10 | 87952151 | T | G | Y | D | PTEN | ENST00000371953 | 0.53529 | 0.92454 | 0.98375 | 0.95141 | 0.82443 | 0.9547074257 | 0.99254 | 0.8348597884 | 0.443611 | 3.229910982 | 0 |
| 10 | 87952152 | A | C | Y | S | PTEN | ENST00000371953 | 0.44869 | 0.8844 | 0.96015 | 0.93319 | 0.79697 | 0.9211650967 | 0.98487 | 0.627795577 | 0.340343 | 3.55174582 | 0 |
| 10 | 87952154 | T | A | Y | N | PTEN | ENST00000371953 | 0.78503 | 0.99056 | 0.9667 | 0.98696 | 0.98071 | 0.9962013703 | 0.96981 | 0.9997779727 | 0.52044 | 6.707859911 | 1 |
| 10 | 87952154 | T | G | Y | D | PTEN | ENST00000371953 | 0.81127 | 0.99329 | 0.96571 | 0.98679 | 0.98799 | 0.9988039168 | 0.99523 | 0.9987387061 | 0.580594 | 7.16372556 | 1 |
| 10 | 87952155 | A | C | Y | S | PTEN | ENST00000371953 | 0.7157 | 0.97448 | 0.93019 | 0.9901 | 0.9681 | 0.9952159289 | 0.99229 | 0.999368608 | 0.584252 | 8.328544113 | 1 |
| 10 | 87952155 | A | G | Y | C | PTEN | ENST00000371953 | 0.92747 | 0.98144 | 0.97224 | 0.99267 | 0.99652 | 0.9955974967 | 0.97954 | 0.9997333884 | 0.589475 | 8.133183928 | 1 |
| 10 | 87952157 | T | A | Y | N | PTEN | ENST00000371953 | 0.84241 | 0.9893 | 0.97023 | 0.99129 | 0.98183 | 0.9917357819 | 0.95677 | 0.9991939664 | 0.479414 | 9.402378812 | 1 |
| 10 | 87952157 | T | C | Y | H | PTEN | ENST00000371953 | 0.8366 | 0.97893 | 0.97359 | 0.98195 | 0.98513 | 0.9750854307 | 0.97131 | 0.995806098 | 0.503839 | 8.547832029 | 0 |
| 10 | 87952157 | T | G | Y | D | PTEN | ENST00000371953 | 0.84891 | 0.99204 | 0.96735 | 0.99276 | 0.99326 | 0.9969639 | 0.99427 | 0.9990065694 | 0.578512 | 9.402378812 | 1 |
| 10 | 87952158 | A | C | Y | S | PTEN | ENST00000371953 | 0.91793 | 0.97725 | 0.93344 | 0.98926 | 0.99202 | 0.9888917878 | 0.97661 | 0.9989004135 | 0.569367 | 8.490514601 | 0 |
| 10 | 87952158 | A | G | Y | C | PTEN | ENST00000371953 | 0.93204 | 0.91105 | 0.97327 | 0.98715 | 0.98834 | 0.9835080432 | 0.93204 | 0.9954118133 | 0.530372 | 7.089147879 | 0 |
| 10 | 87952161 | G | A | S | N | PTEN | ENST00000371953 | 0.4668 | 0.76995 | 0.89865 | 0.93653 | 0.72701 | 0.8285657002 | 0.70887 | 0.8069394231 | 0.0839689 | 3.834960565 | 0 |
| 10 | 87952162 | C | A | S | R | PTEN | ENST00000371953 | 0.56055 | 0.83124 | 0.91017 | 0.94107 | 0.85326 | 0.9281042384 | 0.9719 | 0.5683190227 | 0.391826 | 5.959471686 | 0 |
| 10 | 87952163 | T | A | Y | N | PTEN | ENST00000371953 | 0.52659 | 0.91029 | 0.84202 | 0.85581 | 0.76289 | 0.9208269903 | 0.97171 | 0.7560020089 | 0.262349 | 3.795474802 | 0 |
| 10 | 87952163 | T | C | Y | H | PTEN | ENST00000371953 | 0.49305 | 0.86062 | 0.84511 | 0.83613 | 0.69082 | 0.8621994413 | 0.74136 | 0.5624328852 | 0.063219 | 2.410048023 | 0 |
| 10 | 87952163 | T | G | Y | D | PTEN | ENST00000371953 | 0.54365 | 0.92438 | 0.82877 | 0.85515 | 0.77335 | 0.9552753432 | 0.98945 | 0.6884919405 | 0.303104 | 3.902674582 | 0 |
| 10 | 87952164 | A | C | Y | S | PTEN | ENST00000371953 | 0.51573 | 0.88212 | 0.75145 | 0.86026 | 0.74235 | 0.9211179034 | 0.93457 | 0.760964632 | 0.218301 | 3.741640654 | 0 |
| 10 | 87952164 | A | G | Y | C | PTEN | ENST00000371953 | 0.60149 | 0.85466 | 0.84095 | 0.91513 | 0.88684 | 0.9172484906 | 0.94322 | 0.8829120994 | 0.331966 | 5.489811723 | 0 |
| 10 | 87952164 | A | T | Y | F | PTEN | ENST00000371953 | 0.43629 | 0.76384 | 0.72408 | 0.91031 | 0.68381 | 0.7899790138 | 0.86147 | 0.8605796099 | 0.186643 | 2.600396893 | 0 |
| 10 | 87952166 | C | G | L | V | PTEN | ENST00000371953 | 0.54021 | 0.85315 | 0.90887 | 0.94391 | 0.90274 | 0.8470475164 | 0.96977 | 0.9333271384 | 0.336887 | 5.436203851 | 0 |
| 10 | 87952167 | T | C | L | P | PTEN | ENST00000371953 | 0.92084 | 0.98602 | 0.98192 | 0.99125 | 0.99919 | 0.9930770531 | 0.99756 | 0.9968603339 | 0.571988 | 8.967110943 | 1 |
| 10 | 87952167 | T | G | L | R | PTEN | ENST00000371953 | 0.90645 | 0.97869 | 0.97507 | 0.98638 | 0.99326 | 0.9900377445 | 0.99655 | 0.9955345392 | 0.55849 | 7.629388684 | 1 |
| 10 | 87952169 | T | A | L | I | PTEN | ENST00000371953 | 0.47363 | 0.87158 | 0.94915 | 0.93132 | 0.87733 | 0.7419690288 | 0.87998 | 0.3864653616 | 0.256116 | 3.412138472 | 0 |
| 10 | 87952170 | T | C | L | S | PTEN | ENST00000371953 | 0.91962 | 0.95671 | 0.95245 | 0.98665 | 0.997 | 0.9585264641 | 0.99717 | 0.9915633202 | 0.557375 | 5.907451349 | 0 |
| 10 | 87952172 | A | C | K | Q | PTEN | ENST00000371953 | 0.51659 | 0.81254 | 0.84075 | 0.84121 | 0.71783 | 0.8363251548 | 0.95495 | 0.7061591148 | 0.223712 | 4.441274084 | 0 |
| 10 | 87952172 | A | G | K | E | PTEN | ENST00000371953 | 0.5886 | 0.88002 | 0.87378 | 0.9177 | 0.83779 | 0.8802546033 | 0.99376 | 0.7090821862 | 0.362055 | 5.401432278 | 0 |
| 10 | 87952173 | A | C | K | T | PTEN | ENST00000371953 | 0.53193 | 0.84721 | 0.88393 | 0.91743 | 0.87099 | 0.8990989419 | 0.96473 | 0.7483417392 | 0.30745 | 4.471985583 | 0 |
| 10 | 87952173 | A | T | K | M | PTEN | ENST00000371953 | 0.69548 | 0.88733 | 0.81461 | 0.95345 | 0.86902 | 0.8895906623 | 0.94328 | 0.9338892698 | 0.281134 | 6.701211296 | 0 |
| 10 | 87952175 | A | C | N | H | PTEN | ENST00000371953 | 0.44795 | 0.80076 | 0.7383 | 0.86084 | 0.70508 | 0.7777878062 | 0.88406 | 0.566647768 | 0.0799353 | 3.634792507 | 0 |
| 10 | 87952175 | A | G | N | D | PTEN | ENST00000371953 | 0.47269 | 0.82915 | 0.78589 | 0.80819 | 0.73734 | 0.8234726484 | 0.90813 | 0.7059034109 | 0.09647 | 5.012347017 | 0 |
| 10 | 87952175 | A | T | N | Y | PTEN | ENST00000371953 | 0.62135 | 0.84676 | 0.77932 | 0.90348 | 0.81588 | 0.8710858485 | 0.93889 | 0.930119276 | 0.214247 | 6.029625595 | 0 |
| 10 | 87952176 | A | C | N | T | PTEN | ENST00000371953 | 0.48424 | 0.79751 | 0.71522 | 0.84989 | 0.75211 | 0.8100669732 | 0.88714 | 0.7337324619 | 0.149929 | 5.529831193 | 0 |
| 10 | 87952176 | A | G | N | S | PTEN | ENST00000371953 | 0.42645 | 0.71745 | 0.66338 | 0.80037 | 0.68203 | 0.685658539 | 0.82656 | 0.8853055239 | 0.0558164 | 4.018944334 | 0 |
| 10 | 87952176 | A | T | N | I | PTEN | ENST00000371953 | 0.65649 | 0.86502 | 0.74382 | 0.92259 | 0.83172 | 0.9037069917 | 0.89887 | 0.6938628813 | 0.227985 | 6.100375764 | 0 |
| 10 | 87952177 | T | A | N | K | PTEN | ENST00000371953 | 0.39929 | 0.83166 | 0.81068 | 0.87416 | 0.72626 | 0.826760111 | 0.81464 | 0.3562490046 | 0.0677592 | 4.18995291 | 0 |
| 10 | 87952178 | C | A | H | N | PTEN | ENST00000371953 | 0.36985 | 0.81779 | 0.8273 | 0.77974 | 0.69425 | 0.7284991944 | 0.71977 | 0.7298641205 | 0.0586095 | 1.374767653 | 0 |
| 10 | 87952178 | C | G | H | D | PTEN | ENST00000371953 | 0.39387 | 0.87419 | 0.82687 | 0.77218 | 0.74306 | 0.8687119425 | 0.88136 | 0.6650544405 | 0.19083 | 3.136576294 | 0 |
| 10 | 87952178 | C | T | H | Y | PTEN | ENST00000371953 | 0.53086 | 0.8325 | 0.84148 | 0.76547 | 0.75687 | 0.7821630428 | 0.81579 | 0.6683687568 | 0.168843 | 4.321057686 | 0 |
| 10 | 87952179 | A | G | H | R | PTEN | ENST00000371953 | 0.41991 | 0.8457 | 0.83978 | 0.80644 | 0.69168 | 0.7595433475 | 0.75816 | 0.834158659 | 0.153309 | 1.630825397 | 0 |
| 10 | 87952179 | A | T | H | L | PTEN | ENST00000371953 | 0.49203 | 0.84999 | 0.79249 | 0.85033 | 0.74376 | 0.8321343812 | 0.83005 | 0.678443241 | 0.20016 | 4.194417858 | 0 |
| 10 | 87952180 | T | A | H | Q | PTEN | ENST00000371953 | 0.25246 | 0.79222 | 0.80252 | 0.82452 | 0.66161 | 0.6533631083 | 0.54984 | 0.06835552305 | 0.0329786 | -0.005413378716 | 0 |
| 10 | 87952181 | C | G | L | V | PTEN | ENST00000371953 | 0.41063 | 0.79874 | 0.79528 | 0.82864 | 0.82273 | 0.7260590222 | 0.90474 | 0.6725868583 | 0.173726 | 5.059189953 | 0 |
| 10 | 87952182 | T | A | L | Q | PTEN | ENST00000371953 | 0.7614 | 0.83366 | 0.85694 | 0.96547 | 0.93319 | 0.8893872366 | 0.97942 | 0.9769482613 | 0.416548 | 6.752725196 | 0 |
| 10 | 87952182 | T | C | L | P | PTEN | ENST00000371953 | 0.76816 | 0.91763 | 0.93759 | 0.95969 | 0.95207 | 0.9410248426 | 0.98226 | 0.9791040421 | 0.457703 | 7.088482991 | 0 |
| 10 | 87952182 | T | G | L | R | PTEN | ENST00000371953 | 0.75726 | 0.89993 | 0.92147 | 0.94111 | 0.91442 | 0.9202749371 | 0.99237 | 0.9441759586 | 0.45615 | 6.529501607 | 0 |
| 10 | 87952184 | G | A | D | N | PTEN | ENST00000371953 | 0.47615 | 0.78402 | 0.83734 | 0.80941 | 0.71153 | 0.558626406 | 0.80363 | 0.6563633499 | -0.0068855 | 2.743494734 | 0 |
| 10 | 87952184 | G | C | D | H | PTEN | ENST00000371953 | 0.52792 | 0.8385 | 0.85931 | 0.83934 | 0.72093 | 0.6869478521 | 0.81016 | 0.6754673123 | 0.185283 | 3.113920852 | 0 |
| 10 | 87952184 | G | T | D | Y | PTEN | ENST00000371953 | 0.67024 | 0.86235 | 0.84413 | 0.86885 | 0.825 | 0.8071642457 | 0.91442 | 0.8574232459 | 0.227337 | 4.327643119 | 0 |
| 10 | 87952185 | A | C | D | A | PTEN | ENST00000371953 | 0.50896 | 0.85106 | 0.77326 | 0.72457 | 0.66064 | 0.7256458774 | 0.76886 | 0.768488884 | 0.157662 | 3.565378301 | 0 |
| 10 | 87952185 | A | G | D | G | PTEN | ENST00000371953 | 0.55298 | 0.86384 | 0.7808 | 0.74515 | 0.66449 | 0.7925766853 | 0.82501 | 0.7389039397 | 0.169786 | 3.694992986 | 0 |
| 10 | 87952187 | T | A | Y | N | PTEN | ENST00000371953 | 0.84111 | 0.96871 | 0.91975 | 0.94876 | 0.94129 | 0.9689199871 | 0.93846 | 0.9995682836 | 0.356987 | 8.02091241 | 0 |
| 10 | 87952187 | T | C | Y | H | PTEN | ENST00000371953 | 0.83982 | 0.97972 | 0.92628 | 0.91786 | 0.97997 | 0.9450528151 | 0.98567 | 0.9984450936 | 0.457614 | 7.737918834 | 0 |
| 10 | 87952187 | T | G | Y | D | PTEN | ENST00000371953 | 0.85212 | 0.97491 | 0.9135 | 0.95624 | 0.97104 | 0.9882398258 | 0.98304 | 0.9991703033 | 0.458398 | 8.334081786 | 1 |
| 10 | 87952188 | A | G | Y | C | PTEN | ENST00000371953 | 0.93235 | 0.9362 | 0.92387 | 0.94388 | 0.95608 | 0.9722769246 | 0.93979 | 0.9992941618 | 0.431825 | 7.274443102 | 0 |
| 10 | 87952190 | A | G | R | G | PTEN | ENST00000371953 | 0.58895 | 0.85979 | 0.79 | 0.64172 | 0.67843 | 0.8000772996 | 0.95416 | 0.8479251862 | 0.0947101 | 4.821340661 | 0 |
| 10 | 87952191 | G | A | R | K | PTEN | ENST00000371953 | 0.34098 | 0.79496 | 0.85387 | 0.56656 | 0.67015 | 0.6358776403 | 0.61487 | 0.5987557173 | -0.0657857 | -0.6336296197 | 0 |
| 10 | 87952191 | G | C | R | T | PTEN | ENST00000371953 | 0.45604 | 0.89487 | 0.84305 | 0.68139 | 0.71861 | 0.7192380773 | 0.83556 | 0.6723736151 | 0.112887 | 2.874099139 | 0 |
| 10 | 87952191 | G | T | R | I | PTEN | ENST00000371953 | 0.62023 | 0.94361 | 0.82753 | 0.77343 | 0.72626 | 0.7781206796 | 0.81221 | 0.8432205319 | 0.127991 | 3.99286582 | 0 |
| 10 | 87952192 | A | C | R | S | PTEN | ENST00000371953 | 0.45967 | 0.8284 | 0.80189 | 0.67024 | 0.64775 | 0.7226680289 | 0.8397 | 0.8137413263 | 0.106945 | 3.252081777 | 0 |
| 10 | 87952192 | A | T | R | S | PTEN | ENST00000371953 | 0.4545 | 0.8284 | 0.80189 | 0.66459 | 0.64775 | 0.7226680289 | 0.8398 | 0.802038312 | 0.106943 | 3.252081777 | 0 |
| 10 | 87952193 | C | A | P | T | PTEN | ENST00000371953 | 0.62415 | 0.89093 | 0.91803 | 0.86086 | 0.9248 | 0.9065837746 | 0.88614 | 0.95897609 | 0.28329 | 4.621244869 | 0 |
| 10 | 87952193 | C | G | P | A | PTEN | ENST00000371953 | 0.48398 | 0.84169 | 0.88596 | 0.86616 | 0.89422 | 0.857157122 | 0.79953 | 0.9558730721 | 0.233855 | 4.787836857 | 0 |
| 10 | 87952193 | C | T | P | S | PTEN | ENST00000371953 | 0.52101 | 0.89591 | 0.89399 | 0.8652 | 0.89603 | 0.9109192818 | 0.84518 | 0.9605494738 | 0.262654 | 3.64131642 | 0 |
| 10 | 87952194 | C | A | P | Q | PTEN | ENST00000371953 | 0.47093 | 0.85459 | 0.89838 | 0.85011 | 0.86403 | 0.9189261055 | 0.90402 | 0.9543299675 | 0.270499 | 3.805557121 | 0 |
| 10 | 87952194 | C | T | P | L | PTEN | ENST00000371953 | 0.80337 | 0.87391 | 0.94263 | 0.85733 | 0.94839 | 0.9257583564 | 0.94772 | 0.8751099138 | 0.414915 | 5.581984135 | 0 |
| 10 | 87952196 | G | A | V | M | PTEN | ENST00000371953 | 0.68355 | 0.87103 | 0.77213 | 0.83795 | 0.80889 | 0.7437879896 | 0.90657 | 0.8364942985 | 0.236626 | 6.274385418 | 0 |

| | | | | | | | | | | | | | | | | | | |
|---|---|---|---|---|---|---|---|---|---|---|---|---|---|---|---|---|---|---|
| 10 | 87952197 | T | A | V | E | PTEN | ENST00000371953 | 0.49997 | 0.93926 | 0.73976 | 0.89521 | 0.87148 | 0.9309033251 | 0.98246 | 0.9093694091 | 0.413889 | 4.655182276 | 0 |
| 10 | 87952197 | T | C | V | A | PTEN | ENST00000371953 | 0.46519 | 0.86649 | 0.75002 | 0.85612 | 0.76356 | 0.8233495766 | 0.95998 | 0.7866466641 | 0.267159 | 5.910779995 | 0 |
| 10 | 87952197 | T | G | V | G | PTEN | ENST00000371953 | 0.50537 | 0.88902 | 0.57927 | 0.90221 | 0.88776 | 0.9361739268 | 0.98333 | 0.961591959 | 0.422676 | 6.717776335 | 0 |
| 10 | 87952199 | G | A | A | T | PTEN | ENST00000371953 | 0.43938 | 0.78348 | 0.89347 | 0.67601 | 0.69168 | 0.6915846696 | 0.91857 | 0.8856776357 | 0.153171 | 3.246376625 | 0 |
| 10 | 87952199 | G | C | A | P | PTEN | ENST00000371953 | 0.54521 | 0.86095 | 0.93353 | 0.6603 | 0.62657 | 0.8997523087 | 0.85552 | 0.7155671756 | 0.104883 | 4.38200549 | 0 |
| 10 | 87952199 | G | T | A | S | PTEN | ENST00000371953 | 0.48876 | 0.7642 | 0.86001 | 0.70325 | 0.71783 | 0.6645237193 | 0.9215 | 0.651019156 | 0.158514 | 3.985780151 | 0 |
| 10 | 87952200 | C | A | A | E | PTEN | ENST00000371953 | 0.56041 | 0.83804 | 0.93288 | 0.80013 | 0.8345 | 0.9071333628 | 0.96884 | 0.8467435243 | 0.377006 | 5.5981595 | 0 |
| 10 | 87952200 | C | G | A | G | PTEN | ENST00000371953 | 0.56929 | 0.80121 | 0.87619 | 0.78051 | 0.80536 | 0.735016185 | 0.88511 | 0.8345344482 | 0.174256 | 6.050853775 | 0 |
| 10 | 87952200 | C | T | A | V | PTEN | ENST00000371953 | 0.44466 | 0.79485 | 0.90511 | 0.78374 | 0.76487 | 0.7562391668 | 0.93082 | 0.7156607924 | 0.213868 | 4.474229862 | 0 |
| 10 | 87952202 | C | A | L | M | PTEN | ENST00000371953 | 0.56103 | 0.94482 | 0.84559 | 0.89641 | 0.86302 | 0.7129672715 | 0.88135 | 0.9593598247 | 0.215487 | 5.158463739 | 0 |
| 10 | 87952202 | C | G | L | V | PTEN | ENST00000371953 | 0.53663 | 0.8804 | 0.81083 | 0.8808 | 0.87588 | 0.79015823 | 0.89989 | 0.8929063678 | 0.249785 | 4.354580739 | 0 |
| 10 | 87952203 | T | A | L | Q | PTEN | ENST00000371953 | 0.91665 | 0.95873 | 0.8721 | 0.95849 | 0.98257 | 0.9717824205 | 0.9838 | 0.9990770817 | 0.456243 | 7.137638172 | 0 |
| 10 | 87952203 | T | C | L | P | PTEN | ENST00000371953 | 0.92453 | 0.99134 | 0.94585 | 0.9627 | 0.99552 | 0.990819292 | 0.98839 | 0.9988623857 | 0.461992 | 7.161564895 | 1 |
| 10 | 87952203 | T | G | L | R | PTEN | ENST00000371953 | 0.92237 | 0.98704 | 0.92916 | 0.95945 | 0.9917 | 0.9861967 | 0.98533 | 0.9990770817 | 0.461361 | 6.812412718 | 0 |
| 10 | 87952205 | T | A | L | M | PTEN | ENST00000371953 | 0.60786 | 0.82641 | 0.8806 | 0.80184 | 0.82103 | 0.625891134 | 0.8825 | 0.7679252625 | 0.187171 | 6.190591828 | 0 |
| 10 | 87952205 | T | G | L | V | PTEN | ENST00000371953 | 0.53657 | 0.80585 | 0.85694 | 0.83655 | 0.85586 | 0.7269719729 | 0.92224 | 0.6029408574 | 0.186723 | 5.326772062 | 0 |
| 10 | 87952206 | T | C | L | S | PTEN | ENST00000371953 | 0.91721 | 0.92838 | 0.91188 | 0.9195 | 0.95353 | 0.9371550167 | 0.95521 | 0.9417644739 | 0.413355 | 6.078671181 | 0 |
| 10 | 87952206 | T | G | L | W | PTEN | ENST00000371953 | 0.92068 | 0.9286 | 0.9456 | 0.92648 | 0.93786 | 0.8869070182 | 0.93616 | 0.9603250027 | 0.389813 | 8.263410763 | 0 |
| 10 | 87952207 | G | T | L | F | PTEN | ENST00000371953 | 0.46755 | 0.82335 | 0.8928 | 0.72635 | 0.79151 | 0.768772144 | 0.93633 | 0.7513819337 | 0.202436 | 5.947742306 | 0 |
| 10 | 87952208 | T | A | F | I | PTEN | ENST00000371953 | 0.60662 | 0.88078 | 0.95035 | 0.80165 | 0.81182 | 0.9274227787 | 0.91959 | 0.624561429 | 0.251062 | 3.993022681 | 0 |
| 10 | 87952208 | T | G | F | V | PTEN | ENST00000371953 | 0.63858 | 0.86723 | 0.94843 | 0.85835 | 0.88117 | 0.9266246922 | 0.96107 | 0.7929847836 | 0.388071 | 4.158885013 | 0 |
| 10 | 87952209 | T | A | F | Y | PTEN | ENST00000371953 | 0.90396 | 0.94361 | 0.96387 | 0.87466 | 0.85274 | 0.9124582225 | 0.87176 | 0.9602781534 | 0.199125 | 6.394549976 | 0 |
| 10 | 87952209 | T | C | F | S | PTEN | ENST00000371953 | 0.94439 | 0.94871 | 0.95583 | 0.9043 | 0.96846 | 0.9860225102 | 0.97547 | 0.960483551 | 0.422113 | 6.406677551 | 1 |
| 10 | 87952209 | T | G | F | C | PTEN | ENST00000371953 | 0.94475 | 0.98311 | 0.96735 | 0.92927 | 0.95717 | 0.979915263 | 0.96127 | 0.9710974693 | 0.420864 | 6.847040813 | 0 |
| 10 | 87952210 | T | G | F | L | PTEN | ENST00000371953 | 0.39568 | 0.83485 | 0.97062 | 0.75244 | 0.66733 | 0.8810217463 | 0.95919 | 0.8350108266 | 0.199011 | 2.436323112 | 0 |
| 10 | 87952211 | C | A | H | N | PTEN | ENST00000371953 | 0.50043 | 0.82279 | 0.92254 | 0.83116 | 0.76619 | 0.8503559661 | 0.84371 | 0.8793227673 | 0.121928 | 5.012546393 | 0 |
| 10 | 87952211 | C | G | H | D | PTEN | ENST00000371953 | 0.55663 | 0.88228 | 0.9225 | 0.84989 | 0.91526 | 0.9313347853 | 0.94437 | 0.9648094177 | 0.405581 | 5.641677092 | 0 |
| 10 | 87952211 | C | T | H | Y | PTEN | ENST00000371953 | 0.5014 | 0.83558 | 0.92677 | 0.819 | 0.92397 | 0.8229111895 | 0.9145 | 0.9424421787 | 0.34129 | 6.038736241 | 0 |
| 10 | 87952212 | A | C | H | P | PTEN | ENST00000371953 | 0.75973 | 0.96604 | 0.87688 | 0.88969 | 0.95608 | 0.9799056095 | 0.96183 | 0.9513030052 | 0.416575 | 5.673009201 | 1 |
| 10 | 87952212 | A | G | H | R | PTEN | ENST00000371953 | 0.46407 | 0.84673 | 0.931 | 0.72623 | 0.85996 | 0.8314742529 | 0.97362 | 0.9635745287 | 0.360641 | 3.590792682 | 0 |
| 10 | 87952213 | C | A | H | Q | PTEN | ENST00000371953 | 0.41658 | 0.80461 | 0.90715 | 0.85831 | 0.7675 | 0.7942777248 | 0.94768 | 0.6061332822 | 0.334286 | 3.94003721 | 0 |
| 10 | 87952214 | A | C | K | Q | PTEN | ENST00000371953 | 0.64368 | 0.83979 | 0.84503 | 0.80821 | 0.84588 | 0.8502000091 | 0.93564 | 0.7788609266 | 0.250083 | 4.918052884 | 0 |
| 10 | 87952214 | A | G | K | E | PTEN | ENST00000371953 | 0.57787 | 0.88126 | 0.88101 | 0.81662 | 0.87345 | 0.9180706489 | 0.97345 | 0.8043455482 | 0.372146 | 4.670282017 | 0 |
| 10 | 87952215 | A | C | K | T | PTEN | ENST00000371953 | 0.56741 | 0.84979 | 0.88957 | 0.83004 | 0.84482 | 0.8692765412 | 0.96052 | 0.8591893911 | 0.36949 | 4.816367453 | 0 |
| 10 | 87952215 | A | G | K | R | PTEN | ENST00000371953 | 0.74061 | 0.73533 | 0.81457 | 0.70835 | 0.82273 | 0.7429854688 | 0.93063 | 0.7741033435 | 0.227418 | 4.205535952 | 0 |
| 10 | 87952215 | A | T | K | M | PTEN | ENST00000371953 | 0.9231 | 0.92296 | 0.82176 | 0.88298 | 0.93554 | 0.8785661505 | 0.9372 | 0.9703072309 | 0.374362 | 6.727672866 | 0 |
| 10 | 87952216 | G | T | K | N | PTEN | ENST00000371953 | 0.72092 | 0.90136 | 0.9036 | 0.82442 | 0.76487 | 0.9051532395 | 0.89139 | 0.9431592226 | 0.111596 | 5.351332169 | 0 |
| 10 | 87952217 | A | G | M | V | PTEN | ENST00000371953 | 0.46955 | 0.79636 | 0.85111 | 0.61784 | 0.67108 | 0.7885517562 | 0.98445 | 0.6294527054 | 0.247986 | 3.846254031 | 0 |
| 10 | 87952217 | A | T | M | L | PTEN | ENST00000371953 | 0.48297 | 0.7703 | 0.90114 | 0.63579 | 0.62657 | 0.7730841441 | 0.97241 | 0.6626970489 | 0.214411 | 4.019332346 | 0 |
| 10 | 87952218 | T | A | M | K | PTEN | ENST00000371953 | 0.73781 | 0.9362 | 0.9483 | 0.78399 | 0.94986 | 0.980210032 | 0.97513 | 0.9745457172 | 0.411923 | 6.401191609 | 1 |
| 10 | 87952218 | T | G | M | R | PTEN | ENST00000371953 | 0.88904 | 0.94361 | 0.93939 | 0.78654 | 0.94949 | 0.9864629079 | 0.97619 | 0.9893260002 | 0.431555 | 5.428413924 | 1 |
| 10 | 87952219 | G | T | M | I | PTEN | ENST00000371953 | 0.41582 | 0.81997 | 0.90154 | 0.63002 | 0.66449 | 0.7556966019 | 0.82381 | 0.7929369807 | 0.0839104 | 3.321089429 | 0 |
| 10 | 87952220 | A | G | M | V | PTEN | ENST00000371953 | 0.41961 | 0.79574 | 0.71042 | 0.4371 | 0.57617 | 0.5530149573 | 0.64404 | 0.5897129774 | -0.0513812 | -0.8513635595 | 0 |
| 10 | 87952220 | A | T | M | L | PTEN | ENST00000371953 | 0.44765 | 0.76995 | 0.78502 | 0.49282 | 0.51387 | 0.5492917516 | 0.81105 | 0.23694645 | -0.0276536 | 0.3838567876 | 0 |
| 10 | 87952221 | T | C | M | T | PTEN | ENST00000371953 | 0.42341 | 0.83141 | 0.89063 | 0.53022 | 0.55051 | 0.6846206712 | 0.85061 | 0.4642827213 | 0.0913023 | 1.414170591 | 0 |
| 10 | 87952221 | T | G | M | R | PTEN | ENST00000371953 | 0.47602 | 0.89614 | 0.83959 | 0.58917 | 0.65967 | 0.8816270384 | 0.9718 | 0.4634815454 | 0.284057 | 0.9289906947 | 0 |
| 10 | 87952222 | G | T | M | I | PTEN | ENST00000371953 | 0.50134 | 0.81997 | 0.77158 | 0.51467 | 0.54769 | 0.538172316 | 0.82826 | 0.6963697672 | 0.0661274 | 1.132584166 | 0 |
| 10 | 87952223 | T | A | F | I | PTEN | ENST00000371953 | 0.48215 | 0.88308 | 0.88691 | 0.8037 | 0.73296 | 0.875314286 | 0.94196 | 0.6508274213 | 0.233696 | 4.028319011 | 0 |
| 10 | 87952223 | T | G | F | V | PTEN | ENST00000371953 | 0.62749 | 0.87665 | 0.86971 | 0.88147 | 0.84906 | 0.8727221229 | 0.96716 | 0.8493658304 | 0.379769 | 4.844155043 | 0 |
| 10 | 87952224 | T | A | F | Y | PTEN | ENST00000371953 | 0.89382 | 0.96308 | 0.90221 | 0.85204 | 0.89055 | 0.8232041545 | 0.88573 | 0.944634974 | 0.197464 | 8.50623242 | 0 |
| 10 | 87952225 | T | G | F | L | PTEN | ENST00000371953 | 0.46413 | 0.83677 | 0.91499 | 0.74278 | 0.65277 | 0.8179963486 | 0.91143 | 0.6790732145 | 0.173084 | 3.84385059 | 0 |
| 10 | 87952226 | G | C | E | Q | PTEN | ENST00000371953 | 0.49679 | 0.79938 | 0.87034 | 0.82738 | 0.69339 | 0.735895135 | 0.97173 | 0.8097449541 | 0.249315 | 4.405489387 | 0 |
| 10 | 87952227 | A | C | E | A | PTEN | ENST00000371953 | 0.62143 | 0.81384 | 0.87922 | 0.8297 | 0.87247 | 0.7951860347 | 0.96652 | 0.9079868197 | 0.395433 | 5.80250611 | 0 |
| 10 | 87952227 | A | G | E | G | PTEN | ENST00000371953 | 0.68275 | 0.84606 | 0.84432 | 0.86167 | 0.91694 | 0.8614582154 | 0.96129 | 0.9812491536 | 0.40113 | 6.394611347 | 0 |
| 10 | 87952227 | A | T | E | V | PTEN | ENST00000371953 | 0.81072 | 0.84214 | 0.86394 | 0.87554 | 0.91442 | 0.8414013234 | 0.92666 | 0.9766943455 | 0.26592 | 5.569693296 | 0 |
| 10 | 87952228 | A | T | E | D | PTEN | ENST00000371953 | 0.40746 | 0.76231 | 0.88301 | 0.72553 | 0.66258 | 0.7320301978 | 0.98258 | 0.6592527032 | 0.27044 | 4.005624743 | 0 |
| 10 | 87952229 | A | C | T | P | PTEN | ENST00000371953 | 0.59908 | 0.86323 | 0.77527 | 0.80407 | 0.90096 | 0.8476412821 | 0.96396 | 0.8490806818 | 0.396342 | 4.832613638 | 1 |
| 10 | 87952229 | A | G | T | A | PTEN | ENST00000371953 | 0.54223 | 0.84908 | 0.77879 | 0.82367 | 0.89873 | 0.7608924545 | 0.93736 | 0.9727223516 | 0.278051 | 6.625955554 | 0 |
| 10 | 87952229 | A | T | T | S | PTEN | ENST00000371953 | 0.51521 | 0.81046 | 0.79193 | 0.80446 | 0.87442 | 0.6432354351 | 0.93165 | 0.9797381163 | 0.250075 | 5.018307965 | 0 |
| 10 | 87952230 | C | A | T | N | PTEN | ENST00000371953 | 0.57568 | 0.84503 | 0.85552 | 0.84428 | 0.86098 | 0.8089052625 | 0.85931 | 0.9600981474 | 0.139309 | 5.534162978 | 0 |
| 10 | 87952230 | C | T | T | I | PTEN | ENST00000371953 | 0.64617 | 0.88897 | 0.89341 | 0.87297 | 0.967 | 0.8368505336 | 0.94651 | 0.9930045009 | 0.427629 | 7.756334126 | 0 |
| 10 | 87952232 | A | G | I | V | PTEN | ENST00000371953 | 0.35421 | 0.73554 | 0.67613 | 0.48816 | 0.50616 | 0.5513194951 | 0.64606 | 0.4517690241 | -0.0253486 | 0.7979880768 | 0 |
| 10 | 87952232 | A | T | I | F | PTEN | ENST00000371953 | 0.51692 | 0.90891 | 0.78935 | 0.68356 | 0.76685 | 0.8862747212 | 0.94699 | 0.797988832 | 0.166094 | 4.620112023 | 0 |
| 10 | 87952233 | T | G | I | S | PTEN | ENST00000371953 | 0.78003 | 0.89572 | 0.79913 | 0.75646 | 0.89422 | 0.9614990796 | 0.93582 | 0.8648908734 | 0.322173 | 3.979400173 | 0 |
| 10 | 87952234 | T | G | I | M | PTEN | ENST00000371953 | 0.50759 | 0.85799 | 0.74473 | 0.65593 | 0.70914 | 0.7267309522 | 0.85639 | 0.7316102386 | 0.104415 | 4.085249927 | 0 |
| 10 | 87952235 | C | A | P | T | PTEN | ENST00000371953 | 0.64984 | 0.96121 | 0.92345 | 0.95907 | 0.98109 | 0.9223806659 | 0.97381 | 0.9962349534 | 0.49785 | 6.443463793 | 0 |
| 10 | 87952235 | C | T | P | S | PTEN | ENST00000371953 | 0.67421 | 0.94445 | 0.90269 | 0.94819 | 0.97923 | 0.9320254578 | 0.96345 | 0.9946547747 | 0.486632 | 5.023743028 | 0 |
| 10 | 87952238 | A | G | M | V | PTEN | ENST00000371953 | 0.35841 | 0.79968 | 0.72964 | 0.80911 | 0.81124 | 0.826568129 | 0.98262 | 0.2540254791 | 0.321839 | 5.315582203 | 1 |
| 10 | 87952239 | T | A | M | K | PTEN | ENST00000371953 | 0.43454 | 0.9014 | 0.87614 | 0.82135 | 0.8306 | 0.9557283413 | 0.96001 | 0.6744119525 | 0.385463 | 6.066778634 | 1 |
| 10 | 87952239 | T | G | M | R | PTEN | ENST00000371953 | 0.47583 | 0.90739 | 0.85282 | 0.85235 | 0.87099 | 0.9590854579 | 0.95738 | 0.8220246434 | 0.400279 | 6.836538244 | 0 |
| 10 | 87952240 | G | T | M | I | PTEN | ENST00000371953 | 0.46987 | 0.82495 | 0.79666 | 0.83798 | 0.76223 | 0.8339510973 | 0.92873 | 0.8707153201 | 0.25003 | 4.527802894 | 1 |
| 10 | 87952241 | T | A | F | I | PTEN | ENST00000371953 | 0.67931 | 0.89679 | 0.9538 | 0.78542 | 0.90318 | 0.9148652754 | 0.89125 | 0.9457864165 | 0.254873 | 4.033156965 | 0 |
| 10 | 87952242 | T | A | F | Y | PTEN | ENST00000371953 | 0.89651 | 0.87913 | 0.96582 | 0.8103 | 0.88117 | 0.7941155709 | 0.94482 | 0.959100306 | 0.305897 | 4.537619668 | 0 |
| 10 | 87952242 | T | C | F | S | PTEN | ENST00000371953 | 0.93688 | 0.96003 | 0.95556 | 0.84859 | 0.9458 | 0.9396097724 | 0.98115 | 0.9850971699 | 0.431945 | 5.529196039 | 0 |
| 10 | 87952242 | T | G | F | C | PTEN | ENST00000371953 | 0.94762 | 0.92119 | 0.9677 | 0.89803 | 0.96517 | 0.9396128571 | 0.97837 | 0.9921707511 | 0.441882 | 5.419447049 | 0 |
| 10 | 87952243 | C | G | F | L | PTEN | ENST00000371953 | 0.54196 | 0.83979 | 0.97093 | 0.7617 | 0.82216 | 0.8390440941 | 0.95832 | 0.9751716852 | 0.349868 | 3.773841308 | 0 |
| 10 | 87952244 | A | G | S | G | PTEN | ENST00000371953 | 0.50367 | 0.76935 | 0.72677 | 0.8457 | 0.7337 | 0.5998335412 | 0.89706 | 0.8826389909 | 0.175918 | 4.679346437 | 0 |
| 10 | 87952244 | A | T | S | C | PTEN | ENST00000371953 | 0.84096 | 0.95278 | 0.78902 | 0.88856 | 0.91229 | 0.7170551893 | 0.89824 | 0.9875361404 | 0.282746 | 8.271520103 | 0 |
| 10 | 87952245 | G | A | S | N | PTEN | ENST00000371953 | 0.52161 | 0.77139 | 0.83054 | 0.77395 | 0.7337 | 0.5674217867 | 0.78472 | 0.8230819702 | 0.0478954 | 3.09613155 | 0 |
| 10 | 87952245 | G | C | S | T | PTEN | ENST00000371953 | 0.5647 | 0.76771 | 0.77743 | 0.80316 | 0.59638 | 0.5356908582 | 0.88785 | 0.8665537834 | 0.116095 | 2.499073292 | 0 |
| 10 | 87952245 | G | T | S | I | PTEN | ENST00000371953 | 0.82358 | 0.87616 | 0.80185 | 0.87878 | 0.91652 | 0.7950237431 | 0.92719 | 0.9862537384 | 0.355235 | 5.631790334 | 0 |
| 10 | 87952246 | T | A | S | R | PTEN | ENST00000371953 | 0.53414 | 0.82558 | 0.83728 | 0.87568 | 0.78035 | 0.715712541 | 0.98597 | 0.7308360934 | 0.359656 | 5.41350753 | 0 |
| 10 | 87952247 | G | A | G | S | PTEN | ENST00000371953 | 0.73333 | 0.80071 | 0.68213 | 0.64866 | 0.71073 | 0.735283949 | 0.84462 | 0.9216696024 | 0.106109 | 3.985144533 | 0 |
| 10 | 87952247 | G | T | G | C | PTEN | ENST00000371953 | 0.94387 | 0.96051 | 0.77699 | 0.86096 | 0.86902 | 0.8749789613 | 0.85812 | 0.9929305315 | 0.254538 | 7.646615973 | 0 |
| 10 | 87952248 | G | A | G | D | PTEN | ENST00000371953 | 0.89509 | 0.87272 | 0.83941 | 0.78228 | 0.89558 | 0.8024404642 | 0.80014 | 0.9726062417 | 0.240499 | 5.490731766 | 0 |
| 10 | 87952248 | G | C | G | A | PTEN | ENST00000371953 | 0.58988 | 0.82168 | 0.71577 | 0.80587 | 0.86653 | 0.7384458198 | 0.878 | 0.9889085889 | 0.244926 | 4.796589408 | 0 |
| 10 | 87952248 | G | T | G | V | PTEN | ENST00000371953 | 0.90794 | 0.86419 | 0.73197 | 0.83751 | 0.90798 | 0.864339405 | 0.81974 | 0.987488687 | 0.259258 | 5.733483224 | 0 |
| 10 | 87952250 | G | A | G | R | PTEN | ENST00000371953 | 0.68939 | 0.87608 | 0.94827 | 0.82203 | 0.88543 | 0.8766723028 | 0.97155 | 0.9188120365 | 0.325802 | 5.838382135 | 0 |
| 10 | 87952251 | G | A | G | E | PTEN | ENST00000371953 | 0.90555 | 0.96883 | 0.93768 | 0.86927 | 0.92561 | 0.89352955 | 0.92538 | 0.9922170639 | 0.308971 | 5.96296299 | 0 |
| 10 | 87952251 | G | C | G | A | PTEN | ENST00000371953 | 0.57125 | 0.86018 | 0.87985 | 0.77111 | 0.83004 | 0.8412563282 | 0.93833 | 0.9544557929 | 0.215622 | 4.650274581 | 0 |
| 10 | 87952251 | G | T | G | V | PTEN | ENST00000371953 | 0.90799 | 0.92409 | 0.88875 | 0.83929 | 0.93241 | 0.9288063546 | 0.92535 | 0.9942210913 | 0.362469 | 5.831013751 | 0 |
| 10 | 87952253 | A | C | T | P | PTEN | ENST00000371953 | 0.92276 | 0.87364 | 0.5205 | 0.81096 | 0.93631 | 0.8955685582 | 0.91594 | 0.9677414894 | 0.356627 | 5.370258334 | 0 |
| 10 | 87952253 | A | G | T | A | PTEN | ENST00000371953 | 0.75009 | 0.79184 | 0.51949 | 0.71932 | 0.86603 | 0.7775167509 | 0.90509 | 0.9218074083 | 0.235556 | 5.004077151 | 0 |
| 10 | 87952253 | A | T | T | S | PTEN | ENST00000371953 | 0.53522 | 0.77911 | 0.53299 | 0.70121 | 0.65277 | 0.6444931299 | 0.86315 | 0.7638474107 | 0.125868 | 3.881627217 | 0 |
| 10 | 87952254 | C | A | T | N | PTEN | ENST00000371953 | 0.73669 | 0.85459 | 0.63389 | 0.7782 | 0.79334 | 0.8178704125 | 0.78503 | 0.9462340474 | 0.100081 | 4.082478436 | 0 |
| 10 | 87952256 | T | A | C | S | PTEN | ENST00000371953 | 0.69581 | 0.88413 | 0.9332 | 0.92793 | 0.98513 | 0.9352494635 | 0.95834 | 0.9595829844 | 0.430207 | 4.525168161 | 0 |
| 10 | 87952256 | T | C | C | R | PTEN | ENST00000371953 | 0.88529 | 0.98858 | 0.97584 | 0.95389 | 0.97624 | 0.966800556 | 0.97719 | 0.9958640337 | 0.445519 | 6.557211931 | 0 |
| 10 | 87952257 | G | A | C | Y | PTEN | ENST00000371953 | 0.94562 | 0.98704 | 0.98018 | 0.90915 | 0.97997 | 0.9568677016 | 0.97538 | 0.9907337427 | 0.442177 | 5.491879185 | 0 |
| 10 | 87952257 | G | C | C | S | PTEN | ENST00000371953 | 0.86629 | 0.88413 | 0.9332 | 0.90484 | 0.967 | 0.9352494635 | 0.95341 | 0.9710184336 | 0.438236 | 4.525168161 | 0 |
| 10 | 87952257 | G | T | C | F | PTEN | ENST00000371953 | 0.89474 | 0.91868 | 0.94358 | 0.90362 | 0.97923 | 0.9414579141 | 0.9824 | 0.9863212705 | 0.430893 | 4.454127273 | 0 |
| 10 | 87952258 | C | G | C | W | PTEN | ENST00000371953 | 0.92438 | 0.98329 | 0.96377 | 0.9197 | 0.95645 | 0.9561828819 | 0.98265 | 0.9901365042 | 0.439993 | 6.401348366 | 0 |

| | | | | | | | | | | | | | | | | | |
|---|---|---|---|---|---|---|---|---|---|---|---|---|---|---|---|---|---|
| 10 | 87952259 | A | G | N | D | PTEN | ENST00000371953 | 0.5322 | 0.82772 | 0.89419 | 0.81149 | 0.57617 | 0.7903921448 | 0.85669 | 0.8670738935 | 0.0555547 | 4.221043909 | 0 |
| 10 | 87952259 | A | T | N | Y | PTEN | ENST00000371953 | 0.63689 | 0.84808 | 0.88395 | 0.83473 | 0.88165 | 0.8699350307 | 0.97071 | 0.9393889308 | 0.304004 | 3.491342982 | 0 |
| 10 | 87957853 | A | C | N | T | PTEN | ENST00000371953 | 0.51869 | 0.80146 | 0.84573 | 0.74297 | 0.65178 | 0.7970554113 | 0.84961 | 0.6270135045 | 0.0854594 | 1.976755779 | 0 |
| 10 | 87957853 | A | G | N | S | PTEN | ENST00000371953 | 0.45468 | 0.71963 | 0.8016 | 0.71368 | 0.63418 | 0.703654279 | 0.82682 | 0.7121013999 | -0.0300819 | 2.065016263 | 0 |
| 10 | 87957853 | A | T | N | I | PTEN | ENST00000371953 | 0.59689 | 0.88449 | 0.85364 | 0.87472 | 0.8124 | 0.9115627747 | 0.90659 | 0.6761918068 | 0.227273 | 4.620350975 | 0 |
| 10 | 87957854 | T | A | N | K | PTEN | ENST00000371953 | 0.52022 | 0.83166 | 0.90712 | 0.76206 | 0.70752 | 0.8186727819 | 0.7983 | 0.655220747 | 0.0101867 | 4.088106796 | 0 |
| 10 | 87957855 | C | A | P | T | PTEN | ENST00000371953 | 0.51481 | 0.91213 | 0.8864 | 0.86853 | 0.96517 | 0.9619904852 | 0.9689 | 0.9814649224 | 0.414087 | 5.970037386 | 0 |
| 10 | 87957855 | C | G | P | A | PTEN | ENST00000371953 | 0.33597 | 0.80543 | 0.85324 | 0.78353 | 0.87345 | 0.9300661701 | 0.92834 | 0.7890867591 | 0.279645 | 4.787038453 | 0 |
| 10 | 87957855 | C | T | P | S | PTEN | ENST00000371953 | 0.55264 | 0.87344 | 0.86553 | 0.87991 | 0.95389 | 0.9690262306 | 0.95388 | 0.9707708955 | 0.415092 | 5.154386642 | 0 |
| 10 | 87957856 | C | A | P | H | PTEN | ENST00000371953 | 0.84612 | 0.97311 | 0.90331 | 0.90612 | 0.95645 | 0.9683380507 | 0.93542 | 0.9887666106 | 0.406983 | 5.403785365 | 0 |
| 10 | 87957858 | C | A | Q | K | PTEN | ENST00000371953 | 0.50929 | 0.86227 | 0.64642 | 0.76075 | 0.8367 | 0.9013751425 | 0.92835 | 0.6795316424 | 0.207691 | 3.976420126 | 0 |
| 10 | 87957858 | C | G | Q | E | PTEN | ENST00000371953 | 0.48544 | 0.81823 | 0.57566 | 0.71465 | 0.78535 | 0.9219815951 | 0.89113 | 0.7200055122 | 0.136559 | 4.822832158 | 0 |
| 10 | 87957859 | A | C | Q | P | PTEN | ENST00000371953 | 0.72965 | 0.85125 | 0.5742 | 0.79488 | 0.92723 | 0.9888777414 | 0.94743 | 0.8884414434 | 0.345326 | 5.751247575 | 0 |
| 10 | 87957859 | A | G | Q | R | PTEN | ENST00000371953 | 0.54351 | 0.81651 | 0.63469 | 0.7406 | 0.85586 | 0.9124674353 | 0.94013 | 0.7433533072 | 0.209061 | 4.428410643 | 0 |
| 10 | 87957860 | G | T | Q | H | PTEN | ENST00000371953 | 0.55456 | 0.77738 | 0.62609 | 0.64775 | 0.73223 | 0.8752031262 | 0.887 | 0.4140584271 | 0.0410416 | 4.337976457 | 0 |
| 10 | 87957861 | T | A | F | I | PTEN | ENST00000371953 | 0.72748 | 0.88572 | 0.91992 | 0.84061 | 0.91526 | 0.960181164 | 0.87441 | 0.9820507765 | 0.248424 | 4.445186407 | 0 |
| 10 | 87957861 | T | G | F | V | PTEN | ENST00000371953 | 0.85894 | 0.89307 | 0.90587 | 0.88446 | 0.94839 | 0.9523604834 | 0.95442 | 0.9892610312 | 0.395346 | 3.572853438 | 0 |
| 10 | 87957862 | T | G | F | C | PTEN | ENST00000371953 | 0.95277 | 0.936 | 0.9353 | 0.91514 | 0.97067 | 0.9847864359 | 0.95394 | 0.9892075658 | 0.405799 | 4.50125111 | 0 |
| 10 | 87957864 | G | T | V | L | PTEN | ENST00000371953 | 0.53535 | 0.79379 | 0.86367 | 0.83005 | 0.68292 | 0.895220578 | 0.89076 | 0.9384815693 | 0.105635 | 5.375868848 | 0 |
| 10 | 87957865 | T | A | V | E | PTEN | ENST00000371953 | 0.93457 | 0.91593 | 0.87473 | 0.88702 | 0.93939 | 0.9651118416 | 0.97815 | 0.9663737416 | 0.409219 | 4.522926717 | 0 |
| 10 | 87957865 | T | C | V | A | PTEN | ENST00000371953 | 0.78185 | 0.87895 | 0.88134 | 0.87278 | 0.81876 | 0.8878236045 | 0.93103 | 0.9400146604 | 0.240173 | 5.24451597 | 0 |
| 10 | 87957865 | T | G | V | G | PTEN | ENST00000371953 | 0.9313 | 0.91663 | 0.76799 | 0.91585 | 0.9367 | 0.9736559284 | 0.93292 | 0.983024478 | 0.370937 | 5.723317408 | 0 |
| 10 | 87957867 | G | A | V | I | PTEN | ENST00000371953 | 0.4355 | 0.77177 | 0.69676 | 0.66763 | 0.58263 | 0.7889640748 | 0.77226 | 0.9428349137 | 0.0305239 | 2.91536223 | 0 |
| 10 | 87957867 | G | C | V | L | PTEN | ENST00000371953 | 0.67326 | 0.82425 | 0.86238 | 0.87578 | 0.8887 | 0.928078335 | 0.92107 | 0.9482395649 | 0.227324 | 3.750421335 | 0 |
| 10 | 87957868 | T | A | V | D | PTEN | ENST00000371953 | 0.94015 | 0.99114 | 0.87845 | 0.95581 | 0.98183 | 0.9968177205 | 0.98137 | 0.9974677411 | 0.43493 | 5.600602816 | 1 |
| 10 | 87957868 | T | C | V | A | PTEN | ENST00000371953 | 0.91305 | 0.96994 | 0.87708 | 0.90394 | 0.94618 | 0.9439084283 | 0.91958 | 0.9733299017 | 0.308718 | 5.169485906 | 0 |
| 10 | 87957868 | T | G | V | G | PTEN | ENST00000371953 | 0.93704 | 0.98243 | 0.75782 | 0.95729 | 0.97735 | 0.9934318913 | 0.97684 | 0.996757741 | 0.450038 | 5.775776707 | 1 |
| 10 | 87957870 | T | A | C | S | PTEN | ENST00000371953 | 0.48889 | 0.84751 | 0.75444 | 0.59992 | 0.613 | 0.9243226021 | 0.60857 | 0.5097009881 | 0.0260538 | 0.9707432115 | 0 |
| 10 | 87957870 | T | G | C | G | PTEN | ENST00000371953 | 0.53778 | 0.85069 | 0.71764 | 0.62052 | 0.4903 | 0.9503275069 | 0.71027 | 0.8820835352 | 0.0188164 | 2.97537036 | 0 |
| 10 | 87957871 | G | A | C | Y | PTEN | ENST00000371953 | 0.50582 | 0.91619 | 0.86978 | 0.37638 | 0.66828 | 0.9372008467 | 0.41357 | 0.7660400867 | -0.0772878 | -1.320825447 | 0 |
| 10 | 87957871 | G | C | C | S | PTEN | ENST00000371953 | 0.57294 | 0.84751 | 0.75444 | 0.45741 | 0.62101 | 0.9243226021 | 0.62942 | 0.4762449122 | 0.0203341 | 0.9707432115 | 0 |
| 10 | 87957871 | G | T | C | F | PTEN | ENST00000371953 | 0.5663 | 0.89749 | 0.76844 | 0.49521 | 0.59638 | 0.9058284058 | 0.74915 | 0.8927313089 | 0.0444612 | 1.079956237 | 0 |
| 10 | 87957873 | C | G | Q | E | PTEN | ENST00000371953 | 0.50608 | 0.84214 | 0.78613 | 0.78986 | 0.84482 | 0.9457982274 | 0.91314 | 0.7714598775 | 0.233689 | 4.175231461 | 0 |
| 10 | 87957874 | A | C | Q | P | PTEN | ENST00000371953 | 0.90735 | 0.97332 | 0.79301 | 0.94341 | 0.97327 | 0.9903365897 | 0.9549 | 0.9927555617 | 0.434774 | 5.118659065 | 0 |
| 10 | 87957875 | G | T | Q | H | PTEN | ENST00000371953 | 0.61626 | 0.83608 | 0.83352 | 0.8782 | 0.86803 | 0.9484527045 | 0.95292 | 0.9414803982 | 0.273565 | 3.714631451 | 0 |
| 10 | 87957876 | C | A | L | I | PTEN | ENST00000371953 | 0.43142 | 0.83961 | 0.93554 | 0.79035 | 0.71627 | 0.7533822969 | 0.82885 | 0.6148973703 | 0.0961473 | 4.90675999 | 0 |
| 10 | 87957876 | C | G | L | V | PTEN | ENST00000371953 | 0.40952 | 0.8556 | 0.9037 | 0.78514 | 0.75755 | 0.8334574058 | 0.8847 | 0.6002307534 | 0.145097 | 5.674252651 | 0 |
| 10 | 87957877 | T | A | L | Q | PTEN | ENST00000371953 | 0.53778 | 0.81548 | 0.94116 | 0.85205 | 0.79878 | 0.9004560774 | 0.97552 | 0.546355056 | 0.352129 | 3.654886553 | 0 |
| 10 | 87957877 | T | C | L | P | PTEN | ENST00000371953 | 0.76926 | 0.91251 | 0.9775 | 0.90603 | 0.93939 | 0.9693998938 | 0.96819 | 0.7827728987 | 0.39903 | 5.039036529 | 0 |
| 10 | 87957877 | T | G | L | R | PTEN | ENST00000371953 | 0.71353 | 0.89552 | 0.96851 | 0.8914 | 0.88212 | 0.9390379577 | 0.96203 | 0.8050044775 | 0.348041 | 5.196801891 | 0 |
| 10 | 87957879 | A | C | K | Q | PTEN | ENST00000371953 | 0.79626 | 0.86771 | 0.79219 | 0.80816 | 0.85482 | 0.926224561 | 0.89035 | 0.9060198665 | 0.190342 | 5.429603133 | 0 |
| 10 | 87957879 | A | G | K | E | PTEN | ENST00000371953 | 0.66399 | 0.90455 | 0.83118 | 0.81639 | 0.90229 | 0.9605699341 | 0.95277 | 0.9540936351 | 0.382294 | 5.237052044 | 0 |
| 10 | 87957880 | A | C | K | T | PTEN | ENST00000371953 | 0.58283 | 0.85402 | 0.83319 | 0.83082 | 0.85222 | 0.9586921847 | 0.95629 | 0.8599727154 | 0.298068 | 4.229245836 | 0 |
| 10 | 87957880 | A | G | K | R | PTEN | ENST00000371953 | 0.76084 | 0.78007 | 0.76137 | 0.81021 | 0.81876 | 0.8495405143 | 0.87975 | 0.6116938389 | 0.0838814 | 5.045076512 | 0 |
| 10 | 87957881 | G | T | K | N | PTEN | ENST00000371953 | 0.50237 | 0.84444 | 0.85291 | 0.75518 | 0.75485 | 0.9582785854 | 0.80303 | 0.8019225001 | 0.0569874 | 3.922643586 | 0 |
| 10 | 87957882 | G | A | V | M | PTEN | ENST00000371953 | 0.72043 | 0.85586 | 0.81961 | 0.82782 | 0.79028 | 0.8225270741 | 0.87329 | 0.8951537013 | 0.20968 | 5.201781058 | 0 |
| 10 | 87957883 | T | A | V | E | PTEN | ENST00000371953 | 0.8235 | 0.96171 | 0.78835 | 0.93175 | 0.87733 | 0.9471442221 | 0.93657 | 0.9210624099 | 0.418044 | 4.780762007 | 0 |
| 10 | 87957883 | T | C | V | A | PTEN | ENST00000371953 | 0.77601 | 0.85878 | 0.80239 | 0.84257 | 0.83943 | 0.8780267804 | 0.89855 | 0.8014157414 | 0.266458 | 4.214131583 | 0 |
| 10 | 87957883 | T | G | V | G | PTEN | ENST00000371953 | 0.82342 | 0.8704 | 0.67304 | 0.91128 | 0.89285 | 0.9576656367 | 0.93713 | 0.9483178258 | 0.411733 | 5.100700774 | 0 |
| 10 | 87957886 | A | C | K | T | PTEN | ENST00000371953 | 0.92326 | 0.89805 | 0.88837 | 0.90607 | 0.94394 | 0.9522383894 | 0.91453 | 0.987291038 | 0.313867 | 5.283102152 | 0 |
| 10 | 87957886 | A | G | K | R | PTEN | ENST00000371953 | 0.79524 | 0.87154 | 0.82559 | 0.8153 | 0.84159 | 0.8858712712 | 0.90461 | 0.9233392477 | 0.17929 | 4.628920729 | 0 |
| 10 | 87957886 | A | T | K | M | PTEN | ENST00000371953 | 0.9395 | 0.98311 | 0.82538 | 0.9318 | 0.94802 | 0.9502348829 | 0.93191 | 0.9966707826 | 0.321595 | 5.382834015 | 0 |
| 10 | 87957887 | G | T | K | N | PTEN | ENST00000371953 | 0.93339 | 0.90987 | 0.90114 | 0.84427 | 0.89376 | 0.9673366486 | 0.89992 | 0.9821152687 | 0.134191 | 5.762237042 | 0 |
| 10 | 87957888 | A | C | I | L | PTEN | ENST00000371953 | 0.41375 | 0.79847 | 0.81936 | 0.77929 | 0.73 | 0.8910831533 | 0.90318 | 0.7199286222 | 0.196065 | 2.861728597 | 0 |
| 10 | 87957888 | A | G | I | V | PTEN | ENST00000371953 | 0.51876 | 0.79745 | 0.73529 | 0.74562 | 0.73296 | 0.7827919109 | 0.81425 | 0.7238197923 | 0.102965 | 2.877373952 | 0 |
| 10 | 87957888 | A | T | I | L | PTEN | ENST00000371953 | 0.40875 | 0.79847 | 0.81936 | 0.77929 | 0.73 | 0.8910831533 | 0.90348 | 0.7199286222 | 0.196063 | 2.861728597 | 0 |
| 10 | 87957889 | T | A | I | K | PTEN | ENST00000371953 | 0.87031 | 0.95564 | 0.88065 | 0.92033 | 0.98294 | 0.9876960489 | 0.9604 | 0.9032210112 | 0.423484 | 4.3514167 | 0 |
| 10 | 87957890 | A | G | I | M | PTEN | ENST00000371953 | 0.52879 | 0.94063 | 0.7821 | 0.87448 | 0.86352 | 0.888085502 | 0.91909 | 0.9058699012 | 0.196956 | 5.432758716 | 0 |
| 10 | 87957891 | T | C | Y | H | PTEN | ENST00000371953 | 0.51481 | 0.86366 | 0.59537 | 0.45482 | 0.60717 | 0.8491549942 | 0.55011 | 0.7343814969 | -0.0297415 | -0.3916764239 | 0 |
| 10 | 87957891 | T | G | Y | D | PTEN | ENST00000371953 | 0.63942 | 0.96407 | 0.60179 | 0.7948 | 0.8517 | 0.9734540313 | 0.81552 | 0.9724758863 | 0.113205 | 3.004297077 | 0 |
| 10 | 87957892 | A | C | Y | S | PTEN | ENST00000371953 | 0.57596 | 0.9015 | 0.51218 | 0.75814 | 0.76356 | 0.9404168514 | 0.83982 | 0.8972283602 | 0.0888915 | 3.262200461 | 0 |
| 10 | 87957894 | T | A | S | T | PTEN | ENST00000371953 | 0.39923 | 0.76326 | 0.45422 | 0.53422 | 0.60717 | 0.7032246089 | 0.65474 | 0.7060003281 | -0.0852018 | -1.153995403 | 0 |
| 10 | 87957894 | T | C | S | P | PTEN | ENST00000371953 | 0.62354 | 0.85777 | 0.45489 | 0.81067 | 0.83725 | 0.9602964964 | 0.89123 | 0.9628998041 | 0.148637 | 3.11266762 | 0 |
| 10 | 87957894 | T | G | S | A | PTEN | ENST00000371953 | 0.53886 | 0.73866 | 0.42229 | 0.74662 | 0.62213 | 0.7449435585 | 0.90247 | 0.9235979319 | 0.0184788 | 2.395468613 | 0 |
| 10 | 87957895 | C | A | S | Y | PTEN | ENST00000371953 | 0.798 | 0.87291 | 0.52907 | 0.8338 | 0.87247 | 0.8448794873 | 0.81317 | 0.9659212828 | 0.094426 | 3.370722679 | 0 |
| 10 | 87957895 | C | G | S | C | PTEN | ENST00000371953 | 0.83837 | 0.93482 | 0.46374 | 0.8093 | 0.85792 | 0.8719133913 | 0.80495 | 0.9759943485 | 0.0911276 | 3.519057147 | 0 |
| 10 | 87957895 | C | T | S | F | PTEN | ENST00000371953 | 0.84218 | 0.8286 | 0.50038 | 0.83282 | 0.85012 | 0.8590382177 | 0.76179 | 0.9571980238 | 0.0757144 | 3.908272304 | 0 |
| 10 | 87957897 | T | A | S | T | PTEN | ENST00000371953 | 0.86119 | 0.83331 | 0.62478 | 0.8324 | 0.87198 | 0.9072212082 | 0.89912 | 0.9333279729 | 0.160359 | 4.327853111 | 0 |
| 10 | 87957897 | T | C | S | P | PTEN | ENST00000371953 | 0.93324 | 0.86759 | 0.64224 | 0.89754 | 0.97848 | 0.9847202248 | 0.95743 | 0.9841433764 | 0.42438 | 5.392924815 | 0 |
| 10 | 87957897 | T | G | S | A | PTEN | ENST00000371953 | 0.87503 | 0.86244 | 0.60293 | 0.85258 | 0.84694 | 0.8556396779 | 0.8932 | 0.8970276713 | 0.150299 | 4.513471746 | 0 |
| 10 | 87957898 | C | A | S | Y | PTEN | ENST00000371953 | 0.75559 | 0.87619 | 0.70209 | 0.88838 | 0.90668 | 0.9577832551 | 0.922 | 0.9109390378 | 0.320044 | 5.112702566 | 0 |
| 10 | 87957898 | C | G | S | C | PTEN | ENST00000371953 | 0.79207 | 0.90464 | 0.63054 | 0.86509 | 0.90711 | 0.9435640423 | 0.86781 | 0.9907317758 | 0.267205 | 4.979267669 | 0 |
| 10 | 87957898 | C | T | S | F | PTEN | ENST00000371953 | 0.81127 | 0.94479 | 0.67179 | 0.8972 | 0.94913 | 0.9633963563 | 0.95239 | 0.9901694655 | 0.394324 | 5.587582317 | 0 |
| 10 | 87957900 | A | C | N | H | PTEN | ENST00000371953 | 0.56838 | 0.94397 | 0.18543 | 0.75545 | 0.66922 | 0.7271090222 | 0.93034 | 0.8259280324 | -0.0451017 | 4.949601629 | 0 |
| 10 | 87957900 | A | G | N | D | PTEN | ENST00000371953 | 0.39121 | 0.82827 | 0.21598 | 0.67435 | 0.58135 | 0.7467738804 | 0.90111 | 0.6909219623 | -0.0794278 | 3.122467362 | 0 |
| 10 | 87957900 | A | T | N | Y | PTEN | ENST00000371953 | 0.59248 | 0.8556 | 0.21023 | 0.80069 | 0.68646 | 0.8323633246 | 0.83721 | 0.5788261848 | 0.00310245 | 5.028200071 | 0 |
| 10 | 87957901 | A | C | N | T | PTEN | ENST00000371953 | 0.24103 | 0.79786 | 0.16356 | 0.63706 | 0.42851 | 0.7555496374 | 0.71538 | 0.1328990906 | -0.114128 | 3.118344958 | 0 |
| 10 | 87957901 | A | G | N | S | PTEN | ENST00000371953 | 0.20752 | 0.72049 | 0.13344 | 0.59937 | 0.43033 | 0.6371354177 | 0.77009 | 0.04202516771 | -0.103171 | 3.025724558 | 0 |
| 10 | 87957901 | A | T | N | I | PTEN | ENST00000371953 | 0.31842 | 0.85505 | 0.19371 | 0.79237 | 0.64053 | 0.8804381574 | 0.86043 | 0.5087306499 | -0.012961 | 4.695195442 | 0 |
| 10 | 87957902 | T | A | N | K | PTEN | ENST00000371953 | 0.32178 | 0.83137 | 0.24418 | 0.65794 | 0.36411 | 0.7731735043 | 0.90094 | 0.2342253327 | -0.0741801 | 3.252985463 | 0 |
| 10 | 87957903 | T | A | S | T | PTEN | ENST00000371953 | 0.36766 | 0.76514 | 0.43054 | 0.58144 | 0.43215 | 0.7171837557 | 0.82396 | 0.6013193727 | -0.0252003 | 2.319834002 | 0 |
| 10 | 87957903 | T | C | S | P | PTEN | ENST00000371953 | 0.42782 | 0.85437 | 0.44436 | 0.58035 | 0.47225 | 0.8519381065 | 0.62594 | 0.7172430158 | -0.060375 | -0.1497377083 | 0 |
| 10 | 87957903 | T | G | S | A | PTEN | ENST00000371953 | 0.3853 | 0.73286 | 0.40981 | 0.51542 | 0.44999 | 0.6459821644 | 0.74648 | 0.4695385098 | -0.0813388 | 2.750115021 | 0 |
| 10 | 87957904 | C | T | S | L | PTEN | ENST00000371953 | 0.45506 | 0.7547 | 0.45928 | 0.72714 | 0.67386 | 0.7797960912 | 0.75899 | 0.426916493 | -0.0529429 | 2.357344034 | 0 |
| 10 | 87957906 | G | A | G | R | PTEN | ENST00000371953 | 0.599 | 0.86559 | 0.63153 | 0.73914 | 0.69846 | 0.7782554154 | 0.82612 | 0.4576012302 | 0.0848899 | 3.223222927 | 0 |
| 10 | 87957907 | G | A | G | E | PTEN | ENST00000371953 | 0.49641 | 0.86419 | 0.60621 | 0.68245 | 0.66354 | 0.8346101102 | 0.7079 | 0.7426992059 | 0.0556327 | 3.10337264 | 0 |
| 10 | 87957907 | G | C | G | A | PTEN | ENST00000371953 | 0.42788 | 0.8133 | 0.50059 | 0.50089 | 0.57487 | 0.7244039779 | 0.6475 | 0.6347716451 | -0.0390388 | 0.533958781 | 0 |
| 10 | 87957907 | G | T | G | V | PTEN | ENST00000371953 | 0.46332 | 0.85419 | 0.51252 | 0.74779 | 0.68821 | 0.8219603134 | 0.76894 | 0.7841627598 | 0.0780458 | 2.167709269 | 0 |
| 10 | 87957909 | C | A | P | T | PTEN | ENST00000371953 | 0.3154 | 0.82462 | 0.48405 | 0.70592 | 0.62767 | 0.7733652797 | 0.69019 | 0.5662673116 | 0.0606987 | 2.264596629 | 0 |
| 10 | 87957909 | C | G | P | A | PTEN | ENST00000371953 | 0.2832 | 0.78365 | 0.4449 | 0.6606 | 0.58135 | 0.7188667964 | 0.61829 | 0.793043375 | -0.00284841 | 2.869186858 | 0 |
| 10 | 87957909 | C | T | P | S | PTEN | ENST00000371953 | 0.32068 | 0.80267 | 0.44796 | 0.6861 | 0.60362 | 0.7605201408 | 0.64757 | 0.6321691871 | 0.0425356 | 2.677605599 | 0 |
| 10 | 87957910 | C | A | P | H | PTEN | ENST00000371953 | 0.47917 | 0.83316 | 0.52304 | 0.63394 | 0.62547 | 0.7282578432 | 0.50193 | 0.816849947 | -0.0257071 | -0.2633784502 | 0 |
| 10 | 87957910 | C | G | P | R | PTEN | ENST00000371953 | 0.46655 | 0.8158 | 0.54278 | 0.68825 | 0.64673 | 0.7727856707 | 0.67104 | 0.7304550409 | 0.0720414 | 2.685760085 | 0 |
| 10 | 87957910 | C | T | P | L | PTEN | ENST00000371953 | 0.48818 | 0.7976 | 0.54698 | 0.74463 | 0.56426 | 0.7851645224 | 0.73745 | 0.7274839878 | 0.0414948 | 1.368937856 | 0 |
| 10 | 87957912 | A | C | T | P | PTEN | ENST00000371953 | 0.52068 | 0.86227 | 0.40327 | 0.62517 | 0.7285 | 0.8972850213 | 0.93336 | 0.8652070761 | 0.185833 | 2.922753543 | 0 |
| 10 | 87957912 | A | G | T | A | PTEN | ENST00000371953 | 0.46245 | 0.79808 | 0.41793 | 0.58819 | 0.45696 | 0.7319001166 | 0.90406 | 0.5515084267 | 0.084067 | 3.631374813 | 0 |
| 10 | 87957913 | C | A | T | K | PTEN | ENST00000371953 | 0.41926 | 0.84349 | 0.53325 | 0.6598 | 0.65673 | 0.8548821333 | 0.86918 | 0.8778352141 | 0.143237 | 4.667797698 | 0 |
| 10 | 87957913 | C | G | T | R | PTEN | ENST00000371953 | 0.44819 | 0.83073 | 0.51727 | 0.71754 | 0.78286 | 0.8496739694 | 0.86973 | 0.8388609886 | 0.169454 | 4.396366164 | 0 |
| 10 | 87957913 | C | T | T | I | PTEN | ENST00000371953 | 0.41022 | 0.81392 | 0.56023 | 0.66075 | 0.54769 | 0.7825615903 | 0.78822 | 0.8124408722 | 0.0839563 | 2.660440095 | 0 |
| 10 | 87957915 | C | G | R | G | PTEN | ENST00000371953 | 0.47507 | 0.86489 | 0.66559 | 0.83633 | 0.84159 | 0.9613510409 | 0.95632 | 0.9592823982 | 0.23338 | 4.427153978 | 0 |
| 10 | 87957916 | G | A | R | Q | PTEN | ENST00000371953 | 0.58945 | 0.87015 | 0.60082 | 0.79347 | 0.80238 | 0.8711597439 | 0.91314 | 0.7446089387 | 0.153696 | 4.539396529 | 0 |

| | | | | | | | | | | | | | | | | | | |
|---|---|---|---|---|---|---|---|---|---|---|---|---|---|---|---|---|---|---|
| 10 | 87957916 | G | C | R | P | PTEN | ENST00000371953 | 0.77402 | 0.88818 | 0.71662 | 0.82708 | 0.91399 | 0.9869620252 | 0.90879 | 0.9788703918 | 0.278384 | 4.558085296 | 0 |
| 10 | 87957916 | G | T | R | L | PTEN | ENST00000371953 | 0.59458 | 0.85232 | 0.70537 | 0.81648 | 0.85996 | 0.9416440572 | 0.96615 | 0.9690157771 | 0.257715 | 5.505305153 | 0 |
| 10 | 87957918 | C | G | R | G | PTEN | ENST00000371953 | 0.70776 | 0.87434 | 0.69531 | 0.90357 | 0.86149 | 0.9222932749 | 0.86819 | 0.9695243835 | 0.227264 | 4.64785933 | 0 |
| 10 | 87957918 | C | T | R | W | PTEN | ENST00000371953 | 0.8356 | 0.81811 | 0.78659 | 0.91609 | 0.86852 | 0.8398442188 | 0.81812 | 0.9906001091 | 0.216821 | 7.243283958 | 0 |
| 10 | 87957919 | G | A | R | Q | PTEN | ENST00000371953 | 0.80953 | 0.83701 | 0.6328 | 0.88562 | 0.89193 | 0.7890513556 | 0.97429 | 0.985021174 | 0.385857 | 5.588352888 | 0 |
| 10 | 87957919 | G | C | R | P | PTEN | ENST00000371953 | 0.9408 | 0.92524 | 0.73642 | 0.92273 | 0.94091 | 0.9592578575 | 0.85636 | 0.9867343903 | 0.291684 | 5.936052167 | 0 |
| 10 | 87957919 | G | T | R | L | PTEN | ENST00000371953 | 0.83921 | 0.87637 | 0.72654 | 0.91874 | 0.93162 | 0.8865521937 | 0.91387 | 0.9945994616 | 0.276188 | 6.007482384 | 0 |
| 10 | 87957921 | G | A | E | K | PTEN | ENST00000371953 | 0.54998 | 0.82507 | 0.81909 | 0.71519 | 0.76619 | 0.8904754788 | 0.90818 | 0.9148057699 | 0.138117 | 4.361983711 | 0 |
| 10 | 87957921 | G | C | E | Q | PTEN | ENST00000371953 | 0.54596 | 0.80262 | 0.70587 | 0.69155 | 0.72398 | 0.784318857 | 0.91567 | 0.8944188952 | 0.126661 | 4.288603002 | 0 |
| 10 | 87957922 | A | C | E | A | PTEN | ENST00000371953 | 0.49196 | 0.81168 | 0.71612 | 0.69238 | 0.64977 | 0.8351799143 | 0.90149 | 0.8352763057 | 0.127543 | 4.323735724 | 0 |
| 10 | 87957922 | A | G | E | G | PTEN | ENST00000371953 | 0.5497 | 0.83701 | 0.66757 | 0.75747 | 0.70589 | 0.875736079 | 0.956 | 0.887218833 | 0.162185 | 2.665305293 | 0 |
| 10 | 87957922 | A | T | E | V | PTEN | ENST00000371953 | 0.56623 | 0.8489 | 0.69413 | 0.79746 | 0.77205 | 0.8931494557 | 0.93426 | 0.9336206913 | 0.19415 | 4.71200097 | 0 |
| 10 | 87957924 | G | C | D | H | PTEN | ENST00000371953 | 0.71378 | 0.84827 | 0.80976 | 0.84366 | 0.80476 | 0.904430577 | 0.81364 | 0.9546401501 | 0.154769 | 4.738963735 | 0 |
| 10 | 87957924 | G | T | D | Y | PTEN | ENST00000371953 | 0.84635 | 0.87238 | 0.79048 | 0.86373 | 0.91357 | 0.9429448415 | 0.91921 | 0.9745308757 | 0.25905 | 5.128315327 | 0 |
| 10 | 87957925 | A | C | D | A | PTEN | ENST00000371953 | 0.57273 | 0.8545 | 0.71969 | 0.71496 | 0.73074 | 0.9200221146 | 0.77503 | 0.9268731475 | 0.11417 | 4.045642897 | 0 |
| 10 | 87957925 | A | G | D | G | PTEN | ENST00000371953 | 0.60061 | 0.86473 | 0.72958 | 0.76911 | 0.74517 | 0.9145211647 | 0.88151 | 0.9359555244 | 0.149297 | 4.06081829 | 0 |
| 10 | 87957925 | A | T | D | V | PTEN | ENST00000371953 | 0.60186 | 0.89093 | 0.71861 | 0.84723 | 0.84428 | 0.946499958 | 0.92118 | 0.9176187515 | 0.230443 | 4.091381693 | 0 |
| 10 | 87957926 | C | A | D | E | PTEN | ENST00000371953 | 0.35052 | 0.77184 | 0.75491 | 0.53498 | 0.56829 | 0.8045670424 | 0.74883 | 0.6264117956 | -0.0409909 | 1.847211839 | 0 |
| 10 | 87957927 | A | C | K | Q | PTEN | ENST00000371953 | 0.63397 | 0.82898 | 0.77309 | 0.81354 | 0.83779 | 0.8599653113 | 0.88537 | 0.9267325401 | 0.128103 | 4.829726232 | 1 |
| 10 | 87957927 | A | G | K | E | PTEN | ENST00000371953 | 0.49286 | 0.88559 | 0.81731 | 0.81858 | 0.83888 | 0.9205776657 | 0.94003 | 0.7689402922 | 0.255811 | 4.783726992 | 0 |
| 10 | 87957928 | A | C | K | T | PTEN | ENST00000371953 | 0.59082 | 0.84433 | 0.81704 | 0.88961 | 0.87198 | 0.9191563848 | 0.86693 | 0.9323012233 | 0.223465 | 4.205362128 | 0 |
| 10 | 87957928 | A | G | K | R | PTEN | ENST00000371953 | 0.7242 | 0.75216 | 0.73886 | 0.84258 | 0.78658 | 0.7672962609 | 0.88686 | 0.9190536737 | 0.111254 | 4.246257966 | 0 |
| 10 | 87957928 | A | T | K | M | PTEN | ENST00000371953 | 0.92198 | 0.97392 | 0.75093 | 0.9364 | 0.89467 | 0.8980302683 | 0.86984 | 0.9801871181 | 0.244027 | 7.451403609 | 0 |
| 10 | 87957929 | G | T | K | N | PTEN | ENST00000371953 | 0.511 | 0.84547 | 0.84191 | 0.82196 | 0.70752 | 0.9104652508 | 0.88682 | 0.8111476302 | 0.0239089 | 4.321078099 | 0 |
| 10 | 87957930 | T | A | F | I | PTEN | ENST00000371953 | 0.45248 | 0.87867 | 0.67901 | 0.72355 | 0.40224 | 0.8557958828 | 0.75309 | 0.3728739804 | -0.0185679 | 2.723057977 | 0 |
| 10 | 87957930 | T | G | F | V | PTEN | ENST00000371953 | 0.43774 | 0.86419 | 0.63067 | 0.69275 | 0.47888 | 0.8392622686 | 0.68491 | 0.3476144373 | -0.0141048 | 2.038983539 | 0 |
| 10 | 87957931 | T | A | F | Y | PTEN | ENST00000371953 | 0.34609 | 0.82819 | 0.69965 | 0.56631 | 0.45523 | 0.7322156248 | 0.58984 | 0.1943851858 | -0.0416118 | -0.1241237541 | 0 |
| 10 | 87957931 | T | C | F | S | PTEN | ENST00000371953 | 0.48685 | 0.92813 | 0.67093 | 0.71639 | 0.5108 | 0.8845398187 | 0.7747 | 0.3101680577 | -0.00988878 | 1.467490255 | 0 |
| 10 | 87957931 | T | G | F | C | PTEN | ENST00000371953 | 0.68235 | 0.8861 | 0.70337 | 0.75658 | 0.74446 | 0.9059706606 | 0.84397 | 0.6508636475 | 0.0529225 | 2.001674015 | 0 |
| 10 | 87957932 | C | G | F | L | PTEN | ENST00000371953 | 0.32199 | 0.83485 | 0.70895 | 0.5898 | 0.49511 | 0.7871770076 | 0.54595 | 0.09954319149 | -0.0778057 | 1.062834973 | 0 |
| 10 | 87957933 | A | G | M | V | PTEN | ENST00000371953 | 0.35085 | 0.79601 | 0.68733 | 0.75652 | 0.73296 | 0.8198867123 | 0.96917 | 0.757121861 | 0.242387 | 2.594328742 | 0 |
| 10 | 87957934 | T | A | M | K | PTEN | ENST00000371953 | 0.5108 | 0.90065 | 0.8345 | 0.89919 | 0.97698 | 0.9794865982 | 0.93275 | 0.9369813204 | 0.392838 | 5.154436169 | 0 |
| 10 | 87957934 | T | C | M | T | PTEN | ENST00000371953 | 0.42317 | 0.83141 | 0.87837 | 0.86373 | 0.90185 | 0.905364377 | 0.96821 | 0.5717546344 | 0.239746 | 4.544123965 | 0 |
| 10 | 87957934 | T | G | M | R | PTEN | ENST00000371953 | 0.64027 | 0.89636 | 0.81682 | 0.89779 | 0.97252 | 0.9838729206 | 0.94979 | 0.950838387 | 0.409242 | 4.649165685 | 0 |
| 10 | 87957936 | T | A | Y | N | PTEN | ENST00000371953 | 0.46059 | 0.91441 | 0.88802 | 0.75548 | 0.62101 | 0.8915593557 | 0.84034 | 0.4287872314 | 0.0837851 | 3.27147142 | 0 |
| 10 | 87957936 | T | C | Y | H | PTEN | ENST00000371953 | 0.55978 | 0.86526 | 0.89314 | 0.6018 | 0.61875 | 0.7792995903 | 0.71306 | 0.5322156549 | 0.0327314 | 2.570501856 | 0 |
| 10 | 87957937 | A | T | Y | F | PTEN | ENST00000371953 | 0.38283 | 0.7642 | 0.79633 | 0.43815 | 0.4919 | 0.6778075758 | 0.70559 | 0.3030012597 | -0.00824581 | 1.241406412 | 0 |
| 10 | 87957939 | T | A | F | I | PTEN | ENST00000371953 | 0.42229 | 0.88078 | 0.87387 | 0.83778 | 0.79151 | 0.9265595001 | 0.85496 | 0.8925760388 | 0.17927 | 4.163243114 | 0 |
| 10 | 87957939 | T | G | F | V | PTEN | ENST00000371953 | 0.46899 | 0.86551 | 0.83743 | 0.86942 | 0.79456 | 0.9185222562 | 0.96867 | 0.8558343053 | 0.34838 | 5.39431464 | 0 |
| 10 | 87957940 | T | A | F | Y | PTEN | ENST00000371953 | 0.71072 | 0.83822 | 0.87449 | 0.81865 | 0.89422 | 0.8631937789 | 0.85223 | 0.9287329912 | 0.216767 | 4.953765445 | 0 |
| 10 | 87957940 | T | C | F | S | PTEN | ENST00000371953 | 0.63145 | 0.92959 | 0.85558 | 0.91214 | 0.95717 | 0.9767215811 | 0.97757 | 0.9663241506 | 0.413248 | 6.557842187 | 1 |
| 10 | 87957941 | T | G | F | L | PTEN | ENST00000371953 | 0.34669 | 0.84458 | 0.88986 | 0.77389 | 0.8153 | 0.8970406405 | 0.89391 | 0.2251863033 | 0.160243 | 4.304017663 | 0 |
| 10 | 87957942 | G | C | E | Q | PTEN | ENST00000371953 | 0.6243 | 0.80896 | 0.8562 | 0.8338 | 0.74517 | 0.7840606087 | 0.88445 | 0.9127358198 | 0.118026 | 4.53999549 | 0 |
| 10 | 87957943 | A | C | E | A | PTEN | ENST00000371953 | 0.54277 | 0.8385 | 0.86964 | 0.83865 | 0.74376 | 0.8269916021 | 0.85007 | 0.9554544091 | 0.128903 | 4.528010717 | 0 |
| 10 | 87957943 | A | G | E | G | PTEN | ENST00000371953 | 0.61701 | 0.86159 | 0.83624 | 0.85279 | 0.85843 | 0.882945973 | 0.83244 | 0.9851306081 | 0.159021 | 4.872703844 | 0 |
| 10 | 87957943 | A | T | E | V | PTEN | ENST00000371953 | 0.57907 | 0.83916 | 0.859 | 0.82269 | 0.77335 | 0.8530687773 | 0.92637 | 0.9437974095 | 0.18293 | 3.683843797 | 0 |
| 10 | 87957945 | T | A | F | I | PTEN | ENST00000371953 | 0.51014 | 0.88511 | 0.94707 | 0.82661 | 0.79334 | 0.9434448678 | 0.82952 | 0.859582603 | 0.176438 | 2.86132259 | 0 |
| 10 | 87957945 | T | G | F | V | PTEN | ENST00000371953 | 0.4901 | 0.87188 | 0.93686 | 0.85428 | 0.80889 | 0.9426231502 | 0.96958 | 0.8280827403 | 0.349446 | 3.468296458 | 0 |
| 10 | 87957946 | T | A | F | Y | PTEN | ENST00000371953 | 0.53529 | 0.84119 | 0.95389 | 0.77017 | 0.75211 | 0.8917960246 | 0.95335 | 0.8422718048 | 0.222361 | 5.611667714 | 0 |
| 10 | 87957946 | T | G | F | C | PTEN | ENST00000371953 | 0.93223 | 0.98311 | 0.95983 | 0.92857 | 0.94205 | 0.9770955473 | 0.97306 | 0.9604059458 | 0.420459 | 5.314746461 | 0 |
| 10 | 87957947 | C | G | F | L | PTEN | ENST00000371953 | 0.28101 | 0.83597 | 0.9624 | 0.72299 | 0.74093 | 0.9042980583 | 0.95973 | 0.2011914998 | 0.169442 | 2.035525145 | 0 |
| 10 | 87957948 | C | G | P | A | PTEN | ENST00000371953 | 0.48806 | 0.82763 | 0.80814 | 0.88724 | 0.9248 | 0.8355301943 | 0.82318 | 0.9180243611 | 0.242471 | 5.40087539 | 0 |
| 10 | 87957949 | C | A | P | H | PTEN | ENST00000371953 | 0.82155 | 0.86723 | 0.86502 | 0.9058 | 0.93123 | 0.8527674533 | 0.86097 | 0.974288106 | 0.298108 | 5.003757907 | 1 |
| 10 | 87957949 | C | G | P | R | PTEN | ENST00000371953 | 0.68923 | 0.88315 | 0.87442 | 0.89238 | 0.95462 | 0.895852035 | 0.92263 | 0.9566039443 | 0.391313 | 5.474820042 | 0 |
| 10 | 87957949 | C | T | P | L | PTEN | ENST00000371953 | 0.69708 | 0.80914 | 0.88086 | 0.90853 | 0.94281 | 0.8873987956 | 0.87873 | 0.9875740409 | 0.272884 | 4.386761071 | 0 |
| 10 | 87957951 | C | A | Q | K | PTEN | ENST00000371953 | 0.49049 | 0.8648 | 0.87795 | 0.89689 | 0.86603 | 0.86156271 | 0.94148 | 0.8101456761 | 0.238514 | 4.235298728 | 0 |
| 10 | 87957952 | A | C | Q | P | PTEN | ENST00000371953 | 0.55154 | 0.84874 | 0.83967 | 0.80664 | 0.8306 | 0.9551826503 | 0.97099 | 0.8892106414 | 0.325738 | 4.206431975 | 1 |
| 10 | 87957952 | A | G | Q | R | PTEN | ENST00000371953 | 0.68827 | 0.82305 | 0.87248 | 0.85504 | 0.90972 | 0.8531054495 | 0.98051 | 0.8588424325 | 0.357552 | 4.147343964 | 0 |
| 10 | 87957952 | A | T | Q | L | PTEN | ENST00000371953 | 0.53995 | 0.78843 | 0.82412 | 0.81242 | 0.83943 | 0.8647471791 | 0.97858 | 0.9058896303 | 0.274334 | 5.205495909 | 0 |
| 10 | 87957953 | G | T | Q | H | PTEN | ENST00000371953 | 0.58347 | 0.79045 | 0.87068 | 0.80722 | 0.86553 | 0.874954862 | 0.92446 | 0.5704116821 | 0.206914 | 4.791880894 | 0 |
| 10 | 87957954 | C | A | P | T | PTEN | ENST00000371953 | 0.51113 | 0.85949 | 0.92037 | 0.90176 | 0.95133 | 0.902075069 | 0.90252 | 0.9783747792 | 0.319684 | 4.691259715 | 0 |
| 10 | 87957954 | C | G | P | A | PTEN | ENST00000371953 | 0.48088 | 0.8138 | 0.88562 | 0.8732 | 0.91986 | 0.8556045893 | 0.82822 | 0.9611526728 | 0.218562 | 4.898972889 | 0 |
| 10 | 87957954 | C | T | P | S | PTEN | ENST00000371953 | 0.52646 | 0.81302 | 0.89454 | 0.87711 | 0.88448 | 0.8992299182 | 0.83669 | 0.9638381004 | 0.206424 | 3.622093376 | 0 |
| 10 | 87957955 | C | G | P | R | PTEN | ENST00000371953 | 0.67581 | 0.88976 | 0.9404 | 0.89725 | 0.94129 | 0.912065927 | 0.91924 | 0.991084218 | 0.375448 | 5.516599786 | 0 |
| 10 | 87957957 | T | A | L | I | PTEN | ENST00000371953 | 0.48614 | 0.87572 | 0.88396 | 0.8163 | 0.81646 | 0.7631980148 | 0.81343 | 0.5034524202 | 0.150089 | 3.763700497 | 0 |
| 10 | 87957959 | A | T | L | F | PTEN | ENST00000371953 | 0.71954 | 0.94463 | 0.87578 | 0.93266 | 0.89648 | 0.8828144871 | 0.89648 | 0.9889274836 | 0.265524 | 5.056155916 | 0 |
| 10 | 87957960 | C | A | P | T | PTEN | ENST00000371953 | 0.56006 | 0.83939 | 0.95266 | 0.91715 | 0.96371 | 0.9404682311 | 0.89313 | 0.9973840117 | 0.410527 | 5.650060829 | 0 |
| 10 | 87957960 | C | G | P | A | PTEN | ENST00000371953 | 0.52699 | 0.80193 | 0.93174 | 0.87592 | 0.90755 | 0.909581233 | 0.83514 | 0.9729361534 | 0.238722 | 5.018046574 | 0 |
| 10 | 87957960 | C | T | P | S | PTEN | ENST00000371953 | 0.58681 | 0.87705 | 0.93649 | 0.93093 | 0.90842 | 0.9430770848 | 0.83843 | 0.9767465591 | 0.252145 | 5.311175934 | 0 |
| 10 | 87957961 | C | A | P | H | PTEN | ENST00000371953 | 0.67891 | 0.97295 | 0.96861 | 0.94909 | 0.94091 | 0.9363753007 | 0.8545 | 0.9970280528 | 0.377716 | 5.824738882 | 0 |
| 10 | 87957961 | C | G | P | R | PTEN | ENST00000371953 | 0.81119 | 0.86419 | 0.97396 | 0.94168 | 0.9822 | 0.9641512018 | 0.88832 | 0.9680835009 | 0.374588 | 5.479897287 | 0 |
| 10 | 87957961 | C | T | P | L | PTEN | ENST00000371953 | 0.69997 | 0.80789 | 0.97595 | 0.9145 | 0.94765 | 0.9527503689 | 0.86755 | 0.9915677905 | 0.366333 | 4.395122606 | 0 |
| 10 | 87957963 | G | A | V | M | PTEN | ENST00000371953 | 0.84294 | 0.92554 | 0.96818 | 0.93906 | 0.88354 | 0.8799216885 | 0.85273 | 0.9766659141 | 0.269112 | 5.189470618 | 0 |
| 10 | 87957963 | G | T | V | L | PTEN | ENST00000371953 | 0.41712 | 0.79509 | 0.95445 | 0.80452 | 0.7609 | 0.8872035499 | 0.92054 | 0.8171289563 | 0.219568 | 2.783512465 | 0 |
| 10 | 87957964 | T | A | V | E | PTEN | ENST00000371953 | 0.9236 | 0.97392 | 0.95651 | 0.95487 | 0.99386 | 0.990672176 | 0.97042 | 0.9981964231 | 0.458739 | 7.073971844 | 1 |
| 10 | 87957964 | T | C | V | A | PTEN | ENST00000371953 | 0.74698 | 0.96308 | 0.96094 | 0.91642 | 0.95499 | 0.9298056359 | 0.94438 | 0.9824039936 | 0.354631 | 5.866555949 | 0 |
| 10 | 87957964 | T | G | V | G | PTEN | ENST00000371953 | 0.92128 | 0.98215 | 0.87616 | 0.95839 | 0.99386 | 0.9913165778 | 0.9768 | 0.9976415634 | 0.459077 | 7.20137969 | 1 |
| 10 | 87957966 | T | G | C | G | PTEN | ENST00000371953 | 0.70182 | 0.85361 | 0.99117 | 0.90952 | 0.98404 | 0.946520408 | 0.92926 | 0.9831871986 | 0.404943 | 6.383025218 | 0 |
| 10 | 87957967 | G | A | C | Y | PTEN | ENST00000371953 | 0.77029 | 0.91936 | 0.9993 | 0.87941 | 0.95353 | 0.9608410582 | 0.94083 | 0.9914938211 | 0.412042 | 5.877013941 | 0 |
| 10 | 87957967 | G | C | C | S | PTEN | ENST00000371953 | 0.73757 | 0.85482 | 0.99477 | 0.86627 | 0.94394 | 0.9166239742 | 0.87905 | 0.9408074617 | 0.350241 | 4.670456667 | 0 |
| 10 | 87957967 | G | T | C | F | PTEN | ENST00000371953 | 0.81442 | 0.91315 | 0.99574 | 0.9215 | 0.97661 | 0.9440190098 | 0.95636 | 0.9930005074 | 0.426753 | 6.450528989 | 0 |
| 10 | 87957968 | T | G | C | W | PTEN | ENST00000371953 | 0.72716 | 0.97717 | 0.99827 | 0.94266 | 0.90229 | 0.9591233833 | 0.95528 | 0.9859676957 | 0.370723 | 7.653923618 | 0 |
| 10 | 87957969 | G | A | G | S | PTEN | ENST00000371953 | 0.87275 | 0.9469 | 0.99117 | 0.97772 | 0.99722 | 0.9561054336 | 0.98295 | 0.9948439598 | 0.545027 | 4.269999781 | 0 |
| 10 | 87957969 | G | T | G | C | PTEN | ENST00000371953 | 0.92284 | 0.9727 | 0.99661 | 0.97933 | 0.99806 | 0.9846169007 | 0.98315 | 0.9990122318 | 0.562666 | 7.077111917 | 1 |
| 10 | 87957970 | G | C | G | A | PTEN | ENST00000371953 | 0.90292 | 0.97175 | 0.99197 | 0.96727 | 0.99806 | 0.9708016337 | 0.98531 | 0.9940663576 | 0.53746 | 4.805705207 | 0 |
| 10 | 87957972 | G | T | D | Y | PTEN | ENST00000371953 | 0.90031 | 0.98069 | 0.98999 | 0.98787 | 0.99722 | 0.9961272959 | 0.98262 | 0.9984161854 | 0.574307 | 7.657062146 | 1 |
| 10 | 87957973 | A | T | D | V | PTEN | ENST00000371953 | 0.92947 | 0.98725 | 0.97322 | 0.99396 | 0.99443 | 0.9940087049 | 0.99022 | 0.9990087748 | 0.572157 | 7.330310975 | 1 |
| 10 | 87957974 | T | A | D | E | PTEN | ENST00000371953 | 0.57125 | 0.96308 | 0.98434 | 0.98557 | 0.95608 | 0.9811113324 | 0.99363 | 0.9964603782 | 0.50133 | 5.072229308 | 0 |
| 10 | 87957975 | A | C | I | L | PTEN | ENST00000371953 | 0.80416 | 0.873 | 0.95107 | 0.90652 | 0.93201 | 0.9250346885 | 0.87472 | 0.9743922353 | 0.253417 | 4.990058127 | 0 |
| 10 | 87957975 | A | G | I | V | PTEN | ENST00000371953 | 0.59429 | 0.77385 | 0.89553 | 0.7181 | 0.78906 | 0.7790141719 | 0.73616 | 0.8676537871 | 0.0753301 | 3.191401431 | 0 |
| 10 | 87957975 | A | T | I | F | PTEN | ENST00000371953 | 0.82108 | 0.9232 | 0.95617 | 0.93243 | 0.97512 | 0.9757021665 | 0.93797 | 0.9908179045 | 0.409581 | 4.875428592 | 0 |
| 10 | 87957976 | T | A | I | N | PTEN | ENST00000371953 | 0.9428 | 0.99368 | 0.9858 | 0.96936 | 0.97997 | 0.9943015597 | 0.89654 | 0.998054266 | 0.317054 | 8.296672633 | 1 |
| 10 | 87957977 | C | G | I | M | PTEN | ENST00000371953 | 0.73205 | 0.94115 | 0.92916 | 0.9246 | 0.90007 | 0.9305268663 | 0.88123 | 0.9803738594 | 0.256227 | 5.279305988 | 0 |
| 10 | 87957978 | A | C | K | Q | PTEN | ENST00000371953 | 0.83352 | 0.96209 | 0.98015 | 0.93928 | 0.95717 | 0.9467252797 | 0.92408 | 0.9793586135 | 0.268526 | 5.669638977 | 0 |
| 10 | 87957978 | A | G | K | E | PTEN | ENST00000371953 | 0.85441 | 0.9799 | 0.98873 | 0.94348 | 0.9579 | 0.9755838457 | 0.96275 | 0.9836227298 | 0.376908 | 7.3479769 | 0 |
| 10 | 87957979 | A | C | K | T | PTEN | ENST00000371953 | 0.92135 | 0.97855 | 0.98993 | 0.95577 | 0.98367 | 0.9512522043 | 0.97252 | 0.9982856619 | 0.448415 | 6.624583774 | 0 |
| 10 | 87957979 | A | G | K | R | PTEN | ENST00000371953 | 0.60164 | 0.83431 | 0.96949 | 0.80578 | 0.91694 | 0.9022440259 | 0.92085 | 0.9013283253 | 0.221981 | 4.249117725 | 0 |
| 10 | 87957979 | A | T | K | I | PTEN | ENST00000371953 | 0.93983 | 0.99351 | 0.98323 | 0.96076 | 0.9781 | 0.9605636407 | 0.93481 | 0.9974381924 | 0.399949 | 6.586347092 | 0 |
| 10 | 87957980 | A | T | K | N | PTEN | ENST00000371953 | 0.88696 | 0.97758 | 0.99277 | 0.94718 | 0.93397 | 0.9704395932 | 0.92696 | 0.9925090671 | 0.13918 | 6.397790629 | 0 |
| 10 | 87957981 | G | A | V | I | PTEN | ENST00000371953 | 0.44661 | 0.78954 | 0.88315 | 0.77926 | 0.62101 | 0.7412899791 | 0.83223 | 0.8746429682 | 0.110053 | 3.244309898 | 0 |
| 10 | 87957981 | G | C | V | L | PTEN | ENST00000371953 | 0.61918 | 0.86457 | 0.96768 | 0.89024 | 0.87198 | 0.9064951993 | 0.95345 | 0.9248183966 | 0.272748 | 4.649500585 | 0 |

| | | | | | | | | | | | | | | | | |
|---|---|---|---|---|---|---|---|---|---|---|---|---|---|---|---|---|
| 10 | 87957981 | G | T | V | L | PTEN | ENST00000371953 | 0.61306 | 0.86457 | 0.96768 | 0.89077 | 0.87198 | 0.9064951993 | 0.95324 | 0.9402778745 | 0.272746 | 4.649500585 | 0 |
| 10 | 87957982 | T | A | V | E | PTEN | ENST00000371953 | 0.92287 | 0.99114 | 0.96974 | 0.96134 | 0.99202 | 0.9940265522 | 0.97829 | 0.9972637892 | 0.463569 | 8.388180045 | 1 |
| 10 | 87957982 | T | G | V | G | PTEN | ENST00000371953 | 0.91036 | 0.98196 | 0.91464 | 0.96323 | 0.99006 | 0.9937291667 | 0.98451 | 0.9939960241 | 0.462915 | 7.257833748 | 0 |
| 10 | 87957985 | A | G | E | G | PTEN | ENST00000371953 | 0.93748 | 0.96763 | 0.96968 | 0.95104 | 0.97475 | 0.9608681872 | 0.96241 | 0.9951044321 | 0.437091 | 8.395570037 | 0 |
| 10 | 87957985 | A | T | E | V | PTEN | ENST00000371953 | 0.93226 | 0.915 | 0.97671 | 0.93231 | 0.94728 | 0.9310234498 | 0.93293 | 0.9964818954 | 0.331189 | 5.277933795 | 0 |
| 10 | 87957987 | T | G | F | V | PTEN | ENST00000371953 | 0.52181 | 0.91893 | 0.96268 | 0.90652 | 0.97252 | 0.9295651523 | 0.96393 | 0.9816930294 | 0.38419 | 4.335384884 | 0 |
| 10 | 87957988 | T | A | F | Y | PTEN | ENST00000371953 | 0.92416 | 0.97582 | 0.97366 | 0.92138 | 0.93201 | 0.884311192 | 0.8874 | 0.9728689194 | 0.26433 | 6.918143865 | 0 |
| 10 | 87957988 | T | G | F | C | PTEN | ENST00000371953 | 0.95034 | 0.96895 | 0.98082 | 0.95546 | 0.9844 | 0.9749544406 | 0.93329 | 0.9931510687 | 0.388803 | 4.984423712 | 0 |
| 10 | 87957990 | T | A | F | I | PTEN | ENST00000371953 | 0.83575 | 0.90435 | 0.98748 | 0.86834 | 0.9252 | 0.9669463775 | 0.83862 | 0.9815481901 | 0.233199 | 6.002075846 | 0 |
| 10 | 87957990 | T | G | F | V | PTEN | ENST00000371953 | 0.83813 | 0.87728 | 0.98519 | 0.90121 | 0.97997 | 0.9606619521 | 0.95824 | 0.9724411368 | 0.404332 | 5.320836389 | 0 |
| 10 | 87957991 | T | A | F | Y | PTEN | ENST00000371953 | 0.56124 | 0.83047 | 0.98997 | 0.7807 | 0.80118 | 0.8904221574 | 0.88639 | 0.854378283 | 0.182116 | 3.495643887 | 0 |
| 10 | 87957992 | C | G | F | L | PTEN | ENST00000371953 | 0.64167 | 0.83761 | 0.99282 | 0.89634 | 0.87345 | 0.9401766708 | 0.83295 | 0.979092598 | 0.209766 | 5.450502302 | 0 |
| 10 | 87957993 | C | G | H | D | PTEN | ENST00000371953 | 0.62993 | 0.88228 | 0.96993 | 0.8437 | 0.89558 | 0.9888054572 | 0.96435 | 0.9767342806 | 0.39499 | 5.218160402 | 0 |
| 10 | 87957994 | A | C | H | P | PTEN | ENST00000371953 | 0.86629 | 0.91486 | 0.94948 | 0.84855 | 0.97735 | 0.9937466696 | 0.97726 | 0.9899194241 | 0.432149 | 5.962316501 | 1 |
| 10 | 87957994 | A | G | H | R | PTEN | ENST00000371953 | 0.66842 | 0.8613 | 0.9733 | 0.81018 | 0.9681 | 0.9712922819 | 0.97793 | 0.9821910858 | 0.406753 | 4.646175019 | 0 |
| 10 | 87957994 | A | T | H | L | PTEN | ENST00000371953 | 0.72284 | 0.87129 | 0.95989 | 0.88336 | 0.95717 | 0.9790050114 | 0.96466 | 0.9961909056 | 0.410729 | 5.425939204 | 0 |
| 10 | 87957995 | C | A | H | Q | PTEN | ENST00000371953 | 0.4322 | 0.81243 | 0.96422 | 0.72527 | 0.91186 | 0.9573376657 | 0.97138 | 0.9544219375 | 0.339942 | 4.753993255 | 0 |
| 10 | 87957996 | A | G | K | E | PTEN | ENST00000371953 | 0.62688 | 0.88897 | 0.93946 | 0.8245 | 0.95535 | 0.9774076678 | 0.98227 | 0.9812945127 | 0.393494 | 6.11165074 | 0 |
| 10 | 87957997 | A | C | K | T | PTEN | ENST00000371953 | 0.8861 | 0.95535 | 0.94885 | 0.93103 | 0.93083 | 0.9711419022 | 0.92253 | 0.9928624034 | 0.284319 | 5.701880907 | 0 |
| 10 | 87957997 | A | G | K | R | PTEN | ENST00000371953 | 0.73285 | 0.78596 | 0.89093 | 0.75981 | 0.89603 | 0.8959321549 | 0.9446 | 0.8555200696 | 0.253873 | 4.567255006 | 0 |
| 10 | 87957997 | A | T | K | I | PTEN | ENST00000371953 | 0.91826 | 0.9856 | 0.92116 | 0.93864 | 0.92601 | 0.9748413253 | 0.88573 | 0.9954890153 | 0.280434 | 6.292220831 | 0 |
| 10 | 87957998 | A | T | K | N | PTEN | ENST00000371953 | 0.79263 | 0.87359 | 0.9563 | 0.8179 | 0.81472 | 0.972580452 | 0.90284 | 0.9756788611 | 0.0689803 | 5.231762778 | 0 |
| 10 | 87957999 | C | A | Q | K | PTEN | ENST00000371953 | 0.31629 | 0.8599 | 0.97068 | 0.83491 | 0.75687 | 0.8144886321 | 0.88555 | 0.9184906483 | 0.176944 | 3.617156661 | 0 |
| 10 | 87957999 | C | G | Q | E | PTEN | ENST00000371953 | 0.27755 | 0.82465 | 0.95157 | 0.83286 | 0.67108 | 0.8341423908 | 0.88447 | 0.8244607449 | 0.109082 | 4.674624664 | 0 |
| 10 | 87958000 | A | C | Q | P | PTEN | ENST00000371953 | 0.55367 | 0.84718 | 0.95243 | 0.82268 | 0.7675 | 0.9528726144 | 0.97064 | 0.9749709368 | 0.238659 | 3.29054105 | 0 |
| 10 | 87958000 | A | G | Q | R | PTEN | ENST00000371953 | 0.57772 | 0.81682 | 0.96918 | 0.83666 | 0.85945 | 0.7752282391 | 0.9164 | 0.5273552311 | 0.219713 | 5.615586961 | 0 |
| 10 | 87958000 | A | T | Q | L | PTEN | ENST00000371953 | 0.61873 | 0.80517 | 0.94498 | 0.89466 | 0.81703 | 0.853959832 | 0.87183 | 0.9358674884 | 0.154252 | 5.940780243 | 0 |
| 10 | 87958002 | A | C | N | H | PTEN | ENST00000371953 | 0.77577 | 0.81991 | 0.96584 | 0.87317 | 0.81876 | 0.8573276487 | 0.87397 | 0.8784519434 | 0.124451 | 4.910234727 | 0 |
| 10 | 87958003 | A | C | N | T | PTEN | ENST00000371953 | 0.60288 | 0.82747 | 0.96035 | 0.86253 | 0.74235 | 0.8467717376 | 0.86677 | 0.887375176 | 0.113522 | 4.711699053 | 0 |
| 10 | 87958003 | A | G | N | S | PTEN | ENST00000371953 | 0.53213 | 0.72049 | 0.94317 | 0.74026 | 0.5377 | 0.7782190323 | 0.71772 | 0.4299001669 | -0.0894229 | 3.391570518 | 0 |
| 10 | 87958003 | A | T | N | I | PTEN | ENST00000371953 | 0.81024 | 0.85771 | 0.96626 | 0.9163 | 0.93358 | 0.9483412968 | 0.9057 | 0.9590728879 | 0.245851 | 5.128686288 | 0 |
| 10 | 87958005 | A | C | K | Q | PTEN | ENST00000371953 | 0.69019 | 0.95796 | 0.92326 | 0.87114 | 0.79878 | 0.8545774079 | 0.88177 | 0.9223773479 | 0.140883 | 6.169147377 | 0 |
| 10 | 87958006 | A | C | K | T | PTEN | ENST00000371953 | 0.64035 | 0.86428 | 0.95185 | 0.89289 | 0.82046 | 0.8991564085 | 0.92126 | 0.8596379161 | 0.238051 | 5.589204049 | 0 |
| 10 | 87958006 | A | G | K | R | PTEN | ENST00000371953 | 0.77298 | 0.80347 | 0.90503 | 0.85653 | 0.78411 | 0.7505125097 | 0.85988 | 0.7614589146 | 0.128494 | 3.909795931 | 0 |
| 10 | 87958006 | A | T | K | M | PTEN | ENST00000371953 | 0.66541 | 0.92482 | 0.90829 | 0.89816 | 0.82159 | 0.888604794 | 0.88738 | 0.8733427525 | 0.210263 | 4.08574633 | 0 |
| 10 | 87958007 | G | T | K | N | PTEN | ENST00000371953 | 0.93449 | 0.88476 | 0.95765 | 0.88157 | 0.79637 | 0.9086718765 | 0.86831 | 0.9576888084 | 0.0965197 | 4.645638823 | 0 |
| 10 | 87958008 | A | T | M | L | PTEN | ENST00000371953 | 0.41351 | 0.77066 | 0.96773 | 0.61079 | 0.62876 | 0.6859396471 | 0.89595 | 0.3820599318 | 0.108194 | 3.311826891 | 0 |
| 10 | 87958009 | T | A | M | K | PTEN | ENST00000371953 | 0.62023 | 0.90159 | 0.98507 | 0.89476 | 0.85012 | 0.910954756 | 0.91185 | 0.8498818874 | 0.239903 | 3.361840456 | 0 |
| 10 | 87958009 | T | C | M | T | PTEN | ENST00000371953 | 0.58183 | 0.83561 | 0.99107 | 0.89992 | 0.84428 | 0.8408529195 | 0.89226 | 0.8786407113 | 0.249283 | 4.154035806 | 1 |
| 10 | 87958009 | T | G | M | R | PTEN | ENST00000371953 | 0.77172 | 0.89914 | 0.98184 | 0.89591 | 0.85638 | 0.9252470241 | 0.92606 | 0.9105671644 | 0.282629 | 4.537688448 | 1 |
| 10 | 87958010 | G | T | M | I | PTEN | ENST00000371953 | 0.59089 | 0.82205 | 0.96525 | 0.86316 | 0.62436 | 0.7408208137 | 0.74966 | 0.7882491921 | 0.114004 | 3.458473869 | 0 |
| 10 | 87958011 | C | A | L | I | PTEN | ENST00000371953 | 0.3772 | 0.81631 | 0.96707 | 0.57491 | 0.51539 | 0.6374221857 | 0.60923 | 0.3786233664 | -0.0407751 | 2.262345577 | 0 |
| 10 | 87958011 | C | G | L | V | PTEN | ENST00000371953 | 0.38513 | 0.79874 | 0.94491 | 0.6042 | 0.4983 | 0.7152850055 | 0.70208 | 0.3240306675 | -0.0340354 | 3.445657015 | 0 |
| 10 | 87958012 | T | A | L | Q | PTEN | ENST00000371953 | 0.63981 | 0.83394 | 0.97076 | 0.80125 | 0.67661 | 0.8343914399 | 0.67818 | 0.7554320693 | 0.0831474 | 3.695337661 | 0 |
| 10 | 87958012 | T | C | L | P | PTEN | ENST00000371953 | 0.80203 | 0.97491 | 0.99098 | 0.88313 | 0.78844 | 0.9167028271 | 0.69056 | 0.9129371643 | 0.1131 | 3.421187227 | 0 |
| 10 | 87958012 | T | G | L | R | PTEN | ENST00000371953 | 0.64594 | 0.9147 | 0.98807 | 0.8011 | 0.68908 | 0.8792442638 | 0.69889 | 0.599613387 | 0.0862753 | 3.688580406 | 0 |
| 10 | 87958014 | A | C | K | Q | PTEN | ENST00000371953 | 0.81111 | 0.88762 | 0.92918 | 0.87706 | 0.86803 | 0.8344780598 | 0.88597 | 0.9433214068 | 0.124826 | 5.838975165 | 0 |
| 10 | 87958014 | A | G | K | E | PTEN | ENST00000371953 | 0.82131 | 0.93098 | 0.94937 | 0.87657 | 0.92275 | 0.896843398 | 0.9532 | 0.9541531205 | 0.300697 | 6.536056165 | 0 |
| 10 | 87958015 | A | G | K | R | PTEN | ENST00000371953 | 0.53812 | 0.74065 | 0.91036 | 0.73091 | 0.64876 | 0.7117756501 | 0.81548 | 0.7727690935 | 0.0783487 | 4.073666767 | 0 |
| 10 | 87958015 | A | T | K | I | PTEN | ENST00000371953 | 0.93195 | 0.98069 | 0.93573 | 0.92414 | 0.91694 | 0.9164907119 | 0.84792 | 0.9882909656 | 0.208566 | 6.112886556 | 0 |
| 10 | 87958016 | A | T | K | N | PTEN | ENST00000371953 | 0.71064 | 0.85631 | 0.96151 | 0.86226 | 0.8199 | 0.8871112014 | 0.84788 | 0.9448004365 | 0.0183521 | 5.089490122 | 0 |
| 10 | 87958017 | A | C | K | Q | PTEN | ENST00000371953 | 0.60098 | 0.88868 | 0.91448 | 0.87179 | 0.7609 | 0.8606688046 | 0.85824 | 0.9630497098 | 0.0995105 | 5.651285629 | 0 |
| 10 | 87958017 | A | G | K | E | PTEN | ENST00000371953 | 0.65109 | 0.92349 | 0.93956 | 0.87895 | 0.92151 | 0.9145395491 | 0.95406 | 0.9836697578 | 0.280774 | 5.567048132 | 0 |
| 10 | 87958018 | A | C | K | T | PTEN | ENST00000371953 | 0.79358 | 0.97362 | 0.94354 | 0.92748 | 0.93554 | 0.8949735118 | 0.88827 | 0.9949658513 | 0.240913 | 4.88890864 | 0 |
| 10 | 87958018 | A | G | K | R | PTEN | ENST00000371953 | 0.48646 | 0.80666 | 0.89499 | 0.81506 | 0.70179 | 0.7489024554 | 0.84954 | 0.9500575066 | 0.0529065 | 4.028529593 | 0 |
| 10 | 87958018 | A | T | K | M | PTEN | ENST00000371953 | 0.85055 | 0.97628 | 0.89819 | 0.94234 | 0.93044 | 0.8967554011 | 0.88431 | 0.998583436 | 0.240478 | 5.941804102 | 0 |
| 10 | 87960894 | G | A | D | N | PTEN | ENST00000371953 | 0.85749 | 0.79213 | 0.89403 | 0.86955 | 0.77141 | 0.7575776563 | 0.8592 | 0.9841326475 | -0.0475822 | 4.022898171 | 0 |
| 10 | 87960894 | G | C | D | H | PTEN | ENST00000371953 | 0.88379 | 0.95887 | 0.91433 | 0.8903 | 0.89376 | 0.8076181941 | 0.82357 | 0.9890329838 | 0.137562 | 4.887345665 | 0 |
| 10 | 87960894 | G | T | D | Y | PTEN | ENST00000371953 | 0.89382 | 0.95605 | 0.90133 | 0.89286 | 0.91736 | 0.8922318503 | 0.86944 | 0.996339798 | 0.241668 | 5.245992895 | 0 |
| 10 | 87960895 | A | G | D | G | PTEN | ENST00000371953 | 0.88758 | 0.86686 | 0.85401 | 0.88121 | 0.86352 | 0.9035179016 | 0.8348 | 0.99309057 | 0.128762 | 4.095961317 | 0 |
| 10 | 87960896 | C | A | D | E | PTEN | ENST00000371953 | 0.21729 | 0.77148 | 0.87229 | 0.63719 | 0.54058 | 0.7221042152 | 0.57618 | 0.003510455237 | -0.151575 | 0.7659868341 | 0 |
| 10 | 87960897 | A | G | K | E | PTEN | ENST00000371953 | 0.86734 | 0.98348 | 0.91909 | 0.9354 | 0.89739 | 0.9737237095 | 0.90318 | 0.9757905602 | 0.141151 | 5.563459181 | 0 |
| 10 | 87960898 | A | C | K | T | PTEN | ENST00000371953 | 0.93429 | 0.95189 | 0.92316 | 0.94544 | 0.94281 | 0.9544408873 | 0.88705 | 0.9955189228 | 0.254578 | 5.902354152 | 0 |
| 10 | 87960898 | A | G | K | R | PTEN | ENST00000371953 | 0.82155 | 0.79795 | 0.86705 | 0.83011 | 0.8873 | 0.8952384252 | 0.86626 | 0.9576516151 | 0.107747 | 3.534793635 | 0 |
| 10 | 87960898 | A | T | K | I | PTEN | ENST00000371953 | 0.94673 | 0.98638 | 0.89719 | 0.95134 | 0.92601 | 0.9670642438 | 0.86707 | 0.9987721048 | 0.260793 | 5.600800356 | 0 |
| 10 | 87960899 | A | T | K | N | PTEN | ENST00000371953 | 0.91725 | 0.91337 | 0.9348 | 0.91588 | 0.88684 | 0.9698417099 | 0.90372 | 0.9705690742 | 0.105676 | 5.595277844 | 0 |
| 10 | 87960900 | A | G | M | V | PTEN | ENST00000371953 | 0.43938 | 0.84646 | 0.89678 | 0.86638 | 0.89829 | 0.9101187716 | 0.94293 | 0.871607542 | 0.4018 | 4.03624767 | 0 |
| 10 | 87960900 | A | T | M | L | PTEN | ENST00000371953 | 0.43853 | 0.78684 | 0.93711 | 0.81286 | 0.8543 | 0.8675547074 | 0.91294 | 0.7142344818 | 0.263897 | 3.239644073 | 0 |
| 10 | 87960901 | T | A | M | K | PTEN | ENST00000371953 | 0.84635 | 0.97099 | 0.97028 | 0.95244 | 0.9917 | 0.9870926435 | 0.97007 | 0.9980102806 | 0.446791 | 5.866040445 | 0 |
| 10 | 87960901 | T | G | M | R | PTEN | ENST00000371953 | 0.86762 | 0.95873 | 0.96436 | 0.93033 | 0.98938 | 0.9904877543 | 0.972 | 0.9940741658 | 0.433856 | 6.148727412 | 0 |
| 10 | 87960902 | G | T | M | I | PTEN | ENST00000371953 | 0.55312 | 0.84398 | 0.93397 | 0.88399 | 0.93201 | 0.8883984901 | 0.9161 | 0.3260285556 | 0.377743 | 3.638159323 | 0 |
| 10 | 87960903 | T | A | F | I | PTEN | ENST00000371953 | 0.87536 | 0.98388 | 0.96477 | 0.96172 | 0.99471 | 0.9664018983 | 0.93454 | 0.9912616014 | 0.487659 | 6.266100219 | 0 |
| 10 | 87960903 | T | G | F | V | PTEN | ENST00000371953 | 0.87814 | 0.96506 | 0.95785 | 0.9745 | 0.99578 | 0.956570867 | 0.94446 | 0.9885496117 | 0.479312 | 5.706129577 | 0 |
| 10 | 87960904 | T | A | F | Y | PTEN | ENST00000371953 | 0.93651 | 0.97582 | 0.96921 | 0.93242 | 0.98183 | 0.8922767439 | 0.91621 | 0.9556564402 | 0.395952 | 5.998972774 | 0 |
| 10 | 87960904 | T | C | F | S | PTEN | ENST00000371953 | 0.95307 | 0.99114 | 0.96307 | 0.97523 | 0.99603 | 0.9857264192 | 0.98109 | 0.9961494211 | 0.502971 | 4.96246424 | 1 |
| 10 | 87960904 | T | G | F | C | PTEN | ENST00000371953 | 0.74213 | 0.93986 | 0.97447 | 0.93219 | 0.96298 | 0.9796564915 | 0.96584 | 0.9614534378 | 0.454731 | 3.781049041 | 0 |
| 10 | 87960906 | C | A | H | N | PTEN | ENST00000371953 | 0.74308 | 0.93062 | 0.97501 | 0.93011 | 0.88496 | 0.9418998128 | 0.82267 | 0.9952114409 | 0.16973 | 7.576303747 | 0 |
| 10 | 87960906 | C | G | H | D | PTEN | ENST00000371953 | 0.78503 | 0.98086 | 0.97313 | 0.94355 | 0.98183 | 0.9865586441 | 0.92485 | 0.9926023257 | 0.429682 | 7.666000457 | 0 |
| 10 | 87960906 | C | T | H | Y | PTEN | ENST00000371953 | 0.73717 | 0.86874 | 0.97667 | 0.87642 | 0.96189 | 0.9265616897 | 0.94357 | 0.9835084081 | 0.421528 | 5.409109013 | 0 |
| 10 | 87960907 | A | C | H | P | PTEN | ENST00000371953 | 0.92613 | 0.9893 | 0.9452 | 0.95015 | 0.98367 | 0.9940332862 | 0.95265 | 0.997564435 | 0.440097 | 7.516305651 | 1 |
| 10 | 87960907 | A | G | H | R | PTEN | ENST00000371953 | 0.7214 | 0.92622 | 0.97665 | 0.87396 | 0.97401 | 0.9560282332 | 0.95961 | 0.9718914628 | 0.406738 | 5.703389496 | 0 |
| 10 | 87960907 | A | T | H | L | PTEN | ENST00000371953 | 0.82576 | 0.92844 | 0.96446 | 0.9583 | 0.98621 | 0.9709037872 | 0.93668 | 0.9971068501 | 0.430513 | 5.391522806 | 0 |
| 10 | 87960908 | C | A | H | Q | PTEN | ENST00000371953 | 0.50805 | 0.83613 | 0.96677 | 0.86105 | 0.89467 | 0.9414800028 | 0.93215 | 0.9626953602 | 0.326425 | 3.666372573 | 0 |
| 10 | 87960909 | T | A | F | I | PTEN | ENST00000371953 | 0.89594 | 0.96469 | 0.96925 | 0.93234 | 0.96553 | 0.9637219586 | 0.91477 | 0.9931004047 | 0.309341 | 4.815900929 | 0 |
| 10 | 87960909 | T | G | F | V | PTEN | ENST00000371953 | 0.9026 | 0.94984 | 0.96538 | 0.964 | 0.9844 | 0.953667024 | 0.91119 | 0.9960938143 | 0.40691 | 5.455616332 | 0 |
| 10 | 87960910 | T | A | F | Y | PTEN | ENST00000371953 | 0.94363 | 0.96848 | 0.97379 | 0.90865 | 0.93397 | 0.905920534 | 0.89355 | 0.9881934006 | 0.257395 | 6.133386952 | 0 |
| 10 | 87960911 | T | G | F | L | PTEN | ENST00000371953 | 0.92956 | 0.93659 | 0.98118 | 0.89948 | 0.89918 | 0.9404679144 | 0.89715 | 0.9844656587 | 0.255549 | 4.511954577 | 0 |
| 10 | 87960913 | G | C | W | S | PTEN | ENST00000371953 | 0.95154 | 0.99148 | 0.96032 | 0.95748 | 0.94394 | 0.9743240026 | 0.93398 | 0.9998251796 | 0.430487 | 5.335588298 | 0 |
| 10 | 87960914 | G | T | W | C | PTEN | ENST00000371953 | 0.95612 | 0.98772 | 0.96758 | 0.96551 | 0.95753 | 0.9860967774 | 0.96855 | 0.9998401403 | 0.467643 | 4.28618535 | 0 |
| 10 | 87960915 | G | C | V | L | PTEN | ENST00000371953 | 0.53657 | 0.78709 | 0.8012 | 0.73012 | 0.72926 | 0.8223502402 | 0.96306 | 0.6119667888 | 0.181318 | 3.393704527 | 0 |
| 10 | 87960915 | G | T | V | L | PTEN | ENST00000371953 | 0.53113 | 0.78709 | 0.8012 | 0.73407 | 0.72926 | 0.8223502402 | 0.96329 | 0.6119667888 | 0.181317 | 3.393704527 | 0 |
| 10 | 87960916 | T | G | V | G | PTEN | ENST00000371953 | 0.91423 | 0.94728 | 0.69147 | 0.90597 | 0.98404 | 0.9629255108 | 0.9768 | 0.9551254511 | 0.39869 | 5.024773614 | 1 |
| 10 | 87960918 | A | C | N | H | PTEN | ENST00000371953 | 0.79207 | 0.96441 | 0.86381 | 0.93636 | 0.93241 | 0.9370182565 | 0.95254 | 0.9973599315 | 0.188484 | 4.834691189 | 0 |
| 10 | 87960918 | A | G | N | D | PTEN | ENST00000371953 | 0.81716 | 0.9764 | 0.89187 | 0.95479 | 0.96956 | 0.9440489204 | 0.96289 | 0.9966991402 | 0.228684 | 7.299869298 | 0 |
| 10 | 87960918 | A | T | N | Y | PTEN | ENST00000371953 | 0.83606 | 0.97592 | 0.8888 | 0.97685 | 0.98621 | 0.9699773959 | 0.97151 | 0.9991874099 | 0.450195 | 6.100890827 | 1 |
| 10 | 87960919 | A | C | N | T | PTEN | ENST00000371953 | 0.89895 | 0.96965 | 0.83879 | 0.96867 | 0.9729 | 0.9105304842 | 0.94743 | 0.9975192903 | 0.223991 | 7.470408052 | 1 |
| 10 | 87960919 | A | G | N | S | PTEN | ENST00000371953 | 0.8588 | 0.95159 | 0.80126 | 0.96578 | 0.96773 | 0.8756384837 | 0.96104 | 0.9948621392 | 0.198408 | 5.94641997 | 0 |
| 10 | 87960919 | A | T | N | I | PTEN | ENST00000371953 | 0.92382 | 0.9811 | 0.86416 | 0.98348 | 0.96044 | 0.9692789429 | 0.9404 | 0.9993579984 | 0.283629 | 6.693008556 | 1 |
| 10 | 87960920 | T | A | N | K | PTEN | ENST00000371953 | 0.75822 | 0.9771 | 0.91427 | 0.95305 | 0.93475 | 0.9678595486 | 0.96755 | 0.9957387447 | 0.166274 | 6.951005654 | 1 |
| 10 | 87960921 | A | C | T | P | PTEN | ENST00000371953 | 0.82334 | 0.98329 | 0.82268 | 0.98959 | 0.99892 | 0.9894632858 | 0.99424 | 0.9955255542 | 0.572134 | 5.216542436 | 0 |
| 10 | 87960921 | A | G | T | A | PTEN | ENST00000371953 | 0.77172 | 0.96033 | 0.81658 | 0.98672 | 0.997 | 0.9656999198 | 0.99368 | 0.9941347241 | 0.56941 | 4.934869297 | 0 |

| | | | | | | | | | | | | | | | | | | |
|---|---|---|---|---|---|---|---|---|---|---|---|---|---|---|---|---|---|---|
| 10 | 87960921 | A | T | T | S | PTEN | ENST00000371953 | 0.74252 | 0.95316 | 0.83238 | 0.98425 | 0.98404 | 0.9173337195 | 0.96349 | 0.9918353558 | 0.406415 | 4.033049804 | 0 |
| 10 | 87960922 | C | A | T | K | PTEN | ENST00000371953 | 0.81857 | 0.97954 | 0.90716 | 0.97685 | 0.9986 | 0.9901647547 | 0.99256 | 0.994514823 | 0.563006 | 6.831160217 | 1 |
| 10 | 87960922 | C | G | T | R | PTEN | ENST00000371953 | 0.80716 | 0.97662 | 0.89615 | 0.97869 | 0.99765 | 0.9935963863 | 0.99546 | 0.9941807389 | 0.563258 | 7.800457953 | 1 |
| 10 | 87960924 | T | A | F | I | PTEN | ENST00000371953 | 0.84073 | 0.95467 | 0.9235 | 0.92886 | 0.96664 | 0.9555681016 | 0.90875 | 0.9945616581 | 0.287019 | 7.812206728 | 0 |
| 10 | 87960924 | T | G | F | V | PTEN | ENST00000371953 | 0.68411 | 0.89517 | 0.90744 | 0.93965 | 0.9822 | 0.9484780845 | 0.96506 | 0.9855214953 | 0.405892 | 7.36659351 | 0 |
| 10 | 87960925 | T | A | F | Y | PTEN | ENST00000371953 | 0.81724 | 0.83931 | 0.92929 | 0.83819 | 0.90406 | 0.8671803556 | 0.84561 | 0.8659515977 | 0.194206 | 4.329537269 | 0 |
| 10 | 87960925 | T | C | F | S | PTEN | ENST00000371953 | 0.94999 | 0.93387 | 0.91479 | 0.91092 | 0.95499 | 0.9439694065 | 0.91362 | 0.9842364788 | 0.306511 | 5.689697503 | 0 |
| 10 | 87960925 | T | G | F | C | PTEN | ENST00000371953 | 0.95049 | 0.98348 | 0.94102 | 0.96201 | 0.98764 | 0.9493668808 | 0.95327 | 0.9981902355 | 0.434084 | 6.779446702 | 0 |
| 10 | 87960926 | C | G | F | L | PTEN | ENST00000371953 | 0.75424 | 0.88868 | 0.93937 | 0.91337 | 0.91399 | 0.9140213864 | 0.91536 | 0.986915946 | 0.255452 | 6.062438067 | 0 |
| 10 | 87960927 | T | A | F | I | PTEN | ENST00000371953 | 0.8402 | 0.96723 | 0.8676 | 0.95351 | 0.98693 | 0.980499926 | 0.98313 | 0.9934209252 | 0.470191 | 7.196168056 | 0 |
| 10 | 87960928 | T | A | F | Y | PTEN | ENST00000371953 | 0.92747 | 0.97613 | 0.88158 | 0.93363 | 0.9703 | 0.9196592616 | 0.94811 | 0.987673547 | 0.340924 | 5.908806202 | 0 |
| 10 | 87960928 | T | C | F | S | PTEN | ENST00000371953 | 0.95087 | 0.99392 | 0.86756 | 0.96989 | 0.99138 | 0.9891464694 | 0.98991 | 0.9970756769 | 0.485367 | 6.857940532 | 0 |
| 10 | 87960928 | T | G | F | C | PTEN | ENST00000371953 | 0.95108 | 0.98772 | 0.9006 | 0.97132 | 0.99138 | 0.9824464893 | 0.98648 | 0.9974211454 | 0.492368 | 7.564929984 | 0 |
| 10 | 87960929 | C | G | F | L | PTEN | ENST00000371953 | 0.75822 | 0.96463 | 0.89768 | 0.92076 | 0.9506 | 0.9500678402 | 0.9843 | 0.9868984222 | 0.437965 | 5.303909774 | 0 |
| 10 | 87960930 | A | C | I | L | PTEN | ENST00000371953 | 0.60956 | 0.79586 | 0.78434 | 0.84828 | 0.90052 | 0.8242617975 | 0.92734 | 0.8592782617 | 0.26066 | 3.560451543 | 0 |
| 10 | 87960930 | A | G | I | V | PTEN | ENST00000371953 | 0.32674 | 0.74525 | 0.68159 | 0.5968 | 0.68821 | 0.7178782565 | 0.80932 | 0.4871818125 | 0.0950428 | 1.807434251 | 0 |
| 10 | 87960930 | A | T | I | L | PTEN | ENST00000371953 | 0.60339 | 0.79586 | 0.78434 | 0.84828 | 0.90007 | 0.8242617975 | 0.92738 | 0.8592782617 | 0.26066 | 3.560451543 | 0 |
| 10 | 87960931 | T | A | I | K | PTEN | ENST00000371953 | 0.92072 | 0.99078 | 0.85272 | 0.93561 | 0.98693 | 0.980479655 | 0.98573 | 0.996455431 | 0.461734 | 7.812989759 | 0 |
| 10 | 87960931 | T | C | I | T | PTEN | ENST00000371953 | 0.90519 | 0.91663 | 0.89356 | 0.90415 | 0.96627 | 0.9121748972 | 0.97992 | 0.9587164521 | 0.450382 | 4.075738958 | 0 |
| 10 | 87960931 | T | G | I | R | PTEN | ENST00000371953 | 0.92768 | 0.99078 | 0.8311 | 0.93697 | 0.98972 | 0.9731018233 | 0.98748 | 0.9981983614 | 0.462196 | 7.63107445 | 0 |
| 10 | 87960932 | A | G | I | M | PTEN | ENST00000371953 | 0.82717 | 0.87935 | 0.74412 | 0.87852 | 0.9182 | 0.8375697809 | 0.91085 | 0.9202358723 | 0.259734 | 4.609338829 | 0 |
| 10 | 87960933 | C | A | P | T | PTEN | ENST00000371953 | 0.37036 | 0.82373 | 0.75817 | 0.87749 | 0.82892 | 0.7669889465 | 0.94195 | 0.7311721444 | 0.278959 | 3.114439571 | 0 |
| 10 | 87960933 | C | T | P | S | PTEN | ENST00000371953 | 0.52879 | 0.7978 | 0.725 | 0.83103 | 0.83505 | 0.7411733522 | 0.86416 | 0.6701241732 | 0.206713 | 3.454402522 | 0 |
| 10 | 87960934 | C | A | P | Q | PTEN | ENST00000371953 | 0.6918 | 0.77738 | 0.73571 | 0.85376 | 0.80653 | 0.7511423693 | 0.90001 | 0.7149906158 | 0.230723 | 3.18223631 | 0 |
| 10 | 87960934 | C | G | P | R | PTEN | ENST00000371953 | 0.67255 | 0.80935 | 0.80885 | 0.83112 | 0.88022 | 0.7568703622 | 0.9373 | 0.7252378464 | 0.343526 | 3.067617786 | 0 |
| 10 | 87960934 | C | T | P | L | PTEN | ENST00000371953 | 0.73181 | 0.80534 | 0.81616 | 0.87493 | 0.89648 | 0.7975643887 | 0.91689 | 0.901001215 | 0.287608 | 4.193086102 | 0 |
| 10 | 87960936 | G | A | G | R | PTEN | ENST00000371953 | 0.77521 | 0.85146 | 0.92204 | 0.88936 | 0.90406 | 0.8062030826 | 0.96003 | 0.8068361878 | 0.398952 | 3.495639241 | 0 |
| 10 | 87960937 | G | C | G | A | PTEN | ENST00000371953 | 0.58084 | 0.81509 | 0.85643 | 0.79058 | 0.78411 | 0.7367816453 | 0.94633 | 0.8294281363 | 0.231432 | 2.680804562 | 0 |
| 10 | 87960937 | G | T | G | V | PTEN | ENST00000371953 | 0.84271 | 0.8998 | 0.86126 | 0.90997 | 0.93939 | 0.8268356314 | 0.95127 | 0.8993576169 | 0.402569 | 3.332308938 | 0 |
| 10 | 87960939 | C | A | P | T | PTEN | ENST00000371953 | 0.33758 | 0.82515 | 0.56263 | 0.70132 | 0.7217 | 0.685833982 | 0.70521 | 0.587879777 | 0.0717379 | 3.816931442 | 0 |
| 10 | 87960939 | C | G | P | A | PTEN | ENST00000371953 | 0.30781 | 0.78737 | 0.52416 | 0.61069 | 0.59881 | 0.6369814713 | 0.72203 | 0.7820994854 | 0.0630761 | 2.989677073 | 0 |
| 10 | 87960939 | C | T | P | S | PTEN | ENST00000371953 | 0.32104 | 0.79515 | 0.52678 | 0.62094 | 0.58391 | 0.6906665547 | 0.58122 | 0.8743044734 | 0.00412513 | 2.984234326 | 0 |
| 10 | 87960940 | C | A | P | Q | PTEN | ENST00000371953 | 0.47646 | 0.7777 | 0.54245 | 0.70813 | 0.75417 | 0.6560998811 | 0.77178 | 0.7356240153 | 0.0647044 | 3.565144059 | 0 |
| 10 | 87960940 | C | G | P | R | PTEN | ENST00000371953 | 0.44942 | 0.82115 | 0.61041 | 0.57553 | 0.61068 | 0.7329462883 | 0.81232 | 0.8527896404 | 0.0946955 | 3.992153723 | 0 |
| 10 | 87960942 | G | A | E | K | PTEN | ENST00000371953 | 0.57645 | 0.83657 | 0.669 | 0.82922 | 0.81703 | 0.735839142 | 0.86499 | 0.9832016826 | 0.198832 | 4.560207992 | 0 |
| 10 | 87960943 | A | C | E | A | PTEN | ENST00000371953 | 0.58283 | 0.81688 | 0.53885 | 0.81585 | 0.75687 | 0.7270165084 | 0.76643 | 0.9660228491 | 0.106186 | 3.949589676 | 0 |
| 10 | 87960943 | A | G | E | G | PTEN | ENST00000371953 | 0.62856 | 0.83939 | 0.48012 | 0.81675 | 0.76553 | 0.7457608143 | 0.78508 | 0.9799653292 | 0.131253 | 3.766174024 | 0 |
| 10 | 87960943 | A | T | E | V | PTEN | ENST00000371953 | 0.78883 | 0.84186 | 0.51008 | 0.83622 | 0.848 | 0.7854498032 | 0.85865 | 0.9724232554 | 0.15324 | 4.249354602 | 0 |
| 10 | 87960944 | G | T | E | D | PTEN | ENST00000371953 | 0.33265 | 0.76567 | 0.53972 | 0.41904 | 0.60362 | 0.6655534759 | 0.78313 | 0.2613711655 | -0.0214939 | 3.492636481 | 0 |
| 10 | 87960945 | G | A | E | K | PTEN | ENST00000371953 | 0.82193 | 0.82848 | 0.79507 | 0.78863 | 0.89603 | 0.8237942571 | 0.94118 | 0.960932672 | 0.287152 | 3.236775307 | 0 |
| 10 | 87960945 | G | C | E | Q | PTEN | ENST00000371953 | 0.80258 | 0.80834 | 0.68098 | 0.75641 | 0.83725 | 0.7484446374 | 0.9079 | 0.8971610069 | 0.231714 | 3.76066236 | 0 |
| 10 | 87960946 | A | C | E | A | PTEN | ENST00000371953 | 0.77362 | 0.81688 | 0.69505 | 0.82084 | 0.90007 | 0.7861429682 | 0.92464 | 0.9440504909 | 0.265375 | 3.85297514 | 0 |
| 10 | 87960946 | A | G | E | G | PTEN | ENST00000371953 | 0.66778 | 0.83774 | 0.65027 | 0.82029 | 0.83615 | 0.8035464986 | 0.95449 | 0.9401840568 | 0.340787 | 3.434824704 | 0 |
| 10 | 87960946 | A | T | E | V | PTEN | ENST00000371953 | 0.85338 | 0.83804 | 0.67217 | 0.86328 | 0.89829 | 0.8106268209 | 0.91497 | 0.9393345714 | 0.270986 | 3.437801056 | 0 |
| 10 | 87960948 | A | C | T | P | PTEN | ENST00000371953 | 0.46718 | 0.86227 | 0.49367 | 0.69595 | 0.60952 | 0.8682632134 | 0.85772 | 0.7145286202 | 0.0788012 | 1.32188478 | 0 |
| 10 | 87960948 | A | G | T | A | PTEN | ENST00000371953 | 0.40775 | 0.78576 | 0.47503 | 0.57192 | 0.45523 | 0.578351416 | 0.76695 | 0.7043732405 | -0.0561848 | 1.462218929 | 0 |
| 10 | 87960948 | A | T | T | S | PTEN | ENST00000371953 | 0.33737 | 0.75868 | 0.47474 | 0.50116 | 0.40224 | 0.4814428563 | 0.76469 | 0.5826959014 | -0.0586798 | 0.1869993138 | 0 |
| 10 | 87960949 | C | A | T | N | PTEN | ENST00000371953 | 0.29303 | 0.82996 | 0.54608 | 0.54762 | 0.43397 | 0.7050035511 | 0.68467 | 0.526127696 | -0.0858576 | -0.3098825524 | 0 |
| 10 | 87960949 | C | G | T | S | PTEN | ENST00000371953 | 0.28896 | 0.75868 | 0.47474 | 0.49548 | 0.39643 | 0.4814428563 | 0.7244 | 0.3584737704 | -0.0600861 | 0.1869993138 | 0 |
| 10 | 87960949 | C | T | T | I | PTEN | ENST00000371953 | 0.37323 | 0.8206 | 0.60201 | 0.66143 | 0.45348 | 0.6806471632 | 0.7448 | 0.6498464346 | -0.00453244 | 1.754184661 | 0 |
| 10 | 87960951 | T | A | S | T | PTEN | ENST00000371953 | 0.40559 | 0.7644 | 0.56347 | 0.52439 | 0.39251 | 0.5346784944 | 0.77989 | 0.4598942399 | -0.0303785 | 2.36827374 | 0 |
| 10 | 87960951 | T | C | S | P | PTEN | ENST00000371953 | 0.50147 | 0.85728 | 0.58559 | 0.62066 | 0.68292 | 0.8202100287 | 0.9333 | 0.5086200237 | 0.082958 | 1.611516282 | 0 |
| 10 | 87960951 | T | G | S | A | PTEN | ENST00000371953 | 0.43232 | 0.73866 | 0.55396 | 0.51105 | 0.39053 | 0.4554867836 | 0.8491 | 0.5327374935 | 0.00877276 | 3.31400856 | 0 |
| 10 | 87960952 | C | T | S | L | PTEN | ENST00000371953 | 0.46612 | 0.76413 | 0.59169 | 0.68978 | 0.68113 | 0.6170196408 | 0.89823 | 0.7870517969 | 0.0810011 | 1.911520465 | 0 |
| 10 | 87960954 | G | A | E | K | PTEN | ENST00000371953 | 0.56394 | 0.82904 | 0.69537 | 0.74247 | 0.65575 | 0.6491504525 | 0.78827 | 0.7181569044 | 0.0556828 | 2.840062253 | 0 |
| 10 | 87960954 | G | C | E | Q | PTEN | ENST00000371953 | 0.52055 | 0.84201 | 0.57517 | 0.76668 | 0.65377 | 0.5973150669 | 0.86518 | 0.9539062977 | 0.0238629 | 1.599597948 | 0 |
| 10 | 87960955 | A | C | E | A | PTEN | ENST00000371953 | 0.59871 | 0.81108 | 0.58333 | 0.66628 | 0.60002 | 0.6343048642 | 0.75713 | 0.7434463928 | 0.0478634 | 2.627771042 | 0 |
| 10 | 87960955 | A | T | E | V | PTEN | ENST00000371953 | 0.64571 | 0.83536 | 0.56056 | 0.78179 | 0.66449 | 0.6602523988 | 0.82186 | 0.9276878238 | 0.0829636 | 3.725026837 | 0 |
| 10 | 87960956 | A | T | E | D | PTEN | ENST00000371953 | 0.43196 | 0.76117 | 0.58357 | 0.61577 | 0.54201 | 0.5450587146 | 0.64931 | 0.2396088988 | -0.113383 | 0.3025029252 | 0 |
| 10 | 87960957 | A | C | K | Q | PTEN | ENST00000371953 | 0.58496 | 0.83979 | 0.6405 | 0.70659 | 0.74021 | 0.6880806574 | 0.94035 | 0.8983733654 | 0.131944 | 2.951704528 | 0 |
| 10 | 87960957 | A | G | K | E | PTEN | ENST00000371953 | 0.57118 | 0.88108 | 0.6921 | 0.65988 | 0.79697 | 0.7668783926 | 0.98869 | 0.6584794521 | 0.254267 | 2.691642505 | 0 |
| 10 | 87960958 | A | C | K | T | PTEN | ENST00000371953 | 0.53113 | 0.87551 | 0.69292 | 0.73244 | 0.85274 | 0.765427683 | 0.89334 | 0.8983891606 | 0.209907 | 2.916679891 | 0 |
| 10 | 87960958 | A | G | K | R | PTEN | ENST00000371953 | 0.47018 | 0.72862 | 0.60642 | 0.63156 | 0.65077 | 0.5394843376 | 0.79926 | 0.7219588161 | -0.0175752 | 2.202635191 | 0 |
| 10 | 87960958 | A | T | K | I | PTEN | ENST00000371953 | 0.86692 | 0.93298 | 0.66897 | 0.81106 | 0.9045 | 0.8155722334 | 0.89879 | 0.9690440297 | 0.226494 | 3.802684093 | 0 |
| 10 | 87960959 | A | T | K | N | PTEN | ENST00000371953 | 0.60515 | 0.85239 | 0.71836 | 0.75882 | 0.70097 | 0.7756970326 | 0.88621 | 0.8732761741 | 0.0762637 | 2.096052588 | 0 |
| 10 | 87960960 | G | C | V | L | PTEN | ENST00000371953 | 0.4857 | 0.78643 | 0.67031 | 0.68938 | 0.5519 | 0.584620046 | 0.68508 | 0.7736004591 | -0.0506217 | 1.114749235 | 0 |
| 10 | 87960960 | G | T | V | L | PTEN | ENST00000371953 | 0.4805 | 0.78643 | 0.67031 | 0.69192 | 0.5491 | 0.584620046 | 0.69685 | 0.7736004591 | -0.0506043 | 1.114749235 | 0 |
| 10 | 87960961 | T | A | V | E | PTEN | ENST00000371953 | 0.78399 | 0.91153 | 0.67838 | 0.85863 | 0.77527 | 0.7940666148 | 0.83867 | 0.9534512162 | 0.132562 | 3.053349392 | 0 |
| 10 | 87960961 | T | C | V | A | PTEN | ENST00000371953 | 0.56193 | 0.84369 | 0.69589 | 0.74401 | 0.68908 | 0.595080152 | 0.81142 | 0.8917869925 | 0.083224 | 2.689342145 | 0 |
| 10 | 87960961 | T | G | V | G | PTEN | ENST00000371953 | 0.62589 | 0.86306 | 0.55019 | 0.851 | 0.77399 | 0.7251771251 | 0.76089 | 0.942451179 | 0.124028 | 2.302185121 | 0 |
| 10 | 87960963 | G | A | E | K | PTEN | ENST00000371953 | 0.73477 | 0.85134 | 0.83194 | 0.89388 | 0.91526 | 0.7345764758 | 0.96312 | 0.9173664451 | 0.37834 | 4.746891292 | 0 |
| 10 | 87960963 | G | C | E | Q | PTEN | ENST00000371953 | 0.67637 | 0.8344 | 0.72179 | 0.79245 | 0.83725 | 0.6546775123 | 0.91156 | 0.8884959221 | 0.220128 | 3.672351427 | 0 |
| 10 | 87960964 | A | C | E | A | PTEN | ENST00000371953 | 0.77632 | 0.82163 | 0.74004 | 0.86489 | 0.91484 | 0.7047088648 | 0.95385 | 0.9176463485 | 0.377076 | 3.838855741 | 0 |
| 10 | 87960964 | A | G | E | G | PTEN | ENST00000371953 | 0.85916 | 0.83912 | 0.69561 | 0.88342 | 0.90885 | 0.7124642019 | 0.92883 | 0.9224758744 | 0.375264 | 3.35873573 | 0 |
| 10 | 87960964 | A | T | E | V | PTEN | ENST00000371953 | 0.83298 | 0.84075 | 0.71865 | 0.88491 | 0.94543 | 0.7566930601 | 0.96423 | 0.8743780255 | 0.396076 | 4.352413869 | 0 |
| 10 | 87960965 | A | T | E | D | PTEN | ENST00000371953 | 0.61686 | 0.85687 | 0.73759 | 0.72421 | 0.79273 | 0.6276871385 | 0.90031 | 0.8666340709 | 0.146882 | 3.847453038 | 0 |
| 10 | 87960966 | A | C | N | H | PTEN | ENST00000371953 | 0.65 | 0.81087 | 0.75152 | 0.94392 | 0.91314 | 0.7080495938 | 0.95579 | 0.9789110422 | 0.226637 | 4.85494422 | 0 |
| 10 | 87960966 | A | T | N | Y | PTEN | ENST00000371953 | 0.83644 | 0.92405 | 0.78286 | 0.96379 | 0.96846 | 0.8120267154 | 0.96179 | 0.9937654734 | 0.410285 | 5.305413986 | 0 |
| 10 | 87960967 | A | C | N | T | PTEN | ENST00000371953 | 0.9026 | 0.80465 | 0.72363 | 0.95103 | 0.90668 | 0.7368137456 | 0.93885 | 0.9764600396 | 0.235226 | 4.290111899 | 0 |
| 10 | 87960967 | A | G | N | S | PTEN | ENST00000371953 | 0.73309 | 0.73533 | 0.68314 | 0.94815 | 0.88684 | 0.6112515741 | 0.93712 | 0.9381760955 | 0.152826 | 3.575720743 | 0 |
| 10 | 87960967 | A | T | N | I | PTEN | ENST00000371953 | 0.92871 | 0.86151 | 0.74899 | 0.97912 | 0.95826 | 0.8471134796 | 0.93921 | 0.9849860305 | 0.334614 | 4.067796204 | 0 |
| 10 | 87961968 | T | A | N | K | PTEN | ENST00000371953 | 0.81638 | 0.87029 | 0.81196 | 0.93814 | 0.88637 | 0.741464224 | 0.93265 | 0.9867549539 | 0.179551 | 4.583675962 | 0 |
| 10 | 87960970 | G | A | G | E | PTEN | ENST00000371953 | 0.92504 | 0.9208 | 0.73774 | 0.94217 | 0.97475 | 0.886126476 | 0.92865 | 0.9003981792 | 0.500646 | 3.824649059 | 0 |
| 10 | 87960970 | G | C | G | A | PTEN | ENST00000371953 | 0.90202 | 0.87317 | 0.6451 | 0.92491 | 0.96044 | 0.8320331641 | 0.91239 | 0.9541327357 | 0.356375 | 3.649442225 | 0 |
| 10 | 87960970 | G | T | G | V | PTEN | ENST00000371953 | 0.92438 | 0.86857 | 0.6629 | 0.9433 | 0.97104 | 0.9250536788 | 0.94995 | 0.9136699154 | 0.509962 | 3.954804186 | 0 |
| 10 | 87960972 | A | T | S | C | PTEN | ENST00000371953 | 0.67517 | 0.94615 | 0.44539 | 0.87142 | 0.81703 | 0.7921324868 | 0.83679 | 0.965444386 | 0.165588 | 4.601603135 | 0 |
| 10 | 87960973 | G | A | S | N | PTEN | ENST00000371953 | 0.47036 | 0.77211 | 0.49189 | 0.68962 | 0.64468 | 0.6994684766 | 0.88062 | 0.6736463904 | 0.0221455 | 3.126758503 | 0 |
| 10 | 87960973 | G | C | S | T | PTEN | ENST00000371953 | 0.4614 | 0.76771 | 0.4331 | 0.54945 | 0.55468 | 0.6988919798 | 0.8399 | 0.6718329787 | 0.0102897 | 3.527513526 | 0 |
| 10 | 87960973 | G | T | S | I | PTEN | ENST00000371953 | 0.67795 | 0.91917 | 0.4853 | 0.80173 | 0.76223 | 0.8718298562 | 0.86489 | 0.9649749994 | 0.177517 | 5.001252646 | 0 |
| 10 | 87960975 | C | G | L | V | PTEN | ENST00000371953 | 0.29889 | 0.79478 | 0.36806 | 0.68822 | 0.5377 | 0.5829613662 | 0.6613 | 0.5318233967 | 0.0117078 | 0.9680637023 | 0 |
| 10 | 87960976 | T | A | L | Q | PTEN | ENST00000371953 | 0.4569 | 0.81905 | 0.46894 | 0.83479 | 0.7816 | 0.7429923819 | 0.85156 | 0.9162476063 | 0.174585 | 3.613955504 | 0 |
| 10 | 87960976 | T | C | L | P | PTEN | ENST00000371953 | 0.4725 | 0.90935 | 0.63371 | 0.88304 | 0.81124 | 0.8821842695 | 0.91199 | 0.8787325621 | 0.219556 | 2.942706364 | 0 |
| 10 | 87960976 | T | G | L | R | PTEN | ENST00000371953 | 0.46426 | 0.89054 | 0.5798 | 0.84945 | 0.77463 | 0.8302338407 | 0.86034 | 0.9273590446 | 0.168044 | 2.850037319 | 0 |
| 10 | 87960978 | T | C | C | R | PTEN | ENST00000371953 | 0.42412 | 0.93519 | 0.77776 | 0.78003 | 0.65967 | 0.7717819551 | 0.88473 | 0.4384544194 | 0.118828 | 3.405780171 | 0 |
| 10 | 87960979 | G | A | C | Y | PTEN | ENST00000371953 | 0.41298 | 0.93298 | 0.80112 | 0.69099 | 0.60599 | 0.7444736357 | 0.71736 | 0.7631869912 | 0.0942715 | 2.62800586 | 0 |
| 10 | 87960979 | G | C | C | S | PTEN | ENST00000371953 | 0.43695 | 0.84833 | 0.65598 | 0.64438 | 0.54486 | 0.7179227645 | 0.72344 | 0.4862813354 | 0.0659506 | 1.285885488 | 0 |
| 10 | 87960979 | G | T | C | F | PTEN | ENST00000371953 | 0.44887 | 0.91712 | 0.68882 | 0.69245 | 0.58771 | 0.7236554533 | 0.81261 | 0.6200456619 | 0.0891108 | 2.670986552 | 0 |
| 10 | 87960981 | G | A | D | N | PTEN | ENST00000371953 | 0.47488 | 0.78539 | 0.56898 | 0.76385 | 0.64261 | 0.559468009 | 0.83175 | 0.8794468045 | -0.0175515 | 1.185363795 | 0 |
| 10 | 87960981 | G | T | D | Y | PTEN | ENST00000371953 | 0.64097 | 0.93261 | 0.56989 | 0.77549 | 0.85378 | 0.7711836174 | 0.91435 | 0.9210479453 | 0.257911 | 4.455947502 | 0 |
| 10 | 87960982 | A | C | D | A | PTEN | ENST00000371953 | 0.52532 | 0.85159 | 0.4844 | 0.6861 | 0.54201 | 0.6816875097 | 0.7886 | 0.8615446091 | 0.1438 | 2.334493751 | 0 |
| 10 | 87960982 | A | T | D | V | PTEN | ENST00000371953 | 0.64368 | 0.88395 | 0.47081 | 0.82019 | 0.70589 | 0.7621298921 | 0.925 | 0.8990598917 | 0.19203 | 3.05861706 | 0 |

| | | | | | | | | | | | | | | | | | | |
|---|---|---|---|---|---|---|---|---|---|---|---|---|---|---|---|---|---|---|
| 10 | 87960983 | T | A | D | E | PTEN | ENST00000371953 | 0.39783 | 0.77148 | 0.52953 | 0.61333 | 0.5046 | 0.4805816433 | 0.7766 | 0.4277610481 | 0.00072921 | 1.719784866 | 0 |
| 10 | 87960984 | C | A | Q | K | PTEN | ENST00000371953 | 0.3054 | 0.85629 | 0.53684 | 0.68339 | 0.57487 | 0.5968911807 | 0.76062 | 0.7469841242 | 0.0714879 | 2.1617579 | 0 |
| 10 | 87960984 | C | G | Q | E | PTEN | ENST00000371953 | 0.27189 | 0.81364 | 0.47208 | 0.50329 | 0.53334 | 0.6184094824 | 0.70765 | 0.08562511791 | -0.111224 | 1.531970174 | 0 |
| 10 | 87960985 | A | C | Q | P | PTEN | ENST00000371953 | 0.47344 | 0.87654 | 0.48418 | 0.71897 | 0.67015 | 0.839820023 | 0.83146 | 0.7382846475 | 0.11505 | 2.154035655 | 0 |
| 10 | 87960985 | A | G | Q | R | PTEN | ENST00000371953 | 0.41339 | 0.81229 | 0.52468 | 0.55978 | 0.49511 | 0.5534122654 | 0.82189 | 0.6691964269 | 0.0551398 | 2.185474553 | 0 |
| 10 | 87960985 | A | T | Q | L | PTEN | ENST00000371953 | 0.47677 | 0.79411 | 0.46904 | 0.67573 | 0.56695 | 0.6955694423 | 0.85902 | 0.7250156403 | 0.0957613 | 3.84802815 | 0 |
| 10 | 87960987 | G | A | E | K | PTEN | ENST00000371953 | 0.59966 | 0.82792 | 0.79971 | 0.78053 | 0.80889 | 0.6258935026 | 0.95872 | 0.6648115625 | 0.302841 | 1.915583761 | 0 |
| 10 | 87960987 | G | C | E | Q | PTEN | ENST00000371953 | 0.54433 | 0.84778 | 0.69285 | 0.76799 | 0.73877 | 0.4993273125 | 0.8724 | 0.8709594607 | 0.169569 | 2.341902477 | 0 |
| 10 | 87960988 | A | C | E | A | PTEN | ENST00000371953 | 0.56082 | 0.8126 | 0.70098 | 0.70353 | 0.77205 | 0.5581489229 | 0.98751 | 0.7775473282 | 0.342341 | 2.5214818 | 0 |
| 10 | 87960988 | A | G | E | G | PTEN | ENST00000371953 | 0.61492 | 0.83669 | 0.65687 | 0.69357 | 0.79516 | 0.6468164825 | 0.98513 | 0.7561615579 | 0.358013 | 2.796823615 | 0 |
| 10 | 87960988 | A | T | E | V | PTEN | ENST00000371953 | 0.61514 | 0.85536 | 0.68062 | 0.73203 | 0.86951 | 0.6668348018 | 0.99144 | 0.7679396189 | 0.365703 | 3.182173608 | 0 |
| 10 | 87960989 | A | T | E | D | PTEN | ENST00000371953 | 0.43214 | 0.76117 | 0.70223 | 0.63403 | 0.63947 | 0.4528138671 | 0.90331 | 0.5573758483 | 0.118965 | 2.265913762 | 0 |
| 10 | 87960990 | A | C | I | L | PTEN | ENST00000371953 | 0.38628 | 0.78988 | 0.53198 | 0.56605 | 0.5108 | 0.4346299873 | 0.63937 | 0.8175597787 | -0.00308371 | -0.05987509178 | 0 |
| 10 | 87960990 | A | G | I | V | PTEN | ENST00000371953 | 0.37239 | 0.73472 | 0.44437 | 0.60489 | 0.44293 | 0.3287904986 | 0.72911 | 0.8021666408 | -0.00500653 | 2.110232894 | 0 |
| 10 | 87960990 | A | T | I | F | PTEN | ENST00000371953 | 0.44296 | 0.90445 | 0.53764 | 0.72171 | 0.54628 | 0.4968750769 | 0.82811 | 0.6735252142 | 0.108917 | 1.906397438 | 0 |
| 10 | 87960991 | T | A | I | N | PTEN | ENST00000371953 | 0.45273 | 0.93519 | 0.61929 | 0.77242 | 0.61416 | 0.6424232333 | 0.79256 | 0.7690058947 | 0.111074 | 0.7284641929 | 0 |
| 10 | 87960991 | T | C | I | T | PTEN | ENST00000371953 | 0.38966 | 0.86607 | 0.63643 | 0.70914 | 0.5976 | 0.4537024569 | 0.70862 | 0.6636890769 | 0.108031 | 1.281032199 | 0 |
| 10 | 87960991 | T | G | I | S | PTEN | ENST00000371953 | 0.42526 | 0.88793 | 0.54401 | 0.68641 | 0.54769 | 0.6852657478 | 0.7022 | 0.8517552018 | 0.113117 | 0.2934585216 | 0 |
| 10 | 87960992 | C | G | I | M | PTEN | ENST00000371953 | 0.28198 | 0.88875 | 0.49319 | 0.77582 | 0.60599 | 0.3948514729 | 0.80869 | 0.2330228535 | 0.0193887 | 2.544002882 | 0 |
| 10 | 87960993 | G | A | D | N | PTEN | ENST00000371953 | 0.53187 | 0.78402 | 0.55621 | 0.80767 | 0.65967 | 0.5950978753 | 0.86349 | 0.4497483969 | -0.0194005 | 2.249010798 | 0 |
| 10 | 87960993 | G | C | D | H | PTEN | ENST00000371953 | 0.74165 | 0.9489 | 0.58894 | 0.86849 | 0.84159 | 0.7092469852 | 0.78569 | 0.7379391801 | 0.163322 | 2.62008169 | 0 |
| 10 | 87960993 | G | T | D | Y | PTEN | ENST00000371953 | 0.87315 | 0.92004 | 0.56607 | 0.86978 | 0.87781 | 0.7851716007 | 0.86298 | 0.7899973666 | 0.19538 | 3.53813503 | 0 |
| 10 | 87960994 | A | C | D | A | PTEN | ENST00000371953 | 0.54055 | 0.86267 | 0.48198 | 0.64572 | 0.61988 | 0.702031514 | 0.80174 | 0.9035574198 | 0.150906 | 2.821282462 | 0 |
| 10 | 87960994 | A | G | D | G | PTEN | ENST00000371953 | 0.54136 | 0.86526 | 0.50163 | 0.73498 | 0.5976 | 0.7171334363 | 0.86471 | 0.7614105541 | 0.166202 | 2.98166034 | 0 |
| 10 | 87960994 | A | T | D | V | PTEN | ENST00000371953 | 0.56352 | 0.89851 | 0.48003 | 0.83305 | 0.74446 | 0.7870638235 | 0.92772 | 0.7828245199 | 0.198795 | 3.20427825 | 0 |
| 10 | 87960995 | T | A | D | E | PTEN | ENST00000371953 | 0.37188 | 0.77437 | 0.52718 | 0.67669 | 0.56961 | 0.5276070544 | 0.7901 | 0.7090507746 | 0.0706616 | 2.229248402 | 0 |
| 10 | 87960996 | A | G | S | G | PTEN | ENST00000371953 | 0.4454 | 0.76461 | 0.44459 | 0.543 | 0.57877 | 0.4357542133 | 0.78914 | 0.4781304896 | -0.0152423 | 0.345212867 | 0 |
| 10 | 87960996 | A | T | S | C | PTEN | ENST00000371953 | 0.66208 | 0.94813 | 0.50707 | 0.86944 | 0.65869 | 0.5556882869 | 0.86854 | 0.9080187678 | 0.0887919 | 3.041082303 | 0 |
| 10 | 87960997 | G | A | S | N | PTEN | ENST00000371953 | 0.42126 | 0.77316 | 0.55091 | 0.75223 | 0.6847 | 0.4849246695 | 0.84199 | 0.379339844 | -0.00853542 | 1.357003167 | 0 |
| 10 | 87960997 | G | C | S | T | PTEN | ENST00000371953 | 0.39963 | 0.81783 | 0.50347 | 0.73722 | 0.59146 | 0.437690586 | 0.71938 | 0.8073094487 | 0.0406023 | 2.000762145 | 0 |
| 10 | 87960997 | G | T | S | I | PTEN | ENST00000371953 | 0.4894 | 0.84673 | 0.51621 | 0.78737 | 0.65377 | 0.6105204011 | 0.77709 | 0.8669617176 | 0.10725 | 3.499487468 | 0 |
| 10 | 87960998 | C | A | S | R | PTEN | ENST00000371953 | 0.36164 | 0.82842 | 0.56 | 0.78727 | 0.5348 | 0.5697677099 | 0.64716 | 0.2860965729 | 0.010777 | 1.610038887 | 0 |
| 10 | 87960999 | A | C | I | L | PTEN | ENST00000371953 | 0.44448 | 0.79054 | 0.66771 | 0.76422 | 0.52446 | 0.5682336442 | 0.81121 | 0.6933174133 | 0.121484 | 1.595221036 | 0 |
| 10 | 87960999 | A | G | I | V | PTEN | ENST00000371953 | 0.40517 | 0.73554 | 0.56396 | 0.72889 | 0.56961 | 0.4061290137 | 0.83706 | 0.8127830029 | 0.0922651 | 1.993588717 | 0 |
| 10 | 87960999 | A | T | I | F | PTEN | ENST00000371953 | 0.47564 | 0.90533 | 0.67726 | 0.81787 | 0.68908 | 0.6672338313 | 0.87145 | 0.8727890849 | 0.170709 | 2.534613028 | 0 |
| 10 | 87961000 | T | A | I | N | PTEN | ENST00000371953 | 0.52899 | 0.93894 | 0.76634 | 0.83726 | 0.71627 | 0.7913622591 | 0.85454 | 0.7781178355 | 0.168819 | 2.450372871 | 0 |
| 10 | 87961000 | T | C | I | T | PTEN | ENST00000371953 | 0.45819 | 0.87256 | 0.78218 | 0.77041 | 0.76946 | 0.6199647693 | 0.8614 | 0.6627797484 | 0.175804 | 1.873939444 | 0 |
| 10 | 87961000 | T | G | I | S | PTEN | ENST00000371953 | 0.52986 | 0.88829 | 0.68768 | 0.82251 | 0.77972 | 0.7825567016 | 0.89005 | 0.3449267149 | 0.187778 | 1.632820621 | 0 |
| 10 | 87961001 | T | G | I | M | PTEN | ENST00000371953 | 0.36448 | 0.86111 | 0.62548 | 0.79362 | 0.52446 | 0.5401858341 | 0.85996 | 0.3403647542 | 0.114018 | 2.583988671 | 0 |
| 10 | 87961002 | T | A | C | S | PTEN | ENST00000371953 | 0.4249 | 0.84751 | 0.54036 | 0.63741 | 0.50616 | 0.5634678886 | 0.62564 | 0.3056121469 | 0.0351868 | 1.040441099 | 0 |
| 10 | 87961002 | T | C | C | R | PTEN | ENST00000371953 | 0.48481 | 0.92418 | 0.6537 | 0.71113 | 0.58135 | 0.7327926724 | 0.75452 | 0.3856115341 | 0.077097 | 1.553100003 | 0 |
| 10 | 87961002 | T | G | C | G | PTEN | ENST00000371953 | 0.472 | 0.85069 | 0.50884 | 0.6473 | 0.54486 | 0.6388206205 | 0.79045 | 0.3292928305 | 0.0835115 | 0.4672375713 | 0 |
| 10 | 87961003 | G | A | C | Y | PTEN | ENST00000371953 | 0.38937 | 0.91619 | 0.6741 | 0.56488 | 0.55468 | 0.665903632 | 0.61264 | 0.1793497657 | 0.0463757 | 1.137085457 | 0 |
| 10 | 87961003 | G | C | C | S | PTEN | ENST00000371953 | 0.44077 | 0.84751 | 0.54036 | 0.66716 | 0.53334 | 0.5634678886 | 0.69679 | 0.3116712899 | 0.0416055 | 1.040441099 | 0 |
| 10 | 87961003 | G | T | C | F | PTEN | ENST00000371953 | 0.43896 | 0.89733 | 0.55024 | 0.53648 | 0.57487 | 0.5843632988 | 0.7388 | 0.3326697052 | 0.0442134 | 0.02533040929 | 0 |
| 10 | 87961004 | C | G | C | W | PTEN | ENST00000371953 | 0.39626 | 0.89552 | 0.61261 | 0.86731 | 0.70427 | 0.7074471386 | 0.89856 | 0.2114213258 | 0.13462 | 3.793291811 | 0 |
| 10 | 87961005 | A | G | S | G | PTEN | ENST00000371953 | 0.50244 | 0.76461 | 0.52958 | 0.80199 | 0.5108 | 0.6157551844 | 0.88292 | 0.4365753233 | 0.0588403 | 2.372282626 | 0 |
| 10 | 87961005 | A | T | S | C | PTEN | ENST00000371953 | 0.71 | 0.94008 | 0.59591 | 0.92513 | 0.85274 | 0.6650767502 | 0.84159 | 0.8909626007 | 0.188901 | 6.306010951 | 0 |
| 10 | 87961006 | G | A | S | N | PTEN | ENST00000371953 | 0.47879 | 0.7757 | 0.63653 | 0.78965 | 0.58135 | 0.6018484639 | 0.84584 | 0.768527329 | -0.0101422 | 1.866206123 | 0 |
| 10 | 87961006 | G | C | S | T | PTEN | ENST00000371953 | 0.46382 | 0.76477 | 0.58359 | 0.77611 | 0.51539 | 0.4553636843 | 0.77062 | 0.6430058479 | 0.055405 | 1.151363196 | 0 |
| 10 | 87961006 | G | T | S | I | PTEN | ENST00000371953 | 0.67279 | 0.87969 | 0.61271 | 0.83171 | 0.80831 | 0.7456167004 | 0.89857 | 0.8969962001 | 0.204184 | 4.311767869 | 0 |
| 10 | 87961007 | T | A | S | R | PTEN | ENST00000371953 | 0.4189 | 0.90581 | 0.65592 | 0.81049 | 0.77972 | 0.7427901818 | 0.92808 | 0.8809503317 | 0.155794 | 2.295208897 | 0 |
| 10 | 87961008 | A | C | I | L | PTEN | ENST00000371953 | 0.42353 | 0.79156 | 0.43088 | 0.62892 | 0.52745 | 0.4893323095 | 0.75458 | 0.369625777 | 0.0613127 | 0.8561523967 | 0 |
| 10 | 87961008 | A | G | I | V | PTEN | ENST00000371953 | 0.37611 | 0.73472 | 0.33274 | 0.61237 | 0.52145 | 0.3197317469 | 0.72461 | 0.286916468 | 0.0200967 | 1.382289539 | 0 |
| 10 | 87961008 | A | T | I | L | PTEN | ENST00000371953 | 0.41854 | 0.79156 | 0.43088 | 0.63029 | 0.5304 | 0.4893323095 | 0.75444 | 0.369625777 | 0.0612863 | 0.8561523967 | 0 |
| 10 | 87961009 | T | A | I | K | PTEN | ENST00000371953 | 0.38565 | 0.93519 | 0.51341 | 0.62475 | 0.67843 | 0.7718671564 | 0.85038 | 0.5368831158 | 0.115321 | 1.048175647 | 0 |
| 10 | 87961009 | T | C | I | T | PTEN | ENST00000371953 | 0.41451 | 0.85472 | 0.5425 | 0.51358 | 0.59393 | 0.4681153042 | 0.58731 | 0.309713295 | -0.0140916 | -0.769501099 | 0 |
| 10 | 87961009 | T | G | I | R | PTEN | ENST00000371953 | 0.44417 | 0.93519 | 0.48472 | 0.72149 | 0.70427 | 0.7397624822 | 0.87314 | 0.6205109358 | 0.125156 | 1.839644923 | 0 |
| 10 | 87961010 | A | G | I | M | PTEN | ENST00000371953 | 0.47747 | 0.79973 | 0.38449 | 0.68478 | 0.48544 | 0.4345829592 | 0.8123 | 0.4741397202 | 0.060746 | 1.705434355 | 0 |
| 10 | 87961011 | G | A | E | K | PTEN | ENST00000371953 | 0.5709 | 0.82705 | 0.77143 | 0.84046 | 0.84375 | 0.6222648212 | 0.98767 | 0.9405685067 | 0.444827 | 2.376023712 | 0 |
| 10 | 87961011 | G | C | E | Q | PTEN | ENST00000371953 | 0.55195 | 0.79676 | 0.64138 | 0.79172 | 0.79577 | 0.4782060435 | 0.97717 | 0.5568040013 | 0.315767 | 2.524153246 | 0 |
| 10 | 87961012 | A | C | E | A | PTEN | ENST00000371953 | 0.46016 | 0.85695 | 0.66328 | 0.81371 | 0.84428 | 0.5481064786 | 0.98232 | 0.6391812563 | 0.346515 | 2.861111554 | 0 |
| 10 | 87961012 | A | G | E | G | PTEN | ENST00000371953 | 0.52745 | 0.83669 | 0.60376 | 0.8262 | 0.84535 | 0.6264395654 | 0.98885 | 0.7828611135 | 0.377375 | 0.907386852 | 0 |
| 10 | 87961012 | A | T | E | V | PTEN | ENST00000371953 | 0.53853 | 0.90391 | 0.63248 | 0.87479 | 0.86047 | 0.6368115643 | 0.94591 | 0.8566892147 | 0.283902 | 2.25625801 | 0 |
| 10 | 87961013 | G | T | E | D | PTEN | ENST00000371953 | 0.25843 | 0.81884 | 0.65634 | 0.63984 | 0.78844 | 0.4667665966 | 0.9735 | 0.1185464412 | 0.193143 | 2.421500111 | 0 |
| 10 | 87961014 | C | A | R | S | PTEN | ENST00000371953 | 0.39521 | 0.8267 | 0.38325 | 0.74886 | 0.59146 | 0.6308756701 | 0.69105 | 0.7302384973 | 0.109509 | 2.260349764 | 0 |
| 10 | 87961014 | C | G | R | G | PTEN | ENST00000371953 | 0.46488 | 0.8593 | 0.37725 | 0.78284 | 0.75687 | 0.7303060643 | 0.97327 | 0.752068758 | 0.241252 | 1.87656066 | 0 |
| 10 | 87961014 | C | T | R | C | PTEN | ENST00000371953 | 0.5721 | 0.87267 | 0.48496 | 0.77151 | 0.63525 | 0.6597953382 | 0.77377 | 0.8492737412 | 0.106471 | 4.935093196 | 0 |
| 10 | 87961015 | G | A | R | H | PTEN | ENST00000371953 | 0.61753 | 0.8511 | 0.39252 | 0.7937 | 0.66449 | 0.4595418162 | 0.86021 | 0.5155140162 | 0.151932 | 2.610726195 | 0 |
| 10 | 87961015 | G | C | R | P | PTEN | ENST00000371953 | 0.6426 | 0.92582 | 0.4326 | 0.88259 | 0.86803 | 0.8337324271 | 0.96798 | 0.9389010668 | 0.367403 | 2.948940856 | 0 |
| 10 | 87961015 | G | T | R | L | PTEN | ENST00000371953 | 0.60178 | 0.85232 | 0.4057 | 0.81741 | 0.78658 | 0.6696584423 | 0.97067 | 0.9300262332 | 0.259784 | 2.819546187 | 0 |
| 10 | 87961017 | G | A | A | T | PTEN | ENST00000371953 | 0.38261 | 0.78791 | 0.30429 | 0.60661 | 0.46889 | 0.2563597047 | 0.76214 | 0.1486644 | 0.0565551 | 0.9841848645 | 0 |
| 10 | 87961017 | G | C | A | P | PTEN | ENST00000371953 | 0.42252 | 0.86195 | 0.3781 | 0.689 | 0.51387 | 0.5942175895 | 0.72999 | 0.1383482069 | 0.0704325 | 1.904793193 | 0 |
| 10 | 87961018 | C | A | A | E | PTEN | ENST00000371953 | 0.34593 | 0.83536 | 0.38423 | 0.65057 | 0.5348 | 0.4713315569 | 0.61176 | 0.5822110176 | 0.0740833 | -0.1104978942 | 0 |
| 10 | 87961018 | C | G | A | G | PTEN | ENST00000371953 | 0.41139 | 0.79267 | 0.30817 | 0.67449 | 0.52446 | 0.3186049733 | 0.66253 | 0.4847162366 | 0.031314 | -0.0273358244 | 0 |
| 10 | 87961018 | C | T | A | V | PTEN | ENST00000371953 | 0.43556 | 0.79617 | 0.34553 | 0.65197 | 0.58771 | 0.3470027743 | 0.72656 | 0.6973128915 | 0.0589214 | 1.112248919 | 0 |
| 10 | 87961020 | G | A | D | N | PTEN | ENST00000371953 | 0.58218 | 0.87051 | 0.54327 | 0.80228 | 0.67933 | 0.4345622889 | 0.88789 | 0.941005528 | 0.0854782 | 3.177231558 | 0 |
| 10 | 87961020 | G | C | D | H | PTEN | ENST00000371953 | 0.74866 | 0.96033 | 0.57759 | 0.89472 | 0.84106 | 0.6088043098 | 0.83939 | 0.9611403346 | 0.203204 | 3.402010872 | 0 |
| 10 | 87961020 | G | T | D | Y | PTEN | ENST00000371953 | 0.89006 | 0.96506 | 0.56871 | 0.90862 | 0.91058 | 0.6897569228 | 0.93122 | 0.9830447435 | 0.30532 | 3.911189924 | 0 |
| 10 | 87961021 | A | C | D | A | PTEN | ENST00000371953 | 0.5735 | 0.85232 | 0.48134 | 0.78587 | 0.68995 | 0.6040380446 | 0.87644 | 0.8799943924 | 0.176262 | 2.921263111 | 0 |
| 10 | 87961021 | A | T | D | V | PTEN | ENST00000371953 | 0.58817 | 0.89014 | 0.4629 | 0.88907 | 0.8517 | 0.7027374861 | 0.96526 | 0.8820897937 | 0.304185 | 3.822645228 | 0 |
| 10 | 87961022 | T | A | D | E | PTEN | ENST00000371953 | 0.35536 | 0.774 | 0.50336 | 0.57744 | 0.60717 | 0.3395555366 | 0.73083 | 0.4528569877 | 0.0324147 | 2.165918327 | 0 |
| 10 | 87961023 | A | C | N | H | PTEN | ENST00000371953 | 0.79342 | 0.94091 | 0.58906 | 0.84626 | 0.77399 | 0.4649636653 | 0.9235 | 0.9444534183 | 0.127435 | 3.755960869 | 0 |
| 10 | 87961023 | A | G | N | D | PTEN | ENST00000371953 | 0.50498 | 0.87364 | 0.63764 | 0.68401 | 0.55883 | 0.4440422415 | 0.87656 | 0.7386288047 | 0.0957884 | 3.098532574 | 0 |
| 10 | 87961023 | A | T | N | Y | PTEN | ENST00000371953 | 0.84195 | 0.9303 | 0.62795 | 0.88273 | 0.84321 | 0.5727598803 | 0.89922 | 0.9697468281 | 0.202731 | 4.570456338 | 0 |
| 10 | 87961024 | A | C | N | T | PTEN | ENST00000371953 | 0.46824 | 0.84996 | 0.55197 | 0.69019 | 0.71391 | 0.4642999703 | 0.87188 | 0.8050363064 | 0.128962 | 4.377344436 | 0 |
| 10 | 87961024 | A | G | N | S | PTEN | ENST00000371953 | 0.41022 | 0.72137 | 0.50546 | 0.53444 | 0.55607 | 0.3259834247 | 0.75686 | 0.8198728561 | -0.00708391 | 2.264928733 | 0 |
| 10 | 87961024 | A | T | N | I | PTEN | ENST00000371953 | 0.55628 | 0.93111 | 0.58295 | 0.86908 | 0.8233 | 0.6371628634 | 0.89111 | 0.9600203633 | 0.190982 | 4.111302618 | 0 |
| 10 | 87961025 | T | A | N | K | PTEN | ENST00000371953 | 0.40922 | 0.86111 | 0.67089 | 0.66389 | 0.60717 | 0.4947511289 | 0.89669 | 0.3664288819 | 0.0835755 | 3.633607319 | 0 |
| 10 | 87961026 | G | A | D | N | PTEN | ENST00000371953 | 0.58496 | 0.94947 | 0.76881 | 0.9234 | 0.76685 | 0.5321572796 | 0.92404 | 0.9617212415 | 0.0849017 | 3.206695558 | 0 |
| 10 | 87961026 | G | C | D | H | PTEN | ENST00000371953 | 0.87964 | 0.97539 | 0.79375 | 0.9374 | 0.95645 | 0.6868075253 | 0.96675 | 0.973923564 | 0.47245 | 4.02934081 | 0 |
| 10 | 87961026 | G | T | D | Y | PTEN | ENST00000371953 | 0.89423 | 0.97869 | 0.77595 | 0.9411 | 0.96664 | 0.7877660498 | 0.98777 | 0.9878755212 | 0.508208 | 5.188272736 | 0 |
| 10 | 87961027 | A | G | D | G | PTEN | ENST00000371953 | 0.6132 | 0.97114 | 0.71929 | 0.90716 | 0.88069 | 0.7335910619 | 0.98577 | 0.8676788211 | 0.441502 | 3.121676708 | 0 |
| 10 | 87961027 | A | T | D | V | PTEN | ENST00000371953 | 0.9018 | 0.97507 | 0.70124 | 0.96052 | 0.96262 | 0.7778178821 | 0.97913 | 0.9448677897 | 0.468045 | 3.915217471 | 0 |
| 10 | 87961029 | A | C | K | Q | PTEN | ENST00000371953 | 0.54528 | 0.93628 | 0.60421 | 0.86766 | 0.84906 | 0.4837164403 | 0.87052 | 0.5861059877 | 0.132054 | 4.037357726 | 0 |
| 10 | 87961029 | A | G | K | E | PTEN | ENST00000371953 | 0.5179 | 0.94296 | 0.65516 | 0.77169 | 0.82668 | 0.5656978597 | 0.86344 | 0.5951110721 | 0.147916 | 3.077624339 | 0 |
| 10 | 87961030 | A | C | K | T | PTEN | ENST00000371953 | 0.5025 | 0.95219 | 0.67427 | 0.87133 | 0.76881 | 0.6210260341 | 0.86032 | 0.749745369 | 0.170322 | 3.262255772 | 0 |
| 10 | 87961030 | A | G | K | R | PTEN | ENST00000371953 | 0.45487 | 0.73489 | 0.56375 | 0.65813 | 0.61646 | 0.3461626586 | 0.77381 | 0.5454814434 | 0.0142036 | 1.413660049 | 0 |
| 10 | 87961030 | A | T | K | M | PTEN | ENST00000371953 | 0.72796 | 0.98134 | 0.56707 | 0.90146 | 0.88637 | 0.6245666772 | 0.84771 | 0.8770964914 | 0.183098 | 4.838442627 | 0 |
| 10 | 87961031 | G | T | K | N | PTEN | ENST00000371953 | 0.40477 | 0.95467 | 0.6901 | 0.83401 | 0.68734 | 0.5819150981 | 0.91772 | 0.7359457612 | 0.0454609 | 3.176600745 | 0 |
| 10 | 87961032 | G | C | E | Q | PTEN | ENST00000371953 | 0.63443 | 0.90368 | 0.65597 | 0.78376 | 0.69594 | 0.5133285436 | 0.81956 | 0.8840903044 | 0.0409198 | 2.783854385 | 0 |

| | | | | | | | | | | | | | | | | | | |
|---|---|---|---|---|---|---|---|---|---|---|---|---|---|---|---|---|---|---|
| 10 | 87961033 | A | C | E | A | PTEN | ENST00000371953 | 0.54446 | 0.86978 | 0.66738 | 0.74936 | 0.74164 | 0.5809535081 | 0.76808 | 0.5687602758 | 0.0179634 | 3.285810766 | 0 |
| 10 | 87961033 | A | G | E | G | PTEN | ENST00000371953 | 0.60149 | 0.89172 | 0.63118 | 0.68542 | 0.71153 | 0.6238997735 | 0.72706 | 0.9460273385 | 0.0808354 | 2.581680756 | 0 |
| 10 | 87961033 | A | T | E | V | PTEN | ENST00000371953 | 0.60047 | 0.93871 | 0.64519 | 0.78104 | 0.80238 | 0.6737512901 | 0.79947 | 0.9492245913 | 0.0922449 | 3.580359968 | 0 |
| 10 | 87961034 | A | T | E | D | PTEN | ENST00000371953 | 0.25 | 0.76193 | 0.66293 | 0.48835 | 0.58771 | 0.4249902356 | 0.53093 | 0.06215482578 | -0.130818 | -0.1038299147 | 0 |
| 10 | 87961035 | T | A | Y | N | PTEN | ENST00000371953 | 0.60515 | 0.91093 | 0.90151 | 0.84816 | 0.88259 | 0.7701753711 | 0.97317 | 0.7071460485 | 0.340872 | 4.36501139 | 0 |
| 10 | 87961035 | T | C | Y | H | PTEN | ENST00000371953 | 0.71754 | 0.95441 | 0.90663 | 0.80608 | 0.86703 | 0.6612538835 | 0.80665 | 0.7045371532 | 0.171431 | 3.741025521 | 0 |
| 10 | 87961035 | T | G | Y | D | PTEN | ENST00000371953 | 0.61776 | 0.94057 | 0.90151 | 0.86945 | 0.90755 | 0.8614291172 | 0.97441 | 0.7528155446 | 0.36535 | 3.769831709 | 0 |
| 10 | 87961036 | A | C | Y | S | PTEN | ENST00000371953 | 0.53623 | 0.886 | 0.84965 | 0.75417 | 0.8345 | 0.7777797776 | 0.97686 | 0.4081436241 | 0.335673 | 3.587709087 | 0 |
| 10 | 87961036 | A | G | Y | C | PTEN | ENST00000371953 | 0.77664 | 0.96932 | 0.90499 | 0.83432 | 0.89648 | 0.7662498551 | 0.89039 | 0.8315035795 | 0.242414 | 3.429422035 | 0 |
| 10 | 87961036 | A | T | Y | F | PTEN | ENST00000371953 | 0.44047 | 0.76571 | 0.83161 | 0.7182 | 0.68558 | 0.455894062 | 0.90257 | 0.3672480861 | 0.0964441 | 3.104806931 | 0 |
| 10 | 87961038 | C | A | L | I | PTEN | ENST00000371953 | 0.49782 | 0.96251 | 0.77953 | 0.82956 | 0.77527 | 0.416331078 | 0.83198 | 0.8078786731 | 0.0533019 | 4.489717417 | 0 |
| 10 | 87961038 | C | G | L | V | PTEN | ENST00000371953 | 0.63405 | 0.95687 | 0.72579 | 0.83527 | 0.82443 | 0.5081853376 | 0.87908 | 0.930811286 | 0.0743833 | 4.626586383 | 0 |
| 10 | 87961039 | T | A | L | Q | PTEN | ENST00000371953 | 0.79358 | 0.96994 | 0.79789 | 0.91626 | 0.89422 | 0.8131266767 | 0.89014 | 0.9713250399 | 0.0857712 | 5.42209553 | 0 |
| 10 | 87961039 | T | G | L | R | PTEN | ENST00000371953 | 0.65877 | 0.98712 | 0.85705 | 0.92601 | 0.92397 | 0.8644984742 | 0.90866 | 0.9523135032 | 0.21803 | 4.190171404 | 0 |
| 10 | 87961041 | G | A | V | I | PTEN | ENST00000371953 | 0.40068 | 0.72766 | 0.4708 | 0.39418 | 0.58896 | 0.2456240128 | 0.61267 | 0.343254596 | -0.118553 | 0.02210279465 | 0 |
| 10 | 87961042 | T | A | V | E | PTEN | ENST00000371953 | 0.72124 | 0.9584 | 0.65427 | 0.83041 | 0.80536 | 0.76258844 | 0.89111 | 0.9246652126 | 0.0928151 | 2.712660148 | 0 |
| 10 | 87961042 | T | C | V | A | PTEN | ENST00000371953 | 0.53771 | 0.88282 | 0.66575 | 0.72837 | 0.67933 | 0.5205560782 | 0.86003 | 0.7734666467 | -0.00302384 | 3.583806592 | 0 |
| 10 | 87961044 | C | A | L | I | PTEN | ENST00000371953 | 0.70856 | 0.97332 | 0.88962 | 0.90563 | 0.88307 | 0.540055505 | 0.86269 | 0.9519315958 | 0.140617 | 4.647033082 | 0 |
| 10 | 87961044 | C | G | L | V | PTEN | ENST00000371953 | 0.57645 | 0.96895 | 0.83823 | 0.90311 | 0.86753 | 0.6530493078 | 0.88323 | 0.963134408 | 0.169329 | 4.175706184 | 0 |
| 10 | 87961044 | C | T | L | F | PTEN | ENST00000371953 | 0.74269 | 0.97311 | 0.8707 | 0.90482 | 0.90229 | 0.7632398925 | 0.86006 | 0.9683091044 | 0.176501 | 4.570827654 | 0 |
| 10 | 87961045 | T | A | L | H | PTEN | ENST00000371953 | 0.92052 | 0.97939 | 0.93853 | 0.95492 | 0.96664 | 0.9171892751 | 0.94385 | 0.9968026876 | 0.387455 | 5.779235163 | 0 |
| 10 | 87961045 | T | G | L | R | PTEN | ENST00000371953 | 0.93105 | 0.9882 | 0.93711 | 0.95446 | 0.98657 | 0.9667820064 | 0.94742 | 0.9969468713 | 0.431766 | 6.648155171 | 0 |
| 10 | 87961047 | A | C | T | P | PTEN | ENST00000371953 | 0.57027 | 0.87812 | 0.81343 | 0.78443 | 0.85689 | 0.8643777711 | 0.9493 | 0.8799415827 | 0.236901 | 4.461912751 | 1 |
| 10 | 87961047 | A | G | T | A | PTEN | ENST00000371953 | 0.5306 | 0.80105 | 0.81495 | 0.75494 | 0.77076 | 0.6112944847 | 0.92199 | 0.9099635482 | 0.142433 | 3.900664998 | 0 |
| 10 | 87961047 | A | T | T | S | PTEN | ENST00000371953 | 0.51738 | 0.87508 | 0.83294 | 0.74926 | 0.81006 | 0.5266827013 | 0.86513 | 0.8166257143 | 0.0878361 | 2.763141479 | 0 |
| 10 | 87961048 | C | A | T | N | PTEN | ENST00000371953 | 0.60647 | 0.9584 | 0.89534 | 0.81702 | 0.88165 | 0.7438129014 | 0.87414 | 0.9122230304 | 0.113976 | 4.411942779 | 0 |
| 10 | 87961048 | C | T | T | I | PTEN | ENST00000371953 | 0.62977 | 0.86759 | 0.92939 | 0.81677 | 0.91314 | 0.6857998298 | 0.88148 | 0.8157776594 | 0.114381 | 3.620620596 | 0 |
| 10 | 87961050 | T | A | L | I | PTEN | ENST00000371953 | 0.62038 | 0.95776 | 0.92462 | 0.83792 | 0.82046 | 0.5640603047 | 0.81899 | 0.7358792424 | 0.0711078 | 3.97436903 | 0 |
| 10 | 87961050 | T | G | L | V | PTEN | ENST00000371953 | 0.76187 | 0.96506 | 0.88236 | 0.89819 | 0.84159 | 0.6774281425 | 0.89309 | 0.9342075844 | 0.111086 | 5.771582785 | 0 |
| 10 | 87961051 | T | C | L | S | PTEN | ENST00000371953 | 0.93798 | 0.98602 | 0.92647 | 0.95767 | 0.97735 | 0.9594740627 | 0.95116 | 0.9962984308 | 0.34981 | 5.5205155 | 1 |
| 10 | 87961052 | A | T | L | F | PTEN | ENST00000371953 | 0.5932 | 0.95605 | 0.90676 | 0.87294 | 0.82273 | 0.762228355 | 0.82592 | 0.8908486962 | 0.0984891 | 3.92464194 | 0 |
| 10 | 87961053 | A | C | T | P | PTEN | ENST00000371953 | 0.54324 | 0.90266 | 0.65714 | 0.66281 | 0.74797 | 0.7763254738 | 0.8034 | 0.6297922525 | 0.0675936 | 3.015874335 | 0 |
| 10 | 87961053 | A | T | T | S | PTEN | ENST00000371953 | 0.39104 | 0.75868 | 0.6725 | 0.48495 | 0.54628 | 0.4248422108 | 0.78352 | 0.3012842834 | -0.0893085 | 2.78088431 | 0 |
| 10 | 87961054 | C | G | T | R | PTEN | ENST00000371953 | 0.36609 | 0.82907 | 0.76277 | 0.67805 | 0.5976 | 0.7403976573 | 0.81131 | 0.4070102935 | 0.0673304 | 3.268880664 | 0 |
| 10 | 87961054 | C | T | T | I | PTEN | ENST00000371953 | 0.43028 | 0.81454 | 0.81322 | 0.75714 | 0.73 | 0.675843427 | 0.67364 | 0.8525992632 | 0.035553 | 4.779920212 | 0 |
| 10 | 87961056 | A | C | K | Q | PTEN | ENST00000371953 | 0.83112 | 0.97295 | 0.85997 | 0.89605 | 0.90229 | 0.8673614034 | 0.95828 | 0.9773274093 | 0.167767 | 4.860036427 | 0 |
| 10 | 87961056 | A | G | K | E | PTEN | ENST00000371953 | 0.85537 | 0.98638 | 0.8905 | 0.93574 | 0.96407 | 0.9110650345 | 0.95718 | 0.9888098836 | 0.339791 | 5.925857346 | 0 |
| 10 | 87961057 | A | C | K | T | PTEN | ENST00000371953 | 0.92712 | 0.97924 | 0.90129 | 0.94041 | 0.9458 | 0.8801677907 | 0.93738 | 0.9954701736 | 0.294541 | 5.827955927 | 0 |
| 10 | 87961057 | A | G | K | R | PTEN | ENST00000371953 | 0.92419 | 0.94974 | 0.83019 | 0.82487 | 0.86403 | 0.7604389743 | 0.93968 | 0.9750890705 | 0.100547 | 5.252596107 | 0 |
| 10 | 87961057 | A | T | K | I | PTEN | ENST00000371953 | 0.94162 | 0.99406 | 0.86704 | 0.95094 | 0.93863 | 0.9023548552 | 0.9322 | 0.9986741543 | 0.289562 | 5.373031496 | 0 |
| 10 | 87961058 | A | T | K | N | PTEN | ENST00000371953 | 0.90993 | 0.97822 | 0.91137 | 0.91836 | 0.89239 | 0.901753419 | 0.95271 | 0.9908692837 | 0.102831 | 5.27533555 | 0 |
| 10 | 87961059 | A | C | N | H | PTEN | ENST00000371953 | 0.62893 | 0.93655 | 0.85839 | 0.64442 | 0.7872 | 0.5199827487 | 0.92566 | 0.8063195944 | 0.030063 | 4.094605424 | 0 |
| 10 | 87961059 | A | G | N | D | PTEN | ENST00000371953 | 0.49589 | 0.82772 | 0.88678 | 0.5355 | 0.55607 | 0.5909067849 | 0.85287 | 0.7964686155 | -0.00118247 | 2.62180959 | 0 |
| 10 | 87961059 | A | T | N | Y | PTEN | ENST00000371953 | 0.60178 | 0.91999 | 0.88365 | 0.83851 | 0.85843 | 0.6770400197 | 0.85456 | 0.9432120323 | 0.116925 | 4.498338272 | 0 |
| 10 | 87961060 | A | C | N | T | PTEN | ENST00000371953 | 0.36794 | 0.79751 | 0.83437 | 0.58263 | 0.49351 | 0.4968893161 | 0.79246 | 0.5246742964 | -0.028212 | 4.123896372 | 0 |
| 10 | 87961060 | A | G | N | S | PTEN | ENST00000371953 | 0.33126 | 0.72564 | 0.80187 | 0.51987 | 0.56695 | 0.4012047534 | 0.8923 | 0.8200184107 | -0.0922408 | 3.514282165 | 0 |
| 10 | 87961060 | A | T | N | I | PTEN | ENST00000371953 | 0.43022 | 0.87783 | 0.86212 | 0.77679 | 0.81646 | 0.7039355799 | 0.88639 | 0.8931591511 | 0.106393 | 5.470630484 | 0 |
| 10 | 87961061 | T | A | N | K | PTEN | ENST00000371953 | 0.35361 | 0.83394 | 0.90991 | 0.66303 | 0.71548 | 0.611095328 | 0.8726 | 0.2554459572 | -0.0377321 | 3.984904699 | 0 |
| 10 | 87961062 | G | A | D | N | PTEN | ENST00000371953 | 0.74762 | 0.80201 | 0.92382 | 0.88288 | 0.81414 | 0.7684571178 | 0.94374 | 0.9022991379 | 0.0185268 | 4.786463014 | 0 |
| 10 | 87961062 | G | C | D | H | PTEN | ENST00000371953 | 0.8897 | 0.97143 | 0.93653 | 0.91843 | 0.93709 | 0.8177503704 | 0.89674 | 0.9664647217 | 0.2804 | 6.343141514 | 0 |
| 10 | 87961062 | G | T | D | Y | PTEN | ENST00000371953 | 0.89707 | 0.97628 | 0.92647 | 0.90334 | 0.94281 | 0.8702848701 | 0.93802 | 0.9721975923 | 0.316492 | 6.839695763 | 0 |
| 10 | 87961063 | A | C | D | A | PTEN | ENST00000371953 | 0.92642 | 0.93364 | 0.88237 | 0.87867 | 0.93901 | 0.8115312886 | 0.85335 | 0.9390271902 | 0.252425 | 4.956108859 | 0 |
| 10 | 87961063 | A | G | D | G | PTEN | ENST00000371953 | 0.81473 | 0.94046 | 0.88897 | 0.80976 | 0.91271 | 0.8517744653 | 0.92897 | 0.8958262241 | 0.264147 | 4.795152772 | 0 |
| 10 | 87961063 | A | T | D | V | PTEN | ENST00000371953 | 0.9296 | 0.97734 | 0.88463 | 0.91798 | 0.94281 | 0.856279067 | 0.91908 | 0.9936693311 | 0.293529 | 5.796527639 | 0 |
| 10 | 87961064 | T | A | D | E | PTEN | ENST00000371953 | 0.36626 | 0.77364 | 0.90778 | 0.66885 | 0.63525 | 0.6047739274 | 0.90238 | 0.6991664767 | 0.0635029 | 2.846405822 | 0 |
| 10 | 87961065 | C | A | L | I | PTEN | ENST00000371953 | 0.41392 | 0.84389 | 0.89135 | 0.75825 | 0.70508 | 0.5678339262 | 0.84249 | 0.4620930552 | 0.101327 | 3.649590188 | 0 |
| 10 | 87961065 | C | G | L | V | PTEN | ENST00000371953 | 0.55012 | 0.88126 | 0.83727 | 0.80086 | 0.78782 | 0.7308995959 | 0.90369 | 0.7890514731 | 0.130186 | 4.73729819 | 0 |
| 10 | 87961065 | C | T | L | F | PTEN | ENST00000371953 | 0.77219 | 0.96653 | 0.87033 | 0.86665 | 0.90755 | 0.8084957395 | 0.9037 | 0.9772386743 | 0.180976 | 6.478537712 | 0 |
| 10 | 87961066 | T | A | L | H | PTEN | ENST00000371953 | 0.89376 | 0.89862 | 0.92854 | 0.95032 | 0.9729 | 0.9163961186 | 0.9409 | 0.9935218692 | 0.347372 | 8.955288289 | 1 |
| 10 | 87961066 | T | C | L | P | PTEN | ENST00000371953 | 0.91078 | 0.98981 | 0.94462 | 0.95507 | 0.98071 | 0.9699172716 | 0.9466 | 0.9949869006 | 0.341856 | 6.433154244 | 1 |
| 10 | 87961066 | T | G | L | R | PTEN | ENST00000371953 | 0.71818 | 0.91229 | 0.92752 | 0.93473 | 0.94469 | 0.9667141015 | 0.96839 | 0.9825049043 | 0.370838 | 6.641897785 | 1 |
| 10 | 87961068 | G | A | D | N | PTEN | ENST00000371953 | 0.89359 | 0.96682 | 0.84551 | 0.97109 | 0.97885 | 0.9432884151 | 0.98715 | 0.9974943399 | 0.214912 | 5.571279531 | 0 |
| 10 | 87961068 | G | C | D | H | PTEN | ENST00000371953 | 0.88976 | 0.97869 | 0.86666 | 0.97818 | 0.98404 | 0.9596035931 | 0.96646 | 0.9971701503 | 0.476783 | 6.626073744 | 1 |
| 10 | 87961068 | G | T | D | Y | PTEN | ENST00000371953 | 0.89583 | 0.98069 | 0.85439 | 0.97598 | 0.98549 | 0.977355269 | 0.97372 | 0.9987350106 | 0.492387 | 6.649709684 | 1 |
| 10 | 87961069 | A | C | D | A | PTEN | ENST00000371953 | 0.93283 | 0.98086 | 0.79289 | 0.99024 | 0.98109 | 0.9665103728 | 0.9425 | 0.9993100166 | 0.414616 | 6.495662374 | 1 |
| 10 | 87961071 | A | C | K | Q | PTEN | ENST00000371953 | 0.78566 | 0.95858 | 0.79496 | 0.87822 | 0.86852 | 0.7185074688 | 0.92726 | 0.8793406487 | 0.122408 | 4.836984506 | 0 |
| 10 | 87961071 | A | G | K | E | PTEN | ENST00000371953 | 0.91975 | 0.98069 | 0.83413 | 0.87725 | 0.89829 | 0.7725267339 | 0.9243 | 0.9602161646 | 0.268166 | 5.727148946 | 0 |
| 10 | 87961072 | A | C | K | T | PTEN | ENST00000371953 | 0.90765 | 0.97788 | 0.84236 | 0.91453 | 0.93397 | 0.7777883295 | 0.86856 | 0.9787677149 | 0.221404 | 5.885577084 | 0 |
| 10 | 87961072 | A | G | K | R | PTEN | ENST00000371953 | 0.58803 | 0.87177 | 0.76253 | 0.82866 | 0.80058 | 0.5724842943 | 0.90797 | 0.8786813021 | 0.117674 | 4.160073357 | 0 |
| 10 | 87961072 | A | T | K | I | PTEN | ENST00000371953 | 0.83928 | 0.99392 | 0.81587 | 0.93215 | 0.93044 | 0.8196899931 | 0.88436 | 0.9905299565 | 0.220158 | 5.64986953 | 0 |
| 10 | 87961073 | A | T | K | N | PTEN | ENST00000371953 | 0.81316 | 0.9764 | 0.85434 | 0.87163 | 0.83228 | 0.7789690029 | 0.91662 | 0.9414196014 | 0.055728 | 5.250422232 | 0 |
| 10 | 87961074 | G | A | A | T | PTEN | ENST00000371953 | 0.8504 | 0.96618 | 0.83991 | 0.90396 | 0.89603 | 0.7217602147 | 0.89148 | 0.9787090481 | 0.230311 | 5.824143203 | 0 |
| 10 | 87961074 | G | C | A | P | PTEN | ENST00000371953 | 0.86481 | 0.98329 | 0.88647 | 0.88935 | 0.92028 | 0.915150322 | 0.85221 | 0.9899848279 | 0.270925 | 5.836757642 | 0 |
| 10 | 87961074 | G | T | A | S | PTEN | ENST00000371953 | 0.63842 | 0.95687 | 0.80071 | 0.89295 | 0.8233 | 0.6275535319 | 0.86635 | 0.958796978 | 0.166275 | 5.076665396 | 0 |
| 10 | 87961075 | C | A | A | E | PTEN | ENST00000371953 | 0.81308 | 0.97779 | 0.88945 | 0.9151 | 0.93824 | 0.9250341515 | 0.86863 | 0.9955285411 | 0.295171 | 5.975378797 | 0 |
| 10 | 87961075 | C | G | A | G | PTEN | ENST00000371953 | 0.81552 | 0.96871 | 0.82209 | 0.89703 | 0.87296 | 0.6494855155 | 0.85084 | 0.9808187075 | 0.162282 | 6.453281743 | 0 |
| 10 | 87961077 | A | C | N | H | PTEN | ENST00000371953 | 0.55223 | 0.90852 | 0.74328 | 0.75298 | 0.79758 | 0.7411183231 | 0.92355 | 0.896427393 | 0.077995 | 3.977456729 | 0 |
| 10 | 87961077 | A | T | N | Y | PTEN | ENST00000371953 | 0.65359 | 0.9799 | 0.77752 | 0.90661 | 0.92924 | 0.8391249301 | 0.90244 | 0.9940313907 | 0.260816 | 4.827462915 | 0 |
| 10 | 87961078 | A | C | N | T | PTEN | ENST00000371953 | 0.87775 | 0.96965 | 0.70664 | 0.89156 | 0.92275 | 0.7248395833 | 0.90587 | 0.9933428304 | 0.136334 | 5.586590622 | 0 |
| 10 | 87961078 | A | G | N | S | PTEN | ENST00000371953 | 0.83081 | 0.9503 | 0.66317 | 0.8822 | 0.86951 | 0.6114762072 | 0.92878 | 0.9824436679 | 0.0484565 | 5.353565767 | 0 |
| 10 | 87961078 | A | T | N | I | PTEN | ENST00000371953 | 0.90969 | 0.9813 | 0.73912 | 0.93573 | 0.92884 | 0.8364596428 | 0.89721 | 0.9979112277 | 0.276232 | 6.145557012 | 0 |
| 10 | 87961079 | T | A | N | K | PTEN | ENST00000371953 | 0.73301 | 0.97662 | 0.81873 | 0.88161 | 0.87539 | 0.79790894 | 0.943 | 0.9911279678 | 0.0818233 | 5.981160397 | 0 |
| 10 | 87961080 | A | C | K | Q | PTEN | ENST00000371953 | 0.90329 | 0.97311 | 0.82973 | 0.92805 | 0.94129 | 0.8314555518 | 0.95766 | 0.9924431266 | 0.183902 | 5.840976253 | 0 |
| 10 | 87961080 | A | G | K | E | PTEN | ENST00000371953 | 0.91831 | 0.98652 | 0.86193 | 0.9419 | 0.96846 | 0.8753689432 | 0.94758 | 0.9926235676 | 0.342363 | 5.968796078 | 0 |
| 10 | 87961081 | A | G | K | R | PTEN | ENST00000371953 | 0.89024 | 0.95441 | 0.79994 | 0.90767 | 0.90624 | 0.7170894379 | 0.94692 | 0.9890476846 | 0.182972 | 5.977690741 | 0 |
| 10 | 87961081 | A | T | K | I | PTEN | ENST00000371953 | 0.92198 | 0.99419 | 0.83394 | 0.95891 | 0.93475 | 0.8818789932 | 0.9297 | 0.9991361499 | 0.272537 | 7.890697131 | 0 |
| 10 | 87961082 | A | T | K | N | PTEN | ENST00000371953 | 0.88423 | 0.9783 | 0.88277 | 0.9318 | 0.90229 | 0.8705663914 | 0.95394 | 0.9930223038 | 0.140168 | 5.966725723 | 0 |
| 10 | 87961083 | G | A | D | N | PTEN | ENST00000371953 | 0.88354 | 0.96723 | 0.8835 | 0.89847 | 0.87247 | 0.7938798245 | 0.96222 | 0.9976040554 | 0.022598 | 5.379623357 | 0 |
| 10 | 87961083 | G | C | D | H | PTEN | ENST00000371953 | 0.88067 | 0.97893 | 0.90392 | 0.93115 | 0.9328 | 0.8552916976 | 0.90738 | 0.9973209328 | 0.321258 | 6.877383442 | 0 |
| 10 | 87961083 | G | T | D | Y | PTEN | ENST00000371953 | 0.88342 | 0.98086 | 0.89234 | 0.92207 | 0.94356 | 0.9218073089 | 0.94366 | 0.9984158278 | 0.414445 | 6.145514456 | 0 |
| 10 | 87961084 | A | C | D | A | PTEN | ENST00000371953 | 0.92757 | 0.98134 | 0.83846 | 0.94021 | 0.94318 | 0.8761538371 | 0.87929 | 0.9984611273 | 0.293545 | 6.092833111 | 0 |
| 10 | 87961084 | A | G | D | G | PTEN | ENST00000371953 | 0.92966 | 0.98388 | 0.84705 | 0.92151 | 0.93631 | 0.8885274008 | 0.90962 | 0.9941363335 | 0.300018 | 5.17874022 | 0 |
| 10 | 87961084 | A | T | D | V | PTEN | ENST00000371953 | 0.92367 | 0.9875 | 0.84103 | 0.95177 | 0.96153 | 0.9020627991 | 0.9362 | 0.999050796 | 0.393691 | 6.857502417 | 0 |
| 10 | 87961085 | C | A | D | E | PTEN | ENST00000371953 | 0.6489 | 0.96424 | 0.86298 | 0.8449 | 0.85534 | 0.7500856214 | 0.94705 | 0.9843381517 | 0.136344 | 4.631449203 | 0 |
| 10 | 87961086 | A | C | K | Q | PTEN | ENST00000371953 | 0.80668 | 0.97262 | 0.83098 | 0.9064 | 0.91736 | 0.7011452929 | 0.92963 | 0.9403270483 | 0.148581 | 4.471838657 | 0 |
| 10 | 87961086 | A | G | K | E | PTEN | ENST00000371953 | 0.78756 | 0.98196 | 0.8667 | 0.8754 | 0.93554 | 0.7598267003 | 0.9529 | 0.9684147239 | 0.317577 | 4.648283129 | 0 |
| 10 | 87961087 | A | G | K | R | PTEN | ENST00000371953 | 0.86254 | 0.95331 | 0.80004 | 0.80672 | 0.87877 | 0.5331132235 | 0.92292 | 0.9453144362 | 0.118825 | 4.230877883 | 0 |
| 10 | 87961088 | A | T | K | N | PTEN | ENST00000371953 | 0.72284 | 0.97803 | 0.88834 | 0.87739 | 0.83888 | 0.7568385511 | 0.92333 | 0.980057478 | 0.028544 | 4.528246624 | 0 |
| 10 | 87961089 | G | A | A | T | PTEN | ENST00000371953 | 0.50302 | 0.81277 | 0.79445 | 0.84364 | 0.71391 | 0.3856534134 | 0.82226 | 0.677295506 | 0.125516 | 3.287749862 | 0 |
| 10 | 87961094 | C | A | N | K | PTEN | ENST00000371953 | 0.53515 | 0.9764 | 0.82663 | 0.83828 | 0.80358 | 0.7707053907 | 0.92203 | 0.9813655615 | 0.056624 | 5.309941114 | 0 |
| 10 | 87961096 | G | A | R | Q | PTEN | ENST00000371953 | 0.7497 | 0.85322 | 0.78131 | 0.83571 | 0.83888 | 0.6176238449 | 0.93819 | 0.8782696851 | 0.203733 | 4.867583638 | 0 |

| | | | | | | | | | | | | | | | | | | |
|---|---|---|---|---|---|---|---|---|---|---|---|---|---|---|---|---|---|---|
| 10 | 87961102 | T | C | F | S | PTEN | ENST00000371953 | 0.94986 | 0.99392 | 0.89943 | 0.94995 | 0.99138 | 0.9168549833 | 0.97783 | 0.9962536029 | 0.447454 | 5.792078017 | 0 |
| 10 | 87961102 | T | G | F | C | PTEN | ENST00000371953 | 0.94953 | 0.98759 | 0.92168 | 0.94831 | 0.97438 | 0.9094283195 | 0.90432 | 0.996872548 | 0.342153 | 6.190123444 | 0 |
| 10 | 87961103 | T | G | F | L | PTEN | ENST00000371953 | 0.80787 | 0.9764 | 0.92354 | 0.89544 | 0.90406 | 0.8021910157 | 0.90975 | 0.9861120582 | 0.191688 | 4.857226462 | 0 |
| 10 | 87961104 | T | A | S | T | PTEN | ENST00000371953 | 0.6681 | 0.96209 | 0.84677 | 0.8775 | 0.87148 | 0.7539672353 | 0.86837 | 0.9323583523 | 0.142104 | 4.215364387 | 0 |
| 10 | 87961105 | C | A | S | Y | PTEN | ENST00000371953 | 0.88285 | 0.98273 | 0.89824 | 0.89482 | 0.93044 | 0.8396189384 | 0.89833 | 0.9960741997 | 0.305646 | 8.655723248 | 0 |
| 10 | 87961105 | C | G | S | C | PTEN | ENST00000371953 | 0.90585 | 0.96697 | 0.84824 | 0.9093 | 0.90798 | 0.801824596 | 0.87054 | 0.9942777753 | 0.271515 | 7.477763038 | 0 |
| 10 | 87961105 | C | T | S | F | PTEN | ENST00000371953 | 0.91631 | 0.97448 | 0.87964 | 0.89455 | 0.95207 | 0.8534039367 | 0.9129 | 0.9960741997 | 0.359924 | 7.449402277 | 0 |
| 10 | 87961108 | C | A | P | Q | PTEN | ENST00000371953 | 0.74101 | 0.96302 | 0.85531 | 0.90698 | 0.90972 | 0.6022353336 | 0.89734 | 0.9886056091 | 0.308015 | 4.769972783 | 0 |
| 10 | 87961110 | A | C | N | H | PTEN | ENST00000371953 | 0.80645 | 0.96407 | 0.73482 | 0.90382 | 0.92316 | 0.7889423578 | 0.9535 | 0.9407685995 | 0.162017 | 6.25100783 | 0 |
| 10 | 87961110 | A | T | N | Y | PTEN | ENST00000371953 | 0.85434 | 0.97662 | 0.77275 | 0.93925 | 0.96444 | 0.8548611187 | 0.93603 | 0.9960806966 | 0.372503 | 6.773979269 | 0 |
| 10 | 87961112 | T | A | N | K | PTEN | ENST00000371953 | 0.84876 | 0.94445 | 0.80358 | 0.84116 | 0.84267 | 0.8239731979 | 0.95446 | 0.9810500741 | 0.0797563 | 5.954527211 | 0 |
| 10 | 87961113 | T | G | F | V | PTEN | ENST00000371953 | 0.87268 | 0.97662 | 0.91544 | 0.94169 | 0.97587 | 0.8560768807 | 0.91354 | 0.9968932592 | 0.36464 | 6.024985071 | 0 |
| 10 | 87961114 | T | G | F | C | PTEN | ENST00000371953 | 0.95055 | 0.98725 | 0.94315 | 0.96552 | 0.9844 | 0.9099460415 | 0.9639 | 0.9988891482 | 0.462167 | 6.22616706 | 0 |
| 10 | 87961115 | T | G | F | L | PTEN | ENST00000371953 | 0.88982 | 0.91377 | 0.94692 | 0.93512 | 0.94281 | 0.8088220304 | 0.94249 | 0.9864311814 | 0.272825 | 4.192103746 | 0 |
| 10 | 87961117 | A | G | K | R | PTEN | ENST00000371953 | 0.63881 | 0.80441 | 0.76245 | 0.78834 | 0.81933 | 0.3867346186 | 0.91683 | 0.7879915237 | 0.13374 | 3.467623414 | 0 |
| 10 | 87961117 | A | T | K | M | PTEN | ENST00000371953 | 0.94206 | 0.98373 | 0.76856 | 0.92386 | 0.95353 | 0.6340914958 | 0.9465 | 0.9951145642 | 0.315508 | 6.077795394 | 0 |
| 10 | 87965290 | A | G | K | E | PTEN | ENST00000371953 | 0.48107 | 0.88377 | 0.85811 | 0.66522 | 0.76487 | 0.4741166659 | 0.91004 | 0.7266689539 | 0.187066 | 3.433416436 | 0 |
| 10 | 87965293 | C | A | L | M | PTEN | ENST00000371953 | 0.43562 | 0.87305 | 0.81953 | 0.68219 | 0.66639 | 0.3963498579 | 0.8026 | 0.8340976834 | 0.0767538 | 4.108525233 | 0 |
| 10 | 87965297 | A | C | Y | S | PTEN | ENST00000371953 | 0.5392 | 0.8844 | 0.59605 | 0.62262 | 0.70752 | 0.58727495 | 0.95132 | 0.5745735765 | 0.0980486 | 3.117882334 | 0 |
| 10 | 87965305 | A | C | K | Q | PTEN | ENST00000371953 | 0.56734 | 0.86635 | 0.86892 | 0.75975 | 0.74164 | 0.3148256781 | 0.90369 | 0.9139257073 | 0.152386 | 3.603284482 | 0 |
| 10 | 87965305 | A | G | K | E | PTEN | ENST00000371953 | 0.87409 | 0.90647 | 0.89339 | 0.78467 | 0.89101 | 0.3999494273 | 0.95332 | 0.9097911119 | 0.335918 | 3.354043381 | 0 |
| 10 | 87965307 | A | T | K | N | PTEN | ENST00000371953 | 0.81724 | 0.96723 | 0.91308 | 0.87932 | 0.83116 | 0.4111054249 | 0.90794 | 0.938775003 | 0.076563 | 4.049794937 | 0 |
| 10 | 87965308 | A | T | T | S | PTEN | ENST00000371953 | 0.57645 | 0.8847 | 0.85627 | 0.86383 | 0.8517 | 0.1868888983 | 0.92111 | 0.95664078 | 0.206298 | 3.842989033 | 1 |
| 10 | 87965311 | G | A | V | I | PTEN | ENST00000371953 | 0.71337 | 0.8489 | 0.60954 | 0.74432 | 0.75957 | 0.08062519285 | 0.80417 | 0.8461043239 | 0.167352 | 4.473650984 | 0 |
| 10 | 87965312 | T | A | V | E | PTEN | ENST00000371953 | 0.56602 | 0.95858 | 0.79392 | 0.87127 | 0.83172 | 0.2612643137 | 0.95401 | 0.8578307033 | 0.307918 | 2.782929679 | 0 |
| 10 | 87965312 | T | C | V | A | PTEN | ENST00000371953 | 0.68851 | 0.91879 | 0.79788 | 0.72154 | 0.81818 | 0.1743945764 | 0.86157 | 0.8661197424 | 0.218455 | 3.086749313 | 0 |
| 10 | 87965312 | T | G | V | G | PTEN | ENST00000371953 | 0.75957 | 0.86654 | 0.66532 | 0.83514 | 0.82216 | 0.3199782208 | 0.79986 | 0.8737176061 | 0.198971 | 3.454740697 | 0 |
| 10 | 87965314 | G | A | E | K | PTEN | ENST00000371953 | 0.76235 | 0.83524 | 0.85555 | 0.88352 | 0.80178 | 0.3023581272 | 0.8631 | 0.9630889893 | 0.189107 | 3.840082945 | 0 |
| 10 | 87965314 | G | C | E | Q | PTEN | ENST00000371953 | 0.71946 | 0.91769 | 0.74195 | 0.83077 | 0.77591 | 0.2352282727 | 0.89191 | 0.9346660376 | 0.199763 | 3.077901442 | 0 |
| 10 | 87965315 | A | C | E | A | PTEN | ENST00000371953 | 0.59378 | 0.84092 | 0.74694 | 0.86332 | 0.74587 | 0.2822584072 | 0.83869 | 0.8112204075 | 0.20621 | 3.935983595 | 0 |
| 10 | 87965315 | A | G | E | G | PTEN | ENST00000371953 | 0.80921 | 0.94913 | 0.70473 | 0.92397 | 0.80417 | 0.2853783315 | 0.86254 | 0.9149128199 | 0.298421 | 3.014519701 | 0 |
| 10 | 87965315 | A | T | E | V | PTEN | ENST00000371953 | 0.92147 | 0.93906 | 0.74156 | 0.92339 | 0.86852 | 0.3996098611 | 0.92006 | 0.8543508649 | 0.304587 | 3.880658847 | 0 |
| 10 | 87965316 | G | T | E | D | PTEN | ENST00000371953 | 0.58724 | 0.89821 | 0.75379 | 0.84271 | 0.74093 | 0.1302037675 | 0.8914 | 0.8860912323 | 0.14048 | 2.516256244 | 0 |
| 10 | 87965317 | G | A | E | K | PTEN | ENST00000371953 | 0.57561 | 0.868 | 0.80557 | 0.87491 | 0.74164 | 0.2875114647 | 0.84099 | 0.920112431 | 0.175512 | 3.383923307 | 0 |
| 10 | 87965317 | G | C | E | Q | PTEN | ENST00000371953 | 0.5089 | 0.80291 | 0.69628 | 0.79618 | 0.71233 | 0.2392270919 | 0.88974 | 0.5128036462 | 0.185578 | 2.879205677 | 0 |
| 10 | 87965318 | A | C | E | A | PTEN | ENST00000371953 | 0.63543 | 0.85333 | 0.70122 | 0.87226 | 0.75417 | 0.275481874 | 0.82964 | 0.7612946033 | 0.194352 | 2.514513245 | 0 |
| 10 | 87965318 | A | G | E | G | PTEN | ENST00000371953 | 0.82934 | 0.87665 | 0.65753 | 0.89675 | 0.78473 | 0.276608138 | 0.84362 | 0.9244289994 | 0.276179 | 2.7726213 | 0 |
| 10 | 87965318 | A | T | E | V | PTEN | ENST00000371953 | 0.67231 | 0.88681 | 0.67626 | 0.91028 | 0.79938 | 0.3759931016 | 0.89712 | 0.9015543461 | 0.249201 | 3.540898347 | 0 |
| 10 | 87965319 | G | T | E | D | PTEN | ENST00000371953 | 0.4246 | 0.76154 | 0.70091 | 0.67706 | 0.70345 | 0.1166542952 | 0.89559 | 0.7915587425 | 0.127429 | 1.977228749 | 0 |
| 10 | 87965320 | C | A | P | T | PTEN | ENST00000371953 | 0.37481 | 0.84342 | 0.67574 | 0.69744 | 0.70262 | 0.3144261666 | 0.95657 | 0.5534601212 | 0.17598 | 2.616596639 | 0 |
| 10 | 87965320 | C | G | P | A | PTEN | ENST00000371953 | 0.34375 | 0.79207 | 0.61496 | 0.64211 | 0.56291 | 0.2277154297 | 0.83534 | 0.6677267551 | 0.12242 | 2.190323498 | 0 |
| 10 | 87965320 | C | T | P | S | PTEN | ENST00000371953 | 0.37566 | 0.79515 | 0.63035 | 0.62405 | 0.51843 | 0.2976199411 | 0.6901 | 0.4567449093 | 0.0366759 | 2.215931696 | 0 |
| 10 | 87965321 | C | A | P | Q | PTEN | ENST00000371953 | 0.43496 | 0.7802 | 0.64182 | 0.80765 | 0.78411 | 0.2785142833 | 0.96911 | 0.110358789 | 0.123024 | 3.304082234 | 0 |
| 10 | 87965321 | C | G | P | R | PTEN | ENST00000371953 | 0.43016 | 0.88976 | 0.71104 | 0.85892 | 0.76157 | 0.310187635 | 0.91143 | 0.4586499668 | 0.247901 | 2.316334938 | 0 |
| 10 | 87965321 | C | T | P | L | PTEN | ENST00000371953 | 0.4415 | 0.82952 | 0.72381 | 0.83262 | 0.71861 | 0.2684918759 | 0.87975 | 0.8101576567 | 0.162855 | 3.114397031 | 0 |
| 10 | 87965323 | T | A | S | T | PTEN | ENST00000371953 | 0.37873 | 0.76326 | 0.70912 | 0.63562 | 0.60002 | 0.2019403635 | 0.78269 | 0.5053884983 | 0.0790059 | 1.743168502 | 0 |
| 10 | 87965323 | T | C | S | P | PTEN | ENST00000371953 | 0.50017 | 0.85459 | 0.72479 | 0.76029 | 0.74936 | 0.2308986594 | 0.92714 | 0.7430360317 | 0.187941 | 2.143181971 | 0 |
| 10 | 87965323 | T | G | S | A | PTEN | ENST00000371953 | 0.4001 | 0.73244 | 0.68377 | 0.61661 | 0.62767 | 0.1146458587 | 0.82176 | 0.6638429761 | 0.11062 | 1.158145437 | 0 |
| 10 | 87965324 | C | T | S | L | PTEN | ENST00000371953 | 0.51705 | 0.7725 | 0.72693 | 0.76618 | 0.67108 | 0.2260350881 | 0.93572 | 0.9052033424 | 0.192208 | 2.903831285 | 0 |
| 10 | 87965326 | A | C | N | H | PTEN | ENST00000371953 | 0.55298 | 0.81024 | 0.78573 | 0.87853 | 0.62876 | 0.2724245391 | 0.92676 | 0.3928866982 | 0.119582 | 4.222999403 | 0 |
| 10 | 87965326 | A | G | N | D | PTEN | ENST00000371953 | 0.47143 | 0.87792 | 0.82472 | 0.81791 | 0.70345 | 0.2446137422 | 0.89194 | 0.4151570201 | 0.10606 | 3.111001502 | 0 |
| 10 | 87965326 | A | T | N | Y | PTEN | ENST00000371953 | 0.62574 | 0.94445 | 0.81843 | 0.91508 | 0.77011 | 0.3940457985 | 0.96468 | 0.9609924555 | 0.310107 | 3.846417533 | 0 |
| 10 | 87965327 | A | C | N | T | PTEN | ENST00000371953 | 0.50017 | 0.7985 | 0.75768 | 0.87044 | 0.5976 | 0.2412784114 | 0.86249 | 0.7681484222 | 0.148995 | 3.082583677 | 0 |
| 10 | 87965327 | A | G | N | S | PTEN | ENST00000371953 | 0.43635 | 0.7218 | 0.70974 | 0.76814 | 0.62767 | 0.183494173 | 0.86118 | 0.7065964937 | 0.0289951 | 2.747592062 | 0 |
| 10 | 87965327 | A | T | N | I | PTEN | ENST00000371953 | 0.67421 | 0.90115 | 0.78025 | 0.91183 | 0.76024 | 0.4493882259 | 0.947 | 0.9160073996 | 0.295928 | 3.229602198 | 0 |
| 10 | 87965329 | C | A | P | T | PTEN | ENST00000371953 | 0.33731 | 0.82287 | 0.63736 | 0.69496 | 0.64775 | 0.2930924106 | 0.78153 | 0.2738221586 | 0.124331 | 1.098248238 | 0 |
| 10 | 87965329 | C | G | P | A | PTEN | ENST00000371953 | 0.32104 | 0.78365 | 0.61476 | 0.64923 | 0.58518 | 0.2007201948 | 0.67855 | 0.7011523843 | 0.0564374 | 1.471508783 | 0 |
| 10 | 87965330 | C | A | P | Q | PTEN | ENST00000371953 | 0.43346 | 0.78402 | 0.62725 | 0.74974 | 0.74021 | 0.2335971212 | 0.73309 | 0.7106030583 | 0.110652 | 2.103458057 | 0 |
| 10 | 87965330 | C | G | P | R | PTEN | ENST00000371953 | 0.43768 | 0.80811 | 0.70856 | 0.76323 | 0.74164 | 0.3354662416 | 0.66328 | 0.7147506475 | 0.109619 | 1.443168927 | 0 |
| 10 | 87965330 | C | T | P | L | PTEN | ENST00000371953 | 0.41222 | 0.80377 | 0.71101 | 0.8251 | 0.69082 | 0.2876575203 | 0.75402 | 0.7389445305 | 0.112506 | 1.414808567 | 0 |
| 10 | 87965332 | G | A | E | K | PTEN | ENST00000371953 | 0.54936 | 0.84646 | 0.85304 | 0.82966 | 0.74658 | 0.3415389924 | 0.94443 | 0.9263256788 | 0.297612 | 2.946769188 | 0 |
| 10 | 87965332 | G | C | E | Q | PTEN | ENST00000371953 | 0.51632 | 0.86318 | 0.73997 | 0.76404 | 0.72322 | 0.3114573244 | 0.86614 | 0.9199525118 | 0.180634 | 3.168354702 | 0 |
| 10 | 87965333 | A | C | E | A | PTEN | ENST00000371953 | 0.58796 | 0.8126 | 0.75279 | 0.73779 | 0.75957 | 0.3328757448 | 0.86847 | 0.855324924 | 0.217095 | 2.695409301 | 0 |
| 10 | 87965333 | A | G | E | G | PTEN | ENST00000371953 | 0.6426 | 0.83669 | 0.70038 | 0.80635 | 0.7562 | 0.2891319988 | 0.89028 | 0.9317396879 | 0.288889 | 2.826547388 | 0 |
| 10 | 87965333 | A | T | E | V | PTEN | ENST00000371953 | 0.80803 | 0.87384 | 0.73292 | 0.83893 | 0.81006 | 0.4226563896 | 0.94331 | 0.9247080088 | 0.309576 | 2.890740279 | 0 |
| 10 | 87965334 | G | T | E | D | PTEN | ENST00000371953 | 0.42615 | 0.76193 | 0.75625 | 0.65566 | 0.65077 | 0.1827830919 | 0.83674 | 0.7566874027 | 0.0856947 | 1.35659254 | 0 |
| 10 | 87965335 | G | A | A | T | PTEN | ENST00000371953 | 0.70398 | 0.78688 | 0.75402 | 0.84022 | 0.7255 | 0.2819736085 | 0.8067 | 0.9033406377 | 0.173605 | 2.727082469 | 0 |
| 10 | 87965335 | G | C | A | P | PTEN | ENST00000371953 | 0.86297 | 0.96332 | 0.80741 | 0.84346 | 0.74164 | 0.3802394689 | 0.79982 | 0.8928703666 | 0.242852 | 2.959954257 | 0 |
| 10 | 87965335 | G | T | A | S | PTEN | ENST00000371953 | 0.67477 | 0.78178 | 0.70283 | 0.8281 | 0.73877 | 0.2615832922 | 0.8302 | 0.8349680901 | 0.174805 | 1.856984031 | 0 |
| 10 | 87965336 | C | A | A | D | PTEN | ENST00000371953 | 0.83329 | 0.94339 | 0.84789 | 0.89587 | 0.79758 | 0.4396757632 | 0.83639 | 0.946336627 | 0.272512 | 3.984752494 | 0 |
| 10 | 87965336 | C | G | A | G | PTEN | ENST00000371953 | 0.68483 | 0.88116 | 0.72715 | 0.85375 | 0.74021 | 0.2699368155 | 0.83012 | 0.8359998465 | 0.153379 | 3.669669814 | 0 |
| 10 | 87965336 | C | T | A | V | PTEN | ENST00000371953 | 0.67152 | 0.86329 | 0.76911 | 0.86827 | 0.75349 | 0.3393689115 | 0.81767 | 0.9262815118 | 0.120219 | 3.438570564 | 0 |
| 10 | 87965338 | A | G | S | G | PTEN | ENST00000371953 | 0.76618 | 0.93674 | 0.77258 | 0.86964 | 0.74728 | 0.3416853372 | 0.86739 | 0.3957398571 | 0.0792237 | 3.650464505 | 0 |
| 10 | 87965338 | A | T | S | C | PTEN | ENST00000371953 | 0.677 | 0.96587 | 0.82331 | 0.89841 | 0.77591 | 0.4457334347 | 0.86719 | 0.9294776917 | 0.225625 | 4.642439813 | 0 |
| 10 | 87965339 | G | A | S | N | PTEN | ENST00000371953 | 0.77029 | 0.93298 | 0.86017 | 0.82606 | 0.73661 | 0.3742268012 | 0.85326 | 0.9141686559 | 0.116111 | 4.128870104 | 0 |
| 10 | 87965339 | G | C | S | T | PTEN | ENST00000371953 | 0.74324 | 0.93612 | 0.82067 | 0.81206 | 0.7337 | 0.3393361777 | 0.84456 | 0.9166113734 | 0.174634 | 4.033960912 | 0 |
| 10 | 87965339 | G | T | S | I | PTEN | ENST00000371953 | 0.9344 | 0.97662 | 0.83686 | 0.82841 | 0.78597 | 0.5543297468 | 0.86118 | 0.9284473062 | 0.305267 | 4.674532721 | 0 |
| 10 | 87965340 | C | A | S | R | PTEN | ENST00000371953 | 0.67589 | 0.97175 | 0.86516 | 0.8751 | 0.73661 | 0.5304748311 | 0.82786 | 0.9272546172 | 0.245684 | 4.096621814 | 0 |
| 10 | 87965341 | A | G | S | G | PTEN | ENST00000371953 | 0.52865 | 0.78217 | 0.64638 | 0.70013 | 0.70097 | 0.1622258982 | 0.85802 | 0.8515467048 | 0.126026 | 1.989371274 | 0 |
| 10 | 87965341 | A | T | S | C | PTEN | ENST00000371953 | 0.71971 | 0.78355 | 0.71046 | 0.78742 | 0.71548 | 0.249003645 | 0.78721 | 0.9063662291 | 0.132327 | 3.42574935 | 0 |
| 10 | 87965342 | G | A | S | N | PTEN | ENST00000371953 | 0.504 | 0.79519 | 0.75045 | 0.69885 | 0.73661 | 0.1661907305 | 0.81768 | 0.7935962081 | 0.0392831 | 1.853126572 | 0 |
| 10 | 87965342 | G | C | S | T | PTEN | ENST00000371953 | 0.44997 | 0.76326 | 0.70588 | 0.5718 | 0.65869 | 0.1628683858 | 0.67085 | 0.5425492525 | 0.0156934 | -0.3743010186 | 0 |
| 10 | 87965342 | G | T | S | I | PTEN | ENST00000371953 | 0.69925 | 0.85721 | 0.71639 | 0.75165 | 0.74517 | 0.3444276872 | 0.76926 | 0.8681536317 | 0.162932 | 3.214589362 | 0 |
| 10 | 87965343 | T | A | S | R | PTEN | ENST00000371953 | 0.51132 | 0.88749 | 0.75809 | 0.75794 | 0.71391 | 0.2765054337 | 0.84815 | 0.8309020996 | 0.137154 | 2.110535666 | 0 |
| 10 | 87965344 | T | A | S | T | PTEN | ENST00000371953 | 0.38776 | 0.76552 | 0.87918 | 0.85993 | 0.59393 | 0.3482358465 | 0.84123 | 0.5602980852 | 0.148303 | 3.44269265 | 0 |
| 10 | 87965344 | T | C | S | P | PTEN | ENST00000371953 | 0.48812 | 0.85756 | 0.89646 | 0.88118 | 0.68821 | 0.3685650745 | 0.94652 | 0.873599112 | 0.223762 | 3.391925659 | 0 |
| 10 | 87965344 | T | G | S | A | PTEN | ENST00000371953 | 0.4381 | 0.79035 | 0.87165 | 0.8056 | 0.67478 | 0.2745405962 | 0.86953 | 0.7976073027 | 0.170464 | 3.001432043 | 0 |
| 10 | 87965345 | C | T | S | L | PTEN | ENST00000371953 | 0.63712 | 0.84422 | 0.89682 | 0.88365 | 0.75822 | 0.3766913068 | 0.96187 | 0.7173374891 | 0.296113 | 3.304514551 | 0 |
| 10 | 87965347 | A | C | T | P | PTEN | ENST00000371953 | 0.48506 | 0.86159 | 0.6266 | 0.8085 | 0.65967 | 0.256883183 | 0.97431 | 0.8542055488 | 0.26852 | 2.597856562 | 0 |
| 10 | 87965347 | A | G | T | A | PTEN | ENST00000371953 | 0.44011 | 0.78576 | 0.62983 | 0.69179 | 0.66828 | 0.2787315765 | 0.95858 | 0.3206516504 | 0.134414 | 1.252393642 | 0 |
| 10 | 87965347 | A | T | T | S | PTEN | ENST00000371953 | 0.39823 | 0.75868 | 0.64654 | 0.58269 | 0.59881 | 0.1701198683 | 0.85505 | 0.9217122197 | 0.139477 | 1.633695776 | 0 |
| 10 | 87965348 | C | A | T | N | PTEN | ENST00000371953 | 0.37216 | 0.84885 | 0.73541 | 0.73244 | 0.73516 | 0.2855260121 | 0.80089 | 0.2752990749 | 0.143881 | 2.628748691 | 0 |
| 10 | 87965348 | C | T | T | I | PTEN | ENST00000371953 | 0.41505 | 0.81454 | 0.79809 | 0.77767 | 0.76289 | 0.3493880689 | 0.91235 | 0.3420790572 | 0.187208 | 3.436014045 | 0 |
| 10 | 87965350 | T | A | S | T | PTEN | ENST00000371953 | 0.51028 | 0.82937 | 0.87332 | 0.85398 | 0.72775 | 0.3401794057 | 0.86072 | 0.8734192252 | 0.160808 | 3.962631239 | 0 |
| 10 | 87965350 | T | C | S | P | PTEN | ENST00000371953 | 0.58624 | 0.8556 | 0.88904 | 0.87137 | 0.68821 | 0.3248475898 | 0.94027 | 0.8122804761 | 0.266484 | 3.877430141 | 0 |
| 10 | 87965350 | T | G | S | A | PTEN | ENST00000371953 | 0.54828 | 0.7773 | 0.86588 | 0.81345 | 0.71391 | 0.2525473728 | 0.87704 | 0.8815540075 | 0.181325 | 3.310406967 | 0 |
| 10 | 87965351 | C | A | S | Y | PTEN | ENST00000371953 | 0.54338 | 0.96675 | 0.9213 | 0.90818 | 0.78473 | 0.4694928621 | 0.87276 | 0.8946756721 | 0.258123 | 4.922971458 | 0 |
| 10 | 87965351 | C | G | S | C | PTEN | ENST00000371953 | 0.71289 | 0.94741 | 0.88457 | 0.9117 | 0.74235 | 0.4544287275 | 0.82698 | 0.7012683153 | 0.16076 | 7.553489184 | 0 |
| 10 | 87965351 | C | T | S | F | PTEN | ENST00000371953 | 0.59857 | 0.95327 | 0.90459 | 0.91277 | 0.7872 | 0.4410791969 | 0.8921 | 0.9788662791 | 0.276951 | 5.038015313 | 0 |
| 10 | 87965353 | G | A | V | I | PTEN | ENST00000371953 | 0.54603 | 0.74455 | 0.80199 | 0.74299 | 0.74728 | 0.1194139692 | 0.77732 | 0.3691926599 | 0.153292 | 3.280087496 | 0 |
| 10 | 87965353 | G | C | V | L | PTEN | ENST00000371953 | 0.54501 | 0.78709 | 0.93375 | 0.82316 | 0.6847 | 0.2452049327 | 0.80013 | 0.8164522648 | 0.164471 | 2.323283402 | 0 |

| | | | | | | | | | | | | | | | | | | |
|---|---|---|---|---|---|---|---|---|---|---|---|---|---|---|---|---|---|---|
| 10 | 87965353 | G | T | V | L | PTEN | ENST00000371953 | 0.53947 | 0.78709 | 0.93375 | 0.82316 | 0.68381 | 0.2452049327 | 0.79937 | 0.8164522648 | 0.164467 | 2.323283402 | 0 |
| 10 | 87965354 | T | A | V | E | PTEN | ENST00000371953 | 0.4566 | 0.92089 | 0.93442 | 0.82311 | 0.7337 | 0.4583785494 | 0.90913 | 0.692943275 | 0.274166 | 3.157257427 | 0 |
| 10 | 87965354 | T | C | V | A | PTEN | ENST00000371953 | 0.4334 | 0.85511 | 0.9433 | 0.65157 | 0.74797 | 0.2864739137 | 0.84766 | 0.5312334895 | 0.170868 | 3.266984064 | 0 |
| 10 | 87965354 | T | G | V | G | PTEN | ENST00000371953 | 0.50302 | 0.86259 | 0.85333 | 0.83219 | 0.60952 | 0.4762975847 | 0.67054 | 0.6262210011 | 0.142473 | 3.491003584 | 0 |
| 10 | 87965356 | A | C | T | P | PTEN | ENST00000371953 | 0.73565 | 0.95159 | 0.86121 | 0.89513 | 0.78223 | 0.2981203529 | 0.89907 | 0.8902409673 | 0.276706 | 4.096910802 | 0 |
| 10 | 87965356 | A | G | T | A | PTEN | ENST00000371953 | 0.42591 | 0.78709 | 0.86692 | 0.81276 | 0.67933 | 0.3545446052 | 0.90709 | 0.7828607559 | 0.191231 | 3.571476809 | 0 |
| 10 | 87965356 | A | T | T | S | PTEN | ENST00000371953 | 0.39731 | 0.76434 | 0.87878 | 0.75476 | 0.72398 | 0.2060966545 | 0.82621 | 0.8613105416 | 0.154556 | 2.967537464 | 0 |
| 10 | 87965357 | C | A | T | K | PTEN | ENST00000371953 | 0.55731 | 0.89315 | 0.93703 | 0.87184 | 0.77845 | 0.5090811021 | 0.89039 | 0.7724126577 | 0.281192 | 3.815290713 | 0 |
| 10 | 87965357 | C | G | T | R | PTEN | ENST00000371953 | 0.56387 | 0.92622 | 0.92811 | 0.9019 | 0.7675 | 0.5050097922 | 0.89389 | 0.8089766502 | 0.287871 | 4.145312607 | 0 |
| 10 | 87965357 | C | T | T | I | PTEN | ENST00000371953 | 0.43303 | 0.87051 | 0.94914 | 0.8837 | 0.7816 | 0.3862557998 | 0.88585 | 0.8002746105 | 0.273374 | 4.211164906 | 0 |
| 10 | 87965359 | C | A | P | T | PTEN | ENST00000371953 | 0.38089 | 0.9155 | 0.94835 | 0.88449 | 0.80118 | 0.406665244 | 0.89813 | 0.7898840308 | 0.27058 | 3.580864097 | 0 |
| 10 | 87965359 | C | G | P | A | PTEN | ENST00000371953 | 0.35394 | 0.85721 | 0.92264 | 0.8504 | 0.72398 | 0.319452126 | 0.79845 | 0.817299068 | 0.170618 | 3.532683739 | 0 |
| 10 | 87965360 | C | A | P | Q | PTEN | ENST00000371953 | 0.59798 | 0.94168 | 0.93442 | 0.88938 | 0.78906 | 0.4042804176 | 0.87374 | 0.8873098493 | 0.256398 | 3.486176859 | 0 |
| 10 | 87965360 | C | G | P | R | PTEN | ENST00000371953 | 0.63206 | 0.95441 | 0.96415 | 0.89248 | 0.81298 | 0.4450057732 | 0.9357 | 0.9060242772 | 0.352021 | 3.99652933 | 0 |
| 10 | 87965360 | C | T | P | L | PTEN | ENST00000371953 | 0.52234 | 0.93375 | 0.96624 | 0.89044 | 0.81646 | 0.4053803652 | 0.92756 | 0.9506920576 | 0.294519 | 3.743559764 | 0 |
| 10 | 87965362 | G | A | D | N | PTEN | ENST00000371953 | 0.72965 | 0.95933 | 0.9592 | 0.89964 | 0.74728 | 0.3505419577 | 0.90075 | 0.8621680737 | 0.0686211 | 3.36702606 | 0 |
| 10 | 87965362 | G | C | D | H | PTEN | ENST00000371953 | 0.87248 | 0.97769 | 0.97008 | 0.90981 | 0.79577 | 0.4280762832 | 0.92847 | 0.9447489381 | 0.317891 | 4.798531356 | 0 |
| 10 | 87965362 | G | T | D | Y | PTEN | ENST00000371953 | 0.88067 | 0.97972 | 0.96164 | 0.91639 | 0.84747 | 0.5552321765 | 0.96435 | 0.9707068801 | 0.357871 | 8.118113467 | 0 |
| 10 | 87965363 | A | C | D | A | PTEN | ENST00000371953 | 0.75344 | 0.96791 | 0.92547 | 0.88922 | 0.84321 | 0.5112789868 | 0.87342 | 0.8807720542 | 0.286746 | 4.361468158 | 0 |
| 10 | 87965363 | A | G | D | G | PTEN | ENST00000371953 | 0.57477 | 0.87909 | 0.93446 | 0.83637 | 0.73443 | 0.4595580936 | 0.97016 | 0.8842650056 | 0.315181 | 3.864999483 | 0 |
| 10 | 87965364 | T | A | D | E | PTEN | ENST00000371953 | 0.54008 | 0.95256 | 0.94794 | 0.83946 | 0.7872 | 0.2119824222 | 0.91956 | 0.6091895103 | 0.149128 | 4.101248625 | 0 |
| 10 | 87965365 | G | A | V | I | PTEN | ENST00000371953 | 0.50655 | 0.72679 | 0.72072 | 0.79502 | 0.72926 | 0.09102668928 | 0.83096 | 0.7957395911 | 0.179719 | 2.943442082 | 0 |
| 10 | 87965365 | G | C | V | L | PTEN | ENST00000371953 | 0.54889 | 0.80201 | 0.87609 | 0.84302 | 0.73074 | 0.2054353832 | 0.92283 | 0.6922671795 | 0.251681 | 3.020847261 | 0 |
| 10 | 87965365 | G | T | V | F | PTEN | ENST00000371953 | 0.89405 | 0.92269 | 0.85302 | 0.89442 | 0.78035 | 0.3642947731 | 0.91007 | 0.9206647873 | 0.29815 | 4.508479025 | 0 |
| 10 | 87965366 | T | A | V | D | PTEN | ENST00000371953 | 0.71626 | 0.94533 | 0.89403 | 0.8792 | 0.79151 | 0.4824832717 | 0.97642 | 0.837433219 | 0.368052 | 3.543879994 | 0 |
| 10 | 87965366 | T | C | V | A | PTEN | ENST00000371953 | 0.5001 | 0.84342 | 0.88707 | 0.72642 | 0.67933 | 0.2051987556 | 0.97497 | 0.4464616477 | 0.276917 | 3.20296856 | 0 |
| 10 | 87965366 | T | G | V | G | PTEN | ENST00000371953 | 0.53535 | 0.86306 | 0.7761 | 0.82383 | 0.7147 | 0.3661052097 | 0.97888 | 0.6711878777 | 0.307871 | 3.877765021 | 0 |
| 10 | 87965368 | A | G | S | G | PTEN | ENST00000371953 | 0.45832 | 0.76973 | 0.89058 | 0.85173 | 0.66449 | 0.2967357443 | 0.94973 | 0.7758143544 | 0.198422 | 4.376838857 | 0 |
| 10 | 87965368 | A | T | S | C | PTEN | ENST00000371953 | 0.81253 | 0.95915 | 0.92548 | 0.91212 | 0.80118 | 0.4259069734 | 0.91731 | 0.9158478975 | 0.294157 | 5.101920019 | 0 |
| 10 | 87965369 | G | A | S | N | PTEN | ENST00000371953 | 0.73133 | 0.9155 | 0.94834 | 0.88637 | 0.71391 | 0.2899291489 | 0.86752 | 0.8117592335 | 0.130007 | 4.223889618 | 0 |
| 10 | 87965369 | G | C | S | T | PTEN | ENST00000371953 | 0.59089 | 0.89831 | 0.92329 | 0.88045 | 0.7217 | 0.3039897055 | 0.88205 | 0.7711013556 | 0.18831 | 4.763700878 | 0 |
| 10 | 87965369 | G | T | S | I | PTEN | ENST00000371953 | 0.9308 | 0.9713 | 0.93021 | 0.88829 | 0.80595 | 0.5668981844 | 0.92672 | 0.9377455115 | 0.335335 | 4.78042311 | 0 |
| 10 | 87965370 | T | A | S | R | PTEN | ENST00000371953 | 0.78891 | 0.96362 | 0.95276 | 0.9055 | 0.81298 | 0.4790170886 | 0.96243 | 0.8347071409 | 0.306989 | 3.762576105 | 0 |
| 10 | 87965371 | G | A | D | N | PTEN | ENST00000371953 | 0.7426 | 0.9671 | 0.93946 | 0.89465 | 0.72246 | 0.4461909085 | 0.91172 | 0.9332023263 | 0.0558039 | 3.814949548 | 0 |
| 10 | 87965371 | G | T | D | Y | PTEN | ENST00000371953 | 0.89131 | 0.98086 | 0.93957 | 0.91694 | 0.83339 | 0.6272866906 | 0.96413 | 0.9726886749 | 0.396112 | 6.135398791 | 0 |
| 10 | 87965372 | A | C | D | A | PTEN | ENST00000371953 | 0.84839 | 0.9811 | 0.89522 | 0.9311 | 0.77205 | 0.5765121291 | 0.88209 | 0.9447928667 | 0.312361 | 4.512090458 | 0 |
| 10 | 87965372 | A | G | D | G | PTEN | ENST00000371953 | 0.8389 | 0.98388 | 0.90595 | 0.92615 | 0.80178 | 0.5033440203 | 0.94752 | 0.9420247674 | 0.344946 | 3.725083377 | 0 |
| 10 | 87965372 | A | T | D | V | PTEN | ENST00000371953 | 0.93432 | 0.9875 | 0.89792 | 0.9496 | 0.85689 | 0.6033219508 | 0.96544 | 0.9630534649 | 0.367999 | 5.332754113 | 0 |
| 10 | 87965373 | C | A | D | E | PTEN | ENST00000371953 | 0.75766 | 0.96387 | 0.92175 | 0.81653 | 0.71153 | 0.2673836932 | 0.91798 | 0.8271698356 | 0.230931 | 3.647438692 | 0 |
| 10 | 87965374 | A | G | N | D | PTEN | ENST00000371953 | 0.49958 | 0.92711 | 0.86286 | 0.84139 | 0.73 | 0.2321318005 | 0.88822 | 0.8585134745 | 0.130302 | 3.010614066 | 0 |
| 10 | 87965374 | A | T | N | Y | PTEN | ENST00000371953 | 0.78915 | 0.95965 | 0.84827 | 0.90395 | 0.80178 | 0.4271860409 | 0.95206 | 0.9384793639 | 0.326764 | 5.328428198 | 0 |
| 10 | 87965375 | A | C | N | T | PTEN | ENST00000371953 | 0.70278 | 0.86788 | 0.77786 | 0.84911 | 0.73074 | 0.2951622246 | 0.88323 | 0.8695832491 | 0.175393 | 2.94596692 | 0 |
| 10 | 87965375 | A | G | N | S | PTEN | ENST00000371953 | 0.52334 | 0.8525 | 0.73266 | 0.78272 | 0.69594 | 0.1805578855 | 0.86623 | 0.4860972762 | 0.0427702 | 2.668208571 | 0 |
| 10 | 87965375 | A | T | N | I | PTEN | ENST00000371953 | 0.74189 | 0.91631 | 0.81618 | 0.91222 | 0.80772 | 0.4823496927 | 0.97611 | 0.9398780465 | 0.333339 | 3.8993887 | 0 |
| 10 | 87965376 | T | A | N | K | PTEN | ENST00000371953 | 0.63881 | 0.93962 | 0.88198 | 0.85664 | 0.73805 | 0.3754050233 | 0.88748 | 0.7706554532 | 0.138724 | 3.71079844 | 0 |
| 10 | 87965377 | G | A | E | K | PTEN | ENST00000371953 | 0.79096 | 0.9756 | 0.96512 | 0.91115 | 0.81588 | 0.4142102286 | 0.94714 | 0.9676092267 | 0.33444 | 3.947944954 | 0 |
| 10 | 87965377 | G | C | E | Q | PTEN | ENST00000371953 | 0.71674 | 0.96791 | 0.90359 | 0.88747 | 0.73516 | 0.3201935447 | 0.90343 | 0.9166442156 | 0.213085 | 3.859830186 | 0 |
| 10 | 87965378 | A | C | E | A | PTEN | ENST00000371953 | 0.93146 | 0.97243 | 0.91522 | 0.93775 | 0.78906 | 0.3568997689 | 0.93617 | 0.928560257 | 0.324371 | 3.791552922 | 0 |
| 10 | 87965378 | A | G | E | G | PTEN | ENST00000371953 | 0.94589 | 0.97822 | 0.88088 | 0.93936 | 0.78658 | 0.3677998181 | 0.91357 | 0.9372981787 | 0.33324 | 4.188203697 | 0 |
| 10 | 87965378 | A | T | E | V | PTEN | ENST00000371953 | 0.86734 | 0.97803 | 0.89884 | 0.93653 | 0.79697 | 0.4627144422 | 0.94605 | 0.9219557643 | 0.341325 | 3.9783192 | 0 |
| 10 | 87965379 | A | T | E | D | PTEN | ENST00000371953 | 0.67748 | 0.96121 | 0.91307 | 0.83016 | 0.75143 | 0.1580685744 | 0.91544 | 0.7603652477 | 0.179627 | 3.17188711 | 0 |
| 10 | 87965380 | C | A | P | T | PTEN | ENST00000371953 | 0.67795 | 0.95887 | 0.8984 | 0.90349 | 0.76881 | 0.3373514963 | 0.87588 | 0.9226456881 | 0.275634 | 2.678739005 | 0 |
| 10 | 87965380 | C | G | P | A | PTEN | ENST00000371953 | 0.52759 | 0.91099 | 0.85901 | 0.89537 | 0.76619 | 0.2435896179 | 0.81791 | 0.7534162998 | 0.201715 | 3.095863681 | 0 |
| 10 | 87965380 | C | T | P | S | PTEN | ENST00000371953 | 0.57336 | 0.95497 | 0.87086 | 0.89936 | 0.7609 | 0.3070401412 | 0.83562 | 0.9001399875 | 0.200489 | 2.859597336 | 0 |
| 10 | 87965381 | C | A | P | H | PTEN | ENST00000371953 | 0.86311 | 0.86747 | 0.90867 | 0.89307 | 0.76487 | 0.2928469969 | 0.8525 | 0.8444193602 | 0.251923 | 4.020767226 | 0 |
| 10 | 87965381 | C | G | P | R | PTEN | ENST00000371953 | 0.86524 | 0.96332 | 0.92426 | 0.89571 | 0.79818 | 0.380852335 | 0.90535 | 0.8100438118 | 0.339641 | 4.135706968 | 0 |
| 10 | 87965381 | C | T | P | L | PTEN | ENST00000371953 | 0.74101 | 0.80885 | 0.92313 | 0.81773 | 0.77845 | 0.3268384269 | 0.88729 | 0.6218206286 | 0.274255 | 2.968472164 | 0 |
| 10 | 87965383 | G | A | D | N | PTEN | ENST00000371953 | 0.76634 | 0.79578 | 0.90191 | 0.90186 | 0.74797 | 0.3585439478 | 0.91263 | 0.9016844034 | 0.0704068 | 4.112750076 | 0 |
| 10 | 87965383 | G | C | D | H | PTEN | ENST00000371953 | 0.89095 | 0.97114 | 0.92459 | 0.9061 | 0.80118 | 0.4441284206 | 0.93413 | 0.9125632644 | 0.35606 | 4.526148736 | 0 |
| 10 | 87965383 | G | T | D | Y | PTEN | ENST00000371953 | 0.89801 | 0.97734 | 0.90687 | 0.91518 | 0.85894 | 0.575130798 | 0.97127 | 0.962223649 | 0.399208 | 6.08036699 | 0 |
| 10 | 87965384 | A | C | D | A | PTEN | ENST00000371953 | 0.9381 | 0.93242 | 0.84358 | 0.93384 | 0.81703 | 0.5145755383 | 0.90776 | 0.9442052245 | 0.33049 | 3.878390373 | 0 |
| 10 | 87965384 | A | G | D | G | PTEN | ENST00000371953 | 0.93943 | 0.93938 | 0.8611 | 0.92856 | 0.82836 | 0.472214831 | 0.9431 | 0.9376121759 | 0.351626 | 3.731550764 | 0 |
| 10 | 87965384 | A | T | D | V | PTEN | ENST00000371953 | 0.9359 | 0.98243 | 0.84673 | 0.94997 | 0.88354 | 0.5487564164 | 0.97255 | 0.9824638963 | 0.382383 | 5.305811483 | 0 |
| 10 | 87965385 | T | A | D | E | PTEN | ENST00000371953 | 0.79508 | 0.88583 | 0.87807 | 0.84112 | 0.79089 | 0.1776336615 | 0.91462 | 0.8370096087 | 0.188984 | 3.659842349 | 0 |
| 10 | 87965386 | C | G | H | D | PTEN | ENST00000371953 | 0.59995 | 0.87637 | 0.87957 | 0.85725 | 0.8356 | 0.4562213444 | 0.94875 | 0.7753748298 | 0.434278 | 3.403190582 | 0 |
| 10 | 87965386 | C | T | H | Y | PTEN | ENST00000371953 | 0.6962 | 0.93449 | 0.89419 | 0.90334 | 0.83116 | 0.3509236494 | 0.91528 | 0.956709981 | 0.332547 | 6.375977734 | 0 |
| 10 | 87965387 | A | C | H | P | PTEN | ENST00000371953 | 0.78336 | 0.96803 | 0.81811 | 0.91729 | 0.87829 | 0.4364932771 | 0.94362 | 0.9317550063 | 0.428538 | 4.355596981 | 0 |
| 10 | 87965387 | A | G | H | R | PTEN | ENST00000371953 | 0.86838 | 0.8835 | 0.8941 | 0.85006 | 0.87974 | 0.3683906092 | 0.94867 | 0.9388087392 | 0.342503 | 3.978292536 | 0 |
| 10 | 87965387 | A | T | H | L | PTEN | ENST00000371953 | 0.9074 | 0.89711 | 0.85877 | 0.94134 | 0.88776 | 0.4185671773 | 0.92551 | 0.9671525359 | 0.32366 | 3.762884409 | 0 |
| 10 | 87965388 | T | A | H | Q | PTEN | ENST00000371953 | 0.62962 | 0.79554 | 0.86512 | 0.81373 | 0.86302 | 0.3040322305 | 0.9446 | 0.8014551401 | 0.278781 | 3.363134135 | 0 |
| 10 | 87965389 | T | A | Y | N | PTEN | ENST00000371953 | 0.73853 | 0.94989 | 0.97058 | 0.90264 | 0.80058 | 0.3082543608 | 0.94004 | 0.7499366403 | 0.346326 | 3.668183935 | 0 |
| 10 | 87965389 | T | C | Y | H | PTEN | ENST00000371953 | 0.70174 | 0.86389 | 0.97445 | 0.8449 | 0.77011 | 0.2350969934 | 0.87036 | 0.5428085578 | 0.279937 | 4.531765711 | 0 |
| 10 | 87965389 | T | G | Y | D | PTEN | ENST00000371953 | 0.86983 | 0.97893 | 0.96619 | 0.93516 | 0.82668 | 0.3546670435 | 0.95527 | 0.8836153746 | 0.37556 | 3.155881008 | 0 |
| 10 | 87965390 | A | C | Y | S | PTEN | ENST00000371953 | 0.804 | 0.94104 | 0.93003 | 0.9021 | 0.80713 | 0.3203060265 | 0.94032 | 0.8460598588 | 0.332541 | 2.482316064 | 0 |
| 10 | 87965390 | A | G | Y | C | PTEN | ENST00000371953 | 0.93725 | 0.97295 | 0.97005 | 0.93276 | 0.79089 | 0.2890256175 | 0.91739 | 0.9374729991 | 0.342027 | 5.407539179 | 0 |
| 10 | 87965390 | A | T | Y | F | PTEN | ENST00000371953 | 0.74005 | 0.88727 | 0.9214 | 0.79117 | 0.75957 | 0.08725408685 | 0.88424 | 0.3216204427 | 0.0220969 | 3.943368662 | 0 |
| 10 | 87965392 | A | G | R | G | PTEN | ENST00000371953 | 0.53934 | 0.86002 | 0.92759 | 0.85814 | 0.79697 | 0.3481295066 | 0.97709 | 0.6652588248 | 0.280189 | 3.826045669 | 0 |
| 10 | 87965393 | G | A | R | K | PTEN | ENST00000371953 | 0.62665 | 0.8416 | 0.96295 | 0.77946 | 0.7255 | 0.2700598046 | 0.86982 | 0.8041946888 | 0.182985 | 4.18677295 | 0 |
| 10 | 87965393 | G | C | R | T | PTEN | ENST00000371953 | 0.65704 | 0.95256 | 0.95135 | 0.8546 | 0.76881 | 0.3146069666 | 0.92955 | 0.8819806576 | 0.30518 | 3.124611 | 0 |
| 10 | 87965393 | G | T | R | I | PTEN | ENST00000371953 | 0.93383 | 0.98291 | 0.94151 | 0.91533 | 0.8517 | 0.3175913425 | 0.9444 | 0.8899227977 | 0.323861 | 4.328589567 | 0 |
| 10 | 87965394 | A | C | R | S | PTEN | ENST00000371953 | 0.4937 | 0.88008 | 0.93264 | 0.81783 | 0.79998 | 0.3432481706 | 0.98002 | 0.6205040812 | 0.282749 | 2.081568262 | 0 |
| 10 | 87965394 | A | T | R | S | PTEN | ENST00000371953 | 0.48825 | 0.88008 | 0.93264 | 0.81783 | 0.79818 | 0.3432481706 | 0.97984 | 0.6205040812 | 0.28274 | 2.081568262 | 0 |
| 10 | 87965395 | T | A | Y | N | PTEN | ENST00000371953 | 0.67231 | 0.98719 | 0.97843 | 0.9266 | 0.8153 | 0.2559863031 | 0.94714 | 0.9397402406 | 0.362466 | 4.195075642 | 0 |
| 10 | 87965395 | T | C | Y | H | PTEN | ENST00000371953 | 0.70639 | 0.9813 | 0.98152 | 0.89112 | 0.77654 | 0.2020234855 | 0.84977 | 0.9075986147 | 0.265481 | 4.831617716 | 0 |
| 10 | 87965395 | T | G | Y | D | PTEN | ENST00000371953 | 0.67915 | 0.93532 | 0.97522 | 0.91575 | 0.80889 | 0.2679139194 | 0.95087 | 0.9454779029 | 0.358773 | 3.161465774 | 0 |
| 10 | 87965396 | A | C | Y | S | PTEN | ENST00000371953 | 0.75376 | 0.97924 | 0.95555 | 0.92703 | 0.80178 | 0.2870945604 | 0.92817 | 0.9493911862 | 0.312404 | 2.852000462 | 0 |
| 10 | 87965396 | A | G | Y | C | PTEN | ENST00000371953 | 0.84499 | 0.9811 | 0.9788 | 0.93116 | 0.80948 | 0.2428004825 | 0.87191 | 0.9236215353 | 0.314794 | 4.647444048 | 0 |
| 10 | 87965396 | A | T | Y | F | PTEN | ENST00000371953 | 0.71714 | 0.95057 | 0.94427 | 0.86769 | 0.76816 | 0.07448625848 | 0.8737 | 0.856071353 | 0.202819 | 4.802700054 | 0 |
| 10 | 87965398 | T | A | S | T | PTEN | ENST00000371953 | 0.63765 | 0.84856 | 0.92647 | 0.889 | 0.74235 | 0.2836503008 | 0.87866 | 0.9353901744 | 0.190935 | 3.858492242 | 0 |
| 10 | 87965398 | T | C | S | P | PTEN | ENST00000371953 | 0.86333 | 0.96506 | 0.93549 | 0.93378 | 0.77011 | 0.3716441401 | 0.8972 | 0.9154129624 | 0.284971 | 5.220612825 | 0 |
| 10 | 87965398 | T | G | S | A | PTEN | ENST00000371953 | 0.66668 | 0.81043 | 0.91948 | 0.85252 | 0.74446 | 0.230977292 | 0.84897 | 0.8887504339 | 0.187745 | 3.734342652 | 0 |
| 10 | 87965399 | C | A | S | Y | PTEN | ENST00000371953 | 0.85308 | 0.96675 | 0.95981 | 0.91206 | 0.79089 | 0.4253222691 | 0.8952 | 0.9342187643 | 0.290426 | 5.242815713 | 0 |
| 10 | 87965399 | C | G | S | C | PTEN | ENST00000371953 | 0.88145 | 0.95345 | 0.93488 | 0.91231 | 0.74728 | 0.4254911157 | 0.86286 | 0.8598821759 | 0.234735 | 5.945720682 | 0 |
| 10 | 87965399 | C | T | S | F | PTEN | ENST00000371953 | 0.89463 | 0.90991 | 0.94911 | 0.90662 | 0.78906 | 0.3635853031 | 0.92716 | 0.9407563806 | 0.31389 | 6.070383427 | 0 |
| 10 | 87965401 | G | A | D | N | PTEN | ENST00000371953 | 0.77219 | 0.89783 | 0.99068 | 0.90231 | 0.73949 | 0.3287304156 | 0.90826 | 0.9372284412 | 0.0646453 | 4.086835575 | 0 |
| 10 | 87965401 | G | C | D | H | PTEN | ENST00000371953 | 0.89656 | 0.97175 | 0.99327 | 0.90991 | 0.8356 | 0.4114058262 | 0.94352 | 0.9513357878 | 0.358765 | 6.093089113 | 0 |
| 10 | 87965401 | G | T | D | Y | PTEN | ENST00000371953 | 0.90218 | 0.97434 | 0.99046 | 0.91262 | 0.87588 | 0.5473457942 | 0.96758 | 0.9822622538 | 0.373637 | 5.275737076 | 0 |
| 10 | 87965402 | A | C | D | A | PTEN | ENST00000371953 | 0.94332 | 0.89697 | 0.97899 | 0.93017 | 0.86951 | 0.4984403753 | 0.90814 | 0.9080817699 | 0.328224 | 4.340188935 | 0 |
| 10 | 87965402 | A | G | D | G | PTEN | ENST00000371953 | 0.94392 | 0.93076 | 0.98136 | 0.92406 | 0.87198 | 0.439813749 | 0.95641 | 0.9583169222 | 0.362308 | 5.358929401 | 0 |
| 10 | 87965404 | A | G | T | A | PTEN | ENST00000371953 | 0.46035 | 0.82109 | 0.87787 | 0.87245 | 0.74797 | 0.2499223245 | 0.91695 | 0.9140710235 | 0.213522 | 4.590510414 | 0 |
| 10 | 87965405 | C | A | T | N | PTEN | ENST00000371953 | 0.80037 | 0.96994 | 0.94227 | 0.90609 | 0.7285 | 0.2469346572 | 0.84849 | 0.9018520117 | 0.173457 | 3.670611637 | 0 |

| | | | | | | | | | | | | | | | | | | |
|---|---|---|---|---|---|---|---|---|---|---|---|---|---|---|---|---|---|---|
| 10 | 87965405 | C | T | T | I | PTEN | ENST00000371953 | 0.69804 | 0.96587 | 0.96496 | 0.90182 | 0.77076 | 0.2637000241 | 0.86776 | 0.9330973625 | 0.283337 | 4.215877286 | 0 |
| 10 | 87965407 | A | G | T | A | PTEN | ENST00000371953 | 0.6475 | 0.9671 | 0.91252 | 0.90301 | 0.76289 | 0.305214584 | 0.90633 | 0.9236215353 | 0.24123 | 4.798497291 | 0 |
| 10 | 87965407 | A | T | T | S | PTEN | ENST00000371953 | 0.57723 | 0.96067 | 0.92454 | 0.89587 | 0.73516 | 0.1375597584 | 0.90265 | 0.8916456103 | 0.18452 | 2.77257586 | 0 |
| 10 | 87965408 | C | A | T | N | PTEN | ENST00000371953 | 0.66747 | 0.97662 | 0.9618 | 0.91018 | 0.73 | 0.2456244122 | 0.85658 | 0.9217982292 | 0.177205 | 3.655617399 | 0 |
| 10 | 87965408 | C | T | T | I | PTEN | ENST00000371953 | 0.8402 | 0.97332 | 0.97812 | 0.92446 | 0.77718 | 0.3893145393 | 0.89843 | 0.96582371 | 0.305304 | 4.274948073 | 0 |
| 10 | 87965410 | G | A | D | N | PTEN | ENST00000371953 | 0.77021 | 0.80295 | 0.89237 | 0.9037 | 0.74867 | 0.3609261439 | 0.90701 | 0.9238358736 | 0.0806969 | 3.872738772 | 0 |
| 10 | 87965410 | G | C | D | H | PTEN | ENST00000371953 | 0.77085 | 0.97411 | 0.91249 | 0.91761 | 0.81006 | 0.440314147 | 0.91758 | 0.9481703043 | 0.338406 | 4.840895471 | 0 |
| 10 | 87965410 | G | T | D | Y | PTEN | ENST00000371953 | 0.90196 | 0.97869 | 0.8969 | 0.91749 | 0.86047 | 0.5768662375 | 0.96589 | 0.9577050805 | 0.374527 | 8.191674222 | 0 |
| 10 | 87965411 | A | C | D | A | PTEN | ENST00000371953 | 0.87051 | 0.9725 | 0.83859 | 0.93479 | 0.82892 | 0.5332469127 | 0.9166 | 0.9601668119 | 0.339424 | 5.127216735 | 0 |
| 10 | 87965411 | A | T | D | V | PTEN | ENST00000371953 | 0.86467 | 0.98432 | 0.84703 | 0.95447 | 0.88684 | 0.5388437641 | 0.96694 | 0.9789830446 | 0.372591 | 6.8221266 | 0 |
| 10 | 87965412 | C | A | D | E | PTEN | ENST00000371953 | 0.57645 | 0.92397 | 0.87144 | 0.9019 | 0.74306 | 0.1983644666 | 0.88022 | 0.9075523615 | 0.19448 | 2.920412995 | 0 |
| 10 | 87965413 | T | A | S | T | PTEN | ENST00000371953 | 0.60551 | 0.96171 | 0.89115 | 0.90749 | 0.72016 | 0.2864172734 | 0.87676 | 0.8663796782 | 0.156904 | 3.440655163 | 0 |
| 10 | 87965413 | T | C | S | P | PTEN | ENST00000371953 | 0.86439 | 0.98196 | 0.9028 | 0.93599 | 0.74164 | 0.3593740244 | 0.89668 | 0.9270880818 | 0.236499 | 3.638316804 | 0 |
| 10 | 87965414 | C | G | S | C | PTEN | ENST00000371953 | 0.88566 | 0.96682 | 0.89637 | 0.93955 | 0.76421 | 0.4306167498 | 0.85783 | 0.8897024989 | 0.20054 | 4.543172221 | 0 |
| 10 | 87965414 | C | T | S | F | PTEN | ENST00000371953 | 0.89845 | 0.97448 | 0.91957 | 0.94264 | 0.79212 | 0.4116142922 | 0.88024 | 0.9636505842 | 0.195283 | 4.607154384 | 0 |
| 10 | 87965416 | G | A | D | N | PTEN | ENST00000371953 | 0.72404 | 0.90935 | 0.90694 | 0.90596 | 0.72322 | 0.2920136015 | 0.89819 | 0.899915576 | 0.0843504 | 3.970290088 | 0 |
| 10 | 87965416 | G | T | D | Y | PTEN | ENST00000371953 | 0.76871 | 0.97569 | 0.90304 | 0.91813 | 0.8278 | 0.5106633389 | 0.94859 | 0.9863022566 | 0.354171 | 6.16799366 | 0 |
| 10 | 87965417 | A | C | D | A | PTEN | ENST00000371953 | 0.73934 | 0.91917 | 0.85786 | 0.88802 | 0.77141 | 0.4638523979 | 0.93383 | 0.939694941 | 0.31854 | 5.314940143 | 1 |
| 10 | 87965417 | A | T | D | V | PTEN | ENST00000371953 | 0.88598 | 0.97228 | 0.85535 | 0.93399 | 0.85638 | 0.472895368 | 0.97845 | 0.9638980627 | 0.365941 | 8.105786799 | 0 |
| 10 | 87965418 | T | A | D | E | PTEN | ENST00000371953 | 0.34261 | 0.77545 | 0.88721 | 0.76802 | 0.71153 | 0.1439249508 | 0.91692 | 0.159297213 | 0.118436 | 3.241234051 | 0 |
| 10 | 87965419 | C | T | P | S | PTEN | ENST00000371953 | 0.4074 | 0.80235 | 0.8315 | 0.84202 | 0.72775 | 0.2845702747 | 0.85079 | 0.7685803771 | 0.191783 | 2.685971704 | 0 |
| 10 | 87965420 | C | G | P | R | PTEN | ENST00000371953 | 0.82069 | 0.94054 | 0.88382 | 0.91046 | 0.78597 | 0.3546872181 | 0.90714 | 0.8338621855 | 0.298049 | 4.174962624 | 0 |
| 10 | 87965420 | C | T | P | L | PTEN | ENST00000371953 | 0.69115 | 0.87327 | 0.89014 | 0.89033 | 0.77205 | 0.3307245853 | 0.93352 | 0.9191515446 | 0.278287 | 4.679317428 | 0 |
| 10 | 87965422 | G | C | E | Q | PTEN | ENST00000371953 | 0.87072 | 0.96791 | 0.87763 | 0.91651 | 0.7337 | 0.3647591461 | 0.88751 | 0.8979304433 | 0.228151 | 4.146157114 | 0 |
| 10 | 87965423 | A | C | E | A | PTEN | ENST00000371953 | 0.85249 | 0.97243 | 0.8905 | 0.94202 | 0.76356 | 0.4312532057 | 0.89217 | 0.9436371922 | 0.283093 | 3.816489683 | 0 |
| 10 | 87965423 | A | T | E | V | PTEN | ENST00000371953 | 0.87951 | 0.97803 | 0.86979 | 0.94717 | 0.81876 | 0.5354122564 | 0.89871 | 0.9680508375 | 0.301311 | 4.293654745 | 0 |
| 10 | 87965424 | G | T | E | D | PTEN | ENST00000371953 | 0.69788 | 0.96121 | 0.88792 | 0.88934 | 0.71312 | 0.1788375528 | 0.8402 | 0.8755295277 | 0.171912 | 2.917010293 | 0 |
| 10 | 87965425 | A | C | N | H | PTEN | ENST00000371953 | 0.79682 | 0.97054 | 0.82392 | 0.90192 | 0.76356 | 0.2634041397 | 0.93297 | 0.9336876869 | 0.194969 | 7.724255323 | 0 |
| 10 | 87965425 | A | G | N | D | PTEN | ENST00000371953 | 0.6712 | 0.97582 | 0.8553 | 0.84334 | 0.74164 | 0.1766632993 | 0.88089 | 0.9084843397 | 0.117785 | 2.481487295 | 0 |
| 10 | 87965425 | A | T | N | Y | PTEN | ENST00000371953 | 0.84552 | 0.98028 | 0.84853 | 0.93381 | 0.82216 | 0.387841135 | 0.92745 | 0.9625746608 | 0.297843 | 5.62363087 | 0 |
| 10 | 87965426 | A | C | N | T | PTEN | ENST00000371953 | 0.8944 | 0.96909 | 0.78888 | 0.91426 | 0.76223 | 0.2371466191 | 0.89537 | 0.9387457967 | 0.189646 | 4.744923951 | 0 |
| 10 | 87965426 | A | G | N | S | PTEN | ENST00000371953 | 0.7169 | 0.94989 | 0.74759 | 0.87836 | 0.73074 | 0.1657781917 | 0.8786 | 0.9073016644 | 0.0950143 | 2.56220783 | 0 |
| 10 | 87965426 | A | T | N | I | PTEN | ENST00000371953 | 0.92241 | 0.9811 | 0.8165 | 0.94089 | 0.8124 | 0.4466967323 | 0.91938 | 0.9822385311 | 0.307579 | 7.724255323 | 0 |
| 10 | 87965428 | G | A | E | K | PTEN | ENST00000371953 | 0.80116 | 0.82622 | 0.9285 | 0.90853 | 0.79878 | 0.4152356336 | 0.90475 | 0.9148424864 | 0.291616 | 4.674267127 | 0 |
| 10 | 87965428 | G | C | E | Q | PTEN | ENST00000371953 | 0.8681 | 0.85192 | 0.84484 | 0.86358 | 0.7147 | 0.3050688176 | 0.88838 | 0.9061008096 | 0.205136 | 4.459587058 | 0 |
| 10 | 87965429 | A | C | E | A | PTEN | ENST00000371953 | 0.93204 | 0.85505 | 0.85878 | 0.89422 | 0.77399 | 0.3516364795 | 0.88624 | 0.9250364304 | 0.272073 | 4.335477169 | 0 |
| 10 | 87965429 | A | G | E | G | PTEN | ENST00000371953 | 0.94653 | 0.87601 | 0.81082 | 0.92411 | 0.79151 | 0.3492504274 | 0.87225 | 0.9376121759 | 0.296778 | 4.896974692 | 0 |
| 10 | 87965429 | A | T | E | V | PTEN | ENST00000371953 | 0.94276 | 0.92882 | 0.83393 | 0.93601 | 0.78286 | 0.4773925747 | 0.92761 | 0.928560257 | 0.314907 | 7.89143889 | 0 |
| 10 | 87965430 | A | T | E | D | PTEN | ENST00000371953 | 0.61574 | 0.76267 | 0.85293 | 0.70738 | 0.73148 | 0.1420324527 | 0.91865 | 0.6187386513 | 0.145889 | 2.764394076 | 0 |
| 10 | 87965431 | C | A | P | T | PTEN | ENST00000371953 | 0.3657 | 0.87188 | 0.85841 | 0.75234 | 0.72474 | 0.3009332658 | 0.86648 | 0.7624218464 | 0.177011 | 3.235712256 | 0 |
| 10 | 87965431 | C | T | P | S | PTEN | ENST00000371953 | 0.37154 | 0.82714 | 0.83148 | 0.73862 | 0.69594 | 0.3183981874 | 0.81387 | 0.2325444669 | 0.148139 | 3.153974005 | 0 |
| 10 | 87965432 | C | A | P | H | PTEN | ENST00000371953 | 0.52612 | 0.93443 | 0.87126 | 0.77453 | 0.73223 | 0.2624645558 | 0.77409 | 0.8163957 | 0.180119 | 3.870038985 | 0 |
| 10 | 87965432 | C | G | P | R | PTEN | ENST00000371953 | 0.51008 | 0.86627 | 0.88431 | 0.75727 | 0.76024 | 0.34929466 | 0.91765 | 0.7174755335 | 0.265013 | 3.807815071 | 0 |
| 10 | 87965432 | C | T | P | L | PTEN | ENST00000371953 | 0.51738 | 0.8586 | 0.8878 | 0.79655 | 0.71391 | 0.2620920217 | 0.85252 | 0.8217976689 | 0.172158 | 3.333835061 | 0 |
| 10 | 87965435 | T | G | F | C | PTEN | ENST00000371953 | 0.68475 | 0.96487 | 0.9141 | 0.85259 | 0.80358 | 0.2258433953 | 0.93143 | 0.8910109401 | 0.276094 | 4.569838054 | 0 |
| 10 | 87965437 | G | A | D | N | PTEN | ENST00000371953 | 0.57928 | 0.89418 | 0.8929 | 0.83252 | 0.72322 | 0.1965894766 | 0.85668 | 0.8741156459 | 0.0509822 | 7.688335073 | 0 |
| 10 | 87965437 | G | C | D | H | PTEN | ENST00000371953 | 0.8765 | 0.96723 | 0.91139 | 0.85915 | 0.81933 | 0.2839320011 | 0.95352 | 0.9272360206 | 0.318245 | 4.884582228 | 0 |
| 10 | 87965438 | A | C | D | A | PTEN | ENST00000371953 | 0.78638 | 0.9073 | 0.83777 | 0.85199 | 0.81006 | 0.3401704683 | 0.94211 | 0.9100535512 | 0.296267 | 3.944833552 | 0 |
| 10 | 87965438 | A | T | D | V | PTEN | ENST00000371953 | 0.80898 | 0.96677 | 0.84412 | 0.88059 | 0.87636 | 0.3798133793 | 0.9763 | 0.9660507441 | 0.362926 | 7.688335073 | 0 |
| 10 | 87965440 | G | A | E | K | PTEN | ENST00000371953 | 0.72051 | 0.88967 | 0.95593 | 0.8416 | 0.7528 | 0.2641926416 | 0.97799 | 0.7906720042 | 0.308932 | 7.10025346 | 0 |
| 10 | 87965441 | A | C | E | A | PTEN | ENST00000371953 | 0.76745 | 0.89398 | 0.89905 | 0.80184 | 0.7727 | 0.2748383811 | 0.9459 | 0.8918602467 | 0.263735 | 7.10025346 | 0 |
| 10 | 87965441 | A | T | E | V | PTEN | ENST00000371953 | 0.82685 | 0.95541 | 0.88171 | 0.87072 | 0.81761 | 0.3652717178 | 0.95629 | 0.9195489287 | 0.307245 | 7.10025346 | 0 |
| 10 | 87965442 | A | T | E | D | PTEN | ENST00000371953 | 0.45107 | 0.76231 | 0.89554 | 0.73389 | 0.63525 | 0.0997342934 | 0.884 | 0.712569356 | 0.123149 | 1.228542424 | 0 |
| 10 | 87965443 | G | A | D | N | PTEN | ENST00000371953 | 0.69836 | 0.87051 | 0.79071 | 0.80746 | 0.70994 | 0.2486282879 | 0.83298 | 0.8221899867 | 0.0444668 | 3.522483829 | 0 |
| 10 | 87965443 | G | C | D | H | PTEN | ENST00000371953 | 0.72965 | 0.96033 | 0.81242 | 0.83537 | 0.75074 | 0.3313846172 | 0.85108 | 0.8956211209 | 0.188282 | 4.512519889 | 0 |
| 10 | 87965443 | G | T | D | Y | PTEN | ENST00000371953 | 0.74037 | 0.96895 | 0.79527 | 0.85498 | 0.75755 | 0.4623260164 | 0.87805 | 0.9643880725 | 0.213763 | 7.41206979 | 0 |
| 10 | 87965444 | A | C | D | A | PTEN | ENST00000371953 | 0.77227 | 0.89143 | 0.7256 | 0.80336 | 0.68995 | 0.3734457081 | 0.80709 | 0.9247144461 | 0.179151 | 4.750959033 | 0 |
| 10 | 87965446 | C | G | Q | E | PTEN | ENST00000371953 | 0.44503 | 0.83034 | 0.75185 | 0.7286 | 0.70914 | 0.1198286426 | 0.91336 | 0.6364372969 | 0.177998 | 2.304300534 | 0 |
| 10 | 87965447 | A | G | Q | R | PTEN | ENST00000371953 | 0.52087 | 0.85429 | 0.79786 | 0.77854 | 0.7217 | 0.147001276 | 0.97156 | 0.3363396525 | 0.145304 | 4.284799269 | 0 |
| 10 | 87965447 | A | T | Q | L | PTEN | ENST00000371953 | 0.68179 | 0.78539 | 0.73953 | 0.80267 | 0.74164 | 0.2060995465 | 0.91869 | 0.8424136043 | 0.198848 | 3.96192837 | 0 |
| 10 | 87965449 | C | A | H | N | PTEN | ENST00000371953 | 0.58981 | 0.88205 | 0.94486 | 0.70256 | 0.71073 | 0.1849711716 | 0.89225 | 0.6197591424 | 0.150923 | 7.412709771 | 0 |
| 10 | 87965449 | C | G | H | D | PTEN | ENST00000371953 | 0.62105 | 0.91961 | 0.94143 | 0.6891 | 0.7727 | 0.3416482105 | 0.98863 | 0.661231339 | 0.370734 | 2.531104352 | 0 |
| 10 | 87965450 | A | C | H | P | PTEN | ENST00000371953 | 0.66216 | 0.95933 | 0.90563 | 0.7044 | 0.825 | 0.3542018442 | 0.98466 | 0.8556643128 | 0.345251 | 7.412709771 | 0 |
| 10 | 87965452 | A | C | T | P | PTEN | ENST00000371953 | 0.50062 | 0.86134 | 0.56521 | 0.76409 | 0.75143 | 0.1875928736 | 0.95823 | 0.5393137336 | 0.215905 | 5.24471461 | 0 |
| 10 | 87965452 | A | T | T | S | PTEN | ENST00000371953 | 0.40144 | 0.75753 | 0.58623 | 0.58072 | 0.64775 | 0.1140788364 | 0.7246 | 0.343552053 | 0.0377627 | 1.091870402 | 0 |
| 10 | 87965453 | C | A | T | K | PTEN | ENST00000371953 | 0.47703 | 0.8469 | 0.70649 | 0.77315 | 0.70262 | 0.3265947622 | 0.94173 | 0.7710331678 | 0.249398 | 6.272799701 | 0 |
| 10 | 87965453 | C | G | T | R | PTEN | ENST00000371953 | 0.48126 | 0.83469 | 0.6866 | 0.78353 | 0.72775 | 0.3228566011 | 0.93928 | 0.770516336 | 0.243691 | 6.272799701 | 0 |
| 10 | 87965453 | C | T | T | I | PTEN | ENST00000371953 | 0.59103 | 0.81929 | 0.74437 | 0.76809 | 0.69168 | 0.2507880681 | 0.864 | 0.8867489696 | 0.178143 | 6.272799701 | 0 |
| 10 | 87965455 | C | A | Q | K | PTEN | ENST00000371953 | 0.40981 | 0.88228 | 0.7429 | 0.90925 | 0.70589 | 0.2988362539 | 0.98409 | 0.7851752043 | 0.258646 | 4.288161621 | 0 |
| 10 | 87965455 | C | G | Q | E | PTEN | ENST00000371953 | 0.37543 | 0.8381 | 0.68419 | 0.81968 | 0.71153 | 0.2286690188 | 0.9231 | 0.6364372969 | 0.18377 | 7.943386052 | 0 |
| 10 | 87965456 | A | C | Q | P | PTEN | ENST00000371953 | 0.71891 | 0.94597 | 0.69666 | 0.89873 | 0.75957 | 0.3317877642 | 0.98552 | 0.8752916455 | 0.348201 | 7.943386052 | 0 |
| 10 | 87965459 | T | A | I | N | PTEN | ENST00000371953 | 0.89543 | 0.99392 | 0.97583 | 0.94308 | 0.84641 | 0.5482030824 | 0.98146 | 0.9082522988 | 0.431409 | 7.964654175 | 0 |
| 10 | 87965460 | T | G | I | M | PTEN | ENST00000371953 | 0.55236 | 0.9695 | 0.92076 | 0.86891 | 0.81646 | 0.2401386104 | 0.96979 | 0.7384355664 | 0.276303 | 7.964654175 | 0 |
| 10 | 87965461 | A | C | T | P | PTEN | ENST00000371953 | 0.65956 | 0.86485 | 0.80721 | 0.9171 | 0.77654 | 0.2884916594 | 0.9301 | 0.9363570213 | 0.358018 | 7.831733475 | 0 |
| 10 | 87965462 | C | G | T | R | PTEN | ENST00000371953 | 0.78368 | 0.96564 | 0.8848 | 0.92195 | 0.78035 | 0.4443480273 | 0.95275 | 0.9219950438 | 0.392857 | 7.831733475 | 0 |
| 10 | 87965464 | A | C | K | Q | PTEN | ENST00000371953 | 0.55168 | 0.93824 | 0.73369 | 0.83342 | 0.75822 | 0.6569609701 | 0.89356 | 0.8710054159 | 0.184515 | 7.857675077 | 0 |
| 10 | 87965464 | A | G | K | E | PTEN | ENST00000371953 | 0.54426 | 0.91433 | 0.78118 | 0.81037 | 0.79334 | 0.7088709947 | 0.97007 | 0.8600805402 | 0.302062 | 7.857675077 | 0 |
| 10 | 87965465 | A | G | K | R | PTEN | ENST00000371953 | 0.59559 | 0.84328 | 0.69136 | 0.76891 | 0.74446 | 0.5163948021 | 0.88966 | 0.9369748235 | 0.169632 | 3.298813488 | 0 |
